# Post Induction Hypotension prediction during general anesthesia using Machine Learning Techniques

**DOI:** 10.1101/2025.04.14.25322061

**Authors:** Harikesh Subramanian, Shyam Visweswaran, Senthilkumar Sadhasivam, Aman Mahajan

## Abstract

**Background:** Intraoperative hypotension burden not equally distributed during various periods of a general anesthetic. Post-induction hypotension usually has an iatrogenic cause and is related to the combination of drug doses used for induction of anesthesia. Predicting post-induction blood pressures, prior to induction, may inform anesthesiologists on the most judicious selection of anesthetic drugs and dosages. Our objective was to explore patterns of post-induction hypotension, apply machine learning (ML) to predict the mean blood pressure (MBP) immediately after induction, and develop a prototype of a clinical tool that can implement the ML model.

**Materials and Methods:** We extracted data from preoperative and intraoperative anesthetic episodes over an 8-year period at the University of Pittsburgh Medical Center. The dataset consisted of 93,037 anesthetic episodes, which was partitioned into training, validation, and test sets. Preoperative and pre-induction predictor variables included demographics, comorbidities, laboratory test values, pre-induction MBP, and administrations of induction drugs. The target consisted of 15 minutes of MBP values immediately after induction.

**Results:** The best-performing model was extreme-gradient boosted trees (XGB), with an R-squared value of 0.31 and a mean absolute error of 11.96, which is a moderately good performance. The performance of the model decreased over each minute post-induction.

**Conclusion:** ML modeling to predict post-induction MBP is feasible, and a clinical tool that incorporates ML can potentially aid in preventing hypotension in the operating room.

## BACKGROUND

The general anesthetic episode consists of three key chronological phases: induction, maintenance, and emergence. Induction occurs at the start of the anesthetic episode when a rapid and smooth loss of consciousness, analgesia, and amnesia is achieved. Maintenance of anesthesia follows induction and is the period during which the surgical procedure is performed. Emergence occurs at the end of the anesthetic episode when the effects of anesthetic medications are reversed, and the patient regains consciousness.

### Post-induction hypotension

In the initial few minutes of induction, when anesthetic medications are administered, multiple physiological processes occur in the patient. The induction medications cause amnesia, analgesia, apnea, and a blunting of the sympathetic response and concurrently, there is activation of a stress response from the placement of the endotracheal tube^1,2^. These opposing effects can lead to a period of labile vital signs leading to post-induction hypotension.

Post-induction hypotension has a high prevalence and can occur in up to 18% of all anesthetic episodes, necessitating some form of intervention^3^. Hypotension can occur in as many as 32-51% of patients who have cardiovascular dysfunction documented by an echocardiogram^4,5^. Other risk factors include advanced age, preoperative low blood pressure, concurrent illnesses, and certain preoperative medications^6^. Early identification of post-induction hypotension is important since any hypotension in the intraoperative period has been linked to major postoperative adverse outcomes such as acute kidney injury, myocardial injury, and neurocognitive decline^7^.

An appropriate combination and dosage of medications, as well as pre-treatment with certain vasopressors, can minimize post-induction hypotension, but identifying patients who are at risk of hypotension is not simple. Induction is a complex, high-stakes procedure that requires considering the patient’s entire medical history, current illness, previous medication administration, and a plethora of other factors such as the ejection fraction, comorbidities, and weight. A clinical tool that can accurately predict hypotension after induction based on the patient’s preoperative clinical information and pre-induction vital signs would be invaluable for anesthesiologists as well as trainees to optimize induction.

### Value of intraoperative record data

In the previous decade, preoperative data has been widely utilized to predict outcomes in the field of perioperative research^8,9^. The preoperative electronic health record (EHR) is an excellent source for demographic information, comorbidities, and laboratory test values and provides a comprehensive profile of the patient before surgery. However, the EHR is frequently incomplete and sometimes inaccurate, and it is challenging to translate it into a structured format for analyses. In contrast, many Anesthesia Information Management Systems (AIMS) output intraoperative data in a structured format for the purposes of documentation, retrieval, and medico-legal needs^10^. Intraoperative vital sign data in AIMS are captured automatically and can be output for analyses. Medication, fluid, and blood administration data are manually entered into AIMS systems on the same intraoperative timeline. Thus, the intraoperative anesthesia record is an invaluable source for analyses, and the potential of this high-quality granular data is increasingly being realized ^8,11–13^.

### Modeling induction

Analyzing and modeling induction is challenging since it does not occur instantaneously but occurs gradually over a period of time. Most AIMS systems allow the anesthesiologist to annotate the intraoperative record during induction, and it is common practice in our institution to do the induction annotations at the beginning of this period.

Little prior research has been done in predictive modeling of post-induction hypotension. Two articles by Kang et. al. and Hu et. al. describe training a classification model on a small number of patients to predict post-induction hypotension as a binary target (stable vs. unstable induction)^14,15^. This approach has two main drawbacks. First, it requires the threshold of a stable/safe blood pressure post-induction to be set, which has been a hotly debated topic^16^.

Second, predicting a binary target is not as clinically useful as predicting the actual blood pressure for anesthesiologists in their daily practice.

In this project, we build on this work and develop a regression model on a large dataset targeting post-induction blood pressure, which can eventually be used as a clinical tool.

## GOALS AND OBJECTIVES

The overall goal of this project was to use machine learning (ML) to predict mean blood pressure (MBP) during the post-induction period and eventually develop a clinical tool based on ML that will be useful in predicting post-induction hypotension.

The specific objectives of the project included exploratory analysis, developing and evaluating ML models, and prototyping a user interface of a clinical tool. The first objective was to assess the degree and patterns of post-induction hypotension. The second objective was to build and evaluate ML models that accurately predict MBP in the 15-minute period after induction. The third objective was to build and perform a preliminary usability evaluation of a clinical tool prototype that will deploy an ML model.

## MATERIALS AND METHODS

In this section, we describe details of the dataset, the preprocessing steps, the experimental methods, including the methods used for deriving ML models, the performance measures used in the evaluation of the ML models, and finally, the usability studies performed on the prototype clinical tool.

### Dataset and variables

### Data source

The dataset was created by combining preoperative and intraoperative data from anesthetic episodes at the University of Pittsburgh Medical Center (UPMC) over an 8-year period.

Preoperative data were extracted from the *Cerner^TM^ R3* system and included comorbidities (ICD codes) documented in the EHR in the year preceding the anesthetic episode. Intraoperative anesthetic data were extracted from the *Cerner^TM^ SA-Anesthesia module*, which included timestamped medications, fluid administrations, blood pressure measurements, and annotations made by the anesthesiologist.

The dataset was de-identified by removing patient identifiers, and a unique case number was assigned to each anesthetic episode. Repeat anesthetic episodes for the same patient were assigned unique case numbers.

### Inclusion and exclusion criteria

All anesthetic episodes for patients over the age of 18 years whose procedures occurred between January 1, 2013, and December 31, 2020, and who had the requisite preoperative echocardiogram within one year prior to the anesthetic episode at any of UPMC’s hospitals were included. Episodes that lacked preoperative data or had incomplete intraoperative anesthetic records were excluded. Figure 1A details the selection of anesthetic episodes and gives the total number of anesthetic episodes ultimately used in the study.

**Figure 1:**
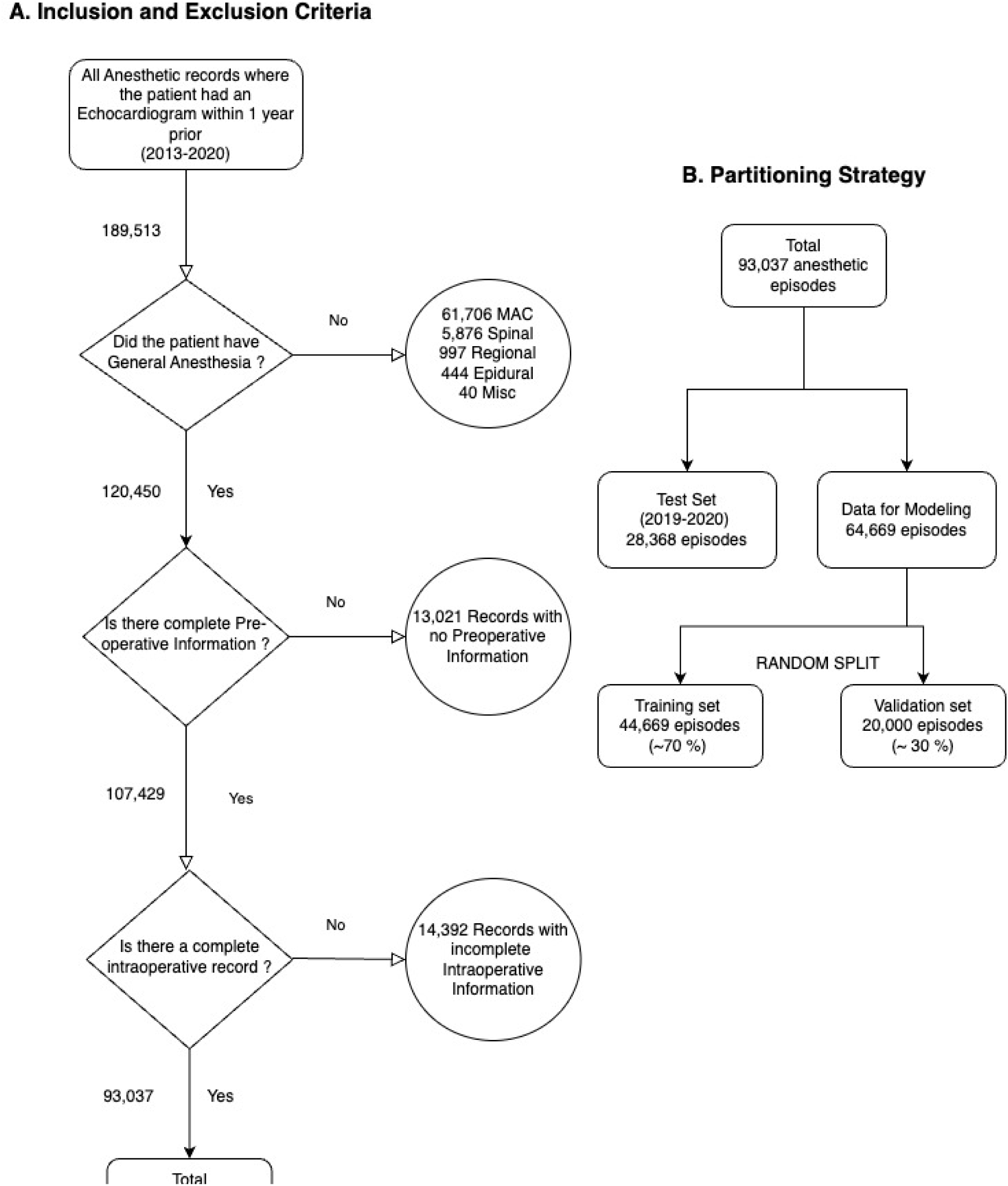
Creation of dataset. A) Flow chart showing the application of inclusion and exclusion criteria for selection of anesthetic episodes. B) Flow chart showing the partitioning of the dataset into training, validation, and test datasets.

**Figure 2.**
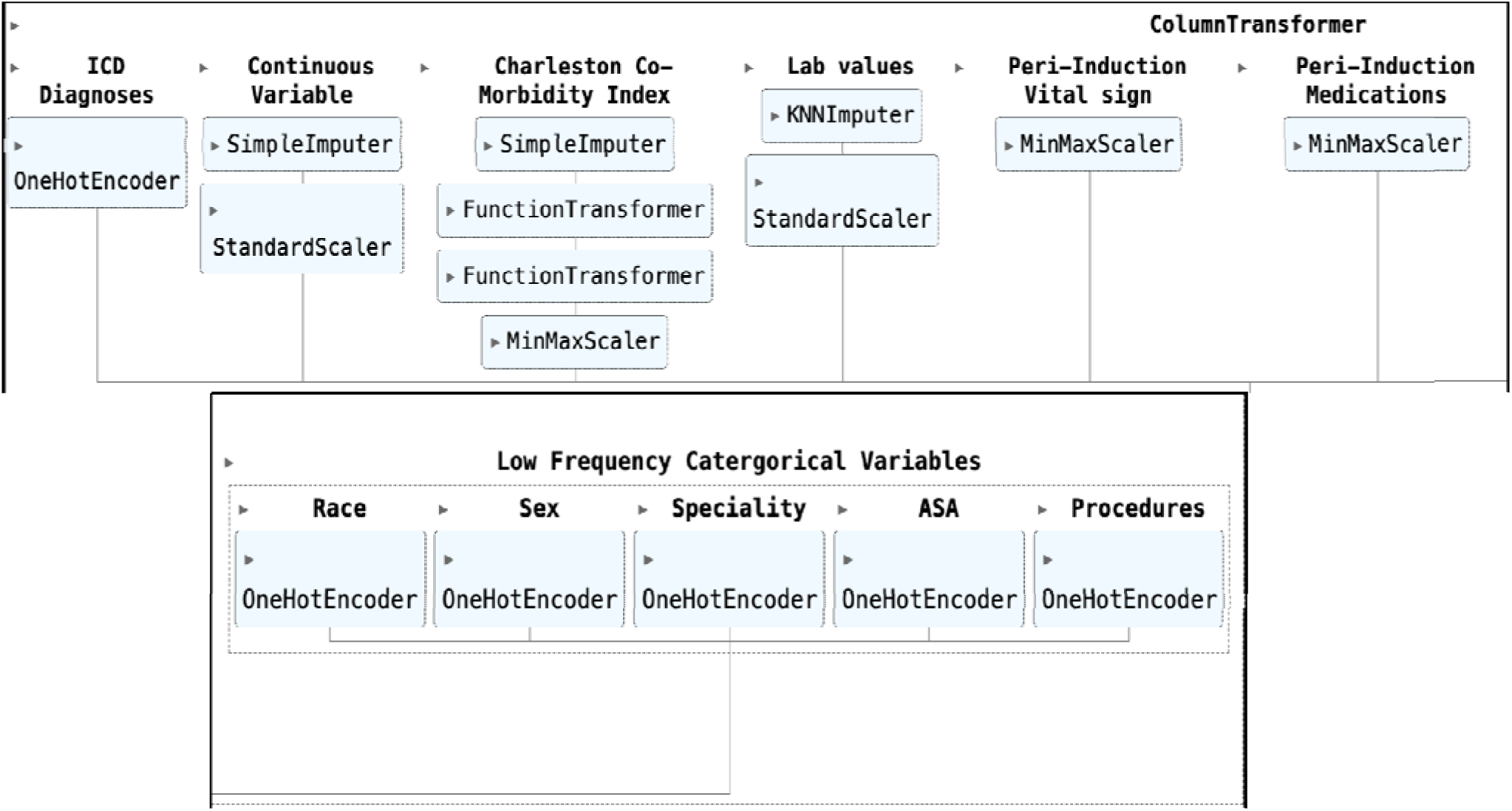
The preprocessing pipeline.

### Predictor and target variables

We chose to limit the preoperative input variables to information that would be available to the anesthesiologist at the time of induction. Induction in the intraoperative record timeline was labeled as t_0,_ and the pre-induction and post-induction periods were constrained to be 10 minutes prior to t_0_ and 15 minutes after t_0,_ respectively. The peri-induction period is defined as the concatenation of pre-induction, induction, and post-induction periods. The predictor and target variables are described below.

- Demographic variables included age (continuous, in years), sex (categorical), race (categorical), and weight (continuous, in kgs) at the start of the anesthetic episode (Table 1).
- Preoperative comorbidity variables included a list of binary-encoded ICD-10 diagnoses (39 diagnoses; present/absent values), the left ventricular ejection fraction (LVEF) (continuous, %), and the severity of illness as calculated by the Charleston Comorbidity Index (CCI) (continuous, no units) (Table 1).
- Preoperative laboratory test result variables included the values of 12 laboratory test results commonly reported which were serum concentrations of sodium, creatinine, blood urea nitrogen (BUN), hemoglobin, white blood cell (WBC) counts, albumin, bilirubin, aspartate transferase (AST), estimated glomerular filtration rate (EGFR), platelet concentration, international normalized ratio (INR), and partial thromboplastin time (PTT). Only the most recent values reported within a year prior to the anesthetic episode were included (Table 1).
- Variables related to the procedure included the American Society of Anesthesiologists (ASA) physical status (categorical, ASA 1-5 with or without E status), the surgical procedure (categorical, 1,855 unique procedures), and the surgeon’s primary specialty (categorical, 13 unique specialties) (Table 1).
- Peri-induction medication variables included 30 anesthetic, ionotropic, and vasopressor drugs that are commonly administered in the peri-induction period. For each drug, all administrations were totaled after conversion to the same dose unit to produce a single value (Table 1).The drug doses in the entire peri-induction period (including the target period) were used as inputs. The reason being that manual documentation of drug administration can be often recorded in the post-induction period, due to competing clinical tasks being performed by the clinicians in the room.
- Variables related to the pre-induction physiological measurements included two readings of MBP, one at t_-1_ and another at t_0_ (see Figure 3).
- The target variable was post-induction MBP at each minute from t_1_ (1 minute post-induction) through t_15_ (15 minutes post-induction) (see Figure 3).

**Figure 3.**
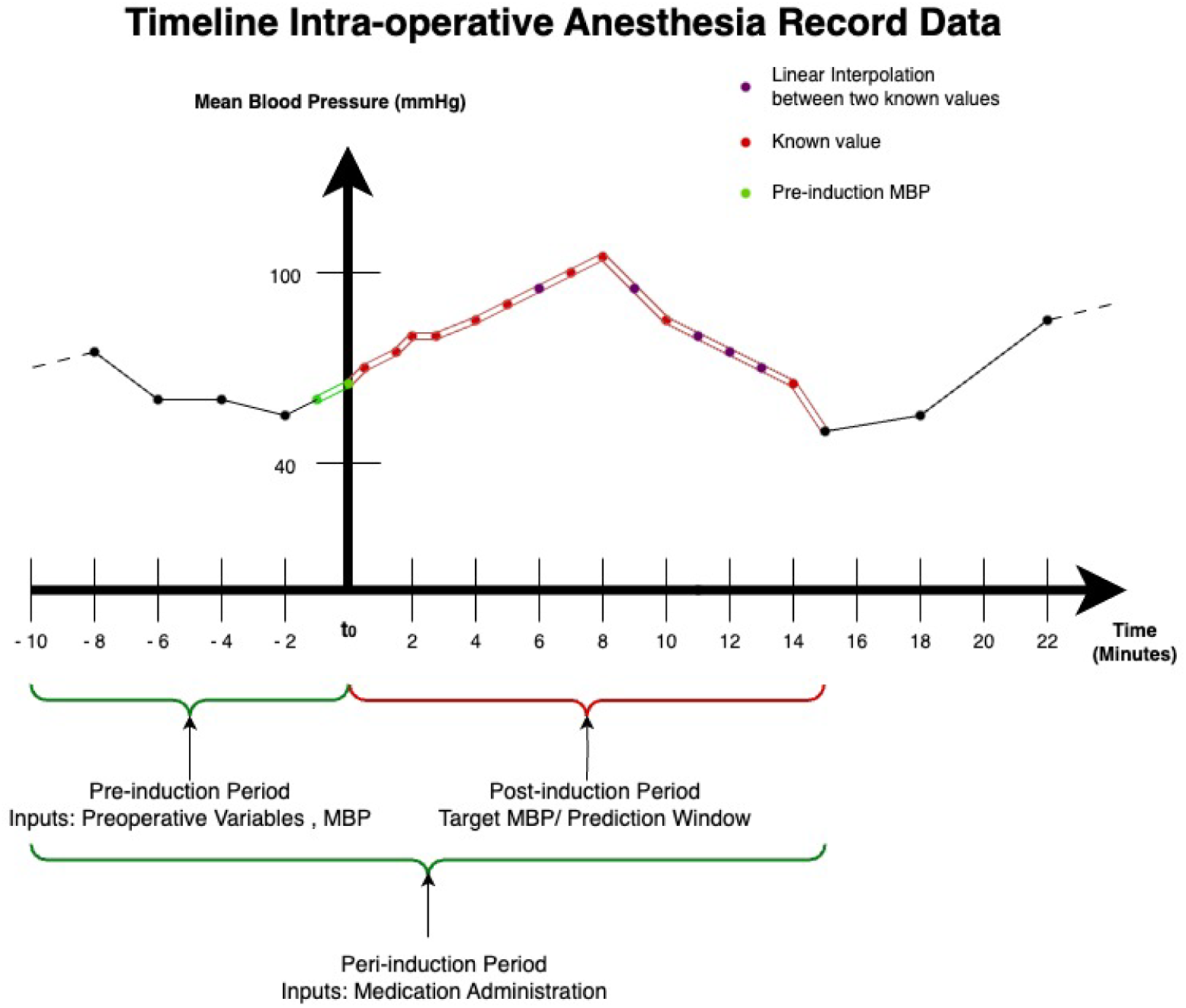
Timeline of induction and the post-induction prediction period.

**Table 1.**
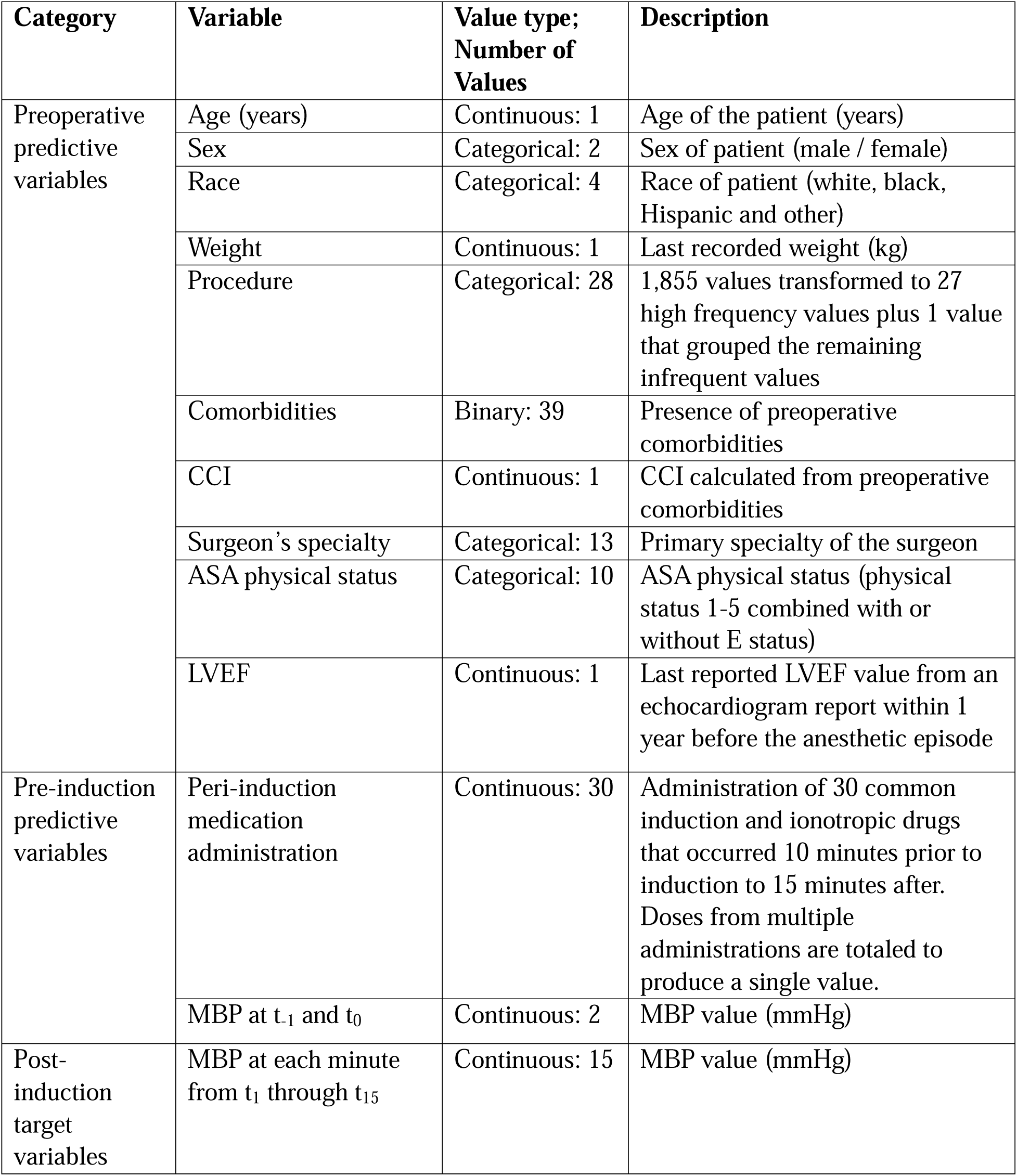
Description of predictor and target variables.

### Preprocessing

Here, we describe the handling of missing data and encoding of variables. Missing categorical values were grouped to the “infrequent/unknown” value within each variable. Missing values for age and weight, CCI were imputed using the median value. Missing laboratory test values were imputed using *k*-nearest neighbor imputation. Imputation was performed within the preprocessing pipeline prior to fitting on the training set alone. When cross-validation was used, imputation was repeated within each fold. The imputation pipelines trained on the training data were used to impute what was missing in the test set.

Categorical variables such as sex, race, ASA physical status, procedure, and surgeon’s primary specialty were one-hot encoded. To limit the number of values generated by the one-hot encoding of variables (e.g., the procedure variable contained 1,855 ICD codes), those values that occurred with a low frequency (less than 0.5% in the data) were mapped to a single “infrequent/unknown” value.

Different groups of continuous variables were preprocessed differentially. Continuous variables such as age, weight, and laboratory test values underwent normal standardization and scaling. Variables such as the CCI, which did not follow a normal distribution, were log-transformed prior to min-max scaling. A summary of preprocessing is shown in Figure 2.

Peri-induction drug administration doses were converted to the same units. Outlier values were removed. Multiple administrations of the same drug were totaled to produce a single value.

Missing or absent administrations were coded as zero values. The totaled drug administrations were subsequently min-max scaled.

The MBP measurements reported from the monitors were primarily from the non-invasive blood pressure (NIBP) cuff. A more accurate measure of MBP via an invasive arterial line (A-line) monitor was also available in some anesthetic episodes. If the A-line pressures were within physiological values (> 25 mmHg and < 285 mmHg), this value was used as the preferred value of MBP.

Since MBP measurements are performed and documented at a frequency ranging between 1-5 minutes, linear interpolation was used to impute the MBP values at 1-minute time points between two measurements. Subsequently, anesthetic episodes without at least one MBP measurement prior to induction and one after induction were dropped. In a small minority of anesthetic episodes, when the last measured MBP was less than 15 minutes after induction, missing MBP values were filled forward until the 15th minute.

After preprocessing, each anesthetic episode consisted of 118 predictor variables, and the target consisted of 15 MBP values.

### Experimental methods

#### Training, validation, and test datasets

The dataset was partitioned, as shown in Figure 1B. A test dataset of anesthetic episodes in 2019 and 2020 (28,368 anesthetic episodes) was used to report the performance of models. The remaining episodes from 2013 to 2018 (64,669 anesthetic episodes) were randomly split 70% / 30% into training (44,669 anesthetic episodes) and validation datasets (20,000 anesthetic episodes). The training dataset was used to train the models, and the validation dataset was used to tune the models.

#### Exploratory analyses

We explored the predictor and target variables with summary statistics as well as graphical tools on the full datasets as well as the training, validation, and test datasets. Conditional exploration of the input space based on the average MBP of the first 15 minutes after induction was performed.

#### Modeling

We used supervised learning methods to predict MBP at each minute in the 15-minute post-induction period. The inputs included preoperative variables and induction variables. The target variable was MBP in the first 15 minutes following induction, i.e., at each minute from t_1_ through t_15_ (see Figure 3). Parameter tuning for select models was performed by grid search and cross-validation using the validation set. The performance of the models is reported on the test dataset.

#### ML algorithms and environment

A brief description of the different models follows. The linear regression model was fit with an objective function of mean squared error loss. A ridge regression model (which uses the L2 regularization penalty) and a bayesian ridge regression model (which adapts the priors for the coefficients to the data at hand) was also trained. Decision tree models, which attempt to divide the input space into regions along each dimension to optimize, at each split, the maximum improvement in purity, were trained. Ensembles of decision trees, which reduce overfitting and are some of the best-performing ML models in healthcare, were also utilized. Hist-gradient boosted trees which is a faster implementation of the X-Gradient boosted tree, was the ensemble implementation we used^17^. Neural network models, which allow for non-linear coefficient fitting and can perform well in large multi-dimensional data, were also trained.

Models that could accommodate multi-output regression were directly fit, while those that could not, were modified using two separate strategies to make predictions in the multi-output space. In the first strategy, individual models were trained to target each minute after induction separately, with each model using only the 118 predictor variables. In the second strategy, we used a composite chained regression model. In this approach, sequential models were trained, where the first model’s prediction (targeting t_1_) was provided as an input variable to the second model (targeting t_2_), whose output was provided as an input variable to the third model, and so on in a chained fashion.

The ML models were trained using the scikit-learn implementation version 1.2.1 using Python version 3.11.4, ScikitLearn version 1.2.1^18^, NumPy package1.24.3 and Pandas package version 2.0.3.

#### Evaluation

The models were evaluated using R-squared value, mean absolute error, mean squared error, and mean absolute percentage error. The R-Squared value was used for tuning. The R-squared value calculates the proportion of variance that is explained by the model. A value closer to 1 indicates better performance. The mean absolute error is the sum of the absolute value of the residuals. The mean squared error s is the sum of the squared value of the residuals. The mean absolute percentage error calculates the mean absolute value of the percentage error of the predictions. Since the target has a dimension of 15 (at each minute from t_1_ through t_15_), the performance values from all 15 predictions were averaged and reported.

### Development and usability of a prototype tool

A series of semi-structured interviews of select anesthesiologists at UPMC was conducted via videoconferencing. The questionnaire used for the interview is in Supplementary File 4.

Questions were formulated to understand the elements that go into the decision-making by anesthesiologists at multiple points in time during their normal workflow (while reviewing *Cerner^TM^ Surgical Powerchart,* after visiting patients, and finally, prior to induction in the operating room). The interviews were transcribed, and qualitative analysis using an affinity diagram was created to help identify key concepts. This exploration was used to inform the design of a prototype user interface for the clinical tool.

## RESULTS

### Exploratory analysis results

A total of 93,037 anesthetic episodes were included in the analysis; these episodes occurred between January 1, 2013, and December 31, 2020. The distributions of predictor and target variables across the training, validation, and test datasets were similar (see Table 2). Detailed distributions of all the predictor variables are presented in Supplementary Files 1, 2, and 3.

**Table 2.**
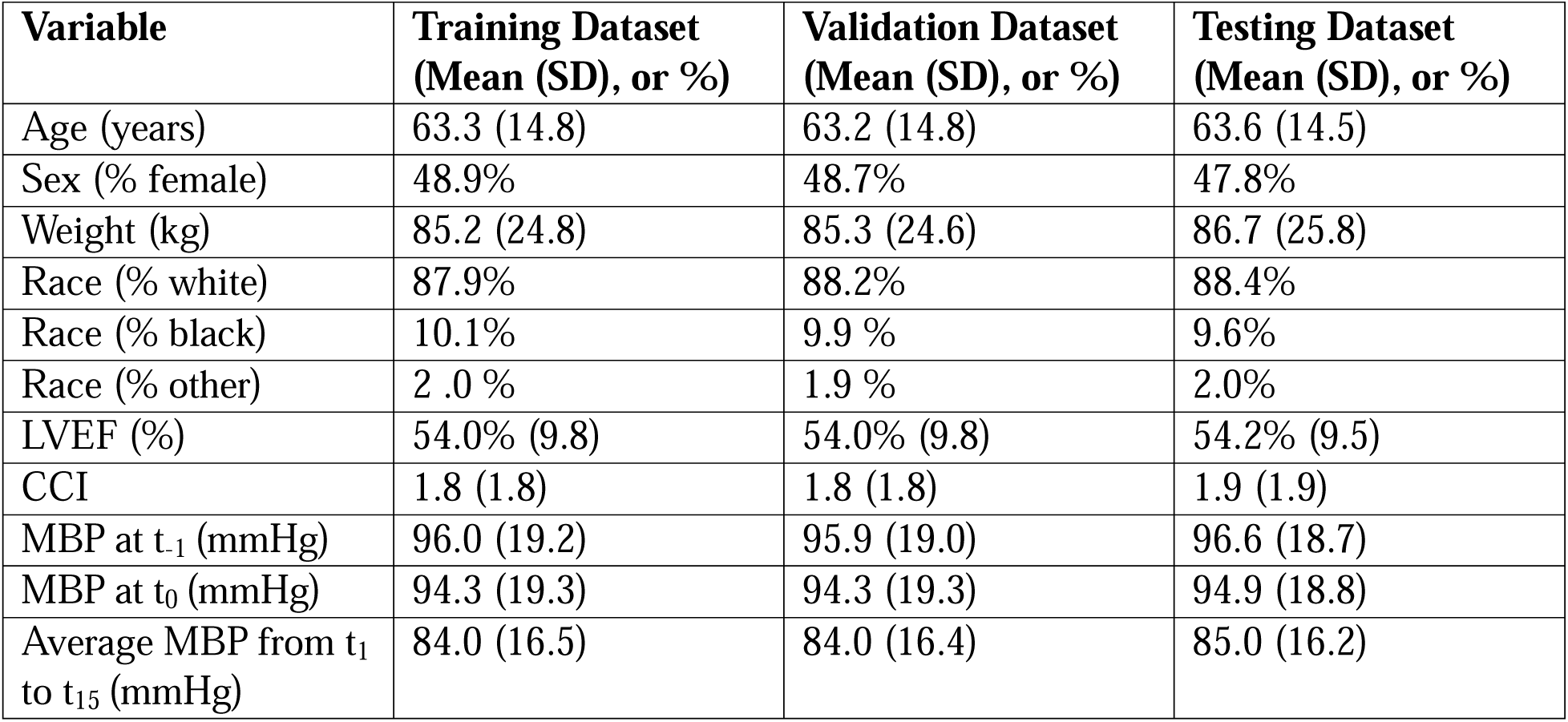
Characteristics of the training, validation, and test datasets.

Post-induction hypotension was explored (see Figure 4). The MBP had a decreasing trend post-induction, as shown in Figure 4A. The mean MBP (SD) at time point t _-10_ (at 10 minutes prior to induction) was 99.2 mmHg (20.2 mmHg), and this gradually dropped to 79.9 mmHg (19.2 mmHg) at time t_15_ (at 15 minutes after induction). This difference was statistically significant (p < 0.001). There was a comparable distribution of MBP across each minute in the pre- and post-induction period (see Figure 4B).

The relationship between age and post-induction MBP in Figure 4C and 4E showed that extremely low post induction MBP and extremely high pre-induction MBPs were more likely in the older age groups. The relationship between ejection fraction and post-induction MBP is shown in Figure 4D and 4F, which revealed that, on average, patients with lower ejection fractions started off with lower MBPs and had a comparably smaller decrease in MBP in the post-induction period.

**Figure 4A.**
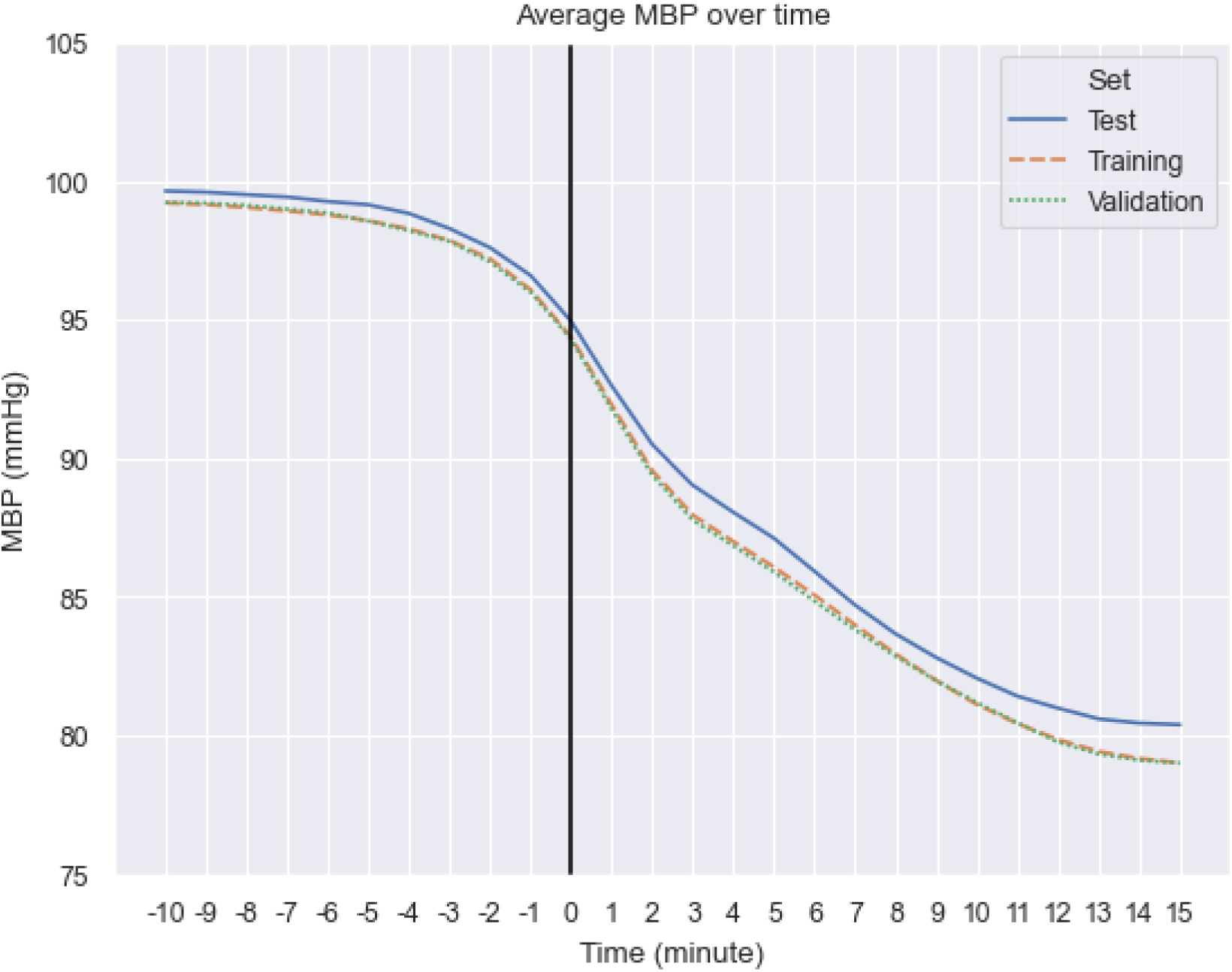
Mean MBP over time for training, validation, and test datasets.

**Figure 4B.**
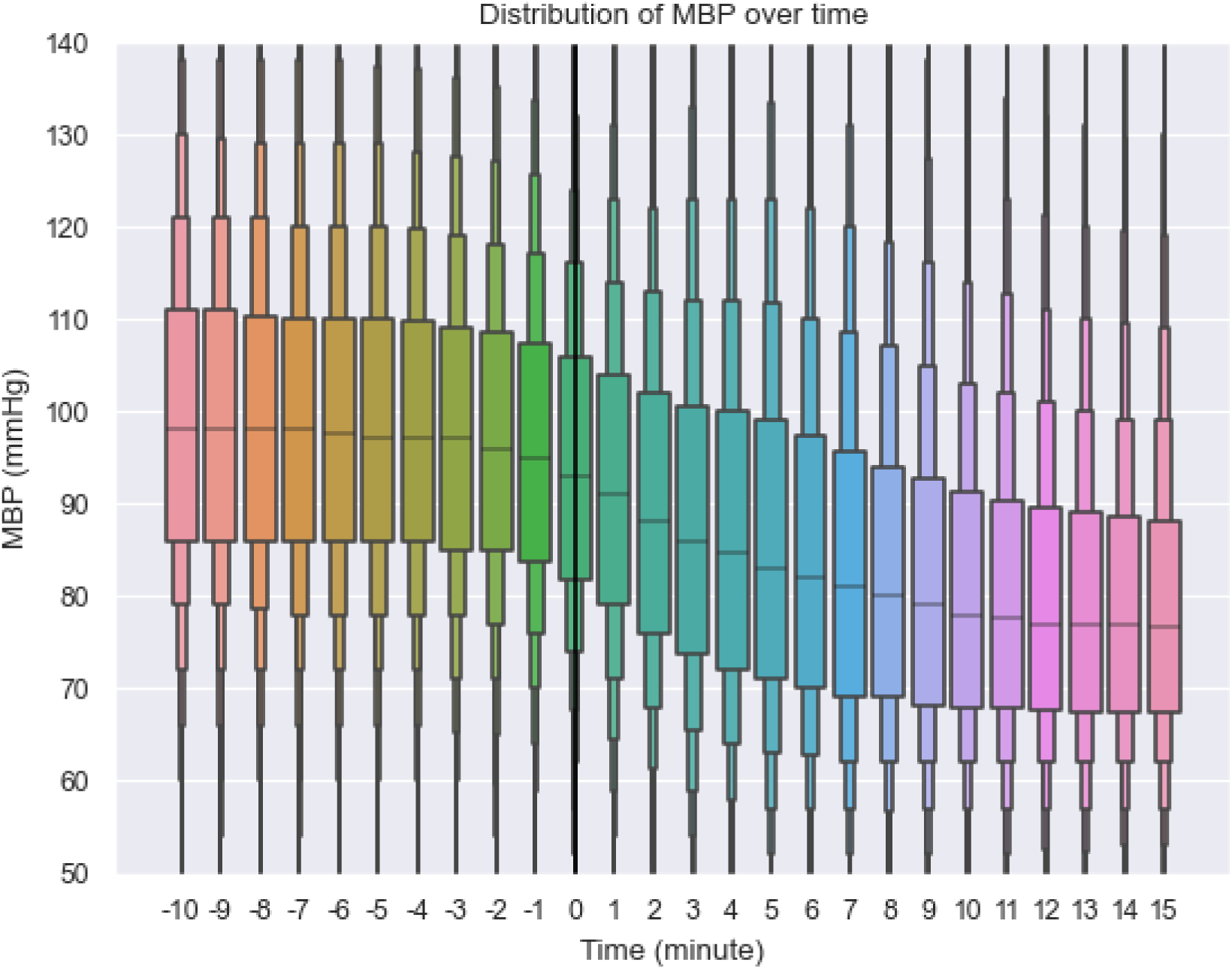
Distribution of MBP over time for the full dataset.

**Figure 4C.**
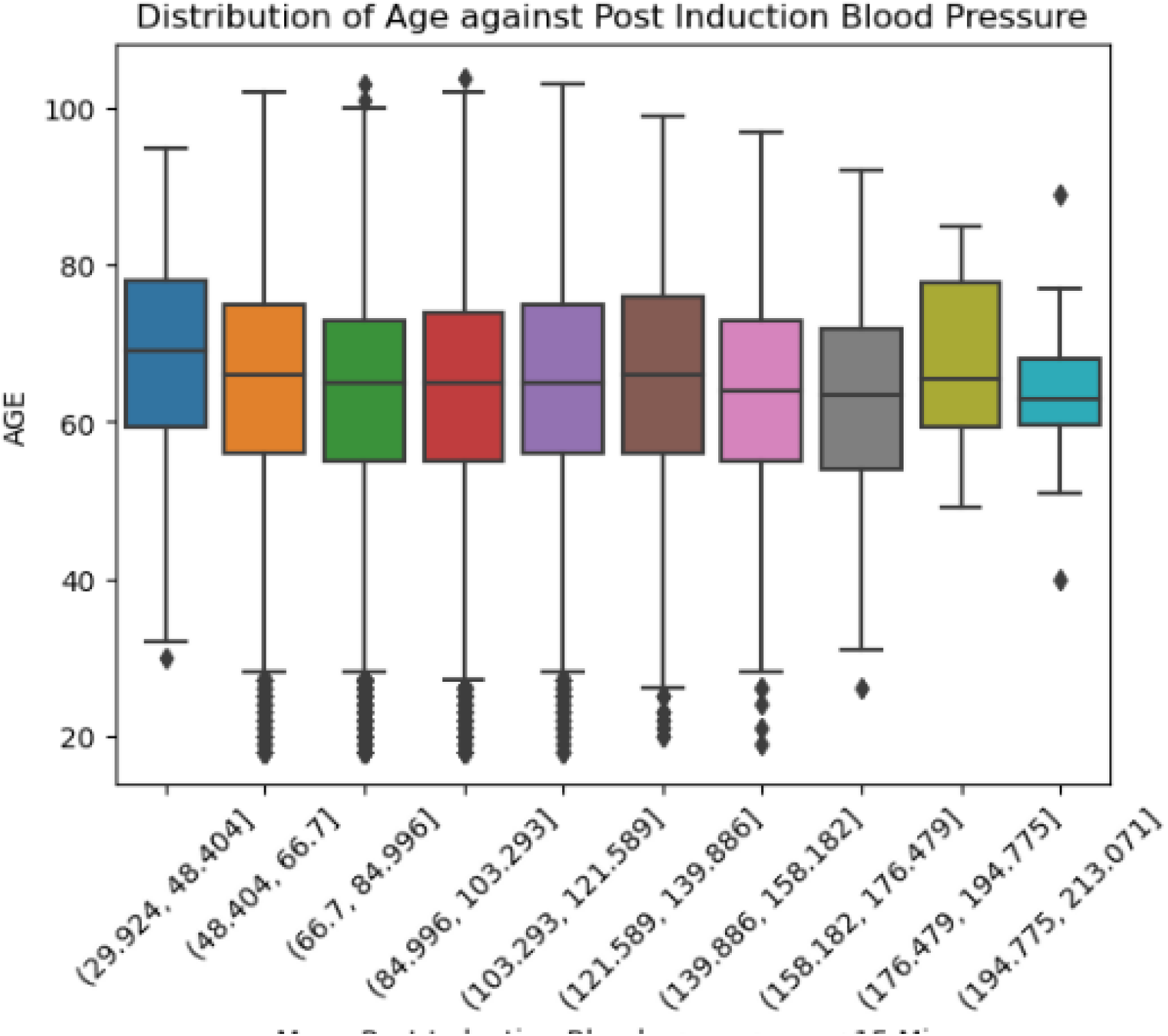
Age and distribution of post-induction MBP for the full dataset.

**Figure 4D.**
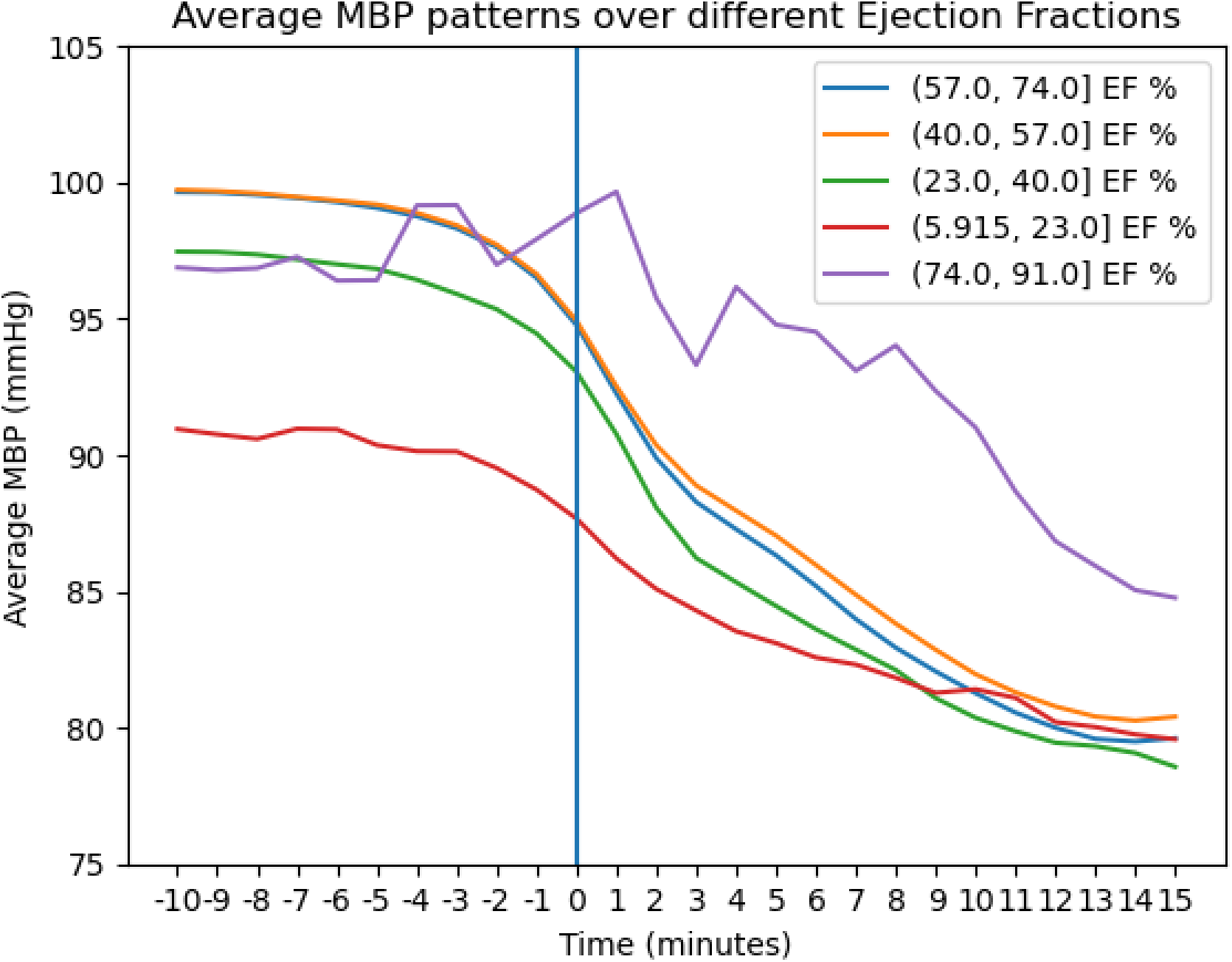
Ejection fractions against mean MBP over time in the full dataset.

**Figure 4E.**
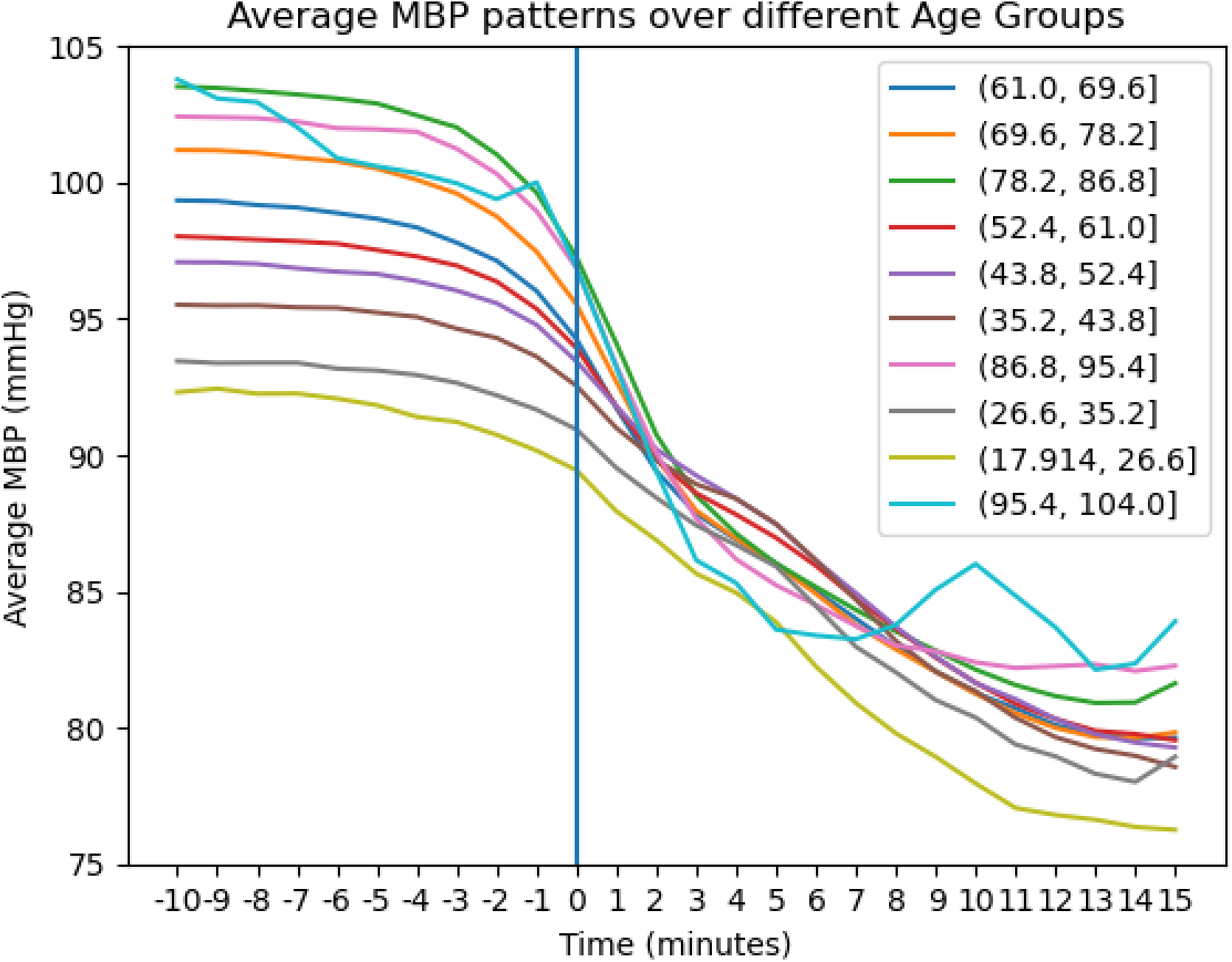
Patterns of MBP over induction, while comparing different groups.

**Figure 4F.**
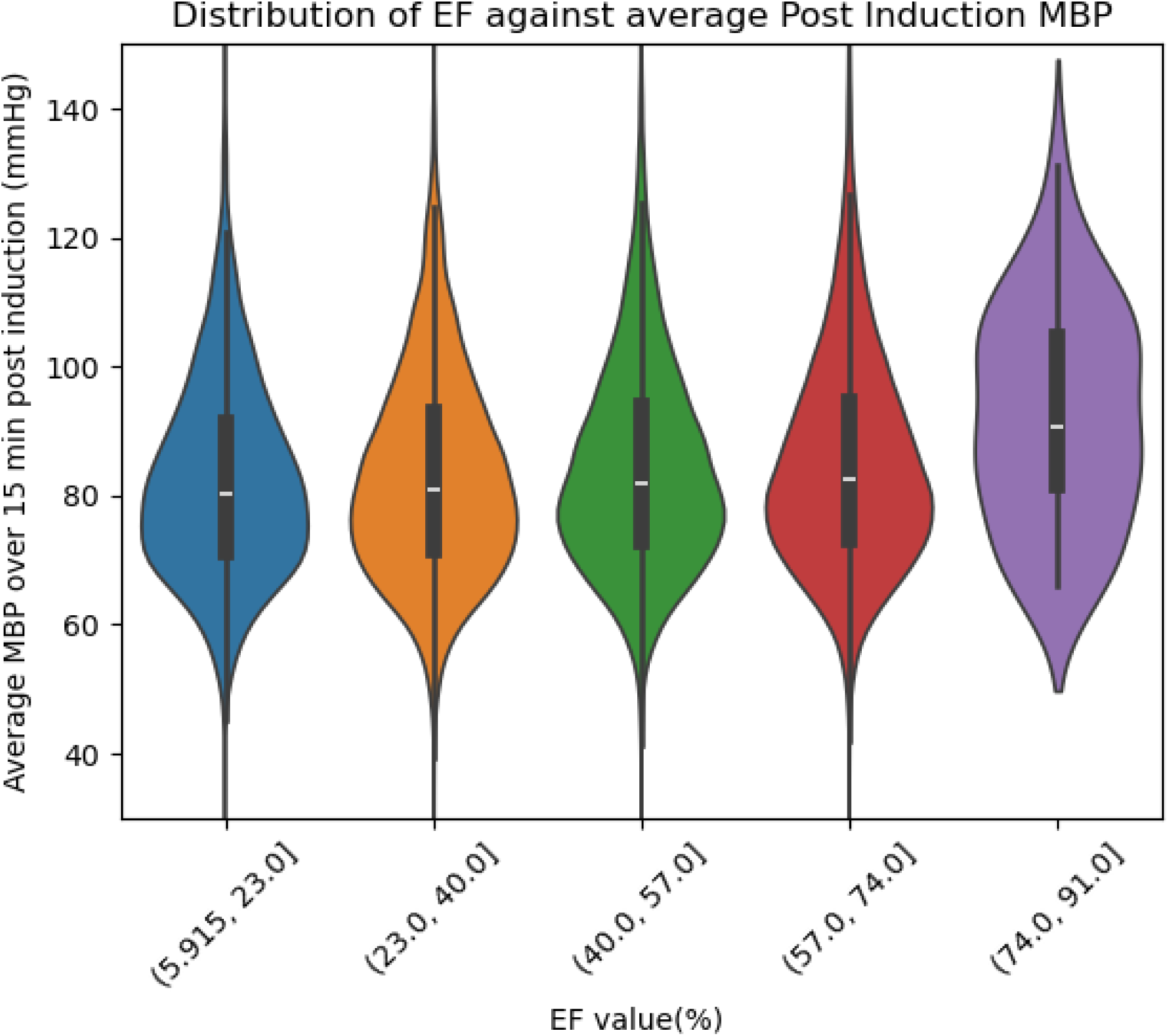
Post-induction MBP distribution against ejection fraction in the full dataset.

### Model performance results

Based on the validation dataset performance, we chose the best-performing models and evaluated them on the test dataset (see Table 3). Overall, the hist-gradient boosted model had the best performance, followed by neural network and linear regression models. Models using a multi-output strategy generally performed better than the chained regression strategy. The hist-gradient boosted model with a multi-output strategy, had a mean R-squared value of 0.32, a mean absolute error of 11.8, a mean percentage error of 14%, and a mean squared error of 285.05.

**Table 3.** Performance of the models on the test dataset.

| **Model** |  | **R-Squared** | **Mean Absolute Error** | **Mean Percentage Error** | **Mean Squared Error** |
| --- | --- | --- | --- | --- | --- |
| Linear Regression: | Multioutput | 0.26 | 12.4 | 0.15 | 299.73 |
|  | Chained | 0.26 | 12.4 | 0.15 | 299.73 |
| Ridge Regression | Multioutput | 0.26 | 12.4 | 0.15 | 299.66 |
|  | Chained | 0.26 | 12.4 | 0.15 | 299.72 |
| Bayesian Ridge Regression | Multioutput | 0.26 | 12.4 | 0.15 | 299.69 |
|  | Chained | 0.25 | 12.5 | 0.15 | 303.16 |
| Tree-based Models: XGB | Multioutput | 0.32 | 11.8 | 0.14 | 285.05 |
|  | Chained | 0.28 | 11.6 | 0.15 | 325.16 |
| Neural Network: | 1 Hidden Layer | 0.27 | 12.3 | 0.15 | 294.75 |
|  | 5 Hidden Layers | 0.14 | 13.3 | 0.16 | 347.59 |

**Table 4.** Key concepts identified via affinity diagramming of the interview transcripts.

| Category | Description | Quote |
| --- | --- | --- |
| Slow performance | Many feel the EHR system currently is very slow | “.. It takes so long for the *** notes from *** to load in ***..” |
| Drug dose Planning | Anesthesiologists decide the drug doses usually in the operating room just before induction | “ .. I decide the dose usually when I am picking up the medication.” |
| Difficulty gathering data | Many of participants had difficulty gathering data from various sources in the EHR system | “It is sometimes hard to find outside results” |
| Summarization | Review involves visiting different tabs in the EHR system in search of common data in different orders | “I first go to Allergies... then I go to labs... then I go to vital signs..” |

Model evaluation was also performed across each minute post-induction. Figure 5 shows the R-squared values of the best-performing models for each post-induction minute. On average, the performance decreased over time for most models. The XGB model with the multi-output strategy was relatively resilient to the drop in performance over time compared to the other models.

**Figure 5.**
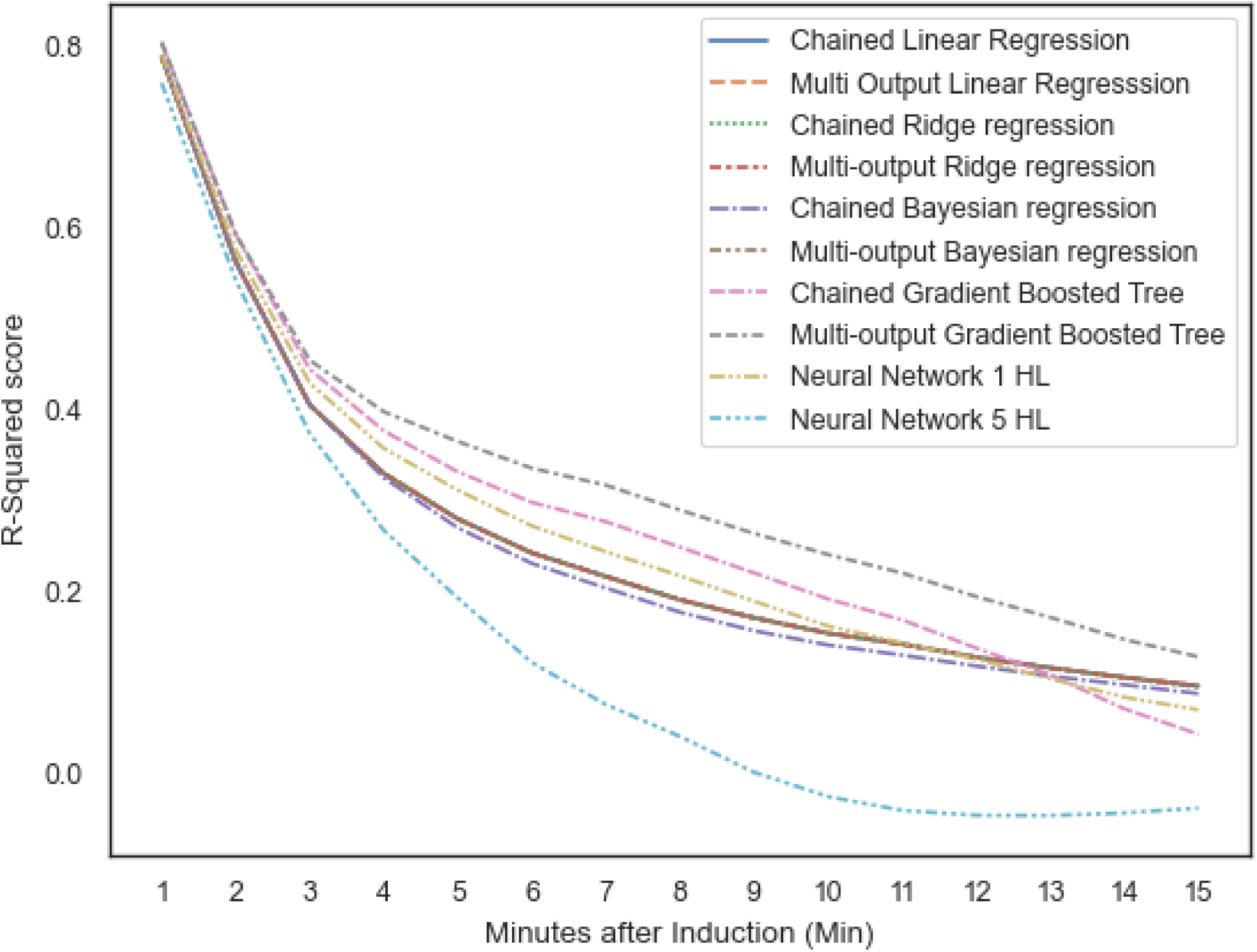
Decrease in R-squared performance over time on the test set. The included models are listed in Table 3.

Visualization of the predictions for some anesthetic episodes was performed. Overall, the model appeared to track the general trend in the MBP, but when there was a higher minute-to-minute variability in the MBP, it was unable to predict well. A sample of four episodes from the test set is shown in Figure 6, illustrating this.

**Figure 6.**
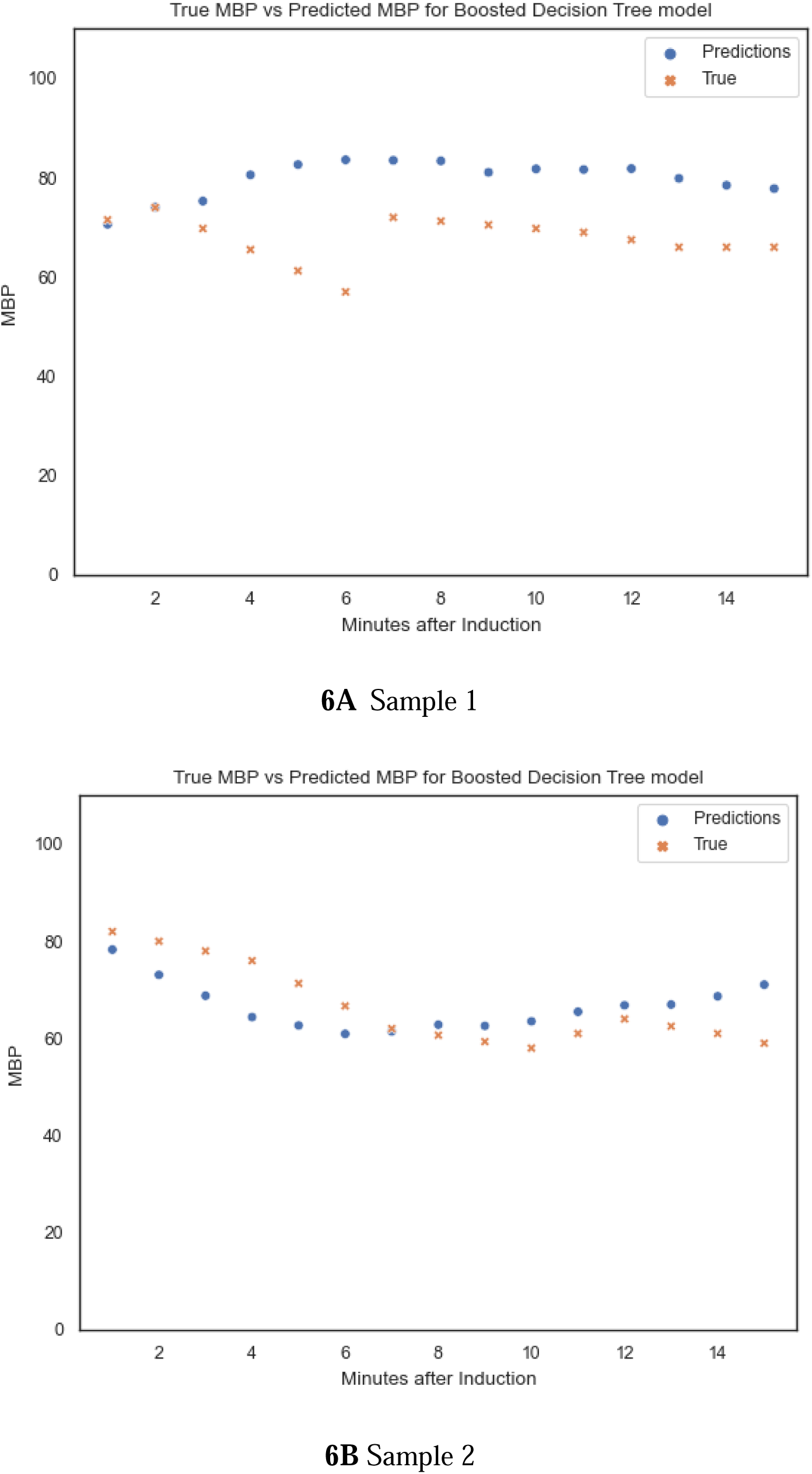
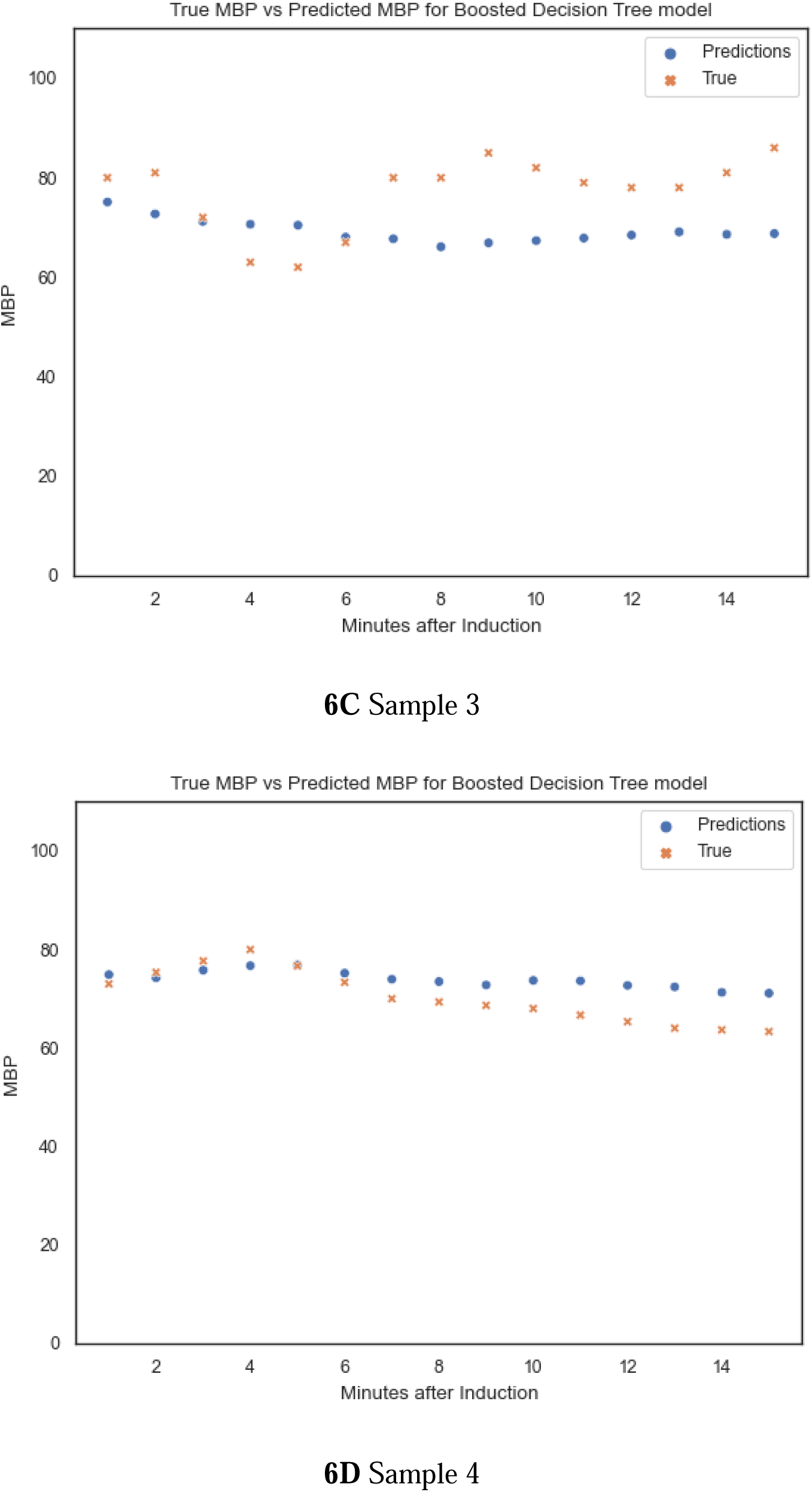
Visualizations of four sample predictions.

In Figure 6, golden dots indicate true values of MBP, while blue dots indicate predicted values by the XGB model. When the minute-to-minute variability is small, as seen in B and D, the model predicts better. When the minute-to-minute variability is high, as seen in A and C, the prediction errors are higher.

### Usability study results

Six anesthesiologists consented to the interviews, each lasting 20-30 minutes. Key design concepts that could impact the prototype were identified from the affinity diagram derived from the interview transcripts. Based on the results from the affinity diagramming, the best place to implement the induction tool appears to be in the operative room, ideally next to the medication cart. The tool should be simple to use and should display preoperative and intraoperative inputs as well as predictions. The speed of performance of the tool was identified as a critical factor for the users.

The prototype included a header displaying preoperative inputs about the patient, including identifiers, surgical information, weight, etc. A medication input section allowed the user to input doses of the medication they plan to administer. The display section on the bottom right displays the predicted MBP over time. A screenshot of the display of the prototype of the tool is shown in Figure 7.

**Figure 7.**
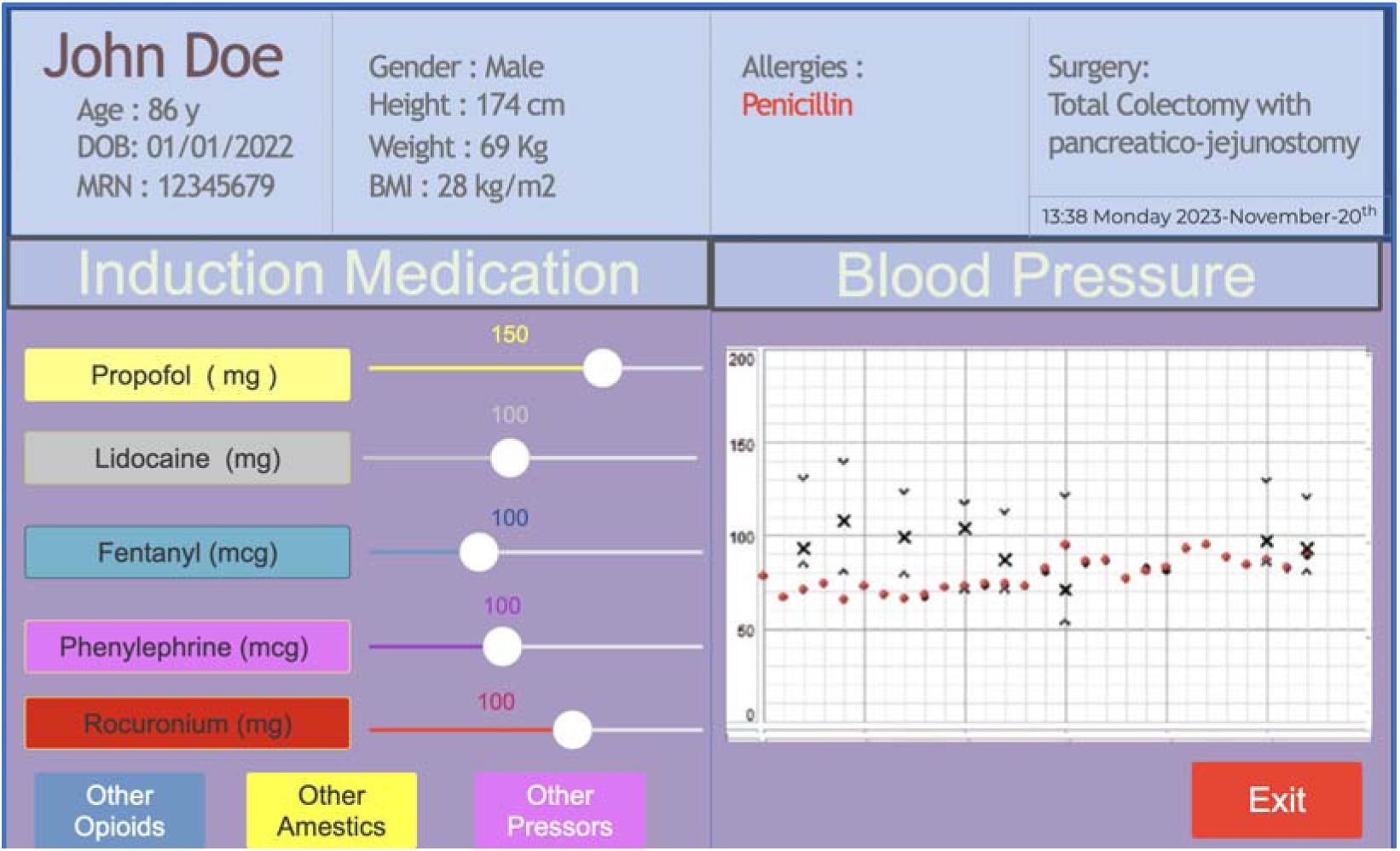
Display of the prototype clinical tool.

## DISCUSSION

Post-induction hypotension remains a significant clinical obstacle in the practice of anesthesiology. In contrast to hypotension that may occur during other phases of the anesthetic episode, post-induction hypotension can be attributed entirely to the induction process and is an iatrogenic problem. Expert anesthesiologists with years of training can identify patients who are at risk of developing hypotension after induction and modify the induction medications accordingly.

Identifying a minimum MBP below which outcomes deteriorate has proven elusive ^6,11^. This is partly because blood pressure is a surrogate of blood flow, which is the determinant of tissue perfusion. Previous studies have employed classification methods to predict post-induction MBP as normal or low^1,9,12^. Even if this approach proves effective in modeling, we believe its applicability as a clinical tool is limited. In contrast, our strategy directly targets MBP, thus avoiding the issue of choosing a threshold. It also allows the anesthesiologist to experiment with the different medication inputs in a continuous way to plan the induction process.

In our exploration of the various models, ensemble decision tree models were found to have the best performance. This aligns with other reports of using ML in healthcare, where data can have high variability^11^. The advantage also extends to the deployment of these models. Decision tree models have a relatively faster prediction time and can allow for near-continuous input and output display, as shown in our prototype^17^.

The challenges we faced in this project were primarily in the area of data preprocessing. To make these models we had to assume induction is an instantaneous point in time as annotated in the intraoperative record while, in fact, it occurs over several minutes as drugs are given gradually.

This issue is reflected in our peri-induction medication preprocessing. We were compelled to use all administrations from the peri-induction period, including the post-induction period, so that all induction drugs were captured. This could be a source of potential overfitting in the model, since theoretically it exposes the model to events a few minutes in the future as it predicts the blood pressure.

The rationale for using this technique was twofold. First, as briefly discussed in the section in preprocessing, we believe that induction medication documentation lags behind administration, since most clinicians are performing multiple tasks during induction and do not prioritize documentation at that time. To be able to capture all induction medications, we had to include medication administration up to 10 minutes after induction annotation. Second, we wanted to keep the first iteration of the CDS tool simple to use. A CDS tool that uses both the medication dose and the timing of administration as an input may perform better but would be unfeasible to deploy. When planning induction, anesthesiologists would have to input not only the doses they plan to administer but also the exact minute that they plan to administer it. This would be cumbersome and impractical and does not reflect how induction of anesthesia in the real world is conducted.

Additionally, our models lacks some key variables that are known to impact MBP, such as daily opioid intake, which were not readily available to us.

Future work is needed in a few areas. Further work is needed in modeling to investigate whether performance can be improved. Additional evaluations are needed to characterize model drift over time and model generalizability across sites. Before deployment, trials are needed to show that post-induction MBP predictions will enable anesthesiologists to prevent post-induction hypotension and that prevention of post-induction hypotension will improve patient outcomes. It is hard to make a case at this time that moving in the needle on induction will move the needle on outcomes.

## CONCLUSION

In this project, we developed an ML model to predict post-induction MBP that had moderately good performance. The usability study provided supported the hypothesis that anesthesiologists will find a clinical tool that predicts post-induction MBP useful during induction.

## Data Availability

All data produced in the present study are available upon reasonable request to the authors after IRB review

## ACKNOWLEDGMENTS, COMPETING INTERESTS, FUNDING AND ALL OTHER REQUIRED STATEMENTS

This research was supported in part by the University of Pittsburgh Center for Research Computing through the resources provided. Specifically, this work used the HTC cluster, which is supported by NIH award number S10OD028483. There are no other competing interests to declare from any of the authors, and no funding was secured for performing the study.

## SUPPLEMENTS

Supplement 1. Prototype HTML implementation and PDF file

