## Supplement File 1 for "Post Induction Hypotension prediction during general anesthesia using Machine Learning Techniques": readme.txt

This is a prototype built with Proto.io.

Use the preview.html file to preview your prototype offline as you would in the Proto.io player: wrapped in its device skin, and in a fit-to-screen mode.

If you have the skills to customize the code you can edit the preview.html file to change the look and feel of the preview page.

We strongly suggest you make no changes to the files in the /src folder.
