## Supplement File 1 for "Post Induction Hypotension prediction during general anesthesia using Machine Learning Techniques": data.js

```
prx.xdata = {"cc":223,"data":{"template":{"id":"","stateid":""},"items":[{"name":"rectangle","type":"rectangle","lib":"common","caption":"Rectangle 1","backgroundColor":{"type":"solid","color":"rgba(193,212,241,1)"},"borderWidth":"8","borderColor":{"type":"solid","color":"rgba(30,77,130,1)"},"borderRadius":0,"imgSrc":{"fileId":"","name":"","assetType":"","bucketsource":"","url":""},"backgroundSizeType":"cover","backgroundSizeValue":35,"width":1440,"height":249,"lineHeight":20,"lineHeightAuto":true,"actions":[],"textFont":"Arial,sans-serif","textFontStyle":"400","textSize":16,"textColor":"#383838","textProperties":[],"textSpacing":0,"textAlign":"center","text":"","typeName":"rectangle","left":0,"top":0,"vpos":"top","hpos":"left","visible":true,"id":"box-193","group":"root","zindex":1,"wtype":"fixed","htype":"fixed","opacity":1,"borderStyle":"solid","rotation":0,"scale":1,"aspectratio":false},{"name":"text","type":"text","lib":"common","caption":"Gender : Male...","text":"Gender : Male","textFont":"Trebuchet MS,sans-serif","textFontStyle":"400","textProperties":[],"textSize":"32","textColor":{"type":"solid","color":"rgba(113,113,113,1)"},"backgroundColor":"none","width":186,"height":39,"lineHeight":39,"textAlign":"left","textSpacing":"-1.6","enableShadow":false,"autoResize":true,"v2":true,"left":379,"top":47,"vpos":"top","hpos":"left","visible":true,"group":"root","id":"box-194","zindex":8,"wtype":"fixed","htype":"fixed","opacity":1,"propagateEvents":false,"lineHeightAuto":true,"rotation":0,"scale":1,"aspectratio":false},{"name":"text","type":"text","lib":"common","caption":"DOB: 01/01/2022...","text":"DOB: 01/01/2022","textFont":"Trebuchet MS,sans-serif","textFontStyle":"400","textProperties":[],"textSize":"32","textColor":{"type":"solid","color":"rgba(113,113,113,1)"},"backgroundColor":"none","width":230,"height":39,"lineHeight":39,"textAlign":"left","textSpacing":"-1.6","enableShadow":false,"autoResize":true,"v2":true,"left":76,"top":130,"vpos":"top","hpos":"left","visible":true,"group":"root","id":"box-195","zindex":8,"wtype":"fixed","htype":"fixed","opacity":1,"propagateEvents":false,"lineHeightAuto":true,"rotation":0,"scale":1,"aspectratio":false},{"name":"text","type":"text","lib":"common","caption":"Height : 174 cm...","text":"Height : 174 cm&nbsp;","textFont":"Trebuchet MS,sans-serif","textFontStyle":"400","textProperties":[],"textSize":"32","textColor":{"type":"solid","color":"rgba(113,113,113,1)"},"backgroundColor":"none","width":216,"height":39,"lineHeight":39,"textAlign":"left","textSpacing":"-1.6","enableShadow":false,"autoResize":true,"v2":true,"left":379,"top":86,"vpos":"top","hpos":"left","visible":true,"group":"root","id":"box-196","zindex":8,"wtype":"fixed","htype":"fixed","opacity":1,"propagateEvents":false,"lineHeightAuto":true,"rotation":0,"scale":1,"aspectratio":false},{"name":"text","type":"text","lib":"common","caption":"Surgery:\nTotal Colec...","text":"Surgery:&nbsp;<br>Total Colectomy with&nbsp;<br>pancreatico-jejunostomy&nbsp;","textFont":"Trebuchet MS,sans-serif","textFontStyle":"400","textProperties":[],"textSize":"32","textColor":{"type":"solid","color":"rgba(113,113,113,1)"},"backgroundColor":"none","width":336,"height":117,"lineHeight":39,"textAlign":"left","textSpacing":"-1.6","enableShadow":false,"autoResize":true,"v2":true,"left":1084,"top":47,"vpos":"top","hpos":"left","visible":true,"group":"root","id":"box-197","zindex":8,"wtype":"fixed","htype":"fixed","opacity":1,"propagateEvents":false,"lineHeightAuto":true,"rotation":0,"scale":1,"aspectratio":false},{"name":"text","type":"text","lib":"common","caption":"Age : 86 y","text":"Age : 86 y&nbsp;","textFont":"Trebuchet MS,sans-serif","textFontStyle":"400","textProperties":[],"textSize":"32","textColor":{"type":"solid","color":"rgba(113,113,113,1)"},"backgroundColor":"none","width":139,"height":39,"lineHeight":39,"textAlign":"left","textSpacing":"-1.6","enableShadow":false,"autoResize":true,"v2":true,"left":76,"top":91,"vpos":"top","hpos":"left","visible":true,"group":"root","id":"box-198","zindex":8,"wtype":"fixed","htype":"fixed","opacity":1,"propagateEvents":false,"lineHeightAuto":true,"rotation":0,"scale":1,"aspectratio":false},{"name":"text","type":"text","lib":"common","caption":"Allergies :\nPenicill...","text":"Allergies :<br><span style=\"color: rgba(249,51,36,1);\">Penicillin</span><br><br>","textFont":"Trebuchet MS,sans-serif","textFontStyle":"400","textProperties":[],"textSize":"32","textColor":{"type":"solid","color":"rgba(113,113,113,1)"},"backgroundColor":"none","width":203,"height":117,"lineHeight":39,"textAlign":"left","textSpacing":"-1.6","enableShadow":false,"autoResize":false,"v2":true,"left":766,"top":47,"vpos":"top","hpos":"left","visible":true,"group":"root","id":"box-199","zindex":8,"wtype":"fixed","htype":"fixed","opacity":1,"propagateEvents":false,"lineHeightAuto":true,"rotation":0,"scale":1,"aspectratio":false},{"name":"text","type":"text","lib":"common","caption":"BMI : 28 kg/m2...","text":"BMI : 28 kg/m2","textFont":"Trebuchet MS,sans-serif","textFontStyle":"400","textProperties":[],"textSize":"32","textColor":{"type":"solid","color":"rgba(113,113,113,1)"},"backgroundColor":"none","width":199,"height":39,"lineHeight":39,"textAlign":"left","textSpacing":"-1.6","enableShadow":false,"autoResize":true,"v2":true,"left":379,"top":171,"vpos":"top","hpos":"left","visible":true,"group":"root","id":"box-200","zindex":8,"wtype":"fixed","htype":"fixed","opacity":1,"propagateEvents":false,"lineHeightAuto":true,"rotation":0,"scale":1,"aspectratio":false},{"name":"text","type":"text","lib":"common","caption":"Weight : 69 Kg...","text":"Weight : 69 Kg","textFont":"Trebuchet MS,sans-serif","textFontStyle":"400","textProperties":[],"textSize":"32","textColor":{"type":"solid","color":"rgba(113,113,113,1)"},"backgroundColor":"none","width":190,"height":39,"lineHeight":39,"textAlign":"left","textSpacing":"-1.6","enableShadow":false,"autoResize":true,"v2":true,"left":379,"top":130,"vpos":"top","hpos":"left","visible":true,"group":"root","id":"box-201","zindex":8,"wtype":"fixed","htype":"fixed","opacity":1,"propagateEvents":false,"lineHeightAuto":true,"rotation":0,"scale":1,"aspectratio":false},{"name":"verticalline","type":"verticalline","lib":"common","caption":"Vertical line 3","width":1,"height":229,"color":"A6A6A6","weight":1,"left":341,"top":10,"vpos":"top","hpos":"left","visible":true,"group":"root","id":"box-202","zindex":7,"wtype":"fixed","htype":"fixed","opacity":1,"borderStyle":"solid","rotation":0,"scale":1,"aspectratio":false},{"name":"verticalline","type":"verticalline","lib":"common","caption":"Vertical line 5","width":1,"height":229,"color":"A6A6A6","weight":1,"left":1064,"top":15,"vpos":"top","hpos":"left","visible":true,"group":"root","id":"box-203","zindex":7,"wtype":"fixed","htype":"fixed","opacity":1,"borderStyle":"solid","rotation":0,"scale":1,"aspectratio":false},{"name":"verticalline","type":"verticalline","lib":"common","caption":"Vertical line 6","width":1,"height":229,"color":"A6A6A6","weight":1,"left":733,"top":10,"vpos":"top","hpos":"left","visible":true,"group":"root","id":"box-204","zindex":7,"wtype":"fixed","htype":"fixed","opacity":1,"borderStyle":"solid","rotation":0,"scale":1,"aspectratio":false},{"name":"datetime","type":"datetime","lib":"common","caption":"Date/Time 1","width":376,"height":41,"backgroundColor":"none","borderWidth":"1","borderColor":{"type":"solid","color":"rgba(102,150,200,1)"},"textFont":"Montserrat","textFontStyle":"400","textSize":"20","textColor":"383838","textProperties":[],"textAlign":"center","dateFormat":"yearmonthday","dayFormat":"superscript","monthFormat":"long","yearFormat":"long","timeFormat":"24","dateGeneralFormat":"timedate","dayNameFormat":"long","dateSeparator":"-","left":1064,"top":201,"vpos":"top","hpos":"left","visible":true,"group":"root","id":"box-205","zindex":5,"wtype":"fixed","htype":"fixed","opacity":1,"rotation":0,"scale":1,"aspectratio":false},{"name":"text","type":"text","lib":"common","caption":"MRN : 12345679...","text":"MRN : 12345679&nbsp;","textFont":"Trebuchet MS,sans-serif","textFontStyle":"400","textProperties":[],"textSize":"32","textColor":{"type":"solid","color":"rgba(113,113,113,1)"},"backgroundColor":"none","width":218,"height":39,"lineHeight":39,"textAlign":"left","textSpacing":"-1.6","enableShadow":false,"autoResize":true,"v2":true,"left":76,"top":171,"vpos":"top","hpos":"left","visible":true,"group":"root","id":"box-206","zindex":4,"wtype":"fixed","htype":"fixed","opacity":1,"propagateEvents":false,"lineHeightAuto":true,"rotation":0,"scale":1,"aspectratio":false},{"name":"verticalline","type":"verticalline","lib":"common","caption":"Vertical line 1","width":1,"height":658,"color":{"type":"solid","color":"rgba(70,127,215,1)"},"weight":1,"left":734,"top":242,"vpos":"top","hpos":"left","visible":true,"group":"root","id":"box-207","zindex":3,"wtype":"fixed","htype":"fixed","opacity":1,"borderStyle":"solid","rotation":0,"scale":1,"aspectratio":false},{"name":"text","type":"text","lib":"common","caption":"John Doe","text":"John Doe&nbsp;","textFont":"Trebuchet MS,sans-serif","textFontStyle":"400","textProperties":[],"textSize":"64","textColor":{"type":"solid","color":"rgba(117,85,88,1)"},"backgroundColor":"none","width":272,"height":78,"lineHeight":78,"textAlign":"left","textSpacing":"-1.6","enableShadow":false,"autoResize":true,"v2":true,"left":40,"top":15,"vpos":"top","hpos":"left","visible":true,"id":"box-208","group":"root","zindex":2,"wtype":"fixed","htype":"fixed","opacity":1,"propagateEvents":false,"lineHeightAuto":true,"rotation":0,"scale":1,"aspectratio":false},{"name":"basic_textfield","type":"basic_textfield","lib":"generic2","caption":"Text Field 2","width":733,"height":77,"value":"Induction Medication","placeholder":"Placeholder","inputtype":"text","backgroundColor":{"type":"solid","color":"rgba(177,190,227,1)"},"textFont":"Arial,sans-serif","textFontStyle":"400","textSize":"64","textColor":{"type":"solid","color":"rgba(233,240,215,1)"},"textProperties":[],"placeholderColor":"c1c8c9","borderWidth":"5","borderStyle":"solid","borderRadius":4,"borderColor":{"type":"solid","color":"rgba(85,85,85,1)"},"textAlign":"center","left":2,"top":242,"vpos":"top","hpos":"left","visible":true,"id":"box-209","group":"root","zindex":11,"wtype":"fixed","htype":"fixed","opacity":1,"rotation":0,"scale":1,"aspectratio":false},{"name":"basic_textfield","type":"basic_textfield","lib":"generic2","caption":"Text Field 3","width":698,"height":77,"value":"Blood Pressure","placeholder":"Placeholder","inputtype":"text","backgroundColor":{"type":"solid","color":"rgba(177,190,227,1)"},"textFont":"Arial,sans-serif","textFontStyle":"400","textSize":"64","textColor":{"type":"solid","color":"rgba(233,240,215,1)"},"textProperties":[],"placeholderColor":"c1c8c9","borderWidth":"5","borderStyle":"solid","borderRadius":4,"borderColor":{"type":"solid","color":"rgba(85,85,85,1)"},"textAlign":"center","left":733,"top":244,"vpos":"top","hpos":"left","visible":true,"id":"box-210","group":"root","zindex":11,"wtype":"fixed","htype":"fixed","opacity":1,"rotation":0,"scale":1,"aspectratio":false},{"name":"basic_textfield","type":"basic_textfield","lib":"generic2","caption":"Text Field 13","width":338,"height":55,"value":"Etomidate (mg)","placeholder":"Placeholder","inputtype":"text","backgroundColor":{"type":"solid","color":"rgba(255, 255, 0,1)"},"textFont":"Arial,sans-serif","textFontStyle":"400","textSize":"28","textColor":{"type":"solid","color":"rgba(56,56,56,1)"},"textProperties":[],"placeholderColor":"c1c8c9","borderWidth":1,"borderStyle":"solid","borderRadius":4,"borderColor":{"type":"solid","color":"rgba(27,77,87,1)"},"textAlign":"center","left":31,"top":374,"vpos":"top","hpos":"left","visible":true,"group":"root","id":"box-211","zindex":10,"wtype":"fixed","htype":"fixed","opacity":1,"rotation":0,"scale":1,"aspectratio":false,"actions":[{"title":"click ","type":"click","actionId":"change-property","callback":true,"pageId":"2","itemId":"box-67","propertyId":"value","valueType":"customValue","newvalue":"Etomidate (mg)","delay":"0","guid":"647d5bf3-f0f4-48d3-af03-d284f2cf0c7d","callbackAction":{"actionId":"go-to-page","callback":false,"pageId":"2","animation":"fade","animationEasing":"Cubic.easeOut","duration":"400","delay":"0","guid":"20355424-a25c-43d7-ac5b-9534c2a54ab6","clb_disabled":false},"sort":0}]},{"name":"basic_textfield","type":"basic_textfield","lib":"generic2","caption":"Text Field 14","width":338,"height":55,"value":"Ketamine (mg)","placeholder":"Placeholder","inputtype":"text","backgroundColor":{"type":"solid","color":"rgba(255, 255, 0,1)"},"textFont":"Arial,sans-serif","textFontStyle":"400","textSize":"28","textColor":{"type":"solid","color":"rgba(56,56,56,1)"},"textProperties":[],"placeholderColor":"c1c8c9","borderWidth":1,"borderStyle":"solid","borderRadius":4,"borderColor":{"type":"solid","color":"rgba(27,77,87,1)"},"textAlign":"center","left":31,"top":458,"vpos":"top","hpos":"left","visible":true,"group":"root","id":"box-212","zindex":10,"wtype":"fixed","htype":"fixed","opacity":1,"rotation":0,"scale":1,"aspectratio":false,"actions":[{"title":"click ","type":"click","actionId":"change-property","callback":true,"pageId":"2","itemId":"box-67","propertyId":"value","valueType":"customValue","newvalue":"Ketamine (mcg)","delay":"0","guid":"b02f8196-d229-45f9-adb2-8d4516b02bc8","callbackAction":{"actionId":"go-to-page","callback":false,"pageId":"2","animation":"fade","animationEasing":"Cubic.easeOut","duration":"400","delay":"0","guid":"201f9a80-15c6-489d-a60f-59a52490e59f","clb_disabled":false},"sort":0}]},{"name":"basic_textfield","type":"basic_textfield","lib":"generic2","caption":"Text Field 15","width":338,"height":55,"value":"Propofol (mg)","placeholder":"Placeholder","inputtype":"text","backgroundColor":{"type":"solid","color":"rgba(255, 255, 0,1)"},"textFont":"Arial,sans-serif","textFontStyle":"400","textSize":"28","textColor":{"type":"solid","color":"rgba(56,56,56,1)"},"textProperties":[],"placeholderColor":"c1c8c9","borderWidth":1,"borderStyle":"solid","borderRadius":4,"borderColor":{"type":"solid","color":"rgba(27,77,87,1)"},"textAlign":"center","left":31,"top":558,"vpos":"top","hpos":"left","visible":true,"group":"root","id":"box-223","zindex":10,"wtype":"fixed","htype":"fixed","opacity":1,"rotation":0,"scale":1,"aspectratio":false,"actions":[{"title":"click ","type":"click","actionId":"change-property","callback":true,"pageId":"2","itemId":"box-67","propertyId":"value","valueType":"customValue","newvalue":"Propofol (mg)","delay":"0","guid":"c1551700-f891-458a-b4de-7814c5c10f5c","callbackAction":{"actionId":"go-to-page","callback":false,"pageId":"2","animation":"fade","animationEasing":"Cubic.easeOut","duration":"400","delay":"0","guid":"bbdc0303-2cc4-405d-ab9a-91330997213c","clb_disabled":false},"sort":0}]},{"name":"image","type":"image","lib":"common","caption":"Screen Shot 2022-12-12 at 7.38.12 PM.png","imgSrc":{"fileId":"68d047acd614373a75ca2548bba92b44.png","assetType":"project","bucketsource":"main","name":"Screen Shot 2022-12-12 at 7.38.12 PM.png"},"repeat":false,"width":658,"height":398,"borderWidth":0,"borderRadius":0,"borderColor":"#D1D1D1","borderPos":"inside","propagateEvents":false,"actions":[],"overlay":false,"autoResize":false,"aspectratio":1.6506849315068493,"assistiveDescription":"","left":762,"top":359,"visible":true,"id":"box-213","group":"root","zindex":12,"hpos":"left","vpos":"top","wtype":"fixed","htype":"fixed","opacity":1,"borderStyle":"solid","rotation":0,"scale":1},{"name":"basic_button_text","type":"basic_button_text","lib":"generic2","caption":"Button with Background 5","width":177,"height":80,"backgroundColor":{"type":"solid","color":"rgba(249,51,36,1)"},"borderWidth":0,"borderColor":"#025abf","borderRadius":4,"text":"Back","textFont":"Arial,sans-serif","textFontStyle":"400","textSize":"36","textColor":"FFFFFF","textProperties":[],"textAlign":"center","iconpos":"none","left":472,"top":652,"vpos":"top","hpos":"left","visible":true,"id":"box-214","group":"root","zindex":13,"wtype":"fixed","htype":"fixed","opacity":1,"rotation":0,"scale":1,"aspectratio":false,"actions":[{"title":"Interaction 1","type":"click","actionId":"go-to-page","callback":false,"pageId":"2","animation":"fade","animationEasing":"Cubic.easeOut","duration":"400","delay":"0","guid":"dad0c4e3-784a-4804-ab16-f6ce57266e06","sort":0}]}]},"fonts":[{"value":"IBM Plex Sans","displayValue":"IBM Plex Sans","type":"googlefonts","variants":["100","100italic","200","200italic","300","300italic","400","400italic","500","500italic","600","600italic","700","700italic"]},{"value":"Open Sans","displayValue":"Open Sans","type":"googlefonts","variants":["300","300italic","400","400italic","600","600italic","700","700italic","800","800italic"]},{"value":"Montserrat","displayValue":"Montserrat","type":"googlefonts","variants":["100","100italic","200","200italic","300","300italic","400","400italic","500","500italic","600","600italic","700","700italic","800","800italic","900","900italic"]},{"value":"Roboto","displayValue":"Roboto","type":"googlefonts","variants":["100","100italic","300","300italic","400","400italic","500","500italic","700","700italic","900","900italic"]},{"value":"Source Sans Pro","displayValue":"Source Sans Pro","type":"googlefonts","variants":["200","200italic","300","300italic","400","400italic","600","600italic","700","700italic","900","900italic"]},{"value":"Anton","displayValue":"Anton","type":"googlefonts","variants":["400"]},{"value":"Dancing Script","displayValue":"Dancing Script","type":"googlefonts","variants":["400","500","600","700"]},{"value":"Pacifico","displayValue":"Pacifico","type":"googlefonts","variants":["400"]}],"custom_fonts_version":0,"grid":{"grid":{"size":50,"subdivisions":5},"gridsystem":{"portrait":{"col_number":3,"col_width":-20,"gutter_width":20,"margins":10},"landscape":{"col_number":3,"col_width":460,"gutter_width":20,"margins":10}},"guides":{"custom":{"landscape":{"horizontal":[{"id":"custom","dim":15}],"vertical":[]}}}},"savedColors":[{"type":"solid","color":"rgba(255, 255, 0,1)"},{"type":"solid","color":"rgba(160,47,201,1)"},{"type":"solid","color":"rgba(234,114,252,1)"}],"iSelectedPage":4,"iSelectedTemplate":-1,"iSelectedSymbol":-1,"iSelectedState":0,"libraries":{"common":true,"icons":true,"sounds":true,"generic2":true},"maxpageid":7,"maxsymbolid":1,"maxtemplateid":0,"project":{"startscreenlandscape":1,"navigationbar":"0","statusbar":"0","startscreen":1,"projectid":"e5f8f28e-1485-4050-8191-3d42cb769275","background":"#ffffff","screenTransition":{"transition":"fade","easing":"Cubic.easeOut","duration":"400"}},"sort":{"pages":{"type":"custom","custom":[]},"containers":{"type":"custom","custom":[]},"templates":{"type":"custom","custom":[]}},"variables":{"propofol_amount":{"type":"numeric","value":"","evaluate":false},"prop_dose":{"type":"numeric","value":"{propofol_amount} * 2.50","evaluate":true},"lido":{"value":"","type":"numeric","evaluate":true},"lido_dose":{"type":"numeric","value":"{lido} * 1.50","evaluate":true},"fent":{"value":"","type":"numeric","evaluate":true},"phen":{"value":"","type":"numeric","evaluate":true},"rocu":{"value":"","type":"numeric","evaluate":true},"screenshot":{"type":"numeric","value":"1","evaluate":false}},"v5":true,"v5x11":true,"richtext":true,"iscrolljs":true,"expandableCompat":true,"dropboxfs":true,"ios9":true,"galleryAssetsToStaticV2":true,"crossProjectTypekitCheck":"NONE","templatesCompatibility":"done","groupsV2":true,"stateTransitionUpgraded":true,"groupToContainerCompatibilityV2":true,"device":{"dpr":1,"portrait_applies":0,"landscape_applies":1,"portrait":[0,0],"landscape":[1440,900],"uuid":"133086fa-e022-48c1-9ac1-c999659222d9","defaultOrientation":"landscape"},"transition":"{\"duration\":250,\"delay\":0,\"easing\":\"out\"}","customEasings":{},"stateTransitionToMSUpgraded":true,"fontStyleCompatibility":true,"customGuidesToObjects":true,"soundEffectsInteraction":true,"pages":[{"navigationbar":0,"statusbar":0,"orientation":"landscape","actions":[],"title":"Screen 1","id":1,"states":[{"data":"[{\"name\":\"text\",\"type\":\"text\",\"lib\":\"common\",\"caption\":\"AIMoL\",\"text\":\"<span style=\\\"font-weight:400;font-style:normal\\\">AIMoL</span>\",\"textFont\":\"Trebuchet MS,sans-serif\",\"textFontStyle\":\"400\",\"textProperties\":[],\"textSize\":\"144\",\"textColor\":{\"type\":\"solid\",\"color\":\"rgba(20,20,20,1)\"},\"backgroundColor\":\"none\",\"width\":379,\"height\":167,\"lineHeight\":\"78\",\"textAlign\":\"left\",\"textSpacing\":\"-1.6\",\"enableShadow\":false,\"autoResize\":false,\"v2\":true,\"left\":539,\"top\":331,\"vpos\":\"top\",\"hpos\":\"left\",\"visible\":true,\"id\":\"box-1\",\"group\":\"root\",\"zindex\":2,\"wtype\":\"fixed\",\"htype\":\"fixed\",\"opacity\":1,\"propagateEvents\":false,\"lineHeightAuto\":false,\"rotation\":0,\"scale\":1,\"aspectratio\":false,\"kitName\":\"sections-kit\",\"kitTemplateName\":\"typography-heading-1\"},{\"name\":\"text\",\"type\":\"text\",\"lib\":\"common\",\"caption\":\"Anesthesia Induction...\",\"text\":\"Anesthesia Induction with Machine optimized learning\",\"textFont\":\"Arial,sans-serif\",\"textFontStyle\":\"400\",\"textProperties\":[],\"textSize\":\"36\",\"textColor\":{\"type\":\"solid\",\"color\":\"rgba(173,66,60,1)\"},\"backgroundColor\":\"none\",\"width\":862,\"height\":44,\"lineHeight\":44,\"textAlign\":\"left\",\"textSpacing\":0,\"enableShadow\":false,\"autoResize\":true,\"v2\":true,\"left\":297,\"top\":454,\"vpos\":\"top\",\"hpos\":\"left\",\"visible\":true,\"id\":\"box-44\",\"group\":\"root\",\"zindex\":3,\"wtype\":\"fixed\",\"htype\":\"fixed\",\"opacity\":1,\"propagateEvents\":false,\"lineHeightAuto\":true,\"rotation\":0,\"scale\":1,\"aspectratio\":false},{\"name\":\"basic_button_texticon\",\"type\":\"basic_button_texticon\",\"lib\":\"generic2\",\"caption\":\"Button - Text & Icon 1\",\"width\":332,\"height\":80,\"backgroundColor\":{\"type\":\"solid\",\"color\":\"rgba(101,180,206,1)\"},\"borderWidth\":\"4\",\"borderColor\":{\"type\":\"solid\",\"color\":\"rgba(177,217,231,1)\"},\"borderRadius\":4,\"text\":\"Plan your induction\",\"textFont\":\"Arial,sans-serif\",\"textFontStyle\":\"400\",\"textSize\":\"28\",\"textColor\":\"FFFFFF\",\"textProperties\":[],\"textAlign\":\"center\",\"iconpos\":\"right\",\"iconSize\":\"2\",\"img\":{\"fileId\":\"04fc9053132a2832640e4ab5705f5ab5.svg\",\"name\":\"arrow_forward.svg\",\"assetType\":\"icon\",\"bucketsource\":\"static\",\"url\":\"f1424098557988/78855ffe7a4c9efea4291339339586d3.svg\",\"targetSrc\":\"generated/04fc9053132a2832640e4ab5705f5ab5_ffffff.svg\",\"color\":\"ffffff\"},\"left\":569,\"top\":720,\"vpos\":\"top\",\"hpos\":\"left\",\"visible\":true,\"id\":\"box-89\",\"group\":\"root\",\"zindex\":4,\"wtype\":\"fixed\",\"htype\":\"fixed\",\"opacity\":1,\"rotation\":0,\"scale\":1,\"aspectratio\":false,\"actions\":[{\"title\":\"clicker\",\"type\":\"click\",\"actionId\":\"go-to-page\",\"callback\":false,\"pageId\":\"2\",\"animation\":\"fade\",\"animationEasing\":\"Cubic.easeOut\",\"duration\":\"400\",\"delay\":\"0\",\"guid\":\"06e0f2d5-ba76-495f-859d-c6ec1629f93d\",\"sort\":0}]}]","actions":[],"title":"State 1","template":{"id":"","stateid":""},"id":"E03587AD-FC9C-ECCB-10CF60A3AD0A5CD9","background":{"type":"solid","color":"rgba(235,191,198,1)"},"history":[],"groups":{}}]},{"id":2,"title":"Induction screen","orientation":"landscape","statusbar":"0","navigationbar":"0","actions":[],"states":[{"title":"State 1","background":{"type":"solid","color":"rgba(169,151,196,1)"},"data":"[{\"name\":\"rectangle\",\"type\":\"rectangle\",\"lib\":\"common\",\"caption\":\"Rectangle 1\",\"backgroundColor\":{\"type\":\"solid\",\"color\":\"rgba(193,212,241,1)\"},\"borderWidth\":\"8\",\"borderColor\":{\"type\":\"solid\",\"color\":\"rgba(30,77,130,1)\"},\"borderRadius\":0,\"imgSrc\":{\"fileId\":\"\",\"name\":\"\",\"assetType\":\"\",\"bucketsource\":\"\",\"url\":\"\"},\"backgroundSizeType\":\"cover\",\"backgroundSizeValue\":35,\"width\":1440,\"height\":249,\"lineHeight\":20,\"lineHeightAuto\":true,\"actions\":[],\"textFont\":\"Arial,sans-serif\",\"textFontStyle\":\"400\",\"textSize\":16,\"textColor\":\"#383838\",\"textProperties\":[],\"textSpacing\":0,\"textAlign\":\"center\",\"text\":\"\",\"typeName\":\"rectangle\",\"left\":0,\"top\":0,\"vpos\":\"top\",\"hpos\":\"left\",\"visible\":true,\"id\":\"box-45\",\"group\":\"root\",\"zindex\":1,\"wtype\":\"fixed\",\"htype\":\"fixed\",\"opacity\":1,\"borderStyle\":\"solid\",\"rotation\":0,\"scale\":1,\"aspectratio\":false},{\"name\":\"text\",\"type\":\"text\",\"lib\":\"common\",\"caption\":\"Gender : Male...\",\"text\":\"Gender : Male\",\"textFont\":\"Trebuchet MS,sans-serif\",\"textFontStyle\":\"400\",\"textProperties\":[],\"textSize\":\"32\",\"textColor\":{\"type\":\"solid\",\"color\":\"rgba(113,113,113,1)\"},\"backgroundColor\":\"none\",\"width\":186,\"height\":39,\"lineHeight\":39,\"textAlign\":\"left\",\"textSpacing\":\"-1.6\",\"enableShadow\":false,\"autoResize\":true,\"v2\":true,\"left\":379,\"top\":47,\"vpos\":\"top\",\"hpos\":\"left\",\"visible\":true,\"group\":\"root\",\"id\":\"box-64\",\"zindex\":8,\"wtype\":\"fixed\",\"htype\":\"fixed\",\"opacity\":1,\"propagateEvents\":false,\"lineHeightAuto\":true,\"rotation\":0,\"scale\":1,\"aspectratio\":false},{\"name\":\"text\",\"type\":\"text\",\"lib\":\"common\",\"caption\":\"DOB: 01/01/2022...\",\"text\":\"DOB: 01/01/2022\",\"textFont\":\"Trebuchet MS,sans-serif\",\"textFontStyle\":\"400\",\"textProperties\":[],\"textSize\":\"32\",\"textColor\":{\"type\":\"solid\",\"color\":\"rgba(113,113,113,1)\"},\"backgroundColor\":\"none\",\"width\":230,\"height\":39,\"lineHeight\":39,\"textAlign\":\"left\",\"textSpacing\":\"-1.6\",\"enableShadow\":false,\"autoResize\":true,\"v2\":true,\"left\":76,\"top\":130,\"vpos\":\"top\",\"hpos\":\"left\",\"visible\":true,\"group\":\"root\",\"id\":\"box-61\",\"zindex\":8,\"wtype\":\"fixed\",\"htype\":\"fixed\",\"opacity\":1,\"propagateEvents\":false,\"lineHeightAuto\":true,\"rotation\":0,\"scale\":1,\"aspectratio\":false},{\"name\":\"text\",\"type\":\"text\",\"lib\":\"common\",\"caption\":\"Height : 174 cm...\",\"text\":\"Height : 174 cm&nbsp;\",\"textFont\":\"Trebuchet MS,sans-serif\",\"textFontStyle\":\"400\",\"textProperties\":[],\"textSize\":\"32\",\"textColor\":{\"type\":\"solid\",\"color\":\"rgba(113,113,113,1)\"},\"backgroundColor\":\"none\",\"width\":216,\"height\":39,\"lineHeight\":39,\"textAlign\":\"left\",\"textSpacing\":\"-1.6\",\"enableShadow\":false,\"autoResize\":true,\"v2\":true,\"left\":379,\"top\":86,\"vpos\":\"top\",\"hpos\":\"left\",\"visible\":true,\"group\":\"root\",\"id\":\"box-62\",\"zindex\":8,\"wtype\":\"fixed\",\"htype\":\"fixed\",\"opacity\":1,\"propagateEvents\":false,\"lineHeightAuto\":true,\"rotation\":0,\"scale\":1,\"aspectratio\":false},{\"name\":\"text\",\"type\":\"text\",\"lib\":\"common\",\"caption\":\"Surgery:\\nTotal Colec...\",\"text\":\"Surgery:&nbsp;<br>Total Colectomy with&nbsp;<br>pancreatico-jejunostomy&nbsp;\",\"textFont\":\"Trebuchet MS,sans-serif\",\"textFontStyle\":\"400\",\"textProperties\":[],\"textSize\":\"32\",\"textColor\":{\"type\":\"solid\",\"color\":\"rgba(113,113,113,1)\"},\"backgroundColor\":\"none\",\"width\":336,\"height\":117,\"lineHeight\":39,\"textAlign\":\"left\",\"textSpacing\":\"-1.6\",\"enableShadow\":false,\"autoResize\":true,\"v2\":true,\"left\":1084,\"top\":47,\"vpos\":\"top\",\"hpos\":\"left\",\"visible\":true,\"group\":\"root\",\"id\":\"box-63\",\"zindex\":8,\"wtype\":\"fixed\",\"htype\":\"fixed\",\"opacity\":1,\"propagateEvents\":false,\"lineHeightAuto\":true,\"rotation\":0,\"scale\":1,\"aspectratio\":false},{\"name\":\"text\",\"type\":\"text\",\"lib\":\"common\",\"caption\":\"Age : 86 y\",\"text\":\"Age : 86 y&nbsp;\",\"textFont\":\"Trebuchet MS,sans-serif\",\"textFontStyle\":\"400\",\"textProperties\":[],\"textSize\":\"32\",\"textColor\":{\"type\":\"solid\",\"color\":\"rgba(113,113,113,1)\"},\"backgroundColor\":\"none\",\"width\":139,\"height\":39,\"lineHeight\":39,\"textAlign\":\"left\",\"textSpacing\":\"-1.6\",\"enableShadow\":false,\"autoResize\":true,\"v2\":true,\"left\":76,\"top\":91,\"vpos\":\"top\",\"hpos\":\"left\",\"visible\":true,\"group\":\"root\",\"id\":\"box-60\",\"zindex\":8,\"wtype\":\"fixed\",\"htype\":\"fixed\",\"opacity\":1,\"propagateEvents\":false,\"lineHeightAuto\":true,\"rotation\":0,\"scale\":1,\"aspectratio\":false},{\"name\":\"text\",\"type\":\"text\",\"lib\":\"common\",\"caption\":\"Allergies :\\nPenicill...\",\"text\":\"Allergies :<br><span style=\\\"color: rgba(249,51,36,1);\\\">Penicillin</span><br><br>\",\"textFont\":\"Trebuchet MS,sans-serif\",\"textFontStyle\":\"400\",\"textProperties\":[],\"textSize\":\"32\",\"textColor\":{\"type\":\"solid\",\"color\":\"rgba(113,113,113,1)\"},\"backgroundColor\":\"none\",\"width\":203,\"height\":117,\"lineHeight\":39,\"textAlign\":\"left\",\"textSpacing\":\"-1.6\",\"enableShadow\":false,\"autoResize\":false,\"v2\":true,\"left\":766,\"top\":47,\"vpos\":\"top\",\"hpos\":\"left\",\"visible\":true,\"group\":\"root\",\"id\":\"box-59\",\"zindex\":8,\"wtype\":\"fixed\",\"htype\":\"fixed\",\"opacity\":1,\"propagateEvents\":false,\"lineHeightAuto\":true,\"rotation\":0,\"scale\":1,\"aspectratio\":false},{\"name\":\"text\",\"type\":\"text\",\"lib\":\"common\",\"caption\":\"BMI : 28 kg/m2...\",\"text\":\"BMI : 28 kg/m2\",\"textFont\":\"Trebuchet MS,sans-serif\",\"textFontStyle\":\"400\",\"textProperties\":[],\"textSize\":\"32\",\"textColor\":{\"type\":\"solid\",\"color\":\"rgba(113,113,113,1)\"},\"backgroundColor\":\"none\",\"width\":199,\"height\":39,\"lineHeight\":39,\"textAlign\":\"left\",\"textSpacing\":\"-1.6\",\"enableShadow\":false,\"autoResize\":true,\"v2\":true,\"left\":379,\"top\":171,\"vpos\":\"top\",\"hpos\":\"left\",\"visible\":true,\"group\":\"root\",\"id\":\"box-56\",\"zindex\":8,\"wtype\":\"fixed\",\"htype\":\"fixed\",\"opacity\":1,\"propagateEvents\":false,\"lineHeightAuto\":true,\"rotation\":0,\"scale\":1,\"aspectratio\":false},{\"name\":\"text\",\"type\":\"text\",\"lib\":\"common\",\"caption\":\"Weight : 69 Kg...\",\"text\":\"Weight : 69 Kg\",\"textFont\":\"Trebuchet MS,sans-serif\",\"textFontStyle\":\"400\",\"textProperties\":[],\"textSize\":\"32\",\"textColor\":{\"type\":\"solid\",\"color\":\"rgba(113,113,113,1)\"},\"backgroundColor\":\"none\",\"width\":190,\"height\":39,\"lineHeight\":39,\"textAlign\":\"left\",\"textSpacing\":\"-1.6\",\"enableShadow\":false,\"autoResize\":true,\"v2\":true,\"left\":379,\"top\":130,\"vpos\":\"top\",\"hpos\":\"left\",\"visible\":true,\"group\":\"root\",\"id\":\"box-58\",\"zindex\":8,\"wtype\":\"fixed\",\"htype\":\"fixed\",\"opacity\":1,\"propagateEvents\":false,\"lineHeightAuto\":true,\"rotation\":0,\"scale\":1,\"aspectratio\":false},{\"name\":\"verticalline\",\"type\":\"verticalline\",\"lib\":\"common\",\"caption\":\"Vertical line 3\",\"width\":1,\"height\":229,\"color\":\"A6A6A6\",\"weight\":1,\"left\":341,\"top\":10,\"vpos\":\"top\",\"hpos\":\"left\",\"visible\":true,\"group\":\"root\",\"id\":\"box-52\",\"zindex\":7,\"wtype\":\"fixed\",\"htype\":\"fixed\",\"opacity\":1,\"borderStyle\":\"solid\",\"rotation\":0,\"scale\":1,\"aspectratio\":false},{\"name\":\"verticalline\",\"type\":\"verticalline\",\"lib\":\"common\",\"caption\":\"Vertical line 5\",\"width\":1,\"height\":229,\"color\":\"A6A6A6\",\"weight\":1,\"left\":1064,\"top\":15,\"vpos\":\"top\",\"hpos\":\"left\",\"visible\":true,\"group\":\"root\",\"id\":\"box-54\",\"zindex\":7,\"wtype\":\"fixed\",\"htype\":\"fixed\",\"opacity\":1,\"borderStyle\":\"solid\",\"rotation\":0,\"scale\":1,\"aspectratio\":false},{\"name\":\"verticalline\",\"type\":\"verticalline\",\"lib\":\"common\",\"caption\":\"Vertical line 6\",\"width\":1,\"height\":229,\"color\":\"A6A6A6\",\"weight\":1,\"left\":733,\"top\":10,\"vpos\":\"top\",\"hpos\":\"left\",\"visible\":true,\"group\":\"root\",\"id\":\"box-55\",\"zindex\":7,\"wtype\":\"fixed\",\"htype\":\"fixed\",\"opacity\":1,\"borderStyle\":\"solid\",\"rotation\":0,\"scale\":1,\"aspectratio\":false},{\"name\":\"datetime\",\"type\":\"datetime\",\"lib\":\"common\",\"caption\":\"Date/Time 1\",\"width\":376,\"height\":41,\"backgroundColor\":\"none\",\"borderWidth\":\"1\",\"borderColor\":{\"type\":\"solid\",\"color\":\"rgba(102,150,200,1)\"},\"textFont\":\"Montserrat\",\"textFontStyle\":\"400\",\"textSize\":\"20\",\"textColor\":\"383838\",\"textProperties\":[],\"textAlign\":\"center\",\"dateFormat\":\"yearmonthday\",\"dayFormat\":\"superscript\",\"monthFormat\":\"long\",\"yearFormat\":\"long\",\"timeFormat\":\"24\",\"dateGeneralFormat\":\"timedate\",\"dayNameFormat\":\"long\",\"dateSeparator\":\"-\",\"left\":1064,\"top\":201,\"vpos\":\"top\",\"hpos\":\"left\",\"visible\":true,\"group\":\"root\",\"id\":\"box-50\",\"zindex\":5,\"wtype\":\"fixed\",\"htype\":\"fixed\",\"opacity\":1,\"rotation\":0,\"scale\":1,\"aspectratio\":false},{\"name\":\"text\",\"type\":\"text\",\"lib\":\"common\",\"caption\":\"MRN : 12345679...\",\"text\":\"MRN : 12345679&nbsp;\",\"textFont\":\"Trebuchet MS,sans-serif\",\"textFontStyle\":\"400\",\"textProperties\":[],\"textSize\":\"32\",\"textColor\":{\"type\":\"solid\",\"color\":\"rgba(113,113,113,1)\"},\"backgroundColor\":\"none\",\"width\":218,\"height\":39,\"lineHeight\":39,\"textAlign\":\"left\",\"textSpacing\":\"-1.6\",\"enableShadow\":false,\"autoResize\":true,\"v2\":true,\"left\":76,\"top\":171,\"vpos\":\"top\",\"hpos\":\"left\",\"visible\":true,\"group\":\"root\",\"id\":\"box-48\",\"zindex\":4,\"wtype\":\"fixed\",\"htype\":\"fixed\",\"opacity\":1,\"propagateEvents\":false,\"lineHeightAuto\":true,\"rotation\":0,\"scale\":1,\"aspectratio\":false},{\"name\":\"verticalline\",\"type\":\"verticalline\",\"lib\":\"common\",\"caption\":\"Vertical line 1\",\"width\":1,\"height\":658,\"color\":{\"type\":\"solid\",\"color\":\"rgba(70,127,215,1)\"},\"weight\":1,\"left\":734,\"top\":242,\"vpos\":\"top\",\"hpos\":\"left\",\"visible\":true,\"group\":\"root\",\"id\":\"box-47\",\"zindex\":3,\"wtype\":\"fixed\",\"htype\":\"fixed\",\"opacity\":1,\"borderStyle\":\"solid\",\"rotation\":0,\"scale\":1,\"aspectratio\":false},{\"name\":\"text\",\"type\":\"text\",\"lib\":\"common\",\"caption\":\"John Doe\",\"text\":\"John Doe&nbsp;\",\"textFont\":\"Trebuchet MS,sans-serif\",\"textFontStyle\":\"400\",\"textProperties\":[],\"textSize\":\"64\",\"textColor\":{\"type\":\"solid\",\"color\":\"rgba(117,85,88,1)\"},\"backgroundColor\":\"none\",\"width\":272,\"height\":78,\"lineHeight\":78,\"textAlign\":\"left\",\"textSpacing\":\"-1.6\",\"enableShadow\":false,\"autoResize\":true,\"v2\":true,\"left\":40,\"top\":15,\"vpos\":\"top\",\"hpos\":\"left\",\"visible\":true,\"id\":\"box-46\",\"group\":\"root\",\"zindex\":2,\"wtype\":\"fixed\",\"htype\":\"fixed\",\"opacity\":1,\"propagateEvents\":false,\"lineHeightAuto\":true,\"rotation\":0,\"scale\":1,\"aspectratio\":false},{\"name\":\"basic_slider\",\"type\":\"basic_slider\",\"lib\":\"generic2\",\"caption\":\"Slider 1\",\"width\":347,\"height\":41,\"barThickness\":4,\"barColor\":\"e3e6e8\",\"fillBarColor\":{\"type\":\"solid\",\"color\":\"rgba(255,255,0,1)\"},\"sliderColor\":\"ffffff\",\"barBorderRadius\":2,\"sliderBorderWidth\":1,\"sliderBorderRadius\":\"50%\",\"borderColor\":\"d4d7d9\",\"sliderPosition\":\"70\",\"left\":379,\"top\":359,\"vpos\":\"top\",\"hpos\":\"left\",\"visible\":true,\"id\":\"box-66\",\"group\":\"root\",\"zindex\":9,\"wtype\":\"fixed\",\"htype\":\"fixed\",\"opacity\":1,\"rotation\":0,\"scale\":1,\"aspectratio\":false,\"blurB\":0,\"saturateB\":1,\"brightnessB\":1,\"contrastB\":1,\"grayscaleB\":0,\"sepiaB\":0,\"invertB\":0,\"hueRotateB\":0,\"actions\":[{\"title\":\"Interaction 1\",\"type\":\"sliderdrag\",\"actionId\":\"set-variable\",\"callback\":true,\"variableName\":\"propofol_amount\",\"valueSource\":\"callbackvalue\",\"callbackvariable\":\"value\",\"delay\":\"0\",\"guid\":\"115a9a30-6b72-488a-9aac-6c0f68f0647e\",\"callbackAction\":{\"actionId\":\"change-property\",\"callback\":true,\"pageId\":\"2\",\"itemId\":\"box-88\",\"propertyId\":\"text\",\"valueType\":\"readFromVariable\",\"variableName\":\"propofol_amount\",\"delay\":\"0\",\"guid\":\"c2a09f50-a737-4a42-9a21-cde40585b144\",\"callbackAction\":{\"actionId\":\"change-screen-state\",\"callback\":false,\"pageId\":\"2\",\"stateId\":\"d36a0c09-b058-464f-be6a-c8a9af1bd182\",\"delay\":\"0\",\"guid\":\"d0676d5b-2df5-42fd-8abd-6c2031d040d4\",\"clb_disabled\":false},\"clb_disabled\":false},\"sort\":0}]},{\"name\":\"basic_slider\",\"type\":\"basic_slider\",\"lib\":\"generic2\",\"caption\":\"Slider 2\",\"width\":347,\"height\":41,\"barThickness\":4,\"barColor\":\"e3e6e8\",\"fillBarColor\":{\"type\":\"solid\",\"color\":\"rgba(198,198,198,1)\"},\"sliderColor\":\"ffffff\",\"barBorderRadius\":2,\"sliderBorderWidth\":1,\"sliderBorderRadius\":\"50%\",\"borderColor\":\"d4d7d9\",\"sliderPosition\":\"44\",\"left\":373,\"top\":452,\"vpos\":\"top\",\"hpos\":\"left\",\"visible\":true,\"id\":\"box-71\",\"group\":\"root\",\"zindex\":9,\"wtype\":\"fixed\",\"htype\":\"fixed\",\"opacity\":1,\"rotation\":0,\"scale\":1,\"aspectratio\":false,\"actions\":[{\"title\":\"Set Val\",\"type\":\"sliderdrag\",\"actionId\":\"set-variable\",\"callback\":true,\"variableName\":\"lido\",\"valueSource\":\"callbackvalue\",\"callbackvariable\":\"value\",\"delay\":\"0\",\"guid\":\"9b245471-98ea-4eee-bfee-54afd95ae238\",\"callbackAction\":{\"actionId\":\"change-property\",\"callback\":true,\"pageId\":\"2\",\"itemId\":\"box-90\",\"propertyId\":\"text\",\"valueType\":\"readFromVariable\",\"variableName\":\"lido\",\"delay\":\"0\",\"guid\":\"60c3c50a-c227-4306-a203-d4050745a2bd\",\"callbackAction\":{\"actionId\":\"change-screen-state\",\"callback\":false,\"pageId\":\"2\",\"stateId\":\"d36a0c09-b058-464f-be6a-c8a9af1bd182\",\"delay\":\"0\",\"guid\":\"5fc4670e-6282-40f1-9ef1-90cc2ef2f621\",\"clb_disabled\":false},\"clb_disabled\":false},\"sort\":0}]},{\"name\":\"basic_slider\",\"type\":\"basic_slider\",\"lib\":\"generic2\",\"caption\":\"Slider 3\",\"width\":347,\"height\":41,\"barThickness\":4,\"barColor\":\"e3e6e8\",\"fillBarColor\":{\"type\":\"solid\",\"color\":\"rgba(249,51,36,1)\"},\"sliderColor\":\"ffffff\",\"barBorderRadius\":2,\"sliderBorderWidth\":1,\"sliderBorderRadius\":\"50%\",\"borderColor\":\"d4d7d9\",\"sliderPosition\":\"61\",\"left\":379,\"top\":735,\"vpos\":\"top\",\"hpos\":\"left\",\"visible\":true,\"id\":\"box-75\",\"group\":\"root\",\"zindex\":9,\"wtype\":\"fixed\",\"htype\":\"fixed\",\"opacity\":1,\"rotation\":0,\"scale\":1,\"aspectratio\":false,\"actions\":[{\"title\":\"r\",\"type\":\"sliderdrag\",\"actionId\":\"set-variable\",\"callback\":true,\"variableName\":\"rocu\",\"valueSource\":\"callbackvalue\",\"callbackvariable\":\"value\",\"delay\":\"0\",\"guid\":\"2b4e0cec-9c01-4ba9-b4dc-0afa28954a63\",\"callbackAction\":{\"actionId\":\"change-property\",\"callback\":true,\"pageId\":\"2\",\"itemId\":\"box-93\",\"propertyId\":\"text\",\"valueType\":\"readFromVariable\",\"variableName\":\"rocu\",\"delay\":\"0\",\"guid\":\"3e75b7c7-86bb-441a-896d-592470061468\",\"callbackAction\":{\"actionId\":\"change-screen-state\",\"callback\":false,\"pageId\":\"2\",\"stateId\":\"d36a0c09-b058-464f-be6a-c8a9af1bd182\",\"delay\":\"0\",\"guid\":\"e9686745-10a0-4404-811b-45fc272155f8\",\"clb_disabled\":false},\"clb_disabled\":false},\"sort\":0}]},{\"name\":\"basic_slider\",\"type\":\"basic_slider\",\"lib\":\"generic2\",\"caption\":\"Slider 4\",\"width\":347,\"height\":41,\"barThickness\":4,\"barColor\":\"e3e6e8\",\"fillBarColor\":{\"type\":\"solid\",\"color\":\"rgba(160,58,207,1)\"},\"sliderColor\":\"ffffff\",\"barBorderRadius\":2,\"sliderBorderWidth\":1,\"sliderBorderRadius\":\"50%\",\"borderColor\":\"d4d7d9\",\"sliderPosition\":\"40\",\"left\":379,\"top\":648,\"vpos\":\"top\",\"hpos\":\"left\",\"visible\":true,\"id\":\"box-76\",\"group\":\"root\",\"zindex\":9,\"wtype\":\"fixed\",\"htype\":\"fixed\",\"opacity\":1,\"rotation\":0,\"scale\":1,\"aspectratio\":false,\"actions\":[{\"title\":\"frt\",\"type\":\"sliderdrag\",\"actionId\":\"set-variable\",\"callback\":true,\"variableName\":\"phen\",\"valueSource\":\"callbackvalue\",\"callbackvariable\":\"value\",\"delay\":\"0\",\"guid\":\"722752a1-347b-4a35-954e-102ac67616a9\",\"callbackAction\":{\"actionId\":\"change-property\",\"callback\":true,\"pageId\":\"2\",\"itemId\":\"box-92\",\"propertyId\":\"text\",\"valueType\":\"readFromVariable\",\"variableName\":\"phen\",\"delay\":\"0\",\"guid\":\"77d08880-f288-4ea2-83c9-7e43a6afdf5e\",\"callbackAction\":{\"actionId\":\"change-screen-state\",\"callback\":false,\"pageId\":\"2\",\"stateId\":\"d36a0c09-b058-464f-be6a-c8a9af1bd182\",\"delay\":\"0\",\"guid\":\"81fc5eaf-47f9-45a4-8ca4-8f748104cfb9\",\"clb_disabled\":false},\"clb_disabled\":false},\"sort\":0}]},{\"name\":\"basic_slider\",\"type\":\"basic_slider\",\"lib\":\"generic2\",\"caption\":\"Slider 5\",\"width\":347,\"height\":41,\"barThickness\":4,\"barColor\":\"e3e6e8\",\"fillBarColor\":{\"type\":\"solid\",\"color\":\"rgba(102,150,200,1)\"},\"sliderColor\":\"ffffff\",\"barBorderRadius\":2,\"sliderBorderWidth\":1,\"sliderBorderRadius\":\"50%\",\"borderColor\":\"d4d7d9\",\"sliderPosition\":\"33\",\"left\":379,\"top\":550,\"vpos\":\"top\",\"hpos\":\"left\",\"visible\":true,\"id\":\"box-77\",\"group\":\"root\",\"zindex\":9,\"wtype\":\"fixed\",\"htype\":\"fixed\",\"opacity\":1,\"rotation\":0,\"scale\":1,\"aspectratio\":false,\"actions\":[{\"title\":\"f\",\"type\":\"sliderdrag\",\"actionId\":\"set-variable\",\"callback\":true,\"variableName\":\"fent\",\"valueSource\":\"callbackvalue\",\"callbackvariable\":\"value\",\"delay\":\"0\",\"guid\":\"01536077-54d1-418a-a1b5-720ab0f5ab67\",\"callbackAction\":{\"actionId\":\"change-property\",\"callback\":true,\"pageId\":\"2\",\"itemId\":\"box-91\",\"propertyId\":\"text\",\"valueType\":\"readFromVariable\",\"variableName\":\"fent\",\"delay\":\"0\",\"guid\":\"41599e71-e3b8-448b-bc1a-608621aa5d23\",\"callbackAction\":{\"actionId\":\"change-screen-state\",\"callback\":false,\"pageId\":\"2\",\"stateId\":\"d36a0c09-b058-464f-be6a-c8a9af1bd182\",\"delay\":\"0\",\"guid\":\"bf3b8f44-6643-4910-affd-fd010dc5aae8\",\"clb_disabled\":false},\"clb_disabled\":false},\"sort\":0}]},{\"name\":\"basic_textfield\",\"type\":\"basic_textfield\",\"lib\":\"generic2\",\"caption\":\"Text Field prop\",\"width\":338,\"height\":55,\"value\":\"Propofol  ( mg ) \",\"placeholder\":\"Placeholder\",\"inputtype\":\"text\",\"backgroundColor\":{\"type\":\"solid\",\"color\":\"rgba(255,255,122,1)\"},\"textFont\":\"Arial,sans-serif\",\"textFontStyle\":\"400\",\"textSize\":\"28\",\"textColor\":{\"type\":\"solid\",\"color\":\"rgba(56,56,56,1)\"},\"textProperties\":[],\"placeholderColor\":\"c1c8c9\",\"borderWidth\":1,\"borderStyle\":\"solid\",\"borderRadius\":4,\"borderColor\":{\"type\":\"solid\",\"color\":\"rgba(255,255,0,1)\"},\"textAlign\":\"center\",\"left\":22,\"top\":359,\"vpos\":\"top\",\"hpos\":\"left\",\"visible\":true,\"group\":\"root\",\"id\":\"box-67\",\"zindex\":10,\"wtype\":\"fixed\",\"htype\":\"fixed\",\"opacity\":1,\"rotation\":0,\"scale\":1,\"aspectratio\":false,\"userCaption\":true},{\"name\":\"basic_textfield\",\"type\":\"basic_textfield\",\"lib\":\"generic2\",\"caption\":\"Text Field lido\",\"width\":338,\"height\":55,\"value\":\"Lidocaine  (mg)\",\"placeholder\":\"Placeholder\",\"inputtype\":\"text\",\"backgroundColor\":{\"type\":\"solid\",\"color\":\"rgba(198,198,198,1)\"},\"textFont\":\"Arial,sans-serif\",\"textFontStyle\":\"400\",\"textSize\":\"28\",\"textColor\":{\"type\":\"solid\",\"color\":\"rgba(56,56,56,1)\"},\"textProperties\":[],\"placeholderColor\":\"c1c8c9\",\"borderWidth\":1,\"borderStyle\":\"solid\",\"borderRadius\":4,\"borderColor\":{\"type\":\"solid\",\"color\":\"rgba(208,212,0,1)\"},\"textAlign\":\"center\",\"left\":22,\"top\":445,\"vpos\":\"top\",\"hpos\":\"left\",\"visible\":true,\"group\":\"root\",\"id\":\"box-72\",\"zindex\":10,\"wtype\":\"fixed\",\"htype\":\"fixed\",\"opacity\":1,\"rotation\":0,\"scale\":1,\"aspectratio\":false,\"userCaption\":true},{\"name\":\"basic_textfield\",\"type\":\"basic_textfield\",\"lib\":\"generic2\",\"caption\":\"Text Field Phenylephrine\",\"width\":338,\"height\":55,\"value\":\"Phenylephrine (mcg) \",\"placeholder\":\"Placeholder\",\"inputtype\":\"text\",\"backgroundColor\":{\"type\":\"solid\",\"color\":\"rgba(234,114,252,1)\"},\"textFont\":\"Arial,sans-serif\",\"textFontStyle\":\"400\",\"textSize\":\"28\",\"textColor\":{\"type\":\"solid\",\"color\":\"rgba(56,56,56,1)\"},\"textProperties\":[],\"placeholderColor\":\"c1c8c9\",\"borderWidth\":1,\"borderStyle\":\"solid\",\"borderRadius\":4,\"borderColor\":{\"type\":\"solid\",\"color\":\"rgba(255,255,0,1)\"},\"textAlign\":\"center\",\"left\":22,\"top\":641,\"vpos\":\"top\",\"hpos\":\"left\",\"visible\":true,\"group\":\"root\",\"id\":\"box-73\",\"zindex\":10,\"wtype\":\"fixed\",\"htype\":\"fixed\",\"opacity\":1,\"rotation\":0,\"scale\":1,\"aspectratio\":false,\"userCaption\":true},{\"name\":\"basic_textfield\",\"type\":\"basic_textfield\",\"lib\":\"generic2\",\"caption\":\"Text Field rocur\",\"width\":338,\"height\":55,\"value\":\"Rocuronium (mg)\",\"placeholder\":\"Placeholder\",\"inputtype\":\"text\",\"backgroundColor\":{\"type\":\"solid\",\"color\":\"rgba(226,15,0,1)\"},\"textFont\":\"Arial,sans-serif\",\"textFontStyle\":\"400\",\"textSize\":\"28\",\"textColor\":{\"type\":\"solid\",\"color\":\"rgba(56,56,56,1)\"},\"textProperties\":[],\"placeholderColor\":\"c1c8c9\",\"borderWidth\":1,\"borderStyle\":\"solid\",\"borderRadius\":4,\"borderColor\":{\"type\":\"solid\",\"color\":\"rgba(255,255,0,1)\"},\"textAlign\":\"center\",\"left\":22,\"top\":721,\"vpos\":\"top\",\"hpos\":\"left\",\"visible\":true,\"group\":\"root\",\"id\":\"box-79\",\"zindex\":10,\"wtype\":\"fixed\",\"htype\":\"fixed\",\"opacity\":1,\"rotation\":0,\"scale\":1,\"aspectratio\":false,\"userCaption\":true},{\"name\":\"basic_textfield\",\"type\":\"basic_textfield\",\"lib\":\"generic2\",\"caption\":\"Text Field 2\",\"width\":733,\"height\":77,\"value\":\"Induction Medication\",\"placeholder\":\"Placeholder\",\"inputtype\":\"text\",\"backgroundColor\":{\"type\":\"solid\",\"color\":\"rgba(177,190,227,1)\"},\"textFont\":\"Arial,sans-serif\",\"textFontStyle\":\"400\",\"textSize\":\"64\",\"textColor\":{\"type\":\"solid\",\"color\":\"rgba(233,240,215,1)\"},\"textProperties\":[],\"placeholderColor\":\"c1c8c9\",\"borderWidth\":\"5\",\"borderStyle\":\"solid\",\"borderRadius\":4,\"borderColor\":{\"type\":\"solid\",\"color\":\"rgba(85,85,85,1)\"},\"textAlign\":\"center\",\"left\":2,\"top\":242,\"vpos\":\"top\",\"hpos\":\"left\",\"visible\":true,\"id\":\"box-69\",\"group\":\"root\",\"zindex\":11,\"wtype\":\"fixed\",\"htype\":\"fixed\",\"opacity\":1,\"rotation\":0,\"scale\":1,\"aspectratio\":false},{\"name\":\"basic_textfield\",\"type\":\"basic_textfield\",\"lib\":\"generic2\",\"caption\":\"Text Field 3\",\"width\":698,\"height\":77,\"value\":\"Blood Pressure\",\"placeholder\":\"Placeholder\",\"inputtype\":\"text\",\"backgroundColor\":{\"type\":\"solid\",\"color\":\"rgba(177,190,227,1)\"},\"textFont\":\"Arial,sans-serif\",\"textFontStyle\":\"400\",\"textSize\":\"64\",\"textColor\":{\"type\":\"solid\",\"color\":\"rgba(233,240,215,1)\"},\"textProperties\":[],\"placeholderColor\":\"c1c8c9\",\"borderWidth\":\"5\",\"borderStyle\":\"solid\",\"borderRadius\":4,\"borderColor\":{\"type\":\"solid\",\"color\":\"rgba(85,85,85,1)\"},\"textAlign\":\"center\",\"left\":733,\"top\":244,\"vpos\":\"top\",\"hpos\":\"left\",\"visible\":true,\"id\":\"box-70\",\"group\":\"root\",\"zindex\":11,\"wtype\":\"fixed\",\"htype\":\"fixed\",\"opacity\":1,\"rotation\":0,\"scale\":1,\"aspectratio\":false},{\"name\":\"basic_textfield\",\"type\":\"basic_textfield\",\"lib\":\"generic2\",\"caption\":\"Text Field fenta\",\"width\":338,\"height\":55,\"value\":\"Fentanyl (mcg)\",\"placeholder\":\"Placeholder\",\"inputtype\":\"text\",\"backgroundColor\":{\"type\":\"solid\",\"color\":\"rgba(101,180,206,1)\"},\"textFont\":\"Arial,sans-serif\",\"textFontStyle\":\"400\",\"textSize\":\"28\",\"textColor\":{\"type\":\"solid\",\"color\":\"rgba(56,56,56,1)\"},\"textProperties\":[],\"placeholderColor\":\"c1c8c9\",\"borderWidth\":1,\"borderStyle\":\"solid\",\"borderRadius\":4,\"borderColor\":{\"type\":\"solid\",\"color\":\"rgba(27,77,87,1)\"},\"textAlign\":\"center\",\"left\":22,\"top\":543,\"vpos\":\"top\",\"hpos\":\"left\",\"visible\":true,\"group\":\"root\",\"id\":\"box-78\",\"zindex\":10,\"wtype\":\"fixed\",\"htype\":\"fixed\",\"opacity\":1,\"rotation\":0,\"scale\":1,\"aspectratio\":false,\"userCaption\":true},{\"name\":\"image\",\"type\":\"image\",\"lib\":\"common\",\"caption\":\"ss4\",\"imgSrc\":{\"fileId\":\"68d047acd614373a75ca2548bba92b44.png\",\"assetType\":\"project\",\"bucketsource\":\"main\",\"name\":\"Screen Shot 2022-12-12 at 7.38.12 PM.png\"},\"repeat\":false,\"width\":658,\"height\":399,\"borderWidth\":0,\"borderRadius\":0,\"borderColor\":\"#D1D1D1\",\"borderPos\":\"inside\",\"propagateEvents\":false,\"actions\":[],\"overlay\":false,\"autoResize\":false,\"aspectratio\":1.6491228070175439,\"assistiveDescription\":\"\",\"left\":762,\"top\":359,\"visible\":true,\"id\":\"box-80\",\"group\":\"root\",\"zindex\":12,\"hpos\":\"left\",\"vpos\":\"top\",\"wtype\":\"fixed\",\"htype\":\"fixed\",\"opacity\":1,\"borderStyle\":\"solid\",\"rotation\":0,\"scale\":1,\"hasMask\":false,\"userCaption\":true},{\"name\":\"basic_button_text\",\"type\":\"basic_button_text\",\"lib\":\"generic2\",\"caption\":\"Button with Background 1\",\"width\":177,\"height\":80,\"backgroundColor\":{\"type\":\"solid\",\"color\":\"rgba(102,150,200,1)\"},\"borderWidth\":0,\"borderColor\":\"#025abf\",\"borderRadius\":4,\"text\":\"Other Opioids\",\"textFont\":\"Arial,sans-serif\",\"textFontStyle\":\"400\",\"textSize\":\"28\",\"textColor\":\"FFFFFF\",\"textProperties\":[],\"textAlign\":\"center\",\"iconpos\":\"none\",\"left\":32,\"top\":800,\"vpos\":\"top\",\"hpos\":\"left\",\"visible\":true,\"id\":\"box-81\",\"group\":\"root\",\"zindex\":13,\"wtype\":\"fixed\",\"htype\":\"fixed\",\"opacity\":1,\"rotation\":0,\"scale\":1,\"aspectratio\":false,\"actions\":[{\"title\":\"Interaction 1\",\"type\":\"tap\",\"actionId\":\"go-to-page\",\"callback\":false,\"pageId\":\"4\",\"animation\":\"fade\",\"animationEasing\":\"Cubic.easeOut\",\"duration\":\"400\",\"delay\":\"0\",\"guid\":\"70e06ba4-aaab-4eb6-a4d5-6ce84bf7e1a8\",\"sort\":0}]},{\"name\":\"basic_button_text\",\"type\":\"basic_button_text\",\"lib\":\"generic2\",\"caption\":\"Button with Background 3\",\"width\":177,\"height\":80,\"backgroundColor\":{\"type\":\"solid\",\"color\":\"rgba(255, 255, 0,1)\"},\"borderWidth\":0,\"borderColor\":\"#025abf\",\"borderRadius\":4,\"text\":\"Other Amestics\",\"textFont\":\"Arial,sans-serif\",\"textFontStyle\":\"400\",\"textSize\":\"28\",\"textColor\":{\"type\":\"solid\",\"color\":\"rgba(56,56,56,1)\"},\"textProperties\":[],\"textAlign\":\"center\",\"iconpos\":\"none\",\"left\":252,\"top\":800,\"vpos\":\"top\",\"hpos\":\"left\",\"visible\":true,\"id\":\"box-83\",\"group\":\"root\",\"zindex\":13,\"wtype\":\"fixed\",\"htype\":\"fixed\",\"opacity\":1,\"rotation\":0,\"scale\":1,\"aspectratio\":false,\"actions\":[{\"title\":\"Interaction 1\",\"type\":\"tap\",\"actionId\":\"go-to-page\",\"callback\":false,\"pageId\":\"7\",\"animation\":\"fade\",\"animationEasing\":\"Cubic.easeOut\",\"duration\":\"400\",\"delay\":\"0\",\"guid\":\"fc07bf54-a966-4bda-8519-360dafb4096d\",\"sort\":0}]},{\"name\":\"basic_button_text\",\"type\":\"basic_button_text\",\"lib\":\"generic2\",\"caption\":\"Button with Background 4\",\"width\":177,\"height\":80,\"backgroundColor\":{\"type\":\"solid\",\"color\":\"rgba(234,114,252,1)\"},\"borderWidth\":0,\"borderColor\":\"#025abf\",\"borderRadius\":4,\"text\":\"Other Pressors\",\"textFont\":\"Arial,sans-serif\",\"textFontStyle\":\"400\",\"textSize\":\"28\",\"textColor\":\"FFFFFF\",\"textProperties\":[],\"textAlign\":\"center\",\"iconpos\":\"none\",\"left\":489,\"top\":800,\"vpos\":\"top\",\"hpos\":\"left\",\"visible\":true,\"id\":\"box-84\",\"group\":\"root\",\"zindex\":13,\"wtype\":\"fixed\",\"htype\":\"fixed\",\"opacity\":1,\"rotation\":0,\"scale\":1,\"aspectratio\":false,\"actions\":[{\"title\":\"Interaction 1\",\"type\":\"tap\",\"actionId\":\"go-to-page\",\"callback\":false,\"pageId\":\"6\",\"animation\":\"fade\",\"animationEasing\":\"Cubic.easeOut\",\"duration\":\"400\",\"delay\":\"0\",\"guid\":\"c4dfad33-4930-4c21-9f4f-50dc187fb243\",\"sort\":0}]},{\"name\":\"basic_button_text\",\"type\":\"basic_button_text\",\"lib\":\"generic2\",\"caption\":\"Button with Background 2\",\"width\":177,\"height\":80,\"backgroundColor\":{\"type\":\"solid\",\"color\":\"rgba(249,51,36,1)\"},\"borderWidth\":0,\"borderColor\":\"#025abf\",\"borderRadius\":4,\"text\":\"Exit\",\"textFont\":\"Arial,sans-serif\",\"textFontStyle\":\"400\",\"textSize\":\"36\",\"textColor\":\"FFFFFF\",\"textProperties\":[],\"textAlign\":\"center\",\"iconpos\":\"none\",\"left\":1233,\"top\":789,\"vpos\":\"top\",\"hpos\":\"left\",\"visible\":true,\"id\":\"box-82\",\"group\":\"root\",\"zindex\":13,\"wtype\":\"fixed\",\"htype\":\"fixed\",\"opacity\":1,\"rotation\":0,\"scale\":1,\"aspectratio\":false,\"actions\":[{\"title\":\"Interaction 1\",\"type\":\"click\",\"actionId\":\"go-to-page\",\"callback\":false,\"pageId\":\"1\",\"animation\":\"fade\",\"animationEasing\":\"Cubic.easeOut\",\"duration\":\"400\",\"delay\":\"0\",\"guid\":\"8e6d1162-0adc-439f-b39c-aef4571472d2\",\"sort\":0}]},{\"name\":\"text\",\"type\":\"text\",\"lib\":\"common\",\"caption\":\"text\",\"text\":\"150\",\"textFont\":\"Arial,sans-serif\",\"textFontStyle\":\"400\",\"textProperties\":[],\"textSize\":\"24\",\"textColor\":{\"type\":\"solid\",\"color\":\"rgba(255,255,0,1)\"},\"backgroundColor\":\"none\",\"width\":275,\"height\":29,\"lineHeight\":29,\"textAlign\":\"left\",\"textSpacing\":0,\"enableShadow\":false,\"autoResize\":false,\"v2\":true,\"left\":505,\"top\":330,\"vpos\":\"top\",\"hpos\":\"left\",\"visible\":true,\"id\":\"box-88\",\"group\":\"root\",\"zindex\":14,\"wtype\":\"fixed\",\"htype\":\"fixed\",\"opacity\":1,\"propagateEvents\":false,\"lineHeightAuto\":true,\"rotation\":0,\"scale\":1,\"aspectratio\":false,\"userCaption\":true,\"actions\":[]},{\"name\":\"text\",\"type\":\"text\",\"lib\":\"common\",\"caption\":\"text 2\",\"text\":\"100\",\"textFont\":\"Arial,sans-serif\",\"textFontStyle\":\"400\",\"textProperties\":[],\"textSize\":\"24\",\"textColor\":{\"type\":\"solid\",\"color\":\"rgba(198,198,198,1)\"},\"backgroundColor\":\"none\",\"width\":275,\"height\":29,\"lineHeight\":29,\"textAlign\":\"left\",\"textSpacing\":0,\"enableShadow\":false,\"autoResize\":false,\"v2\":true,\"left\":505,\"top\":414,\"vpos\":\"top\",\"hpos\":\"left\",\"visible\":true,\"id\":\"box-90\",\"group\":\"root\",\"zindex\":14,\"wtype\":\"fixed\",\"htype\":\"fixed\",\"opacity\":1,\"propagateEvents\":false,\"lineHeightAuto\":true,\"rotation\":0,\"scale\":1,\"aspectratio\":false,\"userCaption\":true,\"actions\":[]},{\"name\":\"text\",\"type\":\"text\",\"lib\":\"common\",\"caption\":\"text 3\",\"text\":\"100\",\"textFont\":\"Arial,sans-serif\",\"textFontStyle\":\"400\",\"textProperties\":[],\"textSize\":\"24\",\"textColor\":{\"type\":\"solid\",\"color\":\"rgba(43,89,162,1)\"},\"backgroundColor\":\"none\",\"width\":275,\"height\":29,\"lineHeight\":29,\"textAlign\":\"left\",\"textSpacing\":0,\"enableShadow\":false,\"autoResize\":false,\"v2\":true,\"left\":505,\"top\":521,\"vpos\":\"top\",\"hpos\":\"left\",\"visible\":true,\"id\":\"box-91\",\"group\":\"root\",\"zindex\":14,\"wtype\":\"fixed\",\"htype\":\"fixed\",\"opacity\":1,\"propagateEvents\":false,\"lineHeightAuto\":true,\"rotation\":0,\"scale\":1,\"aspectratio\":false,\"userCaption\":true,\"actions\":[]},{\"name\":\"text\",\"type\":\"text\",\"lib\":\"common\",\"caption\":\"text 4\",\"text\":\"100\",\"textFont\":\"Arial,sans-serif\",\"textFontStyle\":\"400\",\"textProperties\":[],\"textSize\":\"24\",\"textColor\":{\"type\":\"solid\",\"color\":\"rgba(183,47,214,1)\"},\"backgroundColor\":\"none\",\"width\":275,\"height\":29,\"lineHeight\":29,\"textAlign\":\"left\",\"textSpacing\":0,\"enableShadow\":false,\"autoResize\":false,\"v2\":true,\"left\":497,\"top\":612,\"vpos\":\"top\",\"hpos\":\"left\",\"visible\":true,\"id\":\"box-92\",\"group\":\"root\",\"zindex\":14,\"wtype\":\"fixed\",\"htype\":\"fixed\",\"opacity\":1,\"propagateEvents\":false,\"lineHeightAuto\":true,\"rotation\":0,\"scale\":1,\"aspectratio\":false,\"userCaption\":true,\"actions\":[]},{\"name\":\"text\",\"type\":\"text\",\"lib\":\"common\",\"caption\":\"text 5\",\"text\":\"100\",\"textFont\":\"Arial,sans-serif\",\"textFontStyle\":\"400\",\"textProperties\":[],\"textSize\":\"24\",\"textColor\":{\"type\":\"solid\",\"color\":\"rgba(209,25,105,1)\"},\"backgroundColor\":\"none\",\"width\":275,\"height\":29,\"lineHeight\":29,\"textAlign\":\"left\",\"textSpacing\":0,\"enableShadow\":false,\"autoResize\":false,\"v2\":true,\"left\":491,\"top\":712,\"vpos\":\"top\",\"hpos\":\"left\",\"visible\":true,\"id\":\"box-93\",\"group\":\"root\",\"zindex\":14,\"wtype\":\"fixed\",\"htype\":\"fixed\",\"opacity\":1,\"propagateEvents\":false,\"lineHeightAuto\":true,\"rotation\":0,\"scale\":1,\"aspectratio\":false,\"userCaption\":true,\"actions\":[]},{\"name\":\"image\",\"type\":\"image\",\"lib\":\"common\",\"caption\":\"ss3\",\"imgSrc\":{\"fileId\":\"7902aa96fae82cf51263d633901fa229.png\",\"assetType\":\"project\",\"bucketsource\":\"main\",\"name\":\"Screen Shot 2022-12-12 at 9.52.17 PM.png\"},\"repeat\":false,\"width\":654,\"height\":395,\"borderWidth\":0,\"borderRadius\":0,\"borderColor\":\"#D1D1D1\",\"borderPos\":\"inside\",\"propagateEvents\":false,\"actions\":[],\"overlay\":false,\"autoResize\":false,\"aspectratio\":1.65625,\"assistiveDescription\":\"\",\"left\":764,\"top\":366,\"visible\":true,\"id\":\"box-216\",\"group\":\"root\",\"zindex\":15,\"hpos\":\"left\",\"vpos\":\"top\",\"wtype\":\"fixed\",\"htype\":\"fixed\",\"opacity\":1,\"borderStyle\":\"solid\",\"rotation\":0,\"scale\":1,\"userCaption\":true,\"hasMask\":false},{\"name\":\"image\",\"type\":\"image\",\"lib\":\"common\",\"caption\":\"ss2\",\"imgSrc\":{\"fileId\":\"08d3b5585c3baf0fd8ac52e8c44a8630.png\",\"assetType\":\"project\",\"bucketsource\":\"main\",\"name\":\"Screen Shot 2022-12-12 at 9.52.29 PM.png\"},\"repeat\":false,\"width\":670,\"height\":409,\"borderWidth\":0,\"borderRadius\":0,\"borderColor\":\"#D1D1D1\",\"borderPos\":\"inside\",\"propagateEvents\":false,\"actions\":[],\"overlay\":false,\"autoResize\":false,\"aspectratio\":1.6379310344827587,\"assistiveDescription\":\"\",\"left\":749,\"top\":359,\"visible\":true,\"group\":\"root\",\"id\":\"box-217\",\"zindex\":16,\"hpos\":\"left\",\"vpos\":\"top\",\"wtype\":\"fixed\",\"htype\":\"fixed\",\"opacity\":1,\"borderStyle\":\"solid\",\"rotation\":0,\"scale\":1,\"userCaption\":true,\"hasMask\":false},{\"name\":\"image\",\"type\":\"image\",\"lib\":\"common\",\"caption\":\"ss1\",\"imgSrc\":{\"fileId\":\"68d047acd614373a75ca2548bba92b44.png\",\"assetType\":\"project\",\"bucketsource\":\"main\",\"name\":\"Screen Shot 2022-12-12 at 7.38.12 PM.png\"},\"repeat\":false,\"width\":664,\"height\":402,\"borderWidth\":0,\"borderRadius\":0,\"borderColor\":\"#D1D1D1\",\"borderPos\":\"inside\",\"propagateEvents\":false,\"actions\":[],\"overlay\":false,\"autoResize\":false,\"aspectratio\":1.6506849315068493,\"assistiveDescription\":\"\",\"left\":750,\"top\":357,\"visible\":true,\"id\":\"box-218\",\"group\":\"root\",\"zindex\":17,\"hpos\":\"left\",\"vpos\":\"top\",\"wtype\":\"fixed\",\"htype\":\"fixed\",\"opacity\":1,\"borderStyle\":\"solid\",\"rotation\":0,\"scale\":1,\"userCaption\":true,\"hasMask\":false}]","template":{"id":"","stateid":""},"history":[],"id":"73452cc4-51f4-48ca-9d25-b763541915d3","actions":[],"groups":{}},{"title":"State 2","background":{"type":"solid","color":"rgba(169,151,196,1)"},"data":"[{\"name\":\"rectangle\",\"type\":\"rectangle\",\"lib\":\"common\",\"caption\":\"Rectangle 1\",\"backgroundColor\":{\"type\":\"solid\",\"color\":\"rgba(193,212,241,1)\"},\"borderWidth\":\"8\",\"borderColor\":{\"type\":\"solid\",\"color\":\"rgba(30,77,130,1)\"},\"borderRadius\":0,\"imgSrc\":{\"fileId\":\"\",\"name\":\"\",\"assetType\":\"\",\"bucketsource\":\"\",\"url\":\"\"},\"backgroundSizeType\":\"cover\",\"backgroundSizeValue\":35,\"width\":1440,\"height\":249,\"lineHeight\":20,\"lineHeightAuto\":true,\"actions\":[],\"textFont\":\"Arial,sans-serif\",\"textFontStyle\":\"400\",\"textSize\":16,\"textColor\":\"#383838\",\"textProperties\":[],\"textSpacing\":0,\"textAlign\":\"center\",\"text\":\"\",\"typeName\":\"rectangle\",\"left\":0,\"top\":0,\"vpos\":\"top\",\"hpos\":\"left\",\"visible\":true,\"id\":\"box-45\",\"group\":\"root\",\"zindex\":1,\"wtype\":\"fixed\",\"htype\":\"fixed\",\"opacity\":1,\"borderStyle\":\"solid\",\"rotation\":0,\"scale\":1,\"aspectratio\":false},{\"name\":\"text\",\"type\":\"text\",\"lib\":\"common\",\"caption\":\"Gender : Male...\",\"text\":\"Gender : Male\",\"textFont\":\"Trebuchet MS,sans-serif\",\"textFontStyle\":\"400\",\"textProperties\":[],\"textSize\":\"32\",\"textColor\":{\"type\":\"solid\",\"color\":\"rgba(113,113,113,1)\"},\"backgroundColor\":\"none\",\"width\":186,\"height\":39,\"lineHeight\":39,\"textAlign\":\"left\",\"textSpacing\":\"-1.6\",\"enableShadow\":false,\"autoResize\":true,\"v2\":true,\"left\":379,\"top\":47,\"vpos\":\"top\",\"hpos\":\"left\",\"visible\":true,\"group\":\"root\",\"id\":\"box-64\",\"zindex\":8,\"wtype\":\"fixed\",\"htype\":\"fixed\",\"opacity\":1,\"propagateEvents\":false,\"lineHeightAuto\":true,\"rotation\":0,\"scale\":1,\"aspectratio\":false},{\"name\":\"text\",\"type\":\"text\",\"lib\":\"common\",\"caption\":\"DOB: 01/01/2022...\",\"text\":\"DOB: 01/01/2022\",\"textFont\":\"Trebuchet MS,sans-serif\",\"textFontStyle\":\"400\",\"textProperties\":[],\"textSize\":\"32\",\"textColor\":{\"type\":\"solid\",\"color\":\"rgba(113,113,113,1)\"},\"backgroundColor\":\"none\",\"width\":230,\"height\":39,\"lineHeight\":39,\"textAlign\":\"left\",\"textSpacing\":\"-1.6\",\"enableShadow\":false,\"autoResize\":true,\"v2\":true,\"left\":76,\"top\":130,\"vpos\":\"top\",\"hpos\":\"left\",\"visible\":true,\"group\":\"root\",\"id\":\"box-61\",\"zindex\":8,\"wtype\":\"fixed\",\"htype\":\"fixed\",\"opacity\":1,\"propagateEvents\":false,\"lineHeightAuto\":true,\"rotation\":0,\"scale\":1,\"aspectratio\":false},{\"name\":\"text\",\"type\":\"text\",\"lib\":\"common\",\"caption\":\"Height : 174 cm...\",\"text\":\"Height : 174 cm&nbsp;\",\"textFont\":\"Trebuchet MS,sans-serif\",\"textFontStyle\":\"400\",\"textProperties\":[],\"textSize\":\"32\",\"textColor\":{\"type\":\"solid\",\"color\":\"rgba(113,113,113,1)\"},\"backgroundColor\":\"none\",\"width\":216,\"height\":39,\"lineHeight\":39,\"textAlign\":\"left\",\"textSpacing\":\"-1.6\",\"enableShadow\":false,\"autoResize\":true,\"v2\":true,\"left\":379,\"top\":86,\"vpos\":\"top\",\"hpos\":\"left\",\"visible\":true,\"group\":\"root\",\"id\":\"box-62\",\"zindex\":8,\"wtype\":\"fixed\",\"htype\":\"fixed\",\"opacity\":1,\"propagateEvents\":false,\"lineHeightAuto\":true,\"rotation\":0,\"scale\":1,\"aspectratio\":false},{\"name\":\"text\",\"type\":\"text\",\"lib\":\"common\",\"caption\":\"Surgery:\\nTotal Colec...\",\"text\":\"Surgery:&nbsp;<br>Total Colectomy with&nbsp;<br>pancreatico-jejunostomy&nbsp;\",\"textFont\":\"Trebuchet MS,sans-serif\",\"textFontStyle\":\"400\",\"textProperties\":[],\"textSize\":\"32\",\"textColor\":{\"type\":\"solid\",\"color\":\"rgba(113,113,113,1)\"},\"backgroundColor\":\"none\",\"width\":336,\"height\":117,\"lineHeight\":39,\"textAlign\":\"left\",\"textSpacing\":\"-1.6\",\"enableShadow\":false,\"autoResize\":true,\"v2\":true,\"left\":1084,\"top\":47,\"vpos\":\"top\",\"hpos\":\"left\",\"visible\":true,\"group\":\"root\",\"id\":\"box-63\",\"zindex\":8,\"wtype\":\"fixed\",\"htype\":\"fixed\",\"opacity\":1,\"propagateEvents\":false,\"lineHeightAuto\":true,\"rotation\":0,\"scale\":1,\"aspectratio\":false},{\"name\":\"text\",\"type\":\"text\",\"lib\":\"common\",\"caption\":\"Age : 86 y\",\"text\":\"Age : 86 y&nbsp;\",\"textFont\":\"Trebuchet MS,sans-serif\",\"textFontStyle\":\"400\",\"textProperties\":[],\"textSize\":\"32\",\"textColor\":{\"type\":\"solid\",\"color\":\"rgba(113,113,113,1)\"},\"backgroundColor\":\"none\",\"width\":139,\"height\":39,\"lineHeight\":39,\"textAlign\":\"left\",\"textSpacing\":\"-1.6\",\"enableShadow\":false,\"autoResize\":true,\"v2\":true,\"left\":76,\"top\":91,\"vpos\":\"top\",\"hpos\":\"left\",\"visible\":true,\"group\":\"root\",\"id\":\"box-60\",\"zindex\":8,\"wtype\":\"fixed\",\"htype\":\"fixed\",\"opacity\":1,\"propagateEvents\":false,\"lineHeightAuto\":true,\"rotation\":0,\"scale\":1,\"aspectratio\":false},{\"name\":\"text\",\"type\":\"text\",\"lib\":\"common\",\"caption\":\"Allergies :\\nPenicill...\",\"text\":\"Allergies :<br><span style=\\\"color: rgba(249,51,36,1);\\\">Penicillin</span><br><br>\",\"textFont\":\"Trebuchet MS,sans-serif\",\"textFontStyle\":\"400\",\"textProperties\":[],\"textSize\":\"32\",\"textColor\":{\"type\":\"solid\",\"color\":\"rgba(113,113,113,1)\"},\"backgroundColor\":\"none\",\"width\":203,\"height\":117,\"lineHeight\":39,\"textAlign\":\"left\",\"textSpacing\":\"-1.6\",\"enableShadow\":false,\"autoResize\":false,\"v2\":true,\"left\":766,\"top\":47,\"vpos\":\"top\",\"hpos\":\"left\",\"visible\":true,\"group\":\"root\",\"id\":\"box-59\",\"zindex\":8,\"wtype\":\"fixed\",\"htype\":\"fixed\",\"opacity\":1,\"propagateEvents\":false,\"lineHeightAuto\":true,\"rotation\":0,\"scale\":1,\"aspectratio\":false},{\"name\":\"text\",\"type\":\"text\",\"lib\":\"common\",\"caption\":\"BMI : 28 kg/m2...\",\"text\":\"BMI : 28 kg/m2\",\"textFont\":\"Trebuchet MS,sans-serif\",\"textFontStyle\":\"400\",\"textProperties\":[],\"textSize\":\"32\",\"textColor\":{\"type\":\"solid\",\"color\":\"rgba(113,113,113,1)\"},\"backgroundColor\":\"none\",\"width\":199,\"height\":39,\"lineHeight\":39,\"textAlign\":\"left\",\"textSpacing\":\"-1.6\",\"enableShadow\":false,\"autoResize\":true,\"v2\":true,\"left\":379,\"top\":171,\"vpos\":\"top\",\"hpos\":\"left\",\"visible\":true,\"group\":\"root\",\"id\":\"box-56\",\"zindex\":8,\"wtype\":\"fixed\",\"htype\":\"fixed\",\"opacity\":1,\"propagateEvents\":false,\"lineHeightAuto\":true,\"rotation\":0,\"scale\":1,\"aspectratio\":false},{\"name\":\"text\",\"type\":\"text\",\"lib\":\"common\",\"caption\":\"Weight : 69 Kg...\",\"text\":\"Weight : 69 Kg\",\"textFont\":\"Trebuchet MS,sans-serif\",\"textFontStyle\":\"400\",\"textProperties\":[],\"textSize\":\"32\",\"textColor\":{\"type\":\"solid\",\"color\":\"rgba(113,113,113,1)\"},\"backgroundColor\":\"none\",\"width\":190,\"height\":39,\"lineHeight\":39,\"textAlign\":\"left\",\"textSpacing\":\"-1.6\",\"enableShadow\":false,\"autoResize\":true,\"v2\":true,\"left\":379,\"top\":130,\"vpos\":\"top\",\"hpos\":\"left\",\"visible\":true,\"group\":\"root\",\"id\":\"box-58\",\"zindex\":8,\"wtype\":\"fixed\",\"htype\":\"fixed\",\"opacity\":1,\"propagateEvents\":false,\"lineHeightAuto\":true,\"rotation\":0,\"scale\":1,\"aspectratio\":false},{\"name\":\"verticalline\",\"type\":\"verticalline\",\"lib\":\"common\",\"caption\":\"Vertical line 3\",\"width\":1,\"height\":229,\"color\":\"A6A6A6\",\"weight\":1,\"left\":341,\"top\":10,\"vpos\":\"top\",\"hpos\":\"left\",\"visible\":true,\"group\":\"root\",\"id\":\"box-52\",\"zindex\":7,\"wtype\":\"fixed\",\"htype\":\"fixed\",\"opacity\":1,\"borderStyle\":\"solid\",\"rotation\":0,\"scale\":1,\"aspectratio\":false},{\"name\":\"verticalline\",\"type\":\"verticalline\",\"lib\":\"common\",\"caption\":\"Vertical line 5\",\"width\":1,\"height\":229,\"color\":\"A6A6A6\",\"weight\":1,\"left\":1064,\"top\":15,\"vpos\":\"top\",\"hpos\":\"left\",\"visible\":true,\"group\":\"root\",\"id\":\"box-54\",\"zindex\":7,\"wtype\":\"fixed\",\"htype\":\"fixed\",\"opacity\":1,\"borderStyle\":\"solid\",\"rotation\":0,\"scale\":1,\"aspectratio\":false},{\"name\":\"verticalline\",\"type\":\"verticalline\",\"lib\":\"common\",\"caption\":\"Vertical line 6\",\"width\":1,\"height\":229,\"color\":\"A6A6A6\",\"weight\":1,\"left\":733,\"top\":10,\"vpos\":\"top\",\"hpos\":\"left\",\"visible\":true,\"group\":\"root\",\"id\":\"box-55\",\"zindex\":7,\"wtype\":\"fixed\",\"htype\":\"fixed\",\"opacity\":1,\"borderStyle\":\"solid\",\"rotation\":0,\"scale\":1,\"aspectratio\":false},{\"name\":\"datetime\",\"type\":\"datetime\",\"lib\":\"common\",\"caption\":\"Date/Time 1\",\"width\":376,\"height\":41,\"backgroundColor\":\"none\",\"borderWidth\":\"1\",\"borderColor\":{\"type\":\"solid\",\"color\":\"rgba(102,150,200,1)\"},\"textFont\":\"Montserrat\",\"textFontStyle\":\"400\",\"textSize\":\"20\",\"textColor\":\"383838\",\"textProperties\":[],\"textAlign\":\"center\",\"dateFormat\":\"yearmonthday\",\"dayFormat\":\"superscript\",\"monthFormat\":\"long\",\"yearFormat\":\"long\",\"timeFormat\":\"24\",\"dateGeneralFormat\":\"timedate\",\"dayNameFormat\":\"long\",\"dateSeparator\":\"-\",\"left\":1064,\"top\":201,\"vpos\":\"top\",\"hpos\":\"left\",\"visible\":true,\"group\":\"root\",\"id\":\"box-50\",\"zindex\":5,\"wtype\":\"fixed\",\"htype\":\"fixed\",\"opacity\":1,\"rotation\":0,\"scale\":1,\"aspectratio\":false},{\"name\":\"text\",\"type\":\"text\",\"lib\":\"common\",\"caption\":\"MRN : 12345679...\",\"text\":\"MRN : 12345679&nbsp;\",\"textFont\":\"Trebuchet MS,sans-serif\",\"textFontStyle\":\"400\",\"textProperties\":[],\"textSize\":\"32\",\"textColor\":{\"type\":\"solid\",\"color\":\"rgba(113,113,113,1)\"},\"backgroundColor\":\"none\",\"width\":218,\"height\":39,\"lineHeight\":39,\"textAlign\":\"left\",\"textSpacing\":\"-1.6\",\"enableShadow\":false,\"autoResize\":true,\"v2\":true,\"left\":76,\"top\":171,\"vpos\":\"top\",\"hpos\":\"left\",\"visible\":true,\"group\":\"root\",\"id\":\"box-48\",\"zindex\":4,\"wtype\":\"fixed\",\"htype\":\"fixed\",\"opacity\":1,\"propagateEvents\":false,\"lineHeightAuto\":true,\"rotation\":0,\"scale\":1,\"aspectratio\":false},{\"name\":\"verticalline\",\"type\":\"verticalline\",\"lib\":\"common\",\"caption\":\"Vertical line 1\",\"width\":1,\"height\":658,\"color\":{\"type\":\"solid\",\"color\":\"rgba(70,127,215,1)\"},\"weight\":1,\"left\":734,\"top\":242,\"vpos\":\"top\",\"hpos\":\"left\",\"visible\":true,\"group\":\"root\",\"id\":\"box-47\",\"zindex\":3,\"wtype\":\"fixed\",\"htype\":\"fixed\",\"opacity\":1,\"borderStyle\":\"solid\",\"rotation\":0,\"scale\":1,\"aspectratio\":false},{\"name\":\"text\",\"type\":\"text\",\"lib\":\"common\",\"caption\":\"John Doe\",\"text\":\"John Doe&nbsp;\",\"textFont\":\"Trebuchet MS,sans-serif\",\"textFontStyle\":\"400\",\"textProperties\":[],\"textSize\":\"64\",\"textColor\":{\"type\":\"solid\",\"color\":\"rgba(117,85,88,1)\"},\"backgroundColor\":\"none\",\"width\":272,\"height\":78,\"lineHeight\":78,\"textAlign\":\"left\",\"textSpacing\":\"-1.6\",\"enableShadow\":false,\"autoResize\":true,\"v2\":true,\"left\":40,\"top\":15,\"vpos\":\"top\",\"hpos\":\"left\",\"visible\":true,\"id\":\"box-46\",\"group\":\"root\",\"zindex\":2,\"wtype\":\"fixed\",\"htype\":\"fixed\",\"opacity\":1,\"propagateEvents\":false,\"lineHeightAuto\":true,\"rotation\":0,\"scale\":1,\"aspectratio\":false},{\"name\":\"basic_slider\",\"type\":\"basic_slider\",\"lib\":\"generic2\",\"caption\":\"Slider 1\",\"width\":347,\"height\":41,\"barThickness\":4,\"barColor\":\"e3e6e8\",\"fillBarColor\":{\"type\":\"solid\",\"color\":\"rgba(255,255,0,1)\"},\"sliderColor\":\"ffffff\",\"barBorderRadius\":2,\"sliderBorderWidth\":1,\"sliderBorderRadius\":\"50%\",\"borderColor\":\"d4d7d9\",\"sliderPosition\":\"70\",\"left\":379,\"top\":359,\"vpos\":\"top\",\"hpos\":\"left\",\"visible\":true,\"id\":\"box-66\",\"group\":\"root\",\"zindex\":9,\"wtype\":\"fixed\",\"htype\":\"fixed\",\"opacity\":1,\"rotation\":0,\"scale\":1,\"aspectratio\":false,\"blurB\":0,\"saturateB\":1,\"brightnessB\":1,\"contrastB\":1,\"grayscaleB\":0,\"sepiaB\":0,\"invertB\":0,\"hueRotateB\":0,\"actions\":[{\"title\":\"Interaction 1\",\"type\":\"sliderdrag\",\"actionId\":\"set-variable\",\"callback\":true,\"variableName\":\"propofol_amount\",\"valueSource\":\"callbackvalue\",\"callbackvariable\":\"value\",\"delay\":\"0\",\"guid\":\"9d9286b6-7bab-4ebb-947b-50a5ffb030a0\",\"callbackAction\":{\"actionId\":\"change-property\",\"callback\":true,\"pageId\":\"2\",\"itemId\":\"box-88\",\"propertyId\":\"text\",\"valueType\":\"readFromVariable\",\"variableName\":\"propofol_amount\",\"delay\":\"0\",\"guid\":\"730e44a2-4cd9-46f8-9726-319a49f90ac6\",\"callbackAction\":{\"actionId\":\"change-screen-state\",\"callback\":false,\"pageId\":\"2\",\"stateId\":\"576fe87f-7989-4c31-93a7-292d92d03cfb\",\"delay\":\"0\",\"guid\":\"de37f8cc-6236-4d8d-8f42-062d7ee64e3d\",\"clb_disabled\":false},\"clb_disabled\":false},\"sort\":0}]},{\"name\":\"basic_slider\",\"type\":\"basic_slider\",\"lib\":\"generic2\",\"caption\":\"Slider 2\",\"width\":347,\"height\":41,\"barThickness\":4,\"barColor\":\"e3e6e8\",\"fillBarColor\":{\"type\":\"solid\",\"color\":\"rgba(198,198,198,1)\"},\"sliderColor\":\"ffffff\",\"barBorderRadius\":2,\"sliderBorderWidth\":1,\"sliderBorderRadius\":\"50%\",\"borderColor\":\"d4d7d9\",\"sliderPosition\":\"44\",\"left\":373,\"top\":452,\"vpos\":\"top\",\"hpos\":\"left\",\"visible\":true,\"id\":\"box-71\",\"group\":\"root\",\"zindex\":9,\"wtype\":\"fixed\",\"htype\":\"fixed\",\"opacity\":1,\"rotation\":0,\"scale\":1,\"aspectratio\":false,\"actions\":[{\"title\":\"Set Val\",\"type\":\"sliderdrag\",\"actionId\":\"set-variable\",\"callback\":true,\"variableName\":\"lido\",\"valueSource\":\"callbackvalue\",\"callbackvariable\":\"value\",\"delay\":\"0\",\"guid\":\"f98b8443-5652-42df-a2d0-21803d04df1e\",\"callbackAction\":{\"actionId\":\"change-property\",\"callback\":true,\"pageId\":\"2\",\"itemId\":\"box-90\",\"propertyId\":\"text\",\"valueType\":\"readFromVariable\",\"variableName\":\"lido\",\"delay\":\"0\",\"guid\":\"d6e567aa-4b61-42f2-b2e2-1c17cbe5e14c\",\"callbackAction\":{\"actionId\":\"change-screen-state\",\"callback\":false,\"pageId\":\"2\",\"stateId\":\"576fe87f-7989-4c31-93a7-292d92d03cfb\",\"delay\":\"0\",\"guid\":\"3da0c010-6cc1-4b0e-be98-3639dde14c90\",\"clb_disabled\":false},\"clb_disabled\":false},\"sort\":0}]},{\"name\":\"basic_slider\",\"type\":\"basic_slider\",\"lib\":\"generic2\",\"caption\":\"Slider 3\",\"width\":347,\"height\":41,\"barThickness\":4,\"barColor\":\"e3e6e8\",\"fillBarColor\":{\"type\":\"solid\",\"color\":\"rgba(249,51,36,1)\"},\"sliderColor\":\"ffffff\",\"barBorderRadius\":2,\"sliderBorderWidth\":1,\"sliderBorderRadius\":\"50%\",\"borderColor\":\"d4d7d9\",\"sliderPosition\":\"61\",\"left\":379,\"top\":735,\"vpos\":\"top\",\"hpos\":\"left\",\"visible\":true,\"id\":\"box-75\",\"group\":\"root\",\"zindex\":9,\"wtype\":\"fixed\",\"htype\":\"fixed\",\"opacity\":1,\"rotation\":0,\"scale\":1,\"aspectratio\":false,\"actions\":[{\"title\":\"r\",\"type\":\"sliderdrag\",\"actionId\":\"set-variable\",\"callback\":true,\"variableName\":\"rocu\",\"valueSource\":\"callbackvalue\",\"callbackvariable\":\"value\",\"delay\":\"0\",\"guid\":\"8de41d81-e9d5-44a6-a223-1ac97b1b8b55\",\"callbackAction\":{\"actionId\":\"change-property\",\"callback\":true,\"pageId\":\"2\",\"itemId\":\"box-93\",\"propertyId\":\"text\",\"valueType\":\"readFromVariable\",\"variableName\":\"rocu\",\"delay\":\"0\",\"guid\":\"641f6d09-c333-49dc-b11a-79e14f1b718a\",\"callbackAction\":{\"actionId\":\"change-screen-state\",\"callback\":false,\"pageId\":\"2\",\"stateId\":\"576fe87f-7989-4c31-93a7-292d92d03cfb\",\"delay\":\"0\",\"guid\":\"a2d70c60-be48-4771-8a5e-65d9dc19f16e\",\"clb_disabled\":false},\"clb_disabled\":false},\"sort\":0}]},{\"name\":\"basic_slider\",\"type\":\"basic_slider\",\"lib\":\"generic2\",\"caption\":\"Slider 4\",\"width\":347,\"height\":41,\"barThickness\":4,\"barColor\":\"e3e6e8\",\"fillBarColor\":{\"type\":\"solid\",\"color\":\"rgba(160,58,207,1)\"},\"sliderColor\":\"ffffff\",\"barBorderRadius\":2,\"sliderBorderWidth\":1,\"sliderBorderRadius\":\"50%\",\"borderColor\":\"d4d7d9\",\"sliderPosition\":\"40\",\"left\":379,\"top\":648,\"vpos\":\"top\",\"hpos\":\"left\",\"visible\":true,\"id\":\"box-76\",\"group\":\"root\",\"zindex\":9,\"wtype\":\"fixed\",\"htype\":\"fixed\",\"opacity\":1,\"rotation\":0,\"scale\":1,\"aspectratio\":false,\"actions\":[{\"title\":\"frt\",\"type\":\"sliderdrag\",\"actionId\":\"set-variable\",\"callback\":true,\"variableName\":\"phen\",\"valueSource\":\"callbackvalue\",\"callbackvariable\":\"value\",\"delay\":\"0\",\"guid\":\"d72a3a63-1473-49fb-9762-435b1ac2e7dc\",\"callbackAction\":{\"actionId\":\"change-property\",\"callback\":true,\"pageId\":\"2\",\"itemId\":\"box-92\",\"propertyId\":\"text\",\"valueType\":\"readFromVariable\",\"variableName\":\"phen\",\"delay\":\"0\",\"guid\":\"a97221ea-b9e9-424e-9ba5-5c6db5a3ecfd\",\"callbackAction\":{\"actionId\":\"change-screen-state\",\"callback\":false,\"pageId\":\"2\",\"stateId\":\"576fe87f-7989-4c31-93a7-292d92d03cfb\",\"delay\":\"0\",\"guid\":\"cd6f060b-3d7d-41c3-a4d7-f6fa40818352\",\"clb_disabled\":false},\"clb_disabled\":false},\"sort\":0}]},{\"name\":\"basic_slider\",\"type\":\"basic_slider\",\"lib\":\"generic2\",\"caption\":\"Slider 5\",\"width\":347,\"height\":41,\"barThickness\":4,\"barColor\":\"e3e6e8\",\"fillBarColor\":{\"type\":\"solid\",\"color\":\"rgba(102,150,200,1)\"},\"sliderColor\":\"ffffff\",\"barBorderRadius\":2,\"sliderBorderWidth\":1,\"sliderBorderRadius\":\"50%\",\"borderColor\":\"d4d7d9\",\"sliderPosition\":\"33\",\"left\":379,\"top\":550,\"vpos\":\"top\",\"hpos\":\"left\",\"visible\":true,\"id\":\"box-77\",\"group\":\"root\",\"zindex\":9,\"wtype\":\"fixed\",\"htype\":\"fixed\",\"opacity\":1,\"rotation\":0,\"scale\":1,\"aspectratio\":false,\"actions\":[{\"title\":\"f\",\"type\":\"sliderdrag\",\"actionId\":\"set-variable\",\"callback\":true,\"variableName\":\"fent\",\"valueSource\":\"callbackvalue\",\"callbackvariable\":\"value\",\"delay\":\"0\",\"guid\":\"ced73a24-634c-4d71-8d1b-dc359f0508e7\",\"callbackAction\":{\"actionId\":\"change-property\",\"callback\":true,\"pageId\":\"2\",\"itemId\":\"box-91\",\"propertyId\":\"text\",\"valueType\":\"readFromVariable\",\"variableName\":\"fent\",\"delay\":\"0\",\"guid\":\"06bae57c-5c05-4c3d-987a-f1a51aef7c71\",\"callbackAction\":{\"actionId\":\"change-screen-state\",\"callback\":false,\"pageId\":\"2\",\"stateId\":\"576fe87f-7989-4c31-93a7-292d92d03cfb\",\"delay\":\"0\",\"guid\":\"86c7dbea-2343-421b-b85a-b166e6a16fab\",\"clb_disabled\":false},\"clb_disabled\":false},\"sort\":0}]},{\"name\":\"basic_textfield\",\"type\":\"basic_textfield\",\"lib\":\"generic2\",\"caption\":\"Text Field prop\",\"width\":338,\"height\":55,\"value\":\"Propofol  ( mg ) \",\"placeholder\":\"Placeholder\",\"inputtype\":\"text\",\"backgroundColor\":{\"type\":\"solid\",\"color\":\"rgba(255,255,122,1)\"},\"textFont\":\"Arial,sans-serif\",\"textFontStyle\":\"400\",\"textSize\":\"28\",\"textColor\":{\"type\":\"solid\",\"color\":\"rgba(56,56,56,1)\"},\"textProperties\":[],\"placeholderColor\":\"c1c8c9\",\"borderWidth\":1,\"borderStyle\":\"solid\",\"borderRadius\":4,\"borderColor\":{\"type\":\"solid\",\"color\":\"rgba(255,255,0,1)\"},\"textAlign\":\"center\",\"left\":22,\"top\":359,\"vpos\":\"top\",\"hpos\":\"left\",\"visible\":true,\"group\":\"root\",\"id\":\"box-67\",\"zindex\":10,\"wtype\":\"fixed\",\"htype\":\"fixed\",\"opacity\":1,\"rotation\":0,\"scale\":1,\"aspectratio\":false,\"userCaption\":true},{\"name\":\"basic_textfield\",\"type\":\"basic_textfield\",\"lib\":\"generic2\",\"caption\":\"Text Field lido\",\"width\":338,\"height\":55,\"value\":\"Lidocaine  (mg)\",\"placeholder\":\"Placeholder\",\"inputtype\":\"text\",\"backgroundColor\":{\"type\":\"solid\",\"color\":\"rgba(198,198,198,1)\"},\"textFont\":\"Arial,sans-serif\",\"textFontStyle\":\"400\",\"textSize\":\"28\",\"textColor\":{\"type\":\"solid\",\"color\":\"rgba(56,56,56,1)\"},\"textProperties\":[],\"placeholderColor\":\"c1c8c9\",\"borderWidth\":1,\"borderStyle\":\"solid\",\"borderRadius\":4,\"borderColor\":{\"type\":\"solid\",\"color\":\"rgba(208,212,0,1)\"},\"textAlign\":\"center\",\"left\":22,\"top\":445,\"vpos\":\"top\",\"hpos\":\"left\",\"visible\":true,\"group\":\"root\",\"id\":\"box-72\",\"zindex\":10,\"wtype\":\"fixed\",\"htype\":\"fixed\",\"opacity\":1,\"rotation\":0,\"scale\":1,\"aspectratio\":false,\"userCaption\":true},{\"name\":\"basic_textfield\",\"type\":\"basic_textfield\",\"lib\":\"generic2\",\"caption\":\"Text Field Phenylephrine\",\"width\":338,\"height\":55,\"value\":\"Phenylephrine (mcg) \",\"placeholder\":\"Placeholder\",\"inputtype\":\"text\",\"backgroundColor\":{\"type\":\"solid\",\"color\":\"rgba(234,114,252,1)\"},\"textFont\":\"Arial,sans-serif\",\"textFontStyle\":\"400\",\"textSize\":\"28\",\"textColor\":{\"type\":\"solid\",\"color\":\"rgba(56,56,56,1)\"},\"textProperties\":[],\"placeholderColor\":\"c1c8c9\",\"borderWidth\":1,\"borderStyle\":\"solid\",\"borderRadius\":4,\"borderColor\":{\"type\":\"solid\",\"color\":\"rgba(255,255,0,1)\"},\"textAlign\":\"center\",\"left\":22,\"top\":641,\"vpos\":\"top\",\"hpos\":\"left\",\"visible\":true,\"group\":\"root\",\"id\":\"box-73\",\"zindex\":10,\"wtype\":\"fixed\",\"htype\":\"fixed\",\"opacity\":1,\"rotation\":0,\"scale\":1,\"aspectratio\":false,\"userCaption\":true},{\"name\":\"basic_textfield\",\"type\":\"basic_textfield\",\"lib\":\"generic2\",\"caption\":\"Text Field rocur\",\"width\":338,\"height\":55,\"value\":\"Rocuronium (mg)\",\"placeholder\":\"Placeholder\",\"inputtype\":\"text\",\"backgroundColor\":{\"type\":\"solid\",\"color\":\"rgba(226,15,0,1)\"},\"textFont\":\"Arial,sans-serif\",\"textFontStyle\":\"400\",\"textSize\":\"28\",\"textColor\":{\"type\":\"solid\",\"color\":\"rgba(56,56,56,1)\"},\"textProperties\":[],\"placeholderColor\":\"c1c8c9\",\"borderWidth\":1,\"borderStyle\":\"solid\",\"borderRadius\":4,\"borderColor\":{\"type\":\"solid\",\"color\":\"rgba(255,255,0,1)\"},\"textAlign\":\"center\",\"left\":22,\"top\":721,\"vpos\":\"top\",\"hpos\":\"left\",\"visible\":true,\"group\":\"root\",\"id\":\"box-79\",\"zindex\":10,\"wtype\":\"fixed\",\"htype\":\"fixed\",\"opacity\":1,\"rotation\":0,\"scale\":1,\"aspectratio\":false,\"userCaption\":true},{\"name\":\"basic_textfield\",\"type\":\"basic_textfield\",\"lib\":\"generic2\",\"caption\":\"Text Field 2\",\"width\":733,\"height\":77,\"value\":\"Induction Medication\",\"placeholder\":\"Placeholder\",\"inputtype\":\"text\",\"backgroundColor\":{\"type\":\"solid\",\"color\":\"rgba(177,190,227,1)\"},\"textFont\":\"Arial,sans-serif\",\"textFontStyle\":\"400\",\"textSize\":\"64\",\"textColor\":{\"type\":\"solid\",\"color\":\"rgba(233,240,215,1)\"},\"textProperties\":[],\"placeholderColor\":\"c1c8c9\",\"borderWidth\":\"5\",\"borderStyle\":\"solid\",\"borderRadius\":4,\"borderColor\":{\"type\":\"solid\",\"color\":\"rgba(85,85,85,1)\"},\"textAlign\":\"center\",\"left\":2,\"top\":242,\"vpos\":\"top\",\"hpos\":\"left\",\"visible\":true,\"id\":\"box-69\",\"group\":\"root\",\"zindex\":11,\"wtype\":\"fixed\",\"htype\":\"fixed\",\"opacity\":1,\"rotation\":0,\"scale\":1,\"aspectratio\":false},{\"name\":\"basic_textfield\",\"type\":\"basic_textfield\",\"lib\":\"generic2\",\"caption\":\"Text Field 3\",\"width\":698,\"height\":77,\"value\":\"Blood Pressure\",\"placeholder\":\"Placeholder\",\"inputtype\":\"text\",\"backgroundColor\":{\"type\":\"solid\",\"color\":\"rgba(177,190,227,1)\"},\"textFont\":\"Arial,sans-serif\",\"textFontStyle\":\"400\",\"textSize\":\"64\",\"textColor\":{\"type\":\"solid\",\"color\":\"rgba(233,240,215,1)\"},\"textProperties\":[],\"placeholderColor\":\"c1c8c9\",\"borderWidth\":\"5\",\"borderStyle\":\"solid\",\"borderRadius\":4,\"borderColor\":{\"type\":\"solid\",\"color\":\"rgba(85,85,85,1)\"},\"textAlign\":\"center\",\"left\":733,\"top\":244,\"vpos\":\"top\",\"hpos\":\"left\",\"visible\":true,\"id\":\"box-70\",\"group\":\"root\",\"zindex\":11,\"wtype\":\"fixed\",\"htype\":\"fixed\",\"opacity\":1,\"rotation\":0,\"scale\":1,\"aspectratio\":false},{\"name\":\"basic_textfield\",\"type\":\"basic_textfield\",\"lib\":\"generic2\",\"caption\":\"Text Field fenta\",\"width\":338,\"height\":55,\"value\":\"Fentanyl (mcg)\",\"placeholder\":\"Placeholder\",\"inputtype\":\"text\",\"backgroundColor\":{\"type\":\"solid\",\"color\":\"rgba(101,180,206,1)\"},\"textFont\":\"Arial,sans-serif\",\"textFontStyle\":\"400\",\"textSize\":\"28\",\"textColor\":{\"type\":\"solid\",\"color\":\"rgba(56,56,56,1)\"},\"textProperties\":[],\"placeholderColor\":\"c1c8c9\",\"borderWidth\":1,\"borderStyle\":\"solid\",\"borderRadius\":4,\"borderColor\":{\"type\":\"solid\",\"color\":\"rgba(27,77,87,1)\"},\"textAlign\":\"center\",\"left\":22,\"top\":543,\"vpos\":\"top\",\"hpos\":\"left\",\"visible\":true,\"group\":\"root\",\"id\":\"box-78\",\"zindex\":10,\"wtype\":\"fixed\",\"htype\":\"fixed\",\"opacity\":1,\"rotation\":0,\"scale\":1,\"aspectratio\":false,\"userCaption\":true},{\"name\":\"basic_button_text\",\"type\":\"basic_button_text\",\"lib\":\"generic2\",\"caption\":\"Button with Background 1\",\"width\":177,\"height\":80,\"backgroundColor\":{\"type\":\"solid\",\"color\":\"rgba(102,150,200,1)\"},\"borderWidth\":0,\"borderColor\":\"#025abf\",\"borderRadius\":4,\"text\":\"Other Opioids\",\"textFont\":\"Arial,sans-serif\",\"textFontStyle\":\"400\",\"textSize\":\"28\",\"textColor\":\"FFFFFF\",\"textProperties\":[],\"textAlign\":\"center\",\"iconpos\":\"none\",\"left\":32,\"top\":800,\"vpos\":\"top\",\"hpos\":\"left\",\"visible\":true,\"id\":\"box-81\",\"group\":\"root\",\"zindex\":13,\"wtype\":\"fixed\",\"htype\":\"fixed\",\"opacity\":1,\"rotation\":0,\"scale\":1,\"aspectratio\":false,\"actions\":[{\"title\":\"Interaction 1\",\"type\":\"tap\",\"actionId\":\"go-to-page\",\"callback\":false,\"pageId\":\"4\",\"animation\":\"fade\",\"animationEasing\":\"Cubic.easeOut\",\"duration\":\"400\",\"delay\":\"0\",\"guid\":\"cf1fdbfe-2594-4acf-9780-ee410704e4e7\",\"sort\":0}]},{\"name\":\"basic_button_text\",\"type\":\"basic_button_text\",\"lib\":\"generic2\",\"caption\":\"Button with Background 3\",\"width\":177,\"height\":80,\"backgroundColor\":{\"type\":\"solid\",\"color\":\"rgba(255, 255, 0,1)\"},\"borderWidth\":0,\"borderColor\":\"#025abf\",\"borderRadius\":4,\"text\":\"Other Amestics\",\"textFont\":\"Arial,sans-serif\",\"textFontStyle\":\"400\",\"textSize\":\"28\",\"textColor\":{\"type\":\"solid\",\"color\":\"rgba(56,56,56,1)\"},\"textProperties\":[],\"textAlign\":\"center\",\"iconpos\":\"none\",\"left\":252,\"top\":800,\"vpos\":\"top\",\"hpos\":\"left\",\"visible\":true,\"id\":\"box-83\",\"group\":\"root\",\"zindex\":13,\"wtype\":\"fixed\",\"htype\":\"fixed\",\"opacity\":1,\"rotation\":0,\"scale\":1,\"aspectratio\":false,\"actions\":[{\"title\":\"Interaction 1\",\"type\":\"tap\",\"actionId\":\"go-to-page\",\"callback\":false,\"pageId\":\"7\",\"animation\":\"fade\",\"animationEasing\":\"Cubic.easeOut\",\"duration\":\"400\",\"delay\":\"0\",\"guid\":\"27252ddc-3dfc-45e9-8131-3dfb443a6bc0\",\"sort\":0}]},{\"name\":\"basic_button_text\",\"type\":\"basic_button_text\",\"lib\":\"generic2\",\"caption\":\"Button with Background 4\",\"width\":177,\"height\":80,\"backgroundColor\":{\"type\":\"solid\",\"color\":\"rgba(234,114,252,1)\"},\"borderWidth\":0,\"borderColor\":\"#025abf\",\"borderRadius\":4,\"text\":\"Other Pressors\",\"textFont\":\"Arial,sans-serif\",\"textFontStyle\":\"400\",\"textSize\":\"28\",\"textColor\":\"FFFFFF\",\"textProperties\":[],\"textAlign\":\"center\",\"iconpos\":\"none\",\"left\":489,\"top\":800,\"vpos\":\"top\",\"hpos\":\"left\",\"visible\":true,\"id\":\"box-84\",\"group\":\"root\",\"zindex\":13,\"wtype\":\"fixed\",\"htype\":\"fixed\",\"opacity\":1,\"rotation\":0,\"scale\":1,\"aspectratio\":false,\"actions\":[{\"title\":\"Interaction 1\",\"type\":\"tap\",\"actionId\":\"go-to-page\",\"callback\":false,\"pageId\":\"6\",\"animation\":\"fade\",\"animationEasing\":\"Cubic.easeOut\",\"duration\":\"400\",\"delay\":\"0\",\"guid\":\"e0c7afcc-7cd6-42de-8f44-39e342fc2dcd\",\"sort\":0}]},{\"name\":\"basic_button_text\",\"type\":\"basic_button_text\",\"lib\":\"generic2\",\"caption\":\"Button with Background 2\",\"width\":177,\"height\":80,\"backgroundColor\":{\"type\":\"solid\",\"color\":\"rgba(249,51,36,1)\"},\"borderWidth\":0,\"borderColor\":\"#025abf\",\"borderRadius\":4,\"text\":\"Exit\",\"textFont\":\"Arial,sans-serif\",\"textFontStyle\":\"400\",\"textSize\":\"36\",\"textColor\":\"FFFFFF\",\"textProperties\":[],\"textAlign\":\"center\",\"iconpos\":\"none\",\"left\":1233,\"top\":789,\"vpos\":\"top\",\"hpos\":\"left\",\"visible\":true,\"id\":\"box-82\",\"group\":\"root\",\"zindex\":13,\"wtype\":\"fixed\",\"htype\":\"fixed\",\"opacity\":1,\"rotation\":0,\"scale\":1,\"aspectratio\":false,\"actions\":[{\"title\":\"Interaction 1\",\"type\":\"click\",\"actionId\":\"go-to-page\",\"callback\":false,\"pageId\":\"1\",\"animation\":\"fade\",\"animationEasing\":\"Cubic.easeOut\",\"duration\":\"400\",\"delay\":\"0\",\"guid\":\"24e2c494-0883-41c4-ab86-dcc358a40f14\",\"sort\":0}]},{\"name\":\"text\",\"type\":\"text\",\"lib\":\"common\",\"caption\":\"text\",\"text\":\"150\",\"textFont\":\"Arial,sans-serif\",\"textFontStyle\":\"400\",\"textProperties\":[],\"textSize\":\"24\",\"textColor\":{\"type\":\"solid\",\"color\":\"rgba(255,255,0,1)\"},\"backgroundColor\":\"none\",\"width\":275,\"height\":29,\"lineHeight\":29,\"textAlign\":\"left\",\"textSpacing\":0,\"enableShadow\":false,\"autoResize\":false,\"v2\":true,\"left\":505,\"top\":330,\"vpos\":\"top\",\"hpos\":\"left\",\"visible\":true,\"id\":\"box-88\",\"group\":\"root\",\"zindex\":14,\"wtype\":\"fixed\",\"htype\":\"fixed\",\"opacity\":1,\"propagateEvents\":false,\"lineHeightAuto\":true,\"rotation\":0,\"scale\":1,\"aspectratio\":false,\"userCaption\":true,\"actions\":[]},{\"name\":\"text\",\"type\":\"text\",\"lib\":\"common\",\"caption\":\"text 2\",\"text\":\"100\",\"textFont\":\"Arial,sans-serif\",\"textFontStyle\":\"400\",\"textProperties\":[],\"textSize\":\"24\",\"textColor\":{\"type\":\"solid\",\"color\":\"rgba(198,198,198,1)\"},\"backgroundColor\":\"none\",\"width\":275,\"height\":29,\"lineHeight\":29,\"textAlign\":\"left\",\"textSpacing\":0,\"enableShadow\":false,\"autoResize\":false,\"v2\":true,\"left\":505,\"top\":414,\"vpos\":\"top\",\"hpos\":\"left\",\"visible\":true,\"id\":\"box-90\",\"group\":\"root\",\"zindex\":14,\"wtype\":\"fixed\",\"htype\":\"fixed\",\"opacity\":1,\"propagateEvents\":false,\"lineHeightAuto\":true,\"rotation\":0,\"scale\":1,\"aspectratio\":false,\"userCaption\":true,\"actions\":[]},{\"name\":\"text\",\"type\":\"text\",\"lib\":\"common\",\"caption\":\"text 3\",\"text\":\"100\",\"textFont\":\"Arial,sans-serif\",\"textFontStyle\":\"400\",\"textProperties\":[],\"textSize\":\"24\",\"textColor\":{\"type\":\"solid\",\"color\":\"rgba(43,89,162,1)\"},\"backgroundColor\":\"none\",\"width\":275,\"height\":29,\"lineHeight\":29,\"textAlign\":\"left\",\"textSpacing\":0,\"enableShadow\":false,\"autoResize\":false,\"v2\":true,\"left\":505,\"top\":521,\"vpos\":\"top\",\"hpos\":\"left\",\"visible\":true,\"id\":\"box-91\",\"group\":\"root\",\"zindex\":14,\"wtype\":\"fixed\",\"htype\":\"fixed\",\"opacity\":1,\"propagateEvents\":false,\"lineHeightAuto\":true,\"rotation\":0,\"scale\":1,\"aspectratio\":false,\"userCaption\":true,\"actions\":[]},{\"name\":\"text\",\"type\":\"text\",\"lib\":\"common\",\"caption\":\"text 4\",\"text\":\"100\",\"textFont\":\"Arial,sans-serif\",\"textFontStyle\":\"400\",\"textProperties\":[],\"textSize\":\"24\",\"textColor\":{\"type\":\"solid\",\"color\":\"rgba(183,47,214,1)\"},\"backgroundColor\":\"none\",\"width\":275,\"height\":29,\"lineHeight\":29,\"textAlign\":\"left\",\"textSpacing\":0,\"enableShadow\":false,\"autoResize\":false,\"v2\":true,\"left\":497,\"top\":612,\"vpos\":\"top\",\"hpos\":\"left\",\"visible\":true,\"id\":\"box-92\",\"group\":\"root\",\"zindex\":14,\"wtype\":\"fixed\",\"htype\":\"fixed\",\"opacity\":1,\"propagateEvents\":false,\"lineHeightAuto\":true,\"rotation\":0,\"scale\":1,\"aspectratio\":false,\"userCaption\":true,\"actions\":[]},{\"name\":\"text\",\"type\":\"text\",\"lib\":\"common\",\"caption\":\"text 5\",\"text\":\"100\",\"textFont\":\"Arial,sans-serif\",\"textFontStyle\":\"400\",\"textProperties\":[],\"textSize\":\"24\",\"textColor\":{\"type\":\"solid\",\"color\":\"rgba(209,25,105,1)\"},\"backgroundColor\":\"none\",\"width\":275,\"height\":29,\"lineHeight\":29,\"textAlign\":\"left\",\"textSpacing\":0,\"enableShadow\":false,\"autoResize\":false,\"v2\":true,\"left\":491,\"top\":712,\"vpos\":\"top\",\"hpos\":\"left\",\"visible\":true,\"id\":\"box-93\",\"group\":\"root\",\"zindex\":14,\"wtype\":\"fixed\",\"htype\":\"fixed\",\"opacity\":1,\"propagateEvents\":false,\"lineHeightAuto\":true,\"rotation\":0,\"scale\":1,\"aspectratio\":false,\"userCaption\":true,\"actions\":[]},{\"name\":\"image\",\"type\":\"image\",\"lib\":\"common\",\"caption\":\"ss2\",\"imgSrc\":{\"fileId\":\"08d3b5585c3baf0fd8ac52e8c44a8630.png\",\"assetType\":\"project\",\"bucketsource\":\"main\",\"name\":\"Screen Shot 2022-12-12 at 9.52.29 PM.png\"},\"repeat\":false,\"width\":658.448275862069,\"height\":402,\"borderWidth\":0,\"borderRadius\":0,\"borderColor\":\"#D1D1D1\",\"borderPos\":\"inside\",\"propagateEvents\":false,\"actions\":[],\"overlay\":false,\"autoResize\":false,\"aspectratio\":1.6379310344827587,\"assistiveDescription\":\"\",\"left\":750,\"top\":357,\"visible\":true,\"group\":\"root\",\"id\":\"box-217\",\"zindex\":16,\"hpos\":\"left\",\"vpos\":\"top\",\"wtype\":\"fixed\",\"htype\":\"fixed\",\"opacity\":1,\"borderStyle\":\"solid\",\"rotation\":0,\"scale\":1,\"userCaption\":true,\"hasMask\":false}]","template":{"id":"","stateid":""},"history":[],"id":"d36a0c09-b058-464f-be6a-c8a9af1bd182","actions":[],"groups":{}},{"title":"State 3","background":{"type":"solid","color":"rgba(169,151,196,1)"},"data":"[{\"name\":\"rectangle\",\"type\":\"rectangle\",\"lib\":\"common\",\"caption\":\"Rectangle 1\",\"backgroundColor\":{\"type\":\"solid\",\"color\":\"rgba(193,212,241,1)\"},\"borderWidth\":\"8\",\"borderColor\":{\"type\":\"solid\",\"color\":\"rgba(30,77,130,1)\"},\"borderRadius\":0,\"imgSrc\":{\"fileId\":\"\",\"name\":\"\",\"assetType\":\"\",\"bucketsource\":\"\",\"url\":\"\"},\"backgroundSizeType\":\"cover\",\"backgroundSizeValue\":35,\"width\":1440,\"height\":249,\"lineHeight\":20,\"lineHeightAuto\":true,\"actions\":[],\"textFont\":\"Arial,sans-serif\",\"textFontStyle\":\"400\",\"textSize\":16,\"textColor\":\"#383838\",\"textProperties\":[],\"textSpacing\":0,\"textAlign\":\"center\",\"text\":\"\",\"typeName\":\"rectangle\",\"left\":0,\"top\":0,\"vpos\":\"top\",\"hpos\":\"left\",\"visible\":true,\"id\":\"box-45\",\"group\":\"root\",\"zindex\":1,\"wtype\":\"fixed\",\"htype\":\"fixed\",\"opacity\":1,\"borderStyle\":\"solid\",\"rotation\":0,\"scale\":1,\"aspectratio\":false},{\"name\":\"text\",\"type\":\"text\",\"lib\":\"common\",\"caption\":\"Gender : Male...\",\"text\":\"Gender : Male\",\"textFont\":\"Trebuchet MS,sans-serif\",\"textFontStyle\":\"400\",\"textProperties\":[],\"textSize\":\"32\",\"textColor\":{\"type\":\"solid\",\"color\":\"rgba(113,113,113,1)\"},\"backgroundColor\":\"none\",\"width\":186,\"height\":39,\"lineHeight\":39,\"textAlign\":\"left\",\"textSpacing\":\"-1.6\",\"enableShadow\":false,\"autoResize\":true,\"v2\":true,\"left\":379,\"top\":47,\"vpos\":\"top\",\"hpos\":\"left\",\"visible\":true,\"group\":\"root\",\"id\":\"box-64\",\"zindex\":8,\"wtype\":\"fixed\",\"htype\":\"fixed\",\"opacity\":1,\"propagateEvents\":false,\"lineHeightAuto\":true,\"rotation\":0,\"scale\":1,\"aspectratio\":false},{\"name\":\"text\",\"type\":\"text\",\"lib\":\"common\",\"caption\":\"DOB: 01/01/2022...\",\"text\":\"DOB: 01/01/2022\",\"textFont\":\"Trebuchet MS,sans-serif\",\"textFontStyle\":\"400\",\"textProperties\":[],\"textSize\":\"32\",\"textColor\":{\"type\":\"solid\",\"color\":\"rgba(113,113,113,1)\"},\"backgroundColor\":\"none\",\"width\":230,\"height\":39,\"lineHeight\":39,\"textAlign\":\"left\",\"textSpacing\":\"-1.6\",\"enableShadow\":false,\"autoResize\":true,\"v2\":true,\"left\":76,\"top\":130,\"vpos\":\"top\",\"hpos\":\"left\",\"visible\":true,\"group\":\"root\",\"id\":\"box-61\",\"zindex\":8,\"wtype\":\"fixed\",\"htype\":\"fixed\",\"opacity\":1,\"propagateEvents\":false,\"lineHeightAuto\":true,\"rotation\":0,\"scale\":1,\"aspectratio\":false},{\"name\":\"text\",\"type\":\"text\",\"lib\":\"common\",\"caption\":\"Height : 174 cm...\",\"text\":\"Height : 174 cm&nbsp;\",\"textFont\":\"Trebuchet MS,sans-serif\",\"textFontStyle\":\"400\",\"textProperties\":[],\"textSize\":\"32\",\"textColor\":{\"type\":\"solid\",\"color\":\"rgba(113,113,113,1)\"},\"backgroundColor\":\"none\",\"width\":216,\"height\":39,\"lineHeight\":39,\"textAlign\":\"left\",\"textSpacing\":\"-1.6\",\"enableShadow\":false,\"autoResize\":true,\"v2\":true,\"left\":379,\"top\":86,\"vpos\":\"top\",\"hpos\":\"left\",\"visible\":true,\"group\":\"root\",\"id\":\"box-62\",\"zindex\":8,\"wtype\":\"fixed\",\"htype\":\"fixed\",\"opacity\":1,\"propagateEvents\":false,\"lineHeightAuto\":true,\"rotation\":0,\"scale\":1,\"aspectratio\":false},{\"name\":\"text\",\"type\":\"text\",\"lib\":\"common\",\"caption\":\"Surgery:\\nTotal Colec...\",\"text\":\"Surgery:&nbsp;<br>Total Colectomy with&nbsp;<br>pancreatico-jejunostomy&nbsp;\",\"textFont\":\"Trebuchet MS,sans-serif\",\"textFontStyle\":\"400\",\"textProperties\":[],\"textSize\":\"32\",\"textColor\":{\"type\":\"solid\",\"color\":\"rgba(113,113,113,1)\"},\"backgroundColor\":\"none\",\"width\":336,\"height\":117,\"lineHeight\":39,\"textAlign\":\"left\",\"textSpacing\":\"-1.6\",\"enableShadow\":false,\"autoResize\":true,\"v2\":true,\"left\":1084,\"top\":47,\"vpos\":\"top\",\"hpos\":\"left\",\"visible\":true,\"group\":\"root\",\"id\":\"box-63\",\"zindex\":8,\"wtype\":\"fixed\",\"htype\":\"fixed\",\"opacity\":1,\"propagateEvents\":false,\"lineHeightAuto\":true,\"rotation\":0,\"scale\":1,\"aspectratio\":false},{\"name\":\"text\",\"type\":\"text\",\"lib\":\"common\",\"caption\":\"Age : 86 y\",\"text\":\"Age : 86 y&nbsp;\",\"textFont\":\"Trebuchet MS,sans-serif\",\"textFontStyle\":\"400\",\"textProperties\":[],\"textSize\":\"32\",\"textColor\":{\"type\":\"solid\",\"color\":\"rgba(113,113,113,1)\"},\"backgroundColor\":\"none\",\"width\":139,\"height\":39,\"lineHeight\":39,\"textAlign\":\"left\",\"textSpacing\":\"-1.6\",\"enableShadow\":false,\"autoResize\":true,\"v2\":true,\"left\":76,\"top\":91,\"vpos\":\"top\",\"hpos\":\"left\",\"visible\":true,\"group\":\"root\",\"id\":\"box-60\",\"zindex\":8,\"wtype\":\"fixed\",\"htype\":\"fixed\",\"opacity\":1,\"propagateEvents\":false,\"lineHeightAuto\":true,\"rotation\":0,\"scale\":1,\"aspectratio\":false},{\"name\":\"text\",\"type\":\"text\",\"lib\":\"common\",\"caption\":\"Allergies :\\nPenicill...\",\"text\":\"Allergies :<br><span style=\\\"color: rgba(249,51,36,1);\\\">Penicillin</span><br><br>\",\"textFont\":\"Trebuchet MS,sans-serif\",\"textFontStyle\":\"400\",\"textProperties\":[],\"textSize\":\"32\",\"textColor\":{\"type\":\"solid\",\"color\":\"rgba(113,113,113,1)\"},\"backgroundColor\":\"none\",\"width\":203,\"height\":117,\"lineHeight\":39,\"textAlign\":\"left\",\"textSpacing\":\"-1.6\",\"enableShadow\":false,\"autoResize\":false,\"v2\":true,\"left\":766,\"top\":47,\"vpos\":\"top\",\"hpos\":\"left\",\"visible\":true,\"group\":\"root\",\"id\":\"box-59\",\"zindex\":8,\"wtype\":\"fixed\",\"htype\":\"fixed\",\"opacity\":1,\"propagateEvents\":false,\"lineHeightAuto\":true,\"rotation\":0,\"scale\":1,\"aspectratio\":false},{\"name\":\"text\",\"type\":\"text\",\"lib\":\"common\",\"caption\":\"BMI : 28 kg/m2...\",\"text\":\"BMI : 28 kg/m2\",\"textFont\":\"Trebuchet MS,sans-serif\",\"textFontStyle\":\"400\",\"textProperties\":[],\"textSize\":\"32\",\"textColor\":{\"type\":\"solid\",\"color\":\"rgba(113,113,113,1)\"},\"backgroundColor\":\"none\",\"width\":199,\"height\":39,\"lineHeight\":39,\"textAlign\":\"left\",\"textSpacing\":\"-1.6\",\"enableShadow\":false,\"autoResize\":true,\"v2\":true,\"left\":379,\"top\":171,\"vpos\":\"top\",\"hpos\":\"left\",\"visible\":true,\"group\":\"root\",\"id\":\"box-56\",\"zindex\":8,\"wtype\":\"fixed\",\"htype\":\"fixed\",\"opacity\":1,\"propagateEvents\":false,\"lineHeightAuto\":true,\"rotation\":0,\"scale\":1,\"aspectratio\":false},{\"name\":\"text\",\"type\":\"text\",\"lib\":\"common\",\"caption\":\"Weight : 69 Kg...\",\"text\":\"Weight : 69 Kg\",\"textFont\":\"Trebuchet MS,sans-serif\",\"textFontStyle\":\"400\",\"textProperties\":[],\"textSize\":\"32\",\"textColor\":{\"type\":\"solid\",\"color\":\"rgba(113,113,113,1)\"},\"backgroundColor\":\"none\",\"width\":190,\"height\":39,\"lineHeight\":39,\"textAlign\":\"left\",\"textSpacing\":\"-1.6\",\"enableShadow\":false,\"autoResize\":true,\"v2\":true,\"left\":379,\"top\":130,\"vpos\":\"top\",\"hpos\":\"left\",\"visible\":true,\"group\":\"root\",\"id\":\"box-58\",\"zindex\":8,\"wtype\":\"fixed\",\"htype\":\"fixed\",\"opacity\":1,\"propagateEvents\":false,\"lineHeightAuto\":true,\"rotation\":0,\"scale\":1,\"aspectratio\":false},{\"name\":\"verticalline\",\"type\":\"verticalline\",\"lib\":\"common\",\"caption\":\"Vertical line 3\",\"width\":1,\"height\":229,\"color\":\"A6A6A6\",\"weight\":1,\"left\":341,\"top\":10,\"vpos\":\"top\",\"hpos\":\"left\",\"visible\":true,\"group\":\"root\",\"id\":\"box-52\",\"zindex\":7,\"wtype\":\"fixed\",\"htype\":\"fixed\",\"opacity\":1,\"borderStyle\":\"solid\",\"rotation\":0,\"scale\":1,\"aspectratio\":false},{\"name\":\"verticalline\",\"type\":\"verticalline\",\"lib\":\"common\",\"caption\":\"Vertical line 5\",\"width\":1,\"height\":229,\"color\":\"A6A6A6\",\"weight\":1,\"left\":1064,\"top\":15,\"vpos\":\"top\",\"hpos\":\"left\",\"visible\":true,\"group\":\"root\",\"id\":\"box-54\",\"zindex\":7,\"wtype\":\"fixed\",\"htype\":\"fixed\",\"opacity\":1,\"borderStyle\":\"solid\",\"rotation\":0,\"scale\":1,\"aspectratio\":false},{\"name\":\"verticalline\",\"type\":\"verticalline\",\"lib\":\"common\",\"caption\":\"Vertical line 6\",\"width\":1,\"height\":229,\"color\":\"A6A6A6\",\"weight\":1,\"left\":733,\"top\":10,\"vpos\":\"top\",\"hpos\":\"left\",\"visible\":true,\"group\":\"root\",\"id\":\"box-55\",\"zindex\":7,\"wtype\":\"fixed\",\"htype\":\"fixed\",\"opacity\":1,\"borderStyle\":\"solid\",\"rotation\":0,\"scale\":1,\"aspectratio\":false},{\"name\":\"datetime\",\"type\":\"datetime\",\"lib\":\"common\",\"caption\":\"Date/Time 1\",\"width\":376,\"height\":41,\"backgroundColor\":\"none\",\"borderWidth\":\"1\",\"borderColor\":{\"type\":\"solid\",\"color\":\"rgba(102,150,200,1)\"},\"textFont\":\"Montserrat\",\"textFontStyle\":\"400\",\"textSize\":\"20\",\"textColor\":\"383838\",\"textProperties\":[],\"textAlign\":\"center\",\"dateFormat\":\"yearmonthday\",\"dayFormat\":\"superscript\",\"monthFormat\":\"long\",\"yearFormat\":\"long\",\"timeFormat\":\"24\",\"dateGeneralFormat\":\"timedate\",\"dayNameFormat\":\"long\",\"dateSeparator\":\"-\",\"left\":1064,\"top\":201,\"vpos\":\"top\",\"hpos\":\"left\",\"visible\":true,\"group\":\"root\",\"id\":\"box-50\",\"zindex\":5,\"wtype\":\"fixed\",\"htype\":\"fixed\",\"opacity\":1,\"rotation\":0,\"scale\":1,\"aspectratio\":false},{\"name\":\"text\",\"type\":\"text\",\"lib\":\"common\",\"caption\":\"MRN : 12345679...\",\"text\":\"MRN : 12345679&nbsp;\",\"textFont\":\"Trebuchet MS,sans-serif\",\"textFontStyle\":\"400\",\"textProperties\":[],\"textSize\":\"32\",\"textColor\":{\"type\":\"solid\",\"color\":\"rgba(113,113,113,1)\"},\"backgroundColor\":\"none\",\"width\":218,\"height\":39,\"lineHeight\":39,\"textAlign\":\"left\",\"textSpacing\":\"-1.6\",\"enableShadow\":false,\"autoResize\":true,\"v2\":true,\"left\":76,\"top\":171,\"vpos\":\"top\",\"hpos\":\"left\",\"visible\":true,\"group\":\"root\",\"id\":\"box-48\",\"zindex\":4,\"wtype\":\"fixed\",\"htype\":\"fixed\",\"opacity\":1,\"propagateEvents\":false,\"lineHeightAuto\":true,\"rotation\":0,\"scale\":1,\"aspectratio\":false},{\"name\":\"verticalline\",\"type\":\"verticalline\",\"lib\":\"common\",\"caption\":\"Vertical line 1\",\"width\":1,\"height\":658,\"color\":{\"type\":\"solid\",\"color\":\"rgba(70,127,215,1)\"},\"weight\":1,\"left\":734,\"top\":242,\"vpos\":\"top\",\"hpos\":\"left\",\"visible\":true,\"group\":\"root\",\"id\":\"box-47\",\"zindex\":3,\"wtype\":\"fixed\",\"htype\":\"fixed\",\"opacity\":1,\"borderStyle\":\"solid\",\"rotation\":0,\"scale\":1,\"aspectratio\":false},{\"name\":\"text\",\"type\":\"text\",\"lib\":\"common\",\"caption\":\"John Doe\",\"text\":\"John Doe&nbsp;\",\"textFont\":\"Trebuchet MS,sans-serif\",\"textFontStyle\":\"400\",\"textProperties\":[],\"textSize\":\"64\",\"textColor\":{\"type\":\"solid\",\"color\":\"rgba(117,85,88,1)\"},\"backgroundColor\":\"none\",\"width\":272,\"height\":78,\"lineHeight\":78,\"textAlign\":\"left\",\"textSpacing\":\"-1.6\",\"enableShadow\":false,\"autoResize\":true,\"v2\":true,\"left\":40,\"top\":15,\"vpos\":\"top\",\"hpos\":\"left\",\"visible\":true,\"id\":\"box-46\",\"group\":\"root\",\"zindex\":2,\"wtype\":\"fixed\",\"htype\":\"fixed\",\"opacity\":1,\"propagateEvents\":false,\"lineHeightAuto\":true,\"rotation\":0,\"scale\":1,\"aspectratio\":false},{\"name\":\"basic_slider\",\"type\":\"basic_slider\",\"lib\":\"generic2\",\"caption\":\"Slider 1\",\"width\":347,\"height\":41,\"barThickness\":4,\"barColor\":\"e3e6e8\",\"fillBarColor\":{\"type\":\"solid\",\"color\":\"rgba(255,255,0,1)\"},\"sliderColor\":\"ffffff\",\"barBorderRadius\":2,\"sliderBorderWidth\":1,\"sliderBorderRadius\":\"50%\",\"borderColor\":\"d4d7d9\",\"sliderPosition\":\"70\",\"left\":379,\"top\":359,\"vpos\":\"top\",\"hpos\":\"left\",\"visible\":true,\"id\":\"box-66\",\"group\":\"root\",\"zindex\":9,\"wtype\":\"fixed\",\"htype\":\"fixed\",\"opacity\":1,\"rotation\":0,\"scale\":1,\"aspectratio\":false,\"blurB\":0,\"saturateB\":1,\"brightnessB\":1,\"contrastB\":1,\"grayscaleB\":0,\"sepiaB\":0,\"invertB\":0,\"hueRotateB\":0,\"actions\":[{\"title\":\"Interaction 1\",\"type\":\"sliderdrag\",\"actionId\":\"set-variable\",\"callback\":true,\"variableName\":\"propofol_amount\",\"valueSource\":\"callbackvalue\",\"callbackvariable\":\"value\",\"delay\":\"0\",\"guid\":\"5249fb7c-613a-45ec-8b60-7b50e6c91353\",\"callbackAction\":{\"actionId\":\"change-property\",\"callback\":true,\"pageId\":\"2\",\"itemId\":\"box-88\",\"propertyId\":\"text\",\"valueType\":\"readFromVariable\",\"variableName\":\"propofol_amount\",\"delay\":\"0\",\"guid\":\"782ddea1-581a-4681-bf14-128142baf749\",\"callbackAction\":{\"actionId\":\"change-screen-state\",\"callback\":false,\"pageId\":\"2\",\"stateId\":\"fc0c0693-42f3-489d-adea-e259a6e69adf\",\"delay\":\"0\",\"guid\":\"3d119f38-e48d-424f-82fa-024bee9e8654\",\"clb_disabled\":false},\"clb_disabled\":false},\"sort\":0}]},{\"name\":\"basic_slider\",\"type\":\"basic_slider\",\"lib\":\"generic2\",\"caption\":\"Slider 2\",\"width\":347,\"height\":41,\"barThickness\":4,\"barColor\":\"e3e6e8\",\"fillBarColor\":{\"type\":\"solid\",\"color\":\"rgba(198,198,198,1)\"},\"sliderColor\":\"ffffff\",\"barBorderRadius\":2,\"sliderBorderWidth\":1,\"sliderBorderRadius\":\"50%\",\"borderColor\":\"d4d7d9\",\"sliderPosition\":\"44\",\"left\":373,\"top\":452,\"vpos\":\"top\",\"hpos\":\"left\",\"visible\":true,\"id\":\"box-71\",\"group\":\"root\",\"zindex\":9,\"wtype\":\"fixed\",\"htype\":\"fixed\",\"opacity\":1,\"rotation\":0,\"scale\":1,\"aspectratio\":false,\"actions\":[{\"title\":\"Set Val\",\"type\":\"sliderdrag\",\"actionId\":\"set-variable\",\"callback\":true,\"variableName\":\"lido\",\"valueSource\":\"callbackvalue\",\"callbackvariable\":\"value\",\"delay\":\"0\",\"guid\":\"f46cc9d7-3404-49b7-b2b6-8619e00fdd1b\",\"callbackAction\":{\"actionId\":\"change-property\",\"callback\":true,\"pageId\":\"2\",\"itemId\":\"box-90\",\"propertyId\":\"text\",\"valueType\":\"readFromVariable\",\"variableName\":\"lido\",\"delay\":\"0\",\"guid\":\"3557f991-e23f-4f11-b691-220611bc830c\",\"callbackAction\":{\"actionId\":\"change-screen-state\",\"callback\":false,\"pageId\":\"2\",\"stateId\":\"fc0c0693-42f3-489d-adea-e259a6e69adf\",\"delay\":\"0\",\"guid\":\"9ed71663-0165-4083-ac31-8d7c08459897\",\"clb_disabled\":false},\"clb_disabled\":false},\"sort\":0}]},{\"name\":\"basic_slider\",\"type\":\"basic_slider\",\"lib\":\"generic2\",\"caption\":\"Slider 3\",\"width\":347,\"height\":41,\"barThickness\":4,\"barColor\":\"e3e6e8\",\"fillBarColor\":{\"type\":\"solid\",\"color\":\"rgba(249,51,36,1)\"},\"sliderColor\":\"ffffff\",\"barBorderRadius\":2,\"sliderBorderWidth\":1,\"sliderBorderRadius\":\"50%\",\"borderColor\":\"d4d7d9\",\"sliderPosition\":\"61\",\"left\":379,\"top\":735,\"vpos\":\"top\",\"hpos\":\"left\",\"visible\":true,\"id\":\"box-75\",\"group\":\"root\",\"zindex\":9,\"wtype\":\"fixed\",\"htype\":\"fixed\",\"opacity\":1,\"rotation\":0,\"scale\":1,\"aspectratio\":false,\"actions\":[{\"title\":\"r\",\"type\":\"sliderdrag\",\"actionId\":\"set-variable\",\"callback\":true,\"variableName\":\"rocu\",\"valueSource\":\"callbackvalue\",\"callbackvariable\":\"value\",\"delay\":\"0\",\"guid\":\"62d84234-c58e-4981-95da-e3362d1981e9\",\"callbackAction\":{\"actionId\":\"change-property\",\"callback\":true,\"pageId\":\"2\",\"itemId\":\"box-93\",\"propertyId\":\"text\",\"valueType\":\"readFromVariable\",\"variableName\":\"rocu\",\"delay\":\"0\",\"guid\":\"5015c9b0-b173-421d-b7aa-c1f449381924\",\"callbackAction\":{\"actionId\":\"change-screen-state\",\"callback\":false,\"pageId\":\"2\",\"stateId\":\"fc0c0693-42f3-489d-adea-e259a6e69adf\",\"delay\":\"0\",\"guid\":\"73b66e38-8248-4dc7-9302-f107a378fe46\",\"clb_disabled\":false},\"clb_disabled\":false},\"sort\":0}]},{\"name\":\"basic_slider\",\"type\":\"basic_slider\",\"lib\":\"generic2\",\"caption\":\"Slider 4\",\"width\":347,\"height\":41,\"barThickness\":4,\"barColor\":\"e3e6e8\",\"fillBarColor\":{\"type\":\"solid\",\"color\":\"rgba(160,58,207,1)\"},\"sliderColor\":\"ffffff\",\"barBorderRadius\":2,\"sliderBorderWidth\":1,\"sliderBorderRadius\":\"50%\",\"borderColor\":\"d4d7d9\",\"sliderPosition\":\"40\",\"left\":379,\"top\":648,\"vpos\":\"top\",\"hpos\":\"left\",\"visible\":true,\"id\":\"box-76\",\"group\":\"root\",\"zindex\":9,\"wtype\":\"fixed\",\"htype\":\"fixed\",\"opacity\":1,\"rotation\":0,\"scale\":1,\"aspectratio\":false,\"actions\":[{\"title\":\"frt\",\"type\":\"sliderdrag\",\"actionId\":\"set-variable\",\"callback\":true,\"variableName\":\"phen\",\"valueSource\":\"callbackvalue\",\"callbackvariable\":\"value\",\"delay\":\"0\",\"guid\":\"6e9ee942-8284-4e8b-aeb5-0c83cfbddec8\",\"callbackAction\":{\"actionId\":\"change-property\",\"callback\":true,\"pageId\":\"2\",\"itemId\":\"box-92\",\"propertyId\":\"text\",\"valueType\":\"readFromVariable\",\"variableName\":\"phen\",\"delay\":\"0\",\"guid\":\"bed0b053-6866-4a3e-b93b-7b8073b8872c\",\"callbackAction\":{\"actionId\":\"change-screen-state\",\"callback\":false,\"pageId\":\"2\",\"stateId\":\"fc0c0693-42f3-489d-adea-e259a6e69adf\",\"delay\":\"0\",\"guid\":\"6f0206e6-2c48-460d-9bfb-7e3786908f60\",\"clb_disabled\":false},\"clb_disabled\":false},\"sort\":0}]},{\"name\":\"basic_slider\",\"type\":\"basic_slider\",\"lib\":\"generic2\",\"caption\":\"Slider 5\",\"width\":347,\"height\":41,\"barThickness\":4,\"barColor\":\"e3e6e8\",\"fillBarColor\":{\"type\":\"solid\",\"color\":\"rgba(102,150,200,1)\"},\"sliderColor\":\"ffffff\",\"barBorderRadius\":2,\"sliderBorderWidth\":1,\"sliderBorderRadius\":\"50%\",\"borderColor\":\"d4d7d9\",\"sliderPosition\":\"33\",\"left\":379,\"top\":550,\"vpos\":\"top\",\"hpos\":\"left\",\"visible\":true,\"id\":\"box-77\",\"group\":\"root\",\"zindex\":9,\"wtype\":\"fixed\",\"htype\":\"fixed\",\"opacity\":1,\"rotation\":0,\"scale\":1,\"aspectratio\":false,\"actions\":[{\"title\":\"f\",\"type\":\"sliderdrag\",\"actionId\":\"set-variable\",\"callback\":true,\"variableName\":\"fent\",\"valueSource\":\"callbackvalue\",\"callbackvariable\":\"value\",\"delay\":\"0\",\"guid\":\"6129fa4f-987d-406e-8895-4537b3b72529\",\"callbackAction\":{\"actionId\":\"change-property\",\"callback\":true,\"pageId\":\"2\",\"itemId\":\"box-91\",\"propertyId\":\"text\",\"valueType\":\"readFromVariable\",\"variableName\":\"fent\",\"delay\":\"0\",\"guid\":\"69d8ad42-0ccf-4223-abdb-8fb8d4beac0a\",\"callbackAction\":{\"actionId\":\"change-screen-state\",\"callback\":false,\"pageId\":\"2\",\"stateId\":\"fc0c0693-42f3-489d-adea-e259a6e69adf\",\"delay\":\"0\",\"guid\":\"48916d8c-69ff-4c80-9615-26a98286603b\",\"clb_disabled\":false},\"clb_disabled\":false},\"sort\":0}]},{\"name\":\"basic_textfield\",\"type\":\"basic_textfield\",\"lib\":\"generic2\",\"caption\":\"Text Field prop\",\"width\":338,\"height\":55,\"value\":\"Propofol  ( mg ) \",\"placeholder\":\"Placeholder\",\"inputtype\":\"text\",\"backgroundColor\":{\"type\":\"solid\",\"color\":\"rgba(255,255,122,1)\"},\"textFont\":\"Arial,sans-serif\",\"textFontStyle\":\"400\",\"textSize\":\"28\",\"textColor\":{\"type\":\"solid\",\"color\":\"rgba(56,56,56,1)\"},\"textProperties\":[],\"placeholderColor\":\"c1c8c9\",\"borderWidth\":1,\"borderStyle\":\"solid\",\"borderRadius\":4,\"borderColor\":{\"type\":\"solid\",\"color\":\"rgba(255,255,0,1)\"},\"textAlign\":\"center\",\"left\":22,\"top\":359,\"vpos\":\"top\",\"hpos\":\"left\",\"visible\":true,\"group\":\"root\",\"id\":\"box-67\",\"zindex\":10,\"wtype\":\"fixed\",\"htype\":\"fixed\",\"opacity\":1,\"rotation\":0,\"scale\":1,\"aspectratio\":false,\"userCaption\":true},{\"name\":\"basic_textfield\",\"type\":\"basic_textfield\",\"lib\":\"generic2\",\"caption\":\"Text Field lido\",\"width\":338,\"height\":55,\"value\":\"Lidocaine  (mg)\",\"placeholder\":\"Placeholder\",\"inputtype\":\"text\",\"backgroundColor\":{\"type\":\"solid\",\"color\":\"rgba(198,198,198,1)\"},\"textFont\":\"Arial,sans-serif\",\"textFontStyle\":\"400\",\"textSize\":\"28\",\"textColor\":{\"type\":\"solid\",\"color\":\"rgba(56,56,56,1)\"},\"textProperties\":[],\"placeholderColor\":\"c1c8c9\",\"borderWidth\":1,\"borderStyle\":\"solid\",\"borderRadius\":4,\"borderColor\":{\"type\":\"solid\",\"color\":\"rgba(208,212,0,1)\"},\"textAlign\":\"center\",\"left\":22,\"top\":445,\"vpos\":\"top\",\"hpos\":\"left\",\"visible\":true,\"group\":\"root\",\"id\":\"box-72\",\"zindex\":10,\"wtype\":\"fixed\",\"htype\":\"fixed\",\"opacity\":1,\"rotation\":0,\"scale\":1,\"aspectratio\":false,\"userCaption\":true},{\"name\":\"basic_textfield\",\"type\":\"basic_textfield\",\"lib\":\"generic2\",\"caption\":\"Text Field Phenylephrine\",\"width\":338,\"height\":55,\"value\":\"Phenylephrine (mcg) \",\"placeholder\":\"Placeholder\",\"inputtype\":\"text\",\"backgroundColor\":{\"type\":\"solid\",\"color\":\"rgba(234,114,252,1)\"},\"textFont\":\"Arial,sans-serif\",\"textFontStyle\":\"400\",\"textSize\":\"28\",\"textColor\":{\"type\":\"solid\",\"color\":\"rgba(56,56,56,1)\"},\"textProperties\":[],\"placeholderColor\":\"c1c8c9\",\"borderWidth\":1,\"borderStyle\":\"solid\",\"borderRadius\":4,\"borderColor\":{\"type\":\"solid\",\"color\":\"rgba(255,255,0,1)\"},\"textAlign\":\"center\",\"left\":22,\"top\":641,\"vpos\":\"top\",\"hpos\":\"left\",\"visible\":true,\"group\":\"root\",\"id\":\"box-73\",\"zindex\":10,\"wtype\":\"fixed\",\"htype\":\"fixed\",\"opacity\":1,\"rotation\":0,\"scale\":1,\"aspectratio\":false,\"userCaption\":true},{\"name\":\"basic_textfield\",\"type\":\"basic_textfield\",\"lib\":\"generic2\",\"caption\":\"Text Field rocur\",\"width\":338,\"height\":55,\"value\":\"Rocuronium (mg)\",\"placeholder\":\"Placeholder\",\"inputtype\":\"text\",\"backgroundColor\":{\"type\":\"solid\",\"color\":\"rgba(226,15,0,1)\"},\"textFont\":\"Arial,sans-serif\",\"textFontStyle\":\"400\",\"textSize\":\"28\",\"textColor\":{\"type\":\"solid\",\"color\":\"rgba(56,56,56,1)\"},\"textProperties\":[],\"placeholderColor\":\"c1c8c9\",\"borderWidth\":1,\"borderStyle\":\"solid\",\"borderRadius\":4,\"borderColor\":{\"type\":\"solid\",\"color\":\"rgba(255,255,0,1)\"},\"textAlign\":\"center\",\"left\":22,\"top\":721,\"vpos\":\"top\",\"hpos\":\"left\",\"visible\":true,\"group\":\"root\",\"id\":\"box-79\",\"zindex\":10,\"wtype\":\"fixed\",\"htype\":\"fixed\",\"opacity\":1,\"rotation\":0,\"scale\":1,\"aspectratio\":false,\"userCaption\":true},{\"name\":\"basic_textfield\",\"type\":\"basic_textfield\",\"lib\":\"generic2\",\"caption\":\"Text Field 2\",\"width\":733,\"height\":77,\"value\":\"Induction Medication\",\"placeholder\":\"Placeholder\",\"inputtype\":\"text\",\"backgroundColor\":{\"type\":\"solid\",\"color\":\"rgba(177,190,227,1)\"},\"textFont\":\"Arial,sans-serif\",\"textFontStyle\":\"400\",\"textSize\":\"64\",\"textColor\":{\"type\":\"solid\",\"color\":\"rgba(233,240,215,1)\"},\"textProperties\":[],\"placeholderColor\":\"c1c8c9\",\"borderWidth\":\"5\",\"borderStyle\":\"solid\",\"borderRadius\":4,\"borderColor\":{\"type\":\"solid\",\"color\":\"rgba(85,85,85,1)\"},\"textAlign\":\"center\",\"left\":2,\"top\":242,\"vpos\":\"top\",\"hpos\":\"left\",\"visible\":true,\"id\":\"box-69\",\"group\":\"root\",\"zindex\":11,\"wtype\":\"fixed\",\"htype\":\"fixed\",\"opacity\":1,\"rotation\":0,\"scale\":1,\"aspectratio\":false},{\"name\":\"basic_textfield\",\"type\":\"basic_textfield\",\"lib\":\"generic2\",\"caption\":\"Text Field 3\",\"width\":698,\"height\":77,\"value\":\"Blood Pressure\",\"placeholder\":\"Placeholder\",\"inputtype\":\"text\",\"backgroundColor\":{\"type\":\"solid\",\"color\":\"rgba(177,190,227,1)\"},\"textFont\":\"Arial,sans-serif\",\"textFontStyle\":\"400\",\"textSize\":\"64\",\"textColor\":{\"type\":\"solid\",\"color\":\"rgba(233,240,215,1)\"},\"textProperties\":[],\"placeholderColor\":\"c1c8c9\",\"borderWidth\":\"5\",\"borderStyle\":\"solid\",\"borderRadius\":4,\"borderColor\":{\"type\":\"solid\",\"color\":\"rgba(85,85,85,1)\"},\"textAlign\":\"center\",\"left\":733,\"top\":244,\"vpos\":\"top\",\"hpos\":\"left\",\"visible\":true,\"id\":\"box-70\",\"group\":\"root\",\"zindex\":11,\"wtype\":\"fixed\",\"htype\":\"fixed\",\"opacity\":1,\"rotation\":0,\"scale\":1,\"aspectratio\":false},{\"name\":\"basic_textfield\",\"type\":\"basic_textfield\",\"lib\":\"generic2\",\"caption\":\"Text Field fenta\",\"width\":338,\"height\":55,\"value\":\"Fentanyl (mcg)\",\"placeholder\":\"Placeholder\",\"inputtype\":\"text\",\"backgroundColor\":{\"type\":\"solid\",\"color\":\"rgba(101,180,206,1)\"},\"textFont\":\"Arial,sans-serif\",\"textFontStyle\":\"400\",\"textSize\":\"28\",\"textColor\":{\"type\":\"solid\",\"color\":\"rgba(56,56,56,1)\"},\"textProperties\":[],\"placeholderColor\":\"c1c8c9\",\"borderWidth\":1,\"borderStyle\":\"solid\",\"borderRadius\":4,\"borderColor\":{\"type\":\"solid\",\"color\":\"rgba(27,77,87,1)\"},\"textAlign\":\"center\",\"left\":22,\"top\":543,\"vpos\":\"top\",\"hpos\":\"left\",\"visible\":true,\"group\":\"root\",\"id\":\"box-78\",\"zindex\":10,\"wtype\":\"fixed\",\"htype\":\"fixed\",\"opacity\":1,\"rotation\":0,\"scale\":1,\"aspectratio\":false,\"userCaption\":true},{\"name\":\"basic_button_text\",\"type\":\"basic_button_text\",\"lib\":\"generic2\",\"caption\":\"Button with Background 1\",\"width\":177,\"height\":80,\"backgroundColor\":{\"type\":\"solid\",\"color\":\"rgba(102,150,200,1)\"},\"borderWidth\":0,\"borderColor\":\"#025abf\",\"borderRadius\":4,\"text\":\"Other Opioids\",\"textFont\":\"Arial,sans-serif\",\"textFontStyle\":\"400\",\"textSize\":\"28\",\"textColor\":\"FFFFFF\",\"textProperties\":[],\"textAlign\":\"center\",\"iconpos\":\"none\",\"left\":32,\"top\":800,\"vpos\":\"top\",\"hpos\":\"left\",\"visible\":true,\"id\":\"box-81\",\"group\":\"root\",\"zindex\":13,\"wtype\":\"fixed\",\"htype\":\"fixed\",\"opacity\":1,\"rotation\":0,\"scale\":1,\"aspectratio\":false,\"actions\":[{\"title\":\"Interaction 1\",\"type\":\"tap\",\"actionId\":\"go-to-page\",\"callback\":false,\"pageId\":\"4\",\"animation\":\"fade\",\"animationEasing\":\"Cubic.easeOut\",\"duration\":\"400\",\"delay\":\"0\",\"guid\":\"d7282a4d-f692-4031-87ae-61a83b77dbeb\",\"sort\":0}]},{\"name\":\"basic_button_text\",\"type\":\"basic_button_text\",\"lib\":\"generic2\",\"caption\":\"Button with Background 3\",\"width\":177,\"height\":80,\"backgroundColor\":{\"type\":\"solid\",\"color\":\"rgba(255, 255, 0,1)\"},\"borderWidth\":0,\"borderColor\":\"#025abf\",\"borderRadius\":4,\"text\":\"Other Amestics\",\"textFont\":\"Arial,sans-serif\",\"textFontStyle\":\"400\",\"textSize\":\"28\",\"textColor\":{\"type\":\"solid\",\"color\":\"rgba(56,56,56,1)\"},\"textProperties\":[],\"textAlign\":\"center\",\"iconpos\":\"none\",\"left\":252,\"top\":800,\"vpos\":\"top\",\"hpos\":\"left\",\"visible\":true,\"id\":\"box-83\",\"group\":\"root\",\"zindex\":13,\"wtype\":\"fixed\",\"htype\":\"fixed\",\"opacity\":1,\"rotation\":0,\"scale\":1,\"aspectratio\":false,\"actions\":[{\"title\":\"Interaction 1\",\"type\":\"tap\",\"actionId\":\"go-to-page\",\"callback\":false,\"pageId\":\"7\",\"animation\":\"fade\",\"animationEasing\":\"Cubic.easeOut\",\"duration\":\"400\",\"delay\":\"0\",\"guid\":\"720a7796-198f-4f33-b462-ebdba6f6a88d\",\"sort\":0}]},{\"name\":\"basic_button_text\",\"type\":\"basic_button_text\",\"lib\":\"generic2\",\"caption\":\"Button with Background 4\",\"width\":177,\"height\":80,\"backgroundColor\":{\"type\":\"solid\",\"color\":\"rgba(234,114,252,1)\"},\"borderWidth\":0,\"borderColor\":\"#025abf\",\"borderRadius\":4,\"text\":\"Other Pressors\",\"textFont\":\"Arial,sans-serif\",\"textFontStyle\":\"400\",\"textSize\":\"28\",\"textColor\":\"FFFFFF\",\"textProperties\":[],\"textAlign\":\"center\",\"iconpos\":\"none\",\"left\":489,\"top\":800,\"vpos\":\"top\",\"hpos\":\"left\",\"visible\":true,\"id\":\"box-84\",\"group\":\"root\",\"zindex\":13,\"wtype\":\"fixed\",\"htype\":\"fixed\",\"opacity\":1,\"rotation\":0,\"scale\":1,\"aspectratio\":false,\"actions\":[{\"title\":\"Interaction 1\",\"type\":\"tap\",\"actionId\":\"go-to-page\",\"callback\":false,\"pageId\":\"6\",\"animation\":\"fade\",\"animationEasing\":\"Cubic.easeOut\",\"duration\":\"400\",\"delay\":\"0\",\"guid\":\"575ea204-ba00-4afa-a79d-82740e37e8a3\",\"sort\":0}]},{\"name\":\"basic_button_text\",\"type\":\"basic_button_text\",\"lib\":\"generic2\",\"caption\":\"Button with Background 2\",\"width\":177,\"height\":80,\"backgroundColor\":{\"type\":\"solid\",\"color\":\"rgba(249,51,36,1)\"},\"borderWidth\":0,\"borderColor\":\"#025abf\",\"borderRadius\":4,\"text\":\"Exit\",\"textFont\":\"Arial,sans-serif\",\"textFontStyle\":\"400\",\"textSize\":\"36\",\"textColor\":\"FFFFFF\",\"textProperties\":[],\"textAlign\":\"center\",\"iconpos\":\"none\",\"left\":1233,\"top\":789,\"vpos\":\"top\",\"hpos\":\"left\",\"visible\":true,\"id\":\"box-82\",\"group\":\"root\",\"zindex\":13,\"wtype\":\"fixed\",\"htype\":\"fixed\",\"opacity\":1,\"rotation\":0,\"scale\":1,\"aspectratio\":false,\"actions\":[{\"title\":\"Interaction 1\",\"type\":\"click\",\"actionId\":\"go-to-page\",\"callback\":false,\"pageId\":\"1\",\"animation\":\"fade\",\"animationEasing\":\"Cubic.easeOut\",\"duration\":\"400\",\"delay\":\"0\",\"guid\":\"ba1c28d8-6e98-4cdb-b7e6-46e16b0d28db\",\"sort\":0}]},{\"name\":\"text\",\"type\":\"text\",\"lib\":\"common\",\"caption\":\"text\",\"text\":\"150\",\"textFont\":\"Arial,sans-serif\",\"textFontStyle\":\"400\",\"textProperties\":[],\"textSize\":\"24\",\"textColor\":{\"type\":\"solid\",\"color\":\"rgba(255,255,0,1)\"},\"backgroundColor\":\"none\",\"width\":275,\"height\":29,\"lineHeight\":29,\"textAlign\":\"left\",\"textSpacing\":0,\"enableShadow\":false,\"autoResize\":false,\"v2\":true,\"left\":505,\"top\":330,\"vpos\":\"top\",\"hpos\":\"left\",\"visible\":true,\"id\":\"box-88\",\"group\":\"root\",\"zindex\":14,\"wtype\":\"fixed\",\"htype\":\"fixed\",\"opacity\":1,\"propagateEvents\":false,\"lineHeightAuto\":true,\"rotation\":0,\"scale\":1,\"aspectratio\":false,\"userCaption\":true,\"actions\":[]},{\"name\":\"text\",\"type\":\"text\",\"lib\":\"common\",\"caption\":\"text 2\",\"text\":\"100\",\"textFont\":\"Arial,sans-serif\",\"textFontStyle\":\"400\",\"textProperties\":[],\"textSize\":\"24\",\"textColor\":{\"type\":\"solid\",\"color\":\"rgba(198,198,198,1)\"},\"backgroundColor\":\"none\",\"width\":275,\"height\":29,\"lineHeight\":29,\"textAlign\":\"left\",\"textSpacing\":0,\"enableShadow\":false,\"autoResize\":false,\"v2\":true,\"left\":505,\"top\":414,\"vpos\":\"top\",\"hpos\":\"left\",\"visible\":true,\"id\":\"box-90\",\"group\":\"root\",\"zindex\":14,\"wtype\":\"fixed\",\"htype\":\"fixed\",\"opacity\":1,\"propagateEvents\":false,\"lineHeightAuto\":true,\"rotation\":0,\"scale\":1,\"aspectratio\":false,\"userCaption\":true,\"actions\":[]},{\"name\":\"text\",\"type\":\"text\",\"lib\":\"common\",\"caption\":\"text 3\",\"text\":\"100\",\"textFont\":\"Arial,sans-serif\",\"textFontStyle\":\"400\",\"textProperties\":[],\"textSize\":\"24\",\"textColor\":{\"type\":\"solid\",\"color\":\"rgba(43,89,162,1)\"},\"backgroundColor\":\"none\",\"width\":275,\"height\":29,\"lineHeight\":29,\"textAlign\":\"left\",\"textSpacing\":0,\"enableShadow\":false,\"autoResize\":false,\"v2\":true,\"left\":505,\"top\":521,\"vpos\":\"top\",\"hpos\":\"left\",\"visible\":true,\"id\":\"box-91\",\"group\":\"root\",\"zindex\":14,\"wtype\":\"fixed\",\"htype\":\"fixed\",\"opacity\":1,\"propagateEvents\":false,\"lineHeightAuto\":true,\"rotation\":0,\"scale\":1,\"aspectratio\":false,\"userCaption\":true,\"actions\":[]},{\"name\":\"text\",\"type\":\"text\",\"lib\":\"common\",\"caption\":\"text 4\",\"text\":\"100\",\"textFont\":\"Arial,sans-serif\",\"textFontStyle\":\"400\",\"textProperties\":[],\"textSize\":\"24\",\"textColor\":{\"type\":\"solid\",\"color\":\"rgba(183,47,214,1)\"},\"backgroundColor\":\"none\",\"width\":275,\"height\":29,\"lineHeight\":29,\"textAlign\":\"left\",\"textSpacing\":0,\"enableShadow\":false,\"autoResize\":false,\"v2\":true,\"left\":497,\"top\":612,\"vpos\":\"top\",\"hpos\":\"left\",\"visible\":true,\"id\":\"box-92\",\"group\":\"root\",\"zindex\":14,\"wtype\":\"fixed\",\"htype\":\"fixed\",\"opacity\":1,\"propagateEvents\":false,\"lineHeightAuto\":true,\"rotation\":0,\"scale\":1,\"aspectratio\":false,\"userCaption\":true,\"actions\":[]},{\"name\":\"text\",\"type\":\"text\",\"lib\":\"common\",\"caption\":\"text 5\",\"text\":\"100\",\"textFont\":\"Arial,sans-serif\",\"textFontStyle\":\"400\",\"textProperties\":[],\"textSize\":\"24\",\"textColor\":{\"type\":\"solid\",\"color\":\"rgba(209,25,105,1)\"},\"backgroundColor\":\"none\",\"width\":275,\"height\":29,\"lineHeight\":29,\"textAlign\":\"left\",\"textSpacing\":0,\"enableShadow\":false,\"autoResize\":false,\"v2\":true,\"left\":491,\"top\":712,\"vpos\":\"top\",\"hpos\":\"left\",\"visible\":true,\"id\":\"box-93\",\"group\":\"root\",\"zindex\":14,\"wtype\":\"fixed\",\"htype\":\"fixed\",\"opacity\":1,\"propagateEvents\":false,\"lineHeightAuto\":true,\"rotation\":0,\"scale\":1,\"aspectratio\":false,\"userCaption\":true,\"actions\":[]},{\"name\":\"image\",\"type\":\"image\",\"lib\":\"common\",\"caption\":\"ss3\",\"imgSrc\":{\"fileId\":\"7902aa96fae82cf51263d633901fa229.png\",\"assetType\":\"project\",\"bucketsource\":\"main\",\"name\":\"Screen Shot 2022-12-12 at 9.52.17 PM.png\"},\"repeat\":false,\"width\":665.8125,\"height\":402,\"borderWidth\":0,\"borderRadius\":0,\"borderColor\":\"#D1D1D1\",\"borderPos\":\"inside\",\"propagateEvents\":false,\"actions\":[],\"overlay\":false,\"autoResize\":false,\"aspectratio\":1.65625,\"assistiveDescription\":\"\",\"left\":750,\"top\":357,\"visible\":true,\"id\":\"box-216\",\"group\":\"root\",\"zindex\":15,\"hpos\":\"left\",\"vpos\":\"top\",\"wtype\":\"fixed\",\"htype\":\"fixed\",\"opacity\":1,\"borderStyle\":\"solid\",\"rotation\":0,\"scale\":1,\"userCaption\":true,\"hasMask\":false}]","template":{"id":"","stateid":""},"history":[],"id":"576fe87f-7989-4c31-93a7-292d92d03cfb","actions":[],"groups":{}},{"title":"State 4","background":{"type":"solid","color":"rgba(169,151,196,1)"},"data":"[{\"name\":\"rectangle\",\"type\":\"rectangle\",\"lib\":\"common\",\"caption\":\"Rectangle 1\",\"backgroundColor\":{\"type\":\"solid\",\"color\":\"rgba(193,212,241,1)\"},\"borderWidth\":\"8\",\"borderColor\":{\"type\":\"solid\",\"color\":\"rgba(30,77,130,1)\"},\"borderRadius\":0,\"imgSrc\":{\"fileId\":\"\",\"name\":\"\",\"assetType\":\"\",\"bucketsource\":\"\",\"url\":\"\"},\"backgroundSizeType\":\"cover\",\"backgroundSizeValue\":35,\"width\":1440,\"height\":249,\"lineHeight\":20,\"lineHeightAuto\":true,\"actions\":[],\"textFont\":\"Arial,sans-serif\",\"textFontStyle\":\"400\",\"textSize\":16,\"textColor\":\"#383838\",\"textProperties\":[],\"textSpacing\":0,\"textAlign\":\"center\",\"text\":\"\",\"typeName\":\"rectangle\",\"left\":0,\"top\":0,\"vpos\":\"top\",\"hpos\":\"left\",\"visible\":true,\"id\":\"box-45\",\"group\":\"root\",\"zindex\":1,\"wtype\":\"fixed\",\"htype\":\"fixed\",\"opacity\":1,\"borderStyle\":\"solid\",\"rotation\":0,\"scale\":1,\"aspectratio\":false},{\"name\":\"text\",\"type\":\"text\",\"lib\":\"common\",\"caption\":\"Gender : Male...\",\"text\":\"Gender : Male\",\"textFont\":\"Trebuchet MS,sans-serif\",\"textFontStyle\":\"400\",\"textProperties\":[],\"textSize\":\"32\",\"textColor\":{\"type\":\"solid\",\"color\":\"rgba(113,113,113,1)\"},\"backgroundColor\":\"none\",\"width\":186,\"height\":39,\"lineHeight\":39,\"textAlign\":\"left\",\"textSpacing\":\"-1.6\",\"enableShadow\":false,\"autoResize\":true,\"v2\":true,\"left\":379,\"top\":47,\"vpos\":\"top\",\"hpos\":\"left\",\"visible\":true,\"group\":\"root\",\"id\":\"box-64\",\"zindex\":8,\"wtype\":\"fixed\",\"htype\":\"fixed\",\"opacity\":1,\"propagateEvents\":false,\"lineHeightAuto\":true,\"rotation\":0,\"scale\":1,\"aspectratio\":false},{\"name\":\"text\",\"type\":\"text\",\"lib\":\"common\",\"caption\":\"DOB: 01/01/2022...\",\"text\":\"DOB: 01/01/2022\",\"textFont\":\"Trebuchet MS,sans-serif\",\"textFontStyle\":\"400\",\"textProperties\":[],\"textSize\":\"32\",\"textColor\":{\"type\":\"solid\",\"color\":\"rgba(113,113,113,1)\"},\"backgroundColor\":\"none\",\"width\":230,\"height\":39,\"lineHeight\":39,\"textAlign\":\"left\",\"textSpacing\":\"-1.6\",\"enableShadow\":false,\"autoResize\":true,\"v2\":true,\"left\":76,\"top\":130,\"vpos\":\"top\",\"hpos\":\"left\",\"visible\":true,\"group\":\"root\",\"id\":\"box-61\",\"zindex\":8,\"wtype\":\"fixed\",\"htype\":\"fixed\",\"opacity\":1,\"propagateEvents\":false,\"lineHeightAuto\":true,\"rotation\":0,\"scale\":1,\"aspectratio\":false},{\"name\":\"text\",\"type\":\"text\",\"lib\":\"common\",\"caption\":\"Height : 174 cm...\",\"text\":\"Height : 174 cm&nbsp;\",\"textFont\":\"Trebuchet MS,sans-serif\",\"textFontStyle\":\"400\",\"textProperties\":[],\"textSize\":\"32\",\"textColor\":{\"type\":\"solid\",\"color\":\"rgba(113,113,113,1)\"},\"backgroundColor\":\"none\",\"width\":216,\"height\":39,\"lineHeight\":39,\"textAlign\":\"left\",\"textSpacing\":\"-1.6\",\"enableShadow\":false,\"autoResize\":true,\"v2\":true,\"left\":379,\"top\":86,\"vpos\":\"top\",\"hpos\":\"left\",\"visible\":true,\"group\":\"root\",\"id\":\"box-62\",\"zindex\":8,\"wtype\":\"fixed\",\"htype\":\"fixed\",\"opacity\":1,\"propagateEvents\":false,\"lineHeightAuto\":true,\"rotation\":0,\"scale\":1,\"aspectratio\":false},{\"name\":\"text\",\"type\":\"text\",\"lib\":\"common\",\"caption\":\"Surgery:\\nTotal Colec...\",\"text\":\"Surgery:&nbsp;<br>Total Colectomy with&nbsp;<br>pancreatico-jejunostomy&nbsp;\",\"textFont\":\"Trebuchet MS,sans-serif\",\"textFontStyle\":\"400\",\"textProperties\":[],\"textSize\":\"32\",\"textColor\":{\"type\":\"solid\",\"color\":\"rgba(113,113,113,1)\"},\"backgroundColor\":\"none\",\"width\":336,\"height\":117,\"lineHeight\":39,\"textAlign\":\"left\",\"textSpacing\":\"-1.6\",\"enableShadow\":false,\"autoResize\":true,\"v2\":true,\"left\":1084,\"top\":47,\"vpos\":\"top\",\"hpos\":\"left\",\"visible\":true,\"group\":\"root\",\"id\":\"box-63\",\"zindex\":8,\"wtype\":\"fixed\",\"htype\":\"fixed\",\"opacity\":1,\"propagateEvents\":false,\"lineHeightAuto\":true,\"rotation\":0,\"scale\":1,\"aspectratio\":false},{\"name\":\"text\",\"type\":\"text\",\"lib\":\"common\",\"caption\":\"Age : 86 y\",\"text\":\"Age : 86 y&nbsp;\",\"textFont\":\"Trebuchet MS,sans-serif\",\"textFontStyle\":\"400\",\"textProperties\":[],\"textSize\":\"32\",\"textColor\":{\"type\":\"solid\",\"color\":\"rgba(113,113,113,1)\"},\"backgroundColor\":\"none\",\"width\":139,\"height\":39,\"lineHeight\":39,\"textAlign\":\"left\",\"textSpacing\":\"-1.6\",\"enableShadow\":false,\"autoResize\":true,\"v2\":true,\"left\":76,\"top\":91,\"vpos\":\"top\",\"hpos\":\"left\",\"visible\":true,\"group\":\"root\",\"id\":\"box-60\",\"zindex\":8,\"wtype\":\"fixed\",\"htype\":\"fixed\",\"opacity\":1,\"propagateEvents\":false,\"lineHeightAuto\":true,\"rotation\":0,\"scale\":1,\"aspectratio\":false},{\"name\":\"text\",\"type\":\"text\",\"lib\":\"common\",\"caption\":\"Allergies :\\nPenicill...\",\"text\":\"Allergies :<br><span style=\\\"color: rgba(249,51,36,1);\\\">Penicillin</span><br><br>\",\"textFont\":\"Trebuchet MS,sans-serif\",\"textFontStyle\":\"400\",\"textProperties\":[],\"textSize\":\"32\",\"textColor\":{\"type\":\"solid\",\"color\":\"rgba(113,113,113,1)\"},\"backgroundColor\":\"none\",\"width\":203,\"height\":117,\"lineHeight\":39,\"textAlign\":\"left\",\"textSpacing\":\"-1.6\",\"enableShadow\":false,\"autoResize\":false,\"v2\":true,\"left\":766,\"top\":47,\"vpos\":\"top\",\"hpos\":\"left\",\"visible\":true,\"group\":\"root\",\"id\":\"box-59\",\"zindex\":8,\"wtype\":\"fixed\",\"htype\":\"fixed\",\"opacity\":1,\"propagateEvents\":false,\"lineHeightAuto\":true,\"rotation\":0,\"scale\":1,\"aspectratio\":false},{\"name\":\"text\",\"type\":\"text\",\"lib\":\"common\",\"caption\":\"BMI : 28 kg/m2...\",\"text\":\"BMI : 28 kg/m2\",\"textFont\":\"Trebuchet MS,sans-serif\",\"textFontStyle\":\"400\",\"textProperties\":[],\"textSize\":\"32\",\"textColor\":{\"type\":\"solid\",\"color\":\"rgba(113,113,113,1)\"},\"backgroundColor\":\"none\",\"width\":199,\"height\":39,\"lineHeight\":39,\"textAlign\":\"left\",\"textSpacing\":\"-1.6\",\"enableShadow\":false,\"autoResize\":true,\"v2\":true,\"left\":379,\"top\":171,\"vpos\":\"top\",\"hpos\":\"left\",\"visible\":true,\"group\":\"root\",\"id\":\"box-56\",\"zindex\":8,\"wtype\":\"fixed\",\"htype\":\"fixed\",\"opacity\":1,\"propagateEvents\":false,\"lineHeightAuto\":true,\"rotation\":0,\"scale\":1,\"aspectratio\":false},{\"name\":\"text\",\"type\":\"text\",\"lib\":\"common\",\"caption\":\"Weight : 69 Kg...\",\"text\":\"Weight : 69 Kg\",\"textFont\":\"Trebuchet MS,sans-serif\",\"textFontStyle\":\"400\",\"textProperties\":[],\"textSize\":\"32\",\"textColor\":{\"type\":\"solid\",\"color\":\"rgba(113,113,113,1)\"},\"backgroundColor\":\"none\",\"width\":190,\"height\":39,\"lineHeight\":39,\"textAlign\":\"left\",\"textSpacing\":\"-1.6\",\"enableShadow\":false,\"autoResize\":true,\"v2\":true,\"left\":379,\"top\":130,\"vpos\":\"top\",\"hpos\":\"left\",\"visible\":true,\"group\":\"root\",\"id\":\"box-58\",\"zindex\":8,\"wtype\":\"fixed\",\"htype\":\"fixed\",\"opacity\":1,\"propagateEvents\":false,\"lineHeightAuto\":true,\"rotation\":0,\"scale\":1,\"aspectratio\":false},{\"name\":\"verticalline\",\"type\":\"verticalline\",\"lib\":\"common\",\"caption\":\"Vertical line 3\",\"width\":1,\"height\":229,\"color\":\"A6A6A6\",\"weight\":1,\"left\":341,\"top\":10,\"vpos\":\"top\",\"hpos\":\"left\",\"visible\":true,\"group\":\"root\",\"id\":\"box-52\",\"zindex\":7,\"wtype\":\"fixed\",\"htype\":\"fixed\",\"opacity\":1,\"borderStyle\":\"solid\",\"rotation\":0,\"scale\":1,\"aspectratio\":false},{\"name\":\"verticalline\",\"type\":\"verticalline\",\"lib\":\"common\",\"caption\":\"Vertical line 5\",\"width\":1,\"height\":229,\"color\":\"A6A6A6\",\"weight\":1,\"left\":1064,\"top\":15,\"vpos\":\"top\",\"hpos\":\"left\",\"visible\":true,\"group\":\"root\",\"id\":\"box-54\",\"zindex\":7,\"wtype\":\"fixed\",\"htype\":\"fixed\",\"opacity\":1,\"borderStyle\":\"solid\",\"rotation\":0,\"scale\":1,\"aspectratio\":false},{\"name\":\"verticalline\",\"type\":\"verticalline\",\"lib\":\"common\",\"caption\":\"Vertical line 6\",\"width\":1,\"height\":229,\"color\":\"A6A6A6\",\"weight\":1,\"left\":733,\"top\":10,\"vpos\":\"top\",\"hpos\":\"left\",\"visible\":true,\"group\":\"root\",\"id\":\"box-55\",\"zindex\":7,\"wtype\":\"fixed\",\"htype\":\"fixed\",\"opacity\":1,\"borderStyle\":\"solid\",\"rotation\":0,\"scale\":1,\"aspectratio\":false},{\"name\":\"datetime\",\"type\":\"datetime\",\"lib\":\"common\",\"caption\":\"Date/Time 1\",\"width\":376,\"height\":41,\"backgroundColor\":\"none\",\"borderWidth\":\"1\",\"borderColor\":{\"type\":\"solid\",\"color\":\"rgba(102,150,200,1)\"},\"textFont\":\"Montserrat\",\"textFontStyle\":\"400\",\"textSize\":\"20\",\"textColor\":\"383838\",\"textProperties\":[],\"textAlign\":\"center\",\"dateFormat\":\"yearmonthday\",\"dayFormat\":\"superscript\",\"monthFormat\":\"long\",\"yearFormat\":\"long\",\"timeFormat\":\"24\",\"dateGeneralFormat\":\"timedate\",\"dayNameFormat\":\"long\",\"dateSeparator\":\"-\",\"left\":1064,\"top\":201,\"vpos\":\"top\",\"hpos\":\"left\",\"visible\":true,\"group\":\"root\",\"id\":\"box-50\",\"zindex\":5,\"wtype\":\"fixed\",\"htype\":\"fixed\",\"opacity\":1,\"rotation\":0,\"scale\":1,\"aspectratio\":false},{\"name\":\"text\",\"type\":\"text\",\"lib\":\"common\",\"caption\":\"MRN : 12345679...\",\"text\":\"MRN : 12345679&nbsp;\",\"textFont\":\"Trebuchet MS,sans-serif\",\"textFontStyle\":\"400\",\"textProperties\":[],\"textSize\":\"32\",\"textColor\":{\"type\":\"solid\",\"color\":\"rgba(113,113,113,1)\"},\"backgroundColor\":\"none\",\"width\":218,\"height\":39,\"lineHeight\":39,\"textAlign\":\"left\",\"textSpacing\":\"-1.6\",\"enableShadow\":false,\"autoResize\":true,\"v2\":true,\"left\":76,\"top\":171,\"vpos\":\"top\",\"hpos\":\"left\",\"visible\":true,\"group\":\"root\",\"id\":\"box-48\",\"zindex\":4,\"wtype\":\"fixed\",\"htype\":\"fixed\",\"opacity\":1,\"propagateEvents\":false,\"lineHeightAuto\":true,\"rotation\":0,\"scale\":1,\"aspectratio\":false},{\"name\":\"verticalline\",\"type\":\"verticalline\",\"lib\":\"common\",\"caption\":\"Vertical line 1\",\"width\":1,\"height\":658,\"color\":{\"type\":\"solid\",\"color\":\"rgba(70,127,215,1)\"},\"weight\":1,\"left\":734,\"top\":242,\"vpos\":\"top\",\"hpos\":\"left\",\"visible\":true,\"group\":\"root\",\"id\":\"box-47\",\"zindex\":3,\"wtype\":\"fixed\",\"htype\":\"fixed\",\"opacity\":1,\"borderStyle\":\"solid\",\"rotation\":0,\"scale\":1,\"aspectratio\":false},{\"name\":\"text\",\"type\":\"text\",\"lib\":\"common\",\"caption\":\"John Doe\",\"text\":\"John Doe&nbsp;\",\"textFont\":\"Trebuchet MS,sans-serif\",\"textFontStyle\":\"400\",\"textProperties\":[],\"textSize\":\"64\",\"textColor\":{\"type\":\"solid\",\"color\":\"rgba(117,85,88,1)\"},\"backgroundColor\":\"none\",\"width\":272,\"height\":78,\"lineHeight\":78,\"textAlign\":\"left\",\"textSpacing\":\"-1.6\",\"enableShadow\":false,\"autoResize\":true,\"v2\":true,\"left\":40,\"top\":15,\"vpos\":\"top\",\"hpos\":\"left\",\"visible\":true,\"id\":\"box-46\",\"group\":\"root\",\"zindex\":2,\"wtype\":\"fixed\",\"htype\":\"fixed\",\"opacity\":1,\"propagateEvents\":false,\"lineHeightAuto\":true,\"rotation\":0,\"scale\":1,\"aspectratio\":false},{\"name\":\"basic_slider\",\"type\":\"basic_slider\",\"lib\":\"generic2\",\"caption\":\"Slider 1\",\"width\":347,\"height\":41,\"barThickness\":4,\"barColor\":\"e3e6e8\",\"fillBarColor\":{\"type\":\"solid\",\"color\":\"rgba(255,255,0,1)\"},\"sliderColor\":\"ffffff\",\"barBorderRadius\":2,\"sliderBorderWidth\":1,\"sliderBorderRadius\":\"50%\",\"borderColor\":\"d4d7d9\",\"sliderPosition\":\"70\",\"left\":379,\"top\":359,\"vpos\":\"top\",\"hpos\":\"left\",\"visible\":true,\"id\":\"box-66\",\"group\":\"root\",\"zindex\":9,\"wtype\":\"fixed\",\"htype\":\"fixed\",\"opacity\":1,\"rotation\":0,\"scale\":1,\"aspectratio\":false,\"blurB\":0,\"saturateB\":1,\"brightnessB\":1,\"contrastB\":1,\"grayscaleB\":0,\"sepiaB\":0,\"invertB\":0,\"hueRotateB\":0,\"actions\":[{\"title\":\"Interaction 1\",\"type\":\"sliderdrag\",\"actionId\":\"set-variable\",\"callback\":true,\"variableName\":\"propofol_amount\",\"valueSource\":\"callbackvalue\",\"callbackvariable\":\"value\",\"delay\":\"0\",\"guid\":\"d83221a1-ad70-4aee-a0e3-0ac11141fee4\",\"callbackAction\":{\"actionId\":\"change-property\",\"callback\":true,\"pageId\":\"2\",\"itemId\":\"box-88\",\"propertyId\":\"text\",\"valueType\":\"readFromVariable\",\"variableName\":\"propofol_amount\",\"delay\":\"0\",\"guid\":\"e9419bbd-1c90-43ed-8b1d-353eafc30162\",\"callbackAction\":{\"actionId\":\"change-screen-state\",\"callback\":false,\"pageId\":\"2\",\"stateId\":\"73452cc4-51f4-48ca-9d25-b763541915d3\",\"delay\":\"0\",\"guid\":\"ff6e724f-1e19-4966-b390-e1091ca3671e\",\"clb_disabled\":false},\"clb_disabled\":false},\"sort\":0}]},{\"name\":\"basic_slider\",\"type\":\"basic_slider\",\"lib\":\"generic2\",\"caption\":\"Slider 2\",\"width\":347,\"height\":41,\"barThickness\":4,\"barColor\":\"e3e6e8\",\"fillBarColor\":{\"type\":\"solid\",\"color\":\"rgba(198,198,198,1)\"},\"sliderColor\":\"ffffff\",\"barBorderRadius\":2,\"sliderBorderWidth\":1,\"sliderBorderRadius\":\"50%\",\"borderColor\":\"d4d7d9\",\"sliderPosition\":\"44\",\"left\":373,\"top\":452,\"vpos\":\"top\",\"hpos\":\"left\",\"visible\":true,\"id\":\"box-71\",\"group\":\"root\",\"zindex\":9,\"wtype\":\"fixed\",\"htype\":\"fixed\",\"opacity\":1,\"rotation\":0,\"scale\":1,\"aspectratio\":false,\"actions\":[{\"title\":\"Set Val\",\"type\":\"sliderdrag\",\"actionId\":\"set-variable\",\"callback\":true,\"variableName\":\"lido\",\"valueSource\":\"callbackvalue\",\"callbackvariable\":\"value\",\"delay\":\"0\",\"guid\":\"be2e3f63-f34d-4d26-8881-0f4a471894c9\",\"callbackAction\":{\"actionId\":\"change-property\",\"callback\":true,\"pageId\":\"2\",\"itemId\":\"box-90\",\"propertyId\":\"text\",\"valueType\":\"readFromVariable\",\"variableName\":\"lido\",\"delay\":\"0\",\"guid\":\"16468069-edff-4d16-8428-c35f2b4e6106\",\"callbackAction\":{\"actionId\":\"change-screen-state\",\"callback\":false,\"pageId\":\"2\",\"stateId\":\"73452cc4-51f4-48ca-9d25-b763541915d3\",\"delay\":\"0\",\"guid\":\"c882f6c5-832b-4451-8d02-c26dc34fae3a\",\"clb_disabled\":false},\"clb_disabled\":false},\"sort\":0}]},{\"name\":\"basic_slider\",\"type\":\"basic_slider\",\"lib\":\"generic2\",\"caption\":\"Slider 3\",\"width\":347,\"height\":41,\"barThickness\":4,\"barColor\":\"e3e6e8\",\"fillBarColor\":{\"type\":\"solid\",\"color\":\"rgba(249,51,36,1)\"},\"sliderColor\":\"ffffff\",\"barBorderRadius\":2,\"sliderBorderWidth\":1,\"sliderBorderRadius\":\"50%\",\"borderColor\":\"d4d7d9\",\"sliderPosition\":\"61\",\"left\":379,\"top\":735,\"vpos\":\"top\",\"hpos\":\"left\",\"visible\":true,\"id\":\"box-75\",\"group\":\"root\",\"zindex\":9,\"wtype\":\"fixed\",\"htype\":\"fixed\",\"opacity\":1,\"rotation\":0,\"scale\":1,\"aspectratio\":false,\"actions\":[{\"title\":\"r\",\"type\":\"sliderdrag\",\"actionId\":\"set-variable\",\"callback\":true,\"variableName\":\"rocu\",\"valueSource\":\"callbackvalue\",\"callbackvariable\":\"value\",\"delay\":\"0\",\"guid\":\"aafe5c53-bbdf-4f47-899d-3521f88e75c4\",\"callbackAction\":{\"actionId\":\"change-property\",\"callback\":true,\"pageId\":\"2\",\"itemId\":\"box-93\",\"propertyId\":\"text\",\"valueType\":\"readFromVariable\",\"variableName\":\"rocu\",\"delay\":\"0\",\"guid\":\"e22baf5e-45f4-4986-b532-d0647afcfa88\",\"callbackAction\":{\"actionId\":\"change-screen-state\",\"callback\":false,\"pageId\":\"2\",\"stateId\":\"73452cc4-51f4-48ca-9d25-b763541915d3\",\"delay\":\"0\",\"guid\":\"05c89431-dc3f-4e25-b535-414916d40080\",\"clb_disabled\":false},\"clb_disabled\":false},\"sort\":0}]},{\"name\":\"basic_slider\",\"type\":\"basic_slider\",\"lib\":\"generic2\",\"caption\":\"Slider 4\",\"width\":347,\"height\":41,\"barThickness\":4,\"barColor\":\"e3e6e8\",\"fillBarColor\":{\"type\":\"solid\",\"color\":\"rgba(160,58,207,1)\"},\"sliderColor\":\"ffffff\",\"barBorderRadius\":2,\"sliderBorderWidth\":1,\"sliderBorderRadius\":\"50%\",\"borderColor\":\"d4d7d9\",\"sliderPosition\":\"40\",\"left\":379,\"top\":648,\"vpos\":\"top\",\"hpos\":\"left\",\"visible\":true,\"id\":\"box-76\",\"group\":\"root\",\"zindex\":9,\"wtype\":\"fixed\",\"htype\":\"fixed\",\"opacity\":1,\"rotation\":0,\"scale\":1,\"aspectratio\":false,\"actions\":[{\"title\":\"frt\",\"type\":\"sliderdrag\",\"actionId\":\"set-variable\",\"callback\":true,\"variableName\":\"phen\",\"valueSource\":\"callbackvalue\",\"callbackvariable\":\"value\",\"delay\":\"0\",\"guid\":\"916ca37b-9bd4-4e3b-a014-e443d285ba5e\",\"callbackAction\":{\"actionId\":\"change-property\",\"callback\":true,\"pageId\":\"2\",\"itemId\":\"box-92\",\"propertyId\":\"text\",\"valueType\":\"readFromVariable\",\"variableName\":\"phen\",\"delay\":\"0\",\"guid\":\"e09c9f08-4c05-47b8-9fe3-acc6da0d0773\",\"callbackAction\":{\"actionId\":\"change-screen-state\",\"callback\":false,\"pageId\":\"2\",\"stateId\":\"73452cc4-51f4-48ca-9d25-b763541915d3\",\"delay\":\"0\",\"guid\":\"93e8e3fc-6faf-4452-9589-b630cf120a54\",\"clb_disabled\":false},\"clb_disabled\":false},\"sort\":0}]},{\"name\":\"basic_slider\",\"type\":\"basic_slider\",\"lib\":\"generic2\",\"caption\":\"Slider 5\",\"width\":347,\"height\":41,\"barThickness\":4,\"barColor\":\"e3e6e8\",\"fillBarColor\":{\"type\":\"solid\",\"color\":\"rgba(102,150,200,1)\"},\"sliderColor\":\"ffffff\",\"barBorderRadius\":2,\"sliderBorderWidth\":1,\"sliderBorderRadius\":\"50%\",\"borderColor\":\"d4d7d9\",\"sliderPosition\":\"33\",\"left\":379,\"top\":550,\"vpos\":\"top\",\"hpos\":\"left\",\"visible\":true,\"id\":\"box-77\",\"group\":\"root\",\"zindex\":9,\"wtype\":\"fixed\",\"htype\":\"fixed\",\"opacity\":1,\"rotation\":0,\"scale\":1,\"aspectratio\":false,\"actions\":[{\"title\":\"f\",\"type\":\"sliderdrag\",\"actionId\":\"set-variable\",\"callback\":true,\"variableName\":\"fent\",\"valueSource\":\"callbackvalue\",\"callbackvariable\":\"value\",\"delay\":\"0\",\"guid\":\"3bb6f6e5-0ada-442d-a67e-54d701863f2a\",\"callbackAction\":{\"actionId\":\"change-property\",\"callback\":true,\"pageId\":\"2\",\"itemId\":\"box-91\",\"propertyId\":\"text\",\"valueType\":\"readFromVariable\",\"variableName\":\"fent\",\"delay\":\"0\",\"guid\":\"a542f1a9-fe52-4a0c-912e-efd83adb17b2\",\"callbackAction\":{\"actionId\":\"change-screen-state\",\"callback\":false,\"pageId\":\"2\",\"stateId\":\"73452cc4-51f4-48ca-9d25-b763541915d3\",\"delay\":\"0\",\"guid\":\"9aabd6af-1692-4f5f-af8d-5580a63b22fd\",\"clb_disabled\":false},\"clb_disabled\":false},\"sort\":0}]},{\"name\":\"basic_textfield\",\"type\":\"basic_textfield\",\"lib\":\"generic2\",\"caption\":\"Text Field prop\",\"width\":338,\"height\":55,\"value\":\"Propofol  ( mg ) \",\"placeholder\":\"Placeholder\",\"inputtype\":\"text\",\"backgroundColor\":{\"type\":\"solid\",\"color\":\"rgba(255,255,122,1)\"},\"textFont\":\"Arial,sans-serif\",\"textFontStyle\":\"400\",\"textSize\":\"28\",\"textColor\":{\"type\":\"solid\",\"color\":\"rgba(56,56,56,1)\"},\"textProperties\":[],\"placeholderColor\":\"c1c8c9\",\"borderWidth\":1,\"borderStyle\":\"solid\",\"borderRadius\":4,\"borderColor\":{\"type\":\"solid\",\"color\":\"rgba(255,255,0,1)\"},\"textAlign\":\"center\",\"left\":22,\"top\":359,\"vpos\":\"top\",\"hpos\":\"left\",\"visible\":true,\"group\":\"root\",\"id\":\"box-67\",\"zindex\":10,\"wtype\":\"fixed\",\"htype\":\"fixed\",\"opacity\":1,\"rotation\":0,\"scale\":1,\"aspectratio\":false,\"userCaption\":true},{\"name\":\"basic_textfield\",\"type\":\"basic_textfield\",\"lib\":\"generic2\",\"caption\":\"Text Field lido\",\"width\":338,\"height\":55,\"value\":\"Lidocaine  (mg)\",\"placeholder\":\"Placeholder\",\"inputtype\":\"text\",\"backgroundColor\":{\"type\":\"solid\",\"color\":\"rgba(198,198,198,1)\"},\"textFont\":\"Arial,sans-serif\",\"textFontStyle\":\"400\",\"textSize\":\"28\",\"textColor\":{\"type\":\"solid\",\"color\":\"rgba(56,56,56,1)\"},\"textProperties\":[],\"placeholderColor\":\"c1c8c9\",\"borderWidth\":1,\"borderStyle\":\"solid\",\"borderRadius\":4,\"borderColor\":{\"type\":\"solid\",\"color\":\"rgba(208,212,0,1)\"},\"textAlign\":\"center\",\"left\":22,\"top\":445,\"vpos\":\"top\",\"hpos\":\"left\",\"visible\":true,\"group\":\"root\",\"id\":\"box-72\",\"zindex\":10,\"wtype\":\"fixed\",\"htype\":\"fixed\",\"opacity\":1,\"rotation\":0,\"scale\":1,\"aspectratio\":false,\"userCaption\":true},{\"name\":\"basic_textfield\",\"type\":\"basic_textfield\",\"lib\":\"generic2\",\"caption\":\"Text Field Phenylephrine\",\"width\":338,\"height\":55,\"value\":\"Phenylephrine (mcg) \",\"placeholder\":\"Placeholder\",\"inputtype\":\"text\",\"backgroundColor\":{\"type\":\"solid\",\"color\":\"rgba(234,114,252,1)\"},\"textFont\":\"Arial,sans-serif\",\"textFontStyle\":\"400\",\"textSize\":\"28\",\"textColor\":{\"type\":\"solid\",\"color\":\"rgba(56,56,56,1)\"},\"textProperties\":[],\"placeholderColor\":\"c1c8c9\",\"borderWidth\":1,\"borderStyle\":\"solid\",\"borderRadius\":4,\"borderColor\":{\"type\":\"solid\",\"color\":\"rgba(255,255,0,1)\"},\"textAlign\":\"center\",\"left\":22,\"top\":641,\"vpos\":\"top\",\"hpos\":\"left\",\"visible\":true,\"group\":\"root\",\"id\":\"box-73\",\"zindex\":10,\"wtype\":\"fixed\",\"htype\":\"fixed\",\"opacity\":1,\"rotation\":0,\"scale\":1,\"aspectratio\":false,\"userCaption\":true},{\"name\":\"basic_textfield\",\"type\":\"basic_textfield\",\"lib\":\"generic2\",\"caption\":\"Text Field rocur\",\"width\":338,\"height\":55,\"value\":\"Rocuronium (mg)\",\"placeholder\":\"Placeholder\",\"inputtype\":\"text\",\"backgroundColor\":{\"type\":\"solid\",\"color\":\"rgba(226,15,0,1)\"},\"textFont\":\"Arial,sans-serif\",\"textFontStyle\":\"400\",\"textSize\":\"28\",\"textColor\":{\"type\":\"solid\",\"color\":\"rgba(56,56,56,1)\"},\"textProperties\":[],\"placeholderColor\":\"c1c8c9\",\"borderWidth\":1,\"borderStyle\":\"solid\",\"borderRadius\":4,\"borderColor\":{\"type\":\"solid\",\"color\":\"rgba(255,255,0,1)\"},\"textAlign\":\"center\",\"left\":22,\"top\":721,\"vpos\":\"top\",\"hpos\":\"left\",\"visible\":true,\"group\":\"root\",\"id\":\"box-79\",\"zindex\":10,\"wtype\":\"fixed\",\"htype\":\"fixed\",\"opacity\":1,\"rotation\":0,\"scale\":1,\"aspectratio\":false,\"userCaption\":true},{\"name\":\"basic_textfield\",\"type\":\"basic_textfield\",\"lib\":\"generic2\",\"caption\":\"Text Field 2\",\"width\":733,\"height\":77,\"value\":\"Induction Medication\",\"placeholder\":\"Placeholder\",\"inputtype\":\"text\",\"backgroundColor\":{\"type\":\"solid\",\"color\":\"rgba(177,190,227,1)\"},\"textFont\":\"Arial,sans-serif\",\"textFontStyle\":\"400\",\"textSize\":\"64\",\"textColor\":{\"type\":\"solid\",\"color\":\"rgba(233,240,215,1)\"},\"textProperties\":[],\"placeholderColor\":\"c1c8c9\",\"borderWidth\":\"5\",\"borderStyle\":\"solid\",\"borderRadius\":4,\"borderColor\":{\"type\":\"solid\",\"color\":\"rgba(85,85,85,1)\"},\"textAlign\":\"center\",\"left\":2,\"top\":242,\"vpos\":\"top\",\"hpos\":\"left\",\"visible\":true,\"id\":\"box-69\",\"group\":\"root\",\"zindex\":11,\"wtype\":\"fixed\",\"htype\":\"fixed\",\"opacity\":1,\"rotation\":0,\"scale\":1,\"aspectratio\":false},{\"name\":\"basic_textfield\",\"type\":\"basic_textfield\",\"lib\":\"generic2\",\"caption\":\"Text Field 3\",\"width\":698,\"height\":77,\"value\":\"Blood Pressure\",\"placeholder\":\"Placeholder\",\"inputtype\":\"text\",\"backgroundColor\":{\"type\":\"solid\",\"color\":\"rgba(177,190,227,1)\"},\"textFont\":\"Arial,sans-serif\",\"textFontStyle\":\"400\",\"textSize\":\"64\",\"textColor\":{\"type\":\"solid\",\"color\":\"rgba(233,240,215,1)\"},\"textProperties\":[],\"placeholderColor\":\"c1c8c9\",\"borderWidth\":\"5\",\"borderStyle\":\"solid\",\"borderRadius\":4,\"borderColor\":{\"type\":\"solid\",\"color\":\"rgba(85,85,85,1)\"},\"textAlign\":\"center\",\"left\":733,\"top\":244,\"vpos\":\"top\",\"hpos\":\"left\",\"visible\":true,\"id\":\"box-70\",\"group\":\"root\",\"zindex\":11,\"wtype\":\"fixed\",\"htype\":\"fixed\",\"opacity\":1,\"rotation\":0,\"scale\":1,\"aspectratio\":false},{\"name\":\"basic_textfield\",\"type\":\"basic_textfield\",\"lib\":\"generic2\",\"caption\":\"Text Field fenta\",\"width\":338,\"height\":55,\"value\":\"Fentanyl (mcg)\",\"placeholder\":\"Placeholder\",\"inputtype\":\"text\",\"backgroundColor\":{\"type\":\"solid\",\"color\":\"rgba(101,180,206,1)\"},\"textFont\":\"Arial,sans-serif\",\"textFontStyle\":\"400\",\"textSize\":\"28\",\"textColor\":{\"type\":\"solid\",\"color\":\"rgba(56,56,56,1)\"},\"textProperties\":[],\"placeholderColor\":\"c1c8c9\",\"borderWidth\":1,\"borderStyle\":\"solid\",\"borderRadius\":4,\"borderColor\":{\"type\":\"solid\",\"color\":\"rgba(27,77,87,1)\"},\"textAlign\":\"center\",\"left\":22,\"top\":543,\"vpos\":\"top\",\"hpos\":\"left\",\"visible\":true,\"group\":\"root\",\"id\":\"box-78\",\"zindex\":10,\"wtype\":\"fixed\",\"htype\":\"fixed\",\"opacity\":1,\"rotation\":0,\"scale\":1,\"aspectratio\":false,\"userCaption\":true},{\"name\":\"image\",\"type\":\"image\",\"lib\":\"common\",\"caption\":\"ss4\",\"imgSrc\":{\"fileId\":\"68d047acd614373a75ca2548bba92b44.png\",\"assetType\":\"project\",\"bucketsource\":\"main\",\"name\":\"Screen Shot 2022-12-12 at 7.38.12 PM.png\"},\"repeat\":false,\"width\":664,\"height\":402.63829787234044,\"borderWidth\":0,\"borderRadius\":0,\"borderColor\":\"#D1D1D1\",\"borderPos\":\"inside\",\"propagateEvents\":false,\"actions\":[],\"overlay\":false,\"autoResize\":false,\"aspectratio\":1.6491228070175439,\"assistiveDescription\":\"\",\"left\":750,\"top\":357,\"visible\":true,\"id\":\"box-80\",\"group\":\"root\",\"zindex\":12,\"hpos\":\"left\",\"vpos\":\"top\",\"wtype\":\"fixed\",\"htype\":\"fixed\",\"opacity\":1,\"borderStyle\":\"solid\",\"rotation\":0,\"scale\":1,\"hasMask\":false,\"userCaption\":true},{\"name\":\"basic_button_text\",\"type\":\"basic_button_text\",\"lib\":\"generic2\",\"caption\":\"Button with Background 1\",\"width\":177,\"height\":80,\"backgroundColor\":{\"type\":\"solid\",\"color\":\"rgba(102,150,200,1)\"},\"borderWidth\":0,\"borderColor\":\"#025abf\",\"borderRadius\":4,\"text\":\"Other Opioids\",\"textFont\":\"Arial,sans-serif\",\"textFontStyle\":\"400\",\"textSize\":\"28\",\"textColor\":\"FFFFFF\",\"textProperties\":[],\"textAlign\":\"center\",\"iconpos\":\"none\",\"left\":32,\"top\":800,\"vpos\":\"top\",\"hpos\":\"left\",\"visible\":true,\"id\":\"box-81\",\"group\":\"root\",\"zindex\":13,\"wtype\":\"fixed\",\"htype\":\"fixed\",\"opacity\":1,\"rotation\":0,\"scale\":1,\"aspectratio\":false,\"actions\":[{\"title\":\"Interaction 1\",\"type\":\"tap\",\"actionId\":\"go-to-page\",\"callback\":false,\"pageId\":\"4\",\"animation\":\"fade\",\"animationEasing\":\"Cubic.easeOut\",\"duration\":\"400\",\"delay\":\"0\",\"guid\":\"afa3724f-fd08-4a24-a7dc-0e68229a2ab1\",\"sort\":0}]},{\"name\":\"basic_button_text\",\"type\":\"basic_button_text\",\"lib\":\"generic2\",\"caption\":\"Button with Background 3\",\"width\":177,\"height\":80,\"backgroundColor\":{\"type\":\"solid\",\"color\":\"rgba(255, 255, 0,1)\"},\"borderWidth\":0,\"borderColor\":\"#025abf\",\"borderRadius\":4,\"text\":\"Other Amestics\",\"textFont\":\"Arial,sans-serif\",\"textFontStyle\":\"400\",\"textSize\":\"28\",\"textColor\":{\"type\":\"solid\",\"color\":\"rgba(56,56,56,1)\"},\"textProperties\":[],\"textAlign\":\"center\",\"iconpos\":\"none\",\"left\":252,\"top\":800,\"vpos\":\"top\",\"hpos\":\"left\",\"visible\":true,\"id\":\"box-83\",\"group\":\"root\",\"zindex\":13,\"wtype\":\"fixed\",\"htype\":\"fixed\",\"opacity\":1,\"rotation\":0,\"scale\":1,\"aspectratio\":false,\"actions\":[{\"title\":\"Interaction 1\",\"type\":\"tap\",\"actionId\":\"go-to-page\",\"callback\":false,\"pageId\":\"7\",\"animation\":\"fade\",\"animationEasing\":\"Cubic.easeOut\",\"duration\":\"400\",\"delay\":\"0\",\"guid\":\"21ed5ca2-ee39-46fb-afa5-394bb4aaaa61\",\"sort\":0}]},{\"name\":\"basic_button_text\",\"type\":\"basic_button_text\",\"lib\":\"generic2\",\"caption\":\"Button with Background 4\",\"width\":177,\"height\":80,\"backgroundColor\":{\"type\":\"solid\",\"color\":\"rgba(234,114,252,1)\"},\"borderWidth\":0,\"borderColor\":\"#025abf\",\"borderRadius\":4,\"text\":\"Other Pressors\",\"textFont\":\"Arial,sans-serif\",\"textFontStyle\":\"400\",\"textSize\":\"28\",\"textColor\":\"FFFFFF\",\"textProperties\":[],\"textAlign\":\"center\",\"iconpos\":\"none\",\"left\":489,\"top\":800,\"vpos\":\"top\",\"hpos\":\"left\",\"visible\":true,\"id\":\"box-84\",\"group\":\"root\",\"zindex\":13,\"wtype\":\"fixed\",\"htype\":\"fixed\",\"opacity\":1,\"rotation\":0,\"scale\":1,\"aspectratio\":false,\"actions\":[{\"title\":\"Interaction 1\",\"type\":\"tap\",\"actionId\":\"go-to-page\",\"callback\":false,\"pageId\":\"6\",\"animation\":\"fade\",\"animationEasing\":\"Cubic.easeOut\",\"duration\":\"400\",\"delay\":\"0\",\"guid\":\"b627cf4b-fa30-4f86-9534-4a5e4056a205\",\"sort\":0}]},{\"name\":\"basic_button_text\",\"type\":\"basic_button_text\",\"lib\":\"generic2\",\"caption\":\"Button with Background 2\",\"width\":177,\"height\":80,\"backgroundColor\":{\"type\":\"solid\",\"color\":\"rgba(249,51,36,1)\"},\"borderWidth\":0,\"borderColor\":\"#025abf\",\"borderRadius\":4,\"text\":\"Exit\",\"textFont\":\"Arial,sans-serif\",\"textFontStyle\":\"400\",\"textSize\":\"36\",\"textColor\":\"FFFFFF\",\"textProperties\":[],\"textAlign\":\"center\",\"iconpos\":\"none\",\"left\":1233,\"top\":789,\"vpos\":\"top\",\"hpos\":\"left\",\"visible\":true,\"id\":\"box-82\",\"group\":\"root\",\"zindex\":13,\"wtype\":\"fixed\",\"htype\":\"fixed\",\"opacity\":1,\"rotation\":0,\"scale\":1,\"aspectratio\":false,\"actions\":[{\"title\":\"Interaction 1\",\"type\":\"click\",\"actionId\":\"go-to-page\",\"callback\":false,\"pageId\":\"1\",\"animation\":\"fade\",\"animationEasing\":\"Cubic.easeOut\",\"duration\":\"400\",\"delay\":\"0\",\"guid\":\"12996f29-86d0-4929-9a3f-2ec00fad2b1a\",\"sort\":0}]},{\"name\":\"text\",\"type\":\"text\",\"lib\":\"common\",\"caption\":\"text\",\"text\":\"150\",\"textFont\":\"Arial,sans-serif\",\"textFontStyle\":\"400\",\"textProperties\":[],\"textSize\":\"24\",\"textColor\":{\"type\":\"solid\",\"color\":\"rgba(255,255,0,1)\"},\"backgroundColor\":\"none\",\"width\":275,\"height\":29,\"lineHeight\":29,\"textAlign\":\"left\",\"textSpacing\":0,\"enableShadow\":false,\"autoResize\":false,\"v2\":true,\"left\":505,\"top\":330,\"vpos\":\"top\",\"hpos\":\"left\",\"visible\":true,\"id\":\"box-88\",\"group\":\"root\",\"zindex\":14,\"wtype\":\"fixed\",\"htype\":\"fixed\",\"opacity\":1,\"propagateEvents\":false,\"lineHeightAuto\":true,\"rotation\":0,\"scale\":1,\"aspectratio\":false,\"userCaption\":true,\"actions\":[]},{\"name\":\"text\",\"type\":\"text\",\"lib\":\"common\",\"caption\":\"text 2\",\"text\":\"100\",\"textFont\":\"Arial,sans-serif\",\"textFontStyle\":\"400\",\"textProperties\":[],\"textSize\":\"24\",\"textColor\":{\"type\":\"solid\",\"color\":\"rgba(198,198,198,1)\"},\"backgroundColor\":\"none\",\"width\":275,\"height\":29,\"lineHeight\":29,\"textAlign\":\"left\",\"textSpacing\":0,\"enableShadow\":false,\"autoResize\":false,\"v2\":true,\"left\":505,\"top\":414,\"vpos\":\"top\",\"hpos\":\"left\",\"visible\":true,\"id\":\"box-90\",\"group\":\"root\",\"zindex\":14,\"wtype\":\"fixed\",\"htype\":\"fixed\",\"opacity\":1,\"propagateEvents\":false,\"lineHeightAuto\":true,\"rotation\":0,\"scale\":1,\"aspectratio\":false,\"userCaption\":true,\"actions\":[]},{\"name\":\"text\",\"type\":\"text\",\"lib\":\"common\",\"caption\":\"text 3\",\"text\":\"100\",\"textFont\":\"Arial,sans-serif\",\"textFontStyle\":\"400\",\"textProperties\":[],\"textSize\":\"24\",\"textColor\":{\"type\":\"solid\",\"color\":\"rgba(43,89,162,1)\"},\"backgroundColor\":\"none\",\"width\":275,\"height\":29,\"lineHeight\":29,\"textAlign\":\"left\",\"textSpacing\":0,\"enableShadow\":false,\"autoResize\":false,\"v2\":true,\"left\":505,\"top\":521,\"vpos\":\"top\",\"hpos\":\"left\",\"visible\":true,\"id\":\"box-91\",\"group\":\"root\",\"zindex\":14,\"wtype\":\"fixed\",\"htype\":\"fixed\",\"opacity\":1,\"propagateEvents\":false,\"lineHeightAuto\":true,\"rotation\":0,\"scale\":1,\"aspectratio\":false,\"userCaption\":true,\"actions\":[]},{\"name\":\"text\",\"type\":\"text\",\"lib\":\"common\",\"caption\":\"text 4\",\"text\":\"100\",\"textFont\":\"Arial,sans-serif\",\"textFontStyle\":\"400\",\"textProperties\":[],\"textSize\":\"24\",\"textColor\":{\"type\":\"solid\",\"color\":\"rgba(183,47,214,1)\"},\"backgroundColor\":\"none\",\"width\":275,\"height\":29,\"lineHeight\":29,\"textAlign\":\"left\",\"textSpacing\":0,\"enableShadow\":false,\"autoResize\":false,\"v2\":true,\"left\":497,\"top\":612,\"vpos\":\"top\",\"hpos\":\"left\",\"visible\":true,\"id\":\"box-92\",\"group\":\"root\",\"zindex\":14,\"wtype\":\"fixed\",\"htype\":\"fixed\",\"opacity\":1,\"propagateEvents\":false,\"lineHeightAuto\":true,\"rotation\":0,\"scale\":1,\"aspectratio\":false,\"userCaption\":true,\"actions\":[]},{\"name\":\"text\",\"type\":\"text\",\"lib\":\"common\",\"caption\":\"text 5\",\"text\":\"100\",\"textFont\":\"Arial,sans-serif\",\"textFontStyle\":\"400\",\"textProperties\":[],\"textSize\":\"24\",\"textColor\":{\"type\":\"solid\",\"color\":\"rgba(209,25,105,1)\"},\"backgroundColor\":\"none\",\"width\":275,\"height\":29,\"lineHeight\":29,\"textAlign\":\"left\",\"textSpacing\":0,\"enableShadow\":false,\"autoResize\":false,\"v2\":true,\"left\":491,\"top\":712,\"vpos\":\"top\",\"hpos\":\"left\",\"visible\":true,\"id\":\"box-93\",\"group\":\"root\",\"zindex\":14,\"wtype\":\"fixed\",\"htype\":\"fixed\",\"opacity\":1,\"propagateEvents\":false,\"lineHeightAuto\":true,\"rotation\":0,\"scale\":1,\"aspectratio\":false,\"userCaption\":true,\"actions\":[]}]","template":{"id":"","stateid":""},"history":[],"id":"fc0c0693-42f3-489d-adea-e259a6e69adf","actions":[],"groups":{}}]},{"id":4,"title":"Opioid","orientation":"landscape","statusbar":"0","navigationbar":"0","actions":[],"states":[{"title":"State 1","background":{"type":"solid","color":"rgba(169,151,196,1)"},"data":"[{\"name\":\"rectangle\",\"type\":\"rectangle\",\"lib\":\"common\",\"caption\":\"Rectangle 1\",\"backgroundColor\":{\"type\":\"solid\",\"color\":\"rgba(193,212,241,1)\"},\"borderWidth\":\"8\",\"borderColor\":{\"type\":\"solid\",\"color\":\"rgba(30,77,130,1)\"},\"borderRadius\":0,\"imgSrc\":{\"fileId\":\"\",\"name\":\"\",\"assetType\":\"\",\"bucketsource\":\"\",\"url\":\"\"},\"backgroundSizeType\":\"cover\",\"backgroundSizeValue\":35,\"width\":1440,\"height\":249,\"lineHeight\":20,\"lineHeightAuto\":true,\"actions\":[],\"textFont\":\"Arial,sans-serif\",\"textFontStyle\":\"400\",\"textSize\":16,\"textColor\":\"#383838\",\"textProperties\":[],\"textSpacing\":0,\"textAlign\":\"center\",\"text\":\"\",\"typeName\":\"rectangle\",\"left\":0,\"top\":0,\"vpos\":\"top\",\"hpos\":\"left\",\"visible\":true,\"id\":\"box-94\",\"group\":\"root\",\"zindex\":1,\"wtype\":\"fixed\",\"htype\":\"fixed\",\"opacity\":1,\"borderStyle\":\"solid\",\"rotation\":0,\"scale\":1,\"aspectratio\":false},{\"name\":\"text\",\"type\":\"text\",\"lib\":\"common\",\"caption\":\"Gender : Male...\",\"text\":\"Gender : Male\",\"textFont\":\"Trebuchet MS,sans-serif\",\"textFontStyle\":\"400\",\"textProperties\":[],\"textSize\":\"32\",\"textColor\":{\"type\":\"solid\",\"color\":\"rgba(113,113,113,1)\"},\"backgroundColor\":\"none\",\"width\":186,\"height\":39,\"lineHeight\":39,\"textAlign\":\"left\",\"textSpacing\":\"-1.6\",\"enableShadow\":false,\"autoResize\":true,\"v2\":true,\"left\":379,\"top\":47,\"vpos\":\"top\",\"hpos\":\"left\",\"visible\":true,\"group\":\"root\",\"id\":\"box-95\",\"zindex\":8,\"wtype\":\"fixed\",\"htype\":\"fixed\",\"opacity\":1,\"propagateEvents\":false,\"lineHeightAuto\":true,\"rotation\":0,\"scale\":1,\"aspectratio\":false},{\"name\":\"text\",\"type\":\"text\",\"lib\":\"common\",\"caption\":\"DOB: 01/01/2022...\",\"text\":\"DOB: 01/01/2022\",\"textFont\":\"Trebuchet MS,sans-serif\",\"textFontStyle\":\"400\",\"textProperties\":[],\"textSize\":\"32\",\"textColor\":{\"type\":\"solid\",\"color\":\"rgba(113,113,113,1)\"},\"backgroundColor\":\"none\",\"width\":230,\"height\":39,\"lineHeight\":39,\"textAlign\":\"left\",\"textSpacing\":\"-1.6\",\"enableShadow\":false,\"autoResize\":true,\"v2\":true,\"left\":76,\"top\":130,\"vpos\":\"top\",\"hpos\":\"left\",\"visible\":true,\"group\":\"root\",\"id\":\"box-96\",\"zindex\":8,\"wtype\":\"fixed\",\"htype\":\"fixed\",\"opacity\":1,\"propagateEvents\":false,\"lineHeightAuto\":true,\"rotation\":0,\"scale\":1,\"aspectratio\":false},{\"name\":\"text\",\"type\":\"text\",\"lib\":\"common\",\"caption\":\"Height : 174 cm...\",\"text\":\"Height : 174 cm&nbsp;\",\"textFont\":\"Trebuchet MS,sans-serif\",\"textFontStyle\":\"400\",\"textProperties\":[],\"textSize\":\"32\",\"textColor\":{\"type\":\"solid\",\"color\":\"rgba(113,113,113,1)\"},\"backgroundColor\":\"none\",\"width\":216,\"height\":39,\"lineHeight\":39,\"textAlign\":\"left\",\"textSpacing\":\"-1.6\",\"enableShadow\":false,\"autoResize\":true,\"v2\":true,\"left\":379,\"top\":86,\"vpos\":\"top\",\"hpos\":\"left\",\"visible\":true,\"group\":\"root\",\"id\":\"box-97\",\"zindex\":8,\"wtype\":\"fixed\",\"htype\":\"fixed\",\"opacity\":1,\"propagateEvents\":false,\"lineHeightAuto\":true,\"rotation\":0,\"scale\":1,\"aspectratio\":false},{\"name\":\"text\",\"type\":\"text\",\"lib\":\"common\",\"caption\":\"Surgery:\\nTotal Colec...\",\"text\":\"Surgery:&nbsp;<br>Total Colectomy with&nbsp;<br>pancreatico-jejunostomy&nbsp;\",\"textFont\":\"Trebuchet MS,sans-serif\",\"textFontStyle\":\"400\",\"textProperties\":[],\"textSize\":\"32\",\"textColor\":{\"type\":\"solid\",\"color\":\"rgba(113,113,113,1)\"},\"backgroundColor\":\"none\",\"width\":336,\"height\":117,\"lineHeight\":39,\"textAlign\":\"left\",\"textSpacing\":\"-1.6\",\"enableShadow\":false,\"autoResize\":true,\"v2\":true,\"left\":1084,\"top\":47,\"vpos\":\"top\",\"hpos\":\"left\",\"visible\":true,\"group\":\"root\",\"id\":\"box-98\",\"zindex\":8,\"wtype\":\"fixed\",\"htype\":\"fixed\",\"opacity\":1,\"propagateEvents\":false,\"lineHeightAuto\":true,\"rotation\":0,\"scale\":1,\"aspectratio\":false},{\"name\":\"text\",\"type\":\"text\",\"lib\":\"common\",\"caption\":\"Age : 86 y\",\"text\":\"Age : 86 y&nbsp;\",\"textFont\":\"Trebuchet MS,sans-serif\",\"textFontStyle\":\"400\",\"textProperties\":[],\"textSize\":\"32\",\"textColor\":{\"type\":\"solid\",\"color\":\"rgba(113,113,113,1)\"},\"backgroundColor\":\"none\",\"width\":139,\"height\":39,\"lineHeight\":39,\"textAlign\":\"left\",\"textSpacing\":\"-1.6\",\"enableShadow\":false,\"autoResize\":true,\"v2\":true,\"left\":76,\"top\":91,\"vpos\":\"top\",\"hpos\":\"left\",\"visible\":true,\"group\":\"root\",\"id\":\"box-99\",\"zindex\":8,\"wtype\":\"fixed\",\"htype\":\"fixed\",\"opacity\":1,\"propagateEvents\":false,\"lineHeightAuto\":true,\"rotation\":0,\"scale\":1,\"aspectratio\":false},{\"name\":\"text\",\"type\":\"text\",\"lib\":\"common\",\"caption\":\"Allergies :\\nPenicill...\",\"text\":\"Allergies :<br><span style=\\\"color: rgba(249,51,36,1);\\\">Penicillin</span><br><br>\",\"textFont\":\"Trebuchet MS,sans-serif\",\"textFontStyle\":\"400\",\"textProperties\":[],\"textSize\":\"32\",\"textColor\":{\"type\":\"solid\",\"color\":\"rgba(113,113,113,1)\"},\"backgroundColor\":\"none\",\"width\":203,\"height\":117,\"lineHeight\":39,\"textAlign\":\"left\",\"textSpacing\":\"-1.6\",\"enableShadow\":false,\"autoResize\":false,\"v2\":true,\"left\":766,\"top\":47,\"vpos\":\"top\",\"hpos\":\"left\",\"visible\":true,\"group\":\"root\",\"id\":\"box-100\",\"zindex\":8,\"wtype\":\"fixed\",\"htype\":\"fixed\",\"opacity\":1,\"propagateEvents\":false,\"lineHeightAuto\":true,\"rotation\":0,\"scale\":1,\"aspectratio\":false},{\"name\":\"text\",\"type\":\"text\",\"lib\":\"common\",\"caption\":\"BMI : 28 kg/m2...\",\"text\":\"BMI : 28 kg/m2\",\"textFont\":\"Trebuchet MS,sans-serif\",\"textFontStyle\":\"400\",\"textProperties\":[],\"textSize\":\"32\",\"textColor\":{\"type\":\"solid\",\"color\":\"rgba(113,113,113,1)\"},\"backgroundColor\":\"none\",\"width\":199,\"height\":39,\"lineHeight\":39,\"textAlign\":\"left\",\"textSpacing\":\"-1.6\",\"enableShadow\":false,\"autoResize\":true,\"v2\":true,\"left\":379,\"top\":171,\"vpos\":\"top\",\"hpos\":\"left\",\"visible\":true,\"group\":\"root\",\"id\":\"box-101\",\"zindex\":8,\"wtype\":\"fixed\",\"htype\":\"fixed\",\"opacity\":1,\"propagateEvents\":false,\"lineHeightAuto\":true,\"rotation\":0,\"scale\":1,\"aspectratio\":false},{\"name\":\"text\",\"type\":\"text\",\"lib\":\"common\",\"caption\":\"Weight : 69 Kg...\",\"text\":\"Weight : 69 Kg\",\"textFont\":\"Trebuchet MS,sans-serif\",\"textFontStyle\":\"400\",\"textProperties\":[],\"textSize\":\"32\",\"textColor\":{\"type\":\"solid\",\"color\":\"rgba(113,113,113,1)\"},\"backgroundColor\":\"none\",\"width\":190,\"height\":39,\"lineHeight\":39,\"textAlign\":\"left\",\"textSpacing\":\"-1.6\",\"enableShadow\":false,\"autoResize\":true,\"v2\":true,\"left\":379,\"top\":130,\"vpos\":\"top\",\"hpos\":\"left\",\"visible\":true,\"group\":\"root\",\"id\":\"box-102\",\"zindex\":8,\"wtype\":\"fixed\",\"htype\":\"fixed\",\"opacity\":1,\"propagateEvents\":false,\"lineHeightAuto\":true,\"rotation\":0,\"scale\":1,\"aspectratio\":false},{\"name\":\"verticalline\",\"type\":\"verticalline\",\"lib\":\"common\",\"caption\":\"Vertical line 3\",\"width\":1,\"height\":229,\"color\":\"A6A6A6\",\"weight\":1,\"left\":341,\"top\":10,\"vpos\":\"top\",\"hpos\":\"left\",\"visible\":true,\"group\":\"root\",\"id\":\"box-103\",\"zindex\":7,\"wtype\":\"fixed\",\"htype\":\"fixed\",\"opacity\":1,\"borderStyle\":\"solid\",\"rotation\":0,\"scale\":1,\"aspectratio\":false},{\"name\":\"verticalline\",\"type\":\"verticalline\",\"lib\":\"common\",\"caption\":\"Vertical line 5\",\"width\":1,\"height\":229,\"color\":\"A6A6A6\",\"weight\":1,\"left\":1064,\"top\":15,\"vpos\":\"top\",\"hpos\":\"left\",\"visible\":true,\"group\":\"root\",\"id\":\"box-104\",\"zindex\":7,\"wtype\":\"fixed\",\"htype\":\"fixed\",\"opacity\":1,\"borderStyle\":\"solid\",\"rotation\":0,\"scale\":1,\"aspectratio\":false},{\"name\":\"verticalline\",\"type\":\"verticalline\",\"lib\":\"common\",\"caption\":\"Vertical line 6\",\"width\":1,\"height\":229,\"color\":\"A6A6A6\",\"weight\":1,\"left\":733,\"top\":10,\"vpos\":\"top\",\"hpos\":\"left\",\"visible\":true,\"group\":\"root\",\"id\":\"box-105\",\"zindex\":7,\"wtype\":\"fixed\",\"htype\":\"fixed\",\"opacity\":1,\"borderStyle\":\"solid\",\"rotation\":0,\"scale\":1,\"aspectratio\":false},{\"name\":\"datetime\",\"type\":\"datetime\",\"lib\":\"common\",\"caption\":\"Date/Time 1\",\"width\":376,\"height\":41,\"backgroundColor\":\"none\",\"borderWidth\":\"1\",\"borderColor\":{\"type\":\"solid\",\"color\":\"rgba(102,150,200,1)\"},\"textFont\":\"Montserrat\",\"textFontStyle\":\"400\",\"textSize\":\"20\",\"textColor\":\"383838\",\"textProperties\":[],\"textAlign\":\"center\",\"dateFormat\":\"yearmonthday\",\"dayFormat\":\"superscript\",\"monthFormat\":\"long\",\"yearFormat\":\"long\",\"timeFormat\":\"24\",\"dateGeneralFormat\":\"timedate\",\"dayNameFormat\":\"long\",\"dateSeparator\":\"-\",\"left\":1064,\"top\":201,\"vpos\":\"top\",\"hpos\":\"left\",\"visible\":true,\"group\":\"root\",\"id\":\"box-106\",\"zindex\":5,\"wtype\":\"fixed\",\"htype\":\"fixed\",\"opacity\":1,\"rotation\":0,\"scale\":1,\"aspectratio\":false},{\"name\":\"text\",\"type\":\"text\",\"lib\":\"common\",\"caption\":\"MRN : 12345679...\",\"text\":\"MRN : 12345679&nbsp;\",\"textFont\":\"Trebuchet MS,sans-serif\",\"textFontStyle\":\"400\",\"textProperties\":[],\"textSize\":\"32\",\"textColor\":{\"type\":\"solid\",\"color\":\"rgba(113,113,113,1)\"},\"backgroundColor\":\"none\",\"width\":218,\"height\":39,\"lineHeight\":39,\"textAlign\":\"left\",\"textSpacing\":\"-1.6\",\"enableShadow\":false,\"autoResize\":true,\"v2\":true,\"left\":76,\"top\":171,\"vpos\":\"top\",\"hpos\":\"left\",\"visible\":true,\"group\":\"root\",\"id\":\"box-107\",\"zindex\":4,\"wtype\":\"fixed\",\"htype\":\"fixed\",\"opacity\":1,\"propagateEvents\":false,\"lineHeightAuto\":true,\"rotation\":0,\"scale\":1,\"aspectratio\":false},{\"name\":\"verticalline\",\"type\":\"verticalline\",\"lib\":\"common\",\"caption\":\"Vertical line 1\",\"width\":1,\"height\":658,\"color\":{\"type\":\"solid\",\"color\":\"rgba(70,127,215,1)\"},\"weight\":1,\"left\":734,\"top\":242,\"vpos\":\"top\",\"hpos\":\"left\",\"visible\":true,\"group\":\"root\",\"id\":\"box-108\",\"zindex\":3,\"wtype\":\"fixed\",\"htype\":\"fixed\",\"opacity\":1,\"borderStyle\":\"solid\",\"rotation\":0,\"scale\":1,\"aspectratio\":false},{\"name\":\"text\",\"type\":\"text\",\"lib\":\"common\",\"caption\":\"John Doe\",\"text\":\"John Doe&nbsp;\",\"textFont\":\"Trebuchet MS,sans-serif\",\"textFontStyle\":\"400\",\"textProperties\":[],\"textSize\":\"64\",\"textColor\":{\"type\":\"solid\",\"color\":\"rgba(117,85,88,1)\"},\"backgroundColor\":\"none\",\"width\":272,\"height\":78,\"lineHeight\":78,\"textAlign\":\"left\",\"textSpacing\":\"-1.6\",\"enableShadow\":false,\"autoResize\":true,\"v2\":true,\"left\":40,\"top\":15,\"vpos\":\"top\",\"hpos\":\"left\",\"visible\":true,\"id\":\"box-109\",\"group\":\"root\",\"zindex\":2,\"wtype\":\"fixed\",\"htype\":\"fixed\",\"opacity\":1,\"propagateEvents\":false,\"lineHeightAuto\":true,\"rotation\":0,\"scale\":1,\"aspectratio\":false},{\"name\":\"basic_textfield\",\"type\":\"basic_textfield\",\"lib\":\"generic2\",\"caption\":\"Text Field 2\",\"width\":733,\"height\":77,\"value\":\"Induction Medication\",\"placeholder\":\"Placeholder\",\"inputtype\":\"text\",\"backgroundColor\":{\"type\":\"solid\",\"color\":\"rgba(177,190,227,1)\"},\"textFont\":\"Arial,sans-serif\",\"textFontStyle\":\"400\",\"textSize\":\"64\",\"textColor\":{\"type\":\"solid\",\"color\":\"rgba(233,240,215,1)\"},\"textProperties\":[],\"placeholderColor\":\"c1c8c9\",\"borderWidth\":\"5\",\"borderStyle\":\"solid\",\"borderRadius\":4,\"borderColor\":{\"type\":\"solid\",\"color\":\"rgba(85,85,85,1)\"},\"textAlign\":\"center\",\"left\":2,\"top\":242,\"vpos\":\"top\",\"hpos\":\"left\",\"visible\":true,\"id\":\"box-119\",\"group\":\"root\",\"zindex\":11,\"wtype\":\"fixed\",\"htype\":\"fixed\",\"opacity\":1,\"rotation\":0,\"scale\":1,\"aspectratio\":false},{\"name\":\"basic_textfield\",\"type\":\"basic_textfield\",\"lib\":\"generic2\",\"caption\":\"Text Field 3\",\"width\":698,\"height\":77,\"value\":\"Blood Pressure\",\"placeholder\":\"Placeholder\",\"inputtype\":\"text\",\"backgroundColor\":{\"type\":\"solid\",\"color\":\"rgba(177,190,227,1)\"},\"textFont\":\"Arial,sans-serif\",\"textFontStyle\":\"400\",\"textSize\":\"64\",\"textColor\":{\"type\":\"solid\",\"color\":\"rgba(233,240,215,1)\"},\"textProperties\":[],\"placeholderColor\":\"c1c8c9\",\"borderWidth\":\"5\",\"borderStyle\":\"solid\",\"borderRadius\":4,\"borderColor\":{\"type\":\"solid\",\"color\":\"rgba(85,85,85,1)\"},\"textAlign\":\"center\",\"left\":733,\"top\":244,\"vpos\":\"top\",\"hpos\":\"left\",\"visible\":true,\"id\":\"box-120\",\"group\":\"root\",\"zindex\":11,\"wtype\":\"fixed\",\"htype\":\"fixed\",\"opacity\":1,\"rotation\":0,\"scale\":1,\"aspectratio\":false},{\"name\":\"basic_textfield\",\"type\":\"basic_textfield\",\"lib\":\"generic2\",\"caption\":\"Text Field 13\",\"width\":338,\"height\":55,\"value\":\"Hydromorphone (mg)\",\"placeholder\":\"Placeholder\",\"inputtype\":\"text\",\"backgroundColor\":{\"type\":\"solid\",\"color\":\"rgba(101,180,206,1)\"},\"textFont\":\"Arial,sans-serif\",\"textFontStyle\":\"400\",\"textSize\":\"28\",\"textColor\":{\"type\":\"solid\",\"color\":\"rgba(56,56,56,1)\"},\"textProperties\":[],\"placeholderColor\":\"c1c8c9\",\"borderWidth\":1,\"borderStyle\":\"solid\",\"borderRadius\":4,\"borderColor\":{\"type\":\"solid\",\"color\":\"rgba(27,77,87,1)\"},\"textAlign\":\"center\",\"left\":31,\"top\":374,\"vpos\":\"top\",\"hpos\":\"left\",\"visible\":true,\"group\":\"root\",\"id\":\"box-136\",\"zindex\":10,\"wtype\":\"fixed\",\"htype\":\"fixed\",\"opacity\":1,\"rotation\":0,\"scale\":1,\"aspectratio\":false,\"actions\":[{\"title\":\"click \",\"type\":\"click\",\"actionId\":\"change-property\",\"callback\":true,\"pageId\":\"2\",\"itemId\":\"box-78\",\"propertyId\":\"value\",\"valueType\":\"customValue\",\"newvalue\":\"Hydromorphone (mg)\",\"delay\":\"0\",\"guid\":\"b60c630b-5aa9-4da6-ad36-4690f3b46170\",\"callbackAction\":{\"actionId\":\"go-to-page\",\"callback\":false,\"pageId\":\"2\",\"animation\":\"fade\",\"animationEasing\":\"Cubic.easeOut\",\"duration\":\"400\",\"delay\":\"0\",\"guid\":\"2fe8ad00-3cfb-45aa-9706-b07cb75f893b\",\"clb_disabled\":false},\"sort\":0}]},{\"name\":\"basic_textfield\",\"type\":\"basic_textfield\",\"lib\":\"generic2\",\"caption\":\"Text Field 14\",\"width\":338,\"height\":55,\"value\":\"Morphine (mg)\",\"placeholder\":\"Placeholder\",\"inputtype\":\"text\",\"backgroundColor\":{\"type\":\"solid\",\"color\":\"rgba(101,180,206,1)\"},\"textFont\":\"Arial,sans-serif\",\"textFontStyle\":\"400\",\"textSize\":\"28\",\"textColor\":{\"type\":\"solid\",\"color\":\"rgba(56,56,56,1)\"},\"textProperties\":[],\"placeholderColor\":\"c1c8c9\",\"borderWidth\":1,\"borderStyle\":\"solid\",\"borderRadius\":4,\"borderColor\":{\"type\":\"solid\",\"color\":\"rgba(27,77,87,1)\"},\"textAlign\":\"center\",\"left\":31,\"top\":458,\"vpos\":\"top\",\"hpos\":\"left\",\"visible\":true,\"group\":\"root\",\"id\":\"box-166\",\"zindex\":10,\"wtype\":\"fixed\",\"htype\":\"fixed\",\"opacity\":1,\"rotation\":0,\"scale\":1,\"aspectratio\":false,\"actions\":[{\"title\":\"click \",\"type\":\"click\",\"actionId\":\"change-property\",\"callback\":true,\"pageId\":\"2\",\"itemId\":\"box-78\",\"propertyId\":\"value\",\"valueType\":\"customValue\",\"newvalue\":\"Morphine (mg)\",\"delay\":\"0\",\"guid\":\"cf0f1897-aa38-4a3a-ae25-223469ab0ee5\",\"callbackAction\":{\"actionId\":\"go-to-page\",\"callback\":false,\"pageId\":\"2\",\"animation\":\"fade\",\"animationEasing\":\"Cubic.easeOut\",\"duration\":\"400\",\"delay\":\"0\",\"guid\":\"4c7ab12d-5c3d-491c-8d1b-1d75761e0dec\",\"clb_disabled\":false},\"sort\":0}]},{\"name\":\"image\",\"type\":\"image\",\"lib\":\"common\",\"caption\":\"Screen Shot 2022-12-12 at 7.38.12 PM.png\",\"imgSrc\":{\"fileId\":\"68d047acd614373a75ca2548bba92b44.png\",\"assetType\":\"project\",\"bucketsource\":\"main\",\"name\":\"Screen Shot 2022-12-12 at 7.38.12 PM.png\"},\"repeat\":false,\"width\":658,\"height\":398,\"borderWidth\":0,\"borderRadius\":0,\"borderColor\":\"#D1D1D1\",\"borderPos\":\"inside\",\"propagateEvents\":false,\"actions\":[],\"overlay\":false,\"autoResize\":false,\"aspectratio\":1.6506849315068493,\"assistiveDescription\":\"\",\"left\":762,\"top\":359,\"visible\":true,\"id\":\"box-122\",\"group\":\"root\",\"zindex\":12,\"hpos\":\"left\",\"vpos\":\"top\",\"wtype\":\"fixed\",\"htype\":\"fixed\",\"opacity\":1,\"borderStyle\":\"solid\",\"rotation\":0,\"scale\":1},{\"name\":\"basic_button_text\",\"type\":\"basic_button_text\",\"lib\":\"generic2\",\"caption\":\"Button with Background 5\",\"width\":177,\"height\":80,\"backgroundColor\":{\"type\":\"solid\",\"color\":\"rgba(249,51,36,1)\"},\"borderWidth\":0,\"borderColor\":\"#025abf\",\"borderRadius\":4,\"text\":\"Back\",\"textFont\":\"Arial,sans-serif\",\"textFontStyle\":\"400\",\"textSize\":\"36\",\"textColor\":\"FFFFFF\",\"textProperties\":[],\"textAlign\":\"center\",\"iconpos\":\"none\",\"left\":472,\"top\":652,\"vpos\":\"top\",\"hpos\":\"left\",\"visible\":true,\"id\":\"box-168\",\"group\":\"root\",\"zindex\":13,\"wtype\":\"fixed\",\"htype\":\"fixed\",\"opacity\":1,\"rotation\":0,\"scale\":1,\"aspectratio\":false,\"actions\":[{\"title\":\"Interaction 1\",\"type\":\"click\",\"actionId\":\"go-to-page\",\"callback\":false,\"pageId\":\"2\",\"animation\":\"fade\",\"animationEasing\":\"Cubic.easeOut\",\"duration\":\"400\",\"delay\":\"0\",\"guid\":\"5462dd60-9d70-4dd8-a4dc-97f8bbeea671\",\"sort\":0}]},{\"name\":\"basic_textfield\",\"type\":\"basic_textfield\",\"lib\":\"generic2\",\"caption\":\"Text Field 15\",\"width\":338,\"height\":55,\"value\":\"Sufentanyl (mcg)\",\"placeholder\":\"Placeholder\",\"inputtype\":\"text\",\"backgroundColor\":{\"type\":\"solid\",\"color\":\"rgba(101,180,206,1)\"},\"textFont\":\"Arial,sans-serif\",\"textFontStyle\":\"400\",\"textSize\":\"28\",\"textColor\":{\"type\":\"solid\",\"color\":\"rgba(56,56,56,1)\"},\"textProperties\":[],\"placeholderColor\":\"c1c8c9\",\"borderWidth\":1,\"borderStyle\":\"solid\",\"borderRadius\":4,\"borderColor\":{\"type\":\"solid\",\"color\":\"rgba(27,77,87,1)\"},\"textAlign\":\"center\",\"left\":32,\"top\":543,\"vpos\":\"top\",\"hpos\":\"left\",\"visible\":true,\"group\":\"root\",\"id\":\"box-167\",\"zindex\":10,\"wtype\":\"fixed\",\"htype\":\"fixed\",\"opacity\":1,\"rotation\":0,\"scale\":1,\"aspectratio\":false,\"actions\":[{\"title\":\"click \",\"type\":\"click\",\"actionId\":\"change-property\",\"callback\":true,\"pageId\":\"2\",\"itemId\":\"box-78\",\"propertyId\":\"value\",\"valueType\":\"customValue\",\"newvalue\":\"Sufentanyl (mg)\",\"delay\":\"0\",\"guid\":\"ad66c836-a092-4add-b6af-af49db605d31\",\"callbackAction\":{\"actionId\":\"go-to-page\",\"callback\":false,\"pageId\":\"2\",\"animation\":\"fade\",\"animationEasing\":\"Cubic.easeOut\",\"duration\":\"400\",\"delay\":\"0\",\"guid\":\"f1f70443-b2e3-4a16-ac8b-aa087e8a9113\",\"clb_disabled\":false},\"sort\":0}]},{\"name\":\"basic_textfield\",\"type\":\"basic_textfield\",\"lib\":\"generic2\",\"caption\":\"Text Field 16\",\"width\":338,\"height\":55,\"value\":\"Fentanyl (mcg)\",\"placeholder\":\"Placeholder\",\"inputtype\":\"text\",\"backgroundColor\":{\"type\":\"solid\",\"color\":\"rgba(101,180,206,1)\"},\"textFont\":\"Arial,sans-serif\",\"textFontStyle\":\"400\",\"textSize\":\"28\",\"textColor\":{\"type\":\"solid\",\"color\":\"rgba(56,56,56,1)\"},\"textProperties\":[],\"placeholderColor\":\"c1c8c9\",\"borderWidth\":1,\"borderStyle\":\"solid\",\"borderRadius\":4,\"borderColor\":{\"type\":\"solid\",\"color\":\"rgba(27,77,87,1)\"},\"textAlign\":\"center\",\"left\":40,\"top\":637,\"vpos\":\"top\",\"hpos\":\"left\",\"visible\":true,\"group\":\"root\",\"id\":\"box-221\",\"zindex\":10,\"wtype\":\"fixed\",\"htype\":\"fixed\",\"opacity\":1,\"rotation\":0,\"scale\":1,\"aspectratio\":false,\"actions\":[{\"title\":\"click \",\"type\":\"click\",\"actionId\":\"change-property\",\"callback\":true,\"pageId\":\"2\",\"itemId\":\"box-78\",\"propertyId\":\"value\",\"valueType\":\"customValue\",\"newvalue\":\"Fentanyl (mg)\",\"delay\":\"0\",\"guid\":\"23621eef-4760-445e-b746-c13acb1e5cd3\",\"callbackAction\":{\"actionId\":\"go-to-page\",\"callback\":false,\"pageId\":\"2\",\"animation\":\"fade\",\"animationEasing\":\"Cubic.easeOut\",\"duration\":\"400\",\"delay\":\"0\",\"guid\":\"c002a994-03b7-4332-856f-f0a2332fe0e1\",\"clb_disabled\":false},\"sort\":0}]}]","template":{"id":"","stateid":""},"history":[],"id":"e59b7eb2-bf3b-4228-b34e-61c88119d743","actions":[],"groups":{}}]},{"id":6,"title":"Pressor","orientation":"landscape","statusbar":"0","navigationbar":"0","actions":[],"states":[{"title":"State 1","background":{"type":"solid","color":"rgba(169,151,196,1)"},"data":"[{\"name\":\"rectangle\",\"type\":\"rectangle\",\"lib\":\"common\",\"caption\":\"Rectangle 1\",\"backgroundColor\":{\"type\":\"solid\",\"color\":\"rgba(193,212,241,1)\"},\"borderWidth\":\"8\",\"borderColor\":{\"type\":\"solid\",\"color\":\"rgba(30,77,130,1)\"},\"borderRadius\":0,\"imgSrc\":{\"fileId\":\"\",\"name\":\"\",\"assetType\":\"\",\"bucketsource\":\"\",\"url\":\"\"},\"backgroundSizeType\":\"cover\",\"backgroundSizeValue\":35,\"width\":1440,\"height\":249,\"lineHeight\":20,\"lineHeightAuto\":true,\"actions\":[],\"textFont\":\"Arial,sans-serif\",\"textFontStyle\":\"400\",\"textSize\":16,\"textColor\":\"#383838\",\"textProperties\":[],\"textSpacing\":0,\"textAlign\":\"center\",\"text\":\"\",\"typeName\":\"rectangle\",\"left\":0,\"top\":0,\"vpos\":\"top\",\"hpos\":\"left\",\"visible\":true,\"id\":\"box-170\",\"group\":\"root\",\"zindex\":1,\"wtype\":\"fixed\",\"htype\":\"fixed\",\"opacity\":1,\"borderStyle\":\"solid\",\"rotation\":0,\"scale\":1,\"aspectratio\":false},{\"name\":\"text\",\"type\":\"text\",\"lib\":\"common\",\"caption\":\"Gender : Male...\",\"text\":\"Gender : Male\",\"textFont\":\"Trebuchet MS,sans-serif\",\"textFontStyle\":\"400\",\"textProperties\":[],\"textSize\":\"32\",\"textColor\":{\"type\":\"solid\",\"color\":\"rgba(113,113,113,1)\"},\"backgroundColor\":\"none\",\"width\":186,\"height\":39,\"lineHeight\":39,\"textAlign\":\"left\",\"textSpacing\":\"-1.6\",\"enableShadow\":false,\"autoResize\":true,\"v2\":true,\"left\":379,\"top\":47,\"vpos\":\"top\",\"hpos\":\"left\",\"visible\":true,\"group\":\"root\",\"id\":\"box-171\",\"zindex\":8,\"wtype\":\"fixed\",\"htype\":\"fixed\",\"opacity\":1,\"propagateEvents\":false,\"lineHeightAuto\":true,\"rotation\":0,\"scale\":1,\"aspectratio\":false},{\"name\":\"text\",\"type\":\"text\",\"lib\":\"common\",\"caption\":\"DOB: 01/01/2022...\",\"text\":\"DOB: 01/01/2022\",\"textFont\":\"Trebuchet MS,sans-serif\",\"textFontStyle\":\"400\",\"textProperties\":[],\"textSize\":\"32\",\"textColor\":{\"type\":\"solid\",\"color\":\"rgba(113,113,113,1)\"},\"backgroundColor\":\"none\",\"width\":230,\"height\":39,\"lineHeight\":39,\"textAlign\":\"left\",\"textSpacing\":\"-1.6\",\"enableShadow\":false,\"autoResize\":true,\"v2\":true,\"left\":76,\"top\":130,\"vpos\":\"top\",\"hpos\":\"left\",\"visible\":true,\"group\":\"root\",\"id\":\"box-172\",\"zindex\":8,\"wtype\":\"fixed\",\"htype\":\"fixed\",\"opacity\":1,\"propagateEvents\":false,\"lineHeightAuto\":true,\"rotation\":0,\"scale\":1,\"aspectratio\":false},{\"name\":\"text\",\"type\":\"text\",\"lib\":\"common\",\"caption\":\"Height : 174 cm...\",\"text\":\"Height : 174 cm&nbsp;\",\"textFont\":\"Trebuchet MS,sans-serif\",\"textFontStyle\":\"400\",\"textProperties\":[],\"textSize\":\"32\",\"textColor\":{\"type\":\"solid\",\"color\":\"rgba(113,113,113,1)\"},\"backgroundColor\":\"none\",\"width\":216,\"height\":39,\"lineHeight\":39,\"textAlign\":\"left\",\"textSpacing\":\"-1.6\",\"enableShadow\":false,\"autoResize\":true,\"v2\":true,\"left\":379,\"top\":86,\"vpos\":\"top\",\"hpos\":\"left\",\"visible\":true,\"group\":\"root\",\"id\":\"box-173\",\"zindex\":8,\"wtype\":\"fixed\",\"htype\":\"fixed\",\"opacity\":1,\"propagateEvents\":false,\"lineHeightAuto\":true,\"rotation\":0,\"scale\":1,\"aspectratio\":false},{\"name\":\"text\",\"type\":\"text\",\"lib\":\"common\",\"caption\":\"Surgery:\\nTotal Colec...\",\"text\":\"Surgery:&nbsp;<br>Total Colectomy with&nbsp;<br>pancreatico-jejunostomy&nbsp;\",\"textFont\":\"Trebuchet MS,sans-serif\",\"textFontStyle\":\"400\",\"textProperties\":[],\"textSize\":\"32\",\"textColor\":{\"type\":\"solid\",\"color\":\"rgba(113,113,113,1)\"},\"backgroundColor\":\"none\",\"width\":336,\"height\":117,\"lineHeight\":39,\"textAlign\":\"left\",\"textSpacing\":\"-1.6\",\"enableShadow\":false,\"autoResize\":true,\"v2\":true,\"left\":1084,\"top\":47,\"vpos\":\"top\",\"hpos\":\"left\",\"visible\":true,\"group\":\"root\",\"id\":\"box-174\",\"zindex\":8,\"wtype\":\"fixed\",\"htype\":\"fixed\",\"opacity\":1,\"propagateEvents\":false,\"lineHeightAuto\":true,\"rotation\":0,\"scale\":1,\"aspectratio\":false},{\"name\":\"text\",\"type\":\"text\",\"lib\":\"common\",\"caption\":\"Age : 86 y\",\"text\":\"Age : 86 y&nbsp;\",\"textFont\":\"Trebuchet MS,sans-serif\",\"textFontStyle\":\"400\",\"textProperties\":[],\"textSize\":\"32\",\"textColor\":{\"type\":\"solid\",\"color\":\"rgba(113,113,113,1)\"},\"backgroundColor\":\"none\",\"width\":139,\"height\":39,\"lineHeight\":39,\"textAlign\":\"left\",\"textSpacing\":\"-1.6\",\"enableShadow\":false,\"autoResize\":true,\"v2\":true,\"left\":76,\"top\":91,\"vpos\":\"top\",\"hpos\":\"left\",\"visible\":true,\"group\":\"root\",\"id\":\"box-175\",\"zindex\":8,\"wtype\":\"fixed\",\"htype\":\"fixed\",\"opacity\":1,\"propagateEvents\":false,\"lineHeightAuto\":true,\"rotation\":0,\"scale\":1,\"aspectratio\":false},{\"name\":\"text\",\"type\":\"text\",\"lib\":\"common\",\"caption\":\"Allergies :\\nPenicill...\",\"text\":\"Allergies :<br><span style=\\\"color: rgba(249,51,36,1);\\\">Penicillin</span><br><br>\",\"textFont\":\"Trebuchet MS,sans-serif\",\"textFontStyle\":\"400\",\"textProperties\":[],\"textSize\":\"32\",\"textColor\":{\"type\":\"solid\",\"color\":\"rgba(113,113,113,1)\"},\"backgroundColor\":\"none\",\"width\":203,\"height\":117,\"lineHeight\":39,\"textAlign\":\"left\",\"textSpacing\":\"-1.6\",\"enableShadow\":false,\"autoResize\":false,\"v2\":true,\"left\":766,\"top\":47,\"vpos\":\"top\",\"hpos\":\"left\",\"visible\":true,\"group\":\"root\",\"id\":\"box-176\",\"zindex\":8,\"wtype\":\"fixed\",\"htype\":\"fixed\",\"opacity\":1,\"propagateEvents\":false,\"lineHeightAuto\":true,\"rotation\":0,\"scale\":1,\"aspectratio\":false},{\"name\":\"text\",\"type\":\"text\",\"lib\":\"common\",\"caption\":\"BMI : 28 kg/m2...\",\"text\":\"BMI : 28 kg/m2\",\"textFont\":\"Trebuchet MS,sans-serif\",\"textFontStyle\":\"400\",\"textProperties\":[],\"textSize\":\"32\",\"textColor\":{\"type\":\"solid\",\"color\":\"rgba(113,113,113,1)\"},\"backgroundColor\":\"none\",\"width\":199,\"height\":39,\"lineHeight\":39,\"textAlign\":\"left\",\"textSpacing\":\"-1.6\",\"enableShadow\":false,\"autoResize\":true,\"v2\":true,\"left\":379,\"top\":171,\"vpos\":\"top\",\"hpos\":\"left\",\"visible\":true,\"group\":\"root\",\"id\":\"box-177\",\"zindex\":8,\"wtype\":\"fixed\",\"htype\":\"fixed\",\"opacity\":1,\"propagateEvents\":false,\"lineHeightAuto\":true,\"rotation\":0,\"scale\":1,\"aspectratio\":false},{\"name\":\"text\",\"type\":\"text\",\"lib\":\"common\",\"caption\":\"Weight : 69 Kg...\",\"text\":\"Weight : 69 Kg\",\"textFont\":\"Trebuchet MS,sans-serif\",\"textFontStyle\":\"400\",\"textProperties\":[],\"textSize\":\"32\",\"textColor\":{\"type\":\"solid\",\"color\":\"rgba(113,113,113,1)\"},\"backgroundColor\":\"none\",\"width\":190,\"height\":39,\"lineHeight\":39,\"textAlign\":\"left\",\"textSpacing\":\"-1.6\",\"enableShadow\":false,\"autoResize\":true,\"v2\":true,\"left\":379,\"top\":130,\"vpos\":\"top\",\"hpos\":\"left\",\"visible\":true,\"group\":\"root\",\"id\":\"box-178\",\"zindex\":8,\"wtype\":\"fixed\",\"htype\":\"fixed\",\"opacity\":1,\"propagateEvents\":false,\"lineHeightAuto\":true,\"rotation\":0,\"scale\":1,\"aspectratio\":false},{\"name\":\"verticalline\",\"type\":\"verticalline\",\"lib\":\"common\",\"caption\":\"Vertical line 3\",\"width\":1,\"height\":229,\"color\":\"A6A6A6\",\"weight\":1,\"left\":341,\"top\":10,\"vpos\":\"top\",\"hpos\":\"left\",\"visible\":true,\"group\":\"root\",\"id\":\"box-179\",\"zindex\":7,\"wtype\":\"fixed\",\"htype\":\"fixed\",\"opacity\":1,\"borderStyle\":\"solid\",\"rotation\":0,\"scale\":1,\"aspectratio\":false},{\"name\":\"verticalline\",\"type\":\"verticalline\",\"lib\":\"common\",\"caption\":\"Vertical line 5\",\"width\":1,\"height\":229,\"color\":\"A6A6A6\",\"weight\":1,\"left\":1064,\"top\":15,\"vpos\":\"top\",\"hpos\":\"left\",\"visible\":true,\"group\":\"root\",\"id\":\"box-180\",\"zindex\":7,\"wtype\":\"fixed\",\"htype\":\"fixed\",\"opacity\":1,\"borderStyle\":\"solid\",\"rotation\":0,\"scale\":1,\"aspectratio\":false},{\"name\":\"verticalline\",\"type\":\"verticalline\",\"lib\":\"common\",\"caption\":\"Vertical line 6\",\"width\":1,\"height\":229,\"color\":\"A6A6A6\",\"weight\":1,\"left\":733,\"top\":10,\"vpos\":\"top\",\"hpos\":\"left\",\"visible\":true,\"group\":\"root\",\"id\":\"box-181\",\"zindex\":7,\"wtype\":\"fixed\",\"htype\":\"fixed\",\"opacity\":1,\"borderStyle\":\"solid\",\"rotation\":0,\"scale\":1,\"aspectratio\":false},{\"name\":\"datetime\",\"type\":\"datetime\",\"lib\":\"common\",\"caption\":\"Date/Time 1\",\"width\":376,\"height\":41,\"backgroundColor\":\"none\",\"borderWidth\":\"1\",\"borderColor\":{\"type\":\"solid\",\"color\":\"rgba(102,150,200,1)\"},\"textFont\":\"Montserrat\",\"textFontStyle\":\"400\",\"textSize\":\"20\",\"textColor\":\"383838\",\"textProperties\":[],\"textAlign\":\"center\",\"dateFormat\":\"yearmonthday\",\"dayFormat\":\"superscript\",\"monthFormat\":\"long\",\"yearFormat\":\"long\",\"timeFormat\":\"24\",\"dateGeneralFormat\":\"timedate\",\"dayNameFormat\":\"long\",\"dateSeparator\":\"-\",\"left\":1064,\"top\":201,\"vpos\":\"top\",\"hpos\":\"left\",\"visible\":true,\"group\":\"root\",\"id\":\"box-182\",\"zindex\":5,\"wtype\":\"fixed\",\"htype\":\"fixed\",\"opacity\":1,\"rotation\":0,\"scale\":1,\"aspectratio\":false},{\"name\":\"text\",\"type\":\"text\",\"lib\":\"common\",\"caption\":\"MRN : 12345679...\",\"text\":\"MRN : 12345679&nbsp;\",\"textFont\":\"Trebuchet MS,sans-serif\",\"textFontStyle\":\"400\",\"textProperties\":[],\"textSize\":\"32\",\"textColor\":{\"type\":\"solid\",\"color\":\"rgba(113,113,113,1)\"},\"backgroundColor\":\"none\",\"width\":218,\"height\":39,\"lineHeight\":39,\"textAlign\":\"left\",\"textSpacing\":\"-1.6\",\"enableShadow\":false,\"autoResize\":true,\"v2\":true,\"left\":76,\"top\":171,\"vpos\":\"top\",\"hpos\":\"left\",\"visible\":true,\"group\":\"root\",\"id\":\"box-183\",\"zindex\":4,\"wtype\":\"fixed\",\"htype\":\"fixed\",\"opacity\":1,\"propagateEvents\":false,\"lineHeightAuto\":true,\"rotation\":0,\"scale\":1,\"aspectratio\":false},{\"name\":\"verticalline\",\"type\":\"verticalline\",\"lib\":\"common\",\"caption\":\"Vertical line 1\",\"width\":1,\"height\":658,\"color\":{\"type\":\"solid\",\"color\":\"rgba(70,127,215,1)\"},\"weight\":1,\"left\":734,\"top\":242,\"vpos\":\"top\",\"hpos\":\"left\",\"visible\":true,\"group\":\"root\",\"id\":\"box-184\",\"zindex\":3,\"wtype\":\"fixed\",\"htype\":\"fixed\",\"opacity\":1,\"borderStyle\":\"solid\",\"rotation\":0,\"scale\":1,\"aspectratio\":false},{\"name\":\"text\",\"type\":\"text\",\"lib\":\"common\",\"caption\":\"John Doe\",\"text\":\"John Doe&nbsp;\",\"textFont\":\"Trebuchet MS,sans-serif\",\"textFontStyle\":\"400\",\"textProperties\":[],\"textSize\":\"64\",\"textColor\":{\"type\":\"solid\",\"color\":\"rgba(117,85,88,1)\"},\"backgroundColor\":\"none\",\"width\":272,\"height\":78,\"lineHeight\":78,\"textAlign\":\"left\",\"textSpacing\":\"-1.6\",\"enableShadow\":false,\"autoResize\":true,\"v2\":true,\"left\":40,\"top\":15,\"vpos\":\"top\",\"hpos\":\"left\",\"visible\":true,\"id\":\"box-185\",\"group\":\"root\",\"zindex\":2,\"wtype\":\"fixed\",\"htype\":\"fixed\",\"opacity\":1,\"propagateEvents\":false,\"lineHeightAuto\":true,\"rotation\":0,\"scale\":1,\"aspectratio\":false},{\"name\":\"basic_textfield\",\"type\":\"basic_textfield\",\"lib\":\"generic2\",\"caption\":\"Text Field 2\",\"width\":733,\"height\":77,\"value\":\"Induction Medication\",\"placeholder\":\"Placeholder\",\"inputtype\":\"text\",\"backgroundColor\":{\"type\":\"solid\",\"color\":\"rgba(177,190,227,1)\"},\"textFont\":\"Arial,sans-serif\",\"textFontStyle\":\"400\",\"textSize\":\"64\",\"textColor\":{\"type\":\"solid\",\"color\":\"rgba(233,240,215,1)\"},\"textProperties\":[],\"placeholderColor\":\"c1c8c9\",\"borderWidth\":\"5\",\"borderStyle\":\"solid\",\"borderRadius\":4,\"borderColor\":{\"type\":\"solid\",\"color\":\"rgba(85,85,85,1)\"},\"textAlign\":\"center\",\"left\":2,\"top\":242,\"vpos\":\"top\",\"hpos\":\"left\",\"visible\":true,\"id\":\"box-186\",\"group\":\"root\",\"zindex\":11,\"wtype\":\"fixed\",\"htype\":\"fixed\",\"opacity\":1,\"rotation\":0,\"scale\":1,\"aspectratio\":false},{\"name\":\"basic_textfield\",\"type\":\"basic_textfield\",\"lib\":\"generic2\",\"caption\":\"Text Field 3\",\"width\":698,\"height\":77,\"value\":\"Blood Pressure\",\"placeholder\":\"Placeholder\",\"inputtype\":\"text\",\"backgroundColor\":{\"type\":\"solid\",\"color\":\"rgba(177,190,227,1)\"},\"textFont\":\"Arial,sans-serif\",\"textFontStyle\":\"400\",\"textSize\":\"64\",\"textColor\":{\"type\":\"solid\",\"color\":\"rgba(233,240,215,1)\"},\"textProperties\":[],\"placeholderColor\":\"c1c8c9\",\"borderWidth\":\"5\",\"borderStyle\":\"solid\",\"borderRadius\":4,\"borderColor\":{\"type\":\"solid\",\"color\":\"rgba(85,85,85,1)\"},\"textAlign\":\"center\",\"left\":733,\"top\":244,\"vpos\":\"top\",\"hpos\":\"left\",\"visible\":true,\"id\":\"box-187\",\"group\":\"root\",\"zindex\":11,\"wtype\":\"fixed\",\"htype\":\"fixed\",\"opacity\":1,\"rotation\":0,\"scale\":1,\"aspectratio\":false},{\"name\":\"basic_textfield\",\"type\":\"basic_textfield\",\"lib\":\"generic2\",\"caption\":\"Text Field 13\",\"width\":338,\"height\":55,\"value\":\"Ephedrine (mg)\",\"placeholder\":\"Placeholder\",\"inputtype\":\"text\",\"backgroundColor\":{\"type\":\"solid\",\"color\":\"rgba(234,114,252,1)\"},\"textFont\":\"Arial,sans-serif\",\"textFontStyle\":\"400\",\"textSize\":\"28\",\"textColor\":{\"type\":\"solid\",\"color\":\"rgba(56,56,56,1)\"},\"textProperties\":[],\"placeholderColor\":\"c1c8c9\",\"borderWidth\":1,\"borderStyle\":\"solid\",\"borderRadius\":4,\"borderColor\":{\"type\":\"solid\",\"color\":\"rgba(27,77,87,1)\"},\"textAlign\":\"center\",\"left\":31,\"top\":374,\"vpos\":\"top\",\"hpos\":\"left\",\"visible\":true,\"group\":\"root\",\"id\":\"box-188\",\"zindex\":10,\"wtype\":\"fixed\",\"htype\":\"fixed\",\"opacity\":1,\"rotation\":0,\"scale\":1,\"aspectratio\":false,\"actions\":[{\"title\":\"click \",\"type\":\"click\",\"actionId\":\"change-property\",\"callback\":true,\"pageId\":\"2\",\"itemId\":\"box-73\",\"propertyId\":\"value\",\"valueType\":\"customValue\",\"newvalue\":\"Ephedrine (mg)\",\"delay\":\"0\",\"guid\":\"09d8ca82-c4bb-4046-9cd6-6b55263c675c\",\"callbackAction\":{\"actionId\":\"go-to-page\",\"callback\":false,\"pageId\":\"2\",\"animation\":\"fade\",\"animationEasing\":\"Cubic.easeOut\",\"duration\":\"400\",\"delay\":\"0\",\"guid\":\"c6a1af57-3859-487f-a0f2-10f68fb6c874\",\"clb_disabled\":false},\"sort\":0}]},{\"name\":\"basic_textfield\",\"type\":\"basic_textfield\",\"lib\":\"generic2\",\"caption\":\"Text Field 14\",\"width\":338,\"height\":55,\"value\":\"Norepinephrine (mcg)\",\"placeholder\":\"Placeholder\",\"inputtype\":\"text\",\"backgroundColor\":{\"type\":\"solid\",\"color\":\"rgba(234,114,252,1)\"},\"textFont\":\"Arial,sans-serif\",\"textFontStyle\":\"400\",\"textSize\":\"28\",\"textColor\":{\"type\":\"solid\",\"color\":\"rgba(56,56,56,1)\"},\"textProperties\":[],\"placeholderColor\":\"c1c8c9\",\"borderWidth\":1,\"borderStyle\":\"solid\",\"borderRadius\":4,\"borderColor\":{\"type\":\"solid\",\"color\":\"rgba(27,77,87,1)\"},\"textAlign\":\"center\",\"left\":31,\"top\":458,\"vpos\":\"top\",\"hpos\":\"left\",\"visible\":true,\"group\":\"root\",\"id\":\"box-189\",\"zindex\":10,\"wtype\":\"fixed\",\"htype\":\"fixed\",\"opacity\":1,\"rotation\":0,\"scale\":1,\"aspectratio\":false,\"actions\":[{\"title\":\"click \",\"type\":\"click\",\"actionId\":\"change-property\",\"callback\":true,\"pageId\":\"2\",\"itemId\":\"box-73\",\"propertyId\":\"value\",\"valueType\":\"customValue\",\"newvalue\":\"Norepinephine (mcg)\",\"delay\":\"0\",\"guid\":\"62667815-6c21-41aa-869d-3cbf9270d976\",\"callbackAction\":{\"actionId\":\"go-to-page\",\"callback\":false,\"pageId\":\"2\",\"animation\":\"fade\",\"animationEasing\":\"Cubic.easeOut\",\"duration\":\"400\",\"delay\":\"0\",\"guid\":\"4a53e949-1453-4eeb-9be4-7c32b0d093fe\",\"clb_disabled\":false},\"sort\":0}]},{\"name\":\"basic_textfield\",\"type\":\"basic_textfield\",\"lib\":\"generic2\",\"caption\":\"Text Field 15\",\"width\":338,\"height\":55,\"value\":\"Phenylephrine (mcg)\",\"placeholder\":\"Placeholder\",\"inputtype\":\"text\",\"backgroundColor\":{\"type\":\"solid\",\"color\":\"rgba(234,114,252,1)\"},\"textFont\":\"Arial,sans-serif\",\"textFontStyle\":\"400\",\"textSize\":\"28\",\"textColor\":{\"type\":\"solid\",\"color\":\"rgba(56,56,56,1)\"},\"textProperties\":[],\"placeholderColor\":\"c1c8c9\",\"borderWidth\":1,\"borderStyle\":\"solid\",\"borderRadius\":4,\"borderColor\":{\"type\":\"solid\",\"color\":\"rgba(27,77,87,1)\"},\"textAlign\":\"center\",\"left\":40,\"top\":543,\"vpos\":\"top\",\"hpos\":\"left\",\"visible\":true,\"group\":\"root\",\"id\":\"box-222\",\"zindex\":10,\"wtype\":\"fixed\",\"htype\":\"fixed\",\"opacity\":1,\"rotation\":0,\"scale\":1,\"aspectratio\":false,\"actions\":[{\"title\":\"click \",\"type\":\"click\",\"actionId\":\"change-property\",\"callback\":true,\"pageId\":\"2\",\"itemId\":\"box-73\",\"propertyId\":\"value\",\"valueType\":\"customValue\",\"newvalue\":\"Phenylephrine (mcg)\",\"delay\":\"0\",\"guid\":\"82772c85-ac05-41a2-8997-e36a3cab85e6\",\"callbackAction\":{\"actionId\":\"go-to-page\",\"callback\":false,\"pageId\":\"2\",\"animation\":\"fade\",\"animationEasing\":\"Cubic.easeOut\",\"duration\":\"400\",\"delay\":\"0\",\"guid\":\"b2ecddaf-d786-4cd7-8f28-69df01c9a742\",\"clb_disabled\":false},\"sort\":0}]},{\"name\":\"image\",\"type\":\"image\",\"lib\":\"common\",\"caption\":\"Screen Shot 2022-12-12 at 7.38.12 PM.png\",\"imgSrc\":{\"fileId\":\"68d047acd614373a75ca2548bba92b44.png\",\"assetType\":\"project\",\"bucketsource\":\"main\",\"name\":\"Screen Shot 2022-12-12 at 7.38.12 PM.png\"},\"repeat\":false,\"width\":658,\"height\":398,\"borderWidth\":0,\"borderRadius\":0,\"borderColor\":\"#D1D1D1\",\"borderPos\":\"inside\",\"propagateEvents\":false,\"actions\":[],\"overlay\":false,\"autoResize\":false,\"aspectratio\":1.6506849315068493,\"assistiveDescription\":\"\",\"left\":762,\"top\":359,\"visible\":true,\"id\":\"box-190\",\"group\":\"root\",\"zindex\":12,\"hpos\":\"left\",\"vpos\":\"top\",\"wtype\":\"fixed\",\"htype\":\"fixed\",\"opacity\":1,\"borderStyle\":\"solid\",\"rotation\":0,\"scale\":1},{\"name\":\"basic_button_text\",\"type\":\"basic_button_text\",\"lib\":\"generic2\",\"caption\":\"Button with Background 5\",\"width\":177,\"height\":80,\"backgroundColor\":{\"type\":\"solid\",\"color\":\"rgba(249,51,36,1)\"},\"borderWidth\":0,\"borderColor\":\"#025abf\",\"borderRadius\":4,\"text\":\"Back\",\"textFont\":\"Arial,sans-serif\",\"textFontStyle\":\"400\",\"textSize\":\"36\",\"textColor\":\"FFFFFF\",\"textProperties\":[],\"textAlign\":\"center\",\"iconpos\":\"none\",\"left\":472,\"top\":652,\"vpos\":\"top\",\"hpos\":\"left\",\"visible\":true,\"id\":\"box-191\",\"group\":\"root\",\"zindex\":13,\"wtype\":\"fixed\",\"htype\":\"fixed\",\"opacity\":1,\"rotation\":0,\"scale\":1,\"aspectratio\":false,\"actions\":[{\"title\":\"Interaction 1\",\"type\":\"click\",\"actionId\":\"go-to-page\",\"callback\":false,\"pageId\":\"2\",\"animation\":\"fade\",\"animationEasing\":\"Cubic.easeOut\",\"duration\":\"400\",\"delay\":\"0\",\"guid\":\"4dc61238-d8e0-40c2-a2c2-3758742ba100\",\"sort\":0}]}]","template":{"id":"","stateid":""},"history":[],"id":"6fb2a64b-1351-4eb2-9442-19e2a6110a8b","actions":[],"groups":{}}]},{"id":7,"title":"Amnestics","orientation":"landscape","statusbar":"0","navigationbar":"0","actions":[],"states":[{"title":"State 1","background":{"type":"solid","color":"rgba(169,151,196,1)"},"data":"[{\"name\":\"rectangle\",\"type\":\"rectangle\",\"lib\":\"common\",\"caption\":\"Rectangle 1\",\"backgroundColor\":{\"type\":\"solid\",\"color\":\"rgba(193,212,241,1)\"},\"borderWidth\":\"8\",\"borderColor\":{\"type\":\"solid\",\"color\":\"rgba(30,77,130,1)\"},\"borderRadius\":0,\"imgSrc\":{\"fileId\":\"\",\"name\":\"\",\"assetType\":\"\",\"bucketsource\":\"\",\"url\":\"\"},\"backgroundSizeType\":\"cover\",\"backgroundSizeValue\":35,\"width\":1440,\"height\":249,\"lineHeight\":20,\"lineHeightAuto\":true,\"actions\":[],\"textFont\":\"Arial,sans-serif\",\"textFontStyle\":\"400\",\"textSize\":16,\"textColor\":\"#383838\",\"textProperties\":[],\"textSpacing\":0,\"textAlign\":\"center\",\"text\":\"\",\"typeName\":\"rectangle\",\"left\":0,\"top\":0,\"vpos\":\"top\",\"hpos\":\"left\",\"visible\":true,\"id\":\"box-193\",\"group\":\"root\",\"zindex\":1,\"wtype\":\"fixed\",\"htype\":\"fixed\",\"opacity\":1,\"borderStyle\":\"solid\",\"rotation\":0,\"scale\":1,\"aspectratio\":false},{\"name\":\"text\",\"type\":\"text\",\"lib\":\"common\",\"caption\":\"Gender : Male...\",\"text\":\"Gender : Male\",\"textFont\":\"Trebuchet MS,sans-serif\",\"textFontStyle\":\"400\",\"textProperties\":[],\"textSize\":\"32\",\"textColor\":{\"type\":\"solid\",\"color\":\"rgba(113,113,113,1)\"},\"backgroundColor\":\"none\",\"width\":186,\"height\":39,\"lineHeight\":39,\"textAlign\":\"left\",\"textSpacing\":\"-1.6\",\"enableShadow\":false,\"autoResize\":true,\"v2\":true,\"left\":379,\"top\":47,\"vpos\":\"top\",\"hpos\":\"left\",\"visible\":true,\"group\":\"root\",\"id\":\"box-194\",\"zindex\":8,\"wtype\":\"fixed\",\"htype\":\"fixed\",\"opacity\":1,\"propagateEvents\":false,\"lineHeightAuto\":true,\"rotation\":0,\"scale\":1,\"aspectratio\":false},{\"name\":\"text\",\"type\":\"text\",\"lib\":\"common\",\"caption\":\"DOB: 01/01/2022...\",\"text\":\"DOB: 01/01/2022\",\"textFont\":\"Trebuchet MS,sans-serif\",\"textFontStyle\":\"400\",\"textProperties\":[],\"textSize\":\"32\",\"textColor\":{\"type\":\"solid\",\"color\":\"rgba(113,113,113,1)\"},\"backgroundColor\":\"none\",\"width\":230,\"height\":39,\"lineHeight\":39,\"textAlign\":\"left\",\"textSpacing\":\"-1.6\",\"enableShadow\":false,\"autoResize\":true,\"v2\":true,\"left\":76,\"top\":130,\"vpos\":\"top\",\"hpos\":\"left\",\"visible\":true,\"group\":\"root\",\"id\":\"box-195\",\"zindex\":8,\"wtype\":\"fixed\",\"htype\":\"fixed\",\"opacity\":1,\"propagateEvents\":false,\"lineHeightAuto\":true,\"rotation\":0,\"scale\":1,\"aspectratio\":false},{\"name\":\"text\",\"type\":\"text\",\"lib\":\"common\",\"caption\":\"Height : 174 cm...\",\"text\":\"Height : 174 cm&nbsp;\",\"textFont\":\"Trebuchet MS,sans-serif\",\"textFontStyle\":\"400\",\"textProperties\":[],\"textSize\":\"32\",\"textColor\":{\"type\":\"solid\",\"color\":\"rgba(113,113,113,1)\"},\"backgroundColor\":\"none\",\"width\":216,\"height\":39,\"lineHeight\":39,\"textAlign\":\"left\",\"textSpacing\":\"-1.6\",\"enableShadow\":false,\"autoResize\":true,\"v2\":true,\"left\":379,\"top\":86,\"vpos\":\"top\",\"hpos\":\"left\",\"visible\":true,\"group\":\"root\",\"id\":\"box-196\",\"zindex\":8,\"wtype\":\"fixed\",\"htype\":\"fixed\",\"opacity\":1,\"propagateEvents\":false,\"lineHeightAuto\":true,\"rotation\":0,\"scale\":1,\"aspectratio\":false},{\"name\":\"text\",\"type\":\"text\",\"lib\":\"common\",\"caption\":\"Surgery:\\nTotal Colec...\",\"text\":\"Surgery:&nbsp;<br>Total Colectomy with&nbsp;<br>pancreatico-jejunostomy&nbsp;\",\"textFont\":\"Trebuchet MS,sans-serif\",\"textFontStyle\":\"400\",\"textProperties\":[],\"textSize\":\"32\",\"textColor\":{\"type\":\"solid\",\"color\":\"rgba(113,113,113,1)\"},\"backgroundColor\":\"none\",\"width\":336,\"height\":117,\"lineHeight\":39,\"textAlign\":\"left\",\"textSpacing\":\"-1.6\",\"enableShadow\":false,\"autoResize\":true,\"v2\":true,\"left\":1084,\"top\":47,\"vpos\":\"top\",\"hpos\":\"left\",\"visible\":true,\"group\":\"root\",\"id\":\"box-197\",\"zindex\":8,\"wtype\":\"fixed\",\"htype\":\"fixed\",\"opacity\":1,\"propagateEvents\":false,\"lineHeightAuto\":true,\"rotation\":0,\"scale\":1,\"aspectratio\":false},{\"name\":\"text\",\"type\":\"text\",\"lib\":\"common\",\"caption\":\"Age : 86 y\",\"text\":\"Age : 86 y&nbsp;\",\"textFont\":\"Trebuchet MS,sans-serif\",\"textFontStyle\":\"400\",\"textProperties\":[],\"textSize\":\"32\",\"textColor\":{\"type\":\"solid\",\"color\":\"rgba(113,113,113,1)\"},\"backgroundColor\":\"none\",\"width\":139,\"height\":39,\"lineHeight\":39,\"textAlign\":\"left\",\"textSpacing\":\"-1.6\",\"enableShadow\":false,\"autoResize\":true,\"v2\":true,\"left\":76,\"top\":91,\"vpos\":\"top\",\"hpos\":\"left\",\"visible\":true,\"group\":\"root\",\"id\":\"box-198\",\"zindex\":8,\"wtype\":\"fixed\",\"htype\":\"fixed\",\"opacity\":1,\"propagateEvents\":false,\"lineHeightAuto\":true,\"rotation\":0,\"scale\":1,\"aspectratio\":false},{\"name\":\"text\",\"type\":\"text\",\"lib\":\"common\",\"caption\":\"Allergies :\\nPenicill...\",\"text\":\"Allergies :<br><span style=\\\"color: rgba(249,51,36,1);\\\">Penicillin</span><br><br>\",\"textFont\":\"Trebuchet MS,sans-serif\",\"textFontStyle\":\"400\",\"textProperties\":[],\"textSize\":\"32\",\"textColor\":{\"type\":\"solid\",\"color\":\"rgba(113,113,113,1)\"},\"backgroundColor\":\"none\",\"width\":203,\"height\":117,\"lineHeight\":39,\"textAlign\":\"left\",\"textSpacing\":\"-1.6\",\"enableShadow\":false,\"autoResize\":false,\"v2\":true,\"left\":766,\"top\":47,\"vpos\":\"top\",\"hpos\":\"left\",\"visible\":true,\"group\":\"root\",\"id\":\"box-199\",\"zindex\":8,\"wtype\":\"fixed\",\"htype\":\"fixed\",\"opacity\":1,\"propagateEvents\":false,\"lineHeightAuto\":true,\"rotation\":0,\"scale\":1,\"aspectratio\":false},{\"name\":\"text\",\"type\":\"text\",\"lib\":\"common\",\"caption\":\"BMI : 28 kg/m2...\",\"text\":\"BMI : 28 kg/m2\",\"textFont\":\"Trebuchet MS,sans-serif\",\"textFontStyle\":\"400\",\"textProperties\":[],\"textSize\":\"32\",\"textColor\":{\"type\":\"solid\",\"color\":\"rgba(113,113,113,1)\"},\"backgroundColor\":\"none\",\"width\":199,\"height\":39,\"lineHeight\":39,\"textAlign\":\"left\",\"textSpacing\":\"-1.6\",\"enableShadow\":false,\"autoResize\":true,\"v2\":true,\"left\":379,\"top\":171,\"vpos\":\"top\",\"hpos\":\"left\",\"visible\":true,\"group\":\"root\",\"id\":\"box-200\",\"zindex\":8,\"wtype\":\"fixed\",\"htype\":\"fixed\",\"opacity\":1,\"propagateEvents\":false,\"lineHeightAuto\":true,\"rotation\":0,\"scale\":1,\"aspectratio\":false},{\"name\":\"text\",\"type\":\"text\",\"lib\":\"common\",\"caption\":\"Weight : 69 Kg...\",\"text\":\"Weight : 69 Kg\",\"textFont\":\"Trebuchet MS,sans-serif\",\"textFontStyle\":\"400\",\"textProperties\":[],\"textSize\":\"32\",\"textColor\":{\"type\":\"solid\",\"color\":\"rgba(113,113,113,1)\"},\"backgroundColor\":\"none\",\"width\":190,\"height\":39,\"lineHeight\":39,\"textAlign\":\"left\",\"textSpacing\":\"-1.6\",\"enableShadow\":false,\"autoResize\":true,\"v2\":true,\"left\":379,\"top\":130,\"vpos\":\"top\",\"hpos\":\"left\",\"visible\":true,\"group\":\"root\",\"id\":\"box-201\",\"zindex\":8,\"wtype\":\"fixed\",\"htype\":\"fixed\",\"opacity\":1,\"propagateEvents\":false,\"lineHeightAuto\":true,\"rotation\":0,\"scale\":1,\"aspectratio\":false},{\"name\":\"verticalline\",\"type\":\"verticalline\",\"lib\":\"common\",\"caption\":\"Vertical line 3\",\"width\":1,\"height\":229,\"color\":\"A6A6A6\",\"weight\":1,\"left\":341,\"top\":10,\"vpos\":\"top\",\"hpos\":\"left\",\"visible\":true,\"group\":\"root\",\"id\":\"box-202\",\"zindex\":7,\"wtype\":\"fixed\",\"htype\":\"fixed\",\"opacity\":1,\"borderStyle\":\"solid\",\"rotation\":0,\"scale\":1,\"aspectratio\":false},{\"name\":\"verticalline\",\"type\":\"verticalline\",\"lib\":\"common\",\"caption\":\"Vertical line 5\",\"width\":1,\"height\":229,\"color\":\"A6A6A6\",\"weight\":1,\"left\":1064,\"top\":15,\"vpos\":\"top\",\"hpos\":\"left\",\"visible\":true,\"group\":\"root\",\"id\":\"box-203\",\"zindex\":7,\"wtype\":\"fixed\",\"htype\":\"fixed\",\"opacity\":1,\"borderStyle\":\"solid\",\"rotation\":0,\"scale\":1,\"aspectratio\":false},{\"name\":\"verticalline\",\"type\":\"verticalline\",\"lib\":\"common\",\"caption\":\"Vertical line 6\",\"width\":1,\"height\":229,\"color\":\"A6A6A6\",\"weight\":1,\"left\":733,\"top\":10,\"vpos\":\"top\",\"hpos\":\"left\",\"visible\":true,\"group\":\"root\",\"id\":\"box-204\",\"zindex\":7,\"wtype\":\"fixed\",\"htype\":\"fixed\",\"opacity\":1,\"borderStyle\":\"solid\",\"rotation\":0,\"scale\":1,\"aspectratio\":false},{\"name\":\"datetime\",\"type\":\"datetime\",\"lib\":\"common\",\"caption\":\"Date/Time 1\",\"width\":376,\"height\":41,\"backgroundColor\":\"none\",\"borderWidth\":\"1\",\"borderColor\":{\"type\":\"solid\",\"color\":\"rgba(102,150,200,1)\"},\"textFont\":\"Montserrat\",\"textFontStyle\":\"400\",\"textSize\":\"20\",\"textColor\":\"383838\",\"textProperties\":[],\"textAlign\":\"center\",\"dateFormat\":\"yearmonthday\",\"dayFormat\":\"superscript\",\"monthFormat\":\"long\",\"yearFormat\":\"long\",\"timeFormat\":\"24\",\"dateGeneralFormat\":\"timedate\",\"dayNameFormat\":\"long\",\"dateSeparator\":\"-\",\"left\":1064,\"top\":201,\"vpos\":\"top\",\"hpos\":\"left\",\"visible\":true,\"group\":\"root\",\"id\":\"box-205\",\"zindex\":5,\"wtype\":\"fixed\",\"htype\":\"fixed\",\"opacity\":1,\"rotation\":0,\"scale\":1,\"aspectratio\":false},{\"name\":\"text\",\"type\":\"text\",\"lib\":\"common\",\"caption\":\"MRN : 12345679...\",\"text\":\"MRN : 12345679&nbsp;\",\"textFont\":\"Trebuchet MS,sans-serif\",\"textFontStyle\":\"400\",\"textProperties\":[],\"textSize\":\"32\",\"textColor\":{\"type\":\"solid\",\"color\":\"rgba(113,113,113,1)\"},\"backgroundColor\":\"none\",\"width\":218,\"height\":39,\"lineHeight\":39,\"textAlign\":\"left\",\"textSpacing\":\"-1.6\",\"enableShadow\":false,\"autoResize\":true,\"v2\":true,\"left\":76,\"top\":171,\"vpos\":\"top\",\"hpos\":\"left\",\"visible\":true,\"group\":\"root\",\"id\":\"box-206\",\"zindex\":4,\"wtype\":\"fixed\",\"htype\":\"fixed\",\"opacity\":1,\"propagateEvents\":false,\"lineHeightAuto\":true,\"rotation\":0,\"scale\":1,\"aspectratio\":false},{\"name\":\"verticalline\",\"type\":\"verticalline\",\"lib\":\"common\",\"caption\":\"Vertical line 1\",\"width\":1,\"height\":658,\"color\":{\"type\":\"solid\",\"color\":\"rgba(70,127,215,1)\"},\"weight\":1,\"left\":734,\"top\":242,\"vpos\":\"top\",\"hpos\":\"left\",\"visible\":true,\"group\":\"root\",\"id\":\"box-207\",\"zindex\":3,\"wtype\":\"fixed\",\"htype\":\"fixed\",\"opacity\":1,\"borderStyle\":\"solid\",\"rotation\":0,\"scale\":1,\"aspectratio\":false},{\"name\":\"text\",\"type\":\"text\",\"lib\":\"common\",\"caption\":\"John Doe\",\"text\":\"John Doe&nbsp;\",\"textFont\":\"Trebuchet MS,sans-serif\",\"textFontStyle\":\"400\",\"textProperties\":[],\"textSize\":\"64\",\"textColor\":{\"type\":\"solid\",\"color\":\"rgba(117,85,88,1)\"},\"backgroundColor\":\"none\",\"width\":272,\"height\":78,\"lineHeight\":78,\"textAlign\":\"left\",\"textSpacing\":\"-1.6\",\"enableShadow\":false,\"autoResize\":true,\"v2\":true,\"left\":40,\"top\":15,\"vpos\":\"top\",\"hpos\":\"left\",\"visible\":true,\"id\":\"box-208\",\"group\":\"root\",\"zindex\":2,\"wtype\":\"fixed\",\"htype\":\"fixed\",\"opacity\":1,\"propagateEvents\":false,\"lineHeightAuto\":true,\"rotation\":0,\"scale\":1,\"aspectratio\":false},{\"name\":\"basic_textfield\",\"type\":\"basic_textfield\",\"lib\":\"generic2\",\"caption\":\"Text Field 2\",\"width\":733,\"height\":77,\"value\":\"Induction Medication\",\"placeholder\":\"Placeholder\",\"inputtype\":\"text\",\"backgroundColor\":{\"type\":\"solid\",\"color\":\"rgba(177,190,227,1)\"},\"textFont\":\"Arial,sans-serif\",\"textFontStyle\":\"400\",\"textSize\":\"64\",\"textColor\":{\"type\":\"solid\",\"color\":\"rgba(233,240,215,1)\"},\"textProperties\":[],\"placeholderColor\":\"c1c8c9\",\"borderWidth\":\"5\",\"borderStyle\":\"solid\",\"borderRadius\":4,\"borderColor\":{\"type\":\"solid\",\"color\":\"rgba(85,85,85,1)\"},\"textAlign\":\"center\",\"left\":2,\"top\":242,\"vpos\":\"top\",\"hpos\":\"left\",\"visible\":true,\"id\":\"box-209\",\"group\":\"root\",\"zindex\":11,\"wtype\":\"fixed\",\"htype\":\"fixed\",\"opacity\":1,\"rotation\":0,\"scale\":1,\"aspectratio\":false},{\"name\":\"basic_textfield\",\"type\":\"basic_textfield\",\"lib\":\"generic2\",\"caption\":\"Text Field 3\",\"width\":698,\"height\":77,\"value\":\"Blood Pressure\",\"placeholder\":\"Placeholder\",\"inputtype\":\"text\",\"backgroundColor\":{\"type\":\"solid\",\"color\":\"rgba(177,190,227,1)\"},\"textFont\":\"Arial,sans-serif\",\"textFontStyle\":\"400\",\"textSize\":\"64\",\"textColor\":{\"type\":\"solid\",\"color\":\"rgba(233,240,215,1)\"},\"textProperties\":[],\"placeholderColor\":\"c1c8c9\",\"borderWidth\":\"5\",\"borderStyle\":\"solid\",\"borderRadius\":4,\"borderColor\":{\"type\":\"solid\",\"color\":\"rgba(85,85,85,1)\"},\"textAlign\":\"center\",\"left\":733,\"top\":244,\"vpos\":\"top\",\"hpos\":\"left\",\"visible\":true,\"id\":\"box-210\",\"group\":\"root\",\"zindex\":11,\"wtype\":\"fixed\",\"htype\":\"fixed\",\"opacity\":1,\"rotation\":0,\"scale\":1,\"aspectratio\":false},{\"name\":\"basic_textfield\",\"type\":\"basic_textfield\",\"lib\":\"generic2\",\"caption\":\"Text Field 13\",\"width\":338,\"height\":55,\"value\":\"Etomidate (mg)\",\"placeholder\":\"Placeholder\",\"inputtype\":\"text\",\"backgroundColor\":{\"type\":\"solid\",\"color\":\"rgba(255, 255, 0,1)\"},\"textFont\":\"Arial,sans-serif\",\"textFontStyle\":\"400\",\"textSize\":\"28\",\"textColor\":{\"type\":\"solid\",\"color\":\"rgba(56,56,56,1)\"},\"textProperties\":[],\"placeholderColor\":\"c1c8c9\",\"borderWidth\":1,\"borderStyle\":\"solid\",\"borderRadius\":4,\"borderColor\":{\"type\":\"solid\",\"color\":\"rgba(27,77,87,1)\"},\"textAlign\":\"center\",\"left\":31,\"top\":374,\"vpos\":\"top\",\"hpos\":\"left\",\"visible\":true,\"group\":\"root\",\"id\":\"box-211\",\"zindex\":10,\"wtype\":\"fixed\",\"htype\":\"fixed\",\"opacity\":1,\"rotation\":0,\"scale\":1,\"aspectratio\":false,\"actions\":[{\"title\":\"click \",\"type\":\"click\",\"actionId\":\"change-property\",\"callback\":true,\"pageId\":\"2\",\"itemId\":\"box-67\",\"propertyId\":\"value\",\"valueType\":\"customValue\",\"newvalue\":\"Etomidate (mg)\",\"delay\":\"0\",\"guid\":\"647d5bf3-f0f4-48d3-af03-d284f2cf0c7d\",\"callbackAction\":{\"actionId\":\"go-to-page\",\"callback\":false,\"pageId\":\"2\",\"animation\":\"fade\",\"animationEasing\":\"Cubic.easeOut\",\"duration\":\"400\",\"delay\":\"0\",\"guid\":\"20355424-a25c-43d7-ac5b-9534c2a54ab6\",\"clb_disabled\":false},\"sort\":0}]},{\"name\":\"basic_textfield\",\"type\":\"basic_textfield\",\"lib\":\"generic2\",\"caption\":\"Text Field 14\",\"width\":338,\"height\":55,\"value\":\"Ketamine (mg)\",\"placeholder\":\"Placeholder\",\"inputtype\":\"text\",\"backgroundColor\":{\"type\":\"solid\",\"color\":\"rgba(255, 255, 0,1)\"},\"textFont\":\"Arial,sans-serif\",\"textFontStyle\":\"400\",\"textSize\":\"28\",\"textColor\":{\"type\":\"solid\",\"color\":\"rgba(56,56,56,1)\"},\"textProperties\":[],\"placeholderColor\":\"c1c8c9\",\"borderWidth\":1,\"borderStyle\":\"solid\",\"borderRadius\":4,\"borderColor\":{\"type\":\"solid\",\"color\":\"rgba(27,77,87,1)\"},\"textAlign\":\"center\",\"left\":31,\"top\":458,\"vpos\":\"top\",\"hpos\":\"left\",\"visible\":true,\"group\":\"root\",\"id\":\"box-212\",\"zindex\":10,\"wtype\":\"fixed\",\"htype\":\"fixed\",\"opacity\":1,\"rotation\":0,\"scale\":1,\"aspectratio\":false,\"actions\":[{\"title\":\"click \",\"type\":\"click\",\"actionId\":\"change-property\",\"callback\":true,\"pageId\":\"2\",\"itemId\":\"box-67\",\"propertyId\":\"value\",\"valueType\":\"customValue\",\"newvalue\":\"Ketamine (mcg)\",\"delay\":\"0\",\"guid\":\"b02f8196-d229-45f9-adb2-8d4516b02bc8\",\"callbackAction\":{\"actionId\":\"go-to-page\",\"callback\":false,\"pageId\":\"2\",\"animation\":\"fade\",\"animationEasing\":\"Cubic.easeOut\",\"duration\":\"400\",\"delay\":\"0\",\"guid\":\"201f9a80-15c6-489d-a60f-59a52490e59f\",\"clb_disabled\":false},\"sort\":0}]},{\"name\":\"basic_textfield\",\"type\":\"basic_textfield\",\"lib\":\"generic2\",\"caption\":\"Text Field 15\",\"width\":338,\"height\":55,\"value\":\"Propofol (mg)\",\"placeholder\":\"Placeholder\",\"inputtype\":\"text\",\"backgroundColor\":{\"type\":\"solid\",\"color\":\"rgba(255, 255, 0,1)\"},\"textFont\":\"Arial,sans-serif\",\"textFontStyle\":\"400\",\"textSize\":\"28\",\"textColor\":{\"type\":\"solid\",\"color\":\"rgba(56,56,56,1)\"},\"textProperties\":[],\"placeholderColor\":\"c1c8c9\",\"borderWidth\":1,\"borderStyle\":\"solid\",\"borderRadius\":4,\"borderColor\":{\"type\":\"solid\",\"color\":\"rgba(27,77,87,1)\"},\"textAlign\":\"center\",\"left\":31,\"top\":558,\"vpos\":\"top\",\"hpos\":\"left\",\"visible\":true,\"group\":\"root\",\"id\":\"box-223\",\"zindex\":10,\"wtype\":\"fixed\",\"htype\":\"fixed\",\"opacity\":1,\"rotation\":0,\"scale\":1,\"aspectratio\":false,\"actions\":[{\"title\":\"click \",\"type\":\"click\",\"actionId\":\"change-property\",\"callback\":true,\"pageId\":\"2\",\"itemId\":\"box-67\",\"propertyId\":\"value\",\"valueType\":\"customValue\",\"newvalue\":\"Propofol (mg)\",\"delay\":\"0\",\"guid\":\"c1551700-f891-458a-b4de-7814c5c10f5c\",\"callbackAction\":{\"actionId\":\"go-to-page\",\"callback\":false,\"pageId\":\"2\",\"animation\":\"fade\",\"animationEasing\":\"Cubic.easeOut\",\"duration\":\"400\",\"delay\":\"0\",\"guid\":\"bbdc0303-2cc4-405d-ab9a-91330997213c\",\"clb_disabled\":false},\"sort\":0}]},{\"name\":\"image\",\"type\":\"image\",\"lib\":\"common\",\"caption\":\"Screen Shot 2022-12-12 at 7.38.12 PM.png\",\"imgSrc\":{\"fileId\":\"68d047acd614373a75ca2548bba92b44.png\",\"assetType\":\"project\",\"bucketsource\":\"main\",\"name\":\"Screen Shot 2022-12-12 at 7.38.12 PM.png\"},\"repeat\":false,\"width\":658,\"height\":398,\"borderWidth\":0,\"borderRadius\":0,\"borderColor\":\"#D1D1D1\",\"borderPos\":\"inside\",\"propagateEvents\":false,\"actions\":[],\"overlay\":false,\"autoResize\":false,\"aspectratio\":1.6506849315068493,\"assistiveDescription\":\"\",\"left\":762,\"top\":359,\"visible\":true,\"id\":\"box-213\",\"group\":\"root\",\"zindex\":12,\"hpos\":\"left\",\"vpos\":\"top\",\"wtype\":\"fixed\",\"htype\":\"fixed\",\"opacity\":1,\"borderStyle\":\"solid\",\"rotation\":0,\"scale\":1},{\"name\":\"basic_button_text\",\"type\":\"basic_button_text\",\"lib\":\"generic2\",\"caption\":\"Button with Background 5\",\"width\":177,\"height\":80,\"backgroundColor\":{\"type\":\"solid\",\"color\":\"rgba(249,51,36,1)\"},\"borderWidth\":0,\"borderColor\":\"#025abf\",\"borderRadius\":4,\"text\":\"Back\",\"textFont\":\"Arial,sans-serif\",\"textFontStyle\":\"400\",\"textSize\":\"36\",\"textColor\":\"FFFFFF\",\"textProperties\":[],\"textAlign\":\"center\",\"iconpos\":\"none\",\"left\":472,\"top\":652,\"vpos\":\"top\",\"hpos\":\"left\",\"visible\":true,\"id\":\"box-214\",\"group\":\"root\",\"zindex\":13,\"wtype\":\"fixed\",\"htype\":\"fixed\",\"opacity\":1,\"rotation\":0,\"scale\":1,\"aspectratio\":false,\"actions\":[{\"title\":\"Interaction 1\",\"type\":\"click\",\"actionId\":\"go-to-page\",\"callback\":false,\"pageId\":\"2\",\"animation\":\"fade\",\"animationEasing\":\"Cubic.easeOut\",\"duration\":\"400\",\"delay\":\"0\",\"guid\":\"dad0c4e3-784a-4804-ab16-f6ce57266e06\",\"sort\":0}]}]","template":{"id":"","stateid":""},"history":[],"id":"9fe37c39-5dd6-4727-9a3f-a121dee954b9","actions":[],"groups":{}}]}],"templates":[],"symbols":[{"mode":"component","title":"Propofol_dose","id":1,"states":[{"title":"State 1","data":"[{\"name\":\"text\",\"type\":\"text\",\"lib\":\"common\",\"caption\":\"Propofol_dose\",\"text\":\"Type soething\",\"textFont\":\"Trebuchet MS,sans-serif\",\"textFontStyle\":\"400\",\"textProperties\":[],\"textSize\":\"32\",\"textColor\":{\"type\":\"solid\",\"color\":\"rgba(113,113,113,1)\"},\"backgroundColor\":\"none\",\"width\":180,\"height\":39,\"lineHeight\":39,\"textAlign\":\"left\",\"textSpacing\":\"-1.6\",\"enableShadow\":false,\"autoResize\":true,\"v2\":true,\"left\":0,\"top\":0,\"vpos\":\"top\",\"hpos\":\"left\",\"visible\":true,\"id\":\"box-86\",\"group\":\"root\",\"zindex\":14,\"wtype\":\"fixed\",\"htype\":\"fixed\",\"opacity\":1,\"propagateEvents\":false,\"lineHeightAuto\":true,\"rotation\":0,\"scale\":1,\"aspectratio\":false,\"userCaption\":true}]","history":[],"id":"7e438339-f3ca-47da-bb77-b19e726b1df2","dimensions":[180,39],"groups":{}}]}],"saveversion":170,"_imgs_version":1670899970,"makescrollable":0}; prx.xdata_str = JSON.stringify(prx.xdata);
```
