## Supplement File 1 for "Post Induction Hypotension prediction during general anesthesia using Machine Learning Techniques": components.js

```
/********************************************************************************************/
/********************************************************************************************/
/*******************************            COMMON             ******************************/
/********************************************************************************************/
/********************************************************************************************/

/************************************* COMMON COMPONENT VARIABLES AND PROPERTIES *************************************/
var _library = 'common';

/************************************* COMPONENT TYPES *************************************/

//TYPE: SHAPES
prx.types.shape = {
    name: 'shape'
    ,onDisplay: function(item,containerid,symbol) {

        if(item.typeName == 'oval' || item.typeName == 'rectangle') {
            return prx.types.rectangle.onDisplay(item,containerid,symbol);
        }

        if(typeof(item.lineHeight) == 'undefined') { item.lineHeight = parseInt(prx.componentsHelper.getProp(item.textSize,'num-text-size')*1.231); }
        if(typeof(item.lineHeightAuto) == 'undefined') { item.lineHeightAuto = Boolean(true); }
        if(typeof(item.textSpacing) == 'undefined') { item.textSpacing = 0; }

        var _id = (!containerid) ? item.id : containerid+'-'+item.id;
        var dims = prx.componentsHelper.getRealDims(item, symbol);

        var path;

        var _props = (typeof(prx.richtext)=='undefined') ? prx.componentsHelper.getProp(item.textProperties,'props-text') : '';

        var cR = '<div id="' + _id + '" ' + prx.items.getComponentBaseAttributes(item, containerid, symbol)  + ' class="' + prx.items.getComponentBaseClasses(item, containerid, symbol) + ' box pos type-shapes '+prx.componentsHelper.getProp(item.typeName,'other')+'" data-shape-type="'+prx.componentsHelper.getProp(item.typeName,'other')+'" ';
        cR += 'data-border-width-shape="'+prx.componentsHelper.getProp(item.borderWidthShape,'border-width')+'" data-num-nodes="'+prx.componentsHelper.getProp(item.numOfNodes,'numOfNodes')+'" data-spike-depth="'+prx.componentsHelper.getProp(item.spikeDepth,'spikeDepth')+'" ';
        cR += 'data-skew="'+prx.componentsHelper.getProp(item.skew,'skew')+'" data-base-width="'+prx.componentsHelper.getProp(item.baseWidth,'baseWidth')+'">';

        cR += '<style>';
        cR += prx.items.getComponentBaseStyle(item, containerid, symbol);
        cR += '#'+_id+' .shapes-text-container { width: 100%; height: 100%; '+_props+' '+prx.componentsHelper.getProp(item.textFont + '|' + item.textFontStyle,'font-family') + prx.componentsHelper.getProp(item.textFontStyle,'font-style') + '; font-size: '+prx.componentsHelper.getProp(item.textSize,'num-text-size')+'px; text-align: '+prx.componentsHelper.getProp(item.textAlign,'align')+'; color: '+prx.componentsHelper.getProp(item.textColor,'color-text')+'; line-height: ' + prx.componentsHelper.getProp(item.lineHeight, 'num-text-size') + 'px; letter-spacing: ' + prx.componentsHelper.getProp(item.textSpacing, 'num-text-size') + 'px;  }';
        if( prx.componentsHelper.getProp(item.joinType,'other') == 'miter') {
            cR += '#'+_id+' svg { overflow: visible; }';
        }
        cR += '</style>';
        cR += prx.items.getComponentPrependDivs(item, containerid, symbol);
        cR += '<div id="'+prx.componentsHelper.getProp(item.typeName,'other')+'-' + _id + '" class="shape-wrapper ">';

        cR += '<svg id="svg-'+ _id +'" viewBox="0 0 '+(dims.width)+' '+(dims.height)+'" width="100%" height="100%" preserveAspectRatio="none" xmlns="http://www.w3.org/2000/svg" version="1.1">';

        if(prx.componentsHelper.getProp(item.typeName,'other') == 'polygon' || prx.componentsHelper.getProp(item.typeName,'other') == 'star') {
            path = prx.componentsHelper.drawComplexShape( item, prx.componentsHelper.getProp(item.typeName,'other'), dims.width, dims.height, prx.componentsHelper.getProp(item.borderWidthShape,'num-border-width'));
        }
        else {
            path = prx.componentsHelper.drawShape( item, prx.componentsHelper.getProp(item.typeName,'other'), dims.width, dims.height, prx.componentsHelper.getProp(item.borderWidthShape,'num-border-width'));
        }

        cR += '<path d="'+path+'" id="path-'+ _id +'" class="liveUpdate-backgroundColor-fill liveUpdate-borderColor-stroke changeProperty-backgroundColor changeProperty-backgroundColor-fill changeProperty-borderColor-stroke"';
        cR += 'style="width:100%; height:100%; ';
        cR += 'fill:'+prx.componentsHelper.getProp(item.backgroundColor,'color-background')+'; ';
        cR += 'stroke:'+prx.componentsHelper.getProp(item.borderColor,'color-border')+'; stroke-width:'+prx.componentsHelper.getProp(item.borderWidthShape,'num-border-width')+'px; stroke-linejoin:'+prx.componentsHelper.getProp(item.joinType,'other')+'; stroke-miterlimit:'+prx.componentsHelper.getProp(item.borderWidthShape,'num-border-width')+';" />';

        cR += '</svg>';

        cR += '<div class="shapes-text-container liveUpdate-textColor-color">';
        cR += '<span data-editableproperty="text">' + prx.componentsHelper.getProp(item.text,'text-textarea') + '</span>';
        cR += '</div>';
        cR += '</div>';
        cR += prx.items.getComponentAppendDivs(item, containerid, symbol);
        cR += '</div>';
        return cR;
    }
    ,onResize: function(item,containerid,symbol) {

        if(item.typeName == 'oval' || item.typeName == 'rectangle') {
            return;
        }

        var _id = (!containerid) ? item.id : containerid+'-'+item.id;
        var dims = prx.componentsHelper.getRealDims(item, symbol);

        document.getElementById('svg-'+ _id).setAttribute('viewBox', '0 0 '+(dims.width)+' '+(dims.height));

        if(prx.componentsHelper.getProp(item.typeName,'other') == 'polygon' || prx.componentsHelper.getProp(item.typeName,'other') == 'star') {
            document.getElementById('path-'+ _id).setAttribute('d', prx.componentsHelper.drawComplexShape( item, prx.componentsHelper.getProp(item.typeName,'other'), dims.width, dims.height, prx.componentsHelper.getProp(item.borderWidthShape,'num-border-width')));
        } else {
            document.getElementById('path-'+ _id).setAttribute('d', prx.componentsHelper.drawShape( item, prx.componentsHelper.getProp(item.typeName,'other'), dims.width, dims.height, prx.componentsHelper.getProp(item.borderWidthShape,'num-border-width')));
        }
    }
    ,interactions: [
        prx.commonproperties.actions
    ]
    ,editableProperties: [
        {
	    	caption: 'Text'
	    	,name: 'text'
	    	,type: 'textarea'
	    	,value: function(item,name) {
	    		if(typeof(item.text) == 'undefined') { item.text = ''; }
	    		return item.text;
	    	},
      		changeProperty: {
                caption: 'Text',
                property: 'text',
                selector: '.shapes-text-container span',
                transitionable: false
  			}
	    }
    ]
    ,propertyGroups: [
        {
            caption: 'Style',
            properties: [
                [
                    {
                        caption: 'Background',
                        name: 'backgroundColor',
                        proptype: 'background-color',
                        type: 'solid-colorpicker',
                        value: function (item, name) {
                            return item.backgroundColor;
                        },
                        liveUpdate: 'fill,background-color',
                        changeProperty: {
                            caption: 'Background color',
                            selector: '.changeProperty-backgroundColor',
                            property: 'fill,background-color',
                            transitionable: true
                        }
                    }
                ],
                [
                    {
                    	caption: 'Border',
                    	name: 'borderWidthShape',
                    	proptype: 'border-width',
                    	type: 'combo-select',
                    	value: function(item,name) {
                    		return item.borderWidthShape;
                    	},
                    	values: { min: 0, max: 20, step: 1 },

	                    changeProperty: {
                    		caption: 'Shape\'s Border Width',
                    		//selector:'.changeProperty-borderWidth',
                    		//property:'stroke-width',
                    		//transitionable: true
		                    rerender: true
                    	},
		                onChange: function (item, name) {
                            if(item.borderWidth <= 0) { $('#property-roundJoin').hide();}
                            else { $('#property-roundJoin').show();}
                        },
	                    hiddenByDefault: function(item) {
		                    return (item.typeName=='rectangle' || item.typeName=='oval');
	                    }
                    },
	                {
		                caption: 'Border',
		                name: 'borderWidth',
		                proptype: 'border-width-1',
		                type: 'combo-select',
		                value: function(item,name) { return item.borderWidth; },
		                values: { min: 0, max: 20, step: 1 } ,
                        expandableType: function(item, name){
                            if(item.typeName=='oval'){
                                return '';
                            } else {
                                return 'borderWidth';
                            }
                        },
                        expandedValues: function(item, name){
		                    if(item.typeName=='oval'){
		                        return [];
                            } else {
		                        return ['t', 'r', 'l', 'b'];
                            }
                        },
		                changeProperty: {
			                caption: 'Border Width',
			                selector: '.changeProperty-borderWidth',
			                property: 'border-width',
			                transitionable: true
		                },
		                hiddenByDefault: function(item) {
			                return !(item.typeName=='rectangle' || item.typeName=='oval');
		                }
	                },
	                {
		                caption: false,
		                name: 'borderStyle',
		                proptype: 'border-style',
		                type: 'select',
		                value: function(item,name) {
			                if(typeof(item.borderStyle) == 'undefined') { item.borderStyle = 'solid'; }
			                return item.borderStyle;
		                },
		                values: [{ value: 'solid', displayValue: 'Solid'},{ value: 'dotted', displayValue: 'Dotted'},{ value: 'dashed', displayValue: 'Dashed'},{ value: 'double', displayValue: 'Double'},{ value: 'none', displayValue: 'None'}],
		                changeProperty: {
			                caption: 'Border Style',
			                selector: '.inner-rec',
			                property: 'border-style',
			                transitionable: false
		                },
		                hiddenByDefault: function(item) {
			                return !(item.typeName=='rectangle' || item.typeName=='oval');
		                }
	                },
                    {
	                    caption: false,
	                    name: 'borderColor',
	                    proptype: 'border-color',
	                    type: 'colorpicker',
	                    value: function (item, name) {
	                        return item.borderColor;
	                    },
	                    liveUpdate: 'stroke,border-color',
	                    changeProperty: {
	                        caption: 'Border color',
	                        selector: '.changeProperty-borderColor',
	                        property: 'stroke,border-color',
	                        transitionable: true
	                    }
	                }
                ]
	            ,[
		            {
		            	caption: '<span class="icon icon-border-radius" title="Border radius"></span>',
			            name: 'borderRadius',
			            proptype: 'border-radius',
			            type: 'combo-select',
			            value: function(item,name) {
				            if(item.borderRadius == parseInt(item.borderRadius)) { return item.borderRadius += 'px'; }
				            return item.borderRadius;
			            },
			            values: { min: 0, max: 20, step: 1 },
                        expandableType: 'borderRadius',
                        expandedValues: ['tl', 'tr', 'bl', 'br'],
			            changeProperty: {
				            caption: 'Border Radius',
				            selector: '.inner-rec',
				            property: 'border-radius',
				            transitionable: true
			            },
			            hiddenByDefault: function(item) {
				            return !(item.typeName=='rectangle');
			            }
		            }
	            ],
             	[
	                {
                        caption: 'Rounded Border'
                        ,name: 'roundJoin'
                        ,type: 'onoff'
                        ,value: function(item,name) {
                            if(typeof(item.roundJoin)=='undefined') {
                                return false;
                            }
                            return item.roundJoin;
                        }
		                ,onChange: function (item, name) {
                            if(item.roundJoin) { item.joinType = 'round';}
                            else { item.joinType = 'miter';}
                        }
		                ,hiddenByDefault: function(item) {
				            return (item.borderWidthShape <= 0) || (item.typeName == 'oval' || item.typeName == 'rectangle');
			            }
                        ,changeProperty: {
                            caption: 'Rounded edges',
                            rerender: true
                        }
                    }
                ]
            ]
        },
        {
            caption: 'Text',
            properties:
			[
			    [
			        {
			            caption: { label: 'Font family', class: 'text-properties-label text-fontfamily-label' },
			            name: 'textFont',
			            proptype: 'font-family',
			            type: 'select',
			            relatedEditableProperties: 'text',
			            relatedCSSProperties: 'font-family',
			            value: function(item,name) {
			                if(typeof(item.textFont) == 'undefined') { item.textFont = 'Arial,sans-serif'; }
			                return item.textFont;
			            },
			            values: function(){ return prx.comps.fonts; }
			      		,changeProperty: {
			                caption: ' Text font',
			                selector: '.shapes-text-container',
			                property: 'font-family',
			                transitionable: false
						 }

			        }
			    ],
			    [
			        prx.commonproperties.textFontStyleRichText(['font-weight', 'font-style'],'text', false, {propName: 'textFontStyle', caption: 'Font style', useAsDynProp: false}),
			        {
			            caption: false,
			            name: 'textSize',
			            proptype: 'font-size',
			            type: 'combo-select',
			            relatedEditableProperties: 'text',
			            relatedCSSProperties: 'font-size',
			            value: function(item,name) {
			                if(typeof(item.textSize) == 'undefined') { item.textSize = 16; }
			                return item.textSize;
			            },
			            values: prx.comps.textsize
			            ,onChange: function(item) {
			                if(item.lineHeightAuto==true) {
			                    item.lineHeight = parseInt(prx.componentsHelper.getProp(item.textSize,'num-text-size')*1.231);
			                }
			            }
			            ,changeProperty: {
			                caption: ' Text size',
			                selector: '.shapes-text-container',
			                property: 'font-size',
			                transitionable: false
			            }
			        }
			    ],
			    [
			        prx.commonproperties.textPropertiesRichText(['font-weight','font-style','text-decoration'],'text', false, {propName: 'textProperties', caption: 'Underline', useAsDynProp: false}),
			        {
			            caption: { label: 'Color', class: 'text-properties-label text-color-label' },
			            pffSettings: prx.commonproperties.pffSettingsColor,
			            name: 'textColor',
			            proptype: 'font-color',
			            type: 'colorpicker',
			            relatedEditableProperties: 'text',
			            relatedCSSProperties: 'color',
			            value: function(item,name) {
			                if(typeof(item.textColor) == 'undefined') { item.textColor = '#2E2E2E'; }
			                return item.textColor;
			            },
			            liveUpdate: 'color'
			            ,changeProperty: {
			                caption: ' Text color',
			                selector: '.shapes-text-container',
			                property: 'color',
			                transitionable: true
			            }
			        }
		  		],
			    [
			        {
			            caption: { label: 'Alignment', class: 'text-properties-label text-alignment-label' },
			            name: 'textAlign',
			            proptype: 'text-align',
			            type: 'radio',
			            relatedEditableProperties: 'text',
			            relatedCSSProperties: 'text-align',
			            pffSettings: prx.commonproperties.pffSettingsTextAlignment,
			            value: function(item,name) {
			                if(typeof(item.textAlign) == 'undefined') { item.textAlign = 'center'; }
			                return item.textAlign;
			            },
			            values: [
			                { value: 'left', displayValue: '', icon: 'align-left'},
			                { value: 'center', displayValue: '', icon: 'align-center'},
			                { value: 'right', displayValue: '', icon: 'align-right'}
			            ],
			            changeProperty: {
			                caption: 'Text Align',
			                selector: '.shapes-text-container',
			                property: 'text-align',
			                transitionable: false
			            }
                	}
			    ],
			    [
			        {
			            caption: 'Line',
			            name: 'lineHeight',
			            proptype: 'line-height',
			            relatedEditableProperties: 'text',
			            relatedCSSProperties: 'line-height',
			            type: 'input-numeric',
			            pffSettings: prx.commonproperties.pffSettingsLineHeight,
			            value: function(item,name) {
			                if(typeof(item.lineHeight) == 'undefined') { item.lineHeight = parseInt(prx.componentsHelper.getProp(item.textSize,'num-text-size')*1.231); }
			                return item.lineHeight;
			            }
			            ,changeProperty: {
			                caption: 'Line height',
			                // it sets line-height: 100 instead of line-height: 100px which is different :(
			                // selector: '.changeProperty-lineHeight',
			                // property: 'line-height',
			                // transitionable: false
			                rerender: true
			            },
			            onChange: function(item) {
			                if (item.lineHeight == '') {//auto
			                    item.lineHeight = parseInt(prx.componentsHelper.getProp(item.textSize, 'num-text-size') * 1.231);
			                    item.lineHeightAuto = Boolean(true);
			                } else {
			                    item.lineHeightAuto = Boolean(false);
			                }
			            }
			        },
			        {
			            caption: 'Character',
			            name: 'textSpacing',
			            proptype: 'letter-spacing',
			            type: 'input-numeric',
			            relatedEditableProperties: 'text',
			            relatedCSSProperties: 'letter-spacing',
			            pffSettings: prx.commonproperties.pffSettingsCharacterSpacing,
			            value: function(item,name) {
			                if(typeof(item.textSpacing) == 'undefined') { item.textSpacing = 0; }
			                return item.textSpacing;
			            },
			            changeProperty: {
			                caption: 'Character Spacing',
			                selector: '.shapes-text-container',
			                property: 'letter-spacing',
			                transitionable: true
			            }
			        }
			    ]
			]
        },
	  	{
  		   	caption: 'Type',
            properties: [
     			[
     			 	{
                        caption: false,
                        proptype: 'shape-type',
                        name: 'typeName',
                        type: 'radio',
                        value: function(item, name) {
                            return item.typeName;
                        },
                        values: [
                            {value: 'oval',displayValue: '', icon: 'shape-oval'},
                            {value: 'rectangle',displayValue: '', icon: 'shape-rectangle'},
                            {value: 'triangle',displayValue: '', icon: 'shape-triangle'},
                            {value: 'rhombus',displayValue: '', icon: 'shape-rhombus'},
                            {value: 'trapezoid',displayValue: '', icon: 'shape-trapezoid'},
                            {value: 'parallelogram',displayValue: '', icon: 'shape-parallelogram'},
                            {value: 'polygon',displayValue: '', icon: 'shape-polygon'},
                            {value: 'star',displayValue: '', icon: 'shape-star'}
                        ],
                        onChange: function (item, name) {
                            if(item.typeName == 'rectangle' || item.typeName == 'oval') {
                                switch(true){
                                    case item.typeName == 'oval' && typeof(item.borderWidth) != 'undefined' && typeof(item.borderWidth) == 'object':
                                        item.borderWidth = item.borderWidth.t;
                                        break;
                                    case typeof(item.borderWidth) == 'undefined' && typeof(item.borderWidthShape) == 'undefined':
                                        item.borderWidth = 0;
                                        break;
                                    case typeof(item.borderWidth) == 'undefined' && typeof(item.borderWidthShape) != 'undefined':
                                        item.borderWidth = item.borderWidthShape;
                                        delete item.borderWidthShape;
                                        break;
                                }
                            } else {
                                switch(true){
                                    case typeof(item.borderWidth) == 'undefined' && typeof(item.borderWidthShape) == 'undefined':
                                        item.borderWidthShape = 0;
                                        break;
                                    case typeof(item.borderWidth) != 'undefined' && typeof(item.borderWidth) == 'object' && typeof(item.borderWidthShape) == 'undefined':
                                        item.borderWidthShape = item.borderWidth.t;
                                        delete item.borderWidth;
                                        break;
                                    case typeof(item.borderWidth) != 'undefined' && typeof(item.borderWidth) != 'object' && typeof(item.borderWidthShape) == 'undefined':
                                        item.borderWidthShape = item.borderWidth;
                                        delete item.borderWidth;
                                        break;
                                }
                            }

                            if(item.typeName == 'polygon' || item.typeName == 'star') {
                                $('#property-numOfNodes').show();
                            }
                            else { $('#property-numOfNodes').hide();}

                            if(item.typeName == 'star') { $('#property-spikeDepth').show();}
                            else { $('#property-spikeDepth').hide();}

                            if(item.typeName == 'parallelogram') {
                                $('#property-skew').show();
                            }
                            else { $('#property-skew').hide();}

                            if(item.typeName == 'trapezoid') {
                                $('#property-baseWidth').show();
                            }
                            else { $('#property-baseWidth').hide();}

                            if(item.typeName == 'oval') {
                                item.borderRadius = '50%';
                            }
                            if(item.typeName == 'rectangle' || item.typeName == 'oval') {
                                $('#property-borderStyle').show();
                                $('#property-borderWidth').show();
                                $('#property-borderWidthShape').hide();
                            } else {
                                $('#property-borderStyle').hide();
                                $('#property-borderWidth').hide();
                                $('#property-borderWidthShape').show();
                            }


                            if(item.typeName == 'rectangle') {
                                $('#property-borderRadius').show();
                                item.borderRadius = '0';
                            } else {
                                $('#property-borderRadius').hide();
                            }

                            return item;

                        },
                        changeProperty: {
                            caption: 'Shape type',
                            rerender: true
                        }
                    }
     			]
     		]
  	   },
  	   {
  			caption: 'Dimensions',
  			properties: [
                [
      			 	{
                        caption: 'Nodes',
                        name: 'numOfNodes',
                        proptype: 'number-of-nodes',
                        type: 'slider-input',
		                values: { min: 4, max: 50, step: 1 },
                        value: function(item,name) {
                            return item.numOfNodes;
                        },
                        hiddenByDefault: function (item, name) {
                            return !(item.typeName == 'star' || item.typeName == 'polygon');
                        },
                        changeProperty: {
					 		caption: 'Shape\'s number of nodes',
					 		rerender: true
					 	}
                    }],[
                    {
                        caption: 'Depth',
                        name: 'spikeDepth',
                        proptype: 'star-spike-depth',
                        type: 'slider-input',
		                values: { min: 2, max: 10, step: 1 },
                        value: function(item,name) {
                            return item.spikeDepth;
                        },
                        hiddenByDefault: function (item, name) {
                            return item.typeName != 'star';
                        },
                        changeProperty: {
					 		caption: 'Star Spike Depth',
					 		rerender: true
					 	}
                    }
      			],
      			[
                    {
                        caption: 'Skew',
                        name: 'skew',
                        proptype: 'paralellogram-skew',
                        type: 'slider-input',
		                values: { min: 0, max: 90, step: 10 },
                        value: function(item,name) {
                            return item.skew;
                        },
                        hiddenByDefault: function (item, name) {
                            return item.typeName != 'parallelogram';
                        },
                        changeProperty: {
					 		caption: 'Parallelogram Skew Intensity',
					 		rerender: true
					 	}
                    }
      			],
      			[
                    {
                        caption: 'Top Base % Size',
                        name: 'baseWidth',
                        proptype: 'trapezoid-base-width',
                        type: 'slider-input',
                        values: { min: 2, max: 100, step: 2 },
                        value: function(item,name) {
                            return item.baseWidth;
                        },
                        hiddenByDefault: function (item, name) {
                            return item.typeName != 'trapezoid';
                        },
                        changeProperty: {
					 		caption: 'Trapezoid Top Base Width',
					 		rerender: true
					 	}
                    }
      			]
  	      	]
  	   }
    ]
};

/***** SPECIAL COMPONENTS *****/

// TYPE: SYMBOL
prx.types.symbol = {
    name: 'symbol'
    ,onDisplay: function(item,containerid,symbol) {

        var isAppleWatchCrown = ((typeof(prx.devices[prx.device]) != 'undefined' && prx.devices[prx.device].scrollType == 'crown') || (typeof(prx.deviceExtraParams)!='undefined' && prx.deviceExtraParams.scrollType == 'crown')) ? true : false;
        var _id = (!containerid) ? item.id : containerid+'-'+item.id;
        var _symbolindex = prx.stc.containers.getIndexFromId(prx.componentsHelper.getProp(item.symbolid,'other'));

        if( typeof(item.scrollToX) == 'undefined' ) { item.scrollToX = 0;}
        if( typeof(item.scrollToY) == 'undefined' ) { item.scrollToY = 0;}

        var cReturn ='';

        cReturn += '<style>';
        if( prx.editor ) {
            if( prx.componentsHelper.getProp(item.scroll,'other') == 'omni' ) {
                cReturn += '#' + _id + '-scroll { top: -'+ item.scrollToY +'px; left: -'+ item.scrollToX +'px; }';
            }
            else if( prx.componentsHelper.getProp(item.scroll,'other') == 'vertical' ) {
                cReturn += '#' + _id + '-scroll { top: -'+ item.scrollToY +'px; }';
            }
            else if( prx.componentsHelper.getProp(item.scroll,'other') == 'horizontal' ) {
                cReturn += '#' + _id + '-scroll { left: -'+ item.scrollToX +'px; }';
            }
        }
        cReturn += '</style>';


        if (_symbolindex > -1) {
            var _symbol_width='100%';
            var _symbol_height='100%';

            if(typeof(prxy.symbols[_symbolindex].states) == 'undefined') {
                var _bg = prxy.symbols[_symbolindex].background;
                var dataState = '';
            } else {
                var stateindex = 0;
                if(typeof(item.symbolstateid) != 'undefined') {
                    stateindex = prx.stc.containers.getStateIndexFromId(prx.componentsHelper.getProp(item.symbolstateid,'other'), prx.stc.containers.getIndexFromId(prx.componentsHelper.getProp(item.symbolid,'other')));
                    if(stateindex == -1) {
                        stateindex = 0;
                        item.symbolstateid = prxy.symbols[_symbolindex].states[stateindex].id;
                    }
                }
                var _bg = prxy.symbols[_symbolindex].states[stateindex].background;
                _symbol_width = prxy.symbols[_symbolindex].states[stateindex].dimensions[0]+'px';
                _symbol_height = prxy.symbols[_symbolindex].states[stateindex].dimensions[1]+'px';
                var dataState = 'data-state-id="'+prxy.symbols[_symbolindex].states[stateindex].id+'"';
            }

            if (prx.componentsHelper.getProp(item.scroll,'other')!='') {
                if (prx.componentsHelper.getProp(item.scroll,'other')=='horizontal' || prx.componentsHelper.getProp(item.scroll,'other')=='omni') {
                    if(typeof(prxy.symbols[_symbolindex].dimensions) != 'undefined') {
                        _symbol_width = prxy.symbols[_symbolindex].dimensions[0]+'px';
                    } else {
                        _symbol_width = prxy.symbols[_symbolindex].states[stateindex].dimensions[0]+'px';
                    }
                }
                if (prx.componentsHelper.getProp(item.scroll,'other')=='vertical' || prx.componentsHelper.getProp(item.scroll,'other')=='omni') {
                    if(typeof(prxy.symbols[_symbolindex].dimensions) != 'undefined') {
                        _symbol_height = prxy.symbols[_symbolindex].dimensions[1]+'px';
                    } else {
                        _symbol_height = prxy.symbols[_symbolindex].states[stateindex].dimensions[1]+'px';
                    }
                }
            }

            var osWatchStyle = (prx.componentsHelper.getProp(item.scroll,'other') == 'vertical' && typeof(item.watchOSscrollbars)!== 'undefined' && prx.componentsHelper.getProp(item.watchOSscrollbars,'boolean') == true) ? true : false;
            /* /ADDED THESE LINES IN ORDER TO MAKE SYMBOLS BEHAVE RESPONSIVELY */

            if(typeof(item.propagateevents) == 'undefined') { item.propagateevents = false; }


            cReturn += '<div id="' + _id + '" class="box type-symbol ' + prx.items.getComponentBaseClasses(item, containerid, symbol) + ' ' +((prx.componentsHelper.getProp(item.propagateevents,'boolean') && (prx.componentsHelper.getProp(item.scroll,'other') == '' || prx.componentsHelper.getProp(item.scroll,'other') == 'none')) ? 'type-symbol-transparent' : '')+ ' scroll-' + prx.componentsHelper.getProp(item.scroll,'other') + '" data-symbol-id="'+prx.componentsHelper.getProp(item.symbolid,'other')+'" '+dataState+' style="position: absolute;" ' + prx.items.getComponentBaseAttributes(item, containerid, symbol)  + '>';


            var applewatchScrollbarClass = '';
            if(prx.componentsHelper.getProp(isAppleWatchCrown,'boolean') == true && osWatchStyle==true){
                var applewatchScrollbarClass = 'watchOSscrollbars';
                var appleWatch38mmWH = [272,340];
                var appleWatch42mmWH = [312,390];
                var appleWatch40mmWH = [324,394];
                // var appleWatch44mmWH = [368,448];
                var cssTop = 35;
                var cssLeft = 259;

                //var cssTop = (prx.devices[prx.device]['portrait'][0] == appleWatch38mmWH[0] && prx.devices[prx.device]['portrait'][1] == appleWatch38mmWH[1]) ? '35' : '53';
                if(prx.devices[prx.device]['portrait'][0] == appleWatch38mmWH[0] && prx.devices[prx.device]['portrait'][1] == appleWatch38mmWH[1]){
                    cssTop = 35;
                    cssLeft = 259;
                } else if(prx.devices[prx.device]['portrait'][0] == appleWatch42mmWH[0] && prx.devices[prx.device]['portrait'][1] == appleWatch42mmWH[1]) {
                    cssTop = 53;
                    cssLeft = 299;
                } else if(prx.devices[prx.device]['portrait'][0] == appleWatch40mmWH[0] && prx.devices[prx.device]['portrait'][1] == appleWatch40mmWH[1]) {
                    cssTop = 60;
                    cssLeft = 312;
                } else { //44mm
                    cssTop = 81;
                    cssLeft = 355;
                }

                var scrollbarWrapperHeight = 71;//(_symbol_dims.height*21)/100;//get 21% of screenheight in px = scrollbar container height
                var scrollbarIndicatorHeight = Math.max(Math.round(((item.height/parseInt(_symbol_height)))*scrollbarWrapperHeight),15);
                cReturn += '<style>';
                cReturn += '#'+_id+' .iScrollVerticalScrollbar {height:'+scrollbarWrapperHeight+'px!important;top:'+cssTop+'px!important;right:0!important;left: '+cssLeft+'px!important}';
                cReturn += '#'+_id+' .iScrollIndicator {height:'+scrollbarIndicatorHeight+'px!important;}';
                cReturn += '</style>';

            }
            cReturn += '<style>';
            cReturn += prx.items.getComponentBaseStyle(item, containerid, symbol);
            cReturn += '</style>';
            cReturn += prx.items.getComponentPrependDivs(item, containerid, symbol);
            cReturn += '<div id="'+_id+'-inner" class="'+applewatchScrollbarClass+'" style="position: absolute; overflow: hidden !important; height: 100%; width: 100%;">';
            cReturn += '<div id="' + _id + '-scroll" class="symbol-scroll" style="width: '+_symbol_width+'; height: '+_symbol_height+'; position: absolute; overflow: hidden; background-color: '+prx.componentsHelper.getProp(_bg,'color-background')+';">';
            cReturn += prx.stc.containers.render(item,containerid,stateindex);
            cReturn += '</div>';
            cReturn += '</div>';

            if (prx.editor && symbol === undefined  ) {
                cReturn += '<div class="type-symbol-hover-message">Double-click to edit container</div>';
            }
            cReturn += prx.items.getComponentAppendDivs(item, containerid, symbol);
            cReturn += '</div>';
        } else {
            _appenditem = false;
            if(prx.editor) {
                return prx.componentsHelper.missingComponent(item,containerid);
            }
        }

        return cReturn;
    }
    ,afterDisplayWaitForCB: true
    ,afterDisplay: function(item, containerid, symbol, callback) {
        var _id = (!containerid) ? item.id : containerid+'-'+item.id;

        var stateindex = 0;
        if(typeof(item.symbolstateid) != 'undefined') {
            stateindex = prx.stc.containers.getStateIndexFromId(item.symbolstateid, prx.stc.containers.getIndexFromId(item.symbolid));
            if(stateindex == -1) {
                stateindex = 0;
            }
        }
        prx.stc.containers.loadRecursiveAfterDisplay(item,containerid, stateindex, callback);
    },
    onResize: function(item, containerid, symbol) {

        var _id = (!containerid) ? item.id : containerid+'-'+item.id;
        var _symbolindex = prx.stc.containers.getIndexFromId(prx.componentsHelper.getProp(item.symbolid,'other'));

        if(_symbolindex > -1) {
            var _symbol_width='100%';
            var _symbol_height='100%';

            var stateindex = 0;
            if(typeof(item.symbolstateid) != 'undefined') {
                stateindex = prx.stc.containers.getStateIndexFromId(prx.componentsHelper.getProp(item.symbolstateid,'other'), prx.stc.containers.getIndexFromId(prx.componentsHelper.getProp(item.symbolid,'other')));
                if(stateindex == -1) {
                    stateindex = 0;
                }

                _symbol_width = prxy.symbols[_symbolindex].states[stateindex].dimensions[0]+'px';
                _symbol_height = prxy.symbols[_symbolindex].states[stateindex].dimensions[1]+'px';

            }
            if(!prx.editor) {
                var stateid = $('#' + _id).attr('data-state-id');
                var tempstateindex = prx.stc.containers.getStateIndexFromId(stateid, _symbolindex);
                if(tempstateindex !== -1) stateindex = tempstateindex;
            }

            if (prx.componentsHelper.getProp(item.scroll,'other')!='') {
                if (prx.componentsHelper.getProp(item.scroll,'other')=='horizontal' || prx.componentsHelper.getProp(item.scroll,'other')=='omni') {
                    if(typeof(prxy.symbols[_symbolindex].dimensions) != 'undefined') {
                        _symbol_width = prxy.symbols[_symbolindex].dimensions[0]+'px';
                    } else {
                        _symbol_width = prxy.symbols[_symbolindex].states[stateindex].dimensions[0]+'px';
                    }
                }
                if (prx.componentsHelper.getProp(item.scroll,'other')=='vertical' || prx.componentsHelper.getProp(item.scroll,'other')=='omni') {
                    if(typeof(prxy.symbols[_symbolindex].dimensions) != 'undefined') {
                        _symbol_height = prxy.symbols[_symbolindex].dimensions[1]+'px';
                    } else {
                        _symbol_height = prxy.symbols[_symbolindex].states[stateindex].dimensions[1]+'px';
                    }
                }
            }

            $('#' + _id + '-scroll').css({ 'width': _symbol_width, 'height': _symbol_height});
        }


    }
    ,interactions: [
        prx.commonproperties.actions
    ]
    ,propertyGroups: [
        {
            caption: 'Container State',
            properties: [[
              		{
              			caption: false
              			,name: 'symbolstateid'
              			,type: 'select'
              			,value: function(item,name) {
              				if(typeof(prxy.symbols[0])!='undefined' && typeof(prxy.symbols[0].states) == 'undefined') { return ''; }
		                    if(prx.stc.containers.getIndexFromId(item.symbolid) == -1) { return ''; }
              				if(typeof(item.symbolstateid) == 'undefined') {
              					item.symbolstateid = prxy.symbols[prx.stc.containers.getIndexFromId(item.symbolid)].states[0].id;
              				}
              				return item.symbolstateid;
              			}
      					,values: function(item){
      						if(typeof(prxy.symbols[0])!='undefined' && typeof(prxy.symbols[0].states) == 'undefined') { return []; }
		                if(prx.stc.containers.getIndexFromId(item.symbolid) == -1) { return []; }
      						var rA = [];
      						for(var i=0;i<prxy.symbols[prx.stc.containers.getIndexFromId(item.symbolid)].states.length;i++) {
      							rA.push({ displayValue: prx.utils.escapeHTML(prxy.symbols[prx.stc.containers.getIndexFromId(item.symbolid)].states[i].title), value: prxy.symbols[prx.stc.containers.getIndexFromId(item.symbolid)].states[i].id });
      						}
      						return rA;
      					}
      					,hiddenByDefault: function(item) {
      						if(typeof(prxy.symbols[0])!='undefined' &&  typeof(prxy.symbols[0].states) == 'undefined') { return true; }
		                    if(prx.stc.containers.getIndexFromId(item.symbolid) == -1) { return true; }
      						if(prxy.symbols[prx.stc.containers.getIndexFromId(item.symbolid)].states.length == 1) {
      							return true;
      						}
      						return false;
      					}
      					,onChange: function() {
		                    prx.stc.canvasMap.updateSymbolsMap('attach_cont');
                        	prx.screenshots.addIconToOutdated();
                    }
      					,changeProperty: {
      						caption: 'Container state',
      						selector: '',
      						property: 'container-state',
      						transitionable: true
      					}
	      			}]]
	    },
        {
            caption: 'Scroll properties',
            properties: [[
                {
                    caption: 'Scroll'
                    ,name: 'scroll'
                    ,type: 'select'
                    ,value: function(item,name) {
                        return item.scroll;
                    }
                    ,values: [{value: 'none',displayValue: 'No'},{value: 'horizontal',displayValue: 'Horizontally'},{value: 'vertical',displayValue: 'Vertically'},{value: 'omni',displayValue: 'Omni-directional'}]
                    ,onChange: function(item){

                        if(item.scroll == '' || item.scroll == 'none') {
                            $('#property-scrollsnap, #property-pinchzoom, #property-momentum, #property-scrollbars, #property-lockdirection, #property-bounce, #property-watchOSscrollbars, #property-scrollToX, #property-scrollToY').hide();
                            $('#property-propagateevents').show();
                        }
                        else {

                            $('#property-scrollsnap, #property-pinchzoom, #property-momentum, #property-bounce, #property-scrollbars').show();
                            $('#property-propagateevents').hide();
                            if(item.scroll == 'omni') {
                                $('#property-lockdirection').show();

                                if(typeof(item.scrollbars) == 'undefined') {
                                    if(item.vscrollbar || item.hscrollbar) {
                                        $('#p-'+item.id+'-scrollbars').prop('checked', true);
                                    } else {
                                        $('#p-'+item.id+'-scrollbars').prop('checked', false);
                                    }
                                }
                            } else {
                                $('#property-lockdirection').hide();
                                if(typeof(item.scrollbars) == 'undefined') {
                                    if(item.scroll == 'horizontal') {
                                        if(item.hscrollbar) {
                                            $('#p-'+item.id+'-scrollbars').prop('checked', true);
                                        } else {
                                            $('#p-'+item.id+'-scrollbars').prop('checked', false);
                                        }
                                    }
                                    if(item.scroll == 'vertical') {

                                        if(item.vscrollbar) {
                                            $('#p-'+item.id+'-scrollbars').prop('checked', true);
                                        } else {
                                            $('#p-'+item.id+'-scrollbars').prop('checked', false);
                                        }
                                    }
                                }
                            }

                            if(item.scroll == 'vertical' && prx.devices[prx.device].scrollType == 'crown' && item.scrollbars == true){
                                $('#property-watchOSscrollbars').show();
                            }else{
                                $('#property-watchOSscrollbars').hide();
                            }

                            /* for scroll to position */
                            if( prx.componentsHelper.getProp(item.scroll,'other') == 'omni' ) {
                                $('#property-scrollToX, #property-scrollToY').show();
                            }
                            else if( prx.componentsHelper.getProp(item.scroll,'other') == 'horizontal' ) {
                                $('#property-scrollToX').show();
                                $('#property-scrollToY').hide();
                            }
                            else if( prx.componentsHelper.getProp(item.scroll,'other') == 'vertical' ) {
                                $('#property-scrollToX').hide();
                                $('#property-scrollToY').show();
                            }
                        }

                    }
                    ,changeProperty: {
                        caption: 'Scroll',
                        rerender: true
                    }
                }],[{
                caption: 'Treat scroll container as snap (carousel)'
                ,name: 'scrollsnap'
                ,type: 'onoff'
                ,value: function(item,name) {
                    return item.scrollsnap;
                }
                ,hiddenByDefault: function(item){
                    return (item.scroll=='' || item.scroll=='none');
                }
                ,changeProperty: {
                    caption: 'Snap',
                    rerender: true
                }
            },
            {
                caption: 'Allow scrolling beyond container boundaries'
                ,name: 'bounce'
                ,type: 'onoff'
                ,value: function(item,name) {
                    if(typeof(item.bounce) == 'undefined') {
                        item.bounce = true;
                    }
                    return item.bounce;
                }
                ,hiddenByDefault: function(item){
                    return (item.scroll=='' || item.scroll=='none');
                }
                ,changeProperty: {
                    caption: 'Scroll beyond boundaries',
                    rerender: true
                }
            },{
                caption: 'Enable Inertia'
                ,name: 'momentum'
                ,type: 'onoff'
                ,value: function(item,name) {
                    return item.momentum;
                }
                ,hiddenByDefault: function(item){
                    return (item.scroll=='' || item.scroll=='none');
                }
                ,changeProperty: {
                    caption: 'Inertia',
                    rerender: true
                }
            },
            {
                caption: 'Enable Pinch / Zoom'
                ,name: 'pinchzoom'
                ,type: 'onoff'
                ,value: function(item,name) {
                    return item.pinchzoom;
                }
                ,hiddenByDefault: function(item){
                    return (item.scroll=='' || item.scroll=='none');
                }
                ,changeProperty: {
                    caption: 'Pinch/Zoom',
                    rerender: true
                }
            },
            {
                caption: 'Scrollbars'
                ,name: 'scrollbars'
                ,type: 'select'
                ,values: [{value: 'OFF',displayValue: 'Don\'t show scrollbars'},{value: 'FADE',displayValue: 'Visible on scroll only'},{value: 'ALWAYS',displayValue: 'Always visible'}]
                ,value: function(item,name) {

                    let _scrollbars = item.scrollbars;

                    // previous compatibility - from hscrollbar and vscrollbar to plain scrollbar
                    // it would be a true/false value of whether scrollbar would be visible - on scroll only
                    if(typeof(_scrollbars) == 'undefined') {
                        switch (item.scroll) {
                            case 'omni':
                                _scrollbars = (eval(item.hscrollbar) || eval(item.vscrollbar)) ? true : false;
                                break;
                            case 'horizontal':
                                _scrollbars = (eval(item.hscrollbar)) ? true : false;
                                break;
                            case 'vertical':
                                _scrollbars = (eval(item.vscrollbar)) ? true : false;
                                break;
                            default:
                                break;
                        }
                    }

                    // new compatibility - want to convert scrollbar to select so that you can choose between
                    // "off", "visible on scroll only" and "always visible"
                    switch(_scrollbars) {
                        case(false):
                            return "OFF";
                        case(true):
                            return "FADE";
                        default:
                            return _scrollbars;
                    }
                }
                ,onChange: function(item) {
                    if(item.scroll =='vertical' && prx.devices[prx.device].deviceType == 'applewatch' && (item.scrollbars == 'FADE' || item.scrollbars == 'ALWAYS')) {
                        $('#property-watchOSscrollbars').show();
                    } else {
                        $('#property-watchOSscrollbars').hide();
                    }
                }
                ,hiddenByDefault: function(item){
                    return (item.scroll=='' || item.scroll=='none');
                }
                ,changeProperty: {
                    caption: 'Scrollbars',
                    rerender: true
                }
            },
            {
                caption: 'watchOS styled scrollbars'
                ,name: 'watchOSscrollbars'
                ,type: 'onoff'
                ,value: function(item,name) {
                    if(typeof(item.watchOSscrollbars) == 'undefined') {
                        return false;
                    }
                    return item.watchOSscrollbars;
                }
                ,hiddenByDefault: function(item){
                    return (!(item.scroll == 'vertical' && prx.devices[prx.device].deviceType == 'applewatch' && item.scrollbars == true));
                }
                ,changeProperty: {
                    caption: 'watchOS styled scrollbars',
                    rerender: true
                }
            },
            {
                caption: 'Lock direction on scroll'
                ,name: 'lockdirection'
                ,type: 'onoff'
                ,value: function(item,name) {
                    if(typeof(item.lockdirection) == 'undefined') {
                        return true;
                    }
                    return item.lockdirection;
                }
                ,hiddenByDefault: function(item){
                    return (item.scroll!='omni');
                }
                ,changeProperty: {
                    caption: 'Lock direction',
                    rerender: true
                }
            },{
                caption: 'Background'
                ,name: 'background'
                ,proptype: 'background-color'
                ,type: 'colorpicker'
                ,value: function(item,name) {
                    return item.background;
                }
                ,hiddenByDefault: function(item){
                    return true;
                },
                changeProperty: {
                    caption: 'Background Color',
                    selector: '.symbol-scroll',
                    property: 'background-color',
                    transitionable: true
                }
            },
            {
                caption: 'Initial horizontal position',
                name: 'scrollToX',
                proptype: 'scroll-to',
                type: 'input-numeric',
                value: function(item,name) {
                    return item.scrollToX;
                },
                values: {
                    min: 0,
                    max: function(item) {
                        let symbol = prxy.symbols[prx.stc.containers.getIndexFromId(item.symbolid)];
                        if(typeof(symbol) === "undefined") return 0;
                        let state = symbol.states[prx.stc.containers.getStateIndexFromId(item.symbolstateid)] || symbol.states[0];
                        return parseInt(state.dimensions[0]) - parseInt(item.width);
                    },
                    step: 1
                },
                hiddenByDefault: function(item,name) {
                    return !((prx.componentsHelper.getProp(item.scroll,'other') == 'omni') || (prx.componentsHelper.getProp(item.scroll,'other') == 'horizontal'));
                },
                changeProperty: {
                    caption: 'Horizontal scroll position',
                    transitionable: true,
                    changeFunction: function(item, containerid, duration, easing) {
                        var _id = (!containerid) ? item.id : containerid+'-'+item.id;

                        item.scrollToX = item.scrollToX || 0;
                        item.scrollToY = item.scrollToY || 0;

                        var scrollable = prx.scrollable._scrollables[_id + '-inner'];

                        if(scrollable !== undefined) {

                            if(scrollable.options.snap) {

                                var pageX = Math.floor(Math.abs(item.scrollToX) / scrollable.wrapperWidth);
                                var pageY = Math.floor(Math.abs(item.scrollToY) / scrollable.wrapperHeight);

                                scrollable.goToPage(pageX, pageY, duration * 1000, prx.easing.getIScrollEasing(easing));

                            } else {
                                scrollable.scrollTo(Math.abs(item.scrollToX) * -1, Math.abs(item.scrollToY) * -1, duration * 1000, prx.easing.getIScrollEasing(easing));
                            }

                        }


                    }
                }
            },
            {
                caption: 'Initial vertical position',
                name: 'scrollToY',
                proptype: 'scroll-to',
                type: 'input-numeric',
                value: function(item,name) {

                    return item.scrollToY;
                },
                values: {
                    min: 0,
                    max: function(item) {
                        let symbol = prxy.symbols[prx.stc.containers.getIndexFromId(item.symbolid)];
                        if(typeof(symbol) === "undefined") return 0;
                        let state = symbol.states[prx.stc.containers.getStateIndexFromId(item.symbolstateid)] || symbol.states[0];
                        return parseInt(state.dimensions[1]) - parseInt(item.height);
                    },
                    step: 1
                },
                hiddenByDefault: function(item,name) {
                    return !((prx.componentsHelper.getProp(item.scroll,'other') == 'omni') || (prx.componentsHelper.getProp(item.scroll,'other') == 'vertical'));
                },
                changeProperty: {
                    caption: 'Vertical scroll position',
                    transitionable: true,
                    changeFunction: function(item, containerid, duration, easing) {
                        var _id = (!containerid) ? item.id : containerid+'-'+item.id;

                        item.scrollToX = item.scrollToX || 0;
                        item.scrollToY = item.scrollToY || 0;

                        var scrollable = prx.scrollable._scrollables[_id + '-inner'];

                        if(scrollable !== undefined) {

                            if(scrollable.options.snap) {

                                var pageX = Math.floor(Math.abs(item.scrollToX) / scrollable.wrapperWidth);
                                var pageY = Math.floor(Math.abs(item.scrollToY) / scrollable.wrapperHeight);

                                scrollable.goToPage(pageX, pageY, duration * 1000, prx.easing.getIScrollEasing(easing));

                            } else {
                                scrollable.scrollTo(Math.abs(item.scrollToX) * -1, Math.abs(item.scrollToY) * -1, duration * 1000, prx.easing.getIScrollEasing(easing));
                            }

                        }

                    }
                }
            }
	      	]]
	    },
        {
            caption: 'Advanced',
            properties: [[
                {
                    caption: 'In empty areas, layers behind the container can be interacted with'
                    ,name: 'propagateevents'
                    ,type: 'onoff'
                    ,value: function(item,name) {
                        if(typeof(item.propagateevents) == 'undefined') {
                            return false;
                        }
                        return item.propagateevents;
                    }
                    ,hiddenByDefault: function(item){
                        return (item.scroll!='' && item.scroll!='none');
                    }
                    ,changeProperty: {
                        caption: 'Allow interactions behind empty areas',
                        rerender: true
                    }
                }
            ]]
        }
    ]
};

prx.types.vrsymbol = {
    name: 'vrsymbol'
    ,onBeforeCreate: function (item, containerid, symbol) {

    	// check if
        if (prx.userPreferences.vrFirstTimeModal !== true) {
            // show modal

            prx.vr.showFirstTimeModal();

            // update settings on db
            prx.userPreferences.vrFirstTimeModal = true;
            prx.interface.updateDatabase();

            return false;
        }

        return true;

    }
    ,onCreate: function (item, containerid, symbol) {
    	var _settings = {};

        _settings.title = prx.stc.helper.namingConvention('', 'vrsymbols');
        _settings.widthMultiplier = 1;
        _settings.heightMultiplier = 1;
        _settings.width = 4096;
        _settings.height = 2048;
        _settings.background = 'none';
        _settings.vr = true;

        prx.stc.containers.backgroundNew(_settings);
        prx.stc.pane.updateContainersList();

        item.symbolid = prxy.symbols[prxy.symbols.length - 1].id;

        prx.vr.loadDefaultScene(item.symbolid);

    }
    ,onDisplay: function(item,containerid,symbol) {

        var _id = (!containerid) ? item.id : containerid+'-'+item.id;
        var _symbolindex = prx.stc.containers.getIndexFromId(prx.componentsHelper.getProp(item.symbolid,'other'));

        var cReturn ='';


        if (_symbolindex > -1) {

            if(typeof(prxy.symbols[_symbolindex].states) == 'undefined') {
                var _bg = prxy.symbols[_symbolindex].background;
                var dataState = '';
            } else {
                var stateindex = 0;
                if(typeof(item.symbolstateid) != 'undefined') {
                    stateindex = prx.stc.containers.getStateIndexFromId(prx.componentsHelper.getProp(item.symbolstateid,'other'), prx.stc.containers.getIndexFromId(prx.componentsHelper.getProp(item.symbolid,'other')));
                    if(stateindex == -1) {
                        stateindex = 0;
                        item.symbolstateid = prxy.symbols[_symbolindex].states[stateindex].id;
                    }
                }
                var _bg = prxy.symbols[_symbolindex].states[stateindex].background;
                var dataState = 'data-state-id="'+prxy.symbols[_symbolindex].states[stateindex].id+'"';
            }

            cReturn += '<div id="' + _id + '" class="box type-vrsymbol ' + prx.items.getComponentBaseClasses(item, containerid, symbol) + ' " data-symbol-id="'+prx.componentsHelper.getProp(item.symbolid,'other')+'" '+dataState+' style="position: absolute; background-color: '+prx.componentsHelper.getProp(_bg,'color-background')+';" ' + prx.items.getComponentBaseAttributes(item, containerid, symbol)  + '>';
            cReturn += '<style>';
            cReturn += prx.items.getComponentBaseStyle(item, containerid, symbol);
            cReturn += '</style>';
            cReturn += prx.items.getComponentPrependDivs(item, containerid, symbol);
            cReturn +='<div id="'+_id+'-inner" class="vrsymbol-inner" style="position: absolute; overflow: hidden !important; height: 100%; width: 100%;">';
            cReturn += prx.vr.render(item, containerid, stateindex, _symbolindex);
            cReturn += '</div>';
            if (prx.editor) {
                cReturn += '<div class="type-vrsymbol-hover-message">Double-click to edit VR Scene</div>';
            }
            cReturn += prx.items.getComponentAppendDivs(item, containerid, symbol);
            cReturn += '</div>';
        } else {
            if(prx.editor) {
                return prx.componentsHelper.missingComponent(item,containerid);
            }
        }
        return cReturn;
    }
    ,afterDisplay: function(item, containerid, symbol) {
        var _id = (!containerid) ? item.id : containerid+'-'+item.id;

        // var stateindex = 0;
        // if(typeof(item.symbolstateid) != "undefined") {
        //     stateindex = prx.stc.containers.getStateIndexFromId(item.symbolstateid, prx.stc.containers.getIndexFromId(item.symbolid));
        //     if(stateindex == -1) {
        //         stateindex = 0;
        //     }
        // }
        if(!prx.editor){
        	prx.vr.ini(item, containerid);
        }

        // because actionareas, images, rectangles dont have afterdisplay, this is currenlty not needed
        //prx.stc.containers.loadRecursiveAfterDisplay(item,containerid, stateindex);
    }
    ,propertyGroups: [
        {
            caption: 'Container State',
            properties: [[
                {
                    caption: false
                    ,name: 'symbolstateid'
                    ,type: 'select'
                    ,value: function(item,name) {
                        if(typeof(prxy.symbols[0])!='undefined' && typeof(prxy.symbols[0].states) == 'undefined') { return ''; }
                        if(prx.stc.containers.getIndexFromId(item.symbolid) == -1) { return ''; }
                        if(typeof(item.symbolstateid) == 'undefined') {
                            item.symbolstateid = prxy.symbols[prx.stc.containers.getIndexFromId(item.symbolid)].states[0].id;
                        }
                        return item.symbolstateid;
                    }
                    ,values: function(item){
                        if(typeof(prxy.symbols[0])!='undefined' && typeof(prxy.symbols[0].states) == 'undefined') { return []; }
                        if(prx.stc.containers.getIndexFromId(item.symbolid) == -1) { return []; }
                        var rA = [];
                        for(var i=0;i<prxy.symbols[prx.stc.containers.getIndexFromId(item.symbolid)].states.length;i++) {
                            rA.push({ displayValue: prx.utils.escapeHTML(prxy.symbols[prx.stc.containers.getIndexFromId(item.symbolid)].states[i].title), value: prxy.symbols[prx.stc.containers.getIndexFromId(item.symbolid)].states[i].id });
                        }
                        return rA;
                    }
                    ,hiddenByDefault: function(item) {
                        if(typeof(prxy.symbols[0])!='undefined' &&  typeof(prxy.symbols[0].states) == 'undefined') { return true; }
                        if(prx.stc.containers.getIndexFromId(item.symbolid) == -1) { return true; }
                        if(prxy.symbols[prx.stc.containers.getIndexFromId(item.symbolid)].states.length == 1) {
                            return true;
                        }
                        return false;
                    }
                    ,changeProperty: {
                        caption: 'Container state',
                        selector: '',
                        property: 'container-state',
                        transitionable: false
                    }
                }]]
        }
    ]
};


/***** /SPECIAL COMPONENTS *****/

/***** BASIC COMPONENTS *****/

//TYPE: TEXT
prx.types.text = {
    name: 'text'
    ,onDisplay: function(item,containerid,symbol) {
        var _id = (!containerid) ? item.id : containerid+'-'+item.id;

        var _props = (typeof(prx.richtext)=='undefined') ? prx.componentsHelper.getProp(item.textProperties,'props-text') : '';

        var _shadow = (item.enableShadow) ? ' text-shadow: 0 1px 0 #FFFFFF;' : '';

        if(typeof(item.propagateEvents) == 'undefined') { item.propagateEvents = false; }
        if(typeof(item.lineHeight) == 'undefined') { item.lineHeight = parseInt(prx.componentsHelper.getProp(item.textSize,'num-text-size')*1.231); }
        if(typeof(item.lineHeightAuto) == 'undefined') { item.lineHeightAuto = Boolean(true); }
        if(typeof(item.textSpacing) == 'undefined') { item.textSpacing = 0; }
        var cR = '<div id="' + _id + '" data-width='+item.width+' ' + prx.items.getComponentBaseAttributes(item, containerid, symbol)  + ' class="' + prx.items.getComponentBaseClasses(item, containerid, symbol) + ' box pos type-text '+(prx.componentsHelper.getProp(item.propagateEvents,'boolean') ? 'pointer-events-none ' : '')+ 'liveUpdate-textColor-color changeProperty-textColor changeProperty-textAlign changeProperty-lineHeight changeProperty-textSpacing ' + (item.v2!==undefined && item.v2 == true ? 'v2-text ' : '')+'" style="line-height: '+prx.componentsHelper.getProp(item.lineHeight,'num-other')+'px; letter-spacing: ' + prx.componentsHelper.getProp(item.textSpacing, 'num-text-size') + 'px; color: '+prx.componentsHelper.getProp(item.textColor,'color-text')+'; '+prx.componentsHelper.getProp(item.textFont + '|' + item.textFontStyle,'font-family') + prx.componentsHelper.getProp(item.textFontStyle,'font-style') + ' font-size: '+prx.componentsHelper.getProp(item.textSize,'num-text-size')+'px; '+_props+_shadow+' text-align: '+prx.componentsHelper.getProp(item.textAlign,'align')+';">';
        cR += '<style>';
        cR += prx.items.getComponentBaseStyle(item, containerid, symbol);
        cR += '</style>';
        cR += prx.items.getComponentPrependDivs(item, containerid, symbol);
        cR += '<div class="text-contents '+(prx.componentsHelper.getProp(item.autoResize,'boolean')==true ? ' autoResize-true ' : '')+ ' liveUpdate-backgroundColor-background-color changeProperty-backgroundColor " style="'+prx.gradients.getBgCssByProperty(item.backgroundColor,'color-background')+';">';
        cR += '<span data-editableproperty="text" '+(prx.componentsHelper.getProp(item.autoResize,'boolean')==true ? 'class="' + (prx.editor ? 'autoresize' : '') + '"' : '')+'>'+prx.componentsHelper.getProp(item.text,'text-textarea')+'</span>';
        cR += '</div>';
        cR += prx.items.getComponentAppendDivs(item, containerid, symbol);
        cR += '</div>';
        return cR;
    },
    onResize: function(item,containerid,symbol) {
        if (prx.editor) {

            // disable autoresize
            prx.componentsHelper.setAutoresizeValue(item, containerid, symbol, false);

            var $itemDiv = $('#' + item.id);

            if (item.v2 === undefined) {
                item.v2 = true;
                $itemDiv.addClass('v2-text');
            }

            // var $innerTextDivHeight = $('#' + item.id + ' [data-editableproperty="text"]').parent().height();
            let itemSelector= $('#' + item.id + ' [data-editableproperty="text"]');
            let $innerTextDivHeight = itemSelector.height();
            let textareaIsActive = itemSelector.find('#editable-properties-text').length > 0 ? true : false;

            if($innerTextDivHeight === 0 && textareaIsActive && prxy.iSelectedIndex > -1){
                let selectedIndex = prxy.iSelectedIndex;
                prx.editingtools.select.deselectAll();
                itemSelector= $('#' + item.id + ' [data-editableproperty="text"]');
                $innerTextDivHeight = itemSelector.height();
                prxy.iSelectedIndex = selectedIndex;
                prx.editingtools.select.refreshSelection();
            }

            // var itemHeight = item.height;
            // if ($innerTextDivHeight > itemHeight) {
            item.height = $innerTextDivHeight;
            $itemDiv.height($innerTextDivHeight);
            // }
        }
    }
    ,onCreate: function(item,containerid,symbol) {
        for(var key in prx.componentsHelper.memorisedTextStyle.style){
            if(!item.hasOwnProperty(key)){
                continue;
            }
            item[key] = prx.componentsHelper.memorisedTextStyle.style[key]
        }
        // if(prx.componentsHelper.memorisedTextStyle.style.textProperties.includes('underline')){
        //     item.text = '<u>' + item.text + '</u>'
        // }
    }
    ,editableProperties: [
        {
            caption: 'Text',
            name: 'text',
            type: 'textarea',
            value: function (item, name) {
                return item.text;
            },
            changeProperty: {
                caption: 'Text',
                selector: '.text-contents',
                property: 'text',
                transitionable: false
            },
            onChange: function(item,that,value) {


                if(that===undefined) {
                    return;
                }

                if (prx.editor) {

                    var $item = $('#' + item.id);

                    item.text = value;

                    var $iframeRichTextEL = $('.fr-iframe');

                    if (item.v2 === undefined) {
                        item.v2 = true;
                        $item.addClass('v2-text');
                    }


                    var isAutoResize = item.autoResize == true ? true : false;
                    var wordWrapVal = isAutoResize == true ? 'nowrap' : 'normal';
                    var tempWidth = isAutoResize == true ? 'auto' : item.width;

                    var _$item = $('#'+item.id+' [data-editableproperty="text"]');
                    var $temp = $('<span>'+item.text+'</span>')
                        .css({
                            display: 'inline-block',
                            'font-size': _$item.css('font-size'),
                            'font-family': _$item.css('font-family'),
                            'line-height': _$item.css('line-height'),
                            'text-decoration': _$item.css('text-decoration'),
                            'font-weight': _$item.css('font-weight'),
                            'font-style': _$item.css('font-style'),
                            'letter-spacing': _$item.css('letter-spacing'),
                            width: tempWidth,
                            height: 'auto',
                            marginLeft: '5px',
                            wordWrap: 'break-word',
                            whiteSpace: wordWrapVal
                        })
                        .appendTo('body');


                    var _height = isNaN(parseInt($temp.css('line-height'))) ? $temp.height() : Math.max(parseInt($temp.css('line-height')), $temp.height());
                    var _width = $temp.width()+5;

                    var _index = prx.items.helper.getIndexFromId(item.id);
                    var _dims = prx.toolbox.itemDimensions(prxy.items[_index]);
                    var oldItem = {
                            x: _dims.left,
                            y: _dims.top,
                            w: _dims.width,
                            h: _dims.height
                        },
                        newItem = {
                            x: _dims.left,
                            y: _dims.top,
                            w: _width,
                            h: _height
                        },
                        textAlign = prxy.items[_index].textAlign;

                    if(isAutoResize) {
                        var froalaEditor = $('#editable-properties-text').data('froala.editor');

                        if(typeof(froalaEditor)!='undefined' &&  typeof(froalaEditor.textAlign)!='undefined'){
                            textAlign = froalaEditor.textAlign
                                }
						$item.width(_width)
                            .attr('data-width', _width);
                                $iframeRichTextEL.width(_width).height(_height);
                        prxy.items[_index].width = _width;

                        }else{
						$iframeRichTextEL.width(item.width).height(_height);
						newItem.w = prxy.items[_index].width;
					}
                    let _align = textAlign === 'center' ? 'n' : textAlign === 'right' ? 'ne' : 'nw';
                    var positionWithOffset = prx.items.helper.repositionOnResize(
                            oldItem,
                            newItem,
                            _dims.rotation,
                            _align);
                    $iframeRichTextEL.width(_width).height(_height);
                    prxy.items[_index].left = positionWithOffset.x;
                    prxy.items[_index].top = positionWithOffset.y;

                    $item.css({'left': positionWithOffset.x, 'top': positionWithOffset.y, 'height': _height});

                    item.wtype = "fixed";
                    item.htype = "fixed";
                    item.height = _height;
                    prx.componentsHelper.memorisedTextStyle.update(item);
                    $temp.remove();
                }
            }
        }
    ]
    ,interactions: [
        {
            caption: 'Interactions',
            name: 'actions',
            type: 'action',
            value: function(item,name) {
                if (typeof(item.actions) == 'undefined') {
                    item.actions = [];
                }
                return item.actions.length;
            },
            hiddenByDefault: function(item){
                if(typeof(item.propagateEvents) != 'undefined' && item.propagateEvents) {
                    return true;
                }
                return false;
            },
            onChange: function(item) {
                if(prx.editor && item.v2===undefined){
                    item.v2 = true;
                    return item;
                }
            }
        }
    ]
    ,propertyGroups: [
        {
            caption: 'Text',
            properties: [
                [
                    {
                        caption: { label: 'Font family', class: 'text-properties-label text-fontfamily-label' },
                        name: 'textFont',
                        proptype: 'font-family',
                        type: 'select',
                        relatedEditableProperties: 'text',
                        relatedCSSProperties: 'font-family',
                        value: function (item, name) {
                            return item.textFont;
                        },
                        values: function () {
                            return prx.comps.fonts;
                        }
                        ,onChange: function(item) {
                            var isAutoResize = item.autoResize == true ? true : false;
                            var wordWrapVal = isAutoResize == true ? 'nowrap' : 'normal';
                            var $item = $('#' + item.id);
                            var tempWidth = isAutoResize == true ? 'auto' : item.width;
                            var $iframeRichTextEL = $('.fr-iframe');
                            var _$item = $('#'+item.id+' [data-editableproperty="text"]');


                            var $temp = $('<span>'+prx.componentsHelper.getProp(item.text,'text-textarea')+ '&nbsp;</span>')
                                .css({
                                    display: 'inline-block',
                                    'font-size': _$item.css('font-size'),
                                    'font-family': item.textFont,
                                    'line-height': _$item.css('line-height'),
                                    'text-decoration': _$item.css('text-decoration'),
                                    'font-weight': _$item.css('font-weight'),
                                    'font-style': _$item.css('font-style'),
                                    'letter-spacing': _$item.css('letter-spacing'),
                                    width: tempWidth,
                                    height: 'auto',
                                    marginLeft: '5px',
                                    wordWrap: 'break-word',
                                    whiteSpace: wordWrapVal
                                })
                                .appendTo('body');

                            var _height = isNaN(parseInt($temp.css('line-height'))) ? $temp.height() : Math.max(parseInt($temp.css('line-height')), $temp.height());
                            var _width = $temp.width()+5;

                    var _index = prx.items.helper.getIndexFromId(item.id),
                        _dims = prx.toolbox.itemDimensions(prxy.items[_index]),
                        oldItem = {
					        x: _dims.left,
                            y: _dims.top,
                            w: _dims.width,
                            h: _dims.height
                        },
                        newItem = {
					        x: _dims.left,
                            y: _dims.top,
                            w: _width,
                            h: _height
                        };

                        if(isAutoResize) {
                            $item.width(_width)
                                .attr('data-width',_width);
                                    $iframeRichTextEL.width(_width).height(_height);
                            prxy.items[_index].width = _width;
                        } else {
                            $iframeRichTextEL.width(item.width).height(_height);
                            newItem.w =_dims.width;
                        }
                        let _align = prxy.items[_index].textAlign === 'center' ? 'n' : prxy.items[_index].textAlign === 'right' ? 'ne' : 'nw';
                        positionWithOffset = prx.items.helper.repositionOnResize(
                            oldItem,
                            newItem,
                            _dims.rotation,
                            _align);
                        prxy.items[_index].left = positionWithOffset.x;
                        prxy.items[_index].top = positionWithOffset.y;

                        $item.css({'left': positionWithOffset.x, 'top': positionWithOffset.y, 'height': _height});

                        item.wtype = "fixed";
                        item.htype = "fixed";
                            item.height = _height;
                            $temp.remove();
                            var el = '#' + item.id + ' #editable-properties-text';
                            var isRichTextEditorOpen = $(el).data('froala.editor')!==undefined ? true : false;
                            if(isRichTextEditorOpen==false) {
                                prx.items.update(item,false);
                            }

                            if(prx.editor && item.v2===undefined){
                                item.v2 = true;
                                return item;
                            }
                            prx.componentsHelper.memorisedTextStyle.update(item);
                        }, changeProperty: {
  						caption: 'Text font',
  						selector: '.liveUpdate-textColor-color',
  						property: 'font-family',
  						transitionable: false
  					}
            	}],
                [
                    {

                        caption: { label: 'Font style', class: 'text-properties-label text-fontstyle-label' },
                        name: 'textFontStyle',
                        proptype: 'font-style',
                        type: 'select',
                        relatedEditableProperties: 'text',
                        relatedCSSProperties: ['font-weight', 'font-style'],
                        pffSettings: prx.commonproperties.pffSettingsTextFontStyle,
                        value: function (item, name) {
                            return item.textFontStyle === undefined ? '' : item.textFontStyle;
                        },
                        values:  function() {
                            return prx.comps.fontStyles;
                        },
                        onChange: function(item) {
                            var isAutoResize = item.autoResize == true ? true : false;
                            var wordWrapVal = isAutoResize == true ? 'nowrap' : 'normal';
                            var $item = $('#' + item.id);
                            var tempWidth = isAutoResize == true ? 'auto' : item.width;
                            var $iframeRichTextEL = $(prx.htmlTextEditor.iframeEl());
                            var _$item = $('#'+item.id+' [data-editableproperty="text"]');
                            var fontWeight = prx.componentsHelper.getProp(item.textFontStyle,'font-weight-value');
                            var fontStyle = prx.componentsHelper.getProp(item.textFontStyle,'font-style-value');

                            if(item.lineHeightAuto==true) {
                                item.lineHeight = parseInt(prx.componentsHelper.getProp(item.textSize,'num-text-size')*1.231);
                            }

                            var $temp = $('<span>'+item.text+ '&nbsp;</span>')
                                .css({
                                    display: 'inline-block',
                                    'font-size': item.textSize + 'px',
                                    'font-family': _$item.css('font-family'),
                                    'line-height': item.lineHeight + 'px',
                                    'text-decoration': _$item.css('text-decoration'),
                                    'font-weight': fontWeight || _$item.css('font-weight'),
                                    'font-style': fontStyle || _$item.css('font-style'),
                                    'letter-spacing': _$item.css('letter-spacing'),
                                    width: tempWidth,
                                    height: 'auto',
                                    marginLeft: '5px',
                                    wordWrap: 'break-word',
                                    whiteSpace: wordWrapVal
                                })
                                .appendTo('body');

                            var _height = isNaN(parseInt($temp.css('line-height'))) ? $temp.height() : Math.max(parseInt($temp.css('line-height')), $temp.height());
                            var _width = $temp.width()+5;

                            var _index = prx.items.helper.getIndexFromId(item.id),
                                _dims = prx.toolbox.itemDimensions(prxy.items[_index]),
                                oldItem = {
                                    x: _dims.left,
                                    y: _dims.top,
                                    w: _dims.width,
                                    h: _dims.height
                                },
                                newItem = {
                                    x: _dims.left,
                                    y: _dims.top,
                                    w: _width,
                                    h: _height
                                };

                            if(isAutoResize) {
                                $item.width(_width)
                                    .attr('data-width',_width);
                                $iframeRichTextEL.width(_width).height(_height);
                                prxy.items[_index].width = _width;
                            }else {
                                $iframeRichTextEL.width(item.width).height(_height);
                                newItem.w = _dims.width;
                            }
                            let _align = prxy.items[_index].textAlign === 'center' ? 'n' : prxy.items[_index].textAlign === 'right' ? 'ne' : 'nw';
                            positionWithOffset = prx.items.helper.repositionOnResize(
                                oldItem,
                                newItem,
                                _dims.rotation,
                                _align);
                            prxy.items[_index].left = positionWithOffset.x;
                            prxy.items[_index].top = positionWithOffset.y;

                            $item.css({'left': positionWithOffset.x, 'top': positionWithOffset.y, 'height': _height});

                            item.wtype = "fixed";
                            item.htype = "fixed";
                            item.height = _height;
                            $temp.remove();
                            var el = '#' + item.id + ' #editable-properties-text';
                            var isRichTextEditorOpen = $(el).data('froala.editor')!==undefined ? true : false;
                            if(isRichTextEditorOpen==false) {
                                prx.items.update(item,false);
                            }
                            $('#p-'+item.id+'-lineHeight').val(item.lineHeight);
                            if(prx.editor && item.v2===undefined){
                                item.v2 = true;
                                return item;
                            }
                            prx.componentsHelper.memorisedTextStyle.update(item);
                        },
                        hiddenByDefault: function(item, name, index) {
                            return false;
                        },
                        changeProperty: {
                            caption: 'Font style',
                            transitionable: false,
                            changeFunction: function(item, containerid, duration, easing, dynPropI){prx.componentsHelper.changeFunctionForTextFontStyleProperties(item, containerid, dynPropI, 'textFontStyle');}
                        }
                    },
                    {
                        caption: false,
                        name: 'textSize',
                        proptype: 'font-size',
                        type: 'combo-select',
                        relatedEditableProperties: 'text',
                        relatedCSSProperties: 'font-size',
                        value: function (item, name) {
                            return item.textSize;
                        },
                        values: prx.comps.textsize,
                        onChange: function(item) {
                            var isAutoResize = item.autoResize == true ? true : false;
                            var wordWrapVal = isAutoResize == true ? 'nowrap' : 'normal';
                            var $item = $('#' + item.id);
                            var tempWidth = isAutoResize == true ? 'auto' : item.width;
                            var $iframeRichTextEL = $(prx.htmlTextEditor.iframeEl());
                            var _$item = $('#'+item.id+' [data-editableproperty="text"]');

                            if(item.lineHeightAuto==true) {
                                item.lineHeight = parseInt(prx.componentsHelper.getProp(item.textSize,'num-text-size')*1.231);
                                prx.componentsHelper.memorisedTextStyle.update(item);
                            }

                            var $temp = $('<span>'+item.text+ '&nbsp;</span>')
                                .css({
                                    display: 'inline-block',
                                    'font-size': item.textSize + 'px',
                                    'font-family': _$item.css('font-family'),
                                    'line-height': item.lineHeight + 'px',
                                    'text-decoration': _$item.css('text-decoration'),
                                    'font-weight': _$item.css('font-weight'),
                                    'font-style': _$item.css('font-style'),
                                    'letter-spacing': _$item.css('letter-spacing'),
                                    width: tempWidth,
                                    height: 'auto',
                                    marginLeft: '5px',
                                    wordWrap: 'break-word',
                                    whiteSpace: wordWrapVal
                                })
                                .appendTo('body');

                            var _height = isNaN(parseInt($temp.css('line-height'))) ? $temp.height() : Math.max(parseInt($temp.css('line-height')), $temp.height());
                            var _width = $temp.width()+5;

                            var _index = prx.items.helper.getIndexFromId(item.id),
                                _dims = prx.toolbox.itemDimensions(prxy.items[_index]),
                                oldItem = {
                                    x: _dims.left,
                                    y: _dims.top,
                                    w: _dims.width,
                                    h: _dims.height
                                },
                                newItem = {
                                    x: _dims.left,
                                    y: _dims.top,
                                    w: _width,
                                    h: _height
                                };

                            if(isAutoResize) {
                                $item.width(_width)
                                    .attr('data-width',_width);
                                $iframeRichTextEL.width(_width).height(_height);
                                prxy.items[_index].width = _width;
                            }else {
                                $iframeRichTextEL.width(item.width).height(_height);
                                newItem.w = _dims.width;
                            }
                            let _align = prxy.items[_index].textAlign === 'center' ? 'n' : prxy.items[_index].textAlign === 'right' ? 'ne' : 'nw';
                            positionWithOffset = prx.items.helper.repositionOnResize(
                                oldItem,
                                newItem,
                                _dims.rotation,
                                _align);
                            prxy.items[_index].left = positionWithOffset.x;
                            prxy.items[_index].top = positionWithOffset.y;

                            $item.css({'left': positionWithOffset.x, 'top': positionWithOffset.y, 'height': _height});

                            item.wtype = "fixed";
                            item.htype = "fixed";
                            item.height = _height;
                            $temp.remove();
                            var el = '#' + item.id + ' #editable-properties-text';
                            var isRichTextEditorOpen = $(el).data('froala.editor')!==undefined ? true : false;
                            if(isRichTextEditorOpen==false) {
                                prx.items.update(item,false);
                            }
                            $('#p-'+item.id+'-lineHeight').val(item.lineHeight);
                            if(prx.editor && item.v2===undefined){
                                item.v2 = true;
                                return item;
                            }
                            prx.componentsHelper.memorisedTextStyle.update(item);
                        }
                        ,changeProperty: {
                            caption: 'Text size',
                            selector: '.liveUpdate-textColor-color',
                            property: 'font-size',
                            transitionable: true
                        }
                    }
		      	],
                [
                    {
                        caption: { label: 'Underline', class: 'colorpicker-wrapper text-underline-label' },
                        name: 'textProperties',
                        proptype: 'text-properties',
                        relatedEditableProperties: 'text',
                        relatedCSSProperties: ['font-weight','font-style','text-decoration'],
                        type: 'checkbox',
                        pffSettings: prx.commonproperties.pffSettingsTextProperties,
                        value: function (item, name) {
                            if (typeof(item.textProperties) == 'undefined') {
                                item.textProperties = [];
                            }
                            return item.textProperties;
                        },
                        values: [
                            { value: 'underline', displayValue: '', icon: 'text-underline'}
                        ],
                        onChange: function(item) {
                            var isAutoResize = item.autoResize == true ? true : false;
                            var wordWrapVal = isAutoResize == true ? 'nowrap' : 'normal';
                            var $item = $('#' + item.id);
                            var tempWidth = isAutoResize == true ? 'auto' : item.width;
                            var $iframeRichTextEL = $(prx.htmlTextEditor.iframeEl());
                            var _$item = $('#'+item.id+' [data-editableproperty="text"]');

                            var $temp = $('<span>'+item.text+ '&nbsp;</span>')
                                .css({
                                    display: 'inline-block',
                                    'font-size': _$item.css('font-size'),
                                    'font-family': _$item.css('font-family'),
                                    'line-height': _$item.css('line-height'),
                                    'text-decoration': _$item.css('text-decoration'),
                                    'font-weight': _$item.css('font-weight'),
                                    'font-style': _$item.css('font-style'),
                                    'letter-spacing': _$item.css('letter-spacing'),
                                    width: tempWidth,
                                    height: 'auto',
                                    marginLeft: '5px',
                                    wordWrap: 'break-word',
                                    whiteSpace: wordWrapVal
                                })
                                .appendTo('body');

                            var _height = isNaN(parseInt($temp.css('line-height'))) ? $temp.height() : Math.max(parseInt($temp.css('line-height')), $temp.height());
                            var _width = $temp.width()+5;

                            if(isAutoResize) {
                                $item.width(_width).height(_height);
                                $iframeRichTextEL.width(_width).height(_height);
                                prxy.items[prx.items.helper.getIndexFromId(item.id)].width = _width;

                            }else {
                                $item.height(_height);
                                $iframeRichTextEL.width(item.width).height(_height);
                            }
                            item.wtype = 'fixed';
                            item.htype = 'fixed';
                            item.height = _height;
                            $temp.remove();

                            var el = '#' + item.id + ' #editable-properties-text';
                            var isRichTextEditorOpen = $(el).data('froala.editor')!==undefined ? true : false;
                            if(isRichTextEditorOpen==false) {
                                prx.items.update(item,false);
                            }
                            if(prx.editor && item.v2===undefined){
                                item.v2 = true;
                                return item;
                            }
                        },
                        changeProperty: {
                            caption: 'Underline',
                            transitionable: false,
                            changeFunction: function(item, containerid, duration, easing, dynPropI){prx.componentsHelper.changeFunctionForTextProperties(item, containerid, dynPropI, 'textProperties');}
                        }
                    }
                    ,{
                        caption: {label: 'Color', class: 'text-properties-label text-color-label'},
                        name: 'textColor',
                        proptype: 'font-color',
                        type: 'solid-colorpicker',
                        pffSettings: prx.commonproperties.pffSettingsColor,
                        value: function (item, name) {
                            return typeof item.textColor == 'undefined' ? '' : item.textColor;
                        },
                        onChange: function(item) {
                            prx.componentsHelper.memorisedTextStyle.update(item);
                        },
                        liveUpdate: 'color',
                        relatedEditableProperties: 'text',
                        relatedCSSProperties: 'color',
                        changeProperty: {
                            caption: 'Text color',
                            selector: '.changeProperty-textColor',
                            property: 'color',
                            transitionable: true
                        }
                    }
				],
				[
                    {
                        caption: { label: 'Alignment', class: 'text-properties-label text-alignment-label' },
                        proptype: 'text-align',
                        name: 'textAlign',
                        type: 'radio',
                        pffSettings: prx.commonproperties.pffSettingsTextAlignment,
                        relatedEditableProperties: 'text',
                        relatedCSSProperties: 'text-align',
                        onChange: function(item) {
                            prx.componentsHelper.memorisedTextStyle.update(item);
                        },
                        value: function(item, name) {
                            return typeof item.textAlign == 'undefined' ? '' : item.textAlign;
                        },
                        values: [
                            { value: 'left', displayValue: '', icon: 'align-left'},
                            { value: 'center', displayValue: '', icon: 'align-center'},
                            { value: 'right', displayValue: '', icon: 'align-right'},
                            { value: 'justify', displayValue: '', icon: 'align-justify'}
                        ],
                        changeProperty: {
                            caption: 'Text align',
                            selector: '.changeProperty-textAlign',
                            property: 'text-align',
                            transitionable: false
                        }
                    }
                ],
                [
		  		{
  					caption: 'Line',
  					name: 'lineHeight',
  					proptype: 'line-height',
                        relatedEditableProperties: 'text',
                        relatedCSSProperties: 'line-height',
  					type: 'input-numeric',
				    pffSettings: prx.commonproperties.pffSettingsLineHeight,
  					value: function(item,name) {
  						if(typeof(item.lineHeight) == 'undefined') { item.lineHeight = parseInt(prx.componentsHelper.getProp(item.textSize,'num-text-size')*1.231); }
  						return item.lineHeight;
  					},
  					changeProperty: {
  						caption: 'Line height',
				        // it sets line-height: 100 instead of line-height: 100px which is different :(
  						//selector: '.changeProperty-lineHeight',
  						//property: 'line-height',
  						//transitionable: false,
				        rerender: true
  					 },
				    onChange: function(item) {
                            if(item.lineHeight==''){//auto
                                item.lineHeight = parseInt(prx.componentsHelper.getProp(item.textSize,'num-text-size')*1.231);
                                item.lineHeightAuto = Boolean(true);
                            }else{
                                item.lineHeightAuto = Boolean(false);
                            }
                            prx.componentsHelper.memorisedTextStyle.update(item);
                            var isAutoResize = item.autoResize == true ? true : false;
                            var wordWrapVal = isAutoResize == true ? 'nowrap' : 'normal';
                            var $item = $('#' + item.id);
                            var tempWidth = isAutoResize == true ? 'auto' : item.width;
                            var $iframeRichTextEL = $(prx.htmlTextEditor.iframeEl());
                            var _$item = $('#'+item.id+' [data-editableproperty="text"]');

                            var $temp = $('<span>'+item.text+ '&nbsp;</span>')
                                .css({
                                    display: 'inline-block',
                                    'font-size': _$item.css('font-size'),
                                    'font-family': _$item.css('font-family'),
                                    'line-height': item.lineHeight + 'px',
                                    'text-decoration': _$item.css('text-decoration'),
                                    'font-weight': _$item.css('font-weight'),
                                    'font-style': _$item.css('font-style'),
                                    'letter-spacing': _$item.css('letter-spacing'),
                                    width: tempWidth,
                                    height: 'auto',
                                    marginLeft: '5px',
                                    wordWrap: 'break-word',
                                    whiteSpace: wordWrapVal
                                })
                                .appendTo('body');

                            var _height = isNaN(parseInt($temp.css('line-height'))) ? $temp.height() : Math.max(parseInt($temp.css('line-height')), $temp.height());
                            var _width = $temp.width()+5;

                        var _index = prx.items.helper.getIndexFromId(item.id),
                            _dims = prx.toolbox.itemDimensions(prxy.items[_index]),
                            oldItem = {
                                x: _dims.left,
                                y: _dims.top,
                                w: _dims.width,
                                h: _dims.height
                            },
                            newItem = {
                                x: _dims.left,
                                y: _dims.top,
                                w: _width,
                                h: _height
                            };

                        if(isAutoResize) {
                            $item.width(_width);
                            $item.attr('data-width',_width);
                                $iframeRichTextEL.width(_width).height(_height);
                            prxy.items[_index].width = _width;
                        }else {
                                $iframeRichTextEL.width(item.width).height(_height);
                            newItem.w = _dims.width
                        }
                        let _align = prxy.items[_index].textAlign === 'center' ? 'n' : prxy.items[_index].textAlign === 'right' ? 'ne' : 'nw';
                        positionWithOffset = prx.items.helper.repositionOnResize(
                            oldItem,
                            newItem,
                            _dims.rotation,
                            _align);
                        prxy.items[_index].left = positionWithOffset.x;
                        prxy.items[_index].top = positionWithOffset.y;

                        $item.css({'left': positionWithOffset.x, 'top': positionWithOffset.y, 'height': _height});

                        item.wtype = "fixed";
						item.htype = "fixed";
                        item.height = _height;
                        $temp.remove();
                        var el = '#' + item.id + ' #editable-properties-text';
                        var isRichTextEditorOpen = $(el).data('froala.editor')!==undefined ? true : false;
                        if(isRichTextEditorOpen==false) {
                            prx.items.update(item,false);
                        }
                        if(prx.editor && item.v2===undefined){
                            item.v2 = true;
                            return item;
                        }
				    }
  				},
                    {
                        caption: 'Character',
                        name: 'textSpacing',
                        proptype: 'letter-spacing',
                        type: 'input-numeric',
                        relatedEditableProperties: 'text',
                        relatedCSSProperties: 'letter-spacing',
                        pffSettings: prx.commonproperties.pffSettingsCharacterSpacing,
                        value: function (item, name) {
                            if (typeof(item.textSpacing) == 'undefined') {
                                item.textSpacing = 0;
                            }
                            return item.textSpacing;
                        },
                        changeProperty: {
                            caption: 'Character Spacing',
                            selector: '.changeProperty-textSpacing',
                            property: 'letter-spacing',
                            transitionable: true
                        },
                        onChange: function(item) {
                            if(item.textSpacing==''){
                                item.textSpacing = 0;
                            }
                            var isAutoResize = item.autoResize == true ? true : false;
                            var wordWrapVal = isAutoResize == true ? 'nowrap' : 'normal';
                            var $item = $('#' + item.id);
                            var tempWidth = isAutoResize == true ? 'auto' : item.width;
                            var $iframeRichTextEL = $(prx.htmlTextEditor.iframeEl());
                            var _$item = $('#'+item.id+' [data-editableproperty="text"]');

                            var $temp = $('<span>'+item.text+ '&nbsp;</span>')
                                .css({
                                    display: 'inline-block',
                                    'font-size': _$item.css('font-size'),
                                    'font-family': _$item.css('font-family'),
                                    'line-height': item.lineHeight + 'px',
                                    'text-decoration': _$item.css('text-decoration'),
                                    'font-weight': _$item.css('font-weight'),
                                    'font-style': _$item.css('font-style'),
                                    'letter-spacing': item.textSpacing + 'px',
                                    width: tempWidth,
                                    height: 'auto',
                                    marginLeft: '5px',
                                    wordWrap: 'break-word',
                                    whiteSpace: wordWrapVal
                                })
                                .appendTo('body');

                            var _height = isNaN(parseInt($temp.css('line-height'))) ? $temp.height() : Math.max(parseInt($temp.css('line-height')), $temp.height());
                            var _width = $temp.width()+5;

                            var _index = prx.items.helper.getIndexFromId(item.id),
                                _dims = prx.toolbox.itemDimensions(prxy.items[_index]),
                                oldItem = {
                                    x: _dims.left,
                                    y: _dims.top,
                                    w: _dims.width,
                                    h: _dims.height
                                },
                                newItem = {
                                    x: _dims.left,
                                    y: _dims.top,
                                    w: _width,
                                    h: _height
                                };

                            if(isAutoResize) {
                                $item.width(_width)
                                    .attr('data-width',_width);
                                $iframeRichTextEL.width(_width).height(_height);
                                prxy.items[_index].width = _width;
                            }else {
                                $iframeRichTextEL.width(item.width).height(_height);
                                newItem.w = _dims.width;
                            }
                            let _align = prxy.items[_index].textAlign === 'center' ? 'n' : prxy.items[_index].textAlign === 'right' ? 'ne' : 'nw';
                            positionWithOffset = prx.items.helper.repositionOnResize(
                                oldItem,
                                newItem,
                                _dims.rotation,
                                _align);
                            prxy.items[_index].left = positionWithOffset.x;
                            prxy.items[_index].top = positionWithOffset.y;

                            $item.css({'left': positionWithOffset.x, 'top': positionWithOffset.y, 'height': _height});
                            prx.componentsHelper.memorisedTextStyle.update(item);
                            item.wtype = "fixed";
							item.htype = "fixed";
                            item.height = _height;
                            $temp.remove();
                            var el = '#' + item.id + ' #editable-properties-text';
                            var isRichTextEditorOpen = $(el).data('froala.editor')!==undefined ? true : false;
                            if(isRichTextEditorOpen==false) {
                                prx.items.update(item,false);
                            }
                            if(prx.editor && item.v2===undefined){
                                item.v2 = true;
                                return item;
                            }
                        }
                    }
		  		],
		  		[
                    {
		      		caption: 'Text Shadow',
		      		name: 'enableShadow',
		      		type: 'onoff',
		      		value: function(item,name) {
		      			return item.enableShadow;
		      		},
		      		changeProperty: {
  						caption: 'Text Shadow',
  						rerender: true
	      			},
                        onChange: function(item) {
                            if(prx.editor && item.v2===undefined){
                                item.v2 = true;
                                return item;
                            }
                        }
                    }
                ],
                [
			      	{
                        caption: 'Use lorem ipsum text',
                        name: 'loremIpsum',
                        type: 'link',
                        onClick: function(item) {
                            if(item.text.indexOf(item.caption.replace('...', '')) == 0 || /^Text(\s[0-9]*)?$/.test(item.caption)) {
                                item.caption = 'Lorem ipsum dolor sit amet...';
                                $('#itembrowser #id-' + item.id + ' .item-caption').text(item.caption);
                            }
                            item.text = 'Lorem ipsum dolor sit amet, consectetur adipiscing elit. Fusce convallis pellentesque metus id lacinia. Nunc dapibus pulvinar auctor. Duis nec sem at orci commodo viverra id in ipsum. Fusce tellus nisl, vestibulum sed rhoncus at, pretium non libero. Cras vel lacus ut ipsum vehicula aliquam at quis urna. Nunc ac ornare ante. Fusce lobortis neque in diam vulputate quis semper sem elementum.';
                            var isAutoResize = item.autoResize == true ? true : false;
                            var $item = $('#' + item.id);
                            var canvasWidth =  prx.stc.screens.check() ? Math.round(prx.devices[prx.device][prxy.pages[prxy.iSelectedPage].orientation][0]) : Math.round(prxy.symbols[prxy.iSelectedSymbol].states[prxy.iSelectedState].dimensions[0]);
                            var canvasHeight =  prx.stc.screens.check() ? Math.round(prx.devices[prx.device][prxy.pages[prxy.iSelectedPage].orientation][1]) : Math.round(prxy.symbols[prxy.iSelectedSymbol].states[prxy.iSelectedState].dimensions[1]);
                            var wordWrapVal = isAutoResize == true ? 'nowrap' : 'normal';
                            var convertToFixed = false;
                            var tempWidth = isAutoResize == true ? 'auto' : item.width;
                            var $iframeRichTextEL = $(prx.htmlTextEditor.iframeEl());
                            var _$item = $('#'+item.id+' [data-editableproperty="text"]');

                            var $temp = $('<span>'+item.text+ '&nbsp;</span>')
                                .css({
                                    display: 'inline-block',
                                    'font-size': _$item.css('font-size'),
                                    'font-family': _$item.css('font-family'),
                                    'line-height': _$item.css('line-height'),
                                    'text-decoration': _$item.css('text-decoration'),
                                    'font-weight': _$item.css('font-weight'),
                                    'font-style': _$item.css('font-style'),
                                    'letter-spacing': _$item.css('letter-spacing'),
                                    width: tempWidth,
                                    height: 'auto',
                                    marginLeft: '5px',
                                    wordWrap: 'break-word',
                                    whiteSpace: wordWrapVal,
                                    display: 'block'
                                })
                                .appendTo('body');


                            var _width = $temp.width()+5;


                            if(_width > canvasWidth && isAutoResize){ //in case autoresize true but using lorem ipsum is going to create a very wide textbox, then convert to fixed
                                convertToFixed = true;
                                var isOnCanvas = (item.left >= 0 && item.left < canvasWidth) && (item.top >= 0 && item.top < canvasHeight) ? true : false;
                                _width = !isOnCanvas ? canvasWidth : canvasWidth - item.left;

                                $temp.css({
                                	width: _width,
                                    whiteSpace: 'normal'
                                });


                            }

                            var _height = isNaN(parseInt($temp.css('line-height'))) ? $temp.height() : Math.max(parseInt($temp.css('line-height')), $temp.height());

                            var _index = prx.items.helper.getIndexFromId(item.id),
                                _dims = prx.toolbox.itemDimensions(prxy.items[_index]),
                                oldItem = {
                                    x: _dims.left,
                                    y: _dims.top,
                                    w: _dims.width,
                                    h: _dims.height
                                },
                                newItem = {
                                    x: _dims.left,
                                    y: _dims.top,
                                    w: _width,
                                    h: _height
                                };

                            if(isAutoResize && !convertToFixed) {
                                $item.width(_width)
                                    .attr('data-width',_width);
                                $iframeRichTextEL.width(_width).height(_height);
                                prxy.items[_index].width = _width;
                            }else {
                                if(convertToFixed){
                                    $item.width(_width);
                                    item.width = _width;
                                    prxy.items[_index].autoResize = false;
                                    $iframeRichTextEL.width(_width).height(_height);
                                }else{
                                    $iframeRichTextEL.width(item.width).height(_height);
                                    newItem.w = _dims.width;
                                }
                            }
                            let _align = prxy.items[_index].textAlign === 'center' ? 'n' : prxy.items[_index].textAlign === 'right' ? 'ne' : 'nw';
                            positionWithOffset = prx.items.helper.repositionOnResize(
                                oldItem,
                                newItem,
                                _dims.rotation,
                                _align);
                            prxy.items[_index].left = positionWithOffset.x;
                            prxy.items[_index].top = positionWithOffset.y;

                            $item.css({'left': positionWithOffset.x, 'top': positionWithOffset.y, 'height': _height});
                            item.wtype = 'fixed';
                            item.htype = 'fixed';
                            item.height = _height;
                            $temp.remove();
                            var el = '#' + item.id + ' #editable-properties-text';
                            var isRichTextEditorOpen = $(el).data('froala.editor')!==undefined ? true : false;
                            if(isRichTextEditorOpen==false) {
                                prx.items.update(item,false);

                            }else{
                                $(prx.htmlTextEditor.iframeEl()).contents().find('body').html(item.text);
                            }
                            if(prx.editor && item.v2===undefined){
                                item.v2 = true;
                            }

                            // gs - make sure froala is closed
                            if (typeof prx.htmlTextEditor.currentEditorInstance !== 'undefined') {
                                prx.htmlTextEditor.destroy();
                            }

                            return item;
                        }
                    }
                ]
            ]
        },
        {
            caption: 'Style',
            properties: [
                [
                    prx.commonproperties.backgroundColor
                ],
                [
                    {
                        caption: 'Width',
                        name: 'autoResize',
                        type: 'switcher',
                        value: function(item,name) {
                            if (typeof(item.autoResize) == 'undefined') {
                                item.autoResize = Boolean(false);
                            }
                            return item.autoResize;
                        },
                        values: [{
                            value: Boolean(true),
                            displayValue: 'Auto'
                        }, {
                            value: Boolean(false),
                            displayValue: 'Fixed'
                        }],
                        onChange: function(item) {
                            //oh boolean values
                            if(item.autoResize == 'false'){
                                item.autoResize = Boolean(false);
                            }else{
                                item.autoResize = Boolean(true);
                            }

                            var textAlign = 'left';

                            if(typeof(item.textAlign) !== 'undefined'){
                                textAlign = item.textAlign;
                            }

                            var oldWidth = item.width;
                            var isAutoResize = item.autoResize == true ? true : false;
                            var wordWrapVal = isAutoResize == true ? 'nowrap' : 'normal';
                            var $item = $('#' + item.id);
                            var tempWidth = isAutoResize == true ? 'auto' : item.width;
                            var $iframeRichTextEL = $(prx.htmlTextEditor.iframeEl());
                            var _$item = $('#'+item.id+' [data-editableproperty="text"]');

                            var $temp = $('<span>'+item.text+ '&nbsp;</span>')
                                .css({
                                    display: 'inline-block',
                                    'font-size': _$item.css('font-size'),
                                    'font-family': _$item.css('font-family'),
                                    'line-height': _$item.css('line-height'),
                                    'text-decoration': _$item.css('text-decoration'),
                                    'font-weight': _$item.css('font-weight'),
                                    'font-style': _$item.css('font-style'),
                                    'letter-spacing': _$item.css('letter-spacing'),
                                    width: tempWidth,
                                    height: 'auto',
                                    marginLeft: '5px',
                                    wordWrap: 'break-word',
                                    whiteSpace: wordWrapVal
                                })
                                .appendTo('body');

                            var _height = isNaN(parseInt($temp.css('line-height'))) ? $temp.height() : Math.max(parseInt($temp.css('line-height')), $temp.height());
                            var _width = $temp.width()+5;

                                var _index = prx.items.helper.getIndexFromId(item.id),
                                    _dims = prx.toolbox.itemDimensions(prxy.items[_index]),
                                    oldItem = {
                                        x: _dims.left,
                                        y: _dims.top,
                                        w: _dims.width,
                                        h: _dims.height
                                    },
                                    newItem = {
                                        x: _dims.left,
                                        y: _dims.top,
                                        w: _width,
                                        h: _height
                                    };

                            if(isAutoResize) {
                                    $item.width(_width)
                                        .attr('data-width',_width);
                                $iframeRichTextEL.width(_width).height(_height);
                                    prxy.items[_index].width = _width;
                            }else {
                                $item.height(_height);
                                $iframeRichTextEL.width(item.width).height(_height);
                                    newItem.w = _dims.width;
                            }
                            let _align = prxy.items[_index].textAlign === 'center' ? 'n' : prxy.items[_index].textAlign === 'right' ? 'ne' : 'nw';
                            positionWithOffset = prx.items.helper.repositionOnResize(
                                oldItem,
                                newItem,
                                _dims.rotation,
                                _align);
                            prxy.items[_index].left = positionWithOffset.x;
                            prxy.items[_index].top = positionWithOffset.y;

                            $item.css({'left': positionWithOffset.x, 'top': positionWithOffset.y, 'height': _height});
                            item.wtype = "fixed";
                            item.htype = "fixed";
                            item.height = _height;
                            $temp.remove();
                            var el = '#' + item.id + ' #editable-properties-text';
                            var isRichTextEditorOpen = $(el).data('froala.editor')!==undefined ? true : false;
                            if(isRichTextEditorOpen==false) {
                                prx.items.update(item,false);
                            }
                            if(prx.editor && item.v2===undefined){
                                item.v2 = true;
                                return item;
                            }
                        },
                        changeProperty: {
                            caption: 'Width',
                            selector: '.text-contents',
                            property: 'class',
                            transitionable: false
                        }
                    }
                ]
            ]
        },
        {
            caption: 'Advanced',
            properties: [
                [
                    {
                        caption: 'Is invisible to touch & mouse events'
                        ,help: 'Turning this property on will cause the item to become invisible to touch or mouse events and allow other items below to become interactive'
                        ,name: 'propagateEvents'
                        ,type: 'onoff'
                        ,value: function(item,name) {
                            if(typeof(item.propagateEvents) == 'undefined') {
                                return false;
                            }
                            return item.propagateEvents;
                        }
                        ,onChange: function(item){
                            if(prx.editor && item.v2===undefined){
                                item.v2 = true;
                            }
                            // will force rerender of properties to show/hide the interactions tab
                            return item;
                        }
                        ,changeProperty: {
                            caption: 'Invisible to touch/mouse interactions',
                            rerender: true
                        }
                    }
                ]
            ]
        }]

};

//TYPE: RICHTEXT
prx.types.richtext = {
    name: 'richtext'
    ,onDisplay: function(item,containerid,symbol) {
        var _id = (!containerid) ? item.id : containerid+'-'+item.id;
        if(typeof(item.propagateEvents) == 'undefined') { item.propagateEvents = false; }
        var cR = '';
        cR += '<div id="' + _id + '" ' + prx.items.getComponentBaseAttributes(item, containerid, symbol)  + ' class="' + prx.items.getComponentBaseClasses(item, containerid, symbol) + 'box pos type-richtext '+(prx.componentsHelper.getProp(item.propagateEvents,'boolean') ? 'pointer-events-none ' : '')+'">';
        cR += '<style>';
        cR += prx.items.getComponentBaseStyle(item, containerid, symbol);
        cR += '</style>';
        cR += prx.items.getComponentPrependDivs(item, containerid, symbol);
        cR += '<div style="overflow: hidden; width: 100%; height: 100%;">';
        cR += '<span>' + item.text + '</span>';
        cR += '</div>';
        cR += prx.items.getComponentAppendDivs(item, containerid, symbol);
        cR += '</div>';
        return cR;
    }
    ,interactions: [
        {
            caption: 'Interactions',
            name: 'actions',
            type: 'action',
            value: function(item,name) {
                if (typeof(item.actions) == 'undefined') {
                    item.actions = [];
                }
                return item.actions.length;
            },
            hiddenByDefault: function(item){
                if(typeof(item.propagateEvents) != 'undefined' && item.propagateEvents) {
                    return true;
                }
                return false;
            }
        }
    ]
    ,propertyGroups: [{
        caption: 'Text',
        properties: [
            [
                {
			  		caption: false
			    	,name: 'text'
			    	,type: 'wysiwyg'
			    	,value: function(item,name) {
			    		return item.text;
			    	}
			    	,changeProperty: {
			    		caption: 'Text',
			    		rerender: true
			    	}
                }
            ],
		        [
	  			{
                    caption: 'Use lorem ipsum text',
                    name: 'loremIpsum',
                    type: 'link',
                    onClick: function(item) {
                        item.text = 'Lorem ipsum dolor sit amet, consectetur adipiscing elit. Fusce convallis pellentesque metus id lacinia. Nunc dapibus pulvinar auctor. Duis nec sem at orci commodo viverra id in ipsum. Fusce tellus nisl, vestibulum sed rhoncus at, pretium non libero. Cras vel lacus ut ipsum vehicula aliquam at quis urna. Nunc ac ornare ante. Fusce lobortis neque in diam vulputate quis semper sem elementum.';
                        return item;
                    }
                }
            ]
        ]
    }, {
        caption: 'Advanced',
        properties: [
            [
                {
                    caption: 'Is invisible to touch & mouse events'
                    ,help: 'Turning this property on will cause the item to become invisible to touch or mouse events and allow other items below to become interactive'
                    ,name: 'propagateEvents'
                    ,type: 'onoff'
                    ,value: function(item,name) {
                        if(typeof(item.propagateEvents) == 'undefined') {
                            return false;
                        }
                        return item.propagateEvents;
                    }
                    ,onChange: function(item){
                        // will force rerender of properties to show/hide the interactions tab
                        return item;
                    }
                    ,changeProperty: {
                        caption: 'Invisible to touch/mouse events',
                        rerender: true
                    }
                }
            ]
        ]
    }

    ]
};

//TYPE: RECTANGLE
prx.types.rectangle = {
    name: 'rectangle'
    ,onDisplay: function(item,containerid,symbol) {

        if(item.typeName == 'oval') {
            item.borderRadius = '50%';
        }

        var _id = (!containerid) ? item.id : containerid+'-'+item.id;

        if(typeof(item.borderStyle) == 'undefined') { item.borderStyle = 'solid'; }
        if(typeof(item.text) == 'undefined') { item.text = ''; }
        if(typeof(item.textProperties) == 'undefined') { item.textProperties = []; }
        if(typeof(item.lineHeight) == 'undefined') { item.lineHeight = parseInt(prx.componentsHelper.getProp(item.textSize,'num-text-size')*1.231); }
        if(typeof(item.lineHeightAuto) == 'undefined') { item.lineHeightAuto = Boolean(true); }
        if(typeof(item.textSpacing) == 'undefined') { item.textSpacing = 0; }

        var _props = (typeof(prx.richtext)=='undefined') ? prx.componentsHelper.getProp(item.textProperties,'props-text') : '';

        var cR = '';
        cR += '<div id="' + _id + '" class="box pos type-rectangle '+ prx.items.getComponentBaseClasses(item, containerid, symbol) +'" data-shape-type="'+item.typeName+'" ' + prx.items.getComponentBaseAttributes(item, containerid, symbol)  + '>';
        cR += '<style>';
        cR += prx.items.getComponentBaseStyle(item, containerid, symbol);
        cR += '#'+_id+' .inner-rec { ' +prx.gradients.getBgCssByProperty(item.backgroundColor,'color-background')+'; border-width: '+prx.componentsHelper.getProp(item.borderWidth,'border-width')+'; border-style: '+prx.componentsHelper.getProp(item.borderStyle,'border-type')+'; border-color: ' + prx.componentsHelper.getProp(item.borderColor,'color-border') + '; border-radius: '+prx.componentsHelper.getProp(item.borderRadius,'border-radius')+'; } ';
        cR += '#' + _id + ' .shapes-text-container { ' + _props + ' ' + prx.componentsHelper.getProp(item.textFont + '|' + item.textFontStyle,'font-family') + prx.componentsHelper.getProp(item.textFontStyle,'font-style') + '; font-size: ' + prx.componentsHelper.getProp(item.textSize, 'num-text-size') + 'px; text-align: ' + prx.componentsHelper.getProp(item.textAlign, 'align') + '; line-height: ' + prx.componentsHelper.getProp(item.lineHeight, 'num-text-size') + 'px; letter-spacing: ' + prx.componentsHelper.getProp(item.textSpacing, 'num-text-size') + 'px; color: ' + prx.componentsHelper.getProp(item.textColor, 'color-text') + '; position: absolute; } ';
        cR += '#' + _id + ' .shapes-image-background { ' + ((prx.componentsHelper.getProp(item.imgSrc,'asset') !== '') ?('background-image: url(' + prx.componentsHelper.getProp(item.imgSrc,'asset')+'); background-size: ' + ((prx.componentsHelper.getProp(item.backgroundSizeType,'text') == 'cover') ? (prx.componentsHelper.getProp(item.backgroundSizeType,'text') + ';') : (prx.componentsHelper.getProp(item.backgroundSizeValue,'num-percentage') + '% auto;'))) : '') + ' border-radius: '+prx.componentsHelper.getProp(item.borderRadius,'border-radius')+'; }';

        //cR += '#' + _id + ' .rectangle-text-container span{display:block;}';
        cR += '</style>';
        cR += prx.items.getComponentPrependDivs(item, containerid, symbol);
        cR += '<div id="rec-' + _id + '" class="inner-rec liveUpdate-backgroundColor-background-color changeProperty-borderRadius changeProperty-backgroundColor changeProperty-backgroundColor-background-color changeProperty-borderColor changeProperty-borderWidth changeProperty-borderColor-border-color liveUpdate-borderColor-border-color">';
        cR += '<div class="shapes-image-background changeProperty-borderRadius"></div>';
        cR += '<div class="shapes-text-container liveUpdate-textColor-color changeProperty-textFontStyle">';
        cR += '<span data-editableproperty="text">' + prx.componentsHelper.getProp(item.text,'text-textarea') + '</span>';
        cR += '<span class="fake-text hidden" style="position:absolute;">&nbsp;&nbsp;</span>'; //This element was added to resolve empty rectangle visibility issues when filter is applied on chromium based browsers.
        cR += '</div>';
        cR += '</div>';
        cR += prx.items.getComponentAppendDivs(item, containerid, symbol);
        cR += '</div>';
        return cR;
    }
    ,interactions: [
        prx.commonproperties.actions
    ]
    ,editableProperties: [
        {
	    	caption: 'Text'
	    	,name: 'text'
	    	,type: 'textarea'
	    	,value: function(item,name) {
	    		if(typeof(item.text) == 'undefined') { item.text = ''; }
	    		return item.text;
	    	},
      		changeProperty: {
                caption: 'Text',
                property: 'text',
                selector: '.shapes-text-container span',
                transitionable: false
  			}
	    }
    ]
    ,propertyGroups: [
        {
            caption: 'Style',
            properties: [
	      			[
	      				{
	      					caption: 'Background',
	      					name: 'backgroundColor',
	      					proptype: 'background-color',
	      					type: 'gradients-colorpicker',
	      					value: function(item,name) { return item.backgroundColor; },
	      					liveUpdate: 'background-color',
	      					changeProperty: {
	      						caption: 'Background Color',
	      						selector: '.inner-rec',
	      						property: 'background-color',
	      						transitionable: true
	      					}
	      				}
	      			]
	      			,
	      			[
                        {
                            caption: 'Border',
                            name: 'borderWidth',
                            proptype: 'border-width',
                            type: 'combo-select',
                            value: function(item,name) { return item.borderWidth; },
                            values: { min: 0, max: 20, step: 1 } ,
                            expandableType: 'borderWidth',
                            expandedValues: ['t', 'r', 'l', 'b'],
                            changeProperty: {
                                caption: 'Border Width',
                                selector: '.changeProperty-borderWidth',
                                property: 'border-width',
                                transitionable: true
                            }
                        }
	      				,{
		      				caption: false,
		      				name: 'borderStyle',
		      				proptype: 'border-style',
		      				type: 'select',
		      				value: function(item,name) {
		      					if(typeof(item.borderStyle) == 'undefined') { item.borderStyle = 'solid'; }
		      					return item.borderStyle;
		      				},
		      				values: [{ value: 'solid', displayValue: 'Solid'},{ value: 'dotted', displayValue: 'Dotted'},{ value: 'dashed', displayValue: 'Dashed'},{ value: 'double', displayValue: 'Double'},{ value: 'none', displayValue: 'None'}],
	      					changeProperty: {
	      						caption: 'Border Style',
	      						selector: '.inner-rec',
	      						property: 'border-style',
	      						transitionable: false
	      					}
		      			}
	      				,
	      				{
	      					caption: false,
	      					name: 'borderColor',
	      					proptype: 'border-color',
	      					type: 'solid-colorpicker',
	      					value: function(item,name) { return item.borderColor; },
	      					liveUpdate: 'border-color' ,
	      					changeProperty: {
	      						caption: 'Border Color',
	      						selector: '.inner-rec',
	      						property: 'border-top-color,border-bottom-color,border-left-color,border-right-color',
	      						transitionable: true
	      					}
	      				}
	      			],[{
		      				caption: '<span class="icon icon-border-radius" title="Border radius"></span>',
		      				name: 'borderRadius',
		      				proptype: 'border-radius',
		      				type: 'combo-select',
		      				value: function(item,name) {
		      					return item.borderRadius;
		      				},
		      				values: {min: 0, max: 20, step: 1},
                            expandableType: 'borderRadius',
                            expandedValues: ['tl', 'tr', 'bl', 'br'],
	      					changeProperty: {
	      						caption: 'Border Radius',
	      						selector: '.changeProperty-borderRadius',
	      						property: 'border-radius',
	      						transitionable: true
	      					}
		      			}
		      		]

	      	]
	    },
        {
            caption: 'Text',
            properties: [
                [
                    {
                        caption: { label: 'Font family', class: 'text-properties-label text-fontfamily-label' },
                        name: 'textFont',
                        proptype: 'font-family',
                        type: 'select',
                        relatedEditableProperties: 'text',
                        relatedCSSProperties: 'font-family',
                        value: function(item,name) {
                            if(typeof(item.textFont) == 'undefined') { item.textFont = 'Arial,sans-serif'; }
                            return item.textFont;
                        },
                        values: function(){ return prx.comps.fonts; }
	      		,changeProperty: {
                            caption: ' Text font',
                            selector: '.shapes-text-container',
                            property: 'font-family',
                            transitionable: false
				 }

                    }
                ],
                [
                    prx.commonproperties.textFontStyleRichText(['font-weight', 'font-style'],'text', false, {propName: 'textFontStyle', caption: 'Font style', useAsDynProp: false}),
                    {
                        caption: false,
                        name: 'textSize',
                        proptype: 'font-size',
                        type: 'combo-select',
                        relatedEditableProperties: 'text',
                        relatedCSSProperties: 'font-size',
                        value: function(item,name) {
                            if(typeof(item.textSize) == 'undefined') { item.textSize = 16; }
                            return item.textSize;
                        },
                        values: prx.comps.textsize
                        ,changeProperty: {
                            caption: ' Text size',
                            selector: '.shapes-text-container',
                            property: 'font-size',
                            transitionable: false
                        },
                        onChange: function(item) {
                            if(item.lineHeightAuto==true) {
                                item.lineHeight = parseInt(prx.componentsHelper.getProp(item.textSize,'num-text-size')*1.231);
                            }
                        }
                    }
	      	]
                ,
                [
                    prx.commonproperties.textPropertiesRichText(['font-weight','font-style','text-decoration'],'text', false, {propName: 'textProperties', caption: 'Underline', useAsDynProp: false}),
                    {
                        caption: { label: 'Color', class: 'text-properties-label text-color-label' },
                        name: 'textColor',
                        proptype: 'font-color',
                        type: 'solid-colorpicker',
                        relatedEditableProperties: 'text',
                        relatedCSSProperties: 'color',
                        pffSettings: prx.commonproperties.pffSettingsColor,
                        value: function(item,name) {
                            if(typeof(item.textColor) == 'undefined') { item.textColor = '#2E2E2E'; }
                            return item.textColor;
                        },
                        liveUpdate: 'color'
                        ,changeProperty: {
                            caption: ' Text color',
                            selector: '.shapes-text-container',
                            property: 'color',
                            transitionable: true
                        }
                    }
                ],
                [
                    {
                        caption: { label: 'Alignment', class: 'text-properties-label text-alignment-label' },
	  			name: 'textAlign',
	  			proptype: 'text-align',
	  			type: 'radio',
                        relatedEditableProperties: 'text',
                        relatedCSSProperties: 'text-align',
                        pffSettings: prx.commonproperties.pffSettingsTextAlignment,
	  			value: function(item,name) {
	  				if(typeof(item.textAlign) == 'undefined') { item.textAlign = 'center'; }
	  				return item.textAlign;
	  			},
                        values: [
                            { value: 'left', displayValue: '', icon: 'align-left'},
                            { value: 'center', displayValue: '', icon: 'align-center'},
                            { value: 'right', displayValue: '', icon: 'align-right'},
                            { value: 'justify', displayValue: '', icon: 'align-justify'}
                        ],
	  			changeProperty: {
                            caption: 'Text Align',
                            selector: '.shapes-text-container',
                            property: 'text-align',
                            transitionable: false
      			}
	  		}
	  		]
                ,
                [
                    {
                        caption: 'Line',
                        name: 'lineHeight',
                        proptype: 'line-height',
                        relatedEditableProperties: 'text',
                        relatedCSSProperties: 'line-height',
                        type: 'input-numeric',
                        pffSettings: prx.commonproperties.pffSettingsLineHeight,
                        value: function(item,name) {
                            if(typeof(item.lineHeight) == 'undefined') { item.lineHeight = parseInt(prx.componentsHelper.getProp(item.textSize,'num-text-size')*1.231); }
                            return item.lineHeight;
                        }
                        ,changeProperty: {
                            caption: 'Line height',
                            // it sets line-height: 100 instead of line-height: 100px which is different :(
                            // selector: '.changeProperty-lineHeight',
                            // property: 'line-height',
                            // transitionable: false
                            rerender: true
                        },
                        onChange: function(item) {
                            if (item.lineHeight == '') {//auto
                                item.lineHeight = parseInt(prx.componentsHelper.getProp(item.textSize, 'num-text-size') * 1.231);
                                item.lineHeightAuto = Boolean(true);
                            } else {
                                item.lineHeightAuto = Boolean(false);
                            }
                        }
                    },
                    {
                        caption: 'Character',
                        name: 'textSpacing',
                        proptype: 'letter-spacing',
                        type: 'input-numeric',
                        relatedEditableProperties: 'text',
                        relatedCSSProperties: 'letter-spacing',
                        pffSettings: prx.commonproperties.pffSettingsCharacterSpacing,
                        value: function(item,name) {
                            if(typeof(item.textSpacing) == 'undefined') { item.textSpacing = 0; }
                            return item.textSpacing;
                        },
                        changeProperty: {
                            caption: 'Character Spacing',
                            selector: '.shapes-text-container',
                            property: 'letter-spacing',
                            transitionable: true
                        },
                    },
                ]
            ]
        },
        {
            caption: 'Image',
            properties: [
                [
                    {
                        caption: false
                        ,name: 'imgSrc'
                        ,type: 'combo-asset'
                        ,displayValue: function(item,name) {
                            if((item.imgSrc == undefined ) || (item.imgSrc.fileId == '')) {
                                return 'No asset selected.';
                            }
                            return prx.utils.escapeHTML(item.imgSrc.name);
                        }
                        ,value: function(item,name) {
                            return JSON.stringify({
                                allow: 'image',
                                asset: (item.imgSrc !== undefined) ? item.imgSrc : {fileId: '', name: '', assetType: ''}
                            });
                        },
                        onChange: function(item) {
                            if (item.imgSrc.name.toLowerCase().endsWith('.svg')) {
                                item.backgroundSizeType = 'percentage';
                            } else {
                                item.backgroundSizeType = 'cover';
                            }

                            return item;
                        }
                        ,changeProperty: {
                            caption: 'Image',
                            rerender: true
                        }

                    }
                ],
                [
                    {
                        caption: 'Size',
                        name: 'backgroundSizeType',
                        proptype: 'background-size-type',
                        type: 'select',
                        value: function(item,name) {
                            if(typeof(item.backgroundSizeType) == 'undefined') { item.backgroundSizeType = 'cover'; }
                            return item.backgroundSizeType;
                        },
                        hiddenByDefault: function(item){
                            return ((item.imgSrc == undefined) || (item.imgSrc.fileId === ''));
                        },
                        onChange: function(item) {
                            return item;
                        },
                        values: [{ value: 'cover', displayValue: 'Cover'}, { value: 'percentage', displayValue: 'Percentage' }],
                        changeProperty: {
                            caption: 'Background Size Type',
                            transitionable: false,
                            changeFunction: function (item, containerid, duration, easing, dynPropI) {
                                if(prx.componentsHelper.getProp(item.imgSrc,'asset') !== '') {
                                    if(item.backgroundSizeType === 'cover') {
                                        $('#rec-' + containerid + '-' + item.id + ' .shapes-image-background').css('background-size', 'cover');
                                    }
                                    else {
                                        $('#rec-' + containerid + '-' + item.id + ' .shapes-image-background').css('background-size', item.backgroundSizeValue + '% auto');
                                    }
                                }
                            },
                            changeable: false
                        }
                    },
                    {
                        caption: false,
                        name: 'backgroundSizeValue',
                        proptype: 'num-percentage',
                        type: 'slider-input',
                        value: function(item,name) {
                            if(typeof(item.backgroundSizeValue) == 'undefined') { item.backgroundSizeValue = 35; }
                            return item.backgroundSizeValue; },
                        hiddenByDefault: function(item){
                            if(item.backgroundSizeType === 'percentage') {
                                return ((item.imgSrc == undefined) || (item.imgSrc.fileId === ''));
                            }
                            return true;
                        },
                        values: { min: 0, max: 100, step: 1 } ,
                        changeProperty: {
                            caption: 'Background Size',
                            changeFunction: function (item, containerid, duration, easing, dynPropI) {
                                if(prx.componentsHelper.getProp(item.imgSrc,'asset') !== '') {
                                    if(item.backgroundSizeType === 'cover') {
                                        $('#rec-' + containerid + '-' + item.id + ' .shapes-image-background').css('background-size', 'cover');
                                    }
                                    else {
                                        $('#rec-' + containerid + '-' + item.id + ' .shapes-image-background').css('background-size', item.backgroundSizeValue + '% auto');
                                    }
                                }
                            },
                            transitionable: false
                        }
                    }
                ]
            ]
        }
    ]

};

prx.types.group = {
    name: 'group',
    onDisplay: function () {
        return '';
    }
};

//TYPE: multiselected
prx.types.multiselected = {
    name: 'multiselected'
    , onDisplay: function () {
        return '';
    }
    , editableProperties: [
        {
            caption: 'Text'
            , name: 'text'
            , type: 'textarea'
            , value: function (item, name) {
                if (typeof(item.text) == 'undefined') {
                    item.text = '';
                }
                return item.text;
            },
            changeProperty: {
                caption: 'Text',
                property: 'text',
                selector: '.shapes-text-container span',
                transitionable: false
            }
        }
    ]
    , propertyGroups: [
        {
            caption: 'Text',
            properties: [
                [
                    {
                        caption: false,
                        name: 'textFont',
                        proptype: 'font-family',
                        type: 'select',
                        value: function (item, name) {
                            if (typeof(item.textFont) == 'undefined') {
                                item.textFont = 'Arial,sans-serif';
                            }
                            return item.textFont;
                        },
                        values: function () {
                            return prx.comps.fonts;
                        }
                        , changeProperty: {
                            caption: ' Text font',
                            selector: '.liveUpdate-textColor-color',
                            property: 'font-family',
                            transitionable: false
                        }

                    }
                ],
                [
                    prx.commonproperties.textFontStyleRichText(['font-weight', 'font-style'],'text', false, {propName: 'textFontStyle', caption: 'Font style', useAsDynProp: false}),
                    {
                        caption: false,
                        name: 'textSize',
                        proptype: 'font-size',
                        type: 'combo-select',
                        value: function (item, name) {
                            if (typeof(item.textSize) == 'undefined') {
                                item.textSize = 16;
                            }
                            return item.textSize;
                        },
                        values: prx.comps.textsize
                        ,changeProperty: {
                            caption: ' Text size',
                            selector: '.liveUpdate-textColor-color',
                            property: 'font-size',
                            transitionable: false
                        }
                    }
                ],
                [
                    {
                        caption: false,
                        name: 'textProperties',
                        proptype: 'text-properties',
                        type: 'checkbox',
                        pffSettings: prx.commonproperties.pffSettingsTextProperties,
                        value: function (item, name) {
                            if (typeof(item.textProperties) == 'undefined') {
                                item.textProperties = [];
                            }
                            return item.textProperties;
                        },
                        values: [
                            { value: 'underline', displayValue: '', icon: 'text-underline'}
                        ],
                        changeProperty: {
                            caption: 'Underline',
                            rerender: true
                        }
                    },
                    {
                        caption: false,
                        name: 'textColor',
                        proptype: 'font-color',
                        type: 'colorpicker',
                        pffSettings: prx.commonproperties.pffSettingsColor,
                        value: function (item, name) {
                            if (typeof(item.textColor) == 'undefined') {
                                item.textColor = '#2E2E2E';
                            }
                            return item.textColor;
                        },
                        liveUpdate: 'color'
                        , changeProperty: {
                            caption: ' Text color',
                            selector: '.liveUpdate-textColor-color',
                            property: 'color',
                            transitionable: true
                        }
                    }
                ]
                ,
                [


                    {
                        caption: false,
                        name: 'textAlign',
                        proptype: 'text-align',
                        type: 'radio',
                        value: function (item, name) {
                            if (typeof(item.textAlign) == 'undefined') {
                                item.textAlign = 'center';
                            }
                            return item.textAlign;
                        },
                        values: [
                            { value: 'left', displayValue: '', icon: 'align-left'},
                            { value: 'center', displayValue: '', icon: 'align-center'},
                            { value: 'right', displayValue: '', icon: 'align-right'}
                        ],
                        changeProperty: {
                            caption: 'Text Align',
                            selector: '.liveUpdate-textColor-color',
                            property: 'text-align',
                            transitionable: false
                        }
                    }
                ]
            ]
        },
        {
            caption: 'Style',
            properties: [
                [
                    {
                        caption: 'Background',
                        name: 'backgroundColor',
                        proptype: 'background-color',
                        type: 'colorpicker',
                        value: function (item, name) {
                            return item.backgroundColor;
                        },
                        liveUpdate: 'background-color',
                        changeProperty: {
                            caption: 'Background Color',
                            selector: '.inner-rec',
                            property: 'background-color',
                            transitionable: true
                        }
                    }
                ]
                ,
                [
                    {
                        caption: 'Border',
                        name: 'borderWidth',
                        proptype: 'border-width',
                        type: 'combo-select',
                        value: function (item, name) {
                            return item.borderWidth;
                        },
                        values: {min: 0, max: 20, step: 1},
                        changeProperty: {
                            caption: 'Border Width',
                            selector: '.inner-rec',
                            property: 'border-width',
                            transitionable: true
                        }
                    }
                    , {
                        caption: false,
                        name: 'borderStyle',
                        proptype: 'border-style',
                        type: 'select',
                        value: function (item, name) {
                            if (typeof(item.borderStyle) == 'undefined') {
                                item.borderStyle = 'solid';
                            }
                            return item.borderStyle;
                        },
                        values: [{value: 'solid', displayValue: 'Solid'}, {
                            value: 'dotted',
                            displayValue: 'Dotted'
                        }, {value: 'dashed', displayValue: 'Dashed'}, {
                            value: 'double',
                            displayValue: 'Double'
                        }, {value: 'none', displayValue: 'None'}],
                        changeProperty: {
                            caption: 'Border Style',
                            selector: '.inner-rec',
                            property: 'border-style',
                            transitionable: false
                        }
                    }
                    ,
                    {
                        caption: false,
                        name: 'borderColor',
                        proptype: 'border-color',
                        type: 'colorpicker',
                        value: function (item, name) {
                            return item.borderColor;
                        },
                        liveUpdate: 'border-color',
                        changeProperty: {
                            caption: 'Border Color',
                            selector: '.inner-rec',
                            property: 'border-top-color,border-bottom-color,border-left-color,border-right-color',
                            transitionable: true
                        }
                    }
                ], [{
                    caption: '<span class="icon icon-border-radius" title="Border radius"></span>',
                    name: 'borderRadius',
                    proptype: 'border-radius',
                    type: 'combo-select',
                    value: function (item, name) {
                        if (item.borderRadius == parseInt(item.borderRadius)) {
                            return item.borderRadius += 'px';
                        }
                        return item.borderRadius;
                    },
                    values: [{value: '0px', displayValue: '0px'}, {value: '1px', displayValue: '1px'}, {
                        value: '2px',
                        displayValue: '2px'
                    }, {value: '3px', displayValue: '3px'}, {value: '4px', displayValue: '4px'}, {
                        value: '5px',
                        displayValue: '5px'
                    }, {value: '6px', displayValue: '6px'}, {value: '7px', displayValue: '7px'}, {
                        value: '8px',
                        displayValue: '8px'
                    }, {value: '9px', displayValue: '9px'}, {value: '10px', displayValue: '10px'}, {
                        value: '11px',
                        displayValue: '11px'
                    }, {value: '12px', displayValue: '12px'}, {value: '13px', displayValue: '13px'}, {
                        value: '14px',
                        displayValue: '14px'
                    }, {value: '15px', displayValue: '15px'}, {value: '16px', displayValue: '16px'}, {
                        value: '17px',
                        displayValue: '17px'
                    }, {value: '18px', displayValue: '18px'}, {value: '19px', displayValue: '19px'}, {
                        value: '20px',
                        displayValue: '20px'
                    }],
                    changeProperty: {
                        caption: 'Border Radius',
                        selector: '.inner-rec',
                        property: 'border-radius',
                        transitionable: true
                    }
                }
                ]

            ]
        }
    ]
};

/* TYPE = CIRCLE */
prx.types.circle = prx.componentsHelper.cloneobject(prx.types.rectangle);
prx.types.circle.name = 'circle';
prx.types.circle.propertyGroups = prx.componentsHelper.removeProperties(prx.types.circle.propertyGroups, ['borderRadius']);
prx.types.circle.propertyGroups = prx.componentsHelper.removePropertySetting(prx.types.circle.propertyGroups, 'borderWidth', 'expandableType');
prx.types.circle.propertyGroups = prx.componentsHelper.removePropertySetting(prx.types.circle.propertyGroups, 'borderWidth', 'expandedValues');


//TYPE: HORIZONTAL LINE
prx.types.horizontalline = {
    name: 'horizontalline'
    ,onDisplay: function(item,containerid,symbol) {
        var _id = (!containerid) ? item.id : containerid+'-'+item.id;
        item.height = item.weight;
        if(typeof(item.borderStyle) == 'undefined') { item.borderStyle = 'solid'; }
        if (prx.componentsHelper.getProp(item.locked,'boolean')) {var xtra='onmousedown="if (event.preventDefault) event.preventDefault()"';} else {var xtra='';} // prevent default drag
        var cR = '<div id="' + _id + '" ' + prx.items.getComponentBaseAttributes(item, containerid, symbol)  + ' class="' + prx.items.getComponentBaseClasses(item, containerid, symbol) + ' box pos type-horizontalline" style="height: '+prx.componentsHelper.getProp(item.weight,'num-other')+'px; overflow: visible">';
        cR += '<style>';
        cR += prx.items.getComponentBaseStyle(item, containerid, symbol);
        cR += '</style>';
        cR += prx.items.getComponentPrependDivs(item, containerid, symbol);
        cR += '<div class="inner liveUpdate-color-border-color" style="border-top: '+prx.componentsHelper.getProp(item.weight,'num-border-width')+'px '+prx.componentsHelper.getProp(item.borderStyle,'border-type')+' '+prx.componentsHelper.getProp(item.color,'color-border')+';">';
        cR += '</div>';
        cR += prx.items.getComponentAppendDivs(item, containerid, symbol);
        cR += '</div>';
        return cR;
    }
    ,propertyGroups: [
        {
            caption: 'Style',
            properties: [
                [
	      			{
            			caption: 'Border'
            			,name: 'weight'
            			,proptype: 'border-width'
            			,type: 'combo-select'
            			,value: function(item,name) {
            				return item.weight;
            			}
    					,values: { min: 1, max: 10, step: 1 }
    					,changeProperty: {
                            caption: 'Thickness',
                            selector: '.inner',
                            property: 'border-width',
                            transitionable: true
						 }
	      			},{
	      				caption: false,
	      				name: 'borderStyle',
	      				proptype: 'border-style',
	      				type: 'select',
	      				value: function(item,name) {
	      					if(typeof(item.borderStyle) == 'undefined') { item.borderStyle = 'solid'; }
	      					return item.borderStyle;
	      				},
	      				values: [{ value: 'solid', displayValue: 'Solid'},{ value: 'dotted', displayValue: 'Dotted'},{ value: 'dashed', displayValue: 'Dashed'},{ value: 'double', displayValue: 'Double'},{ value: 'none', displayValue: 'None'}]
	      				,changeProperty: {
                            caption: 'Style',
                            rerender: true
						 }
		      		},
	      			{
	      				caption: false
	      				,name: 'color'
	      				,proptype: 'border-color'
	      				,type: 'colorpicker'
	      				,value: function(item,name) {
	      					return item.color;
	      				}
	      				,liveUpdate: 'border-color'
	      				,changeProperty: {
                            caption: 'color',
                            selector: '.inner',
                            property: 'border-color',
                            transitionable: true
                        }
	      			}
	      		]
            ]
        }
    ]

};

//TYPE: VERTICAL LINE
prx.types.verticalline = {
    name: 'verticalline'
    ,onDisplay: function(item,containerid,symbol) {
        var _id = (!containerid) ? item.id : containerid+'-'+item.id;
        item.width = item.weight;
        if(typeof(item.borderStyle) == 'undefined') { item.borderStyle = 'solid'; }
        if (prx.componentsHelper.getProp(item.locked,'boolean')) {var xtra='onmousedown="if (event.preventDefault) event.preventDefault()"';} else {var xtra='';} // prevent default drag
        var cR = '<div id="' + _id + '" ' + prx.items.getComponentBaseAttributes(item, containerid, symbol)  + ' class="' + prx.items.getComponentBaseClasses(item, containerid, symbol) + ' box pos type-verticalline" style="width: '+prx.componentsHelper.getProp(item.weight,'num-other')+'px;">';
        cR += '<style>';
        cR += prx.items.getComponentBaseStyle(item, containerid, symbol);
        cR += '</style>';
        cR += prx.items.getComponentPrependDivs(item, containerid, symbol);
        cR += '<div class="inner liveUpdate-color-border-color" style="border-left: '+prx.componentsHelper.getProp(item.weight,'num-border-width')+'px '+prx.componentsHelper.getProp(item.borderStyle,'border-type')+' '+prx.componentsHelper.getProp(item.color,'color-border')+';">';
        cR += '</div>';
        cR += prx.items.getComponentAppendDivs(item, containerid, symbol);
        cR += '</div>';
        return cR;
    }
    ,propertyGroups: [
        {
            caption: 'Style',
            properties: [
                [
	      			{
            			caption: 'Border'
            			,name: 'weight'
            			,proptype: 'border-width'
            			,type: 'combo-select'
            			,value: function(item,name) {
            				return item.weight;
            			}
    					,values: { min: 1, max: 10, step: 1 }
    					,changeProperty: {
                            caption: 'Thickness',
                            selector: '.inner',
                            property: 'border-width',
                            transitionable: true
                        }
	      			}
	      			,{
	      				caption: false,
	      				name: 'borderStyle',
	      				proptype: 'border-style',
	      				type: 'select',
	      				value: function(item,name) {
	      					if(typeof(item.borderStyle) == 'undefined') { item.borderStyle = 'solid'; }
	      					return item.borderStyle;
	      				},
	      				values: [{ value: 'solid', displayValue: 'Solid'},{ value: 'dotted', displayValue: 'Dotted'},{ value: 'dashed', displayValue: 'Dashed'},{ value: 'double', displayValue: 'Double'},{ value: 'none', displayValue: 'None'}]
	      				,changeProperty: {
                            caption: 'Style',
                            rerender: true
                        }
		      		},
	      			{
	      				caption: false
	      				,name: 'color'
	      				,proptype: 'border-color'
	      				,type: 'colorpicker'
	      				,value: function(item,name) {
	      					return item.color;
	      				}
	      				,liveUpdate: 'border-color'
	      				,changeProperty: {
                            caption: 'Color',
                            selector: '.inner',
                            property: 'border-color',
                            transitionable: true
						 }
	      			}

	      		]
	      		]
	      	}
	      ]

};

//TYPE: ACTION AREA
prx.types.actionarea = {
    name: 'actionarea'
    ,onDisplay: function(item,containerid,symbol) {
        var _id = (!containerid) ? item.id : containerid+'-'+item.id;
        let assistiveDescripionAttr = item.assistiveDescription ? 'role="button" aria-label="' + prx.utils.escapeHTML(item.assistiveDescription) + '"' : '';

        var cR = '<div id="' + _id + '" ' + prx.items.getComponentBaseAttributes(item, containerid, symbol)  + ' class="' + prx.items.getComponentBaseClasses(item, containerid, symbol) + ' box pos type-actionarea" ' + assistiveDescripionAttr + '>';
        cR += '<style>';
        cR += prx.items.getComponentBaseStyle(item, containerid, symbol);
        cR += '</style>';
        cR += prx.items.getComponentPrependDivs(item, containerid, symbol);
        cR += '<div id="rec-' + _id + '" class="inner-rec" >';
        cR += '<div></div>';
        cR += '</div>';
        cR += prx.items.getComponentAppendDivs(item, containerid, symbol);
        cR += '</div>';
        return cR;
    }
    ,propertyGroups: [
        {
            caption: 'Accessibility',
            properties: [
                [
                    {
                        caption: 'Descriptive text'
                        , hint: 'For assistive devices and screen readers'
                        , name: 'assistiveDescription'
                        , type: 'input'
                        , value: function (item, name) {
                            return item.assistiveDescription ? item.assistiveDescription : '';
                        }
                        , onChange: function (item) {
                            if (item.assistiveDescription !== '') {
                                document.getElementById(item.id).setAttribute('aria-label', item.assistiveDescription);
                            }
                        },
                        changeProperty: {
                            caption: 'Description for assistive devices',
                            transitionable: false,
                            changeFunction: function(item, containerid) {
                                let _id = (!containerid) ? item.id : containerid+'-'+item.id;
                                document.getElementById(_id).setAttribute('aria-label', item.assistiveDescription);
                            }
                        }
                    }
                ]
            ]
        }
    ]
    ,interactions: [
	  prx.commonproperties.actions
    ]

};

//TYPE: IMAGE

prx.imageChecked =  {};
prx.types.image = {
    name: 'image'
    , onDisplay: function (item, containerid, symbol) {
        var _id = (!containerid) ? item.id : containerid + '-' + item.id;
        if (typeof(item.overlay) == 'undefined') {
            item.overlay = false;
        }
        if (typeof(item.repeat) == 'undefined') {
            item.repeat = false;
        }

        var assetUrl = prx.componentsHelper.getProp(item.imgSrc,'asset');
        var thumbUrl = '';

        if (prx.urlCache != undefined)
            thumbUrl = prx.urlCache.thumbs[prx.componentsHelper.getProp(item.imgSrc.fileId,'other')] || '';

        var missing = prx.url.static + '/images/editor/missing.png';

        if (prx.editor) {

            if( $('#image-load-container').length == 0 )
                $('<div id="image-load-container"></div>').appendTo('body');

            var container = document.getElementById('image-load-container');

            var img = document.createElement('img');

            container.appendChild(img);

            img.onload = function (e) {

                container.removeChild(img);

                $('#' + _id).css('background-image', '');
                $('#' + _id + ' .mask-inner').css('background-image', '');

                delete prx.imageChecked[toCheck];

                $('#' + _id + '-img-wrapper').removeClass('missing');
                $('#' + _id + '-img-wrapper').find('div').remove();
                //$('#' + _id + '-img-wrapper').css('background-image', '');


            };

            img.onerror = function (e) {
                $('#' + _id).css('background-image', '');

                prx.imageChecked[toCheck] = missing;
                $('#' + _id + '-img').attr('src', missing);

                var title = $('<div>' + prx.utils.escapeHTML(item.caption) + '</div>');
                $('#' + _id + '-img-wrapper').addClass('missing');
                $('#' + _id + '-img-wrapper').css('background-image', '');

                $('#' + _id + '-img-wrapper').find('div').remove();
                $('#' + _id + '-img-wrapper').prepend($('<div></div>'));
                $('#' + _id + '-img-wrapper').append(title);
                $('#' + _id + '-img-wrapper').append($('<div></div>'));

                container.removeChild(img);
            };

            var toCheck = '' + assetUrl;

            img.style.visibility = 'hidden';
            container.appendChild(img);
            img.setAttribute('src', toCheck);
            img.setAttribute('alt', 'na');

            if (prx.imageChecked[assetUrl] !== undefined && prx.imageChecked[assetUrl] !== missing) {
                assetUrl = prx.imageChecked[assetUrl] || assetUrl;
                prx.items.addAll();
            }

            assetUrl = prx.imageChecked[assetUrl] || assetUrl;

        }

        var missingClass = '', wrapperBackground = assetUrl;
        if (assetUrl == missing) {
            thumbUrl == '';
            missingClass = ' missing';
            wrapperBackground = '';
        }
        //gy check for gifs
        var fileId = prx.componentsHelper.getProp(item.imgSrc.fileId,'other');
        var fileType = fileId.substr(fileId.lastIndexOf('.') + 1);


        if( typeof(item.borderColor) == 'undefined' ) item.borderColor = '#000000';
        if( typeof(item.borderWidth) == 'undefined' ) item.borderWidth = 0;
        if( typeof(item.borderRadius) == 'undefined' ) item.borderRadius = 0;
        if( typeof(item.borderPos) == 'undefined' ) item.borderPos = 'inside';
        if(typeof(item.borderStyle) == 'undefined') { item.borderStyle = 'solid'; }

        if(prx.mask.isActive(item))
        {
            var thumbUrl2 = thumbUrl;
            thumbUrl = '';
        }

        if(thumbUrl != '' ) {thumbUrl = ' style="background-image:  url(' + thumbUrl + ');" ';}

        let ariaAttribute = item.assistiveDescription ? 'role="img" aria-label="' + prx.utils.escapeHTML(item.assistiveDescription) + '"' : '';
        var cR = '<div id="' + _id + '" ' +  ariaAttribute + ' ' + thumbUrl;
        cR +=  ' ' + prx.items.getComponentBaseAttributes(item, containerid, symbol)  + ' class="' + prx.items.getComponentBaseClasses(item, containerid, symbol) + ' box pos type-image ' + (prx.componentsHelper.getProp(item.propagateEvents,'boolean') ? 'pointer-events-none' : '') + ((prx.componentsHelper.getProp(item.repeat,'boolean')) ? ' type-image-repeater' : '') + '" ' + ((prx.componentsHelper.getProp(item.overlay,'boolean')) ? 'data-mpoverlay="1"' : '') + ' >';

        cR += '<style>';
        cR += prx.items.getComponentBaseStyle(item, containerid, symbol);
        cR += '</style>';

        if(prx.mask.isActive(item))
            cR += prx.items.getComponentPrependDivs(item, containerid, symbol, thumbUrl2);
        else
            cR += prx.items.getComponentPrependDivs(item, containerid, symbol, '');

        cR += '<div class="image-inner borderPos-'+prx.componentsHelper.getProp(item.borderPos,'other')+'">';

        cR += '<div id="' + _id + '-img-wrapper" class="type-image-wrapper liveUpdate-borderColor-border-color changeProperty-borderColor changeProperty-borderRadius changeProperty-borderWidth ' + ((item.imgSrc.fileId !== undefined && prx.componentsHelper.getProp(item.imgSrc.fileId.slice(-4),'other') == '.svg') ? ' type-image-svg' : '') + missingClass + ((prx.componentsHelper.getProp(item.repeat,'boolean')) ? ' type-image-repeater' : '')  + (fileType=='gif' ? ' gif' : '') + '" ' +
            'style="background-image: url(' + wrapperBackground + ');">';

        cR += '<div></div>';

        var tempH = item.height, tempW = item.width;

        cR += '<img id="' + _id + '-img" src="' + ((prx.componentsHelper.getProp(item.imgSrc.fileId,'other') == '') ? 'data:image/gif;base64,R0lGODlhAQABAIAAAAAAAP///yH5BAEAAAAALAAAAAABAAEAAAIBRAA7" style="width:'+tempW+'px;height:'+tempH+'px"' : assetUrl) + '" width="100%" height="100%" />';

        if (assetUrl == missing)
            cR += '<div>' + prx.utils.escapeHTML(item.caption) + '</div>';

        cR += '<div></div>';
        cR += '</div>';

        cR += '</div>';

        if (prx.componentsHelper.getProp(item.imgSrc.fileId,'other') == 'd310bece0e91b91b485ed62166d1fc2e.svg' && prx.componentsHelper.getProp(item.imgSrc.assetType,'other') == 'gallery') {
        	if(prx.editor) {
		        cR += '<div class="type-image-hover-message">Double-click to edit image</div>';
	        }
            cR += '<style>#' + _id + ' .type-image-wrapper { background-color: #f9f9f9; }</style>';
        }

        cR += '<style>';
        cR += '#'+_id+', #'+_id+' .type-image-wrapper { border-radius: '+prx.componentsHelper.getProp(item.borderRadius,'border-radius')+';}';
        cR += '#'+_id+' .type-image-wrapper { border-width: '+prx.componentsHelper.getProp(item.borderWidth,'border-width')+'; border-style: ' + prx.componentsHelper.getProp(item.borderStyle,'border-type') +'; border-color: '+prx.componentsHelper.getProp(item.borderColor,'color-border')+';  }';
        cR += '</style>';

        cR += prx.items.getComponentAppendDivs(item, containerid, symbol);
        cR += '</div>';

        return cR;
    }
    , onResize: function(item, containerid, symbol) {
    	// disable autoresize
        prx.componentsHelper.setAutoresizeValue(item, containerid, symbol, false);
    }
    , interactions: [{
        caption: 'Interactions',
        name: 'actions',
        type: 'action',
        value: function (item, name) {
            if (typeof(item.actions) == 'undefined') {
                item.actions = [];
            }
            return item.actions.length;
        },
        hiddenByDefault: function (item) {
            if (typeof(item.propagateEvents) != 'undefined' && item.propagateEvents) {
                return true;
            }
            return false;
        }
    }]
    ,propertyGroups: [
        {
            caption: 'Image',
            properties: [
                [
                    {
                        caption: false
                        ,name: 'imgSrc'
                        ,type: 'combo-asset'
                        ,displayValue: function(item,name) {
                            if(item.imgSrc.fileId == '') {
                                return 'No asset selected.';
                            }
                            return prx.utils.escapeHTML(item.imgSrc.name);
                        }
                        ,value: function(item,name) {
                            return JSON.stringify({
                                allow: 'image',
                                asset: item.imgSrc
                            });
                        }
                        ,changeProperty: {
                            caption: 'Image',
                            rerender: true
                        },
                        hiddenByDefault: function(item){
                            if(prx.mask.isActive(item) || prx.mask.editModeIsMaskSelected(item))
                            //if(prx.mask.editModeIsMaskSelected(item))
                                return true;
                            else
                                return false;
                        }

                    }
                ],
                [
                    {
                        caption: 'Reset to 1x size'
                        ,name: 'resetdimensionsdpr'
                        ,type: 'link'
                        ,onClick: function(item) {

                            return prx.componentsHelper.resetDimensions(item, true);
                        }
                        ,hiddenByDefault: function(item){
                            item.imgSrc.fileId = item.imgSrc.fileId || '';
                            //return value
                            var r = prx.imageFunctions.checkImageMultiplier(item) == false;

                            if(prx.mask.isActive(item) || prx.mask.editModeIsMaskSelected(item))
                            //if(prx.mask.editModeIsMaskSelected(item))
                                r = r || true;
                            else
                                r = r || false;

                            return r;
                        }
                    }
                ],
                [
                    {
                        caption: function(item){

                            var multiplier = prx.imageFunctions.checkImageMultiplier(item);

                            if (multiplier !== false)
                                return 'Reset to ' + multiplier + 'x size (original)';
                            else
                                return 'Reset to original size';
                        }
                        ,name: 'resetdimensions'
                        ,type: 'link'
                        ,onClick: function(item) {

                            return prx.componentsHelper.resetDimensions(item);
                        },
                        hiddenByDefault: function(item){
                            if(prx.mask.isActive(item) || prx.mask.editModeIsMaskSelected(item))
                                return true;
                            else
                                return false;
                        }
                    }
                ],
                [
                    {
                        caption: 'Reset to aspect ratio'
                        , name: 'resetaspectratio'
                        , type: 'link'
                        , onClick: function(item) {
                            if ($.browser.msie) {
                                $('#' + item.id + ' > img').removeAttr('width').removeAttr('height').css({
                                    width: 'auto',
                                    height: 'auto'
                                }).prop('naturalWidth', $('#' + item.id + ' img').width()).prop('naturalHeight', $('#' + item.id + ' img').height());
                            }

                            var _beforeWidth = $('#' + item.id + ' img').prop('naturalWidth');
                            var _beforeHeight = $('#' + item.id + ' img').prop('naturalHeight');


                            if(prx.mask.isActive(item))
                            {
                                return item;
                            }
                            else
                            {
                                var _afterWidth = item.width;
                                var _afterHeight = (_afterWidth / _beforeWidth) * _beforeHeight;

                                _afterHeight = parseInt(Math.round(_afterHeight));

                                item.height = _afterHeight;

                                //if is edit mode we only update the mask view - no resizing of mask area
                                if(prx.mask.isEditing(item)){
                                    prx.mask.editModeRerenderMaskView(item);
                                    prx.mask.editModeRefreshEditMode(item);
                                }
                            }

                            item.aspectratio = (_afterWidth / _beforeWidth);

                            $('#property-aspectratio').prop( 'checked', true ).change();

                            return item;
                        },
                        hiddenByDefault: function(item){
                            item.imgSrc.fileId = item.imgSrc.fileId || '';
                            //return value
                            var r = item.imgSrc.fileId.slice(-4) == '.svg';

                            if(prx.mask.isActive(item) || prx.mask.editModeIsMaskSelected(item))
                            //if(prx.mask.editModeIsMaskSelected(item))
                                r = r || true;
                            else
                                r = r || false;

                            return r;

                        }
                    }
                ],
                [
                    {
                        caption: 'Repeat image instead of stretch'
                        ,name: 'repeat'
                        ,type: 'onoff'
                        ,value: function(item,name) {
                            if(typeof(item.repeat) == 'undefined') {
                                return false;
                            }
                            return item.repeat;
                        }
                        ,changeProperty: {
                            caption: 'Repeat image',
                            rerender: true
                        },
                        hiddenByDefault: function(item){
                            item.imgSrc.fileId = item.imgSrc.fileId || '';
                            var r = !item.repeat && item.imgSrc.fileId.slice(-4) == '.svg';


                            if(prx.mask.isActive(item) || prx.mask.editModeIsMaskSelected(item))
                                r = r || true;
                            else
                                r = r || false;

                            return r;
                        }
                    }
                ],

            ]
        },
        {
            id: 'mask-settings',
            properties: [
                [
                    {
                        caption: '<div class="icon icon-mask"></div>Mask/Crop'
                        , name: 'maskImage'
                        , type: 'onoff'
                        , onChange: function(item) {
                            var tmp = item.maskImage;
                            delete item.maskImage;

                            if(tmp)
                                prx.mask.enable(item);
                            else
                                prx.mask.disable(item);

                            return item;
                        }
                        ,value: function(item,name) {
                            return prx.mask.isEnabled(item);
                        }
                    },

                ],
                [
                    {
                        caption: 'Reset aspect ratio'
                        , name: 'resetaspectratiomask'
                        , type: 'link'
                        , onClick: function(item) {

                            if(prx.mask.isActive(item)) {

                                var mask_h = item.height; //mask original height

                                //calculate mask's height based on last saved ar
                                item.height = parseInt(item.width / item.mask.last_aspectratio);

                                //vertical change ratio
                                var changeRadioH = item.height/mask_h;

                                //calculate image's height based on the above change
                                item.mask.inner_height = parseInt(item.mask.inner_height * (changeRadioH));

                                //calculate image's y
                                item.mask.y = Math.round((item.mask.y) * changeRadioH);

                                item.aspectratio = (item.mask.last_aspectratio); //set aspect ratio
                                $('#property-aspectratio').prop( 'checked', true ).change(); //lock aspect ratio
                            }

                            return item;
                        }
                        , hiddenByDefault: function(item){
                            item.imgSrc.fileId = item.imgSrc.fileId || '';
                            //return value
                            var r = item.imgSrc.fileId.slice(-4) == '.svg';

                            if(!prx.mask.isActive(item))
                                r = r || true;
                            else
                                r = r || false;

                            return r;

                        }
                    }
                ]
            ]
        },
        {
            caption: 'Style',
            properties: [
                [
                    {
                        caption: 'Border',
                        name: 'borderWidth',
                        proptype: 'border-width',
                        type: 'combo-select',
                        value: function(item,name) { return item.borderWidth; },
                        values: { min: 0, max: 20, step: 1 } ,
                        expandableType: 'borderWidth',
                        expandedValues: ['t', 'r', 'l', 'b'],
                        changeProperty: {
                            caption: 'Border Width',
                            selector: '.changeProperty-borderWidth',
                            property: 'border-width',
                            transitionable: true
                        },
                        hiddenByDefault: function(item){
                            if(prx.mask.isActive(item) || prx.mask.editModeIsMaskSelected(item))
                                return true;
                            else
                                return false;
                        }
                    },
                    {
                        caption: false,
                        name: 'borderStyle',
                        proptype: 'border-style',
                        type: 'select',
                        value: function(item,name) {
                            if(typeof(item.borderStyle) == 'undefined') { item.borderStyle = 'solid'; }
                            return item.borderStyle;
                        },
                        values: [{ value: 'solid', displayValue: 'Solid'},{ value: 'dotted', displayValue: 'Dotted'},{ value: 'dashed', displayValue: 'Dashed'},{ value: 'double', displayValue: 'Double'},{ value: 'none', displayValue: 'None'}],
                        changeProperty: {
                            caption: 'Border Style',
                            selector: '.changeProperty-borderWidth',
                            property: 'border-style',
                            transitionable: false
                        },
                        hiddenByDefault: function(item) {
                            if(prx.mask.isActive(item) || prx.mask.editModeIsMaskSelected(item))
                                return true;
                            else
                                return false;
                        }
                    },
                    {
                        caption: false,
                        name: 'borderColor',
                        proptype: 'border-color',
                        type: 'solid-colorpicker',
                        value: function(item, name) {
                            return typeof item.borderColor == 'undefined' ? '' : item.borderColor;
                        },
                        liveUpdate: 'border-color',
                        changeProperty: {
                            caption: 'Border color',
                            selector: '.changeProperty-borderColor',
                            property: 'border-top-color,border-bottom-color,border-left-color,border-right-color',
                            transitionable: true
                        },
                        hiddenByDefault: function(item){
                            if(item.type == 'image')
                                if(prx.mask.isActive(item) || prx.mask.editModeIsMaskSelected(item))
                                    return true;
                                else
                                    return false;
                            return false;
                        }
                    }
                ],
                [
                    {
                        caption: 'Position'
                        ,name: 'borderPos'
                        ,type: 'select'
                        ,value: function(item,name,index) {
                            if(typeof(item.borderPos) == 'undefined') {
                                item.borderPos = 'inside';
                            }
                            return item.borderPos;
                        }
                        ,values: [{ displayValue: 'Inside', value: 'inside' }, { displayValue: 'Outside', value: 'outside' }]
                        ,changeProperty: {
                            caption: 'Border Position',
                            selector: '.image-inner',
                            property: 'class',
                            transitionable: false
                        },
                        hiddenByDefault: function(item){
                            if(prx.mask.isActive(item) || prx.mask.editModeIsMaskSelected(item))
                                return true;
                            else
                                return false;
                        }
                    },
                    {
                        caption: 'Radius',
                        name: 'borderRadius',
                        proptype: 'border-radius',
                        type: 'combo-select',
                        value: function(item, name) {
                            return typeof item.borderRadius == 'undefined' ? '' : item.borderRadius;
                        },
                        values: {min: 0, max: 20, step: 1},
                        expandableType: 'borderRadius',
                        expandedValues: ['tl', 'tr', 'bl', 'br'],
                        changeProperty: {
                            caption: 'Border radius',
                            selector: '.changeProperty-borderRadius',
                            property: 'border-radius',
                            transitionable: true
                        },
                        hiddenByDefault: function(item){
                            if(prx.mask.isActive(item) || prx.mask.editModeIsMaskSelected(item))
                                return true;
                            else
                                return false;
                        }
                    },

                ]
            ]
        },
        {
            caption: 'Advanced',
            properties: [
                [
                    {
                        caption: 'Is invisible to touch & mouse events'
                        ,help: 'Turning this property on will cause the item to become invisible to touch or mouse events and allow other items below to become interactive'
                        ,name: 'propagateEvents'
                        ,type: 'onoff'
                        ,value: function(item,name) {
                            if(typeof(item.propagateEvents) == 'undefined') {
                                return false;
                            }
                            return item.propagateEvents;
                        }
                        ,onChange: function(item){
                            /*
						 if(item.propagateEvents) {
						 ('#property-actions').hide();
						 } else {
						 $('#property-actions').show();
						 }*/
                            // will force rerender of properties to show/hide the interactions tab
                            return item;
                        }
                        ,changeProperty: {
                            caption: 'Invisible to touch/mouse events',
                            rerender: true
                        },
                        hiddenByDefault: function(item){
                            if(prx.mask.isEditing(item))
                                return true;
                            else
                                return false;
                        }
                    }],
                [
                    {
                        caption: 'Keep position during screen animations'
                        ,help: 'Will not animate with the rest of the screen during Screen transitions or "Move screen" interactions'
                        ,name: 'overlay'
                        ,type: 'onoff'
                        ,value: function(item,name) {
                            if(typeof(item.overlay)=='undefined') {
                                return false;
                            }
                            return item.overlay;
                        }
                        ,changeProperty: {
                            caption: 'Maintain position during screen animations',
                            rerender: true
                        },
                        hiddenByDefault: function(item){
                            if(prx.mask.isEditing(item))
                                return true;
                            else
                                return false;
                        }
                    }
                ]
            ]
        },
        {
            caption: 'Accessibility',
            properties: [
                [
                    {
                        caption: 'Descriptive text'
                        , hint: 'For assistive devices and screen readers'
                        , name: 'assistiveDescription'
                        , type: 'input'
                        , value: function (item, name) {
                            return item.assistiveDescription ? item.assistiveDescription : '';
                        }
                        , onChange: function (item) {
                            if (item.assistiveDescription !== '') {
                                document.getElementById(item.id).setAttribute('aria-label', item.assistiveDescription);
                            }
                        },
                        changeProperty: {
                            caption: 'Description for assistive devices',
                            transitionable: false,
                            changeFunction: function (item, containerid) {
                                let _id = ((!containerid) ? item.id : containerid + '-' + item.id);
                                document.getElementById(_id).setAttribute('aria-label', item.assistiveDescription);
                            }
                        }
                    }
                ]
            ]
    }
    ]
};

//TYPE: PLACEHOLDER
prx.types.placeholder = {
    name: 'placeholder'
    ,onDisplay: function(item,containerid,symbol) {
        var _id = (!containerid) ? item.id : containerid+'-'+item.id;

        var _real = prx.componentsHelper.getRealDims(item, symbol);
        var _realh = _real.height;
        var _realw = _real.width;

        var _width = Math.sqrt(Math.pow(_realw,2) + Math.pow(_realh,2)) - prx.componentsHelper.getProp(item.thickness,'num-other');
        var _angle1 = 90 - Math.atan((_realw-prx.componentsHelper.getProp(item.thickness,'num-other')*2)/(_realh-prx.componentsHelper.getProp(item.thickness,'num-other')*2)) * (180/Math.PI);

        if(typeof(item.textColor) == 'undefined') {
            item.textColor = prx.componentsHelper.getProp(item.borderColor,'other');
        }
        if(typeof(item.textFont) == 'undefined') {
            item.textFont = "Helvetica,sans-serif";
        }
        if(typeof(item.textFontStyle) == 'undefined') {
            item.textFontStyle = 400;
        }
        if(typeof(item.textProperties) == 'undefined') {
            item.textProperties = [];
        }

        //needs the width patenta to correctly calculate width on afterdisplay
        var cR = '';

        var _props = (typeof(prx.richtext)=='undefined') ? prx.componentsHelper.getProp(item.textProperties,'props-text') : '';


        cR += '<div id="' + _id + '" ' + prx.items.getComponentBaseAttributes(item, containerid, symbol)  + ' class="' + prx.items.getComponentBaseClasses(item, containerid, symbol) + ' pos box type-placeholder" style="width: '+_realw+'px;">';
        cR += '<style>';
        cR += prx.items.getComponentBaseStyle(item, containerid, symbol);
        cR += '</style>';
        cR += prx.items.getComponentPrependDivs(item, containerid, symbol);
        cR += '<div class="bg liveUpdate-backgroundColor-background-color liveUpdate-borderColor-border-color changeProperty-backgroundColor changeProperty-borderColor" style="'+prx.gradients.getBgCssByProperty(item.backgroundColor,'color-background')+'; border-color: '+prx.componentsHelper.getProp(item.borderColor,'color-border')+'; border-width: '+prx.componentsHelper.getProp(item.thickness,'num-border-width')+'px;"></div>';
        cR += '<div class="diagonal diagonal1 liveUpdate-borderColor-border-color changeProperty-borderColor" style="border-color: '+prx.componentsHelper.getProp(item.borderColor,'color-border')+'; width: '+_width+'px; -moz-transform: rotate('+_angle1+'deg); -webkit-transform: rotate('+_angle1+'deg); -o-transform: rotate('+_angle1+'deg); transform: rotate('+_angle1+'deg);  border-top-width: '+prx.componentsHelper.getProp(item.thickness,'num-border-width')+'px; left: '+prx.componentsHelper.getProp(item.thickness,'num-other')/2+'px;"></div>';
        cR += '<div class="diagonal diagonal2 liveUpdate-borderColor-border-color changeProperty-borderColor" style="border-color: '+prx.componentsHelper.getProp(item.borderColor,'color-border')+'; width: '+_width+'px; -moz-transform: rotate(-'+_angle1+'deg); -webkit-transform: rotate(-'+_angle1+'deg); -o-transform: rotate(-'+_angle1+'deg); transform: rotate(-'+_angle1+'deg); border-top-width: '+prx.componentsHelper.getProp(item.thickness,'num-border-width')+'px; right: '+prx.componentsHelper.getProp(item.thickness,'num-other')/2+'px;"></div>';
        //if(item.text != "") {
        cR += '<div class="contents" style="position:relative;">';
        cR += '<span class="shapes-text-container liveUpdate-backgroundColor-background-color changeProperty-backgroundColor liveUpdate-textColor-color" style="' + _props + ' ' + prx.componentsHelper.getProp(item.textFont + '|' + item.textFontStyle,'font-family') + prx.componentsHelper.getProp(item.textFontStyle,'font-style') + '; font-size: '+prx.componentsHelper.getProp(item.textSize,'num-text-size')+'px; '+prx.gradients.getBgCssByProperty(item.backgroundColor,'color-background')+'; color: '+prx.componentsHelper.getProp(item.textColor,'color-text')+';"><span data-editableproperty="text">' + prx.componentsHelper.getProp(item.text,'text-textarea') + '</span></span></div>';
        //}
        cR += prx.items.getComponentAppendDivs(item, containerid, symbol);
        cR += '</div>';
        return cR;
    }
    ,afterDisplay: function(item,containerid,symbol) {
        var _id = (!containerid) ? item.id : containerid+'-'+item.id;

        var _real = prx.componentsHelper.getRealDims(item, symbol);
        var _realh = _real.height;
        var _realw = _real.width;

        if( _realh <= item.thickness*2 || _realw <= item.thickness*2 ) {
            $('#' + _id + ' .diagonal1, #' + _id + ' .diagonal2').css('display', 'none');
        } else {
            $('#' + _id + ' .diagonal1, #' + _id + ' .diagonal2').css('display', 'block');
        }

    }
    ,onResize: function(item,containerid,symbol) {
        var _id = (!containerid) ? item.id : containerid+'-'+item.id;

        var _real = prx.componentsHelper.getRealDims(item, symbol);
        var _realh = _real.height;
        var _realw = _real.width;

        var _width = Math.sqrt(Math.pow(_realw,2) + Math.pow(_realh,2)) - item.thickness;
        var _angle1 = 90 - Math.atan((_realw-item.thickness*2)/(_realh-item.thickness*2)) * (180/Math.PI);

        if( _realh <= item.thickness*2 || _realw <= item.thickness*2 ) {
            $('#' + _id + ' .diagonal1, #' + _id + ' .diagonal2').css('display', 'none');
        } else {
            $('#' + _id + ' .diagonal1, #' + _id + ' .diagonal2').css('display', 'block');
        }

        $('#' + _id + ' .diagonal1')
            .width(_width)
            .css('-moz-transform', 'rotate(' + _angle1 + 'deg)')
            .css('-webkit-transform', 'rotate(' + _angle1 + 'deg)')
            .css('-o-transform', 'rotate(' + _angle1 + 'deg)')
            .css('transform', 'rotate(' + _angle1 + 'deg)');
        $('#' + _id + ' .diagonal2')
            .width(_width)
            .css('-moz-transform', 'rotate(-' + _angle1 + 'deg)')
            .css('-webkit-transform', 'rotate(-' + _angle1 + 'deg)')
            .css('-o-transform', 'rotate(-' + _angle1 + 'deg)')
            .css('transform', 'rotate(-' + _angle1 + 'deg)');

    }
    ,interactions: [
        prx.commonproperties.actions
    ]
    ,editableProperties: [
        {
	    	caption: 'Text'
	    	,name: 'text'
	    	,type: 'textarea'
	    	,value: function(item,name) {
	    		return item.text;
	    	}
	    	,changeProperty: {
                caption: 'Text',
                property: 'text',
                selector: '.shapes-text-container span',
                transitionable: false
			 }
	    }
    ]
    ,propertyGroups: [
	    {
	    	caption: 'Style',
	    	properties: [
		    	[
		    		prx.commonproperties.backgroundColorSolid
		    	],
                [
                    {
                        caption: 'Border'
                        ,name: 'thickness'
                        ,proptype: 'border-width'
                        ,type: 'combo-select'
                        ,value: function(item,name) {
                            return item.thickness;
                        }
                        ,values: prx.comps.onetoten
                        ,changeProperty: {
                            caption: 'Border Width',
                            rerender: true
                        }
                    }
                    ,prx.commonproperties.borderColor
                ]
            ]
        },
        {
            caption: 'Text',
            properties: [
                [
                    {
                        caption: { label: 'Font family', class: 'text-properties-label text-fontfamily-label' },
                        name: 'textFont',
                        proptype: 'font-family',
                        type: 'select',
                        relatedEditableProperties: 'text',
                        relatedCSSProperties: 'font-family',
                        value: function(item,name) {
                            if(typeof(item.textFont) == 'undefined') { item.textFont = "Helvetica,sans-serif"; }
                            return item.textFont;
                        },
                        values: function(){ return prx.comps.fonts; }
                        ,changeProperty: {
                            caption: ' Text font',
                            selector: '.shapes-text-container',
                            property: 'font-family',
                            transitionable: false
                        }

                    }
                ],
                [
                    prx.commonproperties.textFontStyleRichText(['font-weight', 'font-style'],'text', false, {propName: 'textFontStyle', caption: 'Font style', useAsDynProp: false}),
                    {
                        caption: false,
                        name: 'textSize',
                        proptype: 'font-size',
                        type: 'combo-select',
                        relatedEditableProperties: 'text',
                        relatedCSSProperties: 'font-size',
                        value: function(item,name) {
                            if(typeof(item.textSize) == 'undefined') { item.textSize = 16; }
                            return item.textSize;
                        },
                        values: prx.comps.textsize
                        ,changeProperty: {
                            caption: ' Text size',
                            selector: '.shapes-text-container',
                            property: 'font-size',
                            transitionable: true
                        }
                    }
                ]
                ,
                [
                    prx.commonproperties.textPropertiesRichText(['font-weight','font-style','text-decoration'],'text', false, {propName: 'textProperties', caption: 'Underline', useAsDynProp: false}),
                    {
                        caption: { label: 'Color', class: 'text-properties-label text-color-label' },
                        name: 'textColor',
                        proptype: 'font-color',
                        type: 'solid-colorpicker',
                        relatedEditableProperties: 'text',
                        relatedCSSProperties: 'color',
                        pffSettings: prx.commonproperties.pffSettingsColor,
                        value: function(item,name) {
                            if(typeof(item.textColor) == 'undefined') { item.textColor = '#2E2E2E'; }
                            return item.textColor;
                        },
                        liveUpdate: 'color'
                        ,changeProperty: {
                            caption: ' Text color',
                            selector: '.shapes-text-container',
                            property: 'color',
                            transitionable: true
                        }
                    }
                ]
            ]
        }

    ]
};

//TYPE: WEBVIEW
prx.types.webview = {
    name: 'webview'
    ,onDisplay: function(item,containerid,symbol) {
        var _id = (!containerid) ? item.id : containerid+'-'+item.id;

        if(typeof(item.scrollable) == 'undefined') {
            item.scrollable = false;
        }

        var cR = '';

        cR += '<div id="' + _id + '" ' + prx.items.getComponentBaseAttributes(item, containerid, symbol)  + ' class="' + prx.items.getComponentBaseClasses(item, containerid, symbol) + ' box pos type-webview">';
        cR += '<style>';
        cR += prx.items.getComponentBaseStyle(item, containerid, symbol);
        cR += '</style>';
        cR += prx.items.getComponentPrependDivs(item, containerid, symbol);
        cR += '<div style="overflow: hidden; height: 100%; width: 100%;">';
        if(!prx.editor && prx.iosmobilebrowser) {
            cR += '<div class="webview-scroll-wrapper">';
        }
        cR += '<iframe src='+prx.componentsHelper.getProp(item.url,'url')+' scrolling='+((prx.componentsHelper.getProp(item.scrollable,'boolean')) ? '"yes"' : '"no"') +' style="border: none; width: 100%; height: 100%;"></iframe>';
        if(!prx.editor && prx.iosmobilebrowser) {
            cR += '</div>';
        }
        cR += '</div>';
        cR += prx.items.getComponentAppendDivs(item, containerid, symbol);
        cR += '</div>';

        return cR;
    }
    ,propertyGroups: [
        {
            caption: 'Properties',
            properties: [
	             [
				      {
					    	caption: 'URL'
					    	,name: 'url'
					    	,type: 'input'
					    	,value: function(item,name) {
					    		return prx.componentsHelper.getProp(item.url,'url');
					    	}
				      		,help: 'Works only with https:// URLs, but it will work for http:// if you use Export to HTML'
						    ,changeProperty: {
                            caption: 'URL',
                            rerender: true
							 }
                            , onChange: function (item) {
                                if(prx.editor && item.url.indexOf("<iframe") > -1) {
                                    var p = /src="([^"]+)"/;
                                    var result = item.url.match(p);
                                    if (result && result[1] && result[1].length) {
                                        item.url = result[1];
                                        return item;
                                    }
                                }
                            },
					    }
				 ],
			     [
		              {
                        caption: 'Scrollable?'
                        ,name: 'scrollable'
                        ,type: 'onoff'
                        ,value: function(item,name) {
                            if(typeof(item.scrollable)=='undefined') {
                                return false;
                            }
                            return item.scrollable;
                        }
                        ,changeProperty: {
				        		caption: 'Scrollable',
				        		rerender: true
				        	}
		              }
	             ]
            ]
        }
    ]
};

//TYPE: HTML
prx.types.html = {
    name: 'html'
    ,onDisplay: function(item,containerid,symbol) {
        var _id = (!containerid) ? item.id : containerid+'-'+item.id;
        var cR = '';
        cR += '<div id="' + _id + '" ' + prx.items.getComponentBaseAttributes(item, containerid, symbol)  + ' class="' + prx.items.getComponentBaseClasses(item, containerid, symbol) + ' box pos type-html">';
        cR += '<style>';
        cR += prx.items.getComponentBaseStyle(item, containerid, symbol);
        cR += '</style>';
        cR += prx.items.getComponentPrependDivs(item, containerid, symbol);
        cR += prx.items.getComponentAppendDivs(item, containerid, symbol);
        cR += '</div>';
        return cR;
    }
    ,afterDisplay: function(item, containerid, symbol) {
        var _id = (!containerid) ? item.id : containerid+'-'+item.id;
        var iframe = new prx.componentsHelper.IFrame($('#'+_id).get(0));
        iframe.doc.body.style.margin = '0px';
	    var div = iframe.doc.createElement('div');
	    div.innerHTML = DOMPurify.sanitize(item.html, {ADD_TAGS: ['iframe', 'html', 'body'], FORCE_BODY: true});
	    iframe.doc.body.appendChild(div);
    }
    ,propertyGroups: [
        {
            caption: 'HTML Code',
            properties: [[
		      	{
			    	caption: false
			    	,name: 'html'
			    	,type: 'html-textarea'
			    	,value: function(item,name) {
			    		return DOMPurify.sanitize(item.html, {ADD_TAGS: ['iframe', 'html', 'body'], FORCE_BODY: true});
			    	}
			    	,changeProperty: {
                        caption: 'HTML code',
                        rerender: true,
                        changeable: true
					 }
			    }
            ]]
        }
    ]
};

//TYPE: AUDIO
prx.types.audio = {
    name: 'audio'
    ,onDisplay: function(item,containerid,symbol) {
        let sourceType = '';
        if(item.audioFile) {
            let audioType= prx.componentsHelper.isAudioFile(item.audioFile.name) ? item.audioFile.name.split('.').pop() : "mp3";
            sourceType = prx.componentsHelper.getAudioFileSourceType(audioType);
        }

        var _id = (!containerid) ? item.id : containerid+'-'+item.id;
        if(typeof(item.loop)=='undefined') { item.loop = false; }
        var cR = '';
        cR += '<div id="' + _id + '" ' + prx.items.getComponentBaseAttributes(item, containerid, symbol)  + ' class="' + prx.items.getComponentBaseClasses(item, containerid, symbol) + ' box pos type-audio">';
        cR += '<style>';
        cR += prx.items.getComponentBaseStyle(item, containerid, symbol);
        cR += '</style>';
        cR += prx.items.getComponentPrependDivs(item, containerid, symbol);
        cR += '<audio '+ ((prx.componentsHelper.getProp(item.controls,'boolean')) ? 'controls' : '') +' '+ ((prx.componentsHelper.getProp(item.preload,'boolean') && !prx.editor) ? 'preload' : 'preload="none"') +' '+ ((prx.componentsHelper.getProp(item.autoplay,'boolean') && !prx.editor) ? 'autoplay' : '') +' '+ ((prx.componentsHelper.getProp(item.loop,'boolean')) ? 'loop' : '') +' controlsList="nodownload">';
        if(typeof(item.audioFile) !== 'undefined' && prx.componentsHelper.getProp(item.audioFile.fileId,'other') != '') cR += '<source src="'+prx.componentsHelper.getProp(item.audioFile,'asset')+'" type="audio/'+ sourceType +'" />';
        if(prx.componentsHelper.getProp(item.audioFileMP3.fileId,'other') != '') cR += '<source src="'+prx.componentsHelper.getProp(item.audioFileMP3,'asset')+'" type="audio/mpeg" />';
        if(prx.componentsHelper.getProp(item.audioFileOGG.fileId,'other') != '') cR += '<source src="'+prx.componentsHelper.getProp(item.audioFileOGG,'asset')+'" type="audio/ogg" />';
        if(prx.componentsHelper.getProp(item.audioFileWAV.fileId,'other') != '') cR += '<source src="'+prx.componentsHelper.getProp(item.audioFileWAV,'asset')+'" type="audio/x-wav" />';
        if(prx.componentsHelper.getProp(item.audioFileAAC.fileId,'other') != '') cR += '<source src="'+prx.componentsHelper.getProp(item.audioFileAAC,'asset')+'" type="audio/x-m4a" />';
        cR += '</audio>';
        cR += prx.items.getComponentAppendDivs(item, containerid, symbol);
        cR += '</div>';
        return cR;
    }
    ,interactions: [prx.commonproperties.actions]
    ,mpactions: {
       	specialEvents: ['audioplay','audiopause','audioend']
    }
    ,afterDisplay: function(item,containerid,symbol) {
    	var _id = (!containerid) ? item.id : containerid+'-'+item.id;

        if(!prx.editor){

            /* for special events */
            var audio = $('#'+_id).find('audio').get(0);

            audio.onplay = function() {
                $('#'+_id).trigger('audioplay');
	        };

            audio.onpause = function() {
                $('#'+_id).trigger('audiopause');
	        };

            audio.onended = function() {
                $('#'+_id).trigger('audioend');
	        };
        }
    }
    ,propertyGroups: [
        {
            caption: 'Audio properties',
            properties: [
                [{
                    caption: 'Audio file MP3'
                    ,name: 'audioFileMP3'
                    ,type: 'asset'
                    ,displayValue: function(item,name) {
                        if(item.audioFileMP3.fileId == '') {
                            return 'No asset selected.';
                        }

                        return prx.utils.escapeHTML(item.audioFileMP3.name);
                    }
                    ,hiddenByDefault: function(item) {
                        if(prx.componentsHelper.getAudioFilesTotal(item) <= 1){
                            if(item.audioFileMP3.fileId != '') {
                                item.audioFile = item.audioFileMP3;
                                item.audioFileMP3 = {fileId: '', assetType: '', name: ''};
                            }
                            return true;
                        }
                        return false;
                    }
                    ,value: function(item,name) {
                        return JSON.stringify({
                            allow: 'audio',
                            asset: item.audioFileMP3
                        });
                    }
                    ,changeProperty: {
                    	caption: 'Audio file MP3',
                        rerender: true
					 }
                }],
            [{
                caption: 'Audio file OGG (.oga)'
                ,name: 'audioFileOGG'
                ,type: 'asset'
                ,displayValue: function(item,name) {
                    if(item.audioFileOGG.fileId == '') {
                        return 'No asset selected.';
                    }
                    return prx.utils.escapeHTML(item.audioFileOGG.name);
                }
                ,hiddenByDefault: function(item) {
                    if(prx.componentsHelper.getAudioFilesTotal(item) <= 1){
                        if(item.audioFileOGG.fileId != '') {
                            item.audioFile = item.audioFileOGG;
                            item.audioFileOGG = {fileId: '', assetType: '', name: ''};
                        }
                        return true;
                    }
                    return false;
                },value: function(item,name) {
                    return JSON.stringify({
                        allow: 'audio',
                        asset: item.audioFileOGG
                    });
                }
                ,changeProperty: {
                    caption: 'Audio file OGG',
                    rerender: true
					 }
            }],
            [{
			    	caption: 'Audio file WAV'
			    	,name: 'audioFileWAV'
			    	,type: 'asset'
                ,displayValue: function(item,name) {
                    if(item.audioFileWAV.fileId == '') {
                        return 'No asset selected.';
                    }
                    return prx.utils.escapeHTML(item.audioFileWAV.name);
                }
                ,hiddenByDefault: function(item) {
                    if(prx.componentsHelper.getAudioFilesTotal(item) <= 1){
                        if(item.audioFileWAV.fileId != '') {
                            item.audioFile = item.audioFileWAV;
                            item.audioFileWAV = {fileId: '', assetType: '', name: ''};
                        }
                        return true;
                    }
                    return false;
                }
                ,value: function(item,name) {
                    return JSON.stringify({
                        allow: 'audio',
                        asset: item.audioFileWAV
                    });
                }
                ,changeProperty: {
                    caption: 'Audio file WAV',
                    rerender: true
					 }
			    }],
			    [{
			    	caption: 'Audio file AAC'
			    	,name: 'audioFileAAC'
			    	,type: 'asset'
                ,displayValue: function(item,name) {
                    if(item.audioFileAAC.fileId == '') {
                        return 'No asset selected.';
                    }
                    return prx.utils.escapeHTML(item.audioFileAAC.name);
                }
                    ,hiddenByDefault: function(item) {
                        if(prx.componentsHelper.getAudioFilesTotal(item) <= 1){
                            if(item.audioFileAAC.fileId != '') {
                                item.audioFile = item.audioFileAAC;
                                item.audioFileAAC = {fileId: '', assetType: '', name: ''};
                            }
                            return true;
                        }
                        return false;
                    }
                ,value: function(item,name) {
                    return JSON.stringify({
                        allow: 'audio',
                        asset: item.audioFileAAC
                    });
                }
                ,changeProperty: {
                    caption: 'Audio file AAC',
                    rerender: true
					 }
			    }],
                [{
                    caption: 'Audio file'
                    ,hint: '.mp3 file type recommended'
                    ,name: 'audioFile'
                    ,type: 'asset'
                    ,displayValue: function(item,name) {
                        if(item.audioFile === undefined || item.audioFile.fileId == '') {
                            return 'No asset selected.';
                        }
                        return prx.utils.escapeHTML(item.audioFile.name);
                    }
                    ,hiddenByDefault: function(item) {
                        if(prx.componentsHelper.getAudioFilesTotal(item) > 1) {
                            return true;
                        }
                        return false;
                    }
                    ,value: function(item,name) {
                        return JSON.stringify({
                            allow: 'audio',
                            asset: item.audioFile
                        });
                    }
                    ,changeProperty: {
                        caption: 'Audio file',
                        rerender: true
                    }
                }],
			    [
			    {
			    	caption: 'Preload audio'
			    	,name: 'preload'
			    	,type: 'onoff'
			    	,value: function(item,name){
			    		return item.preload;
			    	}
			    	,changeProperty: {
                    	caption: 'Preload audio',
                        rerender: true
					 }
			    },
			    {
			    	caption: 'Autoplay'
			    	,name: 'autoplay'
			    	,type: 'onoff'
			    	,value: function(item,name){
			    		return item.autoplay;
			    	}
                    ,help: 'Warning: May not work in some browsers/mobile devices due to browser restrictions'
			    	,hiddenByDefault: function(item,name) {
			    		if(typeof(item.autoplayoff) != 'undefined') {
			    			return true;
                        }

                        return false;
                    }
			    	,changeProperty: {
                    	caption: 'Autoplay',
                        rerender: true
					 }
			    },
			    {
			    	caption: 'Loop'
			    	,name: 'loop'
			    	,type: 'onoff'
			    	,value: function(item,name){
			    		return item.loop;
			    	}
			    	,changeProperty: {
                    	caption: 'Loop',
                        rerender: true
					 }
			    },
			    {
			    	caption: 'Display controls'
			    	,name: 'controls'
			    	,type: 'onoff'
			    	,value: function(item,name){
			    		return item.controls;
			    	}
			    	,changeProperty: {
                    	caption: 'Display controls',
                        rerender: true
                    }
			    }]
			    ]
        }
    ]
};

//TYPE: VIDEO
prx.types.video = {
    name: 'video'
    ,onDisplay: function(item,containerid,symbol) {
        var _id = (!containerid) ? item.id : containerid+'-'+item.id;
        let sourceType = '';
        if(item.videoFile) {
            let videoType = prx.componentsHelper.isVideoFile(item.videoFile.name) ? item.videoFile.name.split('.').pop() : "mp4";
            sourceType = prx.componentsHelper.getVideoFileSourceType(videoType);
        }

        if(typeof(item.vimeoid)=='undefined') { item.vimeoid = ''; }
        if(typeof(item.loop)=='undefined') { item.loop = false; }
        if(typeof(item.mute)=='undefined') { item.mute = false; }

        var cR = '';
        cR += '<div id="' + _id + '" ' + prx.items.getComponentBaseAttributes(item, containerid, symbol)  + ' class="' + prx.items.getComponentBaseClasses(item, containerid, symbol) + ' box pos type-video '+prx.componentsHelper.getProp(item.videoType,'other')+'">';
        cR += '<style>';
        cR += prx.items.getComponentBaseStyle(item, containerid, symbol);
        cR += '</style>';
        cR += prx.items.getComponentPrependDivs(item, containerid, symbol);

        switch(prx.componentsHelper.getProp(item.videoType,'other')) {
        case 'html5':
            cR += '<video id="'+_id+'-html5video" class="html5" width="100%" height="100%" '+ ((prx.componentsHelper.getProp(item.controls,'boolean')) ? 'controls' : '') + ' ' + ((prx.componentsHelper.getProp(item.mute,'boolean')) ? 'muted' : '') + ((prx.componentsHelper.getProp(item.placeholder.fileId,'other') != '') ? ' poster='+prx.componentsHelper.getProp(item.placeholder,'asset') : '' )+' '+ ((!prx.componentsHelper.getProp(item.preload,'boolean') && !(prx.editor && prx.componentsHelper.getProp(item.autoplay,'boolean'))) ? 'preload="none"' : '') +' '+ ((prx.componentsHelper.getProp(item.autoplay,'boolean') && !prx.editor) ? 'autoplay' : '') +' '+((prx.componentsHelper.getProp(item.loop,'boolean')) ? 'loop' : '')+' webkit-playsinline="true" playsinline="true" controlsList="nodownload">';
            if(typeof(item.videoFile) !== 'undefined' && prx.componentsHelper.getProp(item.videoFile.fileId,'other') != '') cR += '<source src="'+prx.componentsHelper.getProp(item.videoFile,'asset')+'" type="video/'+sourceType+'" />';
            if(prx.componentsHelper.getProp(item.videoFileMP4.fileId,'other') != '') cR += '<source src="'+prx.componentsHelper.getProp(item.videoFileMP4,'asset')+'" type="video/mp4" />';
            if(prx.componentsHelper.getProp(item.videoFileWEBM.fileId,'other') != '') cR += '<source src="'+prx.componentsHelper.getProp(item.videoFileWEBM,'asset')+'" type="video/webm" />';
            if(prx.componentsHelper.getProp(item.videoFileOGG.fileId,'other') != '') cR += '<source src="'+prx.componentsHelper.getProp(item.videoFileOGG,'asset')+'" type="video/ogg" />';
            cR += '</video>';
            break;
        case 'youtube':
            item.youtubeid = prx.componentsHelper.getProp(item.youtubeid,'video-url');
            if(prx.editor && $.browser.chrome) {
                cR += '<object width="100%" height="100%"><param name="movie" value="//www.youtube.com/v/'+prx.componentsHelper.getProp(item.youtubeid,'other')+'?version=3&amp;hl=en_GB"></param><param name="allowFullScreen" value="true"></param><param name="allowscriptaccess" value="always"></param><embed src="//www.youtube.com/v/'+prx.componentsHelper.getProp(item.youtubeid,'other')+'?version=3&amp;hl=en_GB" type="application/x-shockwave-flash" width="100%" height="100%" allowscriptaccess="always" allowfullscreen="true"></embed></object>';
            } else {
                cR += '<iframe id="'+_id+'-youtube"   width="100%" height="100%" src="https://www.youtube.com/embed/'+prx.componentsHelper.getProp(item.youtubeid,'other')+'?enablejsapi=1&html5=1&wmode=transparent&playsinline=1'+((prx.componentsHelper.getProp(item.autoplay,'boolean') && !prx.editor) ? '&autoplay=1' : '')+((prx.componentsHelper.getProp(item.loop,'boolean') && !prx.editor) ? '&loop=1&playlist='+prx.componentsHelper.getProp(item.youtubeid,'other') : '')+((!prx.componentsHelper.getProp(item.controls,'boolean')) ? '&controls=0' : '')+'" frameborder="0"  webkitAllowFullScreen mozallowfullscreen allowFullScreen allow="autoplay; fullscreen"></iframe>';
            }
            break;
        case 'vimeo':
            item.vimeoid = prx.componentsHelper.getProp(item.vimeoid,'video-url');
            //if(prx.editor && ($.browser.chrome || prx.phantomjs)) {
            //    cR += '<object width="100%" height="100%"><param name="allowfullscreen" value="true" /><param name="allowscriptaccess" value="always" /><param name="movie" value="https://vimeo.com/moogaloop.swf?clip_id='+prx.componentsHelper.getProp(item.vimeoid,'other')+'&amp;force_embed=1&amp;server=vimeo.com&amp;show_title=1&amp;show_byline=1&amp;show_portrait=1&amp;color=00adef&amp;fullscreen=1&amp;autoplay=0&amp;loop=0" '+ ((prx.componentsHelper.getProp(item.mute,'boolean')) ? '&amp;muted=1' : '') + '/><embed src="https://vimeo.com/moogaloop.swf?clip_id='+prx.componentsHelper.getProp(item.vimeoid,'other')+'&amp;force_embed=1&amp;server=vimeo.com&amp;show_title=1&amp;show_byline=1&amp;show_portrait=1&amp;color=00adef&amp;fullscreen=1&amp;autoplay=0&amp;loop=0"'+ ((prx.componentsHelper.getProp(item.mute,'boolean')) ? '&amp;muted=1' : '') + ' type="application/x-shockwave-flash" allowfullscreen="true" allowscriptaccess="always" width="100%" height="100%"></embed></object>';
            //} else {
                cR += '<iframe id="'+_id+'-vimeo"  width="100%" height="100%" src="https://player.vimeo.com/video/'+prx.componentsHelper.getProp(item.vimeoid,'other')+'?api=1&amp;player_id='+_id+'-vimeo&amp;title=0&amp;byline=0&amp;portrait=0&wmode=transparent'+((prx.componentsHelper.getProp(item.autoplay,'boolean') && !prx.editor) ? '&autoplay=1' : '')+((prx.componentsHelper.getProp(item.loop,'boolean') && !prx.editor) ? '&loop=1' : '') + ((prx.componentsHelper.getProp(item.mute,'boolean')) ? '&muted=1' : '') +'" frameborder="0" webkitAllowFullScreen mozallowfullscreen allowFullScreen allow="autoplay; fullscreen"></iframe>';
            //}
            break;
        }


        cR += prx.items.getComponentAppendDivs(item, containerid, symbol);
        cR += '</div>';
        return cR;
    }
    ,interactions: [prx.commonproperties.actions]
    ,mpactions: {
       	specialEvents: ['videoplay','videopause','videoend']
    }
    ,afterDisplay: function(item,containerid,symbol) {
        if(!prx.editor){
            var _id = (!containerid) ? item.id : containerid+'-'+item.id;

            if( item.videoType == 'html5' ) {
                var vid = $('#'+_id+'-html5video');
            }

            /* special events for youtube video in createVideo function */
            if(item.videoType=='youtube'){
                prx.youtube.createVideo(_id, item.mute);
            }

            /* special events for vimeo video in createVideo function */
            if(item.videoType=='vimeo'){
                prx.vimeo.createVideo(_id);
            }

            /* for special events (html5) */
            if( prx.componentsHelper.getProp(item.videoType,'other') == 'html5' ) {

                var video = $('#'+_id).find('video').get(0);

                video.onplay = function() {
                    $('#'+_id).trigger('videoplay');
		        };

                video.onpause = function() {
                    $('#'+_id).trigger('videopause');
		        };

                video.onended = function() {
                    $('#'+_id).trigger('videoend');
		        };

		        // if the video is in a scrollable container we must check if we it's playing so we can pause manually
                $('#'+_id).find('video').on('touchstart', function() {
                    $('#'+_id).parents().each( function(index, parent) {
                        Object.keys(prx.scrollable._scrollables).forEach( function(key) {
                            if ($(parent).attr('id') === key && !video.paused) {
                                video.pause();
                            }
                        });
                    });
                });
		    }

            $('#'+_id).hammer();
            prx.actions.disableFlashActionOnItemTap('#' + _id, '.flashactiontap-afterdisplay');
        }
    }
    //if not editor new youtube save to array prx.actions.players
    /*,afterDisplay: function(item,containerid,symbol) {
		if(prx.editor && $.browser.mozilla) {
			var _id = (!containerid) ? item.id : containerid+'-'+item.id;
			$('<div class="iframe-fix" style="position: absolute; top: 0; bottom: 0; left: 0; right: 0; pointer-events: auto;"><img style="pointer-events: auto;" src="data:image/gif;base64,R0lGODlhAQABAPAAAP///wAAACH5BAEAAAAALAAAAAABAAEAAAICRAEAOw==" width="100%" height="100%" /></div>')
				.appendTo('#'+_id)
				.click(function(){
					$('#'+_id).click();
					return false;
				})
		}
	}*/
    ,propertyGroups: [
        {
            caption: 'Video properties',
            properties: [[
		      {
		    	  	caption: 'Video type'
		    		,name: 'videoType'
		    		,type: 'select'
		    		,value: function(item,name){
		    	  		return item.videoType;
		      		}
		      		,values: [{ displayValue: 'Uploaded video', value: 'html5' }, { displayValue: 'YouTube video', value: 'youtube' }, { displayValue: 'Vimeo video', value: 'vimeo' }]
		      		,onChange: function(item){
		      			//if(item.videoType == 'youtube') {
		      			switch(item.videoType) {
		      			case 'youtube':
		  					$('#property-videoFile, #property-videoFileMP4, #property-videoFileOGG, #property-videoFileWEBM, #property-placeholder, #property-removePlaceholder, #property-preload, #property-vimeoid').hide();
                            $('#property-youtubeid, #property-controls').show();
                            break;
		      			case 'vimeo':
		      				$('#property-videoFile, #property-videoFileMP4, #property-videoFileOGG, #property-videoFileWEBM, #property-placeholder, #property-removePlaceholder, #property-preload, #property-youtubeid, #property-controls').hide();
                            $('#property-vimeoid').show();
		      				break;
		      			case 'html5':
		      			    if(prx.componentsHelper.getVideoFilesTotal(item) > 1){
                                $('#property-videoFileMP4, #property-videoFileOGG, #property-videoFileWEBM, #property-placeholder, #property-removePlaceholder, #property-preload, #property-controls').show();
                            }
		      			    else {
                                $('#property-videoFile').show();
                            }
                            $('#property-youtubeid, #property-vimeoid').hide();
                            break;
		      			}
		  				return false;
		      		}
		      		,changeProperty: {
                        rerender: true,
                        changeable: false
                    }
		      }],
            [{
                caption: 'YouTube Video ID'
                , name: 'youtubeid'
                , type: 'input'
                , value: function (item, name) {
                    return item.youtubeid;
                }
                , hiddenByDefault: function (item) {
                    return !(item.videoType == 'youtube');
                }
                , onChange: function (item) {

                    if (item.videoType == 'youtube') {
                        //var result = item.youtubeid.match("v=([a-zA-Z0-9]+)&?");
                        //Youtube regex: credits to http://stackoverflow.com/questions/2964678/jquery-youtube-url-validation-with-regex
                        var p = /^(?:https?:\/\/)?(?:www\.)?(?:youtu\.be\/|youtube\.com\/(?:embed\/|v\/|watch\?v=|watch\?.+&v=))((\w|-){11})(?:\S+)?$/;
                        var result = (item.youtubeid.match(p)) ? RegExp.$1 : false;
                        if (result) {
                            item.youtubeid = result;
                            return item;
                        }
                    }
                },
                changeProperty: {
                    	caption: 'YouTube Video ID',
                    rerender: true
                }
            }],
            [{
			    	caption: 'Vimeo Video ID'
			    	,name: 'vimeoid'
			    	,type: 'input'
			    	,value: function(item,name) {
			    		return item.vimeoid;
			    	}
			    	,hiddenByDefault: function(item) {
			    		return !(item.videoType == 'vimeo');
			    	}
                ,onChange: function (item) {

                    if (item.videoType == 'vimeo') {
                        //Credits to http://jsbin.com/asuqic/184/edit?js,output for regex
                        var vimeo_Reg = /https?:\/\/(?:www\.)?vimeo.com\/(?:channels\/(?:\w+\/)?|groups\/([^\/]*)\/videos\/|album\/(\d+)\/video\/|)(\d+)(?:$|\/|\?)/;
                        var match = item.vimeoid.match(vimeo_Reg);
                        if (match) {
                            item.vimeoid = match[3];
                            return item;
                        }
                    }
                },
                changeProperty: {
                    caption: 'Vimeo Video ID',
                    rerender: true
                }
			    }],
		      	[{
			    	caption: 'MP4 Video file'
			    	,name: 'videoFileMP4'
			    	,type: 'asset'
                ,displayValue: function(item,name) {
                    if(item.videoFileMP4.fileId == '') {
                        return 'No asset selected.';
                    }
                    return prx.utils.escapeHTML(item.videoFileMP4.name);
                }
                ,value: function(item,name) {
                    return JSON.stringify({
                        allow: 'video',
                        asset: item.videoFileMP4
                    });
                }
			    	,hiddenByDefault: function(item) {
                        if(item.videoType == 'html5' && prx.componentsHelper.getVideoFilesTotal(item) <= 1){
                            if(item.videoFileMP4.fileId != '') {
                                item.videoFile = item.videoFileMP4;
                                item.videoFileMP4 = {fileId: '', assetType: '', name: ''};
                            }
                            return true;
                        }
                        return !(item.videoType == 'html5');
			    	}
			    	,changeProperty: {
			    	caption: 'MP4 Video file',
                    rerender: true
                }
			    }],
			    [{
			    	caption: 'WebM Video file'
			    	,name: 'videoFileWEBM'
			    	,type: 'asset'
                ,displayValue: function(item,name) {
                    if(item.videoFileWEBM.fileId == '') {
                        return 'No asset selected.';
                    }
                    return prx.utils.escapeHTML(item.videoFileWEBM.name);
                }
                ,value: function(item,name) {
                    return JSON.stringify({
                        allow: 'video',
                        asset: item.videoFileWEBM
                    });
                }
                    ,hiddenByDefault: function(item) {
                        if(item.videoType == 'html5' && prx.componentsHelper.getVideoFilesTotal(item) <= 1){
                            if(item.videoFileWEBM.fileId != '') {
                                item.videoFile = item.videoFileWEBM;
                                item.videoFileWEBM = {fileId: '', assetType: '', name: ''};
                            }
                            return true;
                        }
                        return !(item.videoType == 'html5');
                    }
			    	,changeProperty: {
                    caption: 'WebM Video file',
                    rerender: true
                }
			    }],
			    [{
			    	caption: 'OGG Video file (.ogv)'
			    	,name: 'videoFileOGG'
			    	,type: 'asset'
                ,displayValue: function(item,name) {
                    if(item.videoFileOGG.fileId == '') {
                        return 'No asset selected.';
                    }
                    return prx.utils.escapeHTML(item.videoFileOGG.name);
                }
                ,value: function(item,name) {
                    return JSON.stringify({
                        allow: 'video',
                        asset: item.videoFileOGG
                    });
                },hiddenByDefault: function(item) {
                        if(item.videoType == 'html5' && prx.componentsHelper.getVideoFilesTotal(item) <= 1){
                            if(item.videoFileOGG.fileId != '') {
                                item.videoFile = item.videoFileOGG;
                                item.videoFileOGG = {fileId: '', assetType: '', name: ''};
                            }
                            return true;
                        }
                        return !(item.videoType == 'html5');
                    }
                    ,changeProperty: {
                    caption: 'OGG Video file',
                    rerender: true
                }
			    }],
                [{
                    caption: 'Video file'
                    ,hint: '.mp4 or .webm file types recommended'
                    ,name: 'videoFile'
                    ,type: 'asset'
                    ,displayValue: function(item,name) {
                        if(item.videoFile === undefined || item.videoFile.fileId == '') {
                            return 'No asset selected.';
                        }
                        return prx.utils.escapeHTML(item.videoFile.name);
                    }
                    ,value: function(item,name) {
                        return JSON.stringify({
                            allow: 'video',
                            asset: item.videoFile
                        });
                    }
                    ,hiddenByDefault: function(item) {
                        if(item.videoType == 'html5' && prx.componentsHelper.getVideoFilesTotal(item) > 1) {
                            return true;
                        }
                        return !(item.videoType == 'html5');
                    }
                    ,changeProperty: {
                        caption: 'Video file',
                        rerender: true
                    }
                }],
			     [{
			    	caption: 'Placeholder image'
			    	,name: 'placeholder'
			    	,type: 'combo-asset'
                ,displayValue: function(item,name) {
                    if(item.placeholder.fileId == '') {
                        return 'No asset selected.';
                    }
                    return prx.utils.escapeHTML(item.placeholder.name);
                }
                ,value: function(item,name) {
                    return JSON.stringify({
                        allow: 'image',
                        asset: item.placeholder
                    });
                }
			    	,hiddenByDefault: function(item) {
			    		return !(item.videoType == 'html5');
			    	}
			    	,changeProperty: {
                    caption: 'Placeholder image',
                    rerender: true
                }
			    }],
			    [
			    {
			    	caption: 'Preload video'
			    	,name: 'preload'
			    	,type: 'onoff'
			    	,value: function(item,name){
			    		return item.preload;
			    	}
			    	,hiddenByDefault: function(item) {
			    		return !(item.videoType == 'html5');
			    	}
			    	,changeProperty: {
                    	caption: 'Preload video',
                        rerender: true
                    }
			    },
			    {
			    	caption: 'Autoplay'
			    	,name: 'autoplay'
			    	,type: 'onoff'
			    	,value: function(item,name){
			    		return item.autoplay;
			    	}
                    ,help: 'Warning: May not work in some browsers/mobile devices due to browser restrictions'
                    ,hiddenByDefault: function(item,name) {
			    		if(typeof(item.autoplayoff) != 'undefined') {
                            return true;
                        }
                        return false;
                    }
			    	,changeProperty: {
                    	caption: 'Autoplay',
                        rerender: true
                    }
			    },
			    {
			    	caption: 'Loop'
			    	,name: 'loop'
			    	,type: 'onoff'
			    	,value: function(item,name){
			    		return item.loop;
			    	}
			    	,changeProperty: {
                    	caption: 'Loop',
                        rerender: true
                    }
			    },
			    {
			    	caption: 'Display controls'
			    	,name: 'controls'
			    	,type: 'onoff'
			    	,value: function(item,name){
			    		return item.controls;
			    	}
			    	,hiddenByDefault: function(item) {
			    		return (item.videoType == 'vimeo');
			    	}
			    	,changeProperty: {
                    	caption: 'Display controls',
                        rerender: true
                    }
			    },
                {
                    caption: 'Mute audio'
                    ,name: 'mute'
                    ,type: 'onoff'
                    ,value: function(item,name){
                        return item.mute;
                    }
                    //,help: 'Muting audio relaxes video playing restrictions'
                    ,changeProperty: {
                        caption: 'Mute audio',
                        rerender: true
                    }
                }]
			    ]
        }
    ]
};

prx.types.animationtarget = {
    name: 'animationtarget'
    ,onDisplay: function(item,containerid,symbol) {
        var _id = (!containerid) ? item.id : containerid+'-'+item.id;
        var _dims = prx.componentsHelper.getRealDims(item,symbol);
        var cR = '';
        cR += '<div id="' + _id + '" ' + prx.items.getComponentBaseAttributes(item, containerid, symbol)  + ' class="' + prx.items.getComponentBaseClasses(item, containerid, symbol) + ' box pos type-animationtarget '+((typeof(item.fixPositioning) != 'undefined' && prx.componentsHelper.getProp(item.fixPositioning,'boolean')) ? 'type-animatiotarget-fixed-positioning' : '')+'">';
        cR += '<style>';
        cR += prx.items.getComponentBaseStyle(item, containerid, symbol);
        cR += '</style>';
        cR += prx.items.getComponentPrependDivs(item, containerid, symbol);
        cR += '<div class="animationtarget-circle"></div>';
        cR += '<div class="animationtarget-horizontal"></div>';
        cR += '<div class="animationtarget-vertical"></div>';
        cR += prx.items.getComponentAppendDivs(item, containerid, symbol);
        cR += '</div>';
        return cR;
    }
};

//TYPE: tooltip
prx.types.tooltip = {
    name: 'tooltip'
    ,onDisplay: function(item,containerid,symbol) {
        var _id = (!containerid) ? item.id : containerid+'-'+item.id;
        var _height, _width, _pos, _margin;
        var _dims = prx.componentsHelper.getRealDims(item,symbol);

        var _ttBg = item.backgroundColor;
        if(_ttBg == 'none') { _ttBg = item.borderColor; }

        var _props = (typeof(prx.richtext)=='undefined') ? prx.componentsHelper.getProp(item.textProperties,'props-text') : '';

        switch (item.ttDirection) {
        case 'top':
        case 'bottom':
            _width = '100%';
            _height = parseInt(_dims.height) - (12*prx.componentsHelper.getScale(item.lib)) + 'px';
            _margin = (9*prx.componentsHelper.getScale(item.lib))+'px 0 '+(9*prx.componentsHelper.getScale(item.lib))+'px -'+(7*prx.componentsHelper.getScale(item.lib))+'px';
            _pos = 'width: '+(25*prx.componentsHelper.getScale(item.lib))+'px; height: '+( parseInt(12*prx.componentsHelper.getScale(item.lib)) + parseInt(prx.componentsHelper.getProp(item.borderWidth,'num-border-width')) )+'px; left: '+eval((_dims.width * prx.componentsHelper.getProp(item.ttPosition,'num-other')/100) - ((25*prx.componentsHelper.getScale(item.lib)) * prx.componentsHelper.getProp(item.ttPosition,'num-other')/100))+'px;';
            break;
        case 'left':
        case 'right':
            _height = '100%';
            _width = parseInt(_dims.width) - (12*prx.componentsHelper.getScale(item.lib)) + 'px';
            _margin = '-'+(7*prx.componentsHelper.getScale(item.lib))+'px '+(9*prx.componentsHelper.getScale(item.lib))+'px 0';
            _pos = 'width: '+( parseInt(12*prx.componentsHelper.getScale(item.lib)) + parseInt(prx.componentsHelper.getProp(item.borderWidth,'num-border-width')) )+'px; height: '+(25*prx.componentsHelper.getScale(item.lib))+'px; top: '+eval((_dims.height * prx.componentsHelper.getProp(item.ttPosition,'num-other')/100) - ((25*prx.componentsHelper.getScale(item.lib)) * prx.componentsHelper.getProp(item.ttPosition,'num-other')/100))+'px;';
            break;
        case 'none':
            _height = '100%';
            _width = '100%';
            '0';
            _pos = 'width: 0px; height: 0px';
            break;
        }


        var cR = '';
        cR += '<div id="' + _id + '" ' + prx.items.getComponentBaseAttributes(item, containerid, symbol)  + ' class="' + prx.items.getComponentBaseClasses(item, containerid, symbol) + ' box pos type-tooltip '+ (item.verticalTextAlign === 'center' ? 'verticalAlignCenter' : '') +'">';
        cR += '<style>';
        cR += prx.items.getComponentBaseStyle(item, containerid, symbol);
        cR += '#'+_id+' .tooltip-content-outer { padding-'+prx.componentsHelper.getProp(item.ttDirection,'other')+': '+(12*prx.componentsHelper.getScale(item.lib))+'px; }';
        cR += '#'+_id+' .tooltip-content { border-radius: '+prx.componentsHelper.getProp(item.borderRadius,'border-radius')+'; border: '+prx.componentsHelper.getProp(item.borderWidth,'num-border-width')+'px solid '+prx.componentsHelper.getProp(item.borderColor,'color-border')+'; background-color: '+prx.componentsHelper.getProp(item.backgroundColor,'color-background')+'; }';
        cR += '#'+_id+' .tooltip-text { '+_props+' '+prx.componentsHelper.getProp(item.textFont + '|' + item.textFontStyle,'font-family')+';' + prx.componentsHelper.getProp(item.textFontStyle,'font-style') + ';' + 'font-size: '+prx.componentsHelper.getProp(item.textSize,'num-text-size')+'px; text-align: '+prx.componentsHelper.getProp(item.textAlign,'align')+'; color: '+prx.componentsHelper.getProp(item.textColor,'color-text')+'; }';
        cR += '#'+_id+' .tooltip-outer { '+prx.componentsHelper.getProp(item.ttDirection,'other')+': 0; '+_pos+' }';
        cR += '#'+_id+' .tooltip { background: '+prx.componentsHelper.getProp(_ttBg,'color-background')+'; '+prx.componentsHelper.getProp(item.ttDirection,'other')+': 0; border: '+prx.componentsHelper.getProp(item.borderWidth,'num-border-width')+'px solid '+prx.componentsHelper.getProp(item.borderColor,'color-border')+'; margin: '+_margin+'; }';
        cR += '</style>';
        cR += prx.items.getComponentPrependDivs(item, containerid, symbol);
        cR += '<div class="tooltip-content-wrapper">';
        cR += '<div class="tooltip-content-outer">';
        cR += '<div class="tooltip-content liveUpdate-borderColor-border-color liveUpdate-backgroundColor-background-color changeProperty-backgroundColor changeProperty-borderColor changeProperty-borderRadius">';
        cR += '<div class="tooltip-text liveUpdate-textColor-color changeProperty-textColor changeProperty-textSize changeProperty-textFont changeProperty-textAlign"> <span data-editableproperty="text">'+prx.componentsHelper.getProp(item.text,'text-textarea')+'</span></div>';
        cR += '</div>';
        cR += '</div>';
        cR += '<div class="tooltip-outer">';
        cR += '<div class="tooltip liveUpdate-borderColor-border-color liveUpdate-backgroundColor-background-color changeProperty-backgroundColor changeProperty-borderColor"></div>';
        cR += '</div>';
        cR += prx.items.getComponentAppendDivs(item, containerid, symbol);
        cR += '</div>';

        cR += '</div>';

        return cR;
    }
    ,onResize: function(item,containerid,symbol) {
        var _id = (!containerid) ? item.id : containerid+'-'+item.id;

        var _height, _height2, _width;
        var _dims = prx.componentsHelper.getRealDims(item, symbol);

        switch (item.ttDirection) {
        case 'top':
        case 'bottom':
            _width = '100%';
            _height = eval(_dims.height - (12*prx.componentsHelper.getScale(item.lib))) + 'px';
            $('#' + _id + ' .tooltip-outer').first().css('left', eval((_dims.width * item.ttPosition/100) - ((25*prx.componentsHelper.getScale(item.lib)) * item.ttPosition/100)) + 'px');
            break;
        case 'left':
        case 'right':
            _height = '100%';
            _width = eval(_dims.width - (12*prx.componentsHelper.getScale(item.lib))) + 'px';
            $('#' + _id + ' .tooltip-outer').first().css('top', eval((_dims.height * item.ttPosition/100) - ((25*prx.componentsHelper.getScale(item.lib)) * item.ttPosition/100)) + 'px');
            break;
        case 'none':
            _height = '100%';
            _width = '100%';
            break;
        }
    }
    ,interactions: [
        prx.commonproperties.actions
    ]
    ,editableProperties: [
        {
            caption: 'Text',
            name: 'text',
            type: 'textarea',
            value: function(item,name) { return item.text; },
            changeProperty: {
                property: 'text',
                selector: '.tooltip-text',
                transitionable: false
            }
		 }
    ]
    ,propertyGroups: [
        {
            caption: 'Style',
            properties: [
                [
                    {
                        caption: 'Tooltip '
                        ,name: 'ttDirection'
                        ,type: 'select'
                        ,value: function(item,name) {
                            return item.ttDirection;
                        }
                        ,values: [{ value: 'top', displayValue: 'Top' }, { value: 'bottom', displayValue: 'Bottom' }, { value: 'left', displayValue: 'Left' }, { value: 'right', displayValue: 'Right' }, { value: 'none', displayValue: 'No tooltip' }]
                        ,changeProperty: {
                            caption: 'Tooltip direction',
                            rerender: true
                        }
                    },
			       	{
			  			caption: false
			  			,name: 'ttPosition'
			  			,type: 'combo-select'
			  			,value: function(item,name) {
			    			return item.ttPosition;
			    		}
                        ,values: { min: 0, max: 100, step: 5 }
                        ,changeProperty: {
                            caption: 'Tooltip position',
                            rerender: true
                        }
                    }
                ]
                ,[
                    prx.commonproperties.backgroundColorSolid
                ]
                ,[
                    {
                        caption: 'Border',
                        name: 'borderWidth',
                        proptype: 'border-width',
                        type: 'combo-select',
                        value: function(item,name) { return item.borderWidth; },
                        values: { min: 0, max: 10, step: 1 }
                        ,changeProperty: {
                            caption: 'BorderWidth',
                            rerender: true
                        }
                    }
			     	,prx.commonproperties.borderColor
			       	,prx.commonproperties.borderRadiusExpandable
			    ]
            ]
        },
        {
            caption: 'Text',
            properties: [
                [
                    prx.commonproperties.textFontRichText('font-family','text')
                ],
                [
                    prx.commonproperties.textFontStyleRichText(['font-weight', 'font-style'],'text', false, {propName: 'textFontStyle', caption: 'Text Font style', useAsDynProp: false}),
                    prx.commonproperties.textSizeRichText('font-size','text')
                ],
                [
                    prx.commonproperties.textPropertiesRichText(['font-weight','font-style','text-decoration'],'text', false, {propName: 'textProperties', caption: 'Underline', useAsDynProp: false})
                    ,prx.commonproperties.textColorRichText('color','text')
                ],
                [
                    prx.commonproperties.textAlign
                ]
            ]
        }
    ]

};

//TYPE: BASIC_BUTTON_BG
prx.types.basic_button_bg = {
    name: 'basic_button_bg'
    ,onDisplay: function(item,containerid,symbol) {
        var _id = (!containerid) ? item.id : containerid+'-'+item.id;

        var _props = (typeof(prx.richtext)=='undefined') ? prx.componentsHelper.getProp(item.textProperties,'props-text') : '';
        var text = '<span class="basic-button-text liveUpdate-textColor-color changeProperty-textColor changeProperty-textFont changeProperty-textFontStyle changeProperty-textSize"><span data-editableproperty="text">'+prx.componentsHelper.getProp(item.text,'text-textarea')+'</span></span>';
        var icon = '';
        if(prx.componentsHelper.getProp(item.iconpos,'icon-position') != 'none' && prx.componentsHelper.getProp(item.img.fileId,'other') != '') {
            icon = '<img class="basic-button-icon" src="'+prx.componentsHelper.getProp(item.img,'asset')+'">';
        }

        var cR = '';
        cR += '<div id="' + _id + '" ' + prx.items.getComponentBaseAttributes(item, containerid, symbol)  + ' class="' + prx.items.getComponentBaseClasses(item, containerid, symbol) + ' box pos type-basic-button '+(item.name == 'basic_button_texticon' || item.name == 'basic_button_text' || item.name == 'basic_button_icon' ? 'generic' : '')+'">';

        cR += '<style>';
        cR += prx.items.getComponentBaseStyle(item, containerid, symbol);
        cR += '#'+_id+' .basic-button-inner { '+prx.css.flexJustifyContent(prx.componentsHelper.getProp(item.textAlign,'align'))+' text-align: '+prx.componentsHelper.getProp(item.textAlign,'align')+'; background: '+prx.componentsHelper.getProp(item.backgroundColor,'color-background')+'; border-width: '+prx.componentsHelper.getProp(item.borderWidth,'border-width')+'; border-style: solid; border-color: '+prx.componentsHelper.getProp(item.borderColor,'color-border')+'; border-radius: '+prx.componentsHelper.getProp(item.borderRadius,'border-radius')+'; }';
        cR += '#'+_id+' .basic-button-text { '+(item.iconpos != 'none'? '' : 'width: 100%;')+' '+prx.componentsHelper.getProp(item.textFont + '|' + item.textFontStyle,'font-family')+prx.componentsHelper.getProp(item.textFontStyle,'font-style')+' color: '+prx.componentsHelper.getProp(item.textColor,'color-text')+'; font-size: '+prx.componentsHelper.getProp(item.textSize,'num-text-size')+'px; '+_props+' }';
        if(typeof(item.iconSize) != 'undefined') {
            cR += '#' + _id + ' .basic-button-icon { height: ' + prx.componentsHelper.getProp(item.iconSize,'icon-size') * 20 + '%; }';
        }
        cR += '</style>';
        cR += prx.items.getComponentPrependDivs(item, containerid, symbol);

        cR += '<div class="basic-button-inner basic-button-icon-'+prx.componentsHelper.getProp(item.iconpos,'icon-position')+' liveUpdate-backgroundColor-background liveUpdate-borderColor-border-color changeProperty-backgroundColor changeProperty-borderColor changeProperty-borderWidth changeProperty-borderRadius changeProperty-textAlign">';

        switch(item.iconpos) {
        case 'notext':
            cR += icon;
            break;
        case 'left':
            cR += icon + text;
            break;
        case 'right':
            cR += text + icon;
            break;
        default:
            cR += text;
        }

        cR += '</div>';
        cR += prx.items.getComponentAppendDivs(item, containerid, symbol);
        cR += '</div>';
        return cR;
    },
    interactions: [ prx.commonproperties.actions ],
    editableProperties: [{
        caption: 'Text',
        name: 'text',
        type: 'textarea',
        value: function(item,name) { return item.text; },
        changeProperty: {
            caption: 'Text',
            selector: '.basic-button-text',
            property: 'text',
            transitionable: false
        }
    }],
    propertyGroups: [
        {
            caption: 'Style',
            properties: [
                [
                    {
                        caption: 'Background',
                        name: 'backgroundColor',
                        proptype: 'background',
                        type: 'gradients-colorpicker',
                        value: function(item, name) {
                            return typeof item.backgroundColor == 'undefined' ? '' : item.backgroundColor;
                        },
                        liveUpdate: 'background',
                        changeProperty: {
                            caption: 'Background color',
                            selector: '.changeProperty-backgroundColor',
                            property: 'background',
                            transitionable: true
                        }
                    }
                ],
                [
                    {
                        caption: 'Border',
                        name: 'borderWidth',
                        proptype: 'border-width',
                        type: 'combo-select',
                        value: function(item,name) { return item.borderWidth; },
                        values: { min: 0, max: 20, step: 1 } ,
                        expandableType: 'borderWidth',
                        expandedValues: ['t', 'r', 'l', 'b'],
                        changeProperty: {
                            caption: 'Border Width',
                            selector: '.changeProperty-borderWidth',
                            property: 'border-width',
                            transitionable: true
                        }
                    }
                    ,{
                        caption: false,
                        name: 'borderColor',
                        proptype: 'border-color',
                        type: 'colorpicker',
                        value: function(item,name) { return item.borderColor; },
                        liveUpdate: 'border-color' ,
                        changeProperty: {
                            caption: 'Border Color',
                            selector: '.changeProperty-borderColor',
                            property: 'border-top-color,border-bottom-color,border-left-color,border-right-color',
                            transitionable: true
                        }
                    }
                	,{
                        caption: '<span class="icon icon-border-radius" title="Border radius"></span>',
                        name: 'borderRadius',
                        proptype: 'border-radius',
                        type: 'combo-select',
                        value: function(item,name) {
                            return item.borderRadius;
                        },
                        values: {min: 0, max: 20, step: 1},
                        expandableType: 'borderRadius',
                        expandedValues: ['tl', 'tr', 'bl', 'br'],
                        changeProperty: {
                            caption: 'Border Radius',
                            selector: '.changeProperty-borderRadius',
                            property: 'border-radius',
                            transitionable: true
                        }
                    }
                ]
            ]
        },
        {
            caption: 'Text',
            properties: [
                [
                    Object.assign(prx.commonproperties.textFont, { hiddenByDefault: function(item) {
                            return (item.iconpos === 'notext');
                        },
                    })
                ],
                [
                    Object.assign(prx.commonproperties.textFontStyle, { hiddenByDefault: function(item) {
                            return (item.iconpos === 'notext');
                        },
                    }),
                    Object.assign(prx.commonproperties.textSize, { hiddenByDefault: function(item) {
                            return (item.iconpos === 'notext');
                        },
                    }),
                ],
                [
                    Object.assign(prx.commonproperties.textPropertiesRichText(['font-weight','font-style','text-decoration'],'text', false, {propName: 'textProperties', caption: 'Underline', useAsDynProp: false}), { hiddenByDefault: function(item) {
                            return (item.iconpos === 'notext');
                        },
                    }),
                    {
                        caption: { label: 'Color', class: 'text-properties-label text-color-label' },
                        name: 'textColor',
                        proptype: 'font-color',
                        type: 'solid-colorpicker',
                        relatedEditableProperties: 'text',
                        relatedCSSProperties: 'color',
                        pffSettings: prx.commonproperties.pffSettingsColor,
                        value: function(item,name) {
                            if(typeof(item.textColor) == 'undefined') { item.textColor = '#2E2E2E'; }
                            return item.textColor;
                        },
                        liveUpdate: 'color'
                        ,changeProperty: {
                            caption: ' Text color',
                            selector: '.changeProperty-textColor',
                            property: 'color',
                            transitionable: true
                        }
                        ,hiddenByDefault: function(item) {
                            return (item.iconpos === 'notext');
                        }
                    }
                ],
                [
                    {
                        caption: { label: 'Alignment', class: 'text-properties-label text-alignment-label' },
                        proptype: 'text-align',
                        name: 'textAlign',
                        type: 'radio',
                        pffSettings: prx.commonproperties.pffSettingsTextAlignment,
                        value: function(item, name) {
                            return typeof item.textAlign == 'undefined' ? '' : item.textAlign;
                        },
                        values: [
                            { value: 'left', displayValue: '', icon: 'align-left'},
                            { value: 'center', displayValue: '', icon: 'align-center'},
                            { value: 'right', displayValue: '', icon: 'align-right'},
                            { value: 'justify', displayValue: '', icon: 'align-justify'}
                        ],
                        changeProperty: {
                            caption: 'Text align',
                            rerender: true
                        }
                        ,hiddenByDefault: function(item) {
                            return (item.iconpos === 'notext');
                        }
                    }
                ]
            ]
        },
        {
            caption: 'Icon',
            properties: [[
                {
                    caption: 'Position'
                    ,name: 'iconpos'
                    ,type: 'select'
                    ,value: function(item,name) {
                        return item.iconpos;
                    }
                    ,values: [{value: 'none',displayValue: 'No icon'},{value: 'left',displayValue: 'Left'},{value: 'right',displayValue: 'Right'},{value: 'notext',displayValue: 'Icon only (no text)'}]
                    ,onChange: function(item){
                        if(item.iconpos == 'none') {
                            $('#property-img, #property-iconSize').hide();
                        } else {
                            $('#property-img, #property-iconSize').show();
                        }
                        if(item.iconpos == 'notext') {
                            $('#property-textSize, #property-textFont, #property-textColor, #property-textProperties, #property-textAlign, #property-textFontStyle').hide();
                        } else {
                            $('#property-textSize, #property-textFont, #property-textColor, #property-textProperties, #property-textAlign, #property-textFontStyle').show();
                        }
                        return false;
                    }
                    ,changeProperty: {
                        caption: 'Icon position',
                        rerender: true
                    }
                },
                {
                    caption: 'Size'
                    ,name: 'iconSize'
                    ,proptype: 'icon-size'
                    ,type: 'select'
                    ,value: function(item,name) {
                        return item.iconSize;
                    }
                    ,values: [{ value: '1', displayValue: 'Very small'}, { value: '2', displayValue: 'Small'}, { value: '3', displayValue: 'Normal'}, { value: '4', displayValue: 'Large'}, { value: '5', displayValue: 'Full'}]
                    ,changeProperty: {
                        caption: 'Icon size',
                        rerender: true,
                        changeable: false
                    }
                    ,hiddenByDefault: function(item,name,index){
                        return (item.iconpos == 'none');
                    }
                }],
            [
                {
                    caption: false
                    ,name: 'img'
                    ,type: 'combo-asset'
                    ,displayValue: function(item,name,index) {
                        if(item.img.fileId == '') {
                            return 'No icon selected';
                        }
                        return item.img.name;
                    }
                    ,value: function(item,name,index) {
                        return JSON.stringify({
                            allow: 'image',
                            asset: item.img
                        });
                    }
                    ,hiddenByDefault: function(item,name,index){
                        return (item.iconpos == 'none');
                    }
                    ,changeProperty: {
                        caption: 'Icon',
                        rerender: true
                    }
                }
            ]
            ]
        }
    ]
};

//TYPE: SLIDER
prx.types.basic_slider = {
    name: 'basic_slider'
    ,onDisplay: function(item,containerid,symbol) {
        var _id = (!containerid) ? item.id : containerid+'-'+item.id;
        var _dims = prx.componentsHelper.getRealDims(item,symbol);
        if(typeof item.barBorderRadius === 'undefined'){
            item.barBorderRadius = 0;
        }
        if(typeof item.borderColor === 'undefined'){
            item.borderColor = '';
        }
        if(typeof item.sliderBorderWidth === 'undefined'){
            item.sliderBorderWidth = 0;
        }
        var cR = '';
        cR += '<div id="' + _id + '" ' + prx.items.getComponentBaseAttributes(item, containerid, symbol)  + ' class="' + prx.items.getComponentBaseClasses(item, containerid, symbol) + ' box pos '+(item.name === 'ios7_slider'? 'type-ios7-slider' : 'type-basic-slider')+'">';

        cR += '<style>';
        cR += prx.items.getComponentBaseStyle(item, containerid, symbol);
        cR += '#'+_id+' .slider-bar { height: '+prx.componentsHelper.getProp(item.barThickness,'num-other')+'px; ' + prx.gradients.getBgCss(item.barColor) + '; margin-top: -'+(prx.componentsHelper.getProp(item.barThickness,'num-other')/2)+'px; border-radius: '+prx.componentsHelper.getProp(item.barBorderRadius,'border-radius')+' }'
        cR += '#'+_id+' .slider-bar-filled { width: '+prx.componentsHelper.getProp(item.sliderPosition,'num-percentage')+'%; ' + prx.gradients.getBgCss(item.fillBarColor) + '; border-radius: '+prx.componentsHelper.getProp(item.barBorderRadius,'border-radius')+' }'
        cR += '#'+_id+' .slider-button { background: '+prx.componentsHelper.getProp(item.sliderColor,'color-switch')+'; border: '+prx.componentsHelper.getProp(item.sliderBorderWidth,'num-border-width')+'px solid '+prx.componentsHelper.getProp(item.borderColor,'color-border')+'; border-radius: '+prx.componentsHelper.getProp(item.sliderBorderRadius,'border-radius')+'; width: '+_dims.height+'px; box-sizing: border-box; margin-left: -'+(_dims.height/2)+'px; margin-right: -'+(_dims.height/2)+'px; }';
        cR += '</style>';
        cR += prx.items.getComponentPrependDivs(item, containerid, symbol);

        cR += '<div class="slider-bar bar liveUpdate-barColor-background-color">';
        cR += '<div class="slider-bar-filled bar liveUpdate-fillBarColor-background-color"></div>';
        cR += '</div>';
        cR += '<span class="slider-button liveUpdate-sliderColor-background-color liveUpdate-borderColor-border-color changeProperty-borderColor"></span>';
        cR += prx.items.getComponentAppendDivs(item, containerid, symbol);
        cR += '</div>';
        return cR;
    }
    ,onResize: function(item,containerid,symbol) {
        var _id = (!containerid) ? item.id : containerid+'-'+item.id;
        var _dims = prx.componentsHelper.getRealDims(item,symbol);

        $('#'+_id+' .slider-button').css({
            width: (_dims.height - 2 * item.sliderBorderWidth)+'px',
            marginLeft: '-'+(_dims.height/2)+'px',
            marginRight: '-'+(_dims.height/2)+'px'
        });

        if(prx.editor){
            $('#'+_id).find('.slider-button').css({
                left: ((_dims.width)*(prx.componentsHelper.getProp(item.sliderPosition,'num-percentage')*0.01)) + 'px'
            });
        }

    }
    ,afterDisplay: function(item,containerid,symbol) {
        var _id = (!containerid) ? item.id : containerid+'-'+item.id;

        var _dims = prx.componentsHelper.getRealDims(item,symbol);

        if(!prx.editor) {

            prx.draggable._draggables['#'+_id+' .slider-button'] = Draggable.create('#'+_id+' .slider-button', {
                type: 'x',
                bounds: { left: -(_dims.height/2), width: parseInt(_dims.width) + parseInt(_dims.height), top: 0, height: 0 },
                onDragStart: function(){
                    $(this.target).parents('.box').each(function() {
                        if(typeof(prx.scrollable._scrollables[this.id + '-inner']) != 'undefined') {
                            prx.scrollable._scrollables[this.id + '-inner'].disable();
                        }
                    });

                    var pos = this.x - this.minX;
                    var width = this.maxX - this.minX;

                    var progress = Math.ceil((pos / width)*100);

                    if(typeof(prx.variables._triggerData['#'+_id]) == 'undefined') { prx.variables._triggerData['#'+_id] = {}; }
                    prx.variables._triggerData['#'+_id]['sliderdragstart'] = { value: progress };
                    $('#'+_id).trigger('sliderdragstart');

                    // hack instead of ondrag because greensock on drag only triggers if the item has actually moved
                    // so if you are at 0 or 100 it only triggers once, which results in the value not always being updated
                    // because of the "actionIsRunning" check
                    $(document).on('mousemove.prx-sliderdrag touchmove.prx-sliderdrag', function(){
                        var pos = $('#' + _id + ' .slider-button').position().left;
                        var width = $('#' + _id).width();

                        $('#'+_id+' .slider-bar-filled').width(pos);
                        var progress = Math.ceil((pos / width)*100);

                        if(typeof(prx.variables._triggerData['#'+_id]) == 'undefined') { prx.variables._triggerData['#'+_id] = {}; }
                        prx.variables._triggerData['#'+_id]['sliderdrag'] = { value: progress };
                        $('#'+_id).trigger('sliderdrag');
                    });
                },
                onDragEnd: function(){

                    $(document).off('mousemove.prx-sliderdrag touchmove.prx-sliderdrag');

                    $(this.target).parents('.box').each(function() {
                        if(typeof(prx.scrollable._scrollables[this.id + '-inner']) != 'undefined') {
                            //alert('x')
                            prx.scrollable._scrollables[this.id + '-inner'].enable();
                        }
                    });

                    var pos = this.x - this.minX;
                    var width = this.maxX - this.minX;

                    var progress = Math.ceil((pos / width)*100);

                    prx.currentPlayerStatus.helper.set(_id, 'sliderPosition', progress);

                    if(typeof(prx.variables._triggerData['#'+_id]) == 'undefined') { prx.variables._triggerData['#'+_id] = {}; }
                    prx.variables._triggerData['#'+_id]['sliderdrag'] = { value: progress };
                    prx.variables._triggerData['#'+_id]['sliderdragend'] = { value: progress };
                    $('#'+_id).trigger('sliderdrag');
                    $('#'+_id).trigger('sliderdragend');
                }
            });

            TweenLite.set('#'+_id+' .slider-button',{x:((_dims.width)*(item.sliderPosition*0.01))});

            $('#'+_id+' .slider-bar').hammer().on('mousedown touchstart', function(e){

                var _pageXY = prx.utils.getEventPageXY(e);
                var _pos = _pageXY.x - $(this).offset().left;

                var progress = Math.ceil((_pos / $('#'+_id).width())*100);

                prx.currentPlayerStatus.helper.set(_id, 'sliderPosition', progress);

                TweenLite.to($('#'+_id+' .slider-bar-filled'), 1, {width:_pos});
                TweenLite.to($('#'+_id+' .slider-button'), 1, {x:_pos});

                //$(this).find('.slider-bar-filled').width(_pos);
                //$(this).siblings('.slider-button').css({ left: _pos + 'px' });

                prx.draggable._draggables['#'+_id+' .slider-button'][0].update();

                if(typeof(prx.variables._triggerData['#'+_id]) == 'undefined') { prx.variables._triggerData['#'+_id] = {}; }

                prx.variables._triggerData['#'+_id]['sliderdragstart'] = { value: progress };
                prx.variables._triggerData['#'+_id]['sliderdrag'] = { value: progress };
                prx.variables._triggerData['#'+_id]['sliderdragend'] = { value: progress };
                $('#'+_id).trigger('sliderdragstart');
                $('#'+_id).trigger('sliderdrag');
                $('#'+_id).trigger('sliderdragend');
            });

            prx.actions.disableFlashActionOnItemTap('#' + _id + ' .slider-bar', '.flashactiontap-afterdisplay');
        }
        else {
            $('#'+_id).find(' .slider-button').css({
                left: ((_dims.width)*(item.sliderPosition*0.01)) + 'px'
            });
        }
    }
    ,interactions: [ prx.commonproperties.actions ]
    ,mpactions: {
        specialEvents: ['sliderdragstart','sliderdrag','sliderdragend']
    }
    ,propertyGroups:	[

        {
            caption: 'Bar',
            properties: [
                [
                    {
                        caption: 'Thickness'
                        ,name: 'barThickness'
                        ,proptype: 'bar-thickness'
                        ,type: 'combo-select'
                        ,value: function(item,name) {
                            return item.barThickness;
                        }
                        ,values: { min: 2, max: 20, step: 2 }
                        ,changeProperty: {
                            caption: 'Bar thickness',
                            rerender: true,
                        }
                    }
                ],
                [
                    {
                        caption: 'Color'
                        ,name: 'barColor'
                        ,proptype: 'background-color'
                        ,type: 'gradients-colorpicker'
                        ,value: function(item,name) {
                            return item.barColor;
                        }
                        ,values: prx.comps.basicColors //ios7Colors
                        ,liveUpdate:'background-color'
                        ,changeProperty: {
                            caption: 'Bar color',
                            property: 'background-color',
                            selector: '.slider-bar',
                            transitionable: true
                        }
                    },
                    {
                        caption: 'Fill Color'
                        ,name: 'fillBarColor'
                        ,proptype: 'background-color'
                        ,type: 'gradients-colorpicker'
                        ,value: function(item,name) {
                            return item.fillBarColor;
                        }
                        ,values: prx.comps.basicColors //ios7Colors
                        ,liveUpdate:'background-color'
                        ,changeProperty: {
                            caption: 'Fill Color',
                            property: 'background-color',
                            selector: '.slider-bar-filled',
                            transitionable: true
                        }
                    }
                ],
                [{
                    caption: '<span class="icon icon-border-radius" title="Border radius"></span>'
                    ,name: 'barBorderRadius'
                    ,proptype: 'border-radius'
                    ,type: 'combo-select'
                    ,value: function(item,name) {
                        return item.barBorderRadius;
                    }
                    ,values: { min: 0, max: 20, step: 2 },
                    expandableType: 'borderRadius',
                    expandedValues: ['tl', 'tr', 'bl', 'br'],
                    changeProperty: {
                        caption: 'Bar border radius',
                        property: 'border-radius',
                        selector: '.slider-bar',
                        transitionable: true
                    }
                }]
            ]
        },
        {
            caption: 'Slider',
            properties: [
                [
                    {
                        caption: 'Color'
                        ,name: 'sliderColor'
                        ,proptype: 'background-color'
                        ,type: 'gradients-colorpicker'
                        ,value: function(item,name) {
                            return item.sliderColor;
                        }
                        ,values: prx.comps.basicColors //ios7Colors
                        ,liveUpdate:'background-color'
                        ,changeProperty: {
                            caption: 'Slider color',
                            property: 'background-color',
                            selector: '.slider-button',
                            transitionable: true
                        }
                    }
                ],
                [{
                    caption: 'Border',
                    name: 'sliderBorderWidth',
                    proptype: 'border-width',
                    type: 'combo-select',
                    value: function(item, name) {
                        return typeof item.sliderBorderWidth == 'undefined' ? '' : item.sliderBorderWidth;
                    },
                    values: {min: 0, max: 20, step: 1},
                    changeProperty: {
                        caption: 'Slider border width',
                        property: 'border-width',
                        selector: '.slider-button',
                        transitionable: true,
                    }
                }
                ,prx.commonproperties.borderColor
                ,{
                    caption: '<span class="icon icon-border-radius" title="Border radius"></span>'
                    ,name: 'sliderBorderRadius'
                    ,proptype: 'border-radius'
                    ,type: 'combo-select'
                    ,value: function(item,name) {
                        return item.sliderBorderRadius;
                    }
                    ,values: { min: 0, max: 20, step: 2 },
                    expandableType: 'borderRadius',
                    expandedValues: ['tl', 'tr', 'bl', 'br'],
                    changeProperty: {
                        caption: 'Slider border radius',
                        property: 'border-radius',
                        selector: '.slider-button',
                        transitionable: true
                    }
                }],
                [
                    {
                        caption: 'Original position (%)'
                        ,name: 'sliderPosition'
                        ,type: 'slider-input'
                        ,value: function(item,name) {
                            return item.sliderPosition;
                        }
                        ,values: { min: 0, max: 100, step: 1 }
                        ,changeProperty: {
                            caption: 'Slider position',
                            rerender: true
                        }
                    }
                ],

            ]
        }
    ]
};

//TYPE: BODYMOVIN

prx.vectorCreated =  [];
prx.types.vectoranimation = {
    name: 'vectoranimation'
    ,onCreate: function (item, containerid, symbol) {

        var _id = (!containerid) ? item.id : containerid + '-' + item.id;
        prx.vectorCreated.push('#' + _id + '-bodymovin');
    }
    ,afterDisplayWaitForCB: true
    ,afterDisplay: function(item,containerid,symbol, callback) {

        var _id = (!containerid) ? item.id : containerid + '-' + item.id;
        var $div = $('#' + _id + '-bodymovin');
        var _isUploading = $div.hasClass('uploading');
        var _animationData = _isUploading ? prx.componentsHelper.getProp(item.imgSrc,'asset') : false;
        var _onCreate = prx.vectorCreated.indexOf('#' + _id + '-bodymovin') > -1;
        prx.vectorCreated = [];

        if(prx.editor){
            if (prx.componentsHelper.getProp(item.imgSrc.fileId, 'other') != '') {
                if (!$div.hasClass('missing') && !_isUploading) {
                    var anim = lottie.registerAnimation($div[0]);

                    if (anim !== null) {
                        anim.addEventListener('DOMLoaded', function () {
                            $div.find('#playVectorAnimationprop').addClass('disabled');
                            anim.goToAndStop(1, true);

                            if (_onCreate) {
                                prx.bodymovinAnimation.playbackConsole({
                                    playbackType: 'play', target: {search: false, dataName: _id},
                                    onCompleteEvent: function event() {
                                        $('#' + _id).find('#playVectorAnimationprop').removeClass('disabled');
                                        this.goToAndStop(1, true);
                                    },
                                    // onDomLoadedEvent: function event() {
                                    //    $div.find('#playVectorAnimationprop').addClass('pause');
                                    //    if (callback != null) {
                                    //        callback();
                                    //    }
                                    // },
                                    // onDomLoadedFailedEvent: function event() {
                                    //    if (callback != null) {
                                    //        callback();
                                    //    }
                                    // }
                                });
                            }

                            if (callback != null) {
                                callback();
                            }
                        });
                        anim.addEventListener('data_failed', function () {
                            if (callback != null) {
                                callback();
                            }
                        });
                    }

                }
                else if (_isUploading && _animationData) {


                	var animeData = {
                        container: document.getElementById(_id + '-bodymovin'), // the dom element
                        renderer: 'svg',
                        loop: false,
                        autoplay: _onCreate,
                        name: $div.attr('data-name'),
                        animationData: _animationData
                    };

                    var anim = prx.bodymovinAnimation.loadAnimation(animeData);
                    anim.addEventListener('DOMLoaded', function () {

                        anim.goToAndStop(1, true);

                        if (callback != null) {
                            callback();
                        }
                    });
                    anim.addEventListener('data_failed', function () {
                        if (callback != null) {
                            callback();
                        }
                    });
                    anim.addEventListener('complete', function () {
                        $('#' + _id).find('#playVectorAnimationprop').removeClass('disabled');
                        anim.goToAndStop(1,true);
                    });

                } else {
                    if (callback != null) {
                        callback();
                    }
                }
            } else {
                if (callback != null) {
                    callback();
                }
            }
            // return anim;
        }

    }
    , onResize: function(item, containerid, symbol) {
    	// disable autoresize
     	prx.componentsHelper.setAutoresizeValue(item, containerid, symbol, false);
    }
    , onDisplay: function (item, containerid, symbol) {
        var _id = (!containerid) ? item.id : containerid + '-' + item.id;
        var missing = prx.url.static + '/images/editor/missing.png';
        var isUploading = false;
        var uploadingClass = ' uploading';

        if (typeof(item.overlay) == 'undefined') {
            item.overlay = false;
        }

        var assetUrl = prx.componentsHelper.getProp(item.imgSrc,'asset');

        if(typeof(assetUrl) === 'object') { //uploading
            isUploading = true;
        } else {
            var _prgss = prx.editor ? prx.prgss : '';
            var _crossDomainURL = assetUrl;
            if(prx.editor) {
                if (assetUrl.indexOf('?') === -1) {
                    _crossDomainURL += '?prgss=' + _prgss;
                } else {
                    _crossDomainURL += '&prgss=' + _prgss;
                }
            }
        }

        if (prx.editor && !isUploading && prx.componentsHelper.getProp(item.imgSrc.fileId,'other') != '') {

        	var toCheck = '' + _crossDomainURL;
            var request = $.getJSON(toCheck);

            request.success(function(result) {
                // console.log(result);
                if(typeof(result) == 'object') {
                    if (prx.vectorAnimationUtils.isValidBodyMovin(result)) {
                        validRequest();
                        return;
                    }
                }

                if(typeof(result) == 'string' && result.length > 0) { //windows and chrome...
                    try {
                        var animData = JSON.parse(result);
                        if (prx.vectorAnimationUtils.isValidBodyMovin(animData)) {
                            validRequest();
                            return;
                        }
                    } catch (e) {
                        invalidRequest();
                    }
                    invalidRequest();
                }
                invalidRequest();
            });

            request.error(function(jqXHR, textStatus, errorThrown) {
                // if (textStatus == 'timeout')
                //     console.log('The server is not responding');
                //
                // if (textStatus == 'error')
                //     ag(errorThrown);

                invalidRequest();
            });

            function validRequest(){
                $('#' + _id + '-vector-wrapper').removeClass('missing');
            }

            function invalidRequest(){
                $('#' + _id + '-bodymovin').css('background-image', missing);

                var title = $('<div>' + prx.utils.escapeHTML(item.caption) + '</div>');
                $('#' + _id + '-vector-wrapper').addClass('missing');
                $('#' + _id + '-vector-wrapper').find('div').remove();
                $('#' + _id + '-vector-wrapper').find('img').remove();
                $('#' + _id + '-vector-wrapper').append('<img src="'+missing+'">');
                $('#' + _id + '-vector-wrapper').prepend($('<div></div>'));
                $('#' + _id + '-vector-wrapper').append(title);
                $('#' + _id + '-vector-wrapper').append($('<div></div>'));
            }

        }

        var missingClass = '';
        if (assetUrl == missing || prx.componentsHelper.getProp(item.imgSrc.fileId,'other') == '') {
            missingClass = ' missing';
        }

        if( typeof(item.borderColor) == 'undefined' ) item.borderColor = '#000000';
        if( typeof(item.borderWidth) == 'undefined' ) item.borderWidth = 0;
        if( typeof(item.borderRadius) == 'undefined' ) item.borderRadius = 0;
        if( typeof(item.borderPos) == 'undefined' ) item.borderPos = 'inside';
        if( typeof(item.borderStyle) == 'undefined') { item.borderStyle = 'solid'; }


        var cR = '<div id="' + _id + '"';
        cR +=  ' ' + prx.items.getComponentBaseAttributes(item, containerid, symbol)  + ' class="' + prx.items.getComponentBaseClasses(item, containerid, symbol) + ' box pos type-vectoranimation ' + (prx.componentsHelper.getProp(item.propagateEvents,'boolean') ? 'pointer-events-none' : '') + ((prx.componentsHelper.getProp(item.repeat,'boolean')) ? ' type-vectoranimation-repeater' : '') + '" ' + ((prx.componentsHelper.getProp(item.overlay,'boolean')) ? 'data-mpoverlay="1"' : '') + '>';

        cR += '<style>';
        cR += prx.items.getComponentBaseStyle(item, containerid, symbol);
        cR += '</style>';

        cR += prx.items.getComponentPrependDivs(item, containerid, symbol, '');

        cR += '<div class="vector-inner borderPos-'+prx.componentsHelper.getProp(item.borderPos,'other')+'">';

        cR += '<div id="' + _id + '-vector-wrapper" class="type-vectoranimation-wrapper liveUpdate-borderColor-border-color changeProperty-borderColor changeProperty-borderRadius changeProperty-borderWidth ' + missingClass + '">';

        cR += '<div></div>';

        var tempH = item.height, tempW = item.width;

     	if(prx.editor){
            if(!isUploading){
                if(prx.componentsHelper.getProp(item.imgSrc.fileId,'other') == '') {
                    cR += '<img id="' + _id + '-bodymovin" src="'+missing+'" class="bodymovin" data-fileid="'+item.imgSrc.fileId+'" style="width:'+tempW+'px;height:'+tempH+'px"/>';
                }
                else {
                    cR += '<div id="' + _id + '-bodymovin" class="bodymovin" data-fileid="'+item.imgSrc.fileId+'" data-name="' + _id + '" data-speed = "'+prx.componentsHelper.getProp(item.playbackSpeed,'num-other')+'" data-anim-loop = "false" data-anim-autoplay="false" data-bm-path="' + toCheck + '"  data-bm-renderer="svg"></div>';
                }
            } else { // uploading
                cR += '<div id="' + _id + '-bodymovin" class="bodymovin ' + uploadingClass + '" data-fileid="'+item.imgSrc.fileId+'" data-name="' + _id + '"  data-speed = "'+prx.componentsHelper.getProp(item.playbackSpeed,'num-other')+'" data-anim-loop = "false" data-anim-autoplay="false" data-bm-renderer="svg"></div>';
            }

            if (assetUrl == missing || prx.componentsHelper.getProp(item.imgSrc.fileId,'other') == '')
                cR += '<div>' + prx.utils.escapeHTML(item.caption) + '</div>';

        } else {
            //changing the class identifier so that they never start autoplaying on load
            // autoplay will be handled on page load
            if(prx.componentsHelper.getProp(item.imgSrc.fileId,'other') == '') {
                cR += '<img id="' + _id + '-bodymovin" src="data:image/gif;base64,R0lGODlhAQABAIAAAAAAAP///yH5BAEAAAAALAAAAAABAAEAAAIBRAA7" style="width:"+tempW+"px;height:"+tempH+"px""  class="bodymovin-player" data-fileid="'+item.imgSrc.fileId+'" style="width:'+tempW+'px;height:'+tempH+'px"/>';
            } else {
                cR += '<div id="' + _id + '-bodymovin" class="bodymovin-player" data-fileid="'+item.imgSrc.fileId+'"  data-name="' + _id + '"  data-speed = "'+prx.componentsHelper.getProp(item.playbackSpeed,'num-other')+'" data-anim-loop = "'+item.loop+'" data-anim-autoplay="'+item.autoplay+'" data-bm-renderer="svg"></div>';
            }
        }

        cR += '<div></div>';
        cR += '</div>';

        cR += '</div>';

        if (prx.editor && prx.componentsHelper.getProp(item.imgSrc.fileId,'other') == '5222c5ba6594f582d406726697aebb2f.json' && prx.componentsHelper.getProp(item.imgSrc.assetType,'other') == 'gallery') {
            cR += '<div class="type-vectoranimation-hover-message">Double-click to change Lottie JSON file</div>';
            cR += '<style>#' + _id + ' .vector-inner { background-color: #f9f9f9; }</style>';
            cR += '<style>#' + _id + ' .type-vectoranimation-wrapper { border: ' + 1 * prx.componentsHelper.getScale(item.lib) + 'px solid #eee; box-sizing: border-box; }</style>';
        }
        else {
            cR += '<style>';
            cR += '#'+_id+', #'+_id+' .type-vectoranimation-wrapper { border-radius: '+prx.componentsHelper.getProp(item.borderRadius,'border-radius')+';}';
            cR += '#'+_id+' .type-vectoranimation-wrapper { border: '+prx.componentsHelper.getProp(item.borderWidth,'num-border-width')+'px ' + prx.componentsHelper.getProp(item.borderStyle,'border-type') +' '+prx.componentsHelper.getProp(item.borderColor,'color-border')+';  }';
            cR += '</style>';
        }

        cR += prx.items.getComponentAppendDivs(item, containerid, symbol);
        cR += '</div>';

        return cR;
    }
    , interactions: [{
        caption: 'Interactions',
        name: 'actions',
        type: 'action',
        value: function (item, name) {
            if (typeof(item.actions) == 'undefined') {
                item.actions = [];
            }
            return item.actions.length;
        },
        hiddenByDefault: function (item) {
            if (typeof(item.propagateEvents) != 'undefined' && item.propagateEvents) {
                return true;
            }
            return false;
        }
    }]
    ,mpactions: {
        specialEvents: ['vectoranimationend']
    }
    ,propertyGroups: [
        {
            caption: 'Lottie JSON file',
            hint: 'Use the Bodymovin AE plugin to export After Effects animations to Lottie JSON files\n',
            href: 'https://support.proto.io/hc/en-us/articles/115003021171',
            properties: [
                [
                    {
                        caption: false
                        ,name: 'imgSrc'
                        ,type: 'combo-asset'
                        ,displayValue: function(item,name) {
                            if(item.imgSrc.fileId == '') {
                                return 'No asset selected.';
                            }
                            return prx.utils.escapeHTML(item.imgSrc.name);
                        }
                        ,value: function(item,name) {
                            return JSON.stringify({
                                allow: 'bodymovin',
                                asset: item.imgSrc
                            });
                        }
                        ,changeProperty: {
                            caption: 'Lottie JSON file',
                            rerender: true
                        },
                        hiddenByDefault: function(item){
                            return false;
                        }

                    }
                ],
                [
                    {
                        caption: 'Reset to 1x size'
                        ,name: 'resetdimensionsdpr'
                        ,type: 'link'
                        ,onClick: function(item) {

                            return prx.componentsHelper.resetDimensions(item);
                        }
                        ,hiddenByDefault: function(item){
                            item.imgSrc.fileId = item.imgSrc.fileId || '';
                            //return value
                            var r = prx.imageFunctions.checkImageMultiplier(item) == false;

                            r = r || false;

                            return r;
                        }
                    }
                ],
                [
                    {
                        caption: function(item){

                            var multiplier = prx.imageFunctions.checkImageMultiplier(item);

                            if (multiplier !== false)
                                return 'Reset to ' + multiplier + 'x size (original)';
                            else
                                return 'Reset to original size';
                        }
                        ,name: 'resetdimensions'
                        ,type: 'link'
                        ,onClick: function(item) {

                            return prx.componentsHelper.resetDimensions(item);
                        },
                        hiddenByDefault: function(item){
                            return false;
                        }
                    }
                ],

            ]
        },
        {
            caption: 'Style',
            properties: [
                [
                    {
                        caption: 'Border',
                        name: 'borderWidth',
                        proptype: 'border-width',
                        type: 'combo-select',
                        value: function(item, name) {
                            return typeof item.borderWidth == 'undefined' ? '' : item.borderWidth;
                        },
                        values: {min: 0, max: 20, step: 1},
                        changeProperty: {
                            caption: 'Border width',
                            selector: '.changeProperty-borderWidth',
                            property: 'border-width',
                            transitionable: true
                        },
                        hiddenByDefault: function(item){
                            if(prx.mask.isActive(item) || prx.mask.editModeIsMaskSelected(item))
                                return true;
                            else
                                return false;
                        }
                    },
                    {
                        caption: false,
                        name: 'borderStyle',
                        proptype: 'border-style',
                        type: 'select',
                        value: function(item,name) {
                            if(typeof(item.borderStyle) == 'undefined') { item.borderStyle = 'solid'; }
                            return item.borderStyle;
                        },
                        values: [{ value: 'solid', displayValue: 'Solid'},{ value: 'dotted', displayValue: 'Dotted'},{ value: 'dashed', displayValue: 'Dashed'},{ value: 'double', displayValue: 'Double'},{ value: 'none', displayValue: 'None'}],
                        changeProperty: {
                            caption: 'Border Style',
                            selector: '.changeProperty-borderWidth',
                            property: 'border-style',
                            transitionable: false
                        },
                        hiddenByDefault: function(item) {
                            if(prx.mask.isActive(item) || prx.mask.editModeIsMaskSelected(item))
                                return true;
                            else
                                return false;
                        }
                    },
                    prx.commonproperties.borderColor
                ],
                [
                    {
                        caption: 'Position'
                        ,name: 'borderPos'
                        ,type: 'select'
                        ,value: function(item,name,index) {
                            if(typeof(item.borderPos) == 'undefined') {
                                item.borderPos = 'inside';
                            }
                            return item.borderPos;
                        }
                        ,values: [{ displayValue: 'Inside', value: 'inside' }, { displayValue: 'Outside', value: 'outside' }]
                        ,changeProperty: {
                            caption: 'Border Position',
                            selector: '.vector-inner',
                            property: 'class',
                            transitionable: false
                        },
                        hiddenByDefault: function(item){
                            if(prx.mask.isActive(item) || prx.mask.editModeIsMaskSelected(item))
                                return true;
                            else
                                return false;
                        }
                    },
                    {
                        caption: 'Radius',
                        name: 'borderRadius',
                        proptype: 'border-radius',
                        type: 'combo-select',
                        value: function(item, name) {
                            return typeof item.borderRadius == 'undefined' ? '' : item.borderRadius;
                        },
                        values: {min: 0, max: 20, step: 1},
                        expandableType: 'borderRadius',
                        expandedValues: ['tl', 'tr', 'bl', 'br'],
                        changeProperty: {
                            caption: 'Border radius',
                            selector: '.changeProperty-borderRadius',
                            property: 'border-radius',
                            transitionable: true
                        },
                        hiddenByDefault: function(item){
                            if(prx.mask.isActive(item) || prx.mask.editModeIsMaskSelected(item))
                                return true;
                            else
                                return false;
                        }
                    },

                ]
            ]
        },
        {
            caption: 'Playback Options',
            properties: [
                [
                    {
                        caption: 'Autoplay'
                        ,name: 'autoplay'
                        ,type: 'onoff'
                        ,value: function(item,name) {
                            return item.autoplay;
                        },
                        changeProperty: {
                            caption: 'Autoplay',
                            rerender: true
                        },
                        onChange: function(item) {
                            if(prx.editor){
                                if (item.autoplay) {
                                    item.autoplay = true;
                                    $('#' + item.id + '.bodymovin').attr('data-anim-autoplay',true);
                                }else {
                                    item.autoplay = false;
                                    $('#' + item.id + '.bodymovin').attr('data-anim-autoplay',false);
                                }
                                return item;
                            }
                        }
                    }
                ],
                [
                    {
                        caption: 'Loop'
                        ,name: 'loop'
                        ,type: 'onoff'
                        ,value: function(item,name) {
                            return item.loop;
                        },
                        changeProperty: {
                            caption: 'Loop',
                            rerender: true
                        },
                        onChange: function(item) {
                            if(prx.editor){
                                if (item.loop) {
                                    item.loop = true;
                                    $('#' + item.id + '.bodymovin').attr('data-anim-loop',true);
                                }else {
                                    item.loop = false;
                                    $('#' + item.id + '.bodymovin').attr('data-anim-loop',false);
                                }
                                return item;
                            }
                        }
                    }
                ],
                [
                    {
                        caption: 'Speed',
                        name: 'playbackSpeed',
                        proptype: 'playbackSpeed',
                        type: 'input-numeric',
                        // type: 'slider-input',
                        values: { min: 0.1, max: 15, step: 0.1 },
                        value: function(item,name) {
                            return item.playbackSpeed;
                        },
                        changeProperty: {
                            caption: 'Speed',
                            rerender: true
                        },
                    }
                ]
            ]
        },
    ]
};


/***** /BASIC COMPONENTS *****/

/***** TOOLBAR COMPONENTS *****/
//TYPE: BASIC TABBAR
prx.types.basic_tabbar = {
    name: 'basic_tabbar'
    ,onDisplay: function(item,containerid,symbol) {
        var _id = (!containerid) ? item.id : containerid+'-'+item.id;
        var _dims = prx.componentsHelper.getRealDims(item,symbol);
        if(typeof(item.overlay) == 'undefined') { item.overlay = false; }
        if(typeof(item.changeActive) == 'undefined') { item.changeActive = true; }

        var _props = (typeof(prx.richtext)=='undefined') ? prx.componentsHelper.getProp(item.textProperties,'props-text') : '';

        var activebg = prx.componentsHelper.getProp(item.activeBackgroundColor,'other');
        if(activebg == 'as-inactive') { activebg = 'rgba(0,0,0,0)'; } // patenta gia ios7

        var cR = '<div id="' + _id + '" ' + prx.items.getComponentBaseAttributes(item, containerid, symbol)  + ' class="' + prx.items.getComponentBaseClasses(item, containerid, symbol) + ' box pos type-basic-tabbar type-basic-tabbar-icon-'+prx.componentsHelper.getProp(item.iconpos,'other')+'" '+((prx.componentsHelper.getProp(item.overlay,'boolean'))? 'data-mpoverlay="1"': '')+'>';
        var _tabWidth = Math.floor((_dims.width - prx.componentsHelper.getProp(item.borderWidth,'num-border-width')*(item.tabs.length-1))/item.tabs.length);

        cR += '<style>';
        cR += prx.items.getComponentBaseStyle(item, containerid, symbol);
        cR += '#'+_id+' ul { '+prx.gradients.getBgCssByProperty(item.backgroundColor,'color-background')+'; }';
        cR += '#'+_id+' li { width: '+_tabWidth+'px; border-left: '+prx.componentsHelper.getProp(item.borderWidth,'num-border-width')+'px solid '+prx.componentsHelper.getProp(item.borderColor,'color-border')+'; }';
        cR += '#'+_id+' label .caption { color: '+prx.componentsHelper.getProp(item.textColor,'color-text')+'; '+prx.componentsHelper.getProp(item.textFont + '|' + item.textFontStyle,'font-family')+' '+_props+'; ' + prx.componentsHelper.getProp(item.textFontStyle,'font-style') + '; font-size: '+prx.componentsHelper.getProp(item.textSize,'num-text-size')+'px; }';
        cR += '#'+_id+' input:checked + label { ' + prx.gradients.getBgCssByProperty(activebg,'color-background')+'; }';
        cR += '#'+_id+' input:checked + label .caption { color: '+prx.componentsHelper.getProp(item.activeTextColor,'color-border')+'; }';
        if(item.maskEnabled) {
            cR += '#'+_id+' .icon { '+prx.gradients.getBgCssByProperty(item.maskInactive,'color-background')+'; -webkit-mask-size: auto '+(prx.componentsHelper.getProp(item.iconSize,'num-other') * 20)+'%; mask-size: auto '+(prx.componentsHelper.getProp(item.iconSize,'num-other') * 20)+'%; }';
            cR += '#'+_id+' input:checked + label .icon { '+prx.gradients.getBgCssByProperty(item.maskActive,'color-background')+' }';
        }
        switch (item.iconpos) {
        case 'top':
            cR += '#'+_id+' .caption { line-height: '+parseInt(_dims.height*0.25)+'px; }';
            break;
        case '':
            cR += '#'+_id+' .caption { line-height: '+_dims.height+'px; }';
            break;
        default: break;
        }
        cR += '</style>';
        cR += prx.items.getComponentPrependDivs(item, containerid, symbol);

        cR += '<ul class="liveUpdate-backgroundColor-background-color changeProperty-backgroundColor">';
        $.each(item.tabs, function(i,elm){
            if(typeof(elm.linkedscreen) == 'undefined') { elm.linkedscreen = -1; }
            cR += '<li id="'+_id+'-tabs-'+i+'" class="dynamic-property liveUpdate-borderColor-border-color changeProperty-borderColor" data-dynamic-property-index="'+i+'" '+((prx.componentsHelper.getProp(elm.linkedscreen,'num-other') != -1) ? 'data-linked-screen="'+prx.componentsHelper.getProp(elm.linkedscreen,'num-other')+'"' : '')+'>';
            cR += '<input type="radio" name="'+_id+'-radio-input" id="'+_id+'-radio-'+i+'" '+((prx.componentsHelper.getProp(item.activeTab,'num-other') == i) ? 'checked' : '')+((!item.changeActive) ? ' disabled ' : '')+' data-role="none">';
            cR += '<label for="'+_id+'-radio-'+i+'" '+((prx.componentsHelper.getProp(item.activeTab,'num-other') == i) ? 'class="liveUpdate-activeBackgroundColor-background-color"' : '')+' >';
            if(prx.componentsHelper.getProp(item.iconpos,'other') != '') {
                cR += '<style>';
                //fix for Safari bug with -webkit-mask-image
                if(prx.componentsHelper.getProp(item.maskEnabled,'boolean')) {
                    var assetPathWithMaskEnabled = prx.componentsHelper.maskImageFixForSafari11(elm.icon);
                    cR += '#'+_id+' label[for='+_id+'-radio-'+i+'] .icon, #'+_id+' label[for='+_id+'-radio-'+i+'-overlay] .icon { -webkit-mask-image: url("'+assetPathWithMaskEnabled+'"); mask-image: url("'+assetPathWithMaskEnabled+'"); }';
                } else {
                    cR += '#'+_id+' label[for='+_id+'-radio-'+i+'] .icon, #'+_id+' label[for='+_id+'-radio-'+i+'-overlay] .icon { background-image: url('+prx.componentsHelper.getProp(elm.icon,'asset')+'); background-size: auto '+(prx.componentsHelper.getProp(item.iconSize,'num-other') * 20)+'%; }';
                    if(!prx.componentsHelper.getProp(item.maskEnabled,'boolean') && prx.componentsHelper.getProp(elm.activeicon.fileId,'other') != '' && prx.componentsHelper.getProp(elm.activeicon,'asset') != prx.componentsHelper.getProp(elm.icon,'asset') &&  prx.componentsHelper.getProp(elm.activeicon,'asset') != '') {
                        cR += '#'+_id+' input:checked + label[for='+_id+'-radio-'+i+'] .icon, #'+_id+' input:checked + label[for='+_id+'-radio-'+i+'-overlay] .icon { background-image: url('+prx.componentsHelper.getProp(elm.activeicon,'asset')+'); background-size: auto '+(prx.componentsHelper.getProp(item.iconSize,'num-other') * 20)+'%; }';
                    }
                }
                cR += '</style>';
                cR += '<div class="icon-wrapper"><div class="icon '+ ((prx.componentsHelper.getProp(item.maskEnabled,'boolean')) ? ((prx.componentsHelper.getProp(item.activeTab,'num-other') == i) ? 'liveUpdate-maskActive-background-color' : 'liveUpdate-maskInactive-background-color') : '') +'"></div>';
            }
            if(item.iconpos != 'notext') {
                cR += '<span class="caption '+((item.activeTab == i) ? 'liveUpdate-activeTextColor-color' : 'liveUpdate-textColor-color') + '"><span data-editableproperty="text" data-dynamic-property-index="'+i+'">'+prx.componentsHelper.getProp(elm.text,'text-textarea')+'</span></span>';
            }
            if(prx.componentsHelper.getProp(item.iconpos,'other') != '') {
                cR += '</div>';
            }
            cR += '</li>';
        });
        cR += '</ul>';
        cR += prx.items.getComponentAppendDivs(item, containerid, symbol);
        cR += '</div>';
        return cR;
    }
    ,onResize: function(item,containerid,symbol) {
        var _id = (!containerid) ? item.id : containerid+'-'+item.id;
        var _dims = prx.componentsHelper.getRealDims(item, symbol);
        var _tabWidth = Math.floor((_dims.width - item.borderWidth*(item.tabs.length-(1*prx.componentsHelper.getScale(item.lib))))/item.tabs.length);
        switch (item.iconpos) {
        case 'top':
            $('#'+_id+' .caption').css('line-height', parseInt(_dims.height*0.25)+'px');
            break;
        case '':
            $('#'+_id+' .caption').css('line-height', _dims.height+'px');
            break;
        default: break;
        }

        $('#'+_id+' li').width(_tabWidth);
    }
    ,afterDisplay: function(item,containerid,symbol) {
        var _id = (!containerid) ? item.id : containerid+'-'+item.id;

        if(!prx.editor) {
            $('#' + _id + ' [data-linked-screen]').each(function(){
                var screenId = $(this).attr('data-linked-screen');

                if(prx.stc.screens.getIndexFromId(screenId) != -1) {
                    var guid = prx.utils.getGuid();
                    var action = {
                        title: 'Go to screen "'+prxy.pages[prx.stc.screens.getIndexFromId(screenId)].title+'"',
                        type: 'tap',
                        actionId: 'go-to-page',
                        pageId: screenId,
                        animation: 'none',
                        delay: '0',
                        callback: false,
                        guid: guid,
                        bindTo: '#' + $(this).attr('id')

                    };

                    action = prx.actions.recursivelyAddInfo(action, action.bindTo, containerid, containerid+ '',  action.bindTo, '.protoio-actions-afterdisplay');
                    prx.actions.build(action);
                    prx.actions.disableFlashActionOnItemTap(action.bindTo, '.flashactiontap-afterdisplay'); // default one called from actions.build will be disabled in parseaction soon afterwards

                }
            });
        }
    }
    ,propertyGroups: [
        {
            caption: 'Style',
            properties: [[
              				prx.commonproperties.backgroundColor
              				,{
			      				caption: 'Active'
			      				,name: 'activeBackgroundColor'
			      				,proptype: 'background-color-2-active'
			      				,type: 'gradients-colorpicker'
			      				,value: function(item, name){
			      					return item.activeBackgroundColor;
			      				}
              				 	,liveUpdate: 'background-color'
              				 	,changeProperty: {
		      						caption: 'Active background color',
		      						selector: 'input:checked + label',
		      						property: 'background-color',
		      						transitionable: true
		      					 }
			      			}],
			      			[
              					{
              						caption: 'Border',
              						name: 'borderWidth',
              						proptype: 'border-width',
              						type: 'combo-select',
              						value: function(item,name) { return item.borderWidth; },
              						values: { min: 0, max: 20, step: 1 },
              						changeProperty: {
              							caption: 'Border width',
              							rerender: true
                    }
              					}
              					,prx.commonproperties.borderColor
              				]
              			]
              		},
              		{
              			caption: 'Text',
              			properties: [
              				[
			              {
                        caption: { label: 'Font family', class: 'text-properties-label text-fontfamily-label' },
			            	  name: 'textFont',
			            	  proptype: 'font-family',
			            	  type: 'select',
							  relatedEditableProperties: ['tabs.text'],
							  relatedCSSProperties: 'font-family',
			            	  value: function(item,name) { return item.textFont; },
			            	  values:function() { return prx.comps.fonts; },
			            	  hiddenByDefault: function(item) { return (item.iconpos == 'notext'); }
			            	  ,changeProperty: {
	      						caption: 'Text font',
	      						selector: 'label .caption',
	      						property: 'font-family',
	      						transitionable: false
	      					  }
			              }
                ],
                [
                    prx.commonproperties.textFontStyleRichText(['font-weight', 'font-style'],['tabs.text'], function (item) { return (item.iconpos == 'notext'); }, {propName: 'textFontStyle', caption: 'Text Font Style', useAsDynProp: false}),
                    {
                        caption: false,
                        name: 'textSize',
                        proptype: 'font-size',
                        type: 'combo-select',
                        relatedEditableProperties: ['tabs.text'],
                        relatedCSSProperties: 'font-size',
                        value: function(item,name) { return item.textSize; },
                        values: prx.comps.textsize,
                        hiddenByDefault: function(item) { return (item.iconpos == 'notext'); }
                        ,changeProperty: {
                            caption: 'Text size',
                            selector: 'label .caption',
                            property: 'font-size',
                            transitionable: true
                        }
                    }
                ]
			              ,[
                    prx.commonproperties.textPropertiesRichText(['font-weight','font-style','text-decoration'], ['tabs.text'],   function (item) { return (item.iconpos == 'notext'); }, {propName: 'textProperties', caption: 'Underline', useAsDynProp: false})
			              	    ,{
                        caption: { label: 'Default', class: 'text-properties-label text-color-label' },
                        name: 'textColor',
                        proptype: 'font-color',
                        type: 'colorpicker',
                        relatedEditableProperties: ['tabs.text'],
                        relatedCSSProperties: 'color',
                        pffSettings: prx.commonproperties.pffSettingsColor,
                        value: function (item, name) {
                            return item.textColor;
                        },
                        hiddenByDefault: function (item) {
                            return (item.iconpos == 'notext');
                        }
                        , liveUpdate: 'color'
                        , changeProperty: {
                            caption: 'Text color',
                            selector: 'input:not(:checked) + label .caption',
                            property: 'color',
                            transitionable: true
                        }
                    }
                    ,{
              				    caption: { label: 'Active', class: 'text-properties-label text-color-label' }
			      				,name: 'activeTextColor'
			      				,proptype: 'font-color-2-active'
			      				,type: 'colorpicker'
                        ,pffSettings: prx.commonproperties.pffSettingsColor
			      				,value: function(item, name){
			      					return item.activeTextColor;
			      				}
			      				,hiddenByDefault: function(item) { return (item.iconpos == 'notext'); }
			      				,liveUpdate: 'color'
			      				,changeProperty: {
		      						caption: 'Active text color',
		      						selector: 'input:checked + label .caption',
		      						property: 'color',
		      						transitionable: true
		      					  }
			      			}]
			      		]
			      	},
			      	{
			      		caption: 'Icons',
			      		properties: [[
			            	{
			        			caption: false
			        			,name: 'iconpos'
			        			,type: 'select'
			        			,value: function(item,name) {
				      				return item.iconpos;
				            		}
				      			,values: [{value: '',displayValue: 'No icon'},{value: 'top',displayValue: 'Top'},{value: 'notext',displayValue: 'Icon only (no text)'}]
				      		    ,onChange: function(item){
				      				if(item.iconpos == '') {
				      					$('[id=property-icon], [id=property-activeicon], #property-iconSize, #property-maskEnabled, #property-maskInactive, #property-maskActive').hide();
				      				} else {
				      					$('[id=property-icon], [id=property-activeicon], #property-iconSize, #property-maskEnabled, #property-maskInactive, #property-maskActive').show();
				      				}
				      				if(item.iconpos == 'notext') {
				      					$('[id=property-text], #property-textSize, #property-textFont, #property-textFontStyle, #property-textColor, #property-textProperties, #property-activeTextColor').hide();
				      				} else {
				      					$('[id=property-text], #property-textSize, #property-textFont, #property-textFontStyle, #property-textColor, #property-textProperties, #property-activeTextColor').show();
				      				}
				      				return false;
				      			}
				      			,changeProperty: {
		      						caption: 'Icon position',
		      						rerender: true
		      					}
			            	}
				      		,
				      		{
                                caption: function(item, name) {
                                    if(item.name === 'basic_tabbar_icon' || item.name === 'basic_tabbar_texticon')
                                        return 'Size';
                                    return false;
                                }
				      			,name: 'iconSize'
				      			,proptype: 'icon-size'
				      			,type: 'select'
				      			,value: function(item,name) {
				      				return item.iconSize;
				      			}
				      			,values: [{ value: '1', displayValue: 'Very small'}, { value: '2', displayValue: 'Small'}, { value: '3', displayValue: 'Normal'}, { value: '4', displayValue: 'Large'}, { value: '5', displayValue: 'Full'}]
				      			,hiddenByDefault: function(item,name){
				      				return (item.iconpos == '');
				      			}
				      			,changeProperty: {
		      						caption: 'Icon size',
		      						rerender: true
		      					}
				      		}]
				      		,
				      		[{
				      			caption: 'Mask'
				      			,name: 'maskEnabled'
				      			,type: 'onoff'
                                ,help: 'Enabling this option will overwrite the icon\'s original color'
                                ,helpPosition: 'mask-custom'
				      			,value: function(item,name) { return item.maskEnabled; }
					      		,hiddenByDefault: function(item,name){
				      				return (item.iconpos == '');
				      			}
					      		,onChange: function(item) {
				      				if(item.maskEnabled) {
				      					$('#property-maskInactive, #property-maskActive').show();
				      					$('[id=property-activeicon]').hide();
				      				} else {
				      					$('#property-maskInactive, #property-maskActive').hide();
				      					$('[id=property-activeicon]').show();
				      				}
				      			}
				      			,changeProperty: {
		      						caption: 'Mask icons',
		      						rerender: true
		      					}
				      		}
				      		,
				      		{
				      			caption: 'Inactive'
				      			,name: 'maskInactive'
				      			,type: 'colorpicker'
				      			,value: function(item,name) { return item.maskInactive; }
					      		,hiddenByDefault: function(item,name){
				      				return (item.iconpos == '' || !item.maskEnabled);
				      			}
					      		,liveUpdate: 'background-color'
				      			,changeProperty: {
		      						caption: 'Inactive icon mask',
		      						rerender: true
		      					}
				      		}
				      		,
				      		{
				      			caption: 'Active'
				      			,name: 'maskActive'
				      			,type: 'colorpicker'
				      			,value: function(item,name) { return item.maskActive; }
					      		,hiddenByDefault: function(item,name){
				      				return (item.iconpos == '' || !item.maskEnabled);
				      			}
					      		,liveUpdate: 'background-color'
				      			,changeProperty: {
		      						caption: 'Active icon mask',
		      						rerender: true
		      					}
				      		}
				      		]]
				      	}
	      				,
	      				{
	      					caption: 'Advanced',
	      					properties: [[

	  			{
	  	  			caption: 'Active tab'
	  	  			,name: 'activeTab'
	  	  			,type: 'select'
	  	  			,value: function(item,name) {
	  	      			return item.activeTab;
	  	      		}
	  	      		,values: function(item,name) {
	  	      			var _rA = [{value: '999',displayValue: 'None'}];
	  	      			for (var n = 0; n < item.tabs.length; n++) {
	  	      				_rA.push({value: n,displayValue: prx.utils.escapeHTML(item.tabs[n].text.replace(/<br>/g, '\n').replace(/(<([^>]+)>|(&nbsp;))/ig, ''))});
	  	      			}
	  	      			return _rA;
	  	      		}

	      			,changeProperty: {
  						caption: 'Active tab',
  						rerender: true
  					}
	  			}]
	      		,
	      		[
	      			{
		  	  			caption: 'Change active state on click'
		  	  			,name: 'changeActive'
		  	  			,type: 'onoff'
		  	  			,value: function(item,name) {
		  	      			return item.changeActive;
		  	      		}
		      			,changeProperty: {
	  						caption: 'Change active state on click',
	  						rerender: true
	  					}
	  				}
	      		]
	  	      	,[{
                        caption: 'Keep position during screen animations'
                        ,help: 'Will not animate with the rest of the screen during Screen transitions or "Move screen" interactions'
                        ,name: 'overlay'
	  					,type: 'onoff'
	  					,value: function(item,name) {
	  						if(typeof(item.overlay)=='undefined') {
	  							return false;
	  						}
	  						return item.overlay;
	  					}

		      			,changeProperty: {
                            caption: 'Keep position during screen animations',
	  						rerender: true
	  					}
	  	      	}]
	  	      	]
	  	      	}
    ]
    ,dynamicProperties: {
       		data: 'tabs'
       		,propertyCaption: 'Tabs'
        	,propertyName: 'Tab'
       		,addCaption: 'Add tab'
       		,deleteCaption: 'Delete'
       		,blankItem: {
       			text: 'Label',
       			icon: {fileId:'a764f2746aa43431594a25b3e9d5dc34.svg',name:'star.svg',assetType:'icon',bucketsource:'static',url:'f1352971179296/94a90bf9a645dba63ad7a41d18f82ea7.svg',targetSrc:'generated/a764f2746aa43431594a25b3e9d5dc34_6c7075.svg',color:'6c7075'},
                activeicon: {fileId:'a764f2746aa43431594a25b3e9d5dc34.svg',name:'star.svg',assetType:'icon',bucketsource:'static',url:'f1352971179296/94a90bf9a645dba63ad7a41d18f82ea7.svg',targetSrc:'generated/a764f2746aa43431594a25b3e9d5dc34_ffffff.svg',color:'ffffff'},
       			actions: []
       		}
       		,captionProperty: 'text'
       		,interactions: [
       			{
      				caption: 'Interactions'
      				,name: 'actions'
      				,type: 'action'
      				,value: function(item,name,index) {
      					if (typeof(item.tabs[index].actions) == 'undefined') {
      						item.tabs[index].actions = [];
      					}

      					return item.tabs[index].actions.length;
      				}
      			}
       		]
       		,editableProperties: [
       			{
      				caption: 'Text'
      				,name: 'text'
      				,type: 'input'
      				,value: function(item,name,index) {
      					return item.tabs[index].text;
      				}
      				,hiddenByDefault: function(item,name){
      					return (item.iconpos == 'notext');
      				}
      				,changeProperty: {
  						caption: 'Text',
  						selector: '.caption',
  						property: 'text',
  						transitionable: false
  					 }
      			}
       		]
      		,propertyGroups: [
      			{
      				caption: 'Icon',
      				properties: [[
		      			{
		      				caption: false
		      				,name: 'icon'
		      				,type: 'combo-asset'
                            ,hint: function(item,name,index) {
                                if(item.maskEnabled) {
                                    return 'Use Icons only. Uncheck the "Mask icons" property to use images from the Asset manager.';
                                }
                                return false;
                            }
		      				,displayValue: function(item,name,index) {
		      					if(item.tabs[index].icon.fileId == '') {
		      						return 'No icon selected';
		      					}
		      					return item.tabs[index].icon.name;
		      				}
		      				,value: function(item,name,index) {
		      					return JSON.stringify({
		      						allow: 'image',
		      						asset: item.tabs[index].icon
		      					});
		      				}
		    				,hiddenByDefault: function(item,name,index){
		    					return (item.iconpos == '');
		    				}
			      			,changeProperty: {
		  						caption: 'Icon',
		  						rerender: true
		  					 }
		      			}
		      		]]
		      	},
		      	{
		      		caption: 'Active icon (optional)',
		      		properties: [[
		      			{
		      				caption: false
		      				,name: 'activeicon'
		      				,type: 'combo-asset'
		      				,displayValue: function(item,name,index) {
		      					if(item.tabs[index].activeicon.fileId == '') {
		      						return 'No icon selected';
		      					}
		      					return item.tabs[index].activeicon.name;
		      				}
		      				,value: function(item,name,index) {
		      					return JSON.stringify({
		      						allow: 'image',
		      						asset: item.tabs[index].activeicon
		      					});
		      				}
		    				,hiddenByDefault: function(item,name,index){
		    					if(item.iconpos == '') {
		    						return true;
		    					}
		    					return (item.maskEnabled);
		    				}
			      			,changeProperty: {
		  						caption: 'Active icon',
		  						rerender: true
		  					 }
		      			}
		      		]]
		      	},
		      	{
		      		caption: 'Linked screen (optional)',
		      		properties: [[
		      			{
		      				caption: false
		      				,name: 'linkedscreen'
		      				,type: 'select'
		      				,help: 'Will trigger a "Go to screen" action on Tap, and force active state to this tab when the selected screen is active'
		      				,value: function(item,name,index) {
		      					return item.tabs[index].linkedscreen;
		      				}
		      				,values: function(item,name,index) {
		      					var options = [];
		      					for(var i=0; i<prxy.pages.length ; i++) {
		      						options.push({ displayValue: prx.utils.escapeHTML(prxy.pages[i].title), value: prxy.pages[i].id, id: prxy.pages[i].id });
		      					}
                            options.sort(prx.stc.pane.helper.sortCustomPages);
		      					options.unshift({ displayValue: 'None', value: -1, id: -1});
		      					return options;
		      				}
		    				,changeProperty: {
		  						caption: 'Linked Screen',
		  						rerender: true
		  					 }
		      			}
		      		]]
		      	}
        ]
    }
};

/***** /TOOLBAR COMPONENTS *****/


/***** /FORM COMPONENTS *****/
//TYPE: GENERIC ONOFF SWITCH
prx.types.generic_onoffswitch = {
    name: 'generic_onoffswitch'
    ,onDisplay: function(item,containerid,symbol) {
        var _id = (!containerid) ? item.id : containerid+'-'+item.id;

        var _active = '';
        if(item.active) {
            _active = 'checked="checked"';
        }

        var cR = '<div id="' + _id + '" ' + prx.items.getComponentBaseAttributes(item, containerid, symbol)  + ' class="' + prx.items.getComponentBaseClasses(item, containerid, symbol) + ' box pos type-generic-onoffswitch">';

        cR += '<style>';
        cR += prx.items.getComponentBaseStyle(item, containerid, symbol);
        cR += '#'+_id+' label { border-radius: '+prx.componentsHelper.getProp(item.borderRadius,'border-radius')+';}';
        cR += '#'+_id+' .onoffswitch-inner div { line-height: '+(item.height-(4*prx.componentsHelper.getScale(item.lib)))+'px; }';
        cR += '#'+_id+' .onoffswitch-inner .active { background-color: '+prx.componentsHelper.getProp(item.activeLabelColor,'color-background')+'; color: '+prx.componentsHelper.getProp(item.activeLabelTextColor,'color-text')+'; }';
        cR += '#'+_id+' .onoffswitch-inner .inactive { background-color: '+prx.componentsHelper.getProp(item.inactiveLabelColor,'color-background')+'; color: '+prx.componentsHelper.getProp(item.inactiveLabelTextColor,'color-text')+'; }';
        cR += '#'+_id+' .onoffswitch-switch { background-color: '+prx.componentsHelper.getProp(item.switchColor,'color-background')+'; width: '+prx.componentsHelper.getProp(item.switchSize,'num-other')+'px; border-radius: '+prx.componentsHelper.getProp(item.borderRadius,'border-radius')+'; margin: '+((item.height - prx.componentsHelper.getProp(item.switchSize,'num-other'))/2)+'px; right: '+(item.width-(Number(item.switchSize) + 4 + (item.height - Number(item.switchSize))))+'px; }';
        cR += '</style>';
        cR += prx.items.getComponentPrependDivs(item, containerid, symbol);
        cR += '<input type="checkbox" '+_active+ ' id="'+_id+'-onoffswitch" data-role="none" >';
        cR += '<label for="'+_id+'-onoffswitch">';
        cR += '<div class="onoffswitch-inner">';
        cR += '<div class="active">'+prx.componentsHelper.getProp(item.activeLabelText,'other')+'</div>';
        cR += '<div class="inactive">'+prx.componentsHelper.getProp(item.inactiveLabelText,'other')+'</div>';
        cR += '</div>';
        cR += '<div class="onoffswitch-switch"></div>';
        cR += '</label>';
        cR += prx.items.getComponentAppendDivs(item, containerid, symbol);
        cR += '</div>';

        return cR;
    }
    ,properties: [
	        {
	        	caption: 'Border Radius (px)',
	        	name: 'borderRadius',
	        	proptype: 'border-radius',
	        	type: 'combo-select', value: function(item,name) { return item.borderRadius; },
	        	values: { min: 0, max: 50, step: 1 }
	        },
      		{
      			caption: 'Switch color'
      			,name: 'switchColor'
      			,proptype: 'background-color-3-switch'
      			,type: 'colorpicker'
      			,value: function(item,name) {
      				return item.switchColor;
      			}
      		},
	        {
	        	caption: 'Switch size',
	        	name: 'switchSize',
	        	proptype: 'switch-size',
	        	type: 'combo-select', value: function(item,name) { return item.switchSize; },
	        	values: { min: 5, max: 60, step: 1 }
	        }
      		,
      		{
      			caption: 'Active Label Text'
      			,name: 'activeLabelText'
      			,type: 'input'
      			,value: function(item,name) {
      				return item.activeLabelText;
      			}
      		}
      		,
      		{
      			caption: 'Active Label Color'
      			,name: 'activeLabelColor'
      			,proptype: 'background-color-2-active'
      			,type: 'colorpicker'
      			,value: function(item,name) {
      				return item.activeLabelColor;
      			}
      		}
      		,
      		{
      			caption: 'Active Label Text Color'
      			,name: 'activeLabelTextColor'
      			,proptype: 'font-color-2-active'
      			,type: 'colorpicker'
      			,value: function(item,name) {
      				return item.activeLabelTextColor;
      			}
      		}
      		,
      		{
      			caption: 'Inactive Label Text'
      			,name: 'inactiveLabelText'
      			,type: 'input'
      			,value: function(item,name) {
      				return item.inactiveLabelText;
      			}
      		}
      		,
      		{
      			caption: 'Inactive Label Color'
      			,name: 'inactiveLabelColor'
      			,proptype: 'background-color'
      			,type: 'colorpicker'
      			,value: function(item,name) {
      				return item.inactiveLabelColor;
      			}
      		}
      		,
      		{
      			caption: 'Inactive Label Text Color'
      			,name: 'inactiveLabelTextColor'
      			,proptype: 'font-color'
      			,type: 'colorpicker'
      			,value: function(item,name) {
      				return item.inactiveLabelTextColor;
      			}
      		}
      		,
      		{
      			caption: 'Active'
      			,name: 'active'
      			,type: 'onoff'
      			,value: function(item,name) {
      				return item.active;
      			}
      		}
      		,{
      			caption: 'Interactions on activation',
      			name: 'flipswitchActionsOnActive',
      			type: 'action',
      			value: function(item,name) {
	      			if(typeof(item.flipswitchActionsOnActive) == 'undefined') {
	      				item.flipswitchActionsOnActive = [];
	      			}
	      			return item.flipswitchActionsOnActive.length;
      			}
      		}
      		,{
      			caption: 'Interactions on deactivation',
      			name: 'flipswitchActionsOnDeactive',
      			type: 'action',
      			value: function(item,name) {
      				if(typeof(item.flipswitchActionsOnDeactive) == 'undefined') {
  						item.flipswitchActionsOnDeactive = [];
      				}
      				return item.flipswitchActionsOnDeactive.length;
      			}
      		}
      	]
};
/***** /FORM COMPONENTS *****/

//TYPE: DATE/TIME
prx.types.datetime = {
    name: 'datetime'
    , onDisplay: function (item, containerid, symbol) {
        var _id = (!containerid) ? item.id : containerid + '-' + item.id;
        var _props = prx.componentsHelper.getProp(item.textProperties,'props-text');
        	_props += prx.componentsHelper.getProp(item.textFontStyle,'props-text-style');

        var cR = '<div id="' + _id + '" ' + prx.items.getComponentBaseAttributes(item, containerid, symbol) + ' class="' + prx.items.getComponentBaseClasses(item, containerid, symbol) + ' box pos type-datetime">';

        cR += '<style>';
        cR += prx.items.getComponentBaseStyle(item, containerid, symbol);
        cR += '#'+_id+' #background { '+prx.gradients.getBgCssByProperty(item.backgroundColor,'color-background')+';}';
        cR += '#'+_id+' #background { border: '+prx.componentsHelper.getProp(item.borderWidth,'num-border-width')+'px solid '+prx.componentsHelper.getProp(item.borderColor,'color-border')+'; }';
        cR += '#'+_id+' #datetime { color: '+prx.componentsHelper.getProp(item.textColor,'color-text')+'; '+prx.componentsHelper.getProp(item.textFont + '|' + item.textFontStyle,'font-family')+' '+_props+';' + prx.componentsHelper.getProp(item.textFontStyle,'font-style')+'; font-size: '+prx.componentsHelper.getProp(item.textSize,'num-text-size')+'px; text-align: '+prx.componentsHelper.getProp(item.textAlign, 'align')+'; }';
        cR += '#'+_id+' sup {font-size: '+prx.componentsHelper.getProp(item.textSize,'num-text-size')*0.75+'px;}';
        cR += '</style>';
        cR += prx.items.getComponentPrependDivs(item, containerid, symbol);
        cR += '<div id="background" class="liveUpdate-backgroundColor-background-color changeProperty-backgroundColor liveUpdate-borderColor-border-color changeProperty-borderColor">';
        cR += '<div id="datetime" class="liveUpdate-textColor-color changeProperty-textColor changeProperty-textFont  changeProperty-textSize"></div>';
        cR += '</div>';
        cR += prx.items.getComponentAppendDivs(item, containerid, symbol);
        cR += '</div>';

        return cR;


    }
    , afterDisplay: function (item, containerid, symbol) {
        var datetime = new Date();
        var weekdays = ['Sunday', 'Monday', 'Tuesday', 'Wednesday', 'Thursday', 'Friday', 'Saturday'];
        var shortWeekdays = ['Sun', 'Mon', 'Tue', 'Wed', 'Thu', 'Fri', 'Sat'];
        var shortMonths = ['Jan', 'Feb', 'Mar', 'Apr', 'May', 'Jun', 'Jul', 'Aug', 'Sept', 'Oct', 'Nov', 'Dec'];
        var months = ['January', 'February', 'March', 'April', 'May', 'June', 'July', 'August', 'September', 'October', 'November', 'December'];
        var _id = (!containerid) ? item.id : containerid + '-' + item.id;

        var displayDay = function (){
        	switch (item.dayFormat){
            case 'none':
                return '';
                break;
            case 'single':
                return datetime.getDate();
                break;
            case 'double':
                	return ('0' + datetime.getDate()).slice(-2);
                break;
            case 'superscript':
                var lastDigit = datetime.getDate().toString().slice(-2);
                var superscript = '';
                if(datetime.getDate() > 10 && datetime.getDate() < 20){
                    superscript = 'th';
                }else {
                    switch (lastDigit) {
                    case '1':
                        superscript = 'st';
                        break;
                    case '2':
                        superscript = 'nd';
                        break;
                    case '3':
                        superscript = 'rd';
                        break;
                    default:
                        superscript = 'th';
                        break;
                    }
                }
                return datetime.getDate() + superscript.sup();
                break;
            }
        };
        var displayDayName = function (){
            switch (item.dayNameFormat) {
            case 'none':
                return '';
                break;
            case 'short':
                return shortWeekdays[datetime.getDay()]+' ';
                break;
            case 'long':
                return weekdays[datetime.getDay()]+' ';
                break;
            }
        };
        var displayMonth = function (){
        	switch (item.monthFormat){
            case 'none':
                return '';
                break;
            case 'single':
                return datetime.getMonth() + 1;
                break;
            case 'double':
                return ('0' + (datetime.getMonth() + 1)).slice(-2);
                break;
            case 'short':
                	return shortMonths[datetime.getMonth()];
                break;
            case 'long':
                	return months[datetime.getMonth()];
                break;
            }

        };
        var displayYear = function (){
            switch (item.yearFormat) {
            case 'none':
                return '';
                break;
            case 'long':
                return datetime.getFullYear();
                break;
            case 'short':
                return datetime.getFullYear().toString().slice(-2);
                break;
            }
        };
        var displayTime = function (){
            switch (item.timeFormat) {
            case '12a':
                if (datetime.getHours() > 12){
                    	var hour = datetime.getHours() - 12;
                } else{
                    	var hour = datetime.getHours();
                }
                return hour + ':' + ('0' + datetime.getMinutes()).slice(-2);
                break;
            case '12b':
                if (datetime.getHours() > 12){
                    var hour = datetime.getHours() - 12;
                    var ampm = 'PM';
                } else{
                    var hour = datetime.getHours();
                    var ampm = 'AM';
                }
                return hour + ':' + ('0' + datetime.getMinutes()).slice(-2) + ' ' + ampm;
                break;
            case '24':
                return datetime.getHours() + ':' + ('0' + datetime.getMinutes()).slice(-2);
                break;
            }
        };

        var displayDate = function (){
            switch (item.dateFormat){
            case 'daymonthyear':
                return displayDay() + daySeparator() + displayMonth() + yearSeparator() + displayYear();
                break;
            case 'monthdayyear':
                return displayMonth() + daySeparator() + displayDay() + yearSeparator() + displayYear();
                break;
            case 'yearmonthday':
                return displayYear() + yearSeparator() + displayMonth() + daySeparator() + displayDay();
            }
        };

        var daySeparator = function(){
            if (displayDay() == ''){
                return '';
            }else {
                if (displayMonth() == '' && displayYear() == '') {
                    return '';
                } else {
                    return item.dateSeparator;
                }
            }
        };

        var yearSeparator = function () {
            if (displayYear() == ''){
                return '';
            }else {
                if (displayMonth() == '' && displayDay() == '') {
                    return '';
                } else {
                    if (displayDay() != '' && displayMonth() == '') {
                        return '';
                    } else {
                        return item.dateSeparator;
                    }
                }
            }
        };

        var display = function (){
        	switch (item.dateGeneralFormat){
            case 'date':
                return displayDayName() + displayDate();
                break;
            case 'time':
                return displayTime();
                break;
            case 'datetime':
                return displayDayName() + displayDate() + ' ' + displayTime();
                break;
            case 'timedate':
                return displayTime() + ' ' + displayDayName() + displayDate();
                break;
            }
        };

        $('#' + _id + ' #datetime').append(display());
    }
    , interactions: [
        {
            caption: 'Interactions',
            name: 'actions',
            type: 'action',
            value: function(item,name) {
                if (typeof(item.actions) == 'undefined') {
                    item.actions = [];
                }
                return item.actions.length;
            },
            hiddenByDefault: function(item){
            	return false;
            },
            onChange: function(item) {
                if(prx.editor && item.v2===undefined){
                    item.v2 = true;
                    return item;
                }
            }
        }
    ]
    , propertyGroups: [ {
        caption: 'Style',
        properties: [
            [
                prx.commonproperties.backgroundColor
            ],
            [ {
                caption: 'Border',
                name: 'borderWidth',
                proptype: 'border-width',
                type: 'combo-select',
                value: function( item, name ) {
                    return item.borderWidth;
                },
                values: {
                    min: 0,
                    max: 20,
                    step: 1
                },
                changeProperty: {
                    caption: 'Border width',
                    rerender: true
                }
            }, prx.commonproperties.borderColor ]
        ]
    }, {
        caption: 'Text',
        properties: [
            [ 	prx.commonproperties.textFont
            ],
            [
                prx.commonproperties.textFontStyle,
                prx.commonproperties.textSize
            ],
            [
            	prx.commonproperties.textProperties,
                prx.commonproperties.textColor()
            ],
            [
                prx.commonproperties.textAlign
            ]
        ]
    }, {
        caption: 'Format',
        properties: [
            [ {
                caption: { label: 'General Format', class: 'component-datetime-label' },
                name: 'dateGeneralFormat',
                proptype: 'datetime-general-format',
                type: 'select',
                value: function( item, name ) {
                    return typeof item.dateGeneralFormat == 'undefined' ? '' : item.dateGeneralFormat;
                },
                values: prx.comps.dateGeneralFormat,
                changeProperty: {
                    caption: 'General Format',
                    transitionable: false,
                    rerender: true
                }
            } ],
	        [ {
		        caption: { label: 'Date Format', class: 'component-datetime-label' },
		        name: 'dateFormat',
		        proptype: 'datetime-format',
		        type: 'select',
		        value: function( item, name ) {
			        return typeof item.dateFormat == 'undefined' ? '' : item.dateFormat;
		        },
		        values: prx.comps.dateFormat,
		        changeProperty: {
                    caption: 'Date Format',
			        transitionable: false,
                    rerender: true
		        }
	        } ],
	        [ {
		        caption: { label: 'Separator', class: 'component-datetime-label' },
		        name: 'dateSeparator',
		        proptype: 'datetime-separator',
		        type: 'select',
		        value: function( item, name ) {
			        return typeof item.dateSeparator == 'undefined' ? '' : item.dateSeparator;
		        },
		        values: prx.comps.dateSeparator,
		        changeProperty: {
                    caption: 'Separator',
			        transitionable: false,
                    rerender: true
		        }
	        } ]
        ]
    },{
        caption: 'Content',
        properties: [
            [ {
                caption: { label: 'Day Name', class: 'component-datetime-label' },
                name: 'dayNameFormat',
                proptype: 'datetime-day-name-format',
                type: 'select',
                value: function( item, name ) {
                    return typeof item.dayNameFormat == 'undefined' ? '' : item.dayNameFormat;
                },
                values: prx.comps.dayNameFormat,
                changeProperty: {
                    caption: 'Day Name',
                    transitionable: false,
                    rerender: true
                }
            },{
                caption: { label: 'Day', class: 'component-datetime-label' },
                name: 'dayFormat',
                proptype: 'datetime-day-format',
                type: 'select',
                value: function( item, name ) {
                    return typeof item.dayFormat == 'undefined' ? '' : item.dayFormat;
                },
                values: prx.comps.dayFormat,
                changeProperty: {
                    caption: 'Day Format',
                    transitionable: false,
                    rerender: true
                }
            }, {
                caption: { label: 'Month', class: 'component-datetime-label' },
                name: 'monthFormat',
                proptype: 'datetime-month-format',
                type: 'select',
                value: function( item, name ) {
                    return typeof item.monthFormat == 'undefined' ? '' : item.monthFormat;
                },
                values: prx.comps.monthFormat,
                changeProperty: {
                    caption: 'Month Format',
                    transitionable: false,
                    rerender: true
                }
            }, {
                caption: { label: 'Year', class: 'component-datetime-label' },
                name: 'yearFormat',
                proptype: 'datetime-year-format',
                type: 'select',
                value: function( item, name ) {
                    return typeof item.yearFormat == 'undefined' ? '' : item.yearFormat;
                },
                values: prx.comps.yearFormat,
                changeProperty: {
                    caption: 'Year Format',
                    transitionable: false,
                    rerender: true
                }
            } ],
            [ {
                caption: { label: 'Time', class: 'component-datetime-label' },
                name: 'timeFormat',
                proptype: 'datetime-time-format',
                type: 'select',
                value: function( item, name ) {
                    return typeof item.timeFormat == 'undefined' ? '' : item.timeFormat;
                },
                values: prx.comps.timeFormat,
                changeProperty: {
                    caption: 'Time Format',
                    transitionable: false,
                    rerender: true
                }
            } ]
        ]
    } ]
};


/************************************************************************************************/
/************************************* COMPONENTS (OBJECTS) *************************************/
/************************************************************************************************/


/****** GENERAL ******/

prx.components.vrsymbol = {
    name: 'vrsymbol'
    ,type: 'vrsymbol'
    ,lib: _library
    ,caption: 'VR Container (beta)'
    ,scrollToX: 0
    ,scrollToY: 0
    ,width: 'full'
    ,height: 'full'
    ,icon: '-1600px -900px'
    ,properties: 'v,l,o,hpos,vpos,w,h,r,f,ds'
    ,helper: prx.url.devices+'/common/vrsymbol/helper.png'
};


//TYPE: TEXT
prx.components.text = {
    name: 'text'
    ,type: 'text'
    ,lib: _library
    ,caption: 'Text'
    ,icon: '0 0'
    ,helper: prx.url.devices+'/common/text/helper.png'
    ,text: 'Type something'
    ,textFont: 'Arial,sans-serif'
    ,textFontStyle:  '400'
    ,textProperties: []
    ,textSize: 17*prx.componentsHelper.getScale(_library)
    ,textColor:  '000000'
    ,backgroundColor:  'none'
    ,width: 180*prx.componentsHelper.getScale(_library)
    ,height: 40*prx.componentsHelper.getScale(_library)
    ,lineHeight: parseInt(1.231*17*prx.componentsHelper.getScale(_library))
    ,textAlign: 'left'
    ,textSpacing: 0
    ,enableShadow: false
    ,autoResize: true
    ,resizeHandles: 'e,w'
    ,v2: true
};


/****** SHAPES ******/

//TYPE: RECTANGLE
prx.components.rectangle = {
    name: 'rectangle'
    ,type: 'rectangle'
    ,lib: _library
    ,caption: 'Rectangle'
    ,icon: '-100px 0'
    ,helper: prx.url.devices+'/common/rectangle/helper.png'
    ,backgroundColor: 'EDEDED'
    ,borderWidth: 0
    ,borderColor: 'D1D1D1'
    ,borderRadius: 0
    ,imgSrc: {fileId: '', name: '', assetType: '', bucketsource: '', url: ''}
    ,backgroundSizeType: 'cover'
    ,backgroundSizeValue: 35
    ,width: 264*prx.componentsHelper.getScale(_library)
    ,height: 96*prx.componentsHelper.getScale(_library)
    ,lineHeight: parseInt(1.231*17*prx.componentsHelper.getScale(_library))
    ,lineHeightAuto: true
    ,actions:[]
    ,textFont: 'Arial,sans-serif'
    ,textFontStyle:  '400'
    ,textSize: 16*prx.componentsHelper.getScale(_library)
    ,textColor:  '#383838'
    ,textProperties: []
    ,textSpacing: 0
    ,textAlign: 'center'
    ,text: ''
    ,typeName: 'rectangle'
};

//TYPE: CIRCLE
prx.components.circle = {
    name: 'circle'
    ,type: 'circle'
    ,lib: _library
    ,caption: 'Oval'
    ,icon: '-200px 0'
    ,helper: prx.url.devices+'/common/circle/helper.png'
    ,backgroundColor: 'EDEDED'
    ,borderWidth: '0'
    ,borderColor: 'D1D1D1'
    ,borderRadius: '50%'
    ,width: 140*prx.componentsHelper.getScale(_library)
    ,height: 140*prx.componentsHelper.getScale(_library)
    ,lineHeight: parseInt(1.231*17*prx.componentsHelper.getScale(_library))
    ,lineHeightAuto: true
    ,actions:[]
    ,textFont: 'Arial,sans-serif'
    ,textFontStyle:  '400'
    ,textSize: 16*prx.componentsHelper.getScale(_library)
    ,textColor:  '#383838'
    ,textProperties: []
    ,textSpacing: 0
    ,textAlign: 'center'
    ,text: ''
    ,typeName: 'oval'
    ,imgSrc: {fileId: '', name: '', assetType: '', bucketsource: '', url: ''}
    ,backgroundSizeType: 'cover'
    ,backgroundSizeValue: 35
};

//TYPE: SVG SHAPES
prx.components.shape = {
    name: 'shape'
    ,type: 'shape'
    ,lib: _library
    ,caption: 'Shape'
    ,icon: '-300px 0'
    ,width: 140*prx.componentsHelper.getScale(_library)
    ,height: 140*prx.componentsHelper.getScale(_library)
    ,helper: prx.url.devices+'/common/shape/helper.png'
    ,textFont: 'Arial,sans-serif'
    ,textFontStyle:  '400'
    ,textSize: 16*prx.componentsHelper.getScale(_library)
    ,lineHeight: parseInt(1.231*17*prx.componentsHelper.getScale(_library))
    ,lineHeightAuto: true
    ,textColor:  '#000000'
    ,textProperties: []
    ,textSpacing: 0
    ,textAlign: 'center'
    ,text: ''
    ,backgroundColor: 'EDEDED'
    ,borderColor: 'D1D1D1'
    ,borderWidthShape: 0
    ,borderRadius: 0
    ,joinType: 'round'
    ,roundJoin: true
    ,typeName: 'star'
    ,numOfNodes: 5
    ,spikeDepth: 2
    ,skew: 10
    ,baseWidth: 50
    ,actions:[]
};

//TYPE: HORIZONTAL LINE
prx.components.horizontalline = {
    name: 'horizontalline'
    ,type: 'horizontalline'
    ,lib: _library
    ,caption: 'Horizontal line'
    ,icon: '-400px 0'
    ,helper: prx.url.devices+'/common/horizontalline/helper.png'
    ,width: 100*prx.componentsHelper.getScale(_library)
    ,height: 1*prx.componentsHelper.getScale(_library)
    ,resizable : true
    ,resizeHandles : 'e,w'
    ,color: 'A6A6A6'
    ,weight: 1*prx.componentsHelper.getScale(_library)
    ,properties: 'v,l,o,hpos,vpos,w,r,dr,f,ds'
};

//TYPE: VERTICAL LINE
prx.components.verticalline = {
    name: 'verticalline'
    ,type: 'verticalline'
    ,lib: _library
    ,caption: 'Vertical line'
    ,icon: '-500px 0'
    ,helper: prx.url.devices+'/common/verticalline/helper.png'
    ,width: 1*prx.componentsHelper.getScale(_library)
    ,height: 100*prx.componentsHelper.getScale(_library)
    ,resizable : true
    ,resizeHandles : 'n,s'
    ,color: 'A6A6A6'
    ,weight: 1*prx.componentsHelper.getScale(_library)
    ,properties: 'v,l,o,hpos,vpos,h,r,dr,f,ds'
};


/****** ACTIONS ******/

//TYPE: INTERACTION AREA
prx.components.actionarea = {
    name: 'actionarea'
    ,type: 'actionarea'
    ,lib: _library
    ,caption: 'Interaction Area'
    ,assistiveDescription: ''
    ,icon: '-600px 0'
    ,helper: prx.url.devices+'/common/actionarea/helper.png'
    ,width: 100*prx.componentsHelper.getScale(_library)
    ,height: 100*prx.componentsHelper.getScale(_library)
    ,properties: 'v,l,hpos,vpos,w,h,r'
};

//TYPE: ANIMATION TARGET
prx.components.animationtarget = {
    name: 'animationtarget'
    ,type: 'animationtarget'
    ,lib: _library
    ,caption: 'Animation target'
    ,icon: '-700px 0'
    ,helper: prx.url.devices+'/common/animationtarget/helper.png'
    ,width: 20
    ,height: 20
    ,resizable : false
    ,properties: 'v,l,hpos,vpos'
    ,fixPositioning: true
};


/****** ASSETS ******/

//TYPE: IMAGE
prx.components.image = {
    name: 'image'
    ,type: 'image'
    ,lib: _library
    ,caption: 'Image'
    ,icon: '-800px 0'
    ,helper: prx.url.devices+'/common/image/helper.png'
    , imgSrc: {
        fileId: 'd310bece0e91b91b485ed62166d1fc2e.svg',
        assetType: 'gallery',
        bucketsource: 'static',
        name: ' image_placeholder.svg',
        url: 'f1353077251107/01eb56561388a5a9015bcab43ddeeab5.svg'
    }
    ,repeat: false
    ,width: 200 * prx.componentsHelper.getScale(_library)
    ,height: 200 * prx.componentsHelper.getScale(_library)
    ,borderWidth: 0 * prx.componentsHelper.getScale(_library)
    ,borderRadius: 0 * prx.componentsHelper.getScale(_library)
    ,borderColor: '#D1D1D1'
    ,borderPos: 'inside'
    ,propagateEvents: false
    ,actions:[]
    ,overlay: false
    ,autoResize: true
    ,aspectratio: true
    ,assistiveDescription: ''
};

prx.components.vectoranimation = {
    name: 'vectoranimation'
    ,type: 'vectoranimation'
    ,lib: _library
    ,caption: 'AE Lottie Animation'
    ,icon: '-300px -1000px'
    ,helper: prx.url.devices+'/common/vectoranimation/helper.png'
 	,imgSrc: {fileId:'5222c5ba6594f582d406726697aebb2f.json',assetType:'gallery',bucketsource:'static',name:' balloon_animation.json',url:'f1353077251107/5222c5ba6594f582d406726697aebb2f.json', vectorAnimationType: 'bodymovin'}
    ,repeat: false
    , width: 200 * prx.componentsHelper.getScale(_library)
    , height: 200 * prx.componentsHelper.getScale(_library)
    ,borderWidth: 0 * prx.componentsHelper.getScale(_library)
    ,borderRadius: 0 * prx.componentsHelper.getScale(_library)
    ,borderColor: '#000000'
    ,borderPos: 'inside'
    ,propagateEvents: false
    ,actions:[]
    ,overlay: false
    ,autoResize: true
    ,loop: true
    ,autoplay: true
    ,aspectratio: true
    ,playbackSpeed: 1
};

//TYPE: ICON
prx.components.icon = {
    name: 'icon'
    ,type: 'image'
    ,lib: _library
    ,caption: 'Icon'
    ,icon: '-900px 0'
    ,helper: prx.url.devices+'/common/icon/helper.png'
    ,imgSrc: {fileId: '0fe09f6dd3fe341363f4d011b280b36c.svg', name: 'light-on.svg', assetType: 'icon', bucketsource: 'static', url: 'f1352971179296/0fe09f6dd3fe341363f4d011b280b36c.svg'}
    ,repeat: false
    ,width: 100 * prx.componentsHelper.getScale(_library)
    ,height: 100 * prx.componentsHelper.getScale(_library)
    ,propagateEvents: false
    ,actions:[]
    ,overlay: false
    ,autoResize: false
    ,aspectratio: true
};


/****** WEB ******/

//TYPE: PLACEHOLDER
prx.components.placeholder = {
    name: 'placeholder'
    ,type: 'placeholder'
    ,lib: _library
    ,caption: 'Placeholder'
    ,icon: '-1000px 0'
    ,helper: prx.url.devices+'/common/placeholder/helper.png'
    ,width: 200*prx.componentsHelper.getScale(_library)
    ,height: 100*prx.componentsHelper.getScale(_library)
    ,text: 'Placeholder'
    ,textFont: "Helvetica,sans-serif"
    ,textFontStyle:  '400'
    ,textSize: 14*prx.componentsHelper.getScale(_library)
    ,textAlign: 'center'
    ,textColor: '000000'
    ,textProperties: []
    ,backgroundColor: 'EDEDED'
    ,borderColor: 'D1D1D1'
    ,thickness: 1*prx.componentsHelper.getScale(_library)
    ,actions:[]
};

//TYPE: WEB VIEW
prx.components.webview = {
    name: 'webview'
    ,type: 'webview'
    ,lib: _library
    ,caption: 'Web View'
    ,icon: '-1100px 0'
    ,helper: prx.url.devices+'/common/webview/helper.png'
    ,url: 'about:blank'
    ,width: 100*prx.componentsHelper.getScale(_library)
    ,height: 100*prx.componentsHelper.getScale(_library)
    ,scrollable: true
};

//TYPE: HTML CODE
prx.components.html = {
    name: 'html'
    ,type: 'html'
    ,lib: _library
    ,caption: 'HTML Code'
    ,icon: '-1200px 0'
    ,helper: prx.url.devices+'/common/html/helper.png'
    ,width: 100*prx.componentsHelper.getScale(_library)
    ,height: 100*prx.componentsHelper.getScale(_library)
    ,html: ''
};


/****** MEDIA ******/

//TYPE: AUDIO
prx.components.audio = {
    name: 'audio'
    ,type: 'audio'
    ,lib: _library
    ,caption: 'Audio'
    ,icon: '-1300px 0'
    ,helper: prx.url.devices+'/common/audio/helper.png'
    ,width: 300
    ,height: 65
    ,audioFile: { fileId: '', assetType: '', name: ''}
    ,audioFileWAV: { fileId: '', assetType: '', name: ''}
    ,audioFileMP3: { fileId: '', assetType: '', name: ''}
    ,audioFileOGG: { fileId: '', assetType: '', name: ''}
    ,audioFileAAC: { fileId: '', assetType: '', name: ''}
    ,controls: true
    ,preload: false
    ,autoplay: false
    ,autoplayoff: true
    ,loop: false
};

//TYPE: VIDEO
prx.components.video = {
    name: 'video'
    ,type: 'video'
    ,lib: _library
    ,caption: 'Video'
    ,icon: '-1400px 0'
    ,helper: prx.url.devices+'/common/video/helper.png'
    ,width: 250*prx.componentsHelper.getScale(_library)
    ,height: 180*prx.componentsHelper.getScale(_library)
    ,videoType: 'html5'
    ,videoFile: { fileId: '', assetType: '', name: '' }
    ,videoFileMP4: { fileId: '', assetType: '', name: '' }
    ,videoFileOGG: { fileId: '', assetType: '', name: '' }
    ,videoFileWEBM: { fileId: '', assetType: '', name: '' }
    ,controls: true
    ,preload: false
    ,autoplay: false
    ,autoplayoff: true
    ,loop: false
    ,mute: false
    ,placeholder: { fileId: '', assetType: '', name: '' }
    ,youtubeid: ''
    ,vimeoid: ''
};


/****** OTHER ******/

prx.components.tooltip = {
    name: 'tooltip'
    ,type: 'tooltip'
    ,lib: _library
    ,caption: 'Tooltip'
    ,icon: '-1500px 0'
    ,helper: prx.url.devices+ '/common/tooltip/helper.png'
    ,width: 250*prx.componentsHelper.getScale(_library)
    ,height: 150*prx.componentsHelper.getScale(_library)
    ,resizable : true
    ,borderColor: '555555'
    ,backgroundColor: 'ffffff'
    ,borderWidth: 3*prx.componentsHelper.getScale(_library)
    ,borderRadius: 5*prx.componentsHelper.getScale(_library)
    ,ttDirection: 'bottom'
    ,ttPosition: '50'
    ,text: ''
    ,textFont: 'Arial,sans-serif'
    ,textFontStyle:  '400'
    ,textSize: 12*prx.componentsHelper.getScale(_library)
    ,textColor: '383838'
    ,textProperties: []
    ,textAlign: 'left'
};

prx.components.basic_tabbar = {
    name: 'basic_tabbar'
    ,type: 'basic_tabbar'
    ,lib: _library
    ,caption: 'Basic Tabbar'
    ,icon: '-1600px 0'
    ,helper: prx.url.devices+ '/common/basic_tabbar/helper.png'
    ,width:"full"
    ,height: 50*prx.componentsHelper.getScale(_library)
    ,vpos: "bottom"
    ,resizable : true
    ,backgroundColor: 'FFFFFF'
    ,borderWidth: 0
    ,borderColor: 'C6C6C6'
    ,textFont: 'Arial,sans-serif'
    ,textFontStyle:  '700'
    ,textSize: 10*prx.componentsHelper.getScale(_library)
    ,textColor:  '383838'
    ,textProperties: []
    ,iconpos: 'top'
    ,iconSize: 3
    ,activeBackgroundColor: '005fbf'
    ,activeTextColor: 'FFFFFF'
    ,maskEnabled: true
    ,maskInactive: '383838'
    ,maskActive: 'FFFFFF'
    ,activeTab: 0
    ,overlay: false
    ,dynamicAlignment: 'h'
    ,tabs: [
        {
            text: 'Home'
            ,icon: {"fileId":"e4e7dbcddba77b96954aa3602719162c.svg","name":"kub-home.svg","assetType":"icon","bucketsource":"static","url":"f1352971179296/e6a1ba573190139ceda51280e9fdad9c.svg","targetSrc":"generated/e6a1ba573190139ceda51280e9fdad9c_333333.svg","color":"383838"}
            ,activeicon: {"fileId":"e4e7dbcddba77b96954aa3602719162c.svg","name":"kub-home.svg","assetType":"icon","bucketsource":"static","url":"f1352971179296/e6a1ba573190139ceda51280e9fdad9c.svg","targetSrc":"generated/e6a1ba573190139ceda51280e9fdad9c_ffffff.svg","color":"ffffff"}
            ,actions: []
        },
        {
            text: 'Messages'
            ,icon: {"fileId":"e55334acc02f524531f172be5348bfd2.svg","name":"mail-2.svg","assetType":"icon","bucketsource":"static","url":"f1352971179296/1b58b288e91e6a4cb64d90433880003d.svg","targetSrc":"generated/1b58b288e91e6a4cb64d90433880003d_333333.svg","color":"383838"}
            ,activeicon: {"fileId":"e55334acc02f524531f172be5348bfd2.svg","name":"mail-2.svg","assetType":"icon","bucketsource":"static","url":"f1352971179296/1b58b288e91e6a4cb64d90433880003d.svg","targetSrc":"generated/1b58b288e91e6a4cb64d90433880003d_ffffff.svg","color":"ffffff"}
            ,actions: []
        }
    ]
};

prx.components.datetime = {
    name: 'datetime'
    ,type: 'datetime'
    ,lib: _library
    ,caption: 'Date/Time'
    ,icon: '-1700px 0'
    ,helper: prx.url.devices+  '/common/datetime/helper.png'
    ,width: 320*prx.componentsHelper.getScale(_library)
    ,height: 30*prx.componentsHelper.getScale(_library)
    ,resizable : true
    ,backgroundColor: 'none'
    ,borderWidth: 0
    ,borderColor: 'C6C6C6'
    ,textFont: 'Arial,sans-serif'
    ,textFontStyle:  '400'
    ,textSize: 17*prx.componentsHelper.getScale(_library)
    ,textColor:  '383838'
    ,textProperties: []
    ,textAlign: 'center'
    ,dateFormat: 'daymonthyear'
    ,dayFormat: 'superscript'
    ,monthFormat: 'long'
    ,yearFormat: 'long'
    ,timeFormat: '24'
    ,dateGeneralFormat: 'datetime'
    ,dayNameFormat: 'long'
    ,dateSeparator: ' '

};


/****** NON-COMPONENTS/DEPRECATED ******/

prx.components.symbol = {
    name: 'symbol'
    ,type: 'symbol'
    ,lib: _library
    ,caption: 'Container'
    ,scrollToX: 0
    ,scrollToY: 0
};

prx.components.richtext = {
    name: 'richtext'
    ,type: 'richtext'
    ,lib: _library
    ,caption: 'Rich Text'
    ,icon: '0 0'
    ,helper: prx.url.devices+'/common/richtext/helper.png'
    ,text: '<p>Sample text</p>'
    ,width: 200*prx.componentsHelper.getScale(_library)
    ,height: 150*prx.componentsHelper.getScale(_library)
};

prx.components.basic_tabbar_retina = {
    name: 'basic_tabbar_retina'
    ,type: 'basic_tabbar'
    ,lib: _library
    ,caption: 'Basic Tabbar'
    ,icon: '-1600px 0'
    ,helper: prx.url.devices+ '/common/basic_tabbar_retina/helper.png'
    ,width:'full'
    ,height: 96*prx.componentsHelper.getScale(_library)
    ,vpos: 'bottom'
    ,resizable : true
    ,backgroundColor: 'FFFFFF'
    ,borderWidth: 0
    ,borderColor: 'C6C6C6'
    ,textFont: 'Arial,sans-serif'
    ,textFontStyle: '800'
    ,textSize: 10*prx.componentsHelper.getScale(_library)		//was 21, made 10 as basic tabbar above
    ,textColor:  '383838'
    ,textProperties: []
    ,iconpos: 'top'
    ,iconSize: 3
    ,activeBackgroundColor: '005fbf'
    ,activeTextColor: 'FFFFFF'
    ,maskEnabled: true
    ,maskInactive: '383838'
    ,maskActive: 'FFFFFF'
    ,activeTab: 0
    ,changeActive: true
    ,overlay: false
    ,dynamicAlignment: 'h'
    ,tabs: [
        {
            text: 'Home'
            ,icon: {fileId:'e4e7dbcddba77b96954aa3602719162c.svg',name:'kub-home.svg',assetType:'icon',bucketsource:'static',url:'f1352971179296/e6a1ba573190139ceda51280e9fdad9c.svg',targetSrc:'generated/e6a1ba573190139ceda51280e9fdad9c_333333.svg',color:'383838'}
            ,activeicon: {fileId:'e4e7dbcddba77b96954aa3602719162c.svg',name:'kub-home.svg',assetType:'icon',bucketsource:'static',url:'f1352971179296/e6a1ba573190139ceda51280e9fdad9c.svg',targetSrc:'generated/e6a1ba573190139ceda51280e9fdad9c_ffffff.svg',color:'ffffff'}
            ,actions: []
        },
        {
            text: 'Messages'
            ,icon: {fileId:'e55334acc02f524531f172be5348bfd2.svg',name:'mail-2.svg',assetType:'icon',bucketsource:'static',url:'f1352971179296/1b58b288e91e6a4cb64d90433880003d.svg',targetSrc:'generated/1b58b288e91e6a4cb64d90433880003d_333333.svg',color:'383838'}
            ,activeicon: {fileId:'e55334acc02f524531f172be5348bfd2.svg',name:'mail-2.svg',assetType:'icon',bucketsource:'static',url:'f1352971179296/1b58b288e91e6a4cb64d90433880003d.svg',targetSrc:'generated/1b58b288e91e6a4cb64d90433880003d_ffffff.svg',color:'ffffff'}
            ,actions: []
        }
    ]
};

prx.components.generic_onoffswitch = {
    name: 'generic_onoffswitch'
    ,type: 'generic_onoffswitch'
    ,lib: _library
    ,caption: 'Generic On/Off Switch'
    ,icon: '0 0'
    ,helper: 'flipswitch/helper.png'
    ,width: 90*prx.componentsHelper.getScale(_library)
    ,height: 30*prx.componentsHelper.getScale(_library)
    ,resizable : true
    ,borderRadius: 5*prx.componentsHelper.getScale(_library)
    ,switchColor: 'FFFFFF'
    ,switchSize: 17*prx.componentsHelper.getScale(_library)
    ,activeLabelText: 'ON'
    ,activeLabelColor: '6194FD'
    ,activeLabelTextColor: 'FFFFFF'
    ,inactiveLabelText: 'OFF'
    ,inactiveLabelColor: 'FFFFFF'
    ,inactiveLabelTextColor: '666666'
    ,active: true
    ,flipswitchActionsOnActive: []
    ,flipswitchActionsOnDeactive: []
};
```
