## Supplement File 1 for "Post Induction Hypotension prediction during general anesthesia using Machine Learning Techniques": components.js

```
var _library = 'generic2';

//////////////////////////////////////////////////////
//////////////////////////////////////////////////////
//////////////////////////////////////////////////////
//////////////////////////////////////////////////////

//TYPE: BUTTON GROUP
prx.types.basic_buttongroup = {
    name: 'basic_buttongroup'
    ,onDisplay: function(item,containerid,symbol) {
        var _id = (!containerid) ? item.id : containerid+'-'+item.id;
        var _dims = prx.componentsHelper.getRealDims(item,symbol);

        var _props = prx.componentsHelper.getProp(item.textProperties,'props-text');
        var cR = '';
        cR += '<div id="' + _id + '" ' + prx.items.getComponentBaseAttributes(item, containerid, symbol)  + ' class="' + prx.items.getComponentBaseClasses(item, containerid, symbol) + ' box pos type-basic-buttongroup ' + (item.isVertical ? 'vertical' : 'horizontal') + ' iconpos-' + item.iconpos + ' textAlign-' + item.textAlign + ' iconSize-' + item.iconSize + '">';

        cR += '<style>';
        cR += prx.items.getComponentBaseStyle(item, containerid, symbol);

        cR += '#'+_id+' .basic-buttongroup-inner { '+prx.gradients.getBgCssByProperty(item.backgroundColor,'color-background')+'; border: '+prx.componentsHelper.getProp(item.borderWidth,'num-border-width')+'px solid '+prx.componentsHelper.getProp(item.borderColor,'color-border')+'; border-radius: '+prx.componentsHelper.getProp(item.borderRadius,'border-radius')+';  '+prx.componentsHelper.getProp(item.textFont,'font-family')+' color: '+prx.componentsHelper.getProp(item.textColor,'color-text')+'; font-size: '+prx.componentsHelper.getProp(item.textSize,'num-text-size')+'px; '+_props+' }';
        cR += '#'+_id+' .basic-buttongroup-button input:checked + label { '+prx.gradients.getBgCssByProperty(item.activeBackgroundColor,'color-background')+'; color: '+prx.componentsHelper.getProp(item.activeTextColor,'color-text')+'; }';
        cR += '#'+_id+' .icon-wrapper .icon { width: 100%; height: 100%; }';

        if(item.isVertical){
            cR += '#'+_id+' .basic-buttongroup-button { width: 100%; border-top: '+prx.componentsHelper.getProp(item.borderWidth,'num-border-width')+'px solid '+prx.componentsHelper.getProp(item.borderColor,'color-border')+'; }';
            cR += '#'+_id+' .basic-buttongroup-button:first-child {border-top: 0px}';
            cR += '#'+_id+' .basic-buttongroup-button:first-child label { border-top-left-radius: '+(prx.componentsHelper.getProp(parseInt(prx.componentsHelper.getProp(item.borderRadius,'border-radius')) - parseInt(prx.componentsHelper.getProp(item.borderWidth,'num-border-width')) - 1,'border-radius'))+'; border-top-right-radius: '+(prx.componentsHelper.getProp(parseInt(prx.componentsHelper.getProp(item.borderRadius,'border-radius')) - parseInt(prx.componentsHelper.getProp(item.borderWidth,'num-border-width')) - 1,'border-radius'))+'; }';
            cR += '#'+_id+' .basic-buttongroup-button:last-child label { border-bottom-left-radius: '+(prx.componentsHelper.getProp(parseInt(prx.componentsHelper.getProp(item.borderRadius,'border-radius')) - parseInt(prx.componentsHelper.getProp(item.borderWidth,'num-border-width')) - 1,'border-radius'))+'; border-bottom-right-radius: '+(prx.componentsHelper.getProp(parseInt(prx.componentsHelper.getProp(item.borderRadius,'border-radius')) - parseInt(prx.componentsHelper.getProp(item.borderWidth,'num-border-width')) - 1,'border-radius'))+'; }';
            cR += '#'+_id+' .icon-wrapper { '+(item.iconpos == 'right'? 'order: 2; ':'')+'width: '+ (prx.componentsHelper.getProp(_dims.height,'num-other')/item.buttons.length/100*item.iconSize*20*1.1) +'px; }';

        } else {
            cR += '#' + _id + ' .basic-buttongroup-button { height: 100%; border-left: ' + prx.componentsHelper.getProp(item.borderWidth, 'num-border-width') + 'px solid ' + prx.componentsHelper.getProp(item.borderColor, 'color-border') + '; }';
            cR += '#' + _id + ' .basic-buttongroup-button:first-child label { border-top-left-radius: ' + (prx.componentsHelper.getProp(parseInt(prx.componentsHelper.getProp(item.borderRadius, 'border-radius')) - parseInt(prx.componentsHelper.getProp(item.borderWidth, 'num-border-width')) - 1, 'border-radius')) + '; border-bottom-left-radius: ' + (prx.componentsHelper.getProp(parseInt(prx.componentsHelper.getProp(item.borderRadius, 'border-radius')) - parseInt(prx.componentsHelper.getProp(item.borderWidth, 'num-border-width')) - 1, 'border-radius')) + '; }';
            cR += '#' + _id + ' .basic-buttongroup-button:last-child label { border-top-right-radius: ' + (prx.componentsHelper.getProp(parseInt(prx.componentsHelper.getProp(item.borderRadius, 'border-radius')) - parseInt(prx.componentsHelper.getProp(item.borderWidth, 'num-border-width')) - 1, 'border-radius')) + '; border-bottom-right-radius: ' + (prx.componentsHelper.getProp(parseInt(prx.componentsHelper.getProp(item.borderRadius, 'border-radius')) - parseInt(prx.componentsHelper.getProp(item.borderWidth, 'num-border-width')) - 1, 'border-radius')) + '; }';
            cR += '#'+_id+' .icon-wrapper { '+(item.iconpos == 'right'? 'order: 2; ':'')+'width: '+ (prx.componentsHelper.getProp(_dims.height,'num-other')/100*item.iconSize*20*1.1) +'px; }';
        }
        if (item.iconpos !== 'top'){
            cR += '#'+_id+' .icon-wrapper { height: '+ (prx.componentsHelper.getProp(item.iconSize,'num-other')*20) +'%; }';
        }
        if(item.maskEnabled) {
            cR += '#'+_id+' .icon {background-color: '+prx.componentsHelper.getProp(item.maskInactive,'color-background')+'; -webkit-mask-size: auto '+(item.iconSize*20)+'%; mask-size: auto '+(item.iconSize*20)+'%; }';
            if (item.iconpos !== 'top'){
                cR += '#'+_id+' .icon { -webkit-mask-size: auto 100%; mask-size: auto 100%;}';
            }
            else {
                cR += '#'+_id+' .icon {-webkit-mask-size: auto '+(item.iconSize*20)+'%; mask-size: auto '+(item.iconSize*20)+'%;}';
            }
            cR += '#'+_id+' input:checked + label .icon { background-color: '+prx.componentsHelper.getProp(item.maskActive,'color-background')+' }';
        }
        cR += '</style>';
        cR += prx.items.getComponentPrependDivs(item, containerid, symbol);

        cR += '<div class="basic-buttongroup-inner liveUpdate-backgroundColor-background-color liveUpdate-textColor-color liveUpdate-borderColor-border-color changeProperty-backgroundColor changeProperty-borderColor changeProperty-borderRadius changeProperty-borderWidth changeProperty-textColor changeProperty-textSize changeProperty-textFont">';
        $.each(item.buttons, function(i,elm) {

            cR += '<div id="'+_id+'-buttons-'+i+'" class="basic-buttongroup-button dynamic-property liveUpdate-borderColor-border-color changeProperty-borderWidth changeProperty-borderColor'+((i == 0 || i == item.buttons.length-1)? ' changeProperty-borderRadius' : '')+'" data-dynamic-property-index="'+i+'">';
            if( prx.componentsHelper.getProp(item.iconpos,'other') != 'noicon' && prx.componentsHelper.getProp(item.iconpos,'other') != 'none') {
                cR += '<style>';
                //fix for Safari bug with -webkit-mask-image
                if(prx.componentsHelper.getProp(item.maskEnabled,'boolean')) {
                    var assetPathWithMaskEnabled = prx.componentsHelper.maskImageFixForSafari11(elm.icon);
                    cR += '#'+_id+' label[for='+_id+'-radio-'+i+'] .icon, #'+_id+' label[for='+_id+'-radio-'+i+'-overlay] .icon { -webkit-mask-image: url("'+assetPathWithMaskEnabled+'"); mask-image: url("'+assetPathWithMaskEnabled+'"); }';
                } else {
                    if (item.iconpos !== 'top'){
                        cR += '#'+_id+' label[for='+_id+'-radio-'+i+'] .icon, #'+_id+' label[for='+_id+'-radio-'+i+'-overlay] .icon { background-size: auto 100%; }';
                    }
                    else {
                        cR += '#'+_id+' label[for='+_id+'-radio-'+i+'] .icon, #'+_id+' label[for='+_id+'-radio-'+i+'-overlay] .icon { background-size: auto '+(item.iconSize*20)+'%; }';
                    }
                    cR += '#'+_id+' label[for='+_id+'-radio-'+i+'] .icon, #'+_id+' label[for='+_id+'-radio-'+i+'-overlay] .icon { background-image: url('+prx.componentsHelper.getProp(elm.icon,'asset')+'); }';
                    if(!prx.componentsHelper.getProp(item.maskEnabled,'boolean') && prx.componentsHelper.getProp(elm.activeicon.fileId,'other') != '' && prx.componentsHelper.getProp(elm.activeicon,'asset') != prx.componentsHelper.getProp(elm.icon,'asset') &&  prx.componentsHelper.getProp(elm.activeicon,'asset') != '') {
                        if (item.iconpos !== 'top'){
                            cR += '#'+_id+' input:checked + label[for='+_id+'-radio-'+i+'] .icon, #'+_id+' input:checked + label[for='+_id+'-radio-'+i+'-overlay] .icon { background-size: auto 100%; }';
                        }
                        else {
                            cR += '#'+_id+' input:checked + label[for='+_id+'-radio-'+i+'] .icon, #'+_id+' input:checked + label[for='+_id+'-radio-'+i+'-overlay] .icon { background-size: auto '+(item.iconSize*20)+'%; }';
                        }
                        cR += '#'+_id+' input:checked + label[for='+_id+'-radio-'+i+'] .icon, #'+_id+' input:checked + label[for='+_id+'-radio-'+i+'-overlay] .icon { background-image: url('+prx.componentsHelper.getProp(elm.activeicon,'asset')+'); }';
                    }
                }
                cR += '</style>';
                }
            cR += '<input type="radio" name="'+_id+'-radio-input" id="'+_id+'-radio-'+i+'" '+((prx.componentsHelper.getProp(item.selected,'num-other') == i) ? 'checked' : '')+' data-role="none" '+((prx.componentsHelper.getProp(item.active,'num-other') == i) ? 'class="liveUpdate-activeBackgroundColor-background-color liveUpdate-activeTextColor-color changeProperty-activeBackgroundColor"' : '')+' ' +((!prx.componentsHelper.getProp(item.changeActive,'boolean')) ? 'disabled' : '')+ '>';
            cR += '<label for="'+_id+'-radio-'+i+'" '+((prx.componentsHelper.getProp(item.selected,'num-other') == i) ? 'class="changeProperty-activeBackgroundColor liveUpdate-activeBackgroundColor-background-color liveUpdate-activeTextColor-color"' : '')+' >';
            if( prx.componentsHelper.getProp(item.iconpos,'other') != 'noicon' && prx.componentsHelper.getProp(item.iconpos,'other') != 'none') {
                cR += '<div class="icon-wrapper"><div class="icon ' + ((prx.componentsHelper.getProp(item.maskEnabled, 'boolean') && $.browser.webkit) ? ((prx.componentsHelper.getProp(item.activeTab, 'num-other') == i) ? 'liveUpdate-maskActive-background-color' : 'liveUpdate-maskInactive-background-color') : '') + '"></div></div>';
            }
            if(item.iconpos != 'notext') {
                cR += '<div class="caption '+((item.activeTab == i) ? 'liveUpdate-activeTextColor-color' : 'liveUpdate-textColor-color') + '"><span data-editableproperty="text" data-dynamic-property-index="'+i+'">'+prx.componentsHelper.getProp(elm.text,'text-textarea')+'</span></div>';
            }
            cR += '</label>';
            cR += '</div>';
        });

        cR += '</div>';
        cR += prx.items.getComponentAppendDivs(item, containerid, symbol);
        cR += '</div>';
        return cR;
    }
    ,onResize: function(item,containerid,symbol) {
        var _id = (!containerid) ? item.id : containerid+'-'+item.id;
        var _dims = prx.componentsHelper.getRealDims(item,symbol);
        if(item.isVertical) {
            $('#' + _id + ' .icon-wrapper').css({
                width: prx.componentsHelper.getProp(_dims.height, 'num-other') / item.buttons.length / 100 * item.iconSize * 20*1.1 + 'px',
            });
            if (item.iconpos !== 'top') {
                $('#' + _id + ' .icon-wrapper').css({
                    height: prx.componentsHelper.getProp(item.iconSize, 'num-other') * 20 + '%',
                });
            }
        } else {
            $('#' + _id + ' .icon-wrapper').css({
                width: prx.componentsHelper.getProp(_dims.height, 'num-other')  / 100 * item.iconSize * 20*1.1 + 'px',
            });
            if (item.iconpos !== 'top') {
                $('#' + _id + ' .icon-wrapper').css({
                    height: prx.componentsHelper.getProp(item.iconSize, 'num-other') * 20 + '%',
                });
            }
        }
    }
    ,propertyGroups: [
        {
            caption: 'Style',
            properties: [
                [
                    {
                        caption: 'Background',
                        name: 'backgroundColor',
                        proptype: 'background-color',
                        type: 'gradients-colorpicker',
                        value: function(item,name) { return item.backgroundColor; },
                        values: prx.comps.basicColors,
                        liveUpdate: 'background-color',
                        changeProperty: {
                            caption: 'Inactive background color',
                            selector: '.changeProperty-backgroundColor',
                            property: 'background-color',
                            transitionable: true,
                        }
                    },
                    {
                        caption: 'Active',
                        name: 'activeBackgroundColor',
                        proptype: 'background-color-2-active',
                        type: 'gradients-colorpicker',
                        value: function(item,name) { return item.activeBackgroundColor; },
                        values: prx.comps.basicColors,
                        liveUpdate: 'background-color',
                        changeProperty: {
                            caption: 'Active background color',
                            transitionable: false,
                            changeFunction: function(item, containerid) {
                                var _id = (!containerid) ? item.id : containerid+'-'+item.id;
                                $('#' + _id).append(`<style>#${_id} .basic-buttongroup-button input:checked + label { ${prx.gradients.getBgCssByProperty(item.activeBackgroundColor,'color-background')}; }</style>` )
                            }
                        }
                    }
                ]
                ,[
                    prx.commonproperties.borderWidth
                    ,prx.commonproperties.borderColor
                    ,{
                        caption: '<span class="icon icon-border-radius" title="Border radius"></span>',
                        name: 'borderRadius',
                        proptype: 'border-radius',
                        type: 'combo-select',
                        value: function(item, name) {
                            return typeof item.borderRadius == 'undefined' ? '' : item.borderRadius;
                        },
                        values: {min: 0, max: 20, step: 1},
                        changeProperty: {
                            caption: 'Border radius',
                            rerender: true,
                        }
                    }
                ]
            ]
        },
        {
            caption: 'Text',
            properties: [
                [
                    {
                        caption: { label: 'Font family', class: 'text-properties-label text-fontfamily-label' },
                        name: 'textFont',
                        proptype: 'font-family',
                        type: 'select',
                        relatedEditableProperties: 'text',
                        relatedCSSProperties: 'font-family',
                        value: function(item,name) {
                            if(typeof(item.textFont) == 'undefined') { item.textFont = 'Arial,sans-serif'; }
                            return item.textFont;
                        },
                        values: function(){ return prx.comps.fonts; }
                        ,changeProperty: {
                            caption: ' Text font',
                            selector: '.changeProperty-textFont',
                            property: 'font-family',
                            transitionable: false
                        }

                    }
                ],
                [
                    prx.commonproperties.textFontStyleRichText(['font-weight', 'font-style'],['buttons.text'], false, {propName: 'textFontStyle', caption: 'Text font style', useAsDynProp: false}),
                    {
                        caption: false,
                        name: 'textSize',
                        proptype: 'font-size',
                        type: 'combo-select',
                        relatedEditableProperties: 'text',
                        relatedCSSProperties: 'font-size',
                        value: function(item,name) {
                            if(typeof(item.textSize) == 'undefined') { item.textSize = 16; }
                            return item.textSize;
                        },
                        values: prx.comps.textsize
                        ,changeProperty: {
                            caption: ' Text size',
                            selector: '.changeProperty-textSize',
                            property: 'font-size',
                            transitionable: false
                        },
                        onChange: function(item) {
                            if(item.lineHeightAuto==true) {
                                item.lineHeight = parseInt(prx.componentsHelper.getProp(item.textSize,'num-text-size')*1.231);
                            }
                        }
                    }
                ]
                ,
                [
                    prx.commonproperties.textPropertiesRichText(['font-weight','font-style','text-decoration'],['buttons.text'], false, {propName: 'textProperties', caption: 'Underline', useAsDynProp: false}),
                    {
                        caption: { label: 'Inactive', class: 'text-properties-label text-color-label' },
                        name: 'textColor',
                        proptype: 'font-color',
                        type: 'solid-colorpicker',
                        relatedEditableProperties: 'text',
                        relatedCSSProperties: 'color',
                        pffSettings: prx.commonproperties.pffSettingsColor,
                        value: function(item,name) {
                            return item.textColor;
                        },
                        liveUpdate: 'color'
                        ,changeProperty: {
                            caption: 'Inactive text color',
                            selector: 'input:not(:checked) + label .caption',
                            property: 'color',
                            transitionable: true
                        }
                    },
                    {
                        caption: { label: 'Active', class: 'text-properties-label text-color-label' },
                        name: 'activeTextColor',
                        proptype: 'font-color-2-active',
                        type: 'solid-colorpicker',
                        relatedEditableProperties: 'text',
                        relatedCSSProperties: 'color',
                        pffSettings: prx.commonproperties.pffSettingsColor,
                        value: function(item,name) {
                            return item.activeTextColor;
                        },
                        liveUpdate: 'color'
                        ,changeProperty: {
                            caption: 'Active text color',
                            transitionable: false,
                            changeFunction: function(item, containerid) {
                                var _id = (!containerid) ? item.id : containerid+'-'+item.id;
                                $('#' + _id).append(`<style>#${_id} .basic-buttongroup-button input:checked + label { color: ${prx.componentsHelper.getProp(item.activeTextColor,'color-text')}; }</style>` )
                            }
                        }
                    }
                ],
                [
                    {
                        caption: { label: 'Alignment', class: 'text-properties-label text-alignment-label' },
                        name: 'textAlign',
                        proptype: 'text-align',
                        type: 'radio',
                        relatedEditableProperties: 'text',
                        relatedCSSProperties: 'text-align',
                        pffSettings: prx.commonproperties.pffSettingsTextAlignment,
                        value: function(item,name) {
                            if(typeof(item.textAlign) == 'undefined') { item.textAlign = 'center'; }
                            return item.textAlign;
                        },
                        values: [
                            { value: 'left', displayValue: '', icon: 'align-left'},
                            { value: 'center', displayValue: '', icon: 'align-center'},
                            { value: 'right', displayValue: '', icon: 'align-right'},
                            { value: 'justify', displayValue: '', icon: 'align-justify'}
                        ],
                        changeProperty: {
                            caption: 'Text Align',
                            rerender: true,
                        }
                    }
                ]
            ]
        },
        {
            caption: 'Icons',
            properties: [[
                {
                    caption: false
                    ,name: 'iconpos'
                    ,type: 'select'
                    ,value: function(item,name) {
                        return item.iconpos;
                    }
                    ,values: function(item,name) {
                        var possibleValues = [];

                        if(item.name !== 'basic_buttongroup_horizontal_texticon_right' && item.name !== 'basic_buttongroup_horizontal_texticon_top' && item.name !== 'basic_buttongroup_vertical_texticon') {
                            possibleValues.push({value: 'none',displayValue: 'No icon'});
                        }

                        possibleValues.push({value: 'left',displayValue: 'Left'});
                        possibleValues.push({value: 'top',displayValue: 'Top'});
                        possibleValues.push({value: 'right',displayValue: 'Right'});

                        if(item.name !== 'basic_buttongroup_horizontal_texticon_right' && item.name !== 'basic_buttongroup_horizontal_texticon_top' && item.name !== 'basic_buttongroup_vertical_texticon') {
                            possibleValues.push({value: 'notext',displayValue: 'Icon only (no text)'});
                        }

                        return possibleValues;
                    }
                    ,onChange: function(item){
                        if(item.iconpos == 'none') {
                            $('#property-iconSize, #property-maskEnabled, #property-maskInactive, #property-maskActive').hide();
                        } else {
                            $('#property-iconSize, #property-maskEnabled, #property-maskInactive, #property-maskActive').show();
                        }
                        if(item.iconpos == 'notext') {
                            $('#property-textSize, #property-textFont, #property-textFontStyle, #property-textColor, #property-textProperties, #property-textAlign, #property-activeTextColor').hide();
                        } else {
                            $('#property-textSize, #property-textFont, #property-textFontStyle, #property-textColor, #property-textProperties, #property-textAlign, #property-activeTextColor').show();
                        }
                        return false;
                    }
                    ,changeProperty: {
                        caption: 'Icon position',
                        rerender: true
                    }
                },
                {
                    caption: function(item, name) {
                        if(item.name === 'basic_buttongroup_horizontal_icon' || item.name === 'basic_buttongroup_vertical_icon')
                            return 'Size';

                        return false;
                    }
                    ,name: 'iconSize'
                    ,proptype: 'icon-size'
                    ,type: 'select'
                    ,value: function(item,name) {
                        return item.iconSize;
                    }
                    ,values: [{ value: '1', displayValue: 'Very small'}, { value: '2', displayValue: 'Small'}, { value: '3', displayValue: 'Normal'}, { value: '4', displayValue: 'Large'}, { value: '5', displayValue: 'Full'}]
                    ,hiddenByDefault: function(item,name){
                        return (item.iconpos == 'none');
                    }
                    ,changeProperty: {
                        caption: 'Icon size',
                        rerender: true
                    }
                }],
            [{
                caption: 'Mask'
                ,name: 'maskEnabled'
                ,type: 'onoff'
                ,help: 'Enabling this option will overwrite the icon\'s original color'
                ,helpPosition: 'mask-custom'
                ,value: function(item,name) { return item.maskEnabled; }
                ,hiddenByDefault: function(item,name){
                    return (item.iconpos == 'none');
                }
                ,onChange: function(item) {
                    if(item.maskEnabled) {
                        $('#property-maskInactive, #property-maskActive').show();
                        $('[id=property-activeicon]').hide();
                    } else {
                        $('#property-maskInactive, #property-maskActive').hide();
                        $('[id=property-activeicon]').show();
                    }
                }
                ,changeProperty: {
                    caption: 'Mask icons',
                    rerender: true
                }
            }
            ,
            {
                caption: 'Inactive'
                ,name: 'maskInactive'
                ,type: 'colorpicker'
                ,value: function(item,name) { return item.maskInactive; }
                ,hiddenByDefault: function(item,name){
                    return (item.iconpos == 'none' || !item.maskEnabled);
                }
                ,liveUpdate: 'background-color'
                ,changeProperty: {
                    caption: 'Inactive icon mask',
                    transitionable: false,
                    changeFunction (item, containerid) {
                        if(prx.componentsHelper.getProp(item.maskEnabled,'boolean')) {
                            let id = (!containerid) ? item.id : containerid + '-' + item.id;
                            let cssSelector = '#' + id + ' input + label .icon';

                            prx.componentsHelper.addStyleForChangeFunction(id, id + '-maskInactive', cssSelector, 'background-color', item.maskInactive, 'color-background');
                        }
                    }
                }
            }
            ,
            {
                caption: 'Active'
                ,name: 'maskActive'
                ,type: 'colorpicker'
                ,value: function(item,name) { return item.maskActive; }
                ,hiddenByDefault: function(item,name){
                    return (item.iconpos == 'none' || !item.maskEnabled);
                }
                ,liveUpdate: 'background-color'
                ,changeProperty: {
                    caption: 'Active icon mask',
                    transitionable: false,
                    changeFunction (item, containerid) {
                        if(prx.componentsHelper.getProp(item.maskEnabled,'boolean')) {
                            let id = (!containerid) ? item.id : containerid + '-' + item.id;
                            let cssSelector = '#' + id + ' input:checked + label .icon ';

                            prx.componentsHelper.addStyleForChangeFunction(id, id + '-maskActive', cssSelector, 'background-color', item.maskActive, 'color-background');
                        }
                    }
                }
            }
            ]]
        },
        {
            caption: 'Advanced',
            properties: [
                [
                    {
                        caption: 'Active Button'
                        ,name: 'selected'
                        ,type: 'select'
                        ,value: function(item,name) {
                            return item.selected;
                        }
                        ,values: function(item,name) {

                            var _rA = [];
                            _rA.push({value: '999',displayValue: 'None'});

                            for (var n = 0; n < item.buttons.length; n++) {
                                _rA.push({value: n,displayValue: prx.utils.escapeHTML(item.buttons[n].text.replace(/<br>/g, '\n').replace(/(<([^>]+)>|(&nbsp;))/ig, ''))});
                            }

                            return _rA;
                        }
                        ,changeProperty: {
                            caption: 'Active button',
                            transitionable: false,
                            changeFunction (item, containerid) {
                                item.buttons.forEach(function (button, idx) {
                                    let id = (!containerid) ? item.id : containerid + '-' + item.id;
                                    let cssSelector = '#' + id + ' input' + '#' + id + '-radio-' + idx;

                                    if (item.selected == idx) {
                                        var isInitiallyDisabled = $(cssSelector)[0].disabled;

                                        if(isInitiallyDisabled) {
                                            $(cssSelector)[0].disabled = false;
                                        }
                                        $(cssSelector)[0].click();
                                        if(isInitiallyDisabled) {
                                            $(cssSelector)[0].disabled = true;
                                        }
                                    }
                                });
                            }
                        }

                    }
                ],
                [
                    {
                        caption: 'Change active state on click'
                        ,name: 'changeActive'
                        ,type: 'onoff'
                        ,value: function(item,name) {
                            return item.changeActive;
                        }
                        ,changeProperty: {
                            caption: 'Change active state on click',
                            transitionable: false,
                            changeFunction (item, containerid) {
                                item.buttons.forEach(function (button, idx) {
                                    let id = (!containerid) ? item.id : containerid + '-' + item.id;
                                    let cssSelector = '#' + id + ' input' + '#' + id + '-radio-' + idx;

                                    $(cssSelector)[0].disabled = !item.changeActive;
                                });
                            }
                        }
                    }
                ]
            ]
        }
    ]
    ,dynamicProperties: {
        data: 'buttons'
        ,propertyCaption: 'Buttons'
        ,propertyName: 'Button'
        ,addCaption: 'Add button'
        ,deleteCaption: 'Delete'
        ,captionProperty: 'text'
        ,blankItem: {
            text: 'Label'
            ,icon: {fileId:'987371e0c84e1e3877c0b8f6aea57272.svg',name:'home.svg',assetType:'icon',bucketsource:'static',url:'f1352971179296/caeebe7f3a62939528c6a4ed009de42c.svg',targetSrc:'generated/987371e0c84e1e3877c0b8f6aea57272_6c7075.svg',color:'6c7075'}
            ,activeicon: {fileId:'987371e0c84e1e3877c0b8f6aea57272.svg',name:'home.svg',assetType:'icon',bucketsource:'static',url:'f1352971179296/caeebe7f3a62939528c6a4ed009de42c.svg',targetSrc:'generated/987371e0c84e1e3877c0b8f6aea57272_ffffff.svg',color:'ffffff'}
            ,actions: []
        }
        ,interactions: [ prx.commonproperties.actions_buttons ]
        ,editableProperties: [
            {
                caption: 'Text'
                ,name: 'text'
                ,type: 'input'
                ,value: function(item,name,index) {
                    return item.buttons[index].text;
                }
                ,changeProperty: {
                    caption: 'Text',
                    selector: 'label',
                    property: 'text',
                    transitionable: false
                }
            }
        ]
        ,propertyGroups: [
            {
                caption: 'Icon',
                properties: [[
                    {
                        caption: false
                        ,name: 'icon'
                        ,type: 'combo-asset'
                        ,hint: function(item,name,index) {
                            if(item.maskEnabled) {
                                return 'Use Icons only. Uncheck the "Mask icons" property to use images from the Asset manager.';
                            }
                            return false;
                        }
                        ,displayValue: function(item,name,index) {
                            if(item.buttons[index].icon.fileId == '') {
                                return 'No icon selected';
                            }
                            return item.buttons[index].icon.name;
                        }
                        ,value: function(item,name,index) {
                            return JSON.stringify({
                                allow: 'image',
                                asset: item.buttons[index].icon
                            });
                        }
                        ,hiddenByDefault: function(item,name,index){
                            return (item.iconpos == 'none');
                        }
                        ,changeProperty: {
                            caption: 'Icon',
                            transitionable: false,
                            changeFunction (item, containerid, duration, easing, index) {
                                let id = (!containerid) ? item.id : containerid + '-' + item.id;
                                let cssSelector = '#' + id + ' label[for=' + id + '-radio-' + index + '] .icon, ' + '#' + id + ' label[for=' + id + '-radio-' + index + '-overlay] .icon';

                                var assetPathWithMaskEnabled = prx.componentsHelper.maskImageFixForSafari11(item.buttons[index].icon);

                                if (prx.componentsHelper.getProp(item.maskEnabled,'boolean')) {
                                    prx.componentsHelper.changeImageForChangeFunction(cssSelector, assetPathWithMaskEnabled, false, true);
                                } else {
                                    prx.componentsHelper.changeImageForChangeFunction(cssSelector, assetPathWithMaskEnabled, true, true);
                                    prx.componentsHelper.addStyleForChangeFunction(id, id + '-maskicon-background-size-' + index, cssSelector, 'background-size', 'auto 60%', 'other');
                                }
                            }
                        }
                    }
                ]]
            },
            {
                caption: 'Active Icon (optional)',
                properties: [[
                    {
                        caption: false
                        ,name: 'activeicon'
                        ,type: 'combo-asset'
                        ,displayValue: function(item,name,index) {
                            if(item.buttons[index].activeicon.fileId == '') {
                                return 'No icon selected';
                            }
                            return item.buttons[index].activeicon.name;
                        }
                        ,value: function(item,name,index) {
                            return JSON.stringify({
                                allow: 'image',
                                asset: item.buttons[index].activeicon
                            });
                        }
                        ,hiddenByDefault: function(item,name,index){
                            return (item.iconpos == 'none' || item.maskEnabled);
                        }
                        ,changeProperty: {
                            caption: 'Active icon (optional)',
                            transitionable: false,
                            changeFunction (item, containerid, duration, easing, index) {
                                let id = (!containerid) ? item.id : containerid + '-' + item.id;
                                let cssSelector = '#' + id + ' input:checked + label[for=' + id + '-radio-' + index + '] .icon, ' + '#' + id + ' input:checked + label[for=' + id + '-radio-' + index + '-overlay] .icon';

                                var assetPathWithMaskEnabled = prx.componentsHelper.maskImageFixForSafari11(item.buttons[index].activeicon);

                                prx.componentsHelper.changeImageForChangeFunction(cssSelector, assetPathWithMaskEnabled, true, true);
                                prx.componentsHelper.addStyleForChangeFunction(id, id + '-activeicon-background-size-' + index, cssSelector, 'background-size', 'auto 60%', 'other');
                            }
                        }
                    }
                ]]
            }
        ]
    }
};

//TYPE: BUTTON GROUP - TEXT HORIZONTAL
prx.types.basic_buttongroup_horizontal_text = prx.componentsHelper.cloneobject(prx.types.basic_buttongroup);
prx.types.basic_buttongroup_horizontal_text.name = 'basic_buttongroup_horizontal_text';
prx.componentsHelper.removeProperties(prx.types.basic_buttongroup_horizontal_text.propertyGroups, ['iconpos', 'iconSize', 'maskEnabled', 'maskInactive', 'maskActive']);
prx.componentsHelper.removeProperties(prx.types.basic_buttongroup_horizontal_text.dynamicProperties.propertyGroups, ['icon', 'activeicon']);

//TYPE: BUTTON GROUP - ICONS ONLY HORIZONTAL
prx.types.basic_buttongroup_horizontal_icon = prx.componentsHelper.cloneobject(prx.types.basic_buttongroup);
prx.types.basic_buttongroup_horizontal_icon.name = 'basic_buttongroup_horizontal_icon';
prx.componentsHelper.removeProperties(prx.types.basic_buttongroup_horizontal_icon.propertyGroups, ['textFont', 'textSize', 'textFontStyle', 'textAlign', 'textColor', 'textProperties', 'activeTextColor', 'iconpos']);
prx.componentsHelper.removeProperties(prx.types.basic_buttongroup_horizontal_icon.dynamicProperties.editableProperties, ['text']);

//TYPE: BUTTON GROUP - TEXT VERTICAL
prx.types.basic_buttongroup_vertical_text = prx.componentsHelper.cloneobject(prx.types.basic_buttongroup);
prx.types.basic_buttongroup_vertical_text.name = 'basic_buttongroup_vertical_text';
prx.componentsHelper.removeProperties(prx.types.basic_buttongroup_vertical_text.propertyGroups, ['iconpos', 'iconSize', 'maskEnabled', 'maskInactive', 'maskActive']);
prx.componentsHelper.removeProperties(prx.types.basic_buttongroup_vertical_text.dynamicProperties.propertyGroups, ['icon', 'activeicon']);

//TYPE: BUTTON GROUP - TEXT ICON VERTICAL
prx.types.basic_buttongroup_vertical_texticon = prx.componentsHelper.cloneobject(prx.types.basic_buttongroup);
prx.types.basic_buttongroup_vertical_texticon.name = 'basic_buttongroup_vertical_text';

//TYPE: BUTTON GROUP - ICONS ONLY VERTICAL
prx.types.basic_buttongroup_vertical_icon = prx.componentsHelper.cloneobject(prx.types.basic_buttongroup);
prx.types.basic_buttongroup_vertical_icon.name = 'basic_buttongroup_vertical_icon';
prx.componentsHelper.removeProperties(prx.types.basic_buttongroup_vertical_icon.propertyGroups, ['textFont', 'textSize', 'textFontStyle', 'textAlign', 'textColor', 'textProperties', 'activeTextColor', 'iconpos']);
prx.componentsHelper.removeProperties(prx.types.basic_buttongroup_vertical_icon.dynamicProperties.editableProperties, ['text']);

//TYPE: CHECKBOX LIST
prx.types.basic_checkbox_list = {
    name: 'basic_checkbox_list'
    ,onDisplay: function(item,containerid,symbol) {
        var _id = (!containerid) ? item.id : containerid+'-'+item.id;
        var _dims = prx.componentsHelper.getRealDims(item,symbol);

        var _props = prx.componentsHelper.getProp(item.textProperties,'props-text');
        var cR = '';
        cR += '<div id="' + _id + '" ' + prx.items.getComponentBaseAttributes(item, containerid, symbol)  + ' class="' + prx.items.getComponentBaseClasses(item, containerid, symbol) + ' box pos type-basic-checkboxlist ' + (item.isVertical ? 'vertical' : 'horizontal') + ' iconpos-' + item.iconpos + ' textAlign-' + item.textAlign + '">';

        cR += '<style>';
        cR += prx.items.getComponentBaseStyle(item, containerid, symbol);
        if(item.isVertical){
            cR += '#' + _id + ' .basic-checkboxlist-inner { '+prx.gradients.getBgCssByProperty(item.backgroundColor,'color-background')+'; border: '+prx.componentsHelper.getProp(item.borderWidth,'num-border-width')+'px solid '+prx.componentsHelper.getProp(item.borderColor,'color-border')+'; border-radius: '+prx.componentsHelper.getProp(item.borderRadius,'border-radius')+';  '+prx.componentsHelper.getProp(item.textFont,'font-family')+' color: '+prx.componentsHelper.getProp(item.textColor,'color-text')+'; font-size: '+prx.componentsHelper.getProp(item.textSize,'num-text-size')+'px; '+_props+' }';
            cR += '#' + _id + ' .basic-checkboxlist-button { width: 100%; border-top: '+prx.componentsHelper.getProp(item.borderWidth,'num-border-width')+'px solid '+prx.componentsHelper.getProp(item.borderColor,'color-border')+'; }';
            cR += '#' + _id + ' .basic-checkboxlist-button:first-child {border-top: 0px}';
            cR += '#' + _id + ' .basic-checkboxlist-button input:checked + label { '+prx.gradients.getBgCssByProperty(item.activeBackgroundColor,'color-background')+'; color: '+prx.componentsHelper.getProp(item.activeTextColor,'color-text')+'; }';
            cR += '#' + _id + ' .basic-checkboxlist-button:first-child label { border-top-left-radius: '+(prx.componentsHelper.getProp(parseInt(prx.componentsHelper.getProp(item.borderRadius,'border-radius')) - parseInt(prx.componentsHelper.getProp(item.borderWidth,'num-border-width')) - 1,'border-radius'))+'; border-top-right-radius: '+(prx.componentsHelper.getProp(parseInt(prx.componentsHelper.getProp(item.borderRadius,'border-radius')) - parseInt(prx.componentsHelper.getProp(item.borderWidth,'num-border-width')) - 1,'border-radius'))+'; }';
            cR += '#' + _id + ' .basic-checkboxlist-button:last-child label { border-bottom-left-radius: '+(prx.componentsHelper.getProp(parseInt(prx.componentsHelper.getProp(item.borderRadius,'border-radius')) - parseInt(prx.componentsHelper.getProp(item.borderWidth,'num-border-width')) - 1,'border-radius'))+'; border-bottom-right-radius: '+(prx.componentsHelper.getProp(parseInt(prx.componentsHelper.getProp(item.borderRadius,'border-radius')) - parseInt(prx.componentsHelper.getProp(item.borderWidth,'num-border-width')) - 1,'border-radius'))+'; }';
            cR += '#' + _id + ' .icon { height: '+ ((_dims.height/item.listitems.length || 1) * prx.componentsHelper.getProp(item.iconSize,'num-other')*0.2) +'px;}';
        } else {
            cR += '#' + _id + ' .basic-checkboxlist-inner { ' + prx.gradients.getBgCssByProperty(item.backgroundColor,'color-background') + '; border: ' + prx.componentsHelper.getProp(item.borderWidth, 'num-border-width') + 'px solid ' + prx.componentsHelper.getProp(item.borderColor, 'color-border') + '; border-radius: ' + prx.componentsHelper.getProp(item.borderRadius, 'border-radius') + ';  ' + prx.componentsHelper.getProp(item.textFont, 'font-family') + ' color: ' + prx.componentsHelper.getProp(item.textColor, 'color-text') + '; font-size: ' + prx.componentsHelper.getProp(item.textSize, 'num-text-size') + 'px; ' + _props + ' }';
            cR += '#' + _id + ' .basic-checkboxlist-button { height: 100%; border-left: ' + prx.componentsHelper.getProp(item.borderWidth, 'num-border-width') + 'px solid ' + prx.componentsHelper.getProp(item.borderColor, 'color-border') + '; }';
            cR += '#' + _id + ' .basic-checkboxlist-button input:checked + label { ' + prx.gradients.getBgCssByProperty(item.activeBackgroundColor,'color-background') + '; color: ' + prx.componentsHelper.getProp(item.activeTextColor, 'color-text') + '; }';
            cR += '#' + _id + ' .basic-checkboxlist-button:first-child label { border-top-left-radius: ' + (prx.componentsHelper.getProp(parseInt(prx.componentsHelper.getProp(item.borderRadius, 'border-radius')) - parseInt(prx.componentsHelper.getProp(item.borderWidth, 'num-border-width')) - 1, 'border-radius')) + '; border-bottom-left-radius: ' + (prx.componentsHelper.getProp(parseInt(prx.componentsHelper.getProp(item.borderRadius, 'border-radius')) - parseInt(prx.componentsHelper.getProp(item.borderWidth, 'num-border-width')) - 1, 'border-radius')) + '; }';
            cR += '#' + _id + ' .basic-checkboxlist-button:last-child label { border-top-right-radius: ' + (prx.componentsHelper.getProp(parseInt(prx.componentsHelper.getProp(item.borderRadius, 'border-radius')) - parseInt(prx.componentsHelper.getProp(item.borderWidth, 'num-border-width')) - 1, 'border-radius')) + '; border-bottom-right-radius: ' + (prx.componentsHelper.getProp(parseInt(prx.componentsHelper.getProp(item.borderRadius, 'border-radius')) - parseInt(prx.componentsHelper.getProp(item.borderWidth, 'num-border-width')) - 1, 'border-radius')) + '; }';
            cR += '#' + _id + ' .icon { height: '+ (_dims.height * prx.componentsHelper.getProp(item.iconSize,'num-other')*0.2) +'px;}';
        }
        cR += '</style>';
        cR += prx.items.getComponentPrependDivs(item, containerid, symbol);

        cR += '<div class="basic-checkboxlist-inner liveUpdate-backgroundColor-background-color liveUpdate-textColor-color liveUpdate-borderColor-border-color changeProperty-backgroundColor changeProperty-borderColor changeProperty-borderRadius changeProperty-borderWidth changeProperty-textColor changeProperty-textSize changeProperty-textFont">';
        $.each(item.listitems, function(i,elm) {

            cR += '<div id="'+_id+'-buttons-'+i+'" class="basic-checkboxlist-button dynamic-property liveUpdate-borderColor-border-color changeProperty-borderWidth changeProperty-borderColor'+((i == 0 || i == item.listitems.length-1)? ' changeProperty-borderRadius' : '')+'" data-dynamic-property-index="'+i+'">';
            cR += '<input type="checkbox" name="'+_id+'-checkbox" id="'+_id+'-checkbox-'+i+'" '+((prx.componentsHelper.getProp(item.selected,'num-other') == i || (Array.isArray(item.selected) && item.selected.includes(i.toString()))) ? 'checked' : '')+' data-role="none" '+((prx.componentsHelper.getProp(item.active,'num-other') == i) ? 'class="liveUpdate-activeBackgroundColor-background-color liveUpdate-activeTextColor-color changeProperty-activeBackgroundColor"' : '')+' ' +((!prx.componentsHelper.getProp(item.changeActive,'boolean')) ? 'disabled' : '')+ '>';
            cR += '<label for="'+_id+'-checkbox-'+i+'" '+((prx.componentsHelper.getProp(item.selected,'num-other') == i) || (Array.isArray(item.selected) && item.selected.includes(i.toString())) ? 'class="changeProperty-activeBackgroundColor liveUpdate-activeBackgroundColor-background-color liveUpdate-activeTextColor-color"' : '')+' >';

            if( prx.componentsHelper.getProp(item.iconpos,'other') != 'noicon' && prx.componentsHelper.getProp(item.iconpos,'other') != 'none') {
                cR += '<img class="icon inactiveIcon" src="'+prx.componentsHelper.getProp(item.inactiveicon,'asset')+'">';
                cR += '<img class="icon activeIcon" src="'+prx.componentsHelper.getProp(item.activeicon,'asset')+'">';
            }
            cR += '<div class="caption '+((item.activeTab == i) ? 'liveUpdate-activeTextColor-color' : 'liveUpdate-textColor-color') + '"><span data-editableproperty="text" data-dynamic-property-index="'+i+'">'+prx.componentsHelper.getProp(elm.text,'text-textarea')+'</span></div>';
            cR += '</label>';
            cR += '</div>';
        });

        cR += '</div>';
        cR += prx.items.getComponentAppendDivs(item, containerid, symbol);
        cR += '</div>';
        return cR;
    }
    ,afterDisplay: function(item,containerid,symbol){
        var _id = (!containerid) ? item.id : containerid+'-'+item.id;
        if(!prx.editor) {
            $('#' + _id).hammer();
            prx.actions.disableFlashActionOnItemTap('#' + _id, '.flashactiontap-afterdisplay');
        }
    }
    ,onResize: function(item,containerid,symbol) {
        var _id = (!containerid) ? item.id : containerid+'-'+item.id;
        var _dims = prx.componentsHelper.getRealDims(item,symbol);
        // chrome bug. cannot do this with css because it flickers.
        if(item.isVertical) {
            $('#' + _id + ' .icon').height(((_dims.height / item.listitems.length || 1) * prx.componentsHelper.getProp(item.iconSize, 'num-other') * 0.2));
        } else {
            $('#' + _id + ' .icon').height((_dims.height * prx.componentsHelper.getProp(item.iconSize, 'num-other') * 0.2));
        }
    }
    ,propertyGroups: [
        {
            caption: 'Style',
            properties: [
                [
                    {
                        caption: 'Background',
                        name: 'backgroundColor',
                        proptype: 'background-color',
                        type: 'gradients-colorpicker',
                        value: function(item,name) { return item.backgroundColor; },
                        values: prx.comps.basicColors,
                        liveUpdate: 'background-color',
                        changeProperty: {
                            caption: 'Inactive background color',
                            selector: '.changeProperty-backgroundColor',
                            property: 'background-color',
                            transitionable: true
                        }
                    },
                    {
                        caption: 'Active',
                        name: 'activeBackgroundColor',
                        proptype: 'background-color-2-active',
                        type: 'gradients-colorpicker',
                        value: function(item,name) { return item.activeBackgroundColor; },
                        values: prx.comps.basicColors,
                        liveUpdate: 'background-color',
                        changeProperty: {
                            caption: 'Active background color',
                            selector: '.changeProperty-activeBackgroundColor',
                            property: 'background-color',
                            transitionable: true
                        }
                    }
                ]
                ,[
                    prx.commonproperties.borderWidth
                    ,prx.commonproperties.borderColor
                    ,{
                        caption: '<span class="icon icon-border-radius" title="Border radius"></span>',
                        name: 'borderRadius',
                        proptype: 'border-radius',
                        type: 'combo-select',
                        value: function(item, name) {
                            return typeof item.borderRadius == 'undefined' ? '' : item.borderRadius;
                        },
                        values: {min: 0, max: 20, step: 1},
                        changeProperty: {
                            caption: 'Border radius',
                            transitionable: false,
                            changeFunction: function (item, containerid, duration, easing, dynPropI){
                                if (typeof item == 'undefined') return;
                                var _id = (!containerid) ? item.id : containerid+'-'+item.id;
                                $('#'+_id+' .basic-checkboxlist-inner').css('border-radius', item.borderRadius+'px');
                                if(item.isVertical){
                                    $('#'+_id+' .basic-checkboxlist-button:first-child label').css({'border-top-left-radius': item.borderRadius+'px', 'border-top-right-radius': item.borderRadius+'px'});
                                    $('#'+_id+' .basic-checkboxlist-button:last-child label').css({'border-bottom-left-radius': item.borderRadius+'px', 'border-bottom-right-radius': item.borderRadius+'px'});
                                } else {
                                    $('#'+_id+' .basic-checkboxlist-button:first-child label').css({'border-top-left-radius': item.borderRadius+'px', 'border-bottom-left-radius': item.borderRadius+'px'});
                                    $('#'+_id+' .basic-checkboxlist-button:last-child label').css({'border-top-right-radius': item.borderRadius+'px', 'border-bottom-right-radius': item.borderRadius+'px'});
                                }
                            }
                        }
                    }
                ]
            ]
        },
        {
            caption: 'Text',
            properties: [
                [
                    {
                        caption: { label: 'Font family', class: 'text-properties-label text-fontfamily-label' },
                        name: 'textFont',
                        proptype: 'font-family',
                        type: 'select',
                        relatedEditableProperties: 'text',
                        relatedCSSProperties: 'font-family',
                        value: function(item,name) {
                            if(typeof(item.textFont) == 'undefined') { item.textFont = 'Arial,sans-serif'; }
                            return item.textFont;
                        },
                        values: function(){ return prx.comps.fonts; }
                        ,changeProperty: {
                            caption: ' Text font',
                            selector: '.changeProperty-textFont',
                            property: 'font-family',
                            transitionable: false
                        }

                    }
                ],
                [
                    prx.commonproperties.textFontStyleRichText(['font-weight', 'font-style'],['listitems.text'], false, {propName: 'textFontStyle', caption: 'Text Font Style', useAsDynProp: false}),
                    {
                        caption: false,
                        name: 'textSize',
                        proptype: 'font-size',
                        type: 'combo-select',
                        relatedEditableProperties: ['listitems.text'],
                        relatedCSSProperties: 'font-size',
                        value: function(item,name) {
                            if(typeof(item.textSize) == 'undefined') { item.textSize = 16; }
                            return item.textSize;
                        },
                        values: prx.comps.textsize
                        ,changeProperty: {
                            caption: ' Text size',
                            selector: '.changeProperty-textSize',
                            property: 'font-size',
                            transitionable: true
                        },
                        onChange: function(item) {
                            if(item.lineHeightAuto==true) {
                                item.lineHeight = parseInt(prx.componentsHelper.getProp(item.textSize,'num-text-size')*1.231);
                            }
                        }
                    }
                ]
                ,
                [
                    prx.commonproperties.textPropertiesRichText(['font-weight','font-style','text-decoration'],['listitems.text'], false, {propName: 'textProperties', caption: 'Underline', useAsDynProp: false}),
                    {
                        caption: { label: 'Inactive', class: 'text-properties-label text-color-label' },
                        name: 'textColor',
                        proptype: 'font-color',
                        type: 'solid-colorpicker',
                        relatedEditableProperties: ['listitems.text'],
                        relatedCSSProperties: 'color',
                        pffSettings: prx.commonproperties.pffSettingsColor,
                        value: function(item,name) {
                            return item.textColor;
                        },
                        liveUpdate: 'color'
                        ,changeProperty: {
                            caption: 'Inactive text color',
                            selector: 'input:not(:checked) + label .caption',
                            property: 'color',
                            transitionable: true
                        }
                    },
                    {
                        caption: { label: 'Active', class: 'text-properties-label text-color-label' },
                        name: 'activeTextColor',
                        proptype: 'font-color-2-active',
                        type: 'solid-colorpicker',
                        relatedEditableProperties: ['listitems.text'],
                        relatedCSSProperties: 'color',
                        pffSettings: prx.commonproperties.pffSettingsColor,
                        value: function(item,name) {
                            return item.activeTextColor;
                        },
                        liveUpdate: 'color'
                        ,changeProperty: {
                            caption: 'Active text color',
                            selector: 'input:checked + label .caption',
                            property: 'color',
                            transitionable: true
                        }
                    }
                ],
                [
                    {
                        caption: { label: 'Alignment', class: 'text-properties-label text-alignment-label' },
                        name: 'textAlign',
                        proptype: 'text-align',
                        type: 'radio',
                        relatedEditableProperties: ['listitems.text'],
                        relatedCSSProperties: 'text-align',
                        pffSettings: prx.commonproperties.pffSettingsTextAlignment,
                        value: function(item,name) {
                            if(typeof(item.textAlign) == 'undefined') { item.textAlign = 'center'; }
                            return item.textAlign;
                        },
                        values: [
                            { value: 'left', displayValue: '', icon: 'align-left'},
                            { value: 'center', displayValue: '', icon: 'align-center'},
                            { value: 'right', displayValue: '', icon: 'align-right'},
                            { value: 'justify', displayValue: '', icon: 'align-justify'}
                        ],
                        changeProperty: {
                            caption: 'Text Align',
                            rerender: true
                        }
                    }
                ]
            ]
        },
        {
            caption: 'Icons',
            properties: [[
                {
                    caption: false
                    ,name: 'iconpos'
                    ,proptype: 'iconpos'
                    ,type: 'select'
                    ,value: function(item,name) {
                        return item.iconpos;
                    }
                    ,values: [{value: 'none',displayValue: 'No icon'},{value: 'left',displayValue: 'Left'},{value: 'right',displayValue: 'Right'}]
                    ,onChange: function(item){
                        if(item.iconpos == 'none') {
                            $('#property-iconSize, #property-inactiveicon, #property-activeicon').hide();
                        } else {
                            $('#property-iconSize, #property-inactiveicon, #property-activeicon').show();
                        }
                        return false;
                    }
                    ,changeProperty: {
                        caption: 'Icon position',
                        rerender: true
                    }
                },
                {
                    caption: false
                    ,name: 'iconSize'
                    ,proptype: 'icon-size'
                    ,type: 'select'
                    ,value: function(item,name) {
                        return item.iconSize;
                    }
                    ,values: [{ value: '1', displayValue: 'Very small'}, { value: '2', displayValue: 'Small'}, { value: '3', displayValue: 'Normal'}, { value: '4', displayValue: 'Large'}, { value: '5', displayValue: 'Full'}]
                    ,hiddenByDefault: function(item,name){
                        return (item.iconpos == 'none');
                    }
                    ,changeProperty: {
                        caption: 'Icon size',
                        rerender: true
                    }
                }],
            [{
                caption: 'Inactive'
                ,name: 'inactiveicon'
                ,proptype: 'inactiveicon'
                ,type: 'combo-asset'
                ,displayValue: function(item,name,index) {
                    if(item.inactiveicon.fileId == '') {
                        return 'No icon selected';
                    }
                    return item.inactiveicon.name;
                }
                ,value: function(item,name,index) {
                    return JSON.stringify({
                        allow: 'image',
                        asset: item.inactiveicon
                    });
                }
                ,hiddenByDefault: function(item,name,index){
                    return (item.iconpos == 'none');
                }
                ,changeProperty: {
                    caption: 'Inactive icon',
                    rerender: true
                }
            }],
            [{
                caption: 'Active'
                ,name: 'activeicon'
                ,proptype: 'activeicon'
                ,type: 'combo-asset'
                ,displayValue: function(item,name,index) {
                    if(item.activeicon.fileId == '') {
                        return 'No icon selected';
                    }
                    return item.activeicon.name;
                }
                ,value: function(item,name,index) {
                    return JSON.stringify({
                        allow: 'image',
                        asset: item.activeicon
                    });
                }
                ,hiddenByDefault: function(item,name,index){
                    return (item.iconpos == 'none');
                }
                ,changeProperty: {
                    caption: 'Active icon',
                    rerender: true
                }
            }]
            ]
        },
        {
            caption: 'Advanced',
            properties: [
                [
                    {
                        caption: 'Active Item'
                        ,name: 'selected'
                        ,type: 'multiple-select'
                        ,hint: 'Hold {ctrlKeyName} and select to select multiple items'
                        ,value: function(item,name) {
                            return item.selected;
                        }
                        ,values: function(item,name) {

                            var _rA = [];

                            for (var n = 0; n < item.listitems.length; n++) {
                                _rA.push({value: n,displayValue: prx.utils.escapeHTML(item.listitems[n].text.replace(/<br>/g, '\n').replace(/(<([^>]+)>|(&nbsp;))/ig, ''))});
                            }

                            return _rA;
                        }
                        ,changeProperty: {
                            caption: 'Active Item',
                            rerender: true
                        }

                    }
                ],
                [
                    {
                        caption: 'Change active state on click'
                        ,name: 'changeActive'
                        ,type: 'onoff'
                        ,value: function(item,name) {
                            return item.changeActive;
                        }
                        ,changeProperty: {
                            caption: 'Change active state on click',
                            rerender: true
                        }
                    }
                ]
            ]
        }
    ]
    ,dynamicProperties: {
        data: 'listitems'
        ,allowMultipleActiveSelection: true
        ,propertyCaption: 'List items'
        ,propertyName: 'List item'
        ,addCaption: 'Add list item'
        ,deleteCaption: 'Delete'
        ,captionProperty: 'text'
        ,blankItem: {
            text: 'Label'
            ,actions: []
            ,actions1: []
            ,actions2: []
        }
        ,interactions: [
            {
                caption: 'Change',
                name: 'actions',
                type: 'action'
                ,value: function(item,name,index) {
                    if (typeof(item.listitems[index].actions) == 'undefined') {
                        item.listitems[index].actions = [];
                    }
                    return item.listitems[index].actions.length;
                },
                changeProperty: {
                    caption: 'Change',
                    selector: '',
                    property: 'action',
                    transitionable: false,
                    changeable: false
                }
            },
            {
                caption: 'Activation',
                name: 'actions1',
                type: 'action',
                value: function (item, name,index) {
                    if (typeof(item.listitems[index].actions1) == "undefined") {
                        item.listitems[index].actions1 = [];
                    }
                    return item.listitems[index].actions1.length;
                },
                changeProperty: {
                    caption: 'Activation',
                    selector: '',
                    property: 'action',
                    transitionable: false,
                    changeable: false
                }
            },
            {
                caption: 'Deactivation',
                name: 'actions2',
                type: 'action',
                value: function (item, name,index) {
                    if (typeof(item.listitems[index].actions2) == "undefined") {
                        item.listitems[index].actions2 = [];
                    }
                    return item.listitems[index].actions2.length;
                },
                changeProperty: {
                    caption: 'Deactivation',
                    selector: '',
                    property: 'action',
                    transitionable: false,
                    changeable: false
                }
            }

        ]
        ,mpactions: {
            specialEvents: ['checkboxchange']
        }
        ,editableProperties: [
            {
                caption: 'Text'
                ,name: 'text'
                ,type: 'input'
                ,value: function(item,name,index) {
                    return item.listitems[index].text;
                }
                ,changeProperty: {
                    caption: 'Text',
                    selector: 'label .caption',
                    property: 'text',
                    transitionable: false
                }
            }
        ]
    }
};

//TYPE: CHECKBOX LIST HORIZONTAL
prx.types.basic_checkbox_list_horizontal = prx.componentsHelper.cloneobject(prx.types.basic_checkbox_list);
prx.types.basic_checkbox_list_horizontal.name = 'basic_checkbox_list_horizontal';

//TYPE: CHECKBOX LIST VERTICAL
prx.types.basic_checkbox_list_vertical = prx.componentsHelper.cloneobject(prx.types.basic_checkbox_list);
prx.types.basic_checkbox_list_vertical.name = 'basic_checkbox_list_vertical';

//TYPE: BASIC BUTTON text ICON
prx.types.basic_button_texticon = prx.componentsHelper.cloneobject(prx.types.basic_button_bg);
prx.types.basic_button_texticon.name = 'basic_button_texticon';
prx.types.basic_button_texticon.propertyGroups = prx.componentsHelper.editProperty(prx.types.basic_button_texticon.propertyGroups, 'iconpos', 'values', [{value: 'left',displayValue: 'Left'},{value: 'right',displayValue: 'Right'}]);

//TYPE: BASIC BUTTON ICON
prx.types.basic_button_icon = prx.componentsHelper.cloneobject(prx.types.basic_button_bg);
prx.types.basic_button_icon.name = 'basic_button_icon';
prx.componentsHelper.removeProperties(prx.types.basic_button_icon.propertyGroups, ['iconpos', 'textFont', 'textFontStyle', 'textSize', 'textColor', 'textProperties', 'textAlign']);
prx.componentsHelper.removeProperties(prx.types.basic_button_icon.editableProperties, ['text']);

//TYPE: BASIC BUTTON TEXT
prx.types.basic_button_text = prx.componentsHelper.cloneobject(prx.types.basic_button_bg);
prx.types.basic_button_text.name = 'basic_button_text';
prx.componentsHelper.removeProperties(prx.types.basic_button_text.propertyGroups, ['iconpos', 'iconSize', 'img']);

//TYPE: BASIC TABBAR TEXT
prx.types.basic_tabbar_text = prx.componentsHelper.cloneobject(prx.types.basic_tabbar);
prx.types.basic_tabbar_text.name = 'basic_tabbar_text';
prx.componentsHelper.removeProperties(prx.types.basic_tabbar_text.propertyGroups, ['iconpos', 'iconSize', 'maskEnabled', 'maskInactive', 'maskActive']);
prx.componentsHelper.removeProperties(prx.types.basic_tabbar_text.dynamicProperties.propertyGroups, ['icon', 'activeicon']);

//TYPE: BASIC TABBAR ICON
prx.types.basic_tabbar_icon = prx.componentsHelper.cloneobject(prx.types.basic_tabbar);
prx.types.basic_tabbar_icon.name = 'basic_tabbar_icon';
prx.componentsHelper.removeProperties(prx.types.basic_tabbar_icon.propertyGroups, ['textFont', 'textSize', 'textColor', 'textProperties', 'activeTextColor', 'iconpos']);
prx.componentsHelper.removeProperties(prx.types.basic_tabbar_icon.dynamicProperties.editableProperties, ['text']);

//TYPE: BASIC TABBAR TEXT & ICON
prx.types.basic_tabbar_texticon = prx.componentsHelper.cloneobject(prx.types.basic_tabbar);
prx.types.basic_tabbar_texticon.name = 'basic_tabbar_texticon';
prx.componentsHelper.removeProperties(prx.types.basic_tabbar_texticon.propertyGroups, ['iconpos']);

// TYPE: ON OFF SWITCH
prx.types.basic_switch = {
    name: 'basic_switch'
    ,onDisplay: function(item,containerid,symbol) {
        var _id = (!containerid) ? item.id : containerid+'-'+item.id;
        var _dims = prx.componentsHelper.getRealDims(item,symbol);

        var _hasRadioButtonsGroupName = typeof(item.radioButtonsGroupName) != 'undefined' && item.radioButtonsGroupName.length > 0;
        var _type = !_hasRadioButtonsGroupName ? 'checkbox' : 'radio';
        var radioButtonsGroupName = _hasRadioButtonsGroupName ? 'name="' + containerid + '-radioButtonsGroup-' + item.radioButtonsGroupName + '"' : '';

        var cR = '';
        cR += '<div id="' + _id + '" ' + prx.items.getComponentBaseAttributes(item, containerid, symbol)  + ' class="' + prx.items.getComponentBaseClasses(item, containerid, symbol) + ' box pos type-basic-switch">';

        cR += '<style>';
        cR += prx.items.getComponentBaseStyle(item, containerid, symbol);
        cR += '#'+_id+' label .switch-background { width: 100%; height: 100%; border-radius: '+prx.componentsHelper.getProp(item.borderRadius,'border-radius')+'; border: '+prx.componentsHelper.getProp(item.borderWidth,'num-border-width')+'px solid '+prx.componentsHelper.getProp(item.borderColor,'color-border')+'; background: '+prx.componentsHelper.getProp(item.inactiveBarColor,'color-background')+'; }';
        cR += '#'+_id+' input:checked + label .switch-background { background: '+prx.componentsHelper.getProp(item.activeBarColor,'color-background')+'; }';
        cR += '#'+_id+' .switch { left: '+ ((_dims.height - item.switchSize)/2) + 'px; width: '+item.switchSize+'px; height: '+item.switchSize+'px; background: '+prx.componentsHelper.getProp(item.inactiveSwitchColor,'color-background')+'; border-radius: '+prx.componentsHelper.getProp(item.borderRadius,'border-radius')+'; border: '+prx.componentsHelper.getProp(item.borderWidth,'num-border-width')+'px solid '+prx.componentsHelper.getProp(item.borderColor,'color-border')+'; }';
        cR += '#'+_id+' input:checked + label .switch-background .switch { left: '+(_dims.width - item.switchSize - item.borderWidth * 2 - (_dims.height - item.switchSize)/2)+'px; background: '+prx.componentsHelper.getProp(item.activeSwitchColor,'color-background')+' }';
        cR += '</style>';
        cR += prx.items.getComponentPrependDivs(item, containerid, symbol);

        cR += '<input type="'+_type+'" '+((prx.componentsHelper.getProp(item.active,'boolean')) ? 'checked' : '')+ ' id="'+_id+'-flipswitch" data-role="none" ' + radioButtonsGroupName + ' />';

        var labelClasses,
            switchClasses;

        if(prx.componentsHelper.getProp(item.active,'boolean')) {
            labelClasses = ' liveUpdate-activeBarColor-background changeProperty-activeBarColor';
            switchClasses = 'liveUpdate-activeSwitchColor-background changeProperty-activeSwitchColor';
        }
        else {

            labelClasses = ' liveUpdate-inactiveBarColor-background changeProperty-inactiveBarColor';
            switchClasses = 'liveUpdate-inactiveSwitchColor-background changeProperty-inactiveSwitchColor';
        }

        cR += '<label for="'+_id+'-flipswitch"><div class="switch-background liveUpdate-borderColor-border-color changeProperty-borderWidth changeProperty-borderColor changeProperty-borderRadius '+ labelClasses +'" data-clickable="true">'; // data-clickable for greensock draggable - if draggable or in draggable container clicking on the flipswitch wont change state

        cR += '<div class="switch liveUpdate-borderColor-border-color changeProperty-borderWidth changeProperty-borderColor changeProperty-borderRadius '+ switchClasses +'"></div>';
        cR += '</div>';
        cR += '</label>';

        cR += prx.items.getComponentAppendDivs(item, containerid, symbol);
        cR += '</div>';

        return cR;
    }
    ,onResize: function(item,containerid,symbol){
        var _id = (!containerid) ? item.id : containerid+'-'+item.id;
        var _dims = prx.componentsHelper.getRealDims(item,symbol);
        if($('#'+_id+'-flipswitch').is(':checked')){
            $('#' + _id + ' label .switch-background .switch').css('left', (_dims.width - item.switchSize - item.borderWidth * 2 - (_dims.height - item.switchSize) / 2));
        } else {
            $('#' + _id + ' label .switch-background .switch').css('left', ((_dims.height - item.switchSize)/2));
        }
    }
    ,afterDisplay: function(item,containerid,symbol) {
        var _id = (!containerid) ? item.id : containerid+'-'+item.id;

        if(!prx.editor) {

            var _hasRadioButtonsGroupName = typeof(item.radioButtonsGroupName) != 'undefined' && item.radioButtonsGroupName.length > 0;
            //this is checkbox by default
            var _isCheckboxAndRadioGrouped = _hasRadioButtonsGroupName;

            prx.currentPlayerStatus.helper.set(_id, 'active', item.active);

            $('#'+_id+'-flipswitch').on('change.custom-change-event', function(e, updateStatus){

                if(typeof(prx.variables._triggerData['#'+_id+'-flipswitch:checked']) == 'undefined') { prx.variables._triggerData['#'+_id+'-flipswitch:checked'] = {}; }
                prx.variables._triggerData['#'+_id+'-flipswitch:checked']['checkboxchange'] = { state: $(this).is(':checked') };
                if(typeof(prx.variables._triggerData['#'+_id+'-flipswitch:not(:checked)']) == 'undefined') { prx.variables._triggerData['#'+_id+'-flipswitch:not(:checked)'] = {}; }
                prx.variables._triggerData['#'+_id+'-flipswitch:not(:checked)']['checkboxchange'] = { state: $(this).is(':checked') };
                if(typeof(prx.variables._triggerData['#' + _id]) == 'undefined') { prx.variables._triggerData['#' + _id] = {}; }
                prx.variables._triggerData['#' + _id]['checkboxchange'] = { state: $(this).is(':checked') };
                $(this).trigger('checkboxchange');

                if ((updateStatus !== undefined) && !updateStatus) {
                    return;
                }

                if (_isCheckboxAndRadioGrouped) {
                    var screenItems = prx.currentPlayerStatus.helper.get(containerid, 'items');

                    Object.keys(screenItems).forEach(function (key) {
                        if (screenItems[key].radioButtonsGroupName && (screenItems[key].radioButtonsGroupName === item.radioButtonsGroupName)) {
                            prx.currentPlayerStatus.helper.set(containerid + '-' + screenItems[key].id, 'active', false);
                        }
                    });
                }

                prx.currentPlayerStatus.helper.set(_id, 'active', $(this).is(':checked'));
            });

            if(_isCheckboxAndRadioGrouped) {

                $('label[for="'+_id+'-flipswitch"]').on('mousedown', function(event) {
                    var $checkbox = $('#'+_id+'-flipswitch');
                    var checked = $checkbox.prop('checked');
                    $('input[type=\'radio\'][name=\''+$checkbox.attr('name')+'\']').each(function(t){
                        if($(this).is(':checked')){
                            $(this).prop('checked', false).trigger('change', [false]);
                        } else {
                            $(this).prop('checked', false);
                        }
                    });
                    $checkbox.prop('checked', !checked).trigger('change', [true]);
                    event.preventDefault();
                });
                //
                $('label[for="'+_id+'-flipswitch"]').on('click change', function(event) {
                    event.preventDefault();
                });

                $('#'+_id+'-flipswitch').on('click change', function(event) {
                    event.preventDefault();
                });

            }

            $('#'+_id).hammer();
            prx.actions.disableFlashActionOnItemTap('#' + _id, '.flashactiontap-afterdisplay');
        }
    }
    ,interactions: [
        {
            caption: 'Change',
            name: 'actions',
            type: 'action',
            value: function (item, name) {
                if (typeof(item.actions) == 'undefined') {
                    item.actions = [];
                }
                return item.actions.length;
            },
            changeProperty: {
                caption: 'Interactions on change',
                selector: '',
                property: 'action',
                transitionable: false,
                changeable: false
            }
        }, {
            caption: 'Activation',
            name: 'flipswitchActionsOnActive',
            type: 'action',
            value: function (item, name) {
                if (typeof(item.flipswitchActionsOnActive) == 'undefined') {
                    if (typeof(item.actionsOnActive) == 'undefined') {
                        item.flipswitchActionsOnActive = [];
                    } else {
                        item.flipswitchActionsOnActive = item.actionsOnActive;
                    }
                }
                return item.flipswitchActionsOnActive.length;
            }
        }, {
            caption: 'Deactivation',
            name: 'flipswitchActionsOnDeactive',
            type: 'action',
            value: function (item, name) {
                if (typeof(item.flipswitchActionsOnDeactive) == 'undefined') {
                    if (typeof(item.actionsOnDeactive) == 'undefined') {
                        item.flipswitchActionsOnDeactive = [];
                    } else {
                        item.flipswitchActionsOnDeactive = item.actionsOnDeactive;
                    }
                }
                return item.flipswitchActionsOnDeactive.length;
            }
        }
    ]
    ,mpactions: {
        specialEvents: ['checkboxchange'],
    }
    ,propertyGroups: [
        {
            caption: 'Style',
            properties: [
                [
                    {
                        caption: 'Inactive'
                        ,name: 'inactiveBarColor'
                        ,proptype: 'background-2-inactive'
                        ,type: 'gradients-colorpicker'
                        ,value: function(item,name) {
                            return item.inactiveBarColor;
                        }
                        ,values: prx.comps.basicColors
                        ,liveUpdate:'background'
                        ,changeProperty: {
                            transitionable: false,
                            caption: 'Inactive bar color',
                            changeFunction(item, containerid) {
                                let id = (!containerid) ? item.id : containerid + '-' + item.id;
                                let cssSelector = '#' + id + ' input:not(:checked)' + '#' + id + '-flipswitch + label .switch-background ';

                                prx.componentsHelper.addStyleForChangeFunction(id, id + '-inactiveBarColor', cssSelector, 'background', item.inactiveBarColor, 'color-background');
                            }
                        }
                    },
                    {
                        caption: 'Active'
                        ,name: 'activeBarColor'
                        ,proptype: 'background-2-active'
                        ,type: 'gradients-colorpicker'
                        ,value: function(item,name) {
                            return item.activeBarColor;
                        }
                        ,values: prx.comps.basicColors
                        ,liveUpdate:'background'
                        , changeProperty: {
                            transitionable: false,
                            caption: 'Active bar color',
                            changeFunction(item, containerid) {
                                let id = (!containerid) ? item.id : containerid + '-' + item.id;
                                let cssSelector = '#' + id + ' input:checked' + '#' + id + '-flipswitch + label .switch-background ';

                                prx.componentsHelper.addStyleForChangeFunction(id, id + '-activeBarColor', cssSelector, 'background', item.activeBarColor, 'color-background');
                            }
                        }
                    }
                ]
                ,[
                    {
                        caption: 'Border',
                        name: 'borderWidth',
                        proptype: 'border-width',
                        type: 'combo-select',
                        value: function(item, name) {
                            return typeof item.borderWidth == 'undefined' ? '' : item.borderWidth;
                        },
                        values: {min: 0, max: 20, step: 1},
                        changeProperty: {
                            caption: 'Border width',
                            selector: '.changeProperty-borderWidth',
                            property: 'border-width',
                            transitionable: true
                        }
                    }
                    ,prx.commonproperties.borderColor
                    ,{
                        caption: '<span class="icon icon-border-radius" title="Border radius"></span>'
                        ,name: 'borderRadius'
                        ,proptype: 'border-radius'
                        ,type: 'combo-select'
                        ,value: function(item,name) {
                            return item.borderRadius;
                        }
                        ,values: { min: 0, max: 20, step: 2 },
                        expandableType: 'borderRadius',
                        expandedValues: ['tl', 'tr', 'bl', 'br'],
                        changeProperty: {
                            caption: 'Bar border radius',
                            property: 'border-radius',
                            selector: '.changeProperty-borderRadius',
                            transitionable: true
                        }
                    }
                ]
            ]
        }
        ,{
            caption: 'Switch',
            properties: [
                [
                    {
                        caption: 'Size'
                        ,name: 'switchSize'
                        ,proptype: 'switchSize'
                        ,type: 'combo-select'
                        ,value: function(item,name) {
                            return item.switchSize;
                        }
                        ,values: { min: 2, max: 20, step: 2 }
                        ,changeProperty: {
                            caption: 'switchSize',
                            rerender: true,
                            changeable: false
                        }
                    }
                ],[
                    {
                        caption: 'Inactive'
                        ,name: 'inactiveSwitchColor'
                        ,proptype: 'background-inactive'
                        ,type: 'gradients-colorpicker'
                        ,value: function(item,name) {
                            return item.inactiveSwitchColor;
                        }
                        ,values: prx.comps.basicColors
                        ,liveUpdate: 'background'
                        ,changeProperty: {
                            transitionable: false,
                            caption: 'Inactive switch color',
                            changeFunction(item, containerid) {
                                let id = (!containerid) ? item.id : containerid + '-' + item.id;
                                let cssSelector = '#' + id + ' input:not(:checked)' + '#' + id + '-flipswitch + label .switch ';

                                prx.componentsHelper.addStyleForChangeFunction(id, id + '-inactiveSwitchColor', cssSelector, 'background', item.inactiveSwitchColor,'color-background');
                            }
                        }
                    }
                    ,{
                        caption: 'Active'
                        ,name: 'activeSwitchColor'
                        ,proptype: 'background-active'
                        ,type: 'gradients-colorpicker'
                        ,value: function(item,name) {
                            return item.activeSwitchColor;
                        }
                        ,values: prx.comps.basicColors
                        ,liveUpdate: 'background'
                        ,changeProperty: {
                            transitionable: false,
                            caption: 'Active switch color',
                            changeFunction(item, containerid) {
                                let id = (!containerid) ? item.id : containerid + '-' + item.id;
                                let cssSelector = '#' + id + ' input:checked' + '#' + id + '-flipswitch + label .switch ';


                                prx.componentsHelper.addStyleForChangeFunction(id, id + '-activeSwitchColor', cssSelector, 'background', item.activeSwitchColor,'color-background');
                            }
                        }
                    }
                ]
            ]
        }
        ,{
            caption: 'Advanced',
            properties: [
                [
                    {
                        caption: 'Active'
                        ,name: 'active'
                        ,type: 'onoff'
                        ,value: function(item,name) {
                            return item.active;
                        }
                        ,onChange: function(item, name) {
                            return prx.commonproperties.radioButtonsActiveStatusOnChange(item, name);
                        }
                        ,changeProperty: {
                            caption: 'Active',
                            selector: 'input',
                            property: 'checkbox-state',
                            transitionable: false
                        }
                    },
                ],
                [
                    prx.commonproperties.radioButtonsGroupNameInput
                ]
            ]
        }
    ]

};

// TYPE: ON OFF SWITCH 2
prx.types.basic_switch2 = {
    name: 'basic_switch2'
    ,onDisplay: function(item,containerid,symbol) {
        var _id = (!containerid) ? item.id : containerid+'-'+item.id;
        var _dims = prx.componentsHelper.getRealDims(item,symbol);

        var _hasRadioButtonsGroupName = typeof(item.radioButtonsGroupName) != 'undefined' && item.radioButtonsGroupName.length > 0;
        var _type = !_hasRadioButtonsGroupName ? 'checkbox' : 'radio';
        var radioButtonsGroupName = _hasRadioButtonsGroupName ? 'name="' + containerid + '-radioButtonsGroup-' + item.radioButtonsGroupName + '"' : '';

        var cR = '';
        cR += '<div id="' + _id + '" ' + prx.items.getComponentBaseAttributes(item, containerid, symbol)  + ' class="' + prx.items.getComponentBaseClasses(item, containerid, symbol) + ' box pos type-basic-switch">';

        cR += '<style>';
        cR += prx.items.getComponentBaseStyle(item, containerid, symbol);
        cR += '#'+_id+' label .switch-background { width: 100%; height: '+item.barSize+'px; border-radius: '+prx.componentsHelper.getProp(item.borderRadius,'border-radius')+'; border: '+prx.componentsHelper.getProp(item.borderWidth,'num-border-width')+'px solid '+prx.componentsHelper.getProp(item.borderColor,'color-border')+'; background: '+prx.componentsHelper.getProp(item.inactiveBarColor,'color-background')+'; }';
        cR += '#'+_id+' input:checked + label .switch-background { background: '+prx.componentsHelper.getProp(item.activeBarColor,'color-background')+'; }';
        cR += '#'+_id+' .switch { left: '+ ((item.barSize - _dims.height)/2) + 'px; width: '+_dims.height+'px; height: '+_dims.height+'px; background: '+prx.componentsHelper.getProp(item.inactiveSwitchColor,'color-background')+'; border-radius: '+prx.componentsHelper.getProp(item.borderRadius,'border-radius')+'; border: '+prx.componentsHelper.getProp(item.borderWidth,'num-border-width')+'px solid '+prx.componentsHelper.getProp(item.borderColor,'color-border')+'; }';
        cR += '#'+_id+' input:checked + label .switch-background .switch { left: '+(_dims.width - _dims.height - item.borderWidth * 2 - (item.barSize - _dims.height)/2)+'px; background: '+prx.componentsHelper.getProp(item.activeSwitchColor,'color-background')+' }';
        cR += '</style>';
        cR += prx.items.getComponentPrependDivs(item, containerid, symbol);

        cR += '<input type="'+_type+'" '+((prx.componentsHelper.getProp(item.active,'boolean')) ? 'checked' : '')+ ' id="'+_id+'-flipswitch" data-role="none" ' + radioButtonsGroupName + ' />';

        var labelClasses,
            switchClasses;

        if(prx.componentsHelper.getProp(item.active,'boolean')) {
            labelClasses = ' liveUpdate-activeBarColor-background changeProperty-activeBarColor';
            switchClasses = 'liveUpdate-activeSwitchColor-background changeProperty-activeSwitchColor';
        }
        else {

            labelClasses = ' liveUpdate-inactiveBarColor-background changeProperty-inactiveBarColor';
            switchClasses = 'liveUpdate-inactiveSwitchColor-background changeProperty-inactiveSwitchColor';
        }

        cR += '<label for="'+_id+'-flipswitch"><div class="switch-background liveUpdate-borderColor-border-color changeProperty-borderWidth changeProperty-borderColor changeProperty-borderRadius '+ labelClasses +'" data-clickable="true">'; // data-clickable for greensock draggable - if draggable or in draggable container clicking on the flipswitch wont change state

        cR += '<div class="switch liveUpdate-borderColor-border-color changeProperty-borderWidth changeProperty-borderColor changeProperty-borderRadius '+ switchClasses +'"></div>';
        cR += '</div>';
        cR += '</label>';

        cR += prx.items.getComponentAppendDivs(item, containerid, symbol);
        cR += '</div>';

        return cR;
    }
    ,onResize: function(item,containerid,symbol){
        var _id = (!containerid) ? item.id : containerid+'-'+item.id;
        var _dims = prx.componentsHelper.getRealDims(item,symbol);
        $('#' + _id + ' .switch').css('width', _dims.height).css('height', _dims.height);
        if($('#'+_id+'-flipswitch').is(':checked')){
            $('#' + _id + ' label .switch-background .switch').css('left', (_dims.width - _dims.height - item.borderWidth * 2 - (item.barSize - _dims.height)/2));
        } else {
            $('#' + _id + ' label .switch-background .switch').css('left', ((item.barSize - _dims.height)/2));
        }
    }
    ,afterDisplay: function(item,containerid,symbol) {
        var _id = (!containerid) ? item.id : containerid+'-'+item.id;

        if(!prx.editor) {

            var _hasRadioButtonsGroupName = typeof(item.radioButtonsGroupName) != 'undefined' && item.radioButtonsGroupName.length > 0;
            //this is checkbox by default
            var _isCheckboxAndRadioGrouped = _hasRadioButtonsGroupName;

            $('#'+_id+'-flipswitch').on('change.custom-change-event', function(e, updateStatus){

                if(typeof(prx.variables._triggerData['#'+_id+'-flipswitch:checked']) == 'undefined') { prx.variables._triggerData['#'+_id+'-flipswitch:checked'] = {}; }
                prx.variables._triggerData['#'+_id+'-flipswitch:checked']['checkboxchange'] = { state: $(this).is(':checked') };
                if(typeof(prx.variables._triggerData['#'+_id+'-flipswitch:not(:checked)']) == 'undefined') { prx.variables._triggerData['#'+_id+'-flipswitch:not(:checked)'] = {}; }
                prx.variables._triggerData['#'+_id+'-flipswitch:not(:checked)']['checkboxchange'] = { state: $(this).is(':checked') };
                if(typeof(prx.variables._triggerData['#' + _id]) == 'undefined') { prx.variables._triggerData['#' + _id] = {}; }
                prx.variables._triggerData['#' + _id]['checkboxchange'] = { state: $(this).is(':checked') };
                $(this).trigger('checkboxchange');

                if ((updateStatus !== undefined) && !updateStatus) {
                    return;
                }

                if (_isCheckboxAndRadioGrouped) {
                    var screenItems = prx.currentPlayerStatus.helper.get(containerid, 'items');

                    Object.keys(screenItems).forEach(function (key) {
                        if (screenItems[key].radioButtonsGroupName && (screenItems[key].radioButtonsGroupName === item.radioButtonsGroupName)) {
                            prx.currentPlayerStatus.helper.set(containerid + '-' + screenItems[key].id, 'active', false);
                        }
                    });
                }

                prx.currentPlayerStatus.helper.set(_id, 'active', $(this).is(':checked'));
            });

            if(_isCheckboxAndRadioGrouped) {

                $('label[for="'+_id+'-flipswitch"]').on('mousedown', function(event) {
                    var $checkbox = $('#'+_id+'-flipswitch');
                    var checked = $checkbox.prop('checked');
                    $('input[type=\'radio\'][name=\''+$checkbox.attr('name')+'\']').each(function(){
                        if($(this).is(':checked')){
                            $(this).prop('checked', false).trigger('change', [false]);
                        } else {
                            $(this).prop('checked', false);
                        }
                    });
                    $checkbox.prop('checked', !checked).trigger('change', [true]);
                    event.preventDefault();
                });
                //
                $('label[for="'+_id+'-flipswitch"]').on('click change', function(event) {
                    event.preventDefault();
                });

                $('#'+_id+'-flipswitch').on('click change', function(event) {
                    event.preventDefault();
                });

            }

            $('#'+_id).hammer();
            prx.actions.disableFlashActionOnItemTap('#' + _id, '.flashactiontap-afterdisplay');
        }
    }
    ,interactions: [
        {
            caption: 'Change',
            name: 'actions',
            type: 'action',
            value: function (item, name) {
                if (typeof(item.actions) == 'undefined') {
                    item.actions = [];
                }
                return item.actions.length;
            },
            changeProperty: {
                caption: 'Interactions on change',
                selector: '',
                property: 'action',
                transitionable: false,
                changeable: false
            }
        }, {
            caption: 'Activation',
            name: 'flipswitchActionsOnActive',
            type: 'action',
            value: function (item, name) {
                if (typeof(item.flipswitchActionsOnActive) == 'undefined') {
                    if (typeof(item.actionsOnActive) == 'undefined') {
                        item.flipswitchActionsOnActive = [];
                    } else {
                        item.flipswitchActionsOnActive = item.actionsOnActive;
                    }
                }
                return item.flipswitchActionsOnActive.length;
            }
        }, {
            caption: 'Deactivation',
            name: 'flipswitchActionsOnDeactive',
            type: 'action',
            value: function (item, name) {
                if (typeof(item.flipswitchActionsOnDeactive) == 'undefined') {
                    if (typeof(item.actionsOnDeactive) == 'undefined') {
                        item.flipswitchActionsOnDeactive = [];
                    } else {
                        item.flipswitchActionsOnDeactive = item.actionsOnDeactive;
                    }
                }
                return item.flipswitchActionsOnDeactive.length;
            }
        }
    ]
    ,mpactions: {
        specialEvents: ['checkboxchange'],
    }
    ,propertyGroups: [
        {
            caption: 'Style',
            properties: [
                [
                    {
                        caption: 'Size'
                        ,name: 'barSize'
                        ,proptype: 'barSize'
                        ,type: 'combo-select'
                        ,value: function(item,name) {
                            return item.barSize;
                        }
                        ,values: { min: 2, max: 20, step: 2 }
                        ,changeProperty: {
                            caption: 'barSize',
                            rerender: true,
                            changeable: false
                        }
                    }
                ],[
                    {
                        caption: 'Inactive'
                        ,name: 'inactiveBarColor'
                        ,proptype: 'background-2-inactive'
                        ,type: 'gradients-colorpicker'
                        ,value: function(item,name) {
                            return item.inactiveBarColor;
                        }
                        ,values: prx.comps.basicColors
                        ,liveUpdate:'background'
                        ,changeProperty: {
                            transitionable: false,
                            caption: 'Inactive bar color',
                            changeFunction(item, containerid) {
                                let id = (!containerid) ? item.id : containerid + '-' + item.id;
                                let cssSelector = '#' + id + ' input:not(:checked)' + '#' + id + '-flipswitch + label .switch-background ';

                                prx.componentsHelper.addStyleForChangeFunction(id, id + '-inactiveBarColor', cssSelector, 'background',item.inactiveBarColor, 'color-background');
                            }
                        }
                    },
                    {
                        caption: 'Active'
                        ,name: 'activeBarColor'
                        ,proptype: 'background-2-active'
                        ,type: 'gradients-colorpicker'
                        ,value: function(item,name) {
                            return item.activeBarColor;
                        }
                        ,values: prx.comps.basicColors
                        ,liveUpdate:'background'
                        ,changeProperty: {
                            caption: 'Active bar color',
                            transitionable: false,
                            changeFunction(item, containerid) {
                                let id = (!containerid) ? item.id : containerid + '-' + item.id;
                                let cssSelector = '#' + id + ' input:checked' + '#' + id + '-flipswitch + label .switch-background ';

                                prx.componentsHelper.addStyleForChangeFunction(id, id + '-activeBarColor', cssSelector, 'background', item.activeBarColor,'color-background');
                            }
                        }
                    }
                ]
                ,[
                    prx.commonproperties.borderWidth
                    ,prx.commonproperties.borderColor
                    ,{
                        caption: '<span class="icon icon-border-radius" title="Border radius"></span>'
                        ,name: 'borderRadius'
                        ,proptype: 'border-radius'
                        ,type: 'combo-select'
                        ,value: function(item,name) {
                            return item.borderRadius;
                        }
                        ,values: { min: 0, max: 20, step: 2 },
                        expandableType: 'borderRadius',
                        expandedValues: ['tl', 'tr', 'bl', 'br'],
                        changeProperty: {
                            caption: 'Bar border radius',
                            property: 'border-radius',
                            selector: '.changeProperty-borderRadius',
                            transitionable: true
                        }
                    }
                ]
            ]
        }
        ,{
            caption: 'Switch',
            properties: [
                [
                    {
                        caption: 'Inactive'
                        ,name: 'inactiveSwitchColor'
                        ,proptype: 'background-inactive'
                        ,type: 'gradients-colorpicker'
                        ,value: function(item,name) {
                            return item.inactiveSwitchColor;
                        }
                        ,values: prx.comps.basicColors
                        ,liveUpdate: 'background'
                        ,changeProperty: {
                            caption: 'Inactive switch color',
                            transitionable: false,
                            changeFunction(item, containerid) {
                                let id = (!containerid) ? item.id : containerid + '-' + item.id;
                                let cssSelector = '#' + id + ' input:not(:checked)' + '#' + id + '-flipswitch + label .switch ';

                                prx.componentsHelper.addStyleForChangeFunction(id, id + '-inactiveSwitchColor', cssSelector, 'background', item.inactiveSwitchColor,'color-background');
                            }
                        }
                    }
                    ,{
                        caption: 'Active'
                        ,name: 'activeSwitchColor'
                        ,proptype: 'background-active'
                        ,type: 'gradients-colorpicker'
                        ,value: function(item,name) {
                            return item.activeSwitchColor;
                        }
                        ,values: prx.comps.basicColors
                        ,liveUpdate: 'background'
                        ,changeProperty: {
                            caption: 'Active switch color',
                        transitionable: false,
                            changeFunction(item, containerid) {
                                let id = (!containerid) ? item.id : containerid + '-' + item.id;
                                let cssSelector = '#' + id + ' input:checked' + '#' + id + '-flipswitch + label .switch ';

                                prx.componentsHelper.addStyleForChangeFunction(id, id + '-activeSwitchColor', cssSelector, 'background', item.activeSwitchColor, 'color-background');
                            }
                        }
                    }
                ]
            ]
        }
        ,{
            caption: 'Advanced',
            properties: [
                [
                    {
                        caption: 'Active'
                        ,name: 'active'
                        ,type: 'onoff'
                        ,value: function(item,name) {
                            return item.active;
                        }
                        ,onChange: function(item, name) {
                            return prx.commonproperties.radioButtonsActiveStatusOnChange(item, name);
                        }
                        ,changeProperty: {
                            caption: 'Active',
                            selector: 'input',
                            property: 'checkbox-state',
                            transitionable: false
                        }
                    },
                ],
                [
                    prx.commonproperties.radioButtonsGroupNameInput
                ]
            ]
        }
    ]

};

// TYPE: CHECKBOX
prx.types.basic_checkbox = {
    name: 'basic_checkbox'
    ,onDisplay: function(item,containerid,symbol) {
        var _id = (!containerid) ? item.id : containerid+'-'+item.id;
        var _dims = prx.componentsHelper.getRealDims(item,symbol);
        var cR = '';

        if(typeof(item.radioButtonsGroupName) == 'undefined') {
            item.radioButtonsGroupName = '';
        }

        var _hasRadioButtonsGroupName = typeof(item.radioButtonsGroupName) != 'undefined' && item.radioButtonsGroupName.length > 0;
        var _type = !_hasRadioButtonsGroupName ? 'checkbox' : 'radio';
        var radioButtonsGroupName = _hasRadioButtonsGroupName ? 'name="' + containerid + '-radioButtonsGroup-' + item.radioButtonsGroupName + '"' : '';

        var _props = (typeof(prx.richtext)=='undefined') ? prx.componentsHelper.getProp(item.textProperties,'props-text') : '';
        var text = '<div class="basic-checkbox-text liveUpdate-textColor-color "><span data-editableproperty="text" class="changeProperty-textColor changeProperty-textFont changeProperty-textFontStyle changeProperty-textSize changeProperty-textProperties">'+prx.componentsHelper.getProp(item.text,'text-textarea')+'</span></div>';

        cR += '<div id="' + _id + '" ' + prx.items.getComponentBaseAttributes(item, containerid, symbol)  + ' class="' + prx.items.getComponentBaseClasses(item, containerid, symbol) + ' box pos type-basic-checkbox">';

        cR += '<style>';
        cR += prx.items.getComponentBaseStyle(item, containerid, symbol);
        cR += '#'+_id+' label { line-height: '+item.textSize+'px}';
        cR += '#'+_id+' label .basic-checkbox-text { width: calc(100% - '+parseInt(item.checkboxSize,10)+'px - ' + (12 * prx.componentsHelper.getScale(item.lib)) + 'px); margin-left: '+(parseInt(item.checkboxSize,10)+12* prx.componentsHelper.getScale(item.lib))+'px;  '+prx.componentsHelper.getProp(item.textFont + '|' + item.textFontStyle,'font-family') + prx.componentsHelper.getProp(item.textFontStyle,'font-style')+' color: '+prx.componentsHelper.getProp(item.textColor,'color-text')+'; font-size: '+prx.componentsHelper.getProp(item.textSize,'num-text-size')+'px; '+_props+' }';
        cR += '#'+_id+' label .basic-checkbox-inner { top: '+((parseInt(item.textSize,10)-parseInt(item.checkboxSize,10))/2)+'px; width: '+item.checkboxSize+'px; height: '+item.checkboxSize+'px; min-height: '+item.checkboxSize+'px; max-height: '+item.checkboxSize+'px; min-width: '+item.checkboxSize+'px; max-width: '+item.checkboxSize+'px; background: '+prx.componentsHelper.getProp(item.backgroundColor,'color-background')+'; border: '+prx.componentsHelper.getProp(item.borderWidth,'num-border-width')+'px solid '+prx.componentsHelper.getProp(item.borderColor,'color-border')+'; border-radius: '+prx.componentsHelper.getProp(item.borderRadius,'border-radius')+'; }';
        cR += '#'+_id+' input:checked + label .basic-checkbox-inner { background: '+prx.componentsHelper.getProp(item.activeBackgroundColor,'color-background')+'; border: '+prx.componentsHelper.getProp(item.borderWidth,'num-border-width')+'px solid '+prx.componentsHelper.getProp(item.borderColor,'color-border')+'; border-radius: '+prx.componentsHelper.getProp(item.borderRadius,'border-radius')+'; }';
        cR += '</style>';
        cR += prx.items.getComponentPrependDivs(item, containerid, symbol);

        cR += '<input type="'+_type+'" id="'+_id+'-checkbox" '+((prx.componentsHelper.getProp(item.active,'boolean')) ? 'checked="checked"' : '') + ' data-role="none" ' + radioButtonsGroupName + '/>';
        cR += '<label for="'+_id+'-checkbox" data-clickable="true"><div class="basic-checkbox-inner '+((prx.componentsHelper.getProp(item.active,'boolean')) ? 'liveUpdate-activeBackgroundColor-background-color changeProperty-activeBackgroundColor' : 'liveUpdate-backgroundColor-background-color changeProperty-backgroundColor') + ' liveUpdate-borderColor-border-color changeProperty-borderColor changeProperty-borderWidth changeProperty-borderRadius">';
        cR += '<img class="basic-checkbox-icon" src="'+prx.componentsHelper.getProp(item.img,'asset')+'">';
        cR += '</div>';
        cR += text;
        cR += '</label>';
        cR += prx.items.getComponentAppendDivs(item, containerid, symbol);
        cR += '</div>';
        return cR;
    }
    ,afterDisplay: function(item,containerid,symbol) {
        var _id = (!containerid) ? item.id : containerid+'-'+item.id;

        var _hasRadioButtonsGroupName = typeof(item.radioButtonsGroupName) != 'undefined' && item.radioButtonsGroupName.length > 0;
        //checkbox by default
        var _isCheckboxAndRadioGrouped = _hasRadioButtonsGroupName;
        if(!prx.editor) {

            $('#'+_id+'-checkbox').on('change.custom-change-event', function(e, updateStatus){
                if(typeof(prx.variables._triggerData['input:checked[id='+_id+'-checkbox]']) == 'undefined') { prx.variables._triggerData['input:checked[id='+_id+'-checkbox]'] = {}; }
                prx.variables._triggerData['input:checked[id='+_id+'-checkbox]']['checkboxchange'] = { state: $(this).is(':checked') };
                if(typeof(prx.variables._triggerData['input[id='+_id+'-checkbox]:not(:checked)']) == 'undefined') { prx.variables._triggerData['input[id='+_id+'-checkbox]:not(:checked)'] = {}; }
                prx.variables._triggerData['input[id='+_id+'-checkbox]:not(:checked)']['checkboxchange'] = { state: $(this).is(':checked') };
                if(typeof(prx.variables._triggerData['#' + _id]) == 'undefined') { prx.variables._triggerData['#' + _id] = {}; }
                prx.variables._triggerData['#' + _id]['checkboxchange'] = { state: $(this).is(':checked') };
                $(this).trigger('checkboxchange');

                if ((updateStatus !== undefined) && !updateStatus) {
                    return;
                }

                if (_isCheckboxAndRadioGrouped) {
                    var screenItems = prx.currentPlayerStatus.helper.get(containerid, 'items');

                    Object.keys(screenItems).forEach(function (key) {
                        if (screenItems[key].radioButtonsGroupName && (screenItems[key].radioButtonsGroupName === item.radioButtonsGroupName)) {
                            prx.currentPlayerStatus.helper.set(containerid + '-' + screenItems[key].id, 'active', false);
                        }
                    });
                }

                prx.currentPlayerStatus.helper.set(_id, 'active', $(this).is(':checked'));
            });

            if(_isCheckboxAndRadioGrouped) {
                $('label[for="'+_id+'-checkbox"]').on('mousedown touchstart', function(event) {
                    var $checkbox = $('#'+_id+'-checkbox');
                    var checked = $checkbox.prop('checked');
                    $('input[type=\'radio\'][name=\''+$checkbox.attr('name')+'\']').each(function(){
                        if($(this).is(':checked')){
                            $(this).prop('checked', false).trigger('change', [false]);
                        } else {
                            $(this).prop('checked', false);
                        }
                    });
                    $checkbox.prop('checked', !checked).trigger('change', [true]);
                    event.preventDefault();
                });

                $('label[for="'+_id+'-checkbox"]').hammer().on('click tap', function(event) {
                    event.preventDefault();
                });

                $('#'+_id+'-checkbox').on('change', function(event) {
                    event.preventDefault();
                });


            }
            $('#'+_id).hammer();
            prx.actions.disableFlashActionOnItemTap('#' + _id, '.flashactiontap-afterdisplay');
        }
    }
    ,interactions: [
        {
            caption: 'Change',
            name: 'actions',
            type: 'action',
            value: function (item, name) {
                if (typeof(item.actions) == 'undefined') {
                    item.actions = [];
                }
                return item.actions.length;
            },
            changeProperty: {
                caption: 'Interactions on change',
                selector: '',
                property: 'action',
                transitionable: false,
                changeable: false
            }
        },
        {
            caption: 'Activation',
            name: 'checkboxActionsOnActive',
            type: 'action',
            value: function (item, name) {
                if (typeof(item.checkboxActionsOnActive) == 'undefined') {
                    item.checkboxActionsOnActive = [];
                }
                return item.checkboxActionsOnActive.length;
            }
        },
        {
            caption: 'Deactivation',
            name: 'checkboxActionsOnDeactive',
            type: 'action',
            value: function (item, name) {
                if (typeof(item.checkboxActionsOnDeactive) == 'undefined') {
                    item.checkboxActionsOnDeactive = [];
                }
                return item.checkboxActionsOnDeactive.length;
            }
        }
    ]
    ,mpactions: {
        specialEvents: ['checkboxchange']
    }
    ,editableProperties: [
        {
            caption: 'Text'
            ,name: 'text'
            ,type: 'textarea'
            ,isRichText: false
            ,value: function(item,name) {
                return item.text;
            }
            ,changeProperty: {
                caption: 'Text',
                property: 'text',
                selector: '.basic-checkbox-text',
                transitionable: false
            }
        }
    ]
    ,propertyGroups: [
        {
            caption: 'Style',
            properties: [
                [
                    {
                        caption: 'Checkbox size',
                        name: 'checkboxSize',
                        proptype: 'checkbox-size',
                        type: 'combo-select',
                        values: { min: 16, max: 50, step: 2 },
                        value: function(item,name) {
                            return item.checkboxSize;
                        },
                        changeProperty: {
                            caption: 'Checkbox size',
                            rerender: true
                        }
                    }
                ],
                [
                    {
                        caption: 'Background',
                        name: 'backgroundColor',
                        proptype: 'background-color',
                        type: 'gradients-colorpicker',
                        value: function(item,name) { return item.backgroundColor; },
                        values: prx.comps.basicColors,
                        liveUpdate: 'background-color',
                        changeProperty: {
                            caption: 'Background color',
                            transitionable: false,
                            changeFunction(item, containerid) {
                                let id = (!containerid) ? item.id : containerid + '-' + item.id;
                                let cssSelector = '#' + id + ' input:not(:checked)' + '#' + id + '-checkbox + label .basic-checkbox-inner  ';

                                prx.componentsHelper.addStyleForChangeFunction(id, id + '-backgroundColor', cssSelector, 'background', item.backgroundColor,'color-background');
                            }
                        }
                    },
                    {
                        caption: 'Active',
                        name: 'activeBackgroundColor',
                        proptype: 'background-color-2-active',
                        type: 'gradients-colorpicker',
                        value: function(item,name) { return item.activeBackgroundColor; },
                        values: prx.comps.basicColors,
                        liveUpdate: 'background-color',
                        changeProperty: {
                            caption: 'Active background color',
                            transitionable: false,
                            changeFunction(item, containerid) {
                                let id = (!containerid) ? item.id : containerid + '-' + item.id;
                                let cssSelector = '#' + id + ' input:checked' + '#' + id + '-checkbox + label .basic-checkbox-inner ';

                                prx.componentsHelper.addStyleForChangeFunction(id, id + '-activeBackgroundColor', cssSelector, 'background', item.activeBackgroundColor,'color-background');
                            }
                        }
                    }
                ],
                [
                    prx.commonproperties.borderWidth
                    ,prx.commonproperties.borderColor
                    ,prx.commonproperties.borderRadius
                ]
            ]
        },
        {
            caption: 'Text',
            properties: [
                [
                    prx.commonproperties.textFont
                ],
                [
                    prx.commonproperties.textFontStyle,
                    prx.commonproperties.textSize
                ],
                [
                    prx.commonproperties.textPropertiesRichText(['font-weight','font-style','text-decoration'],'text', false, {propName: 'textProperties', caption: 'Underline', useAsDynProp: false}),
                    {
                        caption: { label: 'Color', class: 'text-properties-label text-color-label' },
                        name: 'textColor',
                        proptype: 'font-color',
                        type: 'solid-colorpicker',
                        relatedEditableProperties: 'text',
                        relatedCSSProperties: 'color',
                        pffSettings: prx.commonproperties.pffSettingsColor,
                        value: function(item,name) {
                            if(typeof(item.textColor) == 'undefined') { item.textColor = '#2E2E2E'; }
                            return item.textColor;
                        },
                        liveUpdate: 'color'
                        ,changeProperty: {
                            caption: ' Text color',
                            selector: '.changeProperty-textColor',
                            property: 'color',
                            transitionable: true
                        }
                    }
                ]
            ]
        },
        {
            caption: 'Icon',
            properties: [[
                {
                    caption: false
                    ,name: 'img'
                    ,type: 'combo-asset'
                    ,displayValue: function(item,name,index) {
                        if(item.img.fileId == '') {
                            return 'No icon selected';
                        }
                        return item.img.name;
                    }
                    ,value: function(item,name,index) {
                        return JSON.stringify({
                            allow: 'image',
                            asset: item.img
                        });
                    }
                    ,changeProperty: {
                        caption: 'Icon',
                        transitionable: false,
                        changeFunction(item, containerid) {
                            let id = (!containerid) ? item.id : containerid + '-' + item.id;
                            let cssSelector = '#' + id + ' label img.basic-checkbox-icon ';

                            prx.componentsHelper.changeImageForChangeFunction(cssSelector, item.img);
                        }
                    }
                }
            ]
            ]
        },
        {
            caption: 'Advanced',
            properties: [
                [
                    {
                        caption: 'Active'
                        ,name: 'active'
                        ,type: 'onoff'
                        ,onChange: function(item, name) {
                            return prx.commonproperties.radioButtonsActiveStatusOnChange(item, name);
                        }
                        ,value: function(item,name) {
                            return item.active;
                        }
                        ,changeProperty: {
                            caption: 'Active state',
                            transitionable: false,
                            changeFunction(item, containerid) {
                                let id = (!containerid) ? item.id : containerid + '-' + item.id;
                                $('#' + id + ' input')[0].checked = item.active;
                            }
                        }

                    }
                ],
                [
                    prx.commonproperties.radioButtonsGroupNameInput
                ]
            ]
        }
    ]
};

// TYPE: SEARCH BAR
prx.types.basic_searchbar = {
    name: 'basic_searchbar'
    ,onDisplay: function(item,containerid,symbol) {
        var _id = (!containerid) ? item.id : containerid+'-'+item.id;

        var _props = prx.componentsHelper.getProp(item.textProperties,'props-text');

        var _dims = prx.componentsHelper.getRealDims(item, symbol);

        var cR = '';
        cR += '<div id="' + _id + '" ' + prx.items.getComponentBaseAttributes(item, containerid, symbol)  + ' class="' + prx.items.getComponentBaseClasses(item, containerid, symbol) + ' box pos type-basic-searchbar type-basic-searchbar-text textAlign-'+prx.componentsHelper.getProp(item.textAlign,'align')+'">';

        cR += '<style>';
        cR += prx.items.getComponentBaseStyle(item, containerid, symbol);
        cR += '#'+_id+'  input { color: '+prx.componentsHelper.getProp(item.textColor,'color-text')+'; font-size: '+prx.componentsHelper.getProp(item.textSize,'num-text-size')+'px;  '+prx.componentsHelper.getProp(item.textFont + '|' + item.textFontStyle,'font-family') + prx.componentsHelper.getProp(item.textFontStyle,'font-style')+' '+ _props + ' ;}';
        cR += '#'+_id+' .basic-searchbar-faux-input { color: '+prx.componentsHelper.getProp(item.textColor,'color-text')+'; font-size: '+prx.componentsHelper.getProp(item.textSize,'num-text-size')+'px;  '+prx.componentsHelper.getProp(item.textFont + '|' + item.textFontStyle,'font-family') + prx.componentsHelper.getProp(item.textFontStyle,'font-style')+' line-height: '+_dims.height+'px; '+ _props + ' }';

        cR += '#'+_id+' .basic-searchbar-faux-input-overlay .placeholder-input, #'+_id+' .basic-searchbar-input-overlay .placeholder-input { color: '+prx.componentsHelper.getProp(item.placeholderColor,'color-text')+'; font-size: '+prx.componentsHelper.getProp(item.textSize,'num-text-size')+'px;  '+prx.componentsHelper.getProp(item.textFont + '|' + item.textFontStyle,'font-family') + prx.componentsHelper.getProp(item.textFontStyle,'font-style')+' '+ _props + ' }';
        if(prx.componentsHelper.getProp(item.imgSrc.fileId,'other') != '') {
            cR += '#' + _id + ' .basic-searchbar-icon { width: '+prx.componentsHelper.getProp(item.textSize,'num-text-size')+'px; background-image: url('+prx.componentsHelper.getProp(item.imgSrc,'asset')+');}';
            cR += '#' + _id + ' .basic-searchbar-icon-faux { width: '+prx.componentsHelper.getProp(item.textSize,'num-text-size')+'px;}';
        }
        cR += '#'+_id+' .basic-searchbar-input-flex { background: '+prx.componentsHelper.getProp(item.backgroundColor,'color-background')+'; border-width: '+prx.componentsHelper.getProp(item.borderWidth,'border-width')+'; border-style: '+prx.componentsHelper.getProp(item.borderStyle,'border-type')+'; border-color: '+prx.componentsHelper.getProp(item.borderColor,'color-border')+'; border-radius: '+prx.componentsHelper.getProp(item.borderRadius,'border-radius')+'; text-align: ' + prx.componentsHelper.getProp(item.textAlign, 'align') + '}';
        cR += '#'+_id+' .basic-searchbar-input-flex input { text-align: '+ (prx.componentsHelper.getProp(item.textAlign,'align') === 'right' ? 'right' : 'left') +' }';
        cR += '#' + _id + ' .basic-searchbar-icon-erase { width: '+(_dims.height*0.4)+'px; background-image: url('+prx.componentsHelper.getProp(item.imgSrc2,'asset')+'); color: '+prx.componentsHelper.getProp(item.placeholderColor,'color-text')+';}';
        cR += '</style>';
        cR += prx.items.getComponentPrependDivs(item, containerid, symbol);

        if(prx.editor) {
            cR += '<div class="basic-searchbar-input-flex liveUpdate-backgroundColor-background-color liveUpdate-borderColor-border-color liveUpdate-borderWidth-border-width changeProperty-backgroundColor changeProperty-borderColor changeProperty-borderWidth changeProperty-borderRadius">';
            if(prx.componentsHelper.getProp(item.imgSrc.fileId,'other') != '') {
                cR += '<span class="basic-searchbar-icon-faux"></span>';
            }
            cR += '<div class="basic-searchbar-faux-input liveUpdate-textColor-color changeProperty-textColor changeProperty-textSize changeProperty-textFont" data-editableproperty="value">'+prx.componentsHelper.getProp(item.value,'text')+'</div>';
            cR += '<div class="basic-searchbar-faux-input-overlay liveUpdate-backgroundColor-background-color">'+(prx.componentsHelper.getProp(item.imgSrc.fileId,'other') != '' ? '<span class="basic-searchbar-icon"></span>': '')+'<span class="liveUpdate-placeholderColor-color placeholder-input">'+prx.componentsHelper.getProp(item.placeholder,'other')+'</span></div>';
            cR += '<span class="basic-searchbar-icon-erase"></span>';
            cR += '</div>';
        } else {
            cR += '<div class="basic-searchbar-input-flex liveUpdate-backgroundColor-background-color liveUpdate-borderColor-border-color liveUpdate-borderWidth-border-width changeProperty-backgroundColor  changeProperty-borderColor changeProperty-borderWidth changeProperty-borderRadius">';
            if(prx.componentsHelper.getProp(item.imgSrc.fileId,'other') != '') {
                cR += '<span class="basic-searchbar-icon-faux"></span>';
            }
            cR += '<input autocomplete="new-password" data-lpignore="true" type="text" value="'+prx.componentsHelper.getProp(item.value,'other')+'" data-role="none" required class="real-input changeProperty-textColor changeProperty-textSize changeProperty-textFont changeProperty-textAlign" />';
            cR += '<div class="basic-searchbar-input-overlay">'+(prx.componentsHelper.getProp(item.imgSrc.fileId,'other') != '' ? '<span class="basic-searchbar-icon"></span>': '')+'<span class="placeholder-input changeProperty-textFont changeProperty-textSize changeProperty-textAlign">'+prx.componentsHelper.getProp(item.placeholder,'other')+'</span></div>';
            cR += '<span class="basic-searchbar-icon-erase"></span>';
            cR += '</div>';
        }
        cR += prx.items.getComponentAppendDivs(item, containerid, symbol);
        cR += '</div>';
        return cR;
    }
    ,onResize: function(item,containerid,symbol) {
        var _id = (!containerid) ? item.id : containerid+'-'+item.id;
        var _dims = prx.componentsHelper.getRealDims(item,symbol);

        if(prx.componentsHelper.getProp(item.imgSrc.fileId,'other') != '') {
            $('#' + _id + ' .basic-searchbar-icon, #' + _id + ' .basic-searchbar-icon-faux').css('width', prx.componentsHelper.getProp(item.textSize,'num-text-size')+'px');
        }
        $('#'+_id).find('.basic-searchbar-icon-erase').css('width', (_dims.height*0.4)+'px');
        $('#'+_id).find('.basic-searchbar-faux-input').css('line-height', _dims.height+'px');
    }
    ,afterDisplay: function(item,containerid,symbol) {
        var _id = (!containerid) ? item.id : containerid+'-'+item.id;
        var _dims = prx.componentsHelper.getRealDims(item, symbol);


        if(!prx.editor) {
            $('#'+_id)
                .hammer()
                .find('.real-input')
                .focus(function(){
                    if(typeof(prx.variables._triggerData['#'+_id]) == 'undefined') { prx.variables._triggerData['#'+_id] = {}; }
                    prx.variables._triggerData['#'+_id]['inputfocus'] = { value: $(this).val() };
                    $('#'+_id).trigger('inputfocus');
                })
                .blur(function(){
                    if(typeof(prx.variables._triggerData['#'+_id]) == 'undefined') { prx.variables._triggerData['#'+_id] = {}; }
                    prx.variables._triggerData['#'+_id]['inputblur'] = { value: $(this).val() };
                    $('#'+_id).trigger('inputblur');
                })
                .keyup(function(e){
                    if(typeof(prx.variables._triggerData['#'+_id]) == 'undefined') { prx.variables._triggerData['#'+_id] = {}; }
                    prx.variables._triggerData['#'+_id]['inputkeyup'] = { value: $(this).val() };
                    var event = $.Event('inputkeyup');
                    event.which = e.which;
                    $('#'+_id).trigger(event);
                })
                .on('input', function(e){
                    if($(this).val() == '') {
                        $('#'+_id+' input + div .placeholder-input').css('display', 'block');
                    } else {
                        $('#'+_id+' input + div .placeholder-input').css('display', 'none');
                    }
                    prx.currentPlayerStatus.helper.set(_id, 'value', e.target.value);
                }).trigger('input');

            prx.actions.disableFlashActionOnItemTap('#' + _id, '.flashactiontap-afterdisplay');

            $('#' + _id + ' .basic-searchbar-icon-erase').hammer().on('tap', function() {
                $('#'+_id+' .real-input').val('').focus().trigger('input');
            });
            prx.actions.disableFlashActionOnItemTap('#' + _id + ' .basic-searchbar-icon-erase', '.flashactiontap-afterdisplay');
        }
    }
    ,interactions: [ prx.commonproperties.actions ]
    ,mpactions: {
        specialEvents: ['inputfocus','inputblur','inputkeyup']
    }
    ,editableProperties: [
        {
            caption: 'Value'
            ,name: 'value'
            ,type: 'input'
            ,isRichText: false
            ,value: function(item,name) {
                return item.value;
            }
            ,changeProperty: {
                caption: 'Value',
                property: 'input-value',
                selector: 'input.real-input',
                transitionable: false,
                changeFunction: function(item, containerid) {
                    var _id = (!containerid) ? item.id : containerid + '-' + item.id;

                    if ($.trim(item.value) == '') {
                        $('#' + _id + ' input + div .placeholder-input').css('display', 'block');
                    } else {
                        $('#' + _id + ' input + div .placeholder-input').css('display', 'none');
                    }

                    $('#' + _id + ' input.real-input').val(item.value);
                }
            }
        }
    ]
    ,propertyGroups: [
        {
            caption: 'Style',
            properties: [
                [
                    {
                        caption: 'Background',
                        name: 'backgroundColor',
                        proptype: 'background-color',
                        type: 'gradients-colorpicker',
                        value: function(item, name) {
                            return typeof item.backgroundColor == 'undefined' ? '' : item.backgroundColor;
                        },
                        liveUpdate: 'background-color',
                        changeProperty: {
                            caption: 'Background color',
                            selector: '.changeProperty-backgroundColor',
                            property: 'background-color',
                            transitionable: true
                        }
                    }
                ]
                ,[
                    prx.commonproperties.borderWidthExpandable
                    ,{
                        caption: false,
                        name: 'borderStyle',
                        proptype: 'border-style',
                        type: 'select',
                        value: function(item,name) {
                            return item.borderStyle;
                        },
                        values: [{ value: 'solid', displayValue: 'Solid'},{ value: 'dotted', displayValue: 'Dotted'},{ value: 'dashed', displayValue: 'Dashed'},{ value: 'double', displayValue: 'Double'},{ value: 'none', displayValue: 'None'}],
                        changeProperty: {
                            caption: 'Border Style',
                            selector: '.basic-searchbar-input-flex',
                            property: 'border-style',
                            transitionable: false
                        }
                    }
                    ,prx.commonproperties.borderColor //ios7BorderColor
                    ,prx.commonproperties.borderRadiusExpandable
                ]
            ]
        },
        {
            caption: 'Text',
            properties: [
                [
                    prx.commonproperties.textFont
                ],
                [
                    prx.commonproperties.textFontStyle,
                    prx.commonproperties.textSize
                ],
                [
                    prx.commonproperties.textProperties
                    ,{
                        caption: { label: 'Color', class: 'text-properties-label text-color-label' },
                        name: 'textColor',
                        proptype: 'font-color',
                        type: 'solid-colorpicker',
                        relatedEditableProperties: 'text',
                        relatedCSSProperties: 'color',
                        pffSettings: prx.commonproperties.pffSettingsColor,
                        value: function(item,name) {
                            if(typeof(item.textColor) == 'undefined') { item.textColor = '#2E2E2E'; }
                            return item.textColor;
                        },
                        liveUpdate: 'color'
                        ,changeProperty: {
                            caption: ' Text color',
                            selector: '.changeProperty-textColor',
                            property: 'color',
                            transitionable: true
                        }
                    }
                ],
                [
                    prx.commonproperties.textAlign
                ]
            ]
        },
        {
            caption: 'Placeholder (If field is empty)',
            properties: [
                [
                    {
                        caption: 'Placeholder Text',
                        name: 'placeholder',
                        type: 'input',
                        value: function(item,name) {
                            return item.placeholder;
                        }
                        ,changeProperty: {
                            caption: 'Placeholder text',
                            transitionable: false,
                            changeFunction(item, containerid) {
                                let id = (!containerid) ? item.id : containerid + '-' + item.id;
                                let cssSelector = '#' + id + ' .basic-searchbar-input-overlay .placeholder-input';

                                if (prx.editor) {
                                    $('#' + id + ' .basic-searchbar-faux-input').text(prx.componentsHelper.getProp(item.placeholder, 'other'));
                                } else {
                                    $(cssSelector).text(prx.componentsHelper.getProp(item.placeholder,'other'));
                                }
                            }
                        }
                    }
                ],
                [
                    {
                        caption: 'Placeholder Color',
                        name: 'placeholderColor',
                        proptype: 'placeholder-color',
                        type: 'combo-colorpicker',
                        value: function(item,name) {
                            if(typeof(item.placeholderColor)=='undefined') { return '999999'; }
                            return item.placeholderColor;
                        }
                        ,values: prx.comps.basicColors //ios7Colors
                        ,liveUpdate:'color'
                        ,changeProperty: {
                            caption: 'Placeholder color',
                            transitionable: false,
                            changeFunction(item, containerid) {
                                let id = (!containerid) ? item.id : containerid + '-' + item.id;
                                let cssSelector = '#' + id;

                                if (prx.editor) {
                                    prx.componentsHelper.addStyleForChangeFunction(id, id + '-placeholderColor', cssSelector + ' .basic-searchbar-input-flex .basic-searchbar-faux-input-overlay .placeholder-input', 'color', item.placeholderColor, 'color-text');
                                    //for the scenario where placeholder text is also changed.
                                    prx.componentsHelper.addStyleForChangeFunction(id, id + '-placeholderColor', cssSelector + ' .basic-searchbar-input-flex .basic-searchbar-faux-input', 'color', item.placeholderColor, 'color-text');
                                } else {
                                    prx.componentsHelper.addStyleForChangeFunction(id, id + '-placeholderColor', cssSelector + ' .basic-searchbar-input-overlay .placeholder-input', 'color', item.placeholderColor, 'color-text');
                                }
                            }
                        }
                    }
                ]
            ]
        },
        {
            caption: 'Icon',
            properties: [
                [
                    {
                        caption: false,
                        name: 'imgSrc',
                        type: 'combo-asset',
                        displayValue: function(item,name) {
                            if(item.imgSrc.fileId == '') {
                                return 'No asset selected.';
                            }
                            return item.imgSrc.name;
                        }
                        ,value: function(item,name) {
                            return JSON.stringify({
                                allow: 'image',
                                asset: item.imgSrc
                            });
                        }
                        ,changeProperty: {
                            caption: 'Icon',
                            transitionable: false,
                            changeFunction(item, containerid) {
                                let id = (!containerid) ? item.id : containerid + '-' + item.id;
                                let cssSelector = '#' + id + ' .basic-searchbar-icon ';

                                prx.componentsHelper.changeImageForChangeFunction(cssSelector, item.imgSrc, true);
                            }
                        }
                    }
                ]
            ]
        }
    ]

};

//TYPE: PROGRESSBAR
prx.types.basic_progressbar = {
    name: 'basic_progressbar'
    ,onDisplay: function(item,containerid,symbol) {
        var _id = (!containerid) ? item.id : containerid+'-'+item.id;

        var cR = '';
        cR += '<div id="' + _id + '" ' + prx.items.getComponentBaseAttributes(item, containerid, symbol)  + ' class="' + prx.items.getComponentBaseClasses(item, containerid, symbol) + ' box pos type-basic-progressbar">';
        cR += '<style>';
        cR += prx.items.getComponentBaseStyle(item, containerid, symbol);
        cR += '#'+_id+' .progressbar-bar { background: '+prx.componentsHelper.getProp(item.barColor,'color-background')+'; border-radius: '+prx.componentsHelper.getProp(item.borderRadius,'border-radius')+'; }';
        cR += '#'+_id+' .progressbar-bar-filled { width: '+prx.componentsHelper.getProp(item.progress,'num-percetage')+'%; background: '+prx.componentsHelper.getProp(item.fillBarColor,'color-fill')+'; border-radius: '+prx.componentsHelper.getProp(item.borderRadius,'border-radius')+'; }';
        cR += '</style>';
        cR += prx.items.getComponentPrependDivs(item, containerid, symbol);
        cR += '<div class="progressbar-bar bar liveUpdate-barColor-background-color">';
        cR += '<div class="progressbar-bar-filled bar liveUpdate-fillBarColor-background-color"></div>';
        cR += '</div>';
        cR += prx.items.getComponentAppendDivs(item, containerid, symbol);
        cR += '</div>';
        return cR;
    }
    ,propertyGroups:	[
        {
            caption: 'Style',
            properties: [
                [
                    {
                        caption: 'Color'
                        , name: 'barColor'
                        , proptype: 'background-color'
                        , type: 'solid-colorpicker'
                        , value: function(item, name) {
                            return item.barColor;
                        }
                        , values: prx.comps.basicColors
                        , liveUpdate: 'background-color'
                        , changeProperty: {
                            caption: 'Color',
                            property: 'background-color',
                            selector: '.progressbar-bar',
                            transitionable: true
                        }

                    },
                    {
                        caption: 'Fill Color'
                        , name: 'fillBarColor'
                        , proptype: 'background-color-2-fill'
                        , type: 'solid-colorpicker'
                        , value: function(item, name) {
                            return item.fillBarColor;
                        }
                        , values: prx.comps.basicColors
                        , liveUpdate: 'background-color'
                        , changeProperty: {
                            caption: 'Fill Color',
                            property: 'background-color',
                            selector: '.progressbar-bar-filled',
                            transitionable: true
                        }
                    }
                ],
                [
                    {
                        caption: '<span class="icon icon-border-radius" title="Border radius"></span>',
                        name: 'borderRadius',
                        proptype: 'border-radius',
                        type: 'combo-select',
                        value: function(item, name) {
                            return item.borderRadius;
                        },
                        values: {min: 0, max: 20, step: 1},
                        expandableType: 'borderRadius',
                        expandedValues: ['tl', 'tr', 'bl', 'br'],
                        changeProperty: {
                            caption: 'Border Radius',
                            selector: '.bar',
                            property: 'border-radius',
                            transitionable: true
                        }
                    }
                ]
            ]
        },
        {
            caption: 'Progress (%)',
            properties: [

                [
                    {
                        caption: false
                        ,name: 'progress'
                        ,proptype: 'progress'
                        ,type: 'slider-input'
                        ,value: function(item,name) {
                            return item.progress;
                        }
                        ,values: { min: 0, max: 100, step: 1 }
                        ,changeProperty: {
                            caption: 'Progress',
                            property: 'percent-width',
                            selector: '.progressbar-bar-filled',
                            transitionable: true
                        }
                    }
                ]
            ]
        }
    ]
};

// TYPE: CIRCULAR PROGRESSBAR
prx.types.basic_circularprogressbar = {
    name: 'basic_circularprogressbar'
    ,onDisplay: function(item,containerid,symbol) {
        var _id = (!containerid) ? item.id : containerid+'-'+item.id;
        var _dims = prx.componentsHelper.getRealDims(item,symbol);
        var size = _dims.width < _dims.height? _dims.width : _dims.height;
        var strokeWidth = item.strokeWidth;
        var strokeDash = (size-strokeWidth) * Math.PI * item.progress / 100;
        var strokeOffset = (size-strokeWidth) * Math.PI * (100 - item.progress) / 100;
        var dashOffset = (size-strokeWidth) * Math.PI / 4*3;
        var cR = '';
        cR += '<div id="' + _id + '" ' + prx.items.getComponentBaseAttributes(item, containerid, symbol)  + ' data-stroke-width="'+item.strokeWidth+'" class="' + prx.items.getComponentBaseClasses(item, containerid, symbol) + ' box pos type-basic-circularprogressbar '+(item.loader ? 'type-basic-loader':'')+'">';
        cR += '<style>';
        cR += prx.items.getComponentBaseStyle(item, containerid, symbol);
        cR += ' #'+_id+' .circularprogressbar {height: '+_dims.height+'px; width: '+_dims.width+'px; } ';
        cR += ' #'+_id+' .circularprogressbar .circularprogressbar-bar {fill: none; stroke: '+prx.componentsHelper.getProp(item.barColor,'color-background')+'; stroke-width: '+strokeWidth+';}';
        cR += ' #'+_id+' .circularprogressbar .circularprogressbar-bar-filled {fill: none; stroke: '+prx.componentsHelper.getProp(item.fillBarColor,'color-fill')+'; stroke-dasharray: '+strokeDash+' '+strokeOffset+'; stroke-dashoffset: -'+dashOffset+'; stroke-linecap: '+(item.roundedEnds?'round':'butt')+'; stroke-width: '+strokeWidth+';}';
        cR += '</style>';
        cR += prx.items.getComponentPrependDivs(item, containerid, symbol);
        cR += '<svg class="circularprogressbar" xmlns="http://www.w3.org/2000/svg" viewBox="-'+strokeWidth+' -'+strokeWidth+' '+size+' '+size+'">';
        cR += '<circle cx="'+(size/2-strokeWidth)+'" cy="'+(size/2-strokeWidth)+'" r="'+((size-strokeWidth)/2>0?(size-strokeWidth)/2:0)+'" class="circularprogressbar-bar liveUpdate-barColor-stroke liveUpdate-strokeWidth-stroke-width" />';
        cR += '<circle cx="'+(size/2-strokeWidth)+'" cy="'+(size/2-strokeWidth)+'" r="'+((size-strokeWidth)/2>0?(size-strokeWidth)/2:0)+'" class="circularprogressbar-bar-filled liveUpdate-fillBarColor-stroke liveUpdate-strokeWidth-stroke-width" />';
        cR += '</svg>';
        cR += prx.items.getComponentAppendDivs(item, containerid, symbol);
        cR += '</div>';
        return cR;
    }
    ,onResize: function(item,containerid,symbol){
        var id = containerid ? containerid + '-' + item.id : item.id;
        var width = parseInt($('#'+id).width(),10);
        var height = parseInt($('#'+id).height(),10);
        var size = width < height? width : height;
        var strokeWidth = item.strokeWidth;
        var strokeDash = (size-strokeWidth) * Math.PI * item.progress / 100;
        var strokeOffset = (size-strokeWidth) * Math.PI * (100 - item.progress) / 100;
        var dashOffset = (size-strokeWidth) * Math.PI / 4*3;
        var _dims = prx.componentsHelper.getRealDims(item,symbol);
        $('#'+id+' .circularprogressbar')
            .css({width: _dims.width, height: _dims.height})
            .removeAttr('viewBox')
            .attr('viewBox', '-'+strokeWidth+' -'+strokeWidth+' '+size+' '+size);
        $('#'+id+' .circularprogressbar circle')
            .css({'stroke-width': strokeWidth})
            .removeAttr('cx')
            .removeAttr('cy')
            .removeAttr('r')
            .attr('cx', _dims.width/2)
            .attr('cy', _dims.height/2)
            .attr('r', (size-strokeWidth)/2>0?(size-strokeWidth)/2:0);
        $('#'+id+' .circularprogressbar .circularprogressbar-bar-filled')
            .css({'stroke-dasharray': strokeDash+' '+strokeOffset, 'stroke-dashoffset': -dashOffset});
    }
    ,propertyGroups:	[
        {
            caption: 'Style',
            properties: [
                [
                    {
                        caption: 'Color'
                        , name: 'barColor'
                        , proptype: 'background-color'
                        , type: 'solid-colorpicker'
                        , value: function(item, name) {
                            return item.barColor;
                        }
                        , values: prx.comps.basicColors
                        , liveUpdate: 'stroke'
                        , changeProperty: {
                            caption: 'Color',
                            property: 'stroke',
                            selector: '.circularprogressbar-bar',
                            transitionable: true
                        }

                    },
                    {
                        caption: 'Fill Color'
                        , name: 'fillBarColor'
                        , proptype: 'background-color-2-fill'
                        , type: 'solid-colorpicker'
                        , value: function(item, name) {
                            return item.fillBarColor;
                        }
                        , values: prx.comps.basicColors
                        , liveUpdate: 'stroke'
                        , changeProperty: {
                            caption: 'Fill Color',
                            property: 'stroke',
                            selector: '.circularprogressbar-bar-filled',
                            transitionable: true
                        }
                    }
                ]
                , [
                    {
                        caption: 'Thickness'
                        , name: 'strokeWidth'
                        , proptype: 'stroke-width'
                        , type: 'combo-select'
                        , value: function(item, name) {
                            return item.strokeWidth;
                        }
                        , values: {min: 1, max: 20, step: 1}
                        , changeProperty: {
                            caption: 'Thickness',
                            rerender: true
                        }
                    }
                ]
                , [
                    {
                        caption: 'Rounded ends'
                        , name: 'roundedEnds'
                        , proptype: 'rounded-ends'
                        , type: 'onoff'
                        , value: function(item, name) {
                            return item.roundedEnds;
                        }
                        , changeProperty: {
                            caption: 'Rounded ends',
                            rerender: true
                        }
                    },
                ]
            ]
        },
        {
            caption: 'Progress (%)',
            properties: [
                [
                    {
                        caption: false
                        , name: 'progress'
                        , proptype: 'progress'
                        , type: 'slider-input'
                        , value: function(item, name) {
                            return item.progress;
                        }
                        , values: {min: 0, max: 100, step: 1}
                        , changeProperty: {
                            caption: 'Progress',
                            rerender: true
                        }
                    }
                ]
            ]
        }
    ]
};

//TYPE: BASIC LOADER
prx.types.basic_loader = prx.componentsHelper.cloneobject(prx.types.basic_circularprogressbar);
prx.types.basic_loader.name = 'basic_loader';
prx.componentsHelper.removeProperties(prx.types.basic_loader.propertyGroups, ['progress']);

// TYPE: RADIOBUTTON
prx.types.basic_radiobutton = {
    name: 'basic_radiobutton'
    ,onDisplay: function(item,containerid,symbol) {

        var _id = (!containerid) ? item.id : containerid+'-'+item.id;
        var _dims = prx.componentsHelper.getRealDims(item, symbol);
        var _check = '';
        var _active = '';

        if(typeof(item.radioButtonsGroupName) == 'undefined') {
            item.radioButtonsGroupName = '';
        }

        var _hasRadioButtonsGroupName = typeof(item.radioButtonsGroupName) != 'undefined' && item.radioButtonsGroupName.length > 0;
        var _type = (prx.componentsHelper.getProp(item.actAsCheckbox,'boolean') && !_hasRadioButtonsGroupName) ? 'checkbox' : 'radio';
        var radioButtonsGroupName = _hasRadioButtonsGroupName ? 'name="' + containerid + '-radioButtonsGroup-' + item.radioButtonsGroupName + '"' : '';

        if(prx.componentsHelper.getProp(item.active,'boolean')) { _active = 'checked="checked"';}

        var _props = (typeof(prx.richtext)=='undefined') ? prx.componentsHelper.getProp(item.textProperties,'props-text') : '';
        var text = '<div class="basic-radiobutton-text liveUpdate-textColor-color changeProperty-textColor changeProperty-textFont changeProperty-textFontStyle changeProperty-textSize changeProperty-textProperties"><span data-editableproperty="text">'+prx.componentsHelper.getProp(item.text,'text-textarea')+'</span></div>';

        var cR = '';
        cR += '<div id="' + _id + '" ' + prx.items.getComponentBaseAttributes(item, containerid, symbol)  + ' class="' + prx.items.getComponentBaseClasses(item, containerid, symbol) + ' box pos type-basic-radiobutton">';

        cR += '<style>';
        cR += prx.items.getComponentBaseStyle(item, containerid, symbol);
        cR += '#'+_id+' label { height: 100%; width: 100%; line-height: '+item.textSize+'px;}';
        cR += '#'+_id+' label .basic-radiobutton-text { word-wrap: break-word; width: calc(100% - '+parseInt(item.radiobuttonSize,10)+'px - ' + (12 * prx.componentsHelper.getScale(item.lib)) + 'px); margin-left: '+(parseInt(item.radiobuttonSize,10)+12 * prx.componentsHelper.getScale(item.lib))+'px;  '+prx.componentsHelper.getProp(item.textFont + '|' + item.textFontStyle,'font-family') + prx.componentsHelper.getProp(item.textFontStyle,'font-style')+' color: '+prx.componentsHelper.getProp(item.textColor,'color-text')+'; font-size: '+prx.componentsHelper.getProp(item.textSize,'num-text-size')+'px; '+_props+' }';
        cR += '#'+_id+' label .bordered-circle { position: absolute; top: '+((item.textSize-item.radiobuttonSize)/2)+'px; width: '+item.radiobuttonSize+'px; height: '+item.radiobuttonSize+'px; max-width: '+item.radiobuttonSize+'px; max-height: '+item.radiobuttonSize+'px; min-width: '+item.radiobuttonSize+'px; min-height: '+item.radiobuttonSize+'px; background: '+prx.componentsHelper.getProp(item.backgroundColor,'color-background')+'; border: '+prx.componentsHelper.getProp(item.borderWidth,'num-border-width')+'px solid '+prx.componentsHelper.getProp(item.borderColor,'color-border')+'; }';
        cR += '#'+_id+' label .bordered-circle .circle { background-color: '+prx.componentsHelper.getProp(item.activeColor,'color-background')+'; }';
        cR += '#'+_id+' input:checked + label .bordered-circle .circle {  width: 50%; height: 50%; margin-top: -25%; margin-left: -25%; background: '+prx.componentsHelper.getProp(item.activeColor,'color-background')+';}';
        cR += '</style>';
        cR += prx.items.getComponentPrependDivs(item, containerid, symbol);

        cR += '<input type="'+_type+'" id="'+_id+'-checkbox" '+_active+'  '+radioButtonsGroupName+'  style="display: none;" data-role="none" />';
        cR += '<label for="'+_id+'-checkbox"><div class="bordered-circle liveUpdate-backgroundColor-background-color liveUpdate-borderColor-border-color liveUpdate-borderWidth-border-width changeProperty-backgroundColor changeProperty-borderColor changeProperty-borderWidth">';
        cR += '<div class="circle liveUpdate-activeColor-background-color changeProperty-activeColor"></div>';
        cR += '</div>';
        cR += text;
        cR += '</label>';
        cR += prx.items.getComponentAppendDivs(item, containerid, symbol);
        cR += '</div>';
        return cR;
    }
    ,afterDisplay: function(item,containerid,symbol) {
        var _id = (!containerid) ? item.id : containerid+'-'+item.id;

        if(!prx.editor) {

            var _hasRadioButtonsGroupName = typeof(item.radioButtonsGroupName) != 'undefined' && item.radioButtonsGroupName.length > 0;
            var _isCheckboxAndRadioGrouped = (prx.componentsHelper.getProp(item.actAsCheckbox,'boolean') && _hasRadioButtonsGroupName);

            prx.currentPlayerStatus.helper.set(_id, 'active', item.active);

            $('#'+_id+'-checkbox').on('change.custom-change-event', function(e, updateStatus){

                if(typeof(prx.variables._triggerData['input:checked[id='+_id+'-checkbox]']) == 'undefined') { prx.variables._triggerData['input:checked[id='+_id+'-checkbox]'] = {}; }
                prx.variables._triggerData['input:checked[id='+_id+'-checkbox]']['checkboxchange'] = { state: $(this).is(':checked') };
                if(typeof(prx.variables._triggerData['input[id='+_id+'-checkbox]:not(:checked)']) == 'undefined') { prx.variables._triggerData['input[id='+_id+'-checkbox]:not(:checked)'] = {}; }
                prx.variables._triggerData['input[id='+_id+'-checkbox]:not(:checked)']['checkboxchange'] = { state: $(this).is(':checked') };
                if(typeof(prx.variables._triggerData['#' + _id]) == 'undefined') { prx.variables._triggerData['#' + _id] = {}; }
                prx.variables._triggerData['#' + _id]['checkboxchange'] = { state: $(this).is(':checked') };
                $(this).trigger('checkboxchange');

                if ((updateStatus !== undefined) && !updateStatus) {
                    return;
                }

                if (_isCheckboxAndRadioGrouped) {
                    var screenItems = prx.currentPlayerStatus.helper.get(containerid, 'items');

                    Object.keys(screenItems).forEach(function (key) {
                        if (screenItems[key].radioButtonsGroupName && (screenItems[key].radioButtonsGroupName === item.radioButtonsGroupName)) {
                            prx.currentPlayerStatus.helper.set(containerid + '-' + screenItems[key].id, 'active', false);
                        }
                    });
                }

                prx.currentPlayerStatus.helper.set(_id, 'active', $(this).is(':checked'));
            });

            if(_isCheckboxAndRadioGrouped) {

                $('label[for="'+_id+'-checkbox"]').on('mousedown touchstart', function(event) {

                    var $checkbox = $('#'+_id+'-checkbox');
                    var checked = $checkbox.prop('checked');

                    $('input[type=\'radio\'][name=\''+$checkbox.attr('name')+'\']').each(function(){
                        if($(this).is(':checked')){
                            $(this).prop('checked', false).trigger('change', [false]);
                        } else {
                            $(this).prop('checked', false);
                        }
                    });

                    $checkbox.prop('checked', !checked).trigger('change', [true]);
                    event.preventDefault();

                });

                $('label[for="'+_id+'-checkbox"]').hammer().on('click tap', function(event) {
                    event.preventDefault();
                });

                $('#'+_id+'-checkbox').on('change', function(event) {
                    event.preventDefault();
                });

            }


            $('#'+_id).hammer();
            prx.actions.disableFlashActionOnItemTap('#' + _id, '.flashactiontap-afterdisplay');
        }
    }
    ,interactions: [
        {
            caption: 'Change',
            name: 'actions',
            type: 'action',
            value: function (item, name) {
                if (typeof(item.actions) == 'undefined') {
                    item.actions = [];
                }
                return item.actions.length;
            },
            changeProperty: {
                caption: 'Interactions on change',
                selector: '',
                property: 'action',
                transitionable: false,
                changeable: false
            }
        },
        {
            caption: 'Activation',
            name: 'checkboxActionsOnActive',
            type: 'action',
            value: function(item,name) {
                if(typeof(item.checkboxActionsOnActive) == 'undefined') {
                    item.checkboxActionsOnActive = [];
                }
                return item.checkboxActionsOnActive.length;
            },
            changeProperty: { caption: 'Interactions', selector: '', property: 'action', transitionable: false, changeable: false }

        },
        {
            caption: 'Deactivation',
            name: 'checkboxActionsOnDeactive',
            type: 'action',
            value: function(item,name) {
                if(typeof(item.checkboxActionsOnDeactive) == 'undefined') {
                    item.checkboxActionsOnDeactive = [];
                }
                return item.checkboxActionsOnDeactive.length;
            },
            changeProperty: { caption: 'Interactions', selector: '', property: 'action', transitionable: false, changeable: false }

        }
    ]
    ,editableProperties: [
        {
            caption: 'Text'
            ,name: 'text'
            ,type: 'textarea'
            ,isRichText: false
            ,value: function(item,name) {
                return item.text;
            }
            ,changeProperty: {
                caption: 'Text',
                property: 'text',
                selector: '.basic-radiobutton-text',
                transitionable: false
            }
        }
    ]
    ,mpactions: {
        specialEvents: ['checkboxchange']
    }
    ,propertyGroups: [
        {
            caption: 'Style',
            properties: [
                [
                    {
                        caption: 'Radiobutton size',
                        name: 'radiobuttonSize',
                        proptype: 'radiobutton-size',
                        type: 'combo-select',
                        values: { min: 16, max: 50, step: 2 },
                        value: function(item,name) {
                            return item.radiobuttonSize;
                        },
                        changeProperty: {
                            caption: 'Radiobutton size',
                            rerender: true
                        }
                    }
                ],
                [
                    {
                        caption: 'Background'
                        ,name: 'backgroundColor'
                        ,proptype: 'background-color'
                        ,type: 'gradients-colorpicker'
                        ,value: function(item,name) {
                            return item.backgroundColor;
                        }
                        ,values: prx.comps.basicColors
                        ,liveUpdate: 'background-color'
                        ,changeProperty: {
                            caption: 'Background color',
                            selector: '.changeProperty-backgroundColor',
                            property: 'background-color',
                            transitionable: true
                        }
                    }
                ],
                [
                    prx.commonproperties.borderWidth
                    ,{
                        name: 'borderColor',
                        proptype: 'border-color',
                        type: 'solid-colorpicker',
                        value: function(item,name) { return item.borderColor; },
                        values: prx.comps.basicColors,
                        liveUpdate: 'border-color'
                        ,changeProperty: {
                            caption: 'Border color',
                            selector: '.changeProperty-borderColor',
                            property: 'border-color',
                            transitionable: true
                        }
                    }
                ],
                [
                    {
                        caption: 'Active Dot'
                        ,name: 'activeColor'
                        ,proptype: 'background-color-2-active'
                        ,type: 'gradients-colorpicker'
                        ,value: function(item,name) {
                            return item.activeColor;
                        }
                        ,values: prx.comps.basicColors
                        ,liveUpdate: 'background-color'
                        ,changeProperty: {
                            caption: 'Active color',
                            selector: '.changeProperty-activeColor',
                            property: 'background-color',
                            transitionable: true
                        }
                    }
                ]
            ]
        },
        {
            caption: 'Text',
            properties: [
                [
                    prx.commonproperties.textFont
                ],
                [
                    prx.commonproperties.textFontStyle,
                    prx.commonproperties.textSize
                ],
                [
                    prx.commonproperties.textPropertiesRichText(['font-weight','font-style','text-decoration'],'text', false, {propName: 'textProperties', caption: 'Underline', useAsDynProp: false}),
                    {
                        caption: { label: 'Color', class: 'text-properties-label text-color-label' },
                        name: 'textColor',
                        proptype: 'font-color',
                        type: 'solid-colorpicker',
                        relatedEditableProperties: 'text',
                        relatedCSSProperties: 'color',
                        pffSettings: prx.commonproperties.pffSettingsColor,
                        value: function(item,name) {
                            if(typeof(item.textColor) == 'undefined') { item.textColor = '#2E2E2E'; }
                            return item.textColor;
                        },
                        liveUpdate: 'color'
                        ,changeProperty: {
                            caption: 'Text color',
                            selector: '.changeProperty-textColor',
                            property: 'color',
                            transitionable: true
                        }
                    }
                ]
            ]
        },
        {
            caption: 'Advanced',
            properties: [
                [
                    {
                        caption: 'Active'
                        ,name: 'active'
                        ,proptype: 'active'
                        ,type: 'onoff'
                        ,value: function(item,name) {
                            return item.active;
                        }
                        ,onChange: function(item, name) {
                            return prx.commonproperties.radioButtonsActiveStatusOnChange(item, name);
                        }
                        ,changeProperty: {
                            caption: 'Active',
                            selector: 'input',
                            property: 'checkbox-state',
                            transitionable: false
                        }
                    }
                ],[
                    {
                        caption: 'Can be deactivated after activation'
                        ,name: 'actAsCheckbox'
                        ,proptype: 'radio-as-checkbox'
                        ,type: 'onoff'
                        ,value: function(item,name) {
                            return item.actAsCheckbox;
                        }
                        ,changeProperty: {
                            caption: 'Can be deactivated after activation',
                            transitionable: false,
                            changeFunction(item, containerid) {
                                let id = (!containerid) ? item.id : containerid + '-' + item.id;
                                if (item.actAsCheckbox)
                                    $('#' + id + ' input')[0].type = 'checkbox';
                                else
                                    $('#' + id + ' input')[0].type = 'radio';
                            }
                        }
                    }
                ],
                [
                    prx.commonproperties.radioButtonsGroupNameInput
                ]
            ]
        }
    ]
};

// TYPE: SELECT
prx.types.basic_select = {
    name: 'basic_select'
    ,onDisplay: function(item,containerid,symbol) {
        var _id = (!containerid) ? item.id : containerid+'-'+item.id;
        var cR = '';
        var _dims = prx.componentsHelper.getRealDims(item, symbol);
        var activeTop = parseInt(item.dropdownTextSize, 10)>parseInt(_dims.height, 10)
            ? ((parseInt(_dims.height, 10) - parseInt(item.dropdownTextSize, 10))/2 - item.selected * parseInt(item.dropdownTextSize, 10) - parseInt(item.dropdownBorderWidth, 10)) - 8 * prx.componentsHelper.getScale(item.lib)
            : (-8 * prx.componentsHelper.getScale(item.lib) - item.selected * parseInt(_dims.height, 10) - parseInt(item.dropdownBorderWidth, 10));
        if(!prx.editor && item.top+activeTop<0) activeTop = -item.top;
        cR += '<div id="' + _id + '" ' + prx.items.getComponentBaseAttributes(item, containerid, symbol)  + ' class="' + prx.items.getComponentBaseClasses(item, containerid, symbol) + ' box pos type-basic-select liveUpdate-textColor-color">';

        cR += '<style>';
        cR += prx.items.getComponentBaseStyle(item, containerid, symbol);
        cR += '#'+_id+' .basic-select-active-value { '+prx.componentsHelper.getProp(item.textFont + '|' + item.textFontStyle,'font-family') + prx.componentsHelper.getProp(item.textFontStyle,'font-style')+' '+prx.componentsHelper.getProp(item.textProperties,'props-text')+' color: '+prx.componentsHelper.getProp(item.textColor,'color-text')+'; font-size: '+prx.componentsHelper.getProp(item.textSize,'num-text-size')+'px; text-align: '+prx.componentsHelper.getProp(item.textAlign,'align')+' }';
        cR += '#'+_id+' .basic-select-contextmenu { '+prx.componentsHelper.getProp(item.dropdownTextFont + '|' + item.dropdownTextFontStyle,'font-family') + prx.componentsHelper.getProp(item.dropdownTextFontStyle,'font-style')+' '+prx.componentsHelper.getProp(item.dropdownTextProperties,'props-text')+' color: '+prx.componentsHelper.getProp(item.dropdownTextColor,'color-text')+'; font-size: '+prx.componentsHelper.getProp(item.dropdownTextSize,'num-text-size')+'px; text-align: '+prx.componentsHelper.getProp(item.dropdownTextAlign,'align')+' }';
        cR += '#'+_id+' .select-trigger { line-height: '+(_dims.height-prx.componentsHelper.getProp(item.borderWidth,'num-border-width')*2)+'px; border-width: '+prx.componentsHelper.getProp(item.borderWidth,'border-width')+'; border-style: '+prx.componentsHelper.getProp(item.borderStyle,'border-type')+'; border-color: '+prx.componentsHelper.getProp(item.borderColor,'color-border')+'; '+prx.gradients.getBgCssByProperty(item.backgroundColor,'color-background')+'; border-radius: '+prx.componentsHelper.getProp(item.borderRadius,'border-radius')+'; text-align: '+item.textAlign+';}';
        cR += '#'+_id+' .basic-select-contextmenu { top: '+activeTop+'px; border: '+prx.componentsHelper.getProp(item.dropdownBorderWidth,'num-border-width')+'px '+prx.componentsHelper.getProp(item.dropdownBorderStyle,'border-type')+' '+prx.componentsHelper.getProp(item.dropdownBorderColor,'color-border')+'; '+prx.gradients.getBgCssByProperty(item.dropdownBackgroundColor,'color-background')+'; border-radius: '+prx.componentsHelper.getProp(item.dropdownBorderRadius,'border-radius')+'}';
        cR += '#'+_id+' .basic-select-item {line-height: '+item.dropdownTextSize+'px; min-height: '+_dims.height+'px;}';
        cR += '#'+_id+' .select-trigger-icon { width: '+_dims.height+'px; background-image: url('+prx.componentsHelper.getProp(item.buttonicon,'asset')+'); line-height: '+(prx.componentsHelper.getProp(_dims.height,'num-other')-prx.componentsHelper.getProp(item.borderWidth,'num-border-width')*2)+'px; }';
        if(!prx.editor) {
            cR += '#' + _id + ' .basic-select-item:hover {' + prx.gradients.getBgCssByProperty(item.dropdownHoverBackgroundColor, 'color-background') + ' }';
        } else {
            cR += '#' + _id + ' .active {' + prx.gradients.getBgCssByProperty(item.dropdownHoverBackgroundColor, 'color-background') + ' }';
        }
        cR += '</style>';
        cR += prx.items.getComponentPrependDivs(item, containerid, symbol);

        cR += '<div class="select-trigger liveUpdate-borderColor-border-color liveUpdate-backgroundColor-background changeProperty-backgroundColor changeProperty-borderColor changeProperty-borderWidth changeProperty-borderStyle changeProperty-borderRadius changeProperty-textAlign">';
        cR += '<span class="basic-select-active-value changeProperty-textColor liveUpdate-textColor-color changeProperty-textSize changeProperty-textFont changeProperty-textFontStyle changeProperty-textProperties">';
        cR += (typeof(item.buttons[item.selected]) != 'undefined') ? prx.componentsHelper.getProp(item.buttons[item.selected].text,'text-textarea') : ((typeof(item.buttons[0]) != 'undefined') ? prx.componentsHelper.getProp(item.buttons[0].text,'text-textarea') : '');
        cR += '</span>';
        cR += '<div class="select-trigger-icon"></div>';
        cR += '</div>';
        cR += '<div class="basic-select-contextmenu changeProperty-dropdownBackgroundColor liveUpdate-dropdownBackgroundColor-background-color liveUpdate-dropdownBorderColor-border-color">';
        $.each(item.buttons, function(i,elm){
            cR += '<div class="basic-select-item dynamic-property changeProperty-dropdownTextFont changeProperty-dropdownTextColor changeProperty-dropdownTextFontStyle changeProperty-dropdownTextSize changeProperty-dropdownTextAlign changeProperty-dropdownTextProperties '+ (item.selected == i ? 'active liveUpdate-dropdownHoverBackgroundColor-background-color' : '') +'" data-dynamic-property-index="'+i+'" id="'+_id+'-buttons-'+i+'">';
            cR += '<div class="image"><img src="'+prx.componentsHelper.getProp(item.dropdownActiveIcon,'asset')+'"></div>';
            cR += '<div class="error-msg"><span data-editableproperty="text" data-dynamic-property-index="'+i+'">'+prx.componentsHelper.getProp(elm.text,'text-textarea')+'</span></div>';
            cR += '</div>';
        });
        cR += '</div>';
        cR += prx.items.getComponentAppendDivs(item, containerid, symbol);
        cR += '</div>';
        return cR;
    }
    ,onResize: function(item,containerid,symbol) {
        var _id = (!containerid) ? item.id : containerid+'-'+item.id;
        var _dims = prx.componentsHelper.getRealDims(item,symbol);
        var activeTop = parseInt(item.dropdownTextSize, 10)>parseInt(_dims.height, 10)
            ? ((parseInt(_dims.height, 10) - parseInt(item.dropdownTextSize, 10))/2 - item.selected * parseInt(item.dropdownTextSize, 10) - parseInt(item.dropdownBorderWidth, 10)) - 8 * prx.componentsHelper.getScale(item.lib)
            : (-8 * prx.componentsHelper.getScale(item.lib) - item.selected * parseInt(_dims.height, 10) - parseInt(item.dropdownBorderWidth, 10));

        $('#'+_id+' .basic-select-contextmenu').css('top', activeTop);
        $('#'+_id+' .select-trigger-icon').css('width', _dims.height);
        $('#'+_id+' .basic-select-item').css('min-height', _dims.height);
        $('#'+_id+' .select-trigger').css('line-height', prx.componentsHelper.getProp(_dims.height,'num-other')-prx.componentsHelper.getProp(item.borderWidth,'num-border-width')*2 + 'px');
    }
    ,afterDisplay: function(item,containerid,symbol) {
        var _id = (!containerid) ? item.id : containerid+'-'+item.id;
        var _dims = prx.componentsHelper.getRealDims(item,symbol);
        if(!prx.editor) {
            let selector = ' .basic-select-contextmenu';

            $('#' + _id).hammer().on('click', function (e) {
                e.stopPropagation();
            });

            $('#'+_id+' .select-trigger').hammer({ domEvents: false }).on('tap',function(){
                if(typeof(prx.variables._triggerData['#'+_id]) == 'undefined') { prx.variables._triggerData['#'+_id] = {}; }

                if (!$('#' + _id + selector).is(':visible')) {
                    prx.helper.toggleDropdownOptions(false, '#' + _id, '#' + _id + selector);

                    //Clicked dropdown will open
                    let selectedText = prx.currentPlayerStatus.helper.get(_id, "buttons")[prx.currentPlayerStatus.helper.get(_id, "selected")].text;
                    prx.variables._triggerData['#' + _id]['selectopen'] = {value: selectedText};
                    $('#' + _id).trigger('selectopen');
                }else{
                    // The user clicks on the Dropdown arrow to close the component or opens another dropdown
                    prx.helper.closeDropdowns();
                }

                $(document).one('dummy.close-dropdowns', function() {
                    prx.helper.toggleDropdownOptions(true, '#' + _id, '#' + _id + selector);

                    let selectedText = prx.currentPlayerStatus.helper.get(_id, "buttons")[prx.currentPlayerStatus.helper.get(_id, "selected")].text;
                    // The user clicks outside the dropdown, which triggers the “dropdown-close” event
                    prx.variables._triggerData['#'+_id]['selectclose'] = { value: selectedText };
                    $('#'+_id).trigger('selectclose');
                });
            });

            $('#' + _id + ' .basic-select-item').hammer().on('tap', function (e) {
                prx.helper.toggleDropdownOptions(true, '#' + _id, '#'+_id + selector);

                $('#' + _id + ' .basic-select-item').removeClass('active');
                $(this).addClass('active');
                let activeSelection = parseInt(e.target.dataset.dynamicPropertyIndex);
                let activeText = prx.currentPlayerStatus.helper.get(_id, "buttons")[activeSelection].text;

                $('#'+_id+' .basic-select-active-value').text(activeText);
                var selected = $(this).attr('data-dynamic-property-index');
                var activeTop = parseInt(item.dropdownTextSize, 10)>parseInt(_dims.height, 10)
                    ? ((parseInt(_dims.height, 10) - parseInt(item.dropdownTextSize, 10))/2 - selected * parseInt(item.dropdownTextSize, 10) - parseInt(item.dropdownBorderWidth, 10)) - 8 * prx.componentsHelper.getScale(item.lib)
                    : (-8 * prx.componentsHelper.getScale(item.lib) - selected * parseInt(_dims.height, 10) - parseInt(item.dropdownBorderWidth, 10));
                if(item.top+activeTop<0) activeTop = -item.top;
                $('#' + _id + selector).css('top', activeTop);

                if(typeof(prx.variables._triggerData['#'+_id]) == 'undefined') { prx.variables._triggerData['#'+_id] = {}; }
                prx.variables._triggerData['#'+_id]['selectchange'] = { value: activeText };
                $('#'+_id).trigger('selectchange');
                // The user selects an item from the dropdown, and so the dropdown closes
                prx.variables._triggerData['#'+_id]['selectclose'] = { value: activeText };
                $('#'+_id).trigger('selectclose');

                prx.currentPlayerStatus.helper.set(_id, 'selected', activeSelection);
            });

            prx.actions.disableFlashActionOnItemTap('#' + _id + ' .select-trigger', '.flashactiontap-afterdisplay');
            prx.actions.disableFlashActionOnItemTap('#' + _id + ' .basic-select-item', '.flashactiontap-afterdisplay');

        }
    }
    , mpactions: {
        specialEvents: ['selectchange', 'selectopen', 'selectclose']
    }
    , interactions: [prx.commonproperties.actions]
    ,propertyGroups: [
        {
            caption: 'Closed Button Style',
            properties: [
                [
                    {
	                    caption: 'Background',
	                    name: 'backgroundColor',
	                    proptype: 'background',
	                    type: 'gradients-colorpicker',
	                    value: function (item, name) {
	                        return item.backgroundColor;
	                    },
	                    liveUpdate: 'background',
	                    changeProperty: {
	                        caption: 'Background color',
	                        selector: '.changeProperty-backgroundColor',
	                        property: 'background',
	                        transitionable: true
	                    }
	                }
                ],
	            [
                    {
	                    caption: 'Border',
	                    name: 'borderWidth',
	                    proptype: 'border-width',
	                    type: 'combo-select',
	                    value: function(item,name) { return item.borderWidth; },
	                    values: { min: 0, max: 20, step: 1 } ,
                        expandableType: 'borderWidth',
	                    expandedValues: ['t', 'r', 'l', 'b'],
	                    changeProperty: {
	                        caption: 'Border width',
	                        selector: '.changeProperty-borderWidth',
	                        property: 'border-width',
	                        transitionable: true
	                    }
	                }
                    , {
	                    caption: false,
	                    name: 'borderStyle',
	                    proptype: 'border-style',
	                    type: 'select',
	                    value: function (item, name) {
	                        if (typeof(item.borderStyle) == 'undefined') {
	                            item.borderStyle = 'solid';
	                        }
	                        return item.borderStyle;
	                    },
	                    values: [{value: 'solid', displayValue: 'Solid'}, {
	                        value: 'dotted',
	                        displayValue: 'Dotted'
	                    }, {value: 'dashed', displayValue: 'Dashed'}, {
	                        value: 'double',
	                        displayValue: 'Double'
	                    }, {value: 'none', displayValue: 'None'}],
	                    changeProperty: {
	                        caption: 'Border style',
	                        selector: '.changeProperty-borderStyle',
	                        property: 'border-style',
	                        transitionable: false
	                    }
	                }
                    ,{
                        name: 'borderColor',
                        proptype: 'border-color',
                        type: 'solid-colorpicker',
                        value: function(item,name) { return item.borderColor; },
                        values: prx.comps.basicColors,
                        liveUpdate: 'border-color'
                        ,changeProperty: {
                            caption: 'Border color',
                            selector: '.changeProperty-borderColor',
                            property: 'border-color',
                            transitionable: false,
                            changeFunction(item, containerid) {
                                let id = (!containerid) ? item.id : containerid + '-' + item.id;
                                let cssSelector = '#' + id + ' .select-trigger ';

                                prx.componentsHelper.addStyleForChangeFunction(id, id + '-borderColor', cssSelector, 'border-color', item.borderColor, 'color-border');
                            }
                        }
                    }
                ],
	            [
                    prx.commonproperties.borderRadiusExpandable
                ]
            ]
        },
        {
            caption: 'Dropdown Icon',
            properties: [
                [
                    {
                        caption: false
                        ,name: 'buttonicon'
                        ,proptype: 'buttonicon'
                        ,type: 'combo-asset'
                        ,displayValue: function(item,name) {
                            if(item.buttonicon.fileId == '') {
                                return 'No icon selected';
                            }
                            return item.buttonicon.name;
                        }
                        ,value: function(item,name) {
                            return JSON.stringify({
                                allow: 'image',
                                asset: item.buttonicon
                            });
                        }
                        ,changeProperty: {
                            caption: 'Dropdown icon',
                            transitionable: false,
                            changeFunction(item, containerid) {
                                let id = (!containerid) ? item.id : containerid + '-' + item.id;
                                let cssSelector = '#' + id + ' .select-trigger-icon ';

                                prx.componentsHelper.changeImageForChangeFunction(cssSelector, item.buttonicon, true);
                            }
                        }
                    }
                ]
            ]
        },
        {
            caption: 'Closed Button Text',
            properties: [
                [
                    {
                        caption: { label: 'Font family', class: 'text-properties-label text-fontfamily-label' },
                        name: 'textFont',
                        proptype: 'font-family',
                        type: 'select',
                        relatedEditableProperties: ['buttons.closedText'],
                        relatedCSSProperties: 'font-family',
                        value: function(item,name) {
                            if(typeof(item.textFont) == 'undefined') { item.textFont = 'Arial,sans-serif'; }
                            return item.textFont;
                        },
                        values: function(){ return prx.comps.fonts; }
                        ,changeProperty: {
                            caption: 'Text font',
                            selector: '.changeProperty-textFont',
                            property: 'font-family',
                            transitionable: false
                        }
                    }
                ],
                [
                    prx.commonproperties.textFontStyle,
                    prx.commonproperties.textSize
                ],
                [
                    prx.commonproperties.textPropertiesRichText(['font-weight','font-style','text-decoration'],'closedText', false, {propName: 'textProperties', caption: 'Underline', useAsDynProp: false}),
                    {
                        caption: { label: 'Color', class: 'text-properties-label text-color-label' },
                        name: 'textColor',
                        proptype: 'font-color',
                        type: 'solid-colorpicker',
                        relatedEditableProperties: 'text',
                        relatedCSSProperties: 'color',
                        pffSettings: prx.commonproperties.pffSettingsColor,
                        value: function(item,name) {
                            if(typeof(item.textColor) == 'undefined') { item.textColor = '#2E2E2E'; }
                            return item.textColor;
                        },
                        liveUpdate: 'color'
                        ,changeProperty: {
                            caption: ' Text color',
                            selector: '.changeProperty-textColor',
                            property: 'color',
                            transitionable: true
                        }
                    }
                ],
                [
                    prx.commonproperties.textAlign
                ]
            ]
        },
        {
            caption: 'Popover style',
            properties: [
                [
                    {
                        caption: 'Background',
                        name: 'dropdownBackgroundColor',
                        proptype: 'background-color-2',
                        type: 'gradients-colorpicker',
                        value: function (item, name) {
                            return item.dropdownBackgroundColor;
                        },
                        liveUpdate: 'background-color',
                        changeProperty: {
                            caption: 'Popover background color',
                            selector: '.basic-select-contextmenu',
                            property: 'background-color',
                            transitionable: true
                        }
                    }
                ],
                [
                    {
                        caption: 'Border',
                        name: 'dropdownBorderWidth',
                        proptype: 'border-width-2',
                        type: 'combo-select',
                        value: function(item,name) { return item.dropdownBorderWidth; },
                        values: { min: 0, max: 20, step: 1 } ,
                        changeProperty: {
                            caption: 'Popover border width',
                            selector: '.basic-select-contextmenu',
                            property: 'border-width',
                            transitionable: true
                        }
                    }
                    , {
                        caption: false,
                        name: 'dropdownBorderStyle',
                        proptype: 'border-style-2',
                        type: 'select',
                        value: function (item, name) {
                            return item.dropdownBorderStyle;
                        },
                        values: [{value: 'solid', displayValue: 'Solid'}, {
                            value: 'dotted',
                            displayValue: 'Dotted'
                        }, {value: 'dashed', displayValue: 'Dashed'}, {
                            value: 'double',
                            displayValue: 'Double'
                        }, {value: 'none', displayValue: 'None'}],
                        changeProperty: {
                            caption: 'Popover border style',
                            selector: '.basic-select-contextmenu',
                            property: 'border-style',
                            transitionable: false
                        }
                    }
                    ,{
                        name: 'dropdownBorderColor',
                        proptype: 'border-color-2',
                        type: 'solid-colorpicker',
                        value: function(item,name) { return item.dropdownBorderColor; },
                        values: prx.comps.basicColors,
                        liveUpdate: 'border-color'
                        ,changeProperty: {
                            caption: 'Popover border color',
                            selector: '.basic-select-contextmenu',
                            property: 'border-color',
                        }
                    }
                ],
                [
            	{
                        caption: '<span class="icon icon-border-radius" title="Border radius"></span>',
                        name: 'dropdownBorderRadius',
                        proptype: 'border-radius-2',
                        type: 'combo-select',
                        value: function(item,name) {
                            return item.dropdownBorderRadius;
                        },
                        values: {min: 0, max: 20, step: 1},
                        changeProperty: {
                            caption: 'Popover border radius',
                            selector: '.basic-select-contextmenu',
                            property: 'border-radius',
                            transitionable: true
                        }
                    }
                ],
                [
                    {
                        caption: 'Hover background',
                        name: 'dropdownHoverBackgroundColor',
                        proptype: 'background-color-3',
                        type: 'gradients-colorpicker',
                        value: function (item, name) {
                            return item.dropdownHoverBackgroundColor;
                        },
                        liveUpdate: 'background-color',
                        changeProperty: {
                            caption: 'Hover background color',
                            transitionable: false,
                            changeFunction(item, containerid) {
                                let id = (!containerid) ? item.id : containerid + '-' + item.id;
                                let cssSelector = '#' + id + ' .basic-select-item:hover';

                                prx.componentsHelper.addStyleForChangeFunction(id, id + '-activeBackgroundColor', cssSelector, 'background-color', item.dropdownHoverBackgroundColor, 'color-background');
                            }

                        }
                    }
                ]
            ]

        },
        {
            caption: 'Selected option style',
            properties: [
                [
                    {
                        caption: false
                        ,name: 'dropdownActiveIcon'
                        ,proptype: 'dropdownActiveIcon'
                        ,type: 'combo-asset'
                        ,displayValue: function(item,name) {
                            if(item.dropdownActiveIcon.fileId == '') {
                                return 'No icon selected';
                            }
                            return item.dropdownActiveIcon.name;
                        }
                        ,value: function(item,name) {
                            return JSON.stringify({
                                allow: 'image',
                                asset: item.dropdownActiveIcon
                            });
                        }
                        ,changeProperty: {
                            caption: 'Active icon',
                            transitionable: false,
                            changeFunction(item, containerid) {
                                let id = (!containerid) ? item.id : containerid + '-' + item.id;
                                let cssSelector = '#' + id + ' .basic-select-item .image > img ';

                                prx.componentsHelper.changeImageForChangeFunction(cssSelector, item.dropdownActiveIcon);
                            }
                        }
                    }
                ]
            ]

        },
        {
            caption: 'Popover text',
            properties: [
                [
                    {
                        caption: { label: 'Font family', class: 'text-properties-label text-fontfamily-label' },
                        name: 'dropdownTextFont',
                        proptype: 'font-family-2',
                        type: 'select',
                        relatedEditableProperties: 'buttons.text',
                        relatedCSSProperties: 'font-family',
                        value: function(item,name) {
                            if(typeof(item.dropdownTextFont) == 'undefined') { item.dropdownTextFont = 'Arial,sans-serif'; }
                            return item.dropdownTextFont;
                        },
                        values: function(){ return prx.comps.fonts; }
                        ,changeProperty: {
                            caption: 'Popover text font',
                            selector: '.changeProperty-dropdownTextFont',
                            property: 'font-family',
                            transitionable: false
                        }
                    }
                ],
                [
                    {
                        caption: { label: 'Select font style', class: 'text-properties-label text-fontstyle-label' },
                        name: 'dropdownTextFontStyle',
                        proptype: 'font-style-2',
                        type: 'select',
                        pffSettings: prx.commonproperties.pffSettingsTextFontStyle,
                        value: function(item, name) {
                            return typeof item.dropdownTextFontStyle == 'undefined' ? '' : item.dropdownTextFontStyle;
                        },
                        values: function() {
                            return prx.comps.fontStyles;
                        },
                        hiddenByDefault: function(item) {
                            return false;
                        },
                        changeProperty: {
                            caption: 'Popover font style',
                            transitionable: false,
                            changeFunction: function(item, containerid){
                                let id = (!containerid) ? item.id : containerid + '-' + item.id;
                                let cssSelector = '#' + id + ' .basic-select-contextmenu';
                                prx.componentsHelper.addStyleForChangeFunction(id, id + '-font-style-2', cssSelector, '', item.dropdownTextFontStyle, 'font-style');
                            }

                        }
                    },
                    {
                        caption: false,
                        name: 'dropdownTextSize',
                        proptype: 'font-size-2',
                        type: 'combo-select',
                        relatedEditableProperties: ['buttons.text'],
                        relatedCSSProperties: 'font-size',
                        value: function(item,name) {
                            if(typeof(item.dropdownTextSize) == 'undefined') { item.dropdownTextSize = 16; }
                            return item.dropdownTextSize;
                        },
                        values: prx.comps.textsize
                        ,changeProperty: {
                            caption: 'Popover text size',
                            selector: '.changeProperty-dropdownTextSize',
                            property: 'font-size',
                            transitionable: false
                        },
                        onChange: function(item) {
                            if(item.lineHeightAuto==true) {
                                item.lineHeight = parseInt(prx.componentsHelper.getProp(item.dropdownTextSize,'num-text-size')*1.231);
                            }
                        }
                    }
                ]
                ,
                [
                    prx.commonproperties.textPropertiesRichText(['font-weight','font-style','text-decoration'],'buttons.text', false, {propName: 'dropdownTextProperties', caption: 'Popover underline', useAsDynProp: false}),
                    {
                        caption: { label: 'Color', class: 'text-properties-label text-color-label' },
                        name: 'dropdownTextColor',
                        proptype: 'font-color-2',
                        type: 'solid-colorpicker',
                        relatedEditableProperties: ['buttons.text'],
                        relatedCSSProperties: 'color',
                        pffSettings: prx.commonproperties.pffSettingsColor,
                        value: function(item,name) {
                            return item.dropdownTextColor;
                        },
                        liveUpdate: 'color'
                        ,changeProperty: {
                            caption: 'Popover text color',
                            selector: '.changeProperty-dropdownTextColor',
                            property: 'color',
                            transitionable: true
                        }
                    }
                ],
                [
                    {
                        caption: { label: 'Alignment', class: 'text-properties-label text-alignment-label' },
                        name: 'dropdownTextAlign',
                        proptype: 'text-align-2',
                        type: 'radio',
                        relatedEditableProperties: ['buttons.text'],
                        relatedCSSProperties: 'text-align',
                        pffSettings: prx.commonproperties.pffSettingsTextAlignment,
                        value: function(item,name) {
                            return item.dropdownTextAlign;
                        },
                        values: [
                            { value: 'left', displayValue: '', icon: 'align-left'},
                            { value: 'center', displayValue: '', icon: 'align-center'},
                            { value: 'right', displayValue: '', icon: 'align-right'},
                            { value: 'justify', displayValue: '', icon: 'align-justify'}
                        ],
                        changeProperty: {
                            caption: 'Popover text align',
                            selector: '.changeProperty-dropdownTextAlign',
                            property: 'text-align',
                            transitionable: false
                        }
                    }
                ]
            ]
        },
        {
            caption: 'Advanced',
            properties: [
                [
                    {
                        caption: 'Active option'
                        ,name: 'selected'
                        ,proptype: 'selected'
                        ,type: 'select'
                        ,value: function(item,name) {
                            return item.selected;
                        }
                        ,values: function(item,name) {
                            var _rA = [];
                            for (var n = 0; n < item.buttons.length; n++) {
                                _rA.push({value: n,displayValue: item.buttons[n].text});
                            }
                            return _rA;
                        }
                        ,changeProperty: {
                            caption: 'Active option',
                            transitionable: false,
                            changeFunction(item, containerid) {
                                let id = (!containerid) ? item.id : containerid + '-' + item.id;
                                let cssSelector = '#' + id + ' .basic-select-item';
                                let items = $(cssSelector);

                                $.each(items, function (index, _item) {
                                    $(_item).removeClass('active');
                                    if (index === parseInt(item.selected)){
                                        //select value
                                        $(_item).addClass('active');
                                        //change the dropdown preview to the selected value
                                        $('#' + id).find('.basic-select-active-value').text($(_item).find('span').text());
                                    }
                                });
                            }
                        }
                    }
                ]
            ]
        }
    ]
    ,editableProperties: [
        {
            caption: 'Text'
            ,name: 'closedText'
            ,type: 'input'
            ,hiddenByDefault: function(item){ return true; }
            ,value: function(item,name,index) {
                return item.buttons[index].text;
            }
            ,liveUpdate:'color'
            ,onChange: function(item, index, val){
                if(item.selected == index) {
                    $('#' + item.id).find('.basic-select-active-value').html(val);
                }
            }
            ,changeProperty: {
                caption: 'Text',
                property: 'text',
                selector: '.basic-select-active-value', //do not delete, even it is rerender we need this for the changetextprop and changetextfont style functions
                rerender: true
            }
        }
    ]
    ,dynamicProperties: {
        data: 'buttons'
        ,propertyCaption: 'Menu items'
        ,propertyName: 'Menu item'
        ,addCaption: 'Add menu item'
        ,deleteCaption: 'Delete'
        ,blankItem: {
            text: 'Label',
            actions: []
        }
        ,captionProperty: 'text'

        ,editableProperties:[
            {
                caption: 'Text'
                ,name: 'text'
                ,type: 'input'
                ,value: function(item,name,index) {
                    return item.buttons[index].text;
                }
                ,onChange: function(item, index, val){
                    if(item.selected == index) {
                        $('#' + item.id).find('.basic-select-active-value').html(val);
                    }
                }
                ,changeProperty: {
                    caption: 'Text',
                    transitionable: false,
                    property: 'text',
                    selector: '.basic-select-item', //do not delete, even it is rerender we need this for the changetextprop and changetextfont style functions
                    changeFunction(item, containerid) {
                        let id = (!containerid) ? item.id : containerid + '-' + item.id;
                        let cssSelector = '#' + id + ' .basic-select-item .error-msg span';

                        $.each($(cssSelector), function (key, _item) {
                            $(_item).html(item.buttons[key].text);
                        });
                    }
                }
            },
        ]
        ,interactions:	[ prx.commonproperties.actions_buttons ]
        ,propertyGroups: [
            {
                caption: '',
                properties: [
                    [

                    ]
                ]
            }
        ]

    }
};

// TYPE: DROPDOWN MENU
prx.types.basic_dropdown = {
    name: 'basic_dropdown'
    ,onDisplay: function(item,containerid,symbol) {
        var _id = (!containerid) ? item.id : containerid+'-'+item.id;
        var cR = '';
        var _checked = '';
        var _dims = prx.componentsHelper.getRealDims(item, symbol);

        var _position;
        switch(item.dropdownPosition){
        case 'right':
            _position = 'flex-end';
            break;
        case 'center':
            _position = 'center';
            break;
            _position = 'flex-start';
            break;
        }
        cR += '<div id="' + _id + '" ' + prx.items.getComponentBaseAttributes(item, containerid, symbol)  + ' class="' + prx.items.getComponentBaseClasses(item, containerid, symbol) + ' box pos type-basic-dropdown liveUpdate-textColor-color position-dropdown-' + _position + ' ">';
        cR += '<style>';
        cR += prx.items.getComponentBaseStyle(item, containerid, symbol);
        cR += '#'+_id+' .basic-dropdown-active-value { '+prx.componentsHelper.getProp(item.textFont + '|' + item.textFontStyle,'font-family') + prx.componentsHelper.getProp(item.textFontStyle,'font-style')+' '+prx.componentsHelper.getProp(item.textProperties,'props-text')+' color: '+prx.componentsHelper.getProp(item.textColor,'color-text')+'; font-size: '+prx.componentsHelper.getProp(item.textSize,'num-text-size')+'px; text-align: '+prx.componentsHelper.getProp(item.textAlign,'align')+' }';
        cR += '#'+_id+' .basic-dropdown-contextmenu { '+prx.componentsHelper.getProp(item.dropdownTextFont + '|' + item.dropdownTextFontStyle,'font-family') + prx.componentsHelper.getProp(item.dropdownTextFontStyle,'font-style')+' '+prx.componentsHelper.getProp(item.dropdownTextProperties,'props-text')+' color: '+prx.componentsHelper.getProp(item.dropdownTextColor,'color-text')+'; font-size: '+prx.componentsHelper.getProp(item.dropdownTextSize,'num-text-size')+'px; text-align: '+prx.componentsHelper.getProp(item.dropdownTextAlign,'align')+' }';
        cR += '#'+_id+' .dropdown-trigger { line-height: '+(_dims.height-prx.componentsHelper.getProp(item.borderWidth,'num-border-width')*2)+'px; border-width: '+prx.componentsHelper.getProp(item.borderWidth,'border-width')+'; border-style: '+prx.componentsHelper.getProp(item.borderStyle,'border-type')+'; border-color: '+prx.componentsHelper.getProp(item.borderColor,'color-border')+'; '+prx.gradients.getBgCssByProperty(item.backgroundColor,'color-background')+'; border-radius: '+prx.componentsHelper.getProp(item.borderRadius,'border-radius')+'; text-align: '+item.textAlign+';}';
        cR += '#'+_id+' .basic-dropdown-contextmenu { width: fit-content; margin-top: 5px; border: '+prx.componentsHelper.getProp(item.dropdownBorderWidth,'num-border-width')+'px '+prx.componentsHelper.getProp(item.dropdownBorderStyle,'border-type')+' '+prx.componentsHelper.getProp(item.dropdownBorderColor,'color-border')+'; '+prx.gradients.getBgCssByProperty(item.dropdownBackgroundColor,'color-background')+'; border-radius: '+prx.componentsHelper.getProp(item.dropdownBorderRadius,'border-radius')+'}';
        cR += '#'+_id+' .basic-dropdown-item:hover { '+ prx.gradients.getBgCssByProperty(item.hoverInactiveBackgroundColor,'color-background') +'; color: '+prx.componentsHelper.getProp(item.hoverInactiveTextColor,'color-text')+'; }';
        cR += '#'+_id+' .basic-dropdown-item.active:hover { '+ prx.gradients.getBgCssByProperty(item.hoverActiveBackgroundColor,'color-background') +'; color: '+prx.componentsHelper.getProp(item.hoverActiveTextColor,'color-text')+'; }';
        cR += '#'+_id+' .active { color: '+prx.componentsHelper.getProp(item.dropdownActiveTextColor,'color-text')+'; '+ prx.gradients.getBgCssByProperty(item.dropdownActiveBackgroundColor,'color-background') +' }';
        cR += '</style>';
        cR += prx.items.getComponentPrependDivs(item, containerid, symbol);

        cR += '<div class="dropdown-trigger liveUpdate-textColor-color liveUpdate-borderColor-border-color liveUpdate-backgroundColor-background-color changeProperty-backgroundColor changeProperty-borderRadius">';
        cR += '<div class="basic-dropdown-active-value changeProperty-textColor liveUpdate-textColor-color changeProperty-textSize changeProperty-textFont changeProperty-textFontStyle changeProperty-textProperties changeProperty-textAlign">';
        cR += '<span data-editableproperty="text">'+prx.componentsHelper.getProp(item.text,'text-textarea')+'</span>';
        cR += '</div>';
        cR += '<div class="dropdown-trigger-icon">' + (item.buttonicon.fileId ? '<img src="'+prx.componentsHelper.getProp(item.buttonicon,'asset')+'"/>' : '') + '</div>';
        cR += '</div>';
        cR += '<div class="basic-dropdown-wrapper">';
        cR += '<div class="basic-dropdown-contextmenu liveUpdate-dropdownBackgroundColor-background-color liveUpdate-dropdownBorderColor-border-color liveUpdate-dropdownTextColor-color changeProperty-dropdownTextColor">';
        $.each(item.buttons, function(i,elm){
            cR += '<div class="basic-dropdown-item dynamic-property changeProperty-dropdownTextFont changeProperty-dropdownTextColor changeProperty-dropdownTextFontStyle changeProperty-dropdownTextSize changeProperty-dropdownTextAlign changeProperty-dropdownTextProperties '+ (item.selected == i ? 'active liveUpdate-dropdownTextColor-color liveUpdate-dropdownActiveBackgroundColor-background-color' : '') +'" data-dynamic-property-index="'+i+'" id="'+_id+'-buttons-'+i+'">';
            cR += '<div class="error-msg"><span data-editableproperty="text" data-dynamic-property-index="'+i+'">'+prx.componentsHelper.getProp(elm.text,'text-textarea')+'</span></div>';
            cR += '</div>';
        });
        cR += '</div>';
        cR += '</div>';
        cR += prx.items.getComponentAppendDivs(item, containerid, symbol);
        cR += '</div>';
        return cR;
    }
    ,onResize: function(item,containerid,symbol) {
        var _id = (!containerid) ? item.id : containerid+'-'+item.id;
        var _dims = prx.componentsHelper.getRealDims(item,symbol);

        $('#'+_id+' .dropdown-trigger').css('line-height', prx.componentsHelper.getProp(_dims.height,'num-other')-prx.componentsHelper.getProp(item.borderWidth,'num-border-width')*2 + 'px');

    }
    ,afterDisplay: function(item,containerid,symbol) {
        var _id = (!containerid) ? item.id : containerid+'-'+item.id;

        if(!prx.editor) {
            let selector = ' .basic-dropdown-contextmenu';

            $('#'+_id).hammer().on('click', function(e){
                e.stopPropagation();
            });

            $('#' + _id + ' .dropdown-trigger').hammer({domEvents: false}).on('tap', function () {
                if(typeof(prx.variables._triggerData['#'+_id]) == 'undefined') { prx.variables._triggerData['#'+_id] = {}; }

                if (!$('#' + _id + selector).is(':visible')) {
                    prx.helper.toggleDropdownOptions(false, '#' + _id, '#' + _id + selector);

                    //Clicked dropdown will open
                    let selectedText = prx.currentPlayerStatus.helper.get(_id, "buttons")[prx.currentPlayerStatus.helper.get(_id, "selected")].text;
                    prx.variables._triggerData['#' + _id]['selectopen'] = {value: selectedText};
                    $('#' + _id).trigger('selectopen');
                }else{
                    // The user clicks on the Dropdown arrow to close the component or opens another dropdown
                    prx.helper.closeDropdowns();
                }

                $(document).one('dummy.close-dropdowns', function() {
                    prx.helper.toggleDropdownOptions(true, '#' + _id, '#' + _id + selector);

                    let selectedText = prx.currentPlayerStatus.helper.get(_id, "buttons")[prx.currentPlayerStatus.helper.get(_id, "selected")].text;
                    prx.variables._triggerData['#'+_id]['selectclose'] = { value: selectedText };
                    $('#'+_id).trigger('selectclose');
                });
            });

            $('#' + _id + ' .basic-dropdown-item').hammer().on('tap', function (e) {
                prx.helper.toggleDropdownOptions(true, '#' + _id, '#' + _id + selector);

                let activeSelection = parseInt(e.target.dataset.dynamicPropertyIndex);
                let activeText = prx.currentPlayerStatus.helper.get(_id, "buttons")[activeSelection].text;

                $('#'+_id+' .basic-dropdown-item').removeClass('active');
                $(this).addClass('active');

                if(typeof(prx.variables._triggerData['#'+_id]) == 'undefined') { prx.variables._triggerData['#'+_id] = {}; }
                prx.variables._triggerData['#'+_id]['selectchange'] = { value: activeText };
                $('#'+_id).trigger('selectchange');
                prx.variables._triggerData['#'+_id]['selectclose'] = { value: activeText };
                $('#'+_id).trigger('selectclose');

                prx.currentPlayerStatus.helper.set(_id, 'selected', activeSelection);
            });

            prx.actions.disableFlashActionOnItemTap('#' + _id + ' .dropdown-trigger', '.flashactiontap-afterdisplay');
            prx.actions.disableFlashActionOnItemTap('#' + _id + ' .basic-dropdown-item', '.flashactiontap-afterdisplay');

        }
    }
    , mpactions: {
        specialEvents: ['selectchange', 'selectopen', 'selectclose']
    }
    , interactions: [prx.commonproperties.actions]
    ,propertyGroups: [
        {
            caption: 'Menu item style',
            properties: [
                [
                    {
                        caption: 'Background',
                        name: 'backgroundColor',
                        proptype: 'background',
                        type: 'gradients-colorpicker',
                        value: function (item, name) {
                            return item.backgroundColor;
                        },
                        liveUpdate: 'background-color',
                        changeProperty: {
                            caption: 'Background color',
                            selector: '.dropdown-trigger',
                            property: 'background',
                            transitionable: true
                        }
                    }
                ],
	            [
                    {
	                    caption: 'Border',
	                    name: 'borderWidth',
	                    proptype: 'border-width',
	                    type: 'combo-select',
	                    value: function(item,name) { return item.borderWidth; },
	                    values: { min: 0, max: 20, step: 1 } ,
                        expandableType: 'borderWidth',
	                    expandedValues: ['t', 'r', 'l', 'b'],
	                    changeProperty: {
	                        caption: 'Border width',
	                        selector: '.dropdown-trigger',
	                        property: 'border-width',
	                        transitionable: true
	                    }
	                }
                    ,{
	                    caption: false,
	                    name: 'borderStyle',
	                    proptype: 'border-style',
	                    type: 'select',
	                    value: function (item, name) {
	                        if (typeof(item.borderStyle) == 'undefined') {
	                            item.borderStyle = 'solid';
	                        }
	                        return item.borderStyle;
	                    },
	                    values: [{value: 'solid', displayValue: 'Solid'}, {
	                        value: 'dotted',
	                        displayValue: 'Dotted'
	                    }, {value: 'dashed', displayValue: 'Dashed'}, {
	                        value: 'double',
	                        displayValue: 'Double'
	                    }, {value: 'none', displayValue: 'None'}],
	                    changeProperty: {
	                        caption: 'Border style',
	                        selector: '.dropdown-trigger',
	                        property: 'border-style',
                            transitionable: false,
	                    }
	                }
                    ,{
                        name: 'borderColor',
                        proptype: 'border-color-2-active',
                        type: 'combo-colorpicker',
                        value: function(item,name) { return item.borderColor; },
                        values: prx.comps.basicColors,
                        liveUpdate: 'border-color'
                        ,changeProperty: {
                            caption: 'Border color',
                            selector: '.dropdown-trigger',
                            property: 'border-color',
                            transitionable: true,
                        }
                    }
                ],
	            [
                    prx.commonproperties.borderRadiusExpandable
                ]
            ]
        },
        {
            caption: 'Menu item text',
            properties: [
                [
                    {
                        caption: { label: 'Font family', class: 'text-properties-label text-fontfamily-label' },
                        name: 'textFont',
                        proptype: 'font-family',
                        type: 'select',
                        value: function(item, name) {
                            return typeof item.textFont == 'undefined' ? '' : item.textFont;
                        },
                        values: function() {
                            return prx.comps.fonts;
                        },
                        changeProperty: {
                            caption: 'Text font',
                            selector: '.changeProperty-textFont',
                            property: 'font-family',
                            transitionable: false
                        }
                    }
                ],
                [
                    prx.commonproperties.textFontStyle,
                    prx.commonproperties.textSize
                ],
                [
                    {
                        caption: { label: 'Underline', class: 'colorpicker-wrapper text-underline-label' },
                        name: 'textProperties',
                        proptype: 'text-properties',
                        type: 'checkbox',
                        pffSettings: prx.commonproperties.pffSettingsTextProperties,
                        value: function(item, name) {
                            if (typeof(item.textProperties) == 'undefined') {
                                item.textProperties = [];
                            }
                            return item.textProperties;
                        },
                        values: [
                            { value: 'underline', displayValue: '', icon: 'text-underline'}
                        ],
                        changeProperty: {
                            caption: 'Underline',
                            rerender: true
                        }
                    },
                    {
                        caption: { label: 'Color', class: 'text-properties-label text-color-label' },
                        name: 'textColor',
                        proptype: 'font-color',
                        type: 'solid-colorpicker',
                        relatedEditableProperties: 'text',
                        relatedCSSProperties: 'color',
                        pffSettings: prx.commonproperties.pffSettingsColor,
                        value: function(item,name) {
                            if(typeof(item.textColor) == 'undefined') { item.textColor = '#2E2E2E'; }
                            return item.textColor;
                        },
                        liveUpdate: 'color'
                        ,changeProperty: {
                            caption: ' Text color',
                            selector: '.changeProperty-textColor',
                            property: 'color',
                            transitionable: true
                        }
                    }
                ],
                [
                    prx.commonproperties.textAlign
                ]
            ]
        },
        {
            caption: 'Dropdown Icon',
            properties: [
                [
                    {
                        caption: false
                        ,name: 'buttonicon'
                        ,type: 'combo-asset'
                        ,displayValue: function(item,name) {
                            if(item.buttonicon.fileId == '') {
                                return 'No icon selected';
                            }
                            return item.buttonicon.name;
                        }
                        ,value: function(item,name) {
                            return JSON.stringify({
                                allow: 'image',
                                asset: item.buttonicon
                            });
                        }
                        ,changeProperty: {
                            caption: 'Dropdown icon',
                            transitionable: false,
                            changeFunction(item, containerid) {
                                let id = (!containerid) ? item.id : containerid + '-' + item.id;
                                let cssSelector = '#' + id + ' .dropdown-trigger-icon img';

                                prx.componentsHelper.changeImageForChangeFunction(cssSelector, item.buttonicon, false);
                            }
                        }
                    }
                ]
            ]
        },
        {
            caption: 'Dropdown style',
            properties: [
                [
                    {
                        caption: 'Background',
                        name: 'dropdownBackgroundColor',
                        proptype: 'background-color-2-dropdown',
                        type: 'gradients-colorpicker',
                        value: function (item, name) {
                            return item.dropdownBackgroundColor;
                        },
                        liveUpdate: 'background-color',
                        changeProperty: {
                            caption: 'Dropdown background color',
                            selector: '.basic-dropdown-contextmenu',
                            property: 'background-color',
                            transitionable: true
                        }
                    }
                ],
	            [
                    {
                        caption: 'Border',
                        name: 'dropdownBorderWidth',
                        proptype: 'border-width',
                        type: 'combo-select',
                        value: function(item,name) { return item.dropdownBorderWidth; },
                        values: { min: 0, max: 20, step: 1 } ,
                        changeProperty: {
                            caption: 'Dropdown border width',
                            selector: '.basic-dropdown-contextmenu',
                            property: 'border-width',
                            transitionable: true
                        }
                    }
                    ,{
                        caption: false,
                        name: 'dropdownBorderStyle',
                        proptype: 'border-style',
                        type: 'select',
                        value: function (item, name) {
                            return item.dropdownBorderStyle;
                        },
                        values: [{value: 'solid', displayValue: 'Solid'}, {
                            value: 'dotted',
                            displayValue: 'Dotted'
                        }, {value: 'dashed', displayValue: 'Dashed'}, {
                            value: 'double',
                            displayValue: 'Double'
                        }, {value: 'none', displayValue: 'None'}],
                        changeProperty: {
                            caption: 'Dropdown border style',
                            selector: '.basic-dropdown-contextmenu',
                            property: 'border-style',
                            transitionable: false
                        }
                    }
                    ,{
                        name: 'dropdownBorderColor',
                        proptype: 'border-color-2-active',
                        type: 'combo-colorpicker',
                        value: function(item,name) { return item.dropdownBorderColor; },
                        values: prx.comps.basicColors,
                        liveUpdate: 'border-color'
                        ,changeProperty: {
                            caption: 'Dropdown border color',
                            selector: '.basic-dropdown-contextmenu',
                            property: 'border-color',
                            transitionable: false,
                        changeFunction(item, containerid) {
                            let id = (!containerid) ? item.id : containerid + '-' + item.id;
                            let cssSelector = '#' + id + ' .basic-dropdown-contextmenu ';

                            prx.componentsHelper.addStyleForChangeFunction(id, id + '-border-color-2-active', cssSelector, 'border-color', item.dropdownBorderColor, 'color-border');
                        }
                        }
                    }
                ],
                [
                    {
                        caption: '<span class="icon icon-border-radius" title="Border radius"></span>',
                        name: 'dropdownBorderRadius',
                        proptype: 'border-radius',
                        type: 'combo-select',
                        value: function(item,name) {
                            return item.dropdownBorderRadius;
                        },
                        values: {min: 0, max: 20, step: 1},
                        changeProperty: {
                            caption: 'Dropdown border radius',
                            selector: '.basic-dropdown-contextmenu',
                            property: 'border-radius',
                            transitionable: true
                        }
                    }
                ],
                [
                    {
                        caption: 'Dropdown position'
                        ,name: 'dropdownPosition'
                        ,type: 'select'
                        ,value: function(item,name) {
                            return item.dropdownPosition;
                        }
                        ,values: [{value: 'left', displayValue: 'Left'},{value: 'center', displayValue: 'Center'},{value: 'right', displayValue: 'Right'},]
                        ,changeProperty: {
                            caption: 'Dropdown position',
                            rerender: true
                        }
                    }
                ]
            ]
        },
        {
            caption: 'Dropdown text',
            properties: [
                [
                    {
                        caption: { label: 'Font family', class: 'text-properties-label text-fontfamily-label' },
                        name: 'dropdownTextFont',
                        proptype: 'font-family',
                        type: 'select',
                        relatedEditableProperties: ['buttons.text'],
                        relatedCSSProperties: 'font-family',
                        value: function(item,name) {
                            if(typeof(item.dropdownTextFont) == 'undefined') { item.dropdownTextFont = 'Arial,sans-serif'; }
                            return item.dropdownTextFont;
                        },
                        values: function(){ return prx.comps.fonts; }
                        ,changeProperty: {
                            caption: 'Dropdown text font',
                            selector: '.changeProperty-dropdownTextFont',
                            property: 'font-family',
                            transitionable: false
                        }

                    }
                ],
                [
                    {
                        caption: { label: 'Font style', class: 'text-properties-label text-fontstyle-label' },
                        name: 'dropdownTextFontStyle',
                        proptype: 'font-style',
                        type: 'select',
                        pffSettings: prx.commonproperties.pffSettingsTextFontStyle,
                        value: function(item, name) {
                            return typeof item.dropdownTextFontStyle == 'undefined' ? '' : item.dropdownTextFontStyle;
                        },
                        values: function() {
                            return prx.comps.fontStyles;
                        },
                        hiddenByDefault: function(item) {
                            return false;
                        },
                        changeProperty: {
                            caption: 'Dropdown font style',
                            transitionable: false,
                            changeFunction: function(item, containerid){
                                let id = (!containerid) ? item.id : containerid + '-' + item.id;
                                let cssSelector = '#' + id + ' .basic-dropdown-contextmenu';

                                prx.componentsHelper.addStyleForChangeFunction(id, id + '-font-style-2', cssSelector, '', item.dropdownTextFontStyle, 'font-style');
                            }
                        }
                    },
                    {
                        caption: false,
                        name: 'dropdownTextSize',
                        proptype: 'font-size',
                        type: 'combo-select',
                        relatedEditableProperties: ['buttons.text'],
                        relatedCSSProperties: 'font-size',
                        value: function(item,name) {
                            if(typeof(item.dropdownTextSize) == 'undefined') { item.dropdownTextSize = 16; }
                            return item.dropdownTextSize;
                        },
                        values: prx.comps.textsize
                        ,changeProperty: {
                            caption: 'Dropdown text size',
                            selector: '.changeProperty-dropdownTextSize',
                            property: 'font-size',
                            transitionable: false
                        },
                        onChange: function(item) {
                            if(item.lineHeightAuto==true) {
                                item.lineHeight = parseInt(prx.componentsHelper.getProp(item.dropdownTextSize,'num-text-size')*1.231);
                            }
                        }
                    }
                ]
                ,
                [
                    {
                        caption: { label: 'Underline', class: 'colorpicker-wrapper text-underline-label' },
                        name: 'dropdownTextProperties',
                        proptype: 'text-properties-dropdown',
                        type: 'checkbox',
                        pffSettings: prx.commonproperties.pffSettingsTextProperties,
                        value: function(item, name) {
                            if (typeof(item.dropdownTextProperties) == 'undefined') {
                                item.dropdownTextProperties = [];
                            }
                            return item.dropdownTextProperties;
                        },
                        values: [
                            { value: 'underline', displayValue: '', icon: 'text-underline'}
                        ],
                        changeProperty: {
                            caption: 'Dropdown underline',
                            transitionable: false,
                            changeFunction: function(item, containerid){
                                let id = (!containerid) ? item.id : containerid + '-' + item.id;
                                let cssSelector = '#' + id + ' .basic-dropdown-contextmenu';

                                prx.componentsHelper.addStyleForChangeFunction(id, id + '-dropdownTextProperties', cssSelector, '', item.dropdownTextProperties, 'props-text');
                            }
                        }
                    },
                    {
                        caption: { label: 'Color', class: 'text-properties-label text-color-label' },
                        name: 'dropdownTextColor',
                        proptype: 'font-color',
                        type: 'solid-colorpicker',
                        relatedEditableProperties: ['buttons.text'],
                        relatedCSSProperties: 'color',
                        pffSettings: prx.commonproperties.pffSettingsColor,
                        value: function(item,name) {
                            return item.dropdownTextColor;
                        },
                        liveUpdate: 'color'
                        ,changeProperty: {
                            caption: 'Dropdown text color',
                            selector: '.changeProperty-dropdownTextColor',
                            property: 'color',
                            transitionable: true
                        }
                    }
                ],
                [
                    {
                        caption: { label: 'Alignment', class: 'text-properties-label text-alignment-label' },
                        name: 'dropdownTextAlign',
                        proptype: 'text-align',
                        type: 'radio',
                        relatedEditableProperties: ['buttons.text'],
                        relatedCSSProperties: 'text-align',
                        pffSettings: prx.commonproperties.pffSettingsTextAlignment,
                        value: function(item,name) {
                            return item.dropdownTextAlign;
                        },
                        values: [
                            { value: 'left', displayValue: '', icon: 'align-left'},
                            { value: 'center', displayValue: '', icon: 'align-center'},
                            { value: 'right', displayValue: '', icon: 'align-right'},
                            { value: 'justify', displayValue: '', icon: 'align-justify'}
                        ],
                        changeProperty: {
                            caption: 'Dropdown text align',
                            selector: '.changeProperty-dropdownTextAlign',
                            property: 'text-align',
                            transitionable: false
                        }
                    }
                ]
            ]
        },
        {
            caption: 'Selected option',
            properties: [
                [
                    {
                        caption: 'Background',
                        name: 'dropdownActiveBackgroundColor',
                        proptype: 'background-color-2-active',
                        type: 'gradients-colorpicker',
                        value: function (item, name) {
                            return item.dropdownActiveBackgroundColor;
                        },
                        liveUpdate: 'background-color',
                        changeProperty: {
                            caption: 'Selected background color',
                            selector: '.active',
                            property: 'background-color',
                            transitionable: true
                        }
                    },
                    {
                        caption: 'Text',
                        name: 'dropdownActiveTextColor',
                        proptype: 'font-color-2-active',
                        type: 'solid-colorpicker',
                        relatedEditableProperties: 'text',
                        relatedCSSProperties: 'color',
                        value: function(item,name) {
                            return item.dropdownActiveTextColor;
                        },
                        liveUpdate: 'color'
                        ,changeProperty: {
                            caption: ' Selected text color',
                            selector: '.active',
                            property: 'color',
                            transitionable: true
                        }
                    },
                ],
                [
                    {
                        caption: 'Selected option'
                        ,name: 'selected'
                        ,type: 'select'
                        ,value: function(item,name) {
                            return item.selected;
                        }
                        ,values: function(item,name) {
                            var _rA = [{displayValue: 'None', value: -1}];
                            for (var n = 0; n < item.buttons.length; n++) {
                                _rA.push({value: n,displayValue: item.buttons[n].text});
                            }
                            return _rA;
                        }
                        ,changeProperty: {
                            caption: 'Active option',
                            transitionable: false,
                            changeFunction(item, containerid) {
                                let id = (!containerid) ? item.id : containerid + '-' + item.id;
                                let cssSelector = '#' + id + ' .basic-dropdown-item';
                                let items = $(cssSelector);

                                $.each(items, function (index, _item) {
                                    $(_item).removeClass('active');
                                    if (index === parseInt(item.selected)){
                                        $(_item).addClass('active');
                                    }
                                });
                            }
                        }
                    }
                ]
            ]

        },
        {
            caption: 'Hover styles',
            properties: [
                [
                    {
                        caption: 'Inactive Background',
                        name: 'hoverInactiveBackgroundColor',
                        proptype: 'background-color-3-inactive',
                        type: 'gradients-colorpicker',
                        value: function (item, name) {
                            return item.hoverInactiveBackgroundColor;
                        },
                        liveUpdate: 'background-color',
                        changeProperty: {
                            caption: 'Hover inactive background color',
                            transitionable: false,
                            changeFunction(item, containerid) {
                                let id = (!containerid) ? item.id : containerid + '-' + item.id;
                                let cssSelector = '#' + id + ' .basic-dropdown-item:hover';

                                prx.componentsHelper.addStyleForChangeFunction(id, id + '-hoverInactiveBackgroundColor', cssSelector, 'background-color', item.hoverInactiveBackgroundColor, 'color-background');
                            }
                        }
                    },
                    {
                        caption: 'Text',
                        name: 'hoverInactiveTextColor',
                        proptype: 'font-color-3-inactive',
                        type: 'solid-colorpicker',
                        relatedEditableProperties: 'text',
                        relatedCSSProperties: 'color',
                        value: function(item,name) {
                            return item.hoverInactiveTextColor;
                        },
                        liveUpdate: 'color',
                        changeProperty: {
                            caption: 'Hover inactive text color',
                            transitionable: false,
                            changeFunction(item, containerid) {
                                let id = (!containerid) ? item.id : containerid + '-' + item.id;
                                let cssSelector = '#' + id + ' .basic-dropdown-item:hover';

                                prx.componentsHelper.addStyleForChangeFunction(id, id + '-hoverInactiveTextColor', cssSelector, 'color', item.hoverInactiveTextColor, 'color-text');
                            }
                        }
                    },
                ],
                [
                    {
                        caption: 'Active Background',
                        name: 'hoverActiveBackgroundColor',
                        proptype: 'background-color-3-active',
                        type: 'gradients-colorpicker',
                        value: function (item, name) {
                            return item.hoverActiveBackgroundColor;
                        },
                        liveUpdate: 'background-color',
                        changeProperty: {
                            caption: 'Hover active background color',
                            transitionable: false,
                            changeFunction(item, containerid) {
                                let id = (!containerid) ? item.id : containerid + '-' + item.id;
                                let cssSelector = '#' + id + ' .basic-dropdown-item.active:hover';

                                prx.componentsHelper.addStyleForChangeFunction(id, id + '-hoverActiveBackgroundColor', cssSelector, 'background-color', item.hoverActiveBackgroundColor, 'color-background');
                            }
                        }
                    },
                    {
                        caption: 'Text',
                        name: 'hoverActiveTextColor',
                        proptype: 'font-color-3-active',
                        type: 'solid-colorpicker',
                        relatedEditableProperties: 'text',
                        relatedCSSProperties: 'color',
                        value: function(item,name) {
                            return item.hoverActiveTextColor;
                        },
                        liveUpdate: 'color',
                        changeProperty: {
                            caption: 'Hover active text color',
                            transitionable: false,
                            changeFunction(item, containerid) {
                                let id = (!containerid) ? item.id : containerid + '-' + item.id;
                                let cssSelector = '#' + id + ' .basic-dropdown-item.active:hover';

                                prx.componentsHelper.addStyleForChangeFunction(id, id + '-hoverActiveTextColor', cssSelector, 'color', item.hoverActiveTextColor, 'color-text');
                            }
                        }
                    },
                ],
            ]
        }
    ]
    ,editableProperties: [
        {
            caption: 'Menu Item'
            ,name: 'text'
            ,type: 'input'
            ,value: function(item,name,index) {
                return item.text;
            }
            ,liveUpdate:'color'
            ,changeProperty: {
                caption: 'Text',
                transitionable: false,
                property: 'text',
                selector: '.basic-dropdown-active-value', //do not delete, even it is rerender we need this for the changetextprop and changetextfont style functions
                changeFunction(item, containerid) {
                    let id = (!containerid) ? item.id : containerid + '-' + item.id;
                    let cssSelector = '#' + id + ' .dropdown-trigger .basic-dropdown-active-value';

                    $(cssSelector).html(item.text);
                }
            }
        }
    ]
    ,dynamicProperties: {
        data: 'buttons'
        ,propertyCaption: 'Menu items'
        ,propertyName: 'Menu item'
        ,addCaption: 'Add menu item'
        ,deleteCaption: 'Delete'
        ,blankItem: {
            text: 'Label',
            actions: []
        }
        ,captionProperty: 'text'

        ,editableProperties:[
            {
                caption: 'Text'
                ,name: 'text'
                ,type: 'input'
                ,property: 'text'
                ,value: function(item,name,index) {
                    return item.buttons[index].text;
                }
                ,liveUpdate:'color'
                ,changeProperty: {
                    caption: 'Text',
                    transitionable: false,
                    property: 'text',
                    selector: '.basic-dropdown-item', //do not delete, even it is rerender we need this for the changetextprop and changetextfont style functions
                    changeFunction(item, containerid) {
                        let id = (!containerid) ? item.id : containerid + '-' + item.id;
                        let cssSelector = '#' + id + ' .basic-dropdown-item .error-msg span';

                        $.each($(cssSelector), function (key, _item) {
                            $(_item).html(item.buttons[key].text);
                        });
                    }
                }
            }
        ]
        ,interactions:	[ prx.commonproperties.actions_buttons ]
        ,propertyGroups: [
            {
                caption: '',
                properties: [
                    [

                    ]
                ]
            }
        ]

    }
};

// TYPE: PAGE CONTROLLER
prx.types.basic_pagecontroller = {
    name: 'basic_pagecontroller'
    ,onDisplay: function(item,containerid,symbol) {
        var _id = (!containerid) ? item.id : containerid+'-'+item.id;
        var _dims = prx.componentsHelper.getRealDims(item,symbol);

        if(typeof(item.changeActive) == 'undefined') { item.changeActive = true; }

        var _props = prx.componentsHelper.getProp(item.textProperties,'props-text');

        var cR = '';
        cR += '<div id="' + _id + '" ' + prx.items.getComponentBaseAttributes(item, containerid, symbol)  + ' class="' + prx.items.getComponentBaseClasses(item, containerid, symbol) + ' box pos type-basic-pagecontroller">';

        cR += '<style>';
        cR += prx.items.getComponentBaseStyle(item, containerid, symbol);
        cR += '#'+_id+' .basic-pagecontroller-button label { background: '+prx.componentsHelper.getProp(item.backgroundColor,'color-background')+'; border-radius: '+prx.componentsHelper.getProp(item.borderRadius,'border-radius')+'; width: '+_dims.height+'px; }';
        cR += '#'+_id+' .basic-pagecontroller-button input:checked + label { transform: scale('+(item.activeSize)+'); background: '+prx.componentsHelper.getProp(item.activeBackgroundColor,'color-background')+'; }';
        cR += '</style>';
        cR += prx.items.getComponentPrependDivs(item, containerid, symbol);

        cR += '<div class="basic-pagecontroller-inner">';
        $.each(item.buttons, function(i,elm) {
            cR += '<div class="basic-pagecontroller-button dynamic-property" data-dynamic-property-index="'+i+'" id="'+_id+'-buttons-'+i+'">';
            cR += '<input type="radio" name="'+_id+'-radio-input" id="'+_id+'-radio-'+i+'" '+((prx.componentsHelper.getProp(item.selected,'num-other') == i) ? 'checked' : '')+' data-role="none" ' +((!prx.componentsHelper.getProp(item.changeActive,'num-other')) ? 'disabled' : '')+ '>';
            cR += '<label for="'+_id+'-radio-'+i+'" class="changeProperty-borderRadius '+((prx.componentsHelper.getProp(item.selected,'num-other') == i) ? 'changeProperty-activeBackgroundColor liveUpdate-activeBackgroundColor-background' : 'changeProperty-backgroundColor liveUpdate-backgroundColor-background')+'" ></label>';
            cR += '</div>';
        });
        cR += '</div>';
        cR += prx.items.getComponentAppendDivs(item, containerid, symbol);
        cR += '</div>';
        return cR;
    },
    onResize: function(item,containerid,symbol) {
        var _id = (!containerid) ? item.id : containerid+'-'+item.id;
        var _dims = prx.componentsHelper.getRealDims(item,symbol);

        $('#'+_id+' label').width(_dims.height);
    },
    propertyGroups: [
        {
            caption: 'Style',
            properties: [
                [
                    {
                        caption: 'Inactive',
                        proptype: 'background' ,
                        name: 'backgroundColor',
                        type: 'solid-colorpicker',
                        value: function(item,name) {
                            return item.backgroundColor;
                        },
                        values: prx.comps.basicColors,
                        liveUpdate: 'background',
                        changeProperty: {
                            caption: 'Background color',
                            selector: '.changeProperty-backgroundColor',
                            property: 'background',
                            transitionable: true
                        }
                    },
                    {
                        caption: 'Active',
                        name: 'activeBackgroundColor',
                        proptype: 'background',
                        type: 'solid-colorpicker',
                        value: function(item,name) { return item.activeBackgroundColor; },
                        values: prx.comps.basicColors,
                        liveUpdate: 'background',
                        changeProperty: {
                            caption: 'Active background color',
                            selector: '.changeProperty-activeBackgroundColor',
                            property: 'background',
                            transitionable: true
                        }
                    }
                ]
                ,[
                    {
                        caption: 'Active Button Scale'
                        ,name: 'activeSize'
                        ,type: 'slider-input'
                        ,value: function(item,name) {
                            return item.activeSize;
                        }
                        ,values: { min: 1, max: 2, step: 0.1 }
                        ,changeProperty: {
                            caption: 'Active size',
                            transitionable: false,
                            changeFunction (item, containerid) {
                                    let id = (!containerid) ? item.id : containerid + '-' + item.id;
                                    let cssSelector = '#' + id + ' .basic-pagecontroller-button input:checked + label';

                                    prx.componentsHelper.addStyleForChangeFunction(id, id + '-activeButtonScale', cssSelector, 'transform', 'scale(' + item.activeSize + ')', 'other');
                            }
                        }
                    }
                ]
                ,[
                    {
                        caption: 'Border <span class="icon icon-border-radius" title="Border radius"></span>',
                        name: 'borderRadius',
                        proptype: 'border-radius',
                        type: 'combo-select',
                        value: function(item,name) { return item.borderRadius; },
                        values: { min: 0, max: 20, step: 1 },
                        expandableType: 'borderRadius',
                        expandedValues: ['tl', 'tr', 'bl', 'br'],
                        changeProperty: {
                            caption: 'Border radius',
                            selector: '.changeProperty-borderRadius',
                            property: 'border-radius',
                            transitionable: true
                        }
                    }
                ]
            ]
        },
        {
            caption: 'Advanced',
            properties: [
                [
                    {
                        caption: 'Active Button'
                        ,name: 'selected'
                        ,type: 'select'
                        ,value: function(item,name) {
                            return item.selected;
                        }
                        ,values: function(item,name) {
                            var _rA = [];
                            //var _rA = [{value: '999',displayValue: 'None'}];
                            for (var n = 0; n < item.buttons.length; n++) {
                                _rA.push({value: n,displayValue: 'Button ' + (n+1) });
                            }
                            return _rA;
                        }
                        ,changeProperty: {
                            caption: 'Active button',
                            transitionable: false,
                            changeFunction (item, containerid) {
                                item.buttons.forEach(function (button, idx) {
                                    let id = (!containerid) ? item.id : containerid + '-' + item.id;
                                    let cssSelector = '#' + id + ' .basic-pagecontroller-button input' + '#' + id + '-radio-' + idx;

                                    if (item.selected == idx) {
                                        var isInitiallyDisabled = $(cssSelector)[0].disabled;

                                        if(isInitiallyDisabled) {
                                            $(cssSelector)[0].disabled = false;
                                        }
                                        $(cssSelector)[0].click();
                                        if(isInitiallyDisabled) {
                                            $(cssSelector)[0].disabled = true;
                                        }
                                    }
                                });
                            }
                        }

                    }
                ],
                [
                    {
                        caption: 'Change active state on click'
                        ,name: 'changeActive'
                        ,type: 'onoff'
                        ,value: function(item) {
                            return item.changeActive;
                        }
                        ,changeProperty: {
                            caption: 'Change active state on click',
                            transitionable: false,
                            changeFunction (item, containerid) {
                                item.buttons.forEach(function (button, idx) {
                                    let id = (!containerid) ? item.id : containerid + '-' + item.id;
                                    let cssSelector = '#' + id + ' .basic-pagecontroller-button input' + '#' + id + '-radio-' + idx;

                                    $(cssSelector)[0].disabled = !item.changeActive;
                                });
                            }
                        }
                    }
                ]
            ]
        }
    ]
    ,dynamicProperties: {
        data: 'buttons'
        ,propertyCaption: 'Buttons'
        ,propertyName: 'Button'
        ,addCaption: 'Add button'
        ,deleteCaption: 'Delete'
        ,blankItem: {
            text: 'Label',
            actions: []
        }
        ,interactions: [ prx.commonproperties.actions_buttons ]
    }
};

//TYPE: TEXTFIELD
prx.types.basic_textfield = {
    name: 'basic_textfield'
    ,onDisplay: function(item,containerid,symbol) {
        var _id = (!containerid) ? item.id : containerid+'-'+item.id;
        if (typeof(item.borderStyle) == 'undefined') {
            item.borderStyle = 'solid';
        }
        var _props = prx.componentsHelper.getProp(item.textProperties,'props-text');
        _props += prx.componentsHelper.getProp(item.textFontStyle,'props-text-style');
        var _dims = prx.componentsHelper.getRealDims(item, symbol);
        var cR = '';

        cR += '<div id="' + _id + '" ' + prx.items.getComponentBaseAttributes(item, containerid, symbol)  + ' class="' + prx.items.getComponentBaseClasses(item, containerid, symbol) + ' box pos type-basic-textfield type-basic-textfield-'+prx.componentsHelper.getProp(item.inputtype,'text-other')+'">';
        cR += '<style>';
        cR += prx.items.getComponentBaseStyle(item, containerid, symbol);
        cR += '#'+_id+' input, #'+_id+' .faux-input { '+prx.gradients.getBgCssByProperty(item.backgroundColor,'color-background')+'; color: '+prx.componentsHelper.getProp(item.textColor,'color-text')+'; font-size: '+prx.componentsHelper.getProp(item.textSize,'num-text-size')+'px; '+prx.componentsHelper.getProp(item.textFont + '|' + item.textFontStyle,'font-family')+'; border-width: '+prx.componentsHelper.getProp(item.borderWidth,'border-width')+'; border-style: '+prx.componentsHelper.getProp(item.borderStyle,'border-style')+'; border-color: '+prx.componentsHelper.getProp(item.borderColor,'color-border')+'; border-radius: '+prx.componentsHelper.getProp(item.borderRadius,'border-radius')+'; text-align: '+prx.componentsHelper.getProp(item.textAlign,'align')+'; '+ _props + ' }';
        cR += '#'+_id+' .faux-input.placeholder-input { color: '+prx.componentsHelper.getProp(item.placeholderColor,'color-text')+'; }';
        cR += '#'+_id+' .faux-input { '+prx.css.flexJustifyContent(prx.componentsHelper.getProp(item.textAlign,'align'))+'; }';
        cR += '#'+_id+' input:-moz-placeholder { color: '+prx.componentsHelper.getProp(item.placeholderColor,'color-text')+'!important;'+_props+' }';
        cR += '#'+_id+' input::-moz-placeholder { color: '+prx.componentsHelper.getProp(item.placeholderColor,'color-text')+'!important;'+_props+' }';
        cR += '#'+_id+' input::-webkit-input-placeholder { color: '+prx.componentsHelper.getProp(item.placeholderColor,'color-text')+'!important;'+_props+' }';
        cR += '</style>';
        cR += prx.items.getComponentPrependDivs(item, containerid, symbol);

        if(prx.editor) {
            cR += '<div class="faux-input liveUpdate-textColor-color liveUpdate-borderColor-border-color liveUpdate-borderStyle-border-style liveUpdate-backgroundColor-background-color changeProperty-backgroundColor changeProperty-textColor changeProperty-textSize changeProperty-textFont changeProperty-borderColor changeProperty-borderWidth changeProperty-borderRadius changeProperty-textAlign" data-editableproperty="value">'+prx.componentsHelper.getProp(item.value,'text')+'</div>';
            cR += '<div class="faux-input placeholder-input liveUpdate-placeholderColor-color liveUpdate-borderColor-border-color liveUpdate-borderStyle-border-style liveUpdate-backgroundColor-background-color changeProperty-backgroundColor changeProperty-textColor changeProperty-textSize changeProperty-textFont changeProperty-borderColor changeProperty-borderWidth changeProperty-borderRadius changeProperty-textAlign">'+prx.componentsHelper.getProp(item.placeholder,'other')+'</div>';
        } else {
            cR += '<input autocomplete="new-password" data-lpignore="true" type="'+prx.componentsHelper.getProp(item.inputtype,'other')+'" value="'+prx.componentsHelper.getProp(item.value,'other')+'" placeholder="'+prx.componentsHelper.getProp(item.placeholder,'other')+'" data-role="none" class="real-input changeProperty-backgroundColor changeProperty-textColor changeProperty-textSize changeProperty-textFont changeProperty-borderColor changeProperty-borderWidth changeProperty-borderRadius changeProperty-textAlign" />';
        }
        cR += prx.items.getComponentAppendDivs(item, containerid, symbol);
        cR += '</div>';
        return cR;
    }
    ,afterDisplay: function(item,containerid,symbol) {
        var _id = (!containerid) ? item.id : containerid+'-'+item.id;

        if(!prx.editor) {
            $('#'+_id)
                .hammer()
                .find('.real-input')
                .focus(function(){
                    if(typeof(prx.variables._triggerData['#'+_id]) == 'undefined') { prx.variables._triggerData['#'+_id] = {}; }
                    prx.variables._triggerData['#'+_id]['inputfocus'] = { value: $(this).val() };
                    $('#'+_id).trigger('inputfocus');
                })
                .blur(function(){
                    if(typeof(prx.variables._triggerData['#'+_id]) == 'undefined') { prx.variables._triggerData['#'+_id] = {}; }
                    prx.variables._triggerData['#'+_id]['inputblur'] = { value: $(this).val() };
                    $('#'+_id).trigger('inputblur');
                })
                .keyup(function(e){
                    if(typeof(prx.variables._triggerData['#'+_id]) == 'undefined') { prx.variables._triggerData['#'+_id] = {}; }
                    prx.variables._triggerData['#'+_id]['inputkeyup'] = { value: $(this).val() };
                    var event = $.Event('inputkeyup');
                    event.which = e.which;
                    $('#'+_id).trigger(event);

                    prx.currentPlayerStatus.helper.set(_id, 'value', e.target.value);
                });

            prx.actions.disableFlashActionOnItemTap('#' + _id, '.flashactiontap-afterdisplay');

        }
    }
    ,interactions: [ prx.commonproperties.actions ]
    ,mpactions: {
        specialEvents: ['inputfocus','inputblur','inputkeyup']
    }
    ,editableProperties: [
        {
            caption: 'Value'
            ,name: 'value'
            ,type: 'input'
            ,isRichText: false
            ,value: function(item,name) {
                return item.value;
            }
            ,changeProperty: {
                caption: 'Value',
                property: 'input-value',
                selector: 'input.real-input',
                transitionable: false
            }
        }
    ]
    ,propertyGroups: [
        {
            caption: 'Style',
            properties: [
                [
                    {
                        caption: 'Background',
                        name: 'backgroundColor',
                        proptype: 'background-color',
                        type: 'gradients-colorpicker',
                        value: function(item, name) {
                            return typeof item.backgroundColor == 'undefined' ? '' : item.backgroundColor;
                        },
                        liveUpdate: 'background-color',
                        changeProperty: {
                            caption: 'Background color',
                            selector: '.changeProperty-backgroundColor',
                            property: 'background-color',
                            transitionable: true
                        }
                    }
                ]
                ,[
                    {
                        caption: 'Border',
                        name: 'borderWidth',
                        proptype: 'border-width',
                        type: 'combo-select',
                        value: function(item,name) { return item.borderWidth; },
                        values: { min: 0, max: 20, step: 1 } ,
                        expandableType: 'borderWidth',
                        expandedValues: ['t', 'r', 'l', 'b'],
                        changeProperty: {
                            caption: 'Border Width',
                            selector: 'input',
                            property: 'border-width',
                            transitionable: true
                        }
                    }
                    , {
                        caption: false,
                        name: 'borderStyle',
                        proptype: 'border-style',
                        type: 'select',
                        liveUpdate: 'border-style',
                        value: function (item, name) {
                            if (typeof(item.borderStyle) == 'undefined') {
                                item.borderStyle = 'solid';
                            }
                            return item.borderStyle;
                        },
                        values: [{value: 'solid', displayValue: 'Solid'}, {
                            value: 'dotted',
                            displayValue: 'Dotted'
                        }, {value: 'dashed', displayValue: 'Dashed'}, {
                            value: 'double',
                            displayValue: 'Double'
                        }, {value: 'none', displayValue: 'None'}],
                        changeProperty: {
                            caption: 'Border Style',
                            selector: '.changeProperty-borderWidth',
                            property: 'border-style',
                            transitionable: false
                        }
                    }
                    ,prx.commonproperties.borderColor
                    ,prx.commonproperties.borderRadiusExpandable
                ]
            ]
        },
        {
            caption: 'Text',
            properties: [
                [
                    prx.commonproperties.textFont
                ],
                [
                    prx.commonproperties.textFontStyle,
                    prx.commonproperties.textSize
                ],
                [
                    prx.commonproperties.textProperties,
                    {
                        caption: { label: 'Color', class: 'text-properties-label text-color-label' },
                        name: 'textColor',
                        proptype: 'font-color',
                        type: 'solid-colorpicker',
                        relatedEditableProperties: 'text',
                        relatedCSSProperties: 'color',
                        pffSettings: prx.commonproperties.pffSettingsColor,
                        value: function(item,name) {
                            if(typeof(item.textColor) == 'undefined') { item.textColor = '#2E2E2E'; }
                            return item.textColor;
                        },
                        liveUpdate: 'color'
                        ,changeProperty: {
                            caption: 'Text color',
                            selector: '.changeProperty-textColor',
                            property: 'color',
                            transitionable: true
                        }
                    }
                ],
                [
                    prx.commonproperties.textAlign
                ]
            ]
        },
        {
            caption: 'Placeholder (If field is empty)',
            properties: [
                [
                    {
                        caption: 'Placeholder Text',
                        name: 'placeholder',
                        type: 'input',
                        value: function(item,name) {
                            return item.placeholder;
                        }
                        ,changeProperty: {
                            caption: 'Placeholder text',
                            transitionable: false,
                            changeFunction: function(item, containerid) {
                                var _id = (!containerid) ? item.id : containerid+'-'+item.id;

                                if(prx.editor) {
                                    $('#' + _id + ' .placeholder-input').text(prx.componentsHelper.getProp(item.placeholder,'other'));
                                } else {
                                    $('#' + _id + ' input').attr('placeholder', prx.componentsHelper.getProp(item.placeholder,'other'));
                                }

                            }
                        }
                    }
                ],
                [
                    {
                        caption: 'Placeholder Color',
                        name: 'placeholderColor',
                        proptype: 'placeholder-color',
                        type: 'combo-colorpicker',
                        value: function(item,name) { if(typeof(item.placeholderColor)=='undefined') { return '999999'; } return item.placeholderColor; }
                        ,values: prx.comps.basicColors
                        ,liveUpdate:'color'
                        ,changeProperty: {
                            caption: 'Placeholder color',
                            transitionable: false,
                            changeFunction: function(item, containerid) {
                                var _id = (!containerid) ? item.id : containerid+'-'+item.id;

                                var cR = '';
                                cR += '#'+_id+' .faux-input.placeholder-input { color: '+prx.componentsHelper.getProp(item.placeholderColor,'color-text')+'; }';
                                cR += '#'+_id+' input:-moz-placeholder { color: '+prx.componentsHelper.getProp(item.placeholderColor,'color-text')+'!important; }';
                                cR += '#'+_id+' input::-moz-placeholder { color: '+prx.componentsHelper.getProp(item.placeholderColor,'color-text')+'!important; }';
                                cR += '#'+_id+' input::-webkit-input-placeholder { color: '+prx.componentsHelper.getProp(item.placeholderColor,'color-text')+'!important; }';

                                $('#' + _id).append('<style>' + cR + '</style>')
                            }
                        }
                    }
                ]
            ]
        },
        {
            caption: 'Input type',
            properties: [
                [
                    {
                        caption: false,
                        name: 'inputtype',
                        type: 'select',
                        value: function(item,name) {
                            return item.inputtype;
                        }
                        ,values: [{ value: 'text', displayValue: 'Text' }, { value: 'number', displayValue: 'Numeric' }, { value: 'email', displayValue: 'Email' }, { value: 'password', displayValue: 'Password' }, { value: 'tel', displayValue: 'Telephone' }]
                        ,changeProperty: {
                            caption: 'Input type',
                            transitionable: false,
                            changeFunction(item, containerid) {
                                let id = (!containerid) ? item.id : containerid + '-' + item.id;
                                $('#' + id + ' input')[0].type = item.inputtype;
                            }
                        }
                        ,hiddenByDefault: function(item) {
                            return (item.name == 'basic_passwordfield');
                        }
                    }
                ]
            ]
        }
    ]

};

//TYPE: TEXTAREA
prx.types.basic_textarea = {
    name: 'basic_textarea'
    ,onDisplay: function(item,containerid,symbol) {
        var _id = (!containerid) ? item.id : containerid+'-'+item.id;

        var _props = prx.componentsHelper.getProp(item.textProperties,'props-text');
        _props += prx.componentsHelper.getProp(item.textFontStyle,'props-text-style');
        var _dims = prx.componentsHelper.getRealDims(item, symbol);
        if (typeof(item.borderStyle) == 'undefined') {
            item.borderStyle = 'solid';
        }
        var cR = '';

        cR += '<div id="' + _id + '" ' + prx.items.getComponentBaseAttributes(item, containerid, symbol)  + ' class="' + prx.items.getComponentBaseClasses(item, containerid, symbol) + ' box pos type-basic-textarea">';
        cR += '<style>';
        cR += prx.items.getComponentBaseStyle(item, containerid, symbol);
        cR += '#'+_id+' > textarea, #'+_id+' .faux-input { background: '+prx.componentsHelper.getProp(item.backgroundColor,'color-background')+'; color: '+prx.componentsHelper.getProp(item.textColor,'color-text')+'; font-size: '+prx.componentsHelper.getProp(item.textSize,'num-text-size')+'px;  '+prx.componentsHelper.getProp(item.textFont,'font-family')+'  border-width: '+prx.componentsHelper.getProp(item.borderWidth,'border-width')+'; border-style: '+prx.componentsHelper.getProp(item.borderStyle,'border-type')+'; border-color: '+prx.componentsHelper.getProp(item.borderColor,'color-border')+'; text-align: '+prx.componentsHelper.getProp(item.textAlign,'align')+'; border-radius: '+prx.componentsHelper.getProp(item.borderRadius,'border-radius')+';  '+ _props + '}';
        cR += '#'+_id+' .faux-input { '+prx.css.flexJustifyContent(prx.componentsHelper.getProp(item.textAlign,'align'))+'; }';
        cR += '#'+_id+' .faux-input.placeholder-input { color: '+prx.componentsHelper.getProp(item.placeholderColor,'color-text')+'; }';
        cR += '#'+_id+' textarea:-moz-placeholder { color: '+prx.componentsHelper.getProp(item.placeholderColor,'color-text')+'!important; '+ _props + '}';
        cR += '#'+_id+' textarea::-webkit-input-placeholder { color: '+prx.componentsHelper.getProp(item.placeholderColor,'color-text')+'!important; '+ _props + '}';
        cR += '#'+_id+' textarea::-moz-placeholder { color: '+prx.componentsHelper.getProp(item.placeholderColor,'color-text')+'!important; '+ _props + '}';
        cR += '</style>';
        cR += prx.items.getComponentPrependDivs(item, containerid, symbol);
        if(prx.editor) {
            cR += '<div class="faux-input liveUpdate-textColor-color liveUpdate-borderColor-border-color liveUpdate-backgroundColor-background changeProperty-backgroundColor changeProperty-borderColor changeProperty-borderWidth changeProperty-borderRadius changeProperty-textSize changeProperty-textFont changeProperty-textFontStyle changeProperty-textColor changeProperty-textAlign" data-editableproperty="value">'+prx.componentsHelper.getProp(item.value,'text-textarea')+'</div>';
            cR += '<div class="faux-input placeholder-input liveUpdate-placeholderColor-color liveUpdate-borderColor-border-color liveUpdate-backgroundColor-background changeProperty-backgroundColor changeProperty-borderColor changeProperty-borderWidth changeProperty-borderRadius changeProperty-textSize changeProperty-textFont changeProperty-textFontStyle changeProperty-textColor changeProperty-textAlign">'+prx.componentsHelper.getProp(item.placeholder,'other')+'</div>';
        } else {
            cR += '<textarea class="real-input liveUpdate-placeholderColor-color liveUpdate-backgroundColor-background liveUpdate-textColor-color liveUpdate-borderColor-border-color changeProperty-backgroundColor changeProperty-borderColor changeProperty-borderWidth changeProperty-borderRadius changeProperty-textSize changeProperty-textFont changeProperty-textFontStyle changeProperty-textColor changeProperty-textAlign" placeholder="'+prx.componentsHelper.getProp(item.placeholder,'other')+'" data-role="none">'+prx.componentsHelper.getProp(item.value,'text-textarea').replace(/<br \/>/g, '\n')+'</textarea>';
        }
        cR += prx.items.getComponentAppendDivs(item, containerid, symbol);
        cR += '</div>';
        return cR;
    }
    ,afterDisplay: function(item,containerid,symbol) {
        var _id = (!containerid) ? item.id : containerid+'-'+item.id;

        if(!prx.editor) {
            $('#'+_id)
                .hammer()
                .find('textarea')
                .focus(function(){
                    if(typeof(prx.variables._triggerData['#'+_id]) == 'undefined') { prx.variables._triggerData['#'+_id] = {}; }
                    prx.variables._triggerData['#'+_id]['inputfocus'] = { value: $(this).val() };
                    $('#'+_id).trigger('inputfocus');
                })
                .blur(function(){
                    if(typeof(prx.variables._triggerData['#'+_id]) == 'undefined') { prx.variables._triggerData['#'+_id] = {}; }
                    prx.variables._triggerData['#'+_id]['inputblur'] = { value: $(this).val() };
                    $('#'+_id).trigger('inputblur');
                })
                .keyup(function(e){
                    if(typeof(prx.variables._triggerData['#'+_id]) == 'undefined') { prx.variables._triggerData['#'+_id] = {}; }
                    prx.variables._triggerData['#'+_id]['inputkeyup'] = { value: $(this).val() };
                    var event = $.Event('inputkeyup');
                    event.which = e.which;
                    $('#'+_id).trigger(event);

                    prx.currentPlayerStatus.helper.set(_id, 'value', e.target.value);
                });

            prx.actions.disableFlashActionOnItemTap('#' + _id, '.flashactiontap-afterdisplay');
        }
    }
    ,editableProperties: [
        {
            caption: 'Value'
            ,name: 'value'
            ,type: 'textarea'
            ,isRichText: false
            ,value: function(item,name) {
                return item.value;
            }
            ,changeProperty: {
                caption: 'Value',
                property: 'textarea-value',
                selector: 'textarea',
                transitionable: false
            }
        }
    ]
    ,interactions: [ prx.commonproperties.actions ]
    ,mpactions: {
        specialEvents: ['inputfocus','inputblur','inputkeyup']
    }
    ,propertyGroups:	[
        {
            caption: 'Style',
            properties: [
                [
                    {
                        caption: 'Background',
                        name: 'backgroundColor',
                        proptype: 'background',
                        type: 'gradients-colorpicker',
                        value: function(item, name) {
                            return typeof item.backgroundColor == 'undefined' ? '' : item.backgroundColor;
                        },
                        liveUpdate: 'background',
                        changeProperty: {
                            caption: 'Background color',
                            selector: '.changeProperty-backgroundColor',
                            property: 'background',
                            transitionable: true
                        }
                    }
                ],
                [
                    prx.commonproperties.borderWidthExpandable
                    , {
                    caption: false,
                    name: 'borderStyle',
                    proptype: 'border-style',
                    type: 'select',
                    value: function (item, name) {
                        if (typeof(item.borderStyle) == 'undefined') {
                            item.borderStyle = 'solid';
                        }
                        return item.borderStyle;
                    },
                    values: [{value: 'solid', displayValue: 'Solid'}, {
                        value: 'dotted',
                        displayValue: 'Dotted'
                    }, {value: 'dashed', displayValue: 'Dashed'}, {
                        value: 'double',
                        displayValue: 'Double'
                    }, {value: 'none', displayValue: 'None'}],
                    changeProperty: {
                        caption: 'Border Style',
                        selector: '.inner-rec',
                        property: 'border-style',
                        transitionable: false
                    }
                }
                    ,prx.commonproperties.borderColor
                    ,prx.commonproperties.borderRadiusExpandable

                ]
            ]

        },
        {
            caption: 'Text',
            properties: [
                [
                    prx.commonproperties.textFont
                ],
                [
                    prx.commonproperties.textFontStyle,
                    prx.commonproperties.textSize
                ],
                [
                    prx.commonproperties.textProperties,
                    {
                        caption: { label: 'Color', class: 'text-properties-label text-color-label' },
                        name: 'textColor',
                        proptype: 'font-color',
                        type: 'solid-colorpicker',
                        relatedEditableProperties: 'text',
                        relatedCSSProperties: 'color',
                        pffSettings: prx.commonproperties.pffSettingsColor,
                        value: function(item,name) {
                            if(typeof(item.textColor) == 'undefined') { item.textColor = '#2E2E2E'; }
                            return item.textColor;
                        },
                        liveUpdate: 'color'
                        ,changeProperty: {
                            caption: ' Text color',
                            selector: '.changeProperty-textColor',
                            property: 'color',
                            transitionable: true
                        }
                    }
                ],
                [
                    prx.commonproperties.textAlign
                ]
            ]
        },
        {
            caption: 'Placeholder  (If field is empty)',
            properties: [
                [
                    {
                        caption: 'Placeholder Text'
                        ,name: 'placeholder'
                        ,type: 'input'
                        ,value: function(item,name) {
                            return item.placeholder;
                        }
                        ,changeProperty: {
                            caption: 'Placeholder text',
                            transitionable: false,
                            changeFunction: function(item, containerid) {
                                var _id = (!containerid) ? item.id : containerid+'-'+item.id;

                                if(prx.editor) {
                                    $('#' + _id + ' .placeholder-input').text(prx.componentsHelper.getProp(item.placeholder,'other'));
                                } else {
                                    $('#' + _id + ' textarea').attr('placeholder', prx.componentsHelper.getProp(item.placeholder,'other'));
                                }

                            }
                        }
                    }
                ],
                [
                    {
                        caption: 'Placeholder Color',
                        name: 'placeholderColor',
                        proptype: 'placeholder-color',
                        type: 'combo-colorpicker',
                        value: function(item,name) { if(typeof(item.placeholderColor)=='undefined') { return '999999'; } return item.placeholderColor; }
                        ,values: prx.comps.basicColors
                        ,liveUpdate:'color'
                        ,changeProperty: {
                            caption: 'Placeholder color',
                            changeFunction: function(item, containerid) {
                                var _id = (!containerid) ? item.id : containerid+'-'+item.id;

                                var cR = '';
                                cR += '#'+_id+' .faux-input.placeholder-input { color: '+prx.componentsHelper.getProp(item.placeholderColor,'color-text')+'; }';
                                cR += '#'+_id+' textarea:-moz-placeholder { color: '+prx.componentsHelper.getProp(item.placeholderColor,'color-text')+'!important; }';
                                cR += '#'+_id+' textarea::-moz-placeholder { color: '+prx.componentsHelper.getProp(item.placeholderColor,'color-text')+'!important; }';
                                cR += '#'+_id+' textarea::-webkit-input-placeholder { color: '+prx.componentsHelper.getProp(item.placeholderColor,'color-text')+'!important; }';

                                $('#' + _id).append('<style>' + cR + '</style>')
                            }
                        }
                    }
                ]
            ]
        }
    ]
};


//////////////////////////////////////////////////////
//////////////////////////////////////////////////////
//////////////////////////////////////////////////////
//////////////////////////////////////////////////////

//TYPE: BASIC BUTTON TEXT ICON
prx.components.basic_button_texticon = {
    name: 'basic_button_texticon'
    ,type: 'basic_button_texticon'
    ,lib: _library
    ,caption: 'Button - Text & Icon'
    ,icon: '-500px -1000px'
    ,helper: prx.url.devices+  '/generic2/basic_button_texticon/helper.png'
    ,width: 200*prx.componentsHelper.getScale(_library)
    ,height: 40*prx.componentsHelper.getScale(_library)
    ,resizable : true
    ,backgroundColor: '0076ff'
    ,borderWidth: 0
    ,borderColor: '#025abf'
    ,borderRadius: 4*prx.componentsHelper.getScale(_library)
    ,text: 'Button'
    ,textFont: 'Arial,sans-serif'
    ,textFontStyle:  '400'
    ,textSize: 12*prx.componentsHelper.getScale(_library)
    ,textColor: 'FFFFFF'
    ,textProperties: []
    ,textAlign: 'center'
    ,iconpos: 'right'
    ,iconSize: '2'
    ,img: {fileId:'04fc9053132a2832640e4ab5705f5ab5.svg',name:'arrow_forward.svg',assetType:'icon',bucketsource:'static',url:'f1424098557988/78855ffe7a4c9efea4291339339586d3.svg',targetSrc:'generated/04fc9053132a2832640e4ab5705f5ab5_ffffff.svg',color:'ffffff'}
};

//TYPE: BASIC BUTTON TEXT
prx.components.basic_button_text = {
    name: 'basic_button_text'
    ,type: 'basic_button_text'
    ,lib: _library
    ,caption: 'Button with Background'
    ,icon: '-400px -1000px'
    ,helper: prx.url.devices+  '/generic2/basic_button_text/helper.png'
    ,width: 200*prx.componentsHelper.getScale(_library)
    ,height: 40*prx.componentsHelper.getScale(_library)
    ,resizable : true
    ,backgroundColor: '0076ff'
    ,borderWidth: 0
    ,borderColor: '#025abf'
    ,borderRadius: 4*prx.componentsHelper.getScale(_library)
    ,text: 'Button'
    ,textFont: 'Arial,sans-serif'
    ,textFontStyle:  '400'
    ,textSize: 12*prx.componentsHelper.getScale(_library)
    ,textColor: 'FFFFFF'
    ,textProperties: []
    ,textAlign: 'center'
    ,iconpos: 'none'
};

//TYPE: BASIC BUTTON ICON
prx.components.basic_button_icon = {
    name: 'basic_button_icon'
    ,type: 'basic_button_icon'
    ,lib: _library
    ,caption: 'Button - Icon only'
    ,icon: '-600px -1000px'
    ,helper: prx.url.devices+  '/generic2/basic_button_icon/helper.png'
    ,width: 200*prx.componentsHelper.getScale(_library)
    ,height: 40*prx.componentsHelper.getScale(_library)
    ,resizable : true
    ,backgroundColor: '0076ff'
    ,borderWidth: 0
    ,borderColor: '#025abf'
    ,borderRadius: 4*prx.componentsHelper.getScale(_library)
    ,text: 'Button'
    ,textFont: 'Arial,sans-serif'
    ,textFontStyle:  '400'
    ,textSize: 12*prx.componentsHelper.getScale(_library)
    ,textColor: 'FFFFFF'
    ,textProperties: []
    ,textAlign: 'center'
    ,iconpos: 'notext'
    ,iconSize: '2'
    ,img: {fileId:'04fc9053132a2832640e4ab5705f5ab5.svg',name:'arrow_forward.svg',assetType:'icon',bucketsource:'static',url:'f1424098557988/78855ffe7a4c9efea4291339339586d3.svg',targetSrc:'generated/04fc9053132a2832640e4ab5705f5ab5_ffffff.svg',color:'ffffff'}
};

//TYPE: TEXT FIELD
prx.components.basic_textfield = {
    name: 'basic_textfield'
    ,type: 'basic_textfield'
    ,lib: _library
    ,caption: 'Text Field'
    ,icon: '-1500px -1000px'
    ,helper: prx.url.devices+  '/generic2/basic_textfield/helper.png'
    ,width: 320*prx.componentsHelper.getScale(_library)
    ,height: 40*prx.componentsHelper.getScale(_library)
    ,value: ''
    ,placeholder: 'Placeholder'
    ,inputtype: 'text'
    ,backgroundColor: 'rgba(0,0,0,0)'
    ,textFont: 'Arial,sans-serif'
    ,textFontStyle:  '400'
    ,textSize: 14*prx.componentsHelper.getScale(_library)
    ,textColor:  '6c7075'
    ,textProperties: []
    ,placeholderColor: 'c1c8c9'
    ,borderWidth: 1*prx.componentsHelper.getScale(_library)
    ,borderStyle: 'solid'
    ,borderRadius: 4*prx.componentsHelper.getScale(_library)
    ,borderColor: 'd4d7d9'
    ,textAlign: 'left'
};

//TYPE: PASSWORD FIELD
prx.components.basic_passwordfield = {
    name: 'basic_passwordfield'
    ,type: 'basic_textfield'
    ,lib: _library
    ,caption: 'Password Field'
    ,icon: '-1600px -1000px'
    ,helper: prx.url.devices+  '/generic2/basic_passwordfield/helper.png'
    ,width: 320*prx.componentsHelper.getScale(_library)
    ,height: 40*prx.componentsHelper.getScale(_library)
    ,value: ''
    ,placeholder: 'Placeholder'
    ,inputtype: 'password'
    ,backgroundColor: 'rgba(0,0,0,0)'
    ,textFont: 'Arial,sans-serif'
    ,textFontStyle:  '400'
    ,textSize: 14*prx.componentsHelper.getScale(_library)
    ,textColor: '6c7075'
    ,textProperties: []
    ,placeholderColor: 'c1c8c9'
    ,borderWidth: 1*prx.componentsHelper.getScale(_library)
    ,borderStyle: 'solid'
    ,borderRadius: 4*prx.componentsHelper.getScale(_library)
    ,borderColor: 'd4d7d9'
    ,textAlign: 'left'
};

//TYPE: TEXTAREA
prx.components.basic_textarea = {
    name: 'basic_textarea'
    ,type: 'basic_textarea'
    ,lib: _library
    ,caption: 'Textarea'
    ,icon: '-1700px -1000px'
    ,helper: prx.url.devices+  '/generic2/basic_textarea/helper.png'
    ,width: 320*prx.componentsHelper.getScale(_library)
    ,height: 96*prx.componentsHelper.getScale(_library)
    ,value: ''
    ,placeholder: 'Placeholder'
    ,backgroundColor: 'rgba(0,0,0,0)'
    ,textFont: 'Arial,sans-serif'
    ,textFontStyle:  '400'
    ,textSize: 14*prx.componentsHelper.getScale(_library)
    ,textColor: '6c7075'
    ,textProperties: []
    ,placeholderColor: 'c1c8c9'
    ,borderWidth: 1*prx.componentsHelper.getScale(_library)
    ,borderRadius: 4*prx.componentsHelper.getScale(_library)
    ,borderColor: 'd4d7d9'
    ,textAlign: 'left'
};

//TYPE: CHECKBOX
prx.components.basic_checkbox = {
    name: 'basic_checkbox'
    ,type: 'basic_checkbox'
    ,lib: _library
    ,caption: 'Checkbox'
    ,icon: '-1800px -1000px'
    ,helper: prx.url.devices+  '/generic2/basic_checkbox/helper.png'
    ,width: 100*prx.componentsHelper.getScale(_library)
    ,height: 16*prx.componentsHelper.getScale(_library)
    ,text: 'Select'
    ,textFont: 'Arial,sans-serif'
    ,textFontStyle:  '400'
    ,textSize: 14*prx.componentsHelper.getScale(_library)
    ,textColor: '6c7075'
    ,textProperties: []
    ,resizable : true
    ,backgroundColor: 'rgba(0,0,0,0)'
    ,activeBackgroundColor: '0076ff'
    ,borderWidth: 1*prx.componentsHelper.getScale(_library)
    ,borderColor: 'd4d7d9'
    ,borderRadius: 4*prx.componentsHelper.getScale(_library)
    ,img: {fileId:'498e767b8d1969245862682798acb78f.svg',name:'check.svg',assetType:'icon',bucketsource:'static',url:'f1352971179296/34d6599607c21d5f87e5d30af9449c1c.svg',targetSrc:'generated/34d6599607c21d5f87e5d30af9449c1c_ffffff.svg',color:'ffffff'}
    ,active: true
    ,radioButtonsGroupName: ''
    ,checkboxSize: 22*prx.componentsHelper.getScale(_library)
};

//TYPE: ON/OFF SWITCH
prx.components.basic_switch = {
    name: 'basic_switch'
    ,type: 'basic_switch'
    ,lib: _library
    ,caption: 'On/Off Switch'
    ,icon: '-1900px -1000px'
    ,helper: prx.url.devices+  '/generic2/basic_switch/helper.png'
    ,width: 48*prx.componentsHelper.getScale(_library)
    ,height: 28*prx.componentsHelper.getScale(_library)
    ,inactiveBarColor: 'e3e6e8'
    ,activeBarColor: '0076ff'
    ,borderWidth: 1*prx.componentsHelper.getScale(_library)
    ,borderColor: 'd4d7d9'
    ,borderRadius: 50 * prx.componentsHelper.getScale(_library)
    ,inactiveSwitchColor: 'FFFFFF'
    ,activeSwitchColor: 'FFFFFF'
    ,switchSize: 22 * prx.componentsHelper.getScale(_library)
    ,radioButtonsGroupName: ''
};

//TYPE: ON/OFF SWITCH 2
prx.components.basic_switch2 = {
    name: 'basic_switch2'
    ,type: 'basic_switch2'
    ,lib: _library
    ,caption: 'On/Off Switch 2'
    ,icon: '0px -1100px'
    ,helper: prx.url.devices+  '/generic2/basic_switch2/helper.png'
    ,width: 48*prx.componentsHelper.getScale(_library)
    ,height: 30*prx.componentsHelper.getScale(_library)
    ,inactiveBarColor: 'e3e6e8'
    ,activeBarColor: '0076ff'
    ,borderWidth: 1*prx.componentsHelper.getScale(_library)
    ,borderColor: 'd4d7d9'
    ,borderRadius: 50 * prx.componentsHelper.getScale(_library)
    ,inactiveSwitchColor: 'FFFFFF'
    ,activeSwitchColor: 'FFFFFF'
    ,barSize: 18 * prx.componentsHelper.getScale(_library)
    ,radioButtonsGroupName: ''
};

//TYPE: PROGRESS BAR
prx.components.basic_progressbar = {
    name: 'basic_progressbar'
    ,type: 'basic_progressbar'
    ,lib: _library
    ,caption: 'Progress bar'
    ,icon: '-600px -1100px'
    ,helper: prx.url.devices+  '/generic2/basic_progressbar/helper.png'
    ,width: 328*prx.componentsHelper.getScale(_library)
    ,height: 8*prx.componentsHelper.getScale(_library)
    ,resizable : true
    ,barColor: 'e3e6e8'
    ,fillBarColor: '0076ff'
    ,borderRadius: 100*prx.componentsHelper.getScale(_library)
    ,progress: 70
};

//TYPE: CIRCULAR PROGRESS BAR
prx.components.basic_circularprogressbar = {
    name: 'basic_circularprogressbar'
    ,type: 'basic_circularprogressbar'
    ,lib: _library
    ,caption: 'Circular progress bar'
    ,icon: '-700px -1100px'
    ,helper: prx.url.devices+  '/generic2/basic_circularprogressbar/helper.png'
    ,width: 88*prx.componentsHelper.getScale(_library)
    ,height:88*prx.componentsHelper.getScale(_library)
    ,resizable : true
    ,barColor: 'e3e6e8'
    ,fillBarColor: '0076ff'
    ,progress: 70
    ,strokeWidth: 8*prx.componentsHelper.getScale(_library)
    ,roundedEnds: true
    ,aspectratio: 1
};

//TYPE: LOADER
prx.components.basic_loader = {
    name: 'basic_loader'
    ,type: 'basic_loader'
    ,lib: _library
    ,caption: 'Loader'
    ,icon: '-700px -1100px'
    ,helper: prx.url.devices+  '/generic2/basic_loader/helper.png'
    ,width: 88*prx.componentsHelper.getScale(_library)
    ,height:88*prx.componentsHelper.getScale(_library)
    ,resizable : true
    ,barColor: 'e3e6e8'
    ,fillBarColor: '0076ff'
    ,progress: 60
    ,strokeWidth: 8*prx.componentsHelper.getScale(_library)
    ,roundedEnds: true
    ,aspectratio: 1
    ,loader: true
};

//TYPE: BUTTON GROUP
prx.components.basic_buttongroup = {
    name: 'basic_buttongroup'
    ,type: 'basic_buttongroup'
    ,lib: _library
    ,caption: 'Button Group'
    ,changeActive: true
    ,icon: '-1000px -100px'
    ,iconpos: 'none'
    ,iconSize: '3'
    ,helper: prx.url.devices+  '/generic2/basic_buttongroup_horizontal_texticon_right/helper.png'
    ,width: 320*prx.componentsHelper.getScale(_library)
    ,height: 40*prx.componentsHelper.getScale(_library)
    ,resizable : true
    ,backgroundColor: 'rgba(0,0,0,0)'
    ,activeBackgroundColor: '6c7075'
    ,borderWidth: 1*prx.componentsHelper.getScale(_library)
    ,borderColor: 'd4d7d9'
    ,borderRadius: 4*prx.componentsHelper.getScale(_library)
    ,maskEnabled: true
    ,maskInactive: '6c7075'
    ,maskActive: 'FFFFFF'
    ,textFont: 'Arial, sans-serif'
    ,textFontStyle:  '400'
    ,textSize: 12*prx.componentsHelper.getScale(_library)
    ,textColor: '6c7075'
    ,textProperties: []
    ,activeTextColor: 'FFFFFF'
    ,selected: ['0']
    ,buttons: [{actions:[],icon:{fileId:'324879579bf3ecf9c4b49ef0c151501b.svg',name:'speech-1.svg',assetType:'icon',bucketsource:'static',url:'f1352971179296/693bf32e7424f6bf747696ddab19cdf5.svg',targetSrc:'generated/324879579bf3ecf9c4b49ef0c151501b_6c7075.svg',color:'6c7075'},activeicon:{fileId:'324879579bf3ecf9c4b49ef0c151501b.svg',name:'speech-1.svg',assetType:'icon',bucketsource:'static',url:'f1352971179296/693bf32e7424f6bf747696ddab19cdf5.svg',targetSrc:'generated/324879579bf3ecf9c4b49ef0c151501b_ffffff.svg',color:'ffffff'},text:'First'},{actions:[],icon:{fileId:'f46effc9d8189665443ebcc0f7ca0934.svg',name:'heart.svg',assetType:'icon',bucketsource:'static',url:'f1515489068744/f46effc9d8189665443ebcc0f7ca0934.svg',targetSrc:'generated/f46effc9d8189665443ebcc0f7ca0934_6c7075.svg',color:'6c7075'},activeicon:{fileId:'f46effc9d8189665443ebcc0f7ca0934.svg',name:'heart.svg',assetType:'icon',bucketsource:'static',url:'f1515489068744/f46effc9d8189665443ebcc0f7ca0934.svg',targetSrc:'generated/f46effc9d8189665443ebcc0f7ca0934_ffffff.svg',color:'ffffff'},text:'Second'}]
    ,dynamicSizeExpand: 'h'
    ,dynamicAlignment: 'h'
};

//TYPE: BUTTON GROUP - TEXT HORIZONTAL TEXT ONLY
prx.components.basic_buttongroup_horizontal_text = {
    name: 'basic_buttongroup_horizontal_text'
    ,type: 'basic_buttongroup_horizontal_text'
    ,lib: _library
    ,caption: 'Button Group - Text only'
    ,changeActive: true
    ,icon: '-700px -1000px'
    ,iconpos: 'none'
    ,iconSize: '3'
    ,helper: prx.url.devices+  '/generic2/basic_buttongroup_horizontal_text/helper.png'
    ,width: 320*prx.componentsHelper.getScale(_library)
    ,height: 40*prx.componentsHelper.getScale(_library)
    ,resizable : true
    ,backgroundColor: 'rgba(0,0,0,0)'
    ,activeBackgroundColor: '6c7075'
    ,borderWidth: 1*prx.componentsHelper.getScale(_library)
    ,borderColor: 'd4d7d9'
    ,borderRadius: 4*prx.componentsHelper.getScale(_library)
    ,textFont: 'Arial, sans-serif'
    ,textFontStyle:  '400'
    ,textSize: 12*prx.componentsHelper.getScale(_library)
    ,textColor: '6c7075'
    ,textProperties: []
    ,activeTextColor: 'FFFFFF'
    ,selected: ['0']
    ,buttons: [{
        actions: [],
        text: 'First'
    }, {
        actions: [],
        text: 'Second'
    }]
    ,dynamicSizeExpand: 'h'
    ,dynamicAlignment: 'h'
    ,isVertical: false
};

//TYPE: BUTTON GROUP - TEXT AND ICONS HORIZONTAL RIGHT
prx.components.basic_buttongroup_horizontal_texticon_right = {
    name: 'basic_buttongroup_horizontal_texticon_right'
    ,type: 'basic_buttongroup'
    ,lib: _library
    ,caption: 'Button Group - Text & Icon'
    ,changeActive: true
    ,icon: '-800px -1000px'
    ,iconpos: 'right'
    ,iconSize: '2'
    ,helper: prx.url.devices+  '/generic2/basic_buttongroup_horizontal_texticon_right/helper.png'
    ,width: 320*prx.componentsHelper.getScale(_library)
    ,height: 40*prx.componentsHelper.getScale(_library)
    ,resizable : true
    ,backgroundColor: 'rgba(0,0,0,0)'
    ,activeBackgroundColor: '6c7075'
    ,borderWidth: 1*prx.componentsHelper.getScale(_library)
    ,borderColor: 'd4d7d9'
    ,borderRadius: 4*prx.componentsHelper.getScale(_library)
    ,maskEnabled: true
    ,maskInactive: '6c7075'
    ,maskActive: 'FFFFFF'
    ,textFont: 'Arial, sans-serif'
    ,textFontStyle:  '400'
    ,textSize: 12*prx.componentsHelper.getScale(_library)
    ,textColor: '6c7075'
    ,textProperties: []
    ,activeTextColor: 'FFFFFF'
    ,selected: ['0']
    ,buttons: [{actions:[],icon:{fileId:'324879579bf3ecf9c4b49ef0c151501b.svg',name:'speech-1.svg',assetType:'icon',bucketsource:'static',url:'f1352971179296/693bf32e7424f6bf747696ddab19cdf5.svg',targetSrc:'generated/324879579bf3ecf9c4b49ef0c151501b_6c7075.svg',color:'6c7075'},activeicon:{fileId:'324879579bf3ecf9c4b49ef0c151501b.svg',name:'speech-1.svg',assetType:'icon',bucketsource:'static',url:'f1352971179296/693bf32e7424f6bf747696ddab19cdf5.svg',targetSrc:'generated/324879579bf3ecf9c4b49ef0c151501b_ffffff.svg',color:'ffffff'},text:'First'},{actions:[],icon:{fileId:'f46effc9d8189665443ebcc0f7ca0934.svg',name:'heart.svg',assetType:'icon',bucketsource:'static',url:'f1515489068744/f46effc9d8189665443ebcc0f7ca0934.svg',targetSrc:'generated/f46effc9d8189665443ebcc0f7ca0934_6c7075.svg',color:'6c7075'},activeicon:{fileId:'f46effc9d8189665443ebcc0f7ca0934.svg',name:'heart.svg',assetType:'icon',bucketsource:'static',url:'f1515489068744/f46effc9d8189665443ebcc0f7ca0934.svg',targetSrc:'generated/f46effc9d8189665443ebcc0f7ca0934_ffffff.svg',color:'ffffff'},text:'Second'}]
    ,dynamicSizeExpand: 'h'
    ,dynamicAlignment: 'h'
    ,isVertical: false
};

//TYPE: BUTTON GROUP - TEXT AND ICONS ON TOP HORIZONTAL
prx.components.basic_buttongroup_horizontal_texticon_top = {
    name: 'basic_buttongroup_horizontal_texticon_top'
    ,type: 'basic_buttongroup'
    ,lib: _library
    ,caption: 'Button Group - Text & Icon'
    ,changeActive: true
    ,icon: '-900px -1000px'
    ,iconpos: 'top'
    ,iconSize: '3'
    ,helper: prx.url.devices+  '/generic2/basic_buttongroup_horizontal_texticon_top/helper.png'
    ,width: 320*prx.componentsHelper.getScale(_library)
    ,height: 64*prx.componentsHelper.getScale(_library)
    ,resizable : true
    ,backgroundColor: 'rgba(0,0,0,0)'
    ,activeBackgroundColor: '6c7075'
    ,borderWidth: 1*prx.componentsHelper.getScale(_library)
    ,borderColor: 'd4d7d9'
    ,borderRadius: 4*prx.componentsHelper.getScale(_library)
    ,maskEnabled: true
    ,maskInactive: '6c7075'
    ,maskActive: 'FFFFFF'
    ,textFont: 'Arial, sans-serif'
    ,textFontStyle:  '400'
    ,textSize: 12*prx.componentsHelper.getScale(_library)
    ,textColor: '6c7075'
    ,textProperties: []
    ,activeTextColor: 'FFFFFF'
    ,selected: ['0']
    ,buttons: [{actions:[],icon:{fileId:'324879579bf3ecf9c4b49ef0c151501b.svg',name:'speech-1.svg',assetType:'icon',bucketsource:'static',url:'f1352971179296/693bf32e7424f6bf747696ddab19cdf5.svg',targetSrc:'generated/324879579bf3ecf9c4b49ef0c151501b_6c7075.svg',color:'6c7075'},activeicon:{fileId:'324879579bf3ecf9c4b49ef0c151501b.svg',name:'speech-1.svg',assetType:'icon',bucketsource:'static',url:'f1352971179296/693bf32e7424f6bf747696ddab19cdf5.svg',targetSrc:'generated/324879579bf3ecf9c4b49ef0c151501b_ffffff.svg',color:'ffffff'},text:'First'},{actions:[],icon:{fileId:'f46effc9d8189665443ebcc0f7ca0934.svg',name:'heart.svg',assetType:'icon',bucketsource:'static',url:'f1515489068744/f46effc9d8189665443ebcc0f7ca0934.svg',targetSrc:'generated/f46effc9d8189665443ebcc0f7ca0934_6c7075.svg',color:'6c7075'},activeicon:{fileId:'f46effc9d8189665443ebcc0f7ca0934.svg',name:'heart.svg',assetType:'icon',bucketsource:'static',url:'f1515489068744/f46effc9d8189665443ebcc0f7ca0934.svg',targetSrc:'generated/f46effc9d8189665443ebcc0f7ca0934_ffffff.svg',color:'ffffff'},text:'Second'}]
    ,dynamicSizeExpand: 'h'
    ,dynamicAlignment: 'h'
    ,isVertical: false
};

//TYPE: BUTTON GROUP - ICONS ONLY HORIZONTAL
prx.components.basic_buttongroup_horizontal_icon = {
    name: 'basic_buttongroup_horizontal_icon'
    ,type: 'basic_buttongroup_horizontal_icon'
    ,lib: _library
    ,caption: 'Button Group - Icon only'
    ,changeActive: true
    ,icon: '-1000px -1000px'
    ,iconpos: 'notext'
    ,iconSize: '2'
    ,helper: prx.url.devices+  '/generic2/basic_buttongroup_horizontal_icon/helper.png'
    ,width: 320*prx.componentsHelper.getScale(_library)
    ,height: 40*prx.componentsHelper.getScale(_library)
    ,resizable : true
    ,backgroundColor: 'rgba(0,0,0,0)'
    ,activeBackgroundColor: '6c7075'
    ,borderWidth: 1*prx.componentsHelper.getScale(_library)
    ,borderColor: 'd4d7d9'
    ,borderRadius: 4*prx.componentsHelper.getScale(_library)
    ,maskEnabled: true
    ,maskInactive: '6c7075'
    ,maskActive: 'FFFFFF'
    ,textFont: 'Arial, sans-serif'
    ,textFontStyle:  '400'
    ,textSize: 12*prx.componentsHelper.getScale(_library)
    ,textColor: '6c7075'
    ,textProperties: []
    ,activeTextColor: 'FFFFFF'
    ,selected: ['0']
    ,buttons: [{actions:[],icon:{fileId:'324879579bf3ecf9c4b49ef0c151501b.svg',name:'speech-1.svg',assetType:'icon',bucketsource:'static',url:'f1352971179296/693bf32e7424f6bf747696ddab19cdf5.svg',targetSrc:'generated/324879579bf3ecf9c4b49ef0c151501b_6c7075.svg',color:'6c7075'},activeicon:{fileId:'324879579bf3ecf9c4b49ef0c151501b.svg',name:'speech-1.svg',assetType:'icon',bucketsource:'static',url:'f1352971179296/693bf32e7424f6bf747696ddab19cdf5.svg',targetSrc:'generated/324879579bf3ecf9c4b49ef0c151501b_ffffff.svg',color:'ffffff'},text:'First'},{actions:[],icon:{fileId:'f46effc9d8189665443ebcc0f7ca0934.svg',name:'heart.svg',assetType:'icon',bucketsource:'static',url:'f1515489068744/f46effc9d8189665443ebcc0f7ca0934.svg',targetSrc:'generated/f46effc9d8189665443ebcc0f7ca0934_6c7075.svg',color:'6c7075'},activeicon:{fileId:'f46effc9d8189665443ebcc0f7ca0934.svg',name:'heart.svg',assetType:'icon',bucketsource:'static',url:'f1515489068744/f46effc9d8189665443ebcc0f7ca0934.svg',targetSrc:'generated/f46effc9d8189665443ebcc0f7ca0934_ffffff.svg',color:'ffffff'},text:'Second'}]
    ,dynamicSizeExpand: 'h'
    ,dynamicAlignment: 'h'
    ,isVertical: false
};

//TYPE: BUTTON GROUP - TEXT VERTICAL
prx.components.basic_buttongroup_vertical_text = {
    name: 'basic_buttongroup_vertical_text'
    ,type: 'basic_buttongroup_vertical_text'
    ,lib: _library
    ,caption: 'Button Group - Text only'
    ,changeActive: true
    ,icon: '-1100px -1000px'
    ,iconpos: 'none'
    ,iconSize: '3'
    ,helper: prx.url.devices+  '/generic2/basic_buttongroup_vertical_text/helper.png'
    ,width: 160*prx.componentsHelper.getScale(_library)
    ,height: 80*prx.componentsHelper.getScale(_library)
    ,resizable : true
    ,backgroundColor: 'rgba(0,0,0,0)'
    ,activeBackgroundColor: '6c7075'
    ,borderWidth: 1*prx.componentsHelper.getScale(_library)
    ,borderColor: 'd4d7d9'
    ,borderRadius: 4*prx.componentsHelper.getScale(_library)
    ,textFont: 'Arial, sans-serif'
    ,textFontStyle:  '400'
    ,textSize: 12*prx.componentsHelper.getScale(_library)
    ,textColor: '6c7075'
    ,textAlign: 'left'
    ,textProperties: []
    ,activeTextColor: 'FFFFFF'
    ,selected: ['0']
    ,buttons: [{
        actions: [],
        text: 'First'
    }, {
        actions: [],
        text: 'Second'
    }]
    ,dynamicSizeExpand: 'v'
    ,dynamicAlignment: 'v'
    ,isVertical: true
};

//TYPE: BUTTON GROUP - TEXT AND ICONS VERTICAL
prx.components.basic_buttongroup_vertical_texticon = {
    name: 'basic_buttongroup_vertical_texticon'
    ,type: 'basic_buttongroup_vertical_texticon'
    ,lib: _library
    ,caption: 'Button Group - Text & Icon'
    ,changeActive: true
    ,icon: '-1200px -1000px'
    ,iconpos: 'left'
    ,iconSize: '2'
    ,helper: prx.url.devices+  '/generic2/basic_buttongroup_vertical_texticon/helper.png'
    ,width: 160*prx.componentsHelper.getScale(_library)
    ,height: 80*prx.componentsHelper.getScale(_library)
    ,resizable : true
    ,backgroundColor: 'rgba(0,0,0,0)'
    ,activeBackgroundColor: '6c7075'
    ,borderWidth: 1*prx.componentsHelper.getScale(_library)
    ,borderColor: 'd4d7d9'
    ,borderRadius: 4*prx.componentsHelper.getScale(_library)
    ,maskEnabled: true
    ,maskInactive: '6c7075'
    ,maskActive: 'FFFFFF'
    ,textFont: 'Arial, sans-serif'
    ,textFontStyle:  '400'
    ,textSize: 12*prx.componentsHelper.getScale(_library)
    ,textColor: '6c7075'
    ,textAlign: 'left'
    ,textProperties: []
    ,activeTextColor: 'FFFFFF'
    ,selected: ['0']
    ,buttons: [{actions:[],icon:{fileId:'324879579bf3ecf9c4b49ef0c151501b.svg',name:'speech-1.svg',assetType:'icon',bucketsource:'static',url:'f1352971179296/693bf32e7424f6bf747696ddab19cdf5.svg',targetSrc:'generated/324879579bf3ecf9c4b49ef0c151501b_6c7075.svg',color:'6c7075'},activeicon:{fileId:'324879579bf3ecf9c4b49ef0c151501b.svg',name:'speech-1.svg',assetType:'icon',bucketsource:'static',url:'f1352971179296/693bf32e7424f6bf747696ddab19cdf5.svg',targetSrc:'generated/324879579bf3ecf9c4b49ef0c151501b_ffffff.svg',color:'ffffff'},text:'First'},{actions:[],icon:{fileId:'f46effc9d8189665443ebcc0f7ca0934.svg',name:'heart.svg',assetType:'icon',bucketsource:'static',url:'f1515489068744/f46effc9d8189665443ebcc0f7ca0934.svg',targetSrc:'generated/f46effc9d8189665443ebcc0f7ca0934_6c7075.svg',color:'6c7075'},activeicon:{fileId:'f46effc9d8189665443ebcc0f7ca0934.svg',name:'heart.svg',assetType:'icon',bucketsource:'static',url:'f1515489068744/f46effc9d8189665443ebcc0f7ca0934.svg',targetSrc:'generated/f46effc9d8189665443ebcc0f7ca0934_ffffff.svg',color:'ffffff'},text:'Second'}]
    ,dynamicSizeExpand: 'v'
    ,dynamicAlignment: 'v'
    ,isVertical: true
};

//TYPE: BUTTON GROUP - ICONS ONLY VERTICAL
prx.components.basic_buttongroup_vertical_icon = {
    name: 'basic_buttongroup_vertical_icon'
    ,type: 'basic_buttongroup_vertical_icon'
    ,lib: _library
    ,caption: 'Button Group - Icon only'
    ,changeActive: true
    ,icon: '-1300px -1100px'
    ,iconpos: 'notext'
    ,iconSize: '2'
    ,helper: prx.url.devices+  '/generic2/basic_buttongroup_vertical_icon/helper.png'
    ,width: 160*prx.componentsHelper.getScale(_library)
    ,height: 80*prx.componentsHelper.getScale(_library)
    ,resizable : true
    ,backgroundColor: 'rgba(0,0,0,0)'
    ,activeBackgroundColor: '6c7075'
    ,borderWidth: 1*prx.componentsHelper.getScale(_library)
    ,borderColor: 'd4d7d9'
    ,borderRadius: 4*prx.componentsHelper.getScale(_library)
    ,maskEnabled: true
    ,maskInactive: '6c7075'
    ,maskActive: 'FFFFFF'
    ,textFont: 'Arial, sans-serif'
    ,textFontStyle:  '400'
    ,textSize: 12*prx.componentsHelper.getScale(_library)
    ,textColor: '6c7075'
    ,textProperties: []
    ,activeTextColor: 'FFFFFF'
    ,selected: ['0']
    ,buttons: [{actions:[],icon:{fileId:'324879579bf3ecf9c4b49ef0c151501b.svg',name:'speech-1.svg',assetType:'icon',bucketsource:'static',url:'f1352971179296/693bf32e7424f6bf747696ddab19cdf5.svg',targetSrc:'generated/324879579bf3ecf9c4b49ef0c151501b_6c7075.svg',color:'6c7075'},activeicon:{fileId:'324879579bf3ecf9c4b49ef0c151501b.svg',name:'speech-1.svg',assetType:'icon',bucketsource:'static',url:'f1352971179296/693bf32e7424f6bf747696ddab19cdf5.svg',targetSrc:'generated/324879579bf3ecf9c4b49ef0c151501b_ffffff.svg',color:'ffffff'},text:'First'},{actions:[],icon:{fileId:'f46effc9d8189665443ebcc0f7ca0934.svg',name:'heart.svg',assetType:'icon',bucketsource:'static',url:'f1515489068744/f46effc9d8189665443ebcc0f7ca0934.svg',targetSrc:'generated/f46effc9d8189665443ebcc0f7ca0934_6c7075.svg',color:'6c7075'},activeicon:{fileId:'f46effc9d8189665443ebcc0f7ca0934.svg',name:'heart.svg',assetType:'icon',bucketsource:'static',url:'f1515489068744/f46effc9d8189665443ebcc0f7ca0934.svg',targetSrc:'generated/f46effc9d8189665443ebcc0f7ca0934_ffffff.svg',color:'ffffff'},text:'Second'}]
    ,dynamicSizeExpand: 'v'
    ,dynamicAlignment: 'v'
    ,isVertical: true
};

//TYPE: CHECKBOX LIST - TEXT AND ICONS HORIZONTAL
prx.components.basic_checkbox_list_horizontal = {
    name: 'basic_checkbox_list_horizontal'
    ,type: 'basic_checkbox_list_horizontal'
    ,lib: _library
    ,caption: 'Checkbox list - Horizontal'
    ,changeActive: true
    ,icon: '-1400px -1000px'
    ,iconpos: 'left'
    ,iconSize: '3'
    ,inactiveicon: {fileId:'dd615595951ee619d2e0b0135cadeeba.svg',name:'check_box_outline_blank.svg',assetType:'icon',bucketsource:'static',url:'f1447836421086/dd615595951ee619d2e0b0135cadeeba.svg',targetSrc:'generated/dd615595951ee619d2e0b0135cadeeba_d4d7d9.svg',color:'d4d7d9'}
    ,activeicon: {fileId:'6c2575641c6ad1c8f5278ea330084ff0.svg',name:'check_box.svg',assetType:'icon',bucketsource:'static',url:'f1447836421086/6c2575641c6ad1c8f5278ea330084ff0.svg',targetSrc:'generated/6c2575641c6ad1c8f5278ea330084ff0_0076ff.svg',color:'0076ff'}    ,helper: prx.url.devices+  '/generic2/basic_checkbox_list_horizontal/helper.png'
    ,width: 456*prx.componentsHelper.getScale(_library)
    ,height: 40*prx.componentsHelper.getScale(_library)
    ,resizable : true
    ,backgroundColor: 'rgba(0,0,0,0)'
    ,activeBackgroundColor: 'rgba(0,0,0,0)'
    ,borderWidth: 1*prx.componentsHelper.getScale(_library)
    ,borderColor: 'd4d7d9'
    ,borderRadius: 0
    ,textFont: 'Arial,sans-serif'
    ,textFontStyle:  '400'
    ,textSize: 12*prx.componentsHelper.getScale(_library)
    ,textColor: '6c7075'
    ,textAlign: 'left'
    ,textProperties: []
    ,activeTextColor: '6c7075'
    ,selected: ['1','2']
    ,listitems: [{
        actions1: []
        ,actions2: []
        ,actions: []
        ,text: 'First'
    },{
        actions1: []
        ,actions2: []
        ,actions: []
        ,text: 'Second'
    },{
        actions1: []
        ,actions2: []
        ,actions: []
        ,text: 'Third'
    }]
    ,dynamicSizeExpand: 'h'
    ,dynamicAlignment: 'h'
    ,isVertical: false
    ,closedText: ''
};

//TYPE: CHECKBOX LIST - TEXT AND ICONS VERTICAL
prx.components.basic_checkbox_list_vertical = {
    name: 'basic_checkbox_list_vertical'
    ,type: 'basic_checkbox_list_vertical'
    ,lib: _library
    ,caption: 'Checkbox list - Vertical'
    ,changeActive: true
    ,icon: '-1300px -1000px'
    ,iconpos: 'left'
    ,iconSize: '3'
    ,inactiveicon: {fileId:'dd615595951ee619d2e0b0135cadeeba.svg',name:'check_box_outline_blank.svg',assetType:'icon',bucketsource:'static',url:'f1447836421086/dd615595951ee619d2e0b0135cadeeba.svg',targetSrc:'generated/dd615595951ee619d2e0b0135cadeeba_d4d7d9.svg',color:'d4d7d9'}
    ,activeicon: {fileId:'6c2575641c6ad1c8f5278ea330084ff0.svg',name:'check_box.svg',assetType:'icon',bucketsource:'static',url:'f1447836421086/6c2575641c6ad1c8f5278ea330084ff0.svg',targetSrc:'generated/6c2575641c6ad1c8f5278ea330084ff0_0076ff.svg',color:'0076ff'}
    ,helper: prx.url.devices+  '/generic2/basic_checkbox_list_vertical/helper.png'
    ,width: 152*prx.componentsHelper.getScale(_library)
    ,height: 120*prx.componentsHelper.getScale(_library)
    ,resizable : true
    ,backgroundColor: 'rgba(0,0,0,0)'
    ,activeBackgroundColor: 'rgba(0,0,0,0)'
    ,borderWidth: 1*prx.componentsHelper.getScale(_library)
    ,borderColor: 'd4d7d9'
    ,borderRadius: 0*prx.componentsHelper.getScale(_library)
    ,textFont: 'Arial,sans-serif'
    ,textFontStyle:  '400'
    ,textSize: 12*prx.componentsHelper.getScale(_library)
    ,textColor: '6c7075'
    ,textAlign: 'left'
    ,textProperties: []
    ,activeTextColor: '6c7075'
    ,selected: ['1', '2']
    ,listitems: [{
        actions1: []
        ,actions2: []
        ,actions: []
        ,text: 'First'
    },{
        actions1: []
        ,actions2: []
        ,actions: []
        ,text: 'Second'
    },{
        actions1: []
        ,actions2: []
        ,actions: []
        ,text: 'Third'
    }]
    ,dynamicSizeExpand: 'v'
    ,dynamicAlignment: 'v'
    ,isVertical: true
};

//TYPE: SLIDER
prx.components.basic_slider = {
    name: 'basic_slider'
    ,type: 'basic_slider'
    ,lib: _library
    ,caption: 'Slider'
    ,icon: '-100px -1100px'
    ,helper: prx.url.devices+  '/generic2/basic_slider/helper.png'
    ,width: 320*prx.componentsHelper.getScale(_library)
    ,height: 32*prx.componentsHelper.getScale(_library)
    ,resizable : true
    ,barThickness: 4*prx.componentsHelper.getScale(_library)
    ,barColor: 'e3e6e8'
    ,fillBarColor: '0076ff'
    ,sliderColor: 'ffffff'
    ,barBorderRadius: 2*prx.componentsHelper.getScale(_library)
    ,sliderBorderWidth: 1*prx.componentsHelper.getScale(_library)
    ,sliderBorderRadius: '50%'
    ,borderColor: 'd4d7d9'
    ,sliderPosition: 70
    ,properties: 'v,l,o,hpos,vpos,w,h,dr,f,ds'
};

//TYPE: SEARCH BAR
prx.components.basic_searchbar = {
    name: 'basic_searchbar'
    ,type: 'basic_searchbar'
    ,lib: _library
    ,caption: 'Search Field'
    ,icon: '-200px -1100px'
    ,helper: prx.url.devices+  '/generic2/basic_searchbar/helper.png'
    ,width: 320*prx.componentsHelper.getScale(_library)
    ,height: 40*prx.componentsHelper.getScale(_library)
    ,value: ''
    ,placeholder: 'Search'
    ,backgroundColor: 'ffffff'
    ,textFont: 'Arial,sans-serif'
    ,textFontStyle:  '400'
    ,textSize: 14*prx.componentsHelper.getScale(_library)
    ,textColor:  '6c7075'
    ,textAlign: 'left'
    ,textProperties: []
    ,placeholderColor: 'c1c8c9'
    ,borderWidth: 1*prx.componentsHelper.getScale(_library)
    ,borderRadius: 4*prx.componentsHelper.getScale(_library)
    ,borderColor: 'd4d7d9'
    ,borderStyle: 'solid'
    ,imgSrc: {fileId:'92e25d8093bef9eb0ec44968eff32c0f.svg',name:'search.svg',assetType:'icon',bucketsource:'static',url:'f1352971179296/14870b88002e540a4d1fc9c2d2a88384.svg',targetSrc:'generated/92e25d8093bef9eb0ec44968eff32c0f_6c7075.svg',color:'6c7075'}
    ,imgSrc2: {fileId:'e648f99b268782f39ea1442529369aa3.svg',name:'cr-delete.svg',assetType:'icon',bucketsource:'static',url:'f1352449307873/61a2922cfa9b623512ce97a21983a037.svg',targetSrc:'generated/e648f99b268782f39ea1442529369aa3_6c7075.svg',color:'6c7075'}
};

//TYPE: RADIO BUTTON
prx.components.basic_radiobutton = {
    name: 'basic_radiobutton'
    ,type: 'basic_radiobutton'
    ,lib: _library
    ,caption: 'Radio Button'
    ,icon: '-300px -1100px'
    ,text: 'Option'
    ,textFont: 'Arial,sans-serif'
    ,textFontStyle:  '400'
    ,textSize: 14*prx.componentsHelper.getScale(_library)
    ,textColor: '6c7075'
    ,textProperties: []
    ,helper: prx.url.devices+  '/generic2/basic_radiobutton/helper.png'
    ,width: 100*prx.componentsHelper.getScale(_library)
    ,height: 16*prx.componentsHelper.getScale(_library)
    ,resizable : true
    ,backgroundColor: 'none'
    ,borderWidth: 1*prx.componentsHelper.getScale(_library)
    ,borderColor: 'd4d7d9'
    ,activeColor: '0076ff'
    ,active: true
    ,actAsCheckbox: true
    ,checkboxActionsOnActive: []
    ,checkboxActionsOnDeactive: []
    ,radioButtonsGroupName: ''
    ,radiobuttonSize: 22*prx.componentsHelper.getScale(_library)
};

//TYPE: SELECT
prx.components.basic_select = {
    name: 'basic_select'
    ,type: 'basic_select'
    ,lib: _library
    ,caption: 'Select'
    ,icon: '-400px -1100px'
    ,helper: prx.url.devices+  '/generic2/basic_select/helper.png'
    ,width: 320*prx.componentsHelper.getScale(_library)
    ,height: 40*prx.componentsHelper.getScale(_library)
    ,resizable : true
    ,withSelection: true
    ,text: 'Sort by:'
    ,textFont: 'Arial,sans-serif'
    ,textFontStyle:  '400'
    ,textSize: 14*prx.componentsHelper.getScale(_library)
    ,textColor:  '6c7075'
    ,textProperties: []
    ,textAlign: 'left'
    ,dropdownTextFont: 'Arial,sans-serif'
    ,dropdownTextFontStyle:  '400'
    ,dropdownTextSize: 14*prx.componentsHelper.getScale(_library)
    ,dropdownTextColor:  '6c7075'
    ,dropdownTextProperties: []
    ,dropdownTextAlign: 'left'
    ,dropdownBackgroundColor: 'ffffff'
    ,dropdownHoverBackgroundColor: 'e3e7e8'
    ,dropdownActiveIcon: {fileId:'e55a24db51c6e010c487864432e3d456.svg',name:'check.svg',assetType:'icon',bucketsource:'static',url:'f1424098557988/34d6599607c21d5f87e5d30af9449c1c.svg',targetSrc:'generated/e55a24db51c6e010c487864432e3d456_6c7075.svg',color:'6c7075'}
    ,dropdownBorderWidth: 1*prx.componentsHelper.getScale(_library)
    ,dropdownBorderColor: 'D6E5E8'
    ,dropdownBorderStyle: 'solid'
    ,dropdownBorderRadius: 4*prx.componentsHelper.getScale(_library)
    ,backgroundColor: 'rgba(0,0,0,0)'
    ,borderStyle: 'solid'
    ,borderWidth: 1*prx.componentsHelper.getScale(_library)
    ,borderColor: 'd4d7d9'
    ,borderRadius: 4*prx.componentsHelper.getScale(_library)
    ,selected: 0
    ,dynamicAlignment: 'v'
    ,buttonicon: {fileId:'5be0df1935f57ae652c04aa0cb25f50e.svg',name:'unfold_more.svg',assetType:'icon',bucketsource:'static',url:'f1424098557988/f2496f33c0227f602f3f15f19eec30aa.svg',targetSrc:'generated/5be0df1935f57ae652c04aa0cb25f50e_6c7075.svg',color:'6c7075'}    ,buttons: [
        {
            text: 'Name',
            actions: []
        },
        {
            text: 'Date',
            actions: []
        },
        {
            text: 'File size',
            actions: []
        }
    ]
};

//TYPE: DROPDOWN MENU
prx.components.basic_dropdown = {
    name: 'basic_dropdown'
    ,type: 'basic_dropdown'
    ,lib: _library
    ,caption: 'Dropdown Menu'
    ,icon: '-500px -1100px'
    ,helper: prx.url.devices+  '/generic2/basic_dropdown/helper.png'
    ,width: 150*prx.componentsHelper.getScale(_library)
    ,height: 40*prx.componentsHelper.getScale(_library)
    ,resizable : true
    ,withSelection: true
    ,text: 'Menu Item'
    ,textFont: 'Arial,sans-serif'
    ,textFontStyle:  '400'
    ,textSize: 14*prx.componentsHelper.getScale(_library)
    ,textColor:  '6c7075'
    ,textProperties: []
    ,textAlign: 'right'
    ,dropdownTextFont: 'Arial,sans-serif'
    ,dropdownTextFontStyle:  '400'
    ,dropdownTextSize: 14*prx.componentsHelper.getScale(_library)
    ,dropdownTextColor:  '6c7375'
    ,dropdownTextProperties: []
    ,dropdownTextAlign: 'left'
    ,dropdownBackgroundColor: 'ffffff'
    ,dropdownBorderStyle: 'solid'
    ,dropdownBorderWidth: 1*prx.componentsHelper.getScale(_library)
    ,dropdownBorderColor: 'd4d8d9'
    ,dropdownBorderRadius: 4*prx.componentsHelper.getScale(_library)
    ,backgroundColor: 'rgba(0,0,0,0)'
    ,borderStyle: 'solid'
    ,borderWidth: 0
    ,borderColor: 'd4d8d9'
    ,borderRadius: 0*prx.componentsHelper.getScale(_library)
    ,selected: 0
    ,buttonicon: {fileId:'185f3be0c18a065efec6e893e0a6ce94.svg',name:'keyboard_arrow_down.svg',assetType:'icon',bucketsource:'static',url:'f1424098425738/f9a2843ca34c5ec824da30d7a887f664.svg',targetSrc:'generated/185f3be0c18a065efec6e893e0a6ce94_6c7075.svg',color:'6c7075'}
    ,buttons: [
        {
            text: 'Option 1',
            actions: []
        },
        {
            text: 'Option 2',
            actions: []
        },
        {
            text: 'Option 3',
            actions: []
        }
    ]
    ,dynamicAlignment: 'v'
    ,dropdownPosition: 'right'
    ,dropdownActiveBackgroundColor: 'e3e7e8'
    ,dropdownActiveTextColor: '6c7375'
    ,hoverInactiveBackgroundColor: 'ffffff'
    ,hoverInactiveTextColor: '6c7375'
    ,hoverActiveBackgroundColor: 'e3e7e8'
    ,hoverActiveTextColor: '6c7375'
};

//TYPE: PAGE CONTROLLER
prx.components.basic_pagecontroller = {
    name: 'basic_pagecontroller'
    ,type: 'basic_pagecontroller'
    ,lib: _library
    ,caption: 'Page Controller'
    ,icon: '-800px -1100px'
    ,helper: prx.url.devices+  '/generic2/basic_pagecontroller/helper.png'
    ,width: 123*prx.componentsHelper.getScale(_library)
    ,height: 10*prx.componentsHelper.getScale(_library)
    ,resizable : true
    ,backgroundColor: 'e3e6e8'
    ,activeBackgroundColor: '0076ff'
    ,borderRadius: '50%'
    ,activeSize: 1.6
    ,selected: 1
    ,dynamicSizeExpand: 'h'
    ,dynamicAlignment: 'h'
    ,buttons: [{
        actions: []
    },{
        actions: []
    },{
        actions: []
    }]
};

//TYPE: TOOLTIP
prx.components.tooltip = {
    name: 'tooltip'
    ,type: 'tooltip'
    ,lib: _library
    ,caption: 'Tooltip'
    ,icon: '-900px -1100px'
    ,helper: prx.url.devices+ '/generic2/tooltip/helper.png'
    ,width: 80*prx.componentsHelper.getScale(_library)
    ,height: 42*prx.componentsHelper.getScale(_library)
    ,resizable : true
    ,borderColor: 'rgba(0, 0, 0, 0)'
    ,backgroundColor: '4b4e4f'
    ,borderWidth: 0
    ,borderRadius: 2*prx.componentsHelper.getScale(_library)
    ,ttDirection: 'bottom'
    ,ttPosition: '50'
    ,text: 'Tooltip'
    ,textFont: 'Arial,sans-serif'
    ,textFontStyle:  '400'
    ,textSize: 10*prx.componentsHelper.getScale(_library)
    ,textColor: 'FFFFFF'
    ,textProperties: []
    ,textAlign: 'center'
    ,verticalTextAlign: 'center'
};

//TYPE: TABBAR
prx.components.basic_tabbar = {
    name: 'basic_tabbar'
    ,type: 'basic_tabbar'
    ,lib: _library
    ,caption: 'Tabbar - Text & Icon'
    ,icon: '-1100px -1100px'
    ,helper: prx.url.devices+ '/generic2/basic_tabbar/helper.png'
    ,width:'full'
    ,height: 48*prx.componentsHelper.getScale(_library)
    ,vpos: 'bottom'
    ,resizable : true
    ,backgroundColor: 'e3e6e8'
    ,borderWidth: 1*prx.componentsHelper.getScale(_library)
    ,borderColor: 'd4d7d9'
    ,textFont: 'Arial,sans-serif'
    ,textFontStyle:  '400'
    ,textSize: 10*prx.componentsHelper.getScale(_library)
    ,textColor:  '6c7075'
    ,textProperties: []
    ,iconpos: 'top'
    ,iconSize: '2'
    ,activeBackgroundColor: '0076ff'
    ,activeTextColor: 'FFFFFF'
    ,maskEnabled: true
    ,maskInactive: '6c7075'
    ,maskActive: 'ffffff'
    ,activeTab: 0
    ,overlay: false
    ,dynamicAlignment: 'h'
    ,tabs: [{text:'First',icon:{fileId:'987371e0c84e1e3877c0b8f6aea57272.svg',name:'home.svg',assetType:'icon',bucketsource:'static',url:'f1352971179296/caeebe7f3a62939528c6a4ed009de42c.svg',targetSrc:'generated/987371e0c84e1e3877c0b8f6aea57272_6c7075.svg',color:'6c7075'},activeicon:{fileId:'987371e0c84e1e3877c0b8f6aea57272.svg',name:'home.svg',assetType:'icon',bucketsource:'static',url:'f1352971179296/caeebe7f3a62939528c6a4ed009de42c.svg',targetSrc:'generated/987371e0c84e1e3877c0b8f6aea57272_ffffff.svg',color:'ffffff'},actions:[],linkedscreen:-1},{text:'Second',icon:{fileId:'324879579bf3ecf9c4b49ef0c151501b.svg',name:'speech-1.svg',assetType:'icon',bucketsource:'static',url:'f1352971179296/693bf32e7424f6bf747696ddab19cdf5.svg',targetSrc:'generated/324879579bf3ecf9c4b49ef0c151501b_6c7075.svg',color:'6c7075'},activeicon:{fileId:'324879579bf3ecf9c4b49ef0c151501b.svg',name:'speech-1.svg',assetType:'icon',bucketsource:'static',url:'f1352971179296/693bf32e7424f6bf747696ddab19cdf5.svg',targetSrc:'generated/324879579bf3ecf9c4b49ef0c151501b_ffffff.svg',color:'ffffff'},actions:[],linkedscreen:-1},{text:'Third',icon:{fileId:'f46effc9d8189665443ebcc0f7ca0934.svg',name:'heart.svg',assetType:'icon',bucketsource:'static',url:'f1515489068744/f46effc9d8189665443ebcc0f7ca0934.svg',targetSrc:'generated/f46effc9d8189665443ebcc0f7ca0934_6c7075.svg',color:'6c7075'},activeicon:{fileId:'f46effc9d8189665443ebcc0f7ca0934.svg',name:'heart.svg',assetType:'icon',bucketsource:'static',url:'f1515489068744/f46effc9d8189665443ebcc0f7ca0934.svg',targetSrc:'generated/f46effc9d8189665443ebcc0f7ca0934_ffffff.svg',color:'ffffff'},actions:[],linkedscreen:-1}]
};

//TYPE: TABBAR ICON ONLY
prx.components.basic_tabbar_icon = {
    name: 'basic_tabbar_icon'
    ,type: 'basic_tabbar_icon'
    ,lib: _library
    ,caption: 'Tabbar - Icon Only'
    ,icon: '-1000px -1100px'
    ,helper: prx.url.devices+ '/generic2/basic_tabbar_icon/helper.png'
    ,width:'full'
    ,height: 48*prx.componentsHelper.getScale(_library)
    ,vpos: 'bottom'
    ,resizable : true
    ,backgroundColor: 'e3e6e8'
    ,borderWidth: 1*prx.componentsHelper.getScale(_library)
    ,borderColor: 'd4d7d9'
    ,textFont: 'Arial,sans-serif'
    ,textFontStyle:  '400'
    ,textSize: 10*prx.componentsHelper.getScale(_library)
    ,textColor:  '6c7075'
    ,textProperties: []
    ,iconpos: 'notext'
    ,iconSize: '2'
    ,activeBackgroundColor: '0076ff'
    ,activeTextColor: 'FFFFFF'
    ,maskEnabled: true
    ,maskInactive: '6c7075'
    ,maskActive: 'FFFFFF'
    ,activeTab: 0
    ,overlay: false
    ,dynamicAlignment: 'h'
    ,tabs: [{text:'First',icon:{fileId:'987371e0c84e1e3877c0b8f6aea57272.svg',name:'home.svg',assetType:'icon',bucketsource:'static',url:'f1352971179296/caeebe7f3a62939528c6a4ed009de42c.svg',targetSrc:'generated/987371e0c84e1e3877c0b8f6aea57272_6c7075.svg',color:'6c7075'},activeicon:{fileId:'987371e0c84e1e3877c0b8f6aea57272.svg',name:'home.svg',assetType:'icon',bucketsource:'static',url:'f1352971179296/caeebe7f3a62939528c6a4ed009de42c.svg',targetSrc:'generated/987371e0c84e1e3877c0b8f6aea57272_ffffff.svg',color:'ffffff'},actions:[],linkedscreen:-1},{text:'Second',icon:{fileId:'324879579bf3ecf9c4b49ef0c151501b.svg',name:'speech-1.svg',assetType:'icon',bucketsource:'static',url:'f1352971179296/693bf32e7424f6bf747696ddab19cdf5.svg',targetSrc:'generated/324879579bf3ecf9c4b49ef0c151501b_6c7075.svg',color:'6c7075'},activeicon:{fileId:'324879579bf3ecf9c4b49ef0c151501b.svg',name:'speech-1.svg',assetType:'icon',bucketsource:'static',url:'f1352971179296/693bf32e7424f6bf747696ddab19cdf5.svg',targetSrc:'generated/324879579bf3ecf9c4b49ef0c151501b_ffffff.svg',color:'ffffff'},actions:[],linkedscreen:-1},{text:'Third',icon:{fileId:'f46effc9d8189665443ebcc0f7ca0934.svg',name:'heart.svg',assetType:'icon',bucketsource:'static',url:'f1515489068744/f46effc9d8189665443ebcc0f7ca0934.svg',targetSrc:'generated/f46effc9d8189665443ebcc0f7ca0934_6c7075.svg',color:'6c7075'},activeicon:{fileId:'f46effc9d8189665443ebcc0f7ca0934.svg',name:'heart.svg',assetType:'icon',bucketsource:'static',url:'f1515489068744/f46effc9d8189665443ebcc0f7ca0934.svg',targetSrc:'generated/f46effc9d8189665443ebcc0f7ca0934_ffffff.svg',color:'ffffff'},actions:[],linkedscreen:-1}]
};

//TYPE: TABBAR TEXT
prx.components.basic_tabbar_text = {
    name: 'basic_tabbar_text'
    ,type: 'basic_tabbar_text'
    ,lib: _library
    ,caption: 'Tabbar - Text Only'
    ,icon: '-1200px -1100px'
    ,helper: prx.url.devices+ '/generic2/basic_tabbar_text/helper.png'
    ,width:'full'
    ,height: 48*prx.componentsHelper.getScale(_library)
    ,vpos: 'bottom'
    ,resizable : true
    ,backgroundColor: 'e3e6e8'
    ,borderWidth: 1*prx.componentsHelper.getScale(_library)
    ,borderColor: 'd4d7d9'
    ,textFont: 'Arial,sans-serif'
    ,textFontStyle:  '400'
    ,textSize: 10*prx.componentsHelper.getScale(_library)
    ,textColor:  '6c7075'
    ,textProperties: []
    ,iconpos: ''
    ,iconSize: 3
    ,activeBackgroundColor: '0076ff'
    ,activeTextColor: 'FFFFFF'
    ,maskEnabled: true
    ,maskInactive: '6c7075'
    ,maskActive: 'FFFFFF'
    ,activeTab: 0
    ,overlay: false
    ,dynamicAlignment: 'h'
    ,tabs: [{text:'First',icon:{fileId:'987371e0c84e1e3877c0b8f6aea57272.svg',name:'home.svg',assetType:'icon',bucketsource:'static',url:'f1352971179296/caeebe7f3a62939528c6a4ed009de42c.svg',targetSrc:'generated/987371e0c84e1e3877c0b8f6aea57272_6c7075.svg',color:'6c7075'},activeicon:{fileId:'987371e0c84e1e3877c0b8f6aea57272.svg',name:'home.svg',assetType:'icon',bucketsource:'static',url:'f1352971179296/caeebe7f3a62939528c6a4ed009de42c.svg',targetSrc:'generated/987371e0c84e1e3877c0b8f6aea57272_ffffff.svg',color:'ffffff'},actions:[],linkedscreen:-1},{text:'Second',icon:{fileId:'324879579bf3ecf9c4b49ef0c151501b.svg',name:'speech-1.svg',assetType:'icon',bucketsource:'static',url:'f1352971179296/693bf32e7424f6bf747696ddab19cdf5.svg',targetSrc:'generated/324879579bf3ecf9c4b49ef0c151501b_6c7075.svg',color:'6c7075'},activeicon:{fileId:'324879579bf3ecf9c4b49ef0c151501b.svg',name:'speech-1.svg',assetType:'icon',bucketsource:'static',url:'f1352971179296/693bf32e7424f6bf747696ddab19cdf5.svg',targetSrc:'generated/324879579bf3ecf9c4b49ef0c151501b_ffffff.svg',color:'ffffff'},actions:[],linkedscreen:-1},{text:'Third',icon:{fileId:'f46effc9d8189665443ebcc0f7ca0934.svg',name:'heart.svg',assetType:'icon',bucketsource:'static',url:'f1515489068744/f46effc9d8189665443ebcc0f7ca0934.svg',targetSrc:'generated/f46effc9d8189665443ebcc0f7ca0934_6c7075.svg',color:'6c7075'},activeicon:{fileId:'f46effc9d8189665443ebcc0f7ca0934.svg',name:'heart.svg',assetType:'icon',bucketsource:'static',url:'f1515489068744/f46effc9d8189665443ebcc0f7ca0934.svg',targetSrc:'generated/f46effc9d8189665443ebcc0f7ca0934_ffffff.svg',color:'ffffff'},actions:[],linkedscreen:-1}]
};

//TYPE: TABBAR TEXT & ICON
prx.components.basic_tabbar_texticon = {
    name: 'basic_tabbar_texticon'
    ,type: 'basic_tabbar_texticon'
    ,lib: _library
    ,caption: 'Tabbar - Text & Icon'
    ,icon: '-1100px -1100px'
    ,helper: prx.url.devices+ '/generic2/basic_tabbar/helper.png'
    ,width:'full'
    ,height: 48*prx.componentsHelper.getScale(_library)
    ,vpos: 'bottom'
    ,resizable : true
    ,backgroundColor: 'e3e6e8'
    ,borderWidth: 1*prx.componentsHelper.getScale(_library)
    ,borderColor: 'd4d7d9'
    ,textFont: 'Arial,sans-serif'
    ,textFontStyle:  '400'
    ,textSize: 10*prx.componentsHelper.getScale(_library)
    ,textColor:  '6c7075'
    ,textProperties: []
    ,iconpos: 'top'
    ,iconSize: '2'
    ,activeBackgroundColor: '0076ff'
    ,activeTextColor: 'FFFFFF'
    ,maskEnabled: true
    ,maskInactive: '6c7075'
    ,maskActive: 'ffffff'
    ,activeTab: 0
    ,overlay: false
    ,dynamicAlignment: 'h'
    ,tabs: [{text:'First',icon:{fileId:'987371e0c84e1e3877c0b8f6aea57272.svg',name:'home.svg',assetType:'icon',bucketsource:'static',url:'f1352971179296/caeebe7f3a62939528c6a4ed009de42c.svg',targetSrc:'generated/987371e0c84e1e3877c0b8f6aea57272_6c7075.svg',color:'6c7075'},activeicon:{fileId:'987371e0c84e1e3877c0b8f6aea57272.svg',name:'home.svg',assetType:'icon',bucketsource:'static',url:'f1352971179296/caeebe7f3a62939528c6a4ed009de42c.svg',targetSrc:'generated/987371e0c84e1e3877c0b8f6aea57272_ffffff.svg',color:'ffffff'},actions:[],linkedscreen:-1},{text:'Second',icon:{fileId:'324879579bf3ecf9c4b49ef0c151501b.svg',name:'speech-1.svg',assetType:'icon',bucketsource:'static',url:'f1352971179296/693bf32e7424f6bf747696ddab19cdf5.svg',targetSrc:'generated/324879579bf3ecf9c4b49ef0c151501b_6c7075.svg',color:'6c7075'},activeicon:{fileId:'324879579bf3ecf9c4b49ef0c151501b.svg',name:'speech-1.svg',assetType:'icon',bucketsource:'static',url:'f1352971179296/693bf32e7424f6bf747696ddab19cdf5.svg',targetSrc:'generated/324879579bf3ecf9c4b49ef0c151501b_ffffff.svg',color:'ffffff'},actions:[],linkedscreen:-1},{text:'Third',icon:{fileId:'f46effc9d8189665443ebcc0f7ca0934.svg',name:'heart.svg',assetType:'icon',bucketsource:'static',url:'f1515489068744/f46effc9d8189665443ebcc0f7ca0934.svg',targetSrc:'generated/f46effc9d8189665443ebcc0f7ca0934_6c7075.svg',color:'6c7075'},activeicon:{fileId:'f46effc9d8189665443ebcc0f7ca0934.svg',name:'heart.svg',assetType:'icon',bucketsource:'static',url:'f1515489068744/f46effc9d8189665443ebcc0f7ca0934.svg',targetSrc:'generated/f46effc9d8189665443ebcc0f7ca0934_ffffff.svg',color:'ffffff'},actions:[],linkedscreen:-1}]
};
```
