## Supplement File 1 for "Post Induction Hypotension prediction during general anesthesia using Machine Learning Techniques": player-export.js

```
/*!
 * 
 *                                                                       
 *                              ::;;;;                                   
 *    ::,,,,::    ::,,  ,,,,::::::11ii::,,::,,,,::      ::::,,::,,,,::   
 *  ,,111111ii,,::1111::111111ii::111111::111111ii,,    ii11,,ii111111,, 
 *  11ii::;;11;;ii11::iiii::;;11;;11ii::ii11::::11ii    ii11;;11;;::ii11 
 *  11;;  ::11ii11;;::11ii  ::11ii11ii,,iiii..  iiii    ii11;;11::  ;;11 
 *  1111;;ii11;;11;;,,ii11::ii11;;ii11;;;;11;;;;11;;;;;;ii11::11ii;;11ii 
 *  11111111;;::11;;  ::ii1111;;  ::ii11::ii1111ii::11ii;;11,,;;1111ii:: 
 *  11;;,,::  ,,::,,      ::,,        ::::  ::::    ,,,,::::    ::,,     
 *  ;;;;                                                                 
 *                                                                       
 *  Build: 12/5/2022, 11:54:24 AM
 */!function(r){var i={};function n(t){if(i[t])return i[t].exports;var e=i[t]={i:t,l:!1,exports:{}};return r[t].call(e.exports,e,e.exports,n),e.l=!0,e.exports}n.m=r,n.c=i,n.d=function(t,e,r){n.o(t,e)||Object.defineProperty(t,e,{enumerable:!0,get:r})},n.r=function(t){"undefined"!=typeof Symbol&&Symbol.toStringTag&&Object.defineProperty(t,Symbol.toStringTag,{value:"Module"}),Object.defineProperty(t,"__esModule",{value:!0})},n.t=function(e,t){if(1&t&&(e=n(e)),8&t)return e;if(4&t&&"object"==typeof e&&e&&e.__esModule)return e;var r=Object.create(null);if(n.r(r),Object.defineProperty(r,"default",{enumerable:!0,value:e}),2&t&&"string"!=typeof e)for(var i in e)n.d(r,i,function(t){return e[t]}.bind(null,i));return r},n.n=function(t){var e=t&&t.__esModule?function(){return t.default}:function(){return t};return n.d(e,"a",e),e},n.o=function(t,e){return Object.prototype.hasOwnProperty.call(t,e)},n.p="/",n(n.s=459)}([function(t,v,y){"use strict";(function(t,e){y.d(v,"m",function(){return r}),y.d(v,"k",function(){return i}),y.d(v,"o",function(){return n}),y.d(v,"n",function(){return i}),y.d(v,"j",function(){return o}),y.d(v,"a",function(){return s}),y.d(v,"b",function(){return l}),y.d(v,"d",function(){return p}),y.d(v,"e",function(){return c}),y.d(v,"f",function(){return u}),y.d(v,"g",function(){return h}),y.d(v,"h",function(){return d}),y.d(v,"i",function(){return f}),y.d(v,"l",function(){return m}),y.d(v,"c",function(){return g});
/*!
 * VERSION: 2.0.2
 * DATE: 2018-08-27
 * UPDATES AND DOCS AT: http://greensock.com
 *
 * @license Copyright (c) 2008-2018, GreenSock. All rights reserved.
 * This work is subject to the terms at http://greensock.com/standard-license or for
 * Club GreenSock members, the software agreement that was issued with your membership.
 *
 * @author: Jack Doyle,
 */
var r="undefined"!=typeof window?window:t.exports&&void 0!==e?e:{},i=function(u){var h={},i=u.document,d=u.GreenSockGlobals=u.GreenSockGlobals||u;if(d.TweenLite)return d.TweenLite;function f(t){var e,r=t.split("."),i=d;for(e=0;e<r.length;e++)i[r[e]]=i=i[r[e]]||{};return i}function l(t){var e,r=[],i=t.length;for(e=0;e!==i;r.push(t[e++]));return r}function m(){}var t,e,r,g,v,n,a,y=f("com.greensock"),x=1e-10,b=(n=Object.prototype.toString,a=n.call([]),function(t){return null!=t&&(t instanceof Array||"object"==typeof t&&!!t.push&&n.call(t)===a)}),_={},w=function(s,l,p,c){this.sc=_[s]?_[s].sc:[],(_[s]=this).gsClass=null,this.func=p;var u=[];this.check=function(t){for(var e,r,i,n,a=l.length,o=a;-1<--a;)(e=_[l[a]]||new w(l[a],[])).gsClass?(u[a]=e.gsClass,o--):t&&e.sc.push(this);if(0===o&&p)for(i=(r=("com.greensock."+s).split(".")).pop(),n=f(r.join("."))[i]=this.gsClass=p.apply(p,u),c&&(d[i]=h[i]=n),a=0;a<this.sc.length;a++)this.sc[a].check()},this.check(!0)},o=u._gsDefine=function(t,e,r,i){return new w(t,e,r,i)},S=y._class=function(t,e,r){return e=e||function(){},o(t,[],function(){return e},r),e};o.globals=d;var s=[0,0,1,1],T=S("easing.Ease",function(t,e,r,i){this._func=t,this._type=r||0,this._power=i||0,this._params=e?s.concat(e):s},!0),P=T.map={},p=T.register=function(t,e,r,i){for(var n,a,o,s,l=e.split(","),p=l.length,c=(r||"easeIn,easeOut,easeInOut").split(",");-1<--p;)for(a=l[p],n=i?S("easing."+a,null,!0):y.easing[a]||{},o=c.length;-1<--o;)s=c[o],P[a+"."+s]=P[s+a]=n[s]=t.getRatio?t:t[s]||new t};for((r=T.prototype)._calcEnd=!1,r.getRatio=function(t){if(this._func)return this._params[0]=t,this._func.apply(null,this._params);var e=this._type,r=this._power,i=1===e?1-t:2===e?t:t<.5?2*t:2*(1-t);return 1===r?i*=i:2===r?i*=i*i:3===r?i*=i*i*i:4===r&&(i*=i*i*i*i),1===e?1-i:2===e?i:t<.5?i/2:1-i/2},e=(t=["Linear","Quad","Cubic","Quart","Quint,Strong"]).length;-1<--e;)r=t[e]+",Power"+e,p(new T(null,null,1,e),r,"easeOut",!0),p(new T(null,null,2,e),r,"easeIn"+(0===e?",easeNone":"")),p(new T(null,null,3,e),r,"easeInOut");P.linear=y.easing.Linear.easeIn,P.swing=y.easing.Quad.easeInOut;var k=S("events.EventDispatcher",function(t){this._listeners={},this._eventTarget=t||this});(r=k.prototype).addEventListener=function(t,e,r,i,n){n=n||0;var a,o,s=this._listeners[t],l=0;for(this!==g||v||g.wake(),null==s&&(this._listeners[t]=s=[]),o=s.length;-1<--o;)(a=s[o]).c===e&&a.s===r?s.splice(o,1):0===l&&a.pr<n&&(l=o+1);s.splice(l,0,{c:e,s:r,up:i,pr:n})},r.removeEventListener=function(t,e){var r,i=this._listeners[t];if(i)for(r=i.length;-1<--r;)if(i[r].c===e)return void i.splice(r,1)},r.dispatchEvent=function(t){var e,r,i,n=this._listeners[t];if(n)for(1<(e=n.length)&&(n=n.slice(0)),r=this._eventTarget;-1<--e;)(i=n[e])&&(i.up?i.c.call(i.s||r,{type:t,target:r}):i.c.call(i.s||r))};var I=u.requestAnimationFrame,A=u.cancelAnimationFrame,E=Date.now||function(){return(new Date).getTime()},C=E();for(e=(t=["ms","moz","webkit","o"]).length;-1<--e&&!I;)I=u[t[e]+"RequestAnimationFrame"],A=u[t[e]+"CancelAnimationFrame"]||u[t[e]+"CancelRequestAnimationFrame"];S("Ticker",function(t,e){var n,a,o,s,l,p=this,c=E(),r=!(!1===e||!I)&&"auto",u=500,h=33,d=function(t){var e,r,i=E()-C;u<i&&(c+=i-h),C+=i,p.time=(C-c)/1e3,e=p.time-l,(!n||0<e||!0===t)&&(p.frame++,l+=e+(s<=e?.004:s-e),r=!0),!0!==t&&(o=a(d)),r&&p.dispatchEvent("tick")};k.call(p),p.time=p.frame=0,p.tick=function(){d(!0)},p.lagSmoothing=function(t,e){if(!arguments.length)return u<1/x;u=t||1/x,h=Math.min(e,u,0)},p.sleep=function(){null!=o&&(r&&A?A(o):clearTimeout(o),a=m,o=null,p===g&&(v=!1))},p.wake=function(t){null!==o?p.sleep():t?c+=-C+(C=E()):10<p.frame&&(C=E()-u+5),a=0===n?m:r&&I?I:function(t){return setTimeout(t,1e3*(l-p.time)+1|0)},p===g&&(v=!0),d(2)},p.fps=function(t){if(!arguments.length)return n;s=1/((n=t)||60),l=this.time+s,p.wake()},p.useRAF=function(t){if(!arguments.length)return r;p.sleep(),r=t,p.fps(n)},p.fps(t),setTimeout(function(){"auto"===r&&p.frame<5&&"hidden"!==(i||{}).visibilityState&&p.useRAF(!1)},1500)}),(r=y.Ticker.prototype=new y.events.EventDispatcher).constructor=y.Ticker;var c=S("core.Animation",function(t,e){if(this.vars=e=e||{},this._duration=this._totalDuration=t||0,this._delay=Number(e.delay)||0,this._timeScale=1,this._active=!0===e.immediateRender,this.data=e.data,this._reversed=!0===e.reversed,q){v||g.wake();var r=this.vars.useFrames?U:q;r.add(this,r._time),this.vars.paused&&this.paused(!0)}});g=c.ticker=new y.Ticker,(r=c.prototype)._dirty=r._gc=r._initted=r._paused=!1,r._totalTime=r._time=0,r._rawPrevTime=-1,r._next=r._last=r._onUpdate=r._timeline=r.timeline=null,r._paused=!1;var O=function(){v&&2e3<E()-C&&("hidden"!==(i||{}).visibilityState||!g.lagSmoothing())&&g.wake();var t=setTimeout(O,2e3);t.unref&&t.unref()};O(),r.play=function(t,e){return null!=t&&this.seek(t,e),this.reversed(!1).paused(!1)},r.pause=function(t,e){return null!=t&&this.seek(t,e),this.paused(!0)},r.resume=function(t,e){return null!=t&&this.seek(t,e),this.paused(!1)},r.seek=function(t,e){return this.totalTime(Number(t),!1!==e)},r.restart=function(t,e){return this.reversed(!1).paused(!1).totalTime(t?-this._delay:0,!1!==e,!0)},r.reverse=function(t,e){return null!=t&&this.seek(t||this.totalDuration(),e),this.reversed(!0).paused(!1)},r.render=function(t,e,r){},r.invalidate=function(){return this._time=this._totalTime=0,this._initted=this._gc=!1,this._rawPrevTime=-1,!this._gc&&this.timeline||this._enabled(!0),this},r.isActive=function(){var t,e=this._timeline,r=this._startTime;return!e||!this._gc&&!this._paused&&e.isActive()&&(t=e.rawTime(!0))>=r&&t<r+this.totalDuration()/this._timeScale-1e-7},r._enabled=function(t,e){return v||g.wake(),this._gc=!t,this._active=this.isActive(),!0!==e&&(t&&!this.timeline?this._timeline.add(this,this._startTime-this._delay):!t&&this.timeline&&this._timeline._remove(this,!0)),!1},r._kill=function(t,e){return this._enabled(!1,!1)},r.kill=function(t,e){return this._kill(t,e),this},r._uncache=function(t){for(var e=t?this:this.timeline;e;)e._dirty=!0,e=e.timeline;return this},r._swapSelfInParams=function(t){for(var e=t.length,r=t.concat();-1<--e;)"{self}"===t[e]&&(r[e]=this);return r},r._callback=function(t){var e=this.vars,r=e[t],i=e[t+"Params"],n=e[t+"Scope"]||e.callbackScope||this;switch(i?i.length:0){case 0:r.call(n);break;case 1:r.call(n,i[0]);break;case 2:r.call(n,i[0],i[1]);break;default:r.apply(n,i)}},r.eventCallback=function(t,e,r,i){if("on"===(t||"").substr(0,2)){var n=this.vars;if(1===arguments.length)return n[t];null==e?delete n[t]:(n[t]=e,n[t+"Params"]=b(r)&&-1!==r.join("").indexOf("{self}")?this._swapSelfInParams(r):r,n[t+"Scope"]=i),"onUpdate"===t&&(this._onUpdate=e)}return this},r.delay=function(t){return arguments.length?(this._timeline.smoothChildTiming&&this.startTime(this._startTime+t-this._delay),this._delay=t,this):this._delay},r.duration=function(t){return arguments.length?(this._duration=this._totalDuration=t,this._uncache(!0),this._timeline.smoothChildTiming&&0<this._time&&this._time<this._duration&&0!==t&&this.totalTime(this._totalTime*(t/this._duration),!0),this):(this._dirty=!1,this._duration)},r.totalDuration=function(t){return this._dirty=!1,arguments.length?this.duration(t):this._totalDuration},r.time=function(t,e){return arguments.length?(this._dirty&&this.totalDuration(),this.totalTime(t>this._duration?this._duration:t,e)):this._time},r.totalTime=function(t,e,r){if(v||g.wake(),!arguments.length)return this._totalTime;if(this._timeline){if(t<0&&!r&&(t+=this.totalDuration()),this._timeline.smoothChildTiming){this._dirty&&this.totalDuration();var i=this._totalDuration,n=this._timeline;if(i<t&&!r&&(t=i),this._startTime=(this._paused?this._pauseTime:n._time)-(this._reversed?i-t:t)/this._timeScale,n._dirty||this._uncache(!1),n._timeline)for(;n._timeline;)n._timeline._time!==(n._startTime+n._totalTime)/n._timeScale&&n.totalTime(n._totalTime,!0),n=n._timeline}this._gc&&this._enabled(!0,!1),this._totalTime===t&&0!==this._duration||(B.length&&Q(),this.render(t,e,!1),B.length&&Q())}return this},r.progress=r.totalProgress=function(t,e){var r=this.duration();return arguments.length?this.totalTime(r*t,e):r?this._time/r:this.ratio},r.startTime=function(t){return arguments.length?(t!==this._startTime&&(this._startTime=t,this.timeline&&this.timeline._sortChildren&&this.timeline.add(this,t-this._delay)),this):this._startTime},r.endTime=function(t){return this._startTime+(0!=t?this.totalDuration():this.duration())/this._timeScale},r.timeScale=function(t){if(!arguments.length)return this._timeScale;var e,r;for(t=t||x,this._timeline&&this._timeline.smoothChildTiming&&(r=(e=this._pauseTime)||0===e?e:this._timeline.totalTime(),this._startTime=r-(r-this._startTime)*this._timeScale/t),this._timeScale=t,r=this.timeline;r&&r.timeline;)r._dirty=!0,r.totalDuration(),r=r.timeline;return this},r.reversed=function(t){return arguments.length?(t!=this._reversed&&(this._reversed=t,this.totalTime(this._timeline&&!this._timeline.smoothChildTiming?this.totalDuration()-this._totalTime:this._totalTime,!0)),this):this._reversed},r.paused=function(t){if(!arguments.length)return this._paused;var e,r,i=this._timeline;return t!=this._paused&&i&&(v||t||g.wake(),r=(e=i.rawTime())-this._pauseTime,!t&&i.smoothChildTiming&&(this._startTime+=r,this._uncache(!1)),this._pauseTime=t?e:null,this._paused=t,this._active=this.isActive(),!t&&0!=r&&this._initted&&this.duration()&&(e=i.smoothChildTiming?this._totalTime:(e-this._startTime)/this._timeScale,this.render(e,e===this._totalTime,!0))),this._gc&&!t&&this._enabled(!0,!1),this};var M=S("core.SimpleTimeline",function(t){c.call(this,0,t),this.autoRemoveChildren=this.smoothChildTiming=!0});(r=M.prototype=new c).constructor=M,r.kill()._gc=!1,r._first=r._last=r._recent=null,r._sortChildren=!1,r.add=r.insert=function(t,e,r,i){var n,a;if(t._startTime=Number(e||0)+t._delay,t._paused&&this!==t._timeline&&(t._pauseTime=this.rawTime()-(t._timeline.rawTime()-t._pauseTime)),t.timeline&&t.timeline._remove(t,!0),t.timeline=t._timeline=this,t._gc&&t._enabled(!0,!0),n=this._last,this._sortChildren)for(a=t._startTime;n&&n._startTime>a;)n=n._prev;return n?(t._next=n._next,n._next=t):(t._next=this._first,this._first=t),t._next?t._next._prev=t:this._last=t,t._prev=n,this._recent=t,this._timeline&&this._uncache(!0),this},r._remove=function(t,e){return t.timeline===this&&(e||t._enabled(!1,!0),t._prev?t._prev._next=t._next:this._first===t&&(this._first=t._next),t._next?t._next._prev=t._prev:this._last===t&&(this._last=t._prev),t._next=t._prev=t.timeline=null,t===this._recent&&(this._recent=this._last),this._timeline&&this._uncache(!0)),this},r.render=function(t,e,r){var i,n=this._first;for(this._totalTime=this._time=this._rawPrevTime=t;n;)i=n._next,(n._active||t>=n._startTime&&!n._paused&&!n._gc)&&(n._reversed?n.render((n._dirty?n.totalDuration():n._totalDuration)-(t-n._startTime)*n._timeScale,e,r):n.render((t-n._startTime)*n._timeScale,e,r)),n=i},r.rawTime=function(){return v||g.wake(),this._totalTime};function D(t){return t&&t.length&&t!==u&&t[0]&&(t[0]===u||t[0].nodeType&&t[0].style&&!t.nodeType)}var F=S("TweenLite",function(t,e,r){if(c.call(this,e,r),this.render=F.prototype.render,null==t)throw"Cannot tween a null target.";this.target=t="string"!=typeof t?t:F.selector(t)||t;var i,n,a,o=t.jquery||t.length&&t!==u&&t[0]&&(t[0]===u||t[0].nodeType&&t[0].style&&!t.nodeType),s=this.vars.overwrite;if(this._overwrite=s=null==s?W[F.defaultOverwrite]:"number"==typeof s?s>>0:W[s],(o||t instanceof Array||t.push&&b(t))&&"number"!=typeof t[0])for(this._targets=a=l(t),this._propLookup=[],this._siblings=[],i=0;i<a.length;i++)(n=a[i])?"string"!=typeof n?n.length&&n!==u&&n[0]&&(n[0]===u||n[0].nodeType&&n[0].style&&!n.nodeType)?(a.splice(i--,1),this._targets=a=a.concat(l(n))):(this._siblings[i]=K(n,this,!1),1===s&&1<this._siblings[i].length&&tt(n,this,null,1,this._siblings[i])):"string"==typeof(n=a[i--]=F.selector(n))&&a.splice(i+1,1):a.splice(i--,1);else this._propLookup={},this._siblings=K(t,this,!1),1===s&&1<this._siblings.length&&tt(t,this,null,1,this._siblings);(this.vars.immediateRender||0===e&&0===this._delay&&!1!==this.vars.immediateRender)&&(this._time=-x,this.render(Math.min(0,-this._delay)))},!0);(r=F.prototype=new c).constructor=F,r.kill()._gc=!1,r.ratio=0,r._firstPT=r._targets=r._overwrittenProps=r._startAt=null,r._notifyPluginsOfEnabled=r._lazy=!1,F.version="2.0.2",F.defaultEase=r._ease=new T(null,null,1,1),F.defaultOverwrite="auto",F.ticker=g,F.autoSleep=120,F.lagSmoothing=function(t,e){g.lagSmoothing(t,e)},F.selector=u.$||u.jQuery||function(t){var e=u.$||u.jQuery;return e?(F.selector=e)(t):(i||(i=u.document),i?i.querySelectorAll?i.querySelectorAll(t):i.getElementById("#"===t.charAt(0)?t.substr(1):t):t)};function R(t){for(var e,r=this._firstPT;r;)e=r.blob?1===t&&null!=this.end?this.end:t?this.join(""):this.start:r.c*t+r.s,r.m?e=r.m.call(this._tween,e,this._target||r.t,this._tween):e<1e-6&&-1e-6<e&&!r.blob&&(e=0),r.f?r.fp?r.t[r.p](r.fp,e):r.t[r.p](e):r.t[r.p]=e,r=r._next}function N(t,e,r,i){var n,a,o,s,l,p,c,u=[],h=0,d="",f=0;for(u.start=t,u.end=e,t=u[0]=t+"",e=u[1]=e+"",r&&(r(u),t=u[0],e=u[1]),u.length=0,n=t.match($)||[],a=e.match($)||[],i&&(i._next=null,i.blob=1,u._firstPT=u._applyPT=i),l=a.length,s=0;s<l;s++)c=a[s],d+=(p=e.substr(h,e.indexOf(c,h)-h))||!s?p:",",h+=p.length,f?f=(f+1)%5:"rgba("===p.substr(-5)&&(f=1),c===n[s]||n.length<=s?d+=c:(d&&(u.push(d),d=""),o=parseFloat(n[s]),u.push(o),u._firstPT={_next:u._firstPT,t:u,p:u.length-1,s:o,c:("="===c.charAt(1)?parseInt(c.charAt(0)+"1",10)*parseFloat(c.substr(2)):parseFloat(c)-o)||0,f:0,m:f&&f<4?Math.round:0}),h+=c.length;return(d+=e.substr(h))&&u.push(d),u.setRatio=R,V.test(e)&&(u.end=null),u}function L(t,e,r,i,n,a,o,s,l){"function"==typeof i&&(i=i(l||0,t));var p=typeof t[e],c="function"!=p?"":e.indexOf("set")||"function"!=typeof t["get"+e.substr(3)]?e:"get"+e.substr(3),u="get"!==r?r:c?o?t[c](o):t[c]():t[e],h="string"==typeof i&&"="===i.charAt(1),d={t:t,p:e,s:u,f:"function"==p,pg:0,n:n||e,m:a?"function"==typeof a?a:Math.round:0,pr:0,c:h?parseInt(i.charAt(0)+"1",10)*parseFloat(i.substr(2)):parseFloat(i)-u||0};if("number"==typeof u&&("number"==typeof i||h)||(o||isNaN(u)||!h&&isNaN(i)||"boolean"==typeof u||"boolean"==typeof i?(d.fp=o,d={t:N(u,h?parseFloat(d.s)+d.c+(d.s+"").replace(/[0-9\-\.]/g,""):i,s||F.defaultStringFilter,d),p:"setRatio",s:0,c:1,f:2,pg:0,n:n||e,pr:0,m:0}):(d.s=parseFloat(u),h||(d.c=parseFloat(i)-d.s||0))),d.c)return(d._next=this._firstPT)&&(d._next._prev=d),this._firstPT=d}var B=[],j={},$=/(?:(-|-=|\+=)?\d*\.?\d*(?:e[\-+]?\d+)?)[0-9]/gi,V=/[\+-]=-?[\.\d]/,H=F._internals={isArray:b,isSelector:D,lazyTweens:B,blobDif:N},z=F._plugins={},G=H.tweenLookup={},Y=0,X=H.reservedProps={ease:1,delay:1,overwrite:1,onComplete:1,onCompleteParams:1,onCompleteScope:1,useFrames:1,runBackwards:1,startAt:1,onUpdate:1,onUpdateParams:1,onUpdateScope:1,onStart:1,onStartParams:1,onStartScope:1,onReverseComplete:1,onReverseCompleteParams:1,onReverseCompleteScope:1,onRepeat:1,onRepeatParams:1,onRepeatScope:1,easeParams:1,yoyo:1,immediateRender:1,repeat:1,repeatDelay:1,data:1,paused:1,reversed:1,autoCSS:1,lazy:1,onOverwrite:1,callbackScope:1,stringFilter:1,id:1,yoyoEase:1},W={none:0,all:1,auto:2,concurrent:3,allOnStart:4,preexisting:5,true:1,false:0},U=c._rootFramesTimeline=new M,q=c._rootTimeline=new M,J=30,Q=H.lazyRender=function(){var t,e=B.length;for(j={};-1<--e;)(t=B[e])&&!1!==t._lazy&&(t.render(t._lazy[0],t._lazy[1],!0),t._lazy=!1);B.length=0};q._startTime=g.time,U._startTime=g.frame,q._active=U._active=!0,setTimeout(Q,1),c._updateRoot=F.render=function(){var t,e,r;if(B.length&&Q(),q.render((g.time-q._startTime)*q._timeScale,!1,!1),U.render((g.frame-U._startTime)*U._timeScale,!1,!1),B.length&&Q(),g.frame>=J){for(r in J=g.frame+(parseInt(F.autoSleep,10)||120),G){for(t=(e=G[r].tweens).length;-1<--t;)e[t]._gc&&e.splice(t,1);0===e.length&&delete G[r]}if((!(r=q._first)||r._paused)&&F.autoSleep&&!U._first&&1===g._listeners.tick.length){for(;r&&r._paused;)r=r._next;r||g.sleep()}}},g.addEventListener("tick",c._updateRoot);function Z(t,e,r,i){var n,a,o=t.vars.onOverwrite;return o&&(n=o(t,e,r,i)),(o=F.onOverwrite)&&(a=o(t,e,r,i)),!1!==n&&!1!==a}var K=function(t,e,r){var i,n,a=t._gsTweenID;if(G[a||(t._gsTweenID=a="t"+Y++)]||(G[a]={target:t,tweens:[]}),e&&((i=G[a].tweens)[n=i.length]=e,r))for(;-1<--n;)i[n]===e&&i.splice(n,1);return G[a].tweens},tt=function(t,e,r,i,n){var a,o,s,l;if(1===i||4<=i){for(l=n.length,a=0;a<l;a++)if((s=n[a])!==e)s._gc||s._kill(null,t,e)&&(o=!0);else if(5===i)break;return o}var p,c=e._startTime+x,u=[],h=0,d=0===e._duration;for(a=n.length;-1<--a;)(s=n[a])===e||s._gc||s._paused||(s._timeline!==e._timeline?(p=p||et(e,0,d),0===et(s,p,d)&&(u[h++]=s)):s._startTime<=c&&s._startTime+s.totalDuration()/s._timeScale>c&&((d||!s._initted)&&c-s._startTime<=2e-10||(u[h++]=s)));for(a=h;-1<--a;)if(l=(s=u[a])._firstPT,2===i&&s._kill(r,t,e)&&(o=!0),2!==i||!s._firstPT&&s._initted&&l){if(2!==i&&!Z(s,e))continue;s._enabled(!1,!1)&&(o=!0)}return o},et=function(t,e,r){for(var i=t._timeline,n=i._timeScale,a=t._startTime;i._timeline;){if(a+=i._startTime,n*=i._timeScale,i._paused)return-100;i=i._timeline}return e<(a/=n)?a-e:r&&a===e||!t._initted&&a-e<2*x?x:(a+=t.totalDuration()/t._timeScale/n)>e+x?0:a-e-x};r._init=function(){var t,e,r,i,n,a,o=this.vars,s=this._overwrittenProps,l=this._duration,p=!!o.immediateRender,c=o.ease;if(o.startAt){for(i in this._startAt&&(this._startAt.render(-1,!0),this._startAt.kill()),n={},o.startAt)n[i]=o.startAt[i];if(n.data="isStart",n.overwrite=!1,n.immediateRender=!0,n.lazy=p&&!1!==o.lazy,n.startAt=n.delay=null,n.onUpdate=o.onUpdate,n.onUpdateParams=o.onUpdateParams,n.onUpdateScope=o.onUpdateScope||o.callbackScope||this,this._startAt=F.to(this.target||{},0,n),p)if(0<this._time)this._startAt=null;else if(0!==l)return}else if(o.runBackwards&&0!==l)if(this._startAt)this._startAt.render(-1,!0),this._startAt.kill(),this._startAt=null;else{for(i in 0!==this._time&&(p=!1),r={},o)X[i]&&"autoCSS"!==i||(r[i]=o[i]);if(r.overwrite=0,r.data="isFromStart",r.lazy=p&&!1!==o.lazy,r.immediateRender=p,this._startAt=F.to(this.target,0,r),p){if(0===this._time)return}else this._startAt._init(),this._startAt._enabled(!1),this.vars.immediateRender&&(this._startAt=null)}if(this._ease=c=c?c instanceof T?c:"function"==typeof c?new T(c,o.easeParams):P[c]||F.defaultEase:F.defaultEase,o.easeParams instanceof Array&&c.config&&(this._ease=c.config.apply(c,o.easeParams)),this._easeType=this._ease._type,this._easePower=this._ease._power,this._firstPT=null,this._targets)for(a=this._targets.length,t=0;t<a;t++)this._initProps(this._targets[t],this._propLookup[t]={},this._siblings[t],s?s[t]:null,t)&&(e=!0);else e=this._initProps(this.target,this._propLookup,this._siblings,s,0);if(e&&F._onPluginEvent("_onInitAllProps",this),s&&(this._firstPT||"function"!=typeof this.target&&this._enabled(!1,!1)),o.runBackwards)for(r=this._firstPT;r;)r.s+=r.c,r.c=-r.c,r=r._next;this._onUpdate=o.onUpdate,this._initted=!0},r._initProps=function(t,e,r,i,n){var a,o,s,l,p,c;if(null==t)return!1;for(a in j[t._gsTweenID]&&Q(),this.vars.css||t.style&&t!==u&&t.nodeType&&z.css&&!1!==this.vars.autoCSS&&function(t,e){var r,i={};for(r in t)X[r]||r in e&&"transform"!==r&&"x"!==r&&"y"!==r&&"width"!==r&&"height"!==r&&"className"!==r&&"border"!==r||!(!z[r]||z[r]&&z[r]._autoCSS)||(i[r]=t[r],delete t[r]);t.css=i}(this.vars,t),this.vars)if(c=this.vars[a],X[a])c&&(c instanceof Array||c.push&&b(c))&&-1!==c.join("").indexOf("{self}")&&(this.vars[a]=c=this._swapSelfInParams(c,this));else if(z[a]&&(l=new z[a])._onInitTween(t,this.vars[a],this,n)){for(this._firstPT=p={_next:this._firstPT,t:l,p:"setRatio",s:0,c:1,f:1,n:a,pg:1,pr:l._priority,m:0},o=l._overwriteProps.length;-1<--o;)e[l._overwriteProps[o]]=this._firstPT;(l._priority||l._onInitAllProps)&&(s=!0),(l._onDisable||l._onEnable)&&(this._notifyPluginsOfEnabled=!0),p._next&&(p._next._prev=p)}else e[a]=L.call(this,t,a,"get",c,a,0,null,this.vars.stringFilter,n);return i&&this._kill(i,t)?this._initProps(t,e,r,i,n):1<this._overwrite&&this._firstPT&&1<r.length&&tt(t,this,e,this._overwrite,r)?(this._kill(e,t),this._initProps(t,e,r,i,n)):(this._firstPT&&(!1!==this.vars.lazy&&this._duration||this.vars.lazy&&!this._duration)&&(j[t._gsTweenID]=!0),s)},r.render=function(t,e,r){var i,n,a,o,s=this._time,l=this._duration,p=this._rawPrevTime;if(l-1e-7<=t&&0<=t)this._totalTime=this._time=l,this.ratio=this._ease._calcEnd?this._ease.getRatio(1):1,this._reversed||(i=!0,n="onComplete",r=r||this._timeline.autoRemoveChildren),0===l&&(!this._initted&&this.vars.lazy&&!r||(this._startTime===this._timeline._duration&&(t=0),(p<0||t<=0&&-1e-7<=t||p===x&&"isPause"!==this.data)&&p!==t&&(r=!0,x<p&&(n="onReverseComplete")),this._rawPrevTime=o=!e||t||p===t?t:x));else if(t<1e-7)this._totalTime=this._time=0,this.ratio=this._ease._calcEnd?this._ease.getRatio(0):0,(0!==s||0===l&&0<p)&&(n="onReverseComplete",i=this._reversed),t<0&&(this._active=!1,0===l&&(!this._initted&&this.vars.lazy&&!r||(0<=p&&(p!==x||"isPause"!==this.data)&&(r=!0),this._rawPrevTime=o=!e||t||p===t?t:x))),(!this._initted||this._startAt&&this._startAt.progress())&&(r=!0);else if(this._totalTime=this._time=t,this._easeType){var c=t/l,u=this._easeType,h=this._easePower;(1===u||3===u&&.5<=c)&&(c=1-c),3===u&&(c*=2),1===h?c*=c:2===h?c*=c*c:3===h?c*=c*c*c:4===h&&(c*=c*c*c*c),this.ratio=1===u?1-c:2===u?c:t/l<.5?c/2:1-c/2}else this.ratio=this._ease.getRatio(t/l);if(this._time!==s||r){if(!this._initted){if(this._init(),!this._initted||this._gc)return;if(!r&&this._firstPT&&(!1!==this.vars.lazy&&this._duration||this.vars.lazy&&!this._duration))return this._time=this._totalTime=s,this._rawPrevTime=p,B.push(this),void(this._lazy=[t,e]);this._time&&!i?this.ratio=this._ease.getRatio(this._time/l):i&&this._ease._calcEnd&&(this.ratio=this._ease.getRatio(0===this._time?0:1))}for(!1!==this._lazy&&(this._lazy=!1),this._active||!this._paused&&this._time!==s&&0<=t&&(this._active=!0),0===s&&(this._startAt&&(0<=t?this._startAt.render(t,!0,r):n||(n="_dummyGS")),this.vars.onStart&&(0===this._time&&0!==l||e||this._callback("onStart"))),a=this._firstPT;a;)a.f?a.t[a.p](a.c*this.ratio+a.s):a.t[a.p]=a.c*this.ratio+a.s,a=a._next;this._onUpdate&&(t<0&&this._startAt&&-1e-4!==t&&this._startAt.render(t,!0,r),e||(this._time!==s||i||r)&&this._callback("onUpdate")),n&&(this._gc&&!r||(t<0&&this._startAt&&!this._onUpdate&&-1e-4!==t&&this._startAt.render(t,!0,r),i&&(this._timeline.autoRemoveChildren&&this._enabled(!1,!1),this._active=!1),!e&&this.vars[n]&&this._callback(n),0===l&&this._rawPrevTime===x&&o!==x&&(this._rawPrevTime=0)))}},r._kill=function(t,e,r){if("all"===t&&(t=null),null==t&&(null==e||e===this.target))return this._lazy=!1,this._enabled(!1,!1);e="string"!=typeof e?e||this._targets||this.target:F.selector(e)||e;var i,n,a,o,s,l,p,c,u,h=r&&this._time&&r._startTime===this._startTime&&this._timeline===r._timeline,d=this._firstPT;if((b(e)||D(e))&&"number"!=typeof e[0])for(i=e.length;-1<--i;)this._kill(t,e[i],r)&&(l=!0);else{if(this._targets){for(i=this._targets.length;-1<--i;)if(e===this._targets[i]){s=this._propLookup[i]||{},this._overwrittenProps=this._overwrittenProps||[],n=this._overwrittenProps[i]=t?this._overwrittenProps[i]||{}:"all";break}}else{if(e!==this.target)return!1;s=this._propLookup,n=this._overwrittenProps=t?this._overwrittenProps||{}:"all"}if(s){if(p=t||s,c=t!==n&&"all"!==n&&t!==s&&("object"!=typeof t||!t._tempKill),r&&(F.onOverwrite||this.vars.onOverwrite)){for(a in p)s[a]&&(u||(u=[]),u.push(a));if((u||!t)&&!Z(this,r,e,u))return!1}for(a in p)(o=s[a])&&(h&&(o.f?o.t[o.p](o.s):o.t[o.p]=o.s,l=!0),o.pg&&o.t._kill(p)&&(l=!0),o.pg&&0!==o.t._overwriteProps.length||(o._prev?o._prev._next=o._next:o===this._firstPT&&(this._firstPT=o._next),o._next&&(o._next._prev=o._prev),o._next=o._prev=null),delete s[a]),c&&(n[a]=1);!this._firstPT&&this._initted&&d&&this._enabled(!1,!1)}}return l},r.invalidate=function(){return this._notifyPluginsOfEnabled&&F._onPluginEvent("_onDisable",this),this._firstPT=this._overwrittenProps=this._startAt=this._onUpdate=null,this._notifyPluginsOfEnabled=this._active=this._lazy=!1,this._propLookup=this._targets?{}:[],c.prototype.invalidate.call(this),this.vars.immediateRender&&(this._time=-x,this.render(Math.min(0,-this._delay))),this},r._enabled=function(t,e){if(v||g.wake(),t&&this._gc){var r,i=this._targets;if(i)for(r=i.length;-1<--r;)this._siblings[r]=K(i[r],this,!0);else this._siblings=K(this.target,this,!0)}return c.prototype._enabled.call(this,t,e),!(!this._notifyPluginsOfEnabled||!this._firstPT)&&F._onPluginEvent(t?"_onEnable":"_onDisable",this)},F.to=function(t,e,r){return new F(t,e,r)},F.from=function(t,e,r){return r.runBackwards=!0,r.immediateRender=0!=r.immediateRender,new F(t,e,r)},F.fromTo=function(t,e,r,i){return i.startAt=r,i.immediateRender=0!=i.immediateRender&&0!=r.immediateRender,new F(t,e,i)},F.delayedCall=function(t,e,r,i,n){return new F(e,0,{delay:t,onComplete:e,onCompleteParams:r,callbackScope:i,onReverseComplete:e,onReverseCompleteParams:r,immediateRender:!1,lazy:!1,useFrames:n,overwrite:0})},F.set=function(t,e){return new F(t,0,e)},F.getTweensOf=function(t,e){if(null==t)return[];var r,i,n,a;if(t="string"!=typeof t?t:F.selector(t)||t,(b(t)||D(t))&&"number"!=typeof t[0]){for(r=t.length,i=[];-1<--r;)i=i.concat(F.getTweensOf(t[r],e));for(r=i.length;-1<--r;)for(a=i[r],n=r;-1<--n;)a===i[n]&&i.splice(r,1)}else if(t._gsTweenID)for(r=(i=K(t).concat()).length;-1<--r;)(i[r]._gc||e&&!i[r].isActive())&&i.splice(r,1);return i||[]},F.killTweensOf=F.killDelayedCallsTo=function(t,e,r){"object"==typeof e&&(r=e,e=!1);for(var i=F.getTweensOf(t,e),n=i.length;-1<--n;)i[n]._kill(r,t)};var rt=S("plugins.TweenPlugin",function(t,e){this._overwriteProps=(t||"").split(","),this._propName=this._overwriteProps[0],this._priority=e||0,this._super=rt.prototype},!0);if(r=rt.prototype,rt.version="1.19.0",rt.API=2,r._firstPT=null,r._addTween=L,r.setRatio=R,r._kill=function(t){var e,r=this._overwriteProps,i=this._firstPT;if(null!=t[this._propName])this._overwriteProps=[];else for(e=r.length;-1<--e;)null!=t[r[e]]&&r.splice(e,1);for(;i;)null!=t[i.n]&&(i._next&&(i._next._prev=i._prev),i._prev?(i._prev._next=i._next,i._prev=null):this._firstPT===i&&(this._firstPT=i._next)),i=i._next;return!1},r._mod=r._roundProps=function(t){for(var e,r=this._firstPT;r;)(e=t[this._propName]||null!=r.n&&t[r.n.split(this._propName+"_").join("")])&&"function"==typeof e&&(2===r.f?r.t._applyPT.m=e:r.m=e),r=r._next},F._onPluginEvent=function(t,e){var r,i,n,a,o,s=e._firstPT;if("_onInitAllProps"===t){for(;s;){for(o=s._next,i=n;i&&i.pr>s.pr;)i=i._next;(s._prev=i?i._prev:a)?s._prev._next=s:n=s,(s._next=i)?i._prev=s:a=s,s=o}s=e._firstPT=n}for(;s;)s.pg&&"function"==typeof s.t[t]&&s.t[t]()&&(r=!0),s=s._next;return r},rt.activate=function(t){for(var e=t.length;-1<--e;)t[e].API===rt.API&&(z[(new t[e])._propName]=t[e]);return!0},o.plugin=function(t){if(!(t&&t.propName&&t.init&&t.API))throw"illegal plugin definition.";var e,r=t.propName,i=t.priority||0,n=t.overwriteProps,a={init:"_onInitTween",set:"setRatio",kill:"_kill",round:"_mod",mod:"_mod",initAll:"_onInitAllProps"},o=S("plugins."+r.charAt(0).toUpperCase()+r.substr(1)+"Plugin",function(){rt.call(this,r,i),this._overwriteProps=n||[]},!0===t.global),s=o.prototype=new rt(r);for(e in(s.constructor=o).API=t.API,a)"function"==typeof t[e]&&(s[a[e]]=t[e]);return o.version=t.version,rt.activate([o]),o},t=u._gsQueue){for(e=0;e<t.length;e++)t[e]();for(r in _)_[r].func||u.console.log("GSAP encountered missing dependency: "+r)}return v=!1,F}(r),n=r.GreenSockGlobals,a=n.com.greensock,o=a.core.SimpleTimeline,s=a.core.Animation,l=n.Ease,p=n.Linear,c=p,u=n.Power1,h=n.Power2,d=n.Power3,f=n.Power4,m=n.TweenPlugin,g=a.events.EventDispatcher}).call(this,y(371)(t),y(81))},function(t,e,r){var c=r(3),u=r(16).f,h=r(14),d=r(17),f=r(83),m=r(107),g=r(55);t.exports=function(t,e){var r,i,n,a,o,s=t.target,l=t.global,p=t.stat;if(r=l?c:p?c[s]||f(s,{}):(c[s]||{}).prototype)for(i in e){if(a=e[i],n=t.noTargetGet?(o=u(r,i))&&o.value:r[i],!g(l?i:s+(p?".":"#")+i,t.forced)&&void 0!==n){if(typeof a==typeof n)continue;m(a,n)}(t.sham||n&&n.sham)&&h(a,"sham",!0),d(r,i,a,t)}}},function(t,e){t.exports=function(t){try{return!!t()}catch(t){return!0}}},function(i,t,e){(function(t){function e(t){return t&&t.Math==Math&&t}var r="object";i.exports=e(typeof globalThis==r&&globalThis)||e(typeof window==r&&window)||e(typeof self==r&&self)||e(typeof t==r&&t)||Function("return this")()}).call(this,e(81))},function(t,e){t.exports=function(t){return"object"==typeof t?null!==t:"function"==typeof t}},function(t,e,r){var i=r(4);t.exports=function(t){if(!i(t))throw TypeError(String(t)+" is not an object");return t}},function(t,e,r){"use strict";function i(t){return s(t)&&l(M,p(t))}var n,a=r(7),o=r(3),s=r(4),l=r(12),p=r(62),c=r(14),u=r(17),h=r(10).f,d=r(28),f=r(45),m=r(8),g=r(52),v=o.DataView,y=v&&v.prototype,x=o.Int8Array,b=x&&x.prototype,_=o.Uint8ClampedArray,w=_&&_.prototype,S=x&&d(x),T=b&&d(b),P=Object.prototype,k=P.isPrototypeOf,I=m("toStringTag"),A=g("TYPED_ARRAY_TAG"),E=!(!o.ArrayBuffer||!v),C=E&&!!f,O=!1,M={Int8Array:1,Uint8Array:1,Uint8ClampedArray:1,Int16Array:2,Uint16Array:2,Int32Array:4,Uint32Array:4,Float32Array:4,Float64Array:8};for(n in M)o[n]||(C=!1);if((!C||"function"!=typeof S||S===Function.prototype)&&(S=function(){throw TypeError("Incorrect invocation")},C))for(n in M)o[n]&&f(o[n],S);if((!C||!T||T===P)&&(T=S.prototype,C))for(n in M)o[n]&&f(o[n].prototype,T);if(C&&d(w)!==T&&f(w,T),a&&!l(T,I))for(n in O=!0,h(T,I,{get:function(){return s(this)?this[A]:void 0}}),M)o[n]&&c(o[n],A,n);E&&f&&d(y)!==P&&f(y,P),t.exports={NATIVE_ARRAY_BUFFER:E,NATIVE_ARRAY_BUFFER_VIEWS:C,TYPED_ARRAY_TAG:O&&A,aTypedArray:function(t){if(i(t))return t;throw TypeError("Target is not a typed array")},aTypedArrayConstructor:function(t){if(f){if(k.call(S,t))return t}else for(var e in M)if(l(M,n)){var r=o[e];if(r&&(t===r||k.call(r,t)))return t}throw TypeError("Target is not a typed array constructor")},exportProto:function(t,e,r){if(a){if(r)for(var i in M){var n=o[i];n&&l(n.prototype,t)&&delete n.prototype[t]}T[t]&&!r||u(T,t,r?e:C&&b[t]||e)}},exportStatic:function(t,e,r){var i,n;if(a){if(f){if(r)for(i in M)(n=o[i])&&l(n,t)&&delete n[t];if(S[t]&&!r)return;try{return u(S,t,r?e:C&&x[t]||e)}catch(t){}}for(i in M)!(n=o[i])||n[t]&&!r||u(n,t,e)}},isView:function(t){var e=p(t);return"DataView"===e||l(M,e)},isTypedArray:i,TypedArray:S,TypedArrayPrototype:T}},function(t,e,r){var i=r(2);t.exports=!i(function(){return 7!=Object.defineProperty({},"a",{get:function(){return 7}}).a})},function(t,e,r){var i=r(3),n=r(51),a=r(52),o=r(109),s=i.Symbol,l=n("wks");t.exports=function(t){return l[t]||(l[t]=o&&s[t]||(o?s:a)("Symbol."+t))}},function(t,e,r){var i=r(24),n=Math.min;t.exports=function(t){return 0<t?n(i(t),9007199254740991):0}},function(t,e,r){var i=r(7),n=r(104),a=r(5),o=r(26),s=Object.defineProperty;e.f=i?s:function(t,e,r){if(a(t),e=o(e,!0),a(r),n)try{return s(t,e,r)}catch(t){}if("get"in r||"set"in r)throw TypeError("Accessors not supported");return"value"in r&&(t[e]=r.value),t}},function(t,e,r){var i=r(15);t.exports=function(t){return Object(i(t))}},function(t,e){var r={}.hasOwnProperty;t.exports=function(t,e){return r.call(t,e)}},function(t,e,r){function i(d){var f=1==d,m=2==d,g=3==d,v=4==d,y=6==d,x=5==d||y;return function(t,e,r,i){for(var n,a,o=w(t),s=_(o),l=b(e,r,3),p=S(s.length),c=0,u=i||T,h=f?u(t,p):m?u(t,0):void 0;c<p;c++)if((x||c in s)&&(a=l(n=s[c],c,o),d))if(f)h[c]=a;else if(a)switch(d){case 3:return!0;case 5:return n;case 6:return c;case 2:P.call(h,n)}else if(v)return!1;return y?-1:g||v?v:h}}var b=r(36),_=r(50),w=r(11),S=r(9),T=r(57),P=[].push;t.exports={forEach:i(0),map:i(1),filter:i(2),some:i(3),every:i(4),find:i(5),findIndex:i(6)}},function(t,e,r){var i=r(7),n=r(10),a=r(39);t.exports=i?function(t,e,r){return n.f(t,e,a(1,r))}:function(t,e,r){return t[e]=r,t}},function(t,e){t.exports=function(t){if(null==t)throw TypeError("Can't call method on "+t);return t}},function(t,e,r){var i=r(7),n=r(65),a=r(39),o=r(19),s=r(26),l=r(12),p=r(104),c=Object.getOwnPropertyDescriptor;e.f=i?c:function(t,e){if(t=o(t),e=s(e,!0),p)try{return c(t,e)}catch(t){}if(l(t,e))return a(!n.f.call(t,e),t[e])}},function(t,e,r){var s=r(3),i=r(51),l=r(14),p=r(12),c=r(83),n=r(105),a=r(20),o=a.get,u=a.enforce,h=String(n).split("toString");i("inspectSource",function(t){return n.call(t)}),(t.exports=function(t,e,r,i){var n=!!i&&!!i.unsafe,a=!!i&&!!i.enumerable,o=!!i&&!!i.noTargetGet;"function"==typeof r&&("string"!=typeof e||p(r,"name")||l(r,"name",e),u(r).source=h.join("string"==typeof e?e:"")),t!==s?(n?!o&&t[e]&&(a=!0):delete t[e],a?t[e]=r:l(t,e,r)):a?t[e]=r:c(e,r)})(Function.prototype,"toString",function(){return"function"==typeof this&&o(this).source||n.call(this)})},function(t,e,r){var i=r(44),n=r(12),a=r(112),o=r(10).f;t.exports=function(t){var e=i.Symbol||(i.Symbol={});n(e,t)||o(e,t,{value:a.f(t)})}},function(t,e,r){var i=r(50),n=r(15);t.exports=function(t){return i(n(t))}},function(t,e,r){var i,n,a,o=r(106),s=r(3),l=r(4),p=r(14),c=r(12),u=r(66),h=r(53),d=s.WeakMap;if(o){var f=new d,m=f.get,g=f.has,v=f.set;i=function(t,e){return v.call(f,t,e),e},n=function(t){return m.call(f,t)||{}},a=function(t){return g.call(f,t)}}else{var y=u("state");h[y]=!0,i=function(t,e){return p(t,y,e),e},n=function(t){return c(t,y)?t[y]:{}},a=function(t){return c(t,y)}}t.exports={set:i,get:n,has:a,enforce:function(t){return a(t)?n(t):i(t,{})},getterFor:function(r){return function(t){var e;if(!l(t)||(e=n(t)).type!==r)throw TypeError("Incompatible receiver, "+r+" required");return e}}}},function(t,e){t.exports=function(t){if("function"!=typeof t)throw TypeError(String(t)+" is not a function");return t}},function(t,e,r){var o=r(15),s=/"/g;t.exports=function(t,e,r,i){var n=String(o(t)),a="<"+e;return""!==r&&(a+=" "+r+'="'+String(i).replace(s,"&quot;")+'"'),a+">"+n+"</"+e+">"}},function(t,e,r){var i=r(2);t.exports=function(e){return i(function(){var t=""[e]('"');return t!==t.toLowerCase()||3<t.split('"').length})}},function(t,e){var r=Math.ceil,i=Math.floor;t.exports=function(t){return isNaN(t=+t)?0:(0<t?i:r)(t)}},function(t,e){var r={}.toString;t.exports=function(t){return r.call(t).slice(8,-1)}},function(t,e,r){var n=r(4);t.exports=function(t,e){if(!n(t))return t;var r,i;if(e&&"function"==typeof(r=t.toString)&&!n(i=r.call(t)))return i;if("function"==typeof(r=t.valueOf)&&!n(i=r.call(t)))return i;if(!e&&"function"==typeof(r=t.toString)&&!n(i=r.call(t)))return i;throw TypeError("Can't convert object to primitive value")}},function(t,e,r){var i=r(10).f,n=r(12),a=r(8)("toStringTag");t.exports=function(t,e,r){t&&!n(t=r?t:t.prototype,a)&&i(t,a,{configurable:!0,value:e})}},function(t,e,r){var i=r(12),n=r(11),a=r(66),o=r(89),s=a("IE_PROTO"),l=Object.prototype;t.exports=o?Object.getPrototypeOf:function(t){return t=n(t),i(t,s)?t[s]:"function"==typeof t.constructor&&t instanceof t.constructor?t.constructor.prototype:t instanceof Object?l:null}},function(t,e,r){"use strict";var i=r(2);t.exports=function(t,e){var r=[][t];return!r||!i(function(){r.call(null,e||function(){throw 1},1)})}},function(t,e,r){var n=r(5),a=r(21),o=r(8)("species");t.exports=function(t,e){var r,i=n(t).constructor;return void 0===i||null==(r=n(i)[o])?e:a(r)}},function(t,e,r){"use strict";function f(t,e){for(var r=0,i=e.length,n=new(X(t))(i);r<i;)n[r]=e[r++];return n}function i(t,e){N(t,e,{get:function(){return F(this)[e]}})}function m(t){var e;return t instanceof $||"ArrayBuffer"==(e=S(t))||"SharedArrayBuffer"==e}function n(t,e){return W(t)&&"symbol"!=typeof e&&e in t&&String(+e)==String(e)}function a(t,e){return n(t,e=d(e,!0))?h(2,t[e]):L(t,e)}function o(t,e,r){return!(n(t,e=d(e,!0))&&T(r)&&w(r,"value"))||w(r,"get")||w(r,"set")||r.configurable||w(r,"writable")&&!r.writable||w(r,"enumerable")&&!r.enumerable?N(t,e,r):(t[e]=r.value,t)}var l=r(1),p=r(3),s=r(7),g=r(102),c=r(6),u=r(80),v=r(38),h=r(39),y=r(14),x=r(9),b=r(137),_=r(138),d=r(26),w=r(12),S=r(62),T=r(4),P=r(35),k=r(45),I=r(40).f,A=r(139),E=r(13).forEach,C=r(46),O=r(10),M=r(16),D=r(20),F=D.get,R=D.set,N=O.f,L=M.f,B=Math.round,j=p.RangeError,$=u.ArrayBuffer,V=u.DataView,H=c.NATIVE_ARRAY_BUFFER_VIEWS,z=c.TYPED_ARRAY_TAG,G=c.TypedArray,Y=c.TypedArrayPrototype,X=c.aTypedArrayConstructor,W=c.isTypedArray,U="BYTES_PER_ELEMENT",q="Wrong length";s?(H||(M.f=a,O.f=o,i(Y,"buffer"),i(Y,"byteOffset"),i(Y,"byteLength"),i(Y,"length")),l({target:"Object",stat:!0,forced:!H},{getOwnPropertyDescriptor:a,defineProperty:o}),t.exports=function(t,c,e,n){function u(t,e){N(t,e,{get:function(){return function(t,e){var r=F(t);return r.view[i](e*c+r.byteOffset,!0)}(this,e)},set:function(t){return function(t,e,r){var i=F(t);n&&(r=(r=B(r))<0?0:255<r?255:255&r),i.view[a](e*c+i.byteOffset,r,!0)}(this,e,t)},enumerable:!0})}var h=t+(n?"Clamped":"")+"Array",i="get"+t,a="set"+t,o=p[h],d=o,r=d&&d.prototype,s={};H?g&&(d=e(function(t,e,r,i){return v(t,d,h),T(e)?m(e)?void 0!==i?new o(e,_(r,c),i):void 0!==r?new o(e,_(r,c)):new o(e):W(e)?f(d,e):A.call(d,e):new o(b(e))}),k&&k(d,G),E(I(o),function(t){t in d||y(d,t,o[t])}),d.prototype=r):(d=e(function(t,e,r,i){v(t,d,h);var n,a,o,s=0,l=0;if(T(e)){if(!m(e))return W(e)?f(d,e):A.call(d,e);n=e,l=_(r,c);var p=e.byteLength;if(void 0===i){if(p%c)throw j(q);if((a=p-l)<0)throw j(q)}else if(p<(a=x(i)*c)+l)throw j(q);o=a/c}else o=b(e),n=new $(a=o*c);for(R(t,{buffer:n,byteOffset:l,byteLength:a,length:o,view:new V(n)});s<o;)u(t,s++)}),k&&k(d,G),r=d.prototype=P(Y)),r.constructor!==d&&y(r,"constructor",d),z&&y(r,z,h),s[h]=d,l({global:!0,forced:d!=o,sham:!H},s),U in d||y(d,U,c),U in r||y(r,U,c),C(h)}):t.exports=function(){}},function(t,e){t.exports=!1},function(t,e,r){function i(t){return"function"==typeof t?t:void 0}var n=r(44),a=r(3);t.exports=function(t,e){return arguments.length<2?i(n[t])||i(a[t]):n[t]&&n[t][e]||a[t]&&a[t][e]}},function(t,e,r){var i=r(24),n=Math.max,a=Math.min;t.exports=function(t,e){var r=i(t);return r<0?n(r+e,0):a(r,e)}},function(t,e,r){function i(){}var n=r(5),a=r(87),o=r(85),s=r(53),l=r(110),p=r(82),c=r(66)("IE_PROTO"),u="prototype",h=function(){var t,e=p("iframe"),r=o.length,i="script";for(e.style.display="none",l.appendChild(e),e.src=String("javascript:"),(t=e.contentWindow.document).open(),t.write("<script>document.F=Object</"+i+">"),t.close(),h=t.F;r--;)delete h[u][o[r]];return h()};t.exports=Object.create||function(t,e){var r;return null!==t?(i[u]=n(t),r=new i,i[u]=null,r[c]=t):r=h(),void 0===e?r:a(r,e)},s[c]=!0},function(t,e,r){var a=r(21);t.exports=function(i,n,t){if(a(i),void 0===n)return i;switch(t){case 0:return function(){return i.call(n)};case 1:return function(t){return i.call(n,t)};case 2:return function(t,e){return i.call(n,t,e)};case 3:return function(t,e,r){return i.call(n,t,e,r)}}return function(){return i.apply(n,arguments)}}},function(t,e,r){var i=r(8),n=r(35),a=r(14),o=i("unscopables"),s=Array.prototype;null==s[o]&&a(s,o,n(null)),t.exports=function(t){s[o][t]=!0}},function(t,e){t.exports=function(t,e,r){if(!(t instanceof e))throw TypeError("Incorrect "+(r?r+" ":"")+"invocation");return t}},function(t,e){t.exports=function(t,e){return{enumerable:!(1&t),configurable:!(2&t),writable:!(4&t),value:e}}},function(t,e,r){var i=r(108),n=r(85).concat("length","prototype");e.f=Object.getOwnPropertyNames||function(t){return i(t,n)}},function(t,e,r){var i=r(25);t.exports=Array.isArray||function(t){return"Array"==i(t)}},function(t,e,r){function i(t){s(t,c,{value:{objectID:"O"+ ++u,weakData:{}}})}var n=r(53),a=r(4),o=r(12),s=r(10).f,l=r(52),p=r(58),c=l("meta"),u=0,h=Object.isExtensible||function(){return!0},d=t.exports={REQUIRED:!1,fastKey:function(t,e){if(!a(t))return"symbol"==typeof t?t:("string"==typeof t?"S":"P")+t;if(!o(t,c)){if(!h(t))return"F";if(!e)return"E";i(t)}return t[c].objectID},getWeakData:function(t,e){if(!o(t,c)){if(!h(t))return!0;if(!e)return!1;i(t)}return t[c].weakData},onFreeze:function(t){return p&&d.REQUIRED&&h(t)&&!o(t,c)&&i(t),t}};n[c]=!0},function(t,e,r){"use strict";var n=r(26),a=r(10),o=r(39);t.exports=function(t,e,r){var i=n(e);i in t?a.f(t,i,o(0,r)):t[i]=r}},function(t,e,r){t.exports=r(3)},function(t,e,r){var n=r(5),a=r(117);t.exports=Object.setPrototypeOf||("__proto__"in{}?function(){var r,i=!1,t={};try{(r=Object.getOwnPropertyDescriptor(Object.prototype,"__proto__").set).call(t,[]),i=t instanceof Array}catch(t){}return function(t,e){return n(t),a(e),i?r.call(t,e):t.__proto__=e,t}}():void 0)},function(t,e,r){"use strict";var i=r(33),n=r(10),a=r(8),o=r(7),s=a("species");t.exports=function(t){var e=i(t),r=n.f;o&&e&&!e[s]&&r(e,s,{configurable:!0,get:function(){return this}})}},function(t,e,r){function i(r){return function(t){var e=String(n(t));return 1&r&&(e=e.replace(o,"")),2&r&&(e=e.replace(s,"")),e}}var n=r(15),a="["+r(77)+"]",o=RegExp("^"+a+a+"*"),s=RegExp(a+a+"*$");t.exports={start:i(1),end:i(2),trim:i(3)}},function(t,e,r){var n=r(17);t.exports=function(t,e,r){for(var i in e)n(t,i,e[i],r);return t}},function(t,e,r){"use strict";r.d(e,"a",function(){return i});var ee=r(0);
/*!
 * VERSION: 2.0.2
 * DATE: 2018-08-27
 * UPDATES AND DOCS AT: http://greensock.com
 *
 * @license Copyright (c) 2008-2018, GreenSock. All rights reserved.
 * This work is subject to the terms at http://greensock.com/standard-license or for
 * Club GreenSock members, the software agreement that was issued with your membership.
 * 
 * @author: Jack Doyle,
 */ee.m._gsDefine("plugins.CSSPlugin",["plugins.TweenPlugin","TweenLite"],function(){var f,T,k,m,$=function(){ee.l.call(this,"css"),this._overwriteProps.length=0,this.setRatio=$.prototype.setRatio},p=ee.m._gsDefine.globals,g={},t=$.prototype=new ee.l("css");(t.constructor=$).version="2.0.2",$.API=2,$.defaultTransformPerspective=0,$.defaultSkewType="compensated",$.defaultSmoothOrigin=!0,t="px",$.suffixMap={top:t,right:t,bottom:t,left:t,width:t,height:t,fontSize:t,padding:t,margin:t,perspective:t,lineHeight:""};function o(t,e){return e.toUpperCase()}function s(t,e){return Z.createElementNS?Z.createElementNS(e||"http://www.w3.org/1999/xhtml",t):Z.createElement(t)}function l(t){return B.test("string"==typeof t?t:(t.currentStyle?t.currentStyle.filter:t.style.filter)||"")?parseFloat(RegExp.$1)/100:1}function v(t){ee.m.console&&console.log(t)}function P(t,e){var r,i,n=(e=e||K).style;if(void 0!==n[t])return t;for(t=t.charAt(0).toUpperCase()+t.substr(1),r=["O","Moz","ms","Ms","Webkit"],i=5;-1<--i&&void 0===n[r[i]+t];);return 0<=i?(nt="-"+(at=3===i?"ms":r[i]).toLowerCase()+"-",at+t):null}function y(t,e){var r,i,n,a={};if(e=e||ot(t,null))if(r=e.length)for(;-1<--r;)-1!==(n=e[r]).indexOf("-transform")&&Nt!==n||(a[n.replace(h,o)]=e.getPropertyValue(n));else for(r in e)-1!==r.indexOf("Transform")&&Rt!==r||(a[r]=e[r]);else if(e=t.currentStyle||t.style)for(r in e)"string"==typeof r&&void 0===a[r]&&(a[r.replace(h,o)]=e[r]);return it||(a.opacity=l(t)),i=Wt(t,e,!1),a.rotation=i.rotation,a.skewX=i.skewX,a.scaleX=i.scaleX,a.scaleY=i.scaleY,a.x=i.x,a.y=i.y,Bt&&(a.z=i.z,a.rotationX=i.rotationX,a.rotationY=i.rotationY,a.scaleZ=i.scaleZ),a.filters&&delete a.filters,a}function x(t,e,r,i,n){var a,o,s,l={},p=t.style;for(o in r)"cssText"!==o&&"length"!==o&&isNaN(o)&&(e[o]!==(a=r[o])||n&&n[o])&&-1===o.indexOf("Origin")&&("number"!=typeof a&&"string"!=typeof a||(l[o]="auto"!==a||"left"!==o&&"top"!==o?""!==a&&"auto"!==a&&"none"!==a||"string"!=typeof e[o]||""===e[o].replace(c,"")?a:0:pt(t,o),void 0!==p[o]&&(s=new xt(p,o,p[o],s))));if(i)for(o in i)"className"!==o&&(l[o]=i[o]);return{difs:l,firstMPT:s}}function b(t,e,r){if("svg"===(t.nodeName+"").toLowerCase())return(r||ot(t))[e]||0;if(t.getCTM&&Gt(t))return t.getBBox()[e]||0;var i=parseFloat("width"===e?t.offsetWidth:t.offsetHeight),n=ct[e],a=n.length;for(r=r||ot(t,null);-1<--a;)i-=parseFloat(st(t,"padding"+n[a],r,!0))||0,i-=parseFloat(st(t,"border"+n[a]+"Width",r,!0))||0;return i}function I(t,e){return"function"==typeof t&&(t=t(D,M)),"string"==typeof t&&"="===t.charAt(1)?parseInt(t.charAt(0)+"1",10)*parseFloat(t.substr(2)):parseFloat(t)-parseFloat(e)||0}function A(t,e){"function"==typeof t&&(t=t(D,M));var r="string"==typeof t&&"="===t.charAt(1);return"string"==typeof t&&"v"===t.charAt(t.length-2)&&(t=(r?t.substr(0,2):0)+window["inner"+("vh"===t.substr(-2)?"Height":"Width")]*(parseFloat(r?t.substr(2):t)/100)),null==t?e:r?parseInt(t.charAt(0)+"1",10)*parseFloat(t.substr(2))+e:parseFloat(t)||0}function E(t,e,r,i){var n,a,o,s,l;return"function"==typeof t&&(t=t(D,M)),(s=null==t?e:"number"==typeof t?t:(n=360,a=t.split("_"),o=((l="="===t.charAt(1))?parseInt(t.charAt(0)+"1",10)*parseFloat(a[0].substr(2)):parseFloat(a[0]))*(-1===t.indexOf("rad")?1:J)-(l?0:e),a.length&&(i&&(i[r]=e+o),-1!==t.indexOf("short")&&(o%=n)!==o%180&&(o=o<0?o+n:o-n),-1!==t.indexOf("_cw")&&o<0?o=(o+3599999999640)%n-(o/n|0)*n:-1!==t.indexOf("ccw")&&0<o&&(o=(o-3599999999640)%n-(o/n|0)*n)),e+o))<1e-6&&-1e-6<s&&(s=0),s}function d(t,e,r){return 255*(6*(t=t<0?t+1:1<t?t-1:t)<1?e+(r-e)*t*6:t<.5?r:3*t<2?e+(r-e)*(2/3-t)*6:e)+.5|0}function i(t,e){var r,i,n,a=t.match(mt)||[],o=0,s="";if(!a.length)return t;for(r=0;r<a.length;r++)i=a[r],o+=(n=t.substr(o,t.indexOf(i,o)-o)).length+i.length,3===(i=ft(i,e)).length&&i.push(1),s+=n+(e?"hsla("+i[0]+","+i[1]+"%,"+i[2]+"%,"+i[3]:"rgba("+i.join(","))+")";return s+t.substr(o)}var C,_,w,V,S,O,M,D,e,r,F=/(?:\-|\.|\b)(\d|\.|e\-)+/g,R=/(?:\d|\-\d|\.\d|\-\.\d|\+=\d|\-=\d|\+=.\d|\-=\.\d)+/g,N=/(?:\+=|\-=|\-|\b)[\d\-\.]+[a-zA-Z0-9]*(?:%|\b)/gi,c=/(?![+-]?\d*\.?\d+|[+-]|e[+-]\d+)[^0-9]/g,L=/(?:\d|\-|\+|=|#|\.)*/g,B=/opacity *= *([^)]*)/i,j=/opacity:([^;]*)/i,a=/alpha\(opacity *=.+?\)/i,H=/^(rgb|hsl)/,u=/([A-Z])/g,h=/-([a-z])/gi,z=/(^(?:url\(\"|url\())|(?:(\"\))$|\)$)/gi,G=/(?:Left|Right|Width)/i,Y=/(M11|M12|M21|M22)=[\d\-\.e]+/gi,X=/progid\:DXImageTransform\.Microsoft\.Matrix\(.+?\)/i,W=/,(?=[^\)]*(?:\(|$))/gi,U=/[\s,\(]/i,q=Math.PI/180,J=180/Math.PI,Q={},n={style:{}},Z=ee.m.document||{createElement:function(){return n}},K=s("div"),tt=s("img"),et=$._internals={_specialProps:g},rt=(ee.m.navigator||{}).userAgent||"",it=(e=rt.indexOf("Android"),r=s("a"),w=-1!==rt.indexOf("Safari")&&-1===rt.indexOf("Chrome")&&(-1===e||3<parseFloat(rt.substr(e+8,2))),S=w&&parseFloat(rt.substr(rt.indexOf("Version/")+8,2))<6,V=-1!==rt.indexOf("Firefox"),(/MSIE ([0-9]{1,}[\.0-9]{0,})/.exec(rt)||/Trident\/.*rv:([0-9]{1,}[\.0-9]{0,})/.exec(rt))&&(O=parseFloat(RegExp.$1)),!!r&&(r.style.cssText="top:1px;opacity:.55;",/^0.55/.test(r.style.opacity))),nt="",at="",ot=("undefined"!=typeof window?window:Z.defaultView||{getComputedStyle:function(){}}).getComputedStyle,st=$.getStyle=function(t,e,r,i,n){var a;return it||"opacity"!==e?(!i&&t.style[e]?a=t.style[e]:(r=r||ot(t))?a=r[e]||r.getPropertyValue(e)||r.getPropertyValue(e.replace(u,"-$1").toLowerCase()):t.currentStyle&&(a=t.currentStyle[e]),null==n||a&&"none"!==a&&"auto"!==a&&"auto auto"!==a?a:n):l(t)},lt=et.convertToPixels=function(t,e,r,i,n){if("px"===i||!i&&"lineHeight"!==e)return r;if("auto"===i||!r)return 0;var a,o,s,l=G.test(e),p=t,c=K.style,u=r<0,h=1===r;if(u&&(r=-r),h&&(r*=100),"lineHeight"!==e||i)if("%"===i&&-1!==e.indexOf("border"))a=r/100*(l?t.clientWidth:t.clientHeight);else{if(c.cssText="border:0 solid red;position:"+st(t,"position")+";line-height:0;","%"!==i&&p.appendChild&&"v"!==i.charAt(0)&&"rem"!==i)c[l?"borderLeftWidth":"borderTopWidth"]=r+i;else{if(p=t.parentNode||Z.body,-1!==st(p,"display").indexOf("flex")&&(c.position="absolute"),o=p._gsCache,s=ee.n.ticker.frame,o&&l&&o.time===s)return o.width*r/100;c[l?"width":"height"]=r+i}p.appendChild(K),a=parseFloat(K[l?"offsetWidth":"offsetHeight"]),p.removeChild(K),l&&"%"===i&&!1!==$.cacheWidths&&((o=p._gsCache=p._gsCache||{}).time=s,o.width=a/r*100),0!==a||n||(a=lt(t,e,r,i,!0))}else o=ot(t).lineHeight,t.style.lineHeight=r,a=parseFloat(ot(t).lineHeight),t.style.lineHeight=o;return h&&(a/=100),u?-a:a},pt=et.calculateOffset=function(t,e,r){if("absolute"!==st(t,"position",r))return 0;var i="left"===e?"Left":"Top",n=st(t,"margin"+i,r);return t["offset"+i]-(lt(t,e,parseFloat(n),n.replace(L,""))||0)},ct={width:["Left","Right"],height:["Top","Bottom"]},ut=["marginLeft","marginRight","marginTop","marginBottom"],ht=function(t,e){if("contain"===t||"auto"===t||"auto auto"===t)return t+" ";null!=t&&""!==t||(t="0 0");var r,i=t.split(" "),n=-1!==t.indexOf("left")?"0%":-1!==t.indexOf("right")?"100%":i[0],a=-1!==t.indexOf("top")?"0%":-1!==t.indexOf("bottom")?"100%":i[1];if(3<i.length&&!e){for(i=t.split(", ").join(",").split(","),t=[],r=0;r<i.length;r++)t.push(ht(i[r]));return t.join(",")}return null==a?a="center"===n?"50%":"0":"center"===a&&(a="50%"),("center"===n||isNaN(parseFloat(n))&&-1===(n+"").indexOf("="))&&(n="50%"),t=n+" "+a+(2<i.length?" "+i[2]:""),e&&(e.oxp=-1!==n.indexOf("%"),e.oyp=-1!==a.indexOf("%"),e.oxr="="===n.charAt(1),e.oyr="="===a.charAt(1),e.ox=parseFloat(n.replace(c,"")),e.oy=parseFloat(a.replace(c,"")),e.v=t),e||t},dt={aqua:[0,255,255],lime:[0,255,0],silver:[192,192,192],black:[0,0,0],maroon:[128,0,0],teal:[0,128,128],blue:[0,0,255],navy:[0,0,128],white:[255,255,255],fuchsia:[255,0,255],olive:[128,128,0],yellow:[255,255,0],orange:[255,165,0],gray:[128,128,128],purple:[128,0,128],green:[0,128,0],red:[255,0,0],pink:[255,192,203],cyan:[0,255,255],transparent:[255,255,255,0]},ft=$.parseColor=function(t,e){var r,i,n,a,o,s,l,p,c,u,h;if(t)if("number"==typeof t)r=[t>>16,t>>8&255,255&t];else{if(","===t.charAt(t.length-1)&&(t=t.substr(0,t.length-1)),dt[t])r=dt[t];else if("#"===t.charAt(0))4===t.length&&(t="#"+(i=t.charAt(1))+i+(n=t.charAt(2))+n+(a=t.charAt(3))+a),r=[(t=parseInt(t.substr(1),16))>>16,t>>8&255,255&t];else if("hsl"===t.substr(0,3))if(r=h=t.match(F),e){if(-1!==t.indexOf("="))return t.match(R)}else o=Number(r[0])%360/360,s=Number(r[1])/100,i=2*(l=Number(r[2])/100)-(n=l<=.5?l*(s+1):l+s-l*s),3<r.length&&(r[3]=Number(r[3])),r[0]=d(o+1/3,i,n),r[1]=d(o,i,n),r[2]=d(o-1/3,i,n);else r=t.match(F)||dt.transparent;r[0]=Number(r[0]),r[1]=Number(r[1]),r[2]=Number(r[2]),3<r.length&&(r[3]=Number(r[3]))}else r=dt.black;return e&&!h&&(i=r[0]/255,n=r[1]/255,a=r[2]/255,l=((p=Math.max(i,n,a))+(c=Math.min(i,n,a)))/2,p===c?o=s=0:(u=p-c,s=.5<l?u/(2-p-c):u/(p+c),o=p===i?(n-a)/u+(n<a?6:0):p===n?(a-i)/u+2:(i-n)/u+4,o*=60),r[0]=o+.5|0,r[1]=100*s+.5|0,r[2]=100*l+.5|0),r},mt="(?:\\b(?:(?:rgb|rgba|hsl|hsla)\\(.+?\\))|\\B#(?:[0-9a-f]{3}){1,2}\\b";for(t in dt)mt+="|"+t+"\\b";mt=new RegExp(mt+")","gi"),$.colorStringFilter=function(t){var e,r=t[0]+" "+t[1];mt.test(r)&&(e=-1!==r.indexOf("hsl(")||-1!==r.indexOf("hsla("),t[0]=i(t[0],e),t[1]=i(t[1],e)),mt.lastIndex=0},ee.n.defaultStringFilter||(ee.n.defaultStringFilter=$.colorStringFilter);function gt(t,e,a,o){if(null==t)return function(t){return t};var s,l=e?(t.match(mt)||[""])[0]:"",p=t.split(l).join("").match(N)||[],c=t.substr(0,t.indexOf(p[0])),u=")"===t.charAt(t.length-1)?")":"",h=-1!==t.indexOf(" ")?" ":",",d=p.length,f=0<d?p[0].replace(F,""):"";return d?s=e?function(t){var e,r,i,n;if("number"==typeof t)t+=f;else if(o&&W.test(t)){for(n=t.replace(W,"|").split("|"),i=0;i<n.length;i++)n[i]=s(n[i]);return n.join(",")}if(e=(t.match(mt)||[l])[0],i=(r=t.split(e).join("").match(N)||[]).length,d>i--)for(;++i<d;)r[i]=a?r[(i-1)/2|0]:p[i];return c+r.join(h)+h+e+u+(-1!==t.indexOf("inset")?" inset":"")}:function(t){var e,r,i;if("number"==typeof t)t+=f;else if(o&&W.test(t)){for(r=t.replace(W,"|").split("|"),i=0;i<r.length;i++)r[i]=s(r[i]);return r.join(",")}if(i=(e=t.match(N)||[]).length,d>i--)for(;++i<d;)e[i]=a?e[(i-1)/2|0]:p[i];return c+e.join(h)+u}:function(t){return t}}function vt(p){return p=p.split(","),function(t,e,r,i,n,a,o){var s,l=(e+"").split(" ");for(o={},s=0;s<4;s++)o[p[s]]=l[s]=l[s]||l[(s-1)/2>>0];return i.parse(t,o,n,a)}}et._setPluginRatio=function(t){this.plugin.setRatio(t);for(var e,r,i,n,a,o=this.data,s=o.proxy,l=o.firstMPT;l;)e=s[l.v],l.r?e=l.r(e):e<1e-6&&-1e-6<e&&(e=0),l.t[l.p]=e,l=l._next;if(o.autoRotate&&(o.autoRotate.rotation=o.mod?o.mod.call(this._tween,s.rotation,this.t,this._tween):s.rotation),1===t||0===t)for(l=o.firstMPT,a=1===t?"e":"b";l;){if((r=l.t).type){if(1===r.type){for(n=r.xs0+r.s+r.xs1,i=1;i<r.l;i++)n+=r["xn"+i]+r["xs"+(i+1)];r[a]=n}}else r[a]=r.s+r.xs0;l=l._next}};function yt(t,e,r,i,n,a){var o=new bt(t,e,r,i-r,n,-1,a);return o.b=r,o.e=o.xs0=i,o}var xt=function(t,e,r,i,n){this.t=t,this.p=e,this.v=r,this.r=n,i&&((i._prev=this)._next=i)},bt=(et._parseToProxy=function(t,e,r,i,n,a){var o,s,l,p,c,u=i,h={},d={},f=r._transform,m=Q;for(r._transform=null,Q=e,i=c=r.parse(t,e,i,n),Q=m,a&&(r._transform=f,u&&(u._prev=null,u._prev&&(u._prev._next=null)));i&&i!==u;){if(i.type<=1&&(d[s=i.p]=i.s+i.c,h[s]=i.s,a||(p=new xt(i,"s",s,p,i.r),i.c=0),1===i.type))for(o=i.l;0<--o;)l="xn"+o,d[s=i.p+"_"+l]=i.data[l],h[s]=i[l],a||(p=new xt(i,l,s,p,i.rxp[l]));i=i._next}return{proxy:h,end:d,firstMPT:p,pt:c}},et.CSSPropTween=function(t,e,r,i,n,a,o,s,l,p,c){this.t=t,this.p=e,this.s=r,this.c=i,this.n=o||e,t instanceof bt||m.push(this.n),this.r=s?"function"==typeof s?s:Math.round:s,this.type=a||0,l&&(this.pr=l,f=!0),this.b=void 0===p?r:p,this.e=void 0===c?r+i:c,n&&((this._next=n)._prev=this)}),_t=$.parseComplex=function(t,e,r,i,n,a,o,s,l,p){r=r||a||"","function"==typeof i&&(i=i(D,M)),o=new bt(t,e,0,0,o,p?2:1,null,!1,s,r,i),i+="",n&&mt.test(i+r)&&(i=[r,i],$.colorStringFilter(i),r=i[0],i=i[1]);var c,u,h,d,f,m,g,v,y,x,b,_,w,S=r.split(", ").join(",").split(" "),T=i.split(", ").join(",").split(" "),P=S.length,k=!1!==C;for(-1===i.indexOf(",")&&-1===r.indexOf(",")||(T=-1!==(i+r).indexOf("rgb")||-1!==(i+r).indexOf("hsl")?(S=S.join(" ").replace(W,", ").split(" "),T.join(" ").replace(W,", ").split(" ")):(S=S.join(" ").split(",").join(", ").split(" "),T.join(" ").split(",").join(", ").split(" ")),P=S.length),P!==T.length&&(P=(S=(a||"").split(" ")).length),o.plugin=l,o.setRatio=p,c=mt.lastIndex=0;c<P;c++)if(d=S[c],f=T[c]+"",(v=parseFloat(d))||0===v)o.appendXtra("",v,I(f,v),f.replace(R,""),k&&-1!==f.indexOf("px")&&Math.round,!0);else if(n&&mt.test(d))_=")"+((_=f.indexOf(")")+1)?f.substr(_):""),w=-1!==f.indexOf("hsl")&&it,x=f,d=ft(d,w),f=ft(f,w),(y=6<d.length+f.length)&&!it&&0===f[3]?(o["xs"+o.l]+=o.l?" transparent":"transparent",o.e=o.e.split(T[c]).join("transparent")):(it||(y=!1),w?o.appendXtra(x.substr(0,x.indexOf("hsl"))+(y?"hsla(":"hsl("),d[0],I(f[0],d[0]),",",!1,!0).appendXtra("",d[1],I(f[1],d[1]),"%,",!1).appendXtra("",d[2],I(f[2],d[2]),y?"%,":"%"+_,!1):o.appendXtra(x.substr(0,x.indexOf("rgb"))+(y?"rgba(":"rgb("),d[0],f[0]-d[0],",",Math.round,!0).appendXtra("",d[1],f[1]-d[1],",",Math.round).appendXtra("",d[2],f[2]-d[2],y?",":_,Math.round),y&&(d=d.length<4?1:d[3],o.appendXtra("",d,(f.length<4?1:f[3])-d,_,!1))),mt.lastIndex=0;else if(m=d.match(F)){if(!(g=f.match(R))||g.length!==m.length)return o;for(u=h=0;u<m.length;u++)b=m[u],x=d.indexOf(b,h),o.appendXtra(d.substr(h,x-h),Number(b),I(g[u],b),"",k&&"px"===d.substr(x+b.length,2)&&Math.round,0===u),h=x+b.length;o["xs"+o.l]+=d.substr(h)}else o["xs"+o.l]+=o.l||o["xs"+o.l]?" "+f:f;if(-1!==i.indexOf("=")&&o.data){for(_=o.xs0+o.data.s,c=1;c<o.l;c++)_+=o["xs"+c]+o.data["xn"+c];o.e=_+o["xs"+c]}return o.l||(o.type=-1,o.xs0=o.e),o.xfirst||o},wt=9;for((t=bt.prototype).l=t.pr=0;0<--wt;)t["xn"+wt]=0,t["xs"+wt]="";t.xs0="",t._next=t._prev=t.xfirst=t.data=t.plugin=t.setRatio=t.rxp=null,t.appendXtra=function(t,e,r,i,n,a){var o=this,s=o.l;return o["xs"+s]+=a&&(s||o["xs"+s])?" "+t:t||"",r||0===s||o.plugin?(o.l++,o.type=o.setRatio?2:1,o["xs"+o.l]=i||"",0<s?(o.data["xn"+s]=e+r,o.rxp["xn"+s]=n,o["xn"+s]=e,o.plugin||(o.xfirst=new bt(o,"xn"+s,e,r,o.xfirst||o,0,o.n,n,o.pr),o.xfirst.xs0=0)):(o.data={s:e+r},o.rxp={},o.s=e,o.c=r,o.r=n),o):(o["xs"+s]+=e+(i||""),o)};function St(t,e){e=e||{},this.p=e.prefix&&P(t)||t,g[t]=g[this.p]=this,this.format=e.formatter||gt(e.defaultValue,e.color,e.collapsible,e.multi),e.parser&&(this.parse=e.parser),this.clrs=e.color,this.multi=e.multi,this.keyword=e.keyword,this.dflt=e.defaultValue,this.pr=e.priority||0}var Tt=et._registerComplexSpecialProp=function(t,e,r){"object"!=typeof e&&(e={parser:r});var i,n=t.split(","),a=e.defaultValue;for(r=r||[a],i=0;i<n.length;i++)e.prefix=0===i&&e.prefix,e.defaultValue=r[i]||a,new St(n[i],e)},Pt=et._registerPluginProp=function(t){if(!g[t]){var l=t.charAt(0).toUpperCase()+t.substr(1)+"Plugin";Tt(t,{parser:function(t,e,r,i,n,a,o){var s=p.com.greensock.plugins[l];return s?(s._cssRegister(),g[r].parse(t,e,r,i,n,a,o)):(v("Error: "+l+" js file not loaded."),n)}})}};(t=St.prototype).parseComplex=function(t,e,r,i,n,a){var o,s,l,p,c,u,h=this.keyword;if(this.multi&&(W.test(r)||W.test(e)?(s=e.replace(W,"|").split("|"),l=r.replace(W,"|").split("|")):h&&(s=[e],l=[r])),l){for(p=l.length>s.length?l.length:s.length,o=0;o<p;o++)e=s[o]=s[o]||this.dflt,r=l[o]=l[o]||this.dflt,h&&(c=e.indexOf(h))!==(u=r.indexOf(h))&&(-1===u?s[o]=s[o].split(h).join(""):-1===c&&(s[o]+=" "+h));e=s.join(", "),r=l.join(", ")}return _t(t,this.p,e,r,this.clrs,this.dflt,i,this.pr,n,a)},t.parse=function(t,e,r,i,n,a,o){return this.parseComplex(t.style,this.format(st(t,this.p,k,!1,this.dflt)),this.format(e),n,a)},$.registerSpecialProp=function(t,l,p){Tt(t,{parser:function(t,e,r,i,n,a,o){var s=new bt(t,r,0,0,n,2,r,!1,p);return s.plugin=a,s.setRatio=l(t,e,i._tween,r),s},priority:p})},$.useSVGTransformAttr=!0;function kt(t,e,r){var i,n=Z.createElementNS("http://www.w3.org/2000/svg",t),a=/([a-z])([A-Z])/g;for(i in r)n.setAttributeNS(null,i.replace(a,"$1-$2").toLowerCase(),r[i]);return e.appendChild(n),n}function It(t,e,r,i,n,a){var o,s,l,p,c,u,h,d,f,m,g,v,y,x,b=t._gsTransform,_=Xt(t,!0);b&&(y=b.xOrigin,x=b.yOrigin),(!i||(o=i.split(" ")).length<2)&&(0===(h=t.getBBox()).x&&0===h.y&&h.width+h.height===0&&(h={x:parseFloat(t.hasAttribute("x")?t.getAttribute("x"):t.hasAttribute("cx")?t.getAttribute("cx"):0)||0,y:parseFloat(t.hasAttribute("y")?t.getAttribute("y"):t.hasAttribute("cy")?t.getAttribute("cy"):0)||0,width:0,height:0}),o=[(-1!==(e=ht(e).split(" "))[0].indexOf("%")?parseFloat(e[0])/100*h.width:parseFloat(e[0]))+h.x,(-1!==e[1].indexOf("%")?parseFloat(e[1])/100*h.height:parseFloat(e[1]))+h.y]),r.xOrigin=p=parseFloat(o[0]),r.yOrigin=c=parseFloat(o[1]),i&&_!==Yt&&(u=_[0],h=_[1],d=_[2],f=_[3],m=_[4],g=_[5],(v=u*f-h*d)&&(s=p*(f/v)+c*(-d/v)+(d*g-f*m)/v,l=p*(-h/v)+c*(u/v)-(u*g-h*m)/v,p=r.xOrigin=o[0]=s,c=r.yOrigin=o[1]=l)),b&&(a&&(r.xOffset=b.xOffset,r.yOffset=b.yOffset,b=r),n||!1!==n&&!1!==$.defaultSmoothOrigin?(s=p-y,l=c-x,b.xOffset+=s*_[0]+l*_[2]-s,b.yOffset+=s*_[1]+l*_[3]-l):b.xOffset=b.yOffset=0),a||t.setAttribute("data-svg-origin",o.join(" "))}function At(t){var e,r,i=this.data,n=-i.rotation*q,a=n+i.skewX*q,o=1e5,s=(Math.cos(n)*i.scaleX*o|0)/o,l=(Math.sin(n)*i.scaleX*o|0)/o,p=(Math.sin(a)*-i.scaleY*o|0)/o,c=(Math.cos(a)*i.scaleY*o|0)/o,u=this.t.style,h=this.t.currentStyle;if(h){r=l,l=-p,p=-r,e=h.filter,u.filter="";var d,f,m=this.t.offsetWidth,g=this.t.offsetHeight,v="absolute"!==h.position,y="progid:DXImageTransform.Microsoft.Matrix(M11="+s+", M12="+l+", M21="+p+", M22="+c,x=i.x+m*i.xPercent/100,b=i.y+g*i.yPercent/100;if(null!=i.ox&&(x+=(d=(i.oxp?m*i.ox*.01:i.ox)-m/2)-(d*s+(f=(i.oyp?g*i.oy*.01:i.oy)-g/2)*l),b+=f-(d*p+f*c)),y+=v?", Dx="+((d=m/2)-(d*s+(f=g/2)*l)+x)+", Dy="+(f-(d*p+f*c)+b)+")":", sizingMethod='auto expand')",-1!==e.indexOf("DXImageTransform.Microsoft.Matrix(")?u.filter=e.replace(X,y):u.filter=y+" "+e,0!==t&&1!==t||1==s&&0===l&&0===p&&1==c&&(v&&-1===y.indexOf("Dx=0, Dy=0")||B.test(e)&&100!==parseFloat(RegExp.$1)||-1===e.indexOf(e.indexOf("Alpha"))&&u.removeAttribute("filter")),!v){var _,w,S,T=O<8?1:-1;for(d=i.ieOffsetX||0,f=i.ieOffsetY||0,i.ieOffsetX=Math.round((m-((s<0?-s:s)*m+(l<0?-l:l)*g))/2+x),i.ieOffsetY=Math.round((g-((c<0?-c:c)*g+(p<0?-p:p)*m))/2+b),wt=0;wt<4;wt++)S=(r=-1!==(_=h[w=ut[wt]]).indexOf("px")?parseFloat(_):lt(this.t,w,parseFloat(_),_.replace(L,""))||0)!==i[w]?wt<2?-i.ieOffsetX:-i.ieOffsetY:wt<2?d-i.ieOffsetX:f-i.ieOffsetY,u[w]=(i[w]=Math.round(r-S*(0===wt||2===wt?1:T)))+"px"}}}var Et,Ct,Ot,Mt,Dt,Ft="scaleX,scaleY,scaleZ,x,y,z,skewX,skewY,rotation,rotationX,rotationY,perspective,xPercent,yPercent".split(","),Rt=P("transform"),Nt=nt+"transform",Lt=P("transformOrigin"),Bt=null!==P("perspective"),jt=et.Transform=function(){this.perspective=parseFloat($.defaultTransformPerspective)||0,this.force3D=!(!1===$.defaultForce3D||!Bt)&&($.defaultForce3D||"auto")},$t=ee.m.SVGElement,Vt=Z.documentElement||{},Ht=(Dt=O||/Android/i.test(rt)&&!ee.m.chrome,Z.createElementNS&&!Dt&&(Ct=kt("svg",Vt),Mt=(Ot=kt("rect",Ct,{width:100,height:50,x:100})).getBoundingClientRect().width,Ot.style[Lt]="50% 50%",Ot.style[Rt]="scaleX(0.5)",Dt=Mt===Ot.getBoundingClientRect().width&&!(V&&Bt),Vt.removeChild(Ct)),Dt),zt=function(t){var e,r=s("svg",this.ownerSVGElement&&this.ownerSVGElement.getAttribute("xmlns")||"http://www.w3.org/2000/svg"),i=this.parentNode,n=this.nextSibling,a=this.style.cssText;if(Vt.appendChild(r),r.appendChild(this),this.style.display="block",t)try{e=this.getBBox(),this._originalGetBBox=this.getBBox,this.getBBox=zt}catch(t){}else this._originalGetBBox&&(e=this._originalGetBBox());return n?i.insertBefore(this,n):i.appendChild(this),Vt.removeChild(r),this.style.cssText=a,e},Gt=function(t){return!(!$t||!t.getCTM||t.parentNode&&!t.ownerSVGElement||!function(e){try{return e.getBBox()}catch(t){return zt.call(e,!0)}}(t))},Yt=[1,0,0,1,0,0],Xt=function(t,e){var r,i,n,a,o,s,l=t._gsTransform||new jt,p=t.style;if(Rt?i=st(t,Nt,null,!0):t.currentStyle&&(i=(i=t.currentStyle.filter.match(Y))&&4===i.length?[i[0].substr(4),Number(i[2].substr(4)),Number(i[1].substr(4)),i[3].substr(4),l.x||0,l.y||0].join(","):""),r=!i||"none"===i||"matrix(1, 0, 0, 1, 0, 0)"===i,!Rt||!(s=!ot(t)||"none"===ot(t).display)&&t.parentNode||(s&&(a=p.display,p.display="block"),t.parentNode||(o=1,Vt.appendChild(t)),r=!(i=st(t,Nt,null,!0))||"none"===i||"matrix(1, 0, 0, 1, 0, 0)"===i,a?p.display=a:s&&Qt(p,"display"),o&&Vt.removeChild(t)),(l.svg||t.getCTM&&Gt(t))&&(r&&-1!==(p[Rt]+"").indexOf("matrix")&&(i=p[Rt],r=0),n=t.getAttribute("transform"),r&&n&&(i="matrix("+(n=t.transform.baseVal.consolidate().matrix).a+","+n.b+","+n.c+","+n.d+","+n.e+","+n.f+")",r=0)),r)return Yt;for(n=(i||"").match(F)||[],wt=n.length;-1<--wt;)a=Number(n[wt]),n[wt]=(o=a-(a|=0))?(1e5*o+(o<0?-.5:.5)|0)/1e5+a:a;return e&&6<n.length?[n[0],n[1],n[4],n[5],n[12],n[13]]:n},Wt=et.getTransform=function(t,e,r,i){if(t._gsTransform&&r&&!i)return t._gsTransform;var n,a,o,s,l,p,c=r&&t._gsTransform||new jt,u=c.scaleX<0,h=Bt&&(parseFloat(st(t,Lt,e,!1,"0 0 0").split(" ")[2])||c.zOrigin)||0,d=parseFloat($.defaultTransformPerspective)||0;if(c.svg=!(!t.getCTM||!Gt(t)),c.svg&&(It(t,st(t,Lt,e,!1,"50% 50%")+"",c,t.getAttribute("data-svg-origin")),Et=$.useSVGTransformAttr||Ht),(n=Xt(t))!==Yt){if(16===n.length){var f,m,g,v,y,x=n[0],b=n[1],_=n[2],w=n[3],S=n[4],T=n[5],P=n[6],k=n[7],I=n[8],A=n[9],E=n[10],C=n[12],O=n[13],M=n[14],D=n[11],F=Math.atan2(P,E);c.zOrigin&&(C=I*(M=-c.zOrigin)-n[12],O=A*M-n[13],M=E*M+c.zOrigin-n[14]),c.rotationX=F*J,F&&(f=S*(v=Math.cos(-F))+I*(y=Math.sin(-F)),m=T*v+A*y,g=P*v+E*y,I=S*-y+I*v,A=T*-y+A*v,E=P*-y+E*v,D=k*-y+D*v,S=f,T=m,P=g),F=Math.atan2(-_,E),c.rotationY=F*J,F&&(m=b*(v=Math.cos(-F))-A*(y=Math.sin(-F)),g=_*v-E*y,A=b*y+A*v,E=_*y+E*v,D=w*y+D*v,x=f=x*v-I*y,b=m,_=g),F=Math.atan2(b,x),c.rotation=F*J,F&&(f=x*(v=Math.cos(F))+b*(y=Math.sin(F)),m=S*v+T*y,g=I*v+A*y,b=b*v-x*y,T=T*v-S*y,A=A*v-I*y,x=f,S=m,I=g),c.rotationX&&359.9<Math.abs(c.rotationX)+Math.abs(c.rotation)&&(c.rotationX=c.rotation=0,c.rotationY=180-c.rotationY),F=Math.atan2(S,T),c.scaleX=(1e5*Math.sqrt(x*x+b*b+_*_)+.5|0)/1e5,c.scaleY=(1e5*Math.sqrt(T*T+P*P)+.5|0)/1e5,c.scaleZ=(1e5*Math.sqrt(I*I+A*A+E*E)+.5|0)/1e5,x/=c.scaleX,S/=c.scaleY,b/=c.scaleX,T/=c.scaleY,2e-5<Math.abs(F)?(c.skewX=F*J,S=0,"simple"!==c.skewType&&(c.scaleY*=1/Math.cos(F))):c.skewX=0,c.perspective=D?1/(D<0?-D:D):0,c.x=C,c.y=O,c.z=M,c.svg&&(c.x-=c.xOrigin-(c.xOrigin*x-c.yOrigin*S),c.y-=c.yOrigin-(c.yOrigin*b-c.xOrigin*T))}else if(!Bt||i||!n.length||c.x!==n[4]||c.y!==n[5]||!c.rotationX&&!c.rotationY){var R=6<=n.length,N=R?n[0]:1,L=n[1]||0,B=n[2]||0,j=R?n[3]:1;c.x=n[4]||0,c.y=n[5]||0,o=Math.sqrt(N*N+L*L),s=Math.sqrt(j*j+B*B),l=N||L?Math.atan2(L,N)*J:c.rotation||0,p=B||j?Math.atan2(B,j)*J+l:c.skewX||0,c.scaleX=o,c.scaleY=s,c.rotation=l,c.skewX=p,Bt&&(c.rotationX=c.rotationY=c.z=0,c.perspective=d,c.scaleZ=1),c.svg&&(c.x-=c.xOrigin-(c.xOrigin*N+c.yOrigin*B),c.y-=c.yOrigin-(c.xOrigin*L+c.yOrigin*j))}for(a in 90<Math.abs(c.skewX)&&Math.abs(c.skewX)<270&&(u?(c.scaleX*=-1,c.skewX+=c.rotation<=0?180:-180,c.rotation+=c.rotation<=0?180:-180):(c.scaleY*=-1,c.skewX+=c.skewX<=0?180:-180)),c.zOrigin=h,c)c[a]<2e-5&&-2e-5<c[a]&&(c[a]=0)}return r&&(t._gsTransform=c).svg&&(Et&&t.style[Rt]?ee.n.delayedCall(.001,function(){Qt(t.style,Rt)}):!Et&&t.getAttribute("transform")&&ee.n.delayedCall(.001,function(){t.removeAttribute("transform")})),c},Ut=et.set3DTransformRatio=et.setTransformRatio=function(t){var e,r,i,n,a,o,s,l,p,c,u,h,d,f,m,g,v,y,x,b,_,w,S,T=this.data,P=this.t.style,k=T.rotation,I=T.rotationX,A=T.rotationY,E=T.scaleX,C=T.scaleY,O=T.scaleZ,M=T.x,D=T.y,F=T.z,R=T.svg,N=T.perspective,L=T.force3D,B=T.skewY,j=T.skewX;if(B&&(j+=B,k+=B),!((1!==t&&0!==t||"auto"!==L||this.tween._totalTime!==this.tween._totalDuration&&this.tween._totalTime)&&L||F||N||A||I||1!==O)||Et&&R||!Bt)k||j||R?(k*=q,w=j*q,S=1e5,r=Math.cos(k)*E,a=Math.sin(k)*E,i=Math.sin(k-w)*-C,o=Math.cos(k-w)*C,w&&"simple"===T.skewType&&(e=Math.tan(w-B*q),i*=e=Math.sqrt(1+e*e),o*=e,B&&(e=Math.tan(B*q),r*=e=Math.sqrt(1+e*e),a*=e)),R&&(M+=T.xOrigin-(T.xOrigin*r+T.yOrigin*i)+T.xOffset,D+=T.yOrigin-(T.xOrigin*a+T.yOrigin*o)+T.yOffset,Et&&(T.xPercent||T.yPercent)&&(m=this.t.getBBox(),M+=.01*T.xPercent*m.width,D+=.01*T.yPercent*m.height),M<(m=1e-6)&&-m<M&&(M=0),D<m&&-m<D&&(D=0)),x=(r*S|0)/S+","+(a*S|0)/S+","+(i*S|0)/S+","+(o*S|0)/S+","+M+","+D+")",R&&Et?this.t.setAttribute("transform","matrix("+x):P[Rt]=(T.xPercent||T.yPercent?"translate("+T.xPercent+"%,"+T.yPercent+"%) matrix(":"matrix(")+x):P[Rt]=(T.xPercent||T.yPercent?"translate("+T.xPercent+"%,"+T.yPercent+"%) matrix(":"matrix(")+E+",0,0,"+C+","+M+","+D+")";else{if(V&&(E<(m=1e-4)&&-m<E&&(E=O=2e-5),C<m&&-m<C&&(C=O=2e-5),!N||T.z||T.rotationX||T.rotationY||(N=0)),k||j)k*=q,g=r=Math.cos(k),v=a=Math.sin(k),j&&(k-=j*q,g=Math.cos(k),v=Math.sin(k),"simple"===T.skewType&&(e=Math.tan((j-B)*q),g*=e=Math.sqrt(1+e*e),v*=e,T.skewY&&(e=Math.tan(B*q),r*=e=Math.sqrt(1+e*e),a*=e))),i=-v,o=g;else{if(!(A||I||1!==O||N||R))return void(P[Rt]=(T.xPercent||T.yPercent?"translate("+T.xPercent+"%,"+T.yPercent+"%) translate3d(":"translate3d(")+M+"px,"+D+"px,"+F+"px)"+(1!==E||1!==C?" scale("+E+","+C+")":""));r=o=1,i=a=0}c=1,n=s=l=p=u=h=0,d=N?-1/N:0,f=T.zOrigin,m=1e-6,b=",",_="0",(k=A*q)&&(g=Math.cos(k),u=d*(l=-(v=Math.sin(k))),n=r*v,s=a*v,d*=c=g,r*=g,a*=g),(k=I*q)&&(e=i*(g=Math.cos(k))+n*(v=Math.sin(k)),y=o*g+s*v,p=c*v,h=d*v,n=i*-v+n*g,s=o*-v+s*g,c*=g,d*=g,i=e,o=y),1!==O&&(n*=O,s*=O,c*=O,d*=O),1!==C&&(i*=C,o*=C,p*=C,h*=C),1!==E&&(r*=E,a*=E,l*=E,u*=E),(f||R)&&(f&&(M+=n*-f,D+=s*-f,F+=c*-f+f),R&&(M+=T.xOrigin-(T.xOrigin*r+T.yOrigin*i)+T.xOffset,D+=T.yOrigin-(T.xOrigin*a+T.yOrigin*o)+T.yOffset),M<m&&-m<M&&(M=_),D<m&&-m<D&&(D=_),F<m&&-m<F&&(F=0)),x=T.xPercent||T.yPercent?"translate("+T.xPercent+"%,"+T.yPercent+"%) matrix3d(":"matrix3d(",x+=(r<m&&-m<r?_:r)+b+(a<m&&-m<a?_:a)+b+(l<m&&-m<l?_:l),x+=b+(u<m&&-m<u?_:u)+b+(i<m&&-m<i?_:i)+b+(o<m&&-m<o?_:o),I||A||1!==O?(x+=b+(p<m&&-m<p?_:p)+b+(h<m&&-m<h?_:h)+b+(n<m&&-m<n?_:n),x+=b+(s<m&&-m<s?_:s)+b+(c<m&&-m<c?_:c)+b+(d<m&&-m<d?_:d)+b):x+=",0,0,0,0,1,0,",x+=M+b+D+b+F+b+(N?1+-F/N:1)+")",P[Rt]=x}};(t=jt.prototype).x=t.y=t.z=t.skewX=t.skewY=t.rotation=t.rotationX=t.rotationY=t.zOrigin=t.xPercent=t.yPercent=t.xOffset=t.yOffset=0,t.scaleX=t.scaleY=t.scaleZ=1,Tt("transform,scale,scaleX,scaleY,scaleZ,x,y,z,rotation,rotationX,rotationY,rotationZ,skewX,skewY,shortRotation,shortRotationX,shortRotationY,shortRotationZ,transformOrigin,svgOrigin,transformPerspective,directionalRotation,parseTransform,force3D,skewType,xPercent,yPercent,smoothOrigin",{parser:function(t,e,r,i,n,a,o){if(i._lastParsedTransform===o)return n;var s,l=(i._lastParsedTransform=o).scale&&"function"==typeof o.scale?o.scale:0;"function"==typeof o[r]&&(s=o[r],o[r]=e),l&&(o.scale=l(D,t));var p,c,u,h,d,f,m,g,v,y=t._gsTransform,x=t.style,b=Ft.length,_=o,w={},S="transformOrigin",T=Wt(t,k,!0,_.parseTransform),P=_.transform&&("function"==typeof _.transform?_.transform(D,M):_.transform);if(T.skewType=_.skewType||T.skewType||$.defaultSkewType,i._transform=T,"rotationZ"in _&&(_.rotation=_.rotationZ),P&&"string"==typeof P&&Rt)(c=K.style)[Rt]=P,c.display="block",c.position="absolute",-1!==P.indexOf("%")&&(c.width=st(t,"width"),c.height=st(t,"height")),Z.body.appendChild(K),p=Wt(K,null,!1),"simple"===T.skewType&&(p.scaleY*=Math.cos(p.skewX*q)),T.svg&&(f=T.xOrigin,m=T.yOrigin,p.x-=T.xOffset,p.y-=T.yOffset,(_.transformOrigin||_.svgOrigin)&&(P={},It(t,ht(_.transformOrigin),P,_.svgOrigin,_.smoothOrigin,!0),f=P.xOrigin,m=P.yOrigin,p.x-=P.xOffset-T.xOffset,p.y-=P.yOffset-T.yOffset),(f||m)&&(g=Xt(K,!0),p.x-=f-(f*g[0]+m*g[2]),p.y-=m-(f*g[1]+m*g[3]))),Z.body.removeChild(K),p.perspective||(p.perspective=T.perspective),null!=_.xPercent&&(p.xPercent=A(_.xPercent,T.xPercent)),null!=_.yPercent&&(p.yPercent=A(_.yPercent,T.yPercent));else if("object"==typeof _){if(p={scaleX:A(null!=_.scaleX?_.scaleX:_.scale,T.scaleX),scaleY:A(null!=_.scaleY?_.scaleY:_.scale,T.scaleY),scaleZ:A(_.scaleZ,T.scaleZ),x:A(_.x,T.x),y:A(_.y,T.y),z:A(_.z,T.z),xPercent:A(_.xPercent,T.xPercent),yPercent:A(_.yPercent,T.yPercent),perspective:A(_.transformPerspective,T.perspective)},null!=(d=_.directionalRotation))if("object"==typeof d)for(c in d)_[c]=d[c];else _.rotation=d;"string"==typeof _.x&&-1!==_.x.indexOf("%")&&(p.x=0,p.xPercent=A(_.x,T.xPercent)),"string"==typeof _.y&&-1!==_.y.indexOf("%")&&(p.y=0,p.yPercent=A(_.y,T.yPercent)),p.rotation=E("rotation"in _?_.rotation:"shortRotation"in _?_.shortRotation+"_short":T.rotation,T.rotation,"rotation",w),Bt&&(p.rotationX=E("rotationX"in _?_.rotationX:"shortRotationX"in _?_.shortRotationX+"_short":T.rotationX||0,T.rotationX,"rotationX",w),p.rotationY=E("rotationY"in _?_.rotationY:"shortRotationY"in _?_.shortRotationY+"_short":T.rotationY||0,T.rotationY,"rotationY",w)),p.skewX=E(_.skewX,T.skewX),p.skewY=E(_.skewY,T.skewY)}for(Bt&&null!=_.force3D&&(T.force3D=_.force3D,h=!0),(u=T.force3D||T.z||T.rotationX||T.rotationY||p.z||p.rotationX||p.rotationY||p.perspective)||null==_.scale||(p.scaleZ=1);-1<--b;)(1e-6<(P=p[v=Ft[b]]-T[v])||P<-1e-6||null!=_[v]||null!=Q[v])&&(h=!0,n=new bt(T,v,T[v],P,n),v in w&&(n.e=w[v]),n.xs0=0,n.plugin=a,i._overwriteProps.push(n.n));return P=_.transformOrigin,T.svg&&(P||_.svgOrigin)&&(f=T.xOffset,m=T.yOffset,It(t,ht(P),p,_.svgOrigin,_.smoothOrigin),n=yt(T,"xOrigin",(y?T:p).xOrigin,p.xOrigin,n,S),n=yt(T,"yOrigin",(y?T:p).yOrigin,p.yOrigin,n,S),f===T.xOffset&&m===T.yOffset||(n=yt(T,"xOffset",y?f:T.xOffset,T.xOffset,n,S),n=yt(T,"yOffset",y?m:T.yOffset,T.yOffset,n,S)),P="0px 0px"),(P||Bt&&u&&T.zOrigin)&&(Rt?(h=!0,v=Lt,P=(P||st(t,v,k,!1,"50% 50%"))+"",(n=new bt(x,v,0,0,n,-1,S)).b=x[v],n.plugin=a,Bt?(c=T.zOrigin,P=P.split(" "),T.zOrigin=(2<P.length&&(0===c||"0px"!==P[2])?parseFloat(P[2]):c)||0,n.xs0=n.e=P[0]+" "+(P[1]||"50%")+" 0px",(n=new bt(T,"zOrigin",0,0,n,-1,n.n)).b=c,n.xs0=n.e=T.zOrigin):n.xs0=n.e=P):ht(P+"",T)),h&&(i._transformType=T.svg&&Et||!u&&3!==this._transformType?2:3),s&&(o[r]=s),l&&(o.scale=l),n},prefix:!0}),Tt("boxShadow",{defaultValue:"0px 0px 0px 0px #999",prefix:!0,color:!0,multi:!0,keyword:"inset"}),Tt("borderRadius",{defaultValue:"0px",parser:function(t,e,r,i,n,a){e=this.format(e);var o,s,l,p,c,u,h,d,f,m,g,v,y,x,b,_,w=["borderTopLeftRadius","borderTopRightRadius","borderBottomRightRadius","borderBottomLeftRadius"],S=t.style;for(f=parseFloat(t.offsetWidth),m=parseFloat(t.offsetHeight),o=e.split(" "),s=0;s<w.length;s++)this.p.indexOf("border")&&(w[s]=P(w[s])),-1!==(c=p=st(t,w[s],k,!1,"0px")).indexOf(" ")&&(c=(p=c.split(" "))[0],p=p[1]),u=l=o[s],h=parseFloat(c),v=c.substr((h+"").length),""===(g=(y="="===u.charAt(1))?(d=parseInt(u.charAt(0)+"1",10),u=u.substr(2),d*=parseFloat(u),u.substr((d+"").length-(d<0?1:0))||""):(d=parseFloat(u),u.substr((d+"").length)))&&(g=T[r]||v),g!==v&&(x=lt(t,"borderLeft",h,v),b=lt(t,"borderTop",h,v),p="%"===g?(c=x/f*100+"%",b/m*100+"%"):"em"===g?(c=x/(_=lt(t,"borderLeft",1,"em"))+"em",b/_+"em"):(c=x+"px",b+"px"),y&&(u=parseFloat(c)+d+g,l=parseFloat(p)+d+g)),n=_t(S,w[s],c+" "+p,u+" "+l,!1,"0px",n);return n},prefix:!0,formatter:gt("0px 0px 0px 0px",!1,!0)}),Tt("borderBottomLeftRadius,borderBottomRightRadius,borderTopLeftRadius,borderTopRightRadius",{defaultValue:"0px",parser:function(t,e,r,i,n,a){return _t(t.style,r,this.format(st(t,r,k,!1,"0px 0px")),this.format(e),!1,"0px",n)},prefix:!0,formatter:gt("0px 0px",!1,!0)}),Tt("backgroundPosition",{defaultValue:"0 0",parser:function(t,e,r,i,n,a){var o,s,l,p,c,u,h="background-position",d=k||ot(t,null),f=this.format((d?O?d.getPropertyValue(h+"-x")+" "+d.getPropertyValue(h+"-y"):d.getPropertyValue(h):t.currentStyle.backgroundPositionX+" "+t.currentStyle.backgroundPositionY)||"0 0"),m=this.format(e);if(-1!==f.indexOf("%")!=(-1!==m.indexOf("%"))&&m.split(",").length<2&&(u=st(t,"backgroundImage").replace(z,""))&&"none"!==u){for(o=f.split(" "),s=m.split(" "),tt.setAttribute("src",u),l=2;-1<--l;)(p=-1!==(f=o[l]).indexOf("%"))!=(-1!==s[l].indexOf("%"))&&(c=0===l?t.offsetWidth-tt.width:t.offsetHeight-tt.height,o[l]=p?parseFloat(f)/100*c+"px":parseFloat(f)/c*100+"%");f=o.join(" ")}return this.parseComplex(t.style,f,m,n,a)},formatter:ht}),Tt("backgroundSize",{defaultValue:"0 0",formatter:function(t){return"co"===(t+="").substr(0,2)?t:ht(-1===t.indexOf(" ")?t+" "+t:t)}}),Tt("perspective",{defaultValue:"0px",prefix:!0}),Tt("perspectiveOrigin",{defaultValue:"50% 50%",prefix:!0}),Tt("transformStyle",{prefix:!0}),Tt("backfaceVisibility",{prefix:!0}),Tt("userSelect",{prefix:!0}),Tt("margin",{parser:vt("marginTop,marginRight,marginBottom,marginLeft")}),Tt("padding",{parser:vt("paddingTop,paddingRight,paddingBottom,paddingLeft")}),Tt("clip",{defaultValue:"rect(0px,0px,0px,0px)",parser:function(t,e,r,i,n,a){var o,s,l;return e=O<9?(s=t.currentStyle,l=O<8?" ":",",o="rect("+s.clipTop+l+s.clipRight+l+s.clipBottom+l+s.clipLeft+")",this.format(e).split(",").join(l)):(o=this.format(st(t,this.p,k,!1,this.dflt)),this.format(e)),this.parseComplex(t.style,o,e,n,a)}}),Tt("textShadow",{defaultValue:"0px 0px 0px #999",color:!0,multi:!0}),Tt("autoRound,strictUnits",{parser:function(t,e,r,i,n){return n}}),Tt("border",{defaultValue:"0px solid #000",parser:function(t,e,r,i,n,a){var o=st(t,"borderTopWidth",k,!1,"0px"),s=this.format(e).split(" "),l=s[0].replace(L,"");return"px"!==l&&(o=parseFloat(o)/lt(t,"borderTopWidth",1,l)+l),this.parseComplex(t.style,this.format(o+" "+st(t,"borderTopStyle",k,!1,"solid")+" "+st(t,"borderTopColor",k,!1,"#000")),s.join(" "),n,a)},color:!0,formatter:function(t){var e=t.split(" ");return e[0]+" "+(e[1]||"solid")+" "+(t.match(mt)||["#000"])[0]}}),Tt("borderWidth",{parser:vt("borderTopWidth,borderRightWidth,borderBottomWidth,borderLeftWidth")}),Tt("float,cssFloat,styleFloat",{parser:function(t,e,r,i,n,a){var o=t.style,s="cssFloat"in o?"cssFloat":"styleFloat";return new bt(o,s,0,0,n,-1,r,!1,0,o[s],e)}});function qt(t){var e,r=this.t,i=r.filter||st(this.data,"filter")||"",n=this.s+this.c*t|0;100==n&&(e=-1===i.indexOf("atrix(")&&-1===i.indexOf("radient(")&&-1===i.indexOf("oader(")?(r.removeAttribute("filter"),!st(this.data,"filter")):(r.filter=i.replace(a,""),!0)),e||(this.xn1&&(r.filter=i=i||"alpha(opacity="+n+")"),-1===i.indexOf("pacity")?0==n&&this.xn1||(r.filter=i+" alpha(opacity="+n+")"):r.filter=i.replace(B,"opacity="+n))}Tt("opacity,alpha,autoAlpha",{defaultValue:"1",parser:function(t,e,r,i,n,a){var o=parseFloat(st(t,"opacity",k,!1,"1")),s=t.style,l="autoAlpha"===r;return"string"==typeof e&&"="===e.charAt(1)&&(e=("-"===e.charAt(0)?-1:1)*parseFloat(e.substr(2))+o),l&&1===o&&"hidden"===st(t,"visibility",k)&&0!==e&&(o=0),it?n=new bt(s,"opacity",o,e-o,n):((n=new bt(s,"opacity",100*o,100*(e-o),n)).xn1=l?1:0,s.zoom=1,n.type=2,n.b="alpha(opacity="+n.s+")",n.e="alpha(opacity="+(n.s+n.c)+")",n.data=t,n.plugin=a,n.setRatio=qt),l&&((n=new bt(s,"visibility",0,0,n,-1,null,!1,0,0!==o?"inherit":"hidden",0===e?"hidden":"inherit")).xs0="inherit",i._overwriteProps.push(n.n),i._overwriteProps.push(r)),n}});function Jt(t){if(this.t._gsClassPT=this,1===t||0===t){this.t.setAttribute("class",0===t?this.b:this.e);for(var e=this.data,r=this.t.style;e;)e.v?r[e.p]=e.v:Qt(r,e.p),e=e._next;1===t&&this.t._gsClassPT===this&&(this.t._gsClassPT=null)}else this.t.getAttribute("class")!==this.e&&this.t.setAttribute("class",this.e)}var Qt=function(t,e){e&&(t.removeProperty?("ms"!==e.substr(0,2)&&"webkit"!==e.substr(0,6)||(e="-"+e),t.removeProperty(e.replace(u,"-$1").toLowerCase())):t.removeAttribute(e))};Tt("className",{parser:function(t,e,r,i,n,a,o){var s,l,p,c,u,h=t.getAttribute("class")||"",d=t.style.cssText;if((n=i._classNamePT=new bt(t,r,0,0,n,2)).setRatio=Jt,n.pr=-11,f=!0,n.b=h,l=y(t,k),p=t._gsClassPT){for(c={},u=p.data;u;)c[u.p]=1,u=u._next;p.setRatio(1)}return(t._gsClassPT=n).e="="!==e.charAt(1)?e:h.replace(new RegExp("(?:\\s|^)"+e.substr(2)+"(?![\\w-])"),"")+("+"===e.charAt(0)?" "+e.substr(2):""),t.setAttribute("class",n.e),s=x(t,l,y(t),o,c),t.setAttribute("class",h),n.data=s.firstMPT,t.style.cssText=d,n=n.xfirst=i.parse(t,s.difs,n,a)}});function Zt(t){if((1===t||0===t)&&this.data._totalTime===this.data._totalDuration&&"isFromStart"!==this.data.data){var e,r,i,n,a,o=this.t.style,s=g.transform.parse;if("all"===this.e)n=!(o.cssText="");else for(i=(e=this.e.split(" ").join("").split(",")).length;-1<--i;)r=e[i],g[r]&&(g[r].parse===s?n=!0:r="transformOrigin"===r?Lt:g[r].p),Qt(o,r);n&&(Qt(o,Rt),(a=this.t._gsTransform)&&(a.svg&&(this.t.removeAttribute("data-svg-origin"),this.t.removeAttribute("transform")),delete this.t._gsTransform))}}for(Tt("clearProps",{parser:function(t,e,r,i,n){return(n=new bt(t,r,0,0,n,2)).setRatio=Zt,n.e=e,n.pr=-10,n.data=i._tween,f=!0,n}}),t="bezier,throwProps,physicsProps,physics2D".split(","),wt=t.length;wt--;)Pt(t[wt]);(t=$.prototype)._firstPT=t._lastParsedTransform=t._transform=null,t._onInitTween=function(t,e,r,i){if(!t.nodeType)return!1;this._target=M=t,this._tween=r,this._vars=e,D=i,C=e.autoRound,f=!1,T=e.suffixMap||$.suffixMap,k=ot(t,""),m=this._overwriteProps;var n,a,o,s,l,p,c,u,h,d=t.style;if(_&&""===d.zIndex&&("auto"!==(n=st(t,"zIndex",k))&&""!==n||this._addLazySet(d,"zIndex",0)),"string"==typeof e&&(s=d.cssText,n=y(t,k),d.cssText=s+";"+e,n=x(t,n,y(t)).difs,!it&&j.test(e)&&(n.opacity=parseFloat(RegExp.$1)),e=n,d.cssText=s),e.className?this._firstPT=a=g.className.parse(t,e.className,"className",this,null,null,e):this._firstPT=a=this.parse(t,e,null),this._transformType){for(h=3===this._transformType,Rt?w&&(_=!0,""===d.zIndex&&("auto"!==(c=st(t,"zIndex",k))&&""!==c||this._addLazySet(d,"zIndex",0)),S&&this._addLazySet(d,"WebkitBackfaceVisibility",this._vars.WebkitBackfaceVisibility||(h?"visible":"hidden"))):d.zoom=1,o=a;o&&o._next;)o=o._next;u=new bt(t,"transform",0,0,null,2),this._linkCSSP(u,null,o),u.setRatio=Rt?Ut:At,u.data=this._transform||Wt(t,k,!0),u.tween=r,u.pr=-1,m.pop()}if(f){for(;a;){for(p=a._next,o=s;o&&o.pr>a.pr;)o=o._next;(a._prev=o?o._prev:l)?a._prev._next=a:s=a,(a._next=o)?o._prev=a:l=a,a=p}this._firstPT=s}return!0},t.parse=function(t,e,r,i){var n,a,o,s,l,p,c,u,h,d,f=t.style;for(n in e){if("function"==typeof(p=e[n])&&(p=p(D,M)),a=g[n])r=a.parse(t,p,n,this,r,i,e);else{if("--"===n.substr(0,2)){this._tween._propLookup[n]=this._addTween.call(this._tween,t.style,"setProperty",ot(t).getPropertyValue(n)+"",p+"",n,!1,n);continue}l=st(t,n,k)+"",h="string"==typeof p,"color"===n||"fill"===n||"stroke"===n||-1!==n.indexOf("Color")||h&&H.test(p)?(h||(p=(3<(p=ft(p)).length?"rgba(":"rgb(")+p.join(",")+")"),r=_t(f,n,l,p,!0,"transparent",r,0,i)):h&&U.test(p)?r=_t(f,n,l,p,!0,null,r,0,i):(c=(o=parseFloat(l))||0===o?l.substr((o+"").length):"",""!==l&&"auto"!==l||(c="width"===n||"height"===n?(o=b(t,n,k),"px"):"left"===n||"top"===n?(o=pt(t,n,k),"px"):(o="opacity"!==n?0:1,"")),""===(u=(d=h&&"="===p.charAt(1))?(s=parseInt(p.charAt(0)+"1",10),p=p.substr(2),s*=parseFloat(p),p.replace(L,"")):(s=parseFloat(p),h?p.replace(L,""):""))&&(u=n in T?T[n]:c),p=s||0===s?(d?s+o:s)+u:e[n],c!==u&&(""===u&&"lineHeight"!==n||(s||0===s)&&o&&(o=lt(t,n,o,c),"%"===u?(o/=lt(t,n,100,"%")/100,!0!==e.strictUnits&&(l=o+"%")):"em"===u||"rem"===u||"vw"===u||"vh"===u?o/=lt(t,n,1,u):"px"!==u&&(s=lt(t,n,s,u),u="px"),d&&(!s&&0!==s||(p=s+o+u)))),d&&(s+=o),!o&&0!==o||!s&&0!==s?void 0!==f[n]&&(p||p+""!="NaN"&&null!=p)?(r=new bt(f,n,s||o||0,0,r,-1,n,!1,0,l,p)).xs0="none"!==p||"display"!==n&&-1===n.indexOf("Style")?p:l:v("invalid "+n+" tween value: "+e[n]):(r=new bt(f,n,o,s-o,r,0,n,!1!==C&&("px"===u||"zIndex"===n),0,l,p)).xs0=u)}i&&r&&!r.plugin&&(r.plugin=i)}return r},t.setRatio=function(t){var e,r,i,n=this._firstPT;if(1!==t||this._tween._time!==this._tween._duration&&0!==this._tween._time)if(t||this._tween._time!==this._tween._duration&&0!==this._tween._time||-1e-6===this._tween._rawPrevTime)for(;n;){if(e=n.c*t+n.s,n.r?e=n.r(e):e<1e-6&&-1e-6<e&&(e=0),n.type)if(1===n.type)if(2===(i=n.l))n.t[n.p]=n.xs0+e+n.xs1+n.xn1+n.xs2;else if(3===i)n.t[n.p]=n.xs0+e+n.xs1+n.xn1+n.xs2+n.xn2+n.xs3;else if(4===i)n.t[n.p]=n.xs0+e+n.xs1+n.xn1+n.xs2+n.xn2+n.xs3+n.xn3+n.xs4;else if(5===i)n.t[n.p]=n.xs0+e+n.xs1+n.xn1+n.xs2+n.xn2+n.xs3+n.xn3+n.xs4+n.xn4+n.xs5;else{for(r=n.xs0+e+n.xs1,i=1;i<n.l;i++)r+=n["xn"+i]+n["xs"+(i+1)];n.t[n.p]=r}else-1===n.type?n.t[n.p]=n.xs0:n.setRatio&&n.setRatio(t);else n.t[n.p]=e+n.xs0;n=n._next}else for(;n;)2!==n.type?n.t[n.p]=n.b:n.setRatio(t),n=n._next;else for(;n;){if(2!==n.type)if(n.r&&-1!==n.type)if(e=n.r(n.s+n.c),n.type){if(1===n.type){for(i=n.l,r=n.xs0+e+n.xs1,i=1;i<n.l;i++)r+=n["xn"+i]+n["xs"+(i+1)];n.t[n.p]=r}}else n.t[n.p]=e+n.xs0;else n.t[n.p]=n.e;else n.setRatio(t);n=n._next}},t._enableTransforms=function(t){this._transform=this._transform||Wt(this._target,k,!0),this._transformType=this._transform.svg&&Et||!t&&3!==this._transformType?2:3};function Kt(t){this.t[this.p]=this.e,this.data._linkCSSP(this,this._next,null,!0)}t._addLazySet=function(t,e,r){var i=this._firstPT=new bt(t,e,0,0,this._firstPT,2);i.e=r,i.setRatio=Kt,i.data=this},t._linkCSSP=function(t,e,r,i){return t&&(e&&(e._prev=t),t._next&&(t._next._prev=t._prev),t._prev?t._prev._next=t._next:this._firstPT===t&&(this._firstPT=t._next,i=!0),r?r._next=t:i||null!==this._firstPT||(this._firstPT=t),t._next=e,t._prev=r),t},t._mod=function(t){for(var e=this._firstPT;e;)"function"==typeof t[e.p]&&(e.r=t[e.p]),e=e._next},t._kill=function(t){var e,r,i,n=t;if(t.autoAlpha||t.alpha){for(r in n={},t)n[r]=t[r];n.opacity=1,n.autoAlpha&&(n.visibility=1)}for(t.className&&(e=this._classNamePT)&&((i=e.xfirst)&&i._prev?this._linkCSSP(i._prev,e._next,i._prev._prev):i===this._firstPT&&(this._firstPT=e._next),e._next&&this._linkCSSP(e._next,e._next._next,i._prev),this._classNamePT=null),e=this._firstPT;e;)e.plugin&&e.plugin!==r&&e.plugin._kill&&(e.plugin._kill(t),r=e.plugin),e=e._next;return ee.l.prototype._kill.call(this,n)};var te=function(t,e,r){var i,n,a,o;if(t.slice)for(n=t.length;-1<--n;)te(t[n],e,r);else for(n=(i=t.childNodes).length;-1<--n;)o=(a=i[n]).type,a.style&&(e.push(y(a)),r&&r.push(a)),1!==o&&9!==o&&11!==o||!a.childNodes.length||te(a,e,r)};return $.cascadeTo=function(t,e,r){var i,n,a,o,s=ee.n.to(t,e,r),l=[s],p=[],c=[],u=[],h=ee.n._internals.reservedProps;for(t=s._targets||s.target,te(t,p,u),s.render(e,!0,!0),te(t,c),s.render(0,!0,!0),s._enabled(!0),i=u.length;-1<--i;)if((n=x(u[i],p[i],c[i])).firstMPT){for(a in n=n.difs,r)h[a]&&(n[a]=r[a]);for(a in o={},n)o[a]=p[i][a];l.push(ee.n.fromTo(u[i],e,o,n))}return l},ee.l.activate([$]),$},!0);var i=ee.o.CSSPlugin},function(t,e,r){var i=r(2),n=r(25),a="".split;t.exports=i(function(){return!Object("z").propertyIsEnumerable(0)})?function(t){return"String"==n(t)?a.call(t,""):Object(t)}:Object},function(t,e,r){var i=r(3),n=r(83),a=r(32),o="__core-js_shared__",s=i[o]||n(o,{});(t.exports=function(t,e){return s[t]||(s[t]=void 0!==e?e:{})})("versions",[]).push({version:"3.1.3",mode:a?"pure":"global",copyright:"© 2019 Denis Pushkarev (zloirock.ru)"})},function(t,e){var r=0,i=Math.random();t.exports=function(t){return"Symbol("+String(void 0===t?"":t)+")_"+(++r+i).toString(36)}},function(t,e){t.exports={}},function(t,e,r){function i(s){return function(t,e,r){var i,n=l(t),a=p(n.length),o=c(r,a);if(s&&e!=e){for(;o<a;)if((i=n[o++])!=i)return!0}else for(;o<a;o++)if((s||o in n)&&n[o]===e)return s||o||0;return!s&&-1}}var l=r(19),p=r(9),c=r(34);t.exports={includes:i(!0),indexOf:i(!1)}},function(t,e,r){function i(t,e){var r=s[o(t)];return r==p||r!=l&&("function"==typeof e?n(e):!!e)}var n=r(2),a=/#|\.prototype\./,o=i.normalize=function(t){return String(t).replace(a,".").toLowerCase()},s=i.data={},l=i.NATIVE="N",p=i.POLYFILL="P";t.exports=i},function(t,e,r){var i=r(108),n=r(85);t.exports=Object.keys||function(t){return i(t,n)}},function(t,e,r){var i=r(4),n=r(41),a=r(8)("species");t.exports=function(t,e){var r;return n(t)&&("function"!=typeof(r=t.constructor)||r!==Array&&!n(r.prototype)?i(r)&&null===(r=r[a])&&(r=void 0):r=void 0),new(void 0===r?Array:r)(0===e?0:e)}},function(t,e,r){var i=r(2);t.exports=!i(function(){return Object.isExtensible(Object.preventExtensions({}))})},function(t,e,r){function h(t,e){this.stopped=t,this.result=e}var d=r(5),f=r(88),m=r(9),g=r(36),v=r(61),y=r(115);(t.exports=function(t,e,r,i,n){var a,o,s,l,p,c,u=g(e,r,i?2:1);if(n)a=t;else{if("function"!=typeof(o=v(t)))throw TypeError("Target is not iterable");if(f(o)){for(s=0,l=m(t.length);s<l;s++)if((p=i?u(d(c=t[s])[0],c[1]):u(t[s]))&&p instanceof h)return p;return new h(!1)}a=o.call(t)}for(;!(c=a.next()).done;)if((p=y(a,u,c.value,i))&&p instanceof h)return p;return new h(!1)}).stop=function(t){return new h(!0,t)}},function(t,e){t.exports={}},function(t,e,r){var i=r(62),n=r(60),a=r(8)("iterator");t.exports=function(t){if(null!=t)return t[a]||t["@@iterator"]||n[i(t)]}},function(t,e,r){var n=r(25),a=r(8)("toStringTag"),o="Arguments"==n(function(){return arguments}());t.exports=function(t){var e,r,i;return void 0===t?"Undefined":null===t?"Null":"string"==typeof(r=function(t,e){try{return t[e]}catch(t){}}(e=Object(t),a))?r:o?n(e):"Object"==(i=n(e))&&"function"==typeof e.callee?"Arguments":i}},function(t,e,r){var i=r(2),n=r(8)("species");t.exports=function(e){return!i(function(){var t=[];return(t.constructor={})[n]=function(){return{foo:1}},1!==t[e](Boolean).foo})}},function(t,e,r){"use strict";var i=r(5);t.exports=function(){var t=i(this),e="";return t.global&&(e+="g"),t.ignoreCase&&(e+="i"),t.multiline&&(e+="m"),t.dotAll&&(e+="s"),t.unicode&&(e+="u"),t.sticky&&(e+="y"),e}},function(t,e,r){"use strict";var i={}.propertyIsEnumerable,n=Object.getOwnPropertyDescriptor,a=n&&!i.call({1:2},1);e.f=a?function(t){var e=n(this,t);return!!e&&e.enumerable}:i},function(t,e,r){var i=r(51),n=r(52),a=i("keys");t.exports=function(t){return a[t]||(a[t]=n(t))}},function(t,e,r){"use strict";var i=r(32),n=r(3),a=r(2);t.exports=i||!a(function(){var t=Math.random();__defineSetter__.call(null,t,function(){}),delete n[t]})},function(t,e,r){var n=r(8)("iterator"),a=!1;try{var i=0,o={next:function(){return{done:!!i++}},return:function(){a=!0}};o[n]=function(){return this},Array.from(o,function(){throw 2})}catch(t){}t.exports=function(t,e){if(!e&&!a)return!1;var r=!1;try{var i={};i[n]=function(){return{next:function(){return{done:r=!0}}}},t(i)}catch(t){}return r}},function(t,e,r){function i(p){return function(t,e,r,i){c(e);var n=u(t),a=h(n),o=d(n.length),s=p?o-1:0,l=p?-1:1;if(r<2)for(;;){if(s in a){i=a[s],s+=l;break}if(s+=l,p?s<0:o<=s)throw TypeError("Reduce of empty array with no initial value")}for(;p?0<=s:s<o;s+=l)s in a&&(i=e(i,a[s],s,n));return i}}var c=r(21),u=r(11),h=r(50),d=r(9);t.exports={left:i(!1),right:i(!0)}},function(t,e,r){"use strict";var i=r(19),n=r(37),a=r(60),o=r(20),s=r(91),l="Array Iterator",p=o.set,c=o.getterFor(l);t.exports=s(Array,"Array",function(t,e){p(this,{type:l,target:i(t),index:0,kind:e})},function(){var t=c(this),e=t.target,r=t.kind,i=t.index++;return!e||i>=e.length?{value:t.target=void 0,done:!0}:"keys"==r?{value:i,done:!1}:"values"==r?{value:e[i],done:!1}:{value:[i,e[i]],done:!1}},"values"),a.Arguments=a.Array,n("keys"),n("values"),n("entries")},function(t,e,r){function i(s){return function(t,e){var r,i,n=String(p(t)),a=l(e),o=n.length;return a<0||o<=a?s?"":void 0:(r=n.charCodeAt(a))<55296||56319<r||a+1===o||(i=n.charCodeAt(a+1))<56320||57343<i?s?n.charAt(a):r:s?n.slice(a,a+2):i-56320+(r-55296<<10)+65536}}var l=r(24),p=r(15);t.exports={codeAt:i(!1),charAt:i(!0)}},function(t,e,r){"use strict";var u=r(14),h=r(17),d=r(2),f=r(8),m=r(73),g=f("species"),v=!d(function(){var t=/./;return t.exec=function(){var t=[];return t.groups={a:"7"},t},"7"!=="".replace(t,"$<a>")}),y=!d(function(){var t=/(?:)/,e=t.exec;t.exec=function(){return e.apply(this,arguments)};var r="ab".split(t);return 2!==r.length||"a"!==r[0]||"b"!==r[1]});t.exports=function(r,t,e,i){var n=f(r),a=!d(function(){var t={};return t[n]=function(){return 7},7!=""[r](t)}),o=a&&!d(function(){var t=!1,e=/a/;return e.exec=function(){return t=!0,null},"split"===r&&(e.constructor={},e.constructor[g]=function(){return e}),e[n](""),!t});if(!a||!o||"replace"===r&&!v||"split"===r&&!y){var s=/./[n],l=e(n,""[r],function(t,e,r,i,n){return e.exec===m?a&&!n?{done:!0,value:s.call(e,r,i)}:{done:!0,value:t.call(r,e,i)}:{done:!1}}),p=l[0],c=l[1];h(String.prototype,r,p),h(RegExp.prototype,n,2==t?function(t,e){return c.call(t,this,e)}:function(t){return c.call(t,this)}),i&&u(RegExp.prototype[n],"sham",!0)}}},function(t,e,r){"use strict";var i,n,o=r(64),s=RegExp.prototype.exec,l=String.prototype.replace,a=s,p=(i=/a/,n=/b*/g,s.call(i,"a"),s.call(n,"a"),0!==i.lastIndex||0!==n.lastIndex),c=void 0!==/()??/.exec("")[1];(p||c)&&(a=function(t){var e,r,i,n,a=this;return c&&(r=new RegExp("^"+a.source+"$(?!\\s)",o.call(a))),p&&(e=a.lastIndex),i=s.call(a,t),p&&i&&(a.lastIndex=a.global?i.index+i[0].length:e),c&&i&&1<i.length&&l.call(i[0],r,function(){for(n=1;n<arguments.length-2;n++)void 0===arguments[n]&&(i[n]=void 0)}),i}),t.exports=a},function(t,e,r){"use strict";var i=r(71).charAt;t.exports=function(t,e,r){return e+(r?i(t,e).length:1)}},function(t,e,r){var n=r(25),a=r(73);t.exports=function(t,e){var r=t.exec;if("function"==typeof r){var i=r.call(t,e);if("object"!=typeof i)throw TypeError("RegExp exec method returned something other than an Object or null");return i}if("RegExp"!==n(t))throw TypeError("RegExp#exec called on incompatible receiver");return a.call(t,e)}},function(t,e,r){var i=r(33);t.exports=i("navigator","userAgent")||""},function(t,e){t.exports="\t\n\v\f\r                　\u2028\u2029\ufeff"},function(t,e){var r=Math.expm1,i=Math.exp;t.exports=!r||22025.465794806718<r(10)||r(10)<22025.465794806718||-2e-17!=r(-2e-17)?function(t){return 0==(t=+t)?t:-1e-6<t&&t<1e-6?t+t*t/2:i(t)-1}:r},function(t,e,r){"use strict";var g=r(1),v=r(3),y=r(55),x=r(17),b=r(42),_=r(59),w=r(38),S=r(4),T=r(2),P=r(68),k=r(27),I=r(99);t.exports=function(i,t,e,n,a){function r(t){var r=s[t];x(s,t,"add"==t?function(t){return r.call(this,0===t?0:t),this}:"delete"==t?function(t){return!(a&&!S(t))&&r.call(this,0===t?0:t)}:"get"==t?function(t){return a&&!S(t)?void 0:r.call(this,0===t?0:t)}:"has"==t?function(t){return!(a&&!S(t))&&r.call(this,0===t?0:t)}:function(t,e){return r.call(this,0===t?0:t,e),this})}var o=v[i],s=o&&o.prototype,l=o,p=n?"set":"add",c={};if(y(i,"function"!=typeof o||!(a||s.forEach&&!T(function(){(new o).entries().next()}))))l=e.getConstructor(t,i,n,p),b.REQUIRED=!0;else if(y(i,!0)){var u=new l,h=u[p](a?{}:-0,1)!=u,d=T(function(){u.has(1)}),f=P(function(t){new o(t)}),m=!a&&T(function(){for(var t=new o,e=5;e--;)t[p](e,e);return!t.has(-0)});f||(((l=t(function(t,e){w(t,l,i);var r=I(new o,t,l);return null!=e&&_(e,r[p],r,n),r})).prototype=s).constructor=l),(d||m)&&(r("delete"),r("has"),n&&r("get")),(m||h)&&r(p),a&&s.clear&&delete s.clear}return c[i]=l,g({global:!0,forced:l!=o},c),k(l,i),a||e.setStrong(l,i,n),l}},function(t,e,r){"use strict";function i(t,e,r){var i,n,a,o=new Array(r),s=8*r-e-1,l=(1<<s)-1,p=l>>1,c=23===e?V(2,-24)-V(2,-77):0,u=t<0||0===t&&1/t<0?1:0,h=0;for((t=$(t))!=t||t===1/0?(n=t!=t?1:0,i=l):(i=H(z(t)/G),t*(a=V(2,-i))<1&&(i--,a*=2),2<=(t+=1<=i+p?c/a:c*V(2,1-p))*a&&(i++,a/=2),l<=i+p?(n=0,i=l):1<=i+p?(n=(t*a-1)*V(2,e),i+=p):(n=t*V(2,p-1)*V(2,e),i=0));8<=e;o[h++]=255&n,n/=256,e-=8);for(i=i<<e|n,s+=e;0<s;o[h++]=255&i,i/=256,s-=8);return o[--h]|=128*u,o}function n(t,e){var r,i=t.length,n=8*i-e-1,a=(1<<n)-1,o=a>>1,s=n-7,l=i-1,p=t[l--],c=127&p;for(p>>=7;0<s;c=256*c+t[l],l--,s-=8);for(r=c&(1<<-s)-1,c>>=-s,s+=e;0<s;r=256*r+t[l],l--,s-=8);if(0===c)c=1-o;else{if(c===a)return r?NaN:p?-1/0:1/0;r+=V(2,e),c-=o}return(p?-1:1)*r*V(2,c-e)}function a(t){return t[3]<<24|t[2]<<16|t[1]<<8|t[0]}function o(t){return[255&t]}function s(t){return[255&t,t>>8&255]}function l(t){return[255&t,t>>8&255,t>>16&255,t>>24&255]}function p(t){return i(t,23,4)}function c(t){return i(t,52,8)}function u(t,e){P(t[D],e,{get:function(){return E(this)[e]}})}function h(t,e,r,i){var n=S(+r),a=E(t);if(n+e>a.byteLength)throw j(F);var o=E(a.buffer).bytes,s=n+a.byteOffset,l=o.slice(s,s+e);return i?l:l.reverse()}function d(t,e,r,i,n,a){var o=S(+r),s=E(t);if(o+e>s.byteLength)throw j(F);for(var l=E(s.buffer).bytes,p=o+s.byteOffset,c=i(+n),u=0;u<e;u++)l[p+u]=c[a?u:e-u-1]}var f=r(3),m=r(7),g=r(6).NATIVE_ARRAY_BUFFER,v=r(14),y=r(48),x=r(2),b=r(38),_=r(24),w=r(9),S=r(137),T=r(40).f,P=r(10).f,k=r(90),I=r(27),A=r(20),E=A.get,C=A.set,O="ArrayBuffer",M="DataView",D="prototype",F="Wrong index",R=f[O],N=R,L=f[M],B=f.Math,j=f.RangeError,$=B.abs,V=B.pow,H=B.floor,z=B.log,G=B.LN2;if(g){if(!x(function(){R(1)})||!x(function(){new R(-1)})||x(function(){return new R,new R(1.5),new R(NaN),R.name!=O})){for(var Y,X=(N=function(t){return b(this,N),new R(S(t))})[D]=R[D],W=T(R),U=0;W.length>U;)(Y=W[U++])in N||v(N,Y,R[Y]);X.constructor=N}var q=new L(new N(2)),J=L[D].setInt8;q.setInt8(0,2147483648),q.setInt8(1,2147483649),!q.getInt8(0)&&q.getInt8(1)||y(L[D],{setInt8:function(t,e){J.call(this,t,e<<24>>24)},setUint8:function(t,e){J.call(this,t,e<<24>>24)}},{unsafe:!0})}else N=function(t){b(this,N,O);var e=S(t);C(this,{bytes:k.call(new Array(e),0),byteLength:e}),m||(this.byteLength=e)},L=function(t,e,r){b(this,L,M),b(t,N,M);var i=E(t).byteLength,n=_(e);if(n<0||i<n)throw j("Wrong offset");if(i<n+(r=void 0===r?i-n:w(r)))throw j("Wrong length");C(this,{buffer:t,byteLength:r,byteOffset:n}),m||(this.buffer=t,this.byteLength=r,this.byteOffset=n)},m&&(u(N,"byteLength"),u(L,"buffer"),u(L,"byteLength"),u(L,"byteOffset")),y(L[D],{getInt8:function(t){return h(this,1,t)[0]<<24>>24},getUint8:function(t){return h(this,1,t)[0]},getInt16:function(t,e){var r=h(this,2,t,1<arguments.length?e:void 0);return(r[1]<<8|r[0])<<16>>16},getUint16:function(t,e){var r=h(this,2,t,1<arguments.length?e:void 0);return r[1]<<8|r[0]},getInt32:function(t,e){return a(h(this,4,t,1<arguments.length?e:void 0))},getUint32:function(t,e){return a(h(this,4,t,1<arguments.length?e:void 0))>>>0},getFloat32:function(t,e){return n(h(this,4,t,1<arguments.length?e:void 0),23)},getFloat64:function(t,e){return n(h(this,8,t,1<arguments.length?e:void 0),52)},setInt8:function(t,e){d(this,1,t,o,e)},setUint8:function(t,e){d(this,1,t,o,e)},setInt16:function(t,e,r){d(this,2,t,s,e,2<arguments.length?r:void 0)},setUint16:function(t,e,r){d(this,2,t,s,e,2<arguments.length?r:void 0)},setInt32:function(t,e,r){d(this,4,t,l,e,2<arguments.length?r:void 0)},setUint32:function(t,e,r){d(this,4,t,l,e,2<arguments.length?r:void 0)},setFloat32:function(t,e,r){d(this,4,t,p,e,2<arguments.length?r:void 0)},setFloat64:function(t,e,r){d(this,8,t,c,e,2<arguments.length?r:void 0)}});I(N,O),I(L,M),e[O]=N,e[M]=L},function(t,e){var r;r=function(){return this}();try{r=r||new Function("return this")()}catch(t){"object"==typeof window&&(r=window)}t.exports=r},function(t,e,r){var i=r(3),n=r(4),a=i.document,o=n(a)&&n(a.createElement);t.exports=function(t){return o?a.createElement(t):{}}},function(t,e,r){var i=r(3),n=r(14);t.exports=function(e,r){try{n(i,e,r)}catch(t){i[e]=r}return r}},function(t,e,r){var i=r(33),n=r(40),a=r(86),o=r(5);t.exports=i("Reflect","ownKeys")||function(t){var e=n.f(o(t)),r=a.f;return r?e.concat(r(t)):e}},function(t,e){t.exports=["constructor","hasOwnProperty","isPrototypeOf","propertyIsEnumerable","toLocaleString","toString","valueOf"]},function(t,e){e.f=Object.getOwnPropertySymbols},function(t,e,r){var i=r(7),o=r(10),s=r(5),l=r(56);t.exports=i?Object.defineProperties:function(t,e){s(t);for(var r,i=l(e),n=i.length,a=0;a<n;)o.f(t,r=i[a++],e[r]);return t}},function(t,e,r){var i=r(8),n=r(60),a=i("iterator"),o=Array.prototype;t.exports=function(t){return void 0!==t&&(n.Array===t||o[a]===t)}},function(t,e,r){var i=r(2);t.exports=!i(function(){function t(){}return t.prototype.constructor=null,Object.getPrototypeOf(new t)!==t.prototype})},function(t,e,r){"use strict";var p=r(11),c=r(34),u=r(9);t.exports=function(t,e,r){for(var i=p(this),n=u(i.length),a=arguments.length,o=c(1<a?e:void 0,n),s=2<a?r:void 0,l=void 0===s?n:c(s,n);o<l;)i[o++]=t;return i}},function(t,e,r){"use strict";function v(){return this}var y=r(1),x=r(92),b=r(28),_=r(45),w=r(27),S=r(14),T=r(17),i=r(8),P=r(32),k=r(60),n=r(124),I=n.IteratorPrototype,A=n.BUGGY_SAFARI_ITERATORS,E=i("iterator"),C="values",O="entries";t.exports=function(t,e,r,i,n,a,o){x(r,e,i);function s(t){if(t===n&&m)return m;if(!A&&t in d)return d[t];switch(t){case"keys":case C:case O:return function(){return new r(this,t)}}return function(){return new r(this)}}var l,p,c,u=e+" Iterator",h=!1,d=t.prototype,f=d[E]||d["@@iterator"]||n&&d[n],m=!A&&f||s(n),g="Array"==e&&d.entries||f;if(g&&(l=b(g.call(new t)),I!==Object.prototype&&l.next&&(P||b(l)===I||(_?_(l,I):"function"!=typeof l[E]&&S(l,E,v)),w(l,u,!0,!0),P&&(k[u]=v))),n==C&&f&&f.name!==C&&(h=!0,m=function(){return f.call(this)}),P&&!o||d[E]===m||S(d,E,m),k[e]=m,n)if(p={values:s(C),keys:a?m:s("keys"),entries:s(O)},o)for(c in p)!A&&!h&&c in d||T(d,c,p[c]);else y({target:e,proto:!0,forced:A||h},p);return p}},function(t,e,r){"use strict";function n(){return this}var a=r(124).IteratorPrototype,o=r(35),s=r(39),l=r(27),p=r(60);t.exports=function(t,e,r){var i=e+" Iterator";return t.prototype=o(a,{next:s(1,r)}),l(t,i,!1,!0),p[i]=n,t}},function(t,e,r){var i=r(94);t.exports=function(t){if(i(t))throw TypeError("The method doesn't accept regular expressions");return t}},function(t,e,r){var i=r(4),n=r(25),a=r(8)("match");t.exports=function(t){var e;return i(t)&&(void 0!==(e=t[a])?!!e:"RegExp"==n(t))}},function(t,e,r){var i=r(8)("match");t.exports=function(e){var r=/./;try{"/./"[e](r)}catch(t){try{return r[i]=!1,"/./"[e](r)}catch(t){}}return!1}},function(t,e,r){function i(p){return function(t,e,r){var i,n,a=String(h(t)),o=a.length,s=void 0===r?" ":String(r),l=c(e);return l<=o||""==s?a:(i=l-o,(n=u.call(s,d(i/s.length))).length>i&&(n=n.slice(0,i)),p?a+n:n+a)}}var c=r(9),u=r(97),h=r(15),d=Math.ceil;t.exports={start:i(!1),end:i(!0)}},function(t,e,r){"use strict";var n=r(24),a=r(15);t.exports="".repeat||function(t){var e=String(a(this)),r="",i=n(t);if(i<0||i==1/0)throw RangeError("Wrong number of repetitions");for(;0<i;(i>>>=1)&&(e+=e))1&i&&(r+=e);return r}},function(t,e,r){var i=r(2),n=r(77);t.exports=function(t){return i(function(){return!!n[t]()||"​᠎"!="​᠎"[t]()||n[t].name!==t})}},function(t,e,r){var a=r(4),o=r(45);t.exports=function(t,e,r){var i,n;return o&&"function"==typeof(i=e.constructor)&&i!==r&&a(n=i.prototype)&&n!==r.prototype&&o(t,n),t}},function(t,e){t.exports=Math.sign||function(t){return 0==(t=+t)||t!=t?t:t<0?-1:1}},function(t,e,r){function i(t){if(S.hasOwnProperty(t)){var e=S[t];delete S[t],e()}}function n(t){return function(){i(t)}}function a(t){i(t.data)}function o(t){c.postMessage(t+"",g.protocol+"//"+g.host)}var s,l,p,c=r(3),u=r(2),h=r(25),d=r(36),f=r(110),m=r(82),g=c.location,v=c.setImmediate,y=c.clearImmediate,x=c.process,b=c.MessageChannel,_=c.Dispatch,w=0,S={},T="onreadystatechange";v&&y||(v=function(t){for(var e=[],r=1;r<arguments.length;)e.push(arguments[r++]);return S[++w]=function(){("function"==typeof t?t:Function(t)).apply(void 0,e)},s(w),w},y=function(t){delete S[t]},"process"==h(x)?s=function(t){x.nextTick(n(t))}:_&&_.now?s=function(t){_.now(n(t))}:b?(p=(l=new b).port2,l.port1.onmessage=a,s=d(p.postMessage,p,1)):!c.addEventListener||"function"!=typeof postMessage||c.importScripts||u(o)?s=T in m("script")?function(t){f.appendChild(m("script"))[T]=function(){f.removeChild(this),i(t)}}:function(t){setTimeout(n(t),0)}:(s=o,c.addEventListener("message",a,!1))),t.exports={set:v,clear:y}},function(t,e,r){var i=r(3),n=r(2),a=r(68),o=r(6).NATIVE_ARRAY_BUFFER_VIEWS,s=i.ArrayBuffer,l=i.Int8Array;t.exports=!o||!n(function(){l(1)})||!n(function(){new l(-1)})||!a(function(t){new l,new l(null),new l(1.5),new l(t)},!0)||n(function(){return 1!==new l(new s(2),1,void 0).length})},function(t,e,r){"use strict";var i,n,a,o,s,l;Object.defineProperty(e,"__esModule",{value:!0}),(n=i||(i={})).prx1=prx,n.prxy1=prxy,e.proto=window.proto,e.prx=i.prx1,e.prxy=i.prxy1,o=a||(a={}),"undefined"!=typeof pff&&(o.pff1=pff),e.pff=a.pff1,l=s||(s={}),"undefined"!=typeof plupload&&(l.plupload1=plupload),"undefined"!=typeof mOxie&&(l.moxie1=mOxie),e.plupload=s.plupload1,e.moxie=s.moxie1},function(t,e,r){var i=r(7),n=r(2),a=r(82);t.exports=!i&&!n(function(){return 7!=Object.defineProperty(a("div"),"a",{get:function(){return 7}}).a})},function(t,e,r){var i=r(51);t.exports=i("native-function-to-string",Function.toString)},function(t,e,r){var i=r(3),n=r(105),a=i.WeakMap;t.exports="function"==typeof a&&/native code/.test(n.call(a))},function(t,e,r){var s=r(12),l=r(84),p=r(16),c=r(10);t.exports=function(t,e){for(var r=l(e),i=c.f,n=p.f,a=0;a<r.length;a++){var o=r[a];s(t,o)||i(t,o,n(e,o))}}},function(t,e,r){var o=r(12),s=r(19),l=r(54).indexOf,p=r(53);t.exports=function(t,e){var r,i=s(t),n=0,a=[];for(r in i)!o(p,r)&&o(i,r)&&a.push(r);for(;e.length>n;)o(i,r=e[n++])&&(~l(a,r)||a.push(r));return a}},function(t,e,r){var i=r(2);t.exports=!!Object.getOwnPropertySymbols&&!i(function(){return!String(Symbol())})},function(t,e,r){var i=r(33);t.exports=i("document","documentElement")},function(t,e,r){var i=r(19),n=r(40).f,a={}.toString,o="object"==typeof window&&window&&Object.getOwnPropertyNames?Object.getOwnPropertyNames(window):[];t.exports.f=function(t){return o&&"[object Window]"==a.call(t)?function(t){try{return n(t)}catch(t){return o.slice()}}(t):n(i(t))}},function(t,e,r){e.f=r(8)},function(t,e,r){"use strict";var h=r(7),i=r(2),d=r(56),f=r(86),m=r(65),g=r(11),v=r(50),n=Object.assign;t.exports=!n||i(function(){var t={},e={},r=Symbol(),i="abcdefghijklmnopqrst";return t[r]=7,i.split("").forEach(function(t){e[t]=t}),7!=n({},t)[r]||d(n({},e)).join("")!=i})?function(t,e){for(var r=g(t),i=arguments.length,n=1,a=f.f,o=m.f;n<i;)for(var s,l=v(arguments[n++]),p=a?d(l).concat(a(l)):d(l),c=p.length,u=0;u<c;)s=p[u++],h&&!o.call(l,s)||(r[s]=l[s]);return r}:n},function(t,e,r){function i(s){return function(t){for(var e,r=c(t),i=p(r),n=i.length,a=0,o=[];a<n;)e=i[a++],l&&!u.call(r,e)||o.push(s?[e,r[e]]:r[e]);return o}}var l=r(7),p=r(56),c=r(19),u=r(65).f;t.exports={entries:i(!0),values:i(!1)}},function(t,e,r){var a=r(5);t.exports=function(e,t,r,i){try{return i?t(a(r)[0],r[1]):t(r)}catch(t){var n=e.return;throw void 0!==n&&a(n.call(e)),t}}},function(t,e){t.exports=Object.is||function(t,e){return t===e?0!==t||1/t==1/e:t!=t&&e!=e}},function(t,e,r){var i=r(4);t.exports=function(t){if(!i(t)&&null!==t)throw TypeError("Can't set "+String(t)+" as a prototype");return t}},function(t,e,r){"use strict";var a=r(21),o=r(4),s=[].slice,l={};t.exports=Function.bind||function(e){var r=a(this),i=s.call(arguments,1),n=function(){var t=i.concat(s.call(arguments));return this instanceof n?function(t,e,r){if(!(e in l)){for(var i=[],n=0;n<e;n++)i[n]="a["+n+"]";l[e]=Function("C,a","return new C("+i.join(",")+")")}return l[e](t,r)}(r,t.length,t):r.apply(e,t)};return o(r.prototype)&&(n.prototype=r.prototype),n}},function(t,e,r){"use strict";var f=r(36),m=r(11),g=r(115),v=r(88),y=r(9),x=r(43),b=r(61);t.exports=function(t,e,r){var i,n,a,o,s=m(t),l="function"==typeof this?this:Array,p=arguments.length,c=1<p?e:void 0,u=void 0!==c,h=0,d=b(s);if(u&&(c=f(c,2<p?r:void 0,2)),null==d||l==Array&&v(d))for(n=new l(i=y(s.length));h<i;h++)x(n,h,u?c(s[h],h):s[h]);else for(o=d.call(s),n=new l;!(a=o.next()).done;h++)x(n,h,u?g(o,c,[a.value,h],!0):a.value);return n.length=h,n}},function(t,e,r){"use strict";var c=r(11),u=r(34),h=r(9),d=Math.min;t.exports=[].copyWithin||function(t,e,r){var i=c(this),n=h(i.length),a=u(t,n),o=u(e,n),s=2<arguments.length?r:void 0,l=d((void 0===s?n:u(s,n))-o,n-a),p=1;for(o<a&&a<o+l&&(p=-1,o+=l-1,a+=l-1);0<l--;)o in i?i[a]=i[o]:delete i[a],a+=p,o+=p;return i}},function(t,e,r){"use strict";var h=r(41),d=r(9),f=r(36),m=function(t,e,r,i,n,a,o,s){for(var l,p=n,c=0,u=!!o&&f(o,s,3);c<i;){if(c in r){if(l=u?u(r[c],c,e):r[c],0<a&&h(l))p=m(t,e,l,d(l.length),p,a-1)-1;else{if(9007199254740991<=p)throw TypeError("Exceed the acceptable array length");t[p]=l}p++}c++}return p};t.exports=m},function(t,e,r){"use strict";var i=r(13).forEach,n=r(29);t.exports=n("forEach")?function(t,e){return i(this,t,1<arguments.length?e:void 0)}:[].forEach},function(t,e,r){"use strict";var a=r(19),o=r(24),s=r(9),i=r(29),l=Math.min,p=[].lastIndexOf,c=!!p&&1/[1].lastIndexOf(1,-0)<0,n=i("lastIndexOf");t.exports=c||n?function(t,e){if(c)return p.apply(this,arguments)||0;var r=a(this),i=s(r.length),n=i-1;for(1<arguments.length&&(n=l(n,o(e))),n<0&&(n=i+n);0<=n;n--)if(n in r&&r[n]===t)return n||0;return-1}:p},function(t,e,r){"use strict";var i,n,a,o=r(28),s=r(14),l=r(12),p=r(8),c=r(32),u=p("iterator"),h=!1;[].keys&&("next"in(a=[].keys())?(n=o(o(a)))!==Object.prototype&&(i=n):h=!0),null==i&&(i={}),c||l(i,u)||s(i,u,function(){return this}),t.exports={IteratorPrototype:i,BUGGY_SAFARI_ITERATORS:h}},function(t,e,r){var i=r(76);t.exports=/Version\/10\.\d+(\.\d+)?( Mobile\/\w+)? Safari\//.test(i)},function(t,e,r){"use strict";var n=r(71).charAt,i=r(20),a=r(91),o="String Iterator",s=i.set,l=i.getterFor(o);a(String,"String",function(t){s(this,{type:o,string:String(t),index:0})},function(){var t,e=l(this),r=e.string,i=e.index;return i>=r.length?{value:void 0,done:!0}:(t=n(r,i),e.index+=t.length,{value:t,done:!1})})},function(t,e,r){var i=r(3),n=r(47).trim,a=r(77),o=i.parseInt,s=/^[+-]?0[Xx]/,l=8!==o(a+"08")||22!==o(a+"0x16");t.exports=l?function(t,e){var r=n(String(t));return o(r,e>>>0||(s.test(r)?16:10))}:o},function(t,e,r){var i=r(3),n=r(47).trim,a=r(77),o=i.parseFloat,s=1/o(a+"-0")!=-1/0;t.exports=s?function(t){var e=n(String(t)),r=o(e);return 0===r&&"-"==e.charAt(0)?-0:r}:o},function(t,e,r){var i=r(4),n=Math.floor;t.exports=function(t){return!i(t)&&isFinite(t)&&n(t)===t}},function(t,e,r){var i=r(25);t.exports=function(t){if("number"!=typeof t&&"Number"!=i(t))throw TypeError("Incorrect invocation");return+t}},function(t,e){var r=Math.log;t.exports=Math.log1p||function(t){return-1e-8<(t=+t)&&t<1e-8?t-t*t/2:r(1+t)}},function(t,e,r){var i,n,a,o,s,l,p,c=r(3),u=r(16).f,h=r(25),d=r(101).set,f=r(76),m=c.MutationObserver||c.WebKitMutationObserver,g=c.process,v=c.Promise,y="process"==h(g),x=u(c,"queueMicrotask"),b=x&&x.value;b||(i=function(){var t,e;for(y&&(t=g.domain)&&t.exit();n;){e=n.fn,n=n.next;try{e()}catch(t){throw n?o():a=void 0,t}}a=void 0,t&&t.enter()},o=y?function(){g.nextTick(i)}:m&&!/(iphone|ipod|ipad).*applewebkit/i.test(f)?(s=!0,l=document.createTextNode(""),new m(i).observe(l,{characterData:!0}),function(){l.data=s=!s}):v&&v.resolve?(p=v.resolve(void 0),function(){p.then(i)}):function(){d.call(c,i)}),t.exports=b||function(t){var e={fn:t,next:void 0};a&&(a.next=e),n||(n=e,o()),a=e}},function(t,e,r){var i=r(5),n=r(4),a=r(134);t.exports=function(t,e){if(i(t),n(e)&&e.constructor===t)return e;var r=a.f(t);return(0,r.resolve)(e),r.promise}},function(t,e,r){"use strict";function i(t){var r,i;this.promise=new t(function(t,e){if(void 0!==r||void 0!==i)throw TypeError("Bad Promise constructor");r=t,i=e}),this.resolve=n(r),this.reject=n(i)}var n=r(21);t.exports.f=function(t){return new i(t)}},function(t,e,r){"use strict";var p=r(10).f,c=r(35),u=r(48),h=r(36),d=r(38),f=r(59),o=r(91),s=r(46),m=r(7),g=r(42).fastKey,i=r(20),v=i.set,y=i.getterFor;t.exports={getConstructor:function(t,r,i,n){function a(t,e,r){var i,n,a=s(t),o=l(t,e);return o?o.value=r:(a.last=o={index:n=g(e,!0),key:e,value:r,previous:i=a.last,next:void 0,removed:!1},a.first||(a.first=o),i&&(i.next=o),m?a.size++:t.size++,"F"!==n&&(a.index[n]=o)),t}var o=t(function(t,e){d(t,o,r),v(t,{type:r,index:c(null),first:void 0,last:void 0,size:0}),m||(t.size=0),null!=e&&f(e,t[n],t,i)}),s=y(r),l=function(t,e){var r,i=s(t),n=g(e);if("F"!==n)return i.index[n];for(r=i.first;r;r=r.next)if(r.key==e)return r};return u(o.prototype,{clear:function(){for(var t=s(this),e=t.index,r=t.first;r;)r.removed=!0,r.previous&&(r.previous=r.previous.next=void 0),delete e[r.index],r=r.next;t.first=t.last=void 0,m?t.size=0:this.size=0},delete:function(t){var e=s(this),r=l(this,t);if(r){var i=r.next,n=r.previous;delete e.index[r.index],r.removed=!0,n&&(n.next=i),i&&(i.previous=n),e.first==r&&(e.first=i),e.last==r&&(e.last=n),m?e.size--:this.size--}return!!r},forEach:function(t,e){for(var r,i=s(this),n=h(t,1<arguments.length?e:void 0,3);r=r?r.next:i.first;)for(n(r.value,r.key,this);r&&r.removed;)r=r.previous},has:function(t){return!!l(this,t)}}),u(o.prototype,i?{get:function(t){var e=l(this,t);return e&&e.value},set:function(t,e){return a(this,0===t?0:t,e)}}:{add:function(t){return a(this,t=0===t?0:t,t)}}),m&&p(o.prototype,"size",{get:function(){return s(this).size}}),o},setStrong:function(t,e,r){var i=e+" Iterator",n=y(e),a=y(i);o(t,e,function(t,e){v(this,{type:i,target:t,state:n(t),kind:e,last:void 0})},function(){for(var t=a(this),e=t.kind,r=t.last;r&&r.removed;)r=r.previous;return t.target&&(t.last=r=r?r.next:t.state.first)?"keys"==e?{value:r.key,done:!1}:"values"==e?{value:r.value,done:!1}:{value:[r.key,r.value],done:!1}:{value:t.target=void 0,done:!0}},r?"entries":"values",!r,!0),s(e)}}},function(t,e,r){"use strict";function l(t){return t.frozen||(t.frozen=new x)}function i(t,e){return o(t.entries,function(t){return t[0]===e})}var p=r(48),c=r(42).getWeakData,u=r(5),h=r(4),d=r(38),f=r(59),n=r(13),m=r(12),a=r(20),g=a.set,v=a.getterFor,o=n.find,s=n.findIndex,y=0,x=function(){this.entries=[]};x.prototype={get:function(t){var e=i(this,t);if(e)return e[1]},has:function(t){return!!i(this,t)},set:function(t,e){var r=i(this,t);r?r[1]=e:this.entries.push([t,e])},delete:function(e){var t=s(this.entries,function(t){return t[0]===e});return~t&&this.entries.splice(t,1),!!~t}},t.exports={getConstructor:function(t,r,i,n){function a(t,e,r){var i=s(t),n=c(u(e),!0);return!0===n?l(i).set(e,r):n[i.id]=r,t}var o=t(function(t,e){d(t,o,r),g(t,{type:r,id:y++,frozen:void 0}),null!=e&&f(e,t[n],t,i)}),s=v(r);return p(o.prototype,{delete:function(t){var e=s(this);if(!h(t))return!1;var r=c(t);return!0===r?l(e).delete(t):r&&m(r,e.id)&&delete r[e.id]},has:function(t){var e=s(this);if(!h(t))return!1;var r=c(t);return!0===r?l(e).has(t):r&&m(r,e.id)}}),p(o.prototype,i?{get:function(t){var e=s(this);if(h(t)){var r=c(t);return!0===r?l(e).get(t):r?r[e.id]:void 0}},set:function(t,e){return a(this,t,e)}}:{add:function(t){return a(this,t,!0)}}),o}}},function(t,e,r){var i=r(24),n=r(9);t.exports=function(t){if(void 0===t)return 0;var e=i(t),r=n(e);if(e!==r)throw RangeError("Wrong length or index");return r}},function(t,e,r){var i=r(24);t.exports=function(t,e){var r=i(t);if(r<0||r%e)throw RangeError("Wrong offset");return r}},function(t,e,r){var d=r(11),f=r(9),m=r(61),g=r(88),v=r(36),y=r(6).aTypedArrayConstructor;t.exports=function(t,e,r){var i,n,a,o,s,l=d(t),p=arguments.length,c=1<p?e:void 0,u=void 0!==c,h=m(l);if(null!=h&&!g(h))for(s=h.call(l),l=[];!(o=s.next()).done;)l.push(o.value);for(u&&2<p&&(c=v(c,r,2)),n=f(l.length),a=new(y(this))(n),i=0;i<n;i++)a[i]=u?c(l[i],i):l[i];return a}},function(t,e){t.exports={CSSRuleList:0,CSSStyleDeclaration:0,CSSValueList:0,ClientRectList:0,DOMRectList:0,DOMStringList:0,DOMTokenList:1,DataTransferItemList:0,FileList:0,HTMLAllCollection:0,HTMLCollection:0,HTMLFormElement:0,HTMLSelectElement:0,MediaList:0,MimeTypeArray:0,NamedNodeMap:0,NodeList:1,PaintRequestList:0,Plugin:0,PluginArray:0,SVGLengthList:0,SVGNumberList:0,SVGPathSegList:0,SVGPointList:0,SVGStringList:0,SVGTransformList:0,SourceBufferList:0,StyleSheetList:0,TextTrackCueList:0,TextTrackList:0,TouchList:0}},function(t,e,r){var i=r(2),n=r(8),a=r(32),o=n("iterator");t.exports=!i(function(){var t=new URL("b?e=1","http://a"),e=t.searchParams;return t.pathname="c%20d",a&&!t.toJSON||!e.sort||"http://a/c%20d?e=1"!==t.href||"1"!==e.get("e")||"a=1"!==String(new URLSearchParams("?a=1"))||!e[o]||"a"!==new URL("https://a@b").username||"b"!==new URLSearchParams(new URLSearchParams("a=b")).get("a")||"xn--e1aybc"!==new URL("http://тест").host||"#%D0%B1"!==new URL("http://a#б").hash})},function(t,e,r){"use strict";r(70);function n(e){try{return decodeURIComponent(e)}catch(t){return e}}function o(t){var e,r=t.replace(C," "),i=4;try{return decodeURIComponent(r)}catch(t){for(;i;)r=r.replace((e=i--,O[e-1]||(O[e-1]=RegExp("((?:%[\\da-f]{2}){"+e+"})","gi"))),n);return r}}function i(t){return D[t]}function a(t){return encodeURIComponent(t).replace(M,i)}function c(t,e){if(e)for(var r,i,n=e.split("&"),a=0;a<n.length;)(r=n[a++]).length&&(i=r.split("="),t.push({key:o(i.shift()),value:o(i.join("="))}))}function u(t){this.entries.length=0,c(this.entries,t)}function p(t,e){if(t<e)throw TypeError("Not enough arguments")}var s=r(1),l=r(141),h=r(17),d=r(48),f=r(27),m=r(92),g=r(20),v=r(38),y=r(12),x=r(36),b=r(5),_=r(4),w=r(364),S=r(61),T=r(8)("iterator"),P="URLSearchParams",k=P+"Iterator",I=g.set,A=g.getterFor(P),E=g.getterFor(k),C=/\+/g,O=Array(4),M=/[!'()~]|%20/g,D={"!":"%21","'":"%27","(":"%28",")":"%29","~":"%7E","%20":"+"},F=m(function(t,e){I(this,{type:k,iterator:w(A(t).entries),kind:e})},"Iterator",function(){var t=E(this),e=t.kind,r=t.iterator.next(),i=r.value;return r.done||(r.value="keys"===e?i.key:"values"===e?i.value:[i.key,i.value]),r}),R=function(t){v(this,R,P);var e,r,i,n,a,o,s,l=0<arguments.length?t:void 0,p=[];if(I(this,{type:P,entries:p,updateURL:function(){},updateSearchParams:u}),void 0!==l)if(_(l))if("function"==typeof(e=S(l)))for(r=e.call(l);!(i=r.next()).done;){if((a=(n=w(b(i.value))).next()).done||(o=n.next()).done||!n.next().done)throw TypeError("Expected sequence with length 2");p.push({key:a.value+"",value:o.value+""})}else for(s in l)y(l,s)&&p.push({key:s,value:l[s]+""});else c(p,"string"==typeof l?"?"===l.charAt(0)?l.slice(1):l:l+"")},N=R.prototype;d(N,{append:function(t,e){p(arguments.length,2);var r=A(this);r.entries.push({key:t+"",value:e+""}),r.updateURL()},delete:function(t){p(arguments.length,1);for(var e=A(this),r=e.entries,i=t+"",n=0;n<r.length;)r[n].key===i?r.splice(n,1):n++;e.updateURL()},get:function(t){p(arguments.length,1);for(var e=A(this).entries,r=t+"",i=0;i<e.length;i++)if(e[i].key===r)return e[i].value;return null},getAll:function(t){p(arguments.length,1);for(var e=A(this).entries,r=t+"",i=[],n=0;n<e.length;n++)e[n].key===r&&i.push(e[n].value);return i},has:function(t){p(arguments.length,1);for(var e=A(this).entries,r=t+"",i=0;i<e.length;)if(e[i++].key===r)return!0;return!1},set:function(t,e){p(arguments.length,1);for(var r,i=A(this),n=i.entries,a=!1,o=t+"",s=e+"",l=0;l<n.length;l++)(r=n[l]).key===o&&(a?n.splice(l--,1):(a=!0,r.value=s));a||n.push({key:o,value:s}),i.updateURL()},sort:function(){var t,e,r,i=A(this),n=i.entries,a=n.slice();for(r=n.length=0;r<a.length;r++){for(t=a[r],e=0;e<r;e++)if(n[e].key>t.key){n.splice(e,0,t);break}e===r&&n.push(t)}i.updateURL()},forEach:function(t,e){for(var r,i=A(this).entries,n=x(t,1<arguments.length?e:void 0,3),a=0;a<i.length;)n((r=i[a++]).value,r.key,this)},keys:function(){return new F(this,"keys")},values:function(){return new F(this,"values")},entries:function(){return new F(this,"entries")}},{enumerable:!0}),h(N,T,N.entries),h(N,"toString",function(){for(var t,e=A(this).entries,r=[],i=0;i<e.length;)t=e[i++],r.push(a(t.key)+"="+a(t.value));return r.join("&")},{enumerable:!0}),f(R,P),s({global:!0,forced:!l},{URLSearchParams:R}),t.exports={URLSearchParams:R,getState:A}},function(t,e){prx.utils=window.prx.utils||{}},function(t,e,r){"use strict";var i,n;Object.defineProperty(e,"__esModule",{value:!0}),(n=i||(i={})).prx1=prx,n.prxy1=prxy,e.prx=i.prx1,e.prxy=i.prxy1},function(t,e,r){"use strict";Object.defineProperty(e,"__esModule",{value:!0});var i=(n.tgIsHTMLElement=function(t){return void 0===t.length},n.tgIsElement=function(t){return void 0===t.length&&void 0===t.children},n.tgIsJQuery=function(t){return void 0!==t.ajaxError},n.getElementsArrayFromElement=function(t){if(null==t)return[];var e=[];if(n.tgIsHTMLElement(t))e.push(t);else if(n.tgIsJQuery(t))e=t.get();else for(var r=0,i=t.length;r<i;++r)e.push(t[r]);return e},n.getScrollbarWH=function(){var t=document.createElement("div");t.style.height="1px",t.style.overflow="scroll";var e=document.createElement("div");e.style.height="2px",t.appendChild(e),document.body.appendChild(t);var r=t.offsetWidth,i=e.offsetWidth;return $(t).remove(),r-i},n);function n(){}e.SuperDomHelper=i},function(t,e,r){"use strict";var i=this&&this.__importStar||function(t){if(t&&t.__esModule)return t;var e={};if(null!=t)for(var r in t)Object.hasOwnProperty.call(t,r)&&(e[r]=t[r]);return e.default=t,e},n=this&&this.__importDefault||function(t){return t&&t.__esModule?t:{default:t}};Object.defineProperty(e,"__esModule",{value:!0}),r(147),r(366);var a=i(r(367)),o=n(r(369));window.jsondiffpatch=a.create({textDiff:{minLength:99999}}),window.proto=new o.default,r(370),r(372),r(458),window.BezierEasing=r(373),window.IScroll=r(374),r(375),window.Hammer=r(376),window.propagating=r(377),r(378),r(379),r(380),window.lottie=r(381),window.DOMPurify=r(382),r(383),window.WebFont=r(385),r(386),r(387),r(388),r(389),r(390),r(391),r(392),r(393),r(394),r(143),r(395),r(397),r(398),r(399),r(400),r(401),r(402),r(403),r(404),r(405),r(406),r(407),r(408),r(409),r(410),r(411),r(412),r(413),r(414),r(415),r(416),r(417),r(418),r(419),r(420),r(421),r(422),r(423),r(424),r(425),r(426),r(427),r(428),r(429),r(430),r(431),r(432),r(433),r(434),r(435),r(436),r(437),r(143),r(438),r(439),r(440),r(441),r(442),r(443),r(444),r(445),r(448),r(450),r(453),r(454),r(455),r(456),r(457)},function(t,e,r){r(148),r(356),t.exports=r(44)},function(t,e,r){r(149),r(150),r(151),r(152),r(153),r(154),r(155),r(156),r(157),r(158),r(159),r(160),r(161),r(162),r(163),r(164),r(165),r(166),r(167),r(168),r(169),r(170),r(171),r(172),r(173),r(174),r(175),r(176),r(177),r(178),r(179),r(180),r(181),r(182),r(183),r(184),r(186),r(187),r(188),r(189),r(190),r(191),r(192),r(193),r(194),r(195),r(196),r(197),r(198),r(199),r(200),r(201),r(202),r(203),r(204),r(205),r(206),r(207),r(208),r(209),r(210),r(211),r(212),r(213),r(214),r(215),r(216),r(217),r(218),r(219),r(220),r(70),r(221),r(222),r(223),r(224),r(225),r(226),r(227),r(228),r(229),r(230),r(231),r(232),r(233),r(234),r(235),r(236),r(237),r(126),r(238),r(239),r(240),r(241),r(242),r(243),r(244),r(245),r(246),r(247),r(248),r(249),r(250),r(251),r(252),r(253),r(254),r(255),r(256),r(257),r(258),r(259),r(261),r(262),r(263),r(264),r(265),r(266),r(267),r(268),r(269),r(270),r(271),r(272),r(273),r(274),r(275),r(276),r(277),r(279),r(280),r(281),r(282),r(283),r(284),r(285),r(286),r(287),r(288),r(289),r(290),r(291),r(293),r(294),r(296),r(297),r(300),r(301),r(302),r(303),r(304),r(305),r(306),r(307),r(308),r(309),r(310),r(311),r(312),r(313),r(314),r(315),r(316),r(317),r(318),r(319),r(320),r(321),r(322),r(323),r(324),r(325),r(326),r(327),r(328),r(329),r(330),r(331),r(332),r(333),r(334),r(335),r(336),r(337),r(338),r(339),r(340),r(341),r(342),r(343),r(344),r(345),r(346),r(347),r(348),r(349),r(350),r(351),r(352),r(353),r(354),r(355),t.exports=r(44)},function(t,e,r){"use strict";function n(t,e){var r=et[t]=_(U[z]);return Y(r,{type:H,tag:t,description:e}),c||(r.description=e),r}function i(e,t){g(e);var r=y(t),i=w(r).concat(ht(r));return $(i,function(t){c&&!ut.call(r,t)||ct(e,t,r[t])}),e}function a(t,e){var r=y(t),i=x(e,!0);if(r!==W||!d(et,i)||d(rt,i)){var n=Q(r,i);return!n||!d(et,i)||d(r,V)&&r[V][i]||(n.enumerable=!0),n}}function o(t){var e=K(y(t)),r=[];return $(e,function(t){d(et,t)||d(D,t)||r.push(t)}),r}var s=r(1),l=r(3),p=r(32),c=r(7),u=r(109),h=r(2),d=r(12),f=r(41),m=r(4),g=r(5),v=r(11),y=r(19),x=r(26),b=r(39),_=r(35),w=r(56),S=r(40),T=r(111),P=r(86),k=r(16),I=r(10),A=r(65),E=r(14),C=r(17),O=r(51),M=r(66),D=r(53),F=r(52),R=r(8),N=r(112),L=r(18),B=r(27),j=r(20),$=r(13).forEach,V=M("hidden"),H="Symbol",z="prototype",G=R("toPrimitive"),Y=j.set,X=j.getterFor(H),W=Object[z],U=l.Symbol,q=l.JSON,J=q&&q.stringify,Q=k.f,Z=I.f,K=T.f,tt=A.f,et=O("symbols"),rt=O("op-symbols"),it=O("string-to-symbol-registry"),nt=O("symbol-to-string-registry"),at=O("wks"),ot=l.QObject,st=!ot||!ot[z]||!ot[z].findChild,lt=c&&h(function(){return 7!=_(Z({},"a",{get:function(){return Z(this,"a",{value:7}).a}})).a})?function(t,e,r){var i=Q(W,e);i&&delete W[e],Z(t,e,r),i&&t!==W&&Z(W,e,i)}:Z,pt=u&&"symbol"==typeof U.iterator?function(t){return"symbol"==typeof t}:function(t){return Object(t)instanceof U},ct=function(t,e,r){t===W&&ct(rt,e,r),g(t);var i=x(e,!0);return g(r),d(et,i)?(r.enumerable?(d(t,V)&&t[V][i]&&(t[V][i]=!1),r=_(r,{enumerable:b(0,!1)})):(d(t,V)||Z(t,V,b(1,{})),t[V][i]=!0),lt(t,i,r)):Z(t,i,r)},ut=function(t){var e=x(t,!0),r=tt.call(this,e);return!(this===W&&d(et,e)&&!d(rt,e))&&(!(r||!d(this,e)||!d(et,e)||d(this,V)&&this[V][e])||r)},ht=function(t){var e=t===W,r=K(e?rt:y(t)),i=[];return $(r,function(t){!d(et,t)||e&&!d(W,t)||i.push(et[t])}),i};u||(C((U=function(t){if(this instanceof U)throw TypeError("Symbol is not a constructor");var e=arguments.length&&void 0!==t?String(t):void 0,r=F(e),i=function(t){this===W&&i.call(rt,t),d(this,V)&&d(this[V],r)&&(this[V][r]=!1),lt(this,r,b(1,t))};return c&&st&&lt(W,r,{configurable:!0,set:i}),n(r,e)})[z],"toString",function(){return X(this).tag}),A.f=ut,I.f=ct,k.f=a,S.f=T.f=o,P.f=ht,c&&(Z(U[z],"description",{configurable:!0,get:function(){return X(this).description}}),p||C(W,"propertyIsEnumerable",ut,{unsafe:!0})),N.f=function(t){return n(R(t),t)}),s({global:!0,wrap:!0,forced:!u,sham:!u},{Symbol:U}),$(w(at),function(t){L(t)}),s({target:H,stat:!0,forced:!u},{for:function(t){var e=String(t);if(d(it,e))return it[e];var r=U(e);return it[e]=r,nt[r]=e,r},keyFor:function(t){if(!pt(t))throw TypeError(t+" is not a symbol");if(d(nt,t))return nt[t]},useSetter:function(){st=!0},useSimple:function(){st=!1}}),s({target:"Object",stat:!0,forced:!u,sham:!c},{create:function(t,e){return void 0===e?_(t):i(_(t),e)},defineProperty:ct,defineProperties:i,getOwnPropertyDescriptor:a}),s({target:"Object",stat:!0,forced:!u},{getOwnPropertyNames:o,getOwnPropertySymbols:ht}),s({target:"Object",stat:!0,forced:h(function(){P.f(1)})},{getOwnPropertySymbols:function(t){return P.f(v(t))}}),q&&s({target:"JSON",stat:!0,forced:!u||h(function(){var t=U();return"[null]"!=J([t])||"{}"!=J({a:t})||"{}"!=J(Object(t))})},{stringify:function(t){for(var e,r,i=[t],n=1;n<arguments.length;)i.push(arguments[n++]);if(r=e=i[1],(m(e)||void 0!==t)&&!pt(t))return f(e)||(e=function(t,e){if("function"==typeof r&&(e=r.call(this,t,e)),!pt(e))return e}),i[1]=e,J.apply(q,i)}}),U[z][G]||E(U[z],G,U[z].valueOf),B(U,H),D[V]=!0},function(t,e,r){r(18)("asyncIterator")},function(t,e,r){"use strict";var i=r(1),n=r(7),a=r(3),o=r(12),s=r(4),l=r(10).f,p=r(107),c=a.Symbol;if(n&&"function"==typeof c&&(!("description"in c.prototype)||void 0!==c().description)){var u={},h=function(t){var e=arguments.length<1||void 0===t?void 0:String(t),r=this instanceof h?new c(e):void 0===e?c():c(e);return""===e&&(u[r]=!0),r};p(h,c);var d=h.prototype=c.prototype;d.constructor=h;var f=d.toString,m="Symbol(test)"==String(c("test")),g=/^Symbol\((.*)\)[^)]+$/;l(d,"description",{configurable:!0,get:function(){var t=s(this)?this.valueOf():this,e=f.call(t);if(o(u,t))return"";var r=m?e.slice(7,-1):e.replace(g,"$1");return""===r?void 0:r}}),i({global:!0,forced:!0},{Symbol:h})}},function(t,e,r){r(18)("hasInstance")},function(t,e,r){r(18)("isConcatSpreadable")},function(t,e,r){r(18)("iterator")},function(t,e,r){r(18)("match")},function(t,e,r){r(18)("matchAll")},function(t,e,r){r(18)("replace")},function(t,e,r){r(18)("search")},function(t,e,r){r(18)("species")},function(t,e,r){r(18)("split")},function(t,e,r){r(18)("toPrimitive")},function(t,e,r){r(18)("toStringTag")},function(t,e,r){r(18)("unscopables")},function(t,e,r){var i=r(1),n=r(113);i({target:"Object",stat:!0,forced:Object.assign!==n},{assign:n})},function(t,e,r){r(1)({target:"Object",stat:!0,sham:!r(7)},{create:r(35)})},function(t,e,r){var i=r(1),n=r(7);i({target:"Object",stat:!0,forced:!n,sham:!n},{defineProperty:r(10).f})},function(t,e,r){var i=r(1),n=r(7);i({target:"Object",stat:!0,forced:!n,sham:!n},{defineProperties:r(87)})},function(t,e,r){var i=r(1),n=r(114).entries;i({target:"Object",stat:!0},{entries:function(t){return n(t)}})},function(t,e,r){var i=r(1),n=r(58),a=r(2),o=r(4),s=r(42).onFreeze,l=Object.freeze;i({target:"Object",stat:!0,forced:a(function(){l(1)}),sham:!n},{freeze:function(t){return l&&o(t)?l(s(t)):t}})},function(t,e,r){var i=r(1),n=r(59),a=r(43);i({target:"Object",stat:!0},{fromEntries:function(t){var r={};return n(t,function(t,e){a(r,t,e)},void 0,!0),r}})},function(t,e,r){var i=r(1),n=r(2),a=r(19),o=r(16).f,s=r(7),l=n(function(){o(1)});i({target:"Object",stat:!0,forced:!s||l,sham:!s},{getOwnPropertyDescriptor:function(t,e){return o(a(t),e)}})},function(t,e,r){var i=r(1),n=r(7),l=r(84),p=r(19),c=r(16),u=r(43);i({target:"Object",stat:!0,sham:!n},{getOwnPropertyDescriptors:function(t){for(var e,r,i=p(t),n=c.f,a=l(i),o={},s=0;a.length>s;)void 0!==(r=n(i,e=a[s++]))&&u(o,e,r);return o}})},function(t,e,r){var i=r(1),n=r(2),a=r(111).f;i({target:"Object",stat:!0,forced:n(function(){return!Object.getOwnPropertyNames(1)})},{getOwnPropertyNames:a})},function(t,e,r){var i=r(1),n=r(2),a=r(11),o=r(28),s=r(89);i({target:"Object",stat:!0,forced:n(function(){o(1)}),sham:!s},{getPrototypeOf:function(t){return o(a(t))}})},function(t,e,r){r(1)({target:"Object",stat:!0},{is:r(116)})},function(t,e,r){var i=r(1),n=r(2),a=r(4),o=Object.isExtensible;i({target:"Object",stat:!0,forced:n(function(){o(1)})},{isExtensible:function(t){return!!a(t)&&(!o||o(t))}})},function(t,e,r){var i=r(1),n=r(2),a=r(4),o=Object.isFrozen;i({target:"Object",stat:!0,forced:n(function(){o(1)})},{isFrozen:function(t){return!a(t)||!!o&&o(t)}})},function(t,e,r){var i=r(1),n=r(2),a=r(4),o=Object.isSealed;i({target:"Object",stat:!0,forced:n(function(){o(1)})},{isSealed:function(t){return!a(t)||!!o&&o(t)}})},function(t,e,r){var i=r(1),n=r(11),a=r(56);i({target:"Object",stat:!0,forced:r(2)(function(){a(1)})},{keys:function(t){return a(n(t))}})},function(t,e,r){var i=r(1),n=r(4),a=r(42).onFreeze,o=r(58),s=r(2),l=Object.preventExtensions;i({target:"Object",stat:!0,forced:s(function(){l(1)}),sham:!o},{preventExtensions:function(t){return l&&n(t)?l(a(t)):t}})},function(t,e,r){var i=r(1),n=r(4),a=r(42).onFreeze,o=r(58),s=r(2),l=Object.seal;i({target:"Object",stat:!0,forced:s(function(){l(1)}),sham:!o},{seal:function(t){return l&&n(t)?l(a(t)):t}})},function(t,e,r){r(1)({target:"Object",stat:!0},{setPrototypeOf:r(45)})},function(t,e,r){var i=r(1),n=r(114).values;i({target:"Object",stat:!0},{values:function(t){return n(t)}})},function(t,e,r){var i=r(17),n=r(185),a=Object.prototype;n!==a.toString&&i(a,"toString",n,{unsafe:!0})},function(t,e,r){"use strict";var i=r(62),n={};n[r(8)("toStringTag")]="z",t.exports="[object z]"!==String(n)?function(){return"[object "+i(this)+"]"}:n.toString},function(t,e,r){"use strict";var i=r(1),n=r(7),a=r(67),o=r(11),s=r(21),l=r(10);n&&i({target:"Object",proto:!0,forced:a},{__defineGetter__:function(t,e){l.f(o(this),t,{get:s(e),enumerable:!0,configurable:!0})}})},function(t,e,r){"use strict";var i=r(1),n=r(7),a=r(67),o=r(11),s=r(21),l=r(10);n&&i({target:"Object",proto:!0,forced:a},{__defineSetter__:function(t,e){l.f(o(this),t,{set:s(e),enumerable:!0,configurable:!0})}})},function(t,e,r){"use strict";var i=r(1),n=r(7),a=r(67),o=r(11),s=r(26),l=r(28),p=r(16).f;n&&i({target:"Object",proto:!0,forced:a},{__lookupGetter__:function(t){var e,r=o(this),i=s(t,!0);do{if(e=p(r,i))return e.get}while(r=l(r))}})},function(t,e,r){"use strict";var i=r(1),n=r(7),a=r(67),o=r(11),s=r(26),l=r(28),p=r(16).f;n&&i({target:"Object",proto:!0,forced:a},{__lookupSetter__:function(t){var e,r=o(this),i=s(t,!0);do{if(e=p(r,i))return e.set}while(r=l(r))}})},function(t,e,r){r(1)({target:"Function",proto:!0},{bind:r(118)})},function(t,e,r){var i=r(7),n=r(10).f,a=Function.prototype,o=a.toString,s=/^\s*function ([^ (]*)/;!i||"name"in a||n(a,"name",{configurable:!0,get:function(){try{return o.call(this).match(s)[1]}catch(t){return""}}})},function(t,e,r){"use strict";var i=r(4),n=r(10),a=r(28),o=r(8)("hasInstance"),s=Function.prototype;o in s||n.f(s,o,{value:function(t){if("function"!=typeof this||!i(t))return!1;if(!i(this.prototype))return t instanceof this;for(;t=a(t);)if(this.prototype===t)return!0;return!1}})},function(t,e,r){var i=r(1),n=r(119);i({target:"Array",stat:!0,forced:!r(68)(function(t){Array.from(t)})},{from:n})},function(t,e,r){r(1)({target:"Array",stat:!0},{isArray:r(41)})},function(t,e,r){"use strict";var i=r(1),n=r(2),a=r(43);i({target:"Array",stat:!0,forced:n(function(){function t(){}return!(Array.of.call(t)instanceof t)})},{of:function(){for(var t=0,e=arguments.length,r=new("function"==typeof this?this:Array)(e);t<e;)a(r,t,arguments[t++]);return r.length=e,r}})},function(t,e,r){"use strict";function p(t){if(!o(t))return!1;var e=t[l];return void 0!==e?!!e:a(t)}var i=r(1),n=r(2),a=r(41),o=r(4),c=r(11),u=r(9),h=r(43),d=r(57),s=r(63),l=r(8)("isConcatSpreadable"),f=9007199254740991,m="Maximum allowed index exceeded",g=!n(function(){var t=[];return t[l]=!1,t.concat()[0]!==t}),v=s("concat");i({target:"Array",proto:!0,forced:!g||!v},{concat:function(t){var e,r,i,n,a,o=c(this),s=d(o,0),l=0;for(e=-1,i=arguments.length;e<i;e++)if(p(a=-1===e?o:arguments[e])){if(n=u(a.length),f<l+n)throw TypeError(m);for(r=0;r<n;r++,l++)r in a&&h(s,l,a[r])}else{if(f<=l)throw TypeError(m);h(s,l++,a)}return s.length=l,s}})},function(t,e,r){var i=r(1),n=r(120),a=r(37);i({target:"Array",proto:!0},{copyWithin:n}),a("copyWithin")},function(t,e,r){"use strict";var i=r(1),n=r(13).every;i({target:"Array",proto:!0,forced:r(29)("every")},{every:function(t,e){return n(this,t,1<arguments.length?e:void 0)}})},function(t,e,r){var i=r(1),n=r(90),a=r(37);i({target:"Array",proto:!0},{fill:n}),a("fill")},function(t,e,r){"use strict";var i=r(1),n=r(13).filter;i({target:"Array",proto:!0,forced:!r(63)("filter")},{filter:function(t,e){return n(this,t,1<arguments.length?e:void 0)}})},function(t,e,r){"use strict";var i=r(1),n=r(13).find,a=r(37),o=!0;"find"in[]&&Array(1).find(function(){o=!1}),i({target:"Array",proto:!0,forced:o},{find:function(t,e){return n(this,t,1<arguments.length?e:void 0)}}),a("find")},function(t,e,r){"use strict";var i=r(1),n=r(13).findIndex,a=r(37),o="findIndex",s=!0;o in[]&&Array(1)[o](function(){s=!1}),i({target:"Array",proto:!0,forced:s},{findIndex:function(t,e){return n(this,t,1<arguments.length?e:void 0)}}),a(o)},function(t,e,r){"use strict";var i=r(1),a=r(121),o=r(11),s=r(9),l=r(24),p=r(57);i({target:"Array",proto:!0},{flat:function(t){var e=arguments.length?t:void 0,r=o(this),i=s(r.length),n=p(r,0);return n.length=a(n,r,r,i,0,void 0===e?1:l(e)),n}})},function(t,e,r){"use strict";var i=r(1),a=r(121),o=r(11),s=r(9),l=r(21),p=r(57);i({target:"Array",proto:!0},{flatMap:function(t,e){var r,i=o(this),n=s(i.length);return l(t),(r=p(i,0)).length=a(r,i,i,n,0,1,t,1<arguments.length?e:void 0),r}})},function(t,e,r){"use strict";var i=r(1),n=r(122);i({target:"Array",proto:!0,forced:[].forEach!=n},{forEach:n})},function(t,e,r){"use strict";var i=r(1),n=r(54).includes,a=r(37);i({target:"Array",proto:!0},{includes:function(t,e){return n(this,t,1<arguments.length?e:void 0)}}),a("includes")},function(t,e,r){"use strict";var i=r(1),n=r(54).indexOf,a=r(29),o=[].indexOf,s=!!o&&1/[1].indexOf(1,-0)<0,l=a("indexOf");i({target:"Array",proto:!0,forced:s||l},{indexOf:function(t,e){return s?o.apply(this,arguments)||0:n(this,t,1<arguments.length?e:void 0)}})},function(t,e,r){"use strict";var i=r(1),n=r(50),a=r(19),o=r(29),s=[].join,l=n!=Object,p=o("join",",");i({target:"Array",proto:!0,forced:l||p},{join:function(t){return s.call(a(this),void 0===t?",":t)}})},function(t,e,r){var i=r(1),n=r(123);i({target:"Array",proto:!0,forced:n!==[].lastIndexOf},{lastIndexOf:n})},function(t,e,r){"use strict";var i=r(1),n=r(13).map;i({target:"Array",proto:!0,forced:!r(63)("map")},{map:function(t,e){return n(this,t,1<arguments.length?e:void 0)}})},function(t,e,r){"use strict";var i=r(1),n=r(69).left;i({target:"Array",proto:!0,forced:r(29)("reduce")},{reduce:function(t,e){return n(this,t,arguments.length,1<arguments.length?e:void 0)}})},function(t,e,r){"use strict";var i=r(1),n=r(69).right;i({target:"Array",proto:!0,forced:r(29)("reduceRight")},{reduceRight:function(t,e){return n(this,t,arguments.length,1<arguments.length?e:void 0)}})},function(t,e,r){"use strict";var i=r(1),n=r(41),a=[].reverse,o=[1,2];i({target:"Array",proto:!0,forced:String(o)===String(o.reverse())},{reverse:function(){return n(this)&&(this.length=this.length),a.call(this)}})},function(t,e,r){"use strict";var i=r(1),p=r(4),c=r(41),u=r(34),h=r(9),d=r(19),f=r(43),n=r(63),m=r(8)("species"),g=[].slice,v=Math.max;i({target:"Array",proto:!0,forced:!n("slice")},{slice:function(t,e){var r,i,n,a=d(this),o=h(a.length),s=u(t,o),l=u(void 0===e?o:e,o);if(c(a)&&("function"!=typeof(r=a.constructor)||r!==Array&&!c(r.prototype)?p(r)&&null===(r=r[m])&&(r=void 0):r=void 0,r===Array||void 0===r))return g.call(a,s,l);for(i=new(void 0===r?Array:r)(v(l-s,0)),n=0;s<l;s++,n++)s in a&&f(i,n,a[s]);return i.length=n,i}})},function(t,e,r){"use strict";var i=r(1),n=r(13).some;i({target:"Array",proto:!0,forced:r(29)("some")},{some:function(t,e){return n(this,t,1<arguments.length?e:void 0)}})},function(t,e,r){"use strict";var i=r(1),n=r(21),a=r(11),o=r(2),s=r(29),l=[].sort,p=[1,2,3],c=o(function(){p.sort(void 0)}),u=o(function(){p.sort(null)}),h=s("sort");i({target:"Array",proto:!0,forced:c||!u||h},{sort:function(t){return void 0===t?l.call(a(this)):l.call(a(this),n(t))}})},function(t,e,r){"use strict";var i=r(1),h=r(34),d=r(24),f=r(9),m=r(11),g=r(57),v=r(43),n=r(63),y=Math.max,x=Math.min;i({target:"Array",proto:!0,forced:!n("splice")},{splice:function(t,e){var r,i,n,a,o,s,l=m(this),p=f(l.length),c=h(t,p),u=arguments.length;if(0===u?r=i=0:i=1===u?(r=0,p-c):(r=u-2,x(y(d(e),0),p-c)),9007199254740991<p+r-i)throw TypeError("Maximum allowed length exceeded");for(n=g(l,i),a=0;a<i;a++)(o=c+a)in l&&v(n,a,l[o]);if(r<(n.length=i)){for(a=c;a<p-i;a++)s=a+r,(o=a+i)in l?l[s]=l[o]:delete l[s];for(a=p;p-i+r<a;a--)delete l[a-1]}else if(i<r)for(a=p-i;c<a;a--)s=a+r-1,(o=a+i-1)in l?l[s]=l[o]:delete l[s];for(a=0;a<r;a++)l[a+c]=arguments[a+2];return l.length=p-i+r,n}})},function(t,e,r){r(46)("Array")},function(t,e,r){r(37)("flat")},function(t,e,r){r(37)("flatMap")},function(t,e,r){var i=r(1),a=r(34),o=String.fromCharCode,n=String.fromCodePoint;i({target:"String",stat:!0,forced:!!n&&1!=n.length},{fromCodePoint:function(t){for(var e,r=[],i=arguments.length,n=0;n<i;){if(e=+arguments[n++],a(e,1114111)!==e)throw RangeError(e+" is not a valid code point");r.push(e<65536?o(e):o(55296+((e-=65536)>>10),e%1024+56320))}return r.join("")}})},function(t,e,r){var i=r(1),o=r(19),s=r(9);i({target:"String",stat:!0},{raw:function(t){for(var e=o(t.raw),r=s(e.length),i=arguments.length,n=[],a=0;a<r;)n.push(String(e[a++])),a<i&&n.push(String(arguments[a]));return n.join("")}})},function(t,e,r){"use strict";var i=r(1),n=r(71).codeAt;i({target:"String",proto:!0},{codePointAt:function(t){return n(this,t)}})},function(t,e,r){"use strict";var i=r(1),s=r(9),l=r(93),p=r(15),n=r(95),c="".endsWith,u=Math.min;i({target:"String",proto:!0,forced:!n("endsWith")},{endsWith:function(t,e){var r=String(p(this));l(t);var i=1<arguments.length?e:void 0,n=s(r.length),a=void 0===i?n:u(s(i),n),o=String(t);return c?c.call(r,o,a):r.slice(a-o.length,a)===o}})},function(t,e,r){"use strict";var i=r(1),n=r(93),a=r(15);i({target:"String",proto:!0,forced:!r(95)("includes")},{includes:function(t,e){return!!~String(a(this)).indexOf(n(t),1<arguments.length?e:void 0)}})},function(t,e,r){"use strict";var i=r(72),u=r(5),h=r(9),n=r(15),d=r(74),f=r(75);i("match",1,function(i,p,c){return[function(t){var e=n(this),r=null==t?void 0:t[i];return void 0!==r?r.call(t,e):new RegExp(t)[i](String(e))},function(t){var e=c(p,t,this);if(e.done)return e.value;var r=u(t),i=String(this);if(!r.global)return f(r,i);for(var n,a=r.unicode,o=[],s=r.lastIndex=0;null!==(n=f(r,i));){var l=String(n[0]);""===(o[s]=l)&&(r.lastIndex=d(i,h(r.lastIndex),a)),s++}return 0===s?null:o}]})},function(t,e,r){"use strict";function a(t){var e,r,i,n,a,o,s=c(this),l=String(t);return e=f(s,RegExp),void 0===(r=s.flags)&&s instanceof RegExp&&!("flags"in S)&&(r=u.call(s)),i=void 0===r?"":String(r),n=new e(e===RegExp?s.source:s,i),a=!!~i.indexOf("g"),o=!!~i.indexOf("u"),n.lastIndex=p(s.lastIndex),new P(n,l,a,o)}var i=r(1),n=r(92),o=r(15),p=r(9),s=r(21),c=r(5),l=r(62),u=r(64),h=r(14),d=r(8),f=r(30),m=r(74),g=r(20),v=r(32),y=d("matchAll"),x="RegExp String",b=x+" Iterator",_=g.set,w=g.getterFor(b),S=RegExp.prototype,T=S.exec,P=n(function(t,e,r,i){_(this,{type:b,regexp:t,string:e,global:r,unicode:i,done:!1})},x,function(){var t=w(this);if(t.done)return{value:void 0,done:!0};var e=t.regexp,r=t.string,i=function(t,e){var r,i=t.exec;if("function"!=typeof i)return T.call(t,e);if("object"!=typeof(r=i.call(t,e)))throw TypeError("Incorrect exec result");return r}(e,r);return null===i?{value:void 0,done:t.done=!0}:t.global?(""==String(i[0])&&(e.lastIndex=m(r,p(e.lastIndex),t.unicode)),{value:i,done:!1}):{value:i,done:!(t.done=!0)}});i({target:"String",proto:!0},{matchAll:function(t){var e,r,i,n=o(this);return null!=t&&(void 0===(r=t[y])&&v&&"RegExp"==l(t)&&(r=a),null!=r)?s(r).call(t,n):(e=String(n),i=new RegExp(t,"g"),v?a.call(i,e):i[y](e))}}),v||y in S||h(S,y,a)},function(t,e,r){"use strict";var i=r(1),n=r(96).end;i({target:"String",proto:!0,forced:r(125)},{padEnd:function(t,e){return n(this,t,1<arguments.length?e:void 0)}})},function(t,e,r){"use strict";var i=r(1),n=r(96).start;i({target:"String",proto:!0,forced:r(125)},{padStart:function(t,e){return n(this,t,1<arguments.length?e:void 0)}})},function(t,e,r){r(1)({target:"String",proto:!0},{repeat:r(97)})},function(t,e,r){"use strict";var i=r(72),T=r(5),h=r(11),P=r(9),k=r(24),a=r(15),I=r(74),A=r(75),E=Math.max,C=Math.min,d=Math.floor,f=/\$([$&'`]|\d\d?|<[^>]*>)/g,m=/\$([$&'`]|\d\d?)/g;i("replace",2,function(n,_,w){return[function(t,e){var r=a(this),i=null==t?void 0:t[n];return void 0!==i?i.call(t,r,e):_.call(String(r),t,e)},function(t,e){var r=w(_,t,this,e);if(r.done)return r.value;var i=T(t),n=String(this),a="function"==typeof e;a||(e=String(e));var o=i.global;if(o){var s=i.unicode;i.lastIndex=0}for(var l=[];;){var p=A(i,n);if(null===p)break;if(l.push(p),!o)break;""===String(p[0])&&(i.lastIndex=I(n,P(i.lastIndex),s))}for(var c,u="",h=0,d=0;d<l.length;d++){p=l[d];for(var f=String(p[0]),m=E(C(k(p.index),n.length),0),g=[],v=1;v<p.length;v++)g.push(void 0===(c=p[v])?c:String(c));var y=p.groups;if(a){var x=[f].concat(g,m,n);void 0!==y&&x.push(y);var b=String(e.apply(void 0,x))}else b=S(f,n,m,g,y,e);h<=m&&(u+=n.slice(h,m)+b,h=m+f.length)}return u+n.slice(h)}];function S(a,o,s,l,p,t){var c=s+a.length,u=l.length,e=m;return void 0!==p&&(p=h(p),e=f),_.call(t,e,function(t,e){var r;switch(e.charAt(0)){case"$":return"$";case"&":return a;case"`":return o.slice(0,s);case"'":return o.slice(c);case"<":r=p[e.slice(1,-1)];break;default:var i=+e;if(0==i)return t;if(u<i){var n=d(i/10);return 0===n?t:n<=u?void 0===l[n-1]?e.charAt(1):l[n-1]+e.charAt(1):t}r=l[i-1]}return void 0===r?"":r})}})},function(t,e,r){"use strict";var i=r(72),l=r(5),n=r(15),p=r(116),c=r(75);i("search",1,function(i,o,s){return[function(t){var e=n(this),r=null==t?void 0:t[i];return void 0!==r?r.call(t,e):new RegExp(t)[i](String(e))},function(t){var e=s(o,t,this);if(e.done)return e.value;var r=l(t),i=String(this),n=r.lastIndex;p(n,0)||(r.lastIndex=0);var a=c(r,i);return p(r.lastIndex,n)||(r.lastIndex=n),null===a?-1:a.index}]})},function(t,e,r){"use strict";var i=r(72),u=r(94),x=r(5),h=r(15),b=r(30),_=r(74),w=r(9),S=r(75),d=r(73),n=r(2),f=[].push,T=Math.min,P=4294967295,k=!n(function(){return!RegExp(P,"y")});i("split",2,function(n,g,v){var y;return y="c"=="abbc".split(/(b)*/)[1]||4!="test".split(/(?:)/,-1).length||2!="ab".split(/(?:ab)*/).length||4!=".".split(/(.?)(.?)/).length||1<".".split(/()()/).length||"".split(/.?/).length?function(t,e){var r=String(h(this)),i=void 0===e?P:e>>>0;if(0==i)return[];if(void 0===t)return[r];if(!u(t))return g.call(r,t,i);for(var n,a,o,s=[],l=(t.ignoreCase?"i":"")+(t.multiline?"m":"")+(t.unicode?"u":"")+(t.sticky?"y":""),p=0,c=new RegExp(t.source,l+"g");(n=d.call(c,r))&&!(p<(a=c.lastIndex)&&(s.push(r.slice(p,n.index)),1<n.length&&n.index<r.length&&f.apply(s,n.slice(1)),o=n[0].length,p=a,s.length>=i));)c.lastIndex===n.index&&c.lastIndex++;return p===r.length?!o&&c.test("")||s.push(""):s.push(r.slice(p)),s.length>i?s.slice(0,i):s}:"0".split(void 0,0).length?function(t,e){return void 0===t&&0===e?[]:g.call(this,t,e)}:g,[function(t,e){var r=h(this),i=null==t?void 0:t[n];return void 0!==i?i.call(t,r,e):y.call(String(r),t,e)},function(t,e){var r=v(y,t,this,e,y!==g);if(r.done)return r.value;var i=x(t),n=String(this),a=b(i,RegExp),o=i.unicode,s=(i.ignoreCase?"i":"")+(i.multiline?"m":"")+(i.unicode?"u":"")+(k?"y":"g"),l=new a(k?i:"^(?:"+i.source+")",s),p=void 0===e?P:e>>>0;if(0==p)return[];if(0===n.length)return null===S(l,n)?[n]:[];for(var c=0,u=0,h=[];u<n.length;){l.lastIndex=k?u:0;var d,f=S(l,k?n:n.slice(u));if(null===f||(d=T(w(l.lastIndex+(k?0:u)),n.length))===c)u=_(n,u,o);else{if(h.push(n.slice(c,u)),h.length===p)return h;for(var m=1;m<=f.length-1;m++)if(h.push(f[m]),h.length===p)return h;u=c=d}}return h.push(n.slice(c)),h}]},!k)},function(t,e,r){"use strict";var i=r(1),a=r(9),o=r(93),s=r(15),n=r(95),l="".startsWith,p=Math.min;i({target:"String",proto:!0,forced:!n("startsWith")},{startsWith:function(t,e){var r=String(s(this));o(t);var i=a(p(1<arguments.length?e:void 0,r.length)),n=String(t);return l?l.call(r,n,i):r.slice(i,i+n.length)===n}})},function(t,e,r){"use strict";var i=r(1),n=r(47).trim;i({target:"String",proto:!0,forced:r(98)("trim")},{trim:function(){return n(this)}})},function(t,e,r){"use strict";var i=r(1),n=r(47).start,a=r(98)("trimStart"),o=a?function(){return n(this)}:"".trimStart;i({target:"String",proto:!0,forced:a},{trimStart:o,trimLeft:o})},function(t,e,r){"use strict";var i=r(1),n=r(47).end,a=r(98)("trimEnd"),o=a?function(){return n(this)}:"".trimEnd;i({target:"String",proto:!0,forced:a},{trimEnd:o,trimRight:o})},function(t,e,r){"use strict";var i=r(1),n=r(22);i({target:"String",proto:!0,forced:r(23)("anchor")},{anchor:function(t){return n(this,"a","name",t)}})},function(t,e,r){"use strict";var i=r(1),n=r(22);i({target:"String",proto:!0,forced:r(23)("big")},{big:function(){return n(this,"big","","")}})},function(t,e,r){"use strict";var i=r(1),n=r(22);i({target:"String",proto:!0,forced:r(23)("blink")},{blink:function(){return n(this,"blink","","")}})},function(t,e,r){"use strict";var i=r(1),n=r(22);i({target:"String",proto:!0,forced:r(23)("bold")},{bold:function(){return n(this,"b","","")}})},function(t,e,r){"use strict";var i=r(1),n=r(22);i({target:"String",proto:!0,forced:r(23)("fixed")},{fixed:function(){return n(this,"tt","","")}})},function(t,e,r){"use strict";var i=r(1),n=r(22);i({target:"String",proto:!0,forced:r(23)("fontcolor")},{fontcolor:function(t){return n(this,"font","color",t)}})},function(t,e,r){"use strict";var i=r(1),n=r(22);i({target:"String",proto:!0,forced:r(23)("fontsize")},{fontsize:function(t){return n(this,"font","size",t)}})},function(t,e,r){"use strict";var i=r(1),n=r(22);i({target:"String",proto:!0,forced:r(23)("italics")},{italics:function(){return n(this,"i","","")}})},function(t,e,r){"use strict";var i=r(1),n=r(22);i({target:"String",proto:!0,forced:r(23)("link")},{link:function(t){return n(this,"a","href",t)}})},function(t,e,r){"use strict";var i=r(1),n=r(22);i({target:"String",proto:!0,forced:r(23)("small")},{small:function(){return n(this,"small","","")}})},function(t,e,r){"use strict";var i=r(1),n=r(22);i({target:"String",proto:!0,forced:r(23)("strike")},{strike:function(){return n(this,"strike","","")}})},function(t,e,r){"use strict";var i=r(1),n=r(22);i({target:"String",proto:!0,forced:r(23)("sub")},{sub:function(){return n(this,"sub","","")}})},function(t,e,r){"use strict";var i=r(1),n=r(22);i({target:"String",proto:!0,forced:r(23)("sup")},{sup:function(){return n(this,"sup","","")}})},function(t,e,r){var i=r(7),n=r(3),a=r(55),o=r(99),s=r(10).f,l=r(40).f,p=r(94),c=r(64),u=r(17),h=r(2),d=r(46),f=r(8)("match"),m=n.RegExp,g=m.prototype,v=/a/g,y=/a/g,x=new m(v)!==v;if(i&&a("RegExp",!x||h(function(){return y[f]=!1,m(v)!=v||m(y)==y||"/a/i"!=m(v,"i")}))){function b(e){e in _||s(_,e,{configurable:!0,get:function(){return m[e]},set:function(t){m[e]=t}})}for(var _=function(t,e){var r=this instanceof _,i=p(t),n=void 0===e;return!r&&i&&t.constructor===_&&n?t:o(x?new m(i&&!n?t.source:t,e):m((i=t instanceof _)?t.source:t,i&&n?c.call(t):e),r?this:g,_)},w=l(m),S=0;w.length>S;)b(w[S++]);(g.constructor=_).prototype=g,u(n,"RegExp",_)}d("RegExp")},function(t,e,r){"use strict";var i=r(1),n=r(73);i({target:"RegExp",proto:!0,forced:/./.exec!==n},{exec:n})},function(t,e,r){var i=r(7),n=r(10),a=r(64);i&&"g"!=/./g.flags&&n.f(RegExp.prototype,"flags",{configurable:!0,get:a})},function(t,e,r){"use strict";var i=r(17),n=r(5),a=r(2),o=r(64),s="toString",l=RegExp.prototype,p=l[s],c=a(function(){return"/a/b"!=p.call({source:"a",flags:"b"})}),u=p.name!=s;(c||u)&&i(RegExp.prototype,s,function(){var t=n(this),e=String(t.source),r=t.flags;return"/"+e+"/"+String(void 0===r&&t instanceof RegExp&&!("flags"in l)?o.call(t):r)},{unsafe:!0})},function(t,e,r){var i=r(1),n=r(127);i({global:!0,forced:parseInt!=n},{parseInt:n})},function(t,e,r){var i=r(1),n=r(128);i({global:!0,forced:parseFloat!=n},{parseFloat:n})},function(t,e,r){"use strict";function i(t){var e,r,i,n,a,o,s,l,p=u(t,!1);if("string"==typeof p&&2<p.length)if(43===(e=(p=v(p)).charCodeAt(0))||45===e){if(88===(r=p.charCodeAt(2))||120===r)return NaN}else if(48===e){switch(p.charCodeAt(1)){case 66:case 98:i=2,n=49;break;case 79:case 111:i=8,n=55;break;default:return+p}for(o=(a=p.slice(2)).length,s=0;s<o;s++)if((l=a.charCodeAt(s))<48||n<l)return NaN;return parseInt(a,i)}return+p}var n=r(7),a=r(3),o=r(55),s=r(17),l=r(12),p=r(25),c=r(99),u=r(26),h=r(2),d=r(35),f=r(40).f,m=r(16).f,g=r(10).f,v=r(47).trim,y="Number",x=a[y],b=x.prototype,_=p(d(b))==y;if(o(y,!x(" 0o1")||!x("0b1")||x("+0x1"))){for(var w,S=function(t){var e=arguments.length<1?0:t,r=this;return r instanceof S&&(_?h(function(){b.valueOf.call(r)}):p(r)!=y)?c(new x(i(e)),r,S):i(e)},T=n?f(x):"MAX_VALUE,MIN_VALUE,NaN,NEGATIVE_INFINITY,POSITIVE_INFINITY,EPSILON,isFinite,isInteger,isNaN,isSafeInteger,MAX_SAFE_INTEGER,MIN_SAFE_INTEGER,parseFloat,parseInt,isInteger".split(","),P=0;T.length>P;P++)l(x,w=T[P])&&!l(S,w)&&g(S,w,m(x,w));(S.prototype=b).constructor=S,s(a,y,S)}},function(t,e,r){r(1)({target:"Number",stat:!0},{EPSILON:Math.pow(2,-52)})},function(t,e,r){r(1)({target:"Number",stat:!0},{isFinite:r(260)})},function(t,e,r){var i=r(3).isFinite;t.exports=Number.isFinite||function(t){return"number"==typeof t&&i(t)}},function(t,e,r){r(1)({target:"Number",stat:!0},{isInteger:r(129)})},function(t,e,r){r(1)({target:"Number",stat:!0},{isNaN:function(t){return t!=t}})},function(t,e,r){var i=r(1),n=r(129),a=Math.abs;i({target:"Number",stat:!0},{isSafeInteger:function(t){return n(t)&&a(t)<=9007199254740991}})},function(t,e,r){r(1)({target:"Number",stat:!0},{MAX_SAFE_INTEGER:9007199254740991})},function(t,e,r){r(1)({target:"Number",stat:!0},{MIN_SAFE_INTEGER:-9007199254740991})},function(t,e,r){var i=r(1),n=r(128);i({target:"Number",stat:!0,forced:Number.parseFloat!=n},{parseFloat:n})},function(t,e,r){var i=r(1),n=r(127);i({target:"Number",stat:!0,forced:Number.parseInt!=n},{parseInt:n})},function(t,e,r){"use strict";var i=r(1),d=r(24),f=r(130),m=r(97),n=r(2),a=1..toFixed,g=Math.floor,v=function(t,e,r){return 0===e?r:e%2==1?v(t,e-1,r*t):v(t*t,e/2,r)};i({target:"Number",proto:!0,forced:a&&("0.000"!==8e-5.toFixed(3)||"1"!==.9.toFixed(0)||"1.25"!==1.255.toFixed(2)||"1000000000000000128"!==(0xde0b6b3a7640080).toFixed(0))||!n(function(){a.call({})})},{toFixed:function(t){function e(t,e){for(var r=-1,i=e;++r<6;)i+=t*c[r],c[r]=i%1e7,i=g(i/1e7)}function r(t){for(var e=6,r=0;0<=--e;)r+=c[e],c[e]=g(r/t),r=r%t*1e7}function i(){for(var t=6,e="";0<=--t;)if(""!==e||0===t||0!==c[t]){var r=String(c[t]);e=""===e?r:e+m.call("0",7-r.length)+r}return e}var n,a,o,s,l=f(this),p=d(t),c=[0,0,0,0,0,0],u="",h="0";if(p<0||20<p)throw RangeError("Incorrect fraction digits");if(l!=l)return"NaN";if(l<=-1e21||1e21<=l)return String(l);if(l<0&&(u="-",l=-l),1e-21<l)if(a=(n=function(t){for(var e=0,r=t;4096<=r;)e+=12,r/=4096;for(;2<=r;)e+=1,r/=2;return e}(l*v(2,69,1))-69)<0?l*v(2,-n,1):l/v(2,n,1),a*=4503599627370496,0<(n=52-n)){for(e(0,a),o=p;7<=o;)e(1e7,0),o-=7;for(e(v(10,o,1),0),o=n-1;23<=o;)r(1<<23),o-=23;r(1<<o),e(1,1),r(2),h=i()}else e(0,a),e(1<<-n,0),h=i()+m.call("0",p);return h=0<p?u+((s=h.length)<=p?"0."+m.call("0",p-s)+h:h.slice(0,s-p)+"."+h.slice(s-p)):u+h}})},function(t,e,r){"use strict";var i=r(1),n=r(2),a=r(130),o=1..toPrecision;i({target:"Number",proto:!0,forced:n(function(){return"1"!==o.call(1,void 0)})||!n(function(){o.call({})})},{toPrecision:function(t){return void 0===t?o.call(a(this)):o.call(a(this),t)}})},function(t,e,r){var i=r(1),n=r(131),a=Math.acosh,o=Math.log,s=Math.sqrt,l=Math.LN2;i({target:"Math",stat:!0,forced:!a||710!=Math.floor(a(Number.MAX_VALUE))||a(1/0)!=1/0},{acosh:function(t){return(t=+t)<1?NaN:94906265.62425156<t?o(t)+l:n(t-1+s(t-1)*s(t+1))}})},function(t,e,r){var i=r(1),n=Math.asinh,a=Math.log,o=Math.sqrt;i({target:"Math",stat:!0,forced:!(n&&0<1/n(0))},{asinh:function t(e){return isFinite(e=+e)&&0!=e?e<0?-t(-e):a(e+o(e*e+1)):e}})},function(t,e,r){var i=r(1),n=Math.atanh,a=Math.log;i({target:"Math",stat:!0,forced:!(n&&1/n(-0)<0)},{atanh:function(t){return 0==(t=+t)?t:a((1+t)/(1-t))/2}})},function(t,e,r){var i=r(1),n=r(100),a=Math.abs,o=Math.pow;i({target:"Math",stat:!0},{cbrt:function(t){return n(t=+t)*o(a(t),1/3)}})},function(t,e,r){var i=r(1),n=Math.floor,a=Math.log,o=Math.LOG2E;i({target:"Math",stat:!0},{clz32:function(t){return(t>>>=0)?31-n(a(t+.5)*o):32}})},function(t,e,r){var i=r(1),n=r(78),a=Math.cosh,o=Math.abs,s=Math.E;i({target:"Math",stat:!0,forced:!a||a(710)===1/0},{cosh:function(t){var e=n(o(t)-1)+1;return(e+1/(e*s*s))*(s/2)}})},function(t,e,r){var i=r(1),n=r(78);i({target:"Math",stat:!0,forced:n!=Math.expm1},{expm1:n})},function(t,e,r){r(1)({target:"Math",stat:!0},{fround:r(278)})},function(t,e,r){var a=r(100),o=Math.abs,i=Math.pow,s=i(2,-52),l=i(2,-23),p=i(2,127)*(2-l),c=i(2,-126);t.exports=Math.fround||function(t){var e,r,i=o(t),n=a(t);return i<c?n*function(t){return t+1/s-1/s}(i/c/l)*c*l:p<(r=(e=(1+l/s)*i)-(e-i))||r!=r?n*(1/0):n*r}},function(t,e,r){var i=r(1),l=Math.abs,p=Math.sqrt;i({target:"Math",stat:!0},{hypot:function(t,e){for(var r,i,n=0,a=0,o=arguments.length,s=0;a<o;)s<(r=l(arguments[a++]))?(n=n*(i=s/r)*i+1,s=r):n+=0<r?(i=r/s)*i:r;return s===1/0?1/0:s*p(n)}})},function(t,e,r){var i=r(1),n=r(2),a=Math.imul;i({target:"Math",stat:!0,forced:n(function(){return-5!=a(4294967295,5)||2!=a.length})},{imul:function(t,e){var r=+t,i=+e,n=65535&r,a=65535&i;return 0|n*a+((65535&r>>>16)*a+n*(65535&i>>>16)<<16>>>0)}})},function(t,e,r){var i=r(1),n=Math.log,a=Math.LOG10E;i({target:"Math",stat:!0},{log10:function(t){return n(t)*a}})},function(t,e,r){r(1)({target:"Math",stat:!0},{log1p:r(131)})},function(t,e,r){var i=r(1),n=Math.log,a=Math.LN2;i({target:"Math",stat:!0},{log2:function(t){return n(t)/a}})},function(t,e,r){r(1)({target:"Math",stat:!0},{sign:r(100)})},function(t,e,r){var i=r(1),n=r(2),a=r(78),o=Math.abs,s=Math.exp,l=Math.E;i({target:"Math",stat:!0,forced:n(function(){return-2e-17!=Math.sinh(-2e-17)})},{sinh:function(t){return o(t=+t)<1?(a(t)-a(-t))/2:(s(t-1)-s(-t-1))*(l/2)}})},function(t,e,r){var i=r(1),n=r(78),a=Math.exp;i({target:"Math",stat:!0},{tanh:function(t){var e=n(t=+t),r=n(-t);return e==1/0?1:r==1/0?-1:(e-r)/(a(t)+a(-t))}})},function(t,e,r){r(27)(Math,"Math",!0)},function(t,e,r){var i=r(1),n=Math.ceil,a=Math.floor;i({target:"Math",stat:!0},{trunc:function(t){return(0<t?a:n)(t)}})},function(t,e,r){r(1)({target:"Date",stat:!0},{now:function(){return(new Date).getTime()}})},function(t,e,r){"use strict";var i=r(1),n=r(2),a=r(11),o=r(26);i({target:"Date",proto:!0,forced:n(function(){return null!==new Date(NaN).toJSON()||1!==Date.prototype.toJSON.call({toISOString:function(){return 1}})})},{toJSON:function(t){var e=a(this),r=o(e);return"number"!=typeof r||isFinite(r)?e.toISOString():null}})},function(t,e,r){var i=r(1),n=r(292);i({target:"Date",proto:!0,forced:Date.prototype.toISOString!==n},{toISOString:n})},function(t,e,r){"use strict";var i=r(2),n=r(96).start,a=Math.abs,o=Date.prototype,s=o.getTime,l=o.toISOString;t.exports=i(function(){return"0385-07-25T07:06:39.999Z"!=l.call(new Date(-5e13-1))})||!i(function(){l.call(new Date(NaN))})?function(){if(!isFinite(s.call(this)))throw RangeError("Invalid time value");var t=this.getUTCFullYear(),e=this.getUTCMilliseconds(),r=t<0?"-":9999<t?"+":"";return r+n(a(t),r?6:4,0)+"-"+n(this.getUTCMonth()+1,2,0)+"-"+n(this.getUTCDate(),2,0)+"T"+n(this.getUTCHours(),2,0)+":"+n(this.getUTCMinutes(),2,0)+":"+n(this.getUTCSeconds(),2,0)+"."+n(e,3,0)+"Z"}:l},function(t,e,r){var i=r(17),n=Date.prototype,a="Invalid Date",o=n.toString,s=n.getTime;new Date(NaN)+""!=a&&i(n,"toString",function(){var t=s.call(this);return t==t?o.call(this):a})},function(t,e,r){var i=r(14),n=r(295),a=r(8)("toPrimitive"),o=Date.prototype;a in o||i(o,a,n)},function(t,e,r){"use strict";var i=r(5),n=r(26);t.exports=function(t){if("string"!==t&&"number"!==t&&"default"!==t)throw TypeError("Incorrect hint");return n(i(this),"number"!==t)}},function(t,e,r){var i=r(3);r(27)(i.JSON,"JSON",!0)},function(t,e,r){"use strict";function m(t){var e;return!(!y(t)||"function"!=typeof(e=t.then))&&e}function a(u,h,d){if(!h.notified){h.notified=!0;var f=h.reactions;k(function(){for(var t=h.value,e=1==h.state,r=0;f.length>r;){var i,n,a,o=f[r++],s=e?o.ok:o.fail,l=o.resolve,p=o.reject,c=o.domain;try{s?(e||(2===h.rejection&&et(u,h),h.rejection=1),!0===s?i=t:(c&&c.enter(),i=s(t),c&&(c.exit(),a=!0)),i===o.promise?p($("Promise-chain cycle")):(n=m(i))?n.call(i,l,p):l(i)):p(t)}catch(t){c&&!a&&c.exit(),p(t)}}h.reactions=[],h.notified=!1,d&&!h.rejection&&K(u,h)})}}function n(t,e,r){var i,n;q?((i=V.createEvent("Event")).promise=e,i.reason=r,i.initEvent(t,!1,!0),h.dispatchEvent(i)):i={promise:e,reason:r},(n=h["on"+t])?n(i):t===J&&A("Unhandled promise rejection",r)}function o(e,r,i,n){return function(t){e(r,i,t,n)}}function s(t,e,r,i){e.done||(e.done=!0,i&&(e=i),e.value=r,e.state=2,a(t,e,!0))}var i,l,p,c=r(1),u=r(32),h=r(3),d=r(44),f=r(48),g=r(27),v=r(46),y=r(4),x=r(21),b=r(38),_=r(25),w=r(59),S=r(68),T=r(30),P=r(101).set,k=r(132),I=r(133),A=r(298),E=r(134),C=r(299),O=r(76),M=r(20),D=r(55),F=r(8)("species"),R="Promise",N=M.get,L=M.set,B=M.getterFor(R),j=h[R],$=h.TypeError,V=h.document,H=h.process,z=h.fetch,G=H&&H.versions,Y=G&&G.v8||"",X=E.f,W=X,U="process"==_(H),q=!!(V&&V.createEvent&&h.dispatchEvent),J="unhandledrejection",Q=D(R,function(){function e(){}var t=j.resolve(1),r=(t.constructor={})[F]=function(t){t(e,e)};return!((U||"function"==typeof PromiseRejectionEvent)&&(!u||t.finally)&&t.then(e)instanceof r&&0!==Y.indexOf("6.6")&&-1===O.indexOf("Chrome/66"))}),Z=Q||!S(function(t){j.all(t).catch(function(){})}),K=function(r,i){P.call(h,function(){var t,e=i.value;if(tt(i)&&(t=C(function(){U?H.emit("unhandledRejection",e,r):n(J,r,e)}),i.rejection=U||tt(i)?2:1,t.error))throw t.value})},tt=function(t){return 1!==t.rejection&&!t.parent},et=function(t,e){P.call(h,function(){U?H.emit("rejectionHandled",t):n("rejectionhandled",t,e.value)})},rt=function(r,i,t,e){if(!i.done){i.done=!0,e&&(i=e);try{if(r===t)throw $("Promise can't be resolved itself");var n=m(t);n?k(function(){var e={done:!1};try{n.call(t,o(rt,r,e,i),o(s,r,e,i))}catch(t){s(r,e,t,i)}}):(i.value=t,i.state=1,a(r,i,!1))}catch(t){s(r,{done:!1},t,i)}}};Q&&(j=function(t){b(this,j,R),x(t),i.call(this);var e=N(this);try{t(o(rt,this,e),o(s,this,e))}catch(t){s(this,e,t)}},(i=function(t){L(this,{type:R,done:!1,notified:!1,parent:!1,reactions:[],rejection:!1,state:0,value:void 0})}).prototype=f(j.prototype,{then:function(t,e){var r=B(this),i=X(T(this,j));return i.ok="function"!=typeof t||t,i.fail="function"==typeof e&&e,i.domain=U?H.domain:void 0,r.parent=!0,r.reactions.push(i),0!=r.state&&a(this,r,!1),i.promise},catch:function(t){return this.then(void 0,t)}}),l=function(){var t=new i,e=N(t);this.promise=t,this.resolve=o(rt,t,e),this.reject=o(s,t,e)},E.f=X=function(t){return t===j||t===p?new l(t):W(t)},u||"function"!=typeof z||c({global:!0,enumerable:!0,forced:!0},{fetch:function(t){return I(j,z.apply(h,arguments))}})),c({global:!0,wrap:!0,forced:Q},{Promise:j}),g(j,R,!1,!0),v(R),p=d[R],c({target:R,stat:!0,forced:Q},{reject:function(t){var e=X(this);return e.reject.call(void 0,t),e.promise}}),c({target:R,stat:!0,forced:u||Q},{resolve:function(t){return I(u&&this===p?j:this,t)}}),c({target:R,stat:!0,forced:Z},{all:function(t){var s=this,e=X(s),l=e.resolve,p=e.reject,r=C(function(){var i=x(s.resolve),n=[],a=0,o=1;w(t,function(t){var e=a++,r=!1;n.push(void 0),o++,i.call(s,t).then(function(t){r||(r=!0,n[e]=t,--o||l(n))},p)}),--o||l(n)});return r.error&&p(r.value),e.promise},race:function(t){var r=this,i=X(r),n=i.reject,e=C(function(){var e=x(r.resolve);w(t,function(t){e.call(r,t).then(i.resolve,n)})});return e.error&&n(e.value),i.promise}})},function(t,e,r){var i=r(3);t.exports=function(t,e){var r=i.console;r&&r.error&&(1===arguments.length?r.error(t):r.error(t,e))}},function(t,e){t.exports=function(t){try{return{error:!1,value:t()}}catch(t){return{error:!0,value:t}}}},function(t,e,r){"use strict";var i=r(1),n=r(33),a=r(30),o=r(133);i({target:"Promise",proto:!0,real:!0},{finally:function(e){var r=a(this,n("Promise")),t="function"==typeof e;return this.then(t?function(t){return o(r,e()).then(function(){return t})}:e,t?function(t){return o(r,e()).then(function(){throw t})}:e)}})},function(t,e,r){"use strict";var i=r(79),n=r(135);t.exports=i("Map",function(e){return function(t){return e(this,arguments.length?t:void 0)}},n,!0)},function(t,e,r){"use strict";var i=r(79),n=r(135);t.exports=i("Set",function(e){return function(t){return e(this,arguments.length?t:void 0)}},n)},function(t,e,r){"use strict";function i(e){return function(t){return e(this,arguments.length?t:void 0)}}var n,a=r(3),o=r(48),s=r(42),l=r(79),p=r(136),c=r(4),u=r(20).enforce,h=r(106),d=!a.ActiveXObject&&"ActiveXObject"in a,f=Object.isExtensible,m=t.exports=l("WeakMap",i,p,!0,!0);if(h&&d){n=p.getConstructor(i,"WeakMap",!0),s.REQUIRED=!0;var g=m.prototype,v=g.delete,y=g.has,x=g.get,b=g.set;o(g,{delete:function(t){if(!c(t)||f(t))return v.call(this,t);var e=u(this);return e.frozen||(e.frozen=new n),v.call(this,t)||e.frozen.delete(t)},has:function(t){if(!c(t)||f(t))return y.call(this,t);var e=u(this);return e.frozen||(e.frozen=new n),y.call(this,t)||e.frozen.has(t)},get:function(t){if(!c(t)||f(t))return x.call(this,t);var e=u(this);return e.frozen||(e.frozen=new n),y.call(this,t)?x.call(this,t):e.frozen.get(t)},set:function(t,e){if(c(t)&&!f(t)){var r=u(this);r.frozen||(r.frozen=new n),y.call(this,t)?b.call(this,t,e):r.frozen.set(t,e)}else b.call(this,t,e);return this}})}},function(t,e,r){"use strict";r(79)("WeakSet",function(e){return function(t){return e(this,arguments.length?t:void 0)}},r(136),!1,!0)},function(t,e,r){"use strict";var i=r(1),n=r(3),a=r(80),o=r(46),s="ArrayBuffer",l=a[s];i({global:!0,forced:n[s]!==l},{ArrayBuffer:l}),o(s)},function(t,e,r){var i=r(1),n=r(6);i({target:"ArrayBuffer",stat:!0,forced:!n.NATIVE_ARRAY_BUFFER_VIEWS},{isView:n.isView})},function(t,e,r){"use strict";var i=r(1),n=r(2),a=r(80),p=r(5),c=r(34),u=r(9),h=r(30),d=a.ArrayBuffer,f=a.DataView,m=d.prototype.slice;i({target:"ArrayBuffer",proto:!0,unsafe:!0,forced:n(function(){return!new d(2).slice(1,void 0).byteLength})},{slice:function(t,e){if(void 0!==m&&void 0===e)return m.call(p(this),t);for(var r=p(this).byteLength,i=c(t,r),n=c(void 0===e?r:e,r),a=new(h(this,d))(u(n-i)),o=new f(this),s=new f(a),l=0;i<n;)s.setUint8(l++,o.getUint8(i++));return a}})},function(t,e,r){var i=r(1),n=r(80);i({global:!0,forced:!r(6).NATIVE_ARRAY_BUFFER},{DataView:n.DataView})},function(t,e,r){r(31)("Int8",1,function(i){return function(t,e,r){return i(this,t,e,r)}})},function(t,e,r){r(31)("Uint8",1,function(i){return function(t,e,r){return i(this,t,e,r)}})},function(t,e,r){r(31)("Uint8",1,function(i){return function(t,e,r){return i(this,t,e,r)}},!0)},function(t,e,r){r(31)("Int16",2,function(i){return function(t,e,r){return i(this,t,e,r)}})},function(t,e,r){r(31)("Uint16",2,function(i){return function(t,e,r){return i(this,t,e,r)}})},function(t,e,r){r(31)("Int32",4,function(i){return function(t,e,r){return i(this,t,e,r)}})},function(t,e,r){r(31)("Uint32",4,function(i){return function(t,e,r){return i(this,t,e,r)}})},function(t,e,r){r(31)("Float32",4,function(i){return function(t,e,r){return i(this,t,e,r)}})},function(t,e,r){r(31)("Float64",8,function(i){return function(t,e,r){return i(this,t,e,r)}})},function(t,e,r){"use strict";var i=r(102),n=r(6),a=r(139);n.exportStatic("from",a,i)},function(t,e,r){"use strict";var i=r(6),n=r(102),a=i.aTypedArrayConstructor;i.exportStatic("of",function(){for(var t=0,e=arguments.length,r=new(a(this))(e);t<e;)r[t]=arguments[t++];return r},n)},function(t,e,r){"use strict";var i=r(6),n=r(120),a=i.aTypedArray;i.exportProto("copyWithin",function(t,e,r){return n.call(a(this),t,e,2<arguments.length?r:void 0)})},function(t,e,r){"use strict";var i=r(6),n=r(13).every,a=i.aTypedArray;i.exportProto("every",function(t,e){return n(a(this),t,1<arguments.length?e:void 0)})},function(t,e,r){"use strict";var i=r(6),n=r(90),a=i.aTypedArray;i.exportProto("fill",function(t){return n.apply(a(this),arguments)})},function(t,e,r){"use strict";var i=r(6),s=r(13).filter,l=r(30),p=i.aTypedArray,c=i.aTypedArrayConstructor;i.exportProto("filter",function(t,e){for(var r=s(p(this),t,1<arguments.length?e:void 0),i=l(this,this.constructor),n=0,a=r.length,o=new(c(i))(a);n<a;)o[n]=r[n++];return o})},function(t,e,r){"use strict";var i=r(6),n=r(13).find,a=i.aTypedArray;i.exportProto("find",function(t,e){return n(a(this),t,1<arguments.length?e:void 0)})},function(t,e,r){"use strict";var i=r(6),n=r(13).findIndex,a=i.aTypedArray;i.exportProto("findIndex",function(t,e){return n(a(this),t,1<arguments.length?e:void 0)})},function(t,e,r){"use strict";var i=r(6),n=r(13).forEach,a=i.aTypedArray;i.exportProto("forEach",function(t,e){n(a(this),t,1<arguments.length?e:void 0)})},function(t,e,r){"use strict";var i=r(6),n=r(54).includes,a=i.aTypedArray;i.exportProto("includes",function(t,e){return n(a(this),t,1<arguments.length?e:void 0)})},function(t,e,r){"use strict";var i=r(6),n=r(54).indexOf,a=i.aTypedArray;i.exportProto("indexOf",function(t,e){return n(a(this),t,1<arguments.length?e:void 0)})},function(t,e,r){"use strict";function i(){return p.call(h(this))}var n=r(3),a=r(6),o=r(70),s=r(8)("iterator"),l=n.Uint8Array,p=o.values,c=o.keys,u=o.entries,h=a.aTypedArray,d=a.exportProto,f=l&&l.prototype[s],m=!!f&&("values"==f.name||null==f.name);d("entries",function(){return u.call(h(this))}),d("keys",function(){return c.call(h(this))}),d("values",i,!m),d(s,i,!m)},function(t,e,r){"use strict";var i=r(6),n=i.aTypedArray,a=[].join;i.exportProto("join",function(t){return a.apply(n(this),arguments)})},function(t,e,r){"use strict";var i=r(6),n=r(123),a=i.aTypedArray;i.exportProto("lastIndexOf",function(t){return n.apply(a(this),arguments)})},function(t,e,r){"use strict";var i=r(6),n=r(13).map,a=r(30),o=i.aTypedArray,s=i.aTypedArrayConstructor;i.exportProto("map",function(t,e){return n(o(this),t,1<arguments.length?e:void 0,function(t,e){return new(s(a(t,t.constructor)))(e)})})},function(t,e,r){"use strict";var i=r(6),n=r(69).left,a=i.aTypedArray;i.exportProto("reduce",function(t,e){return n(a(this),t,arguments.length,1<arguments.length?e:void 0)})},function(t,e,r){"use strict";var i=r(6),n=r(69).right,a=i.aTypedArray;i.exportProto("reduceRight",function(t,e){return n(a(this),t,arguments.length,1<arguments.length?e:void 0)})},function(t,e,r){"use strict";var i=r(6),n=i.aTypedArray,a=Math.floor;i.exportProto("reverse",function(){for(var t,e=n(this).length,r=a(e/2),i=0;i<r;)t=this[i],this[i++]=this[--e],this[e]=t;return this})},function(t,e,r){"use strict";var i=r(6),s=r(9),l=r(138),p=r(11),n=r(2),c=i.aTypedArray,a=n(function(){new Int8Array(1).set({})});i.exportProto("set",function(t,e){c(this);var r=l(1<arguments.length?e:void 0,1),i=this.length,n=p(t),a=s(n.length),o=0;if(i<a+r)throw RangeError("Wrong length");for(;o<a;)this[r+o]=n[o++]},a)},function(t,e,r){"use strict";var i=r(6),s=r(30),n=r(2),l=i.aTypedArray,p=i.aTypedArrayConstructor,c=[].slice,a=n(function(){new Int8Array(1).slice()});i.exportProto("slice",function(t,e){for(var r=c.call(l(this),t,e),i=s(this,this.constructor),n=0,a=r.length,o=new(p(i))(a);n<a;)o[n]=r[n++];return o},a)},function(t,e,r){"use strict";var i=r(6),n=r(13).some,a=i.aTypedArray;i.exportProto("some",function(t,e){return n(a(this),t,1<arguments.length?e:void 0)})},function(t,e,r){"use strict";var i=r(6),n=i.aTypedArray,a=[].sort;i.exportProto("sort",function(t){return a.call(n(this),t)})},function(t,e,r){"use strict";var i=r(6),a=r(9),o=r(34),s=r(30),l=i.aTypedArray;i.exportProto("subarray",function(t,e){var r=l(this),i=r.length,n=o(t,i);return new(s(r,r.constructor))(r.buffer,r.byteOffset+n*r.BYTES_PER_ELEMENT,a((void 0===e?i:o(e,i))-n))})},function(t,e,r){"use strict";var i=r(3),n=r(6),a=r(2),o=i.Int8Array,s=n.aTypedArray,l=[].toLocaleString,p=[].slice,c=!!o&&a(function(){l.call(new o(1))}),u=a(function(){return[1,2].toLocaleString()!=new o([1,2]).toLocaleString()})||!a(function(){o.prototype.toLocaleString.call([1,2])});n.exportProto("toLocaleString",function(){return l.apply(c?p.call(s(this)):s(this),arguments)},u)},function(t,e,r){"use strict";var i=r(3),n=r(6),a=r(2),o=i.Uint8Array,s=o&&o.prototype,l=[].toString,p=[].join;a(function(){l.call({})})&&(l=function(){return p.call(this)}),n.exportProto("toString",l,(s||{}).toString!=l)},function(t,e,r){var i=r(1),n=r(33),a=r(21),o=r(5),s=r(2),l=n("Reflect","apply"),p=Function.apply;i({target:"Reflect",stat:!0,forced:!s(function(){l(function(){})})},{apply:function(t,e,r){return a(t),o(r),l?l(t,e,r):p.call(t,e,r)}})},function(t,e,r){var i=r(1),n=r(33),l=r(21),p=r(5),c=r(4),u=r(35),h=r(118),a=r(2),d=n("Reflect","construct"),f=a(function(){function t(){}return!(d(function(){},[],t)instanceof t)}),m=!a(function(){d(function(){})}),o=f||m;i({target:"Reflect",stat:!0,forced:o,sham:o},{construct:function(t,e,r){l(t),p(e);var i=arguments.length<3?t:l(r);if(m&&!f)return d(t,e,i);if(t==i){switch(e.length){case 0:return new t;case 1:return new t(e[0]);case 2:return new t(e[0],e[1]);case 3:return new t(e[0],e[1],e[2]);case 4:return new t(e[0],e[1],e[2],e[3])}var n=[null];return n.push.apply(n,e),new(h.apply(t,n))}var a=i.prototype,o=u(c(a)?a:Object.prototype),s=Function.apply.call(t,o,e);return c(s)?s:o}})},function(t,e,r){var i=r(1),n=r(7),a=r(5),o=r(26),s=r(10);i({target:"Reflect",stat:!0,forced:r(2)(function(){Reflect.defineProperty(s.f({},1,{value:1}),1,{value:2})}),sham:!n},{defineProperty:function(t,e,r){a(t);var i=o(e,!0);a(r);try{return s.f(t,i,r),!0}catch(t){return!1}}})},function(t,e,r){var i=r(1),n=r(5),a=r(16).f;i({target:"Reflect",stat:!0},{deleteProperty:function(t,e){var r=a(n(t),e);return!(r&&!r.configurable)&&delete t[e]}})},function(t,e,r){var i=r(1),o=r(4),s=r(5),l=r(12),p=r(16),c=r(28);i({target:"Reflect",stat:!0},{get:function t(e,r){var i,n,a=arguments.length<3?e:arguments[2];return s(e)===a?e[r]:(i=p.f(e,r))?l(i,"value")?i.value:void 0===i.get?void 0:i.get.call(a):o(n=c(e))?t(n,r,a):void 0}})},function(t,e,r){var i=r(1),n=r(7),a=r(5),o=r(16);i({target:"Reflect",stat:!0,sham:!n},{getOwnPropertyDescriptor:function(t,e){return o.f(a(t),e)}})},function(t,e,r){var i=r(1),n=r(5),a=r(28);i({target:"Reflect",stat:!0,sham:!r(89)},{getPrototypeOf:function(t){return a(n(t))}})},function(t,e,r){r(1)({target:"Reflect",stat:!0},{has:function(t,e){return e in t}})},function(t,e,r){var i=r(1),n=r(5),a=Object.isExtensible;i({target:"Reflect",stat:!0},{isExtensible:function(t){return n(t),!a||a(t)}})},function(t,e,r){r(1)({target:"Reflect",stat:!0},{ownKeys:r(84)})},function(t,e,r){var i=r(1),n=r(33),a=r(5);i({target:"Reflect",stat:!0,sham:!r(58)},{preventExtensions:function(t){a(t);try{var e=n("Object","preventExtensions");return e&&e(t),!0}catch(t){return!1}}})},function(t,e,r){var i=r(1),l=r(5),p=r(4),c=r(12),u=r(10),h=r(16),d=r(28),f=r(39);i({target:"Reflect",stat:!0},{set:function t(e,r,i){var n,a,o=arguments.length<4?e:arguments[3],s=h.f(l(e),r);if(!s){if(p(a=d(e)))return t(a,r,i,o);s=f(0)}if(c(s,"value")){if(!1===s.writable||!p(o))return!1;if(n=h.f(o,r)){if(n.get||n.set||!1===n.writable)return!1;n.value=i,u.f(o,r,n)}else u.f(o,r,f(0,i));return!0}return void 0!==s.set&&(s.set.call(o,i),!0)}})},function(t,e,r){var i=r(1),n=r(5),a=r(117),o=r(45);o&&i({target:"Reflect",stat:!0},{setPrototypeOf:function(t,e){n(t),a(e);try{return o(t,e),!0}catch(t){return!1}}})},function(t,e,r){r(357),r(358),r(359),r(360),r(361),r(362),r(365),r(142),t.exports=r(44)},function(t,e,r){var i=r(3),n=r(140),a=r(122),o=r(14);for(var s in n){var l=i[s],p=l&&l.prototype;if(p&&p.forEach!==a)try{o(p,"forEach",a)}catch(t){p.forEach=a}}},function(t,e,r){var i=r(3),n=r(140),a=r(70),o=r(14),s=r(8),l=s("iterator"),p=s("toStringTag"),c=a.values;for(var u in n){var h=i[u],d=h&&h.prototype;if(d){if(d[l]!==c)try{o(d,l,c)}catch(t){d[l]=c}if(d[p]||o(d,p,u),n[u])for(var f in a)if(d[f]!==a[f])try{o(d,f,a[f])}catch(t){d[f]=a[f]}}}},function(t,e,r){var i=r(3),n=r(101),a=!i.setImmediate||!i.clearImmediate;r(1)({global:!0,bind:!0,enumerable:!0,forced:a},{setImmediate:n.set,clearImmediate:n.clear})},function(t,e,r){var i=r(1),n=r(3),a=r(132),o=r(25),s=n.process,l="process"==o(s);i({global:!0,enumerable:!0,noTargetGet:!0},{queueMicrotask:function(t){var e=l&&s.domain;a(e?e.bind(t):t)}})},function(t,e,r){function i(n){return function(t,e){var r=2<arguments.length,i=r?s.call(arguments,2):void 0;return n(r?function(){("function"==typeof t?t:Function(t)).apply(this,i)}:t,e)}}var n=r(1),a=r(3),o=r(76),s=[].slice;n({global:!0,bind:!0,forced:/MSIE .\./.test(o)},{setTimeout:i(a.setTimeout),setInterval:i(a.setInterval)})},function(t,e,r){"use strict";r(126);function b(t,e){var r,i,n;if("["==e.charAt(0)){if("]"!=e.charAt(e.length-1))return N;if(!(r=J(e.slice(1,-1))))return N;t.host=r}else if(it(t)){if(e=g(e),Y.test(e))return N;if(null===(r=q(e)))return N;t.host=r}else{if(X.test(e))return N;for(r="",i=I(e),n=0;n<i.length;n++)r+=et(i[n],Q);t.host=r}}function c(t){var e,r,i,n;if("number"==typeof t){for(e=[],r=0;r<4;r++)e.unshift(t%256),t=D(t/256);return e.join(".")}if("object"!=typeof t)return t;for(e="",i=function(t){for(var e=null,r=1,i=null,n=0,a=0;a<8;a++)0!==t[a]?(r<n&&(e=i,r=n),i=null,n=0):(null===i&&(i=a),++n);return r<n&&(e=i,r=n),e}(t),r=0;r<8;r++)n&&0===t[r]||(n&&(n=!1),i===r?(e+=r?":":"::",n=!0):(e+=t[r].toString(16),r<7&&(e+=":")));return"["+e+"]"}function _(t){return""!=t.username||""!=t.password}function n(t){return!t.host||t.cannotBeABaseURL||"file"==t.scheme}function w(t,e){var r;return 2==t.length&&B.test(t.charAt(0))&&(":"==(r=t.charAt(1))||!e&&"|"==r)}function S(t){var e;return 1<t.length&&w(t.slice(0,2))&&(2==t.length||"/"===(e=t.charAt(2))||"\\"===e||"?"===e||"#"===e)}function T(t){var e=t.path,r=e.length;!r||"file"==t.scheme&&1==r&&w(e[0],!0)||e.pop()}function u(t,e,r,i){var n,a,o,s,l,p,c=r||nt,u=0,h="",d=!1,f=!1,m=!1;for(r||(t.scheme="",t.username="",t.password="",t.host=null,t.port=null,t.path=[],t.query=null,t.fragment=null,t.cannotBeABaseURL=!1,e=e.replace(W,"")),e=e.replace(U,""),n=I(e);u<=n.length;){switch(a=n[u],c){case nt:if(!a||!B.test(a)){if(r)return R;c=ot;continue}h+=a.toLowerCase(),c=at;break;case at:if(a&&(j.test(a)||"+"==a||"-"==a||"."==a))h+=a.toLowerCase();else{if(":"!=a){if(r)return R;h="",c=ot,u=0;continue}if(r&&(it(t)!=k(rt,h)||"file"==h&&(_(t)||null!==t.port)||"file"==t.scheme&&!t.host))return;if(t.scheme=h,r)return void(it(t)&&rt[t.scheme]==t.port&&(t.port=null));h="","file"==t.scheme?c=vt:it(t)&&i&&i.scheme==t.scheme?c=st:it(t)?c=ut:"/"==n[u+1]?(c=lt,u++):(t.cannotBeABaseURL=!0,t.path.push(""),c=wt)}break;case ot:if(!i||i.cannotBeABaseURL&&"#"!=a)return R;if(i.cannotBeABaseURL&&"#"==a){t.scheme=i.scheme,t.path=i.path.slice(),t.query=i.query,t.fragment="",t.cannotBeABaseURL=!0,c=Tt;break}c="file"==i.scheme?vt:pt;continue;case st:if("/"!=a||"/"!=n[u+1]){c=pt;continue}c=ht,u++;break;case lt:if("/"==a){c=dt;break}c=_t;continue;case pt:if(t.scheme=i.scheme,a==P)t.username=i.username,t.password=i.password,t.host=i.host,t.port=i.port,t.path=i.path.slice(),t.query=i.query;else if("/"==a||"\\"==a&&it(t))c=ct;else if("?"==a)t.username=i.username,t.password=i.password,t.host=i.host,t.port=i.port,t.path=i.path.slice(),t.query="",c=St;else{if("#"!=a){t.username=i.username,t.password=i.password,t.host=i.host,t.port=i.port,t.path=i.path.slice(),t.path.pop(),c=_t;continue}t.username=i.username,t.password=i.password,t.host=i.host,t.port=i.port,t.path=i.path.slice(),t.query=i.query,t.fragment="",c=Tt}break;case ct:if(!it(t)||"/"!=a&&"\\"!=a){if("/"!=a){t.username=i.username,t.password=i.password,t.host=i.host,t.port=i.port,c=_t;continue}c=dt}else c=ht;break;case ut:if(c=ht,"/"!=a||"/"!=h.charAt(u+1))continue;u++;break;case ht:if("/"==a||"\\"==a)break;c=dt;continue;case dt:if("@"==a){d&&(h="%40"+h),d=!0,o=I(h);for(var g=0;g<o.length;g++){var v=o[g];if(":"!=v||m){var y=et(v,tt);m?t.password+=y:t.username+=y}else m=!0}h=""}else if(a==P||"/"==a||"?"==a||"#"==a||"\\"==a&&it(t)){if(d&&""==h)return"Invalid authority";u-=I(h).length+1,h="",c=ft}else h+=a;break;case ft:case mt:if(r&&"file"==t.scheme){c=xt;continue}if(":"!=a||f){if(a==P||"/"==a||"?"==a||"#"==a||"\\"==a&&it(t)){if(it(t)&&""==h)return N;if(r&&""==h&&(_(t)||null!==t.port))return;if(s=b(t,h))return s;if(h="",c=bt,r)return;continue}"["==a?f=!0:"]"==a&&(f=!1),h+=a}else{if(""==h)return N;if(s=b(t,h))return s;if(h="",c=gt,r==mt)return}break;case gt:if(!$.test(a)){if(a==P||"/"==a||"?"==a||"#"==a||"\\"==a&&it(t)||r){if(""!=h){var x=parseInt(h,10);if(65535<x)return L;t.port=it(t)&&x===rt[t.scheme]?null:x,h=""}if(r)return;c=bt;continue}return L}h+=a;break;case vt:if(t.scheme="file","/"==a||"\\"==a)c=yt;else{if(!i||"file"!=i.scheme){c=_t;continue}if(a==P)t.host=i.host,t.path=i.path.slice(),t.query=i.query;else if("?"==a)t.host=i.host,t.path=i.path.slice(),t.query="",c=St;else{if("#"!=a){S(n.slice(u).join(""))||(t.host=i.host,t.path=i.path.slice(),T(t)),c=_t;continue}t.host=i.host,t.path=i.path.slice(),t.query=i.query,t.fragment="",c=Tt}}break;case yt:if("/"==a||"\\"==a){c=xt;break}i&&"file"==i.scheme&&!S(n.slice(u).join(""))&&(w(i.path[0],!0)?t.path.push(i.path[0]):t.host=i.host),c=_t;continue;case xt:if(a==P||"/"==a||"\\"==a||"?"==a||"#"==a){if(!r&&w(h))c=_t;else if(""==h){if(t.host="",r)return;c=bt}else{if(s=b(t,h))return s;if("localhost"==t.host&&(t.host=""),r)return;h="",c=bt}continue}h+=a;break;case bt:if(it(t)){if(c=_t,"/"!=a&&"\\"!=a)continue}else if(r||"?"!=a)if(r||"#"!=a){if(a!=P&&(c=_t,"/"!=a))continue}else t.fragment="",c=Tt;else t.query="",c=St;break;case _t:if(a==P||"/"==a||"\\"==a&&it(t)||!r&&("?"==a||"#"==a)){if(".."===(p=(p=h).toLowerCase())||"%2e."===p||".%2e"===p||"%2e%2e"===p?(T(t),"/"==a||"\\"==a&&it(t)||t.path.push("")):"."===(l=h)||"%2e"===l.toLowerCase()?"/"==a||"\\"==a&&it(t)||t.path.push(""):("file"==t.scheme&&!t.path.length&&w(h)&&(t.host&&(t.host=""),h=h.charAt(0)+":"),t.path.push(h)),h="","file"==t.scheme&&(a==P||"?"==a||"#"==a))for(;1<t.path.length&&""===t.path[0];)t.path.shift();"?"==a?(t.query="",c=St):"#"==a&&(t.fragment="",c=Tt)}else h+=et(a,K);break;case wt:"?"==a?(t.query="",c=St):"#"==a?(t.fragment="",c=Tt):a!=P&&(t.path[0]+=et(a,Q));break;case St:r||"#"!=a?a!=P&&("'"==a&&it(t)?t.query+="%27":t.query+="#"==a?"%23":et(a,Q)):(t.fragment="",c=Tt);break;case Tt:a!=P&&(t.fragment+=et(a,Z))}u++}}function i(t,e){return{get:t,set:e,configurable:!0,enumerable:!0}}var P,a=r(1),h=r(7),o=r(141),s=r(3),l=r(87),p=r(17),d=r(38),k=r(12),f=r(113),I=r(119),m=r(71).codeAt,g=r(363),v=r(27),y=r(142),x=r(20),A=s.URL,E=y.URLSearchParams,C=y.getState,O=x.set,M=x.getterFor("URL"),D=Math.floor,F=Math.pow,R="Invalid scheme",N="Invalid host",L="Invalid port",B=/[A-Za-z]/,j=/[\d+\-.A-Za-z]/,$=/\d/,V=/^(0x|0X)/,H=/^[0-7]+$/,z=/^\d+$/,G=/^[\dA-Fa-f]+$/,Y=/[\u0000\u0009\u000A\u000D #%/:?@[\\]]/,X=/[\u0000\u0009\u000A\u000D #/:?@[\\]]/,W=/^[\u0000-\u001F ]+|[\u0000-\u001F ]+$/g,U=/[\u0009\u000A\u000D]/g,q=function(t){var e,r,i,n,a,o,s,l=t.split(".");if(l.length&&""==l[l.length-1]&&l.pop(),4<(e=l.length))return t;for(r=[],i=0;i<e;i++){if(""==(n=l[i]))return t;if(a=10,1<n.length&&"0"==n.charAt(0)&&(a=V.test(n)?16:8,n=n.slice(8==a?1:2)),""===n)o=0;else{if(!(10==a?z:8==a?H:G).test(n))return t;o=parseInt(n,a)}r.push(o)}for(i=0;i<e;i++)if(o=r[i],i==e-1){if(o>=F(256,5-e))return null}else if(255<o)return null;for(s=r.pop(),i=0;i<r.length;i++)s+=r[i]*F(256,3-i);return s},J=function(t){function e(){return t.charAt(h)}var r,i,n,a,o,s,l,p=[0,0,0,0,0,0,0,0],c=0,u=null,h=0;if(":"==e()){if(":"!=t.charAt(1))return;h+=2,u=++c}for(;e();){if(8==c)return;if(":"!=e()){for(r=i=0;i<4&&G.test(e());)r=16*r+parseInt(e(),16),h++,i++;if("."==e()){if(0==i)return;if(h-=i,6<c)return;for(n=0;e();){if(a=null,0<n){if(!("."==e()&&n<4))return;h++}if(!$.test(e()))return;for(;$.test(e());){if(o=parseInt(e(),10),null===a)a=o;else{if(0==a)return;a=10*a+o}if(255<a)return;h++}p[c]=256*p[c]+a,2!=++n&&4!=n||c++}if(4!=n)return;break}if(":"==e()){if(h++,!e())return}else if(e())return;p[c++]=r}else{if(null!==u)return;h++,u=++c}}if(null!==u)for(s=c-u,c=7;0!=c&&0<s;)l=p[c],p[c--]=p[u+s-1],p[u+--s]=l;else if(8!=c)return;return p},Q={},Z=f({},Q,{" ":1,'"':1,"<":1,">":1,"`":1}),K=f({},Z,{"#":1,"?":1,"{":1,"}":1}),tt=f({},K,{"/":1,":":1,";":1,"=":1,"@":1,"[":1,"\\":1,"]":1,"^":1,"|":1}),et=function(t,e){var r=m(t,0);return 32<r&&r<127&&!k(e,t)?t:encodeURIComponent(t)},rt={ftp:21,file:null,gopher:70,http:80,https:443,ws:80,wss:443},it=function(t){return k(rt,t.scheme)},nt={},at={},ot={},st={},lt={},pt={},ct={},ut={},ht={},dt={},ft={},mt={},gt={},vt={},yt={},xt={},bt={},_t={},wt={},St={},Tt={},Pt=function(t,e){var r,i,n=d(this,Pt,"URL"),a=1<arguments.length?e:void 0,o=String(t),s=O(n,{type:"URL"});if(void 0!==a)if(a instanceof Pt)r=M(a);else if(i=u(r={},String(a)))throw TypeError(i);if(i=u(s,o,null,r))throw TypeError(i);var l=s.searchParams=new E,p=C(l);p.updateSearchParams(s.query),p.updateURL=function(){s.query=String(l)||null},h||(n.href=It.call(n),n.origin=At.call(n),n.protocol=Et.call(n),n.username=Ct.call(n),n.password=Ot.call(n),n.host=Mt.call(n),n.hostname=Dt.call(n),n.port=Ft.call(n),n.pathname=Rt.call(n),n.search=Nt.call(n),n.searchParams=Lt.call(n),n.hash=Bt.call(n))},kt=Pt.prototype,It=function(){var t=M(this),e=t.scheme,r=t.username,i=t.password,n=t.host,a=t.port,o=t.path,s=t.query,l=t.fragment,p=e+":";return null!==n?(p+="//",_(t)&&(p+=r+(i?":"+i:"")+"@"),p+=c(n),null!==a&&(p+=":"+a)):"file"==e&&(p+="//"),p+=t.cannotBeABaseURL?o[0]:o.length?"/"+o.join("/"):"",null!==s&&(p+="?"+s),null!==l&&(p+="#"+l),p},At=function(){var t=M(this),e=t.scheme,r=t.port;if("blob"==e)try{return new URL(e.path[0]).origin}catch(t){return"null"}return"file"!=e&&it(t)?e+"://"+c(t.host)+(null!==r?":"+r:""):"null"},Et=function(){return M(this).scheme+":"},Ct=function(){return M(this).username},Ot=function(){return M(this).password},Mt=function(){var t=M(this),e=t.host,r=t.port;return null===e?"":null===r?c(e):c(e)+":"+r},Dt=function(){var t=M(this).host;return null===t?"":c(t)},Ft=function(){var t=M(this).port;return null===t?"":String(t)},Rt=function(){var t=M(this),e=t.path;return t.cannotBeABaseURL?e[0]:e.length?"/"+e.join("/"):""},Nt=function(){var t=M(this).query;return t?"?"+t:""},Lt=function(){return M(this).searchParams},Bt=function(){var t=M(this).fragment;return t?"#"+t:""};if(h&&l(kt,{href:i(It,function(t){var e=M(this),r=String(t),i=u(e,r);if(i)throw TypeError(i);C(e.searchParams).updateSearchParams(e.query)}),origin:i(At),protocol:i(Et,function(t){var e=M(this);u(e,String(t)+":",nt)}),username:i(Ct,function(t){var e=M(this),r=I(String(t));if(!n(e)){e.username="";for(var i=0;i<r.length;i++)e.username+=et(r[i],tt)}}),password:i(Ot,function(t){var e=M(this),r=I(String(t));if(!n(e)){e.password="";for(var i=0;i<r.length;i++)e.password+=et(r[i],tt)}}),host:i(Mt,function(t){var e=M(this);e.cannotBeABaseURL||u(e,String(t),ft)}),hostname:i(Dt,function(t){var e=M(this);e.cannotBeABaseURL||u(e,String(t),mt)}),port:i(Ft,function(t){var e=M(this);n(e)||(""==(t=String(t))?e.port=null:u(e,t,gt))}),pathname:i(Rt,function(t){var e=M(this);e.cannotBeABaseURL||(e.path=[],u(e,t+"",bt))}),search:i(Nt,function(t){var e=M(this);""==(t=String(t))?e.query=null:("?"==t.charAt(0)&&(t=t.slice(1)),e.query="",u(e,t,St)),C(e.searchParams).updateSearchParams(e.query)}),searchParams:i(Lt),hash:i(Bt,function(t){var e=M(this);""!=(t=String(t))?("#"==t.charAt(0)&&(t=t.slice(1)),e.fragment="",u(e,t,Tt)):e.fragment=null})}),p(kt,"toJSON",function(){return It.call(this)},{enumerable:!0}),p(kt,"toString",function(){return It.call(this)},{enumerable:!0}),A){var jt=A.createObjectURL,$t=A.revokeObjectURL;jt&&p(Pt,"createObjectURL",function(t){return jt.apply(A,arguments)}),$t&&p(Pt,"revokeObjectURL",function(t){return $t.apply(A,arguments)})}v(Pt,"URL"),a({global:!0,forced:!o,sham:!h},{URL:Pt})},function(t,e,r){"use strict";function v(t){return t+22+75*(t<26)}function y(t,e,r){var i=0;for(t=r?_(t/700):t>>1,t+=_(t/e);455<t;i+=36)t=_(t/35);return _(i+36*t/(t+38))}function a(t){var e,r,i=[],n=(t=function(t){for(var e=[],r=0,i=t.length;r<i;){var n=t.charCodeAt(r++);if(55296<=n&&n<=56319&&r<i){var a=t.charCodeAt(r++);56320==(64512&a)?e.push(((1023&n)<<10)+(1023&a)+65536):(e.push(n),r--)}else e.push(n)}return e}(t)).length,a=128,o=0,s=72;for(e=0;e<t.length;e++)(r=t[e])<128&&i.push(w(r));var l=i.length,p=l;for(l&&i.push("-");p<n;){var c=x;for(e=0;e<t.length;e++)a<=(r=t[e])&&r<c&&(c=r);var u=p+1;if(c-a>_((x-o)/u))throw RangeError(b);for(o+=(c-a)*u,a=c,e=0;e<t.length;e++){if((r=t[e])<a&&++o>x)throw RangeError(b);if(r==a){for(var h=o,d=36;;d+=36){var f=d<=s?1:s+26<=d?26:d-s;if(h<f)break;var m=h-f,g=36-f;i.push(w(v(f+m%g))),h=_(m/g)}i.push(w(v(h))),s=y(o,u,p==l),o=0,++p}}++o,++a}return i.join("")}var x=2147483647,o=/[^\0-\u007E]/,s=/[.\u3002\uFF0E\uFF61]/g,b="Overflow: input needs wider integers to process",_=Math.floor,w=String.fromCharCode;t.exports=function(t){var e,r,i=[],n=t.toLowerCase().replace(s,".").split(".");for(e=0;e<n.length;e++)r=n[e],i.push(o.test(r)?"xn--"+a(r):r);return i.join(".")}},function(t,e,r){var i=r(5),n=r(61);t.exports=function(t){var e=n(t);if("function"!=typeof e)throw TypeError(String(t)+" is not iterable");return i(e.call(t))}},function(t,e,r){"use strict";r(1)({target:"URL",proto:!0,enumerable:!0},{toJSON:function(){return URL.prototype.toString.call(this)}})},function(t,e,r){var i=function(a){"use strict";var l,t=Object.prototype,p=t.hasOwnProperty,e="function"==typeof Symbol?Symbol:{},n=e.iterator||"@@iterator",r=e.asyncIterator||"@@asyncIterator",i=e.toStringTag||"@@toStringTag";function o(t,e,r,i){var n=e&&e.prototype instanceof s?e:s,a=Object.create(n.prototype),o=new I(i||[]);return a._invoke=function(a,o,s){var l=u;return function(t,e){if(l===d)throw new Error("Generator is already running");if(l===f){if("throw"===t)throw e;return E()}for(s.method=t,s.arg=e;;){var r=s.delegate;if(r){var i=T(r,s);if(i){if(i===m)continue;return i}}if("next"===s.method)s.sent=s._sent=s.arg;else if("throw"===s.method){if(l===u)throw l=f,s.arg;s.dispatchException(s.arg)}else"return"===s.method&&s.abrupt("return",s.arg);l=d;var n=c(a,o,s);if("normal"===n.type){if(l=s.done?f:h,n.arg===m)continue;return{value:n.arg,done:s.done}}"throw"===n.type&&(l=f,s.method="throw",s.arg=n.arg)}}}(t,r,o),a}function c(t,e,r){try{return{type:"normal",arg:t.call(e,r)}}catch(t){return{type:"throw",arg:t}}}a.wrap=o;var u="suspendedStart",h="suspendedYield",d="executing",f="completed",m={};function s(){}function g(){}function v(){}var y={};y[n]=function(){return this};var x=Object.getPrototypeOf,b=x&&x(x(A([])));b&&b!==t&&p.call(b,n)&&(y=b);var _=v.prototype=s.prototype=Object.create(y);function w(t){["next","throw","return"].forEach(function(e){t[e]=function(t){return this._invoke(e,t)}})}function S(l){var e;this._invoke=function(r,i){function t(){return new Promise(function(t,e){!function e(t,r,i,n){var a=c(l[t],l,r);if("throw"!==a.type){var o=a.arg,s=o.value;return s&&"object"==typeof s&&p.call(s,"__await")?Promise.resolve(s.__await).then(function(t){e("next",t,i,n)},function(t){e("throw",t,i,n)}):Promise.resolve(s).then(function(t){o.value=t,i(o)},function(t){return e("throw",t,i,n)})}n(a.arg)}(r,i,t,e)})}return e=e?e.then(t,t):t()}}function T(t,e){var r=t.iterator[e.method];if(r===l){if(e.delegate=null,"throw"===e.method){if(t.iterator.return&&(e.method="return",e.arg=l,T(t,e),"throw"===e.method))return m;e.method="throw",e.arg=new TypeError("The iterator does not provide a 'throw' method")}return m}var i=c(r,t.iterator,e.arg);if("throw"===i.type)return e.method="throw",e.arg=i.arg,e.delegate=null,m;var n=i.arg;return n?n.done?(e[t.resultName]=n.value,e.next=t.nextLoc,"return"!==e.method&&(e.method="next",e.arg=l),e.delegate=null,m):n:(e.method="throw",e.arg=new TypeError("iterator result is not an object"),e.delegate=null,m)}function P(t){var e={tryLoc:t[0]};1 in t&&(e.catchLoc=t[1]),2 in t&&(e.finallyLoc=t[2],e.afterLoc=t[3]),this.tryEntries.push(e)}function k(t){var e=t.completion||{};e.type="normal",delete e.arg,t.completion=e}function I(t){this.tryEntries=[{tryLoc:"root"}],t.forEach(P,this),this.reset(!0)}function A(e){if(e){var t=e[n];if(t)return t.call(e);if("function"==typeof e.next)return e;if(!isNaN(e.length)){var r=-1,i=function t(){for(;++r<e.length;)if(p.call(e,r))return t.value=e[r],t.done=!1,t;return t.value=l,t.done=!0,t};return i.next=i}}return{next:E}}function E(){return{value:l,done:!0}}return g.prototype=_.constructor=v,v.constructor=g,v[i]=g.displayName="GeneratorFunction",a.isGeneratorFunction=function(t){var e="function"==typeof t&&t.constructor;return!!e&&(e===g||"GeneratorFunction"===(e.displayName||e.name))},a.mark=function(t){return Object.setPrototypeOf?Object.setPrototypeOf(t,v):(t.__proto__=v,i in t||(t[i]="GeneratorFunction")),t.prototype=Object.create(_),t},a.awrap=function(t){return{__await:t}},w(S.prototype),S.prototype[r]=function(){return this},a.AsyncIterator=S,a.async=function(t,e,r,i){var n=new S(o(t,e,r,i));return a.isGeneratorFunction(e)?n:n.next().then(function(t){return t.done?t.value:n.next()})},w(_),_[i]="Generator",_[n]=function(){return this},_.toString=function(){return"[object Generator]"},a.keys=function(r){var i=[];for(var t in r)i.push(t);return i.reverse(),function t(){for(;i.length;){var e=i.pop();if(e in r)return t.value=e,t.done=!1,t}return t.done=!0,t}},a.values=A,I.prototype={constructor:I,reset:function(t){if(this.prev=0,this.next=0,this.sent=this._sent=l,this.done=!1,this.delegate=null,this.method="next",this.arg=l,this.tryEntries.forEach(k),!t)for(var e in this)"t"===e.charAt(0)&&p.call(this,e)&&!isNaN(+e.slice(1))&&(this[e]=l)},stop:function(){this.done=!0;var t=this.tryEntries[0].completion;if("throw"===t.type)throw t.arg;return this.rval},dispatchException:function(r){if(this.done)throw r;var i=this;function t(t,e){return a.type="throw",a.arg=r,i.next=t,e&&(i.method="next",i.arg=l),!!e}for(var e=this.tryEntries.length-1;0<=e;--e){var n=this.tryEntries[e],a=n.completion;if("root"===n.tryLoc)return t("end");if(n.tryLoc<=this.prev){var o=p.call(n,"catchLoc"),s=p.call(n,"finallyLoc");if(o&&s){if(this.prev<n.catchLoc)return t(n.catchLoc,!0);if(this.prev<n.finallyLoc)return t(n.finallyLoc)}else if(o){if(this.prev<n.catchLoc)return t(n.catchLoc,!0)}else{if(!s)throw new Error("try statement without catch or finally");if(this.prev<n.finallyLoc)return t(n.finallyLoc)}}}},abrupt:function(t,e){for(var r=this.tryEntries.length-1;0<=r;--r){var i=this.tryEntries[r];if(i.tryLoc<=this.prev&&p.call(i,"finallyLoc")&&this.prev<i.finallyLoc){var n=i;break}}n&&("break"===t||"continue"===t)&&n.tryLoc<=e&&e<=n.finallyLoc&&(n=null);var a=n?n.completion:{};return a.type=t,a.arg=e,n?(this.method="next",this.next=n.finallyLoc,m):this.complete(a)},complete:function(t,e){if("throw"===t.type)throw t.arg;return"break"===t.type||"continue"===t.type?this.next=t.arg:"return"===t.type?(this.rval=this.arg=t.arg,this.method="return",this.next="end"):"normal"===t.type&&e&&(this.next=e),m},finish:function(t){for(var e=this.tryEntries.length-1;0<=e;--e){var r=this.tryEntries[e];if(r.finallyLoc===t)return this.complete(r.completion,r.afterLoc),k(r),m}},catch:function(t){for(var e=this.tryEntries.length-1;0<=e;--e){var r=this.tryEntries[e];if(r.tryLoc===t){var i=r.completion;if("throw"===i.type){var n=i.arg;k(r)}return n}}throw new Error("illegal catch attempt")},delegateYield:function(t,e,r){return this.delegate={iterator:A(t),resultName:e,nextLoc:r},"next"===this.method&&(this.arg=l),m}},a}(t.exports);try{regeneratorRuntime=i}catch(t){Function("r","regeneratorRuntime = r")(i)}},function(t,e,r){!function(t,e){"use strict";e=e&&e.hasOwnProperty("default")?e.default:e;var c="function"==typeof Symbol&&"symbol"==typeof Symbol.iterator?function(t){return typeof t}:function(t){return t&&"function"==typeof Symbol&&t.constructor===Symbol&&t!==Symbol.prototype?"symbol":typeof t},n=function(t,e){if(!(t instanceof e))throw new TypeError("Cannot call a class as a function")},r=function(){function i(t,e){for(var r=0;r<e.length;r++){var i=e[r];i.enumerable=i.enumerable||!1,i.configurable=!0,"value"in i&&(i.writable=!0),Object.defineProperty(t,i.key,i)}}return function(t,e,r){return e&&i(t.prototype,e),r&&i(t,r),t}}(),i=function t(e,r,i){null===e&&(e=Function.prototype);var n=Object.getOwnPropertyDescriptor(e,r);if(void 0===n){var a=Object.getPrototypeOf(e);return null===a?void 0:t(a,r,i)}if("value"in n)return n.value;var o=n.get;return void 0!==o?o.call(i):void 0},a=function(t,e){if("function"!=typeof e&&null!==e)throw new TypeError("Super expression must either be null or a function, not "+typeof e);t.prototype=Object.create(e&&e.prototype,{constructor:{value:t,enumerable:!1,writable:!0,configurable:!0}}),e&&(Object.setPrototypeOf?Object.setPrototypeOf(t,e):t.__proto__=e)},o=function(t,e){if(!t)throw new ReferenceError("this hasn't been initialised - super() hasn't been called");return!e||"object"!=typeof e&&"function"!=typeof e?t:e},s=function(t,e){if(Array.isArray(t))return t;if(Symbol.iterator in Object(t))return function(t,e){var r=[],i=!0,n=!1,a=void 0;try{for(var o,s=t[Symbol.iterator]();!(i=(o=s.next()).done)&&(r.push(o.value),!e||r.length!==e);i=!0);}catch(t){n=!0,a=t}finally{try{!i&&s.return&&s.return()}finally{if(n)throw a}}return r}(t,e);throw new TypeError("Invalid attempt to destructure non-iterable instance")},l=function(t){if(Array.isArray(t)){for(var e=0,r=Array(t.length);e<t.length;e++)r[e]=t[e];return r}return Array.from(t)},p=function(){function e(t){n(this,e),this.selfOptions=t||{},this.pipes={}}return r(e,[{key:"options",value:function(t){return t&&(this.selfOptions=t),this.selfOptions}},{key:"pipe",value:function(t,e){var r=e;if("string"==typeof t){if(void 0===r)return this.pipes[t];this.pipes[t]=r}if(t&&t.name){if(r=t,r.processor===this)return r;this.pipes[r.name]=r}return r.processor=this,r}},{key:"process",value:function(t,e){var r=t;r.options=this.options();for(var i=e||t.pipe||"default",n=void 0,a=void 0;i;)void 0!==r.nextAfterChildren&&(r.next=r.nextAfterChildren,r.nextAfterChildren=null),"string"==typeof i&&(i=this.pipe(i)),i.process(r),n=i,i=null,(a=r)&&r.next&&(r=r.next,i=a.nextPipe||r.pipe||n);return r.hasResult?r.result:void 0}}]),e}(),u=function(){function e(t){n(this,e),this.name=t,this.filters=[]}return r(e,[{key:"process",value:function(t){if(!this.processor)throw new Error("add this pipe to a processor before using it");for(var e=this.debug,r=this.filters.length,i=t,n=0;n<r;n++){var a=this.filters[n];if(e&&this.log("filter: "+a.filterName),a(i),"object"===(void 0===i?"undefined":c(i))&&i.exiting){i.exiting=!1;break}}!i.next&&this.resultCheck&&this.resultCheck(i)}},{key:"log",value:function(t){console.log("[jsondiffpatch] "+this.name+" pipe, "+t)}},{key:"append",value:function(){var t;return(t=this.filters).push.apply(t,arguments),this}},{key:"prepend",value:function(){var t;return(t=this.filters).unshift.apply(t,arguments),this}},{key:"indexOf",value:function(t){if(!t)throw new Error("a filter name is required");for(var e=0;e<this.filters.length;e++){var r=this.filters[e];if(r.filterName===t)return e}throw new Error("filter not found: "+t)}},{key:"list",value:function(){return this.filters.map(function(t){return t.filterName})}},{key:"after",value:function(t){var e=this.indexOf(t),r=Array.prototype.slice.call(arguments,1);if(!r.length)throw new Error("a filter is required");return r.unshift(e+1,0),Array.prototype.splice.apply(this.filters,r),this}},{key:"before",value:function(t){var e=this.indexOf(t),r=Array.prototype.slice.call(arguments,1);if(!r.length)throw new Error("a filter is required");return r.unshift(e,0),Array.prototype.splice.apply(this.filters,r),this}},{key:"replace",value:function(t){var e=this.indexOf(t),r=Array.prototype.slice.call(arguments,1);if(!r.length)throw new Error("a filter is required");return r.unshift(e,1),Array.prototype.splice.apply(this.filters,r),this}},{key:"remove",value:function(t){var e=this.indexOf(t);return this.filters.splice(e,1),this}},{key:"clear",value:function(){return this.filters.length=0,this}},{key:"shouldHaveResult",value:function(t){if(!1!==t){if(!this.resultCheck){var r=this;return this.resultCheck=function(t){if(!t.hasResult){console.log(t);var e=new Error(r.name+" failed");throw e.noResult=!0,e}},this}}else this.resultCheck=null}}]),e}(),h=function(){function t(){n(this,t)}return r(t,[{key:"setResult",value:function(t){return this.result=t,this.hasResult=!0,this}},{key:"exit",value:function(){return this.exiting=!0,this}},{key:"switchTo",value:function(t,e){return"string"==typeof t||t instanceof u?this.nextPipe=t:(this.next=t,e&&(this.nextPipe=e)),this}},{key:"push",value:function(t,e){return t.parent=this,void 0!==e&&(t.childName=e),t.root=this.root||this,t.options=t.options||this.options,this.children?(this.children[this.children.length-1].next=t,this.children.push(t)):(this.children=[t],this.nextAfterChildren=this.next||null,this.next=t),t.next=this}}]),t}(),d="function"==typeof Array.isArray?Array.isArray:function(t){return t instanceof Array};function f(t){if("object"!==(void 0===t?"undefined":c(t)))return t;if(null===t)return null;if(d(t))return t.map(f);if(t instanceof Date)return new Date(t.getTime());if(t instanceof RegExp)return function(t){var e=/^\/(.*)\/([gimyu]*)$/.exec(t.toString());return new RegExp(e[1],e[2])}(t);var e={};for(var r in t)Object.prototype.hasOwnProperty.call(t,r)&&(e[r]=f(t[r]));return e}var S=function(t){function i(t,e){n(this,i);var r=o(this,(i.__proto__||Object.getPrototypeOf(i)).call(this));return r.left=t,r.right=e,r.pipe="diff",r}return a(i,t),r(i,[{key:"setResult",value:function(t){if(this.options.cloneDiffValues&&"object"===(void 0===t?"undefined":c(t))){var e="function"==typeof this.options.cloneDiffValues?this.options.cloneDiffValues:f;"object"===c(t[0])&&(t[0]=e(t[0])),"object"===c(t[1])&&(t[1]=e(t[1]))}return h.prototype.setResult.apply(this,arguments)}}]),i}(h),m=function(t){function i(t,e){n(this,i);var r=o(this,(i.__proto__||Object.getPrototypeOf(i)).call(this));return r.left=t,r.delta=e,r.pipe="patch",r}return a(i,t),i}(h),g=function(t){function r(t){n(this,r);var e=o(this,(r.__proto__||Object.getPrototypeOf(r)).call(this));return e.delta=t,e.pipe="reverse",e}return a(r,t),r}(h),v="function"==typeof Array.isArray?Array.isArray:function(t){return t instanceof Array},y=function(t){if(t.left!==t.right)if(void 0!==t.left)if(void 0!==t.right){if("function"==typeof t.left||"function"==typeof t.right)throw new Error("functions are not supported");t.leftType=null===t.left?"null":c(t.left),t.rightType=null===t.right?"null":c(t.right),t.leftType===t.rightType&&"boolean"!==t.leftType&&"number"!==t.leftType?("object"===t.leftType&&(t.leftIsArray=v(t.left)),"object"===t.rightType&&(t.rightIsArray=v(t.right)),t.leftIsArray===t.rightIsArray?t.left instanceof RegExp&&(t.right instanceof RegExp?t.setResult([t.left.toString(),t.right.toString()]).exit():t.setResult([t.left,t.right]).exit()):t.setResult([t.left,t.right]).exit()):t.setResult([t.left,t.right]).exit()}else t.setResult([t.left,0,0]).exit();else{if("function"==typeof t.right)throw new Error("functions are not supported");t.setResult([t.right]).exit()}else t.setResult(void 0).exit()},x=function(t){if(void 0!==t.delta){if(t.nested=!v(t.delta),!t.nested)if(1!==t.delta.length)if(2!==t.delta.length)3===t.delta.length&&0===t.delta[2]&&t.setResult(void 0).exit();else{if(t.left instanceof RegExp){var e=/^\/(.*)\/([gimyu]+)$/.exec(t.delta[1]);if(e)return void t.setResult(new RegExp(e[1],e[2])).exit()}t.setResult(t.delta[1]).exit()}else t.setResult(t.delta[0]).exit()}else t.setResult(t.left).exit()},b=function(t){void 0!==t.delta?(t.nested=!v(t.delta),t.nested||(1!==t.delta.length?2!==t.delta.length?3===t.delta.length&&0===t.delta[2]&&t.setResult([t.delta[0]]).exit():t.setResult([t.delta[1],t.delta[0]]).exit():t.setResult([t.delta[0],0,0]).exit())):t.setResult(t.delta).exit()};function _(t){if(t&&t.children){for(var e=t.children.length,r=void 0,i=t.result,n=0;n<e;n++)void 0!==(r=t.children[n]).result&&((i=i||{})[r.childName]=r.result);i&&t.leftIsArray&&(i._t="a"),t.setResult(i).exit()}}function w(t){if(!t.leftIsArray&&"object"===t.leftType){var e=void 0,r=void 0,i=t.options.propertyFilter;for(e in t.left)Object.prototype.hasOwnProperty.call(t.left,e)&&(i&&!i(e,t)||(r=new S(t.left[e],t.right[e]),t.push(r,e)));for(e in t.right)Object.prototype.hasOwnProperty.call(t.right,e)&&(i&&!i(e,t)||void 0===t.left[e]&&(r=new S(void 0,t.right[e]),t.push(r,e)));t.children&&0!==t.children.length?t.exit():t.setResult(void 0).exit()}}b.filterName=x.filterName=y.filterName="trivial",_.filterName="collectChildren";var T=function(t){if(t.nested&&!t.delta._t){var e=void 0,r=void 0;for(e in t.delta)r=new m(t.left[e],t.delta[e]),t.push(r,e);t.exit()}};T.filterName=w.filterName="objects";var P=function(t){if(t&&t.children&&!t.delta._t){for(var e=t.children.length,r=void 0,i=0;i<e;i++)r=t.children[i],Object.prototype.hasOwnProperty.call(t.left,r.childName)&&void 0===r.result?delete t.left[r.childName]:t.left[r.childName]!==r.result&&(t.left[r.childName]=r.result);t.setResult(t.left).exit()}};P.filterName="collectChildren";var k=function(t){if(t.nested&&!t.delta._t){var e=void 0,r=void 0;for(e in t.delta)r=new g(t.delta[e]),t.push(r,e);t.exit()}};function I(t){if(t&&t.children&&!t.delta._t){for(var e=t.children.length,r=void 0,i={},n=0;n<e;n++)r=t.children[n],i[r.childName]!==r.result&&(i[r.childName]=r.result);t.setResult(i).exit()}}k.filterName="objects",I.filterName="collectChildren";var A=function(t,e,r,i){return t[r]===e[i]},E={get:function(t,e,r,i){var n=i||{},a=function(t,e,r,i){var n=t.length,a=e.length,o=void 0,s=void 0,l=[n+1];for(o=0;o<n+1;o++){l[o]=[a+1];for(s=0;s<a+1;s++){l[o][s]=0}}for(l.match=r,o=1;o<n+1;o++){for(s=1;s<a+1;s++){if(r(t,e,o-1,s-1,i)){l[o][s]=l[o-1][s-1]+1}else{l[o][s]=Math.max(l[o-1][s],l[o][s-1])}}}return l}(t,e,r||A,n),o=function t(e,r,i,n,a,o){if(n===0||a===0){return{sequence:[],indices1:[],indices2:[]}}if(e.match(r,i,n-1,a-1,o)){var s=t(e,r,i,n-1,a-1,o);s.sequence.push(r[n-1]);s.indices1.push(n-1);s.indices2.push(a-1);return s}if(e[n][a-1]>e[n-1][a]){return t(e,r,i,n,a-1,o)}else{return t(e,r,i,n-1,a,o)}}(a,t,e,t.length,e.length,n);return"string"==typeof t&&"string"==typeof e&&(o.sequence=o.sequence.join("")),o}},C="function"==typeof Array.isArray?Array.isArray:function(t){return t instanceof Array},O="function"==typeof Array.prototype.indexOf?function(t,e){return t.indexOf(e)}:function(t,e){for(var r=t.length,i=0;i<r;i++)if(t[i]===e)return i;return-1};function M(t,e,r,i,n){var a=t[r],o=e[i];if(a===o)return!0;if("object"!==(void 0===a?"undefined":c(a))||"object"!==(void 0===o?"undefined":c(o)))return!1;var s=n.objectHash;if(!s)return n.matchByPosition&&r===i;var l=void 0,p=void 0;return"number"==typeof r?(n.hashCache1=n.hashCache1||[],void 0===(l=n.hashCache1[r])&&(n.hashCache1[r]=l=s(a,r))):l=s(a),void 0!==l&&("number"==typeof i?(n.hashCache2=n.hashCache2||[],void 0===(p=n.hashCache2[i])&&(n.hashCache2[i]=p=s(o,i))):p=s(o),void 0!==p&&l===p)}var D=function(t){if(t.leftIsArray){var e={objectHash:t.options&&t.options.objectHash,matchByPosition:t.options&&t.options.matchByPosition},r=0,i=0,n=void 0,a=void 0,o=void 0,s=t.left,l=t.right,p=s.length,c=l.length,u=void 0;for(0<p&&0<c&&!e.objectHash&&"boolean"!=typeof e.matchByPosition&&(e.matchByPosition=!function(t,e,r,i){for(var n=0;n<r;n++)for(var a=t[n],o=0;o<i;o++){var s=e[o];if(n!==o&&a===s)return!0}}(s,l,p,c));r<p&&r<c&&M(s,l,r,r,e);)n=r,u=new S(t.left[n],t.right[n]),t.push(u,n),r++;for(;i+r<p&&i+r<c&&M(s,l,p-1-i,c-1-i,e);)a=p-1-i,o=c-1-i,u=new S(t.left[a],t.right[o]),t.push(u,o),i++;var h=void 0;if(r+i!==p)if(r+i!==c){delete e.hashCache1,delete e.hashCache2;var d=s.slice(r,p-i),f=l.slice(r,c-i),m=E.get(d,f,M,e),g=[];for(h=h||{_t:"a"},n=r;n<p-i;n++)O(m.indices1,n-r)<0&&(h["_"+n]=[s[n],0,0],g.push(n));var v=!0;t.options&&t.options.arrays&&!1===t.options.arrays.detectMove&&(v=!1);var y=!1;t.options&&t.options.arrays&&t.options.arrays.includeValueOnMove&&(y=!0);var x=g.length;for(n=r;n<c-i;n++){var b=O(m.indices2,n-r);if(b<0){var _=!1;if(v&&0<x)for(var w=0;w<x;w++)if(a=g[w],M(d,f,a-r,n-r,e)){h["_"+a].splice(1,2,n,3),y||(h["_"+a][0]=""),o=n,u=new S(t.left[a],t.right[o]),t.push(u,o),g.splice(w,1),_=!0;break}_||(h[n]=[l[n]])}else a=m.indices1[b]+r,o=m.indices2[b]+r,u=new S(t.left[a],t.right[o]),t.push(u,o)}t.setResult(h).exit()}else{for(h=h||{_t:"a"},n=r;n<p-i;n++)h["_"+n]=[s[n],0,0];t.setResult(h).exit()}else{if(p===c)return void t.setResult(void 0).exit();for(h=h||{_t:"a"},n=r;n<c-i;n++)h[n]=[l[n]];t.setResult(h).exit()}}},F={numerically:function(t,e){return t-e},numericallyBy:function(r){return function(t,e){return t[r]-e[r]}}},R=function(t){if(t.nested&&"a"===t.delta._t){var e=void 0,r=void 0,i=t.delta,n=t.left,a=[],o=[],s=[];for(e in i)if("_t"!==e)if("_"===e[0]){if(0!==i[e][2]&&3!==i[e][2])throw new Error("only removal or move can be applied at original array indices, invalid diff type: "+i[e][2]);a.push(parseInt(e.slice(1),10))}else 1===i[e].length?o.push({index:parseInt(e,10),value:i[e][0]}):s.push({index:parseInt(e,10),delta:i[e]});for(a=a.sort(F.numerically),e=a.length-1;0<=e;e--){r=a[e];var l=i["_"+r],p=n.splice(r,1)[0];3===l[2]&&o.push({index:l[1],value:p})}var c=(o=o.sort(F.numericallyBy("index"))).length;for(e=0;e<c;e++){var u=o[e];n.splice(u.index,0,u.value)}var h=s.length,d=void 0;if(0<h)for(e=0;e<h;e++){var f=s[e];d=new m(t.left[f.index],f.delta),t.push(d,f.index)}t.children?t.exit():t.setResult(t.left).exit()}};R.filterName=D.filterName="arrays";var N=function(t){if(t&&t.children&&"a"===t.delta._t){for(var e=t.children.length,r=void 0,i=0;i<e;i++)r=t.children[i],t.left[r.childName]=r.result;t.setResult(t.left).exit()}};N.filterName="arraysCollectChildren";var L=function(t){if(t.nested){if("a"===t.delta._t){var e=void 0,r=void 0;for(e in t.delta)"_t"!==e&&(r=new g(t.delta[e]),t.push(r,e));t.exit()}}else 3===t.delta[2]&&(t.newName="_"+t.delta[1],t.setResult([t.delta[0],parseInt(t.childName.substr(1),10),3]).exit())};L.filterName="arrays";var B=function(t,e,r){if("string"==typeof e&&"_"===e[0])return parseInt(e.substr(1),10);if(C(r)&&0===r[2])return"_"+e;var i=+e;for(var n in t){var a=t[n];if(C(a))if(3===a[2]){var o=parseInt(n.substr(1),10),s=a[1];if(s===+e)return o;o<=i&&i<s?i++:i<=o&&s<i&&i--}else if(0===a[2]){var l=parseInt(n.substr(1),10);l<=i&&i++}else 1===a.length&&n<=i&&i--}return i};function j(t){if(t&&t.children&&"a"===t.delta._t){for(var e=t.children.length,r=void 0,i={_t:"a"},n=0;n<e;n++){var a=(r=t.children[n]).newName;void 0===a&&(a=B(t.delta,r.childName,r.result)),i[a]!==r.result&&(i[a]=r.result)}t.setResult(i).exit()}}j.filterName="arraysCollectChildren";var $=function(t){t.left instanceof Date?(t.right instanceof Date?t.left.getTime()!==t.right.getTime()?t.setResult([t.left,t.right]):t.setResult(void 0):t.setResult([t.left,t.right]),t.exit()):t.right instanceof Date&&t.setResult([t.left,t.right]).exit()};$.filterName="dates";var V=function(t,e){return t(e={exports:{}},e.exports),e.exports}(function(t){function m(){this.Diff_Timeout=1,this.Diff_EditCost=4,this.Match_Threshold=.5,this.Match_Distance=1e3,this.Patch_DeleteThreshold=.5,this.Patch_Margin=4,this.Match_MaxBits=32}m.prototype.diff_main=function(t,e,r,i){void 0===i&&(i=this.Diff_Timeout<=0?Number.MAX_VALUE:(new Date).getTime()+1e3*this.Diff_Timeout);var n=i;if(null==t||null==e)throw new Error("Null input. (diff_main)");if(t==e)return t?[[0,t]]:[];void 0===r&&(r=!0);var a=r,o=this.diff_commonPrefix(t,e),s=t.substring(0,o);t=t.substring(o),e=e.substring(o),o=this.diff_commonSuffix(t,e);var l=t.substring(t.length-o);t=t.substring(0,t.length-o),e=e.substring(0,e.length-o);var p=this.diff_compute_(t,e,a,n);return s&&p.unshift([0,s]),l&&p.push([0,l]),this.diff_cleanupMerge(p),p},m.prototype.diff_compute_=function(t,e,r,i){var n;if(!t)return[[1,e]];if(!e)return[[-1,t]];var a=t.length>e.length?t:e,o=t.length>e.length?e:t,s=a.indexOf(o);if(-1!=s)return n=[[1,a.substring(0,s)],[0,o],[1,a.substring(s+o.length)]],t.length>e.length&&(n[0][0]=n[2][0]=-1),n;if(1==o.length)return[[-1,t],[1,e]];var l=this.diff_halfMatch_(t,e);if(l){var p=l[0],c=l[1],u=l[2],h=l[3],d=l[4],f=this.diff_main(p,u,r,i),m=this.diff_main(c,h,r,i);return f.concat([[0,d]],m)}return r&&100<t.length&&100<e.length?this.diff_lineMode_(t,e,i):this.diff_bisect_(t,e,i)},m.prototype.diff_lineMode_=function(t,e,r){var i=this.diff_linesToChars_(t,e);t=i.chars1,e=i.chars2;var n=i.lineArray,a=this.diff_main(t,e,!1,r);this.diff_charsToLines_(a,n),this.diff_cleanupSemantic(a),a.push([0,""]);for(var o=0,s=0,l=0,p="",c="";o<a.length;){switch(a[o][0]){case 1:l++,c+=a[o][1];break;case-1:s++,p+=a[o][1];break;case 0:if(1<=s&&1<=l){a.splice(o-s-l,s+l),o=o-s-l;for(var i=this.diff_main(p,c,!1,r),u=i.length-1;0<=u;u--)a.splice(o,0,i[u]);o+=i.length}s=l=0,c=p=""}o++}return a.pop(),a},m.prototype.diff_bisect_=function(t,e,r){for(var i=t.length,n=e.length,a=Math.ceil((i+n)/2),o=a,s=2*a,l=new Array(s),p=new Array(s),c=0;c<s;c++)l[c]=-1,p[c]=-1;l[o+1]=0,p[o+1]=0;for(var u=i-n,h=u%2!=0,d=0,f=0,m=0,g=0,v=0;v<a&&!((new Date).getTime()>r);v++){for(var y=-v+d;y<=v-f;y+=2){for(var x=o+y,b=(P=y==-v||y!=v&&l[x-1]<l[x+1]?l[x+1]:l[x-1]+1)-y;P<i&&b<n&&t.charAt(P)==e.charAt(b);)P++,b++;if(l[x]=P,i<P)f+=2;else if(n<b)d+=2;else if(h){var _=o+u-y;if(0<=_&&_<s&&-1!=p[_]){var w=i-p[_];if(w<=P)return this.diff_bisectSplit_(t,e,P,b,r)}}}for(var S=-v+m;S<=v-g;S+=2){for(var _=o+S,T=(w=S==-v||S!=v&&p[_-1]<p[_+1]?p[_+1]:p[_-1]+1)-S;w<i&&T<n&&t.charAt(i-w-1)==e.charAt(n-T-1);)w++,T++;if(p[_]=w,i<w)g+=2;else if(n<T)m+=2;else if(!h){var x=o+u-S;if(0<=x&&x<s&&-1!=l[x]){var P=l[x],b=o+P-x;if((w=i-w)<=P)return this.diff_bisectSplit_(t,e,P,b,r)}}}}return[[-1,t],[1,e]]},m.prototype.diff_bisectSplit_=function(t,e,r,i,n){var a=t.substring(0,r),o=e.substring(0,i),s=t.substring(r),l=e.substring(i),p=this.diff_main(a,o,!1,n),c=this.diff_main(s,l,!1,n);return p.concat(c)},m.prototype.diff_linesToChars_=function(t,e){var o=[],s={};function r(t){for(var e="",r=0,i=-1,n=o.length;i<t.length-1;){-1==(i=t.indexOf("\n",r))&&(i=t.length-1);var a=t.substring(r,i+1);r=i+1,(s.hasOwnProperty?s.hasOwnProperty(a):void 0!==s[a])?e+=String.fromCharCode(s[a]):(e+=String.fromCharCode(n),s[a]=n,o[n++]=a)}return e}o[0]="";var i=r(t),n=r(e);return{chars1:i,chars2:n,lineArray:o}},m.prototype.diff_charsToLines_=function(t,e){for(var r=0;r<t.length;r++){for(var i=t[r][1],n=[],a=0;a<i.length;a++)n[a]=e[i.charCodeAt(a)];t[r][1]=n.join("")}},m.prototype.diff_commonPrefix=function(t,e){if(!t||!e||t.charAt(0)!=e.charAt(0))return 0;for(var r=0,i=Math.min(t.length,e.length),n=i,a=0;r<n;)t.substring(a,n)==e.substring(a,n)?a=r=n:i=n,n=Math.floor((i-r)/2+r);return n},m.prototype.diff_commonSuffix=function(t,e){if(!t||!e||t.charAt(t.length-1)!=e.charAt(e.length-1))return 0;for(var r=0,i=Math.min(t.length,e.length),n=i,a=0;r<n;)t.substring(t.length-n,t.length-a)==e.substring(e.length-n,e.length-a)?a=r=n:i=n,n=Math.floor((i-r)/2+r);return n},m.prototype.diff_commonOverlap_=function(t,e){var r=t.length,i=e.length;if(0==r||0==i)return 0;i<r?t=t.substring(r-i):r<i&&(e=e.substring(0,r));var n=Math.min(r,i);if(t==e)return n;for(var a=0,o=1;;){var s=t.substring(n-o),l=e.indexOf(s);if(-1==l)return a;o+=l,0!=l&&t.substring(n-o)!=e.substring(0,o)||(a=o,o++)}},m.prototype.diff_halfMatch_=function(t,e){if(this.Diff_Timeout<=0)return null;var r=t.length>e.length?t:e,i=t.length>e.length?e:t;if(r.length<4||2*i.length<r.length)return null;var h=this;function n(t,e,r){for(var i,n,a,o,s=t.substring(r,r+Math.floor(t.length/4)),l=-1,p="";-1!=(l=e.indexOf(s,l+1));){var c=h.diff_commonPrefix(t.substring(r),e.substring(l)),u=h.diff_commonSuffix(t.substring(0,r),e.substring(0,l));p.length<u+c&&(p=e.substring(l-u,l)+e.substring(l,l+c),i=t.substring(0,r-u),n=t.substring(r+c),a=e.substring(0,l-u),o=e.substring(l+c))}return 2*p.length>=t.length?[i,n,a,o,p]:null}var a,o,s,l,p,c=n(r,i,Math.ceil(r.length/4)),u=n(r,i,Math.ceil(r.length/2));if(!c&&!u)return null;a=u?c&&c[4].length>u[4].length?c:u:c,t.length>e.length?(o=a[0],s=a[1],l=a[2],p=a[3]):(l=a[0],p=a[1],o=a[2],s=a[3]);var d=a[4];return[o,s,l,p,d]},m.prototype.diff_cleanupSemantic=function(t){for(var e=!1,r=[],i=0,n=null,a=0,o=0,s=0,l=0,p=0;a<t.length;)0==t[a][0]?(r[i++]=a,o=l,s=p,p=l=0,n=t[a][1]):(1==t[a][0]?l+=t[a][1].length:p+=t[a][1].length,n&&n.length<=Math.max(o,s)&&n.length<=Math.max(l,p)&&(t.splice(r[i-1],0,[-1,n]),t[r[i-1]+1][0]=1,i--,a=0<--i?r[i-1]:-1,p=l=s=o=0,e=!(n=null))),a++;for(e&&this.diff_cleanupMerge(t),this.diff_cleanupSemanticLossless(t),a=1;a<t.length;){if(-1==t[a-1][0]&&1==t[a][0]){var c=t[a-1][1],u=t[a][1],h=this.diff_commonOverlap_(c,u),d=this.diff_commonOverlap_(u,c);d<=h?(h>=c.length/2||h>=u.length/2)&&(t.splice(a,0,[0,u.substring(0,h)]),t[a-1][1]=c.substring(0,c.length-h),t[a+1][1]=u.substring(h),a++):(d>=c.length/2||d>=u.length/2)&&(t.splice(a,0,[0,c.substring(0,d)]),t[a-1][0]=1,t[a-1][1]=u.substring(0,u.length-d),t[a+1][0]=-1,t[a+1][1]=c.substring(d),a++),a++}a++}},m.prototype.diff_cleanupSemanticLossless=function(t){function e(t,e){if(!t||!e)return 6;var r=t.charAt(t.length-1),i=e.charAt(0),n=r.match(m.nonAlphaNumericRegex_),a=i.match(m.nonAlphaNumericRegex_),o=n&&r.match(m.whitespaceRegex_),s=a&&i.match(m.whitespaceRegex_),l=o&&r.match(m.linebreakRegex_),p=s&&i.match(m.linebreakRegex_),c=l&&t.match(m.blanklineEndRegex_),u=p&&e.match(m.blanklineStartRegex_);return c||u?5:l||p?4:n&&!o&&s?3:o||s?2:n||a?1:0}for(var r=1;r<t.length-1;){if(0==t[r-1][0]&&0==t[r+1][0]){var i=t[r-1][1],n=t[r][1],a=t[r+1][1],o=this.diff_commonSuffix(i,n);if(o){var s=n.substring(n.length-o);i=i.substring(0,i.length-o),n=s+n.substring(0,n.length-o),a=s+a}for(var l=i,p=n,c=a,u=e(i,n)+e(n,a);n.charAt(0)===a.charAt(0);){i+=n.charAt(0),n=n.substring(1)+a.charAt(0),a=a.substring(1);var h=e(i,n)+e(n,a);u<=h&&(u=h,l=i,p=n,c=a)}t[r-1][1]!=l&&(l?t[r-1][1]=l:(t.splice(r-1,1),r--),t[r][1]=p,c?t[r+1][1]=c:(t.splice(r+1,1),r--))}r++}},m.nonAlphaNumericRegex_=/[^a-zA-Z0-9]/,m.whitespaceRegex_=/\s/,m.linebreakRegex_=/[\r\n]/,m.blanklineEndRegex_=/\n\r?\n$/,m.blanklineStartRegex_=/^\r?\n\r?\n/,m.prototype.diff_cleanupEfficiency=function(t){for(var e=!1,r=[],i=0,n=null,a=0,o=!1,s=!1,l=!1,p=!1;a<t.length;)0==t[a][0]?(n=t[a][1].length<this.Diff_EditCost&&(l||p)?(r[i++]=a,o=l,s=p,t[a][1]):(i=0,null),l=p=!1):(-1==t[a][0]?p=!0:l=!0,n&&(o&&s&&l&&p||n.length<this.Diff_EditCost/2&&o+s+l+p==3)&&(t.splice(r[i-1],0,[-1,n]),t[r[i-1]+1][0]=1,i--,n=null,o&&s?(l=p=!0,i=0):(a=0<--i?r[i-1]:-1,l=p=!1),e=!0)),a++;e&&this.diff_cleanupMerge(t)},m.prototype.diff_cleanupMerge=function(t){t.push([0,""]);for(var e,r=0,i=0,n=0,a="",o="";r<t.length;)switch(t[r][0]){case 1:n++,o+=t[r][1],r++;break;case-1:i++,a+=t[r][1],r++;break;case 0:1<i+n?(0!==i&&0!==n&&(0!==(e=this.diff_commonPrefix(o,a))&&(0<r-i-n&&0==t[r-i-n-1][0]?t[r-i-n-1][1]+=o.substring(0,e):(t.splice(0,0,[0,o.substring(0,e)]),r++),o=o.substring(e),a=a.substring(e)),0!==(e=this.diff_commonSuffix(o,a))&&(t[r][1]=o.substring(o.length-e)+t[r][1],o=o.substring(0,o.length-e),a=a.substring(0,a.length-e))),0===i?t.splice(r-n,i+n,[1,o]):0===n?t.splice(r-i,i+n,[-1,a]):t.splice(r-i-n,i+n,[-1,a],[1,o]),r=r-i-n+(i?1:0)+(n?1:0)+1):0!==r&&0==t[r-1][0]?(t[r-1][1]+=t[r][1],t.splice(r,1)):r++,i=n=0,o=a=""}""===t[t.length-1][1]&&t.pop();var s=!1;for(r=1;r<t.length-1;)0==t[r-1][0]&&0==t[r+1][0]&&(t[r][1].substring(t[r][1].length-t[r-1][1].length)==t[r-1][1]?(t[r][1]=t[r-1][1]+t[r][1].substring(0,t[r][1].length-t[r-1][1].length),t[r+1][1]=t[r-1][1]+t[r+1][1],t.splice(r-1,1),s=!0):t[r][1].substring(0,t[r+1][1].length)==t[r+1][1]&&(t[r-1][1]+=t[r+1][1],t[r][1]=t[r][1].substring(t[r+1][1].length)+t[r+1][1],t.splice(r+1,1),s=!0)),r++;s&&this.diff_cleanupMerge(t)},m.prototype.diff_xIndex=function(t,e){var r,i=0,n=0,a=0,o=0;for(r=0;r<t.length&&(1!==t[r][0]&&(i+=t[r][1].length),-1!==t[r][0]&&(n+=t[r][1].length),!(e<i));r++)a=i,o=n;return t.length!=r&&-1===t[r][0]?o:o+(e-a)},m.prototype.diff_prettyHtml=function(t){for(var e=[],r=/&/g,i=/</g,n=/>/g,a=/\n/g,o=0;o<t.length;o++){var s=t[o][0],l=t[o][1],p=l.replace(r,"&amp;").replace(i,"&lt;").replace(n,"&gt;").replace(a,"&para;<br>");switch(s){case 1:e[o]='<ins style="background:#e6ffe6;">'+p+"</ins>";break;case-1:e[o]='<del style="background:#ffe6e6;">'+p+"</del>";break;case 0:e[o]="<span>"+p+"</span>"}}return e.join("")},m.prototype.diff_text1=function(t){for(var e=[],r=0;r<t.length;r++)1!==t[r][0]&&(e[r]=t[r][1]);return e.join("")},m.prototype.diff_text2=function(t){for(var e=[],r=0;r<t.length;r++)-1!==t[r][0]&&(e[r]=t[r][1]);return e.join("")},m.prototype.diff_levenshtein=function(t){for(var e=0,r=0,i=0,n=0;n<t.length;n++){var a=t[n][0],o=t[n][1];switch(a){case 1:r+=o.length;break;case-1:i+=o.length;break;case 0:e+=Math.max(r,i),i=r=0}}return e+=Math.max(r,i)},m.prototype.diff_toDelta=function(t){for(var e=[],r=0;r<t.length;r++)switch(t[r][0]){case 1:e[r]="+"+encodeURI(t[r][1]);break;case-1:e[r]="-"+t[r][1].length;break;case 0:e[r]="="+t[r][1].length}return e.join("\t").replace(/%20/g," ")},m.prototype.diff_fromDelta=function(t,e){for(var r=[],i=0,n=0,a=e.split(/\t/g),o=0;o<a.length;o++){var s=a[o].substring(1);switch(a[o].charAt(0)){case"+":try{r[i++]=[1,decodeURI(s)]}catch(t){throw new Error("Illegal escape in diff_fromDelta: "+s)}break;case"-":case"=":var l=parseInt(s,10);if(isNaN(l)||l<0)throw new Error("Invalid number in diff_fromDelta: "+s);var p=t.substring(n,n+=l);"="==a[o].charAt(0)?r[i++]=[0,p]:r[i++]=[-1,p];break;default:if(a[o])throw new Error("Invalid diff operation in diff_fromDelta: "+a[o])}}if(n!=t.length)throw new Error("Delta length ("+n+") does not equal source text length ("+t.length+").");return r},m.prototype.match_main=function(t,e,r){if(null==t||null==e||null==r)throw new Error("Null input. (match_main)");return r=Math.max(0,Math.min(r,t.length)),t==e?0:t.length?t.substring(r,r+e.length)==e?r:this.match_bitap_(t,e,r):-1},m.prototype.match_bitap_=function(t,n,a){if(n.length>this.Match_MaxBits)throw new Error("Pattern too long for this browser.");var e=this.match_alphabet_(n),o=this;function r(t,e){var r=t/n.length,i=Math.abs(a-e);return o.Match_Distance?r+i/o.Match_Distance:i?1:r}var i=this.Match_Threshold,s=t.indexOf(n,a);-1!=s&&(i=Math.min(r(0,s),i),-1!=(s=t.lastIndexOf(n,a+n.length))&&(i=Math.min(r(0,s),i)));var l,p,c=1<<n.length-1;s=-1;for(var u,h=n.length+t.length,d=0;d<n.length;d++){for(l=0,p=h;l<p;)r(d,a+p)<=i?l=p:h=p,p=Math.floor((h-l)/2+l);h=p;var f=Math.max(1,a-p+1),m=Math.min(a+p,t.length)+n.length,g=Array(m+2);g[m+1]=(1<<d)-1;for(var v=m;f<=v;v--){var y=e[t.charAt(v-1)];if(g[v]=0===d?(g[v+1]<<1|1)&y:(g[v+1]<<1|1)&y|(u[v+1]|u[v])<<1|1|u[v+1],g[v]&c){var x=r(d,v-1);if(x<=i){if(i=x,!(a<(s=v-1)))break;f=Math.max(1,2*a-s)}}}if(r(d+1,a)>i)break;u=g}return s},m.prototype.match_alphabet_=function(t){for(var e={},r=0;r<t.length;r++)e[t.charAt(r)]=0;for(var r=0;r<t.length;r++)e[t.charAt(r)]|=1<<t.length-r-1;return e},m.prototype.patch_addContext_=function(t,e){if(0!=e.length){for(var r=e.substring(t.start2,t.start2+t.length1),i=0;e.indexOf(r)!=e.lastIndexOf(r)&&r.length<this.Match_MaxBits-this.Patch_Margin-this.Patch_Margin;)i+=this.Patch_Margin,r=e.substring(t.start2-i,t.start2+t.length1+i);i+=this.Patch_Margin;var n=e.substring(t.start2-i,t.start2);n&&t.diffs.unshift([0,n]);var a=e.substring(t.start2+t.length1,t.start2+t.length1+i);a&&t.diffs.push([0,a]),t.start1-=n.length,t.start2-=n.length,t.length1+=n.length+a.length,t.length2+=n.length+a.length}},m.prototype.patch_make=function(t,e,r){var i,n;if("string"==typeof t&&"string"==typeof e&&void 0===r)i=t,2<(n=this.diff_main(i,e,!0)).length&&(this.diff_cleanupSemantic(n),this.diff_cleanupEfficiency(n));else if(t&&"object"==typeof t&&void 0===e&&void 0===r)n=t,i=this.diff_text1(n);else if("string"==typeof t&&e&&"object"==typeof e&&void 0===r)i=t,n=e;else{if("string"!=typeof t||"string"!=typeof e||!r||"object"!=typeof r)throw new Error("Unknown call format to patch_make.");i=t,n=r}if(0===n.length)return[];for(var a=[],o=new m.patch_obj,s=0,l=0,p=0,c=i,u=i,h=0;h<n.length;h++){var d=n[h][0],f=n[h][1];switch(s||0===d||(o.start1=l,o.start2=p),d){case 1:o.diffs[s++]=n[h],o.length2+=f.length,u=u.substring(0,p)+f+u.substring(p);break;case-1:o.length1+=f.length,o.diffs[s++]=n[h],u=u.substring(0,p)+u.substring(p+f.length);break;case 0:f.length<=2*this.Patch_Margin&&s&&n.length!=h+1?(o.diffs[s++]=n[h],o.length1+=f.length,o.length2+=f.length):f.length>=2*this.Patch_Margin&&s&&(this.patch_addContext_(o,c),a.push(o),o=new m.patch_obj,s=0,c=u,l=p)}1!==d&&(l+=f.length),-1!==d&&(p+=f.length)}return s&&(this.patch_addContext_(o,c),a.push(o)),a},m.prototype.patch_deepCopy=function(t){for(var e=[],r=0;r<t.length;r++){var i=t[r],n=new m.patch_obj;n.diffs=[];for(var a=0;a<i.diffs.length;a++)n.diffs[a]=i.diffs[a].slice();n.start1=i.start1,n.start2=i.start2,n.length1=i.length1,n.length2=i.length2,e[r]=n}return e},m.prototype.patch_apply=function(t,e){if(0==t.length)return[e,[]];t=this.patch_deepCopy(t);var r=this.patch_addPadding(t);e=r+e+r,this.patch_splitMax(t);for(var i=0,n=[],a=0;a<t.length;a++){var o,s,l=t[a].start2+i,p=this.diff_text1(t[a].diffs),c=-1;if(p.length>this.Match_MaxBits?-1!=(o=this.match_main(e,p.substring(0,this.Match_MaxBits),l))&&(-1==(c=this.match_main(e,p.substring(p.length-this.Match_MaxBits),l+p.length-this.Match_MaxBits))||c<=o)&&(o=-1):o=this.match_main(e,p,l),-1==o)n[a]=!1,i-=t[a].length2-t[a].length1;else if(n[a]=!0,i=o-l,s=-1==c?e.substring(o,o+p.length):e.substring(o,c+this.Match_MaxBits),p==s)e=e.substring(0,o)+this.diff_text2(t[a].diffs)+e.substring(o+p.length);else{var u=this.diff_main(p,s,!1);if(p.length>this.Match_MaxBits&&this.diff_levenshtein(u)/p.length>this.Patch_DeleteThreshold)n[a]=!1;else{this.diff_cleanupSemanticLossless(u);for(var h,d=0,f=0;f<t[a].diffs.length;f++){var m=t[a].diffs[f];0!==m[0]&&(h=this.diff_xIndex(u,d)),1===m[0]?e=e.substring(0,o+h)+m[1]+e.substring(o+h):-1===m[0]&&(e=e.substring(0,o+h)+e.substring(o+this.diff_xIndex(u,d+m[1].length))),-1!==m[0]&&(d+=m[1].length)}}}}return[e=e.substring(r.length,e.length-r.length),n]},m.prototype.patch_addPadding=function(t){for(var e=this.Patch_Margin,r="",i=1;i<=e;i++)r+=String.fromCharCode(i);for(var i=0;i<t.length;i++)t[i].start1+=e,t[i].start2+=e;var n=t[0],a=n.diffs;if(0==a.length||0!=a[0][0])a.unshift([0,r]),n.start1-=e,n.start2-=e,n.length1+=e,n.length2+=e;else if(e>a[0][1].length){var o=e-a[0][1].length;a[0][1]=r.substring(a[0][1].length)+a[0][1],n.start1-=o,n.start2-=o,n.length1+=o,n.length2+=o}if(n=t[t.length-1],0==(a=n.diffs).length||0!=a[a.length-1][0])a.push([0,r]),n.length1+=e,n.length2+=e;else if(e>a[a.length-1][1].length){var o=e-a[a.length-1][1].length;a[a.length-1][1]+=r.substring(0,o),n.length1+=o,n.length2+=o}return r},m.prototype.patch_splitMax=function(t){for(var e=this.Match_MaxBits,r=0;r<t.length;r++)if(!(t[r].length1<=e)){var i=t[r];t.splice(r--,1);for(var n=i.start1,a=i.start2,o="";0!==i.diffs.length;){var s=new m.patch_obj,l=!0;for(s.start1=n-o.length,s.start2=a-o.length,""!==o&&(s.length1=s.length2=o.length,s.diffs.push([0,o]));0!==i.diffs.length&&s.length1<e-this.Patch_Margin;){var p=i.diffs[0][0],c=i.diffs[0][1];1===p?(s.length2+=c.length,a+=c.length,s.diffs.push(i.diffs.shift()),l=!1):-1===p&&1==s.diffs.length&&0==s.diffs[0][0]&&c.length>2*e?(s.length1+=c.length,n+=c.length,l=!1,s.diffs.push([p,c]),i.diffs.shift()):(c=c.substring(0,e-s.length1-this.Patch_Margin),s.length1+=c.length,n+=c.length,0===p?(s.length2+=c.length,a+=c.length):l=!1,s.diffs.push([p,c]),c==i.diffs[0][1]?i.diffs.shift():i.diffs[0][1]=i.diffs[0][1].substring(c.length))}o=(o=this.diff_text2(s.diffs)).substring(o.length-this.Patch_Margin);var u=this.diff_text1(i.diffs).substring(0,this.Patch_Margin);""!==u&&(s.length1+=u.length,s.length2+=u.length,0!==s.diffs.length&&0===s.diffs[s.diffs.length-1][0]?s.diffs[s.diffs.length-1][1]+=u:s.diffs.push([0,u])),l||t.splice(++r,0,s)}}},m.prototype.patch_toText=function(t){for(var e=[],r=0;r<t.length;r++)e[r]=t[r];return e.join("")},m.prototype.patch_fromText=function(t){var e=[];if(!t)return e;for(var r=t.split("\n"),i=0,n=/^@@ -(\d+),?(\d*) \+(\d+),?(\d*) @@$/;i<r.length;){var a=r[i].match(n);if(!a)throw new Error("Invalid patch string: "+r[i]);var o=new m.patch_obj;for(e.push(o),o.start1=parseInt(a[1],10),""===a[2]?(o.start1--,o.length1=1):"0"==a[2]?o.length1=0:(o.start1--,o.length1=parseInt(a[2],10)),o.start2=parseInt(a[3],10),""===a[4]?(o.start2--,o.length2=1):"0"==a[4]?o.length2=0:(o.start2--,o.length2=parseInt(a[4],10)),i++;i<r.length;){var s=r[i].charAt(0);try{var l=decodeURI(r[i].substring(1))}catch(t){throw new Error("Illegal escape in patch_fromText: "+l)}if("-"==s)o.diffs.push([-1,l]);else if("+"==s)o.diffs.push([1,l]);else if(" "==s)o.diffs.push([0,l]);else{if("@"==s)break;if(""!==s)throw new Error('Invalid patch mode "'+s+'" in: '+l)}i++}}return e},(m.patch_obj=function(){this.diffs=[],this.start1=null,this.start2=null,this.length1=0,this.length2=0}).prototype.toString=function(){var t,e;t=0===this.length1?this.start1+",0":1==this.length1?this.start1+1:this.start1+1+","+this.length1,e=0===this.length2?this.start2+",0":1==this.length2?this.start2+1:this.start2+1+","+this.length2;for(var r,i=["@@ -"+t+" +"+e+" @@\n"],n=0;n<this.diffs.length;n++){switch(this.diffs[n][0]){case 1:r="+";break;case-1:r="-";break;case 0:r=" "}i[n+1]=r+encodeURI(this.diffs[n][1])+"\n"}return i.join("").replace(/%20/g," ")},t.exports=m,t.exports.diff_match_patch=m,t.exports.DIFF_DELETE=-1,t.exports.DIFF_INSERT=1,t.exports.DIFF_EQUAL=0}),H=null,z=function(t){if(!H){var a=void 0;if("undefined"!=typeof diff_match_patch)a="function"==typeof diff_match_patch?new diff_match_patch:new diff_match_patch.diff_match_patch;else if(V)try{a=V&&new V}catch(t){a=null}if(!a){if(!t)return null;var e=new Error("text diff_match_patch library not found");throw e.diff_match_patch_not_found=!0,e}H={diff:function(t,e){return a.patch_toText(a.patch_make(t,e))},patch:function(t,e){for(var r=a.patch_apply(a.patch_fromText(e),t),i=0;i<r[1].length;i++)if(!r[1][i]){var n=new Error("text patch failed");n.textPatchFailed=!0}return r[0]}}}return H},G=function(t){if("string"===t.leftType){var e=t.options&&t.options.textDiff&&t.options.textDiff.minLength||60;if(t.left.length<e||t.right.length<e)t.setResult([t.left,t.right]).exit();else{var r=z();if(r){var i=r.diff;t.setResult([i(t.left,t.right),0,2]).exit()}else t.setResult([t.left,t.right]).exit()}}},Y=function(t){if(!t.nested&&2===t.delta[2]){var e=z(!0).patch;t.setResult(e(t.left,t.delta[0])).exit()}},X=function(t){t.nested||2===t.delta[2]&&t.setResult([function(t){var e=void 0,r=void 0,i=void 0,n=void 0,a=void 0,o=null,s=/^@@ +-(\d+),(\d+) +\+(\d+),(\d+) +@@$/,l=void 0;for(i=t.split("\n"),e=0,r=i.length;e<r;e++){n=i[e];var p=n.slice(0,1);if(p==="@"){o=s.exec(n);l=e;i[l]="@@ -"+o[3]+","+o[4]+" +"+o[1]+","+o[2]+" @@"}else if(p==="+"){i[e]="-"+i[e].slice(1);if(i[e-1].slice(0,1)==="+"){a=i[e];i[e]=i[e-1];i[e-1]=a}}else if(p==="-"){i[e]="+"+i[e].slice(1)}}return i.join("\n")}(t.delta[0]),0,2]).exit()};X.filterName=Y.filterName=G.filterName="texts";var W=function(){function e(t){n(this,e),this.processor=new p(t),this.processor.pipe(new u("diff").append(_,y,$,G,w,D).shouldHaveResult()),this.processor.pipe(new u("patch").append(P,N,x,Y,T,R).shouldHaveResult()),this.processor.pipe(new u("reverse").append(I,j,b,X,k,L).shouldHaveResult())}return r(e,[{key:"options",value:function(){var t;return(t=this.processor).options.apply(t,arguments)}},{key:"diff",value:function(t,e){return this.processor.process(new S(t,e))}},{key:"patch",value:function(t,e){return this.processor.process(new m(t,e))}},{key:"reverse",value:function(t){return this.processor.process(new g(t))}},{key:"unpatch",value:function(t,e){return this.patch(t,this.reverse(e))}},{key:"clone",value:function(t){return f(t)}}]),e}(),U="function"==typeof Array.isArray?Array.isArray:function(t){return t instanceof Array},q="function"==typeof Object.keys?function(t){return Object.keys(t)}:function(t){var e=[];for(var r in t)Object.prototype.hasOwnProperty.call(t,r)&&e.push(r);return e},J=function(t){return"_"===t.substr(0,1)?t.slice(1):t},Q=function(t){return"_t"===t?-1:"_"===t.substr(0,1)?parseInt(t.slice(1),10):parseInt(t,10)+.1},Z=function(t,e){return Q(t)-Q(e)},K=function(){function t(){n(this,t)}return r(t,[{key:"format",value:function(t,e){var r={};return this.prepareContext(r),this.recurse(r,t,e),this.finalize(r)}},{key:"prepareContext",value:function(t){t.buffer=[],t.out=function(){var t;(t=this.buffer).push.apply(t,arguments)}}},{key:"typeFormattterNotFound",value:function(t,e){throw new Error("cannot format delta type: "+e)}},{key:"typeFormattterErrorFormatter",value:function(t,e){return e.toString()}},{key:"finalize",value:function(t){var e=t.buffer;if(U(e))return e.join("")}},{key:"recurse",value:function(e,r,t,i,n,a,o){var s=r&&a,l=s?a.value:t;if(void 0!==r||void 0!==i){var p=this.getDeltaType(r,a),c="node"===p?"a"===r._t?"array":"object":"";void 0!==i?this.nodeBegin(e,i,n,p,c,o):this.rootBegin(e,p,c);try{(this["format_"+p]||this.typeFormattterNotFound(e,p)).call(this,e,r,l,i,n,a)}catch(t){this.typeFormattterErrorFormatter(e,t,r,l,i,n,a),"undefined"!=typeof console&&console.error&&console.error(t.stack)}void 0!==i?this.nodeEnd(e,i,n,p,c,o):this.rootEnd(e,p,c)}}},{key:"formatDeltaChildren",value:function(n,a,o){var s=this;this.forEachDeltaKey(a,o,function(t,e,r,i){s.recurse(n,a[t],o?o[e]:void 0,t,e,r,i)})}},{key:"forEachDeltaKey",value:function(t,e,r){var i=q(t),n="a"===t._t,a={},o=void 0;if(void 0!==e)for(o in e)Object.prototype.hasOwnProperty.call(e,o)&&(void 0!==t[o]||n&&void 0!==t["_"+o]||i.push(o));for(o in t)if(Object.prototype.hasOwnProperty.call(t,o)){var s=t[o];U(s)&&3===s[2]&&!(a[s[1].toString()]={key:o,value:e&&e[parseInt(o.substr(1))]})!==this.includeMoveDestinations&&void 0===e&&void 0===t[s[1]]&&i.push(s[1].toString())}n?i.sort(Z):i.sort();for(var l=0,p=i.length;l<p;l++){var c=i[l];if(!n||"_t"!==c){var u=n?"number"==typeof c?c:parseInt(J(c),10):c,h=l===p-1;r(c,u,a[u],h)}}}},{key:"getDeltaType",value:function(t,e){if(void 0===t)return void 0!==e?"movedestination":"unchanged";if(U(t)){if(1===t.length)return"added";if(2===t.length)return"modified";if(3===t.length&&0===t[2])return"deleted";if(3===t.length&&2===t[2])return"textdiff";if(3===t.length&&3===t[2])return"moved"}else if("object"===(void 0===t?"undefined":c(t)))return"node";return"unknown"}},{key:"parseTextDiff",value:function(t){for(var e=[],r=t.split("\n@@ "),i=0,n=r.length;i<n;i++){var a=r[i],o={pieces:[]},s=/^(?:@@ )?[-+]?(\d+),(\d+)/.exec(a).slice(1);o.location={line:s[0],chr:s[1]};for(var l=a.split("\n").slice(1),p=0,c=l.length;p<c;p++){var u=l[p];if(u.length){var h={type:"context"};"+"===u.substr(0,1)?h.type="added":"-"===u.substr(0,1)&&(h.type="deleted"),h.text=u.slice(1),o.pieces.push(h)}}e.push(o)}return e}}]),t}(),tt=Object.freeze({default:K}),et=function(t){function e(){return n(this,e),o(this,(e.__proto__||Object.getPrototypeOf(e)).apply(this,arguments))}return a(e,t),r(e,[{key:"typeFormattterErrorFormatter",value:function(t,e){t.out('<pre class="jsondiffpatch-error">'+e+"</pre>")}},{key:"formatValue",value:function(t,e){t.out("<pre>"+rt(JSON.stringify(e,null,2))+"</pre>")}},{key:"formatTextDiffString",value:function(t,e){var r=this.parseTextDiff(e);t.out('<ul class="jsondiffpatch-textdiff">');for(var i=0,n=r.length;i<n;i++){var a=r[i];t.out('<li><div class="jsondiffpatch-textdiff-location"><span class="jsondiffpatch-textdiff-line-number">'+a.location.line+'</span><span class="jsondiffpatch-textdiff-char">'+a.location.chr+'</span></div><div class="jsondiffpatch-textdiff-line">');for(var o=a.pieces,s=0,l=o.length;s<l;s++){var p=o[s];t.out('<span class="jsondiffpatch-textdiff-'+p.type+'">'+rt(decodeURI(p.text))+"</span>")}t.out("</div></li>")}t.out("</ul>")}},{key:"rootBegin",value:function(t,e,r){var i="jsondiffpatch-"+e+(r?" jsondiffpatch-child-node-type-"+r:"");t.out('<div class="jsondiffpatch-delta '+i+'">')}},{key:"rootEnd",value:function(t){t.out("</div>"+(t.hasArrows?'<script type="text/javascript">setTimeout('+it.toString()+",10);<\/script>":""))}},{key:"nodeBegin",value:function(t,e,r,i,n){var a="jsondiffpatch-"+i+(n?" jsondiffpatch-child-node-type-"+n:"");t.out('<li class="'+a+'" data-key="'+r+'"><div class="jsondiffpatch-property-name">'+r+"</div>")}},{key:"nodeEnd",value:function(t){t.out("</li>")}},{key:"format_unchanged",value:function(t,e,r){void 0!==r&&(t.out('<div class="jsondiffpatch-value">'),this.formatValue(t,r),t.out("</div>"))}},{key:"format_movedestination",value:function(t,e,r){void 0!==r&&(t.out('<div class="jsondiffpatch-value">'),this.formatValue(t,r),t.out("</div>"))}},{key:"format_node",value:function(t,e,r){var i="a"===e._t?"array":"object";t.out('<ul class="jsondiffpatch-node jsondiffpatch-node-type-'+i+'">'),this.formatDeltaChildren(t,e,r),t.out("</ul>")}},{key:"format_added",value:function(t,e){t.out('<div class="jsondiffpatch-value">'),this.formatValue(t,e[0]),t.out("</div>")}},{key:"format_modified",value:function(t,e){t.out('<div class="jsondiffpatch-value jsondiffpatch-left-value">'),this.formatValue(t,e[0]),t.out('</div><div class="jsondiffpatch-value jsondiffpatch-right-value">'),this.formatValue(t,e[1]),t.out("</div>")}},{key:"format_deleted",value:function(t,e){t.out('<div class="jsondiffpatch-value">'),this.formatValue(t,e[0]),t.out("</div>")}},{key:"format_moved",value:function(t,e){t.out('<div class="jsondiffpatch-value">'),this.formatValue(t,e[0]),t.out('</div><div class="jsondiffpatch-moved-destination">'+e[1]+"</div>"),t.out('<div class="jsondiffpatch-arrow" style="position: relative; left: -34px;">\n          <svg width="30" height="60" style="position: absolute; display: none;">\n          <defs>\n              <marker id="markerArrow" markerWidth="8" markerHeight="8"\n                 refx="2" refy="4"\n                     orient="auto" markerUnits="userSpaceOnUse">\n                  <path d="M1,1 L1,7 L7,4 L1,1" style="fill: #339;" />\n              </marker>\n          </defs>\n          <path d="M30,0 Q-10,25 26,50"\n            style="stroke: #88f; stroke-width: 2px; fill: none; stroke-opacity: 0.5; marker-end: url(#markerArrow);"\n          ></path>\n          </svg>\n      </div>'),t.hasArrows=!0}},{key:"format_textdiff",value:function(t,e){t.out('<div class="jsondiffpatch-value">'),this.formatTextDiffString(t,e[0]),t.out("</div>")}}]),e}(K);function rt(t){for(var e=t,r=[[/&/g,"&amp;"],[/</g,"&lt;"],[/>/g,"&gt;"],[/'/g,"&apos;"],[/"/g,"&quot;"]],i=0;i<r.length;i++)e=e.replace(r[i][0],r[i][1]);return e}var it=function(t){var e=t||document;!function(t,e,r){for(var i=t.querySelectorAll(e),n=0,a=i.length;n<a;n++){r(i[n])}}(e,".jsondiffpatch-arrow",function(t){var e=t.parentNode,r=t.children,i=t.style,n=e,a=r[0],o=a.children[1];a.style.display="none";var s=function(t){var e=t.textContent,r=t.innerText;return e||r}(n.querySelector(".jsondiffpatch-moved-destination")),l=n.parentNode,p=void 0;if(function(t,e){for(var r=t.children,i=0,n=r.length;i<n;i++){e(r[i],i)}}(l,function(t){t.getAttribute("data-key")===s&&(p=t)}),p)try{var c=p.offsetTop-n.offsetTop;a.setAttribute("height",Math.abs(c)+6),i.top=(0<c?0:c)-8+"px";var u=0<c?"M30,0 Q-10,"+Math.round(c/2)+" 26,"+(c-4):"M30,"+-c+" Q-10,"+Math.round(-c/2)+" 26,4";o.setAttribute("d",u),a.style.display=""}catch(t){}})},nt=function(t,e,r){var i=e||document.body,n="jsondiffpatch-unchanged-",a={showing:n+"showing",hiding:n+"hiding",visible:n+"visible",hidden:n+"hidden"},o=i.classList;if(o){if(!r)return o.remove(a.showing),o.remove(a.hiding),o.remove(a.visible),o.remove(a.hidden),void(!1===t&&o.add(a.hidden));!1===t?(o.remove(a.showing),o.add(a.visible),setTimeout(function(){o.add(a.hiding)},10)):(o.remove(a.hiding),o.add(a.showing),o.remove(a.hidden));var s=setInterval(function(){it(i)},100);setTimeout(function(){o.remove(a.showing),o.remove(a.hiding),!1===t?(o.add(a.hidden),o.remove(a.visible)):(o.add(a.visible),o.remove(a.hidden)),setTimeout(function(){o.remove(a.visible),clearInterval(s)},r+400)},r)}},at=void 0;var ot=Object.freeze({showUnchanged:nt,hideUnchanged:function(t,e){return nt(false,t,e)},default:et,format:function(t,e){at||(at=new et);return at.format(t,e)}}),st=function(t){function e(){n(this,e);var t=o(this,(e.__proto__||Object.getPrototypeOf(e)).call(this));return t.includeMoveDestinations=!1,t}return a(e,t),r(e,[{key:"prepareContext",value:function(r){i(e.prototype.__proto__||Object.getPrototypeOf(e.prototype),"prepareContext",this).call(this,r),r.indent=function(t){this.indentLevel=(this.indentLevel||0)+(void 0===t?1:t),this.indentPad=new Array(this.indentLevel+1).join("&nbsp;&nbsp;")},r.row=function(t,e){r.out('<tr><td style="white-space: nowrap;"><pre class="jsondiffpatch-annotated-indent" style="display: inline-block">'),r.out(r.indentPad),r.out('</pre><pre style="display: inline-block">'),r.out(t),r.out('</pre></td><td class="jsondiffpatch-delta-note"><div>'),r.out(e),r.out("</div></td></tr>")}}},{key:"typeFormattterErrorFormatter",value:function(t,e){t.row("",'<pre class="jsondiffpatch-error">'+e+"</pre>")}},{key:"formatTextDiffString",value:function(t,e){var r=this.parseTextDiff(e);t.out('<ul class="jsondiffpatch-textdiff">');for(var i=0,n=r.length;i<n;i++){var a=r[i];t.out('<li><div class="jsondiffpatch-textdiff-location"><span class="jsondiffpatch-textdiff-line-number">'+a.location.line+'</span><span class="jsondiffpatch-textdiff-char">'+a.location.chr+'</span></div><div class="jsondiffpatch-textdiff-line">');for(var o=a.pieces,s=0,l=o.length;s<l;s++){var p=o[s];t.out('<span class="jsondiffpatch-textdiff-'+p.type+'">'+p.text+"</span>")}t.out("</div></li>")}t.out("</ul>")}},{key:"rootBegin",value:function(t,e,r){t.out('<table class="jsondiffpatch-annotated-delta">'),"node"===e&&(t.row("{"),t.indent()),"array"===r&&t.row('"_t": "a",',"Array delta (member names indicate array indices)")}},{key:"rootEnd",value:function(t,e){"node"===e&&(t.indent(-1),t.row("}")),t.out("</table>")}},{key:"nodeBegin",value:function(t,e,r,i,n){t.row("&quot;"+e+"&quot;: {"),"node"===i&&t.indent(),"array"===n&&t.row('"_t": "a",',"Array delta (member names indicate array indices)")}},{key:"nodeEnd",value:function(t,e,r,i,n,a){"node"===i&&t.indent(-1),t.row("}"+(a?"":","))}},{key:"format_unchanged",value:function(){}},{key:"format_movedestination",value:function(){}},{key:"format_node",value:function(t,e,r){this.formatDeltaChildren(t,e,r)}}]),e}(K),lt=function(t){return'<pre style="display:inline-block">&quot;'+t+"&quot;</pre>"},pt={added:function(t,e,r,i){var n=" <pre>([newValue])</pre>";return void 0===i?"new value"+n:"number"==typeof i?"insert at index "+i+n:"add property "+lt(i)+n},modified:function(t,e,r,i){var n=" <pre>([previousValue, newValue])</pre>";return void 0===i?"modify value"+n:"number"==typeof i?"modify at index "+i+n:"modify property "+lt(i)+n},deleted:function(t,e,r,i){var n=" <pre>([previousValue, 0, 0])</pre>";return void 0===i?"delete value"+n:"number"==typeof i?"remove index "+i+n:"delete property "+lt(i)+n},moved:function(t,e,r,i){return'move from <span title="(position to remove at original state)">index '+i+'</span> to <span title="(position to insert at final state)">index '+t[1]+"</span>"},textdiff:function(t,e,r,i){var n=void 0===i?"":"number"==typeof i?" at index "+i:" at property "+lt(i);return"text diff"+n+', format is <a href="https://code.google.com/p/google-diff-match-patch/wiki/Unidiff">a variation of Unidiff</a>'}},ct=function(t,e){var r=this.getDeltaType(e),i=pt[r],n=i&&i.apply(i,Array.prototype.slice.call(arguments,1)),a=JSON.stringify(e,null,2);"textdiff"===r&&(a=a.split("\\n").join('\\n"+\n   "')),t.indent(),t.row(a,n),t.indent(-1)};st.prototype.format_added=ct,st.prototype.format_modified=ct,st.prototype.format_deleted=ct,st.prototype.format_moved=ct,st.prototype.format_textdiff=ct;var ut=void 0;var ht=Object.freeze({default:st,format:function(t,e){ut||(ut=new st);return ut.format(t,e)}}),dt={add:"add",remove:"remove",replace:"replace",move:"move"},ft=function(t){function e(){n(this,e);var t=o(this,(e.__proto__||Object.getPrototypeOf(e)).call(this));return t.includeMoveDestinations=!0,t}return a(e,t),r(e,[{key:"prepareContext",value:function(t){i(e.prototype.__proto__||Object.getPrototypeOf(e.prototype),"prepareContext",this).call(this,t),t.result=[],t.path=[],t.pushCurrentOp=function(t){var e=t.op,r=t.value,i={op:e,path:this.currentPath()};void 0!==r&&(i.value=r),this.result.push(i)},t.pushMoveOp=function(t){var e=this.currentPath();this.result.push({op:dt.move,from:e,path:this.toPath(t)})},t.currentPath=function(){return"/"+this.path.join("/")},t.toPath=function(t){var e=this.path.slice();return e[e.length-1]=t,"/"+e.join("/")}}},{key:"typeFormattterErrorFormatter",value:function(t,e){t.out("[ERROR] "+e)}},{key:"rootBegin",value:function(){}},{key:"rootEnd",value:function(){}},{key:"nodeBegin",value:function(t,e,r){var i=t.path;i.push(r)}},{key:"nodeEnd",value:function(t){var e=t.path;e.pop()}},{key:"format_unchanged",value:function(){}},{key:"format_movedestination",value:function(){}},{key:"format_node",value:function(t,e,r){this.formatDeltaChildren(t,e,r)}},{key:"format_added",value:function(t,e){t.pushCurrentOp({op:dt.add,value:e[0]})}},{key:"format_modified",value:function(t,e){t.pushCurrentOp({op:dt.replace,value:e[1]})}},{key:"format_deleted",value:function(t){t.pushCurrentOp({op:dt.remove})}},{key:"format_moved",value:function(t,e){var r=e[1];t.pushMoveOp(r)}},{key:"format_textdiff",value:function(){throw new Error("Not implemented")}},{key:"format",value:function(t,e){var r={};return this.prepareContext(r),this.recurse(r,t,e),r.result}}]),e}(K),mt=function(t){return t[t.length-1]},gt=function(t){return function(t,e){return t.sort(e),t}(t,function(t,e){var r=t.path.split("/"),i=e.path.split("/");return r.length!==i.length?r.length-i.length:function(t,e){var r=parseInt(t,10),i=parseInt(e,10);if(!(isNaN(r)||isNaN(i))){return i-r}else{return 0}}(mt(r),mt(i))})},vt=function(t,r){var e=Array(r.length+1).fill().map(function(){return[]});return t.map(function(e){var t=r.map(function(t){return t(e)}).indexOf(!0);return t<0&&(t=r.length),{item:e,position:t}}).reduce(function(t,e){return t[e.position].push(e.item),t},e)},yt=function(t){var e=t.op;return"move"===e},xt=function(t){var e=t.op;return"remove"===e},bt=void 0,_t=function(t,e){return bt||(bt=new ft),function(t){var e=vt(t,[yt,xt]),r=s(e,3),i=r[0],n=r[1],a=r[2],o=gt(n);return[].concat(l(o),l(i),l(a))}(bt.format(t,e))},wt=Object.freeze({default:ft,partitionOps:vt,format:_t,log:function(t,e){console.log(_t(t,e))}});function St(t){return e&&e[t]||function(){for(var t=arguments.length,e=Array(t),r=0;r<t;r++)e[r]=arguments[r];return e}}var Tt={added:St("green"),deleted:St("red"),movedestination:St("gray"),moved:St("yellow"),unchanged:St("gray"),error:St("white.bgRed"),textDiffLine:St("gray")},Pt=function(t){function e(){n(this,e);var t=o(this,(e.__proto__||Object.getPrototypeOf(e)).call(this));return t.includeMoveDestinations=!1,t}return a(e,t),r(e,[{key:"prepareContext",value:function(t){i(e.prototype.__proto__||Object.getPrototypeOf(e.prototype),"prepareContext",this).call(this,t),t.indent=function(t){this.indentLevel=(this.indentLevel||0)+(void 0===t?1:t),this.indentPad=new Array(this.indentLevel+1).join("  "),this.outLine()},t.outLine=function(){this.buffer.push("\n"+(this.indentPad||""))},t.out=function(){for(var t=arguments.length,e=Array(t),r=0;r<t;r++)e[r]=arguments[r];for(var i=0,n=e.length;i<n;i++){var a=e[i].split("\n"),o=a.join("\n"+(this.indentPad||""));this.color&&this.color[0]&&(o=this.color[0](o)),this.buffer.push(o)}},t.pushColor=function(t){this.color=this.color||[],this.color.unshift(t)},t.popColor=function(){this.color=this.color||[],this.color.shift()}}},{key:"typeFormattterErrorFormatter",value:function(t,e){t.pushColor(Tt.error),t.out("[ERROR]"+e),t.popColor()}},{key:"formatValue",value:function(t,e){t.out(JSON.stringify(e,null,2))}},{key:"formatTextDiffString",value:function(t,e){var r=this.parseTextDiff(e);t.indent();for(var i=0,n=r.length;i<n;i++){var a=r[i];t.pushColor(Tt.textDiffLine),t.out(a.location.line+","+a.location.chr+" "),t.popColor();for(var o=a.pieces,s=0,l=o.length;s<l;s++){var p=o[s];t.pushColor(Tt[p.type]),t.out(p.text),t.popColor()}i<n-1&&t.outLine()}t.indent(-1)}},{key:"rootBegin",value:function(t,e,r){t.pushColor(Tt[e]),"node"===e&&(t.out("array"===r?"[":"{"),t.indent())}},{key:"rootEnd",value:function(t,e,r){"node"===e&&(t.indent(-1),t.out("array"===r?"]":"}")),t.popColor()}},{key:"nodeBegin",value:function(t,e,r,i,n){t.pushColor(Tt[i]),t.out(r+": "),"node"===i&&(t.out("array"===n?"[":"{"),t.indent())}},{key:"nodeEnd",value:function(t,e,r,i,n,a){"node"===i&&(t.indent(-1),t.out("array"===n?"]":"}"+(a?"":","))),a||t.outLine(),t.popColor()}},{key:"format_unchanged",value:function(t,e,r){void 0!==r&&this.formatValue(t,r)}},{key:"format_movedestination",value:function(t,e,r){void 0!==r&&this.formatValue(t,r)}},{key:"format_node",value:function(t,e,r){this.formatDeltaChildren(t,e,r)}},{key:"format_added",value:function(t,e){this.formatValue(t,e[0])}},{key:"format_modified",value:function(t,e){t.pushColor(Tt.deleted),this.formatValue(t,e[0]),t.popColor(),t.out(" => "),t.pushColor(Tt.added),this.formatValue(t,e[1]),t.popColor()}},{key:"format_deleted",value:function(t,e){this.formatValue(t,e[0])}},{key:"format_moved",value:function(t,e){t.out("==> "+e[1])}},{key:"format_textdiff",value:function(t,e){this.formatTextDiffString(t,e[0])}}]),e}(K),kt=void 0,It=function(t,e){return kt||(kt=new Pt),kt.format(t,e)};var At=Object.freeze({default:Pt,format:It,log:function(t,e){console.log(It(t,e))}}),Et=Object.freeze({base:tt,html:ot,annotated:ht,jsonpatch:wt,console:At});var Ct=void 0;t.DiffPatcher=W,t.formatters=Et,t.console=At,t.create=function(t){return new W(t)},t.dateReviver=function(t,e){var r=void 0;if("string"==typeof e&&(r=/^(\d{4})-(\d{2})-(\d{2})T(\d{2}):(\d{2}):(\d{2})(?:\.(\d*))?(Z|([+-])(\d{2}):(\d{2}))$/.exec(e)))return new Date(Date.UTC(+r[1],+r[2]-1,+r[3],+r[4],+r[5],+r[6],+(r[7]||0)));return e},t.diff=function(){Ct||(Ct=new W);return Ct.diff.apply(Ct,arguments)},t.patch=function(){Ct||(Ct=new W);return Ct.patch.apply(Ct,arguments)},t.unpatch=function(){Ct||(Ct=new W);return Ct.unpatch.apply(Ct,arguments)},t.reverse=function(){Ct||(Ct=new W);return Ct.reverse.apply(Ct,arguments)},t.clone=function(){Ct||(Ct=new W);return Ct.clone.apply(Ct,arguments)},Object.defineProperty(t,"__esModule",{value:!0})}(e,r(368))},function(t,e){},function(t,e,r){"use strict";Object.defineProperty(e,"__esModule",{value:!0});function i(){this.data={}}e.default=i},function(t,e,r){"use strict";r.r(e),r.d(e,"ThrowPropsPlugin",function(){return i}),r.d(e,"default",function(){return i}),r.d(e,"VelocityTracker",function(){return n});var M=r(0);
/*!
 * VERSION: 0.11.1
 * DATE: 2018-08-27
 * UPDATES AND DOCS AT: http://greensock.com
 *
 * @license Copyright (c) 2008-2018, GreenSock. All rights reserved.
 * ThrowPropsPlugin is a Club GreenSock membership benefit; You must have a valid membership to use
 * this code without violating the terms of use. Visit http://greensock.com/club/ to sign up or get more details.
 * This work is subject to the software agreement that was issued with your membership.
 *
 * @author: Jack Doyle,
 */M.m._gsDefine("plugins.ThrowPropsPlugin",["utils.VelocityTracker","plugins.TweenPlugin","TweenLite","easing.Ease"],function(P){function k(t,e){M.l.call(this,"throwProps"),this._overwriteProps.length=0}function p(t,e,r,i,n){var a,o,s,l,p=e.length,c=0,u=h;if("object"==typeof t){for(;-1<--p;){for(s in a=e[p],o=0,t)o+=(l=a[s]-t[s])*l;o<u&&(c=p,u=o)}if((n||h)<h&&n<Math.sqrt(u))return t}else for(;-1<--p;)(o=(a=e[p])-t)<0&&(o=-o),o<u&&i<=a&&a<=r&&(c=p,u=o);return e[c]}function I(t,e,r,i,n,a){if("auto"===t.end)return t;var o,s,l=t.end;if(r=isNaN(r)?h:r,i=isNaN(i)?-h:i,"object"==typeof e){if(o=e.calculated?e:("function"==typeof l?l(e):p(e,l,r,i,a))||e,!e.calculated){for(s in o)e[s]=o[s];e.calculated=!0}o=o[n]}else o="function"==typeof l?l(e):l instanceof Array?p(e,l,r,i,a):Number(l);return r<o?o=r:o<i&&(o=i),{max:o,min:o,unitFactor:t.unitFactor}}function A(t,e,r){for(var i in e)void 0===t[i]&&i!==r&&(t[i]=e[i]);return t}var x,b,_,w,h=999999999999999,r=M.m._gsDefine.globals,E=!1,S={x:1,y:1,z:2,scale:1,scaleX:1,scaleY:1,rotation:1,rotationZ:1,rotationX:2,rotationY:2,skewX:1,skewY:1,xPercent:1,yPercent:1},C=k.calculateChange=function(t,e,r,i){return null==i&&(i=.05),r*i*t/(e instanceof M.b?e:e?new M.b(e):M.k.defaultEase).getRatio(i)},O=k.calculateDuration=function(t,e,r,i,n){n=n||.05;var a=i instanceof M.b?i:i?new M.b(i):M.k.defaultEase;return Math.abs((e-t)*a.getRatio(n)/r/n)},o=k.calculateTweenDuration=function(t,e,r,i,n,a){if("string"==typeof t&&(t=M.k.selector(t)),!t)return 0;null==r&&(r=10),null==i&&(i=.2),null==n&&(n=1),t.length&&(t=t[0]||t);var o,s,l,p,c,u,h,d,f,m,g,v,y,x=0,b=9999999999,_=e.throwProps||e,w=e.ease instanceof M.b?e.ease:e.ease?new M.b(e.ease):M.k.defaultEase,S=isNaN(_.checkpoint)?.05:Number(_.checkpoint),T=isNaN(_.resistance)?k.defaultResistance:Number(_.resistance);if(_.linkedProps)for(v=_.linkedProps.split(","),g={},y=0;y<v.length;y++)(s=_[o=v[y]])&&(l=0<(p=void 0!==s.velocity&&"number"==typeof s.velocity?Number(s.velocity)||0:(f=f||P.getByTarget(t))&&f.isTrackingProp(o)?f.getVelocity(o):0)*(c=isNaN(s.resistance)?T:Number(s.resistance))?p/c:p/-c,u="function"==typeof t[o]?t[o.indexOf("set")||"function"!=typeof t["get"+o.substr(3)]?o:"get"+o.substr(3)]():t[o]||0,g[o]=u+C(p,w,l,S));for(o in _)"resistance"!==o&&"checkpoint"!==o&&"preventOvershoot"!==o&&"linkedProps"!==o&&"radius"!==o&&("object"!=typeof(s=_[o])&&((f=f||P.getByTarget(t))&&f.isTrackingProp(o)?s="number"==typeof s?{velocity:s}:{velocity:f.getVelocity(o)}:l=0<(p=Number(s)||0)*T?p/T:p/-T),"object"==typeof s&&(l=0<(p=void 0!==s.velocity&&"number"==typeof s.velocity?Number(s.velocity)||0:(f=f||P.getByTarget(t))&&f.isTrackingProp(o)?f.getVelocity(o):0)*(c=isNaN(s.resistance)?T:Number(s.resistance))?p/c:p/-c,h=(u="function"==typeof t[o]?t[o.indexOf("set")||"function"!=typeof t["get"+o.substr(3)]?o:"get"+o.substr(3)]():t[o]||0)+C(p,w,l,S),void 0!==s.end&&(s=I(s,g&&o in g?g:h,s.max,s.min,o,_.radius),(a||E)&&(_[o]=A(s,_[o],"end"))),void 0!==s.max&&h>Number(s.max)+1e-10?(m=s.unitFactor||k.defaultUnitFactors[o]||1,(d=u>s.max&&s.min!==s.max||-15<p*m&&p*m<45?i+.1*(r-i):O(u,s.max,p,w,S))+n<b&&(b=d+n)):void 0!==s.min&&h<Number(s.min)-1e-10&&(m=s.unitFactor||k.defaultUnitFactors[o]||1,(d=u<s.min&&s.min!==s.max||-45<p*m&&p*m<15?i+.1*(r-i):O(u,s.min,p,w,S))+n<b&&(b=d+n)),x<d&&(x=d)),x<l&&(x=l));return b<x&&(x=b),r<x?r:x<i?i:x},t=k.prototype=new M.l("throwProps");return(t.constructor=k).version="0.11.1",k.API=2,k._autoCSS=!0,k.defaultResistance=100,k.defaultUnitFactors={time:1e3,totalTime:1e3},k.track=function(t,e,r){return P.track(t,e,r)},k.untrack=function(t,e){P.untrack(t,e)},k.isTracking=function(t,e){return P.isTracking(t,e)},k.getVelocity=function(t,e){var r=P.getByTarget(t);return r?r.getVelocity(e):NaN},k._cssRegister=function(){var t=r.com.greensock.plugins.CSSPlugin;if(t){var e=t._internals,g=e._parseToProxy,v=e._setPluginRatio,y=e.CSSPropTween;e._registerComplexSpecialProp("throwProps",{parser:function(t,e,r,i,n,a){a=new k;var o,s,l,p,c={},u={},h={},d={},f={},m={};for(s in b={},e)"resistance"!==s&&"preventOvershoot"!==s&&"linkedProps"!==s&&"radius"!==s&&("object"==typeof(o=e[s])?(void 0!==o.velocity&&"number"==typeof o.velocity?c[s]=Number(o.velocity)||0:(p=p||P.getByTarget(t),c[s]=p&&p.isTrackingProp(s)?p.getVelocity(s):0),void 0!==o.end&&(d[s]=o.end),void 0!==o.min&&(u[s]=o.min),void 0!==o.max&&(h[s]=o.max),o.preventOvershoot&&(m[s]=!0),void 0!==o.resistance&&(!0,f[s]=o.resistance)):"number"==typeof o?c[s]=o:(p=p||P.getByTarget(t))&&p.isTrackingProp(s)?c[s]=p.getVelocity(s):c[s]=o||0,S[s]&&i._enableTransforms(2===S[s]));for(s in l=g(t,c,i,n,a),x=l.proxy,c=l.end,x)b[s]={velocity:c[s],min:u[s],max:h[s],end:d[s],resistance:f[s],preventOvershoot:m[s]};return null!=e.resistance&&(b.resistance=e.resistance),null!=e.linkedProps&&(b.linkedProps=e.linkedProps),null!=e.radius&&(b.radius=e.radius),e.preventOvershoot&&(b.preventOvershoot=!0),n=new y(t,"throwProps",0,0,l.pt,2),i._overwriteProps.pop(),n.plugin=a,n.setRatio=v,n.data=l,a._onInitTween(x,b,i._tween),n}})}},k.to=function(t,e,r,i,n){e.throwProps||(e={throwProps:e}),0===n&&(e.throwProps.preventOvershoot=!0),E=!0;var a=new M.k(t,i||1,e);return a.render(0,!0,!0),E=(a.vars.css?(a.duration(o(x,{throwProps:b,ease:e.ease},r,i,n)),a._delay&&!a.vars.immediateRender?a.invalidate():_._onInitTween(x,w,a)):(a.kill(),a=new M.k(t,o(t,e,r,i,n),e)),!1),a},t._onInitTween=function(t,e,r,i){this.target=t,this._props=[],_=this,w=e;var n,a,o,s,l,p,c,u,h,d,f,m,g=r._ease,v=isNaN(e.checkpoint)?.05:Number(e.checkpoint),y=r._duration,x=e.preventOvershoot,b=0;if(e.linkedProps)for(f=e.linkedProps.split(","),d={},m=0;m<f.length;m++)(a=e[n=f[m]])&&(l=void 0!==a.velocity&&"number"==typeof a.velocity?Number(a.velocity)||0:(h=h||P.getByTarget(t))&&h.isTrackingProp(n)?h.getVelocity(n):0,o="function"==typeof t[n]?t[n.indexOf("set")||"function"!=typeof t["get"+n.substr(3)]?n:"get"+n.substr(3)]():t[n]||0,d[n]=o+C(l,g,y,v));for(n in e)if("resistance"!==n&&"checkpoint"!==n&&"preventOvershoot"!==n&&"linkedProps"!==n&&"radius"!==n){if("function"==typeof(a=e[n])&&(a=a(i,t)),"number"==typeof a)l=Number(a)||0;else if("object"!=typeof a||isNaN(a.velocity)){if(!(h=h||P.getByTarget(t))||!h.isTrackingProp(n))throw"ERROR: No velocity was defined in the throwProps tween of "+t+" property: "+n;l=h.getVelocity(n)}else l=Number(a.velocity);p=C(l,g,y,v),u=0,o=(s="function"==typeof t[n])?t[n.indexOf("set")||"function"!=typeof t["get"+n.substr(3)]?n:"get"+n.substr(3)]():t[n],"object"==typeof a&&(c=o+p,void 0!==a.end&&(a=I(a,d&&n in d?d:c,a.max,a.min,n,e.radius),E&&(e[n]=A(a,e[n],"end"))),void 0!==a.max&&Number(a.max)<c?x||a.preventOvershoot?p=a.max-o:u=a.max-o-p:void 0!==a.min&&Number(a.min)>c&&(x||a.preventOvershoot?p=a.min-o:u=a.min-o-p)),this._overwriteProps[b]=n,this._props[b++]={p:n,s:o,c1:p,c2:u,f:s,r:!1}}return!0},t._kill=function(t){for(var e=this._props.length;-1<--e;)null!=t[this._props[e].p]&&this._props.splice(e,1);return M.l.prototype._kill.call(this,t)},t._mod=function(t){for(var e,r=this._props,i=r.length;-1<--i;)"function"==typeof(e=t[r[i].p]||t.throwProps)&&(r[i].m=e)},t.setRatio=function(t){for(var e,r,i=this._props.length;-1<--i;)r=(e=this._props[i]).s+e.c1*t+e.c2*t*t,e.m?r=e.m(r,this.target):1===t&&(r=(1e4*r+(r<0?-.5:.5)|0)/1e4),e.f?this.target[e.p](r):this.target[e.p]=r},M.l.activate([k]),k},!0),M.m._gsDefine("utils.VelocityTracker",["TweenLite"],function(){function o(t,e,r){var i=(t._gsTransform||c)[e];return i||0===i?i:(t.style[e]?i=t.style[e]:(r=r||h(t,null))?i=r[e]||r.getPropertyValue(e)||r.getPropertyValue(e.replace(p,"-$1").toLowerCase()):t.currentStyle&&(i=t.currentStyle[e]),parseFloat(i)||0)}function s(t,e,r){this.p=t,this.f=e,this.v1=this.v2=0,this.t1=this.t2=d.time,this.css=!1,this.type="",this._prev=null,r&&((this._next=r)._prev=this)}function e(){var t,e,r=a,i=d.time;if(.03<=i-n)for(n,n=i;r;){for(e=r._firstVP;e;)((t=e.css?o(r.target,e.p):e.f?r.target[e.p]():r.target[e.p])!==e.v1||.15<i-e.t1)&&(e.v2=e.v1,e.v1=t,e.t2=e.t1,e.t1=i),e=e._next;r=r._next}}function l(t){this._lookup={},this.target=t,this.elem=!(!t.style||!t.nodeType),r||(d.addEventListener("tick",e,null,!1,-100),n=d.time,r=!0),a&&((this._next=a)._prev=this),a=this}var a,r,n,p=/([A-Z])/g,c={},t=M.m.document,u={x:1,y:1,z:2,scale:1,scaleX:1,scaleY:1,rotation:1,rotationZ:1,rotationX:2,rotationY:2,skewX:1,skewY:1,xPercent:1,yPercent:1},h=t.defaultView?t.defaultView.getComputedStyle:function(){},d=M.k.ticker,f=l.getByTarget=function(t){for(var e=a;e;){if(e.target===t)return e;e=e._next}},i=l.prototype;return i.addProp=function(t,e){if(!this._lookup[t]){var r=this.target,i="function"==typeof r[t],n=i?this._altProp(t):t,a=this._firstVP;this._firstVP=this._lookup[t]=this._lookup[n]=a=new s(n!==t&&0===t.indexOf("set")?n:t,i,a),a.css=this.elem&&(void 0!==this.target.style[a.p]||u[a.p]),a.css&&u[a.p]&&!r._gsTransform&&M.k.set(r,{x:"+=0",overwrite:!1}),a.type=e||a.css&&0===t.indexOf("rotation")?"deg":"",a.v1=a.v2=a.css?o(r,a.p):i?r[a.p]():r[a.p]}},i.removeProp=function(t){var e=this._lookup[t];e&&(e._prev?e._prev._next=e._next:e===this._firstVP&&(this._firstVP=e._next),e._next&&(e._next._prev=e._prev),this._lookup[t]=0,e.f&&(this._lookup[this._altProp(t)]=0))},i.isTrackingProp=function(t){return this._lookup[t]instanceof s},i.getVelocity=function(t){var e,r,i=this._lookup[t],n=this.target;if(!i)throw"The velocity of "+t+" is not being tracked.";return e=(i.css?o(n,i.p):i.f?n[i.p]():n[i.p])-i.v2,"rad"!==i.type&&"deg"!==i.type||(e%=r="rad"===i.type?2*Math.PI:360)!==e%(r/2)&&(e=e<0?e+r:e-r),e/(d.time-i.t2)},i._altProp=function(t){var e=t.substr(0,3),r=("get"===e?"set":"set"===e?"get":e)+t.substr(3);return"function"==typeof this.target[r]?r:t},l.getByTarget=function(t){var e=a;for("string"==typeof t&&(t=M.k.selector(t)),t.length&&t!==window&&t[0]&&t[0].style&&!t.nodeType&&(t=t[0]);e;){if(e.target===t)return e;e=e._next}},l.track=function(t,e,r){var i=f(t),n=e.split(","),a=n.length;for(r=(r||"").split(","),i||(i=new l(t));-1<--a;)i.addProp(n[a],r[a]||r[0]);return i},l.untrack=function(t,e){var r=f(t),i=(e||"").split(","),n=i.length;if(r){for(;-1<--n;)r.removeProp(i[n]);r._firstVP&&e||(r._prev?r._prev._next=r._next:r===a&&(a=r._next),r._next&&(r._next._prev=r._prev))}},l.isTracking=function(t,e){var r=f(t);return!!r&&(!(e||!r._firstVP)||r.isTrackingProp(e))},l},!0);var i=M.o.ThrowPropsPlugin,n=M.o.com.greensock.utils.VelocityTracker},function(t,e){t.exports=function(t){if(!t.webpackPolyfill){var e=Object.create(t);e.children||(e.children=[]),Object.defineProperty(e,"loaded",{enumerable:!0,get:function(){return e.l}}),Object.defineProperty(e,"id",{enumerable:!0,get:function(){return e.i}}),Object.defineProperty(e,"exports",{enumerable:!0}),e.webpackPolyfill=1}return e}},function(t,e,r){"use strict";r.r(e),r.d(e,"Draggable",function(){return i}),r.d(e,"default",function(){return i});var me=r(0),ge=r(49);
/*!
 * VERSION: 0.16.5
 * DATE: 2018-08-27
 * UPDATES AND DOCS AT: http://greensock.com
 *
 * Requires TweenLite and CSSPlugin version 1.17.0 or later (TweenMax contains both TweenLite and CSSPlugin). ThrowPropsPlugin is required for momentum-based continuation of movement after the mouse/touch is released (ThrowPropsPlugin is a membership benefit of Club GreenSock - http://greensock.com/club/).
 *
 * @license Copyright (c) 2008-2018, GreenSock. All rights reserved.
 * This work is subject to the terms at http://greensock.com/standard-license or for
 * Club GreenSock members, the software agreement that was issued with your membership.
 *
 * @author: Jack Doyle,
 */
me.m._gsDefine("utils.Draggable",["events.EventDispatcher","TweenLite","plugins.CSSPlugin"],function(){function n(){return!1}function c(t){return $t.createElementNS?$t.createElementNS("http://www.w3.org/1999/xhtml",t):$t.createElement(t)}function bt(t,e){var r,i={};if(e)for(r in t)i[r]=t[r]*e;else for(r in t)i[r]=t[r];return i}function _t(){for(var t=Ut.length;-1<--t;)Ut[t]()}function wt(t){for(var e=Ut.length;-1<--e;)Ut[e]===t&&Ut.splice(e,1);me.n.to(v,0,{overwrite:"all",delay:15,onComplete:v,data:"_draggable"})}function k(){return null!=window.pageYOffset?window.pageYOffset:null!=$t.scrollTop?$t.scrollTop:Vt.scrollTop||$t.body.scrollTop||0}function I(){return null!=window.pageXOffset?window.pageXOffset:null!=$t.scrollLeft?$t.scrollLeft:Vt.scrollLeft||$t.body.scrollLeft||0}function a(t,e){var r="x"===e?"Width":"Height",i="scroll"+r,n="client"+r,a=$t.body;return Math.max(0,ne(t)?Math.max(Vt[i],a[i])-(window["inner"+r]||Vt[n]||a[n]):t[i]-t[n])}function St(t,e){return t=t||window.event,u.pageX=t.clientX+$t.body.scrollLeft+Vt.scrollLeft,u.pageY=t.clientY+$t.body.scrollTop+Vt.scrollTop,e&&(t.returnValue=!1),u}function Tt(t){return t?("string"==typeof t&&(t=me.n.selector(t)),t.length&&t!==window&&t[0]&&t[0].style&&!t.nodeType&&(t=t[0]),t===window||t.nodeType&&t.style?t:null):t}function o(t,e){var r,i,n,a=t.style;if(void 0===a[e]){for(n=["O","Moz","ms","Ms","Webkit"],i=5,r=e.charAt(0).toUpperCase()+e.substr(1);-1<--i&&void 0===a[n[i]+r];);if(i<0)return"";e=(3===i?"ms":n[i])+r}return e}function Pt(t,e,r){var i=t.style;i&&(void 0===i[e]&&(e=o(t,e)),null==r?i.removeProperty?i.removeProperty(e.replace(/([A-Z])/g,"-$1").toLowerCase()):i.removeAttribute(e):void 0!==i[e]&&(i[e]=r))}function A(t,e){return y(t instanceof Element?t:t.host||(t.parentNode||{}).host||t,e)}function kt(t,e,r){var i=t.vars,n=i[r],a=t._listeners[e];"function"==typeof n&&n.apply(i[r+"Scope"]||i.callbackScope||t,i[r+"Params"]||[t.pointerEvent]),a&&t.dispatchEvent(e)}function It(t,e){var r,i,n,a=Tt(t);return a?z(a,e):void 0!==t.left?(n=se(e),{left:t.left-n.x,top:t.top-n.y,width:t.width,height:t.height}):{left:i=t.min||t.minX||t.minRotation||0,top:r=t.min||t.minY||0,width:(t.max||t.maxX||t.maxRotation||0)-i,height:(t.max||t.maxY||0)-r}}function T(){if(!$t.createElementNS)return P=0,void(f=!1);var t,e,r,i,n=c("div"),a=$t.createElementNS("http://www.w3.org/2000/svg","svg"),o=c("div"),s=n.style,l=$t.body||Vt,p="flex"===oe(l,"display",!0);$t.body&&M&&(s.position="absolute",l.appendChild(o),o.appendChild(n),i=n.offsetParent,o.style[M]="rotate(1deg)",O=n.offsetParent===i,o.style.position="absolute",s.height="10px",i=n.offsetTop,o.style.border="5px solid red",C=i!==n.offsetTop,l.removeChild(o)),s=a.style,a.setAttributeNS(null,"width","400px"),a.setAttributeNS(null,"height","400px"),a.setAttributeNS(null,"viewBox","0 0 400 400"),s.display="block",s.boxSizing="border-box",s.border="0px solid red",s.transform="none",n.style.cssText="width:100px;height:100px;overflow:scroll;-ms-overflow-style:none;",l.appendChild(n),n.appendChild(a),e=(r=a.createSVGPoint().matrixTransform(a.getScreenCTM())).y,n.scrollTop=100,r.x=r.y=0,r=r.matrixTransform(a.getScreenCTM()),m=e-r.y<100.1?0:e-r.y-150,n.removeChild(a),l.removeChild(n),l.appendChild(a),p&&(l.style.display="block"),e=(t=a.getScreenCTM()).e,s.border="50px solid red",t=a.getScreenCTM(),f=0===e&&0===t.e&&0===t.f&&1===t.a?(P=1,!0):(P=e!==t.e?1:0,1!==t.a),p&&(l.style.display="flex"),l.removeChild(a)}function At(t){return!!(N&&"function"==typeof t.getBBox&&t.getCTM&&(!t.parentNode||t.parentNode.getBBox&&t.parentNode.getCTM))}function Et(t,e){if(!t||t===window||!t.parentNode)return[1,0,0,1,0,0];for(var r,i,n,a,o,s,l,p,c=se(t,F),u=se(t.parentNode,R),h=H(t,c,u,!1,!e);(t=t.parentNode)&&t.parentNode&&t!==Vt;)c=u,u=se(t.parentNode,c===F?R:F),l=H(t,c,u),r=h[0],i=h[1],n=h[2],a=h[3],o=h[4],s=h[5],h[0]=r*l[0]+i*l[2],h[1]=r*l[1]+i*l[3],h[2]=n*l[0]+a*l[2],h[3]=n*l[1]+a*l[3],h[4]=o*l[0]+s*l[2]+l[4],h[5]=o*l[1]+s*l[3]+l[5];return e&&(r=h[0],i=h[1],n=h[2],a=h[3],o=h[4],s=h[5],p=r*a-i*n,h[0]=a/p,h[1]=-i/p,h[2]=-n/p,h[3]=r/p,h[4]=(n*s-a*o)/p,h[5]=-(r*s-i*o)/p),h}function E(t,e,r){var i=t.x*e[0]+t.y*e[2]+e[4],n=t.x*e[1]+t.y*e[3]+e[5];return t.x=i*r[0]+n*r[2]+r[4],t.y=i*r[1]+n*r[3]+r[5],t}function s(t){return!!(t&&t.length&&t[0]&&(t[0].nodeType&&t[0].style&&!t.nodeType||t[0].length&&t[0][0]))}function Ct(t){Dt=t.touches&&Qt<t.touches.length,ce(t.target,"touchend",G)}function Ot(n,a,t,e,o,r){var i,s,l,p={};if(a)if(1!==o&&a instanceof Array){if(p.end=i=[],l=a.length,"object"==typeof a[0])for(s=0;s<l;s++)i[s]=bt(a[s],o);else for(s=0;s<l;s++)i[s]=a[s]*o;t+=1.1,e-=1.1}else p.end="function"==typeof a?function(t){var e,r,i=a.call(n,t);if(1!==o)if("object"==typeof i){for(r in e={},i)e[r]=i[r]*o;i=e}else i*=o;return i}:a;return!t&&0!==t||(p.max=t),!e&&0!==e||(p.min=e),r&&(p.velocity=0),p}function Mt(t,e){for(var r,i=t.length;-1<--i;)(r=t[i]).ondragstart=r.onselectstart=e?null:n,Pt(r,"userSelect",e?"text":"none")}function r(n,a){n=Tt(n),a=a||{};var o,s,l,t,p,c,u=$t.createElement("div"),h=u.style,e=n.firstChild,d=0,f=0,m=n.scrollTop,g=n.scrollLeft,v=n.scrollWidth,y=n.scrollHeight,x=0,b=0,_=0;w&&!1!==a.force3D?(p="translate3d(",c="px,0px)"):M&&(p="translate(",c="px)"),this.scrollTop=function(t,e){if(!arguments.length)return-this.top();this.top(-t,e)},this.scrollLeft=function(t,e){if(!arguments.length)return-this.left();this.left(-t,e)},this.left=function(t,e){if(!arguments.length)return-(n.scrollLeft+f);var r=n.scrollLeft-g,i=f;if((2<r||r<-2)&&!e)return g=n.scrollLeft,me.n.killTweensOf(this,!0,{left:1,scrollLeft:1}),this.left(-g),void(a.onKill&&a.onKill());(t=-t)<0?(f=t-.5|0,t=0):b<t?(f=t-b|0,t=b):f=0,(f||i)&&(p?this._suspendTransforms||(h[M]=p+-f+"px,"+-d+c):h.left=-f+"px",0<=f+x&&(h.paddingRight=f+x+"px")),n.scrollLeft=0|t,g=n.scrollLeft},this.top=function(t,e){if(!arguments.length)return-(n.scrollTop+d);var r=n.scrollTop-m,i=d;if((2<r||r<-2)&&!e)return m=n.scrollTop,me.n.killTweensOf(this,!0,{top:1,scrollTop:1}),this.top(-m),void(a.onKill&&a.onKill());(t=-t)<0?(d=t-.5|0,t=0):_<t?(d=t-_|0,t=_):d=0,(d||i)&&(p?this._suspendTransforms||(h[M]=p+-f+"px,"+-d+c):h.top=-d+"px"),n.scrollTop=0|t,m=n.scrollTop},this.maxScrollTop=function(){return _},this.maxScrollLeft=function(){return b},this.disable=function(){for(e=u.firstChild;e;)t=e.nextSibling,n.appendChild(e),e=t;n===u.parentNode&&n.removeChild(u)},this.enable=function(){if((e=n.firstChild)!==u){for(;e;)t=e.nextSibling,u.appendChild(e),e=t;n.appendChild(u),this.calibrate()}},this.calibrate=function(t){var e,r,i=n.clientWidth===o;m=n.scrollTop,g=n.scrollLeft,i&&n.clientHeight===s&&u.offsetHeight===l&&v===n.scrollWidth&&y===n.scrollHeight&&!t||((d||f)&&(e=this.left(),r=this.top(),this.left(-n.scrollLeft),this.top(-n.scrollTop)),i&&!t||(h.display="block",h.width="auto",h.paddingRight="0px",(x=Math.max(0,n.scrollWidth-n.clientWidth))&&(x+=oe(n,"paddingLeft")+(Y?oe(n,"paddingRight"):0))),h.display="inline-block",h.position="relative",h.overflow="visible",h.verticalAlign="top",h.width="100%",h.paddingRight=x+"px",Y&&(h.paddingBottom=oe(n,"paddingBottom",!0)),Xt&&(h.zoom="1"),o=n.clientWidth,s=n.clientHeight,v=n.scrollWidth,y=n.scrollHeight,b=n.scrollWidth-o,_=n.scrollHeight-s,l=u.offsetHeight,h.display="block",(e||r)&&(this.left(e),this.top(r)))},this.content=u,this.element=n,this._suspendTransforms=!1,this.enable()}var Dt,Ft,P,f,m,C,O,t,e,i,l,p,Rt={css:{},data:"_draggable"},Nt={css:{},data:"_draggable"},Lt={css:{},data:"_draggable"},Bt={css:{}},jt=me.m._gsDefine.globals,u={},h={style:{},appendChild:n,removeChild:n},$t=me.m.document||{createElement:function(){return h}},Vt=$t.documentElement||{},d=c("div"),Ht=[],zt=180/Math.PI,Gt=999999999999999,Yt=Date.now||function(){return(new Date).getTime()},Xt=!($t.addEventListener||!$t.all),Wt=$t.createElement("div"),Ut=[],qt={},Jt=0,g=/^(?:a|input|textarea|button|select)$/i,Qt=0,Zt=me.m.navigator&&-1!==me.m.navigator.userAgent.toLowerCase().indexOf("android"),Kt=0,te={},ee={},v=function(){Ut.length||me.n.ticker.removeEventListener("tick",_t)},re=function(t,e){ce(t,"scroll",e),ne(t.parentNode)||re(t.parentNode,e)},ie=function(t,e){ue(t,"scroll",e),ne(t.parentNode)||ie(t.parentNode,e)},ne=function(t){return!(t&&t!==Vt&&t!==$t&&t!==$t.body&&t!==window&&t.nodeType&&t.parentNode)},ae=function(t){var e=ne(t),r=a(t,"x"),i=a(t,"y");e?t=ee:ae(t.parentNode),t._gsMaxScrollX=r,t._gsMaxScrollY=i,t._gsScrollX=t.scrollLeft||0,t._gsScrollY=t.scrollTop||0},y=("undefined"!=typeof window?window:$t.defaultView||{getComputedStyle:function(){}}).getComputedStyle,x=/(?:Left|Right|Width)/i,b=/(?:\d|\-|\+|=|#|\.)*/g,_=function(t,e,r,i,n){if("px"===i||!i)return r;if("auto"===i||!r)return 0;var a,o=x.test(e),s=t,l=d.style,p=r<0;return p&&(r=-r),"%"===i&&-1!==e.indexOf("border")?a=r/100*(o?t.clientWidth:t.clientHeight):(l.cssText="border:0 solid red;position:"+oe(t,"position",!0)+";line-height:0;","%"!==i&&s.appendChild?l[o?"borderLeftWidth":"borderTopWidth"]=r+i:(s=t.parentNode||$t.body,l[o?"width":"height"]=r+i),s.appendChild(d),a=parseFloat(d[o?"offsetWidth":"offsetHeight"]),s.removeChild(d),0!==a||n||(a=_(t,e,r,i,!0))),p?-a:a},oe=function(t,e,r){var i,n=(t._gsTransform||{})[e];return n||0===n?n:(t.style&&t.style[e]?n=t.style[e]:(i=A(t))?n=(n=i.getPropertyValue(e.replace(/([A-Z])/g,"-$1").toLowerCase()))||i.length?n:i[e]:t.currentStyle&&(n=t.currentStyle[e]),"auto"!==n||"top"!==e&&"left"!==e||(n=function(t,e){if("absolute"!==oe(t,"position",!0))return 0;var r="left"===e?"Left":"Top",i=oe(t,"margin"+r,!0);return t["offset"+r]-(_(t,e,parseFloat(i),(i+"").replace(b,""))||0)}(t,e)),r?n:parseFloat(n)||0)},w=""!==o(d,"perspective"),S=o(d,"transformOrigin").replace(/^ms/g,"Ms").replace(/([A-Z])/g,"-$1").toLowerCase(),M=o(d,"transform"),D=M.replace(/^ms/g,"Ms").replace(/([A-Z])/g,"-$1").toLowerCase(),F={},R={},N=me.m.SVGElement,L=me.m.navigator&&(/MSIE ([0-9]{1,}[\.0-9]{0,})/.exec(me.m.navigator.userAgent)||/Trident\/.*rv:([0-9]{1,}[\.0-9]{0,})/.exec(me.m.navigator.userAgent))&&parseFloat(RegExp.$1)<11,B=[],j=[],$=function(t){if(!t.getBoundingClientRect||!t.parentNode||!M)return{offsetTop:0,offsetLeft:0,scaleX:1,scaleY:1,offsetParent:Vt};if(!1!==de.cacheSVGData&&t._dCache&&t._dCache.lastUpdate===me.n.ticker.frame)return t._dCache;var e,r,i,n,a,o,s,l,p,c,u,h=t,d=V(t);if(d.lastUpdate=me.n.ticker.frame,t.getBBox&&!d.isSVGRoot){for(h=t.parentNode,e=t.getBBox();h&&"svg"!==(h.nodeName+"").toLowerCase();)h=h.parentNode;return n=$(h),d.offsetTop=e.y*n.scaleY,d.offsetLeft=e.x*n.scaleX,d.scaleX=n.scaleX,d.scaleY=n.scaleY,d.offsetParent=h||Vt,d}for((i=d.offsetParent)===$t.body&&(i=Vt),j.length=B.length=0;h&&("matrix(1, 0, 0, 1, 0, 0)"!==(a=oe(h,M,!0))&&"none"!==a&&"translate3d(0px, 0px, 0px)"!==a&&(j.push(h),B.push(h.style[M]),h.style[M]="none"),h!==i);)h=h.parentNode;for(r=i.getBoundingClientRect(),a=t.getScreenCTM(),s=t.createSVGPoint().matrixTransform(a),d.scaleX=Math.sqrt(a.a*a.a+a.b*a.b),d.scaleY=Math.sqrt(a.d*a.d+a.c*a.c),void 0===P&&T(),d.borderBox&&!f&&t.getAttribute("width")&&(n=A(t)||{},l=parseFloat(n.borderLeftWidth)+parseFloat(n.borderRightWidth)||0,p=parseFloat(n.borderTopWidth)+parseFloat(n.borderBottomWidth)||0,c=parseFloat(n.width)||0,u=parseFloat(n.height)||0,d.scaleX*=(c-l)/c,d.scaleY*=(u-p)/u),m?(e=t.getBoundingClientRect(),d.offsetLeft=e.left-r.left,d.offsetTop=e.top-r.top):(d.offsetLeft=s.x-r.left,d.offsetTop=s.y-r.top),d.offsetParent=i,o=j.length;-1<--o;)j[o].style[M]=B[o];return d},se=function(t,e){if(e=e||{},!t||t===Vt||!t.parentNode||t===window)return{x:0,y:0};var r=A(t),i=S&&r?r.getPropertyValue(S):"50% 50%",n=i.split(" "),a=-1!==i.indexOf("left")?"0%":-1!==i.indexOf("right")?"100%":n[0],o=-1!==i.indexOf("top")?"0%":-1!==i.indexOf("bottom")?"100%":n[1];return"center"!==o&&null!=o||(o="50%"),"center"!==a&&!isNaN(parseFloat(a))||(a="50%"),t.getBBox&&At(t)?(t._gsTransform||(me.n.set(t,{x:"+=0",overwrite:!1}),void 0===t._gsTransform.xOrigin&&console.log("Draggable requires at least GSAP 1.17.0")),i=t.getBBox(),e.x=t._gsTransform.xOrigin-i.x,e.y=t._gsTransform.yOrigin-i.y):(t.getBBox&&-1!==(a+o).indexOf("%")&&(t={offsetWidth:(t=t.getBBox()).width,offsetHeight:t.height}),e.x=-1!==a.indexOf("%")?t.offsetWidth*parseFloat(a)/100:parseFloat(a),e.y=-1!==o.indexOf("%")?t.offsetHeight*parseFloat(o)/100:parseFloat(o)),e},V=function(t){if(!1!==de.cacheSVGData&&t._dCache&&t._dCache.lastUpdate===me.n.ticker.frame)return t._dCache;var e,r=t._dCache=t._dCache||{},i=A(t),n=t.getBBox&&At(t),a="svg"===(t.nodeName+"").toLowerCase();if(r.isSVG=n,r.isSVGRoot=a,r.borderBox="border-box"===i.boxSizing,r.computedStyle=i,a)(e=t.parentNode||Vt).insertBefore(d,t),r.offsetParent=d.offsetParent||Vt,e.removeChild(d);else if(n){for(e=t.parentNode;e&&"svg"!==(e.nodeName+"").toLowerCase();)e=e.parentNode;r.offsetParent=e}else r.offsetParent=t.offsetParent;return r},H=function(t,e,r,i,n){if(t===window||!t||!t.style||!t.parentNode)return[1,0,0,1,0,0];var a,o,s,l,p,c,u,h,d,f,m,g,v,y,x=t._dCache||V(t),b=t.parentNode,_=b._dCache||V(b),w=x.computedStyle,S=x.isSVG?_.offsetParent:b.offsetParent;return a=x.isSVG&&-1!==(t.style[M]+"").indexOf("matrix")?t.style[M]:w?w.getPropertyValue(D):t.currentStyle?t.currentStyle[M]:"1,0,0,1,0,0",t.getBBox&&-1!==(t.getAttribute("transform")+"").indexOf("matrix")&&(a=t.getAttribute("transform")),6<(a=(a+"").match(/(?:\-|\.|\b)(\d|\.|e\-)+/g)||[1,0,0,1,0,0]).length&&(a=[a[0],a[1],a[4],a[5],a[12],a[13]]),i?a[4]=a[5]=0:x.isSVG&&(p=t._gsTransform)&&(p.xOrigin||p.yOrigin)&&(a[0]=parseFloat(a[0]),a[1]=parseFloat(a[1]),a[2]=parseFloat(a[2]),a[3]=parseFloat(a[3]),a[4]=parseFloat(a[4])-(p.xOrigin-(p.xOrigin*a[0]+p.yOrigin*a[2])),a[5]=parseFloat(a[5])-(p.yOrigin-(p.xOrigin*a[1]+p.yOrigin*a[3]))),e&&(void 0===P&&T(),s=x.isSVG||x.isSVGRoot?$(t):t,x.isSVG?(l=t.getBBox(),f=_.isSVGRoot?{x:0,y:0}:b.getBBox(),s={offsetLeft:l.x-f.x,offsetTop:l.y-f.y,offsetParent:x.offsetParent}):x.isSVGRoot?(m=parseInt(w.borderTopWidth,10)||0,g=parseInt(w.borderLeftWidth,10)||0,v=(a[0]-P)*g+a[2]*m,y=a[1]*g+(a[3]-P)*m,c=e.x,u=e.y,h=c-(c*a[0]+u*a[2]),d=u-(c*a[1]+u*a[3]),a[4]=parseFloat(a[4])+h,a[5]=parseFloat(a[5])+d,e.x-=h,e.y-=d,c=s.scaleX,u=s.scaleY,n||(e.x*=c,e.y*=u),a[0]*=c,a[1]*=u,a[2]*=c,a[3]*=u,L||(e.x+=v,e.y+=y),S===$t.body&&s.offsetParent===Vt&&(S=Vt)):!C&&t.offsetParent&&(e.x+=parseInt(oe(t.offsetParent,"borderLeftWidth"),10)||0,e.y+=parseInt(oe(t.offsetParent,"borderTopWidth"),10)||0),o=b===Vt||b===$t.body,a[4]=Number(a[4])+e.x+(s.offsetLeft||0)-r.x-(o?0:b.scrollLeft||0),a[5]=Number(a[5])+e.y+(s.offsetTop||0)-r.y-(o?0:b.scrollTop||0),b&&"fixed"===oe(t,"position",w)&&(a[4]+=I(),a[5]+=k()),!b||b===Vt||S!==s.offsetParent||_.isSVG||O&&"100100"!==H(b).join("")||(s=_.isSVGRoot?$(b):b,a[4]-=s.offsetLeft||0,a[5]-=s.offsetTop||0,C||!_.offsetParent||x.isSVG||x.isSVGRoot||(a[4]-=parseInt(oe(_.offsetParent,"borderLeftWidth"),10)||0,a[5]-=parseInt(oe(_.offsetParent,"borderTopWidth"),10)||0))),a},z=function(t,e,r){if(!(t=Tt(t)))return null;e=Tt(e);var i,n,a,o,s,l,p,c,u,h,d,f,m,g,v,y,x,b,_,w,S,T,P=t.getBBox&&At(t);if(t===window)o=k(),a=(n=I())+(Vt.clientWidth||t.innerWidth||$t.body.clientWidth||0),s=o+((t.innerHeight||0)-20<Vt.clientHeight?Vt.clientHeight:t.innerHeight||$t.body.clientHeight||0);else{if(void 0===e||e===window)return t.getBoundingClientRect();n=-(i=se(t)).x,o=-i.y,g=P?(m=(f=t.getBBox()).width,f.height):"svg"!==(t.nodeName+"").toLowerCase()&&t.offsetWidth?(m=t.offsetWidth,t.offsetHeight):(S=A(t),m=parseFloat(S.width),parseFloat(S.height)),a=n+m,s=o+g,"svg"!==t.nodeName.toLowerCase()||Xt||(T=(v=$(t)).computedStyle||{},b=(t.getAttribute("viewBox")||"0 0").split(" "),_=parseFloat(b[0]),w=parseFloat(b[1]),y=parseFloat(T.borderLeftWidth)||0,x=parseFloat(T.borderTopWidth)||0,a-=m-(m-y)/v.scaleX-_,s-=g-(g-x)/v.scaleY-w,n-=y/v.scaleX-_,o-=x/v.scaleY-w,S&&(a+=(parseFloat(T.borderRightWidth)+y)/v.scaleX,s+=(x+parseFloat(T.borderBottomWidth))/v.scaleY))}return t===e?{left:n,top:o,width:a-n,height:s-o}:(l=Et(t),p=Et(e,!0),c=E({x:n,y:o},l,p),u=E({x:a,y:o},l,p),h=E({x:a,y:s},l,p),d=E({x:n,y:s},l,p),n=Math.min(c.x,u.x,h.x,d.x),o=Math.min(c.y,u.y,h.y,d.y),te.x=te.y=0,r&&se(e,te),{left:n+te.x,top:o+te.y,width:Math.max(c.x,u.x,h.x,d.x)-n,height:Math.max(c.y,u.y,h.y,d.y)-o})},le="undefined"!=typeof window&&"ontouchstart"in Vt&&"orientation"in window,pe=function(t){for(var e=t.split(","),r=t.split(","),i={},n=4;-1<--n;)i[e[n]]=r[n],i[r[n]]=e[n];return i}("touchstart,touchmove,touchend,touchcancel"),ce=function(t,e,r,i){if(t.addEventListener){var n=pe[e];i=i||{passive:!1},t.addEventListener(n||e,r,i),n&&e!==n&&t.addEventListener(e,r,i)}else t.attachEvent&&t.attachEvent("on"+e,r)},ue=function(t,e,r){if(t.removeEventListener){var i=pe[e];t.removeEventListener(i||e,r),i&&e!==i&&t.removeEventListener(e,r)}else t.detachEvent&&t.detachEvent("on"+e,r)},G=function(t){Dt=t.touches&&Qt<t.touches.length,ue(t.target,"touchend",G)},he=function(t){var e;return!(!t||!t.getAttribute||"BODY"===t.nodeName)&&(!("true"!==(e=t.getAttribute("data-clickable"))&&("false"===e||!t.onclick&&!g.test(t.nodeName+"")&&"true"!==t.getAttribute("contentEditable")))||he(t.parentNode))},Y=(e=$t.createElement("div"),i=$t.createElement("div"),l=i.style,p=$t.body||d,l.display="inline-block",l.position="relative",e.style.cssText=i.innerHTML="width:90px; height:40px; padding:10px; overflow:auto; visibility: hidden",e.appendChild(i),p.appendChild(e),t=i.offsetHeight+18>e.scrollHeight,p.removeChild(e),t),de=function(d,u){me.c.call(this,d),d=Tt(d),Ft||(Ft=jt.com.greensock.plugins.ThrowPropsPlugin),this.vars=u=bt(u||{}),this.target=d,this.x=this.y=this.rotation=0,this.dragResistance=parseFloat(u.dragResistance)||0,this.edgeResistance=isNaN(u.edgeResistance)?1:parseFloat(u.edgeResistance)||0,this.lockAxis=u.lockAxis,this.autoScroll=u.autoScroll||0,this.lockedAxis=null,this.allowEventDefault=!!u.allowEventDefault;function n(t){if(!(nt.isPressed&&t.which<2))return t.preventDefault(),t.stopPropagation(),!1;nt.endDrag()}function h(t){if(nt.autoScroll&&nt.isDragging&&(st||L)){var e,r,i,n,a,o,s,l,p=d,c=15*nt.autoScroll;for(st=!1,ee.scrollTop=null!=window.pageYOffset?window.pageYOffset:null!=Vt.scrollTop?Vt.scrollTop:$t.body.scrollTop,ee.scrollLeft=null!=window.pageXOffset?window.pageXOffset:null!=Vt.scrollLeft?Vt.scrollLeft:$t.body.scrollLeft,n=nt.pointerX-ee.scrollLeft,a=nt.pointerY-ee.scrollTop;p&&!r;)e=(r=ne(p.parentNode))?ee:p.parentNode,i=r?{bottom:Math.max(Vt.clientHeight,window.innerHeight||0),right:Math.max(Vt.clientWidth,window.innerWidth||0),left:0,top:0}:e.getBoundingClientRect(),o=s=0,rt&&((l=e._gsMaxScrollY-e.scrollTop)<0?s=l:a>i.bottom-ct&&l?(st=!0,s=Math.min(l,c*(1-Math.max(0,i.bottom-a)/ct)|0)):a<i.top+lt&&e.scrollTop&&(st=!0,s=-Math.min(e.scrollTop,c*(1-Math.max(0,a-i.top)/lt)|0)),s&&(e.scrollTop+=s)),et&&((l=e._gsMaxScrollX-e.scrollLeft)<0?o=l:n>i.right-pt&&l?(st=!0,o=Math.min(l,c*(1-Math.max(0,i.right-n)/pt)|0)):n<i.left+ut&&e.scrollLeft&&(st=!0,o=-Math.min(e.scrollLeft,c*(1-Math.max(0,n-i.left)/ut)|0)),o&&(e.scrollLeft+=o)),r&&(o||s)&&(window.scrollTo(e.scrollLeft,e.scrollTop),yt(nt.pointerX+o,nt.pointerY+s)),p=e}if(L){var u=nt.x,h=nt.y;u<1e-6&&-1e-6<u&&(u=0),h<1e-6&&-1e-6<h&&(h=0),Z?(nt.deltaX=u-Y.data.rotation,Y.data.rotation=nt.rotation=u,Y.setRatio(1)):_?(rt&&(nt.deltaY=h-_.top(),_.top(h)),et&&(nt.deltaX=u-_.left(),_.left(u))):Q?(rt&&(nt.deltaY=h-Y.data.y,Y.data.y=h),et&&(nt.deltaX=u-Y.data.x,Y.data.x=u),Y.setRatio(1)):(rt&&(nt.deltaY=h-parseFloat(d.style.top||0),d.style.top=h+"px"),et&&(nt.deltaY=u-parseFloat(d.style.left||0),d.style.left=u+"px")),!I||t||U||(U=!0,kt(nt,"drag","onDrag"),U=!1)}L=!1}function s(t,e){var r,i=nt.x,n=nt.y;d._gsTransform||!Q&&!Z||me.n.set(d,{x:"+=0",overwrite:!1,data:"_draggable"}),Q?(nt.y=d._gsTransform.y,nt.x=d._gsTransform.x):Z?nt.x=nt.rotation=d._gsTransform.rotation:_?(nt.y=_.top(),nt.x=_.left()):(nt.y=parseInt(d.style.top,10)||0,nt.x=parseInt(d.style.left,10)||0),(B||j||$)&&!e&&(nt.isDragging||nt.isThrowing)&&($&&(te.x=nt.x,te.y=nt.y,(r=$(te)).x!==nt.x&&(nt.x=r.x,L=!0),r.y!==nt.y&&(nt.y=r.y,L=!0)),B&&(r=B(nt.x))!==nt.x&&(nt.x=r,Z&&(nt.rotation=r),L=!0),j&&((r=j(nt.y))!==nt.y&&(nt.y=r),L=!0)),L&&h(!0),t||(nt.deltaX=nt.x-i,nt.deltaY=nt.y-n,kt(nt,"throwupdate","onThrowUpdate"))}function l(){var t,e,r,i;k=!1,_?(_.calibrate(),nt.minX=E=-_.maxScrollLeft(),nt.minY=O=-_.maxScrollTop(),nt.maxX=A=nt.maxY=C=0,k=!0):u.bounds&&(t=It(u.bounds,d.parentNode),Z?(nt.minX=E=t.left,nt.maxX=A=t.left+t.width,nt.minY=O=nt.maxY=C=0):void 0!==u.bounds.maxX||void 0!==u.bounds.maxY?(t=u.bounds,nt.minX=E=t.minX,nt.minY=O=t.minY,nt.maxX=A=t.maxX,nt.maxY=C=t.maxY):(e=It(d,d.parentNode),nt.minX=E=oe(d,K)+t.left-e.left,nt.minY=O=oe(d,tt)+t.top-e.top,nt.maxX=A=E+(t.width-e.width),nt.maxY=C=O+(t.height-e.height)),A<E&&(nt.minX=A,nt.maxX=A=E,E=nt.minX),C<O&&(nt.minY=C,nt.maxY=C=O,O=nt.minY),Z&&(nt.minRotation=E,nt.maxRotation=A),k=!0),u.liveSnap&&(i=(r=!0===u.liveSnap?u.snap||{}:u.liveSnap)instanceof Array||"function"==typeof r,Z?(B=mt(i?r:r.rotation,E,A,1),j=null):r.points?$=gt(i?r:r.points,E,A,O,C,r.radius,_?-1:1):(et&&(B=mt(i?r:r.x||r.left||r.scrollLeft,E,A,_?-1:1)),rt&&(j=mt(i?r:r.y||r.top||r.scrollTop,O,C,_?-1:1))))}function o(){nt.isThrowing=!1,kt(nt,"throwcomplete","onThrowComplete")}function p(){nt.isThrowing=!1}function f(t,e){var r,i,n,a;t&&Ft?(!0===t&&(i=(r=u.snap||u.liveSnap||{})instanceof Array||"function"==typeof r,t={resistance:(u.throwResistance||u.resistance||1e3)/(Z?10:1)},Z?t.rotation=Ot(nt,i?r:r.rotation,A,E,1,e):(et&&(t[K]=Ot(nt,i?r:r.points||r.x||r.left||r.scrollLeft,A,E,_?-1:1,e||"x"===nt.lockedAxis)),rt&&(t[tt]=Ot(nt,i?r:r.points||r.y||r.top||r.scrollTop,C,O,_?-1:1,e||"y"===nt.lockedAxis)),(r.points||r instanceof Array&&"object"==typeof r[0])&&(t.linkedProps=K+","+tt,t.radius=r.radius))),nt.isThrowing=!0,a=isNaN(u.overshootTolerance)?1===u.edgeResistance?0:1-nt.edgeResistance+.2:u.overshootTolerance,nt.tween=n=Ft.to(_||d,{throwProps:t,data:"_draggable",ease:u.ease||jt.Power3.easeOut,onComplete:o,onOverwrite:p,onUpdate:u.fastMode?kt:s,onUpdateParams:u.fastMode?[nt,"onthrowupdate","onThrowUpdate"]:r&&r.radius?[!1,!0]:Ht},Math.max(u.minDuration||0,u.maxDuration||0)||2,isNaN(u.minDuration)?0===a||"object"==typeof t&&1e3<t.resistance?0:.5:u.minDuration,a),u.fastMode||(_&&(_._suspendTransforms=!0),n.render(n.duration(),!0,!0),s(!0,!0),nt.endX=nt.x,nt.endY=nt.y,Z&&(nt.endRotation=nt.x),n.play(0),s(!0,!0),_&&(_._suspendTransforms=!1))):k&&nt.applyBounds()}function a(t){var e,r,i,n,a,o,s,l,p,c=z||[1,0,0,1,0,0];z=Et(d.parentNode,!0),t&&nt.isPressed&&c.join(",")!==z.join(",")&&(e=c[0],r=c[1],i=c[2],n=c[3],a=c[4],o=c[5],p=w*(-r/(s=e*n-r*i))+S*(e/s)+-(e*o-r*a)/s,S=(l=w*(n/s)+S*(-i/s)+(i*o-n*a)/s)*z[1]+p*z[3]+z[5],w=l*z[0]+p*z[2]+z[4]),z[1]||z[2]||1!=z[0]||1!=z[3]||0!=z[4]||0!=z[5]||(z=null)}function c(){var t=1-nt.edgeResistance;a(!1),z&&(w=nt.pointerX*z[0]+nt.pointerY*z[2]+z[4],S=nt.pointerX*z[1]+nt.pointerY*z[3]+z[5]),L&&(yt(nt.pointerX,nt.pointerY),h(!0)),_?(l(),P=_.top(),T=_.left()):(ft()?(s(!0,!0),l()):nt.applyBounds(),Z?(N=nt.rotationOrigin=function(t,e,r,i){t=Tt(t);var n=Et(t,!1),a=e.x,o=e.y;return r&&(se(t,e),a-=e.x,o-=e.y),(i=!0===i?e:i||{}).x=a*n[0]+o*n[2]+n[4],i.y=a*n[1]+o*n[3]+n[5],i}(d,{x:0,y:0}),s(!0,!0),T=nt.x,P=nt.y=Math.atan2(N.y-nt.pointerY,nt.pointerX-N.x)*zt):(d.parentNode&&d.parentNode.scrollTop,d.parentNode&&d.parentNode.scrollLeft,P=oe(d,tt),T=oe(d,K))),k&&t&&(A<T?T=A+(T-A)/t:T<E&&(T=E-(E-T)/t),Z||(C<P?P=C+(P-C)/t:P<O&&(P=O-(O-P)/t))),nt.startX=T,nt.startY=P}function m(){!Wt.parentNode||ft()||nt.isDragging||Wt.parentNode.removeChild(Wt)}function g(t,e){var r;if(b&&!nt.isPressed&&t&&("mousedown"!==t.type&&"pointerdown"!==t.type||e||!(Yt()-dt<30)||!pe[nt.pointerEvent.type])){if(G=ft(),nt.pointerEvent=t,pe[t.type]?(H=-1!==t.type.indexOf("touch")?t.currentTarget||t.target:$t,ce(H,"touchend",xt),ce(H,"touchmove",vt),ce(H,"touchcancel",xt),ce($t,"touchstart",Ct)):(H=null,ce($t,"mousemove",vt)),W=null,ce($t,"mouseup",xt),t&&t.target&&ce(t.target,"mouseup",xt),V=ht.call(nt,t.target)&&!u.dragClickables&&!e)return ce(t.target,"change",xt),kt(nt,"pressInit","onPressInit"),kt(nt,"press","onPress"),void Mt(at,!0);if(X=!(!H||et==rt||!1===nt.vars.allowNativeTouchScrolling||nt.vars.allowContextMenu&&t&&(t.ctrlKey||2<t.which))&&(et?"y":"x"),Xt?t=St(t,!0):X||nt.allowEventDefault||(t.preventDefault(),t.preventManipulation&&t.preventManipulation()),t.changedTouches?(t=F=t.changedTouches[0],R=t.identifier):t.pointerId?R=t.pointerId:F=R=null,Qt++,function(t){Ut.push(t),1===Ut.length&&me.n.ticker.addEventListener("tick",_t,this,!1,1)}(h),S=nt.pointerY=t.pageY,w=nt.pointerX=t.pageX,kt(nt,"pressInit","onPressInit"),(X||nt.autoScroll)&&ae(d.parentNode),!d.parentNode||!nt.autoScroll||_||Z||!d.parentNode._gsMaxScrollX||Wt.parentNode||d.getBBox||(Wt.style.width=d.parentNode.scrollWidth+"px",d.parentNode.appendChild(Wt)),c(),nt.tween&&nt.tween.kill(),nt.isThrowing=!1,me.n.killTweensOf(_||d,!0,i),_&&me.n.killTweensOf(d,!0,{scrollTo:1}),nt.tween=nt.lockedAxis=null,!u.zIndexBoost&&(Z||_||!1===u.zIndexBoost)||(d.style.zIndex=de.zIndex++),nt.isPressed=!0,I=!(!u.onDrag&&!nt._listeners.drag),!Z)for(r=at.length;-1<--r;)Pt(at[r],"cursor",u.cursor||"move");kt(nt,"press","onPress")}}function v(t){if(t&&nt.isDragging&&!_){var e=t.target||t.srcElement||d.parentNode,r=e.scrollLeft-e._gsScrollX,i=e.scrollTop-e._gsScrollY;(r||i)&&(z?(w-=r*z[0]+i*z[2],S-=i*z[3]+r*z[1]):(w-=r,S-=i),e._gsScrollX+=r,e._gsScrollY+=i,yt(nt.pointerX,nt.pointerY))}}function y(t){var e=Yt(),r=e-dt<40,i=e-ot<40,n=r&&q===dt,a=!!t.preventDefault,o=nt.pointerEvent&&nt.pointerEvent.defaultPrevented,s=r&&J===dt,l=t.isTrusted||null==t.isTrusted&&r&&n;if(a&&(n||i&&!1!==nt.vars.suppressClickOnDrag)&&t.stopImmediatePropagation(),!(!r||nt.pointerEvent&&nt.pointerEvent.defaultPrevented||n&&l===s))return l&&n&&(J=dt),void(q=dt);(nt.isPressed||i||r)&&(a?l&&t.detail&&r&&!o||(t.preventDefault(),t.preventManipulation&&t.preventManipulation()):t.returnValue=!1)}function x(t){return z?{x:t.x*z[0]+t.y*z[2]+z[4],y:t.x*z[1]+t.y*z[3]+z[5]}:{x:t.x,y:t.y}}var b,_,w,S,T,P,k,I,A,E,C,O,M,D,F,R,N,L,t,B,j,$,V,H,z,G,Y,X,W,U,q,J,e=(u.type||(Xt?"top,left":"x,y")).toLowerCase(),Q=-1!==e.indexOf("x")||-1!==e.indexOf("y"),Z=-1!==e.indexOf("rotation"),K=Z?"rotation":Q?"x":"left",tt=Q?"y":"top",et=-1!==e.indexOf("x")||-1!==e.indexOf("left")||"scroll"===e,rt=-1!==e.indexOf("y")||-1!==e.indexOf("top")||"scroll"===e,it=u.minimumMovement||2,nt=this,at=function(t){if("string"==typeof t&&(t=me.n.selector(t)),!t||t.nodeType)return[t];var e,r=[],i=t.length;for(e=0;e!==i;r.push(t[e++]));return r}(u.trigger||u.handle||d),i={},ot=0,st=!1,lt=u.autoScrollMarginTop||40,pt=u.autoScrollMarginRight||40,ct=u.autoScrollMarginBottom||40,ut=u.autoScrollMarginLeft||40,ht=u.clickableTest||he,dt=0,ft=function(){return nt.tween&&nt.tween.isActive()},mt=function(o,s,l,r){return"function"==typeof o?function(t){var e=nt.isPressed?1-nt.edgeResistance:1;return o.call(nt,l<t?l+(t-l)*e:t<s?s+(t-s)*e:t)*r}:o instanceof Array?function(t){for(var e,r,i=o.length,n=0,a=Gt;-1<--i;)(r=(e=o[i])-t)<0&&(r=-r),r<a&&s<=e&&e<=l&&(n=i,a=r);return o[n]}:isNaN(o)?function(t){return t}:function(){return o*r}},gt=function(l,s,p,c,u,h,d){return h=h&&h<Gt?h*h:Gt,"function"==typeof l?function(t){var e,r,i,n=nt.isPressed?1-nt.edgeResistance:1,a=t.x,o=t.y;return t.x=a=p<a?p+(a-p)*n:a<s?s+(a-s)*n:a,t.y=o=u<o?u+(o-u)*n:o<c?c+(o-c)*n:o,(e=l.call(nt,t))!==t&&(t.x=e.x,t.y=e.y),1!==d&&(t.x*=d,t.y*=d),h<Gt&&(r=t.x-a,i=t.y-o,h<r*r+i*i&&(t.x=a,t.y=o)),t}:l instanceof Array?function(t){for(var e,r,i,n,a=l.length,o=0,s=Gt;-1<--a;)(n=(e=(i=l[a]).x-t.x)*e+(r=i.y-t.y)*r)<s&&(o=a,s=n);return s<=h?l[o]:t}:function(t){return t}},vt=function(t){var e,r,i,n,a,o,s=t;if(b&&!Dt&&nt.isPressed&&t){if(e=(nt.pointerEvent=t).changedTouches){if((t=e[0])!==F&&t.identifier!==R){for(n=e.length;-1<--n&&(t=e[n]).identifier!==R;);if(n<0)return}}else if(t.pointerId&&R&&t.pointerId!==R)return;if(Xt)t=St(t,!0);else{if(H&&X&&!W&&(r=t.pageX,i=t.pageY,z&&(n=r*z[0]+i*z[2]+z[4],i=r*z[1]+i*z[3]+z[5],r=n),((a=Math.abs(r-w))!==(o=Math.abs(i-S))&&(it<a||it<o)||Zt&&X===W)&&(W=o<a&&et?"x":"y",!1!==nt.vars.lockAxisOnTouchScroll&&(nt.lockedAxis="x"===W?"y":"x","function"==typeof nt.vars.onLockAxis&&nt.vars.onLockAxis.call(nt,s)),Zt&&X===W)))return void xt(s);nt.allowEventDefault||X&&(!W||X===W)||!1===s.cancelable||(s.preventDefault(),s.preventManipulation&&s.preventManipulation())}nt.autoScroll&&(st=!0),yt(t.pageX,t.pageY)}},yt=function(t,e){var r,i,n,a,o,s,l=1-nt.dragResistance,p=1-nt.edgeResistance;nt.pointerX=t,nt.pointerY=e,Z?(a=Math.atan2(N.y-e,t-N.x)*zt,180<(o=nt.y-a)?(P-=360,nt.y=a):o<-180&&(P+=360,nt.y=a),n=nt.x!==T||Math.abs(P-a)>it?(nt.y=a,T+(P-a)*l):T):(z&&(s=t*z[0]+e*z[2]+z[4],e=t*z[1]+e*z[3]+z[5],t=s),(i=e-S)<it&&-it<i&&(i=0),(r=t-w)<it&&-it<r&&(r=0),(nt.lockAxis||nt.lockedAxis)&&(r||i)&&((s=nt.lockedAxis)||(nt.lockedAxis=s=et&&Math.abs(r)>Math.abs(i)?"y":rt?"x":null,s&&"function"==typeof nt.vars.onLockAxis&&nt.vars.onLockAxis.call(nt,nt.pointerEvent)),"y"===s?i=0:"x"===s&&(r=0)),n=T+r*l,a=P+i*l),(B||j||$)&&(nt.x!==n||nt.y!==a&&!Z)?($&&(te.x=n,te.y=a,n=(s=$(te)).x,a=s.y),B&&(n=B(n)),j&&(a=j(a))):k&&(A<n?n=A+(n-A)*p:n<E&&(n=E+(n-E)*p),Z||(C<a?a=C+(a-C)*p:a<O&&(a=O+(a-O)*p))),Z||z||(n=Math.round(n),a=Math.round(a)),nt.x===n&&(nt.y===a||Z)||(Z?(nt.endRotation=nt.x=nt.endX=n,L=!0):(rt&&(nt.y=nt.endY=a,L=!0),et&&(nt.x=nt.endX=n,L=!0)),!nt.isDragging&&nt.isPressed&&(nt.isDragging=!0,kt(nt,"dragstart","onDragStart")))},xt=function(t,e){if(b&&nt.isPressed&&(!t||null==R||e||!(t.pointerId&&t.pointerId!==R||t.changedTouches&&!function(t,e){for(var r=t.length;-1<--r;)if(t[r].identifier===e)return!0;return!1}(t.changedTouches,R)))){nt.isPressed=!1;var r,i,n,a,o,s=t,l=nt.isDragging,p=nt.vars.allowContextMenu&&t&&(t.ctrlKey||2<t.which),c=me.n.delayedCall(.001,m);if(H?(ue(H,"touchend",xt),ue(H,"touchmove",vt),ue(H,"touchcancel",xt),ue($t,"touchstart",Ct)):ue($t,"mousemove",vt),ue($t,"mouseup",xt),t&&t.target&&ue(t.target,"mouseup",xt),L=!1,V&&!p)return t&&(ue(t.target,"change",xt),nt.pointerEvent=s),Mt(at,!1),kt(nt,"release","onRelease"),kt(nt,"click","onClick"),void(V=!1);if(wt(h),!Z)for(i=at.length;-1<--i;)Pt(at[i],"cursor",u.cursor||"move");if(l&&(ot=Kt=Yt(),nt.isDragging=!1),Qt--,t){if(Xt&&(t=St(t,!1)),(r=t.changedTouches)&&(t=r[0])!==F&&t.identifier!==R){for(i=r.length;-1<--i&&(t=r[i]).identifier!==R;);if(i<0)return}nt.pointerEvent=s,nt.pointerX=t.pageX,nt.pointerY=t.pageY}return p&&s?(s.preventDefault(),s.preventManipulation&&s.preventManipulation(),kt(nt,"release","onRelease")):s&&!l?(G&&(u.snap||u.bounds)&&f(u.throwProps),kt(nt,"release","onRelease"),Zt&&"touchmove"===s.type||-1!==s.type.indexOf("cancel")||(kt(nt,"click","onClick"),Yt()-dt<300&&kt(nt,"doubleclick","onDoubleClick"),a=s.target||s.srcElement||d,dt=Yt(),o=function(){dt!==q&&nt.enabled()&&!nt.isPressed&&(a.click?a.click():$t.createEvent&&((n=$t.createEvent("MouseEvents")).initMouseEvent("click",!0,!0,window,1,nt.pointerEvent.screenX,nt.pointerEvent.screenY,nt.pointerX,nt.pointerY,!1,!1,!1,!1,0,null),a.dispatchEvent(n)))},Zt||s.defaultPrevented||me.n.delayedCall(1e-5,o))):(f(u.throwProps),Xt||nt.allowEventDefault||!s||!u.dragClickables&&ht.call(nt,s.target)||!l||X&&(!W||X!==W)||!1===s.cancelable||(s.preventDefault(),s.preventManipulation&&s.preventManipulation()),kt(nt,"release","onRelease")),ft()&&c.duration(nt.tween.duration()),l&&kt(nt,"dragend","onDragEnd"),!0}};(t=de.get(this.target))&&t.kill(),this.startDrag=function(t,e){var r,i,n,a;g(t||nt.pointerEvent,!0),e&&!nt.hitTest(t||nt.pointerEvent)&&(r=fe(t||nt.pointerEvent),i=fe(d),n=x({x:r.left+r.width/2,y:r.top+r.height/2}),a=x({x:i.left+i.width/2,y:i.top+i.height/2}),w-=n.x-a.x,S-=n.y-a.y),nt.isDragging||(nt.isDragging=!0,kt(nt,"dragstart","onDragStart"))},this.drag=vt,this.endDrag=function(t){xt(t||nt.pointerEvent,!0)},this.timeSinceDrag=function(){return nt.isDragging?0:(Yt()-ot)/1e3},this.timeSinceClick=function(){return(Yt()-dt)/1e3},this.hitTest=function(t,e){return de.hitTest(nt.target,t,e)},this.getDirection=function(t,e){var r,i,n,a,o,s,l="velocity"===t&&Ft?t:"object"!=typeof t||Z?"start":"element";return"element"===l&&(o=fe(nt.target),s=fe(t)),r="start"===l?nt.x-T:"velocity"===l?Ft.getVelocity(this.target,K):o.left+o.width/2-(s.left+s.width/2),Z?r<0?"counter-clockwise":"clockwise":(e=e||2,i="start"===l?nt.y-P:"velocity"===l?Ft.getVelocity(this.target,tt):o.top+o.height/2-(s.top+s.height/2),a=(n=Math.abs(r/i))<1/e?"":r<0?"left":"right",n<e&&(""!==a&&(a+="-"),a+=i<0?"up":"down"),a)},this.applyBounds=function(t){var e,r,i,n,a,o;if(t&&u.bounds!==t)return u.bounds=t,nt.update(!0);if(s(!0),l(),k){if(e=nt.x,r=nt.y,A<e?e=A:e<E&&(e=E),C<r?r=C:r<O&&(r=O),(nt.x!==e||nt.y!==r)&&(i=!0,nt.x=nt.endX=e,Z?nt.endRotation=e:nt.y=nt.endY=r,h(L=!0),nt.autoScroll&&!nt.isDragging))for(ae(d.parentNode),n=d,ee.scrollTop=null!=window.pageYOffset?window.pageYOffset:null!=Vt.scrollTop?Vt.scrollTop:$t.body.scrollTop,ee.scrollLeft=null!=window.pageXOffset?window.pageXOffset:null!=Vt.scrollLeft?Vt.scrollLeft:$t.body.scrollLeft;n&&!o;)a=(o=ne(n.parentNode))?ee:n.parentNode,rt&&a.scrollTop>a._gsMaxScrollY&&(a.scrollTop=a._gsMaxScrollY),et&&a.scrollLeft>a._gsMaxScrollX&&(a.scrollLeft=a._gsMaxScrollX),n=a;nt.isThrowing&&(i||nt.endX>A||nt.endX<E||nt.endY>C||nt.endY<O)&&f(u.throwProps,i)}return nt},this.update=function(t,e,r){var i=nt.x,n=nt.y;return a(!e),t?nt.applyBounds():(L&&r&&h(!0),s(!0)),e&&(yt(nt.pointerX,nt.pointerY),L&&h(!0)),nt.isPressed&&!e&&(et&&.01<Math.abs(i-nt.x)||rt&&.01<Math.abs(n-nt.y)&&!Z)&&c(),nt.autoScroll&&(ae(d.parentNode),st=nt.isDragging,h(!0)),nt.autoScroll&&(ie(d,v),re(d,v)),nt},this.enable=function(t){var e,r,i;if("soft"!==t){for(r=at.length;-1<--r;)i=at[r],ce(i,"mousedown",g),ce(i,"touchstart",g),ce(i,"click",y,!0),Z||Pt(i,"cursor",u.cursor||"move"),Pt(i,"touchCallout","none"),Pt(i,"touchAction",et==rt?"none":et?"pan-y":"pan-x"),At(i)&&Pt(i.ownerSVGElement||i,"touchAction",et==rt?"none":et?"pan-y":"pan-x"),this.vars.allowContextMenu||ce(i,"contextmenu",n);Mt(at,!1)}return re(d,v),b=!0,Ft&&"soft"!==t&&Ft.track(_||d,Q?"x,y":Z?"rotation":"top,left"),_&&_.enable(),d._gsDragID=e="d"+Jt++,qt[e]=this,_&&(_.element._gsDragID=e),me.n.set(d,{x:"+=0",overwrite:!1,data:"_draggable"}),Y={t:d,data:Xt?D:d._gsTransform,tween:{},setRatio:Xt?function(){me.n.set(d,M)}:ge.a._internals.setTransformRatio||ge.a._internals.set3DTransformRatio},c(),nt.update(!0),nt},this.disable=function(t){var e,r,i=nt.isDragging;if(!Z)for(e=at.length;-1<--e;)Pt(at[e],"cursor",null);if("soft"!==t){for(e=at.length;-1<--e;)r=at[e],Pt(r,"touchCallout",null),Pt(r,"touchAction",null),ue(r,"mousedown",g),ue(r,"touchstart",g),ue(r,"click",y),ue(r,"contextmenu",n);Mt(at,!0),H&&(ue(H,"touchcancel",xt),ue(H,"touchend",xt),ue(H,"touchmove",vt)),ue($t,"mouseup",xt),ue($t,"mousemove",vt)}return ie(d,v),b=!1,Ft&&"soft"!==t&&Ft.untrack(_||d,Q?"x,y":Z?"rotation":"top,left"),_&&_.disable(),wt(h),nt.isDragging=nt.isPressed=V=!1,i&&kt(nt,"dragend","onDragEnd"),nt},this.enabled=function(t,e){return arguments.length?t?nt.enable(e):nt.disable(e):b},this.kill=function(){return nt.isThrowing=!1,me.n.killTweensOf(_||d,!0,i),nt.disable(),me.n.set(at,{clearProps:"userSelect"}),delete qt[d._gsDragID],nt},-1!==e.indexOf("scroll")&&(_=this.scrollProxy=new r(d,function(t,e){var r;for(r in e)void 0===t[r]&&(t[r]=e[r]);return t}({onKill:function(){nt.isPressed&&xt(null)}},u)),d.style.overflowY=rt&&!le?"auto":"hidden",d.style.overflowX=et&&!le?"auto":"hidden",d=_.content),!1!==u.force3D&&me.n.set(d,{force3D:!0}),Z?i.rotation=1:(et&&(i[K]=1),rt&&(i[tt]=1)),Z?(D=(M=Bt).css,M.overwrite=!1):Q&&(D=(M=et&&rt?Rt:et?Nt:Lt).css,M.overwrite=!1),this.enable()},X=de.prototype=new me.c;X.constructor=de,X.pointerX=X.pointerY=X.startX=X.startY=X.deltaX=X.deltaY=0,X.isDragging=X.isPressed=!1,de.version="0.16.5",de.zIndex=1e3,ce($t,"touchcancel",function(){}),ce($t,"contextmenu",function(t){var e;for(e in qt)qt[e].isPressed&&qt[e].endDrag()}),de.create=function(t,e){"string"==typeof t&&(t=me.n.selector(t));for(var r=t&&0!==t.length?s(t)?function(t){var e,r,i,n=[],a=t.length;for(e=0;e<a;e++)if(r=t[e],s(r))for(i=r.length,i=0;i<r.length;i++)n.push(r[i]);else r&&0!==r.length&&n.push(r);return n}(t):[t]:[],i=r.length;-1<--i;)r[i]=new de(r[i],e);return r},de.get=function(t){return qt[(Tt(t)||{})._gsDragID]},de.timeSinceDrag=function(){return(Yt()-Kt)/1e3};var W={},fe=function(t,e){if(t===window)return W.left=W.top=0,W.width=W.right=Vt.clientWidth||t.innerWidth||$t.body.clientWidth||0,W.height=W.bottom=(t.innerHeight||0)-20<Vt.clientHeight?Vt.clientHeight:t.innerHeight||$t.body.clientHeight||0,W;var r=t.pageX!==e?{left:t.pageX-I(),top:t.pageY-k(),right:t.pageX-I()+1,bottom:t.pageY-k()+1}:t.nodeType||t.left===e||t.top===e?Xt?function(t){var e,r,i=0,n=0;for(e=(t=Tt(t)).offsetWidth,r=t.offsetHeight;t;)i+=t.offsetTop,n+=t.offsetLeft,t=t.offsetParent;return{top:i,left:n,width:e,height:r}}(t):Tt(t).getBoundingClientRect():t;return r.right===e&&r.width!==e?(r.right=r.left+r.width,r.bottom=r.top+r.height):r.width===e&&(r={width:r.right-r.left,height:r.bottom-r.top,right:r.right,left:r.left,bottom:r.bottom,top:r.top}),r};return de.hitTest=function(t,e,r){if(t===e)return!1;var i,n,a,o=fe(t),s=fe(e),l=s.left>o.right||s.right<o.left||s.top>o.bottom||s.bottom<o.top;return l||!r?!l:(a=-1!==(r+"").indexOf("%"),r=parseFloat(r)||0,(i={left:Math.max(o.left,s.left),top:Math.max(o.top,s.top)}).width=Math.min(o.right,s.right)-i.left,i.height=Math.min(o.bottom,s.bottom)-i.top,!(i.width<0||i.height<0)&&(a?(r*=.01,(n=i.width*i.height)>=o.width*o.height*r||n>=s.width*s.height*r):i.width>r&&i.height>r))},Wt.style.cssText="visibility:hidden;height:1px;top:-1px;pointer-events:none;position:relative;clear:both;",de},!0);var i=me.o.Draggable},function(t,e,r){function i(t){return(i="function"==typeof Symbol&&"symbol"==typeof Symbol.iterator?function(t){return typeof t}:function(t){return t&&"function"==typeof Symbol&&t.constructor===Symbol&&t!==Symbol.prototype?"symbol":typeof t})(t)}var n;n=function(){var f="function"==typeof Float32Array;function t(o,e,s,r){if(4!==arguments.length)throw new Error("BezierEasing requires 4 arguments.");for(var t=0;t<4;++t)if("number"!=typeof arguments[t]||isNaN(arguments[t])||!isFinite(arguments[t]))throw new Error("BezierEasing arguments should be integers.");if(o<0||1<o||s<0||1<s)throw new Error("BezierEasing x values must be in [0, 1] range.");var a=f?new Float32Array(11):new Array(11);function i(t,e){return 1-3*e+3*t}function n(t,e){return 3*e-6*t}function l(t){return 3*t}function p(t,e,r){return((i(e,r)*t+n(e,r))*t+l(e))*t}function c(t,e,r){return 3*i(e,r)*t*t+2*n(e,r)*t+l(e)}function u(t){for(var e=0,r=1;10!=r&&a[r]<=t;++r)e+=.1;var i=e+.1*((t-a[--r])/(a[r+1]-a[r])),n=c(i,o,s);return.001<=n?function(t,e){for(var r=0;r<4;++r){var i=c(e,o,s);if(0===i)return e;e-=(p(e,o,s)-t)/i}return e}(t,i):0==n?i:function(t,e,r){for(var i,n,a=0;0<(i=p(n=e+(r-e)/2,o,s)-t)?r=n:e=n,1e-7<Math.abs(i)&&++a<10;);return n}(t,e,e+.1)}o==e&&s==r||function(){for(var t=0;t<11;++t)a[t]=p(.1*t,o,s)}();function h(t){return o===e&&s===r?t:0===t?0:1===t?1:p(u(t),e,r)}var d="BezierEasing("+[o,e,s,r]+")";return h.toString=function(){return d},h}return t.css={ease:t(.25,.1,.25,1),linear:t(0,0,1,1),"ease-in":t(.42,0,1,1),"ease-out":t(0,0,.58,1),"ease-in-out":t(.42,0,.58,1)},t},"object"===i(e)?t.exports=n():"function"==typeof window.define&&window.define.amd?window.define([],n):window.BezierEasing=n()},function(t,e,r){function i(t){return(i="function"==typeof Symbol&&"symbol"==typeof Symbol.iterator?function(t){return typeof t}:function(t){return t&&"function"==typeof Symbol&&t.constructor===Symbol&&t!==Symbol.prototype?"symbol":typeof t})(t)}
/*! iScroll v5.1.2 ~ (c) 2008-2014 Matteo Spinelli ~ http://cubiq.org/license */function n(t,e){for(var r in this.wrapper="string"==typeof t?o.querySelector(t):t,this.scroller=this.wrapper.children[0],this.scrollerStyle=this.scroller.style,this.options={zoomMin:1,zoomMax:4,startZoom:1,resizeScrollbars:!0,mouseWheelSpeed:20,snapThreshold:.334,startX:0,startY:0,scrollY:!0,directionLockThreshold:5,momentum:!0,bounce:!0,bounceTime:600,bounceEasing:"",preventDefault:!0,preventDefaultException:{tagName:/^(INPUT|TEXTAREA|BUTTON|SELECT|VIDEO)$/},HWCompositing:!0,useTransition:!0,useTransform:!0},e)this.options[r]=e[r];this.translateZ=this.options.HWCompositing&&m.hasPerspective?" translateZ(0)":"",$.browser.safari&&(this.translateZ=""),this.options.useTransition=m.hasTransition&&this.options.useTransition,this.options.useTransform=m.hasTransform&&this.options.useTransform,this.options.eventPassthrough=!0===this.options.eventPassthrough?"vertical":this.options.eventPassthrough,this.options.preventDefault=!this.options.eventPassthrough&&this.options.preventDefault,this.options.scrollY="vertical"!=this.options.eventPassthrough&&this.options.scrollY,this.options.scrollX="horizontal"!=this.options.eventPassthrough&&this.options.scrollX,this.options.freeScroll=this.options.freeScroll&&!this.options.eventPassthrough,this.options.freeScroll=this.options.twoWayScroll||this.options.freeScroll,this.options.directionLockThreshold=this.options.eventPassthrough?0:this.options.directionLockThreshold,this.options.bounceEasing="string"==typeof this.options.bounceEasing?m.ease[this.options.bounceEasing]||m.ease.circular:this.options.bounceEasing,this.options.resizePolling=void 0===this.options.resizePolling?60:this.options.resizePolling,!0===this.options.tap&&(this.options.tap="tap"),"scale"==this.options.shrinkScrollbars&&(this.options.useTransition=!1),this.options.invertWheelDirection=this.options.invertWheelDirection?-1:1,3==this.options.probeType&&(this.options.useTransition=!1),this.x=0,this.y=0,this.directionX=0,this.directionY=0,this._events={},this.scale=u.min(u.max(this.options.startZoom,this.options.zoomMin),this.options.zoomMax),this._init(),this.refresh(),this.scrollTo(this.options.startX,this.options.startY),this.enable()}function s(t,e,r){var i=o.createElement("div"),n=o.createElement("div");return!0===r&&(i.style.cssText="position:absolute;z-index:9999",n.style.cssText="-webkit-box-sizing:border-box;-moz-box-sizing:border-box;box-sizing:border-box;position:absolute;background:rgba(0,0,0,0.5);border:1px solid rgba(255,255,255,0.9);border-radius:3px"),n.className="iScrollIndicator","h"==t?(!0===r&&(i.style.cssText+=";height:7px;left:2px;right:2px;bottom:0",n.style.height="100%"),i.className="iScrollHorizontalScrollbar"):(!0===r&&(i.style.cssText+=";width:7px;bottom:2px;top:2px;right:1px",n.style.width="100%"),i.className="iScrollVerticalScrollbar"),i.style.cssText+=";overflow:hidden",e||(i.style.pointerEvents="none"),i.appendChild(n),i}function l(t,e){for(var r in this.wrapper="string"==typeof e.el?o.querySelector(e.el):e.el,this.wrapperStyle=this.wrapper.style,this.indicator=this.wrapper.children[0],this.indicatorStyle=this.indicator.style,this.scroller=t,this.options={listenX:!0,listenY:!0,interactive:!1,resize:!0,defaultScrollbars:!1,shrink:!1,fade:!1,speedRatioX:0,speedRatioY:0},e)this.options[r]=e[r];this.sizeRatioX=1,this.sizeRatioY=1,this.maxPosX=0,this.maxPosY=0,this.options.interactive&&(this.options.disableTouch||(m.addEvent(this.indicator,"touchstart",this),m.addEvent(a,"touchend",this)),this.options.disablePointer||(m.addEvent(this.indicator,m.prefixPointerEvent("pointerdown"),this),m.addEvent(a,m.prefixPointerEvent("pointerup"),this)),this.options.disableMouse||(m.addEvent(this.indicator,"mousedown",this),m.addEvent(a,"mouseup",this))),this.options.fade&&(this.wrapperStyle[m.style.transform]=this.scroller.translateZ,this.wrapperStyle[m.style.transitionDuration]=m.isBadAndroid?"0.001s":"0ms",this.wrapperStyle.opacity="0")}
/*! iScroll v5.1.2 ~ (c) 2008-2014 Matteo Spinelli ~ http://cubiq.org/license */
var a,o,u,f,m;a=window,o=document,u=Math,f=a.requestAnimationFrame||a.webkitRequestAnimationFrame||a.mozRequestAnimationFrame||a.oRequestAnimationFrame||a.msRequestAnimationFrame||function(t){a.setTimeout(t,1e3/60)},m=function(){var i={},n=o.createElement("div").style,e=function(){for(var t=["t","webkitT","MozT","msT","OT"],e=0,r=t.length;e<r;e++)if(t[e]+"ransform"in n)return t[e].substr(0,t[e].length-1);return!1}();function t(t){return!1!==e&&(""===e?t:e+t.charAt(0).toUpperCase()+t.substr(1))}i.getTime=Date.now||function(){return(new Date).getTime()},i.extend=function(t,e){for(var r in e)t[r]=e[r]},i.addEvent=function(t,e,r,i){t.addEventListener(e,r,!!i)},i.removeEvent=function(t,e,r,i){t.removeEventListener(e,r,!!i)},i.prefixPointerEvent=function(t){return a.MSPointerEvent?"MSPointer"+t.charAt(9).toUpperCase()+t.substr(10):t},i.momentum=function(t,e,r,i,n,a){var o,s,l=t-e,p=u.abs(l)/r;return s=p/(a=void 0===a?6e-4:a),(o=t+p*p/(2*a)*(l<0?-1:1))<i?(o=n?i-n/2.5*(p/8):i,s=(l=u.abs(o-t))/p):0<o&&(o=n?n/2.5*(p/8):0,s=(l=u.abs(t)+o)/p),{destination:u.round(o),duration:s}};var r=t("transform");return i.extend(i,{hasTransform:!1!==r,hasPerspective:t("perspective")in n,hasTouch:"ontouchstart"in a,hasPointer:a.PointerEvent||a.MSPointerEvent,hasTransition:t("transition")in n}),i.isBadAndroid=/Android /.test(a.navigator.appVersion)&&!/Chrome\/\d/.test(a.navigator.appVersion),i.extend(i.style={},{transform:r,transitionTimingFunction:t("transitionTimingFunction"),transitionDuration:t("transitionDuration"),transitionDelay:t("transitionDelay"),transformOrigin:t("transformOrigin")}),i.hasClass=function(t,e){return new RegExp("(^|\\s)"+e+"(\\s|$)").test(t.className)},i.addClass=function(t,e){if(!i.hasClass(t,e)){var r=t.className.split(" ");r.push(e),t.className=r.join(" ")}},i.removeClass=function(t,e){if(i.hasClass(t,e)){var r=new RegExp("(^|\\s)"+e+"(\\s|$)","g");t.className=t.className.replace(r," ")}},i.offset=function(t){for(var e=-t.offsetLeft,r=-t.offsetTop;t=t.offsetParent;)e-=t.offsetLeft,r-=t.offsetTop;return{left:e,top:r}},i.preventDefaultException=function(t,e){for(var r in e)if(e[r].test(t[r]))return!0;return!1},i.extend(i.eventType={},{touchstart:1,touchmove:1,touchend:1,mousedown:2,mousemove:2,mouseup:2,pointerdown:3,pointermove:3,pointerup:3,MSPointerDown:3,MSPointerMove:3,MSPointerUp:3}),i.extend(i.ease={},{quadratic:{style:"cubic-bezier(0.25, 0.46, 0.45, 0.94)",fn:function(t){return t*(2-t)}},circular:{style:"cubic-bezier(0.1, 0.57, 0.1, 1)",fn:function(t){return u.sqrt(1- --t*t)}},back:{style:"cubic-bezier(0.175, 0.885, 0.32, 1.275)",fn:function(t){return(t-=1)*t*(5*t+4)+1}},bounce:{style:"",fn:function(t){return(t/=1)<1/2.75?7.5625*t*t:t<2/2.75?7.5625*(t-=1.5/2.75)*t+.75:t<2.5/2.75?7.5625*(t-=2.25/2.75)*t+.9375:7.5625*(t-=2.625/2.75)*t+.984375}},elastic:{style:"",fn:function(t){return 0===t?0:1==t?1:.4*u.pow(2,-10*t)*u.sin((t-.055)*(2*u.PI)/.22)+1}}}),i.tap=function(t,e){var r=o.createEvent("Event");r.initEvent(e,!0,!0),r.pageX=t.pageX,r.pageY=t.pageY,t.target.dispatchEvent(r)},i.click=function(t){var e,r=t.target;/(SELECT|INPUT|TEXTAREA)/i.test(r.tagName)||((e=o.createEvent("MouseEvents")).initMouseEvent("click",!0,!0,t.view,1,r.screenX,r.screenY,r.clientX,r.clientY,t.ctrlKey,t.altKey,t.shiftKey,t.metaKey,0,null),e._constructed=!0,r.dispatchEvent(e))},i}(),n.prototype={version:"5.1.2",_init:function(){this._initEvents(),this.options.zoom&&this._initZoom(),(this.options.scrollbars||this.options.indicators)&&this._initIndicators(),this.options.mouseWheel&&this._initWheel(),this.options.snap&&this._initSnap(),this.options.keyBindings&&this._initKeys()},destroy:function(){this._initEvents(!0),this._execEvent("destroy")},_transitionEnd:function(t){t.target==this.scroller&&this.isInTransition&&(this._transitionTime(),this.resetPosition(this.options.bounceTime)||(this.isInTransition=!1,this._execEvent("scrollEnd")))},_start:function(t){if((1==m.eventType[t.type]||0===t.button)&&this.enabled&&(!this.initiated||m.eventType[t.type]===this.initiated)){!this.options.preventDefault||m.isBadAndroid||m.preventDefaultException(t.target,this.options.preventDefaultException)||t.preventDefault();var e,r=t.touches?t.touches[0]:t;this.initiated=m.eventType[t.type],this.twoWayOnEndCheck=!0,this.moved=!1,this.distX=0,this.distY=0,this.directionX=0,this.directionY=0,this.directionLocked=0,this._transitionTime(),this.startTime=m.getTime(),this.options.useTransition&&this.isInTransition?(this.isInTransition=!1,e=this.getComputedPosition(),this._translate(u.round(e.x),u.round(e.y)),this._execEvent("scrollEnd")):!this.options.useTransition&&this.isAnimating&&(this.isAnimating=!1,this._execEvent("scrollEnd")),this.startX=this.x,this.startY=this.y,this.absStartX=this.x,this.absStartY=this.y,this.pointX=r.pageX,this.pointY=r.pageY,this._execEvent("beforeScrollStart")}},_move:function(t){if(this.enabled&&m.eventType[t.type]===this.initiated){this.options.preventDefault&&t.preventDefault();var e,r,i,n,a=t.touches?t.touches[0]:t,o=a.pageX-this.pointX,s=a.pageY-this.pointY,l=m.getTime();if(this.pointX=a.pageX,this.pointY=a.pageY,this.distX+=o,this.distY+=s,i=u.abs(this.distX),n=u.abs(this.distY),!(300<l-this.endTime&&i<10&&n<10)){if(this.options.twoWayScroll&&this.options.scrollX){if(!this.directionLocked&&n<=i&&(this.directionLocked=!0),!this.directionLocked)return void(this.initiated=!1);t.preventDefault(),t.stopPropagation()}else if(this.options.twoWayScroll&&this.options.scrollY){if(!this.directionLocked&&i<n&&(this.directionLocked=!0),!this.directionLocked)return void(this.initiated=!1);t.preventDefault(),t.stopPropagation()}if(this.directionLocked||this.options.freeScroll||(i>n+this.options.directionLockThreshold?this.directionLocked="h":n>=i+this.options.directionLockThreshold?this.directionLocked="v":this.directionLocked="n"),"h"==this.directionLocked){if("vertical"==this.options.eventPassthrough)t.preventDefault();else if("horizontal"==this.options.eventPassthrough)return void(this.initiated=!1);s=0}else if("v"==this.directionLocked){if("horizontal"==this.options.eventPassthrough)t.preventDefault();else if("vertical"==this.options.eventPassthrough)return void(this.initiated=!1);o=0}o=this.hasHorizontalScroll?o:0,s=this.hasVerticalScroll?s:0,e=this.x+o,r=this.y+s,(0<e||e<this.maxScrollX)&&(e=this.options.bounce?this.x+o/3:0<e?0:this.maxScrollX),(0<r||r<this.maxScrollY)&&(r=this.options.bounce?this.y+s/3:0<r?0:this.maxScrollY),this.directionX=0<o?-1:o<0?1:0,this.directionY=0<s?-1:s<0?1:0,this.moved||this._execEvent("scrollStart"),this.moved=!0,this._translate(e,r),300<l-this.startTime&&(this.startTime=l,this.startX=this.x,this.startY=this.y,1==this.options.probeType&&this._execEvent("scroll")),1<this.options.probeType&&this._execEvent("scroll")}}},_end:function(t){if(this.twoWayOnEndCheck||this.enabled&&m.eventType[t.type]===this.initiated){this.twoWayOnEndCheck=!1,this.options.preventDefault&&!m.preventDefaultException(t.target,this.options.preventDefaultException)&&t.preventDefault(),t.changedTouches&&t.changedTouches[0];var e,r,i=m.getTime()-this.startTime,n=u.round(this.x),a=u.round(this.y),o=u.abs(n-this.startX),s=u.abs(a-this.startY),l=0,p="";if(this.isInTransition=0,this.initiated=0,this.endTime=m.getTime(),!this.resetPosition(this.options.bounceTime)){if(this.scrollTo(n,a),!this.moved&&!this.options.snap)return this.options.tap&&m.tap(t,this.options.tap),this.options.click&&m.click(t),void this._execEvent("scrollCancel");if(this.moved&&this._events.flick&&i<200&&o<100&&s<100)return 0==o&&0==s?void this._execEvent("scrollCancel"):void this._execEvent("flick");if(this.options.momentum&&i<300&&(e=this.hasHorizontalScroll?m.momentum(this.x,this.startX,i,this.maxScrollX,this.options.bounce?this.wrapperWidth:0,this.options.deceleration):{destination:n,duration:0},r=this.hasVerticalScroll?m.momentum(this.y,this.startY,i,this.maxScrollY,this.options.bounce?this.wrapperHeight:0,this.options.deceleration):{destination:a,duration:0},n=e.destination,a=r.destination,l=u.max(e.duration,r.duration),this.isInTransition=1),this.options.snap){var c=this._nearestSnap(n,a);this.currentPage=c,l=this.options.snapSpeed||u.max(u.max(u.min(u.abs(n-c.x),1e3),u.min(u.abs(a-c.y),1e3)),300),n=c.x,a=c.y,this.directionX=0,this.directionY=0,p=this.options.bounceEasing}if(n!=this.x||a!=this.y)return(0<n||n<this.maxScrollX||0<a||a<this.maxScrollY)&&(p=m.ease.quadratic),void this.scrollTo(n,a,l,p);if(!this.moved&&this.options.snap)return this.options.tap&&m.tap(t,this.options.tap),this.options.click&&m.click(t),void this._execEvent("scrollCancel");this.moved=!1,this._execEvent("scrollEnd")}}},_resize:function(){var t=this;clearTimeout(this.resizeTimeout),this.resizeTimeout=setTimeout(function(){t.refresh()},this.options.resizePolling)},resetPosition:function(t){var e=this.x,r=this.y;return t=t||0,!this.hasHorizontalScroll||0<this.x?e=0:this.x<this.maxScrollX&&(e=this.maxScrollX),!this.hasVerticalScroll||0<this.y?r=0:this.y<this.maxScrollY&&(r=this.maxScrollY),(e!=this.x||r!=this.y)&&(this.scrollTo(e,r,t,this.options.bounceEasing),!0)},disable:function(){this.enabled=!1},enable:function(){this.enabled=!0},refresh:function(){this.wrapper.offsetHeight,this.wrapperWidth=this.wrapper.clientWidth,this.wrapperHeight=this.wrapper.clientHeight,this.scrollerWidth=u.round(this.scroller.offsetWidth*this.scale),this.scrollerHeight=u.round(this.scroller.offsetHeight*this.scale),this.maxScrollX=this.wrapperWidth-this.scrollerWidth,this.maxScrollY=this.wrapperHeight-this.scrollerHeight,this.hasHorizontalScroll=this.options.scrollX&&this.maxScrollX<0,this.hasVerticalScroll=this.options.scrollY&&this.maxScrollY<0,this.hasHorizontalScroll||(this.maxScrollX=0,this.scrollerWidth=this.wrapperWidth),this.hasVerticalScroll||(this.maxScrollY=0,this.scrollerHeight=this.wrapperHeight),this.endTime=0,this.directionX=0,this.directionY=0,this.wrapperOffset=m.offset(this.wrapper),this._execEvent("refresh"),this.resetPosition()},on:function(t,e){this._events[t]||(this._events[t]=[]),this._events[t].push(e)},off:function(t,e){if(this._events[t]){var r=this._events[t].indexOf(e);-1<r&&this._events[t].splice(r,1)}},_execEvent:function(t){if(this._events[t]){var e=0,r=this._events[t].length;if(r)for(;e<r;e++)this._events[t][e].apply(this,[].slice.call(arguments,1))}},scrollBy:function(t,e,r,i){t=this.x+t,e=this.y+e,r=r||0,this.scrollTo(t,e,r,i)},scrollTo:function(t,e,r,i){i=i||m.ease.circular,this.isInTransition=this.options.useTransition&&0<r,void 0===r||this.options.useTransition&&i.style?(this._transitionTimingFunction(i.style),this._transitionTime(r),this._translate(t,e)):this._animate(t,e,r,i.fn)},scrollToElement:function(t,e,r,i,n){if(t=t.nodeType?t:this.scroller.querySelector(t)){var a=m.offset(t);a.left-=this.wrapperOffset.left,a.top-=this.wrapperOffset.top,!0===r&&(r=u.round(t.offsetWidth/2-this.wrapper.offsetWidth/2)),!0===i&&(i=u.round(t.offsetHeight/2-this.wrapper.offsetHeight/2)),a.left-=r||0,a.top-=i||0,a.left=0<a.left?0:a.left<this.maxScrollX?this.maxScrollX:a.left,a.top=0<a.top?0:a.top<this.maxScrollY?this.maxScrollY:a.top,e=null==e||"auto"===e?u.max(u.abs(this.x-a.left),u.abs(this.y-a.top)):e,this.scrollTo(a.left,a.top,e,n)}},_transitionTime:function(t){if(t=t||0,this.scrollerStyle[m.style.transitionDuration]=t+"ms",!t&&m.isBadAndroid&&(this.scrollerStyle[m.style.transitionDuration]="0.001s"),this.indicators)for(var e=this.indicators.length;e--;)this.indicators[e].transitionTime(t)},_transitionTimingFunction:function(t){if(this.scrollerStyle[m.style.transitionTimingFunction]=t,this.indicators)for(var e=this.indicators.length;e--;)this.indicators[e].transitionTimingFunction(t)},_translate:function(t,e){if(this.options.useTransform?this.scrollerStyle[m.style.transform]="translate("+t+"px,"+e+"px) scale("+this.scale+") "+this.translateZ:(t=u.round(t),e=u.round(e),this.scrollerStyle.left=t+"px",this.scrollerStyle.top=e+"px"),this.x=t,this.y=e,this.indicators)for(var r=this.indicators.length;r--;)this.indicators[r].updatePosition()},_initEvents:function(t){var e=t?m.removeEvent:m.addEvent,r=this.options.bindToWrapper?this.wrapper:a;e(a,"orientationchange",this),e(a,"resize",this),this.options.click&&e(this.wrapper,"click",this,!0),this.options.disableMouse||(e(this.wrapper,"mousedown",this),e(r,"mousemove",this),e(r,"mousecancel",this),e(r,"mouseup",this)),m.hasPointer&&!this.options.disablePointer&&(e(this.wrapper,m.prefixPointerEvent("pointerdown"),this),e(r,m.prefixPointerEvent("pointermove"),this),e(r,m.prefixPointerEvent("pointercancel"),this),e(r,m.prefixPointerEvent("pointerup"),this)),m.hasTouch&&!this.options.disableTouch&&(e(this.wrapper,"touchstart",this),e(r,"touchmove",this),e(r,"touchcancel",this),e(r,"touchend",this)),e(this.scroller,"transitionend",this),e(this.scroller,"webkitTransitionEnd",this),e(this.scroller,"oTransitionEnd",this),e(this.scroller,"MSTransitionEnd",this)},getComputedPosition:function(){var t,e,r=a.getComputedStyle(this.scroller,null);return e=this.options.useTransform?(t=+((r=r[m.style.transform].split(")")[0].split(", "))[12]||r[4]),+(r[13]||r[5])):(t=+r.left.replace(/[^-\d.]/g,""),+r.top.replace(/[^-\d.]/g,"")),{x:t,y:e}},_initIndicators:function(){var t,e=this.options.interactiveScrollbars,r="string"!=typeof this.options.scrollbars,i=[],n=this;this.indicators=[],this.options.scrollbars&&(this.options.scrollY&&(t={el:s("v",e,this.options.scrollbars),interactive:e,defaultScrollbars:!0,customStyle:r,resize:this.options.resizeScrollbars,shrink:this.options.shrinkScrollbars,fade:this.options.fadeScrollbars,listenX:!1},this.wrapper.appendChild(t.el),i.push(t)),this.options.scrollX&&(t={el:s("h",e,this.options.scrollbars),interactive:e,defaultScrollbars:!0,customStyle:r,resize:this.options.resizeScrollbars,shrink:this.options.shrinkScrollbars,fade:this.options.fadeScrollbars,listenY:!1},this.wrapper.appendChild(t.el),i.push(t))),this.options.indicators&&(i=i.concat(this.options.indicators));for(var a=i.length;a--;)this.indicators.push(new l(this,i[a]));function o(t){for(var e=n.indicators.length;e--;)t.call(n.indicators[e])}this.options.fadeScrollbars&&(this.on("scrollEnd",function(){o(function(){this.fade()})}),this.on("scrollCancel",function(){o(function(){this.fade()})}),this.on("scrollStart",function(){o(function(){this.fade(1)})}),this.on("beforeScrollStart",function(){o(function(){this.fade(1,!0)})})),this.on("refresh",function(){o(function(){this.refresh()})}),this.on("destroy",function(){o(function(){this.destroy()}),delete this.indicators})},_initZoom:function(){this.scrollerStyle[m.style.transformOrigin]="0 0"},_zoomStart:function(t){var e=u.abs(t.touches[0].pageX-t.touches[1].pageX),r=u.abs(t.touches[0].pageY-t.touches[1].pageY);this.touchesDistanceStart=u.sqrt(e*e+r*r),this.startScale=this.scale,this.originX=u.abs(t.touches[0].pageX+t.touches[1].pageX)/2+this.wrapperOffset.left-this.x,this.originY=u.abs(t.touches[0].pageY+t.touches[1].pageY)/2+this.wrapperOffset.top-this.y,this._execEvent("zoomStart")},_zoom:function(t){if(this.enabled&&m.eventType[t.type]===this.initiated){this.options.preventDefault&&t.preventDefault();var e,r,i,n=u.abs(t.touches[0].pageX-t.touches[1].pageX),a=u.abs(t.touches[0].pageY-t.touches[1].pageY),o=u.sqrt(n*n+a*a),s=1/this.touchesDistanceStart*o*this.startScale;this.scaled=!0,s<this.options.zoomMin?s=.5*this.options.zoomMin*u.pow(2,s/this.options.zoomMin):s>this.options.zoomMax&&(s=2*this.options.zoomMax*u.pow(.5,this.options.zoomMax/s)),e=s/this.startScale,r=this.originX-this.originX*e+this.startX,i=this.originY-this.originY*e+this.startY,this.scale=s,this.scrollTo(r,i)}},_zoomEnd:function(t){var e,r,i;this.enabled&&m.eventType[t.type]===this.initiated&&(this.options.preventDefault&&t.preventDefault(),this.isInTransition=0,this.initiated=0,this.scale>this.options.zoomMax?this.scale=this.options.zoomMax:this.scale<this.options.zoomMin&&(this.scale=this.options.zoomMin),this.refresh(),i=this.scale/this.startScale,e=this.originX-this.originX*i+this.startX,r=this.originY-this.originY*i+this.startY,0<e?e=0:e<this.maxScrollX&&(e=this.maxScrollX),0<r?r=0:r<this.maxScrollY&&(r=this.maxScrollY),this.x==e&&this.y==r||this.scrollTo(e,r,this.options.bounceTime),this.scaled=!1,this._execEvent("zoomEnd"))},zoom:function(t,e,r,i){if(t<this.options.zoomMin?t=this.options.zoomMin:t>this.options.zoomMax&&(t=this.options.zoomMax),t!=this.scale){var n=t/this.scale;e=void 0===e?this.wrapperWidth/2:e,r=void 0===r?this.wrapperHeight/2:r,i=void 0===i?300:i,e=e+this.wrapperOffset.left-this.x,r=r+this.wrapperOffset.top-this.y,e=e-e*n+this.x,r=r-r*n+this.y,this.scale=t,this.refresh(),0<e?e=0:e<this.maxScrollX&&(e=this.maxScrollX),0<r?r=0:r<this.maxScrollY&&(r=this.maxScrollY),this.scrollTo(e,r,i)}},_wheelZoom:function(t){var e,r,i=this;if(clearTimeout(this.wheelTimeout),this.wheelTimeout=setTimeout(function(){i._execEvent("zoomEnd")},400),"deltaX"in t)e=-t.deltaY/u.abs(t.deltaY);else if("wheelDeltaX"in t)e=t.wheelDeltaY/u.abs(t.wheelDeltaY);else if("wheelDelta"in t)e=t.wheelDelta/u.abs(t.wheelDelta);else{if(!("detail"in t))return;e=-t.detail/u.abs(t.wheelDelta)}r=this.scale+e/5,this.zoom(r,t.pageX,t.pageY,0)},_initWheel:function(){m.addEvent(this.wrapper,"wheel",this),m.addEvent(this.wrapper,"mousewheel",this),m.addEvent(this.wrapper,"DOMMouseScroll",this),this.on("destroy",function(){m.removeEvent(this.wrapper,"wheel",this),m.removeEvent(this.wrapper,"mousewheel",this),m.removeEvent(this.wrapper,"DOMMouseScroll",this)})},_wheel:function(t){if(this.enabled){t.preventDefault(),t.stopPropagation();var e,r,i,n,a=this;if(void 0===this.wheelTimeout&&a._execEvent("scrollStart"),clearTimeout(this.wheelTimeout),this.wheelTimeout=setTimeout(function(){a._execEvent("scrollEnd"),a.wheelTimeout=void 0},400),"deltaX"in t)e=-t.deltaX,r=-t.deltaY;else if("wheelDeltaX"in t)e=t.wheelDeltaX/120*this.options.mouseWheelSpeed,r=t.wheelDeltaY/120*this.options.mouseWheelSpeed;else if("wheelDelta"in t)e=r=t.wheelDelta/120*this.options.mouseWheelSpeed;else{if(!("detail"in t))return;e=r=-t.detail/3*this.options.mouseWheelSpeed}if(e*=this.options.invertWheelDirection,r*=this.options.invertWheelDirection,this.hasVerticalScroll||(e=r,r=0),this.options.snap)return i=this.currentPage.pageX,n=this.currentPage.pageY,0<e?i--:e<0&&i++,0<r?n--:r<0&&n++,void this.goToPage(i,n);i=this.x+u.round(this.hasHorizontalScroll?e:0),n=this.y+u.round(this.hasVerticalScroll?r:0),0<i?i=0:i<this.maxScrollX&&(i=this.maxScrollX),0<n?n=0:n<this.maxScrollY&&(n=this.maxScrollY),this.scrollTo(i,n),1<this.options.probeType&&this._execEvent("scroll")}},_initSnap:function(){this.currentPage={},"string"==typeof this.options.snap&&(this.options.snap=this.scroller.querySelectorAll(this.options.snap)),this.on("refresh",function(){var t,e,r,i,n,a,o=0,s=0,l=0,p=this.options.snapStepX||this.wrapperWidth,c=this.options.snapStepY||this.wrapperHeight;if(this.pages=[],this.wrapperWidth&&this.wrapperHeight&&this.scrollerWidth&&this.scrollerHeight){if(!0===this.options.snap)for(r=u.round(p/2),i=u.round(c/2);l>-this.scrollerWidth;){for(this.pages[o]=[],n=t=0;n>-this.scrollerHeight;)this.pages[o][t]={x:u.max(l,this.maxScrollX),y:u.max(n,this.maxScrollY),width:p,height:c,cx:l-r,cy:n-i},n-=c,t++;l-=p,o++}else for(t=(a=this.options.snap).length,e=-1;o<t;o++)(0===o||a[o].offsetLeft<=a[o-1].offsetLeft)&&(s=0,e++),this.pages[s]||(this.pages[s]=[]),l=u.max(-a[o].offsetLeft,this.maxScrollX),n=u.max(-a[o].offsetTop,this.maxScrollY),r=l-u.round(a[o].offsetWidth/2),i=n-u.round(a[o].offsetHeight/2),this.pages[s][e]={x:l,y:n,width:a[o].offsetWidth,height:a[o].offsetHeight,cx:r,cy:i},l>this.maxScrollX&&s++;"{}"===JSON.stringify(this.currentPage)&&(this.currentPage=this._nearestSnap(this.options.startX,this.options.startY)),this.goToPage(this.currentPage.pageX,this.currentPage.pageY,0),this.options.snapThreshold%1==0?(this.snapThresholdX=this.options.snapThreshold,this.snapThresholdY=this.options.snapThreshold):(this.snapThresholdX=u.round(this.pages[this.currentPage.pageX][this.currentPage.pageY].width*this.options.snapThreshold),this.snapThresholdY=u.round(this.pages[this.currentPage.pageX][this.currentPage.pageY].height*this.options.snapThreshold))}}),this.on("flick",function(){var t=this.options.snapSpeed||u.max(u.max(u.min(u.abs(this.x-this.startX),1e3),u.min(u.abs(this.y-this.startY),1e3)),300);this.goToPage(this.currentPage.pageX+this.directionX,this.currentPage.pageY+this.directionY,t)})},_nearestSnap:function(t,e){if(!this.pages.length)return{x:0,y:0,pageX:0,pageY:0};var r=0,i=this.pages.length,n=0;if(u.abs(t-this.absStartX)<this.snapThresholdX&&u.abs(e-this.absStartY)<this.snapThresholdY)return this.currentPage;for(0<t?t=0:t<this.maxScrollX&&(t=this.maxScrollX),0<e?e=0:e<this.maxScrollY&&(e=this.maxScrollY);r<i;r++)if(t>=this.pages[r][0].cx){t=this.pages[r][0].x;break}for(i=this.pages[r].length;n<i;n++)if(e>=this.pages[0][n].cy){e=this.pages[0][n].y;break}return r==this.currentPage.pageX&&((r+=this.directionX)<0?r=0:r>=this.pages.length&&(r=this.pages.length-1),t=this.pages[r][0].x),n==this.currentPage.pageY&&((n+=this.directionY)<0?n=0:n>=this.pages[0].length&&(n=this.pages[0].length-1),e=this.pages[0][n].y),{x:t,y:e,pageX:r,pageY:n}},goToPage:function(t,e,r,i){if(i=i||this.options.bounceEasing,void 0===this.pages||void 0!==this.pages&&0==this.pages.length)return!1;t>=this.pages.length?t=this.pages.length-1:t<0&&(t=0),e>=this.pages[t].length?e=this.pages[t].length-1:e<0&&(e=0);var n=this.pages[t][e].x,a=this.pages[t][e].y;r=void 0===r?this.options.snapSpeed||u.max(u.max(u.min(u.abs(n-this.x),1e3),u.min(u.abs(a-this.y),1e3)),300):r,this.currentPage={x:n,y:a,pageX:t,pageY:e},this.scrollTo(n,a,r,i)},next:function(t,e){var r=this.currentPage.pageX,i=this.currentPage.pageY;++r>=this.pages.length&&this.hasVerticalScroll&&(r=0,i++),this.goToPage(r,i,t,e)},prev:function(t,e){var r=this.currentPage.pageX,i=this.currentPage.pageY;--r<0&&this.hasVerticalScroll&&(r=0,i--),this.goToPage(r,i,t,e)},_initKeys:function(t){var e,r={pageUp:33,pageDown:34,end:35,home:36,left:37,up:38,right:39,down:40};if("object"==i(this.options.keyBindings))for(e in this.options.keyBindings)"string"==typeof this.options.keyBindings[e]&&(this.options.keyBindings[e]=this.options.keyBindings[e].toUpperCase().charCodeAt(0));else this.options.keyBindings={};for(e in r)this.options.keyBindings[e]=this.options.keyBindings[e]||r[e];m.addEvent(a,"keydown",this),this.on("destroy",function(){m.removeEvent(a,"keydown",this)})},_key:function(t){if(this.enabled){var e,r=this.options.snap,i=r?this.currentPage.pageX:this.x,n=r?this.currentPage.pageY:this.y,a=m.getTime(),o=this.keyTime||0;switch(this.options.useTransition&&this.isInTransition&&(e=this.getComputedPosition(),this._translate(u.round(e.x),u.round(e.y)),this.isInTransition=!1),this.keyAcceleration=a-o<200?u.min(this.keyAcceleration+.25,50):0,t.keyCode){case this.options.keyBindings.pageUp:this.hasHorizontalScroll&&!this.hasVerticalScroll?i+=r?1:this.wrapperWidth:n+=r?1:this.wrapperHeight;break;case this.options.keyBindings.pageDown:this.hasHorizontalScroll&&!this.hasVerticalScroll?i-=r?1:this.wrapperWidth:n-=r?1:this.wrapperHeight;break;case this.options.keyBindings.end:i=r?this.pages.length-1:this.maxScrollX,n=r?this.pages[0].length-1:this.maxScrollY;break;case this.options.keyBindings.home:n=i=0;break;case this.options.keyBindings.left:i+=r?-1:5+this.keyAcceleration>>0;break;case this.options.keyBindings.up:n+=r?1:5+this.keyAcceleration>>0;break;case this.options.keyBindings.right:i-=r?-1:5+this.keyAcceleration>>0;break;case this.options.keyBindings.down:n-=r?1:5+this.keyAcceleration>>0;break;default:return}r?this.goToPage(i,n):(0<i?(i=0,this.keyAcceleration=0):i<this.maxScrollX&&(i=this.maxScrollX,this.keyAcceleration=0),0<n?(n=0,this.keyAcceleration=0):n<this.maxScrollY&&(n=this.maxScrollY,this.keyAcceleration=0),this.scrollTo(i,n),this.keyTime=a)}},_animate:function(a,o,s,l){var p=this,c=this.x,u=this.y,h=m.getTime(),d=h+s;this.isAnimating=!0,function t(){var e,r,i,n=m.getTime();if(d<=n)return p.isAnimating=!1,p._translate(a,o),void(p.resetPosition(p.options.bounceTime)||p._execEvent("scrollEnd"));i=l(n=(n-h)/s),e=(a-c)*i+c,r=(o-u)*i+u,p._translate(e,r),p.isAnimating&&f(t),3==p.options.probeType&&p._execEvent("scroll")}()},handleEvent:function(t){switch(t.type){case"touchstart":case"pointerdown":case"MSPointerDown":case"mousedown":if(t.touches&&3==t.touches.length)return;this._start(t),this.options.zoom&&t.touches&&1<t.touches.length&&this._zoomStart(t);break;case"touchmove":case"pointermove":case"MSPointerMove":case"mousemove":if(t.touches&&3==t.touches.length)return;if(this.options.zoom&&t.touches&&t.touches[1])return void this._zoom(t);this._move(t);break;case"touchend":case"pointerup":case"MSPointerUp":case"mouseup":case"touchcancel":case"pointercancel":case"MSPointerCancel":case"mousecancel":if(t.touches&&3==t.touches.length)return;if(this.scaled)return void this._zoomEnd(t);this._end(t);break;case"orientationchange":case"resize":this._resize();break;case"transitionend":case"webkitTransitionEnd":case"oTransitionEnd":case"MSTransitionEnd":this._transitionEnd(t);break;case"wheel":case"DOMMouseScroll":case"mousewheel":if("zoom"==this.options.wheelAction)return void this._wheelZoom(t);this._wheel(t);break;case"keydown":this._key(t)}}},l.prototype={handleEvent:function(t){switch(t.type){case"touchstart":case"pointerdown":case"MSPointerDown":case"mousedown":this._start(t);break;case"touchmove":case"pointermove":case"MSPointerMove":case"mousemove":this._move(t);break;case"touchend":case"pointerup":case"MSPointerUp":case"mouseup":case"touchcancel":case"pointercancel":case"MSPointerCancel":case"mousecancel":this._end(t)}},destroy:function(){this.options.interactive&&(m.removeEvent(this.indicator,"touchstart",this),m.removeEvent(this.indicator,m.prefixPointerEvent("pointerdown"),this),m.removeEvent(this.indicator,"mousedown",this),m.removeEvent(a,"touchmove",this),m.removeEvent(a,m.prefixPointerEvent("pointermove"),this),m.removeEvent(a,"mousemove",this),m.removeEvent(a,"touchend",this),m.removeEvent(a,m.prefixPointerEvent("pointerup"),this),m.removeEvent(a,"mouseup",this)),this.options.defaultScrollbars&&this.wrapper.parentNode.removeChild(this.wrapper)},_start:function(t){var e=t.touches?t.touches[0]:t;t.preventDefault(),t.stopPropagation(),this.transitionTime(),this.initiated=!0,this.moved=!1,this.lastPointX=e.pageX,this.lastPointY=e.pageY,this.startTime=m.getTime(),this.options.disableTouch||m.addEvent(a,"touchmove",this),this.options.disablePointer||m.addEvent(a,m.prefixPointerEvent("pointermove"),this),this.options.disableMouse||m.addEvent(a,"mousemove",this),this.scroller._execEvent("beforeScrollStart")},_move:function(t){var e,r,i,n,a=t.touches?t.touches[0]:t,o=m.getTime();this.moved||this.scroller._execEvent("scrollStart"),this.moved=!0,e=a.pageX-this.lastPointX,this.lastPointX=a.pageX,r=a.pageY-this.lastPointY,this.lastPointY=a.pageY,i=this.x+e,n=this.y+r,this._pos(i,n),1==this.scroller.options.probeType&&300<o-this.startTime?(this.startTime=o,this.scroller._execEvent("scroll")):1<this.scroller.options.probeType&&this.scroller._execEvent("scroll"),t.preventDefault(),t.stopPropagation()},_end:function(t){if(this.initiated){if(this.initiated=!1,t.preventDefault(),t.stopPropagation(),m.removeEvent(a,"touchmove",this),m.removeEvent(a,m.prefixPointerEvent("pointermove"),this),m.removeEvent(a,"mousemove",this),this.scroller.options.snap){var e=this.scroller._nearestSnap(this.scroller.x,this.scroller.y),r=this.options.snapSpeed||u.max(u.max(u.min(u.abs(this.scroller.x-e.x),1e3),u.min(u.abs(this.scroller.y-e.y),1e3)),300);this.scroller.x==e.x&&this.scroller.y==e.y||(this.scroller.directionX=0,this.scroller.directionY=0,this.scroller.currentPage=e,this.scroller.scrollTo(e.x,e.y,r,this.scroller.options.bounceEasing))}this.moved&&this.scroller._execEvent("scrollEnd")}},transitionTime:function(t){t=t||0,this.indicatorStyle[m.style.transitionDuration]=t+"ms",!t&&m.isBadAndroid&&(this.indicatorStyle[m.style.transitionDuration]="0.001s")},transitionTimingFunction:function(t){this.indicatorStyle[m.style.transitionTimingFunction]=t},refresh:function(){if(this.transitionTime(),this.options.listenX&&!this.options.listenY?this.indicatorStyle.display=this.scroller.hasHorizontalScroll?"block":"none":this.options.listenY&&!this.options.listenX?this.indicatorStyle.display=this.scroller.hasVerticalScroll?"block":"none":this.indicatorStyle.display=this.scroller.hasHorizontalScroll||this.scroller.hasVerticalScroll?"block":"none",this.scroller.hasHorizontalScroll&&this.scroller.hasVerticalScroll?(m.addClass(this.wrapper,"iScrollBothScrollbars"),m.removeClass(this.wrapper,"iScrollLoneScrollbar"),this.options.defaultScrollbars&&this.options.customStyle&&(this.options.listenX?this.wrapper.style.right="8px":this.wrapper.style.bottom="8px")):(m.removeClass(this.wrapper,"iScrollBothScrollbars"),m.addClass(this.wrapper,"iScrollLoneScrollbar"),this.options.defaultScrollbars&&this.options.customStyle&&(this.options.listenX?this.wrapper.style.right="2px":this.wrapper.style.bottom="2px")),this.wrapper.offsetHeight,this.options.listenX&&(this.wrapperWidth=this.wrapper.clientWidth,this.options.resize?(this.indicatorWidth=u.max(u.round(this.wrapperWidth*this.wrapperWidth/(this.scroller.scrollerWidth||this.wrapperWidth||1)),8),this.indicatorStyle.width=this.indicatorWidth+"px"):this.indicatorWidth=this.indicator.clientWidth,this.maxPosX=this.wrapperWidth-this.indicatorWidth,"clip"==this.options.shrink?(this.minBoundaryX=8-this.indicatorWidth,this.maxBoundaryX=this.wrapperWidth-8):(this.minBoundaryX=0,this.maxBoundaryX=this.maxPosX),this.sizeRatioX=this.options.speedRatioX||this.scroller.maxScrollX&&this.maxPosX/this.scroller.maxScrollX),this.options.listenY){var t=void 0!==prx.devices[prx.device]&&"crown"==prx.devices[prx.device].scrollType||void 0!==prx.deviceExtraParams&&"crown"==prx.deviceExtraParams.scrollType;this.wrapperHeight=this.wrapper.clientHeight,this.options.resize?(this.indicatorHeight=u.max(u.round(this.wrapperHeight*this.wrapperHeight/(this.scroller.scrollerHeight||this.wrapperHeight||1)),8),this.indicatorStyle.height=this.indicatorHeight+"px"):this.indicatorHeight=this.indicator.offsetHeight,this.maxPosY=this.wrapperHeight-this.indicatorHeight-(1==t?3:0),"clip"==this.options.shrink?(this.minBoundaryY=8-this.indicatorHeight,this.maxBoundaryY=this.wrapperHeight-8):(this.minBoundaryY=1==t?3:0,this.maxBoundaryY=this.maxPosY),this.maxPosY=this.wrapperHeight-this.indicatorHeight,this.sizeRatioY=this.options.speedRatioY||this.scroller.maxScrollY&&this.maxPosY/this.scroller.maxScrollY}this.updatePosition()},updatePosition:function(){var t=this.options.listenX&&u.round(this.sizeRatioX*this.scroller.x)||0,e=this.options.listenY&&u.round(this.sizeRatioY*this.scroller.y)||0;this.options.ignoreBoundaries||(t<this.minBoundaryX?("scale"==this.options.shrink&&(this.width=u.max(this.indicatorWidth+t,8),this.indicatorStyle.width=this.width+"px"),t=this.minBoundaryX):t>this.maxBoundaryX?t="scale"==this.options.shrink?(this.width=u.max(this.indicatorWidth-(t-this.maxPosX),8),this.indicatorStyle.width=this.width+"px",this.maxPosX+this.indicatorWidth-this.width):this.maxBoundaryX:"scale"==this.options.shrink&&this.width!=this.indicatorWidth&&(this.width=this.indicatorWidth,this.indicatorStyle.width=this.width+"px"),e<this.minBoundaryY?("scale"==this.options.shrink&&(this.height=u.max(this.indicatorHeight+3*e,8),this.indicatorStyle.height=this.height+"px"),e=this.minBoundaryY):e>this.maxBoundaryY?e="scale"==this.options.shrink?(this.height=u.max(this.indicatorHeight-3*(e-this.maxPosY),8),this.indicatorStyle.height=this.height+"px",this.maxPosY+this.indicatorHeight-this.height):this.maxBoundaryY:"scale"==this.options.shrink&&this.height!=this.indicatorHeight&&(this.height=this.indicatorHeight,this.indicatorStyle.height=this.height+"px")),this.x=t,this.y=e,this.scroller.options.useTransform?this.indicatorStyle[m.style.transform]="translate("+t+"px,"+e+"px)"+this.scroller.translateZ:(this.indicatorStyle.left=t+"px",this.indicatorStyle.top=e+"px")},_pos:function(t,e){t<0?t=0:t>this.maxPosX&&(t=this.maxPosX),e<0?e=0:e>this.maxPosY&&(e=this.maxPosY),t=this.options.listenX?u.round(t/this.sizeRatioX):this.scroller.x,e=this.options.listenY?u.round(e/this.sizeRatioY):this.scroller.y,this.scroller.scrollTo(t,e)},fade:function(t,e){if(!e||this.visible){clearTimeout(this.fadeTimeout),this.fadeTimeout=null;var r=t?250:500,i=t?0:300;t=t?"1":"0",this.wrapperStyle[m.style.transitionDuration]=r+"ms",this.fadeTimeout=setTimeout(function(t){this.wrapperStyle.opacity=t,this.visible=+t}.bind(this,t),i)}}},n.utils=m,t.exports?t.exports=n:a.IScroll=n},function(E,t){function F(t){return(F="function"==typeof Symbol&&"symbol"==typeof Symbol.iterator?function(t){return typeof t}:function(t){return t&&"function"==typeof Symbol&&t.constructor===Symbol&&t!==Symbol.prototype?"symbol":typeof t})(t)}(function(){function c(t){return!t||"loaded"==t||"complete"==t}function u(){for(var t=1,e=-1;y.length-++e&&(!y[e].s||(t=y[e].r)););t&&h()}function h(){var t=y.shift();x=1,t?t.t?g(function(){"c"==t.t?function(t){var i,e=f.createElement("link");if(e.href=t.s,e.rel="stylesheet",e.type="text/css",t.e||!l&&!a)e.onload=function(){i||(i=1,g(function(){u()},0))},t.e&&e.onload();else{!function e(r){g(function(){if(!i)try{r.sheet.cssRules.length?(i=1,u()):e(r)}catch(t){1e3==t.code||"security"==t.message||"denied"==t.message?(i=1,g(function(){u()},0)):e(r)}},0)}(e)}g(function(){i||(i=1,u())},m.errorTimeout),t.e||v.parentNode.insertBefore(e,v)}(t):function(t){var e,r=f.createElement("script");r.src=t.s,r.onreadystatechange=r.onload=function(){!e&&c(r.readyState)&&(e=1,u(),r.onload=r.onreadystatechange=null)},g(function(){e||(e=1,u())},m.errorTimeout),t.e?r.onload():v.parentNode.insertBefore(r,v)}(t)},0):(t(),u()):x=0}function e(t,e,r){var i="c"==e?T:S;return x=0,e=e||"j",k(t)?function(t,e,r,i,n,a){function o(){!l&&c(s.readyState)&&(p.r=l=1,x||u(),s.onload=s.onreadystatechange=null,g(function(){_.removeChild(s)},0))}var s=f.createElement(t),l=0,p={t:r,s:e,e:a};s.src=s.data=e,b||(s.style.display="none"),s.width=s.height="0","object"!=t&&(s.type=r),s.onload=s.onreadystatechange=o,"img"==t?s.onerror=o:"script"==t&&(s.onerror=function(){p.e=p.r=1,h()}),y.splice(i,0,p),_.insertBefore(s,b?null:v),g(function(){l||(_.removeChild(s),p.r=p.e=l=1,u())},m.errorTimeout)}(i,t,e,this.i++,0,r):(y.splice(this.i++,0,t),1==y.length&&h()),this}function s(){var t=m;return t.loader={load:e,i:0},t}function p(t){return Object(t)===t}function d(t){return"[object Function]"==n.call(t)}var t,f,r,m,i,g,v,n,y,x,a,b,_,o,l,w,S,T,P,k,I,A;window.Modernizr=function(o,p,y){function t(t,e){var r=t.charAt(0).toUpperCase()+t.substr(1);return i((t+" "+_.join(r+" ")+r).split(" "),e)}function i(t,e){for(var r in t)if(m[t[r]]!==y)return"pfx"!=e||t[r];return!1}function e(t,e){return!!~(""+t).indexOf(e)}function n(t,e){return F(t)===e}function r(t){m.cssText=t}function a(t,e,r,i){var n,a,o,s=p.createElement("div");if(parseInt(r,10))for(;r--;)(o=p.createElement("div")).id=i?i[r]:d+(r+1),s.appendChild(o);return n=["&shy;","<style>",t,"</style>"].join(""),s.id=d,s.innerHTML+=n,h.appendChild(s),a=e(s,t),s.parentNode.removeChild(s),!!a}var s,l,c,u={},h=p.documentElement,d=(p.head||p.getElementsByTagName("head")[0],"modernizr"),f=p.createElement(d),m=f.style,g=p.createElement("input"),v=":)",x=Object.prototype.toString,b=" -webkit- -moz- -o- -ms- -khtml- ".split(" "),_="Webkit Moz O ms Khtml".split(" "),w="http://www.w3.org/2000/svg",S={},T={},P={},k=[],I=(c={select:"input",change:"input",submit:"form",reset:"form",error:"img",load:"img",abort:"img"},function(t,e){e=e||p.createElement(c[t]||"div");var r=(t="on"+t)in e;return r||(e.setAttribute||(e=p.createElement("div")),e.setAttribute&&e.removeAttribute&&(e.setAttribute(t,""),r=n(e[t],"function"),n(e[t],y)||(e[t]=y),e.removeAttribute(t))),e=null,r}),A={}.hasOwnProperty;l=n(A,y)||n(A.call,y)?function(t,e){return e in t&&n(t.constructor.prototype[e],y)}:function(t,e){return A.call(t,e)};var E,C,O,M;E=["fontface","touch","csstransforms3d","generatedcontent"],C=['@font-face {font-family:"font";src:url("https://")}',["@media (",b.join("touch-enabled),("),d,")","{#touch{top:9px;position:absolute}}"].join(""),["@media (",b.join("transform-3d),("),d,")","{#csstransforms3d{left:9px;position:absolute}}"].join(""),['#generatedcontent:after{content:"',v,'";visibility:hidden}'].join("")].join(""),a(C,function(t,e){for(var r=p.styleSheets[p.styleSheets.length-1],i=r.cssRules&&r.cssRules[0]?r.cssRules[0].cssText:r.cssText||"",n=t.childNodes,a={};O--;)a[n[O].id]=n[O];u.touch="ontouchstart"in o||9===a.touch.offsetTop,u.csstransforms3d=9===a.csstransforms3d.offsetLeft,u.generatedcontent=1<=a.generatedcontent.offsetHeight,u.fontface=/src/i.test(i)&&0===i.indexOf(e.split(" ")[0])},O=E.length,E);for(var D in S.flexbox=function(){var t,e,r,i,n,a,o=p.createElement("div"),s=p.createElement("div");i="display",n="box",a="width:42px;padding:0;",i+=":",o.style.cssText=(i+b.join(n+";"+i)).slice(0,-i.length)+(a||""),t="box-flex",e="1",r="width:10px;",s.style.cssText=b.join(t+":"+e+";")+(r||""),o.appendChild(s),h.appendChild(o);var l=42===s.offsetWidth;return o.removeChild(s),h.removeChild(o),l},S.canvas=function(){var t=p.createElement("canvas");return!!t.getContext&&!!t.getContext("2d")},S.canvastext=function(){return!!u.canvas&&!!n(p.createElement("canvas").getContext("2d").fillText,"function")},S.webgl=function(){return!!o.WebGLRenderingContext},S.touch=function(){return u.touch},S.geolocation=function(){return!!navigator.geolocation},S.postmessage=function(){return!!o.postMessage},S.websqldatabase=function(){return!!o.openDatabase},S.indexedDB=function(){for(var t=-1,e=_.length;++t<e;)if(o[_[t].toLowerCase()+"IndexedDB"])return!0;return!!o.indexedDB},S.hashchange=function(){return I("hashchange",o)&&(p.documentMode===y||7<p.documentMode)},S.history=function(){return!!o.history&&!!history.pushState},S.draganddrop=function(){return I("dragstart")&&I("drop")},S.websockets=function(){for(var t=-1,e=_.length;++t<e;)if(o[_[t]+"WebSocket"])return!0;return"WebSocket"in o},S.rgba=function(){return r("background-color:rgba(150,255,150,.5)"),e(m.backgroundColor,"rgba")},S.hsla=function(){return r("background-color:hsla(120,40%,100%,.5)"),e(m.backgroundColor,"rgba")||e(m.backgroundColor,"hsla")},S.multiplebgs=function(){return r("background:url(https://),url(https://),red url(https://)"),/(url\s*\(.*?){3}/.test(m.background)},S.backgroundsize=function(){return t("backgroundSize")},S.borderimage=function(){return t("borderImage")},S.borderradius=function(){return t("borderRadius")},S.boxshadow=function(){return t("boxShadow")},S.textshadow=function(){return""===p.createElement("div").style.textShadow},S.opacity=function(){return function(t,e){r(b.join(t+";")+(e||""))}("opacity:.55"),/^0.55$/.test(m.opacity)},S.cssanimations=function(){return t("animationName")},S.csscolumns=function(){return t("columnCount")},S.cssgradients=function(){var t="background-image:";return r((t+b.join("gradient(linear,left top,right bottom,from(#9f9),to(white));"+t)+b.join("linear-gradient(left top,#9f9, white);"+t)).slice(0,-t.length)),e(m.backgroundImage,"gradient")},S.cssreflections=function(){return t("boxReflect")},S.csstransforms=function(){return!!i(["transformProperty","WebkitTransform","MozTransform","OTransform","msTransform"])},S.csstransforms3d=function(){var t=!!i(["perspectiveProperty","WebkitPerspective","MozPerspective","OPerspective","msPerspective"]);return t&&"webkitPerspective"in h.style&&(t=u.csstransforms3d),t},S.csstransitions=function(){return t("transitionProperty")},S.fontface=function(){return u.fontface},S.generatedcontent=function(){return u.generatedcontent},S.video=function(){var t=p.createElement("video"),e=!1;try{if(e=!!t.canPlayType){(e=new Boolean(e)).ogg=t.canPlayType('video/ogg; codecs="theora"');var r='video/mp4; codecs="avc1.42E01E';e.h264=t.canPlayType(r+'"')||t.canPlayType(r+', mp4a.40.2"'),e.webm=t.canPlayType('video/webm; codecs="vp8, vorbis"')}}catch(t){}return e},S.audio=function(){var t=p.createElement("audio"),e=!1;try{(e=!!t.canPlayType)&&((e=new Boolean(e)).ogg=t.canPlayType('audio/ogg; codecs="vorbis"'),e.mp3=t.canPlayType("audio/mpeg;"),e.wav=t.canPlayType('audio/wav; codecs="1"'),e.m4a=t.canPlayType("audio/x-m4a;")||t.canPlayType("audio/aac;"))}catch(t){}return e},S.localstorage=function(){try{return!!localStorage.getItem}catch(t){return!1}},S.sessionstorage=function(){try{return!!sessionStorage.getItem}catch(t){return!1}},S.webworkers=function(){return!!o.Worker},S.applicationcache=function(){return!!o.applicationCache},S.svg=function(){return!!p.createElementNS&&!!p.createElementNS(w,"svg").createSVGRect},S.inlinesvg=function(){var t=p.createElement("div");return t.innerHTML="<svg/>",(t.firstChild&&t.firstChild.namespaceURI)==w},S.smil=function(){return!!p.createElementNS&&/SVG/.test(x.call(p.createElementNS(w,"animate")))},S.svgclippaths=function(){return!!p.createElementNS&&/SVG/.test(x.call(p.createElementNS(w,"clipPath")))},S)l(S,D)&&(s=D.toLowerCase(),u[s]=S[D](),k.push((u[s]?"":"no-")+s));return u.input||(u.input=function(t){for(var e=0,r=t.length;e<r;e++)P[t[e]]=t[e]in g;return P}("autocomplete autofocus list placeholder max min multiple pattern required step".split(" ")),u.inputtypes=function(t){for(var e,r,i,n=0,a=t.length;n<a;n++)g.setAttribute("type",r=t[n]),(e="text"!==g.type)&&(g.value=v,g.style.cssText="position:absolute;visibility:hidden;",/^range$/.test(r)&&g.style.WebkitAppearance!==y?(h.appendChild(g),e=(i=p.defaultView).getComputedStyle&&"textfield"!==i.getComputedStyle(g,null).WebkitAppearance&&0!==g.offsetHeight,h.removeChild(g)):/^(search|tel)$/.test(r)||(/^(url|email)$/.test(r)?e=g.checkValidity&&!1===g.checkValidity():/^color$/.test(r)?(h.appendChild(g),h.offsetWidth,e=g.value!=v,h.removeChild(g)):e=g.value!=v)),T[t[n]]=!!e;return T}("search tel url email datetime date month week time datetime-local number range color".split(" "))),r(""),f=g=null,o.attachEvent&&((M=p.createElement("div")).innerHTML="<elem></elem>",1!==M.childNodes.length)&&function(t,n){function e(t){for(var e=-1;++e<l;)t.createElement(s[e])}t.iepp=t.iepp||{};var a,o=t.iepp,r=o.html5elements||"abbr|article|aside|audio|canvas|datalist|details|figcaption|figure|footer|header|hgroup|mark|meter|nav|output|progress|section|summary|time|video",s=r.split("|"),l=s.length,i=new RegExp("(^|\\s)("+r+")","gi"),p=new RegExp("<(/*)("+r+")","gi"),c=/^\s*[\{\}]\s*$/,u=new RegExp("(^|[^\\n]*?\\s)("+r+")([^\\n]*)({[\\n\\w\\W]*?})","gi"),h=n.createDocumentFragment(),d=n.documentElement,f=d.firstChild,m=n.createElement("body"),g=n.createElement("style"),v=/print|all/;o.getCSS=function(t,e){if(t+""===y)return"";for(var r,i=-1,n=t.length,a=[];++i<n;)(r=t[i]).disabled||(e=r.media||e,v.test(e)&&a.push(o.getCSS(r.imports,e),r.cssText),e="all");return a.join("")},o.parseCSS=function(t){for(var e,r=[];null!=(e=u.exec(t));)r.push(((c.exec(e[1])?"\n":e[1])+e[2]+e[3]).replace(i,"$1.iepp_$2")+e[4]);return r.join("\n")},o.writeHTML=function(){var t=-1;for(a=a||n.body;++t<l;)for(var e=n.getElementsByTagName(s[t]),r=e.length,i=-1;++i<r;)e[i].className.indexOf("iepp_")<0&&(e[i].className+=" iepp_"+s[t]);h.appendChild(a),d.appendChild(m),m.className=a.className,m.id=a.id,m.innerHTML=a.innerHTML.replace(p,"<$1font")},o._beforePrint=function(){g.styleSheet.cssText=o.parseCSS(o.getCSS(n.styleSheets,"all")),o.writeHTML()},o.restoreHTML=function(){m.innerHTML="",d.removeChild(m),d.appendChild(a)},o._afterPrint=function(){o.restoreHTML(),g.styleSheet.cssText=""},e(n),e(h),o.disablePP||(f.insertBefore(g,f.firstChild),g.media="print",g.className="iepp-printshim",t.attachEvent("onbeforeprint",o._beforePrint),t.attachEvent("onafterprint",o._afterPrint))}(o,p),u._version="2.0.6",u._prefixes=b,u._domPrefixes=_,u.mq=function(t){return o.matchMedia?matchMedia(t).matches:(a("@media "+t+" { #"+d+" { position: absolute; } }",function(t){e="absolute"==(o.getComputedStyle?getComputedStyle(t,null):t.currentStyle).position}),e);var e},u.hasEvent=I,u.testProp=function(t){return i([t])},u.testAllProps=t,u.testStyles=a,h.className=h.className.replace(/\bno-js\b/,"")+" js "+k.join(" "),u}(this,this.document),f=(t=this).document,i=f.documentElement,g=t.setTimeout,v=f.getElementsByTagName("script")[0],n={}.toString,y=[],x=0,a="MozAppearance"in i.style,b=a&&!!f.createRange().compareNode,_=b?i:v.parentNode,o=t.opera&&"[object Opera]"==n.call(t.opera),l="webkitAppearance"in i.style,w=l&&"async"in f.createElement("script"),S=a?"object":o||w?"img":"script",T=l?"img":S,P=Array.isArray||function(t){return"[object Array]"==n.call(t)},k=function(t){return"string"==typeof t},I=[],A={},(m=function(t){function l(t,e,r,i,n){var a=function(t){var e,r,i=t.split("!"),n=I.length,a=i.pop(),o=i.length,s={url:a,origUrl:a,prefixes:i};for(r=0;r<o;r++)(e=A[i[r]])&&(s=e(s));for(r=0;r<n;r++)s=I[r](s);return s}(t),o=a.autoCallback;if(!a.bypass){if(e&&(e=d(e)?e:e[t]||e[i]||e[t.split("/").pop().split("?")[0]]),a.instead)return a.instead(t,e,r,i,n);r.load(a.url,a.forceCSS||!a.forceJS&&/css$/.test(a.url)?"c":void 0,a.noexec),(d(e)||d(o))&&r.load(function(){s(),e&&e(a.origUrl,n,i),o&&o(a.origUrl,n,i)})}}function e(t,e){function r(t){if(k(t))l(t,s,e,0,n);else if(p(t))for(i in t)t.hasOwnProperty(i)&&l(t[i],s,e,i,n)}var i,n=!!t.test,a=n?t.yep:t.nope,o=t.load||t.both,s=t.callback;r(a),r(o),t.complete&&e.load(t.complete)}var r,i,n=this.yepnope.loader;if(k(t))l(t,0,n,0);else if(P(t))for(r=0;r<t.length;r++)i=t[r],k(i)?l(i,0,n,0):P(i)?m(i):p(i)&&e(i,n);else p(t)&&e(t,n)}).addPrefix=function(t,e){A[t]=e},m.addFilter=function(t){I.push(t)},m.errorTimeout=1e4,null==f.readyState&&f.addEventListener&&(f.readyState="loading",f.addEventListener("DOMContentLoaded",r=function(){f.removeEventListener("DOMContentLoaded",r,0),f.readyState="complete"},0)),t.yepnope=s(),Modernizr.load=function(){yepnope.apply(window,[].slice.call(arguments,0))},E.exports=window.Modernizr}).call(window)},function(zt,Gt,Yt){var Xt;function Wt(t){return(Wt="function"==typeof Symbol&&"symbol"==typeof Symbol.iterator?function(t){return typeof t}:function(t){return t&&"function"==typeof Symbol&&t.constructor===Symbol&&t!==Symbol.prototype?"symbol":typeof t})(t)}!function(a,n,t,u){"use strict";var o,s=["","webkit","Moz","MS","ms","o"],e=n.createElement("div"),r="function",l=Math.round,h=Math.abs,d=Date.now;function p(t,e,r){return setTimeout(y(t,r),e)}function i(t,e,r){return!!Array.isArray(t)&&(c(t,r[e],r),!0)}function c(t,e,r){var i;if(t)if(t.forEach)t.forEach(e,r);else if(t.length!==u)for(i=0;i<t.length;)e.call(r,t[i],i,t),i++;else for(i in t)t.hasOwnProperty(i)&&e.call(r,t[i],i,t)}function f(i,t,e){var n="DEPRECATED METHOD: "+t+"\n"+e+" AT \n";return function(){var t=new Error("get-stack-trace"),e=t&&t.stack?t.stack.replace(/^[^\(]+?[\n$]/gm,"").replace(/^\s+at\s+/gm,"").replace(/^Object.<anonymous>\s*\(/gm,"{anonymous}()@"):"Unknown Stack Trace",r=a.console&&(a.console.warn||a.console.log);return r&&r.call(a.console,n,e),i.apply(this,arguments)}}o="function"!=typeof Object.assign?function(t){if(t===u||null===t)throw new TypeError("Cannot convert undefined or null to object");for(var e=Object(t),r=1;r<arguments.length;r++){var i=arguments[r];if(i!==u&&null!==i)for(var n in i)i.hasOwnProperty(n)&&(e[n]=i[n])}return e}:Object.assign;var m=f(function(t,e,r){for(var i=Object.keys(e),n=0;n<i.length;)(!r||r&&t[i[n]]===u)&&(t[i[n]]=e[i[n]]),n++;return t},"extend","Use `assign`."),g=f(function(t,e){return m(t,e,!0)},"merge","Use `assign`.");function v(t,e,r){var i,n=e.prototype;(i=t.prototype=Object.create(n)).constructor=t,i._super=n,r&&o(i,r)}function y(t,e){return function(){return t.apply(e,arguments)}}function x(t,e){return Wt(t)==r?t.apply(e&&e[0]||u,e):t}function b(t,e){return t===u?e:t}function _(e,t,r){c(P(t),function(t){e.addEventListener(t,r,!1)})}function w(e,t,r){c(P(t),function(t){e.removeEventListener(t,r,!1)})}function S(t,e){for(;t;){if(t==e)return!0;t=t.parentNode}return!1}function T(t,e){return-1<t.indexOf(e)}function P(t){return t.trim().split(/\s+/g)}function k(t,e,r){if(t.indexOf&&!r)return t.indexOf(e);for(var i=0;i<t.length;){if(r&&t[i][r]==e||!r&&t[i]===e)return i;i++}return-1}function I(t){return Array.prototype.slice.call(t,0)}function A(t,r,e){for(var i=[],n=[],a=0;a<t.length;){var o=r?t[a][r]:t[a];k(n,o)<0&&i.push(t[a]),n[a]=o,a++}return e&&(i=r?i.sort(function(t,e){return t[r]>e[r]}):i.sort()),i}function E(t,e){for(var r,i,n=e[0].toUpperCase()+e.slice(1),a=0;a<s.length;){if((i=(r=s[a])?r+n:e)in t)return i;a++}return u}var C=1;function O(t){var e=t.ownerDocument||t;return e.defaultView||e.parentWindow||a}var M="ontouchstart"in a,D=!1,F=M&&/mobile|tablet|ip(ad|hone|od)|android/i.test(navigator.userAgent),R="touch",N=25,L=1,B=4,j=8,$=1,V=2,H=4,z=8,G=16,Y=V|H,X=z|G,W=Y|X,U=["x","y"],q=["clientX","clientY"];function J(e,t){var r=this;this.manager=e,this.callback=t,this.element=e.element,this.target=e.options.inputTarget,this.domHandler=function(t){x(e.options.enable,[e])&&r.handler(t)},this.init()}function Q(t,e,r){var i=r.pointers.length,n=r.changedPointers.length,a=e&L&&i-n==0,o=e&(B|j)&&i-n==0;r.isFirst=!!a,r.isFinal=!!o,a&&(t.session={}),r.eventType=e,function(t,e){var r=t.session,i=e.pointers,n=i.length;r.firstInput||(r.firstInput=Z(e));1<n&&!r.firstMultiple?r.firstMultiple=Z(e):1===n&&(r.firstMultiple=!1);var a=r.firstInput,o=r.firstMultiple,s=o?o.center:a.center,l=e.center=K(i);e.timeStamp=d(),e.deltaTime=e.timeStamp-a.timeStamp,e.angle=it(s,l),e.distance=rt(s,l),function(t,e){var r=e.center,i=t.offsetDelta||{},n=t.prevDelta||{},a=t.prevInput||{};e.eventType!==L&&a.eventType!==B||(n=t.prevDelta={x:a.deltaX||0,y:a.deltaY||0},i=t.offsetDelta={x:r.x,y:r.y});e.deltaX=n.x+(r.x-i.x),e.deltaY=n.y+(r.y-i.y)}(r,e),e.offsetDirection=et(e.deltaX,e.deltaY);var p=tt(e.deltaTime,e.deltaX,e.deltaY);e.overallVelocityX=p.x,e.overallVelocityY=p.y,e.overallVelocity=h(p.x)>h(p.y)?p.x:p.y,e.scale=o?function(t,e){return rt(e[0],e[1],q)/rt(t[0],t[1],q)}(o.pointers,i):1,e.rotation=o?function(t,e){return it(e[1],e[0],q)+it(t[1],t[0],q)}(o.pointers,i):0,e.maxPointers=r.prevInput?e.pointers.length>r.prevInput.maxPointers?e.pointers.length:r.prevInput.maxPointers:e.pointers.length,function(t,e){var r,i,n,a,o=t.lastInterval||e,s=e.timeStamp-o.timeStamp;if(e.eventType!=j&&(N<s||o.velocity===u)){var l=e.deltaX-o.deltaX,p=e.deltaY-o.deltaY,c=tt(s,l,p);i=c.x,n=c.y,r=h(c.x)>h(c.y)?c.x:c.y,a=et(l,p),t.lastInterval=e}else r=o.velocity,i=o.velocityX,n=o.velocityY,a=o.direction;e.velocity=r,e.velocityX=i,e.velocityY=n,e.direction=a}(r,e);var c=t.element;S(e.srcEvent.target,c)&&(c=e.srcEvent.target);e.target=c}(t,r),t.emit("hammer.input",r),t.recognize(r),t.session.prevInput=r}function Z(t){for(var e=[],r=0;r<t.pointers.length;)e[r]={clientX:l(t.pointers[r].clientX),clientY:l(t.pointers[r].clientY)},r++;return{timeStamp:d(),pointers:e,center:K(e),deltaX:t.deltaX,deltaY:t.deltaY}}function K(t){var e=t.length;if(1===e)return{x:l(t[0].clientX),y:l(t[0].clientY)};for(var r=0,i=0,n=0;n<e;)r+=t[n].clientX,i+=t[n].clientY,n++;return{x:l(r/e),y:l(i/e)}}function tt(t,e,r){return{x:e/t||0,y:r/t||0}}function et(t,e){return t===e?$:h(t)>=h(e)?t<0?V:H:e<0?z:G}function rt(t,e,r){r||(r=U);var i=e[r[0]]-t[r[0]],n=e[r[1]]-t[r[1]];return Math.sqrt(i*i+n*n)}function it(t,e,r){r||(r=U);var i=e[r[0]]-t[r[0]],n=e[r[1]]-t[r[1]];return 180*Math.atan2(n,i)/Math.PI}J.prototype={handler:function(){},init:function(){this.evEl&&_(this.element,this.evEl,this.domHandler),this.evTarget&&_(this.target,this.evTarget,this.domHandler),this.evWin&&_(O(this.element),this.evWin,this.domHandler)},destroy:function(){this.evEl&&w(this.element,this.evEl,this.domHandler),this.evTarget&&w(this.target,this.evTarget,this.domHandler),this.evWin&&w(O(this.element),this.evWin,this.domHandler)}};var nt={mousedown:L,mousemove:2,mouseup:B},at="mousedown",ot="mousemove mouseup";function st(){this.evEl=at,this.evWin=ot,this.pressed=!1,J.apply(this,arguments)}v(st,J,{handler:function(t){var e=nt[t.type];e&L&&0===t.button&&(this.pressed=!0),2&e&&1!==t.which&&(e=B),this.pressed&&(e&B&&(this.pressed=!1),this.callback(this.manager,e,{pointers:[t],changedPointers:[t],pointerType:"mouse",srcEvent:t}))}});var lt={pointerdown:L,pointermove:2,pointerup:B,pointercancel:j,pointerout:j},pt={2:R,3:"pen",4:"mouse",5:"kinect"},ct="pointerdown",ut="pointermove pointerup pointercancel";function ht(){this.evEl=ct,this.evWin=ut,J.apply(this,arguments),this.store=this.manager.session.pointerEvents=[]}a.MSPointerEvent&&!a.PointerEvent&&(ct="MSPointerDown",ut="MSPointerMove MSPointerUp MSPointerCancel"),v(ht,J,{handler:function(t){var e=this.store,r=!1,i=t.type.toLowerCase().replace("ms",""),n=lt[i],a=pt[t.pointerType]||t.pointerType,o=a==R,s=k(e,t.pointerId,"pointerId");n&L&&(0===t.button||o)?s<0&&(e.push(t),s=e.length-1):n&(B|j)&&(r=!0),s<0||(e[s]=t,this.callback(this.manager,n,{pointers:e,changedPointers:[t],pointerType:a,srcEvent:t}),r&&e.splice(s,1))}});var dt={touchstart:L,touchmove:2,touchend:B,touchcancel:j};function ft(){this.evTarget="touchstart",this.evWin="touchstart touchmove touchend touchcancel",this.started=!1,J.apply(this,arguments)}v(ft,J,{handler:function(t){var e=dt[t.type];if(e===L&&(this.started=!0),this.started){var r=function(t,e){var r=I(t.touches),i=I(t.changedTouches);e&(B|j)&&(r=A(r.concat(i),"identifier",!0));return[r,i]}.call(this,t,e);e&(B|j)&&r[0].length-r[1].length==0&&(this.started=!1),this.callback(this.manager,e,{pointers:r[0],changedPointers:r[1],pointerType:R,srcEvent:t})}}});var mt={touchstart:L,touchmove:2,touchend:B,touchcancel:j},gt="touchstart touchmove touchend touchcancel";function vt(){this.evTarget=gt,this.targetIds={},J.apply(this,arguments)}v(vt,J,{handler:function(t){var e=mt[t.type],r=function(t,e){var r=I(t.touches),i=this.targetIds;if(e&(2|L)&&1===r.length)return i[r[0].identifier]=!0,[r,r];var n,a,o=I(t.changedTouches),s=[],l=this.target;if(a=r.filter(function(t){return S(t.target,l)}),e===L)for(n=0;n<a.length;)i[a[n].identifier]=!0,n++;n=0;for(;n<o.length;)i[o[n].identifier]&&s.push(o[n]),e&(B|j)&&delete i[o[n].identifier],n++;return s.length?[A(a.concat(s),"identifier",!0),s]:void 0}.call(this,t,e);r&&this.callback(this.manager,e,{pointers:r[0],changedPointers:r[1],pointerType:R,srcEvent:t})}});var yt=2500;function xt(){J.apply(this,arguments);var t=y(this.handler,this);this.touch=new vt(this.manager,t),this.mouse=new st(this.manager,t),this.primaryTouch=null,this.lastTouches=[]}function bt(t){var e=t.changedPointers[0];if(e.identifier===this.primaryTouch){var r={x:e.clientX,y:e.clientY};this.lastTouches.push(r);var i=this.lastTouches;setTimeout(function(){var t=i.indexOf(r);-1<t&&i.splice(t,1)},yt)}}v(xt,J,{handler:function(t,e,r){var i=r.pointerType==R,n="mouse"==r.pointerType;if(!(n&&r.sourceCapabilities&&r.sourceCapabilities.firesTouchEvents)){if(i)(function(t,e){t&L?(this.primaryTouch=e.changedPointers[0].identifier,bt.call(this,e)):t&(B|j)&&bt.call(this,e)}).call(this,e,r);else if(n&&function(t){for(var e=t.srcEvent.clientX,r=t.srcEvent.clientY,i=0;i<this.lastTouches.length;i++){var n=this.lastTouches[i],a=Math.abs(e-n.x),o=Math.abs(r-n.y);if(a<=25&&o<=25)return!0}return!1}.call(this,r))return;this.callback(t,e,r)}},destroy:function(){this.touch.destroy(),this.mouse.destroy()}});var _t=E(e.style,"touchAction"),wt=_t!==u,St="manipulation",Tt="none",Pt="pan-x",kt="pan-y";function It(t,e){this.manager=t,this.set(e)}It.prototype={set:function(t){"compute"==t&&(t=this.compute()),wt&&this.manager.element.style&&(this.manager.element.style[_t]=t),this.actions=t.toLowerCase().trim()},update:function(){this.set(this.manager.options.touchAction)},compute:function(){var e=[];return c(this.manager.recognizers,function(t){x(t.options.enable,[t])&&(e=e.concat(t.getTouchAction()))}),function(t){if(T(t,Tt))return Tt;var e=T(t,Pt),r=T(t,kt);if(e&&r)return Tt;if(e||r)return e?Pt:kt;if(T(t,St))return St;return"auto"}(e.join(" "))},preventDefaults:function(t){if(!wt){var e=t.srcEvent,r=t.offsetDirection;if(this.manager.session.prevented)e.preventDefault();else{var i=this.actions,n=T(i,Tt),a=T(i,kt),o=T(i,Pt);if(n){var s=1===t.pointers.length,l=t.distance<2,p=t.deltaTime<250;if(s&&l&&p)return}if(!o||!a)return n||a&&r&Y||o&&r&X?this.preventSrc(e):void 0}}},preventSrc:function(t){this.manager.session.prevented=!0,t.preventDefault()}};var At=1;function Et(t){this.options=o({},this.defaults,t||{}),this.id=C++,this.manager=null,this.options.enable=b(this.options.enable,!0),this.state=At,this.simultaneous={},this.requireFail=[]}function Ct(t){return 16&t?"cancel":8&t?"end":4&t?"move":2&t?"start":""}function Ot(t){return t==G?"down":t==z?"up":t==V?"left":t==H?"right":""}function Mt(t,e){var r=e.manager;return r?r.get(t):t}function Dt(){Et.apply(this,arguments)}function Ft(){Dt.apply(this,arguments),this.pX=null,this.pY=null}function Rt(){Dt.apply(this,arguments)}function Nt(){Et.apply(this,arguments),this._timer=null,this._input=null}function Lt(){Dt.apply(this,arguments)}function Bt(){Dt.apply(this,arguments)}function jt(){Et.apply(this,arguments),this.pTime=!1,this.pCenter=!1,this._timer=null,this._input=null,this.count=0}function $t(t,e){return(e=e||{}).recognizers=b(e.recognizers,$t.defaults.preset),new Vt(t,e)}Et.prototype={defaults:{},set:function(t){return o(this.options,t),this.manager&&this.manager.touchAction.update(),this},recognizeWith:function(t){if(i(t,"recognizeWith",this))return this;var e=this.simultaneous;return e[(t=Mt(t,this)).id]||(e[t.id]=t).recognizeWith(this),this},dropRecognizeWith:function(t){return i(t,"dropRecognizeWith",this)||(t=Mt(t,this),delete this.simultaneous[t.id]),this},requireFailure:function(t){if(i(t,"requireFailure",this))return this;var e=this.requireFail;return-1===k(e,t=Mt(t,this))&&(e.push(t),t.requireFailure(this)),this},dropRequireFailure:function(t){if(i(t,"dropRequireFailure",this))return this;t=Mt(t,this);var e=k(this.requireFail,t);return-1<e&&this.requireFail.splice(e,1),this},hasRequireFailures:function(){return 0<this.requireFail.length},canRecognizeWith:function(t){return!!this.simultaneous[t.id]},emit:function(e){var r=this,t=this.state;function i(t){r.manager.emit(t,e)}t<8&&i(r.options.event+Ct(t)),i(r.options.event),e.additionalEvent&&i(e.additionalEvent),8<=t&&i(r.options.event+Ct(t))},tryEmit:function(t){if(this.canEmit())return this.emit(t);this.state=32},canEmit:function(){for(var t=0;t<this.requireFail.length;){if(!(this.requireFail[t].state&(32|At)))return!1;t++}return!0},recognize:function(t){var e=o({},t);if(!x(this.options.enable,[this,e]))return this.reset(),void(this.state=32);56&this.state&&(this.state=At),this.state=this.process(e),30&this.state&&this.tryEmit(e)},process:function(t){},getTouchAction:function(){},reset:function(){}},v(Dt,Et,{defaults:{pointers:1},attrTest:function(t){var e=this.options.pointers;return 0===e||t.pointers.length===e},process:function(t){var e=this.state,r=t.eventType,i=6&e,n=this.attrTest(t);return i&&(r&j||!n)?16|e:i||n?r&B?8|e:2&e?4|e:2:32}}),v(Ft,Dt,{defaults:{event:"pan",threshold:10,pointers:1,direction:W},getTouchAction:function(){var t=this.options.direction,e=[];return t&Y&&e.push(kt),t&X&&e.push(Pt),e},directionTest:function(t){var e=this.options,r=!0,i=t.distance,n=t.direction,a=t.deltaX,o=t.deltaY;return n&e.direction||(i=e.direction&Y?(n=0===a?$:a<0?V:H,r=a!=this.pX,Math.abs(t.deltaX)):(n=0===o?$:o<0?z:G,r=o!=this.pY,Math.abs(t.deltaY))),t.direction=n,r&&i>e.threshold&&n&e.direction},attrTest:function(t){return Dt.prototype.attrTest.call(this,t)&&(2&this.state||!(2&this.state)&&this.directionTest(t))},emit:function(t){this.pX=t.deltaX,this.pY=t.deltaY;var e=Ot(t.direction);e&&(t.additionalEvent=this.options.event+e),this._super.emit.call(this,t)}}),v(Rt,Dt,{defaults:{event:"pinch",threshold:0,pointers:2},getTouchAction:function(){return[Tt]},attrTest:function(t){return this._super.attrTest.call(this,t)&&(Math.abs(t.scale-1)>this.options.threshold||2&this.state)},emit:function(t){if(1!==t.scale){var e=t.scale<1?"in":"out";t.additionalEvent=this.options.event+e}this._super.emit.call(this,t)}}),v(Nt,Et,{defaults:{event:"press",pointers:1,time:251,threshold:9},getTouchAction:function(){return["auto"]},process:function(t){var e=this.options,r=t.pointers.length===e.pointers,i=t.distance<e.threshold,n=t.deltaTime>e.time;if(this._input=t,!i||!r||t.eventType&(B|j)&&!n)this.reset();else if(t.eventType&L)this.reset(),this._timer=p(function(){this.state=8,this.tryEmit()},e.time,this);else if(t.eventType&B)return 8;return 32},reset:function(){clearTimeout(this._timer)},emit:function(t){8===this.state&&(t&&t.eventType&B?this.manager.emit(this.options.event+"up",t):(this._input.timeStamp=d(),this.manager.emit(this.options.event,this._input)))}}),v(Lt,Dt,{defaults:{event:"rotate",threshold:0,pointers:2},getTouchAction:function(){return[Tt]},attrTest:function(t){return this._super.attrTest.call(this,t)&&(Math.abs(t.rotation)>this.options.threshold||2&this.state)}}),v(Bt,Dt,{defaults:{event:"swipe",threshold:10,velocity:.3,direction:Y|X,pointers:1},getTouchAction:function(){return Ft.prototype.getTouchAction.call(this)},attrTest:function(t){var e,r=this.options.direction;return r&(Y|X)?e=t.overallVelocity:r&Y?e=t.overallVelocityX:r&X&&(e=t.overallVelocityY),this._super.attrTest.call(this,t)&&r&t.offsetDirection&&t.distance>this.options.threshold&&t.maxPointers==this.options.pointers&&h(e)>this.options.velocity&&t.eventType&B},emit:function(t){var e=Ot(t.offsetDirection);e&&this.manager.emit(this.options.event+e,t),this.manager.emit(this.options.event,t)}}),v(jt,Et,{defaults:{event:"tap",pointers:1,taps:1,interval:300,time:250,threshold:9,posThreshold:10},getTouchAction:function(){return[St]},process:function(t){var e=this.options,r=t.pointers.length===e.pointers,i=t.distance<e.threshold,n=t.deltaTime<e.time;if(this.reset(),t.eventType&L&&0===this.count)return this.failTimeout();if(i&&n&&r){if(t.eventType!=B)return this.failTimeout();var a=!this.pTime||t.timeStamp-this.pTime<e.interval,o=!this.pCenter||rt(this.pCenter,t.center)<e.posThreshold;if(this.pTime=t.timeStamp,this.pCenter=t.center,o&&a?this.count+=1:this.count=1,this._input=t,0==this.count%e.taps)return this.hasRequireFailures()?(this._timer=p(function(){this.state=8,this.tryEmit()},e.interval,this),2):8}return 32},failTimeout:function(){return this._timer=p(function(){this.state=32},this.options.interval,this),32},reset:function(){clearTimeout(this._timer)},emit:function(){8==this.state&&(this._input.tapCount=this.count,this.manager.emit(this.options.event,this._input))}}),$t.VERSION="2.0.7",$t.defaults={domEvents:!1,touchAction:"compute",enable:!0,inputTarget:null,inputClass:null,preset:[[Lt,{enable:!1}],[Rt,{enable:!1},["rotate"]],[Bt,{direction:Y}],[Ft,{direction:Y},["swipe"]],[jt],[jt,{event:"doubletap",taps:2},["tap"]],[Nt]],cssProps:{userSelect:"none",touchSelect:"none",touchCallout:"none",contentZooming:"none",userDrag:"none",tapHighlightColor:"rgba(0,0,0,0)"}};function Vt(t,e){this.options=o({},$t.defaults,e||{}),this.options.inputTarget=this.options.inputTarget||t,this.handlers={},this.session={},this.recognizers=[],this.element=t,this.input=function(t){var e=t.options.inputClass;return new(e||(D?ht:F?vt:M?xt:st))(t,Q)}(this),this.touchAction=new It(this,this.options.touchAction),Ht(this,!0),c(this.options.recognizers,function(t){var e=this.add(new t[0](t[1]));t[2]&&e.recognizeWith(t[2]),t[3]&&e.requireFailure(t[3])},this)}function Ht(t,r){var i=t.element;i.style&&c(t.options.cssProps,function(t,e){i.style[E(i.style,e)]=r?t:""})}Vt.prototype={set:function(t){return o(this.options,t),t.touchAction&&this.touchAction.update(),t.inputTarget&&(this.input.destroy(),this.input.target=t.inputTarget,this.input.init()),this},stop:function(t){this.session.stopped=t?2:1},recognize:function(t){var e=this.session;if(!e.stopped){var r;this.touchAction.preventDefaults(t);var i=this.recognizers,n=e.curRecognizer;(!n||n&&8&n.state)&&(n=e.curRecognizer=null);for(var a=0;a<i.length;)r=i[a],2===e.stopped||n&&r!=n&&!r.canRecognizeWith(n)?r.reset():r.recognize(t),!n&&14&r.state&&(n=e.curRecognizer=r),a++}},get:function(t){if(t instanceof Et)return t;for(var e=this.recognizers,r=0;r<e.length;r++)if(e[r].options.event==t)return e[r];return null},add:function(t){if(i(t,"add",this))return this;var e=this.get(t.options.event);return e&&this.remove(e),this.recognizers.push(t),(t.manager=this).touchAction.update(),t},remove:function(t){if(i(t,"remove",this))return this;if(t=this.get(t)){var e=this.recognizers,r=k(e,t);-1!==r&&(e.splice(r,1),this.touchAction.update())}return this},on:function(t,e){var r=this.handlers;return c(P(t),function(t){r[t]=r[t]||[],r[t].push(e)}),this},off:function(t,e){var r=this.handlers;return c(P(t),function(t){e?r[t]&&r[t].splice(k(r[t],e),1):delete r[t]}),this},emit:function(t,e){this.options.domEvents&&function(t,e){var r=n.createEvent("Event");r.initEvent(t,!0,!0),(r.gesture=e).target.dispatchEvent(r)}(t,e);var r=this.handlers[t]&&this.handlers[t].slice();if(r&&r.length){e.type=t,e.preventDefault=function(){e.srcEvent.preventDefault()};for(var i=0;i<r.length;)r[i](e),i++}},destroy:function(){this.element&&Ht(this,!1),this.handlers={},this.session={},this.input.destroy(),this.element=null}},o($t,{INPUT_START:L,INPUT_MOVE:2,INPUT_END:B,INPUT_CANCEL:j,STATE_POSSIBLE:At,STATE_BEGAN:2,STATE_CHANGED:4,STATE_ENDED:8,STATE_RECOGNIZED:8,STATE_CANCELLED:16,STATE_FAILED:32,DIRECTION_NONE:$,DIRECTION_LEFT:V,DIRECTION_RIGHT:H,DIRECTION_UP:z,DIRECTION_DOWN:G,DIRECTION_HORIZONTAL:Y,DIRECTION_VERTICAL:X,DIRECTION_ALL:W,Manager:Vt,Input:J,TouchAction:It,TouchInput:vt,MouseInput:st,PointerEventInput:ht,TouchMouseInput:xt,SingleTouchInput:ft,Recognizer:Et,AttrRecognizer:Dt,Tap:jt,Pan:Ft,Swipe:Bt,Pinch:Rt,Rotate:Lt,Press:Nt,on:_,off:w,each:c,merge:g,extend:m,assign:o,inherit:v,bindFn:y,prefixed:E}),(void 0!==a?a:"undefined"!=typeof self?self:{}).Hammer=$t,(Xt=function(){return $t}.call(Gt,Yt,Gt,zt))===u||(zt.exports=Xt)}(window,document)},function(t,e,r){var i,n,a;n=[],void 0===(a="function"==typeof(i=function(){var c=null;return function i(n,t){var a=t||{preventDefault:!1};if(n.Manager){var o=n,e=function(t,e){var r=Object.create(a);return e&&o.assign(r,e),i(new o(t,r),r)};return o.assign(e,o),e.Manager=function(t,e){var r=Object.create(a);return e&&o.assign(r,e),i(new o.Manager(t,r),r)},e}var s=Object.create(n),r=n.element;function l(t){return t.match(/[^ ]+/g)}function p(t){if("hammer.input"!==t.type){if(t.srcEvent._handled||(t.srcEvent._handled={}),t.srcEvent._handled[t.type])return;t.srcEvent._handled[t.type]=!0}var e=!1;t.stopPropagation=function(){e=!0};var r=t.srcEvent.stopPropagation.bind(t.srcEvent);"function"==typeof r&&(t.srcEvent.stopPropagation=function(){r(),t.stopPropagation()});for(var i=t.firstTarget=c;i&&!e;){var n=i.hammer;if(n)for(var a,o=0;o<n.length;o++)if(a=n[o]._handlers[t.type])for(var s=0;s<a.length&&!e;s++)a[s](t);i=i.parentNode}}return r.hammer||(r.hammer=[]),r.hammer.push(s),n.on("hammer.input",function(t){!0!==a.preventDefault&&a.preventDefault!==t.pointerType||t.preventDefault(),t.isFirst&&(c=t.target)}),s._handlers={},s.on=function(t,r){return l(t).forEach(function(t){var e=s._handlers[t];e||(s._handlers[t]=e=[],n.on(t,p)),e.push(r)}),s},s.off=function(t,r){return l(t).forEach(function(t){var e=s._handlers[t];e&&(0<(e=r?e.filter(function(t){return t!==r}):[]).length?s._handlers[t]=e:(n.off(t,p),delete s._handlers[t]))}),s},s.emit=function(t,e){c=e.target,n.emit(t,e)},s.destroy=function(){var t=n.element.hammer,e=t.indexOf(s);-1!==e&&t.splice(e,1),t.length||delete n.element.hammer,s._handlers={},n.destroy()},s}})?i.apply(e,n):i)||(t.exports=a)},function(t,e){var i,n,r;i=jQuery,n=Hammer,i.fn.hammer=function(t){return this.each(function(){!function(t,e){var r=i(t);r.data("hammer")||r.data("hammer",new n(r[0],e))}(this,t)})},n.Manager.prototype.emit=(r=n.Manager.prototype.emit,function(t,e){r.call(this,t,e),jQuery(this.element).triggerHandler({type:t,gesture:e})})},function(t,e,r){function l(t){return(l="function"==typeof Symbol&&"symbol"==typeof Symbol.iterator?function(t){return typeof t}:function(t){return t&&"function"==typeof Symbol&&t.constructor===Symbol&&t!==Symbol.prototype?"symbol":typeof t})(t)}
/*!
 * Waves v0.5.5
 * http://fian.my.id/Waves 
 * 
 * Copyright 2014 Alfiana E. Sibuea and other contributors 
 * Released under the MIT license 
 * https://github.com/fians/Waves/blob/master/LICENSE 
 */!function(r){"use strict";var t=t||{};var s={duration:500,show:function(t){if(2===t.button)return!1;var e=$(this).find(".waves-ripple.waves-ripple-ripple").get(0),r=$(this).find(".waves-ripple.waves-ripple-static").get(0),i=function(t){var e,r,i={top:0,left:0},n=t&&t.ownerDocument;return e=n.documentElement,"undefined"!==l(t.getBoundingClientRect)&&(i=t.getBoundingClientRect()),r=function(t){return function(t){return null!==t&&t===t.window}(t)?t:9===t.nodeType&&t.defaultView}(n),{top:i.top+r.pageYOffset-e.clientTop,left:i.left+r.pageXOffset-e.clientLeft}}(this),n=t.pageY-i.top,a=t.pageX-i.left;"touches"in t&&(n=t.touches[0].pageY-i.top,a=t.touches[0].pageX-i.left);var o=Math.max(this.clientWidth,this.clientHeight)/100*3;(new TimelineMax).set(e,{scale:0,top:n,left:a,opacity:0}).to(e,s.duration/1e3,{scale:o,opacity:1,force3D:prx.greensockForce3D}),(new TimelineMax).set(r,{scale:o,opacity:0}).to(r,.2,{opacity:.3,force3D:prx.greensockForce3D})},hide:function(){var t=$(this).find(".waves-ripple.waves-ripple-ripple").get(0),e=$(this).find(".waves-ripple.waves-ripple-static").get(0);if(!t)return!1;(new TimelineMax).to(t,.2,{opacity:0,delay:.2,force3D:prx.greensockForce3D}),(new TimelineMax).to(e,.2,{opacity:0,delay:.2,force3D:prx.greensockForce3D})},wrapInput:function(t){for(var e=0;e<t.length;e++){var r=t[e];if("input"===r.tagName.toLowerCase()){var i=r.parentNode;if("i"===i.tagName.toLowerCase()&&-1!==i.className.indexOf("waves-effect"))return!1;var n=document.createElement("i");n.className=r.className+" waves-input-wrapper";var a=r.getAttribute("style");a||(a=""),n.setAttribute("style",a),r.className="waves-button-input",r.removeAttribute("style"),i.replaceChild(n,r),n.appendChild(r)}}},create:function(t){for(var e=0;e<t.length;e++){var r=$(t[e]).find(".prx-material-ripple-roundoverflow").get(0)||t[e],i=document.createElement("div");i.className="waves-ripple waves-ripple-ripple",r.appendChild(i);var n=document.createElement("div");n.className="waves-ripple waves-ripple-static waves-notransition",r.appendChild(n)}}};t.displayEffect=function(t){"duration"in(t=t||{})&&(s.duration=t.duration);var e=void 0!==t.itemid?t.itemid:".waves-effect";s.wrapInput($(e).get()),s.create($(e).get()),Array.prototype.forEach.call($(e).get(),function(t){"ontouchstart"in r?(t.addEventListener("touchstart",s.show,!1),t.addEventListener("touchend",s.hide,!1),t.addEventListener("touchcancel",s.hide,!1)):(t.addEventListener("mousedown",s.show,!1),t.addEventListener("mouseup",s.hide,!1),t.addEventListener("mouseleave",s.hide,!1))})},r.Waves=t}(window)},function(t,e,s){var l;!function n(a,o,s){function l(r,t){if(!o[r]){if(!a[r]){if(p)return p(r,!0);var e=new Error("Cannot find module '"+r+"'");throw e.code="MODULE_NOT_FOUND",e}var i=o[r]={exports:{}};a[r][0].call(i.exports,function(t){var e=a[r][1][t];return l(e||t)},i,i.exports,n,a,o,s)}return o[r].exports}for(var p=!1,t=0;t<s.length;t++)l(s[t]);return l}({1:[function(t,e,r){var i=t("cssfilter").FilterCSS,c=t("./util"),n=new i;function a(t){return t.replace(o,"&lt;").replace(s,"&gt;")}var o=/</g,s=/>/g,l=/"/g,p=/&quot;/g,u=/&#([a-zA-Z0-9]*);?/gim,h=/&colon;?/gim,d=/&newline;?/gim,f=/((j\s*a\s*v\s*a|v\s*b|l\s*i\s*v\s*e)\s*s\s*c\s*r\s*i\s*p\s*t\s*|m\s*o\s*c\s*h\s*a)\:/gi,m=/e\s*x\s*p\s*r\s*e\s*s\s*s\s*i\s*o\s*n\s*\(.*/gi,g=/u\s*r\s*l\s*\(.*/gi;function v(t){return t.replace(l,"&quot;")}function y(t){return t.replace(p,'"')}function x(t){return t.replace(u,function(t,e){return"x"===e[0]||"X"===e[0]?String.fromCharCode(parseInt(e.substr(1),16)):String.fromCharCode(parseInt(e,10))})}function b(t){return t.replace(h,":").replace(d," ")}function _(t){for(var e="",r=0,i=t.length;r<i;r++)e+=t.charCodeAt(r)<32?" ":t.charAt(r);return c.trim(e)}function w(t){return t=_(t=b(t=x(t=y(t))))}function S(t){return t=a(t=v(t))}var T=/<!--[\s\S]*?-->/g;r.whiteList={a:["target","href","title","style"],abbr:["title"],address:["style"],area:["shape","coords","href","alt","style"],article:["style"],aside:["style"],audio:["autoplay","controls","loop","preload","src"],b:[],bdi:["dir"],bdo:["dir"],big:[],blockquote:["cite","style"],br:[],caption:["style"],center:[],cite:[],code:[],col:["align","valign","span","width","style"],colgroup:["align","valign","span","width","style"],dd:[],del:["datetime"],details:["open"],div:["style"],dl:[],dt:[],em:[],font:["color","size","face","style","family"],footer:["style"],h1:["style"],h2:["style"],h3:["style"],h4:["style"],h5:["style"],h6:["style"],header:["style"],hr:["style"],i:[],img:["src","alt","title","width","height"],ins:["datetime"],li:["style"],mark:[],nav:["style"],ol:[],p:["style"],pre:["style"],s:[],section:["style"],small:[],span:["style"],sub:[],sup:[],strong:[],strike:[],table:["width","border","align","valign","style"],tbody:["align","valign","style"],td:["width","colspan","align","valign","style","style"],tfoot:["align","valign","style"],th:["width","colspan","align","valign","style"],thead:["align","valign","style"],tr:["rowspan","align","valign","style"],tt:[],u:[],ul:["style"],video:["autoplay","controls","loop","preload","src","height","width","style"]},r.onTag=function(t,e,r){},r.onIgnoreTag=function(t,e,r){},r.onTagAttr=function(t,e,r){},r.onIgnoreTagAttr=function(t,e,r){},r.safeAttrValue=function(t,e,r,i){if(i=i||n,r=w(r),"href"===e||"src"===e){if("#"===(r=c.trim(r)))return"#";if("http://"!==r.substr(0,7)&&"https://"!==r.substr(0,8)&&"mailto:"!==r.substr(0,7)&&"/"!==r[0])return""}else if("background"===e){if(f.lastIndex=0,f.test(r))return""}else if("style"===e){if(m.lastIndex=0,m.test(r))return"";if(g.lastIndex=0,g.test(r)&&(f.lastIndex=0,f.test(r)))return"";r=i.process(r)}return r=S(r)},r.escapeHtml=a,r.escapeQuote=v,r.unescapeQuote=y,r.escapeHtmlEntities=x,r.escapeDangerHtml5Entities=b,r.clearNonPrintableCharacter=_,r.friendlyAttrValue=w,r.escapeAttrValue=S,r.onIgnoreTagStripAll=function(){return""},r.StripTagBody=function(a,o){"function"!=typeof o&&(o=function(){});var s=!Array.isArray(a),l=[],p=!1;return{onIgnoreTag:function(t,e,r){if(function(t){return s||-1!==c.indexOf(a,t)}(t)){if(r.isClosing){var i="[/removed]",n=r.position+i.length;return l.push([!1!==p?p:r.position,n]),p=!1,i}return p||(p=r.position),"[removed]"}return o(t,e,r)},remove:function(e){var r="",i=0;return c.forEach(l,function(t){r+=e.slice(i,t[0]),i=t[1]}),r+=e.slice(i)}}},r.stripCommentTag=function(t){return t.replace(T,"")},r.stripBlankChar=function(t){var e=t.split("");return(e=e.filter(function(t){var e=t.charCodeAt(0);return 127!==e&&(!(e<=31)||(10===e||13===e))})).join("")},r.cssFilter=n},{"./util":4,cssfilter:8}],2:[function(t,e,r){var i=t("./default"),n=t("./parser"),a=t("./xss");for(var o in(r=e.exports=function(t,e){return new a(e).process(t)}).FilterXSS=a,i)r[o]=i[o];for(var o in n)r[o]=n[o];void 0===(l=function(){return e.exports}.call(r,s,r,e))||(e.exports=l),"undefined"!=typeof window&&(window.filterXSS=e.exports)},{"./default":1,"./parser":3,"./xss":5}],3:[function(t,e,r){var c=t("./util");function h(t){var e=t.indexOf(" ");if(-1===e)var r=t.slice(1,-1);else r=t.slice(1,e+1);return"/"===(r=c.trim(r).toLowerCase()).slice(0,1)&&(r=r.slice(1)),"/"===r.slice(-1)&&(r=r.slice(0,-1)),r}var u=/[^a-zA-Z0-9_:\.\-]/gim;function d(t,e){for(;e<t.length;e++){var r=t[e];if(" "!==r)return"="===r?e:-1}}function f(t,e){for(;0<e;e--){var r=t[e];if(" "!==r)return"="===r?e:-1}}function m(t){return function(t){return'"'===t[0]&&'"'===t[t.length-1]||"'"===t[0]&&"'"===t[t.length-1]}(t)?t.substr(1,t.length-2):t}r.parseTag=function(t,e,r){var i="",n=0,a=!1,o=!1,s=0,l=t.length,p="",c="";for(s=0;s<l;s++){var u=t.charAt(s);if(!1===a){if("<"===u){a=s;continue}}else if(!1===o){if("<"===u){i+=r(t.slice(n,s)),n=a=s;continue}if(">"===u){i+=r(t.slice(n,a)),c=h(p=t.slice(a,s+1)),i+=e(a,i.length,c,p,"</"===p.slice(0,2)),n=s+1,a=!1;continue}if(('"'===u||"'"===u)&&"="===t.charAt(s-1)){o=u;continue}}else if(u===o){o=!1;continue}}return n<t.length&&(i+=r(t.substr(n))),i},r.parseAttr=function(t,i){var e=0,n=[],r=!1,a=t.length;function o(t,e){if(!((t=(t=c.trim(t)).replace(u,"").toLowerCase()).length<1)){var r=i(t,e||"");r&&n.push(r)}}for(var s=0;s<a;s++){var l,p=t.charAt(s);if(!1!==r||"="!==p)if(!1===r||s!==e||'"'!==p&&"'"!==p||"="!==t.charAt(s-1))if(" "!==p);else{if(!1===r){if(-1===(l=d(t,s))){o(c.trim(t.slice(e,s))),r=!1,e=s+1;continue}s=l-1;continue}if(-1!==(l=f(t,s-1)))continue;o(r,m(c.trim(t.slice(e,s)))),r=!1,e=s+1}else{if(-1===(l=t.indexOf(p,s+1)))break;o(r,c.trim(t.slice(e+1,l))),r=!1,e=(s=l)+1}else r=t.slice(e,s),e=s+1}return e<t.length&&(!1===r?o(t.slice(e)):o(r,m(c.trim(t.slice(e))))),c.trim(n.join(" "))}},{"./util":4}],4:[function(t,e,r){e.exports={indexOf:function(t,e){var r,i;if(Array.prototype.indexOf)return t.indexOf(e);for(r=0,i=t.length;r<i;r++)if(t[r]===e)return r;return-1},forEach:function(t,e,r){var i,n;if(Array.prototype.forEach)return t.forEach(e,r);for(i=0,n=t.length;i<n;i++)e.call(r,t[i],i,t)},trim:function(t){return String.prototype.trim?t.trim():t.replace(/(^\s*)|(\s*$)/g,"")}}},{}],5:[function(t,e,r){var i=t("cssfilter").FilterCSS,n=t("./default"),a=t("./parser"),o=a.parseTag,y=a.parseAttr,x=t("./util");function b(t){return null==t}function s(t){(t=t||{}).stripIgnoreTag&&(t.onIgnoreTag&&console.error('Notes: cannot use these two options "stripIgnoreTag" and "onIgnoreTag" at the same time'),t.onIgnoreTag=n.onIgnoreTagStripAll),t.whiteList=t.whiteList||n.whiteList,t.onTag=t.onTag||n.onTag,t.onTagAttr=t.onTagAttr||n.onTagAttr,t.onIgnoreTag=t.onIgnoreTag||n.onIgnoreTag,t.onIgnoreTagAttr=t.onIgnoreTagAttr||n.onIgnoreTagAttr,t.safeAttrValue=t.safeAttrValue||n.safeAttrValue,t.escapeHtml=t.escapeHtml||n.escapeHtml,t.css=t.css||{},this.options=t,this.cssFilter=new i(t.css)}s.prototype.process=function(t){if(!(t=(t=t||"").toString()))return"";var e=this.options,c=e.whiteList,u=e.onTag,h=e.onIgnoreTag,d=e.onTagAttr,f=e.onIgnoreTagAttr,m=e.safeAttrValue,g=e.escapeHtml,v=this.cssFilter;if(e.stripBlankChar&&(t=n.stripBlankChar(t)),e.allowCommentTag||(t=n.stripCommentTag(t)),e.stripIgnoreTagBody){var r=n.StripTagBody(e.stripIgnoreTagBody,h);h=r.onIgnoreTag}else r=!1;var i=o(t,function(t,e,n,r,i){var a,o={sourcePosition:t,position:e,isClosing:i,isWhite:n in c};if(!b(a=u(n,r,o)))return a;if(o.isWhite){if(o.isClosing)return"</"+n+">";var s=function(t){var e=t.indexOf(" ");if(-1===e)return{html:"",closing:"/"===t[t.length-2]};var r="/"===(t=x.trim(t.slice(e+1,-1)))[t.length-1];return r&&(t=x.trim(t.slice(0,-1))),{html:t,closing:r}}(r),l=c[n],p=y(s.html,function(t,e){var r,i=-1!==x.indexOf(l,t);return b(r=d(n,t,e,i))?i?(e=m(n,t,e,v))?t+'="'+e+'"':t:b(r=f(n,t,e,i))?void 0:r:r});r="<"+n;return p&&(r+=" "+p),s.closing&&(r+=" /"),r+=">"}return b(a=h(n,r,o))?g(r):a},g);return r&&(i=r.remove(i)),i},e.exports=s},{"./default":1,"./parser":3,"./util":4,cssfilter:8}],6:[function(t,e,r){var i=t("./default"),n=t("./parser");t("./util");function h(t){return null==t}function a(t){(t=t||{}).whiteList=t.whiteList||i.whiteList,t.onAttr=t.onAttr||i.onAttr,t.onIgnoreAttr=t.onIgnoreAttr||i.onIgnoreAttr,this.options=t}a.prototype.process=function(t){if(!(t=(t=t||"").toString()))return"";var e=this.options,p=e.whiteList,c=e.onAttr,u=e.onIgnoreAttr;return n(t,function(t,e,r,i,n){var a=p[r],o=!1;!0===a?o=a:"function"==typeof a?o=a(i):a instanceof RegExp&&(o=a.test(i)),!0!==o&&(o=!1);var s,l={position:e,sourcePosition:t,source:n,isWhite:o};return o?h(s=c(r,i,l))?r+":"+i:s:h(s=u(r,i,l))?void 0:s})},e.exports=a},{"./default":7,"./parser":9,"./util":10}],7:[function(t,e,r){var i={};i["align-content"]=!1,i["align-items"]=!1,i["align-self"]=!1,i["alignment-adjust"]=!1,i["alignment-baseline"]=!1,i.all=!1,i["anchor-point"]=!1,i.animation=!1,i["animation-delay"]=!1,i["animation-direction"]=!1,i["animation-duration"]=!1,i["animation-fill-mode"]=!1,i["animation-iteration-count"]=!1,i["animation-name"]=!1,i["animation-play-state"]=!1,i["animation-timing-function"]=!1,i.azimuth=!1,i["backface-visibility"]=!1,i.background=!0,i["background-attachment"]=!0,i["background-clip"]=!0,i["background-color"]=!0,i["background-image"]=!0,i["background-origin"]=!0,i["background-position"]=!0,i["background-repeat"]=!0,i["background-size"]=!0,i["baseline-shift"]=!1,i.binding=!1,i.bleed=!1,i["bookmark-label"]=!1,i["bookmark-level"]=!1,i["bookmark-state"]=!1,i.border=!0,i["border-bottom"]=!0,i["border-bottom-color"]=!0,i["border-bottom-left-radius"]=!0,i["border-bottom-right-radius"]=!0,i["border-bottom-style"]=!0,i["border-bottom-width"]=!0,i["border-collapse"]=!0,i["border-color"]=!0,i["border-image"]=!0,i["border-image-outset"]=!0,i["border-image-repeat"]=!0,i["border-image-slice"]=!0,i["border-image-source"]=!0,i["border-image-width"]=!0,i["border-left"]=!0,i["border-left-color"]=!0,i["border-left-style"]=!0,i["border-left-width"]=!0,i["border-radius"]=!0,i["border-right"]=!0,i["border-right-color"]=!0,i["border-right-style"]=!0,i["border-right-width"]=!0,i["border-spacing"]=!0,i["border-style"]=!0,i["border-top"]=!0,i["border-top-color"]=!0,i["border-top-left-radius"]=!0,i["border-top-right-radius"]=!0,i["border-top-style"]=!0,i["border-top-width"]=!0,i["border-width"]=!0,i.bottom=!1,i["box-decoration-break"]=!0,i["box-shadow"]=!0,i["box-sizing"]=!0,i["box-snap"]=!0,i["box-suppress"]=!0,i["break-after"]=!0,i["break-before"]=!0,i["break-inside"]=!0,i["caption-side"]=!1,i.chains=!1,i.clear=!0,i.clip=!1,i["clip-path"]=!1,i["clip-rule"]=!1,i.color=!0,i["color-interpolation-filters"]=!0,i["column-count"]=!1,i["column-fill"]=!1,i["column-gap"]=!1,i["column-rule"]=!1,i["column-rule-color"]=!1,i["column-rule-style"]=!1,i["column-rule-width"]=!1,i["column-span"]=!1,i["column-width"]=!1,i.columns=!1,i.contain=!1,i.content=!1,i["counter-increment"]=!1,i["counter-reset"]=!1,i["counter-set"]=!1,i.crop=!1,i.cue=!1,i["cue-after"]=!1,i["cue-before"]=!1,i.cursor=!1,i.direction=!1,i.display=!0,i["display-inside"]=!0,i["display-list"]=!0,i["display-outside"]=!0,i["dominant-baseline"]=!1,i.elevation=!1,i["empty-cells"]=!1,i.filter=!1,i.flex=!1,i["flex-basis"]=!1,i["flex-direction"]=!1,i["flex-flow"]=!1,i["flex-grow"]=!1,i["flex-shrink"]=!1,i["flex-wrap"]=!1,i.float=!1,i["float-offset"]=!1,i["flood-color"]=!1,i["flood-opacity"]=!1,i["flow-from"]=!1,i["flow-into"]=!1,i.font=!0,i["font-family"]=!0,i["font-feature-settings"]=!0,i["font-kerning"]=!0,i["font-language-override"]=!0,i["font-size"]=!0,i["font-size-adjust"]=!0,i["font-stretch"]=!0,i["font-style"]=!0,i["font-synthesis"]=!0,i["font-variant"]=!0,i["font-variant-alternates"]=!0,i["font-variant-caps"]=!0,i["font-variant-east-asian"]=!0,i["font-variant-ligatures"]=!0,i["font-variant-numeric"]=!0,i["font-variant-position"]=!0,i["font-weight"]=!0,i.grid=!1,i["grid-area"]=!1,i["grid-auto-columns"]=!1,i["grid-auto-flow"]=!1,i["grid-auto-rows"]=!1,i["grid-column"]=!1,i["grid-column-end"]=!1,i["grid-column-start"]=!1,i["grid-row"]=!1,i["grid-row-end"]=!1,i["grid-row-start"]=!1,i["grid-template"]=!1,i["grid-template-areas"]=!1,i["grid-template-columns"]=!1,i["grid-template-rows"]=!1,i["hanging-punctuation"]=!1,i.height=!0,i.hyphens=!1,i.icon=!1,i["image-orientation"]=!1,i["image-resolution"]=!1,i["ime-mode"]=!1,i["initial-letters"]=!1,i["inline-box-align"]=!1,i["justify-content"]=!1,i["justify-items"]=!1,i["justify-self"]=!1,i.left=!1,i["letter-spacing"]=!0,i["lighting-color"]=!0,i["line-box-contain"]=!1,i["line-break"]=!1,i["line-grid"]=!1,i["line-height"]=!0,i["line-snap"]=!1,i["line-stacking"]=!1,i["line-stacking-ruby"]=!1,i["line-stacking-shift"]=!1,i["line-stacking-strategy"]=!1,i["list-style"]=!0,i["list-style-image"]=!0,i["list-style-position"]=!0,i["list-style-type"]=!0,i.margin=!0,i["margin-bottom"]=!0,i["margin-left"]=!0,i["margin-right"]=!0,i["margin-top"]=!0,i["marker-offset"]=!1,i["marker-side"]=!1,i.marks=!1,i.mask=!1,i["mask-box"]=!1,i["mask-box-outset"]=!1,i["mask-box-repeat"]=!1,i["mask-box-slice"]=!1,i["mask-box-source"]=!1,i["mask-box-width"]=!1,i["mask-clip"]=!1,i["mask-image"]=!1,i["mask-origin"]=!1,i["mask-position"]=!1,i["mask-repeat"]=!1,i["mask-size"]=!1,i["mask-source-type"]=!1,i["mask-type"]=!1,i["max-height"]=!0,i["max-lines"]=!1,i["max-width"]=!0,i["min-height"]=!0,i["min-width"]=!0,i["move-to"]=!1,i["nav-down"]=!1,i["nav-index"]=!1,i["nav-left"]=!1,i["nav-right"]=!1,i["nav-up"]=!1,i["object-fit"]=!1,i["object-position"]=!1,i.opacity=!1,i.order=!1,i.orphans=!1,i.outline=!1,i["outline-color"]=!1,i["outline-offset"]=!1,i["outline-style"]=!1,i["outline-width"]=!1,i.overflow=!1,i["overflow-wrap"]=!1,i["overflow-x"]=!1,i["overflow-y"]=!1,i.padding=!0,i["padding-bottom"]=!0,i["padding-left"]=!0,i["padding-right"]=!0,i["padding-top"]=!0,i.page=!1,i["page-break-after"]=!1,i["page-break-before"]=!1,i["page-break-inside"]=!1,i["page-policy"]=!1,i.pause=!1,i["pause-after"]=!1,i["pause-before"]=!1,i.perspective=!1,i["perspective-origin"]=!1,i.pitch=!1,i["pitch-range"]=!1,i["play-during"]=!1,i.position=!1,i["presentation-level"]=!1,i.quotes=!1,i["region-fragment"]=!1,i.resize=!1,i.rest=!1,i["rest-after"]=!1,i["rest-before"]=!1,i.richness=!1,i.right=!1,i.rotation=!1,i["rotation-point"]=!1,i["ruby-align"]=!1,i["ruby-merge"]=!1,i["ruby-position"]=!1,i["shape-image-threshold"]=!1,i["shape-outside"]=!1,i["shape-margin"]=!1,i.size=!1,i.speak=!1,i["speak-as"]=!1,i["speak-header"]=!1,i["speak-numeral"]=!1,i["speak-punctuation"]=!1,i["speech-rate"]=!1,i.stress=!1,i["string-set"]=!1,i["tab-size"]=!1,i["table-layout"]=!1,i["text-align"]=!0,i["text-align-last"]=!0,i["text-combine-upright"]=!0,i["text-decoration"]=!0,i["text-decoration-color"]=!0,i["text-decoration-line"]=!0,i["text-decoration-skip"]=!0,i["text-decoration-style"]=!0,i["text-emphasis"]=!0,i["text-emphasis-color"]=!0,i["text-emphasis-position"]=!0,i["text-emphasis-style"]=!0,i["text-height"]=!0,i["text-indent"]=!0,i["text-justify"]=!0,i["text-orientation"]=!0,i["text-overflow"]=!0,i["text-shadow"]=!0,i["text-space-collapse"]=!0,i["text-transform"]=!0,i["text-underline-position"]=!0,i["text-wrap"]=!0,i.top=!1,i.transform=!1,i["transform-origin"]=!1,i["transform-style"]=!1,i.transition=!1,i["transition-delay"]=!1,i["transition-duration"]=!1,i["transition-property"]=!1,i["transition-timing-function"]=!1,i["unicode-bidi"]=!1,i["vertical-align"]=!1,i.visibility=!1,i["voice-balance"]=!1,i["voice-duration"]=!1,i["voice-family"]=!1,i["voice-pitch"]=!1,i["voice-range"]=!1,i["voice-rate"]=!1,i["voice-stress"]=!1,i["voice-volume"]=!1,i.volume=!1,i["white-space"]=!1,i.widows=!1,i.width=!0,i["will-change"]=!1,i["word-break"]=!0,i["word-spacing"]=!0,i["word-wrap"]=!0,i["wrap-flow"]=!1,i["wrap-through"]=!1,i["writing-mode"]=!1,i["z-index"]=!1,r.whiteList=i,r.onAttr=function(t,e,r){},r.onIgnoreAttr=function(t,e,r){}},{}],8:[function(t,e,r){var i=t("./default"),n=t("./css");for(var a in(r=e.exports=function(t,e){return new n(e).process(t)}).FilterCSS=n,i)r[a]=i[a];void 0===(l=function(){return e.exports}.call(r,s,r,e))||(e.exports=l),"undefined"!=typeof window&&(window.filterCSS=e.exports)},{"./css":6,"./default":7}],9:[function(t,e,r){var u=t("./util");e.exports=function(a,o){";"!==(a=u.trimRight(a))[a.length-1]&&(a+=";");var t=a.length,s=!1,l=0,p=0,c="";function e(){if(!s){var t=u.trim(a.slice(l,p)),e=t.indexOf(":");if(-1!==e){var r=u.trim(t.slice(0,e)),i=u.trim(t.slice(e+1));if(r){var n=o(l,c.length,r,i,t);n&&(c+=n+"; ")}}}l=p+1}for(;p<t;p++){var r=a[p];if("/"===r&&"*"===a[p+1]){var i=a.indexOf("*/",p+2);if(-1===i)break;l=(p=i+1)+1,s=!1}else"("===r?s=!0:")"===r?s=!1:";"===r?s||e():"\n"===r&&e()}return u.trim(c)}},{"./util":10}],10:[function(t,e,r){e.exports={indexOf:function(t,e){var r,i;if(Array.prototype.indexOf)return t.indexOf(e);for(r=0,i=t.length;r<i;r++)if(t[r]===e)return r;return-1},forEach:function(t,e,r){var i,n;if(Array.prototype.forEach)return t.forEach(e,r);for(i=0,n=t.length;i<n;i++)e.call(r,t[i],i,t)},trim:function(t){return String.prototype.trim?t.trim():t.replace(/(^\s*)|(\s*$)/g,"")},trimRight:function(t){return String.prototype.trimRight?t.trimRight():t.replace(/(\s*$)/g,"")}}},{}]},{},[2])},function(module,exports,__webpack_require__){var __WEBPACK_AMD_DEFINE_RESULT__,D5b,E5b;function _typeof(t){return(_typeof="function"==typeof Symbol&&"symbol"==typeof Symbol.iterator?function(t){return typeof t}:function(t){return t&&"function"==typeof Symbol&&t.constructor===Symbol&&t!==Symbol.prototype?"symbol":typeof t})(t)}"undefined"!=typeof navigator&&(D5b=window||{},E5b=function(window){"use strict";var svgNS="http://www.w3.org/2000/svg",locationHref="",initialDefaultFrame=-999999,subframeEnabled=!0,expressionsPlugin,isSafari=/^((?!chrome|android).)*safari/i.test(navigator.userAgent),cachedColors={},bm_rounder=Math.round,bm_rnd,bm_pow=Math.pow,bm_sqrt=Math.sqrt,bm_abs=Math.abs,bm_floor=Math.floor,bm_max=Math.max,bm_min=Math.min,blitter=10,BMMath={};function ProjectInterface(){return{}}!function(){var t,e=["abs","acos","acosh","asin","asinh","atan","atanh","atan2","ceil","cbrt","expm1","clz32","cos","cosh","exp","floor","fround","hypot","imul","log","log1p","log2","log10","max","min","pow","random","round","sign","sin","sinh","sqrt","tan","tanh","trunc","E","LN10","LN2","LOG10E","LOG2E","PI","SQRT1_2","SQRT2"],r=e.length;for(t=0;t<r;t+=1)BMMath[e[t]]=Math[e[t]]}(),BMMath.random=Math.random,BMMath.abs=function(t){if("object"===_typeof(t)&&t.length){var e,r=createSizedArray(t.length),i=t.length;for(e=0;e<i;e+=1)r[e]=Math.abs(t[e]);return r}return Math.abs(t)};var defaultCurveSegments=150,degToRads=Math.PI/180,roundCorner=.5519;function roundValues(t){bm_rnd=t?Math.round:function(t){return t}}function styleDiv(t){t.style.position="absolute",t.style.top=0,t.style.left=0,t.style.display="block",t.style.transformOrigin=t.style.webkitTransformOrigin="0 0",t.style.backfaceVisibility=t.style.webkitBackfaceVisibility="visible",t.style.transformStyle=t.style.webkitTransformStyle=t.style.mozTransformStyle="preserve-3d"}function BMEnterFrameEvent(t,e,r,i){this.type=t,this.currentTime=e,this.totalTime=r,this.direction=i<0?-1:1}function BMCompleteEvent(t,e){this.type=t,this.direction=e<0?-1:1}function BMCompleteLoopEvent(t,e,r,i){this.type=t,this.currentLoop=r,this.totalLoops=e,this.direction=i<0?-1:1}function BMSegmentStartEvent(t,e,r){this.type=t,this.firstFrame=e,this.totalFrames=r}function BMDestroyEvent(t,e){this.type=t,this.target=e}roundValues(!1);var createElementID=(e6b=0,function(){return"__lottie_element_"+ ++e6b}),e6b;function HSVtoRGB(t,e,r){var i,n,a,o,s,l,p,c;switch(l=r*(1-e),p=r*(1-(s=6*t-(o=Math.floor(6*t)))*e),c=r*(1-(1-s)*e),o%6){case 0:i=r,n=c,a=l;break;case 1:i=p,n=r,a=l;break;case 2:i=l,n=r,a=c;break;case 3:i=l,n=p,a=r;break;case 4:i=c,n=l,a=r;break;case 5:i=r,n=l,a=p}return[i,n,a]}function RGBtoHSV(t,e,r){var i,n=Math.max(t,e,r),a=Math.min(t,e,r),o=n-a,s=0===n?0:o/n,l=n/255;switch(n){case a:i=0;break;case t:i=e-r+o*(e<r?6:0),i/=6*o;break;case e:i=r-t+2*o,i/=6*o;break;case r:i=t-e+4*o,i/=6*o}return[i,s,l]}function addSaturationToRGB(t,e){var r=RGBtoHSV(255*t[0],255*t[1],255*t[2]);return r[1]+=e,1<r[1]?r[1]=1:r[1]<=0&&(r[1]=0),HSVtoRGB(r[0],r[1],r[2])}function addBrightnessToRGB(t,e){var r=RGBtoHSV(255*t[0],255*t[1],255*t[2]);return r[2]+=e,1<r[2]?r[2]=1:r[2]<0&&(r[2]=0),HSVtoRGB(r[0],r[1],r[2])}function addHueToRGB(t,e){var r=RGBtoHSV(255*t[0],255*t[1],255*t[2]);return r[0]+=e/360,1<r[0]?r[0]-=1:r[0]<0&&(r[0]+=1),HSVtoRGB(r[0],r[1],r[2])}var rgbToHex=function(){var t,e,i=[];for(t=0;t<256;t+=1)e=t.toString(16),i[t]=1==e.length?"0"+e:e;return function(t,e,r){return t<0&&(t=0),e<0&&(e=0),r<0&&(r=0),"#"+i[t]+i[e]+i[r]}}();function BaseEvent(){}BaseEvent.prototype={triggerEvent:function(t,e){if(this._cbs[t])for(var r=this._cbs[t].length,i=0;i<r;i++)this._cbs[t][i](e)},addEventListener:function(t,e){return this._cbs[t]||(this._cbs[t]=[]),this._cbs[t].push(e),function(){this.removeEventListener(t,e)}.bind(this)},removeEventListener:function(t,e){if(e){if(this._cbs[t]){for(var r=0,i=this._cbs[t].length;r<i;)this._cbs[t][r]===e&&(this._cbs[t].splice(r,1),r-=1,i-=1),r+=1;this._cbs[t].length||(this._cbs[t]=null)}}else this._cbs[t]=null}};var createTypedArray="function"==typeof Uint8ClampedArray&&"function"==typeof Float32Array?function(t,e){return"float32"===t?new Float32Array(e):"int16"===t?new Int16Array(e):"uint8c"===t?new Uint8ClampedArray(e):void 0}:function(t,e){var r,i=0,n=[];switch(t){case"int16":case"uint8c":r=1;break;default:r=1.1}for(i=0;i<e;i+=1)n.push(r);return n};function createSizedArray(t){return Array.apply(null,{length:t})}function createNS(t){return document.createElementNS(svgNS,t)}function createTag(t){return document.createElement(t)}function DynamicPropertyContainer(){}DynamicPropertyContainer.prototype={addDynamicProperty:function(t){-1===this.dynamicProperties.indexOf(t)&&(this.dynamicProperties.push(t),this.container.addDynamicProperty(this),this._isAnimated=!0)},iterateDynamicProperties:function(){this._mdf=!1;var t,e=this.dynamicProperties.length;for(t=0;t<e;t+=1)this.dynamicProperties[t].getValue(),this.dynamicProperties[t]._mdf&&(this._mdf=!0)},initDynamicPropertyContainer:function(t){this.container=t,this.dynamicProperties=[],this._mdf=!1,this._isAnimated=!1}};var getBlendMode=(t7b={0:"source-over",1:"multiply",2:"screen",3:"overlay",4:"darken",5:"lighten",6:"color-dodge",7:"color-burn",8:"hard-light",9:"soft-light",10:"difference",11:"exclusion",12:"hue",13:"saturation",14:"color",15:"luminosity"},function(t){return t7b[t]||""}),t7b,Matrix=(v7b=Math.cos,w7b=Math.sin,x7b=Math.tan,y7b=Math.round,function(){this.reset=z7b,this.rotate=A7b,this.rotateX=B7b,this.rotateY=C7b,this.rotateZ=D7b,this.skew=F7b,this.skewFromAxis=G7b,this.shear=E7b,this.scale=H7b,this.setTransform=I7b,this.translate=J7b,this.transform=K7b,this.applyToPoint=P7b,this.applyToX=Q7b,this.applyToY=R7b,this.applyToZ=S7b,this.applyToPointArray=W7b,this.applyToTriplePoints=V7b,this.applyToPointStringified=X7b,this.toCSS=Y7b,this.to2dCSS=$7b,this.clone=N7b,this.cloneFromProps=O7b,this.equals=M7b,this.inversePoints=U7b,this.inversePoint=T7b,this._t=this.transform,this.isIdentity=L7b,this._identity=!0,this._identityCalculated=!1,this.props=createTypedArray("float32",16),this.reset()}),v7b,w7b,x7b,y7b;
/*!
   Transformation Matrix v2.0
   (c) Epistemex 2014-2015
   www.epistemex.com
   By Ken Fyrstenberg
   Contributions by leeoniya.
   License: MIT, header required.
```
