## Supplement File 1 for "Post Induction Hypotension prediction during general anesthesia using Machine Learning Techniques": aframe-gif-shader.protoio.custom.js

```
/******/ (function(modules) { // webpackBootstrap
    /******/ 	// The module cache
    /******/ 	var installedModules = {};

    /******/ 	// The require function
    /******/ 	function __webpack_require__(moduleId) {

        /******/ 		// Check if module is in cache
        /******/ 		if(installedModules[moduleId])
        /******/ 			return installedModules[moduleId].exports;

        /******/ 		// Create a new module (and put it into the cache)
        /******/ 		var module = installedModules[moduleId] = {
            /******/ 			exports: {},
            /******/ 			id: moduleId,
            /******/ 			loaded: false
            /******/ 		};

        /******/ 		// Execute the module function
        /******/ 		modules[moduleId].call(module.exports, module, module.exports, __webpack_require__);

        /******/ 		// Flag the module as loaded
        /******/ 		module.loaded = true;

        /******/ 		// Return the exports of the module
        /******/ 		return module.exports;
        /******/ 	}


    /******/ 	// expose the modules object (__webpack_modules__)
    /******/ 	__webpack_require__.m = modules;

    /******/ 	// expose the module cache
    /******/ 	__webpack_require__.c = installedModules;

    /******/ 	// __webpack_public_path__
    /******/ 	__webpack_require__.p = "";

    /******/ 	// Load entry module and return exports
    /******/ 	return __webpack_require__(0);
    /******/ })
/************************************************************************/
/******/ ([
    /* 0 */
    /***/ function(module, exports, __webpack_require__) {

        'use strict';

        var _typeof = typeof Symbol === "function" && typeof Symbol.iterator === "symbol" ? function (obj) { return typeof obj; } : function (obj) { return obj && typeof Symbol === "function" && obj.constructor === Symbol ? "symbol" : typeof obj; };

        var _gifsparser = __webpack_require__(1);

        if (typeof AFRAME === 'undefined') {
            throw 'Component attempted to register before AFRAME was available.';
        }

        /* get util from AFRAME */
        var parseUrl = AFRAME.utils.srcLoader.parseUrl;
        var debug = AFRAME.utils.debug;
        // debug.enable('shader:gif:*')

        debug.enable('shader:gif:warn');
        var warn = debug('shader:gif:warn');
        var log = debug('shader:gif:debug');

        /* store data so that you won't load same data */
        var gifData = {};

        /* create error message */
        function createError(err, src) {
            return { status: 'error', src: src, message: err, timestamp: Date.now() };
        }

        AFRAME.registerShader('gif', {

            /**
             * For material component:
             * @see https://github.com/aframevr/aframe/blob/60d198ef8e2bfbc57a13511ae5fca7b62e01691b/src/components/material.js
             * For example of `registerShader`:
             * @see https://github.com/aframevr/aframe/blob/41a50cd5ac65e462120ecc2e5091f5daefe3bd1e/src/shaders/flat.js
             * For MeshBasicMaterial
             * @see http://threejs.org/docs/#Reference/Materials/MeshBasicMaterial
             */

            schema: {

                /* For material */
                color: { type: 'color' },
                fog: { default: true },

                /* For texuture */
                src: { default: null },
                autoplay: { default: true }

            },

            /**
             * Initialize material. Called once.
             * @protected
             */
            init: function init(data) {
                log('init', data);
                log(this.el.components);
                this.__cnv = document.createElement('canvas');
                this.__cnv.width = 2;
                this.__cnv.height = 2;
                this.__ctx = this.__cnv.getContext('2d');
                this.__texture = new THREE.Texture(this.__cnv); //renders straight from a canvas
                this.__material = {};
                this.__reset();
                this.material = new THREE.MeshBasicMaterial({ map: this.__texture });
                this.el.sceneEl.addBehavior(this);
                this.__addPublicFunctions();
                return this.material;
            },


            /**
             * Update or create material.
             * @param {object|null} oldData
             */
            update: function update(oldData) {
                log('update', oldData);
                this.__updateMaterial(oldData);
                this.__updateTexture(oldData);
                return this.material;
            },


            /**
             * Called on each scene tick.
             * @protected
             */
            tick: function tick(t) {
                if (!this.__frames || this.paused()) return;
                if (Date.now() - this.__startTime >= this.__nextFrameTime) {
                    this.nextFrame();
                }
            },


            /*================================
             =            material            =
             ================================*/

            /**
             * Updating existing material.
             * @param {object} data - Material component data.
             */
            __updateMaterial: function __updateMaterial(data) {
                var material = this.material;

                var newData = this.__getMaterialData(data);
                Object.keys(newData).forEach(function (key) {
                    material[key] = newData[key];
                });
            },


            /**
             * Builds and normalize material data, normalizing stuff along the way.
             * @param {Object} data - Material data.
             * @return {Object} data - Processed material data.
             */
            __getMaterialData: function __getMaterialData(data) {
                return {
                    fog: data.fog,
                    color: new THREE.Color(data.color)
                };
            },


            /*==============================
             =            texure            =
             ==============================*/

            /**
             * set texure
             * @private
             * @param {Object} data
             * @property {string} status - success / error
             * @property {string} src - src url
             * @property {array} times - array of time length of each image
             * @property {number} cnt - total counts of gif images
             * @property {array} frames - array of each image
             * @property {Date} timestamp - created at the texure
             */

            __setTexure: function __setTexure(data) {
                log('__setTexure', data);
                if (data.status === 'error') {
                    warn('Error: ' + data.message + '\nsrc: ' + data.src);
                    this.__reset();
                } else if (data.status === 'success' && data.src !== this.__textureSrc) {
                    this.__reset();
                    /* Texture added or changed */
                    this.__ready(data);
                }
            },


            /**
             * Update or create texure.
             * @param {Object} data - Material component data.
             */
            __updateTexture: function __updateTexture(data) {
                var src = data.src;
                var autoplay = data.autoplay;

                /* autoplay */

                if (typeof autoplay === 'boolean') {
                    this.__autoplay = autoplay;
                } else if (typeof autoplay === 'undefined') {
                    this.__autoplay = true;
                }
                if (this.__autoplay && this.__frames) {
                    this.play();
                }

                /* src */
                if (src) {
                    this.__validateSrc(src, this.__setTexure.bind(this));
                } else {
                    /* Texture removed */
                    this.__reset();
                }
            },


            /*=============================================
             =            varidation for texure            =
             =============================================*/

            __validateSrc: function __validateSrc(src, cb) {

                /* check if src is a url */
                var url = parseUrl(src);
                if (url) {
                    this.__getImageSrc(url, cb);
                    return;
                }

                var message = void 0;

                /* check if src is a query selector */
                var el = this.__validateAndGetQuerySelector(src);
                if (!el || (typeof el === 'undefined' ? 'undefined' : _typeof(el)) !== 'object') {
                    return;
                }
                if (el.error) {
                    message = el.error;
                } else {
                    var tagName = el.tagName.toLowerCase();
                    if (tagName === 'video') {
                        src = el.src;
                        message = 'For video, please use `aframe-video-shader`';
                    } else if (tagName === 'img') {
                        this.__getImageSrc(el.src, cb);
                        return;
                    } else {
                        message = 'For <' + tagName + '> element, please use `aframe-html-shader`';
                    }
                }

                /* if there is message, create error data */
                if (message) {
                    (function () {
                        var srcData = gifData[src];
                        var errData = createError(message, src);
                        /* callbacks */
                        if (srcData && srcData.callbacks) {
                            srcData.callbacks.forEach(function (cb) {
                                return cb(errData);
                            });
                        } else {
                            cb(errData);
                        }
                        /* overwrite */
                        gifData[src] = errData;
                    })();
                }
            },


            /**
             * Validate src is a valid image url
             * @param  {string} src - url that will be tested
             * @param  {function} cb - callback with the test result
             */
            __getImageSrc: function __getImageSrc(src, cb) {
                var _this = this;

                /* if src is same as previous, ignore this */
                if (src === this.__textureSrc) {
                    return;
                }

                /* check if we already get the srcData */
                var srcData = gifData[src];
                if (!srcData || !srcData.callbacks) {
                    /* create callback */
                    srcData = gifData[src] = { callbacks: [] };
                    srcData.callbacks.push(cb);
                } else if (srcData.src) {
                    cb(srcData);
                    return;
                } else if (srcData.callbacks) {
                    /* add callback */
                    srcData.callbacks.push(cb);
                    return;
                }
                var tester = new Image();
                tester.crossOrigin = 'Anonymous';
                tester.addEventListener('load', function (e) {
                    /* check if it is gif */
                    // proto.io pass flag to know if image is png
                    _this.__getUnit8Array(src, function (arr, isPng) {
                        if (!arr) {
                            onError('This is not gif. Please use `shader:flat` instead');
                            return;
                        }
                        /* parse data */

                        // proto.io if not png then handle as gif else parse as png
                        if(!isPng) {
                            // parse gif
                            (0, _gifsparser.parseGIF)(arr, function (times, cnt, frames) {
                                /* store data */
                                var newData = { status: 'success', src: src, times: times, cnt: cnt, frames: frames, timestamp: Date.now() };
                                /* callbacks */
                                if (srcData.callbacks) {
                                    srcData.callbacks.forEach(function (cb) {
                                        return cb(newData);
                                    });
                                    /* overwrite */
                                    gifData[src] = newData;
                                }
                            }, function (err) {
                                return onError(err);
                            });
                        } else {
                            // parse png
                            (0, _gifsparser.parseAPNG)(arr, function (times, cnt, frames) {
                                /* store data */
                                var newData = { status: 'success', src: src, times: times, cnt: cnt, frames: frames, timestamp: Date.now() };
                                /* callbacks */
                                if (srcData.callbacks) {
                                    srcData.callbacks.forEach(function (cb) {
                                        return cb(newData);
                                    });
                                    /* overwrite */
                                    gifData[src] = newData;
                                }
                            }, function (err) {
                                return onError(err);
                            });

                        }

                    });
                });
                tester.addEventListener('error', function (e) {
                    return onError('Could be the following issue\n - Not Image\n - Not Found\n - Server Error\n - Cross-Origin Issue');
                });
                function onError(message) {
                    /* create error data */
                    var errData = createError(message, src);
                    /* callbacks */
                    if (srcData.callbacks) {
                        srcData.callbacks.forEach(function (cb) {
                            return cb(errData);
                        });
                        /* overwrite */
                        gifData[src] = errData;
                    }
                }
                tester.src = src;
            },


            /**
             *
             * get mine type
             *
             */
            __getUnit8Array: function __getUnit8Array(src, cb) {
                if (typeof cb !== 'function') {
                    return;
                }

                var xhr = new XMLHttpRequest();
                xhr.open('GET', src);
                xhr.responseType = 'arraybuffer';
                xhr.addEventListener('load', function (e) {
                    var uint8Array = new Uint8Array(e.target.response);
                    var arr = uint8Array.subarray(0, 4);
                    // proto.io check if image header for png
                    var header = '';
                    var isPng = false;
                    for (var i = 0; i < arr.length; i++) {
                        header += arr[i].toString(16);
                    }

                    const PNGSignature = new Uint8Array([0x89, 0x50, 0x4e, 0x47, 0x0d, 0x0a, 0x1a, 0x0a]);

                    if (PNGSignature.some((b, i) => b === uint8Array[i])) {
                        isPng = true;
                    }

                    if (header === '47494638' || isPng) {
                        cb(uint8Array, isPng);
                    } else {
                        cb();
                    }
                });
                xhr.addEventListener('error', function (e) {
                    log(e);
                    cb();
                });
                xhr.send();
            },


            /**
             * Query and validate a query selector,
             *
             * @param  {string} selector - DOM selector.
             * @return {object} Selected DOM element | error message object.
             */
            __validateAndGetQuerySelector: function __validateAndGetQuerySelector(selector) {
                try {
                    var el = document.querySelector(selector);
                    if (!el) {
                        return { error: 'No element was found matching the selector' };
                    }
                    return el;
                } catch (e) {
                    // Capture exception if it's not a valid selector.
                    return { error: 'no valid selector' };
                }
            },


            /*================================
             =            playback            =
             ================================*/

            /**
             * add public functions
             * @private
             */
            __addPublicFunctions: function __addPublicFunctions() {
                this.el.gif = {
                    play: this.play.bind(this),
                    pause: this.pause.bind(this),
                    togglePlayback: this.togglePlayback.bind(this),
                    paused: this.paused.bind(this),
                    nextFrame: this.nextFrame.bind(this)
                };
            },


            /**
             * Pause gif
             * @public
             */
            pause: function pause() {
                log('pause');
                this.__paused = true;
            },


            /**
             * Play gif
             * @public
             */
            play: function play() {
                log('play');
                this.__paused = false;
            },


            /**
             * Toggle playback. play if paused and pause if played.
             * @public
             */

            togglePlayback: function togglePlayback() {

                if (this.paused()) {
                    this.play();
                } else {
                    this.pause();
                }
            },


            /**
             * Return if the playback is paused.
             * @public
             * @return {boolean}
             */
            paused: function paused() {
                return this.__paused;
            },


            /**
             * Go to next frame
             * @public
             */
            nextFrame: function nextFrame() {
                this.__draw();

                /* update next frame time */
                while (Date.now() - this.__startTime >= this.__nextFrameTime) {

                    this.__nextFrameTime += this.__delayTimes[this.__frameIdx++];
                    if ((this.__infinity || this.__loopCnt) && this.__frameCnt <= this.__frameIdx) {
                        /* go back to the first */
                        this.__frameIdx = 0;
                    }
                }
            },


            /*==============================
             =            canvas            =
             ==============================*/

            /**
             * clear canvas
             * @private
             */
            __clearCanvas: function __clearCanvas() {
                this.__ctx.clearRect(0, 0, this.__width, this.__height);
                this.__texture.needsUpdate = true;
            },


            /**
             * draw
             * @private
             */
            __draw: function __draw() {

                // proto.io change to fix transparent animted gifs
                // reference here: https://github.com/mayognaise/aframe-gif-shader/issues/2

                // check if the first frame of the gif has a fully transparent pixel
                if(!this.hasOwnProperty('hasTransparencyProtoio')){

                    // draw image on canvas
                    this.__ctx.drawImage(this.__frames[this.__frameIdx], 0, 0, this.__width, this.__height);

                    // get the data from the canvas
                    var data = this.__ctx.getImageData( 0, 0, this.__width, this.__height).data;
                    var hasTransp = false;

                    // loop pixels to find a transparent pixel
                    for(var i=0;i<data.length;i+=4){
                        if(data[i+3]<255){
                            hasTransp = true;
                            break;
                        }
                    }

                    this.hasTransparencyProtoio = hasTransp;
                    this.__texture.needsUpdate = true;
                    return;
                }

                if(this.hasTransparencyProtoio)
                    this.__clearCanvas(); // for gifs with transparencies we need to clear the canvas
                
                this.__ctx.drawImage(this.__frames[this.__frameIdx], 0, 0, this.__width, this.__height);
                this.__texture.needsUpdate = true;


                // original code
                // this.__ctx.drawImage(this.__frames[this.__frameIdx], 0, 0, this.__width, this.__height);
                // this.__texture.needsUpdate = true;
            },


            /*============================
             =            ready            =
             ============================*/

            /**
             * setup gif animation and play if autoplay is true
             * @private
             * @property {string} src - src url
             * @param {array} times - array of time length of each image
             * @param {number} cnt - total counts of gif images
             * @param {array} frames - array of each image
             */
            __ready: function __ready(_ref) {
                var src = _ref.src;
                var times = _ref.times;
                var cnt = _ref.cnt;
                var frames = _ref.frames;

                log('__ready');
                this.__textureSrc = src;
                this.__delayTimes = times;
                cnt ? this.__loopCnt = cnt : this.__infinity = true;
                this.__frames = frames;
                this.__frameCnt = times.length;
                this.__startTime = Date.now();
                this.__width = THREE.Math.nearestPowerOfTwo(frames[0].width);
                this.__height = THREE.Math.nearestPowerOfTwo(frames[0].height);
                this.__cnv.width = this.__width;
                this.__cnv.height = this.__height;
                this.__draw();
                if (this.__autoplay) {
                    this.play();
                } else {
                    this.pause();
                }
            },


            /*=============================
             =            reset            =
             =============================*/

            /**
             * @private
             */

            __reset: function __reset() {
                this.pause();
                this.__clearCanvas();
                this.__startTime = 0;
                this.__nextFrameTime = 0;
                this.__frameIdx = 0;
                this.__frameCnt = 0;
                this.__delayTimes = null;
                this.__infinity = false;
                this.__loopCnt = 0;
                this.__frames = null;
                this.__textureSrc = null;

                // protoio on image reset delete this property to force re check for transparency when src changes
                if(this.hasOwnProperty("hasTransparencyProtoio"))
                    delete this.hasTransparencyProtoio;
            }
        });

        /***/ },
    /* 1 */
    /***/ function(module, exports) {

        'use strict';

        /**
         *
         * Gif parser by @gtk2k
         * https://github.com/gtk2k/gtk2k.github.io/tree/master/animation_gif
         *
         */

        exports.parseGIF = function (gif, successCB, errorCB) {

            var pos = 0;
            var delayTimes = [];
            var loadCnt = 0;
            var graphicControl = null;
            var imageData = null;
            var frames = [];
            var loopCnt = 0;
            if (gif[0] === 0x47 && gif[1] === 0x49 && gif[2] === 0x46 && // 'GIF'
                gif[3] === 0x38 && gif[4] === 0x39 && gif[5] === 0x61) {
                // '89a'
                pos += 13 + +!!(gif[10] & 0x80) * Math.pow(2, (gif[10] & 0x07) + 1) * 3;
                var gifHeader = gif.subarray(0, pos);
                while (gif[pos] && gif[pos] !== 0x3b) {
                    var offset = pos,
                        blockId = gif[pos];
                    if (blockId === 0x21) {
                        var label = gif[++pos];
                        if ([0x01, 0xfe, 0xf9, 0xff].indexOf(label) !== -1) {
                            label === 0xf9 && delayTimes.push((gif[pos + 3] + (gif[pos + 4] << 8)) * 10);
                            label === 0xff && (loopCnt = gif[pos + 15] + (gif[pos + 16] << 8));
                            while (gif[++pos]) {
                                pos += gif[pos];
                            }label === 0xf9 && (graphicControl = gif.subarray(offset, pos + 1));
                        } else {
                            errorCB && errorCB('parseGIF: unknown label');break;
                        }
                    } else if (blockId === 0x2c) {
                        pos += 9;
                        pos += 1 + +!!(gif[pos] & 0x80) * (Math.pow(2, (gif[pos] & 0x07) + 1) * 3);
                        while (gif[++pos]) {
                            pos += gif[pos];
                        }var imageData = gif.subarray(offset, pos + 1);
                        frames.push(URL.createObjectURL(new Blob([gifHeader, graphicControl, imageData])));
                    } else {
                        errorCB && errorCB('parseGIF: unknown blockId');break;
                    }
                    pos++;
                }
            } else {
                errorCB && errorCB('parseGIF: no GIF89a');
            }
            if (frames.length) {

                var cnv = document.createElement('canvas');
                var loadImg = function loadImg() {
                    frames.forEach(function (src, i) {
                        var img = new Image();
                        img.onload = function (e, i) {
                            if (i === 0) {
                                cnv.width = img.width;
                                cnv.height = img.height;
                            }
                            loadCnt++;
                            frames[i] = this;
                            if (loadCnt === frames.length) {
                                loadCnt = 0;
                                imageFix(1);
                            }
                        }.bind(img, null, i);
                        img.src = src;
                    });
                };
                var imageFix = function imageFix(i) {
                    var img = new Image();
                    img.onload = function (e, i) {
                        loadCnt++;
                        frames[i] = this;
                        if (loadCnt === frames.length) {
                            cnv = null;
                            successCB && successCB(delayTimes, loopCnt, frames);
                        } else {
                            imageFix(++i);
                        }
                    }.bind(img);
                    img.src = cnv.toDataURL('image/gif');
                };
                loadImg();
            }
        };

        //proto.io the next code till the end of the file is added for proto.io to support apng & png parcing

        // functions & variables for png parse
        const PNGSignature = new Uint8Array([0x89, 0x50, 0x4e, 0x47, 0x0d, 0x0a, 0x1a, 0x0a]);
        var crc32Table = new Uint32Array(256);

        for (let i = 0; i < 256; i++) {
            let c = i;
            for (let k = 0; k < 8; k++) {
                c = ((c & 1) !== 0) ? 0xEDB88320 ^ (c >>> 1) : c >>> 1;
            }
            crc32Table[i] = c;
        }

        var crc32 = function (bytes, start = 0, length = bytes.length - start) {
            let crc = -1;
            for (let i = start, l = start + length; i < l; i++) {
                crc = ( crc >>> 8 ) ^ crc32Table[( crc ^ bytes[i] ) & 0xFF];
            }
            return crc ^ (-1);
        };

        var readString = function(bytes, off, length) {
            var chars = Array.prototype.slice.call(bytes.subarray(off, off + length));
            return String.fromCharCode.apply(String, chars);
        }

        var eachChunk = function(bytes, callback) {
            var dv = new DataView(bytes.buffer);
            let off = 8, type, length, res;
            do {
                length = dv.getUint32(off);
                type = readString(bytes, off + 4, 4);
                res = callback(type, bytes, off, length);
                off += 12 + length;
            } while (res !== false && type != 'IEND' && off < bytes.length);
        }

        var makeStringArray = function(x) {
            var res = new Uint8Array(x.length);
            for (let i = 0; i < x.length; i++) {
                res[i] = x.charCodeAt(i);
            }
            return res;
        }

        var subBuffer = function(bytes, start, length) {
            var a = new Uint8Array(length);
            a.set(bytes.subarray(start, start + length));
            return a;
        }

        var makeChunkBytes = function (type, dataBytes) {
            var crcLen = type.length + dataBytes.length;
            var bytes = new Uint8Array(crcLen + 8);
            var dv = new DataView(bytes.buffer);

            dv.setUint32(0, dataBytes.length);
            bytes.set(makeStringArray(type), 4);
            bytes.set(dataBytes, 8);
            var crc = crc32(bytes, 4, crcLen);
            dv.setUint32(crcLen + 4, crc);
            return bytes;
        };

        var makeDWordArray = function (x) {
            return new Uint8Array([(x >>> 24) & 0xff, (x >>> 16) & 0xff, (x >>> 8) & 0xff, x & 0xff]);
        };


        exports.parseAPNG = function (pngData, successCB, errorCB) {

            var preDataParts = [],
                postDataParts = [];
            var headerDataBytes = null,
                frame = null,
                frameNumber = 0,
                apng = {
                    frames: []
                };
            var delayTimes = [];
            var loopCnt = 0;


            // check if image is animated
            var isAnimated = false;
            eachChunk(pngData, type => !(isAnimated = (type === 'acTL')));

            eachChunk(pngData, (type, bytes, off, length) => {
                var dv = new DataView(bytes.buffer);
                switch (type) {
                    case 'IHDR':
                        headerDataBytes = bytes.subarray(off + 8, off + 8 + length);
                        apng.width = dv.getUint32(off + 8);
                        apng.height = dv.getUint32(off + 12);

                        // if the png is not animated initialize the first frame here
                        if(!isAnimated) {
                            frame = {};
                            frame.dataParts = [];
                            frame.width = apng.width;
                            frame.height = apng.height;
                            frame.left = 0;
                            frame.top = 0;
                        }

                        break;
                    case 'acTL':
                        apng.numPlays = dv.getUint32(off + 8 + 4);
                        break;
                    case 'fcTL':
                        if (frame) {
                            apng.frames.push(frame);
                            frameNumber++;
                        }
                        frame = {};
                        frame.width = dv.getUint32(off + 8 + 4);
                        frame.height = dv.getUint32(off + 8 + 8);
                        frame.left = dv.getUint32(off + 8 + 12);
                        frame.top = dv.getUint32(off + 8 + 16);
                        var delayN = dv.getUint16(off + 8 + 20);
                        var delayD = dv.getUint16(off + 8 + 22);
                        if (delayD === 0) {
                            delayD = 100;
                        }
                        frame.delay = 1000 * delayN / delayD;

                        if (frame.delay <= 10) {
                            frame.delay = 100;
                        }

                        delayTimes.push(frame.delay);

                        frame.disposeOp = dv.getUint8(off + 8 + 24);
                        frame.blendOp = dv.getUint8(off + 8 + 25);

                        frame.dataParts = [];

                        if (frameNumber === 0 && frame.disposeOp === 2) {
                            frame.disposeOp = 1;
                        }

                        break;
                    case 'fdAT':
                        if (frame) {
                            frame.dataParts.push(bytes.subarray(off + 8 + 4, off + 8 + length));
                        }
                        break;
                    case 'IDAT':
                        if (frame) {
                            frame.dataParts.push(bytes.subarray(off + 8, off + 8 + length));
                        }
                        break;
                    case 'IEND':
                        postDataParts.push(subBuffer(bytes, off, 12 + length));
                        break;
                    default:
                        preDataParts.push(subBuffer(bytes, off, 12 + length));
                }
            });

            if (frame) {
                apng.frames.push(frame);
            }

            if (apng.frames.length == 0) {
                console.log('no frames')
            }

            var preBlob = new Blob(preDataParts),
                postBlob = new Blob(postDataParts);

            var frames = [];

            // prosses frames
            apng.frames.forEach(frame => {
                var bb = [];
                bb.push(PNGSignature);
                headerDataBytes.set(makeDWordArray(frame.width), 0);
                headerDataBytes.set(makeDWordArray(frame.height), 4);
                bb.push(makeChunkBytes('IHDR', headerDataBytes));
                bb.push(preBlob);
                frame.dataParts.forEach(p => bb.push(makeChunkBytes('IDAT', p)));
                bb.push(postBlob);

                // store blob on new frame array
                frame.imageData = new Blob(bb, {'type': 'image/png'});
                frames.push(URL.createObjectURL(frame.imageData));

                delete frame.dataParts;

                bb = null;
            });

            var clearNextFrame = 0;
            var prevFrameToRerender = null;

            if (frames.length) {

                // further frames processing using canvas
                var loadCnt = 0;
                var cnv = document.createElement('canvas');

                var loadImg = function loadImg() {
                    frames.forEach(function (src, i) {
                        var img = new Image();
                        img.onload = function (e, i) {
                            if (i === 0) {
                                cnv.width = img.width;
                                cnv.height = img.height;
                            }
                            loadCnt++;
                            frames[i] = this;
                            if (loadCnt === frames.length) {
                                loadCnt = 0;
                                imageFix(1);
                            }
                        }.bind(img, null, i);
                        img.src = src;
                    });
                };

                // fix frames placement: same frames dose not have the same size as image but are smaller and use top, left to correctly placed in image
                // here we place them in the correct possition in the images size
                var imageFix = function imageFix(i) {
                    var img = new Image();
                    img.onload = function (e, i) {

                        frames[loadCnt] = this;
                        loadCnt++;
                        if (loadCnt === frames.length) {
                            cnv = null;
                            successCB && successCB(delayTimes, loopCnt, frames);
                        } else {
                            imageFix(++i);
                        }
                    }.bind(img);

                    // the following is done to get the frames in their correct possition (top, left) in the images
                    // frames that are not using the full image size are writen in the file with there bounding box size and use top & left for correct placement


                    // use 2d context on canvas to draw the frame
                    var ctx=cnv.getContext("2d");

                    // clear canvas as is directed by dispose param of the previews frame
                    if(clearNextFrame == 1) // clear everything
                        ctx.clearRect(0,0, apng.width, apng.height);
                    else if(clearNextFrame == 0 && apng.frames[loadCnt].blendOp == 0) // clear only the area that is occupied by the current frame -- if blending option is not 0 then do not clear
                        ctx.clearRect(apng.frames[loadCnt].left, apng.frames[loadCnt].top, apng.frames[loadCnt].width, apng.frames[loadCnt].height);
                    else if(clearNextFrame == 2) { // render previus canvas first
                        ctx.clearRect(0,0, apng.width, apng.height);
                        if(prevFrameToRerender)
                            ctx.drawImage(prevFrameToRerender , 0, 0);
                    }

                    // if disposeOp == 2 (APNG_DISPOSE_OP_PREVIOUS) we save the current frame to use it on the next frame
                    if(apng.frames[loadCnt].disposeOp == 2) {
                        prevFrameToRerender = new Image();
                        prevFrameToRerender.src = cnv.toDataURL('image/png');
                    } else {
                        prevFrameToRerender = null;
                    }

                    // draw current frame on canavas
                    ctx.drawImage(frames[loadCnt] , apng.frames[loadCnt].left, apng.frames[loadCnt].top);

                    img.src = cnv.toDataURL('image/png');

                    // set clear next frame setting based on current dispose option
                    clearNextFrame = apng.frames[loadCnt].disposeOp
                };

                loadImg();
            }
            else {
                errorCB && errorCB('parsePNG: unexpected error');
            }

        }


        /***/ }
    /******/ ]);
```
