## Supplement File 1 for "Post Induction Hypotension prediction during general anesthesia using Machine Learning Techniques": aframe.protoio.custom.js

```
(function(f){if(typeof exports==="object"&&typeof module!=="undefined"){module.exports=f()}else if(typeof define==="function"&&define.amd){define([],f)}else{var g;if(typeof window!=="undefined"){g=window}else if(typeof global!=="undefined"){g=global}else if(typeof self!=="undefined"){g=self}else{g=this}g.AFRAME = f()}})(function(){var define,module,exports;return (function e(t,n,r){function s(o,u){if(!n[o]){if(!t[o]){var a=typeof require=="function"&&require;if(!u&&a)return a(o,!0);if(i)return i(o,!0);var f=new Error("Cannot find module '"+o+"'");throw f.code="MODULE_NOT_FOUND",f}var l=n[o]={exports:{}};t[o][0].call(l.exports,function(e){var n=t[o][1][e];return s(n?n:e)},l,l.exports,e,t,n,r)}return n[o].exports}var i=typeof require=="function"&&require;for(var o=0;o<r.length;o++)s(r[o]);return s})({1:[function(_dereq_,module,exports){
var str = Object.prototype.toString

module.exports = anArray

function anArray(arr) {
  return (
       arr.BYTES_PER_ELEMENT
    && str.call(arr.buffer) === '[object ArrayBuffer]'
    || Array.isArray(arr)
  )
}

},{}],2:[function(_dereq_,module,exports){
module.exports = function numtype(num, def) {
	return typeof num === 'number'
		? num
		: (typeof def === 'number' ? def : 0)
}
},{}],3:[function(_dereq_,module,exports){
'use strict';
// For more information about browser field, check out the browser field at https://github.com/substack/browserify-handbook#browser-field.

module.exports = {
    // Create a <link> tag with optional data attributes
    createLink: function(href, attributes) {
        var head = document.head || document.getElementsByTagName('head')[0];
        var link = document.createElement('link');

        link.href = href;
        link.rel = 'stylesheet';

        for (var key in attributes) {
            if ( ! attributes.hasOwnProperty(key)) {
                continue;
            }
            var value = attributes[key];
            link.setAttribute('data-' + key, value);
        }

        head.appendChild(link);
    },
    // Create a <style> tag with optional data attributes
    createStyle: function(cssText, attributes) {
        var head = document.head || document.getElementsByTagName('head')[0],
            style = document.createElement('style');

        style.type = 'text/css';

        for (var key in attributes) {
            if ( ! attributes.hasOwnProperty(key)) {
                continue;
            }
            var value = attributes[key];
            style.setAttribute('data-' + key, value);
        }

        if (style.sheet) { // for jsdom and IE9+
            style.innerHTML = cssText;
            style.sheet.cssText = cssText;
            head.appendChild(style);
        } else if (style.styleSheet) { // for IE8 and below
            head.appendChild(style);
            style.styleSheet.cssText = cssText;
        } else { // for Chrome, Firefox, and Safari
            style.appendChild(document.createTextNode(cssText));
            head.appendChild(style);
        }
    }
};

},{}],4:[function(_dereq_,module,exports){
(function (global){
/*!
 * The buffer module from node.js, for the browser.
 *
 * @author   Feross Aboukhadijeh <> <http://feross.org>
 * @license  MIT
 */
/* eslint-disable no-proto */

'use strict'

var base64 = _dereq_('base64-js')
var ieee754 = _dereq_('ieee754')
var isArray = _dereq_('isarray')

exports.Buffer = Buffer
exports.SlowBuffer = SlowBuffer
exports.INSPECT_MAX_BYTES = 50

/**
 * If `Buffer.TYPED_ARRAY_SUPPORT`:
 *   === true    Use Uint8Array implementation (fastest)
 *   === false   Use Object implementation (most compatible, even IE6)
 *
 * Browsers that support typed arrays are IE 10+, Firefox 4+, Chrome 7+, Safari 5.1+,
 * Opera 11.6+, iOS 4.2+.
 *
 * Due to various browser bugs, sometimes the Object implementation will be used even
 * when the browser supports typed arrays.
 *
 * Note:
 *
 *   - Firefox 4-29 lacks support for adding new properties to `Uint8Array` instances,
 *     See: https://bugzilla.mozilla.org/show_bug.cgi?id=695438.
 *
 *   - Chrome 9-10 is missing the `TypedArray.prototype.subarray` function.
 *
 *   - IE10 has a broken `TypedArray.prototype.subarray` function which returns arrays of
 *     incorrect length in some situations.

 * We detect these buggy browsers and set `Buffer.TYPED_ARRAY_SUPPORT` to `false` so they
 * get the Object implementation, which is slower but behaves correctly.
 */
Buffer.TYPED_ARRAY_SUPPORT = global.TYPED_ARRAY_SUPPORT !== undefined
  ? global.TYPED_ARRAY_SUPPORT
  : typedArraySupport()

/*
 * Export kMaxLength after typed array support is determined.
 */
exports.kMaxLength = kMaxLength()

function typedArraySupport () {
  try {
    var arr = new Uint8Array(1)
    arr.__proto__ = {__proto__: Uint8Array.prototype, foo: function () { return 42 }}
    return arr.foo() === 42 && // typed array instances can be augmented
        typeof arr.subarray === 'function' && // chrome 9-10 lack `subarray`
        arr.subarray(1, 1).byteLength === 0 // ie10 has broken `subarray`
  } catch (e) {
    return false
  }
}

function kMaxLength () {
  return Buffer.TYPED_ARRAY_SUPPORT
    ? 0x7fffffff
    : 0x3fffffff
}

function createBuffer (that, length) {
  if (kMaxLength() < length) {
    throw new RangeError('Invalid typed array length')
  }
  if (Buffer.TYPED_ARRAY_SUPPORT) {
    // Return an augmented `Uint8Array` instance, for best performance
    that = new Uint8Array(length)
    that.__proto__ = Buffer.prototype
  } else {
    // Fallback: Return an object instance of the Buffer class
    if (that === null) {
      that = new Buffer(length)
    }
    that.length = length
  }

  return that
}

/**
 * The Buffer constructor returns instances of `Uint8Array` that have their
 * prototype changed to `Buffer.prototype`. Furthermore, `Buffer` is a subclass of
 * `Uint8Array`, so the returned instances will have all the node `Buffer` methods
 * and the `Uint8Array` methods. Square bracket notation works as expected -- it
 * returns a single octet.
 *
 * The `Uint8Array` prototype remains unmodified.
 */

function Buffer (arg, encodingOrOffset, length) {
  if (!Buffer.TYPED_ARRAY_SUPPORT && !(this instanceof Buffer)) {
    return new Buffer(arg, encodingOrOffset, length)
  }

  // Common case.
  if (typeof arg === 'number') {
    if (typeof encodingOrOffset === 'string') {
      throw new Error(
        'If encoding is specified then the first argument must be a string'
      )
    }
    return allocUnsafe(this, arg)
  }
  return from(this, arg, encodingOrOffset, length)
}

Buffer.poolSize = 8192 // not used by this implementation

// TODO: Legacy, not needed anymore. Remove in next major version.
Buffer._augment = function (arr) {
  arr.__proto__ = Buffer.prototype
  return arr
}

function from (that, value, encodingOrOffset, length) {
  if (typeof value === 'number') {
    throw new TypeError('"value" argument must not be a number')
  }

  if (typeof ArrayBuffer !== 'undefined' && value instanceof ArrayBuffer) {
    return fromArrayBuffer(that, value, encodingOrOffset, length)
  }

  if (typeof value === 'string') {
    return fromString(that, value, encodingOrOffset)
  }

  return fromObject(that, value)
}

/**
 * Functionally equivalent to Buffer(arg, encoding) but throws a TypeError
 * if value is a number.
 * Buffer.from(str[, encoding])
 * Buffer.from(array)
 * Buffer.from(buffer)
 * Buffer.from(arrayBuffer[, byteOffset[, length]])
 **/
Buffer.from = function (value, encodingOrOffset, length) {
  return from(null, value, encodingOrOffset, length)
}

if (Buffer.TYPED_ARRAY_SUPPORT) {
  Buffer.prototype.__proto__ = Uint8Array.prototype
  Buffer.__proto__ = Uint8Array
  if (typeof Symbol !== 'undefined' && Symbol.species &&
      Buffer[Symbol.species] === Buffer) {
    // Fix subarray() in ES2016. See: https://github.com/feross/buffer/pull/97
    Object.defineProperty(Buffer, Symbol.species, {
      value: null,
      configurable: true
    })
  }
}

function assertSize (size) {
  if (typeof size !== 'number') {
    throw new TypeError('"size" argument must be a number')
  } else if (size < 0) {
    throw new RangeError('"size" argument must not be negative')
  }
}

function alloc (that, size, fill, encoding) {
  assertSize(size)
  if (size <= 0) {
    return createBuffer(that, size)
  }
  if (fill !== undefined) {
    // Only pay attention to encoding if it's a string. This
    // prevents accidentally sending in a number that would
    // be interpretted as a start offset.
    return typeof encoding === 'string'
      ? createBuffer(that, size).fill(fill, encoding)
      : createBuffer(that, size).fill(fill)
  }
  return createBuffer(that, size)
}

/**
 * Creates a new filled Buffer instance.
 * alloc(size[, fill[, encoding]])
 **/
Buffer.alloc = function (size, fill, encoding) {
  return alloc(null, size, fill, encoding)
}

function allocUnsafe (that, size) {
  assertSize(size)
  that = createBuffer(that, size < 0 ? 0 : checked(size) | 0)
  if (!Buffer.TYPED_ARRAY_SUPPORT) {
    for (var i = 0; i < size; ++i) {
      that[i] = 0
    }
  }
  return that
}

/**
 * Equivalent to Buffer(num), by default creates a non-zero-filled Buffer instance.
 * */
Buffer.allocUnsafe = function (size) {
  return allocUnsafe(null, size)
}
/**
 * Equivalent to SlowBuffer(num), by default creates a non-zero-filled Buffer instance.
 */
Buffer.allocUnsafeSlow = function (size) {
  return allocUnsafe(null, size)
}

function fromString (that, string, encoding) {
  if (typeof encoding !== 'string' || encoding === '') {
    encoding = 'utf8'
  }

  if (!Buffer.isEncoding(encoding)) {
    throw new TypeError('"encoding" must be a valid string encoding')
  }

  var length = byteLength(string, encoding) | 0
  that = createBuffer(that, length)

  var actual = that.write(string, encoding)

  if (actual !== length) {
    // Writing a hex string, for example, that contains invalid characters will
    // cause everything after the first invalid character to be ignored. (e.g.
    // 'abxxcd' will be treated as 'ab')
    that = that.slice(0, actual)
  }

  return that
}

function fromArrayLike (that, array) {
  var length = array.length < 0 ? 0 : checked(array.length) | 0
  that = createBuffer(that, length)
  for (var i = 0; i < length; i += 1) {
    that[i] = array[i] & 255
  }
  return that
}

function fromArrayBuffer (that, array, byteOffset, length) {
  array.byteLength // this throws if `array` is not a valid ArrayBuffer

  if (byteOffset < 0 || array.byteLength < byteOffset) {
    throw new RangeError('\'offset\' is out of bounds')
  }

  if (array.byteLength < byteOffset + (length || 0)) {
    throw new RangeError('\'length\' is out of bounds')
  }

  if (byteOffset === undefined && length === undefined) {
    array = new Uint8Array(array)
  } else if (length === undefined) {
    array = new Uint8Array(array, byteOffset)
  } else {
    array = new Uint8Array(array, byteOffset, length)
  }

  if (Buffer.TYPED_ARRAY_SUPPORT) {
    // Return an augmented `Uint8Array` instance, for best performance
    that = array
    that.__proto__ = Buffer.prototype
  } else {
    // Fallback: Return an object instance of the Buffer class
    that = fromArrayLike(that, array)
  }
  return that
}

function fromObject (that, obj) {
  if (Buffer.isBuffer(obj)) {
    var len = checked(obj.length) | 0
    that = createBuffer(that, len)

    if (that.length === 0) {
      return that
    }

    obj.copy(that, 0, 0, len)
    return that
  }

  if (obj) {
    if ((typeof ArrayBuffer !== 'undefined' &&
        obj.buffer instanceof ArrayBuffer) || 'length' in obj) {
      if (typeof obj.length !== 'number' || isnan(obj.length)) {
        return createBuffer(that, 0)
      }
      return fromArrayLike(that, obj)
    }

    if (obj.type === 'Buffer' && isArray(obj.data)) {
      return fromArrayLike(that, obj.data)
    }
  }

  throw new TypeError('First argument must be a string, Buffer, ArrayBuffer, Array, or array-like object.')
}

function checked (length) {
  // Note: cannot use `length < kMaxLength()` here because that fails when
  // length is NaN (which is otherwise coerced to zero.)
  if (length >= kMaxLength()) {
    throw new RangeError('Attempt to allocate Buffer larger than maximum ' +
                         'size: 0x' + kMaxLength().toString(16) + ' bytes')
  }
  return length | 0
}

function SlowBuffer (length) {
  if (+length != length) { // eslint-disable-line eqeqeq
    length = 0
  }
  return Buffer.alloc(+length)
}

Buffer.isBuffer = function isBuffer (b) {
  return !!(b != null && b._isBuffer)
}

Buffer.compare = function compare (a, b) {
  if (!Buffer.isBuffer(a) || !Buffer.isBuffer(b)) {
    throw new TypeError('Arguments must be Buffers')
  }

  if (a === b) return 0

  var x = a.length
  var y = b.length

  for (var i = 0, len = Math.min(x, y); i < len; ++i) {
    if (a[i] !== b[i]) {
      x = a[i]
      y = b[i]
      break
    }
  }

  if (x < y) return -1
  if (y < x) return 1
  return 0
}

Buffer.isEncoding = function isEncoding (encoding) {
  switch (String(encoding).toLowerCase()) {
    case 'hex':
    case 'utf8':
    case 'utf-8':
    case 'ascii':
    case 'latin1':
    case 'binary':
    case 'base64':
    case 'ucs2':
    case 'ucs-2':
    case 'utf16le':
    case 'utf-16le':
      return true
    default:
      return false
  }
}

Buffer.concat = function concat (list, length) {
  if (!isArray(list)) {
    throw new TypeError('"list" argument must be an Array of Buffers')
  }

  if (list.length === 0) {
    return Buffer.alloc(0)
  }

  var i
  if (length === undefined) {
    length = 0
    for (i = 0; i < list.length; ++i) {
      length += list[i].length
    }
  }

  var buffer = Buffer.allocUnsafe(length)
  var pos = 0
  for (i = 0; i < list.length; ++i) {
    var buf = list[i]
    if (!Buffer.isBuffer(buf)) {
      throw new TypeError('"list" argument must be an Array of Buffers')
    }
    buf.copy(buffer, pos)
    pos += buf.length
  }
  return buffer
}

function byteLength (string, encoding) {
  if (Buffer.isBuffer(string)) {
    return string.length
  }
  if (typeof ArrayBuffer !== 'undefined' && typeof ArrayBuffer.isView === 'function' &&
      (ArrayBuffer.isView(string) || string instanceof ArrayBuffer)) {
    return string.byteLength
  }
  if (typeof string !== 'string') {
    string = '' + string
  }

  var len = string.length
  if (len === 0) return 0

  // Use a for loop to avoid recursion
  var loweredCase = false
  for (;;) {
    switch (encoding) {
      case 'ascii':
      case 'latin1':
      case 'binary':
        return len
      case 'utf8':
      case 'utf-8':
      case undefined:
        return utf8ToBytes(string).length
      case 'ucs2':
      case 'ucs-2':
      case 'utf16le':
      case 'utf-16le':
        return len * 2
      case 'hex':
        return len >>> 1
      case 'base64':
        return base64ToBytes(string).length
      default:
        if (loweredCase) return utf8ToBytes(string).length // assume utf8
        encoding = ('' + encoding).toLowerCase()
        loweredCase = true
    }
  }
}
Buffer.byteLength = byteLength

function slowToString (encoding, start, end) {
  var loweredCase = false

  // No need to verify that "this.length <= MAX_UINT32" since it's a read-only
  // property of a typed array.

  // This behaves neither like String nor Uint8Array in that we set start/end
  // to their upper/lower bounds if the value passed is out of range.
  // undefined is handled specially as per ECMA-262 6th Edition,
  // Section 13.3.3.7 Runtime Semantics: KeyedBindingInitialization.
  if (start === undefined || start < 0) {
    start = 0
  }
  // Return early if start > this.length. Done here to prevent potential uint32
  // coercion fail below.
  if (start > this.length) {
    return ''
  }

  if (end === undefined || end > this.length) {
    end = this.length
  }

  if (end <= 0) {
    return ''
  }

  // Force coersion to uint32. This will also coerce falsey/NaN values to 0.
  end >>>= 0
  start >>>= 0

  if (end <= start) {
    return ''
  }

  if (!encoding) encoding = 'utf8'

  while (true) {
    switch (encoding) {
      case 'hex':
        return hexSlice(this, start, end)

      case 'utf8':
      case 'utf-8':
        return utf8Slice(this, start, end)

      case 'ascii':
        return asciiSlice(this, start, end)

      case 'latin1':
      case 'binary':
        return latin1Slice(this, start, end)

      case 'base64':
        return base64Slice(this, start, end)

      case 'ucs2':
      case 'ucs-2':
      case 'utf16le':
      case 'utf-16le':
        return utf16leSlice(this, start, end)

      default:
        if (loweredCase) throw new TypeError('Unknown encoding: ' + encoding)
        encoding = (encoding + '').toLowerCase()
        loweredCase = true
    }
  }
}

// The property is used by `Buffer.isBuffer` and `is-buffer` (in Safari 5-7) to detect
// Buffer instances.
Buffer.prototype._isBuffer = true

function swap (b, n, m) {
  var i = b[n]
  b[n] = b[m]
  b[m] = i
}

Buffer.prototype.swap16 = function swap16 () {
  var len = this.length
  if (len % 2 !== 0) {
    throw new RangeError('Buffer size must be a multiple of 16-bits')
  }
  for (var i = 0; i < len; i += 2) {
    swap(this, i, i + 1)
  }
  return this
}

Buffer.prototype.swap32 = function swap32 () {
  var len = this.length
  if (len % 4 !== 0) {
    throw new RangeError('Buffer size must be a multiple of 32-bits')
  }
  for (var i = 0; i < len; i += 4) {
    swap(this, i, i + 3)
    swap(this, i + 1, i + 2)
  }
  return this
}

Buffer.prototype.swap64 = function swap64 () {
  var len = this.length
  if (len % 8 !== 0) {
    throw new RangeError('Buffer size must be a multiple of 64-bits')
  }
  for (var i = 0; i < len; i += 8) {
    swap(this, i, i + 7)
    swap(this, i + 1, i + 6)
    swap(this, i + 2, i + 5)
    swap(this, i + 3, i + 4)
  }
  return this
}

Buffer.prototype.toString = function toString () {
  var length = this.length | 0
  if (length === 0) return ''
  if (arguments.length === 0) return utf8Slice(this, 0, length)
  return slowToString.apply(this, arguments)
}

Buffer.prototype.equals = function equals (b) {
  if (!Buffer.isBuffer(b)) throw new TypeError('Argument must be a Buffer')
  if (this === b) return true
  return Buffer.compare(this, b) === 0
}

Buffer.prototype.inspect = function inspect () {
  var str = ''
  var max = exports.INSPECT_MAX_BYTES
  if (this.length > 0) {
    str = this.toString('hex', 0, max).match(/.{2}/g).join(' ')
    if (this.length > max) str += ' ... '
  }
  return '<Buffer ' + str + '>'
}

Buffer.prototype.compare = function compare (target, start, end, thisStart, thisEnd) {
  if (!Buffer.isBuffer(target)) {
    throw new TypeError('Argument must be a Buffer')
  }

  if (start === undefined) {
    start = 0
  }
  if (end === undefined) {
    end = target ? target.length : 0
  }
  if (thisStart === undefined) {
    thisStart = 0
  }
  if (thisEnd === undefined) {
    thisEnd = this.length
  }

  if (start < 0 || end > target.length || thisStart < 0 || thisEnd > this.length) {
    throw new RangeError('out of range index')
  }

  if (thisStart >= thisEnd && start >= end) {
    return 0
  }
  if (thisStart >= thisEnd) {
    return -1
  }
  if (start >= end) {
    return 1
  }

  start >>>= 0
  end >>>= 0
  thisStart >>>= 0
  thisEnd >>>= 0

  if (this === target) return 0

  var x = thisEnd - thisStart
  var y = end - start
  var len = Math.min(x, y)

  var thisCopy = this.slice(thisStart, thisEnd)
  var targetCopy = target.slice(start, end)

  for (var i = 0; i < len; ++i) {
    if (thisCopy[i] !== targetCopy[i]) {
      x = thisCopy[i]
      y = targetCopy[i]
      break
    }
  }

  if (x < y) return -1
  if (y < x) return 1
  return 0
}

// Finds either the first index of `val` in `buffer` at offset >= `byteOffset`,
// OR the last index of `val` in `buffer` at offset <= `byteOffset`.
//
// Arguments:
// - buffer - a Buffer to search
// - val - a string, Buffer, or number
// - byteOffset - an index into `buffer`; will be clamped to an int32
// - encoding - an optional encoding, relevant is val is a string
// - dir - true for indexOf, false for lastIndexOf
function bidirectionalIndexOf (buffer, val, byteOffset, encoding, dir) {
  // Empty buffer means no match
  if (buffer.length === 0) return -1

  // Normalize byteOffset
  if (typeof byteOffset === 'string') {
    encoding = byteOffset
    byteOffset = 0
  } else if (byteOffset > 0x7fffffff) {
    byteOffset = 0x7fffffff
  } else if (byteOffset < -0x80000000) {
    byteOffset = -0x80000000
  }
  byteOffset = +byteOffset  // Coerce to Number.
  if (isNaN(byteOffset)) {
    // byteOffset: it it's undefined, null, NaN, "foo", etc, search whole buffer
    byteOffset = dir ? 0 : (buffer.length - 1)
  }

  // Normalize byteOffset: negative offsets start from the end of the buffer
  if (byteOffset < 0) byteOffset = buffer.length + byteOffset
  if (byteOffset >= buffer.length) {
    if (dir) return -1
    else byteOffset = buffer.length - 1
  } else if (byteOffset < 0) {
    if (dir) byteOffset = 0
    else return -1
  }

  // Normalize val
  if (typeof val === 'string') {
    val = Buffer.from(val, encoding)
  }

  // Finally, search either indexOf (if dir is true) or lastIndexOf
  if (Buffer.isBuffer(val)) {
    // Special case: looking for empty string/buffer always fails
    if (val.length === 0) {
      return -1
    }
    return arrayIndexOf(buffer, val, byteOffset, encoding, dir)
  } else if (typeof val === 'number') {
    val = val & 0xFF // Search for a byte value [0-255]
    if (Buffer.TYPED_ARRAY_SUPPORT &&
        typeof Uint8Array.prototype.indexOf === 'function') {
      if (dir) {
        return Uint8Array.prototype.indexOf.call(buffer, val, byteOffset)
      } else {
        return Uint8Array.prototype.lastIndexOf.call(buffer, val, byteOffset)
      }
    }
    return arrayIndexOf(buffer, [ val ], byteOffset, encoding, dir)
  }

  throw new TypeError('val must be string, number or Buffer')
}

function arrayIndexOf (arr, val, byteOffset, encoding, dir) {
  var indexSize = 1
  var arrLength = arr.length
  var valLength = val.length

  if (encoding !== undefined) {
    encoding = String(encoding).toLowerCase()
    if (encoding === 'ucs2' || encoding === 'ucs-2' ||
        encoding === 'utf16le' || encoding === 'utf-16le') {
      if (arr.length < 2 || val.length < 2) {
        return -1
      }
      indexSize = 2
      arrLength /= 2
      valLength /= 2
      byteOffset /= 2
    }
  }

  function read (buf, i) {
    if (indexSize === 1) {
      return buf[i]
    } else {
      return buf.readUInt16BE(i * indexSize)
    }
  }

  var i
  if (dir) {
    var foundIndex = -1
    for (i = byteOffset; i < arrLength; i++) {
      if (read(arr, i) === read(val, foundIndex === -1 ? 0 : i - foundIndex)) {
        if (foundIndex === -1) foundIndex = i
        if (i - foundIndex + 1 === valLength) return foundIndex * indexSize
      } else {
        if (foundIndex !== -1) i -= i - foundIndex
        foundIndex = -1
      }
    }
  } else {
    if (byteOffset + valLength > arrLength) byteOffset = arrLength - valLength
    for (i = byteOffset; i >= 0; i--) {
      var found = true
      for (var j = 0; j < valLength; j++) {
        if (read(arr, i + j) !== read(val, j)) {
          found = false
          break
        }
      }
      if (found) return i
    }
  }

  return -1
}

Buffer.prototype.includes = function includes (val, byteOffset, encoding) {
  return this.indexOf(val, byteOffset, encoding) !== -1
}

Buffer.prototype.indexOf = function indexOf (val, byteOffset, encoding) {
  return bidirectionalIndexOf(this, val, byteOffset, encoding, true)
}

Buffer.prototype.lastIndexOf = function lastIndexOf (val, byteOffset, encoding) {
  return bidirectionalIndexOf(this, val, byteOffset, encoding, false)
}

function hexWrite (buf, string, offset, length) {
  offset = Number(offset) || 0
  var remaining = buf.length - offset
  if (!length) {
    length = remaining
  } else {
    length = Number(length)
    if (length > remaining) {
      length = remaining
    }
  }

  // must be an even number of digits
  var strLen = string.length
  if (strLen % 2 !== 0) throw new TypeError('Invalid hex string')

  if (length > strLen / 2) {
    length = strLen / 2
  }
  for (var i = 0; i < length; ++i) {
    var parsed = parseInt(string.substr(i * 2, 2), 16)
    if (isNaN(parsed)) return i
    buf[offset + i] = parsed
  }
  return i
}

function utf8Write (buf, string, offset, length) {
  return blitBuffer(utf8ToBytes(string, buf.length - offset), buf, offset, length)
}

function asciiWrite (buf, string, offset, length) {
  return blitBuffer(asciiToBytes(string), buf, offset, length)
}

function latin1Write (buf, string, offset, length) {
  return asciiWrite(buf, string, offset, length)
}

function base64Write (buf, string, offset, length) {
  return blitBuffer(base64ToBytes(string), buf, offset, length)
}

function ucs2Write (buf, string, offset, length) {
  return blitBuffer(utf16leToBytes(string, buf.length - offset), buf, offset, length)
}

Buffer.prototype.write = function write (string, offset, length, encoding) {
  // Buffer#write(string)
  if (offset === undefined) {
    encoding = 'utf8'
    length = this.length
    offset = 0
  // Buffer#write(string, encoding)
  } else if (length === undefined && typeof offset === 'string') {
    encoding = offset
    length = this.length
    offset = 0
  // Buffer#write(string, offset[, length][, encoding])
  } else if (isFinite(offset)) {
    offset = offset | 0
    if (isFinite(length)) {
      length = length | 0
      if (encoding === undefined) encoding = 'utf8'
    } else {
      encoding = length
      length = undefined
    }
  // legacy write(string, encoding, offset, length) - remove in v0.13
  } else {
    throw new Error(
      'Buffer.write(string, encoding, offset[, length]) is no longer supported'
    )
  }

  var remaining = this.length - offset
  if (length === undefined || length > remaining) length = remaining

  if ((string.length > 0 && (length < 0 || offset < 0)) || offset > this.length) {
    throw new RangeError('Attempt to write outside buffer bounds')
  }

  if (!encoding) encoding = 'utf8'

  var loweredCase = false
  for (;;) {
    switch (encoding) {
      case 'hex':
        return hexWrite(this, string, offset, length)

      case 'utf8':
      case 'utf-8':
        return utf8Write(this, string, offset, length)

      case 'ascii':
        return asciiWrite(this, string, offset, length)

      case 'latin1':
      case 'binary':
        return latin1Write(this, string, offset, length)

      case 'base64':
        // Warning: maxLength not taken into account in base64Write
        return base64Write(this, string, offset, length)

      case 'ucs2':
      case 'ucs-2':
      case 'utf16le':
      case 'utf-16le':
        return ucs2Write(this, string, offset, length)

      default:
        if (loweredCase) throw new TypeError('Unknown encoding: ' + encoding)
        encoding = ('' + encoding).toLowerCase()
        loweredCase = true
    }
  }
}

Buffer.prototype.toJSON = function toJSON () {
  return {
    type: 'Buffer',
    data: Array.prototype.slice.call(this._arr || this, 0)
  }
}

function base64Slice (buf, start, end) {
  if (start === 0 && end === buf.length) {
    return base64.fromByteArray(buf)
  } else {
    return base64.fromByteArray(buf.slice(start, end))
  }
}

function utf8Slice (buf, start, end) {
  end = Math.min(buf.length, end)
  var res = []

  var i = start
  while (i < end) {
    var firstByte = buf[i]
    var codePoint = null
    var bytesPerSequence = (firstByte > 0xEF) ? 4
      : (firstByte > 0xDF) ? 3
      : (firstByte > 0xBF) ? 2
      : 1

    if (i + bytesPerSequence <= end) {
      var secondByte, thirdByte, fourthByte, tempCodePoint

      switch (bytesPerSequence) {
        case 1:
          if (firstByte < 0x80) {
            codePoint = firstByte
          }
          break
        case 2:
          secondByte = buf[i + 1]
          if ((secondByte & 0xC0) === 0x80) {
            tempCodePoint = (firstByte & 0x1F) << 0x6 | (secondByte & 0x3F)
            if (tempCodePoint > 0x7F) {
              codePoint = tempCodePoint
            }
          }
          break
        case 3:
          secondByte = buf[i + 1]
          thirdByte = buf[i + 2]
          if ((secondByte & 0xC0) === 0x80 && (thirdByte & 0xC0) === 0x80) {
            tempCodePoint = (firstByte & 0xF) << 0xC | (secondByte & 0x3F) << 0x6 | (thirdByte & 0x3F)
            if (tempCodePoint > 0x7FF && (tempCodePoint < 0xD800 || tempCodePoint > 0xDFFF)) {
              codePoint = tempCodePoint
            }
          }
          break
        case 4:
          secondByte = buf[i + 1]
          thirdByte = buf[i + 2]
          fourthByte = buf[i + 3]
          if ((secondByte & 0xC0) === 0x80 && (thirdByte & 0xC0) === 0x80 && (fourthByte & 0xC0) === 0x80) {
            tempCodePoint = (firstByte & 0xF) << 0x12 | (secondByte & 0x3F) << 0xC | (thirdByte & 0x3F) << 0x6 | (fourthByte & 0x3F)
            if (tempCodePoint > 0xFFFF && tempCodePoint < 0x110000) {
              codePoint = tempCodePoint
            }
          }
      }
    }

    if (codePoint === null) {
      // we did not generate a valid codePoint so insert a
      // replacement char (U+FFFD) and advance only 1 byte
      codePoint = 0xFFFD
      bytesPerSequence = 1
    } else if (codePoint > 0xFFFF) {
      // encode to utf16 (surrogate pair dance)
      codePoint -= 0x10000
      res.push(codePoint >>> 10 & 0x3FF | 0xD800)
      codePoint = 0xDC00 | codePoint & 0x3FF
    }

    res.push(codePoint)
    i += bytesPerSequence
  }

  return decodeCodePointsArray(res)
}

// Based on http://stackoverflow.com/a/22747272/680742, the browser with
// the lowest limit is Chrome, with 0x10000 args.
// We go 1 magnitude less, for safety
var MAX_ARGUMENTS_LENGTH = 0x1000

function decodeCodePointsArray (codePoints) {
  var len = codePoints.length
  if (len <= MAX_ARGUMENTS_LENGTH) {
    return String.fromCharCode.apply(String, codePoints) // avoid extra slice()
  }

  // Decode in chunks to avoid "call stack size exceeded".
  var res = ''
  var i = 0
  while (i < len) {
    res += String.fromCharCode.apply(
      String,
      codePoints.slice(i, i += MAX_ARGUMENTS_LENGTH)
    )
  }
  return res
}

function asciiSlice (buf, start, end) {
  var ret = ''
  end = Math.min(buf.length, end)

  for (var i = start; i < end; ++i) {
    ret += String.fromCharCode(buf[i] & 0x7F)
  }
  return ret
}

function latin1Slice (buf, start, end) {
  var ret = ''
  end = Math.min(buf.length, end)

  for (var i = start; i < end; ++i) {
    ret += String.fromCharCode(buf[i])
  }
  return ret
}

function hexSlice (buf, start, end) {
  var len = buf.length

  if (!start || start < 0) start = 0
  if (!end || end < 0 || end > len) end = len

  var out = ''
  for (var i = start; i < end; ++i) {
    out += toHex(buf[i])
  }
  return out
}

function utf16leSlice (buf, start, end) {
  var bytes = buf.slice(start, end)
  var res = ''
  for (var i = 0; i < bytes.length; i += 2) {
    res += String.fromCharCode(bytes[i] + bytes[i + 1] * 256)
  }
  return res
}

Buffer.prototype.slice = function slice (start, end) {
  var len = this.length
  start = ~~start
  end = end === undefined ? len : ~~end

  if (start < 0) {
    start += len
    if (start < 0) start = 0
  } else if (start > len) {
    start = len
  }

  if (end < 0) {
    end += len
    if (end < 0) end = 0
  } else if (end > len) {
    end = len
  }

  if (end < start) end = start

  var newBuf
  if (Buffer.TYPED_ARRAY_SUPPORT) {
    newBuf = this.subarray(start, end)
    newBuf.__proto__ = Buffer.prototype
  } else {
    var sliceLen = end - start
    newBuf = new Buffer(sliceLen, undefined)
    for (var i = 0; i < sliceLen; ++i) {
      newBuf[i] = this[i + start]
    }
  }

  return newBuf
}

/*
 * Need to make sure that buffer isn't trying to write out of bounds.
 */
function checkOffset (offset, ext, length) {
  if ((offset % 1) !== 0 || offset < 0) throw new RangeError('offset is not uint')
  if (offset + ext > length) throw new RangeError('Trying to access beyond buffer length')
}

Buffer.prototype.readUIntLE = function readUIntLE (offset, byteLength, noAssert) {
  offset = offset | 0
  byteLength = byteLength | 0
  if (!noAssert) checkOffset(offset, byteLength, this.length)

  var val = this[offset]
  var mul = 1
  var i = 0
  while (++i < byteLength && (mul *= 0x100)) {
    val += this[offset + i] * mul
  }

  return val
}

Buffer.prototype.readUIntBE = function readUIntBE (offset, byteLength, noAssert) {
  offset = offset | 0
  byteLength = byteLength | 0
  if (!noAssert) {
    checkOffset(offset, byteLength, this.length)
  }

  var val = this[offset + --byteLength]
  var mul = 1
  while (byteLength > 0 && (mul *= 0x100)) {
    val += this[offset + --byteLength] * mul
  }

  return val
}

Buffer.prototype.readUInt8 = function readUInt8 (offset, noAssert) {
  if (!noAssert) checkOffset(offset, 1, this.length)
  return this[offset]
}

Buffer.prototype.readUInt16LE = function readUInt16LE (offset, noAssert) {
  if (!noAssert) checkOffset(offset, 2, this.length)
  return this[offset] | (this[offset + 1] << 8)
}

Buffer.prototype.readUInt16BE = function readUInt16BE (offset, noAssert) {
  if (!noAssert) checkOffset(offset, 2, this.length)
  return (this[offset] << 8) | this[offset + 1]
}

Buffer.prototype.readUInt32LE = function readUInt32LE (offset, noAssert) {
  if (!noAssert) checkOffset(offset, 4, this.length)

  return ((this[offset]) |
      (this[offset + 1] << 8) |
      (this[offset + 2] << 16)) +
      (this[offset + 3] * 0x1000000)
}

Buffer.prototype.readUInt32BE = function readUInt32BE (offset, noAssert) {
  if (!noAssert) checkOffset(offset, 4, this.length)

  return (this[offset] * 0x1000000) +
    ((this[offset + 1] << 16) |
    (this[offset + 2] << 8) |
    this[offset + 3])
}

Buffer.prototype.readIntLE = function readIntLE (offset, byteLength, noAssert) {
  offset = offset | 0
  byteLength = byteLength | 0
  if (!noAssert) checkOffset(offset, byteLength, this.length)

  var val = this[offset]
  var mul = 1
  var i = 0
  while (++i < byteLength && (mul *= 0x100)) {
    val += this[offset + i] * mul
  }
  mul *= 0x80

  if (val >= mul) val -= Math.pow(2, 8 * byteLength)

  return val
}

Buffer.prototype.readIntBE = function readIntBE (offset, byteLength, noAssert) {
  offset = offset | 0
  byteLength = byteLength | 0
  if (!noAssert) checkOffset(offset, byteLength, this.length)

  var i = byteLength
  var mul = 1
  var val = this[offset + --i]
  while (i > 0 && (mul *= 0x100)) {
    val += this[offset + --i] * mul
  }
  mul *= 0x80

  if (val >= mul) val -= Math.pow(2, 8 * byteLength)

  return val
}

Buffer.prototype.readInt8 = function readInt8 (offset, noAssert) {
  if (!noAssert) checkOffset(offset, 1, this.length)
  if (!(this[offset] & 0x80)) return (this[offset])
  return ((0xff - this[offset] + 1) * -1)
}

Buffer.prototype.readInt16LE = function readInt16LE (offset, noAssert) {
  if (!noAssert) checkOffset(offset, 2, this.length)
  var val = this[offset] | (this[offset + 1] << 8)
  return (val & 0x8000) ? val | 0xFFFF0000 : val
}

Buffer.prototype.readInt16BE = function readInt16BE (offset, noAssert) {
  if (!noAssert) checkOffset(offset, 2, this.length)
  var val = this[offset + 1] | (this[offset] << 8)
  return (val & 0x8000) ? val | 0xFFFF0000 : val
}

Buffer.prototype.readInt32LE = function readInt32LE (offset, noAssert) {
  if (!noAssert) checkOffset(offset, 4, this.length)

  return (this[offset]) |
    (this[offset + 1] << 8) |
    (this[offset + 2] << 16) |
    (this[offset + 3] << 24)
}

Buffer.prototype.readInt32BE = function readInt32BE (offset, noAssert) {
  if (!noAssert) checkOffset(offset, 4, this.length)

  return (this[offset] << 24) |
    (this[offset + 1] << 16) |
    (this[offset + 2] << 8) |
    (this[offset + 3])
}

Buffer.prototype.readFloatLE = function readFloatLE (offset, noAssert) {
  if (!noAssert) checkOffset(offset, 4, this.length)
  return ieee754.read(this, offset, true, 23, 4)
}

Buffer.prototype.readFloatBE = function readFloatBE (offset, noAssert) {
  if (!noAssert) checkOffset(offset, 4, this.length)
  return ieee754.read(this, offset, false, 23, 4)
}

Buffer.prototype.readDoubleLE = function readDoubleLE (offset, noAssert) {
  if (!noAssert) checkOffset(offset, 8, this.length)
  return ieee754.read(this, offset, true, 52, 8)
}

Buffer.prototype.readDoubleBE = function readDoubleBE (offset, noAssert) {
  if (!noAssert) checkOffset(offset, 8, this.length)
  return ieee754.read(this, offset, false, 52, 8)
}

function checkInt (buf, value, offset, ext, max, min) {
  if (!Buffer.isBuffer(buf)) throw new TypeError('"buffer" argument must be a Buffer instance')
  if (value > max || value < min) throw new RangeError('"value" argument is out of bounds')
  if (offset + ext > buf.length) throw new RangeError('Index out of range')
}

Buffer.prototype.writeUIntLE = function writeUIntLE (value, offset, byteLength, noAssert) {
  value = +value
  offset = offset | 0
  byteLength = byteLength | 0
  if (!noAssert) {
    var maxBytes = Math.pow(2, 8 * byteLength) - 1
    checkInt(this, value, offset, byteLength, maxBytes, 0)
  }

  var mul = 1
  var i = 0
  this[offset] = value & 0xFF
  while (++i < byteLength && (mul *= 0x100)) {
    this[offset + i] = (value / mul) & 0xFF
  }

  return offset + byteLength
}

Buffer.prototype.writeUIntBE = function writeUIntBE (value, offset, byteLength, noAssert) {
  value = +value
  offset = offset | 0
  byteLength = byteLength | 0
  if (!noAssert) {
    var maxBytes = Math.pow(2, 8 * byteLength) - 1
    checkInt(this, value, offset, byteLength, maxBytes, 0)
  }

  var i = byteLength - 1
  var mul = 1
  this[offset + i] = value & 0xFF
  while (--i >= 0 && (mul *= 0x100)) {
    this[offset + i] = (value / mul) & 0xFF
  }

  return offset + byteLength
}

Buffer.prototype.writeUInt8 = function writeUInt8 (value, offset, noAssert) {
  value = +value
  offset = offset | 0
  if (!noAssert) checkInt(this, value, offset, 1, 0xff, 0)
  if (!Buffer.TYPED_ARRAY_SUPPORT) value = Math.floor(value)
  this[offset] = (value & 0xff)
  return offset + 1
}

function objectWriteUInt16 (buf, value, offset, littleEndian) {
  if (value < 0) value = 0xffff + value + 1
  for (var i = 0, j = Math.min(buf.length - offset, 2); i < j; ++i) {
    buf[offset + i] = (value & (0xff << (8 * (littleEndian ? i : 1 - i)))) >>>
      (littleEndian ? i : 1 - i) * 8
  }
}

Buffer.prototype.writeUInt16LE = function writeUInt16LE (value, offset, noAssert) {
  value = +value
  offset = offset | 0
  if (!noAssert) checkInt(this, value, offset, 2, 0xffff, 0)
  if (Buffer.TYPED_ARRAY_SUPPORT) {
    this[offset] = (value & 0xff)
    this[offset + 1] = (value >>> 8)
  } else {
    objectWriteUInt16(this, value, offset, true)
  }
  return offset + 2
}

Buffer.prototype.writeUInt16BE = function writeUInt16BE (value, offset, noAssert) {
  value = +value
  offset = offset | 0
  if (!noAssert) checkInt(this, value, offset, 2, 0xffff, 0)
  if (Buffer.TYPED_ARRAY_SUPPORT) {
    this[offset] = (value >>> 8)
    this[offset + 1] = (value & 0xff)
  } else {
    objectWriteUInt16(this, value, offset, false)
  }
  return offset + 2
}

function objectWriteUInt32 (buf, value, offset, littleEndian) {
  if (value < 0) value = 0xffffffff + value + 1
  for (var i = 0, j = Math.min(buf.length - offset, 4); i < j; ++i) {
    buf[offset + i] = (value >>> (littleEndian ? i : 3 - i) * 8) & 0xff
  }
}

Buffer.prototype.writeUInt32LE = function writeUInt32LE (value, offset, noAssert) {
  value = +value
  offset = offset | 0
  if (!noAssert) checkInt(this, value, offset, 4, 0xffffffff, 0)
  if (Buffer.TYPED_ARRAY_SUPPORT) {
    this[offset + 3] = (value >>> 24)
    this[offset + 2] = (value >>> 16)
    this[offset + 1] = (value >>> 8)
    this[offset] = (value & 0xff)
  } else {
    objectWriteUInt32(this, value, offset, true)
  }
  return offset + 4
}

Buffer.prototype.writeUInt32BE = function writeUInt32BE (value, offset, noAssert) {
  value = +value
  offset = offset | 0
  if (!noAssert) checkInt(this, value, offset, 4, 0xffffffff, 0)
  if (Buffer.TYPED_ARRAY_SUPPORT) {
    this[offset] = (value >>> 24)
    this[offset + 1] = (value >>> 16)
    this[offset + 2] = (value >>> 8)
    this[offset + 3] = (value & 0xff)
  } else {
    objectWriteUInt32(this, value, offset, false)
  }
  return offset + 4
}

Buffer.prototype.writeIntLE = function writeIntLE (value, offset, byteLength, noAssert) {
  value = +value
  offset = offset | 0
  if (!noAssert) {
    var limit = Math.pow(2, 8 * byteLength - 1)

    checkInt(this, value, offset, byteLength, limit - 1, -limit)
  }

  var i = 0
  var mul = 1
  var sub = 0
  this[offset] = value & 0xFF
  while (++i < byteLength && (mul *= 0x100)) {
    if (value < 0 && sub === 0 && this[offset + i - 1] !== 0) {
      sub = 1
    }
    this[offset + i] = ((value / mul) >> 0) - sub & 0xFF
  }

  return offset + byteLength
}

Buffer.prototype.writeIntBE = function writeIntBE (value, offset, byteLength, noAssert) {
  value = +value
  offset = offset | 0
  if (!noAssert) {
    var limit = Math.pow(2, 8 * byteLength - 1)

    checkInt(this, value, offset, byteLength, limit - 1, -limit)
  }

  var i = byteLength - 1
  var mul = 1
  var sub = 0
  this[offset + i] = value & 0xFF
  while (--i >= 0 && (mul *= 0x100)) {
    if (value < 0 && sub === 0 && this[offset + i + 1] !== 0) {
      sub = 1
    }
    this[offset + i] = ((value / mul) >> 0) - sub & 0xFF
  }

  return offset + byteLength
}

Buffer.prototype.writeInt8 = function writeInt8 (value, offset, noAssert) {
  value = +value
  offset = offset | 0
  if (!noAssert) checkInt(this, value, offset, 1, 0x7f, -0x80)
  if (!Buffer.TYPED_ARRAY_SUPPORT) value = Math.floor(value)
  if (value < 0) value = 0xff + value + 1
  this[offset] = (value & 0xff)
  return offset + 1
}

Buffer.prototype.writeInt16LE = function writeInt16LE (value, offset, noAssert) {
  value = +value
  offset = offset | 0
  if (!noAssert) checkInt(this, value, offset, 2, 0x7fff, -0x8000)
  if (Buffer.TYPED_ARRAY_SUPPORT) {
    this[offset] = (value & 0xff)
    this[offset + 1] = (value >>> 8)
  } else {
    objectWriteUInt16(this, value, offset, true)
  }
  return offset + 2
}

Buffer.prototype.writeInt16BE = function writeInt16BE (value, offset, noAssert) {
  value = +value
  offset = offset | 0
  if (!noAssert) checkInt(this, value, offset, 2, 0x7fff, -0x8000)
  if (Buffer.TYPED_ARRAY_SUPPORT) {
    this[offset] = (value >>> 8)
    this[offset + 1] = (value & 0xff)
  } else {
    objectWriteUInt16(this, value, offset, false)
  }
  return offset + 2
}

Buffer.prototype.writeInt32LE = function writeInt32LE (value, offset, noAssert) {
  value = +value
  offset = offset | 0
  if (!noAssert) checkInt(this, value, offset, 4, 0x7fffffff, -0x80000000)
  if (Buffer.TYPED_ARRAY_SUPPORT) {
    this[offset] = (value & 0xff)
    this[offset + 1] = (value >>> 8)
    this[offset + 2] = (value >>> 16)
    this[offset + 3] = (value >>> 24)
  } else {
    objectWriteUInt32(this, value, offset, true)
  }
  return offset + 4
}

Buffer.prototype.writeInt32BE = function writeInt32BE (value, offset, noAssert) {
  value = +value
  offset = offset | 0
  if (!noAssert) checkInt(this, value, offset, 4, 0x7fffffff, -0x80000000)
  if (value < 0) value = 0xffffffff + value + 1
  if (Buffer.TYPED_ARRAY_SUPPORT) {
    this[offset] = (value >>> 24)
    this[offset + 1] = (value >>> 16)
    this[offset + 2] = (value >>> 8)
    this[offset + 3] = (value & 0xff)
  } else {
    objectWriteUInt32(this, value, offset, false)
  }
  return offset + 4
}

function checkIEEE754 (buf, value, offset, ext, max, min) {
  if (offset + ext > buf.length) throw new RangeError('Index out of range')
  if (offset < 0) throw new RangeError('Index out of range')
}

function writeFloat (buf, value, offset, littleEndian, noAssert) {
  if (!noAssert) {
    checkIEEE754(buf, value, offset, 4, 3.4028234663852886e+38, -3.4028234663852886e+38)
  }
  ieee754.write(buf, value, offset, littleEndian, 23, 4)
  return offset + 4
}

Buffer.prototype.writeFloatLE = function writeFloatLE (value, offset, noAssert) {
  return writeFloat(this, value, offset, true, noAssert)
}

Buffer.prototype.writeFloatBE = function writeFloatBE (value, offset, noAssert) {
  return writeFloat(this, value, offset, false, noAssert)
}

function writeDouble (buf, value, offset, littleEndian, noAssert) {
  if (!noAssert) {
    checkIEEE754(buf, value, offset, 8, 1.7976931348623157E+308, -1.7976931348623157E+308)
  }
  ieee754.write(buf, value, offset, littleEndian, 52, 8)
  return offset + 8
}

Buffer.prototype.writeDoubleLE = function writeDoubleLE (value, offset, noAssert) {
  return writeDouble(this, value, offset, true, noAssert)
}

Buffer.prototype.writeDoubleBE = function writeDoubleBE (value, offset, noAssert) {
  return writeDouble(this, value, offset, false, noAssert)
}

// copy(targetBuffer, targetStart=0, sourceStart=0, sourceEnd=buffer.length)
Buffer.prototype.copy = function copy (target, targetStart, start, end) {
  if (!start) start = 0
  if (!end && end !== 0) end = this.length
  if (targetStart >= target.length) targetStart = target.length
  if (!targetStart) targetStart = 0
  if (end > 0 && end < start) end = start

  // Copy 0 bytes; we're done
  if (end === start) return 0
  if (target.length === 0 || this.length === 0) return 0

  // Fatal error conditions
  if (targetStart < 0) {
    throw new RangeError('targetStart out of bounds')
  }
  if (start < 0 || start >= this.length) throw new RangeError('sourceStart out of bounds')
  if (end < 0) throw new RangeError('sourceEnd out of bounds')

  // Are we oob?
  if (end > this.length) end = this.length
  if (target.length - targetStart < end - start) {
    end = target.length - targetStart + start
  }

  var len = end - start
  var i

  if (this === target && start < targetStart && targetStart < end) {
    // descending copy from end
    for (i = len - 1; i >= 0; --i) {
      target[i + targetStart] = this[i + start]
    }
  } else if (len < 1000 || !Buffer.TYPED_ARRAY_SUPPORT) {
    // ascending copy from start
    for (i = 0; i < len; ++i) {
      target[i + targetStart] = this[i + start]
    }
  } else {
    Uint8Array.prototype.set.call(
      target,
      this.subarray(start, start + len),
      targetStart
    )
  }

  return len
}

// Usage:
//    buffer.fill(number[, offset[, end]])
//    buffer.fill(buffer[, offset[, end]])
//    buffer.fill(string[, offset[, end]][, encoding])
Buffer.prototype.fill = function fill (val, start, end, encoding) {
  // Handle string cases:
  if (typeof val === 'string') {
    if (typeof start === 'string') {
      encoding = start
      start = 0
      end = this.length
    } else if (typeof end === 'string') {
      encoding = end
      end = this.length
    }
    if (val.length === 1) {
      var code = val.charCodeAt(0)
      if (code < 256) {
        val = code
      }
    }
    if (encoding !== undefined && typeof encoding !== 'string') {
      throw new TypeError('encoding must be a string')
    }
    if (typeof encoding === 'string' && !Buffer.isEncoding(encoding)) {
      throw new TypeError('Unknown encoding: ' + encoding)
    }
  } else if (typeof val === 'number') {
    val = val & 255
  }

  // Invalid ranges are not set to a default, so can range check early.
  if (start < 0 || this.length < start || this.length < end) {
    throw new RangeError('Out of range index')
  }

  if (end <= start) {
    return this
  }

  start = start >>> 0
  end = end === undefined ? this.length : end >>> 0

  if (!val) val = 0

  var i
  if (typeof val === 'number') {
    for (i = start; i < end; ++i) {
      this[i] = val
    }
  } else {
    var bytes = Buffer.isBuffer(val)
      ? val
      : utf8ToBytes(new Buffer(val, encoding).toString())
    var len = bytes.length
    for (i = 0; i < end - start; ++i) {
      this[i + start] = bytes[i % len]
    }
  }

  return this
}

// HELPER FUNCTIONS
// ================

var INVALID_BASE64_RE = /[^+\/0-9A-Za-z-_]/g

function base64clean (str) {
  // Node strips out invalid characters like \n and \t from the string, base64-js does not
  str = stringtrim(str).replace(INVALID_BASE64_RE, '')
  // Node converts strings with length < 2 to ''
  if (str.length < 2) return ''
  // Node allows for non-padded base64 strings (missing trailing ===), base64-js does not
  while (str.length % 4 !== 0) {
    str = str + '='
  }
  return str
}

function stringtrim (str) {
  if (str.trim) return str.trim()
  return str.replace(/^\s+|\s+$/g, '')
}

function toHex (n) {
  if (n < 16) return '0' + n.toString(16)
  return n.toString(16)
}

function utf8ToBytes (string, units) {
  units = units || Infinity
  var codePoint
  var length = string.length
  var leadSurrogate = null
  var bytes = []

  for (var i = 0; i < length; ++i) {
    codePoint = string.charCodeAt(i)

    // is surrogate component
    if (codePoint > 0xD7FF && codePoint < 0xE000) {
      // last char was a lead
      if (!leadSurrogate) {
        // no lead yet
        if (codePoint > 0xDBFF) {
          // unexpected trail
          if ((units -= 3) > -1) bytes.push(0xEF, 0xBF, 0xBD)
          continue
        } else if (i + 1 === length) {
          // unpaired lead
          if ((units -= 3) > -1) bytes.push(0xEF, 0xBF, 0xBD)
          continue
        }

        // valid lead
        leadSurrogate = codePoint

        continue
      }

      // 2 leads in a row
      if (codePoint < 0xDC00) {
        if ((units -= 3) > -1) bytes.push(0xEF, 0xBF, 0xBD)
        leadSurrogate = codePoint
        continue
      }

      // valid surrogate pair
      codePoint = (leadSurrogate - 0xD800 << 10 | codePoint - 0xDC00) + 0x10000
    } else if (leadSurrogate) {
      // valid bmp char, but last char was a lead
      if ((units -= 3) > -1) bytes.push(0xEF, 0xBF, 0xBD)
    }

    leadSurrogate = null

    // encode utf8
    if (codePoint < 0x80) {
      if ((units -= 1) < 0) break
      bytes.push(codePoint)
    } else if (codePoint < 0x800) {
      if ((units -= 2) < 0) break
      bytes.push(
        codePoint >> 0x6 | 0xC0,
        codePoint & 0x3F | 0x80
      )
    } else if (codePoint < 0x10000) {
      if ((units -= 3) < 0) break
      bytes.push(
        codePoint >> 0xC | 0xE0,
        codePoint >> 0x6 & 0x3F | 0x80,
        codePoint & 0x3F | 0x80
      )
    } else if (codePoint < 0x110000) {
      if ((units -= 4) < 0) break
      bytes.push(
        codePoint >> 0x12 | 0xF0,
        codePoint >> 0xC & 0x3F | 0x80,
        codePoint >> 0x6 & 0x3F | 0x80,
        codePoint & 0x3F | 0x80
      )
    } else {
      throw new Error('Invalid code point')
    }
  }

  return bytes
}

function asciiToBytes (str) {
  var byteArray = []
  for (var i = 0; i < str.length; ++i) {
    // Node's code seems to be doing this and not & 0x7F..
    byteArray.push(str.charCodeAt(i) & 0xFF)
  }
  return byteArray
}

function utf16leToBytes (str, units) {
  var c, hi, lo
  var byteArray = []
  for (var i = 0; i < str.length; ++i) {
    if ((units -= 2) < 0) break

    c = str.charCodeAt(i)
    hi = c >> 8
    lo = c % 256
    byteArray.push(lo)
    byteArray.push(hi)
  }

  return byteArray
}

function base64ToBytes (str) {
  return base64.toByteArray(base64clean(str))
}

function blitBuffer (src, dst, offset, length) {
  for (var i = 0; i < length; ++i) {
    if ((i + offset >= dst.length) || (i >= src.length)) break
    dst[i + offset] = src[i]
  }
  return i
}

function isnan (val) {
  return val !== val // eslint-disable-line no-self-compare
}

}).call(this,typeof global !== "undefined" ? global : typeof self !== "undefined" ? self : typeof window !== "undefined" ? window : {})

},{"base64-js":5,"ieee754":6,"isarray":7}],5:[function(_dereq_,module,exports){
'use strict'

exports.byteLength = byteLength
exports.toByteArray = toByteArray
exports.fromByteArray = fromByteArray

var lookup = []
var revLookup = []
var Arr = typeof Uint8Array !== 'undefined' ? Uint8Array : Array

var code = 'ABCDEFGHIJKLMNOPQRSTUVWXYZabcdefghijklmnopqrstuvwxyz0123456789+/'
for (var i = 0, len = code.length; i < len; ++i) {
  lookup[i] = code[i]
  revLookup[code.charCodeAt(i)] = i
}

revLookup['-'.charCodeAt(0)] = 62
revLookup['_'.charCodeAt(0)] = 63

function placeHoldersCount (b64) {
  var len = b64.length
  if (len % 4 > 0) {
    throw new Error('Invalid string. Length must be a multiple of 4')
  }

  // the number of equal signs (place holders)
  // if there are two placeholders, than the two characters before it
  // represent one byte
  // if there is only one, then the three characters before it represent 2 bytes
  // this is just a cheap hack to not do indexOf twice
  return b64[len - 2] === '=' ? 2 : b64[len - 1] === '=' ? 1 : 0
}

function byteLength (b64) {
  // base64 is 4/3 + up to two characters of the original data
  return b64.length * 3 / 4 - placeHoldersCount(b64)
}

function toByteArray (b64) {
  var i, j, l, tmp, placeHolders, arr
  var len = b64.length
  placeHolders = placeHoldersCount(b64)

  arr = new Arr(len * 3 / 4 - placeHolders)

  // if there are placeholders, only get up to the last complete 4 chars
  l = placeHolders > 0 ? len - 4 : len

  var L = 0

  for (i = 0, j = 0; i < l; i += 4, j += 3) {
    tmp = (revLookup[b64.charCodeAt(i)] << 18) | (revLookup[b64.charCodeAt(i + 1)] << 12) | (revLookup[b64.charCodeAt(i + 2)] << 6) | revLookup[b64.charCodeAt(i + 3)]
    arr[L++] = (tmp >> 16) & 0xFF
    arr[L++] = (tmp >> 8) & 0xFF
    arr[L++] = tmp & 0xFF
  }

  if (placeHolders === 2) {
    tmp = (revLookup[b64.charCodeAt(i)] << 2) | (revLookup[b64.charCodeAt(i + 1)] >> 4)
    arr[L++] = tmp & 0xFF
  } else if (placeHolders === 1) {
    tmp = (revLookup[b64.charCodeAt(i)] << 10) | (revLookup[b64.charCodeAt(i + 1)] << 4) | (revLookup[b64.charCodeAt(i + 2)] >> 2)
    arr[L++] = (tmp >> 8) & 0xFF
    arr[L++] = tmp & 0xFF
  }

  return arr
}

function tripletToBase64 (num) {
  return lookup[num >> 18 & 0x3F] + lookup[num >> 12 & 0x3F] + lookup[num >> 6 & 0x3F] + lookup[num & 0x3F]
}

function encodeChunk (uint8, start, end) {
  var tmp
  var output = []
  for (var i = start; i < end; i += 3) {
    tmp = (uint8[i] << 16) + (uint8[i + 1] << 8) + (uint8[i + 2])
    output.push(tripletToBase64(tmp))
  }
  return output.join('')
}

function fromByteArray (uint8) {
  var tmp
  var len = uint8.length
  var extraBytes = len % 3 // if we have 1 byte left, pad 2 bytes
  var output = ''
  var parts = []
  var maxChunkLength = 16383 // must be multiple of 3

  // go through the array every three bytes, we'll deal with trailing stuff later
  for (var i = 0, len2 = len - extraBytes; i < len2; i += maxChunkLength) {
    parts.push(encodeChunk(uint8, i, (i + maxChunkLength) > len2 ? len2 : (i + maxChunkLength)))
  }

  // pad the end with zeros, but make sure to not forget the extra bytes
  if (extraBytes === 1) {
    tmp = uint8[len - 1]
    output += lookup[tmp >> 2]
    output += lookup[(tmp << 4) & 0x3F]
    output += '=='
  } else if (extraBytes === 2) {
    tmp = (uint8[len - 2] << 8) + (uint8[len - 1])
    output += lookup[tmp >> 10]
    output += lookup[(tmp >> 4) & 0x3F]
    output += lookup[(tmp << 2) & 0x3F]
    output += '='
  }

  parts.push(output)

  return parts.join('')
}

},{}],6:[function(_dereq_,module,exports){
exports.read = function (buffer, offset, isLE, mLen, nBytes) {
  var e, m
  var eLen = nBytes * 8 - mLen - 1
  var eMax = (1 << eLen) - 1
  var eBias = eMax >> 1
  var nBits = -7
  var i = isLE ? (nBytes - 1) : 0
  var d = isLE ? -1 : 1
  var s = buffer[offset + i]

  i += d

  e = s & ((1 << (-nBits)) - 1)
  s >>= (-nBits)
  nBits += eLen
  for (; nBits > 0; e = e * 256 + buffer[offset + i], i += d, nBits -= 8) {}

  m = e & ((1 << (-nBits)) - 1)
  e >>= (-nBits)
  nBits += mLen
  for (; nBits > 0; m = m * 256 + buffer[offset + i], i += d, nBits -= 8) {}

  if (e === 0) {
    e = 1 - eBias
  } else if (e === eMax) {
    return m ? NaN : ((s ? -1 : 1) * Infinity)
  } else {
    m = m + Math.pow(2, mLen)
    e = e - eBias
  }
  return (s ? -1 : 1) * m * Math.pow(2, e - mLen)
}

exports.write = function (buffer, value, offset, isLE, mLen, nBytes) {
  var e, m, c
  var eLen = nBytes * 8 - mLen - 1
  var eMax = (1 << eLen) - 1
  var eBias = eMax >> 1
  var rt = (mLen === 23 ? Math.pow(2, -24) - Math.pow(2, -77) : 0)
  var i = isLE ? 0 : (nBytes - 1)
  var d = isLE ? 1 : -1
  var s = value < 0 || (value === 0 && 1 / value < 0) ? 1 : 0

  value = Math.abs(value)

  if (isNaN(value) || value === Infinity) {
    m = isNaN(value) ? 1 : 0
    e = eMax
  } else {
    e = Math.floor(Math.log(value) / Math.LN2)
    if (value * (c = Math.pow(2, -e)) < 1) {
      e--
      c *= 2
    }
    if (e + eBias >= 1) {
      value += rt / c
    } else {
      value += rt * Math.pow(2, 1 - eBias)
    }
    if (value * c >= 2) {
      e++
      c /= 2
    }

    if (e + eBias >= eMax) {
      m = 0
      e = eMax
    } else if (e + eBias >= 1) {
      m = (value * c - 1) * Math.pow(2, mLen)
      e = e + eBias
    } else {
      m = value * Math.pow(2, eBias - 1) * Math.pow(2, mLen)
      e = 0
    }
  }

  for (; mLen >= 8; buffer[offset + i] = m & 0xff, i += d, m /= 256, mLen -= 8) {}

  e = (e << mLen) | m
  eLen += mLen
  for (; eLen > 0; buffer[offset + i] = e & 0xff, i += d, e /= 256, eLen -= 8) {}

  buffer[offset + i - d] |= s * 128
}

},{}],7:[function(_dereq_,module,exports){
var toString = {}.toString;

module.exports = Array.isArray || function (arr) {
  return toString.call(arr) == '[object Array]';
};

},{}],8:[function(_dereq_,module,exports){
(function (process){
// Copyright Joyent, Inc. and other Node contributors.
//
// Permission is hereby granted, free of charge, to any person obtaining a
// copy of this software and associated documentation files (the
// "Software"), to deal in the Software without restriction, including
// without limitation the rights to use, copy, modify, merge, publish,
// distribute, sublicense, and/or sell copies of the Software, and to permit
// persons to whom the Software is furnished to do so, subject to the
// following conditions:
//
// The above copyright notice and this permission notice shall be included
// in all copies or substantial portions of the Software.
//
// THE SOFTWARE IS PROVIDED "AS IS", WITHOUT WARRANTY OF ANY KIND, EXPRESS
// OR IMPLIED, INCLUDING BUT NOT LIMITED TO THE WARRANTIES OF
// MERCHANTABILITY, FITNESS FOR A PARTICULAR PURPOSE AND NONINFRINGEMENT. IN
// NO EVENT SHALL THE AUTHORS OR COPYRIGHT HOLDERS BE LIABLE FOR ANY CLAIM,
// DAMAGES OR OTHER LIABILITY, WHETHER IN AN ACTION OF CONTRACT, TORT OR
// OTHERWISE, ARISING FROM, OUT OF OR IN CONNECTION WITH THE SOFTWARE OR THE
// USE OR OTHER DEALINGS IN THE SOFTWARE.

// resolves . and .. elements in a path array with directory names there
// must be no slashes, empty elements, or device names (c:\) in the array
// (so also no leading and trailing slashes - it does not distinguish
// relative and absolute paths)
function normalizeArray(parts, allowAboveRoot) {
  // if the path tries to go above the root, `up` ends up > 0
  var up = 0;
  for (var i = parts.length - 1; i >= 0; i--) {
    var last = parts[i];
    if (last === '.') {
      parts.splice(i, 1);
    } else if (last === '..') {
      parts.splice(i, 1);
      up++;
    } else if (up) {
      parts.splice(i, 1);
      up--;
    }
  }

  // if the path is allowed to go above the root, restore leading ..s
  if (allowAboveRoot) {
    for (; up--; up) {
      parts.unshift('..');
    }
  }

  return parts;
}

// Split a filename into [root, dir, basename, ext], unix version
// 'root' is just a slash, or nothing.
var splitPathRe =
    /^(\/?|)([\s\S]*?)((?:\.{1,2}|[^\/]+?|)(\.[^.\/]*|))(?:[\/]*)$/;
var splitPath = function(filename) {
  return splitPathRe.exec(filename).slice(1);
};

// path.resolve([from ...], to)
// posix version
exports.resolve = function() {
  var resolvedPath = '',
      resolvedAbsolute = false;

  for (var i = arguments.length - 1; i >= -1 && !resolvedAbsolute; i--) {
    var path = (i >= 0) ? arguments[i] : process.cwd();

    // Skip empty and invalid entries
    if (typeof path !== 'string') {
      throw new TypeError('Arguments to path.resolve must be strings');
    } else if (!path) {
      continue;
    }

    resolvedPath = path + '/' + resolvedPath;
    resolvedAbsolute = path.charAt(0) === '/';
  }

  // At this point the path should be resolved to a full absolute path, but
  // handle relative paths to be safe (might happen when process.cwd() fails)

  // Normalize the path
  resolvedPath = normalizeArray(filter(resolvedPath.split('/'), function(p) {
    return !!p;
  }), !resolvedAbsolute).join('/');

  return ((resolvedAbsolute ? '/' : '') + resolvedPath) || '.';
};

// path.normalize(path)
// posix version
exports.normalize = function(path) {
  var isAbsolute = exports.isAbsolute(path),
      trailingSlash = substr(path, -1) === '/';

  // Normalize the path
  path = normalizeArray(filter(path.split('/'), function(p) {
    return !!p;
  }), !isAbsolute).join('/');

  if (!path && !isAbsolute) {
    path = '.';
  }
  if (path && trailingSlash) {
    path += '/';
  }

  return (isAbsolute ? '/' : '') + path;
};

// posix version
exports.isAbsolute = function(path) {
  return path.charAt(0) === '/';
};

// posix version
exports.join = function() {
  var paths = Array.prototype.slice.call(arguments, 0);
  return exports.normalize(filter(paths, function(p, index) {
    if (typeof p !== 'string') {
      throw new TypeError('Arguments to path.join must be strings');
    }
    return p;
  }).join('/'));
};


// path.relative(from, to)
// posix version
exports.relative = function(from, to) {
  from = exports.resolve(from).substr(1);
  to = exports.resolve(to).substr(1);

  function trim(arr) {
    var start = 0;
    for (; start < arr.length; start++) {
      if (arr[start] !== '') break;
    }

    var end = arr.length - 1;
    for (; end >= 0; end--) {
      if (arr[end] !== '') break;
    }

    if (start > end) return [];
    return arr.slice(start, end - start + 1);
  }

  var fromParts = trim(from.split('/'));
  var toParts = trim(to.split('/'));

  var length = Math.min(fromParts.length, toParts.length);
  var samePartsLength = length;
  for (var i = 0; i < length; i++) {
    if (fromParts[i] !== toParts[i]) {
      samePartsLength = i;
      break;
    }
  }

  var outputParts = [];
  for (var i = samePartsLength; i < fromParts.length; i++) {
    outputParts.push('..');
  }

  outputParts = outputParts.concat(toParts.slice(samePartsLength));

  return outputParts.join('/');
};

exports.sep = '/';
exports.delimiter = ':';

exports.dirname = function(path) {
  var result = splitPath(path),
      root = result[0],
      dir = result[1];

  if (!root && !dir) {
    // No dirname whatsoever
    return '.';
  }

  if (dir) {
    // It has a dirname, strip trailing slash
    dir = dir.substr(0, dir.length - 1);
  }

  return root + dir;
};


exports.basename = function(path, ext) {
  var f = splitPath(path)[2];
  // TODO: make this comparison case-insensitive on windows?
  if (ext && f.substr(-1 * ext.length) === ext) {
    f = f.substr(0, f.length - ext.length);
  }
  return f;
};


exports.extname = function(path) {
  return splitPath(path)[3];
};

function filter (xs, f) {
    if (xs.filter) return xs.filter(f);
    var res = [];
    for (var i = 0; i < xs.length; i++) {
        if (f(xs[i], i, xs)) res.push(xs[i]);
    }
    return res;
}

// String.prototype.substr - negative index don't work in IE8
var substr = 'ab'.substr(-1) === 'b'
    ? function (str, start, len) { return str.substr(start, len) }
    : function (str, start, len) {
        if (start < 0) start = str.length + start;
        return str.substr(start, len);
    }
;

}).call(this,_dereq_('_process'))

},{"_process":9}],9:[function(_dereq_,module,exports){
// shim for using process in browser
var process = module.exports = {};

// cached from whatever global is present so that test runners that stub it
// don't break things.  But we need to wrap it in a try catch in case it is
// wrapped in strict mode code which doesn't define any globals.  It's inside a
// function because try/catches deoptimize in certain engines.

var cachedSetTimeout;
var cachedClearTimeout;

function defaultSetTimout() {
    throw new Error('setTimeout has not been defined');
}
function defaultClearTimeout () {
    throw new Error('clearTimeout has not been defined');
}
(function () {
    try {
        if (typeof setTimeout === 'function') {
            cachedSetTimeout = setTimeout;
        } else {
            cachedSetTimeout = defaultSetTimout;
        }
    } catch (e) {
        cachedSetTimeout = defaultSetTimout;
    }
    try {
        if (typeof clearTimeout === 'function') {
            cachedClearTimeout = clearTimeout;
        } else {
            cachedClearTimeout = defaultClearTimeout;
        }
    } catch (e) {
        cachedClearTimeout = defaultClearTimeout;
    }
} ())
function runTimeout(fun) {
    if (cachedSetTimeout === setTimeout) {
        //normal enviroments in sane situations
        return setTimeout(fun, 0);
    }
    // if setTimeout wasn't available but was latter defined
    if ((cachedSetTimeout === defaultSetTimout || !cachedSetTimeout) && setTimeout) {
        cachedSetTimeout = setTimeout;
        return setTimeout(fun, 0);
    }
    try {
        // when when somebody has screwed with setTimeout but no I.E. maddness
        return cachedSetTimeout(fun, 0);
    } catch(e){
        try {
            // When we are in I.E. but the script has been evaled so I.E. doesn't trust the global object when called normally
            return cachedSetTimeout.call(null, fun, 0);
        } catch(e){
            // same as above but when it's a version of I.E. that must have the global object for 'this', hopfully our context correct otherwise it will throw a global error
            return cachedSetTimeout.call(this, fun, 0);
        }
    }


}
function runClearTimeout(marker) {
    if (cachedClearTimeout === clearTimeout) {
        //normal enviroments in sane situations
        return clearTimeout(marker);
    }
    // if clearTimeout wasn't available but was latter defined
    if ((cachedClearTimeout === defaultClearTimeout || !cachedClearTimeout) && clearTimeout) {
        cachedClearTimeout = clearTimeout;
        return clearTimeout(marker);
    }
    try {
        // when when somebody has screwed with setTimeout but no I.E. maddness
        return cachedClearTimeout(marker);
    } catch (e){
        try {
            // When we are in I.E. but the script has been evaled so I.E. doesn't  trust the global object when called normally
            return cachedClearTimeout.call(null, marker);
        } catch (e){
            // same as above but when it's a version of I.E. that must have the global object for 'this', hopfully our context correct otherwise it will throw a global error.
            // Some versions of I.E. have different rules for clearTimeout vs setTimeout
            return cachedClearTimeout.call(this, marker);
        }
    }


}
var queue = [];
var draining = false;
var currentQueue;
var queueIndex = -1;

function cleanUpNextTick() {
    if (!draining || !currentQueue) {
        return;
    }
    draining = false;
    if (currentQueue.length) {
        queue = currentQueue.concat(queue);
    } else {
        queueIndex = -1;
    }
    if (queue.length) {
        drainQueue();
    }
}

function drainQueue() {
    if (draining) {
        return;
    }
    var timeout = runTimeout(cleanUpNextTick);
    draining = true;

    var len = queue.length;
    while(len) {
        currentQueue = queue;
        queue = [];
        while (++queueIndex < len) {
            if (currentQueue) {
                currentQueue[queueIndex].run();
            }
        }
        queueIndex = -1;
        len = queue.length;
    }
    currentQueue = null;
    draining = false;
    runClearTimeout(timeout);
}

process.nextTick = function (fun) {
    var args = new Array(arguments.length - 1);
    if (arguments.length > 1) {
        for (var i = 1; i < arguments.length; i++) {
            args[i - 1] = arguments[i];
        }
    }
    queue.push(new Item(fun, args));
    if (queue.length === 1 && !draining) {
        runTimeout(drainQueue);
    }
};

// v8 likes predictible objects
function Item(fun, array) {
    this.fun = fun;
    this.array = array;
}
Item.prototype.run = function () {
    this.fun.apply(null, this.array);
};
process.title = 'browser';
process.browser = true;
process.env = {};
process.argv = [];
process.version = ''; // empty string to avoid regexp issues
process.versions = {};

function noop() {}

process.on = noop;
process.addListener = noop;
process.once = noop;
process.off = noop;
process.removeListener = noop;
process.removeAllListeners = noop;
process.emit = noop;

process.binding = function (name) {
    throw new Error('process.binding is not supported');
};

process.cwd = function () { return '/' };
process.chdir = function (dir) {
    throw new Error('process.chdir is not supported');
};
process.umask = function() { return 0; };

},{}],10:[function(_dereq_,module,exports){
var Buffer = _dereq_('buffer').Buffer; // for use with browserify

module.exports = function (a, b) {
    if (!Buffer.isBuffer(a)) return undefined;
    if (!Buffer.isBuffer(b)) return undefined;
    if (typeof a.equals === 'function') return a.equals(b);
    if (a.length !== b.length) return false;

    for (var i = 0; i < a.length; i++) {
        if (a[i] !== b[i]) return false;
    }

    return true;
};

},{"buffer":4}],11:[function(_dereq_,module,exports){
(function (process){

/**
 * This is the web browser implementation of `debug()`.
 *
 * Expose `debug()` as the module.
 */

exports = module.exports = _dereq_('./debug');
exports.log = log;
exports.formatArgs = formatArgs;
exports.save = save;
exports.load = load;
exports.useColors = useColors;
exports.storage = 'undefined' != typeof chrome
               && 'undefined' != typeof chrome.storage
                  ? chrome.storage.local
                  : localstorage();

/**
 * Colors.
 */

exports.colors = [
  'lightseagreen',
  'forestgreen',
  'goldenrod',
  'dodgerblue',
  'darkorchid',
  'crimson'
];

/**
 * Currently only WebKit-based Web Inspectors, Firefox >= v31,
 * and the Firebug extension (any Firefox version) are known
 * to support "%c" CSS customizations.
 *
 * TODO: add a `localStorage` variable to explicitly enable/disable colors
 */

function useColors() {
  // is webkit? http://stackoverflow.com/a/16459606/376773
  // document is undefined in react-native: https://github.com/facebook/react-native/pull/1632
  return (typeof document !== 'undefined' && 'WebkitAppearance' in document.documentElement.style) ||
    // is firebug? http://stackoverflow.com/a/398120/376773
    (window.console && (console.firebug || (console.exception && console.table))) ||
    // is firefox >= v31?
    // https://developer.mozilla.org/en-US/docs/Tools/Web_Console#Styling_messages
    (navigator.userAgent.toLowerCase().match(/firefox\/(\d+)/) && parseInt(RegExp.$1, 10) >= 31);
}

/**
 * Map %j to `JSON.stringify()`, since no Web Inspectors do that by default.
 */

exports.formatters.j = function(v) {
  try {
    return JSON.stringify(v);
  } catch (err) {
    return '[UnexpectedJSONParseError]: ' + err.message;
  }
};


/**
 * Colorize log arguments if enabled.
 *
 * @api public
 */

function formatArgs() {
  var args = arguments;
  var useColors = this.useColors;

  args[0] = (useColors ? '%c' : '')
    + this.namespace
    + (useColors ? ' %c' : ' ')
    + args[0]
    + (useColors ? '%c ' : ' ')
    + '+' + exports.humanize(this.diff);

  if (!useColors) return args;

  var c = 'color: ' + this.color;
  args = [args[0], c, 'color: inherit'].concat(Array.prototype.slice.call(args, 1));

  // the final "%c" is somewhat tricky, because there could be other
  // arguments passed either before or after the %c, so we need to
  // figure out the correct index to insert the CSS into
  var index = 0;
  var lastC = 0;
  args[0].replace(/%[a-z%]/g, function(match) {
    if ('%%' === match) return;
    index++;
    if ('%c' === match) {
      // we only are interested in the *last* %c
      // (the user may have provided their own)
      lastC = index;
    }
  });

  args.splice(lastC, 0, c);
  return args;
}

/**
 * Invokes `console.log()` when available.
 * No-op when `console.log` is not a "function".
 *
 * @api public
 */

function log() {
  // this hackery is required for IE8/9, where
  // the `console.log` function doesn't have 'apply'
  return 'object' === typeof console
    && console.log
    && Function.prototype.apply.call(console.log, console, arguments);
}

/**
 * Save `namespaces`.
 *
 * @param {String} namespaces
 * @api private
 */

function save(namespaces) {
  try {
    if (null == namespaces) {
      exports.storage.removeItem('debug');
    } else {
      exports.storage.debug = namespaces;
    }
  } catch(e) {}
}

/**
 * Load `namespaces`.
 *
 * @return {String} returns the previously persisted debug modes
 * @api private
 */

function load() {
  var r;
  try {
    return exports.storage.debug;
  } catch(e) {}

  // If debug isn't set in LS, and we're in Electron, try to load $DEBUG
  if (typeof process !== 'undefined' && 'env' in process) {
    return process.env.DEBUG;
  }
}

/**
 * Enable namespaces listed in `localStorage.debug` initially.
 */

exports.enable(load());

/**
 * Localstorage attempts to return the localstorage.
 *
 * This is necessary because safari throws
 * when a user disables cookies/localstorage
 * and you attempt to access it.
 *
 * @return {LocalStorage}
 * @api private
 */

function localstorage(){
  try {
    return window.localStorage;
  } catch (e) {}
}

}).call(this,_dereq_('_process'))

},{"./debug":12,"_process":9}],12:[function(_dereq_,module,exports){

/**
 * This is the common logic for both the Node.js and web browser
 * implementations of `debug()`.
 *
 * Expose `debug()` as the module.
 */

exports = module.exports = debug.debug = debug;
exports.coerce = coerce;
exports.disable = disable;
exports.enable = enable;
exports.enabled = enabled;
exports.humanize = _dereq_('ms');

/**
 * The currently active debug mode names, and names to skip.
 */

exports.names = [];
exports.skips = [];

/**
 * Map of special "%n" handling functions, for the debug "format" argument.
 *
 * Valid key names are a single, lowercased letter, i.e. "n".
 */

exports.formatters = {};

/**
 * Previously assigned color.
 */

var prevColor = 0;

/**
 * Previous log timestamp.
 */

var prevTime;

/**
 * Select a color.
 *
 * @return {Number}
 * @api private
 */

function selectColor() {
  return exports.colors[prevColor++ % exports.colors.length];
}

/**
 * Create a debugger with the given `namespace`.
 *
 * @param {String} namespace
 * @return {Function}
 * @api public
 */

function debug(namespace) {

  // define the `disabled` version
  function disabled() {
  }
  disabled.enabled = false;

  // define the `enabled` version
  function enabled() {

    var self = enabled;

    // set `diff` timestamp
    var curr = +new Date();
    var ms = curr - (prevTime || curr);
    self.diff = ms;
    self.prev = prevTime;
    self.curr = curr;
    prevTime = curr;

    // add the `color` if not set
    if (null == self.useColors) self.useColors = exports.useColors();
    if (null == self.color && self.useColors) self.color = selectColor();

    var args = new Array(arguments.length);
    for (var i = 0; i < args.length; i++) {
      args[i] = arguments[i];
    }

    args[0] = exports.coerce(args[0]);

    if ('string' !== typeof args[0]) {
      // anything else let's inspect with %o
      args = ['%o'].concat(args);
    }

    // apply any `formatters` transformations
    var index = 0;
    args[0] = args[0].replace(/%([a-z%])/g, function(match, format) {
      // if we encounter an escaped % then don't increase the array index
      if (match === '%%') return match;
      index++;
      var formatter = exports.formatters[format];
      if ('function' === typeof formatter) {
        var val = args[index];
        match = formatter.call(self, val);

        // now we need to remove `args[index]` since it's inlined in the `format`
        args.splice(index, 1);
        index--;
      }
      return match;
    });

    // apply env-specific formatting
    args = exports.formatArgs.apply(self, args);

    var logFn = enabled.log || exports.log || console.log.bind(console);
    logFn.apply(self, args);
  }
  enabled.enabled = true;

  var fn = exports.enabled(namespace) ? enabled : disabled;

  fn.namespace = namespace;

  return fn;
}

/**
 * Enables a debug mode by namespaces. This can include modes
 * separated by a colon and wildcards.
 *
 * @param {String} namespaces
 * @api public
 */

function enable(namespaces) {
  exports.save(namespaces);

  var split = (namespaces || '').split(/[\s,]+/);
  var len = split.length;

  for (var i = 0; i < len; i++) {
    if (!split[i]) continue; // ignore empty strings
    namespaces = split[i].replace(/[\\^$+?.()|[\]{}]/g, '\\$&').replace(/\*/g, '.*?');
    if (namespaces[0] === '-') {
      exports.skips.push(new RegExp('^' + namespaces.substr(1) + '$'));
    } else {
      exports.names.push(new RegExp('^' + namespaces + '$'));
    }
  }
}

/**
 * Disable debug output.
 *
 * @api public
 */

function disable() {
  exports.enable('');
}

/**
 * Returns true if the given mode name is enabled, false otherwise.
 *
 * @param {String} name
 * @return {Boolean}
 * @api public
 */

function enabled(name) {
  var i, len;
  for (i = 0, len = exports.skips.length; i < len; i++) {
    if (exports.skips[i].test(name)) {
      return false;
    }
  }
  for (i = 0, len = exports.names.length; i < len; i++) {
    if (exports.names[i].test(name)) {
      return true;
    }
  }
  return false;
}

/**
 * Coerce `val`.
 *
 * @param {Mixed} val
 * @return {Mixed}
 * @api private
 */

function coerce(val) {
  if (val instanceof Error) return val.stack || val.message;
  return val;
}

},{"ms":13}],13:[function(_dereq_,module,exports){
/**
 * Helpers.
 */

var s = 1000
var m = s * 60
var h = m * 60
var d = h * 24
var y = d * 365.25

/**
 * Parse or format the given `val`.
 *
 * Options:
 *
 *  - `long` verbose formatting [false]
 *
 * @param {String|Number} val
 * @param {Object} options
 * @throws {Error} throw an error if val is not a non-empty string or a number
 * @return {String|Number}
 * @api public
 */

module.exports = function (val, options) {
  options = options || {}
  var type = typeof val
  if (type === 'string' && val.length > 0) {
    return parse(val)
  } else if (type === 'number' && isNaN(val) === false) {
    return options.long ?
			fmtLong(val) :
			fmtShort(val)
  }
  throw new Error('val is not a non-empty string or a valid number. val=' + JSON.stringify(val))
}

/**
 * Parse the given `str` and return milliseconds.
 *
 * @param {String} str
 * @return {Number}
 * @api private
 */

function parse(str) {
  str = String(str)
  if (str.length > 10000) {
    return
  }
  var match = /^((?:\d+)?\.?\d+) *(milliseconds?|msecs?|ms|seconds?|secs?|s|minutes?|mins?|m|hours?|hrs?|h|days?|d|years?|yrs?|y)?$/i.exec(str)
  if (!match) {
    return
  }
  var n = parseFloat(match[1])
  var type = (match[2] || 'ms').toLowerCase()
  switch (type) {
    case 'years':
    case 'year':
    case 'yrs':
    case 'yr':
    case 'y':
      return n * y
    case 'days':
    case 'day':
    case 'd':
      return n * d
    case 'hours':
    case 'hour':
    case 'hrs':
    case 'hr':
    case 'h':
      return n * h
    case 'minutes':
    case 'minute':
    case 'mins':
    case 'min':
    case 'm':
      return n * m
    case 'seconds':
    case 'second':
    case 'secs':
    case 'sec':
    case 's':
      return n * s
    case 'milliseconds':
    case 'millisecond':
    case 'msecs':
    case 'msec':
    case 'ms':
      return n
    default:
      return undefined
  }
}

/**
 * Short format for `ms`.
 *
 * @param {Number} ms
 * @return {String}
 * @api private
 */

function fmtShort(ms) {
  if (ms >= d) {
    return Math.round(ms / d) + 'd'
  }
  if (ms >= h) {
    return Math.round(ms / h) + 'h'
  }
  if (ms >= m) {
    return Math.round(ms / m) + 'm'
  }
  if (ms >= s) {
    return Math.round(ms / s) + 's'
  }
  return ms + 'ms'
}

/**
 * Long format for `ms`.
 *
 * @param {Number} ms
 * @return {String}
 * @api private
 */

function fmtLong(ms) {
  return plural(ms, d, 'day') ||
    plural(ms, h, 'hour') ||
    plural(ms, m, 'minute') ||
    plural(ms, s, 'second') ||
    ms + ' ms'
}

/**
 * Pluralization helper.
 */

function plural(ms, n, name) {
  if (ms < n) {
    return
  }
  if (ms < n * 1.5) {
    return Math.floor(ms / n) + ' ' + name
  }
  return Math.ceil(ms / n) + ' ' + name + 's'
}

},{}],14:[function(_dereq_,module,exports){
'use strict';
var isObj = _dereq_('is-obj');
var hasOwnProperty = Object.prototype.hasOwnProperty;
var propIsEnumerable = Object.prototype.propertyIsEnumerable;

function toObject(val) {
	if (val === null || val === undefined) {
		throw new TypeError('Sources cannot be null or undefined');
	}

	return Object(val);
}

function assignKey(to, from, key) {
	var val = from[key];

	if (val === undefined || val === null) {
		return;
	}

	if (hasOwnProperty.call(to, key)) {
		if (to[key] === undefined || to[key] === null) {
			throw new TypeError('Cannot convert undefined or null to object (' + key + ')');
		}
	}

	if (!hasOwnProperty.call(to, key) || !isObj(val)) {
		to[key] = val;
	} else {
		to[key] = assign(Object(to[key]), from[key]);
	}
}

function assign(to, from) {
	if (to === from) {
		return to;
	}

	from = Object(from);

	for (var key in from) {
		if (hasOwnProperty.call(from, key)) {
			assignKey(to, from, key);
		}
	}

	if (Object.getOwnPropertySymbols) {
		var symbols = Object.getOwnPropertySymbols(from);

		for (var i = 0; i < symbols.length; i++) {
			if (propIsEnumerable.call(from, symbols[i])) {
				assignKey(to, from, symbols[i]);
			}
		}
	}

	return to;
}

module.exports = function deepAssign(target) {
	target = toObject(target);

	for (var s = 1; s < arguments.length; s++) {
		assign(target, arguments[s]);
	}

	return target;
};

},{"is-obj":15}],15:[function(_dereq_,module,exports){
'use strict';
module.exports = function (x) {
	var type = typeof x;
	return x !== null && (type === 'object' || type === 'function');
};

},{}],16:[function(_dereq_,module,exports){
/*! (C) WebReflection Mit Style License */
(function(t,n,r,i){"use strict";function st(e,t){for(var n=0,r=e.length;n<r;n++)gt(e[n],t)}function ot(e){for(var t=0,n=e.length,r;t<n;t++)r=e[t],it(r,w[at(r)])}function ut(e){return function(t){F(t)&&(gt(t,e),st(t.querySelectorAll(E),e))}}function at(e){var t=R.call(e,"is"),n=e.nodeName.toUpperCase(),r=x.call(b,t?m+t.toUpperCase():v+n);return t&&-1<r&&!ft(n,t)?-1:r}function ft(e,t){return-1<E.indexOf(e+'[is="'+t+'"]')}function lt(e){var t=e.currentTarget,n=e.attrChange,r=e.attrName,i=e.target;Y&&(!i||i===t)&&t.attributeChangedCallback&&r!=="style"&&e.prevValue!==e.newValue&&t.attributeChangedCallback(r,n===e[f]?null:e.prevValue,n===e[c]?null:e.newValue)}function ct(e){var t=ut(e);return function(e){$.push(t,e.target)}}function ht(e){G&&(G=!1,e.currentTarget.removeEventListener(p,ht)),st((e.target||n).querySelectorAll(E),e.detail===u?u:o),j&&vt()}function pt(e,t){var n=this;U.call(n,e,t),Z.call(n,{target:n})}function dt(e,t){P(e,t),nt?nt.observe(e,X):(Q&&(e.setAttribute=pt,e[s]=tt(e),e.addEventListener(d,Z)),e.addEventListener(h,lt)),e.createdCallback&&Y&&(e.created=!0,e.createdCallback(),e.created=!1)}function vt(){for(var e,t=0,n=I.length;t<n;t++)e=I[t],S.contains(e)||(n--,I.splice(t--,1),gt(e,u))}function mt(e){throw new Error("A "+e+" type is already registered")}function gt(e,t){var n,r=at(e);-1<r&&(rt(e,w[r]),r=0,t===o&&!e[o]?(e[u]=!1,e[o]=!0,r=1,j&&x.call(I,e)<0&&I.push(e)):t===u&&!e[u]&&(e[o]=!1,e[u]=!0,r=1),r&&(n=e[t+"Callback"])&&n.call(e))}if(i in n)return;var s="__"+i+(Math.random()*1e5>>0),o="attached",u="detached",a="extends",f="ADDITION",l="MODIFICATION",c="REMOVAL",h="DOMAttrModified",p="DOMContentLoaded",d="DOMSubtreeModified",v="<",m="=",g=/^[A-Z][A-Z0-9]*(?:-[A-Z0-9]+)+$/,y=["ANNOTATION-XML","COLOR-PROFILE","FONT-FACE","FONT-FACE-SRC","FONT-FACE-URI","FONT-FACE-FORMAT","FONT-FACE-NAME","MISSING-GLYPH"],b=[],w=[],E="",S=n.documentElement,x=b.indexOf||function(e){for(var t=this.length;t--&&this[t]!==e;);return t},T=r.prototype,N=T.hasOwnProperty,C=T.isPrototypeOf,k=r.defineProperty,L=r.getOwnPropertyDescriptor,A=r.getOwnPropertyNames,O=r.getPrototypeOf,M=r.setPrototypeOf,_=!!r.__proto__,D=r.create||function yt(e){return e?(yt.prototype=e,new yt):this},P=M||(_?function(e,t){return e.__proto__=t,e}:A&&L?function(){function e(e,t){for(var n,r=A(t),i=0,s=r.length;i<s;i++)n=r[i],N.call(e,n)||k(e,n,L(t,n))}return function(t,n){do e(t,n);while((n=O(n))&&!C.call(n,t));return t}}():function(e,t){for(var n in t)e[n]=t[n];return e}),H=t.MutationObserver||t.WebKitMutationObserver,B=(t.HTMLElement||t.Element||t.Node).prototype,j=!C.call(B,S),F=j?function(e){return e.nodeType===1}:function(e){return C.call(B,e)},I=j&&[],q=B.cloneNode,R=B.getAttribute,U=B.setAttribute,z=B.removeAttribute,W=n.createElement,X=H&&{attributes:!0,characterData:!0,attributeOldValue:!0},V=H||function(e){Q=!1,S.removeEventListener(h,V)},$,J=t.requestAnimationFrame||t.webkitRequestAnimationFrame||t.mozRequestAnimationFrame||t.msRequestAnimationFrame||function(e){setTimeout(e,10)},K=!1,Q=!0,G=!0,Y=!0,Z,et,tt,nt,rt,it;M||_?(rt=function(e,t){C.call(t,e)||dt(e,t)},it=dt):(rt=function(e,t){e[s]||(e[s]=r(!0),dt(e,t))},it=rt),j?(Q=!1,function(){var t=L(B,"addEventListener"),n=t.value,r=function(e){var t=new CustomEvent(h,{bubbles:!0});t.attrName=e,t.prevValue=R.call(this,e),t.newValue=null,t[c]=t.attrChange=2,z.call(this,e),this.dispatchEvent(t)},i=function(t,n){var r=this.hasAttribute(t),i=r&&R.call(this,t);e=new CustomEvent(h,{bubbles:!0}),U.call(this,t,n),e.attrName=t,e.prevValue=r?i:null,e.newValue=n,r?e[l]=e.attrChange=1:e[f]=e.attrChange=0,this.dispatchEvent(e)},o=function(e){var t=e.currentTarget,n=t[s],r=e.propertyName,i;n.hasOwnProperty(r)&&(n=n[r],i=new CustomEvent(h,{bubbles:!0}),i.attrName=n.name,i.prevValue=n.value||null,i.newValue=n.value=t[r]||null,i.prevValue==null?i[f]=i.attrChange=0:i[l]=i.attrChange=1,t.dispatchEvent(i))};t.value=function(e,t,u){e===h&&this.attributeChangedCallback&&this.setAttribute!==i&&(this[s]={className:{name:"class",value:this.className}},this.setAttribute=i,this.removeAttribute=r,n.call(this,"propertychange",o)),n.call(this,e,t,u)},k(B,"addEventListener",t)}()):H||(S.addEventListener(h,V),S.setAttribute(s,1),S.removeAttribute(s),Q&&(Z=function(e){var t=this,n,r,i;if(t===e.target){n=t[s],t[s]=r=tt(t);for(i in r){if(!(i in n))return et(0,t,i,n[i],r[i],f);if(r[i]!==n[i])return et(1,t,i,n[i],r[i],l)}for(i in n)if(!(i in r))return et(2,t,i,n[i],r[i],c)}},et=function(e,t,n,r,i,s){var o={attrChange:e,currentTarget:t,attrName:n,prevValue:r,newValue:i};o[s]=e,lt(o)},tt=function(e){for(var t,n,r={},i=e.attributes,s=0,o=i.length;s<o;s++)t=i[s],n=t.name,n!=="setAttribute"&&(r[n]=t.value);return r})),n[i]=function(t,r){c=t.toUpperCase(),K||(K=!0,H?(nt=function(e,t){function n(e,t){for(var n=0,r=e.length;n<r;t(e[n++]));}return new H(function(r){for(var i,s,o,u=0,a=r.length;u<a;u++)i=r[u],i.type==="childList"?(n(i.addedNodes,e),n(i.removedNodes,t)):(s=i.target,Y&&s.attributeChangedCallback&&i.attributeName!=="style"&&(o=R.call(s,i.attributeName),o!==i.oldValue&&s.attributeChangedCallback(i.attributeName,i.oldValue,o)))})}(ut(o),ut(u)),nt.observe(n,{childList:!0,subtree:!0})):($=[],J(function d(){while($.length)$.shift().call(null,$.shift());J(d)}),n.addEventListener("DOMNodeInserted",ct(o)),n.addEventListener("DOMNodeRemoved",ct(u))),n.addEventListener(p,ht),n.addEventListener("readystatechange",ht),n.createElement=function(e,t){var r=W.apply(n,arguments),i=""+e,s=x.call(b,(t?m:v)+(t||i).toUpperCase()),o=-1<s;return t&&(r.setAttribute("is",t=t.toLowerCase()),o&&(o=ft(i.toUpperCase(),t))),Y=!n.createElement.innerHTMLHelper,o&&it(r,w[s]),r},B.cloneNode=function(e){var t=q.call(this,!!e),n=at(t);return-1<n&&it(t,w[n]),e&&ot(t.querySelectorAll(E)),t}),-2<x.call(b,m+c)+x.call(b,v+c)&&mt(t);if(!g.test(c)||-1<x.call(y,c))throw new Error("The type "+t+" is invalid");var i=function(){return f?n.createElement(l,c):n.createElement(l)},s=r||T,f=N.call(s,a),l=f?r[a].toUpperCase():c,c,h;return f&&-1<x.call(b,v+l)&&mt(l),h=b.push((f?m:v)+c)-1,E=E.concat(E.length?",":"",f?l+'[is="'+t.toLowerCase()+'"]':l),i.prototype=w[h]=N.call(s,"prototype")?s.prototype:D(B),st(n.querySelectorAll(E),o),i}})(window,document,Object,"registerElement");
},{}],17:[function(_dereq_,module,exports){
module.exports = function(dtype) {
  switch (dtype) {
    case 'int8':
      return Int8Array
    case 'int16':
      return Int16Array
    case 'int32':
      return Int32Array
    case 'uint8':
      return Uint8Array
    case 'uint16':
      return Uint16Array
    case 'uint32':
      return Uint32Array
    case 'float32':
      return Float32Array
    case 'float64':
      return Float64Array
    case 'array':
      return Array
    case 'uint8_clamped':
      return Uint8ClampedArray
  }
}

},{}],18:[function(_dereq_,module,exports){
/*eslint new-cap:0*/
var dtype = _dereq_('dtype')
module.exports = flattenVertexData
function flattenVertexData (data, output, offset) {
  if (!data) throw new TypeError('must specify data as first parameter')
  offset = +(offset || 0) | 0

  if (Array.isArray(data) && Array.isArray(data[0])) {
    var dim = data[0].length
    var length = data.length * dim

    // no output specified, create a new typed array
    if (!output || typeof output === 'string') {
      output = new (dtype(output || 'float32'))(length + offset)
    }

    var dstLength = output.length - offset
    if (length !== dstLength) {
      throw new Error('source length ' + length + ' (' + dim + 'x' + data.length + ')' +
        ' does not match destination length ' + dstLength)
    }

    for (var i = 0, k = offset; i < data.length; i++) {
      for (var j = 0; j < dim; j++) {
        output[k++] = data[i][j]
      }
    }
  } else {
    if (!output || typeof output === 'string') {
      // no output, create a new one
      var Ctor = dtype(output || 'float32')
      if (offset === 0) {
        output = new Ctor(data)
      } else {
        output = new Ctor(data.length + offset)
        output.set(data, offset)
      }
    } else {
      // store output in existing array
      output.set(data, offset)
    }
  }

  return output
}

},{"dtype":17}],19:[function(_dereq_,module,exports){
var isFunction = _dereq_('is-function')

module.exports = forEach

var toString = Object.prototype.toString
var hasOwnProperty = Object.prototype.hasOwnProperty

function forEach(list, iterator, context) {
    if (!isFunction(iterator)) {
        throw new TypeError('iterator must be a function')
    }

    if (arguments.length < 3) {
        context = this
    }

    if (toString.call(list) === '[object Array]')
        forEachArray(list, iterator, context)
    else if (typeof list === 'string')
        forEachString(list, iterator, context)
    else
        forEachObject(list, iterator, context)
}

function forEachArray(array, iterator, context) {
    for (var i = 0, len = array.length; i < len; i++) {
        if (hasOwnProperty.call(array, i)) {
            iterator.call(context, array[i], i, array)
        }
    }
}

function forEachString(string, iterator, context) {
    for (var i = 0, len = string.length; i < len; i++) {
        // no such thing as a sparse string.
        iterator.call(context, string.charAt(i), i, string)
    }
}

function forEachObject(object, iterator, context) {
    for (var k in object) {
        if (hasOwnProperty.call(object, k)) {
            iterator.call(context, object[k], k, object)
        }
    }
}

},{"is-function":24}],20:[function(_dereq_,module,exports){
(function (global){
if (typeof window !== "undefined") {
    module.exports = window;
} else if (typeof global !== "undefined") {
    module.exports = global;
} else if (typeof self !== "undefined"){
    module.exports = self;
} else {
    module.exports = {};
}

}).call(this,typeof global !== "undefined" ? global : typeof self !== "undefined" ? self : typeof window !== "undefined" ? window : {})

},{}],21:[function(_dereq_,module,exports){
module.exports = function compile(property) {
	if (!property || typeof property !== 'string')
		throw new Error('must specify property for indexof search')

	return new Function('array', 'value', 'start', [
		'start = start || 0',
		'for (var i=start; i<array.length; i++)',
		'  if (array[i]["' + property +'"] === value)',
		'      return i',
		'return -1'
	].join('\n'))
}
},{}],22:[function(_dereq_,module,exports){
if (typeof Object.create === 'function') {
  // implementation from standard node.js 'util' module
  module.exports = function inherits(ctor, superCtor) {
    ctor.super_ = superCtor
    ctor.prototype = Object.create(superCtor.prototype, {
      constructor: {
        value: ctor,
        enumerable: false,
        writable: true,
        configurable: true
      }
    });
  };
} else {
  // old school shim for old browsers
  module.exports = function inherits(ctor, superCtor) {
    ctor.super_ = superCtor
    var TempCtor = function () {}
    TempCtor.prototype = superCtor.prototype
    ctor.prototype = new TempCtor()
    ctor.prototype.constructor = ctor
  }
}

},{}],23:[function(_dereq_,module,exports){
/*!
 * Determine if an object is a Buffer
 *
 * @author   Feross Aboukhadijeh <> <http://feross.org>
 * @license  MIT
 */

// The _isBuffer check is for Safari 5-7 support, because it's missing
// Object.prototype.constructor. Remove this eventually
module.exports = function (obj) {
  return obj != null && (isBuffer(obj) || isSlowBuffer(obj) || !!obj._isBuffer)
}

function isBuffer (obj) {
  return !!obj.constructor && typeof obj.constructor.isBuffer === 'function' && obj.constructor.isBuffer(obj)
}

// For Node v0.10 support. Remove this eventually.
function isSlowBuffer (obj) {
  return typeof obj.readFloatLE === 'function' && typeof obj.slice === 'function' && isBuffer(obj.slice(0, 0))
}

},{}],24:[function(_dereq_,module,exports){
module.exports = isFunction

var toString = Object.prototype.toString

function isFunction (fn) {
  var string = toString.call(fn)
  return string === '[object Function]' ||
    (typeof fn === 'function' && string !== '[object RegExp]') ||
    (typeof window !== 'undefined' &&
     // IE8 and below
     (fn === window.setTimeout ||
      fn === window.alert ||
      fn === window.confirm ||
      fn === window.prompt))
};

},{}],25:[function(_dereq_,module,exports){
var wordWrap = _dereq_('word-wrapper')
var xtend = _dereq_('xtend')
var findChar = _dereq_('indexof-property')('id')
var number = _dereq_('as-number')

var X_HEIGHTS = ['x', 'e', 'a', 'o', 'n', 's', 'r', 'c', 'u', 'm', 'v', 'w', 'z']
var M_WIDTHS = ['m', 'w']
var CAP_HEIGHTS = ['H', 'I', 'N', 'E', 'F', 'K', 'L', 'T', 'U', 'V', 'W', 'X', 'Y', 'Z']


var TAB_ID = '\t'.charCodeAt(0)
var SPACE_ID = ' '.charCodeAt(0)
var ALIGN_LEFT = 0,
    ALIGN_CENTER = 1,
    ALIGN_RIGHT = 2

module.exports = function createLayout(opt) {
  return new TextLayout(opt)
}

function TextLayout(opt) {
  this.glyphs = []
  this._measure = this.computeMetrics.bind(this)
  this.update(opt)
}

TextLayout.prototype.update = function(opt) {
  opt = xtend({
    measure: this._measure
  }, opt)
  this._opt = opt
  this._opt.tabSize = number(this._opt.tabSize, 4)

  if (!opt.font)
    throw new Error('must provide a valid bitmap font')

  var glyphs = this.glyphs
  var text = opt.text||''
  var font = opt.font
  this._setupSpaceGlyphs(font)

  var lines = wordWrap.lines(text, opt)
  var minWidth = opt.width || 0

  //clear glyphs
  glyphs.length = 0

  //get max line width
  var maxLineWidth = lines.reduce(function(prev, line) {
    return Math.max(prev, line.width, minWidth)
  }, 0)

  //the pen position
  var x = 0
  var y = 0
  var lineHeight = number(opt.lineHeight, font.common.lineHeight)
  var baseline = font.common.base
  var descender = lineHeight-baseline
  var letterSpacing = opt.letterSpacing || 0
  var height = lineHeight * lines.length - descender
  var align = getAlignType(this._opt.align)

  //draw text along baseline
  y -= height

  //the metrics for this text layout
  this._width = maxLineWidth
  this._height = height
  this._descender = lineHeight - baseline
  this._baseline = baseline
  this._xHeight = getXHeight(font)
  this._capHeight = getCapHeight(font)
  this._lineHeight = lineHeight
  this._ascender = lineHeight - descender - this._xHeight

  //layout each glyph
  var self = this
  lines.forEach(function(line, lineIndex) {
    var start = line.start
    var end = line.end
    var lineWidth = line.width
    var lastGlyph

    //for each glyph in that line...
    for (var i=start; i<end; i++) {
      var id = text.charCodeAt(i)
      var glyph = self.getGlyph(font, id)
      if (glyph) {
        if (lastGlyph)
          x += getKerning(font, lastGlyph.id, glyph.id)

        var tx = x
        if (align === ALIGN_CENTER)
          tx += (maxLineWidth-lineWidth)/2
        else if (align === ALIGN_RIGHT)
          tx += (maxLineWidth-lineWidth)

        glyphs.push({
          position: [tx, y],
          data: glyph,
          index: i,
          line: lineIndex
        })

        //move pen forward
        x += glyph.xadvance + letterSpacing
        lastGlyph = glyph
      }
    }

    //next line down
    y += lineHeight
    x = 0
  })
  this._linesTotal = lines.length;
}

TextLayout.prototype._setupSpaceGlyphs = function(font) {
  //These are fallbacks, when the font doesn't include
  //' ' or '\t' glyphs
  this._fallbackSpaceGlyph = null
  this._fallbackTabGlyph = null

  if (!font.chars || font.chars.length === 0)
    return

  //try to get space glyph
  //then fall back to the 'm' or 'w' glyphs
  //then fall back to the first glyph available
  var space = getGlyphById(font, SPACE_ID)
          || getMGlyph(font)
          || font.chars[0]

  //and create a fallback for tab
  var tabWidth = this._opt.tabSize * space.xadvance
  this._fallbackSpaceGlyph = space
  this._fallbackTabGlyph = xtend(space, {
    x: 0, y: 0, xadvance: tabWidth, id: TAB_ID,
    xoffset: 0, yoffset: 0, width: 0, height: 0
  })
}

TextLayout.prototype.getGlyph = function(font, id) {
  var glyph = getGlyphById(font, id)
  if (glyph)
    return glyph
  else if (id === TAB_ID)
    return this._fallbackTabGlyph
  else if (id === SPACE_ID)
    return this._fallbackSpaceGlyph
  return null
}

TextLayout.prototype.computeMetrics = function(text, start, end, width) {
  var letterSpacing = this._opt.letterSpacing || 0
  var font = this._opt.font
  var curPen = 0
  var curWidth = 0
  var count = 0
  var glyph
  var lastGlyph

  if (!font.chars || font.chars.length === 0) {
    return {
      start: start,
      end: start,
      width: 0
    }
  }

  end = Math.min(text.length, end)
  for (var i=start; i < end; i++) {
    var id = text.charCodeAt(i)
    var glyph = this.getGlyph(font, id)

    if (glyph) {
      //move pen forward
      var xoff = glyph.xoffset
      var kern = lastGlyph ? getKerning(font, lastGlyph.id, glyph.id) : 0
      curPen += kern

      var nextPen = curPen + glyph.xadvance + letterSpacing
      var nextWidth = curPen + glyph.width

      //we've hit our limit; we can't move onto the next glyph
      if (nextWidth >= width || nextPen >= width)
        break

      //otherwise continue along our line
      curPen = nextPen
      curWidth = nextWidth
      lastGlyph = glyph
    }
    count++
  }

  //make sure rightmost edge lines up with rendered glyphs
  if (lastGlyph)
    curWidth += lastGlyph.xoffset

  return {
    start: start,
    end: start + count,
    width: curWidth
  }
}

//getters for the private vars
;['width', 'height',
  'descender', 'ascender',
  'xHeight', 'baseline',
  'capHeight',
  'lineHeight' ].forEach(addGetter)

function addGetter(name) {
  Object.defineProperty(TextLayout.prototype, name, {
    get: wrapper(name),
    configurable: true
  })
}

//create lookups for private vars
function wrapper(name) {
  return (new Function([
    'return function '+name+'() {',
    '  return this._'+name,
    '}'
  ].join('\n')))()
}

function getGlyphById(font, id) {
  if (!font.chars || font.chars.length === 0)
    return null

  var glyphIdx = findChar(font.chars, id)
  if (glyphIdx >= 0)
    return font.chars[glyphIdx]
  return null
}

function getXHeight(font) {
  for (var i=0; i<X_HEIGHTS.length; i++) {
    var id = X_HEIGHTS[i].charCodeAt(0)
    var idx = findChar(font.chars, id)
    if (idx >= 0)
      return font.chars[idx].height
  }
  return 0
}

function getMGlyph(font) {
  for (var i=0; i<M_WIDTHS.length; i++) {
    var id = M_WIDTHS[i].charCodeAt(0)
    var idx = findChar(font.chars, id)
    if (idx >= 0)
      return font.chars[idx]
  }
  return 0
}

function getCapHeight(font) {
  for (var i=0; i<CAP_HEIGHTS.length; i++) {
    var id = CAP_HEIGHTS[i].charCodeAt(0)
    var idx = findChar(font.chars, id)
    if (idx >= 0)
      return font.chars[idx].height
  }
  return 0
}

function getKerning(font, left, right) {
  if (!font.kernings || font.kernings.length === 0)
    return 0

  var table = font.kernings
  for (var i=0; i<table.length; i++) {
    var kern = table[i]
    if (kern.first === left && kern.second === right)
      return kern.amount
  }
  return 0
}

function getAlignType(align) {
  if (align === 'center')
    return ALIGN_CENTER
  else if (align === 'right')
    return ALIGN_RIGHT
  return ALIGN_LEFT
}
},{"as-number":2,"indexof-property":21,"word-wrapper":51,"xtend":54}],26:[function(_dereq_,module,exports){
(function (Buffer){
var xhr = _dereq_('xhr')
var noop = function(){}
var parseASCII = _dereq_('parse-bmfont-ascii')
var parseXML = _dereq_('parse-bmfont-xml')
var readBinary = _dereq_('parse-bmfont-binary')
var isBinaryFormat = _dereq_('./lib/is-binary')
var xtend = _dereq_('xtend')

var xml2 = (function hasXML2() {
  return self.XMLHttpRequest && "withCredentials" in new XMLHttpRequest
})()

module.exports = function(opt, cb) {
  cb = typeof cb === 'function' ? cb : noop

  if (typeof opt === 'string')
    opt = { uri: opt }
  else if (!opt)
    opt = {}

  var expectBinary = opt.binary
  if (expectBinary)
    opt = getBinaryOpts(opt)

  xhr(opt, function(err, res, body) {
    if (err)
      return cb(err)
    if (!/^2/.test(res.statusCode))
      return cb(new Error('http status code: '+res.statusCode))
    if (!body)
      return cb(new Error('no body result'))

    var binary = false

    //if the response type is an array buffer,
    //we need to convert it into a regular Buffer object
    if (isArrayBuffer(body)) {
      var array = new Uint8Array(body)
      body = new Buffer(array, 'binary')
    }

    //now check the string/Buffer response
    //and see if it has a binary BMF header
    if (isBinaryFormat(body)) {
      binary = true
      //if we have a string, turn it into a Buffer
      if (typeof body === 'string')
        body = new Buffer(body, 'binary')
    }

    //we are not parsing a binary format, just ASCII/XML/etc
    if (!binary) {
      //might still be a buffer if responseType is 'arraybuffer'
      if (Buffer.isBuffer(body))
        body = body.toString(opt.encoding)
      body = body.trim()
    }

    var result
    try {
      var type = res.headers['content-type']
      if (binary)
        result = readBinary(body)
      else if (/json/.test(type) || body.charAt(0) === '{')
        result = JSON.parse(body)
      else if (/xml/.test(type)  || body.charAt(0) === '<')
        result = parseXML(body)
      else
        result = parseASCII(body)
    } catch (e) {
      cb(new Error('error parsing font '+e.message))
      cb = noop
    }
    cb(null, result)
  })
}

function isArrayBuffer(arr) {
  var str = Object.prototype.toString
  return str.call(arr) === '[object ArrayBuffer]'
}

function getBinaryOpts(opt) {
  //IE10+ and other modern browsers support array buffers
  if (xml2)
    return xtend(opt, { responseType: 'arraybuffer' })

  if (typeof self.XMLHttpRequest === 'undefined')
    throw new Error('your browser does not support XHR loading')

  //IE9 and XML1 browsers could still use an override
  var req = new self.XMLHttpRequest()
  req.overrideMimeType('text/plain; charset=x-user-defined')
  return xtend({
    xhr: req
  }, opt)
}

}).call(this,_dereq_("buffer").Buffer)

},{"./lib/is-binary":27,"buffer":4,"parse-bmfont-ascii":29,"parse-bmfont-binary":30,"parse-bmfont-xml":31,"xhr":52,"xtend":54}],27:[function(_dereq_,module,exports){
(function (Buffer){
var equal = _dereq_('buffer-equal')
var HEADER = new Buffer([66, 77, 70, 3])

module.exports = function(buf) {
  if (typeof buf === 'string')
    return buf.substring(0, 3) === 'BMF'
  return buf.length > 4 && equal(buf.slice(0, 4), HEADER)
}
}).call(this,_dereq_("buffer").Buffer)

},{"buffer":4,"buffer-equal":10}],28:[function(_dereq_,module,exports){
'use strict';
/* eslint-disable no-unused-vars */
var hasOwnProperty = Object.prototype.hasOwnProperty;
var propIsEnumerable = Object.prototype.propertyIsEnumerable;

function toObject(val) {
	if (val === null || val === undefined) {
		throw new TypeError('Object.assign cannot be called with null or undefined');
	}

	return Object(val);
}

function shouldUseNative() {
	try {
		if (!Object.assign) {
			return false;
		}

		// Detect buggy property enumeration order in older V8 versions.

		// https://bugs.chromium.org/p/v8/issues/detail?id=4118
		var test1 = new String('abc');  // eslint-disable-line
		test1[5] = 'de';
		if (Object.getOwnPropertyNames(test1)[0] === '5') {
			return false;
		}

		// https://bugs.chromium.org/p/v8/issues/detail?id=3056
		var test2 = {};
		for (var i = 0; i < 10; i++) {
			test2['_' + String.fromCharCode(i)] = i;
		}
		var order2 = Object.getOwnPropertyNames(test2).map(function (n) {
			return test2[n];
		});
		if (order2.join('') !== '0123456789') {
			return false;
		}

		// https://bugs.chromium.org/p/v8/issues/detail?id=3056
		var test3 = {};
		'abcdefghijklmnopqrst'.split('').forEach(function (letter) {
			test3[letter] = letter;
		});
		if (Object.keys(Object.assign({}, test3)).join('') !==
				'abcdefghijklmnopqrst') {
			return false;
		}

		return true;
	} catch (e) {
		// We don't expect any of the above to throw, but better to be safe.
		return false;
	}
}

module.exports = shouldUseNative() ? Object.assign : function (target, source) {
	var from;
	var to = toObject(target);
	var symbols;

	for (var s = 1; s < arguments.length; s++) {
		from = Object(arguments[s]);

		for (var key in from) {
			if (hasOwnProperty.call(from, key)) {
				to[key] = from[key];
			}
		}

		if (Object.getOwnPropertySymbols) {
			symbols = Object.getOwnPropertySymbols(from);
			for (var i = 0; i < symbols.length; i++) {
				if (propIsEnumerable.call(from, symbols[i])) {
					to[symbols[i]] = from[symbols[i]];
				}
			}
		}
	}

	return to;
};

},{}],29:[function(_dereq_,module,exports){
module.exports = function parseBMFontAscii(data) {
  if (!data)
    throw new Error('no data provided')
  data = data.toString().trim()

  var output = {
    pages: [],
    chars: [],
    kernings: []
  }

  var lines = data.split(/\r\n?|\n/g)

  if (lines.length === 0)
    throw new Error('no data in BMFont file')

  for (var i = 0; i < lines.length; i++) {
    var lineData = splitLine(lines[i], i)
    if (!lineData) //skip empty lines
      continue

    if (lineData.key === 'page') {
      if (typeof lineData.data.id !== 'number')
        throw new Error('malformed file at line ' + i + ' -- needs page id=N')
      if (typeof lineData.data.file !== 'string')
        throw new Error('malformed file at line ' + i + ' -- needs page file="path"')
      output.pages[lineData.data.id] = lineData.data.file
    } else if (lineData.key === 'chars' || lineData.key === 'kernings') {
      //... do nothing for these two ...
    } else if (lineData.key === 'char') {
      output.chars.push(lineData.data)
    } else if (lineData.key === 'kerning') {
      output.kernings.push(lineData.data)
    } else {
      output[lineData.key] = lineData.data
    }
  }

  return output
}

function splitLine(line, idx) {
  line = line.replace(/\t+/g, ' ').trim()
  if (!line)
    return null

  var space = line.indexOf(' ')
  if (space === -1)
    throw new Error("no named row at line " + idx)

  var key = line.substring(0, space)

  line = line.substring(space + 1)
  //clear "letter" field as it is non-standard and
  //requires additional complexity to parse " / = symbols
  line = line.replace(/letter=[\'\"]\S+[\'\"]/gi, '')
  line = line.split("=")
  line = line.map(function(str) {
    return str.trim().match((/(".*?"|[^"\s]+)+(?=\s*|\s*$)/g))
  })

  var data = []
  for (var i = 0; i < line.length; i++) {
    var dt = line[i]
    if (i === 0) {
      data.push({
        key: dt[0],
        data: ""
      })
    } else if (i === line.length - 1) {
      data[data.length - 1].data = parseData(dt[0])
    } else {
      data[data.length - 1].data = parseData(dt[0])
      data.push({
        key: dt[1],
        data: ""
      })
    }
  }

  var out = {
    key: key,
    data: {}
  }

  data.forEach(function(v) {
    out.data[v.key] = v.data;
  })

  return out
}

function parseData(data) {
  if (!data || data.length === 0)
    return ""

  if (data.indexOf('"') === 0 || data.indexOf("'") === 0)
    return data.substring(1, data.length - 1)
  if (data.indexOf(',') !== -1)
    return parseIntList(data)
  return parseInt(data, 10)
}

function parseIntList(data) {
  return data.split(',').map(function(val) {
    return parseInt(val, 10)
  })
}
},{}],30:[function(_dereq_,module,exports){
var HEADER = [66, 77, 70]

module.exports = function readBMFontBinary(buf) {
  if (buf.length < 6)
    throw new Error('invalid buffer length for BMFont')

  var header = HEADER.every(function(byte, i) {
    return buf.readUInt8(i) === byte
  })

  if (!header)
    throw new Error('BMFont missing BMF byte header')

  var i = 3
  var vers = buf.readUInt8(i++)
  if (vers > 3)
    throw new Error('Only supports BMFont Binary v3 (BMFont App v1.10)')

  var target = { kernings: [], chars: [] }
  for (var b=0; b<5; b++)
    i += readBlock(target, buf, i)
  return target
}

function readBlock(target, buf, i) {
  if (i > buf.length-1)
    return 0

  var blockID = buf.readUInt8(i++)
  var blockSize = buf.readInt32LE(i)
  i += 4

  switch(blockID) {
    case 1:
      target.info = readInfo(buf, i)
      break
    case 2:
      target.common = readCommon(buf, i)
      break
    case 3:
      target.pages = readPages(buf, i, blockSize)
      break
    case 4:
      target.chars = readChars(buf, i, blockSize)
      break
    case 5:
      target.kernings = readKernings(buf, i, blockSize)
      break
  }
  return 5 + blockSize
}

function readInfo(buf, i) {
  var info = {}
  info.size = buf.readInt16LE(i)

  var bitField = buf.readUInt8(i+2)
  info.smooth = (bitField >> 7) & 1
  info.unicode = (bitField >> 6) & 1
  info.italic = (bitField >> 5) & 1
  info.bold = (bitField >> 4) & 1

  //fixedHeight is only mentioned in binary spec
  if ((bitField >> 3) & 1)
    info.fixedHeight = 1

  info.charset = buf.readUInt8(i+3) || ''
  info.stretchH = buf.readUInt16LE(i+4)
  info.aa = buf.readUInt8(i+6)
  info.padding = [
    buf.readInt8(i+7),
    buf.readInt8(i+8),
    buf.readInt8(i+9),
    buf.readInt8(i+10)
  ]
  info.spacing = [
    buf.readInt8(i+11),
    buf.readInt8(i+12)
  ]
  info.outline = buf.readUInt8(i+13)
  info.face = readStringNT(buf, i+14)
  return info
}

function readCommon(buf, i) {
  var common = {}
  common.lineHeight = buf.readUInt16LE(i)
  common.base = buf.readUInt16LE(i+2)
  common.scaleW = buf.readUInt16LE(i+4)
  common.scaleH = buf.readUInt16LE(i+6)
  common.pages = buf.readUInt16LE(i+8)
  var bitField = buf.readUInt8(i+10)
  common.packed = 0
  common.alphaChnl = buf.readUInt8(i+11)
  common.redChnl = buf.readUInt8(i+12)
  common.greenChnl = buf.readUInt8(i+13)
  common.blueChnl = buf.readUInt8(i+14)
  return common
}

function readPages(buf, i, size) {
  var pages = []
  var text = readNameNT(buf, i)
  var len = text.length+1
  var count = size / len
  for (var c=0; c<count; c++) {
    pages[c] = buf.slice(i, i+text.length).toString('utf8')
    i += len
  }
  return pages
}

function readChars(buf, i, blockSize) {
  var chars = []

  var count = blockSize / 20
  for (var c=0; c<count; c++) {
    var char = {}
    var off = c*20
    char.id = buf.readUInt32LE(i + 0 + off)
    char.x = buf.readUInt16LE(i + 4 + off)
    char.y = buf.readUInt16LE(i + 6 + off)
    char.width = buf.readUInt16LE(i + 8 + off)
    char.height = buf.readUInt16LE(i + 10 + off)
    char.xoffset = buf.readInt16LE(i + 12 + off)
    char.yoffset = buf.readInt16LE(i + 14 + off)
    char.xadvance = buf.readInt16LE(i + 16 + off)
    char.page = buf.readUInt8(i + 18 + off)
    char.chnl = buf.readUInt8(i + 19 + off)
    chars[c] = char
  }
  return chars
}

function readKernings(buf, i, blockSize) {
  var kernings = []
  var count = blockSize / 10
  for (var c=0; c<count; c++) {
    var kern = {}
    var off = c*10
    kern.first = buf.readUInt32LE(i + 0 + off)
    kern.second = buf.readUInt32LE(i + 4 + off)
    kern.amount = buf.readInt16LE(i + 8 + off)
    kernings[c] = kern
  }
  return kernings
}

function readNameNT(buf, offset) {
  var pos=offset
  for (; pos<buf.length; pos++) {
    if (buf[pos] === 0x00)
      break
  }
  return buf.slice(offset, pos)
}

function readStringNT(buf, offset) {
  return readNameNT(buf, offset).toString('utf8')
}
},{}],31:[function(_dereq_,module,exports){
var parseAttributes = _dereq_('./parse-attribs')
var parseFromString = _dereq_('xml-parse-from-string')

//In some cases element.attribute.nodeName can return
//all lowercase values.. so we need to map them to the correct
//case
var NAME_MAP = {
  scaleh: 'scaleH',
  scalew: 'scaleW',
  stretchh: 'stretchH',
  lineheight: 'lineHeight',
  alphachnl: 'alphaChnl',
  redchnl: 'redChnl',
  greenchnl: 'greenChnl',
  bluechnl: 'blueChnl'
}

module.exports = function parse(data) {
  data = data.toString()

  var xmlRoot = parseFromString(data)
  var output = {
    pages: [],
    chars: [],
    kernings: []
  }

  //get config settings
  ;['info', 'common'].forEach(function(key) {
    var element = xmlRoot.getElementsByTagName(key)[0]
    if (element)
      output[key] = parseAttributes(getAttribs(element))
  })

  //get page info
  var pageRoot = xmlRoot.getElementsByTagName('pages')[0]
  if (!pageRoot)
    throw new Error('malformed file -- no <pages> element')
  var pages = pageRoot.getElementsByTagName('page')
  for (var i=0; i<pages.length; i++) {
    var p = pages[i]
    var id = parseInt(p.getAttribute('id'), 10)
    var file = p.getAttribute('file')
    if (isNaN(id))
      throw new Error('malformed file -- page "id" attribute is NaN')
    if (!file)
      throw new Error('malformed file -- needs page "file" attribute')
    output.pages[parseInt(id, 10)] = file
  }

  //get kernings / chars
  ;['chars', 'kernings'].forEach(function(key) {
    var element = xmlRoot.getElementsByTagName(key)[0]
    if (!element)
      return
    var childTag = key.substring(0, key.length-1)
    var children = element.getElementsByTagName(childTag)
    for (var i=0; i<children.length; i++) {
      var child = children[i]
      output[key].push(parseAttributes(getAttribs(child)))
    }
  })
  return output
}

function getAttribs(element) {
  var attribs = getAttribList(element)
  return attribs.reduce(function(dict, attrib) {
    var key = mapName(attrib.nodeName)
    dict[key] = attrib.nodeValue
    return dict
  }, {})
}

function getAttribList(element) {
  //IE8+ and modern browsers
  var attribs = []
  for (var i=0; i<element.attributes.length; i++)
    attribs.push(element.attributes[i])
  return attribs
}

function mapName(nodeName) {
  return NAME_MAP[nodeName.toLowerCase()] || nodeName
}
},{"./parse-attribs":32,"xml-parse-from-string":53}],32:[function(_dereq_,module,exports){
//Some versions of GlyphDesigner have a typo
//that causes some bugs with parsing.
//Need to confirm with recent version of the software
//to see whether this is still an issue or not.
var GLYPH_DESIGNER_ERROR = 'chasrset'

module.exports = function parseAttributes(obj) {
  if (GLYPH_DESIGNER_ERROR in obj) {
    obj['charset'] = obj[GLYPH_DESIGNER_ERROR]
    delete obj[GLYPH_DESIGNER_ERROR]
  }

  for (var k in obj) {
    if (k === 'face' || k === 'charset')
      continue
    else if (k === 'padding' || k === 'spacing')
      obj[k] = parseIntList(obj[k])
    else
      obj[k] = parseInt(obj[k], 10)
  }
  return obj
}

function parseIntList(data) {
  return data.split(',').map(function(val) {
    return parseInt(val, 10)
  })
}
},{}],33:[function(_dereq_,module,exports){
var trim = _dereq_('trim')
  , forEach = _dereq_('for-each')
  , isArray = function(arg) {
      return Object.prototype.toString.call(arg) === '[object Array]';
    }

module.exports = function (headers) {
  if (!headers)
    return {}

  var result = {}

  forEach(
      trim(headers).split('\n')
    , function (row) {
        var index = row.indexOf(':')
          , key = trim(row.slice(0, index)).toLowerCase()
          , value = trim(row.slice(index + 1))

        if (typeof(result[key]) === 'undefined') {
          result[key] = value
        } else if (isArray(result[key])) {
          result[key].push(value)
        } else {
          result[key] = [ result[key], value ]
        }
      }
  )

  return result
}
},{"for-each":19,"trim":48}],34:[function(_dereq_,module,exports){
(function (global){
var performance = global.performance || {};

var present = (function () {
  var names = ['now', 'webkitNow', 'msNow', 'mozNow', 'oNow'];
  while (names.length) {
    var name = names.shift();
    if (name in performance) {
      return performance[name].bind(performance);
    }
  }

  var dateNow = Date.now || function () { return new Date().getTime(); };
  var navigationStart = (performance.timing || {}).navigationStart || dateNow();
  return function () {
    return dateNow() - navigationStart;
  };
}());

present.performanceNow = performance.now;
present.noConflict = function () {
  performance.now = present.performanceNow;
};
present.conflict = function () {
  performance.now = present;
};
present.conflict();

module.exports = present;

}).call(this,typeof global !== "undefined" ? global : typeof self !== "undefined" ? self : typeof window !== "undefined" ? window : {})

},{}],35:[function(_dereq_,module,exports){
(function(root) {

	// Store setTimeout reference so promise-polyfill will be unaffected by
	// other code modifying setTimeout (like sinon.useFakeTimers())
	var setTimeoutFunc = setTimeout;

	// Use polyfill for setImmediate for performance gains
	var asap = (typeof setImmediate === 'function' && setImmediate) ||
		function(fn) { setTimeoutFunc(fn, 1); };

	// Polyfill for Function.prototype.bind
	function bind(fn, thisArg) {
		return function() {
			fn.apply(thisArg, arguments);
		}
	}

	var isArray = Array.isArray || function(value) { return Object.prototype.toString.call(value) === "[object Array]" };

	function Promise(fn) {
		if (typeof this !== 'object') throw new TypeError('Promises must be constructed via new');
		if (typeof fn !== 'function') throw new TypeError('not a function');
		this._state = null;
		this._value = null;
		this._deferreds = []

		doResolve(fn, bind(resolve, this), bind(reject, this))
	}

	function handle(deferred) {
		var me = this;
		if (this._state === null) {
			this._deferreds.push(deferred);
			return
		}
		asap(function() {
			var cb = me._state ? deferred.onFulfilled : deferred.onRejected
			if (cb === null) {
				(me._state ? deferred.resolve : deferred.reject)(me._value);
				return;
			}
			var ret;
			try {
				ret = cb(me._value);
			}
			catch (e) {
				deferred.reject(e);
				return;
			}
			deferred.resolve(ret);
		})
	}

	function resolve(newValue) {
		try { //Promise Resolution Procedure: https://github.com/promises-aplus/promises-spec#the-promise-resolution-procedure
			if (newValue === this) throw new TypeError('A promise cannot be resolved with itself.');
			if (newValue && (typeof newValue === 'object' || typeof newValue === 'function')) {
				var then = newValue.then;
				if (typeof then === 'function') {
					doResolve(bind(then, newValue), bind(resolve, this), bind(reject, this));
					return;
				}
			}
			this._state = true;
			this._value = newValue;
			finale.call(this);
		} catch (e) { reject.call(this, e); }
	}

	function reject(newValue) {
		this._state = false;
		this._value = newValue;
		finale.call(this);
	}

	function finale() {
		for (var i = 0, len = this._deferreds.length; i < len; i++) {
			handle.call(this, this._deferreds[i]);
		}
		this._deferreds = null;
	}

	function Handler(onFulfilled, onRejected, resolve, reject){
		this.onFulfilled = typeof onFulfilled === 'function' ? onFulfilled : null;
		this.onRejected = typeof onRejected === 'function' ? onRejected : null;
		this.resolve = resolve;
		this.reject = reject;
	}

	/**
	 * Take a potentially misbehaving resolver function and make sure
	 * onFulfilled and onRejected are only called once.
	 *
	 * Makes no guarantees about asynchrony.
	 */
	function doResolve(fn, onFulfilled, onRejected) {
		var done = false;
		try {
			fn(function (value) {
				if (done) return;
				done = true;
				onFulfilled(value);
			}, function (reason) {
				if (done) return;
				done = true;
				onRejected(reason);
			})
		} catch (ex) {
			if (done) return;
			done = true;
			onRejected(ex);
		}
	}

	Promise.prototype['catch'] = function (onRejected) {
		return this.then(null, onRejected);
	};

	Promise.prototype.then = function(onFulfilled, onRejected) {
		var me = this;
		return new Promise(function(resolve, reject) {
			handle.call(me, new Handler(onFulfilled, onRejected, resolve, reject));
		})
	};

	Promise.all = function () {
		var args = Array.prototype.slice.call(arguments.length === 1 && isArray(arguments[0]) ? arguments[0] : arguments);

		return new Promise(function (resolve, reject) {
			if (args.length === 0) return resolve([]);
			var remaining = args.length;
			function res(i, val) {
				try {
					if (val && (typeof val === 'object' || typeof val === 'function')) {
						var then = val.then;
						if (typeof then === 'function') {
							then.call(val, function (val) { res(i, val) }, reject);
							return;
						}
					}
					args[i] = val;
					if (--remaining === 0) {
						resolve(args);
					}
				} catch (ex) {
					reject(ex);
				}
			}
			for (var i = 0; i < args.length; i++) {
				res(i, args[i]);
			}
		});
	};

	Promise.resolve = function (value) {
		if (value && typeof value === 'object' && value.constructor === Promise) {
			return value;
		}

		return new Promise(function (resolve) {
			resolve(value);
		});
	};

	Promise.reject = function (value) {
		return new Promise(function (resolve, reject) {
			reject(value);
		});
	};

	Promise.race = function (values) {
		return new Promise(function (resolve, reject) {
			for(var i = 0, len = values.length; i < len; i++) {
				values[i].then(resolve, reject);
			}
		});
	};

	/**
	 * Set the immediate function to execute callbacks
	 * @param fn {function} Function to execute
	 * @private
	 */
	Promise._setImmediateFn = function _setImmediateFn(fn) {
		asap = fn;
	};

	if (typeof module !== 'undefined' && module.exports) {
		module.exports = Promise;
	} else if (!root.Promise) {
		root.Promise = Promise;
	}

})(this);

},{}],36:[function(_dereq_,module,exports){
var dtype = _dereq_('dtype')
var anArray = _dereq_('an-array')
var isBuffer = _dereq_('is-buffer')

var CW = [0, 2, 3]
var CCW = [2, 1, 3]

module.exports = function createQuadElements(array, opt) {
    //if user didn't specify an output array
    if (!array || !(anArray(array) || isBuffer(array))) {
        opt = array || {}
        array = null
    }

    if (typeof opt === 'number') //backwards-compatible
        opt = { count: opt }
    else
        opt = opt || {}

    var type = typeof opt.type === 'string' ? opt.type : 'uint16'
    var count = typeof opt.count === 'number' ? opt.count : 1
    var start = (opt.start || 0)

    var dir = opt.clockwise !== false ? CW : CCW,
        a = dir[0],
        b = dir[1],
        c = dir[2]

    var numIndices = count * 6

    var indices = array || new (dtype(type))(numIndices)
    for (var i = 0, j = 0; i < numIndices; i += 6, j += 4) {
        var x = i + start
        indices[x + 0] = j + 0
        indices[x + 1] = j + 1
        indices[x + 2] = j + 2
        indices[x + 3] = j + a
        indices[x + 4] = j + b
        indices[x + 5] = j + c
    }
    return indices
}
},{"an-array":1,"dtype":17,"is-buffer":23}],37:[function(_dereq_,module,exports){


/*:: type Attr = { [key: string]: string } */

/*

style-attr
====

Very simple parsing and stringifying of style attributes.

`parse`
----

Convert a style attribute string to an object.

*/

/*:: declare function parse (raw: string): Attr */
function parse(raw) {
  var trim = function (s) {
    return s.trim();
  };
  var obj = {};

  getKeyValueChunks(raw).map(trim).filter(Boolean).forEach(function (item) {
    // split with `.indexOf` rather than `.split` because the value may also contain colons.
    var pos = item.indexOf(':');
    var key = item.substr(0, pos).trim();
    var val = item.substr(pos + 1).trim();

    obj[key] = val;
  });

  return obj;
}

/*

`getKeyValueChunks`
----

Split a string into chunks matching `<key>: <value>`

*/
/*:: declare function getKeyValueChunks (raw: string): Array<string> */
function getKeyValueChunks(raw) {
  var chunks = [];
  var offset = 0;
  var sep = ';';
  var hasUnclosedUrl = /url\([^\)]+$/;
  var chunk = '';
  var nextSplit;
  while (offset < raw.length) {
    nextSplit = raw.indexOf(sep, offset);
    if (nextSplit === -1) {
      nextSplit = raw.length;
    }

    chunk += raw.substring(offset, nextSplit);

    // data URIs can contain semicolons, so make sure we get the whole thing
    if (hasUnclosedUrl.test(chunk)) {
      chunk += ';';
      offset = nextSplit + 1;
      continue;
    }

    chunks.push(chunk);
    chunk = '';
    offset = nextSplit + 1;
  }

  return chunks;
}

/*

`stringify`
----

Convert an object into an attribute string

*/
/*:: declare function stringify (obj: Attr): string */
function stringify(obj) {
  return Object.keys(obj).map(function (key) {
    return key + ':' + obj[key];
  }).join(';');
}

/*

`normalize`
----

Normalize an attribute string (eg. collapse duplicates)

*/
/*:: declare function normalize (str: string): string */
function normalize(str) {
  return stringify(parse(str));
}

module.exports.parse = parse;
module.exports.stringify = stringify;
module.exports.normalize = normalize;
},{}],38:[function(_dereq_,module,exports){
var createLayout = _dereq_('layout-bmfont-text')
var inherits = _dereq_('inherits')
var createIndices = _dereq_('quad-indices')
var buffer = _dereq_('three-buffer-vertex-data')
var assign = _dereq_('object-assign')

var vertices = _dereq_('./lib/vertices')
var utils = _dereq_('./lib/utils')

var Base = THREE.BufferGeometry

module.exports = function createTextGeometry (opt) {
  return new TextGeometry(opt)
}

function TextGeometry (opt) {
  Base.call(this)

  if (typeof opt === 'string') {
    opt = { text: opt }
  }

  // use these as default values for any subsequent
  // calls to update()
  this._opt = assign({}, opt)

  // also do an initial setup...
  if (opt) this.update(opt)
}

inherits(TextGeometry, Base)

TextGeometry.prototype.update = function (opt) {
  if (typeof opt === 'string') {
    opt = { text: opt }
  }

  // use constructor defaults
  opt = assign({}, this._opt, opt)

  if (!opt.font) {
    throw new TypeError('must specify a { font } in options')
  }

  this.layout = createLayout(opt)

  // get vec2 texcoords
  var flipY = opt.flipY !== false

  // the desired BMFont data
  var font = opt.font

  // determine texture size from font file
  var texWidth = font.common.scaleW
  var texHeight = font.common.scaleH

  // get visible glyphs
  var glyphs = this.layout.glyphs.filter(function (glyph) {
    var bitmap = glyph.data
    return bitmap.width * bitmap.height > 0
  })

  // provide visible glyphs for convenience
  this.visibleGlyphs = glyphs

  // get common vertex data
  var positions = vertices.positions(glyphs)
  var uvs = vertices.uvs(glyphs, texWidth, texHeight, flipY)
  var indices = createIndices({
    clockwise: true,
    type: 'uint16',
    count: glyphs.length
  })

  // update vertex data
  buffer.index(this, indices, 1, 'uint16')
  buffer.attr(this, 'position', positions, 2)
  buffer.attr(this, 'uv', uvs, 2)

  // update multipage data
  if (!opt.multipage && 'page' in this.attributes) {
    // disable multipage rendering
    this.removeAttribute('page')
  } else if (opt.multipage) {
    var pages = vertices.pages(glyphs)
    // enable multipage rendering
    buffer.attr(this, 'page', pages, 1)
  }
}

TextGeometry.prototype.computeBoundingSphere = function () {
  if (this.boundingSphere === null) {
    this.boundingSphere = new THREE.Sphere()
  }

  var positions = this.attributes.position.array
  var itemSize = this.attributes.position.itemSize
  if (!positions || !itemSize || positions.length < 2) {
    this.boundingSphere.radius = 0
    this.boundingSphere.center.set(0, 0, 0)
    return
  }
  utils.computeSphere(positions, this.boundingSphere)
  if (isNaN(this.boundingSphere.radius)) {
    console.error('THREE.BufferGeometry.computeBoundingSphere(): ' +
      'Computed radius is NaN. The ' +
      '"position" attribute is likely to have NaN values.')
  }
}

TextGeometry.prototype.computeBoundingBox = function () {
  if (this.boundingBox === null) {
    this.boundingBox = new THREE.Box3()
  }

  var bbox = this.boundingBox
  var positions = this.attributes.position.array
  var itemSize = this.attributes.position.itemSize
  if (!positions || !itemSize || positions.length < 2) {
    bbox.makeEmpty()
    return
  }
  utils.computeBox(positions, bbox)
}

},{"./lib/utils":39,"./lib/vertices":40,"inherits":22,"layout-bmfont-text":25,"object-assign":28,"quad-indices":36,"three-buffer-vertex-data":41}],39:[function(_dereq_,module,exports){
var itemSize = 2
var box = { min: [0, 0], max: [0, 0] }

function bounds (positions) {
  var count = positions.length / itemSize
  box.min[0] = positions[0]
  box.min[1] = positions[1]
  box.max[0] = positions[0]
  box.max[1] = positions[1]

  for (var i = 0; i < count; i++) {
    var x = positions[i * itemSize + 0]
    var y = positions[i * itemSize + 1]
    box.min[0] = Math.min(x, box.min[0])
    box.min[1] = Math.min(y, box.min[1])
    box.max[0] = Math.max(x, box.max[0])
    box.max[1] = Math.max(y, box.max[1])
  }
}

module.exports.computeBox = function (positions, output) {
  bounds(positions)
  output.min.set(box.min[0], box.min[1], 0)
  output.max.set(box.max[0], box.max[1], 0)
}

module.exports.computeSphere = function (positions, output) {
  bounds(positions)
  var minX = box.min[0]
  var minY = box.min[1]
  var maxX = box.max[0]
  var maxY = box.max[1]
  var width = maxX - minX
  var height = maxY - minY
  var length = Math.sqrt(width * width + height * height)
  output.center.set(minX + width / 2, minY + height / 2, 0)
  output.radius = length / 2
}

},{}],40:[function(_dereq_,module,exports){
module.exports.pages = function pages (glyphs) {
  var pages = new Float32Array(glyphs.length * 4 * 1)
  var i = 0
  glyphs.forEach(function (glyph) {
    var id = glyph.data.page || 0
    pages[i++] = id
    pages[i++] = id
    pages[i++] = id
    pages[i++] = id
  })
  return pages
}

module.exports.uvs = function uvs (glyphs, texWidth, texHeight, flipY) {
  var uvs = new Float32Array(glyphs.length * 4 * 2)
  var i = 0
  glyphs.forEach(function (glyph) {
    var bitmap = glyph.data
    var bw = (bitmap.x + bitmap.width)
    var bh = (bitmap.y + bitmap.height)

    // top left position
    var u0 = bitmap.x / texWidth
    var v1 = bitmap.y / texHeight
    var u1 = bw / texWidth
    var v0 = bh / texHeight

    if (flipY) {
      v1 = (texHeight - bitmap.y) / texHeight
      v0 = (texHeight - bh) / texHeight
    }

    // BL
    uvs[i++] = u0
    uvs[i++] = v1
    // TL
    uvs[i++] = u0
    uvs[i++] = v0
    // TR
    uvs[i++] = u1
    uvs[i++] = v0
    // BR
    uvs[i++] = u1
    uvs[i++] = v1
  })
  return uvs
}

module.exports.positions = function positions (glyphs) {
  var positions = new Float32Array(glyphs.length * 4 * 2)
  var i = 0
  glyphs.forEach(function (glyph) {
    var bitmap = glyph.data

    // bottom left position
    var x = glyph.position[0] + bitmap.xoffset
    var y = glyph.position[1] + bitmap.yoffset

    // quad size
    var w = bitmap.width
    var h = bitmap.height

    // BL
    positions[i++] = x
    positions[i++] = y
    // TL
    positions[i++] = x
    positions[i++] = y + h
    // TR
    positions[i++] = x + w
    positions[i++] = y + h
    // BR
    positions[i++] = x + w
    positions[i++] = y
  })
  return positions
}

},{}],41:[function(_dereq_,module,exports){
var flatten = _dereq_('flatten-vertex-data')
var warned = false;

module.exports.attr = setAttribute
module.exports.index = setIndex

function setIndex (geometry, data, itemSize, dtype) {
  if (typeof itemSize !== 'number') itemSize = 1
  if (typeof dtype !== 'string') dtype = 'uint16'

  var isR69 = !geometry.index && typeof geometry.setIndex !== 'function'
  var attrib = isR69 ? geometry.getAttribute('index') : geometry.index
  var newAttrib = updateAttribute(attrib, data, itemSize, dtype)
  if (newAttrib) {
    if (isR69) geometry.addAttribute('index', newAttrib)
    else geometry.index = newAttrib
  }
}

function setAttribute (geometry, key, data, itemSize, dtype) {
  if (typeof itemSize !== 'number') itemSize = 3
  if (typeof dtype !== 'string') dtype = 'float32'
  if (Array.isArray(data) &&
    Array.isArray(data[0]) &&
    data[0].length !== itemSize) {
    throw new Error('Nested vertex array has unexpected size; expected ' +
      itemSize + ' but found ' + data[0].length)
  }

  var attrib = geometry.getAttribute(key)
  var newAttrib = updateAttribute(attrib, data, itemSize, dtype)
  if (newAttrib) {
    geometry.addAttribute(key, newAttrib)
  }
}

function updateAttribute (attrib, data, itemSize, dtype) {
  data = data || []
  if (!attrib || rebuildAttribute(attrib, data, itemSize)) {
    // create a new array with desired type
    data = flatten(data, dtype)

    var needsNewBuffer = attrib && typeof attrib.setArray !== 'function'
    if (!attrib || needsNewBuffer) {
      // We are on an old version of ThreeJS which can't
      // support growing / shrinking buffers, so we need
      // to build a new buffer
      if (needsNewBuffer && !warned) {
        warned = true
        console.warn([
          'A WebGL buffer is being updated with a new size or itemSize, ',
          'however this version of ThreeJS only supports fixed-size buffers.',
          '\nThe old buffer may still be kept in memory.\n',
          'To avoid memory leaks, it is recommended that you dispose ',
          'your geometries and create new ones, or update to ThreeJS r82 or newer.\n',
          'See here for discussion:\n',
          'https://github.com/mrdoob/three.js/pull/9631'
        ].join(''))
      }

      // Build a new attribute
      attrib = new THREE.BufferAttribute(data, itemSize);
    }

    attrib.itemSize = itemSize
    attrib.needsUpdate = true

    // New versions of ThreeJS suggest using setArray
    // to change the data. It will use bufferData internally,
    // so you can change the array size without any issues
    if (typeof attrib.setArray === 'function') {
      attrib.setArray(data)
    }

    return attrib
  } else {
    // copy data into the existing array
    flatten(data, attrib.array)
    attrib.needsUpdate = true
    return null
  }
}

// Test whether the attribute needs to be re-created,
// returns false if we can re-use it as-is.
function rebuildAttribute (attrib, data, itemSize) {
  if (attrib.itemSize !== itemSize) return true
  if (!attrib.array) return true
  var attribLength = attrib.array.length
  if (Array.isArray(data) && Array.isArray(data[0])) {
    // [ [ x, y, z ] ]
    return attribLength !== data.length * itemSize
  } else {
    // [ x, y, z ]
    return attribLength !== data.length
  }
  return false
}

},{"flatten-vertex-data":18}],42:[function(_dereq_,module,exports){
(function (global, factory) {
	typeof exports === 'object' && typeof module !== 'undefined' ? factory(exports) :
	typeof define === 'function' && define.amd ? define(['exports'], factory) :
	(factory((global.THREE = global.THREE || {})));
}(this, (function (exports) { 'use strict';

	// Polyfills

	if ( Number.EPSILON === undefined ) {

		Number.EPSILON = Math.pow( 2, - 52 );

	}

	//

	if ( Math.sign === undefined ) {

		// https://developer.mozilla.org/en-US/docs/Web/JavaScript/Reference/Global_Objects/Math/sign

		Math.sign = function ( x ) {

			return ( x < 0 ) ? - 1 : ( x > 0 ) ? 1 : + x;

		};

	}

	if ( Function.prototype.name === undefined ) {

		// Missing in IE9-11.
		// https://developer.mozilla.org/en-US/docs/Web/JavaScript/Reference/Global_Objects/Function/name

		Object.defineProperty( Function.prototype, 'name', {

			get: function () {

				return this.toString().match( /^\s*function\s*([^\(\s]*)/ )[ 1 ];

			}

		} );

	}

	if ( Object.assign === undefined ) {

		// Missing in IE.
		// https://developer.mozilla.org/en-US/docs/Web/JavaScript/Reference/Global_Objects/Object/assign

		( function () {

			Object.assign = function ( target ) {

				'use strict';

				if ( target === undefined || target === null ) {

					throw new TypeError( 'Cannot convert undefined or null to object' );

				}

				var output = Object( target );

				for ( var index = 1; index < arguments.length; index ++ ) {

					var source = arguments[ index ];

					if ( source !== undefined && source !== null ) {

						for ( var nextKey in source ) {

							if ( Object.prototype.hasOwnProperty.call( source, nextKey ) ) {

								output[ nextKey ] = source[ nextKey ];

							}

						}

					}

				}

				return output;

			};

		} )();

	}

	/**
	 * https://github.com/mrdoob/eventdispatcher.js/
	 */

	function EventDispatcher() {}

	Object.assign( EventDispatcher.prototype, {

		addEventListener: function ( type, listener ) {

			if ( this._listeners === undefined ) this._listeners = {};

			var listeners = this._listeners;

			if ( listeners[ type ] === undefined ) {

				listeners[ type ] = [];

			}

			if ( listeners[ type ].indexOf( listener ) === - 1 ) {

				listeners[ type ].push( listener );

			}

		},

		hasEventListener: function ( type, listener ) {

			if ( this._listeners === undefined ) return false;

			var listeners = this._listeners;

			return listeners[ type ] !== undefined && listeners[ type ].indexOf( listener ) !== - 1;

		},

		removeEventListener: function ( type, listener ) {

			if ( this._listeners === undefined ) return;

			var listeners = this._listeners;
			var listenerArray = listeners[ type ];

			if ( listenerArray !== undefined ) {

				var index = listenerArray.indexOf( listener );

				if ( index !== - 1 ) {

					listenerArray.splice( index, 1 );

				}

			}

		},

		dispatchEvent: function ( event ) {

			if ( this._listeners === undefined ) return;

			var listeners = this._listeners;
			var listenerArray = listeners[ event.type ];

			if ( listenerArray !== undefined ) {

				event.target = this;

				var array = [], i = 0;
				var length = listenerArray.length;

				for ( i = 0; i < length; i ++ ) {

					array[ i ] = listenerArray[ i ];

				}

				for ( i = 0; i < length; i ++ ) {

					array[ i ].call( this, event );

				}

			}

		}

	} );

	var REVISION = '83';
	var MOUSE = { LEFT: 0, MIDDLE: 1, RIGHT: 2 };
	var CullFaceNone = 0;
	var CullFaceBack = 1;
	var CullFaceFront = 2;
	var CullFaceFrontBack = 3;
	var FrontFaceDirectionCW = 0;
	var FrontFaceDirectionCCW = 1;
	var BasicShadowMap = 0;
	var PCFShadowMap = 1;
	var PCFSoftShadowMap = 2;
	var FrontSide = 0;
	var BackSide = 1;
	var DoubleSide = 2;
	var FlatShading = 1;
	var SmoothShading = 2;
	var NoColors = 0;
	var FaceColors = 1;
	var VertexColors = 2;
	var NoBlending = 0;
	var NormalBlending = 1;
	var AdditiveBlending = 2;
	var SubtractiveBlending = 3;
	var MultiplyBlending = 4;
	var CustomBlending = 5;
	var BlendingMode = {
		NoBlending: NoBlending,
		NormalBlending: NormalBlending,
		AdditiveBlending: AdditiveBlending,
		SubtractiveBlending: SubtractiveBlending,
		MultiplyBlending: MultiplyBlending,
		CustomBlending: CustomBlending
	};
	var AddEquation = 100;
	var SubtractEquation = 101;
	var ReverseSubtractEquation = 102;
	var MinEquation = 103;
	var MaxEquation = 104;
	var ZeroFactor = 200;
	var OneFactor = 201;
	var SrcColorFactor = 202;
	var OneMinusSrcColorFactor = 203;
	var SrcAlphaFactor = 204;
	var OneMinusSrcAlphaFactor = 205;
	var DstAlphaFactor = 206;
	var OneMinusDstAlphaFactor = 207;
	var DstColorFactor = 208;
	var OneMinusDstColorFactor = 209;
	var SrcAlphaSaturateFactor = 210;
	var NeverDepth = 0;
	var AlwaysDepth = 1;
	var LessDepth = 2;
	var LessEqualDepth = 3;
	var EqualDepth = 4;
	var GreaterEqualDepth = 5;
	var GreaterDepth = 6;
	var NotEqualDepth = 7;
	var MultiplyOperation = 0;
	var MixOperation = 1;
	var AddOperation = 2;
	var NoToneMapping = 0;
	var LinearToneMapping = 1;
	var ReinhardToneMapping = 2;
	var Uncharted2ToneMapping = 3;
	var CineonToneMapping = 4;
	var UVMapping = 300;
	var CubeReflectionMapping = 301;
	var CubeRefractionMapping = 302;
	var EquirectangularReflectionMapping = 303;
	var EquirectangularRefractionMapping = 304;
	var SphericalReflectionMapping = 305;
	var CubeUVReflectionMapping = 306;
	var CubeUVRefractionMapping = 307;
	var TextureMapping = {
		UVMapping: UVMapping,
		CubeReflectionMapping: CubeReflectionMapping,
		CubeRefractionMapping: CubeRefractionMapping,
		EquirectangularReflectionMapping: EquirectangularReflectionMapping,
		EquirectangularRefractionMapping: EquirectangularRefractionMapping,
		SphericalReflectionMapping: SphericalReflectionMapping,
		CubeUVReflectionMapping: CubeUVReflectionMapping,
		CubeUVRefractionMapping: CubeUVRefractionMapping
	};
	var RepeatWrapping = 1000;
	var ClampToEdgeWrapping = 1001;
	var MirroredRepeatWrapping = 1002;
	var TextureWrapping = {
		RepeatWrapping: RepeatWrapping,
		ClampToEdgeWrapping: ClampToEdgeWrapping,
		MirroredRepeatWrapping: MirroredRepeatWrapping
	};
	var NearestFilter = 1003;
	var NearestMipMapNearestFilter = 1004;
	var NearestMipMapLinearFilter = 1005;
	var LinearFilter = 1006;
	var LinearMipMapNearestFilter = 1007;
	var LinearMipMapLinearFilter = 1008;
	var TextureFilter = {
		NearestFilter: NearestFilter,
		NearestMipMapNearestFilter: NearestMipMapNearestFilter,
		NearestMipMapLinearFilter: NearestMipMapLinearFilter,
		LinearFilter: LinearFilter,
		LinearMipMapNearestFilter: LinearMipMapNearestFilter,
		LinearMipMapLinearFilter: LinearMipMapLinearFilter
	};
	var UnsignedByteType = 1009;
	var ByteType = 1010;
	var ShortType = 1011;
	var UnsignedShortType = 1012;
	var IntType = 1013;
	var UnsignedIntType = 1014;
	var FloatType = 1015;
	var HalfFloatType = 1016;
	var UnsignedShort4444Type = 1017;
	var UnsignedShort5551Type = 1018;
	var UnsignedShort565Type = 1019;
	var UnsignedInt248Type = 1020;
	var AlphaFormat = 1021;
	var RGBFormat = 1022;
	var RGBAFormat = 1023;
	var LuminanceFormat = 1024;
	var LuminanceAlphaFormat = 1025;
	var RGBEFormat = RGBAFormat;
	var DepthFormat = 1026;
	var DepthStencilFormat = 1027;
	var RGB_S3TC_DXT1_Format = 2001;
	var RGBA_S3TC_DXT1_Format = 2002;
	var RGBA_S3TC_DXT3_Format = 2003;
	var RGBA_S3TC_DXT5_Format = 2004;
	var RGB_PVRTC_4BPPV1_Format = 2100;
	var RGB_PVRTC_2BPPV1_Format = 2101;
	var RGBA_PVRTC_4BPPV1_Format = 2102;
	var RGBA_PVRTC_2BPPV1_Format = 2103;
	var RGB_ETC1_Format = 2151;
	var LoopOnce = 2200;
	var LoopRepeat = 2201;
	var LoopPingPong = 2202;
	var InterpolateDiscrete = 2300;
	var InterpolateLinear = 2301;
	var InterpolateSmooth = 2302;
	var ZeroCurvatureEnding = 2400;
	var ZeroSlopeEnding = 2401;
	var WrapAroundEnding = 2402;
	var TrianglesDrawMode = 0;
	var TriangleStripDrawMode = 1;
	var TriangleFanDrawMode = 2;
	var LinearEncoding = 3000;
	var sRGBEncoding = 3001;
	var GammaEncoding = 3007;
	var RGBEEncoding = 3002;
	var LogLuvEncoding = 3003;
	var RGBM7Encoding = 3004;
	var RGBM16Encoding = 3005;
	var RGBDEncoding = 3006;
	var BasicDepthPacking = 3200;
	var RGBADepthPacking = 3201;

	/**
	 * @author alteredq / http://alteredqualia.com/
	 * @author mrdoob / http://mrdoob.com/
	 */

	var _Math = {

		DEG2RAD: Math.PI / 180,
		RAD2DEG: 180 / Math.PI,

		generateUUID: function () {

			// http://www.broofa.com/Tools/Math.uuid.htm

			var chars = '0123456789ABCDEFGHIJKLMNOPQRSTUVWXYZabcdefghijklmnopqrstuvwxyz'.split( '' );
			var uuid = new Array( 36 );
			var rnd = 0, r;

			return function generateUUID() {

				for ( var i = 0; i < 36; i ++ ) {

					if ( i === 8 || i === 13 || i === 18 || i === 23 ) {

						uuid[ i ] = '-';

					} else if ( i === 14 ) {

						uuid[ i ] = '4';

					} else {

						if ( rnd <= 0x02 ) rnd = 0x2000000 + ( Math.random() * 0x1000000 ) | 0;
						r = rnd & 0xf;
						rnd = rnd >> 4;
						uuid[ i ] = chars[ ( i === 19 ) ? ( r & 0x3 ) | 0x8 : r ];

					}

				}

				return uuid.join( '' );

			};

		}(),

		clamp: function ( value, min, max ) {

			return Math.max( min, Math.min( max, value ) );

		},

		// compute euclidian modulo of m % n
		// https://en.wikipedia.org/wiki/Modulo_operation

		euclideanModulo: function ( n, m ) {

			return ( ( n % m ) + m ) % m;

		},

		// Linear mapping from range <a1, a2> to range <b1, b2>

		mapLinear: function ( x, a1, a2, b1, b2 ) {

			return b1 + ( x - a1 ) * ( b2 - b1 ) / ( a2 - a1 );

		},

		// https://en.wikipedia.org/wiki/Linear_interpolation

		lerp: function ( x, y, t ) {

			return ( 1 - t ) * x + t * y;

		},

		// http://en.wikipedia.org/wiki/Smoothstep

		smoothstep: function ( x, min, max ) {

			if ( x <= min ) return 0;
			if ( x >= max ) return 1;

			x = ( x - min ) / ( max - min );

			return x * x * ( 3 - 2 * x );

		},

		smootherstep: function ( x, min, max ) {

			if ( x <= min ) return 0;
			if ( x >= max ) return 1;

			x = ( x - min ) / ( max - min );

			return x * x * x * ( x * ( x * 6 - 15 ) + 10 );

		},

		// Random integer from <low, high> interval

		randInt: function ( low, high ) {

			return low + Math.floor( Math.random() * ( high - low + 1 ) );

		},

		// Random float from <low, high> interval

		randFloat: function ( low, high ) {

			return low + Math.random() * ( high - low );

		},

		// Random float from <-range/2, range/2> interval

		randFloatSpread: function ( range ) {

			return range * ( 0.5 - Math.random() );

		},

		degToRad: function ( degrees ) {

			return degrees * _Math.DEG2RAD;

		},

		radToDeg: function ( radians ) {

			return radians * _Math.RAD2DEG;

		},

		isPowerOfTwo: function ( value ) {

			return ( value & ( value - 1 ) ) === 0 && value !== 0;

		},

		nearestPowerOfTwo: function ( value ) {

			return Math.pow( 2, Math.round( Math.log( value ) / Math.LN2 ) );

		},

		nextPowerOfTwo: function ( value ) {

			value --;
			value |= value >> 1;
			value |= value >> 2;
			value |= value >> 4;
			value |= value >> 8;
			value |= value >> 16;
			value ++;

			return value;

		}

	};

	/**
	 * @author mrdoob / http://mrdoob.com/
	 * @author philogb / http://blog.thejit.org/
	 * @author egraether / http://egraether.com/
	 * @author zz85 / http://www.lab4games.net/zz85/blog
	 */

	function Vector2( x, y ) {

		this.x = x || 0;
		this.y = y || 0;

	}

	Vector2.prototype = {

		constructor: Vector2,

		isVector2: true,

		get width() {

			return this.x;

		},

		set width( value ) {

			this.x = value;

		},

		get height() {

			return this.y;

		},

		set height( value ) {

			this.y = value;

		},

		//

		set: function ( x, y ) {

			this.x = x;
			this.y = y;

			return this;

		},

		setScalar: function ( scalar ) {

			this.x = scalar;
			this.y = scalar;

			return this;

		},

		setX: function ( x ) {

			this.x = x;

			return this;

		},

		setY: function ( y ) {

			this.y = y;

			return this;

		},

		setComponent: function ( index, value ) {

			switch ( index ) {

				case 0: this.x = value; break;
				case 1: this.y = value; break;
				default: throw new Error( 'index is out of range: ' + index );

			}

			return this;

		},

		getComponent: function ( index ) {

			switch ( index ) {

				case 0: return this.x;
				case 1: return this.y;
				default: throw new Error( 'index is out of range: ' + index );

			}

		},

		clone: function () {

			return new this.constructor( this.x, this.y );

		},

		copy: function ( v ) {

			this.x = v.x;
			this.y = v.y;

			return this;

		},

		add: function ( v, w ) {

			if ( w !== undefined ) {

				console.warn( 'THREE.Vector2: .add() now only accepts one argument. Use .addVectors( a, b ) instead.' );
				return this.addVectors( v, w );

			}

			this.x += v.x;
			this.y += v.y;

			return this;

		},

		addScalar: function ( s ) {

			this.x += s;
			this.y += s;

			return this;

		},

		addVectors: function ( a, b ) {

			this.x = a.x + b.x;
			this.y = a.y + b.y;

			return this;

		},

		addScaledVector: function ( v, s ) {

			this.x += v.x * s;
			this.y += v.y * s;

			return this;

		},

		sub: function ( v, w ) {

			if ( w !== undefined ) {

				console.warn( 'THREE.Vector2: .sub() now only accepts one argument. Use .subVectors( a, b ) instead.' );
				return this.subVectors( v, w );

			}

			this.x -= v.x;
			this.y -= v.y;

			return this;

		},

		subScalar: function ( s ) {

			this.x -= s;
			this.y -= s;

			return this;

		},

		subVectors: function ( a, b ) {

			this.x = a.x - b.x;
			this.y = a.y - b.y;

			return this;

		},

		multiply: function ( v ) {

			this.x *= v.x;
			this.y *= v.y;

			return this;

		},

		multiplyScalar: function ( scalar ) {

			if ( isFinite( scalar ) ) {

				this.x *= scalar;
				this.y *= scalar;

			} else {

				this.x = 0;
				this.y = 0;

			}

			return this;

		},

		divide: function ( v ) {

			this.x /= v.x;
			this.y /= v.y;

			return this;

		},

		divideScalar: function ( scalar ) {

			return this.multiplyScalar( 1 / scalar );

		},

		min: function ( v ) {

			this.x = Math.min( this.x, v.x );
			this.y = Math.min( this.y, v.y );

			return this;

		},

		max: function ( v ) {

			this.x = Math.max( this.x, v.x );
			this.y = Math.max( this.y, v.y );

			return this;

		},

		clamp: function ( min, max ) {

			// This function assumes min < max, if this assumption isn't true it will not operate correctly

			this.x = Math.max( min.x, Math.min( max.x, this.x ) );
			this.y = Math.max( min.y, Math.min( max.y, this.y ) );

			return this;

		},

		clampScalar: function () {

			var min, max;

			return function clampScalar( minVal, maxVal ) {

				if ( min === undefined ) {

					min = new Vector2();
					max = new Vector2();

				}

				min.set( minVal, minVal );
				max.set( maxVal, maxVal );

				return this.clamp( min, max );

			};

		}(),

		clampLength: function ( min, max ) {

			var length = this.length();

			return this.multiplyScalar( Math.max( min, Math.min( max, length ) ) / length );

		},

		floor: function () {

			this.x = Math.floor( this.x );
			this.y = Math.floor( this.y );

			return this;

		},

		ceil: function () {

			this.x = Math.ceil( this.x );
			this.y = Math.ceil( this.y );

			return this;

		},

		round: function () {

			this.x = Math.round( this.x );
			this.y = Math.round( this.y );

			return this;

		},

		roundToZero: function () {

			this.x = ( this.x < 0 ) ? Math.ceil( this.x ) : Math.floor( this.x );
			this.y = ( this.y < 0 ) ? Math.ceil( this.y ) : Math.floor( this.y );

			return this;

		},

		negate: function () {

			this.x = - this.x;
			this.y = - this.y;

			return this;

		},

		dot: function ( v ) {

			return this.x * v.x + this.y * v.y;

		},

		lengthSq: function () {

			return this.x * this.x + this.y * this.y;

		},

		length: function () {

			return Math.sqrt( this.x * this.x + this.y * this.y );

		},

		lengthManhattan: function() {

			return Math.abs( this.x ) + Math.abs( this.y );

		},

		normalize: function () {

			return this.divideScalar( this.length() );

		},

		angle: function () {

			// computes the angle in radians with respect to the positive x-axis

			var angle = Math.atan2( this.y, this.x );

			if ( angle < 0 ) angle += 2 * Math.PI;

			return angle;

		},

		distanceTo: function ( v ) {

			return Math.sqrt( this.distanceToSquared( v ) );

		},

		distanceToSquared: function ( v ) {

			var dx = this.x - v.x, dy = this.y - v.y;
			return dx * dx + dy * dy;

		},

		distanceToManhattan: function ( v ) {

			return Math.abs( this.x - v.x ) + Math.abs( this.y - v.y );

		},

		setLength: function ( length ) {

			return this.multiplyScalar( length / this.length() );

		},

		lerp: function ( v, alpha ) {

			this.x += ( v.x - this.x ) * alpha;
			this.y += ( v.y - this.y ) * alpha;

			return this;

		},

		lerpVectors: function ( v1, v2, alpha ) {

			return this.subVectors( v2, v1 ).multiplyScalar( alpha ).add( v1 );

		},

		equals: function ( v ) {

			return ( ( v.x === this.x ) && ( v.y === this.y ) );

		},

		fromArray: function ( array, offset ) {

			if ( offset === undefined ) offset = 0;

			this.x = array[ offset ];
			this.y = array[ offset + 1 ];

			return this;

		},

		toArray: function ( array, offset ) {

			if ( array === undefined ) array = [];
			if ( offset === undefined ) offset = 0;

			array[ offset ] = this.x;
			array[ offset + 1 ] = this.y;

			return array;

		},

		fromAttribute: function ( attribute, index, offset ) {

			if ( offset !== undefined ) {

				console.warn( 'THREE.Vector2: offset has been removed from .fromAttribute().' );

			}

			this.x = attribute.getX( index );
			this.y = attribute.getY( index );

			return this;

		},

		rotateAround: function ( center, angle ) {

			var c = Math.cos( angle ), s = Math.sin( angle );

			var x = this.x - center.x;
			var y = this.y - center.y;

			this.x = x * c - y * s + center.x;
			this.y = x * s + y * c + center.y;

			return this;

		}

	};

	/**
	 * @author mrdoob / http://mrdoob.com/
	 * @author alteredq / http://alteredqualia.com/
	 * @author szimek / https://github.com/szimek/
	 */

	var textureId = 0;

	function Texture( image, mapping, wrapS, wrapT, magFilter, minFilter, format, type, anisotropy, encoding ) {

		Object.defineProperty( this, 'id', { value: textureId ++ } );

		this.uuid = _Math.generateUUID();

		this.name = '';

		this.image = image !== undefined ? image : Texture.DEFAULT_IMAGE;
		this.mipmaps = [];

		this.mapping = mapping !== undefined ? mapping : Texture.DEFAULT_MAPPING;

		this.wrapS = wrapS !== undefined ? wrapS : ClampToEdgeWrapping;
		this.wrapT = wrapT !== undefined ? wrapT : ClampToEdgeWrapping;

		this.magFilter = magFilter !== undefined ? magFilter : LinearFilter;
		this.minFilter = minFilter !== undefined ? minFilter : LinearMipMapLinearFilter;

		this.anisotropy = anisotropy !== undefined ? anisotropy : 1;

		this.format = format !== undefined ? format : RGBAFormat;
		this.type = type !== undefined ? type : UnsignedByteType;

		this.offset = new Vector2( 0, 0 );
		this.repeat = new Vector2( 1, 1 );

		this.generateMipmaps = true;
		this.premultiplyAlpha = false;
		this.flipY = true;
		this.unpackAlignment = 4;	// valid values: 1, 2, 4, 8 (see http://www.khronos.org/opengles/sdk/docs/man/xhtml/glPixelStorei.xml)


		// Values of encoding !== THREE.LinearEncoding only supported on map, envMap and emissiveMap.
		//
		// Also changing the encoding after already used by a Material will not automatically make the Material
		// update.  You need to explicitly call Material.needsUpdate to trigger it to recompile.
		this.encoding = encoding !== undefined ? encoding : LinearEncoding;

		this.version = 0;
		this.onUpdate = null;

	}

	Texture.DEFAULT_IMAGE = undefined;
	Texture.DEFAULT_MAPPING = UVMapping;

	Texture.prototype = {

		constructor: Texture,

		isTexture: true,

		set needsUpdate( value ) {

			if ( value === true ) this.version ++;

		},

		clone: function () {

			return new this.constructor().copy( this );

		},

		copy: function ( source ) {

			this.image = source.image;
			this.mipmaps = source.mipmaps.slice( 0 );

			this.mapping = source.mapping;

			this.wrapS = source.wrapS;
			this.wrapT = source.wrapT;

			this.magFilter = source.magFilter;
			this.minFilter = source.minFilter;

			this.anisotropy = source.anisotropy;

			this.format = source.format;
			this.type = source.type;

			this.offset.copy( source.offset );
			this.repeat.copy( source.repeat );

			this.generateMipmaps = source.generateMipmaps;
			this.premultiplyAlpha = source.premultiplyAlpha;
			this.flipY = source.flipY;
			this.unpackAlignment = source.unpackAlignment;
			this.encoding = source.encoding;

			return this;

		},

		toJSON: function ( meta ) {

			if ( meta.textures[ this.uuid ] !== undefined ) {

				return meta.textures[ this.uuid ];

			}

			function getDataURL( image ) {

				var canvas;

				if ( image.toDataURL !== undefined ) {

					canvas = image;

				} else {

					canvas = document.createElementNS( 'http://www.w3.org/1999/xhtml', 'canvas' );
					canvas.width = image.width;
					canvas.height = image.height;

					canvas.getContext( '2d' ).drawImage( image, 0, 0, image.width, image.height );

				}

				if ( canvas.width > 2048 || canvas.height > 2048 ) {

					return canvas.toDataURL( 'image/jpeg', 0.6 );

				} else {

					return canvas.toDataURL( 'image/png' );

				}

			}

			var output = {
				metadata: {
					version: 4.4,
					type: 'Texture',
					generator: 'Texture.toJSON'
				},

				uuid: this.uuid,
				name: this.name,

				mapping: this.mapping,

				repeat: [ this.repeat.x, this.repeat.y ],
				offset: [ this.offset.x, this.offset.y ],
				wrap: [ this.wrapS, this.wrapT ],

				minFilter: this.minFilter,
				magFilter: this.magFilter,
				anisotropy: this.anisotropy,

				flipY: this.flipY
			};

			if ( this.image !== undefined ) {

				// TODO: Move to THREE.Image

				var image = this.image;

				if ( image.uuid === undefined ) {

					image.uuid = _Math.generateUUID(); // UGH

				}

				if ( meta.images[ image.uuid ] === undefined ) {

					meta.images[ image.uuid ] = {
						uuid: image.uuid,
						url: getDataURL( image )
					};

				}

				output.image = image.uuid;

			}

			meta.textures[ this.uuid ] = output;

			return output;

		},

		dispose: function () {

			this.dispatchEvent( { type: 'dispose' } );

		},

		transformUv: function ( uv ) {

			if ( this.mapping !== UVMapping ) return;

			uv.multiply( this.repeat );
			uv.add( this.offset );

			if ( uv.x < 0 || uv.x > 1 ) {

				switch ( this.wrapS ) {

					case RepeatWrapping:

						uv.x = uv.x - Math.floor( uv.x );
						break;

					case ClampToEdgeWrapping:

						uv.x = uv.x < 0 ? 0 : 1;
						break;

					case MirroredRepeatWrapping:

						if ( Math.abs( Math.floor( uv.x ) % 2 ) === 1 ) {

							uv.x = Math.ceil( uv.x ) - uv.x;

						} else {

							uv.x = uv.x - Math.floor( uv.x );

						}
						break;

				}

			}

			if ( uv.y < 0 || uv.y > 1 ) {

				switch ( this.wrapT ) {

					case RepeatWrapping:

						uv.y = uv.y - Math.floor( uv.y );
						break;

					case ClampToEdgeWrapping:

						uv.y = uv.y < 0 ? 0 : 1;
						break;

					case MirroredRepeatWrapping:

						if ( Math.abs( Math.floor( uv.y ) % 2 ) === 1 ) {

							uv.y = Math.ceil( uv.y ) - uv.y;

						} else {

							uv.y = uv.y - Math.floor( uv.y );

						}
						break;

				}

			}

			if ( this.flipY ) {

				uv.y = 1 - uv.y;

			}

		}

	};

	Object.assign( Texture.prototype, EventDispatcher.prototype );

	/**
	 * @author supereggbert / http://www.paulbrunt.co.uk/
	 * @author philogb / http://blog.thejit.org/
	 * @author mikael emtinger / http://gomo.se/
	 * @author egraether / http://egraether.com/
	 * @author WestLangley / http://github.com/WestLangley
	 */

	function Vector4( x, y, z, w ) {

		this.x = x || 0;
		this.y = y || 0;
		this.z = z || 0;
		this.w = ( w !== undefined ) ? w : 1;

	}

	Vector4.prototype = {

		constructor: Vector4,

		isVector4: true,

		set: function ( x, y, z, w ) {

			this.x = x;
			this.y = y;
			this.z = z;
			this.w = w;

			return this;

		},

		setScalar: function ( scalar ) {

			this.x = scalar;
			this.y = scalar;
			this.z = scalar;
			this.w = scalar;

			return this;

		},

		setX: function ( x ) {

			this.x = x;

			return this;

		},

		setY: function ( y ) {

			this.y = y;

			return this;

		},

		setZ: function ( z ) {

			this.z = z;

			return this;

		},

		setW: function ( w ) {

			this.w = w;

			return this;

		},

		setComponent: function ( index, value ) {

			switch ( index ) {

				case 0: this.x = value; break;
				case 1: this.y = value; break;
				case 2: this.z = value; break;
				case 3: this.w = value; break;
				default: throw new Error( 'index is out of range: ' + index );

			}

			return this;

		},

		getComponent: function ( index ) {

			switch ( index ) {

				case 0: return this.x;
				case 1: return this.y;
				case 2: return this.z;
				case 3: return this.w;
				default: throw new Error( 'index is out of range: ' + index );

			}

		},

		clone: function () {

			return new this.constructor( this.x, this.y, this.z, this.w );

		},

		copy: function ( v ) {

			this.x = v.x;
			this.y = v.y;
			this.z = v.z;
			this.w = ( v.w !== undefined ) ? v.w : 1;

			return this;

		},

		add: function ( v, w ) {

			if ( w !== undefined ) {

				console.warn( 'THREE.Vector4: .add() now only accepts one argument. Use .addVectors( a, b ) instead.' );
				return this.addVectors( v, w );

			}

			this.x += v.x;
			this.y += v.y;
			this.z += v.z;
			this.w += v.w;

			return this;

		},

		addScalar: function ( s ) {

			this.x += s;
			this.y += s;
			this.z += s;
			this.w += s;

			return this;

		},

		addVectors: function ( a, b ) {

			this.x = a.x + b.x;
			this.y = a.y + b.y;
			this.z = a.z + b.z;
			this.w = a.w + b.w;

			return this;

		},

		addScaledVector: function ( v, s ) {

			this.x += v.x * s;
			this.y += v.y * s;
			this.z += v.z * s;
			this.w += v.w * s;

			return this;

		},

		sub: function ( v, w ) {

			if ( w !== undefined ) {

				console.warn( 'THREE.Vector4: .sub() now only accepts one argument. Use .subVectors( a, b ) instead.' );
				return this.subVectors( v, w );

			}

			this.x -= v.x;
			this.y -= v.y;
			this.z -= v.z;
			this.w -= v.w;

			return this;

		},

		subScalar: function ( s ) {

			this.x -= s;
			this.y -= s;
			this.z -= s;
			this.w -= s;

			return this;

		},

		subVectors: function ( a, b ) {

			this.x = a.x - b.x;
			this.y = a.y - b.y;
			this.z = a.z - b.z;
			this.w = a.w - b.w;

			return this;

		},

		multiplyScalar: function ( scalar ) {

			if ( isFinite( scalar ) ) {

				this.x *= scalar;
				this.y *= scalar;
				this.z *= scalar;
				this.w *= scalar;

			} else {

				this.x = 0;
				this.y = 0;
				this.z = 0;
				this.w = 0;

			}

			return this;

		},

		applyMatrix4: function ( m ) {

			var x = this.x, y = this.y, z = this.z, w = this.w;
			var e = m.elements;

			this.x = e[ 0 ] * x + e[ 4 ] * y + e[ 8 ] * z + e[ 12 ] * w;
			this.y = e[ 1 ] * x + e[ 5 ] * y + e[ 9 ] * z + e[ 13 ] * w;
			this.z = e[ 2 ] * x + e[ 6 ] * y + e[ 10 ] * z + e[ 14 ] * w;
			this.w = e[ 3 ] * x + e[ 7 ] * y + e[ 11 ] * z + e[ 15 ] * w;

			return this;

		},

		divideScalar: function ( scalar ) {

			return this.multiplyScalar( 1 / scalar );

		},

		setAxisAngleFromQuaternion: function ( q ) {

			// http://www.euclideanspace.com/maths/geometry/rotations/conversions/quaternionToAngle/index.htm

			// q is assumed to be normalized

			this.w = 2 * Math.acos( q.w );

			var s = Math.sqrt( 1 - q.w * q.w );

			if ( s < 0.0001 ) {

				 this.x = 1;
				 this.y = 0;
				 this.z = 0;

			} else {

				 this.x = q.x / s;
				 this.y = q.y / s;
				 this.z = q.z / s;

			}

			return this;

		},

		setAxisAngleFromRotationMatrix: function ( m ) {

			// http://www.euclideanspace.com/maths/geometry/rotations/conversions/matrixToAngle/index.htm

			// assumes the upper 3x3 of m is a pure rotation matrix (i.e, unscaled)

			var angle, x, y, z,		// variables for result
				epsilon = 0.01,		// margin to allow for rounding errors
				epsilon2 = 0.1,		// margin to distinguish between 0 and 180 degrees

				te = m.elements,

				m11 = te[ 0 ], m12 = te[ 4 ], m13 = te[ 8 ],
				m21 = te[ 1 ], m22 = te[ 5 ], m23 = te[ 9 ],
				m31 = te[ 2 ], m32 = te[ 6 ], m33 = te[ 10 ];

			if ( ( Math.abs( m12 - m21 ) < epsilon ) &&
			     ( Math.abs( m13 - m31 ) < epsilon ) &&
			     ( Math.abs( m23 - m32 ) < epsilon ) ) {

				// singularity found
				// first check for identity matrix which must have +1 for all terms
				// in leading diagonal and zero in other terms

				if ( ( Math.abs( m12 + m21 ) < epsilon2 ) &&
				     ( Math.abs( m13 + m31 ) < epsilon2 ) &&
				     ( Math.abs( m23 + m32 ) < epsilon2 ) &&
				     ( Math.abs( m11 + m22 + m33 - 3 ) < epsilon2 ) ) {

					// this singularity is identity matrix so angle = 0

					this.set( 1, 0, 0, 0 );

					return this; // zero angle, arbitrary axis

				}

				// otherwise this singularity is angle = 180

				angle = Math.PI;

				var xx = ( m11 + 1 ) / 2;
				var yy = ( m22 + 1 ) / 2;
				var zz = ( m33 + 1 ) / 2;
				var xy = ( m12 + m21 ) / 4;
				var xz = ( m13 + m31 ) / 4;
				var yz = ( m23 + m32 ) / 4;

				if ( ( xx > yy ) && ( xx > zz ) ) {

					// m11 is the largest diagonal term

					if ( xx < epsilon ) {

						x = 0;
						y = 0.707106781;
						z = 0.707106781;

					} else {

						x = Math.sqrt( xx );
						y = xy / x;
						z = xz / x;

					}

				} else if ( yy > zz ) {

					// m22 is the largest diagonal term

					if ( yy < epsilon ) {

						x = 0.707106781;
						y = 0;
						z = 0.707106781;

					} else {

						y = Math.sqrt( yy );
						x = xy / y;
						z = yz / y;

					}

				} else {

					// m33 is the largest diagonal term so base result on this

					if ( zz < epsilon ) {

						x = 0.707106781;
						y = 0.707106781;
						z = 0;

					} else {

						z = Math.sqrt( zz );
						x = xz / z;
						y = yz / z;

					}

				}

				this.set( x, y, z, angle );

				return this; // return 180 deg rotation

			}

			// as we have reached here there are no singularities so we can handle normally

			var s = Math.sqrt( ( m32 - m23 ) * ( m32 - m23 ) +
			                   ( m13 - m31 ) * ( m13 - m31 ) +
			                   ( m21 - m12 ) * ( m21 - m12 ) ); // used to normalize

			if ( Math.abs( s ) < 0.001 ) s = 1;

			// prevent divide by zero, should not happen if matrix is orthogonal and should be
			// caught by singularity test above, but I've left it in just in case

			this.x = ( m32 - m23 ) / s;
			this.y = ( m13 - m31 ) / s;
			this.z = ( m21 - m12 ) / s;
			this.w = Math.acos( ( m11 + m22 + m33 - 1 ) / 2 );

			return this;

		},

		min: function ( v ) {

			this.x = Math.min( this.x, v.x );
			this.y = Math.min( this.y, v.y );
			this.z = Math.min( this.z, v.z );
			this.w = Math.min( this.w, v.w );

			return this;

		},

		max: function ( v ) {

			this.x = Math.max( this.x, v.x );
			this.y = Math.max( this.y, v.y );
			this.z = Math.max( this.z, v.z );
			this.w = Math.max( this.w, v.w );

			return this;

		},

		clamp: function ( min, max ) {

			// This function assumes min < max, if this assumption isn't true it will not operate correctly

			this.x = Math.max( min.x, Math.min( max.x, this.x ) );
			this.y = Math.max( min.y, Math.min( max.y, this.y ) );
			this.z = Math.max( min.z, Math.min( max.z, this.z ) );
			this.w = Math.max( min.w, Math.min( max.w, this.w ) );

			return this;

		},

		clampScalar: function () {

			var min, max;

			return function clampScalar( minVal, maxVal ) {

				if ( min === undefined ) {

					min = new Vector4();
					max = new Vector4();

				}

				min.set( minVal, minVal, minVal, minVal );
				max.set( maxVal, maxVal, maxVal, maxVal );

				return this.clamp( min, max );

			};

		}(),

		floor: function () {

			this.x = Math.floor( this.x );
			this.y = Math.floor( this.y );
			this.z = Math.floor( this.z );
			this.w = Math.floor( this.w );

			return this;

		},

		ceil: function () {

			this.x = Math.ceil( this.x );
			this.y = Math.ceil( this.y );
			this.z = Math.ceil( this.z );
			this.w = Math.ceil( this.w );

			return this;

		},

		round: function () {

			this.x = Math.round( this.x );
			this.y = Math.round( this.y );
			this.z = Math.round( this.z );
			this.w = Math.round( this.w );

			return this;

		},

		roundToZero: function () {

			this.x = ( this.x < 0 ) ? Math.ceil( this.x ) : Math.floor( this.x );
			this.y = ( this.y < 0 ) ? Math.ceil( this.y ) : Math.floor( this.y );
			this.z = ( this.z < 0 ) ? Math.ceil( this.z ) : Math.floor( this.z );
			this.w = ( this.w < 0 ) ? Math.ceil( this.w ) : Math.floor( this.w );

			return this;

		},

		negate: function () {

			this.x = - this.x;
			this.y = - this.y;
			this.z = - this.z;
			this.w = - this.w;

			return this;

		},

		dot: function ( v ) {

			return this.x * v.x + this.y * v.y + this.z * v.z + this.w * v.w;

		},

		lengthSq: function () {

			return this.x * this.x + this.y * this.y + this.z * this.z + this.w * this.w;

		},

		length: function () {

			return Math.sqrt( this.x * this.x + this.y * this.y + this.z * this.z + this.w * this.w );

		},

		lengthManhattan: function () {

			return Math.abs( this.x ) + Math.abs( this.y ) + Math.abs( this.z ) + Math.abs( this.w );

		},

		normalize: function () {

			return this.divideScalar( this.length() );

		},

		setLength: function ( length ) {

			return this.multiplyScalar( length / this.length() );

		},

		lerp: function ( v, alpha ) {

			this.x += ( v.x - this.x ) * alpha;
			this.y += ( v.y - this.y ) * alpha;
			this.z += ( v.z - this.z ) * alpha;
			this.w += ( v.w - this.w ) * alpha;

			return this;

		},

		lerpVectors: function ( v1, v2, alpha ) {

			return this.subVectors( v2, v1 ).multiplyScalar( alpha ).add( v1 );

		},

		equals: function ( v ) {

			return ( ( v.x === this.x ) && ( v.y === this.y ) && ( v.z === this.z ) && ( v.w === this.w ) );

		},

		fromArray: function ( array, offset ) {

			if ( offset === undefined ) offset = 0;

			this.x = array[ offset ];
			this.y = array[ offset + 1 ];
			this.z = array[ offset + 2 ];
			this.w = array[ offset + 3 ];

			return this;

		},

		toArray: function ( array, offset ) {

			if ( array === undefined ) array = [];
			if ( offset === undefined ) offset = 0;

			array[ offset ] = this.x;
			array[ offset + 1 ] = this.y;
			array[ offset + 2 ] = this.z;
			array[ offset + 3 ] = this.w;

			return array;

		},

		fromAttribute: function ( attribute, index, offset ) {

			if ( offset !== undefined ) {

				console.warn( 'THREE.Vector4: offset has been removed from .fromAttribute().' );

			}

			this.x = attribute.getX( index );
			this.y = attribute.getY( index );
			this.z = attribute.getZ( index );
			this.w = attribute.getW( index );

			return this;

		}

	};

	/**
	 * @author szimek / https://github.com/szimek/
	 * @author alteredq / http://alteredqualia.com/
	 * @author Marius Kintel / https://github.com/kintel
	 */

	/*
	 In options, we can specify:
	 * Texture parameters for an auto-generated target texture
	 * depthBuffer/stencilBuffer: Booleans to indicate if we should generate these buffers
	*/
	function WebGLRenderTarget( width, height, options ) {

		this.uuid = _Math.generateUUID();

		this.width = width;
		this.height = height;

		this.scissor = new Vector4( 0, 0, width, height );
		this.scissorTest = false;

		this.viewport = new Vector4( 0, 0, width, height );

		options = options || {};

		if ( options.minFilter === undefined ) options.minFilter = LinearFilter;

		this.texture = new Texture( undefined, undefined, options.wrapS, options.wrapT, options.magFilter, options.minFilter, options.format, options.type, options.anisotropy, options.encoding );

		this.depthBuffer = options.depthBuffer !== undefined ? options.depthBuffer : true;
		this.stencilBuffer = options.stencilBuffer !== undefined ? options.stencilBuffer : true;
		this.depthTexture = options.depthTexture !== undefined ? options.depthTexture : null;

	}

	Object.assign( WebGLRenderTarget.prototype, EventDispatcher.prototype, {

		isWebGLRenderTarget: true,

		setSize: function ( width, height ) {

			if ( this.width !== width || this.height !== height ) {

				this.width = width;
				this.height = height;

				this.dispose();

			}

			this.viewport.set( 0, 0, width, height );
			this.scissor.set( 0, 0, width, height );

		},

		clone: function () {

			return new this.constructor().copy( this );

		},

		copy: function ( source ) {

			this.width = source.width;
			this.height = source.height;

			this.viewport.copy( source.viewport );

			this.texture = source.texture.clone();

			this.depthBuffer = source.depthBuffer;
			this.stencilBuffer = source.stencilBuffer;
			this.depthTexture = source.depthTexture;

			return this;

		},

		dispose: function () {

			this.dispatchEvent( { type: 'dispose' } );

		}

	} );

	/**
	 * @author alteredq / http://alteredqualia.com
	 */

	function WebGLRenderTargetCube( width, height, options ) {

		WebGLRenderTarget.call( this, width, height, options );

		this.activeCubeFace = 0; // PX 0, NX 1, PY 2, NY 3, PZ 4, NZ 5
		this.activeMipMapLevel = 0;

	}

	WebGLRenderTargetCube.prototype = Object.create( WebGLRenderTarget.prototype );
	WebGLRenderTargetCube.prototype.constructor = WebGLRenderTargetCube;

	WebGLRenderTargetCube.prototype.isWebGLRenderTargetCube = true;

	/**
	 * @author mikael emtinger / http://gomo.se/
	 * @author alteredq / http://alteredqualia.com/
	 * @author WestLangley / http://github.com/WestLangley
	 * @author bhouston / http://clara.io
	 */

	function Quaternion( x, y, z, w ) {

		this._x = x || 0;
		this._y = y || 0;
		this._z = z || 0;
		this._w = ( w !== undefined ) ? w : 1;

	}

	Quaternion.prototype = {

		constructor: Quaternion,

		get x () {

			return this._x;

		},

		set x ( value ) {

			this._x = value;
			this.onChangeCallback();

		},

		get y () {

			return this._y;

		},

		set y ( value ) {

			this._y = value;
			this.onChangeCallback();

		},

		get z () {

			return this._z;

		},

		set z ( value ) {

			this._z = value;
			this.onChangeCallback();

		},

		get w () {

			return this._w;

		},

		set w ( value ) {

			this._w = value;
			this.onChangeCallback();

		},

		set: function ( x, y, z, w ) {

			this._x = x;
			this._y = y;
			this._z = z;
			this._w = w;

			this.onChangeCallback();

			return this;

		},

		clone: function () {

			return new this.constructor( this._x, this._y, this._z, this._w );

		},

		copy: function ( quaternion ) {

			this._x = quaternion.x;
			this._y = quaternion.y;
			this._z = quaternion.z;
			this._w = quaternion.w;

			this.onChangeCallback();

			return this;

		},

		setFromEuler: function ( euler, update ) {

			if ( (euler && euler.isEuler) === false ) {

				throw new Error( 'THREE.Quaternion: .setFromEuler() now expects an Euler rotation rather than a Vector3 and order.' );

			}

			// http://www.mathworks.com/matlabcentral/fileexchange/
			// 	20696-function-to-convert-between-dcm-euler-angles-quaternions-and-euler-vectors/
			//	content/SpinCalc.m

			var c1 = Math.cos( euler._x / 2 );
			var c2 = Math.cos( euler._y / 2 );
			var c3 = Math.cos( euler._z / 2 );
			var s1 = Math.sin( euler._x / 2 );
			var s2 = Math.sin( euler._y / 2 );
			var s3 = Math.sin( euler._z / 2 );

			var order = euler.order;

			if ( order === 'XYZ' ) {

				this._x = s1 * c2 * c3 + c1 * s2 * s3;
				this._y = c1 * s2 * c3 - s1 * c2 * s3;
				this._z = c1 * c2 * s3 + s1 * s2 * c3;
				this._w = c1 * c2 * c3 - s1 * s2 * s3;

			} else if ( order === 'YXZ' ) {

				this._x = s1 * c2 * c3 + c1 * s2 * s3;
				this._y = c1 * s2 * c3 - s1 * c2 * s3;
				this._z = c1 * c2 * s3 - s1 * s2 * c3;
				this._w = c1 * c2 * c3 + s1 * s2 * s3;

			} else if ( order === 'ZXY' ) {

				this._x = s1 * c2 * c3 - c1 * s2 * s3;
				this._y = c1 * s2 * c3 + s1 * c2 * s3;
				this._z = c1 * c2 * s3 + s1 * s2 * c3;
				this._w = c1 * c2 * c3 - s1 * s2 * s3;

			} else if ( order === 'ZYX' ) {

				this._x = s1 * c2 * c3 - c1 * s2 * s3;
				this._y = c1 * s2 * c3 + s1 * c2 * s3;
				this._z = c1 * c2 * s3 - s1 * s2 * c3;
				this._w = c1 * c2 * c3 + s1 * s2 * s3;

			} else if ( order === 'YZX' ) {

				this._x = s1 * c2 * c3 + c1 * s2 * s3;
				this._y = c1 * s2 * c3 + s1 * c2 * s3;
				this._z = c1 * c2 * s3 - s1 * s2 * c3;
				this._w = c1 * c2 * c3 - s1 * s2 * s3;

			} else if ( order === 'XZY' ) {

				this._x = s1 * c2 * c3 - c1 * s2 * s3;
				this._y = c1 * s2 * c3 - s1 * c2 * s3;
				this._z = c1 * c2 * s3 + s1 * s2 * c3;
				this._w = c1 * c2 * c3 + s1 * s2 * s3;

			}

			if ( update !== false ) this.onChangeCallback();

			return this;

		},

		setFromAxisAngle: function ( axis, angle ) {

			// http://www.euclideanspace.com/maths/geometry/rotations/conversions/angleToQuaternion/index.htm

			// assumes axis is normalized

			var halfAngle = angle / 2, s = Math.sin( halfAngle );

			this._x = axis.x * s;
			this._y = axis.y * s;
			this._z = axis.z * s;
			this._w = Math.cos( halfAngle );

			this.onChangeCallback();

			return this;

		},

		setFromRotationMatrix: function ( m ) {

			// http://www.euclideanspace.com/maths/geometry/rotations/conversions/matrixToQuaternion/index.htm

			// assumes the upper 3x3 of m is a pure rotation matrix (i.e, unscaled)

			var te = m.elements,

				m11 = te[ 0 ], m12 = te[ 4 ], m13 = te[ 8 ],
				m21 = te[ 1 ], m22 = te[ 5 ], m23 = te[ 9 ],
				m31 = te[ 2 ], m32 = te[ 6 ], m33 = te[ 10 ],

				trace = m11 + m22 + m33,
				s;

			if ( trace > 0 ) {

				s = 0.5 / Math.sqrt( trace + 1.0 );

				this._w = 0.25 / s;
				this._x = ( m32 - m23 ) * s;
				this._y = ( m13 - m31 ) * s;
				this._z = ( m21 - m12 ) * s;

			} else if ( m11 > m22 && m11 > m33 ) {

				s = 2.0 * Math.sqrt( 1.0 + m11 - m22 - m33 );

				this._w = ( m32 - m23 ) / s;
				this._x = 0.25 * s;
				this._y = ( m12 + m21 ) / s;
				this._z = ( m13 + m31 ) / s;

			} else if ( m22 > m33 ) {

				s = 2.0 * Math.sqrt( 1.0 + m22 - m11 - m33 );

				this._w = ( m13 - m31 ) / s;
				this._x = ( m12 + m21 ) / s;
				this._y = 0.25 * s;
				this._z = ( m23 + m32 ) / s;

			} else {

				s = 2.0 * Math.sqrt( 1.0 + m33 - m11 - m22 );

				this._w = ( m21 - m12 ) / s;
				this._x = ( m13 + m31 ) / s;
				this._y = ( m23 + m32 ) / s;
				this._z = 0.25 * s;

			}

			this.onChangeCallback();

			return this;

		},

		setFromUnitVectors: function () {

			// http://lolengine.net/blog/2014/02/24/quaternion-from-two-vectors-final

			// assumes direction vectors vFrom and vTo are normalized

			var v1, r;

			var EPS = 0.000001;

			return function setFromUnitVectors( vFrom, vTo ) {

				if ( v1 === undefined ) v1 = new Vector3();

				r = vFrom.dot( vTo ) + 1;

				if ( r < EPS ) {

					r = 0;

					if ( Math.abs( vFrom.x ) > Math.abs( vFrom.z ) ) {

						v1.set( - vFrom.y, vFrom.x, 0 );

					} else {

						v1.set( 0, - vFrom.z, vFrom.y );

					}

				} else {

					v1.crossVectors( vFrom, vTo );

				}

				this._x = v1.x;
				this._y = v1.y;
				this._z = v1.z;
				this._w = r;

				return this.normalize();

			};

		}(),

		inverse: function () {

			return this.conjugate().normalize();

		},

		conjugate: function () {

			this._x *= - 1;
			this._y *= - 1;
			this._z *= - 1;

			this.onChangeCallback();

			return this;

		},

		dot: function ( v ) {

			return this._x * v._x + this._y * v._y + this._z * v._z + this._w * v._w;

		},

		lengthSq: function () {

			return this._x * this._x + this._y * this._y + this._z * this._z + this._w * this._w;

		},

		length: function () {

			return Math.sqrt( this._x * this._x + this._y * this._y + this._z * this._z + this._w * this._w );

		},

		normalize: function () {

			var l = this.length();

			if ( l === 0 ) {

				this._x = 0;
				this._y = 0;
				this._z = 0;
				this._w = 1;

			} else {

				l = 1 / l;

				this._x = this._x * l;
				this._y = this._y * l;
				this._z = this._z * l;
				this._w = this._w * l;

			}

			this.onChangeCallback();

			return this;

		},

		multiply: function ( q, p ) {

			if ( p !== undefined ) {

				console.warn( 'THREE.Quaternion: .multiply() now only accepts one argument. Use .multiplyQuaternions( a, b ) instead.' );
				return this.multiplyQuaternions( q, p );

			}

			return this.multiplyQuaternions( this, q );

		},

		premultiply: function ( q ) {

			return this.multiplyQuaternions( q, this );

		},

		multiplyQuaternions: function ( a, b ) {

			// from http://www.euclideanspace.com/maths/algebra/realNormedAlgebra/quaternions/code/index.htm

			var qax = a._x, qay = a._y, qaz = a._z, qaw = a._w;
			var qbx = b._x, qby = b._y, qbz = b._z, qbw = b._w;

			this._x = qax * qbw + qaw * qbx + qay * qbz - qaz * qby;
			this._y = qay * qbw + qaw * qby + qaz * qbx - qax * qbz;
			this._z = qaz * qbw + qaw * qbz + qax * qby - qay * qbx;
			this._w = qaw * qbw - qax * qbx - qay * qby - qaz * qbz;

			this.onChangeCallback();

			return this;

		},

		slerp: function ( qb, t ) {

			if ( t === 0 ) return this;
			if ( t === 1 ) return this.copy( qb );

			var x = this._x, y = this._y, z = this._z, w = this._w;

			// http://www.euclideanspace.com/maths/algebra/realNormedAlgebra/quaternions/slerp/

			var cosHalfTheta = w * qb._w + x * qb._x + y * qb._y + z * qb._z;

			if ( cosHalfTheta < 0 ) {

				this._w = - qb._w;
				this._x = - qb._x;
				this._y = - qb._y;
				this._z = - qb._z;

				cosHalfTheta = - cosHalfTheta;

			} else {

				this.copy( qb );

			}

			if ( cosHalfTheta >= 1.0 ) {

				this._w = w;
				this._x = x;
				this._y = y;
				this._z = z;

				return this;

			}

			var sinHalfTheta = Math.sqrt( 1.0 - cosHalfTheta * cosHalfTheta );

			if ( Math.abs( sinHalfTheta ) < 0.001 ) {

				this._w = 0.5 * ( w + this._w );
				this._x = 0.5 * ( x + this._x );
				this._y = 0.5 * ( y + this._y );
				this._z = 0.5 * ( z + this._z );

				return this;

			}

			var halfTheta = Math.atan2( sinHalfTheta, cosHalfTheta );
			var ratioA = Math.sin( ( 1 - t ) * halfTheta ) / sinHalfTheta,
			ratioB = Math.sin( t * halfTheta ) / sinHalfTheta;

			this._w = ( w * ratioA + this._w * ratioB );
			this._x = ( x * ratioA + this._x * ratioB );
			this._y = ( y * ratioA + this._y * ratioB );
			this._z = ( z * ratioA + this._z * ratioB );

			this.onChangeCallback();

			return this;

		},

		equals: function ( quaternion ) {

			return ( quaternion._x === this._x ) && ( quaternion._y === this._y ) && ( quaternion._z === this._z ) && ( quaternion._w === this._w );

		},

		fromArray: function ( array, offset ) {

			if ( offset === undefined ) offset = 0;

			this._x = array[ offset ];
			this._y = array[ offset + 1 ];
			this._z = array[ offset + 2 ];
			this._w = array[ offset + 3 ];

			this.onChangeCallback();

			return this;

		},

		toArray: function ( array, offset ) {

			if ( array === undefined ) array = [];
			if ( offset === undefined ) offset = 0;

			array[ offset ] = this._x;
			array[ offset + 1 ] = this._y;
			array[ offset + 2 ] = this._z;
			array[ offset + 3 ] = this._w;

			return array;

		},

		onChange: function ( callback ) {

			this.onChangeCallback = callback;

			return this;

		},

		onChangeCallback: function () {}

	};

	Object.assign( Quaternion, {

		slerp: function( qa, qb, qm, t ) {

			return qm.copy( qa ).slerp( qb, t );

		},

		slerpFlat: function(
				dst, dstOffset, src0, srcOffset0, src1, srcOffset1, t ) {

			// fuzz-free, array-based Quaternion SLERP operation

			var x0 = src0[ srcOffset0 + 0 ],
				y0 = src0[ srcOffset0 + 1 ],
				z0 = src0[ srcOffset0 + 2 ],
				w0 = src0[ srcOffset0 + 3 ],

				x1 = src1[ srcOffset1 + 0 ],
				y1 = src1[ srcOffset1 + 1 ],
				z1 = src1[ srcOffset1 + 2 ],
				w1 = src1[ srcOffset1 + 3 ];

			if ( w0 !== w1 || x0 !== x1 || y0 !== y1 || z0 !== z1 ) {

				var s = 1 - t,

					cos = x0 * x1 + y0 * y1 + z0 * z1 + w0 * w1,

					dir = ( cos >= 0 ? 1 : - 1 ),
					sqrSin = 1 - cos * cos;

				// Skip the Slerp for tiny steps to avoid numeric problems:
				if ( sqrSin > Number.EPSILON ) {

					var sin = Math.sqrt( sqrSin ),
						len = Math.atan2( sin, cos * dir );

					s = Math.sin( s * len ) / sin;
					t = Math.sin( t * len ) / sin;

				}

				var tDir = t * dir;

				x0 = x0 * s + x1 * tDir;
				y0 = y0 * s + y1 * tDir;
				z0 = z0 * s + z1 * tDir;
				w0 = w0 * s + w1 * tDir;

				// Normalize in case we just did a lerp:
				if ( s === 1 - t ) {

					var f = 1 / Math.sqrt( x0 * x0 + y0 * y0 + z0 * z0 + w0 * w0 );

					x0 *= f;
					y0 *= f;
					z0 *= f;
					w0 *= f;

				}

			}

			dst[ dstOffset ] = x0;
			dst[ dstOffset + 1 ] = y0;
			dst[ dstOffset + 2 ] = z0;
			dst[ dstOffset + 3 ] = w0;

		}

	} );

	/**
	 * @author mrdoob / http://mrdoob.com/
	 * @author *kile / http://kile.stravaganza.org/
	 * @author philogb / http://blog.thejit.org/
	 * @author mikael emtinger / http://gomo.se/
	 * @author egraether / http://egraether.com/
	 * @author WestLangley / http://github.com/WestLangley
	 */

	function Vector3( x, y, z ) {

		this.x = x || 0;
		this.y = y || 0;
		this.z = z || 0;

	}

	Vector3.prototype = {

		constructor: Vector3,

		isVector3: true,

		set: function ( x, y, z ) {

			this.x = x;
			this.y = y;
			this.z = z;

			return this;

		},

		setScalar: function ( scalar ) {

			this.x = scalar;
			this.y = scalar;
			this.z = scalar;

			return this;

		},

		setX: function ( x ) {

			this.x = x;

			return this;

		},

		setY: function ( y ) {

			this.y = y;

			return this;

		},

		setZ: function ( z ) {

			this.z = z;

			return this;

		},

		setComponent: function ( index, value ) {

			switch ( index ) {

				case 0: this.x = value; break;
				case 1: this.y = value; break;
				case 2: this.z = value; break;
				default: throw new Error( 'index is out of range: ' + index );

			}

			return this;

		},

		getComponent: function ( index ) {

			switch ( index ) {

				case 0: return this.x;
				case 1: return this.y;
				case 2: return this.z;
				default: throw new Error( 'index is out of range: ' + index );

			}

		},

		clone: function () {

			return new this.constructor( this.x, this.y, this.z );

		},

		copy: function ( v ) {

			this.x = v.x;
			this.y = v.y;
			this.z = v.z;

			return this;

		},

		add: function ( v, w ) {

			if ( w !== undefined ) {

				console.warn( 'THREE.Vector3: .add() now only accepts one argument. Use .addVectors( a, b ) instead.' );
				return this.addVectors( v, w );

			}

			this.x += v.x;
			this.y += v.y;
			this.z += v.z;

			return this;

		},

		addScalar: function ( s ) {

			this.x += s;
			this.y += s;
			this.z += s;

			return this;

		},

		addVectors: function ( a, b ) {

			this.x = a.x + b.x;
			this.y = a.y + b.y;
			this.z = a.z + b.z;

			return this;

		},

		addScaledVector: function ( v, s ) {

			this.x += v.x * s;
			this.y += v.y * s;
			this.z += v.z * s;

			return this;

		},

		sub: function ( v, w ) {

			if ( w !== undefined ) {

				console.warn( 'THREE.Vector3: .sub() now only accepts one argument. Use .subVectors( a, b ) instead.' );
				return this.subVectors( v, w );

			}

			this.x -= v.x;
			this.y -= v.y;
			this.z -= v.z;

			return this;

		},

		subScalar: function ( s ) {

			this.x -= s;
			this.y -= s;
			this.z -= s;

			return this;

		},

		subVectors: function ( a, b ) {

			this.x = a.x - b.x;
			this.y = a.y - b.y;
			this.z = a.z - b.z;

			return this;

		},

		multiply: function ( v, w ) {

			if ( w !== undefined ) {

				console.warn( 'THREE.Vector3: .multiply() now only accepts one argument. Use .multiplyVectors( a, b ) instead.' );
				return this.multiplyVectors( v, w );

			}

			this.x *= v.x;
			this.y *= v.y;
			this.z *= v.z;

			return this;

		},

		multiplyScalar: function ( scalar ) {

			if ( isFinite( scalar ) ) {

				this.x *= scalar;
				this.y *= scalar;
				this.z *= scalar;

			} else {

				this.x = 0;
				this.y = 0;
				this.z = 0;

			}

			return this;

		},

		multiplyVectors: function ( a, b ) {

			this.x = a.x * b.x;
			this.y = a.y * b.y;
			this.z = a.z * b.z;

			return this;

		},

		applyEuler: function () {

			var quaternion;

			return function applyEuler( euler ) {

				if ( (euler && euler.isEuler) === false ) {

					console.error( 'THREE.Vector3: .applyEuler() now expects an Euler rotation rather than a Vector3 and order.' );

				}

				if ( quaternion === undefined ) quaternion = new Quaternion();

				return this.applyQuaternion( quaternion.setFromEuler( euler ) );

			};

		}(),

		applyAxisAngle: function () {

			var quaternion;

			return function applyAxisAngle( axis, angle ) {

				if ( quaternion === undefined ) quaternion = new Quaternion();

				return this.applyQuaternion( quaternion.setFromAxisAngle( axis, angle ) );

			};

		}(),

		applyMatrix3: function ( m ) {

			var x = this.x, y = this.y, z = this.z;
			var e = m.elements;

			this.x = e[ 0 ] * x + e[ 3 ] * y + e[ 6 ] * z;
			this.y = e[ 1 ] * x + e[ 4 ] * y + e[ 7 ] * z;
			this.z = e[ 2 ] * x + e[ 5 ] * y + e[ 8 ] * z;

			return this;

		},

		applyMatrix4: function ( m ) {

			// input: THREE.Matrix4 affine matrix

			var x = this.x, y = this.y, z = this.z;
			var e = m.elements;

			this.x = e[ 0 ] * x + e[ 4 ] * y + e[ 8 ]  * z + e[ 12 ];
			this.y = e[ 1 ] * x + e[ 5 ] * y + e[ 9 ]  * z + e[ 13 ];
			this.z = e[ 2 ] * x + e[ 6 ] * y + e[ 10 ] * z + e[ 14 ];

			return this;

		},

		applyProjection: function ( m ) {

			// input: THREE.Matrix4 projection matrix

			var x = this.x, y = this.y, z = this.z;
			var e = m.elements;
			var d = 1 / ( e[ 3 ] * x + e[ 7 ] * y + e[ 11 ] * z + e[ 15 ] ); // perspective divide

			this.x = ( e[ 0 ] * x + e[ 4 ] * y + e[ 8 ]  * z + e[ 12 ] ) * d;
			this.y = ( e[ 1 ] * x + e[ 5 ] * y + e[ 9 ]  * z + e[ 13 ] ) * d;
			this.z = ( e[ 2 ] * x + e[ 6 ] * y + e[ 10 ] * z + e[ 14 ] ) * d;

			return this;

		},

		applyQuaternion: function ( q ) {

			var x = this.x, y = this.y, z = this.z;
			var qx = q.x, qy = q.y, qz = q.z, qw = q.w;

			// calculate quat * vector

			var ix =  qw * x + qy * z - qz * y;
			var iy =  qw * y + qz * x - qx * z;
			var iz =  qw * z + qx * y - qy * x;
			var iw = - qx * x - qy * y - qz * z;

			// calculate result * inverse quat

			this.x = ix * qw + iw * - qx + iy * - qz - iz * - qy;
			this.y = iy * qw + iw * - qy + iz * - qx - ix * - qz;
			this.z = iz * qw + iw * - qz + ix * - qy - iy * - qx;

			return this;

		},

		project: function () {

			var matrix;

			return function project( camera ) {

				if ( matrix === undefined ) matrix = new Matrix4();

				matrix.multiplyMatrices( camera.projectionMatrix, matrix.getInverse( camera.matrixWorld ) );
				return this.applyProjection( matrix );

			};

		}(),

		unproject: function () {

			var matrix;

			return function unproject( camera ) {

				if ( matrix === undefined ) matrix = new Matrix4();

				matrix.multiplyMatrices( camera.matrixWorld, matrix.getInverse( camera.projectionMatrix ) );
				return this.applyProjection( matrix );

			};

		}(),

		transformDirection: function ( m ) {

			// input: THREE.Matrix4 affine matrix
			// vector interpreted as a direction

			var x = this.x, y = this.y, z = this.z;
			var e = m.elements;

			this.x = e[ 0 ] * x + e[ 4 ] * y + e[ 8 ]  * z;
			this.y = e[ 1 ] * x + e[ 5 ] * y + e[ 9 ]  * z;
			this.z = e[ 2 ] * x + e[ 6 ] * y + e[ 10 ] * z;

			return this.normalize();

		},

		divide: function ( v ) {

			this.x /= v.x;
			this.y /= v.y;
			this.z /= v.z;

			return this;

		},

		divideScalar: function ( scalar ) {

			return this.multiplyScalar( 1 / scalar );

		},

		min: function ( v ) {

			this.x = Math.min( this.x, v.x );
			this.y = Math.min( this.y, v.y );
			this.z = Math.min( this.z, v.z );

			return this;

		},

		max: function ( v ) {

			this.x = Math.max( this.x, v.x );
			this.y = Math.max( this.y, v.y );
			this.z = Math.max( this.z, v.z );

			return this;

		},

		clamp: function ( min, max ) {

			// This function assumes min < max, if this assumption isn't true it will not operate correctly

			this.x = Math.max( min.x, Math.min( max.x, this.x ) );
			this.y = Math.max( min.y, Math.min( max.y, this.y ) );
			this.z = Math.max( min.z, Math.min( max.z, this.z ) );

			return this;

		},

		clampScalar: function () {

			var min, max;

			return function clampScalar( minVal, maxVal ) {

				if ( min === undefined ) {

					min = new Vector3();
					max = new Vector3();

				}

				min.set( minVal, minVal, minVal );
				max.set( maxVal, maxVal, maxVal );

				return this.clamp( min, max );

			};

		}(),

		clampLength: function ( min, max ) {

			var length = this.length();

			return this.multiplyScalar( Math.max( min, Math.min( max, length ) ) / length );

		},

		floor: function () {

			this.x = Math.floor( this.x );
			this.y = Math.floor( this.y );
			this.z = Math.floor( this.z );

			return this;

		},

		ceil: function () {

			this.x = Math.ceil( this.x );
			this.y = Math.ceil( this.y );
			this.z = Math.ceil( this.z );

			return this;

		},

		round: function () {

			this.x = Math.round( this.x );
			this.y = Math.round( this.y );
			this.z = Math.round( this.z );

			return this;

		},

		roundToZero: function () {

			this.x = ( this.x < 0 ) ? Math.ceil( this.x ) : Math.floor( this.x );
			this.y = ( this.y < 0 ) ? Math.ceil( this.y ) : Math.floor( this.y );
			this.z = ( this.z < 0 ) ? Math.ceil( this.z ) : Math.floor( this.z );

			return this;

		},

		negate: function () {

			this.x = - this.x;
			this.y = - this.y;
			this.z = - this.z;

			return this;

		},

		dot: function ( v ) {

			return this.x * v.x + this.y * v.y + this.z * v.z;

		},

		lengthSq: function () {

			return this.x * this.x + this.y * this.y + this.z * this.z;

		},

		length: function () {

			return Math.sqrt( this.x * this.x + this.y * this.y + this.z * this.z );

		},

		lengthManhattan: function () {

			return Math.abs( this.x ) + Math.abs( this.y ) + Math.abs( this.z );

		},

		normalize: function () {

			return this.divideScalar( this.length() );

		},

		setLength: function ( length ) {

			return this.multiplyScalar( length / this.length() );

		},

		lerp: function ( v, alpha ) {

			this.x += ( v.x - this.x ) * alpha;
			this.y += ( v.y - this.y ) * alpha;
			this.z += ( v.z - this.z ) * alpha;

			return this;

		},

		lerpVectors: function ( v1, v2, alpha ) {

			return this.subVectors( v2, v1 ).multiplyScalar( alpha ).add( v1 );

		},

		cross: function ( v, w ) {

			if ( w !== undefined ) {

				console.warn( 'THREE.Vector3: .cross() now only accepts one argument. Use .crossVectors( a, b ) instead.' );
				return this.crossVectors( v, w );

			}

			var x = this.x, y = this.y, z = this.z;

			this.x = y * v.z - z * v.y;
			this.y = z * v.x - x * v.z;
			this.z = x * v.y - y * v.x;

			return this;

		},

		crossVectors: function ( a, b ) {

			var ax = a.x, ay = a.y, az = a.z;
			var bx = b.x, by = b.y, bz = b.z;

			this.x = ay * bz - az * by;
			this.y = az * bx - ax * bz;
			this.z = ax * by - ay * bx;

			return this;

		},

		projectOnVector: function ( vector ) {

			var scalar = vector.dot( this ) / vector.lengthSq();

			return this.copy( vector ).multiplyScalar( scalar );

		},

		projectOnPlane: function () {

			var v1;

			return function projectOnPlane( planeNormal ) {

				if ( v1 === undefined ) v1 = new Vector3();

				v1.copy( this ).projectOnVector( planeNormal );

				return this.sub( v1 );

			};

		}(),

		reflect: function () {

			// reflect incident vector off plane orthogonal to normal
			// normal is assumed to have unit length

			var v1;

			return function reflect( normal ) {

				if ( v1 === undefined ) v1 = new Vector3();

				return this.sub( v1.copy( normal ).multiplyScalar( 2 * this.dot( normal ) ) );

			};

		}(),

		angleTo: function ( v ) {

			var theta = this.dot( v ) / ( Math.sqrt( this.lengthSq() * v.lengthSq() ) );

			// clamp, to handle numerical problems

			return Math.acos( _Math.clamp( theta, - 1, 1 ) );

		},

		distanceTo: function ( v ) {

			return Math.sqrt( this.distanceToSquared( v ) );

		},

		distanceToSquared: function ( v ) {

			var dx = this.x - v.x, dy = this.y - v.y, dz = this.z - v.z;

			return dx * dx + dy * dy + dz * dz;

		},

		distanceToManhattan: function ( v ) {

			return Math.abs( this.x - v.x ) + Math.abs( this.y - v.y ) + Math.abs( this.z - v.z );

		},

		setFromSpherical: function( s ) {

			var sinPhiRadius = Math.sin( s.phi ) * s.radius;

			this.x = sinPhiRadius * Math.sin( s.theta );
			this.y = Math.cos( s.phi ) * s.radius;
			this.z = sinPhiRadius * Math.cos( s.theta );

			return this;

		},

		setFromCylindrical: function( c ) {

			this.x = c.radius * Math.sin( c.theta );
			this.y = c.y;
			this.z = c.radius * Math.cos( c.theta );

			return this;

		},

		setFromMatrixPosition: function ( m ) {

			return this.setFromMatrixColumn( m, 3 );

		},

		setFromMatrixScale: function ( m ) {

			var sx = this.setFromMatrixColumn( m, 0 ).length();
			var sy = this.setFromMatrixColumn( m, 1 ).length();
			var sz = this.setFromMatrixColumn( m, 2 ).length();

			this.x = sx;
			this.y = sy;
			this.z = sz;

			return this;

		},

		setFromMatrixColumn: function ( m, index ) {

			if ( typeof m === 'number' ) {

				console.warn( 'THREE.Vector3: setFromMatrixColumn now expects ( matrix, index ).' );
				var temp = m;
				m = index;
				index = temp;

			}

			return this.fromArray( m.elements, index * 4 );

		},

		equals: function ( v ) {

			return ( ( v.x === this.x ) && ( v.y === this.y ) && ( v.z === this.z ) );

		},

		fromArray: function ( array, offset ) {

			if ( offset === undefined ) offset = 0;

			this.x = array[ offset ];
			this.y = array[ offset + 1 ];
			this.z = array[ offset + 2 ];

			return this;

		},

		toArray: function ( array, offset ) {

			if ( array === undefined ) array = [];
			if ( offset === undefined ) offset = 0;

			array[ offset ] = this.x;
			array[ offset + 1 ] = this.y;
			array[ offset + 2 ] = this.z;

			return array;

		},

		fromAttribute: function ( attribute, index, offset ) {

			if ( offset !== undefined ) {

				console.warn( 'THREE.Vector3: offset has been removed from .fromAttribute().' );

			}

			this.x = attribute.getX( index );
			this.y = attribute.getY( index );
			this.z = attribute.getZ( index );

			return this;

		}

	};

	/**
	 * @author mrdoob / http://mrdoob.com/
	 * @author supereggbert / http://www.paulbrunt.co.uk/
	 * @author philogb / http://blog.thejit.org/
	 * @author jordi_ros / http://plattsoft.com
	 * @author D1plo1d / http://github.com/D1plo1d
	 * @author alteredq / http://alteredqualia.com/
	 * @author mikael emtinger / http://gomo.se/
	 * @author timknip / http://www.floorplanner.com/
	 * @author bhouston / http://clara.io
	 * @author WestLangley / http://github.com/WestLangley
	 */

	function Matrix4() {

		this.elements = new Float32Array( [

			1, 0, 0, 0,
			0, 1, 0, 0,
			0, 0, 1, 0,
			0, 0, 0, 1

		] );

		if ( arguments.length > 0 ) {

			console.error( 'THREE.Matrix4: the constructor no longer reads arguments. use .set() instead.' );

		}

	}

	Matrix4.prototype = {

		constructor: Matrix4,

		isMatrix4: true,

		set: function ( n11, n12, n13, n14, n21, n22, n23, n24, n31, n32, n33, n34, n41, n42, n43, n44 ) {

			var te = this.elements;

			te[ 0 ] = n11; te[ 4 ] = n12; te[ 8 ] = n13; te[ 12 ] = n14;
			te[ 1 ] = n21; te[ 5 ] = n22; te[ 9 ] = n23; te[ 13 ] = n24;
			te[ 2 ] = n31; te[ 6 ] = n32; te[ 10 ] = n33; te[ 14 ] = n34;
			te[ 3 ] = n41; te[ 7 ] = n42; te[ 11 ] = n43; te[ 15 ] = n44;

			return this;

		},

		identity: function () {

			this.set(

				1, 0, 0, 0,
				0, 1, 0, 0,
				0, 0, 1, 0,
				0, 0, 0, 1

			);

			return this;

		},

		clone: function () {

			return new Matrix4().fromArray( this.elements );

		},

		copy: function ( m ) {

			this.elements.set( m.elements );

			return this;

		},

		copyPosition: function ( m ) {

			var te = this.elements;
			var me = m.elements;

			te[ 12 ] = me[ 12 ];
			te[ 13 ] = me[ 13 ];
			te[ 14 ] = me[ 14 ];

			return this;

		},

		extractBasis: function ( xAxis, yAxis, zAxis ) {

			xAxis.setFromMatrixColumn( this, 0 );
			yAxis.setFromMatrixColumn( this, 1 );
			zAxis.setFromMatrixColumn( this, 2 );

			return this;

		},

		makeBasis: function ( xAxis, yAxis, zAxis ) {

			this.set(
				xAxis.x, yAxis.x, zAxis.x, 0,
				xAxis.y, yAxis.y, zAxis.y, 0,
				xAxis.z, yAxis.z, zAxis.z, 0,
				0,       0,       0,       1
			);

			return this;

		},

		extractRotation: function () {

			var v1;

			return function extractRotation( m ) {

				if ( v1 === undefined ) v1 = new Vector3();

				var te = this.elements;
				var me = m.elements;

				var scaleX = 1 / v1.setFromMatrixColumn( m, 0 ).length();
				var scaleY = 1 / v1.setFromMatrixColumn( m, 1 ).length();
				var scaleZ = 1 / v1.setFromMatrixColumn( m, 2 ).length();

				te[ 0 ] = me[ 0 ] * scaleX;
				te[ 1 ] = me[ 1 ] * scaleX;
				te[ 2 ] = me[ 2 ] * scaleX;

				te[ 4 ] = me[ 4 ] * scaleY;
				te[ 5 ] = me[ 5 ] * scaleY;
				te[ 6 ] = me[ 6 ] * scaleY;

				te[ 8 ] = me[ 8 ] * scaleZ;
				te[ 9 ] = me[ 9 ] * scaleZ;
				te[ 10 ] = me[ 10 ] * scaleZ;

				return this;

			};

		}(),

		makeRotationFromEuler: function ( euler ) {

			if ( (euler && euler.isEuler) === false ) {

				console.error( 'THREE.Matrix: .makeRotationFromEuler() now expects a Euler rotation rather than a Vector3 and order.' );

			}

			var te = this.elements;

			var x = euler.x, y = euler.y, z = euler.z;
			var a = Math.cos( x ), b = Math.sin( x );
			var c = Math.cos( y ), d = Math.sin( y );
			var e = Math.cos( z ), f = Math.sin( z );

			if ( euler.order === 'XYZ' ) {

				var ae = a * e, af = a * f, be = b * e, bf = b * f;

				te[ 0 ] = c * e;
				te[ 4 ] = - c * f;
				te[ 8 ] = d;

				te[ 1 ] = af + be * d;
				te[ 5 ] = ae - bf * d;
				te[ 9 ] = - b * c;

				te[ 2 ] = bf - ae * d;
				te[ 6 ] = be + af * d;
				te[ 10 ] = a * c;

			} else if ( euler.order === 'YXZ' ) {

				var ce = c * e, cf = c * f, de = d * e, df = d * f;

				te[ 0 ] = ce + df * b;
				te[ 4 ] = de * b - cf;
				te[ 8 ] = a * d;

				te[ 1 ] = a * f;
				te[ 5 ] = a * e;
				te[ 9 ] = - b;

				te[ 2 ] = cf * b - de;
				te[ 6 ] = df + ce * b;
				te[ 10 ] = a * c;

			} else if ( euler.order === 'ZXY' ) {

				var ce = c * e, cf = c * f, de = d * e, df = d * f;

				te[ 0 ] = ce - df * b;
				te[ 4 ] = - a * f;
				te[ 8 ] = de + cf * b;

				te[ 1 ] = cf + de * b;
				te[ 5 ] = a * e;
				te[ 9 ] = df - ce * b;

				te[ 2 ] = - a * d;
				te[ 6 ] = b;
				te[ 10 ] = a * c;

			} else if ( euler.order === 'ZYX' ) {

				var ae = a * e, af = a * f, be = b * e, bf = b * f;

				te[ 0 ] = c * e;
				te[ 4 ] = be * d - af;
				te[ 8 ] = ae * d + bf;

				te[ 1 ] = c * f;
				te[ 5 ] = bf * d + ae;
				te[ 9 ] = af * d - be;

				te[ 2 ] = - d;
				te[ 6 ] = b * c;
				te[ 10 ] = a * c;

			} else if ( euler.order === 'YZX' ) {

				var ac = a * c, ad = a * d, bc = b * c, bd = b * d;

				te[ 0 ] = c * e;
				te[ 4 ] = bd - ac * f;
				te[ 8 ] = bc * f + ad;

				te[ 1 ] = f;
				te[ 5 ] = a * e;
				te[ 9 ] = - b * e;

				te[ 2 ] = - d * e;
				te[ 6 ] = ad * f + bc;
				te[ 10 ] = ac - bd * f;

			} else if ( euler.order === 'XZY' ) {

				var ac = a * c, ad = a * d, bc = b * c, bd = b * d;

				te[ 0 ] = c * e;
				te[ 4 ] = - f;
				te[ 8 ] = d * e;

				te[ 1 ] = ac * f + bd;
				te[ 5 ] = a * e;
				te[ 9 ] = ad * f - bc;

				te[ 2 ] = bc * f - ad;
				te[ 6 ] = b * e;
				te[ 10 ] = bd * f + ac;

			}

			// last column
			te[ 3 ] = 0;
			te[ 7 ] = 0;
			te[ 11 ] = 0;

			// bottom row
			te[ 12 ] = 0;
			te[ 13 ] = 0;
			te[ 14 ] = 0;
			te[ 15 ] = 1;

			return this;

		},

		makeRotationFromQuaternion: function ( q ) {

			var te = this.elements;

			var x = q.x, y = q.y, z = q.z, w = q.w;
			var x2 = x + x, y2 = y + y, z2 = z + z;
			var xx = x * x2, xy = x * y2, xz = x * z2;
			var yy = y * y2, yz = y * z2, zz = z * z2;
			var wx = w * x2, wy = w * y2, wz = w * z2;

			te[ 0 ] = 1 - ( yy + zz );
			te[ 4 ] = xy - wz;
			te[ 8 ] = xz + wy;

			te[ 1 ] = xy + wz;
			te[ 5 ] = 1 - ( xx + zz );
			te[ 9 ] = yz - wx;

			te[ 2 ] = xz - wy;
			te[ 6 ] = yz + wx;
			te[ 10 ] = 1 - ( xx + yy );

			// last column
			te[ 3 ] = 0;
			te[ 7 ] = 0;
			te[ 11 ] = 0;

			// bottom row
			te[ 12 ] = 0;
			te[ 13 ] = 0;
			te[ 14 ] = 0;
			te[ 15 ] = 1;

			return this;

		},

		lookAt: function () {

			var x, y, z;

			return function lookAt( eye, target, up ) {

				if ( x === undefined ) {

					x = new Vector3();
					y = new Vector3();
					z = new Vector3();

				}

				var te = this.elements;

				z.subVectors( eye, target ).normalize();

				if ( z.lengthSq() === 0 ) {

					z.z = 1;

				}

				x.crossVectors( up, z ).normalize();

				if ( x.lengthSq() === 0 ) {

					z.z += 0.0001;
					x.crossVectors( up, z ).normalize();

				}

				y.crossVectors( z, x );


				te[ 0 ] = x.x; te[ 4 ] = y.x; te[ 8 ] = z.x;
				te[ 1 ] = x.y; te[ 5 ] = y.y; te[ 9 ] = z.y;
				te[ 2 ] = x.z; te[ 6 ] = y.z; te[ 10 ] = z.z;

				return this;

			};

		}(),

		multiply: function ( m, n ) {

			if ( n !== undefined ) {

				console.warn( 'THREE.Matrix4: .multiply() now only accepts one argument. Use .multiplyMatrices( a, b ) instead.' );
				return this.multiplyMatrices( m, n );

			}

			return this.multiplyMatrices( this, m );

		},

		premultiply: function ( m ) {

			return this.multiplyMatrices( m, this );

		},

		multiplyMatrices: function ( a, b ) {

			var ae = a.elements;
			var be = b.elements;
			var te = this.elements;

			var a11 = ae[ 0 ], a12 = ae[ 4 ], a13 = ae[ 8 ], a14 = ae[ 12 ];
			var a21 = ae[ 1 ], a22 = ae[ 5 ], a23 = ae[ 9 ], a24 = ae[ 13 ];
			var a31 = ae[ 2 ], a32 = ae[ 6 ], a33 = ae[ 10 ], a34 = ae[ 14 ];
			var a41 = ae[ 3 ], a42 = ae[ 7 ], a43 = ae[ 11 ], a44 = ae[ 15 ];

			var b11 = be[ 0 ], b12 = be[ 4 ], b13 = be[ 8 ], b14 = be[ 12 ];
			var b21 = be[ 1 ], b22 = be[ 5 ], b23 = be[ 9 ], b24 = be[ 13 ];
			var b31 = be[ 2 ], b32 = be[ 6 ], b33 = be[ 10 ], b34 = be[ 14 ];
			var b41 = be[ 3 ], b42 = be[ 7 ], b43 = be[ 11 ], b44 = be[ 15 ];

			te[ 0 ] = a11 * b11 + a12 * b21 + a13 * b31 + a14 * b41;
			te[ 4 ] = a11 * b12 + a12 * b22 + a13 * b32 + a14 * b42;
			te[ 8 ] = a11 * b13 + a12 * b23 + a13 * b33 + a14 * b43;
			te[ 12 ] = a11 * b14 + a12 * b24 + a13 * b34 + a14 * b44;

			te[ 1 ] = a21 * b11 + a22 * b21 + a23 * b31 + a24 * b41;
			te[ 5 ] = a21 * b12 + a22 * b22 + a23 * b32 + a24 * b42;
			te[ 9 ] = a21 * b13 + a22 * b23 + a23 * b33 + a24 * b43;
			te[ 13 ] = a21 * b14 + a22 * b24 + a23 * b34 + a24 * b44;

			te[ 2 ] = a31 * b11 + a32 * b21 + a33 * b31 + a34 * b41;
			te[ 6 ] = a31 * b12 + a32 * b22 + a33 * b32 + a34 * b42;
			te[ 10 ] = a31 * b13 + a32 * b23 + a33 * b33 + a34 * b43;
			te[ 14 ] = a31 * b14 + a32 * b24 + a33 * b34 + a34 * b44;

			te[ 3 ] = a41 * b11 + a42 * b21 + a43 * b31 + a44 * b41;
			te[ 7 ] = a41 * b12 + a42 * b22 + a43 * b32 + a44 * b42;
			te[ 11 ] = a41 * b13 + a42 * b23 + a43 * b33 + a44 * b43;
			te[ 15 ] = a41 * b14 + a42 * b24 + a43 * b34 + a44 * b44;

			return this;

		},

		multiplyToArray: function ( a, b, r ) {

			var te = this.elements;

			this.multiplyMatrices( a, b );

			r[ 0 ] = te[ 0 ]; r[ 1 ] = te[ 1 ]; r[ 2 ] = te[ 2 ]; r[ 3 ] = te[ 3 ];
			r[ 4 ] = te[ 4 ]; r[ 5 ] = te[ 5 ]; r[ 6 ] = te[ 6 ]; r[ 7 ] = te[ 7 ];
			r[ 8 ]  = te[ 8 ]; r[ 9 ]  = te[ 9 ]; r[ 10 ] = te[ 10 ]; r[ 11 ] = te[ 11 ];
			r[ 12 ] = te[ 12 ]; r[ 13 ] = te[ 13 ]; r[ 14 ] = te[ 14 ]; r[ 15 ] = te[ 15 ];

			return this;

		},

		multiplyScalar: function ( s ) {

			var te = this.elements;

			te[ 0 ] *= s; te[ 4 ] *= s; te[ 8 ] *= s; te[ 12 ] *= s;
			te[ 1 ] *= s; te[ 5 ] *= s; te[ 9 ] *= s; te[ 13 ] *= s;
			te[ 2 ] *= s; te[ 6 ] *= s; te[ 10 ] *= s; te[ 14 ] *= s;
			te[ 3 ] *= s; te[ 7 ] *= s; te[ 11 ] *= s; te[ 15 ] *= s;

			return this;

		},

		applyToVector3Array: function () {

			var v1;

			return function applyToVector3Array( array, offset, length ) {

				if ( v1 === undefined ) v1 = new Vector3();
				if ( offset === undefined ) offset = 0;
				if ( length === undefined ) length = array.length;

				for ( var i = 0, j = offset; i < length; i += 3, j += 3 ) {

					v1.fromArray( array, j );
					v1.applyMatrix4( this );
					v1.toArray( array, j );

				}

				return array;

			};

		}(),

		applyToBufferAttribute: function () {

			var v1;

			return function applyToBufferAttribute( attribute ) {

				if ( v1 === undefined ) v1 = new Vector3();

				for ( var i = 0, l = attribute.count; i < l; i ++ ) {

					v1.x = attribute.getX( i );
					v1.y = attribute.getY( i );
					v1.z = attribute.getZ( i );

					v1.applyMatrix4( this );

					attribute.setXYZ( i, v1.x, v1.y, v1.z );

				}

				return attribute;

			};

		}(),

		determinant: function () {

			var te = this.elements;

			var n11 = te[ 0 ], n12 = te[ 4 ], n13 = te[ 8 ], n14 = te[ 12 ];
			var n21 = te[ 1 ], n22 = te[ 5 ], n23 = te[ 9 ], n24 = te[ 13 ];
			var n31 = te[ 2 ], n32 = te[ 6 ], n33 = te[ 10 ], n34 = te[ 14 ];
			var n41 = te[ 3 ], n42 = te[ 7 ], n43 = te[ 11 ], n44 = te[ 15 ];

			//TODO: make this more efficient
			//( based on http://www.euclideanspace.com/maths/algebra/matrix/functions/inverse/fourD/index.htm )

			return (
				n41 * (
					+ n14 * n23 * n32
					 - n13 * n24 * n32
					 - n14 * n22 * n33
					 + n12 * n24 * n33
					 + n13 * n22 * n34
					 - n12 * n23 * n34
				) +
				n42 * (
					+ n11 * n23 * n34
					 - n11 * n24 * n33
					 + n14 * n21 * n33
					 - n13 * n21 * n34
					 + n13 * n24 * n31
					 - n14 * n23 * n31
				) +
				n43 * (
					+ n11 * n24 * n32
					 - n11 * n22 * n34
					 - n14 * n21 * n32
					 + n12 * n21 * n34
					 + n14 * n22 * n31
					 - n12 * n24 * n31
				) +
				n44 * (
					- n13 * n22 * n31
					 - n11 * n23 * n32
					 + n11 * n22 * n33
					 + n13 * n21 * n32
					 - n12 * n21 * n33
					 + n12 * n23 * n31
				)

			);

		},

		transpose: function () {

			var te = this.elements;
			var tmp;

			tmp = te[ 1 ]; te[ 1 ] = te[ 4 ]; te[ 4 ] = tmp;
			tmp = te[ 2 ]; te[ 2 ] = te[ 8 ]; te[ 8 ] = tmp;
			tmp = te[ 6 ]; te[ 6 ] = te[ 9 ]; te[ 9 ] = tmp;

			tmp = te[ 3 ]; te[ 3 ] = te[ 12 ]; te[ 12 ] = tmp;
			tmp = te[ 7 ]; te[ 7 ] = te[ 13 ]; te[ 13 ] = tmp;
			tmp = te[ 11 ]; te[ 11 ] = te[ 14 ]; te[ 14 ] = tmp;

			return this;

		},

		setPosition: function ( v ) {

			var te = this.elements;

			te[ 12 ] = v.x;
			te[ 13 ] = v.y;
			te[ 14 ] = v.z;

			return this;

		},

		getInverse: function ( m, throwOnDegenerate ) {

			// based on http://www.euclideanspace.com/maths/algebra/matrix/functions/inverse/fourD/index.htm
			var te = this.elements,
				me = m.elements,

				n11 = me[ 0 ], n21 = me[ 1 ], n31 = me[ 2 ], n41 = me[ 3 ],
				n12 = me[ 4 ], n22 = me[ 5 ], n32 = me[ 6 ], n42 = me[ 7 ],
				n13 = me[ 8 ], n23 = me[ 9 ], n33 = me[ 10 ], n43 = me[ 11 ],
				n14 = me[ 12 ], n24 = me[ 13 ], n34 = me[ 14 ], n44 = me[ 15 ],

				t11 = n23 * n34 * n42 - n24 * n33 * n42 + n24 * n32 * n43 - n22 * n34 * n43 - n23 * n32 * n44 + n22 * n33 * n44,
				t12 = n14 * n33 * n42 - n13 * n34 * n42 - n14 * n32 * n43 + n12 * n34 * n43 + n13 * n32 * n44 - n12 * n33 * n44,
				t13 = n13 * n24 * n42 - n14 * n23 * n42 + n14 * n22 * n43 - n12 * n24 * n43 - n13 * n22 * n44 + n12 * n23 * n44,
				t14 = n14 * n23 * n32 - n13 * n24 * n32 - n14 * n22 * n33 + n12 * n24 * n33 + n13 * n22 * n34 - n12 * n23 * n34;

			var det = n11 * t11 + n21 * t12 + n31 * t13 + n41 * t14;

			if ( det === 0 ) {

				var msg = "THREE.Matrix4.getInverse(): can't invert matrix, determinant is 0";

				if ( throwOnDegenerate === true ) {

					throw new Error( msg );

				} else {

					console.warn( msg );

				}

				return this.identity();

			}

			var detInv = 1 / det;

			te[ 0 ] = t11 * detInv;
			te[ 1 ] = ( n24 * n33 * n41 - n23 * n34 * n41 - n24 * n31 * n43 + n21 * n34 * n43 + n23 * n31 * n44 - n21 * n33 * n44 ) * detInv;
			te[ 2 ] = ( n22 * n34 * n41 - n24 * n32 * n41 + n24 * n31 * n42 - n21 * n34 * n42 - n22 * n31 * n44 + n21 * n32 * n44 ) * detInv;
			te[ 3 ] = ( n23 * n32 * n41 - n22 * n33 * n41 - n23 * n31 * n42 + n21 * n33 * n42 + n22 * n31 * n43 - n21 * n32 * n43 ) * detInv;

			te[ 4 ] = t12 * detInv;
			te[ 5 ] = ( n13 * n34 * n41 - n14 * n33 * n41 + n14 * n31 * n43 - n11 * n34 * n43 - n13 * n31 * n44 + n11 * n33 * n44 ) * detInv;
			te[ 6 ] = ( n14 * n32 * n41 - n12 * n34 * n41 - n14 * n31 * n42 + n11 * n34 * n42 + n12 * n31 * n44 - n11 * n32 * n44 ) * detInv;
			te[ 7 ] = ( n12 * n33 * n41 - n13 * n32 * n41 + n13 * n31 * n42 - n11 * n33 * n42 - n12 * n31 * n43 + n11 * n32 * n43 ) * detInv;

			te[ 8 ] = t13 * detInv;
			te[ 9 ] = ( n14 * n23 * n41 - n13 * n24 * n41 - n14 * n21 * n43 + n11 * n24 * n43 + n13 * n21 * n44 - n11 * n23 * n44 ) * detInv;
			te[ 10 ] = ( n12 * n24 * n41 - n14 * n22 * n41 + n14 * n21 * n42 - n11 * n24 * n42 - n12 * n21 * n44 + n11 * n22 * n44 ) * detInv;
			te[ 11 ] = ( n13 * n22 * n41 - n12 * n23 * n41 - n13 * n21 * n42 + n11 * n23 * n42 + n12 * n21 * n43 - n11 * n22 * n43 ) * detInv;

			te[ 12 ] = t14 * detInv;
			te[ 13 ] = ( n13 * n24 * n31 - n14 * n23 * n31 + n14 * n21 * n33 - n11 * n24 * n33 - n13 * n21 * n34 + n11 * n23 * n34 ) * detInv;
			te[ 14 ] = ( n14 * n22 * n31 - n12 * n24 * n31 - n14 * n21 * n32 + n11 * n24 * n32 + n12 * n21 * n34 - n11 * n22 * n34 ) * detInv;
			te[ 15 ] = ( n12 * n23 * n31 - n13 * n22 * n31 + n13 * n21 * n32 - n11 * n23 * n32 - n12 * n21 * n33 + n11 * n22 * n33 ) * detInv;

			return this;

		},

		scale: function ( v ) {

			var te = this.elements;
			var x = v.x, y = v.y, z = v.z;

			te[ 0 ] *= x; te[ 4 ] *= y; te[ 8 ] *= z;
			te[ 1 ] *= x; te[ 5 ] *= y; te[ 9 ] *= z;
			te[ 2 ] *= x; te[ 6 ] *= y; te[ 10 ] *= z;
			te[ 3 ] *= x; te[ 7 ] *= y; te[ 11 ] *= z;

			return this;

		},

		getMaxScaleOnAxis: function () {

			var te = this.elements;

			var scaleXSq = te[ 0 ] * te[ 0 ] + te[ 1 ] * te[ 1 ] + te[ 2 ] * te[ 2 ];
			var scaleYSq = te[ 4 ] * te[ 4 ] + te[ 5 ] * te[ 5 ] + te[ 6 ] * te[ 6 ];
			var scaleZSq = te[ 8 ] * te[ 8 ] + te[ 9 ] * te[ 9 ] + te[ 10 ] * te[ 10 ];

			return Math.sqrt( Math.max( scaleXSq, scaleYSq, scaleZSq ) );

		},

		makeTranslation: function ( x, y, z ) {

			this.set(

				1, 0, 0, x,
				0, 1, 0, y,
				0, 0, 1, z,
				0, 0, 0, 1

			);

			return this;

		},

		makeRotationX: function ( theta ) {

			var c = Math.cos( theta ), s = Math.sin( theta );

			this.set(

				1, 0,  0, 0,
				0, c, - s, 0,
				0, s,  c, 0,
				0, 0,  0, 1

			);

			return this;

		},

		makeRotationY: function ( theta ) {

			var c = Math.cos( theta ), s = Math.sin( theta );

			this.set(

				 c, 0, s, 0,
				 0, 1, 0, 0,
				- s, 0, c, 0,
				 0, 0, 0, 1

			);

			return this;

		},

		makeRotationZ: function ( theta ) {

			var c = Math.cos( theta ), s = Math.sin( theta );

			this.set(

				c, - s, 0, 0,
				s,  c, 0, 0,
				0,  0, 1, 0,
				0,  0, 0, 1

			);

			return this;

		},

		makeRotationAxis: function ( axis, angle ) {

			// Based on http://www.gamedev.net/reference/articles/article1199.asp

			var c = Math.cos( angle );
			var s = Math.sin( angle );
			var t = 1 - c;
			var x = axis.x, y = axis.y, z = axis.z;
			var tx = t * x, ty = t * y;

			this.set(

				tx * x + c, tx * y - s * z, tx * z + s * y, 0,
				tx * y + s * z, ty * y + c, ty * z - s * x, 0,
				tx * z - s * y, ty * z + s * x, t * z * z + c, 0,
				0, 0, 0, 1

			);

			 return this;

		},

		makeScale: function ( x, y, z ) {

			this.set(

				x, 0, 0, 0,
				0, y, 0, 0,
				0, 0, z, 0,
				0, 0, 0, 1

			);

			return this;

		},

		makeShear: function ( x, y, z ) {

			this.set(

				1, y, z, 0,
				x, 1, z, 0,
				x, y, 1, 0,
				0, 0, 0, 1

			);

			return this;

		},

		compose: function ( position, quaternion, scale ) {

			this.makeRotationFromQuaternion( quaternion );
			this.scale( scale );
			this.setPosition( position );

			return this;

		},

		decompose: function () {

			var vector, matrix;

			return function decompose( position, quaternion, scale ) {

				if ( vector === undefined ) {

					vector = new Vector3();
					matrix = new Matrix4();

				}

				var te = this.elements;

				var sx = vector.set( te[ 0 ], te[ 1 ], te[ 2 ] ).length();
				var sy = vector.set( te[ 4 ], te[ 5 ], te[ 6 ] ).length();
				var sz = vector.set( te[ 8 ], te[ 9 ], te[ 10 ] ).length();

				// if determine is negative, we need to invert one scale
				var det = this.determinant();
				if ( det < 0 ) {

					sx = - sx;

				}

				position.x = te[ 12 ];
				position.y = te[ 13 ];
				position.z = te[ 14 ];

				// scale the rotation part

				matrix.elements.set( this.elements ); // at this point matrix is incomplete so we can't use .copy()

				var invSX = 1 / sx;
				var invSY = 1 / sy;
				var invSZ = 1 / sz;

				matrix.elements[ 0 ] *= invSX;
				matrix.elements[ 1 ] *= invSX;
				matrix.elements[ 2 ] *= invSX;

				matrix.elements[ 4 ] *= invSY;
				matrix.elements[ 5 ] *= invSY;
				matrix.elements[ 6 ] *= invSY;

				matrix.elements[ 8 ] *= invSZ;
				matrix.elements[ 9 ] *= invSZ;
				matrix.elements[ 10 ] *= invSZ;

				quaternion.setFromRotationMatrix( matrix );

				scale.x = sx;
				scale.y = sy;
				scale.z = sz;

				return this;

			};

		}(),

		makeFrustum: function ( left, right, bottom, top, near, far ) {

			var te = this.elements;
			var x = 2 * near / ( right - left );
			var y = 2 * near / ( top - bottom );

			var a = ( right + left ) / ( right - left );
			var b = ( top + bottom ) / ( top - bottom );
			var c = - ( far + near ) / ( far - near );
			var d = - 2 * far * near / ( far - near );

			te[ 0 ] = x;	te[ 4 ] = 0;	te[ 8 ] = a;	te[ 12 ] = 0;
			te[ 1 ] = 0;	te[ 5 ] = y;	te[ 9 ] = b;	te[ 13 ] = 0;
			te[ 2 ] = 0;	te[ 6 ] = 0;	te[ 10 ] = c;	te[ 14 ] = d;
			te[ 3 ] = 0;	te[ 7 ] = 0;	te[ 11 ] = - 1;	te[ 15 ] = 0;

			return this;

		},

		makePerspective: function ( fov, aspect, near, far ) {

			var ymax = near * Math.tan( _Math.DEG2RAD * fov * 0.5 );
			var ymin = - ymax;
			var xmin = ymin * aspect;
			var xmax = ymax * aspect;

			return this.makeFrustum( xmin, xmax, ymin, ymax, near, far );

		},

		makeOrthographic: function ( left, right, top, bottom, near, far ) {

			var te = this.elements;
			var w = 1.0 / ( right - left );
			var h = 1.0 / ( top - bottom );
			var p = 1.0 / ( far - near );

			var x = ( right + left ) * w;
			var y = ( top + bottom ) * h;
			var z = ( far + near ) * p;

			te[ 0 ] = 2 * w;	te[ 4 ] = 0;	te[ 8 ] = 0;	te[ 12 ] = - x;
			te[ 1 ] = 0;	te[ 5 ] = 2 * h;	te[ 9 ] = 0;	te[ 13 ] = - y;
			te[ 2 ] = 0;	te[ 6 ] = 0;	te[ 10 ] = - 2 * p;	te[ 14 ] = - z;
			te[ 3 ] = 0;	te[ 7 ] = 0;	te[ 11 ] = 0;	te[ 15 ] = 1;

			return this;

		},

		equals: function ( matrix ) {

			var te = this.elements;
			var me = matrix.elements;

			for ( var i = 0; i < 16; i ++ ) {

				if ( te[ i ] !== me[ i ] ) return false;

			}

			return true;

		},

		fromArray: function ( array, offset ) {

			if ( offset === undefined ) offset = 0;

			for( var i = 0; i < 16; i ++ ) {

				this.elements[ i ] = array[ i + offset ];

			}

			return this;

		},

		toArray: function ( array, offset ) {

			if ( array === undefined ) array = [];
			if ( offset === undefined ) offset = 0;

			var te = this.elements;

			array[ offset ] = te[ 0 ];
			array[ offset + 1 ] = te[ 1 ];
			array[ offset + 2 ] = te[ 2 ];
			array[ offset + 3 ] = te[ 3 ];

			array[ offset + 4 ] = te[ 4 ];
			array[ offset + 5 ] = te[ 5 ];
			array[ offset + 6 ] = te[ 6 ];
			array[ offset + 7 ] = te[ 7 ];

			array[ offset + 8 ]  = te[ 8 ];
			array[ offset + 9 ]  = te[ 9 ];
			array[ offset + 10 ] = te[ 10 ];
			array[ offset + 11 ] = te[ 11 ];

			array[ offset + 12 ] = te[ 12 ];
			array[ offset + 13 ] = te[ 13 ];
			array[ offset + 14 ] = te[ 14 ];
			array[ offset + 15 ] = te[ 15 ];

			return array;

		}

	};

	/**
	 * @author mrdoob / http://mrdoob.com/
	 */

	function CubeTexture( images, mapping, wrapS, wrapT, magFilter, minFilter, format, type, anisotropy, encoding ) {

		images = images !== undefined ? images : [];
		mapping = mapping !== undefined ? mapping : CubeReflectionMapping;

		Texture.call( this, images, mapping, wrapS, wrapT, magFilter, minFilter, format, type, anisotropy, encoding );

		this.flipY = false;

	}

	CubeTexture.prototype = Object.create( Texture.prototype );
	CubeTexture.prototype.constructor = CubeTexture;

	CubeTexture.prototype.isCubeTexture = true;

	Object.defineProperty( CubeTexture.prototype, 'images', {

		get: function () {

			return this.image;

		},

		set: function ( value ) {

			this.image = value;

		}

	} );

	/**
	 * @author tschw
	 *
	 * Uniforms of a program.
	 * Those form a tree structure with a special top-level container for the root,
	 * which you get by calling 'new WebGLUniforms( gl, program, renderer )'.
	 *
	 *
	 * Properties of inner nodes including the top-level container:
	 *
	 * .seq - array of nested uniforms
	 * .map - nested uniforms by name
	 *
	 *
	 * Methods of all nodes except the top-level container:
	 *
	 * .setValue( gl, value, [renderer] )
	 *
	 * 		uploads a uniform value(s)
	 *  	the 'renderer' parameter is needed for sampler uniforms
	 *
	 *
	 * Static methods of the top-level container (renderer factorizations):
	 *
	 * .upload( gl, seq, values, renderer )
	 *
	 * 		sets uniforms in 'seq' to 'values[id].value'
	 *
	 * .seqWithValue( seq, values ) : filteredSeq
	 *
	 * 		filters 'seq' entries with corresponding entry in values
	 *
	 *
	 * Methods of the top-level container (renderer factorizations):
	 *
	 * .setValue( gl, name, value )
	 *
	 * 		sets uniform with  name 'name' to 'value'
	 *
	 * .set( gl, obj, prop )
	 *
	 * 		sets uniform from object and property with same name than uniform
	 *
	 * .setOptional( gl, obj, prop )
	 *
	 * 		like .set for an optional property of the object
	 *
	 */

	var emptyTexture = new Texture();
	var emptyCubeTexture = new CubeTexture();

	// --- Base for inner nodes (including the root) ---

	function UniformContainer() {

		this.seq = [];
		this.map = {};

	}

	// --- Utilities ---

	// Array Caches (provide typed arrays for temporary by size)

	var arrayCacheF32 = [];
	var arrayCacheI32 = [];

	// Flattening for arrays of vectors and matrices

	function flatten( array, nBlocks, blockSize ) {

		var firstElem = array[ 0 ];

		if ( firstElem <= 0 || firstElem > 0 ) return array;
		// unoptimized: ! isNaN( firstElem )
		// see http://jacksondunstan.com/articles/983

		var n = nBlocks * blockSize,
			r = arrayCacheF32[ n ];

		if ( r === undefined ) {

			r = new Float32Array( n );
			arrayCacheF32[ n ] = r;

		}

		if ( nBlocks !== 0 ) {

			firstElem.toArray( r, 0 );

			for ( var i = 1, offset = 0; i !== nBlocks; ++ i ) {

				offset += blockSize;
				array[ i ].toArray( r, offset );

			}

		}

		return r;

	}

	// Texture unit allocation

	function allocTexUnits( renderer, n ) {

		var r = arrayCacheI32[ n ];

		if ( r === undefined ) {

			r = new Int32Array( n );
			arrayCacheI32[ n ] = r;

		}

		for ( var i = 0; i !== n; ++ i )
			r[ i ] = renderer.allocTextureUnit();

		return r;

	}

	// --- Setters ---

	// Note: Defining these methods externally, because they come in a bunch
	// and this way their names minify.

	// Single scalar

	function setValue1f( gl, v ) { gl.uniform1f( this.addr, v ); }
	function setValue1i( gl, v ) { gl.uniform1i( this.addr, v ); }

	// Single float vector (from flat array or THREE.VectorN)

	function setValue2fv( gl, v ) {

		if ( v.x === undefined ) gl.uniform2fv( this.addr, v );
		else gl.uniform2f( this.addr, v.x, v.y );

	}

	function setValue3fv( gl, v ) {

		if ( v.x !== undefined )
			gl.uniform3f( this.addr, v.x, v.y, v.z );
		else if ( v.r !== undefined )
			gl.uniform3f( this.addr, v.r, v.g, v.b );
		else
			gl.uniform3fv( this.addr, v );

	}

	function setValue4fv( gl, v ) {

		if ( v.x === undefined ) gl.uniform4fv( this.addr, v );
		else gl.uniform4f( this.addr, v.x, v.y, v.z, v.w );

	}

	// Single matrix (from flat array or MatrixN)

	function setValue2fm( gl, v ) {

		gl.uniformMatrix2fv( this.addr, false, v.elements || v );

	}

	function setValue3fm( gl, v ) {

		gl.uniformMatrix3fv( this.addr, false, v.elements || v );

	}

	function setValue4fm( gl, v ) {

		gl.uniformMatrix4fv( this.addr, false, v.elements || v );

	}

	// Single texture (2D / Cube)

	function setValueT1( gl, v, renderer ) {

		var unit = renderer.allocTextureUnit();
		gl.uniform1i( this.addr, unit );
		renderer.setTexture2D( v || emptyTexture, unit );

	}

	function setValueT6( gl, v, renderer ) {

		var unit = renderer.allocTextureUnit();
		gl.uniform1i( this.addr, unit );
		renderer.setTextureCube( v || emptyCubeTexture, unit );

	}

	// Integer / Boolean vectors or arrays thereof (always flat arrays)

	function setValue2iv( gl, v ) { gl.uniform2iv( this.addr, v ); }
	function setValue3iv( gl, v ) { gl.uniform3iv( this.addr, v ); }
	function setValue4iv( gl, v ) { gl.uniform4iv( this.addr, v ); }

	// Helper to pick the right setter for the singular case

	function getSingularSetter( type ) {

		switch ( type ) {

			case 0x1406: return setValue1f; // FLOAT
			case 0x8b50: return setValue2fv; // _VEC2
			case 0x8b51: return setValue3fv; // _VEC3
			case 0x8b52: return setValue4fv; // _VEC4

			case 0x8b5a: return setValue2fm; // _MAT2
			case 0x8b5b: return setValue3fm; // _MAT3
			case 0x8b5c: return setValue4fm; // _MAT4

			case 0x8b5e: return setValueT1; // SAMPLER_2D
			case 0x8b60: return setValueT6; // SAMPLER_CUBE

			case 0x1404: case 0x8b56: return setValue1i; // INT, BOOL
			case 0x8b53: case 0x8b57: return setValue2iv; // _VEC2
			case 0x8b54: case 0x8b58: return setValue3iv; // _VEC3
			case 0x8b55: case 0x8b59: return setValue4iv; // _VEC4

		}

	}

	// Array of scalars

	function setValue1fv( gl, v ) { gl.uniform1fv( this.addr, v ); }
	function setValue1iv( gl, v ) { gl.uniform1iv( this.addr, v ); }

	// Array of vectors (flat or from THREE classes)

	function setValueV2a( gl, v ) {

		gl.uniform2fv( this.addr, flatten( v, this.size, 2 ) );

	}

	function setValueV3a( gl, v ) {

		gl.uniform3fv( this.addr, flatten( v, this.size, 3 ) );

	}

	function setValueV4a( gl, v ) {

		gl.uniform4fv( this.addr, flatten( v, this.size, 4 ) );

	}

	// Array of matrices (flat or from THREE clases)

	function setValueM2a( gl, v ) {

		gl.uniformMatrix2fv( this.addr, false, flatten( v, this.size, 4 ) );

	}

	function setValueM3a( gl, v ) {

		gl.uniformMatrix3fv( this.addr, false, flatten( v, this.size, 9 ) );

	}

	function setValueM4a( gl, v ) {

		gl.uniformMatrix4fv( this.addr, false, flatten( v, this.size, 16 ) );

	}

	// Array of textures (2D / Cube)

	function setValueT1a( gl, v, renderer ) {

		var n = v.length,
			units = allocTexUnits( renderer, n );

		gl.uniform1iv( this.addr, units );

		for ( var i = 0; i !== n; ++ i ) {

			renderer.setTexture2D( v[ i ] || emptyTexture, units[ i ] );

		}

	}

	function setValueT6a( gl, v, renderer ) {

		var n = v.length,
			units = allocTexUnits( renderer, n );

		gl.uniform1iv( this.addr, units );

		for ( var i = 0; i !== n; ++ i ) {

			renderer.setTextureCube( v[ i ] || emptyCubeTexture, units[ i ] );

		}

	}

	// Helper to pick the right setter for a pure (bottom-level) array

	function getPureArraySetter( type ) {

		switch ( type ) {

			case 0x1406: return setValue1fv; // FLOAT
			case 0x8b50: return setValueV2a; // _VEC2
			case 0x8b51: return setValueV3a; // _VEC3
			case 0x8b52: return setValueV4a; // _VEC4

			case 0x8b5a: return setValueM2a; // _MAT2
			case 0x8b5b: return setValueM3a; // _MAT3
			case 0x8b5c: return setValueM4a; // _MAT4

			case 0x8b5e: return setValueT1a; // SAMPLER_2D
			case 0x8b60: return setValueT6a; // SAMPLER_CUBE

			case 0x1404: case 0x8b56: return setValue1iv; // INT, BOOL
			case 0x8b53: case 0x8b57: return setValue2iv; // _VEC2
			case 0x8b54: case 0x8b58: return setValue3iv; // _VEC3
			case 0x8b55: case 0x8b59: return setValue4iv; // _VEC4

		}

	}

	// --- Uniform Classes ---

	function SingleUniform( id, activeInfo, addr ) {

		this.id = id;
		this.addr = addr;
		this.setValue = getSingularSetter( activeInfo.type );

		// this.path = activeInfo.name; // DEBUG

	}

	function PureArrayUniform( id, activeInfo, addr ) {

		this.id = id;
		this.addr = addr;
		this.size = activeInfo.size;
		this.setValue = getPureArraySetter( activeInfo.type );

		// this.path = activeInfo.name; // DEBUG

	}

	function StructuredUniform( id ) {

		this.id = id;

		UniformContainer.call( this ); // mix-in

	}

	StructuredUniform.prototype.setValue = function( gl, value ) {

		// Note: Don't need an extra 'renderer' parameter, since samplers
		// are not allowed in structured uniforms.

		var seq = this.seq;

		for ( var i = 0, n = seq.length; i !== n; ++ i ) {

			var u = seq[ i ];
			u.setValue( gl, value[ u.id ] );

		}

	};

	// --- Top-level ---

	// Parser - builds up the property tree from the path strings

	var RePathPart = /([\w\d_]+)(\])?(\[|\.)?/g;

	// extracts
	// 	- the identifier (member name or array index)
	//  - followed by an optional right bracket (found when array index)
	//  - followed by an optional left bracket or dot (type of subscript)
	//
	// Note: These portions can be read in a non-overlapping fashion and
	// allow straightforward parsing of the hierarchy that WebGL encodes
	// in the uniform names.

	function addUniform( container, uniformObject ) {

		container.seq.push( uniformObject );
		container.map[ uniformObject.id ] = uniformObject;

	}

	function parseUniform( activeInfo, addr, container ) {

		var path = activeInfo.name,
			pathLength = path.length;

		// reset RegExp object, because of the early exit of a previous run
		RePathPart.lastIndex = 0;

		for (; ;) {

			var match = RePathPart.exec( path ),
				matchEnd = RePathPart.lastIndex,

				id = match[ 1 ],
				idIsIndex = match[ 2 ] === ']',
				subscript = match[ 3 ];

			if ( idIsIndex ) id = id | 0; // convert to integer

			if ( subscript === undefined ||
					subscript === '[' && matchEnd + 2 === pathLength ) {
				// bare name or "pure" bottom-level array "[0]" suffix

				addUniform( container, subscript === undefined ?
						new SingleUniform( id, activeInfo, addr ) :
						new PureArrayUniform( id, activeInfo, addr ) );

				break;

			} else {
				// step into inner node / create it in case it doesn't exist

				var map = container.map,
					next = map[ id ];

				if ( next === undefined ) {

					next = new StructuredUniform( id );
					addUniform( container, next );

				}

				container = next;

			}

		}

	}

	// Root Container

	function WebGLUniforms( gl, program, renderer ) {

		UniformContainer.call( this );

		this.renderer = renderer;

		var n = gl.getProgramParameter( program, gl.ACTIVE_UNIFORMS );

		for ( var i = 0; i !== n; ++ i ) {

			var info = gl.getActiveUniform( program, i ),
				path = info.name,
				addr = gl.getUniformLocation( program, path );

			parseUniform( info, addr, this );

		}

	}

	WebGLUniforms.prototype.setValue = function( gl, name, value ) {

		var u = this.map[ name ];

		if ( u !== undefined ) u.setValue( gl, value, this.renderer );

	};

	WebGLUniforms.prototype.set = function( gl, object, name ) {

		var u = this.map[ name ];

		if ( u !== undefined ) u.setValue( gl, object[ name ], this.renderer );

	};

	WebGLUniforms.prototype.setOptional = function( gl, object, name ) {

		var v = object[ name ];

		if ( v !== undefined ) this.setValue( gl, name, v );

	};


	// Static interface

	WebGLUniforms.upload = function( gl, seq, values, renderer ) {

		for ( var i = 0, n = seq.length; i !== n; ++ i ) {

			var u = seq[ i ],
				v = values[ u.id ];

			if ( v.needsUpdate !== false ) {
				// note: always updating when .needsUpdate is undefined

				u.setValue( gl, v.value, renderer );

			}

		}

	};

	WebGLUniforms.seqWithValue = function( seq, values ) {

		var r = [];

		for ( var i = 0, n = seq.length; i !== n; ++ i ) {

			var u = seq[ i ];
			if ( u.id in values ) r.push( u );

		}

		return r;

	};

	/**
	 * Uniform Utilities
	 */

	var UniformsUtils = {

		merge: function ( uniforms ) {

			var merged = {};

			for ( var u = 0; u < uniforms.length; u ++ ) {

				var tmp = this.clone( uniforms[ u ] );

				for ( var p in tmp ) {

					merged[ p ] = tmp[ p ];

				}

			}

			return merged;

		},

		clone: function ( uniforms_src ) {

			var uniforms_dst = {};

			for ( var u in uniforms_src ) {

				uniforms_dst[ u ] = {};

				for ( var p in uniforms_src[ u ] ) {

					var parameter_src = uniforms_src[ u ][ p ];

					if ( parameter_src && ( parameter_src.isColor ||
						parameter_src.isMatrix3 || parameter_src.isMatrix4 ||
						parameter_src.isVector2 || parameter_src.isVector3 || parameter_src.isVector4 ||
						parameter_src.isTexture ) ) {

						uniforms_dst[ u ][ p ] = parameter_src.clone();

					} else if ( Array.isArray( parameter_src ) ) {

						uniforms_dst[ u ][ p ] = parameter_src.slice();

					} else {

						uniforms_dst[ u ][ p ] = parameter_src;

					}

				}

			}

			return uniforms_dst;

		}

	};

	var alphamap_fragment = "#ifdef USE_ALPHAMAP\n\tdiffuseColor.a *= texture2D( alphaMap, vUv ).g;\n#endif\n";

	var alphamap_pars_fragment = "#ifdef USE_ALPHAMAP\n\tuniform sampler2D alphaMap;\n#endif\n";

	var alphatest_fragment = "#ifdef ALPHATEST\n\tif ( diffuseColor.a < ALPHATEST ) discard;\n#endif\n";

	var aomap_fragment = "#ifdef USE_AOMAP\n\tfloat ambientOcclusion = ( texture2D( aoMap, vUv2 ).r - 1.0 ) * aoMapIntensity + 1.0;\n\treflectedLight.indirectDiffuse *= ambientOcclusion;\n\t#if defined( USE_ENVMAP ) && defined( PHYSICAL )\n\t\tfloat dotNV = saturate( dot( geometry.normal, geometry.viewDir ) );\n\t\treflectedLight.indirectSpecular *= computeSpecularOcclusion( dotNV, ambientOcclusion, material.specularRoughness );\n\t#endif\n#endif\n";

	var aomap_pars_fragment = "#ifdef USE_AOMAP\n\tuniform sampler2D aoMap;\n\tuniform float aoMapIntensity;\n#endif";

	var begin_vertex = "\nvec3 transformed = vec3( position );\n";

	var beginnormal_vertex = "\nvec3 objectNormal = vec3( normal );\n";

	var bsdfs = "float punctualLightIntensityToIrradianceFactor( const in float lightDistance, const in float cutoffDistance, const in float decayExponent ) {\n\t\tif( decayExponent > 0.0 ) {\n#if defined ( PHYSICALLY_CORRECT_LIGHTS )\n\t\t\tfloat distanceFalloff = 1.0 / max( pow( lightDistance, decayExponent ), 0.01 );\n\t\t\tfloat maxDistanceCutoffFactor = pow2( saturate( 1.0 - pow4( lightDistance / cutoffDistance ) ) );\n\t\t\treturn distanceFalloff * maxDistanceCutoffFactor;\n#else\n\t\t\treturn pow( saturate( -lightDistance / cutoffDistance + 1.0 ), decayExponent );\n#endif\n\t\t}\n\t\treturn 1.0;\n}\nvec3 BRDF_Diffuse_Lambert( const in vec3 diffuseColor ) {\n\treturn RECIPROCAL_PI * diffuseColor;\n}\nvec3 F_Schlick( const in vec3 specularColor, const in float dotLH ) {\n\tfloat fresnel = exp2( ( -5.55473 * dotLH - 6.98316 ) * dotLH );\n\treturn ( 1.0 - specularColor ) * fresnel + specularColor;\n}\nfloat G_GGX_Smith( const in float alpha, const in float dotNL, const in float dotNV ) {\n\tfloat a2 = pow2( alpha );\n\tfloat gl = dotNL + sqrt( a2 + ( 1.0 - a2 ) * pow2( dotNL ) );\n\tfloat gv = dotNV + sqrt( a2 + ( 1.0 - a2 ) * pow2( dotNV ) );\n\treturn 1.0 / ( gl * gv );\n}\nfloat G_GGX_SmithCorrelated( const in float alpha, const in float dotNL, const in float dotNV ) {\n\tfloat a2 = pow2( alpha );\n\tfloat gv = dotNL * sqrt( a2 + ( 1.0 - a2 ) * pow2( dotNV ) );\n\tfloat gl = dotNV * sqrt( a2 + ( 1.0 - a2 ) * pow2( dotNL ) );\n\treturn 0.5 / max( gv + gl, EPSILON );\n}\nfloat D_GGX( const in float alpha, const in float dotNH ) {\n\tfloat a2 = pow2( alpha );\n\tfloat denom = pow2( dotNH ) * ( a2 - 1.0 ) + 1.0;\n\treturn RECIPROCAL_PI * a2 / pow2( denom );\n}\nvec3 BRDF_Specular_GGX( const in IncidentLight incidentLight, const in GeometricContext geometry, const in vec3 specularColor, const in float roughness ) {\n\tfloat alpha = pow2( roughness );\n\tvec3 halfDir = normalize( incidentLight.direction + geometry.viewDir );\n\tfloat dotNL = saturate( dot( geometry.normal, incidentLight.direction ) );\n\tfloat dotNV = saturate( dot( geometry.normal, geometry.viewDir ) );\n\tfloat dotNH = saturate( dot( geometry.normal, halfDir ) );\n\tfloat dotLH = saturate( dot( incidentLight.direction, halfDir ) );\n\tvec3 F = F_Schlick( specularColor, dotLH );\n\tfloat G = G_GGX_SmithCorrelated( alpha, dotNL, dotNV );\n\tfloat D = D_GGX( alpha, dotNH );\n\treturn F * ( G * D );\n}\nvec2 ltcTextureCoords( const in GeometricContext geometry, const in float roughness ) {\n\tconst float LUT_SIZE  = 64.0;\n\tconst float LUT_SCALE = (LUT_SIZE - 1.0)/LUT_SIZE;\n\tconst float LUT_BIAS  = 0.5/LUT_SIZE;\n\tvec3 N = geometry.normal;\n\tvec3 V = geometry.viewDir;\n\tvec3 P = geometry.position;\n\tfloat theta = acos( dot( N, V ) );\n\tvec2 uv = vec2(\n\t\tsqrt( saturate( roughness ) ),\n\t\tsaturate( theta / ( 0.5 * PI ) ) );\n\tuv = uv * LUT_SCALE + LUT_BIAS;\n\treturn uv;\n}\nvoid clipQuadToHorizon( inout vec3 L[5], out int n ) {\n\tint config = 0;\n\tif ( L[0].z > 0.0 ) config += 1;\n\tif ( L[1].z > 0.0 ) config += 2;\n\tif ( L[2].z > 0.0 ) config += 4;\n\tif ( L[3].z > 0.0 ) config += 8;\n\tn = 0;\n\tif ( config == 0 ) {\n\t} else if ( config == 1 ) {\n\t\tn = 3;\n\t\tL[1] = -L[1].z * L[0] + L[0].z * L[1];\n\t\tL[2] = -L[3].z * L[0] + L[0].z * L[3];\n\t} else if ( config == 2 ) {\n\t\tn = 3;\n\t\tL[0] = -L[0].z * L[1] + L[1].z * L[0];\n\t\tL[2] = -L[2].z * L[1] + L[1].z * L[2];\n\t} else if ( config == 3 ) {\n\t\tn = 4;\n\t\tL[2] = -L[2].z * L[1] + L[1].z * L[2];\n\t\tL[3] = -L[3].z * L[0] + L[0].z * L[3];\n\t} else if ( config == 4 ) {\n\t\tn = 3;\n\t\tL[0] = -L[3].z * L[2] + L[2].z * L[3];\n\t\tL[1] = -L[1].z * L[2] + L[2].z * L[1];\n\t} else if ( config == 5 ) {\n\t\tn = 0;\n\t} else if ( config == 6 ) {\n\t\tn = 4;\n\t\tL[0] = -L[0].z * L[1] + L[1].z * L[0];\n\t\tL[3] = -L[3].z * L[2] + L[2].z * L[3];\n\t} else if ( config == 7 ) {\n\t\tn = 5;\n\t\tL[4] = -L[3].z * L[0] + L[0].z * L[3];\n\t\tL[3] = -L[3].z * L[2] + L[2].z * L[3];\n\t} else if ( config == 8 ) {\n\t\tn = 3;\n\t\tL[0] = -L[0].z * L[3] + L[3].z * L[0];\n\t\tL[1] = -L[2].z * L[3] + L[3].z * L[2];\n\t\tL[2] =  L[3];\n\t} else if ( config == 9 ) {\n\t\tn = 4;\n\t\tL[1] = -L[1].z * L[0] + L[0].z * L[1];\n\t\tL[2] = -L[2].z * L[3] + L[3].z * L[2];\n\t} else if ( config == 10 ) {\n\t\tn = 0;\n\t} else if ( config == 11 ) {\n\t\tn = 5;\n\t\tL[4] = L[3];\n\t\tL[3] = -L[2].z * L[3] + L[3].z * L[2];\n\t\tL[2] = -L[2].z * L[1] + L[1].z * L[2];\n\t} else if ( config == 12 ) {\n\t\tn = 4;\n\t\tL[1] = -L[1].z * L[2] + L[2].z * L[1];\n\t\tL[0] = -L[0].z * L[3] + L[3].z * L[0];\n\t} else if ( config == 13 ) {\n\t\tn = 5;\n\t\tL[4] = L[3];\n\t\tL[3] = L[2];\n\t\tL[2] = -L[1].z * L[2] + L[2].z * L[1];\n\t\tL[1] = -L[1].z * L[0] + L[0].z * L[1];\n\t} else if ( config == 14 ) {\n\t\tn = 5;\n\t\tL[4] = -L[0].z * L[3] + L[3].z * L[0];\n\t\tL[0] = -L[0].z * L[1] + L[1].z * L[0];\n\t} else if ( config == 15 ) {\n\t\tn = 4;\n\t}\n\tif ( n == 3 )\n\t\tL[3] = L[0];\n\tif ( n == 4 )\n\t\tL[4] = L[0];\n}\nfloat integrateLtcBrdfOverRectEdge( vec3 v1, vec3 v2 ) {\n\tfloat cosTheta = dot( v1, v2 );\n\tfloat theta = acos( cosTheta );\n\tfloat res = cross( v1, v2 ).z * ( ( theta > 0.001 ) ? theta / sin( theta ) : 1.0 );\n\treturn res;\n}\nvoid initRectPoints( const in vec3 pos, const in vec3 halfWidth, const in vec3 halfHeight, out vec3 rectPoints[4] ) {\n\trectPoints[0] = pos - halfWidth - halfHeight;\n\trectPoints[1] = pos + halfWidth - halfHeight;\n\trectPoints[2] = pos + halfWidth + halfHeight;\n\trectPoints[3] = pos - halfWidth + halfHeight;\n}\nvec3 integrateLtcBrdfOverRect( const in GeometricContext geometry, const in mat3 brdfMat, const in vec3 rectPoints[4] ) {\n\tvec3 N = geometry.normal;\n\tvec3 V = geometry.viewDir;\n\tvec3 P = geometry.position;\n\tvec3 T1, T2;\n\tT1 = normalize(V - N * dot( V, N ));\n\tT2 = - cross( N, T1 );\n\tmat3 brdfWrtSurface = brdfMat * transpose( mat3( T1, T2, N ) );\n\tvec3 clippedRect[5];\n\tclippedRect[0] = brdfWrtSurface * ( rectPoints[0] - P );\n\tclippedRect[1] = brdfWrtSurface * ( rectPoints[1] - P );\n\tclippedRect[2] = brdfWrtSurface * ( rectPoints[2] - P );\n\tclippedRect[3] = brdfWrtSurface * ( rectPoints[3] - P );\n\tint n;\n\tclipQuadToHorizon(clippedRect, n);\n\tif ( n == 0 )\n\t\treturn vec3( 0, 0, 0 );\n\tclippedRect[0] = normalize( clippedRect[0] );\n\tclippedRect[1] = normalize( clippedRect[1] );\n\tclippedRect[2] = normalize( clippedRect[2] );\n\tclippedRect[3] = normalize( clippedRect[3] );\n\tclippedRect[4] = normalize( clippedRect[4] );\n\tfloat sum = 0.0;\n\tsum += integrateLtcBrdfOverRectEdge( clippedRect[0], clippedRect[1] );\n\tsum += integrateLtcBrdfOverRectEdge( clippedRect[1], clippedRect[2] );\n\tsum += integrateLtcBrdfOverRectEdge( clippedRect[2], clippedRect[3] );\n\tif (n >= 4)\n\t\tsum += integrateLtcBrdfOverRectEdge( clippedRect[3], clippedRect[4] );\n\tif (n == 5)\n\t\tsum += integrateLtcBrdfOverRectEdge( clippedRect[4], clippedRect[0] );\n\tsum = max( 0.0, sum );\n\tvec3 Lo_i = vec3( sum, sum, sum );\n\treturn Lo_i;\n}\nvec3 Rect_Area_Light_Specular_Reflectance(\n\t\tconst in GeometricContext geometry,\n\t\tconst in vec3 lightPos, const in vec3 lightHalfWidth, const in vec3 lightHalfHeight,\n\t\tconst in float roughness,\n\t\tconst in sampler2D ltcMat, const in sampler2D ltcMag ) {\n\tvec3 rectPoints[4];\n\tinitRectPoints( lightPos, lightHalfWidth, lightHalfHeight, rectPoints );\n\tvec2 uv = ltcTextureCoords( geometry, roughness );\n\tvec4 brdfLtcApproxParams, t;\n\tbrdfLtcApproxParams = texture2D( ltcMat, uv );\n\tt = texture2D( ltcMat, uv );\n\tfloat brdfLtcScalar = texture2D( ltcMag, uv ).a;\n\tmat3 brdfLtcApproxMat = mat3(\n\t\tvec3(   1,   0, t.y ),\n\t\tvec3(   0, t.z,   0 ),\n\t\tvec3( t.w,   0, t.x )\n\t);\n\tvec3 specularReflectance = integrateLtcBrdfOverRect( geometry, brdfLtcApproxMat, rectPoints );\n\tspecularReflectance *= brdfLtcScalar;\n\treturn specularReflectance;\n}\nvec3 Rect_Area_Light_Diffuse_Reflectance(\n\t\tconst in GeometricContext geometry,\n\t\tconst in vec3 lightPos, const in vec3 lightHalfWidth, const in vec3 lightHalfHeight ) {\n\tvec3 rectPoints[4];\n\tinitRectPoints( lightPos, lightHalfWidth, lightHalfHeight, rectPoints );\n\tmat3 diffuseBrdfMat = mat3(1);\n\tvec3 diffuseReflectance = integrateLtcBrdfOverRect( geometry, diffuseBrdfMat, rectPoints );\n\treturn diffuseReflectance;\n}\nvec3 BRDF_Specular_GGX_Environment( const in GeometricContext geometry, const in vec3 specularColor, const in float roughness ) {\n\tfloat dotNV = saturate( dot( geometry.normal, geometry.viewDir ) );\n\tconst vec4 c0 = vec4( - 1, - 0.0275, - 0.572, 0.022 );\n\tconst vec4 c1 = vec4( 1, 0.0425, 1.04, - 0.04 );\n\tvec4 r = roughness * c0 + c1;\n\tfloat a004 = min( r.x * r.x, exp2( - 9.28 * dotNV ) ) * r.x + r.y;\n\tvec2 AB = vec2( -1.04, 1.04 ) * a004 + r.zw;\n\treturn specularColor * AB.x + AB.y;\n}\nfloat G_BlinnPhong_Implicit( ) {\n\treturn 0.25;\n}\nfloat D_BlinnPhong( const in float shininess, const in float dotNH ) {\n\treturn RECIPROCAL_PI * ( shininess * 0.5 + 1.0 ) * pow( dotNH, shininess );\n}\nvec3 BRDF_Specular_BlinnPhong( const in IncidentLight incidentLight, const in GeometricContext geometry, const in vec3 specularColor, const in float shininess ) {\n\tvec3 halfDir = normalize( incidentLight.direction + geometry.viewDir );\n\tfloat dotNH = saturate( dot( geometry.normal, halfDir ) );\n\tfloat dotLH = saturate( dot( incidentLight.direction, halfDir ) );\n\tvec3 F = F_Schlick( specularColor, dotLH );\n\tfloat G = G_BlinnPhong_Implicit( );\n\tfloat D = D_BlinnPhong( shininess, dotNH );\n\treturn F * ( G * D );\n}\nfloat GGXRoughnessToBlinnExponent( const in float ggxRoughness ) {\n\treturn ( 2.0 / pow2( ggxRoughness + 0.0001 ) - 2.0 );\n}\nfloat BlinnExponentToGGXRoughness( const in float blinnExponent ) {\n\treturn sqrt( 2.0 / ( blinnExponent + 2.0 ) );\n}\n";

	var bumpmap_pars_fragment = "#ifdef USE_BUMPMAP\n\tuniform sampler2D bumpMap;\n\tuniform float bumpScale;\n\tvec2 dHdxy_fwd() {\n\t\tvec2 dSTdx = dFdx( vUv );\n\t\tvec2 dSTdy = dFdy( vUv );\n\t\tfloat Hll = bumpScale * texture2D( bumpMap, vUv ).x;\n\t\tfloat dBx = bumpScale * texture2D( bumpMap, vUv + dSTdx ).x - Hll;\n\t\tfloat dBy = bumpScale * texture2D( bumpMap, vUv + dSTdy ).x - Hll;\n\t\treturn vec2( dBx, dBy );\n\t}\n\tvec3 perturbNormalArb( vec3 surf_pos, vec3 surf_norm, vec2 dHdxy ) {\n\t\tvec3 vSigmaX = dFdx( surf_pos );\n\t\tvec3 vSigmaY = dFdy( surf_pos );\n\t\tvec3 vN = surf_norm;\n\t\tvec3 R1 = cross( vSigmaY, vN );\n\t\tvec3 R2 = cross( vN, vSigmaX );\n\t\tfloat fDet = dot( vSigmaX, R1 );\n\t\tvec3 vGrad = sign( fDet ) * ( dHdxy.x * R1 + dHdxy.y * R2 );\n\t\treturn normalize( abs( fDet ) * surf_norm - vGrad );\n\t}\n#endif\n";

	var clipping_planes_fragment = "#if NUM_CLIPPING_PLANES > 0\n\tfor ( int i = 0; i < UNION_CLIPPING_PLANES; ++ i ) {\n\t\tvec4 plane = clippingPlanes[ i ];\n\t\tif ( dot( vViewPosition, plane.xyz ) > plane.w ) discard;\n\t}\n\t\t\n\t#if UNION_CLIPPING_PLANES < NUM_CLIPPING_PLANES\n\t\tbool clipped = true;\n\t\tfor ( int i = UNION_CLIPPING_PLANES; i < NUM_CLIPPING_PLANES; ++ i ) {\n\t\t\tvec4 plane = clippingPlanes[ i ];\n\t\t\tclipped = ( dot( vViewPosition, plane.xyz ) > plane.w ) && clipped;\n\t\t}\n\t\tif ( clipped ) discard;\n\t\n\t#endif\n#endif\n";

	var clipping_planes_pars_fragment = "#if NUM_CLIPPING_PLANES > 0\n\t#if ! defined( PHYSICAL ) && ! defined( PHONG )\n\t\tvarying vec3 vViewPosition;\n\t#endif\n\tuniform vec4 clippingPlanes[ NUM_CLIPPING_PLANES ];\n#endif\n";

	var clipping_planes_pars_vertex = "#if NUM_CLIPPING_PLANES > 0 && ! defined( PHYSICAL ) && ! defined( PHONG )\n\tvarying vec3 vViewPosition;\n#endif\n";

	var clipping_planes_vertex = "#if NUM_CLIPPING_PLANES > 0 && ! defined( PHYSICAL ) && ! defined( PHONG )\n\tvViewPosition = - mvPosition.xyz;\n#endif\n";

	var color_fragment = "#ifdef USE_COLOR\n\tdiffuseColor.rgb *= vColor;\n#endif";

	var color_pars_fragment = "#ifdef USE_COLOR\n\tvarying vec3 vColor;\n#endif\n";

	var color_pars_vertex = "#ifdef USE_COLOR\n\tvarying vec3 vColor;\n#endif";

	var color_vertex = "#ifdef USE_COLOR\n\tvColor.xyz = color.xyz;\n#endif";

	var common = "#define PI 3.14159265359\n#define PI2 6.28318530718\n#define PI_HALF 1.5707963267949\n#define RECIPROCAL_PI 0.31830988618\n#define RECIPROCAL_PI2 0.15915494\n#define LOG2 1.442695\n#define EPSILON 1e-6\n#define saturate(a) clamp( a, 0.0, 1.0 )\n#define whiteCompliment(a) ( 1.0 - saturate( a ) )\nfloat pow2( const in float x ) { return x*x; }\nfloat pow3( const in float x ) { return x*x*x; }\nfloat pow4( const in float x ) { float x2 = x*x; return x2*x2; }\nfloat average( const in vec3 color ) { return dot( color, vec3( 0.3333 ) ); }\nhighp float rand( const in vec2 uv ) {\n\tconst highp float a = 12.9898, b = 78.233, c = 43758.5453;\n\thighp float dt = dot( uv.xy, vec2( a,b ) ), sn = mod( dt, PI );\n\treturn fract(sin(sn) * c);\n}\nstruct IncidentLight {\n\tvec3 color;\n\tvec3 direction;\n\tbool visible;\n};\nstruct ReflectedLight {\n\tvec3 directDiffuse;\n\tvec3 directSpecular;\n\tvec3 indirectDiffuse;\n\tvec3 indirectSpecular;\n};\nstruct GeometricContext {\n\tvec3 position;\n\tvec3 normal;\n\tvec3 viewDir;\n};\nvec3 transformDirection( in vec3 dir, in mat4 matrix ) {\n\treturn normalize( ( matrix * vec4( dir, 0.0 ) ).xyz );\n}\nvec3 inverseTransformDirection( in vec3 dir, in mat4 matrix ) {\n\treturn normalize( ( vec4( dir, 0.0 ) * matrix ).xyz );\n}\nvec3 projectOnPlane(in vec3 point, in vec3 pointOnPlane, in vec3 planeNormal ) {\n\tfloat distance = dot( planeNormal, point - pointOnPlane );\n\treturn - distance * planeNormal + point;\n}\nfloat sideOfPlane( in vec3 point, in vec3 pointOnPlane, in vec3 planeNormal ) {\n\treturn sign( dot( point - pointOnPlane, planeNormal ) );\n}\nvec3 linePlaneIntersect( in vec3 pointOnLine, in vec3 lineDirection, in vec3 pointOnPlane, in vec3 planeNormal ) {\n\treturn lineDirection * ( dot( planeNormal, pointOnPlane - pointOnLine ) / dot( planeNormal, lineDirection ) ) + pointOnLine;\n}\nmat3 transpose( const in mat3 v ) {\n\tmat3 tmp;\n\ttmp[0] = vec3(v[0].x, v[1].x, v[2].x);\n\ttmp[1] = vec3(v[0].y, v[1].y, v[2].y);\n\ttmp[2] = vec3(v[0].z, v[1].z, v[2].z);\n\treturn tmp;\n}\n";

	var cube_uv_reflection_fragment = "#ifdef ENVMAP_TYPE_CUBE_UV\n#define cubeUV_textureSize (1024.0)\nint getFaceFromDirection(vec3 direction) {\n\tvec3 absDirection = abs(direction);\n\tint face = -1;\n\tif( absDirection.x > absDirection.z ) {\n\t\tif(absDirection.x > absDirection.y )\n\t\t\tface = direction.x > 0.0 ? 0 : 3;\n\t\telse\n\t\t\tface = direction.y > 0.0 ? 1 : 4;\n\t}\n\telse {\n\t\tif(absDirection.z > absDirection.y )\n\t\t\tface = direction.z > 0.0 ? 2 : 5;\n\t\telse\n\t\t\tface = direction.y > 0.0 ? 1 : 4;\n\t}\n\treturn face;\n}\n#define cubeUV_maxLods1  (log2(cubeUV_textureSize*0.25) - 1.0)\n#define cubeUV_rangeClamp (exp2((6.0 - 1.0) * 2.0))\nvec2 MipLevelInfo( vec3 vec, float roughnessLevel, float roughness ) {\n\tfloat scale = exp2(cubeUV_maxLods1 - roughnessLevel);\n\tfloat dxRoughness = dFdx(roughness);\n\tfloat dyRoughness = dFdy(roughness);\n\tvec3 dx = dFdx( vec * scale * dxRoughness );\n\tvec3 dy = dFdy( vec * scale * dyRoughness );\n\tfloat d = max( dot( dx, dx ), dot( dy, dy ) );\n\td = clamp(d, 1.0, cubeUV_rangeClamp);\n\tfloat mipLevel = 0.5 * log2(d);\n\treturn vec2(floor(mipLevel), fract(mipLevel));\n}\n#define cubeUV_maxLods2 (log2(cubeUV_textureSize*0.25) - 2.0)\n#define cubeUV_rcpTextureSize (1.0 / cubeUV_textureSize)\nvec2 getCubeUV(vec3 direction, float roughnessLevel, float mipLevel) {\n\tmipLevel = roughnessLevel > cubeUV_maxLods2 - 3.0 ? 0.0 : mipLevel;\n\tfloat a = 16.0 * cubeUV_rcpTextureSize;\n\tvec2 exp2_packed = exp2( vec2( roughnessLevel, mipLevel ) );\n\tvec2 rcp_exp2_packed = vec2( 1.0 ) / exp2_packed;\n\tfloat powScale = exp2_packed.x * exp2_packed.y;\n\tfloat scale = rcp_exp2_packed.x * rcp_exp2_packed.y * 0.25;\n\tfloat mipOffset = 0.75*(1.0 - rcp_exp2_packed.y) * rcp_exp2_packed.x;\n\tbool bRes = mipLevel == 0.0;\n\tscale =  bRes && (scale < a) ? a : scale;\n\tvec3 r;\n\tvec2 offset;\n\tint face = getFaceFromDirection(direction);\n\tfloat rcpPowScale = 1.0 / powScale;\n\tif( face == 0) {\n\t\tr = vec3(direction.x, -direction.z, direction.y);\n\t\toffset = vec2(0.0+mipOffset,0.75 * rcpPowScale);\n\t\toffset.y = bRes && (offset.y < 2.0*a) ?  a : offset.y;\n\t}\n\telse if( face == 1) {\n\t\tr = vec3(direction.y, direction.x, direction.z);\n\t\toffset = vec2(scale+mipOffset, 0.75 * rcpPowScale);\n\t\toffset.y = bRes && (offset.y < 2.0*a) ?  a : offset.y;\n\t}\n\telse if( face == 2) {\n\t\tr = vec3(direction.z, direction.x, direction.y);\n\t\toffset = vec2(2.0*scale+mipOffset, 0.75 * rcpPowScale);\n\t\toffset.y = bRes && (offset.y < 2.0*a) ?  a : offset.y;\n\t}\n\telse if( face == 3) {\n\t\tr = vec3(direction.x, direction.z, direction.y);\n\t\toffset = vec2(0.0+mipOffset,0.5 * rcpPowScale);\n\t\toffset.y = bRes && (offset.y < 2.0*a) ?  0.0 : offset.y;\n\t}\n\telse if( face == 4) {\n\t\tr = vec3(direction.y, direction.x, -direction.z);\n\t\toffset = vec2(scale+mipOffset, 0.5 * rcpPowScale);\n\t\toffset.y = bRes && (offset.y < 2.0*a) ?  0.0 : offset.y;\n\t}\n\telse {\n\t\tr = vec3(direction.z, -direction.x, direction.y);\n\t\toffset = vec2(2.0*scale+mipOffset, 0.5 * rcpPowScale);\n\t\toffset.y = bRes && (offset.y < 2.0*a) ?  0.0 : offset.y;\n\t}\n\tr = normalize(r);\n\tfloat texelOffset = 0.5 * cubeUV_rcpTextureSize;\n\tvec2 s = ( r.yz / abs( r.x ) + vec2( 1.0 ) ) * 0.5;\n\tvec2 base = offset + vec2( texelOffset );\n\treturn base + s * ( scale - 2.0 * texelOffset );\n}\n#define cubeUV_maxLods3 (log2(cubeUV_textureSize*0.25) - 3.0)\nvec4 textureCubeUV(vec3 reflectedDirection, float roughness ) {\n\tfloat roughnessVal = roughness* cubeUV_maxLods3;\n\tfloat r1 = floor(roughnessVal);\n\tfloat r2 = r1 + 1.0;\n\tfloat t = fract(roughnessVal);\n\tvec2 mipInfo = MipLevelInfo(reflectedDirection, r1, roughness);\n\tfloat s = mipInfo.y;\n\tfloat level0 = mipInfo.x;\n\tfloat level1 = level0 + 1.0;\n\tlevel1 = level1 > 5.0 ? 5.0 : level1;\n\tlevel0 += min( floor( s + 0.5 ), 5.0 );\n\tvec2 uv_10 = getCubeUV(reflectedDirection, r1, level0);\n\tvec4 color10 = envMapTexelToLinear(texture2D(envMap, uv_10));\n\tvec2 uv_20 = getCubeUV(reflectedDirection, r2, level0);\n\tvec4 color20 = envMapTexelToLinear(texture2D(envMap, uv_20));\n\tvec4 result = mix(color10, color20, t);\n\treturn vec4(result.rgb, 1.0);\n}\n#endif\n";

	var defaultnormal_vertex = "#ifdef FLIP_SIDED\n\tobjectNormal = -objectNormal;\n#endif\nvec3 transformedNormal = normalMatrix * objectNormal;\n";

	var displacementmap_pars_vertex = "#ifdef USE_DISPLACEMENTMAP\n\tuniform sampler2D displacementMap;\n\tuniform float displacementScale;\n\tuniform float displacementBias;\n#endif\n";

	var displacementmap_vertex = "#ifdef USE_DISPLACEMENTMAP\n\ttransformed += normal * ( texture2D( displacementMap, uv ).x * displacementScale + displacementBias );\n#endif\n";

	var emissivemap_fragment = "#ifdef USE_EMISSIVEMAP\n\tvec4 emissiveColor = texture2D( emissiveMap, vUv );\n\temissiveColor.rgb = emissiveMapTexelToLinear( emissiveColor ).rgb;\n\ttotalEmissiveRadiance *= emissiveColor.rgb;\n#endif\n";

	var emissivemap_pars_fragment = "#ifdef USE_EMISSIVEMAP\n\tuniform sampler2D emissiveMap;\n#endif\n";

	var encodings_fragment = "  gl_FragColor = linearToOutputTexel( gl_FragColor );\n";

	var encodings_pars_fragment = "\nvec4 LinearToLinear( in vec4 value ) {\n  return value;\n}\nvec4 GammaToLinear( in vec4 value, in float gammaFactor ) {\n  return vec4( pow( value.xyz, vec3( gammaFactor ) ), value.w );\n}\nvec4 LinearToGamma( in vec4 value, in float gammaFactor ) {\n  return vec4( pow( value.xyz, vec3( 1.0 / gammaFactor ) ), value.w );\n}\nvec4 sRGBToLinear( in vec4 value ) {\n  return vec4( mix( pow( value.rgb * 0.9478672986 + vec3( 0.0521327014 ), vec3( 2.4 ) ), value.rgb * 0.0773993808, vec3( lessThanEqual( value.rgb, vec3( 0.04045 ) ) ) ), value.w );\n}\nvec4 LinearTosRGB( in vec4 value ) {\n  return vec4( mix( pow( value.rgb, vec3( 0.41666 ) ) * 1.055 - vec3( 0.055 ), value.rgb * 12.92, vec3( lessThanEqual( value.rgb, vec3( 0.0031308 ) ) ) ), value.w );\n}\nvec4 RGBEToLinear( in vec4 value ) {\n  return vec4( value.rgb * exp2( value.a * 255.0 - 128.0 ), 1.0 );\n}\nvec4 LinearToRGBE( in vec4 value ) {\n  float maxComponent = max( max( value.r, value.g ), value.b );\n  float fExp = clamp( ceil( log2( maxComponent ) ), -128.0, 127.0 );\n  return vec4( value.rgb / exp2( fExp ), ( fExp + 128.0 ) / 255.0 );\n}\nvec4 RGBMToLinear( in vec4 value, in float maxRange ) {\n  return vec4( value.xyz * value.w * maxRange, 1.0 );\n}\nvec4 LinearToRGBM( in vec4 value, in float maxRange ) {\n  float maxRGB = max( value.x, max( value.g, value.b ) );\n  float M      = clamp( maxRGB / maxRange, 0.0, 1.0 );\n  M            = ceil( M * 255.0 ) / 255.0;\n  return vec4( value.rgb / ( M * maxRange ), M );\n}\nvec4 RGBDToLinear( in vec4 value, in float maxRange ) {\n    return vec4( value.rgb * ( ( maxRange / 255.0 ) / value.a ), 1.0 );\n}\nvec4 LinearToRGBD( in vec4 value, in float maxRange ) {\n    float maxRGB = max( value.x, max( value.g, value.b ) );\n    float D      = max( maxRange / maxRGB, 1.0 );\n    D            = min( floor( D ) / 255.0, 1.0 );\n    return vec4( value.rgb * ( D * ( 255.0 / maxRange ) ), D );\n}\nconst mat3 cLogLuvM = mat3( 0.2209, 0.3390, 0.4184, 0.1138, 0.6780, 0.7319, 0.0102, 0.1130, 0.2969 );\nvec4 LinearToLogLuv( in vec4 value )  {\n  vec3 Xp_Y_XYZp = value.rgb * cLogLuvM;\n  Xp_Y_XYZp = max(Xp_Y_XYZp, vec3(1e-6, 1e-6, 1e-6));\n  vec4 vResult;\n  vResult.xy = Xp_Y_XYZp.xy / Xp_Y_XYZp.z;\n  float Le = 2.0 * log2(Xp_Y_XYZp.y) + 127.0;\n  vResult.w = fract(Le);\n  vResult.z = (Le - (floor(vResult.w*255.0))/255.0)/255.0;\n  return vResult;\n}\nconst mat3 cLogLuvInverseM = mat3( 6.0014, -2.7008, -1.7996, -1.3320, 3.1029, -5.7721, 0.3008, -1.0882, 5.6268 );\nvec4 LogLuvToLinear( in vec4 value ) {\n  float Le = value.z * 255.0 + value.w;\n  vec3 Xp_Y_XYZp;\n  Xp_Y_XYZp.y = exp2((Le - 127.0) / 2.0);\n  Xp_Y_XYZp.z = Xp_Y_XYZp.y / value.y;\n  Xp_Y_XYZp.x = value.x * Xp_Y_XYZp.z;\n  vec3 vRGB = Xp_Y_XYZp.rgb * cLogLuvInverseM;\n  return vec4( max(vRGB, 0.0), 1.0 );\n}\n";

	var envmap_fragment = "#ifdef USE_ENVMAP\n\t#if defined( USE_BUMPMAP ) || defined( USE_NORMALMAP ) || defined( PHONG )\n\t\tvec3 cameraToVertex = normalize( vWorldPosition - cameraPosition );\n\t\tvec3 worldNormal = inverseTransformDirection( normal, viewMatrix );\n\t\t#ifdef ENVMAP_MODE_REFLECTION\n\t\t\tvec3 reflectVec = reflect( cameraToVertex, worldNormal );\n\t\t#else\n\t\t\tvec3 reflectVec = refract( cameraToVertex, worldNormal, refractionRatio );\n\t\t#endif\n\t#else\n\t\tvec3 reflectVec = vReflect;\n\t#endif\n\t#ifdef ENVMAP_TYPE_CUBE\n\t\tvec4 envColor = textureCube( envMap, flipNormal * vec3( flipEnvMap * reflectVec.x, reflectVec.yz ) );\n\t#elif defined( ENVMAP_TYPE_EQUIREC )\n\t\tvec2 sampleUV;\n\t\tsampleUV.y = saturate( flipNormal * reflectVec.y * 0.5 + 0.5 );\n\t\tsampleUV.x = atan( flipNormal * reflectVec.z, flipNormal * reflectVec.x ) * RECIPROCAL_PI2 + 0.5;\n\t\tvec4 envColor = texture2D( envMap, sampleUV );\n\t#elif defined( ENVMAP_TYPE_SPHERE )\n\t\tvec3 reflectView = flipNormal * normalize( ( viewMatrix * vec4( reflectVec, 0.0 ) ).xyz + vec3( 0.0, 0.0, 1.0 ) );\n\t\tvec4 envColor = texture2D( envMap, reflectView.xy * 0.5 + 0.5 );\n\t#else\n\t\tvec4 envColor = vec4( 0.0 );\n\t#endif\n\tenvColor = envMapTexelToLinear( envColor );\n\t#ifdef ENVMAP_BLENDING_MULTIPLY\n\t\toutgoingLight = mix( outgoingLight, outgoingLight * envColor.xyz, specularStrength * reflectivity );\n\t#elif defined( ENVMAP_BLENDING_MIX )\n\t\toutgoingLight = mix( outgoingLight, envColor.xyz, specularStrength * reflectivity );\n\t#elif defined( ENVMAP_BLENDING_ADD )\n\t\toutgoingLight += envColor.xyz * specularStrength * reflectivity;\n\t#endif\n#endif\n";

	var envmap_pars_fragment = "#if defined( USE_ENVMAP ) || defined( PHYSICAL )\n\tuniform float reflectivity;\n\tuniform float envMapIntensity;\n#endif\n#ifdef USE_ENVMAP\n\t#if ! defined( PHYSICAL ) && ( defined( USE_BUMPMAP ) || defined( USE_NORMALMAP ) || defined( PHONG ) )\n\t\tvarying vec3 vWorldPosition;\n\t#endif\n\t#ifdef ENVMAP_TYPE_CUBE\n\t\tuniform samplerCube envMap;\n\t#else\n\t\tuniform sampler2D envMap;\n\t#endif\n\tuniform float flipEnvMap;\n\t#if defined( USE_BUMPMAP ) || defined( USE_NORMALMAP ) || defined( PHONG ) || defined( PHYSICAL )\n\t\tuniform float refractionRatio;\n\t#else\n\t\tvarying vec3 vReflect;\n\t#endif\n#endif\n";

	var envmap_pars_vertex = "#ifdef USE_ENVMAP\n\t#if defined( USE_BUMPMAP ) || defined( USE_NORMALMAP ) || defined( PHONG )\n\t\tvarying vec3 vWorldPosition;\n\t#else\n\t\tvarying vec3 vReflect;\n\t\tuniform float refractionRatio;\n\t#endif\n#endif\n";

	var envmap_vertex = "#ifdef USE_ENVMAP\n\t#if defined( USE_BUMPMAP ) || defined( USE_NORMALMAP ) || defined( PHONG )\n\t\tvWorldPosition = worldPosition.xyz;\n\t#else\n\t\tvec3 cameraToVertex = normalize( worldPosition.xyz - cameraPosition );\n\t\tvec3 worldNormal = inverseTransformDirection( transformedNormal, viewMatrix );\n\t\t#ifdef ENVMAP_MODE_REFLECTION\n\t\t\tvReflect = reflect( cameraToVertex, worldNormal );\n\t\t#else\n\t\t\tvReflect = refract( cameraToVertex, worldNormal, refractionRatio );\n\t\t#endif\n\t#endif\n#endif\n";

	var fog_fragment = "#ifdef USE_FOG\n\t#ifdef USE_LOGDEPTHBUF_EXT\n\t\tfloat depth = gl_FragDepthEXT / gl_FragCoord.w;\n\t#else\n\t\tfloat depth = gl_FragCoord.z / gl_FragCoord.w;\n\t#endif\n\t#ifdef FOG_EXP2\n\t\tfloat fogFactor = whiteCompliment( exp2( - fogDensity * fogDensity * depth * depth * LOG2 ) );\n\t#else\n\t\tfloat fogFactor = smoothstep( fogNear, fogFar, depth );\n\t#endif\n\tgl_FragColor.rgb = mix( gl_FragColor.rgb, fogColor, fogFactor );\n#endif\n";

	var fog_pars_fragment = "#ifdef USE_FOG\n\tuniform vec3 fogColor;\n\t#ifdef FOG_EXP2\n\t\tuniform float fogDensity;\n\t#else\n\t\tuniform float fogNear;\n\t\tuniform float fogFar;\n\t#endif\n#endif";

	var gradientmap_pars_fragment = "#ifdef TOON\n\tuniform sampler2D gradientMap;\n\tvec3 getGradientIrradiance( vec3 normal, vec3 lightDirection ) {\n\t\tfloat dotNL = dot( normal, lightDirection );\n\t\tvec2 coord = vec2( dotNL * 0.5 + 0.5, 0.0 );\n\t\t#ifdef USE_GRADIENTMAP\n\t\t\treturn texture2D( gradientMap, coord ).rgb;\n\t\t#else\n\t\t\treturn ( coord.x < 0.7 ) ? vec3( 0.7 ) : vec3( 1.0 );\n\t\t#endif\n\t}\n#endif\n";

	var lightmap_fragment = "#ifdef USE_LIGHTMAP\n\treflectedLight.indirectDiffuse += PI * texture2D( lightMap, vUv2 ).xyz * lightMapIntensity;\n#endif\n";

	var lightmap_pars_fragment = "#ifdef USE_LIGHTMAP\n\tuniform sampler2D lightMap;\n\tuniform float lightMapIntensity;\n#endif";

	var lights_lambert_vertex = "vec3 diffuse = vec3( 1.0 );\nGeometricContext geometry;\ngeometry.position = mvPosition.xyz;\ngeometry.normal = normalize( transformedNormal );\ngeometry.viewDir = normalize( -mvPosition.xyz );\nGeometricContext backGeometry;\nbackGeometry.position = geometry.position;\nbackGeometry.normal = -geometry.normal;\nbackGeometry.viewDir = geometry.viewDir;\nvLightFront = vec3( 0.0 );\n#ifdef DOUBLE_SIDED\n\tvLightBack = vec3( 0.0 );\n#endif\nIncidentLight directLight;\nfloat dotNL;\nvec3 directLightColor_Diffuse;\n#if NUM_POINT_LIGHTS > 0\n\tfor ( int i = 0; i < NUM_POINT_LIGHTS; i ++ ) {\n\t\tgetPointDirectLightIrradiance( pointLights[ i ], geometry, directLight );\n\t\tdotNL = dot( geometry.normal, directLight.direction );\n\t\tdirectLightColor_Diffuse = PI * directLight.color;\n\t\tvLightFront += saturate( dotNL ) * directLightColor_Diffuse;\n\t\t#ifdef DOUBLE_SIDED\n\t\t\tvLightBack += saturate( -dotNL ) * directLightColor_Diffuse;\n\t\t#endif\n\t}\n#endif\n#if NUM_SPOT_LIGHTS > 0\n\tfor ( int i = 0; i < NUM_SPOT_LIGHTS; i ++ ) {\n\t\tgetSpotDirectLightIrradiance( spotLights[ i ], geometry, directLight );\n\t\tdotNL = dot( geometry.normal, directLight.direction );\n\t\tdirectLightColor_Diffuse = PI * directLight.color;\n\t\tvLightFront += saturate( dotNL ) * directLightColor_Diffuse;\n\t\t#ifdef DOUBLE_SIDED\n\t\t\tvLightBack += saturate( -dotNL ) * directLightColor_Diffuse;\n\t\t#endif\n\t}\n#endif\n#if NUM_RECT_AREA_LIGHTS > 0\n\tfor ( int i = 0; i < NUM_RECT_AREA_LIGHTS; i ++ ) {\n\t}\n#endif\n#if NUM_DIR_LIGHTS > 0\n\tfor ( int i = 0; i < NUM_DIR_LIGHTS; i ++ ) {\n\t\tgetDirectionalDirectLightIrradiance( directionalLights[ i ], geometry, directLight );\n\t\tdotNL = dot( geometry.normal, directLight.direction );\n\t\tdirectLightColor_Diffuse = PI * directLight.color;\n\t\tvLightFront += saturate( dotNL ) * directLightColor_Diffuse;\n\t\t#ifdef DOUBLE_SIDED\n\t\t\tvLightBack += saturate( -dotNL ) * directLightColor_Diffuse;\n\t\t#endif\n\t}\n#endif\n#if NUM_HEMI_LIGHTS > 0\n\tfor ( int i = 0; i < NUM_HEMI_LIGHTS; i ++ ) {\n\t\tvLightFront += getHemisphereLightIrradiance( hemisphereLights[ i ], geometry );\n\t\t#ifdef DOUBLE_SIDED\n\t\t\tvLightBack += getHemisphereLightIrradiance( hemisphereLights[ i ], backGeometry );\n\t\t#endif\n\t}\n#endif\n";

	var lights_pars = "uniform vec3 ambientLightColor;\nvec3 getAmbientLightIrradiance( const in vec3 ambientLightColor ) {\n\tvec3 irradiance = ambientLightColor;\n\t#ifndef PHYSICALLY_CORRECT_LIGHTS\n\t\tirradiance *= PI;\n\t#endif\n\treturn irradiance;\n}\n#if NUM_DIR_LIGHTS > 0\n\tstruct DirectionalLight {\n\t\tvec3 direction;\n\t\tvec3 color;\n\t\tint shadow;\n\t\tfloat shadowBias;\n\t\tfloat shadowRadius;\n\t\tvec2 shadowMapSize;\n\t};\n\tuniform DirectionalLight directionalLights[ NUM_DIR_LIGHTS ];\n\tvoid getDirectionalDirectLightIrradiance( const in DirectionalLight directionalLight, const in GeometricContext geometry, out IncidentLight directLight ) {\n\t\tdirectLight.color = directionalLight.color;\n\t\tdirectLight.direction = directionalLight.direction;\n\t\tdirectLight.visible = true;\n\t}\n#endif\n#if NUM_POINT_LIGHTS > 0\n\tstruct PointLight {\n\t\tvec3 position;\n\t\tvec3 color;\n\t\tfloat distance;\n\t\tfloat decay;\n\t\tint shadow;\n\t\tfloat shadowBias;\n\t\tfloat shadowRadius;\n\t\tvec2 shadowMapSize;\n\t};\n\tuniform PointLight pointLights[ NUM_POINT_LIGHTS ];\n\tvoid getPointDirectLightIrradiance( const in PointLight pointLight, const in GeometricContext geometry, out IncidentLight directLight ) {\n\t\tvec3 lVector = pointLight.position - geometry.position;\n\t\tdirectLight.direction = normalize( lVector );\n\t\tfloat lightDistance = length( lVector );\n\t\tdirectLight.color = pointLight.color;\n\t\tdirectLight.color *= punctualLightIntensityToIrradianceFactor( lightDistance, pointLight.distance, pointLight.decay );\n\t\tdirectLight.visible = ( directLight.color != vec3( 0.0 ) );\n\t}\n#endif\n#if NUM_SPOT_LIGHTS > 0\n\tstruct SpotLight {\n\t\tvec3 position;\n\t\tvec3 direction;\n\t\tvec3 color;\n\t\tfloat distance;\n\t\tfloat decay;\n\t\tfloat coneCos;\n\t\tfloat penumbraCos;\n\t\tint shadow;\n\t\tfloat shadowBias;\n\t\tfloat shadowRadius;\n\t\tvec2 shadowMapSize;\n\t};\n\tuniform SpotLight spotLights[ NUM_SPOT_LIGHTS ];\n\tvoid getSpotDirectLightIrradiance( const in SpotLight spotLight, const in GeometricContext geometry, out IncidentLight directLight  ) {\n\t\tvec3 lVector = spotLight.position - geometry.position;\n\t\tdirectLight.direction = normalize( lVector );\n\t\tfloat lightDistance = length( lVector );\n\t\tfloat angleCos = dot( directLight.direction, spotLight.direction );\n\t\tif ( angleCos > spotLight.coneCos ) {\n\t\t\tfloat spotEffect = smoothstep( spotLight.coneCos, spotLight.penumbraCos, angleCos );\n\t\t\tdirectLight.color = spotLight.color;\n\t\t\tdirectLight.color *= spotEffect * punctualLightIntensityToIrradianceFactor( lightDistance, spotLight.distance, spotLight.decay );\n\t\t\tdirectLight.visible = true;\n\t\t} else {\n\t\t\tdirectLight.color = vec3( 0.0 );\n\t\t\tdirectLight.visible = false;\n\t\t}\n\t}\n#endif\n#if NUM_RECT_AREA_LIGHTS > 0\n\tstruct RectAreaLight {\n\t\tvec3 color;\n\t\tvec3 position;\n\t\tvec3 halfWidth;\n\t\tvec3 halfHeight;\n\t};\n\tuniform sampler2D ltcMat;\tuniform sampler2D ltcMag;\n\tuniform RectAreaLight rectAreaLights[ NUM_RECT_AREA_LIGHTS ];\n#endif\n#if NUM_HEMI_LIGHTS > 0\n\tstruct HemisphereLight {\n\t\tvec3 direction;\n\t\tvec3 skyColor;\n\t\tvec3 groundColor;\n\t};\n\tuniform HemisphereLight hemisphereLights[ NUM_HEMI_LIGHTS ];\n\tvec3 getHemisphereLightIrradiance( const in HemisphereLight hemiLight, const in GeometricContext geometry ) {\n\t\tfloat dotNL = dot( geometry.normal, hemiLight.direction );\n\t\tfloat hemiDiffuseWeight = 0.5 * dotNL + 0.5;\n\t\tvec3 irradiance = mix( hemiLight.groundColor, hemiLight.skyColor, hemiDiffuseWeight );\n\t\t#ifndef PHYSICALLY_CORRECT_LIGHTS\n\t\t\tirradiance *= PI;\n\t\t#endif\n\t\treturn irradiance;\n\t}\n#endif\n#if defined( USE_ENVMAP ) && defined( PHYSICAL )\n\tvec3 getLightProbeIndirectIrradiance( const in GeometricContext geometry, const in int maxMIPLevel ) {\n\t\tvec3 worldNormal = inverseTransformDirection( geometry.normal, viewMatrix );\n\t\t#ifdef ENVMAP_TYPE_CUBE\n\t\t\tvec3 queryVec = vec3( flipEnvMap * worldNormal.x, worldNormal.yz );\n\t\t\t#ifdef TEXTURE_LOD_EXT\n\t\t\t\tvec4 envMapColor = textureCubeLodEXT( envMap, queryVec, float( maxMIPLevel ) );\n\t\t\t#else\n\t\t\t\tvec4 envMapColor = textureCube( envMap, queryVec, float( maxMIPLevel ) );\n\t\t\t#endif\n\t\t\tenvMapColor.rgb = envMapTexelToLinear( envMapColor ).rgb;\n\t\t#elif defined( ENVMAP_TYPE_CUBE_UV )\n\t\t\tvec3 queryVec = vec3( flipEnvMap * worldNormal.x, worldNormal.yz );\n\t\t\tvec4 envMapColor = textureCubeUV( queryVec, 1.0 );\n\t\t#else\n\t\t\tvec4 envMapColor = vec4( 0.0 );\n\t\t#endif\n\t\treturn PI * envMapColor.rgb * envMapIntensity;\n\t}\n\tfloat getSpecularMIPLevel( const in float blinnShininessExponent, const in int maxMIPLevel ) {\n\t\tfloat maxMIPLevelScalar = float( maxMIPLevel );\n\t\tfloat desiredMIPLevel = maxMIPLevelScalar - 0.79248 - 0.5 * log2( pow2( blinnShininessExponent ) + 1.0 );\n\t\treturn clamp( desiredMIPLevel, 0.0, maxMIPLevelScalar );\n\t}\n\tvec3 getLightProbeIndirectRadiance( const in GeometricContext geometry, const in float blinnShininessExponent, const in int maxMIPLevel ) {\n\t\t#ifdef ENVMAP_MODE_REFLECTION\n\t\t\tvec3 reflectVec = reflect( -geometry.viewDir, geometry.normal );\n\t\t#else\n\t\t\tvec3 reflectVec = refract( -geometry.viewDir, geometry.normal, refractionRatio );\n\t\t#endif\n\t\treflectVec = inverseTransformDirection( reflectVec, viewMatrix );\n\t\tfloat specularMIPLevel = getSpecularMIPLevel( blinnShininessExponent, maxMIPLevel );\n\t\t#ifdef ENVMAP_TYPE_CUBE\n\t\t\tvec3 queryReflectVec = vec3( flipEnvMap * reflectVec.x, reflectVec.yz );\n\t\t\t#ifdef TEXTURE_LOD_EXT\n\t\t\t\tvec4 envMapColor = textureCubeLodEXT( envMap, queryReflectVec, specularMIPLevel );\n\t\t\t#else\n\t\t\t\tvec4 envMapColor = textureCube( envMap, queryReflectVec, specularMIPLevel );\n\t\t\t#endif\n\t\t\tenvMapColor.rgb = envMapTexelToLinear( envMapColor ).rgb;\n\t\t#elif defined( ENVMAP_TYPE_CUBE_UV )\n\t\t\tvec3 queryReflectVec = vec3( flipEnvMap * reflectVec.x, reflectVec.yz );\n\t\t\tvec4 envMapColor = textureCubeUV(queryReflectVec, BlinnExponentToGGXRoughness(blinnShininessExponent));\n\t\t#elif defined( ENVMAP_TYPE_EQUIREC )\n\t\t\tvec2 sampleUV;\n\t\t\tsampleUV.y = saturate( reflectVec.y * 0.5 + 0.5 );\n\t\t\tsampleUV.x = atan( reflectVec.z, reflectVec.x ) * RECIPROCAL_PI2 + 0.5;\n\t\t\t#ifdef TEXTURE_LOD_EXT\n\t\t\t\tvec4 envMapColor = texture2DLodEXT( envMap, sampleUV, specularMIPLevel );\n\t\t\t#else\n\t\t\t\tvec4 envMapColor = texture2D( envMap, sampleUV, specularMIPLevel );\n\t\t\t#endif\n\t\t\tenvMapColor.rgb = envMapTexelToLinear( envMapColor ).rgb;\n\t\t#elif defined( ENVMAP_TYPE_SPHERE )\n\t\t\tvec3 reflectView = normalize( ( viewMatrix * vec4( reflectVec, 0.0 ) ).xyz + vec3( 0.0,0.0,1.0 ) );\n\t\t\t#ifdef TEXTURE_LOD_EXT\n\t\t\t\tvec4 envMapColor = texture2DLodEXT( envMap, reflectView.xy * 0.5 + 0.5, specularMIPLevel );\n\t\t\t#else\n\t\t\t\tvec4 envMapColor = texture2D( envMap, reflectView.xy * 0.5 + 0.5, specularMIPLevel );\n\t\t\t#endif\n\t\t\tenvMapColor.rgb = envMapTexelToLinear( envMapColor ).rgb;\n\t\t#endif\n\t\treturn envMapColor.rgb * envMapIntensity;\n\t}\n#endif\n";

	var lights_phong_fragment = "BlinnPhongMaterial material;\nmaterial.diffuseColor = diffuseColor.rgb;\nmaterial.specularColor = specular;\nmaterial.specularShininess = shininess;\nmaterial.specularStrength = specularStrength;\n";

	var lights_phong_pars_fragment = "varying vec3 vViewPosition;\n#ifndef FLAT_SHADED\n\tvarying vec3 vNormal;\n#endif\nstruct BlinnPhongMaterial {\n\tvec3\tdiffuseColor;\n\tvec3\tspecularColor;\n\tfloat\tspecularShininess;\n\tfloat\tspecularStrength;\n};\n#if NUM_RECT_AREA_LIGHTS > 0\n    void RE_Direct_RectArea_BlinnPhong( const in RectAreaLight rectAreaLight, const in GeometricContext geometry, const in BlinnPhongMaterial material, inout ReflectedLight reflectedLight ) {\n        vec3 matDiffColor = material.diffuseColor;\n        vec3 matSpecColor = material.specularColor;\n        vec3 lightColor   = rectAreaLight.color;\n        float roughness = BlinnExponentToGGXRoughness( material.specularShininess );\n        vec3 spec = Rect_Area_Light_Specular_Reflectance(\n                geometry,\n                rectAreaLight.position, rectAreaLight.halfWidth, rectAreaLight.halfHeight,\n                roughness,\n                ltcMat, ltcMag );\n        vec3 diff = Rect_Area_Light_Diffuse_Reflectance(\n                geometry,\n                rectAreaLight.position, rectAreaLight.halfWidth, rectAreaLight.halfHeight );\n        reflectedLight.directSpecular += lightColor * matSpecColor * spec / PI2;\n        reflectedLight.directDiffuse  += lightColor * matDiffColor * diff / PI2;\n    }\n#endif\nvoid RE_Direct_BlinnPhong( const in IncidentLight directLight, const in GeometricContext geometry, const in BlinnPhongMaterial material, inout ReflectedLight reflectedLight ) {\n\t#ifdef TOON\n\t\tvec3 irradiance = getGradientIrradiance( geometry.normal, directLight.direction ) * directLight.color;\n\t#else\n\t\tfloat dotNL = saturate( dot( geometry.normal, directLight.direction ) );\n\t\tvec3 irradiance = dotNL * directLight.color;\n\t#endif\n\t#ifndef PHYSICALLY_CORRECT_LIGHTS\n\t\tirradiance *= PI;\n\t#endif\n\treflectedLight.directDiffuse += irradiance * BRDF_Diffuse_Lambert( material.diffuseColor );\n\treflectedLight.directSpecular += irradiance * BRDF_Specular_BlinnPhong( directLight, geometry, material.specularColor, material.specularShininess ) * material.specularStrength;\n}\nvoid RE_IndirectDiffuse_BlinnPhong( const in vec3 irradiance, const in GeometricContext geometry, const in BlinnPhongMaterial material, inout ReflectedLight reflectedLight ) {\n\treflectedLight.indirectDiffuse += irradiance * BRDF_Diffuse_Lambert( material.diffuseColor );\n}\n#define RE_Direct\t\t\t\tRE_Direct_BlinnPhong\n#define RE_Direct_RectArea\t\tRE_Direct_RectArea_BlinnPhong\n#define RE_IndirectDiffuse\t\tRE_IndirectDiffuse_BlinnPhong\n#define Material_LightProbeLOD( material )\t(0)\n";

	var lights_physical_fragment = "PhysicalMaterial material;\nmaterial.diffuseColor = diffuseColor.rgb * ( 1.0 - metalnessFactor );\nmaterial.specularRoughness = clamp( roughnessFactor, 0.04, 1.0 );\n#ifdef STANDARD\n\tmaterial.specularColor = mix( vec3( DEFAULT_SPECULAR_COEFFICIENT ), diffuseColor.rgb, metalnessFactor );\n#else\n\tmaterial.specularColor = mix( vec3( MAXIMUM_SPECULAR_COEFFICIENT * pow2( reflectivity ) ), diffuseColor.rgb, metalnessFactor );\n\tmaterial.clearCoat = saturate( clearCoat );\tmaterial.clearCoatRoughness = clamp( clearCoatRoughness, 0.04, 1.0 );\n#endif\n";

	var lights_physical_pars_fragment = "struct PhysicalMaterial {\n\tvec3\tdiffuseColor;\n\tfloat\tspecularRoughness;\n\tvec3\tspecularColor;\n\t#ifndef STANDARD\n\t\tfloat clearCoat;\n\t\tfloat clearCoatRoughness;\n\t#endif\n};\n#define MAXIMUM_SPECULAR_COEFFICIENT 0.16\n#define DEFAULT_SPECULAR_COEFFICIENT 0.04\nfloat clearCoatDHRApprox( const in float roughness, const in float dotNL ) {\n\treturn DEFAULT_SPECULAR_COEFFICIENT + ( 1.0 - DEFAULT_SPECULAR_COEFFICIENT ) * ( pow( 1.0 - dotNL, 5.0 ) * pow( 1.0 - roughness, 2.0 ) );\n}\n#if NUM_RECT_AREA_LIGHTS > 0\n    void RE_Direct_RectArea_Physical( const in RectAreaLight rectAreaLight, const in GeometricContext geometry, const in PhysicalMaterial material, inout ReflectedLight reflectedLight ) {\n        vec3 matDiffColor = material.diffuseColor;\n        vec3 matSpecColor = material.specularColor;\n        vec3 lightColor   = rectAreaLight.color;\n        float roughness = material.specularRoughness;\n        vec3 spec = Rect_Area_Light_Specular_Reflectance(\n                geometry,\n                rectAreaLight.position, rectAreaLight.halfWidth, rectAreaLight.halfHeight,\n                roughness,\n                ltcMat, ltcMag );\n        vec3 diff = Rect_Area_Light_Diffuse_Reflectance(\n                geometry,\n                rectAreaLight.position, rectAreaLight.halfWidth, rectAreaLight.halfHeight );\n        reflectedLight.directSpecular += lightColor * matSpecColor * spec;\n        reflectedLight.directDiffuse  += lightColor * matDiffColor * diff;\n    }\n#endif\nvoid RE_Direct_Physical( const in IncidentLight directLight, const in GeometricContext geometry, const in PhysicalMaterial material, inout ReflectedLight reflectedLight ) {\n\tfloat dotNL = saturate( dot( geometry.normal, directLight.direction ) );\n\tvec3 irradiance = dotNL * directLight.color;\n\t#ifndef PHYSICALLY_CORRECT_LIGHTS\n\t\tirradiance *= PI;\n\t#endif\n\t#ifndef STANDARD\n\t\tfloat clearCoatDHR = material.clearCoat * clearCoatDHRApprox( material.clearCoatRoughness, dotNL );\n\t#else\n\t\tfloat clearCoatDHR = 0.0;\n\t#endif\n\treflectedLight.directSpecular += ( 1.0 - clearCoatDHR ) * irradiance * BRDF_Specular_GGX( directLight, geometry, material.specularColor, material.specularRoughness );\n\treflectedLight.directDiffuse += ( 1.0 - clearCoatDHR ) * irradiance * BRDF_Diffuse_Lambert( material.diffuseColor );\n\t#ifndef STANDARD\n\t\treflectedLight.directSpecular += irradiance * material.clearCoat * BRDF_Specular_GGX( directLight, geometry, vec3( DEFAULT_SPECULAR_COEFFICIENT ), material.clearCoatRoughness );\n\t#endif\n}\nvoid RE_IndirectDiffuse_Physical( const in vec3 irradiance, const in GeometricContext geometry, const in PhysicalMaterial material, inout ReflectedLight reflectedLight ) {\n\treflectedLight.indirectDiffuse += irradiance * BRDF_Diffuse_Lambert( material.diffuseColor );\n}\nvoid RE_IndirectSpecular_Physical( const in vec3 radiance, const in vec3 clearCoatRadiance, const in GeometricContext geometry, const in PhysicalMaterial material, inout ReflectedLight reflectedLight ) {\n\t#ifndef STANDARD\n\t\tfloat dotNV = saturate( dot( geometry.normal, geometry.viewDir ) );\n\t\tfloat dotNL = dotNV;\n\t\tfloat clearCoatDHR = material.clearCoat * clearCoatDHRApprox( material.clearCoatRoughness, dotNL );\n\t#else\n\t\tfloat clearCoatDHR = 0.0;\n\t#endif\n\treflectedLight.indirectSpecular += ( 1.0 - clearCoatDHR ) * radiance * BRDF_Specular_GGX_Environment( geometry, material.specularColor, material.specularRoughness );\n\t#ifndef STANDARD\n\t\treflectedLight.indirectSpecular += clearCoatRadiance * material.clearCoat * BRDF_Specular_GGX_Environment( geometry, vec3( DEFAULT_SPECULAR_COEFFICIENT ), material.clearCoatRoughness );\n\t#endif\n}\n#define RE_Direct\t\t\t\tRE_Direct_Physical\n#define RE_Direct_RectArea\t\tRE_Direct_RectArea_Physical\n#define RE_IndirectDiffuse\t\tRE_IndirectDiffuse_Physical\n#define RE_IndirectSpecular\t\tRE_IndirectSpecular_Physical\n#define Material_BlinnShininessExponent( material )   GGXRoughnessToBlinnExponent( material.specularRoughness )\n#define Material_ClearCoat_BlinnShininessExponent( material )   GGXRoughnessToBlinnExponent( material.clearCoatRoughness )\nfloat computeSpecularOcclusion( const in float dotNV, const in float ambientOcclusion, const in float roughness ) {\n\treturn saturate( pow( dotNV + ambientOcclusion, exp2( - 16.0 * roughness - 1.0 ) ) - 1.0 + ambientOcclusion );\n}\n";

	var lights_template = "\nGeometricContext geometry;\ngeometry.position = - vViewPosition;\ngeometry.normal = normal;\ngeometry.viewDir = normalize( vViewPosition );\nIncidentLight directLight;\n#if ( NUM_POINT_LIGHTS > 0 ) && defined( RE_Direct )\n\tPointLight pointLight;\n\tfor ( int i = 0; i < NUM_POINT_LIGHTS; i ++ ) {\n\t\tpointLight = pointLights[ i ];\n\t\tgetPointDirectLightIrradiance( pointLight, geometry, directLight );\n\t\t#ifdef USE_SHADOWMAP\n\t\tdirectLight.color *= all( bvec2( pointLight.shadow, directLight.visible ) ) ? getPointShadow( pointShadowMap[ i ], pointLight.shadowMapSize, pointLight.shadowBias, pointLight.shadowRadius, vPointShadowCoord[ i ] ) : 1.0;\n\t\t#endif\n\t\tRE_Direct( directLight, geometry, material, reflectedLight );\n\t}\n#endif\n#if ( NUM_SPOT_LIGHTS > 0 ) && defined( RE_Direct )\n\tSpotLight spotLight;\n\tfor ( int i = 0; i < NUM_SPOT_LIGHTS; i ++ ) {\n\t\tspotLight = spotLights[ i ];\n\t\tgetSpotDirectLightIrradiance( spotLight, geometry, directLight );\n\t\t#ifdef USE_SHADOWMAP\n\t\tdirectLight.color *= all( bvec2( spotLight.shadow, directLight.visible ) ) ? getShadow( spotShadowMap[ i ], spotLight.shadowMapSize, spotLight.shadowBias, spotLight.shadowRadius, vSpotShadowCoord[ i ] ) : 1.0;\n\t\t#endif\n\t\tRE_Direct( directLight, geometry, material, reflectedLight );\n\t}\n#endif\n#if ( NUM_DIR_LIGHTS > 0 ) && defined( RE_Direct )\n\tDirectionalLight directionalLight;\n\tfor ( int i = 0; i < NUM_DIR_LIGHTS; i ++ ) {\n\t\tdirectionalLight = directionalLights[ i ];\n\t\tgetDirectionalDirectLightIrradiance( directionalLight, geometry, directLight );\n\t\t#ifdef USE_SHADOWMAP\n\t\tdirectLight.color *= all( bvec2( directionalLight.shadow, directLight.visible ) ) ? getShadow( directionalShadowMap[ i ], directionalLight.shadowMapSize, directionalLight.shadowBias, directionalLight.shadowRadius, vDirectionalShadowCoord[ i ] ) : 1.0;\n\t\t#endif\n\t\tRE_Direct( directLight, geometry, material, reflectedLight );\n\t}\n#endif\n#if ( NUM_RECT_AREA_LIGHTS > 0 ) && defined( RE_Direct_RectArea )\n\tRectAreaLight rectAreaLight;\n\tfor ( int i = 0; i < NUM_RECT_AREA_LIGHTS; i ++ ) {\n\t\trectAreaLight = rectAreaLights[ i ];\n\t\tRE_Direct_RectArea( rectAreaLight, geometry, material, reflectedLight );\n\t}\n#endif\n#if defined( RE_IndirectDiffuse )\n\tvec3 irradiance = getAmbientLightIrradiance( ambientLightColor );\n\t#ifdef USE_LIGHTMAP\n\t\tvec3 lightMapIrradiance = texture2D( lightMap, vUv2 ).xyz * lightMapIntensity;\n\t\t#ifndef PHYSICALLY_CORRECT_LIGHTS\n\t\t\tlightMapIrradiance *= PI;\n\t\t#endif\n\t\tirradiance += lightMapIrradiance;\n\t#endif\n\t#if ( NUM_HEMI_LIGHTS > 0 )\n\t\tfor ( int i = 0; i < NUM_HEMI_LIGHTS; i ++ ) {\n\t\t\tirradiance += getHemisphereLightIrradiance( hemisphereLights[ i ], geometry );\n\t\t}\n\t#endif\n\t#if defined( USE_ENVMAP ) && defined( PHYSICAL ) && defined( ENVMAP_TYPE_CUBE_UV )\n\t \tirradiance += getLightProbeIndirectIrradiance( geometry, 8 );\n\t#endif\n\tRE_IndirectDiffuse( irradiance, geometry, material, reflectedLight );\n#endif\n#if defined( USE_ENVMAP ) && defined( RE_IndirectSpecular )\n\tvec3 radiance = getLightProbeIndirectRadiance( geometry, Material_BlinnShininessExponent( material ), 8 );\n\t#ifndef STANDARD\n\t\tvec3 clearCoatRadiance = getLightProbeIndirectRadiance( geometry, Material_ClearCoat_BlinnShininessExponent( material ), 8 );\n\t#else\n\t\tvec3 clearCoatRadiance = vec3( 0.0 );\n\t#endif\n\t\t\n\tRE_IndirectSpecular( radiance, clearCoatRadiance, geometry, material, reflectedLight );\n#endif\n";

	var logdepthbuf_fragment = "#if defined(USE_LOGDEPTHBUF) && defined(USE_LOGDEPTHBUF_EXT)\n\tgl_FragDepthEXT = log2(vFragDepth) * logDepthBufFC * 0.5;\n#endif";

	var logdepthbuf_pars_fragment = "#ifdef USE_LOGDEPTHBUF\n\tuniform float logDepthBufFC;\n\t#ifdef USE_LOGDEPTHBUF_EXT\n\t\tvarying float vFragDepth;\n\t#endif\n#endif\n";

	var logdepthbuf_pars_vertex = "#ifdef USE_LOGDEPTHBUF\n\t#ifdef USE_LOGDEPTHBUF_EXT\n\t\tvarying float vFragDepth;\n\t#endif\n\tuniform float logDepthBufFC;\n#endif";

	var logdepthbuf_vertex = "#ifdef USE_LOGDEPTHBUF\n\tgl_Position.z = log2(max( EPSILON, gl_Position.w + 1.0 )) * logDepthBufFC;\n\t#ifdef USE_LOGDEPTHBUF_EXT\n\t\tvFragDepth = 1.0 + gl_Position.w;\n\t#else\n\t\tgl_Position.z = (gl_Position.z - 1.0) * gl_Position.w;\n\t#endif\n#endif\n";

	var map_fragment = "#ifdef USE_MAP\n\tvec4 texelColor = texture2D( map, vUv );\n\ttexelColor = mapTexelToLinear( texelColor );\n\tdiffuseColor *= texelColor;\n#endif\n";

	var map_pars_fragment = "#ifdef USE_MAP\n\tuniform sampler2D map;\n#endif\n";

	var map_particle_fragment = "#ifdef USE_MAP\n\tvec4 mapTexel = texture2D( map, vec2( gl_PointCoord.x, 1.0 - gl_PointCoord.y ) * offsetRepeat.zw + offsetRepeat.xy );\n\tdiffuseColor *= mapTexelToLinear( mapTexel );\n#endif\n";

	var map_particle_pars_fragment = "#ifdef USE_MAP\n\tuniform vec4 offsetRepeat;\n\tuniform sampler2D map;\n#endif\n";

	var metalnessmap_fragment = "float metalnessFactor = metalness;\n#ifdef USE_METALNESSMAP\n\tvec4 texelMetalness = texture2D( metalnessMap, vUv );\n\tmetalnessFactor *= texelMetalness.r;\n#endif\n";

	var metalnessmap_pars_fragment = "#ifdef USE_METALNESSMAP\n\tuniform sampler2D metalnessMap;\n#endif";

	var morphnormal_vertex = "#ifdef USE_MORPHNORMALS\n\tobjectNormal += ( morphNormal0 - normal ) * morphTargetInfluences[ 0 ];\n\tobjectNormal += ( morphNormal1 - normal ) * morphTargetInfluences[ 1 ];\n\tobjectNormal += ( morphNormal2 - normal ) * morphTargetInfluences[ 2 ];\n\tobjectNormal += ( morphNormal3 - normal ) * morphTargetInfluences[ 3 ];\n#endif\n";

	var morphtarget_pars_vertex = "#ifdef USE_MORPHTARGETS\n\t#ifndef USE_MORPHNORMALS\n\tuniform float morphTargetInfluences[ 8 ];\n\t#else\n\tuniform float morphTargetInfluences[ 4 ];\n\t#endif\n#endif";

	var morphtarget_vertex = "#ifdef USE_MORPHTARGETS\n\ttransformed += ( morphTarget0 - position ) * morphTargetInfluences[ 0 ];\n\ttransformed += ( morphTarget1 - position ) * morphTargetInfluences[ 1 ];\n\ttransformed += ( morphTarget2 - position ) * morphTargetInfluences[ 2 ];\n\ttransformed += ( morphTarget3 - position ) * morphTargetInfluences[ 3 ];\n\t#ifndef USE_MORPHNORMALS\n\ttransformed += ( morphTarget4 - position ) * morphTargetInfluences[ 4 ];\n\ttransformed += ( morphTarget5 - position ) * morphTargetInfluences[ 5 ];\n\ttransformed += ( morphTarget6 - position ) * morphTargetInfluences[ 6 ];\n\ttransformed += ( morphTarget7 - position ) * morphTargetInfluences[ 7 ];\n\t#endif\n#endif\n";

	var normal_flip = "#ifdef DOUBLE_SIDED\n\tfloat flipNormal = ( float( gl_FrontFacing ) * 2.0 - 1.0 );\n#else\n\tfloat flipNormal = 1.0;\n#endif\n";

	var normal_fragment = "#ifdef FLAT_SHADED\n\tvec3 fdx = vec3( dFdx( vViewPosition.x ), dFdx( vViewPosition.y ), dFdx( vViewPosition.z ) );\n\tvec3 fdy = vec3( dFdy( vViewPosition.x ), dFdy( vViewPosition.y ), dFdy( vViewPosition.z ) );\n\tvec3 normal = normalize( cross( fdx, fdy ) );\n#else\n\tvec3 normal = normalize( vNormal ) * flipNormal;\n#endif\n#ifdef USE_NORMALMAP\n\tnormal = perturbNormal2Arb( -vViewPosition, normal );\n#elif defined( USE_BUMPMAP )\n\tnormal = perturbNormalArb( -vViewPosition, normal, dHdxy_fwd() );\n#endif\n";

	var normalmap_pars_fragment = "#ifdef USE_NORMALMAP\n\tuniform sampler2D normalMap;\n\tuniform vec2 normalScale;\n\tvec3 perturbNormal2Arb( vec3 eye_pos, vec3 surf_norm ) {\n\t\tvec3 q0 = dFdx( eye_pos.xyz );\n\t\tvec3 q1 = dFdy( eye_pos.xyz );\n\t\tvec2 st0 = dFdx( vUv.st );\n\t\tvec2 st1 = dFdy( vUv.st );\n\t\tvec3 S = normalize( q0 * st1.t - q1 * st0.t );\n\t\tvec3 T = normalize( -q0 * st1.s + q1 * st0.s );\n\t\tvec3 N = normalize( surf_norm );\n\t\tvec3 mapN = texture2D( normalMap, vUv ).xyz * 2.0 - 1.0;\n\t\tmapN.xy = normalScale * mapN.xy;\n\t\tmat3 tsn = mat3( S, T, N );\n\t\treturn normalize( tsn * mapN );\n\t}\n#endif\n";

	var packing = "vec3 packNormalToRGB( const in vec3 normal ) {\n  return normalize( normal ) * 0.5 + 0.5;\n}\nvec3 unpackRGBToNormal( const in vec3 rgb ) {\n  return 1.0 - 2.0 * rgb.xyz;\n}\nconst float PackUpscale = 256. / 255.;const float UnpackDownscale = 255. / 256.;\nconst vec3 PackFactors = vec3( 256. * 256. * 256., 256. * 256.,  256. );\nconst vec4 UnpackFactors = UnpackDownscale / vec4( PackFactors, 1. );\nconst float ShiftRight8 = 1. / 256.;\nvec4 packDepthToRGBA( const in float v ) {\n\tvec4 r = vec4( fract( v * PackFactors ), v );\n\tr.yzw -= r.xyz * ShiftRight8;\treturn r * PackUpscale;\n}\nfloat unpackRGBAToDepth( const in vec4 v ) {\n\treturn dot( v, UnpackFactors );\n}\nfloat viewZToOrthographicDepth( const in float viewZ, const in float near, const in float far ) {\n  return ( viewZ + near ) / ( near - far );\n}\nfloat orthographicDepthToViewZ( const in float linearClipZ, const in float near, const in float far ) {\n  return linearClipZ * ( near - far ) - near;\n}\nfloat viewZToPerspectiveDepth( const in float viewZ, const in float near, const in float far ) {\n  return (( near + viewZ ) * far ) / (( far - near ) * viewZ );\n}\nfloat perspectiveDepthToViewZ( const in float invClipZ, const in float near, const in float far ) {\n  return ( near * far ) / ( ( far - near ) * invClipZ - far );\n}\n";

	var premultiplied_alpha_fragment = "#ifdef PREMULTIPLIED_ALPHA\n\tgl_FragColor.rgb *= gl_FragColor.a;\n#endif\n";

	var project_vertex = "#ifdef USE_SKINNING\n\tvec4 mvPosition = modelViewMatrix * skinned;\n#else\n\tvec4 mvPosition = modelViewMatrix * vec4( transformed, 1.0 );\n#endif\ngl_Position = projectionMatrix * mvPosition;\n";

	var roughnessmap_fragment = "float roughnessFactor = roughness;\n#ifdef USE_ROUGHNESSMAP\n\tvec4 texelRoughness = texture2D( roughnessMap, vUv );\n\troughnessFactor *= texelRoughness.r;\n#endif\n";

	var roughnessmap_pars_fragment = "#ifdef USE_ROUGHNESSMAP\n\tuniform sampler2D roughnessMap;\n#endif";

	var shadowmap_pars_fragment = "#ifdef USE_SHADOWMAP\n\t#if NUM_DIR_LIGHTS > 0\n\t\tuniform sampler2D directionalShadowMap[ NUM_DIR_LIGHTS ];\n\t\tvarying vec4 vDirectionalShadowCoord[ NUM_DIR_LIGHTS ];\n\t#endif\n\t#if NUM_SPOT_LIGHTS > 0\n\t\tuniform sampler2D spotShadowMap[ NUM_SPOT_LIGHTS ];\n\t\tvarying vec4 vSpotShadowCoord[ NUM_SPOT_LIGHTS ];\n\t#endif\n\t#if NUM_POINT_LIGHTS > 0\n\t\tuniform sampler2D pointShadowMap[ NUM_POINT_LIGHTS ];\n\t\tvarying vec4 vPointShadowCoord[ NUM_POINT_LIGHTS ];\n\t#endif\n    #if NUM_RECT_AREA_LIGHTS > 0\n    #endif\n\tfloat texture2DCompare( sampler2D depths, vec2 uv, float compare ) {\n\t\treturn step( compare, unpackRGBAToDepth( texture2D( depths, uv ) ) );\n\t}\n\tfloat texture2DShadowLerp( sampler2D depths, vec2 size, vec2 uv, float compare ) {\n\t\tconst vec2 offset = vec2( 0.0, 1.0 );\n\t\tvec2 texelSize = vec2( 1.0 ) / size;\n\t\tvec2 centroidUV = floor( uv * size + 0.5 ) / size;\n\t\tfloat lb = texture2DCompare( depths, centroidUV + texelSize * offset.xx, compare );\n\t\tfloat lt = texture2DCompare( depths, centroidUV + texelSize * offset.xy, compare );\n\t\tfloat rb = texture2DCompare( depths, centroidUV + texelSize * offset.yx, compare );\n\t\tfloat rt = texture2DCompare( depths, centroidUV + texelSize * offset.yy, compare );\n\t\tvec2 f = fract( uv * size + 0.5 );\n\t\tfloat a = mix( lb, lt, f.y );\n\t\tfloat b = mix( rb, rt, f.y );\n\t\tfloat c = mix( a, b, f.x );\n\t\treturn c;\n\t}\n\tfloat getShadow( sampler2D shadowMap, vec2 shadowMapSize, float shadowBias, float shadowRadius, vec4 shadowCoord ) {\n\t\tshadowCoord.xyz /= shadowCoord.w;\n\t\tshadowCoord.z += shadowBias;\n\t\tbvec4 inFrustumVec = bvec4 ( shadowCoord.x >= 0.0, shadowCoord.x <= 1.0, shadowCoord.y >= 0.0, shadowCoord.y <= 1.0 );\n\t\tbool inFrustum = all( inFrustumVec );\n\t\tbvec2 frustumTestVec = bvec2( inFrustum, shadowCoord.z <= 1.0 );\n\t\tbool frustumTest = all( frustumTestVec );\n\t\tif ( frustumTest ) {\n\t\t#if defined( SHADOWMAP_TYPE_PCF )\n\t\t\tvec2 texelSize = vec2( 1.0 ) / shadowMapSize;\n\t\t\tfloat dx0 = - texelSize.x * shadowRadius;\n\t\t\tfloat dy0 = - texelSize.y * shadowRadius;\n\t\t\tfloat dx1 = + texelSize.x * shadowRadius;\n\t\t\tfloat dy1 = + texelSize.y * shadowRadius;\n\t\t\treturn (\n\t\t\t\ttexture2DCompare( shadowMap, shadowCoord.xy + vec2( dx0, dy0 ), shadowCoord.z ) +\n\t\t\t\ttexture2DCompare( shadowMap, shadowCoord.xy + vec2( 0.0, dy0 ), shadowCoord.z ) +\n\t\t\t\ttexture2DCompare( shadowMap, shadowCoord.xy + vec2( dx1, dy0 ), shadowCoord.z ) +\n\t\t\t\ttexture2DCompare( shadowMap, shadowCoord.xy + vec2( dx0, 0.0 ), shadowCoord.z ) +\n\t\t\t\ttexture2DCompare( shadowMap, shadowCoord.xy, shadowCoord.z ) +\n\t\t\t\ttexture2DCompare( shadowMap, shadowCoord.xy + vec2( dx1, 0.0 ), shadowCoord.z ) +\n\t\t\t\ttexture2DCompare( shadowMap, shadowCoord.xy + vec2( dx0, dy1 ), shadowCoord.z ) +\n\t\t\t\ttexture2DCompare( shadowMap, shadowCoord.xy + vec2( 0.0, dy1 ), shadowCoord.z ) +\n\t\t\t\ttexture2DCompare( shadowMap, shadowCoord.xy + vec2( dx1, dy1 ), shadowCoord.z )\n\t\t\t) * ( 1.0 / 9.0 );\n\t\t#elif defined( SHADOWMAP_TYPE_PCF_SOFT )\n\t\t\tvec2 texelSize = vec2( 1.0 ) / shadowMapSize;\n\t\t\tfloat dx0 = - texelSize.x * shadowRadius;\n\t\t\tfloat dy0 = - texelSize.y * shadowRadius;\n\t\t\tfloat dx1 = + texelSize.x * shadowRadius;\n\t\t\tfloat dy1 = + texelSize.y * shadowRadius;\n\t\t\treturn (\n\t\t\t\ttexture2DShadowLerp( shadowMap, shadowMapSize, shadowCoord.xy + vec2( dx0, dy0 ), shadowCoord.z ) +\n\t\t\t\ttexture2DShadowLerp( shadowMap, shadowMapSize, shadowCoord.xy + vec2( 0.0, dy0 ), shadowCoord.z ) +\n\t\t\t\ttexture2DShadowLerp( shadowMap, shadowMapSize, shadowCoord.xy + vec2( dx1, dy0 ), shadowCoord.z ) +\n\t\t\t\ttexture2DShadowLerp( shadowMap, shadowMapSize, shadowCoord.xy + vec2( dx0, 0.0 ), shadowCoord.z ) +\n\t\t\t\ttexture2DShadowLerp( shadowMap, shadowMapSize, shadowCoord.xy, shadowCoord.z ) +\n\t\t\t\ttexture2DShadowLerp( shadowMap, shadowMapSize, shadowCoord.xy + vec2( dx1, 0.0 ), shadowCoord.z ) +\n\t\t\t\ttexture2DShadowLerp( shadowMap, shadowMapSize, shadowCoord.xy + vec2( dx0, dy1 ), shadowCoord.z ) +\n\t\t\t\ttexture2DShadowLerp( shadowMap, shadowMapSize, shadowCoord.xy + vec2( 0.0, dy1 ), shadowCoord.z ) +\n\t\t\t\ttexture2DShadowLerp( shadowMap, shadowMapSize, shadowCoord.xy + vec2( dx1, dy1 ), shadowCoord.z )\n\t\t\t) * ( 1.0 / 9.0 );\n\t\t#else\n\t\t\treturn texture2DCompare( shadowMap, shadowCoord.xy, shadowCoord.z );\n\t\t#endif\n\t\t}\n\t\treturn 1.0;\n\t}\n\tvec2 cubeToUV( vec3 v, float texelSizeY ) {\n\t\tvec3 absV = abs( v );\n\t\tfloat scaleToCube = 1.0 / max( absV.x, max( absV.y, absV.z ) );\n\t\tabsV *= scaleToCube;\n\t\tv *= scaleToCube * ( 1.0 - 2.0 * texelSizeY );\n\t\tvec2 planar = v.xy;\n\t\tfloat almostATexel = 1.5 * texelSizeY;\n\t\tfloat almostOne = 1.0 - almostATexel;\n\t\tif ( absV.z >= almostOne ) {\n\t\t\tif ( v.z > 0.0 )\n\t\t\t\tplanar.x = 4.0 - v.x;\n\t\t} else if ( absV.x >= almostOne ) {\n\t\t\tfloat signX = sign( v.x );\n\t\t\tplanar.x = v.z * signX + 2.0 * signX;\n\t\t} else if ( absV.y >= almostOne ) {\n\t\t\tfloat signY = sign( v.y );\n\t\t\tplanar.x = v.x + 2.0 * signY + 2.0;\n\t\t\tplanar.y = v.z * signY - 2.0;\n\t\t}\n\t\treturn vec2( 0.125, 0.25 ) * planar + vec2( 0.375, 0.75 );\n\t}\n\tfloat getPointShadow( sampler2D shadowMap, vec2 shadowMapSize, float shadowBias, float shadowRadius, vec4 shadowCoord ) {\n\t\tvec2 texelSize = vec2( 1.0 ) / ( shadowMapSize * vec2( 4.0, 2.0 ) );\n\t\tvec3 lightToPosition = shadowCoord.xyz;\n\t\tvec3 bd3D = normalize( lightToPosition );\n\t\tfloat dp = ( length( lightToPosition ) - shadowBias ) / 1000.0;\n\t\t#if defined( SHADOWMAP_TYPE_PCF ) || defined( SHADOWMAP_TYPE_PCF_SOFT )\n\t\t\tvec2 offset = vec2( - 1, 1 ) * shadowRadius * texelSize.y;\n\t\t\treturn (\n\t\t\t\ttexture2DCompare( shadowMap, cubeToUV( bd3D + offset.xyy, texelSize.y ), dp ) +\n\t\t\t\ttexture2DCompare( shadowMap, cubeToUV( bd3D + offset.yyy, texelSize.y ), dp ) +\n\t\t\t\ttexture2DCompare( shadowMap, cubeToUV( bd3D + offset.xyx, texelSize.y ), dp ) +\n\t\t\t\ttexture2DCompare( shadowMap, cubeToUV( bd3D + offset.yyx, texelSize.y ), dp ) +\n\t\t\t\ttexture2DCompare( shadowMap, cubeToUV( bd3D, texelSize.y ), dp ) +\n\t\t\t\ttexture2DCompare( shadowMap, cubeToUV( bd3D + offset.xxy, texelSize.y ), dp ) +\n\t\t\t\ttexture2DCompare( shadowMap, cubeToUV( bd3D + offset.yxy, texelSize.y ), dp ) +\n\t\t\t\ttexture2DCompare( shadowMap, cubeToUV( bd3D + offset.xxx, texelSize.y ), dp ) +\n\t\t\t\ttexture2DCompare( shadowMap, cubeToUV( bd3D + offset.yxx, texelSize.y ), dp )\n\t\t\t) * ( 1.0 / 9.0 );\n\t\t#else\n\t\t\treturn texture2DCompare( shadowMap, cubeToUV( bd3D, texelSize.y ), dp );\n\t\t#endif\n\t}\n#endif\n";

	var shadowmap_pars_vertex = "#ifdef USE_SHADOWMAP\n\t#if NUM_DIR_LIGHTS > 0\n\t\tuniform mat4 directionalShadowMatrix[ NUM_DIR_LIGHTS ];\n\t\tvarying vec4 vDirectionalShadowCoord[ NUM_DIR_LIGHTS ];\n\t#endif\n\t#if NUM_SPOT_LIGHTS > 0\n\t\tuniform mat4 spotShadowMatrix[ NUM_SPOT_LIGHTS ];\n\t\tvarying vec4 vSpotShadowCoord[ NUM_SPOT_LIGHTS ];\n\t#endif\n\t#if NUM_POINT_LIGHTS > 0\n\t\tuniform mat4 pointShadowMatrix[ NUM_POINT_LIGHTS ];\n\t\tvarying vec4 vPointShadowCoord[ NUM_POINT_LIGHTS ];\n\t#endif\n    #if NUM_RECT_AREA_LIGHTS > 0\n    #endif\n#endif\n";

	var shadowmap_vertex = "#ifdef USE_SHADOWMAP\n\t#if NUM_DIR_LIGHTS > 0\n\tfor ( int i = 0; i < NUM_DIR_LIGHTS; i ++ ) {\n\t\tvDirectionalShadowCoord[ i ] = directionalShadowMatrix[ i ] * worldPosition;\n\t}\n\t#endif\n\t#if NUM_SPOT_LIGHTS > 0\n\tfor ( int i = 0; i < NUM_SPOT_LIGHTS; i ++ ) {\n\t\tvSpotShadowCoord[ i ] = spotShadowMatrix[ i ] * worldPosition;\n\t}\n\t#endif\n\t#if NUM_POINT_LIGHTS > 0\n\tfor ( int i = 0; i < NUM_POINT_LIGHTS; i ++ ) {\n\t\tvPointShadowCoord[ i ] = pointShadowMatrix[ i ] * worldPosition;\n\t}\n\t#endif\n    #if NUM_RECT_AREA_LIGHTS > 0\n    #endif\n#endif\n";

	var shadowmask_pars_fragment = "float getShadowMask() {\n\tfloat shadow = 1.0;\n\t#ifdef USE_SHADOWMAP\n\t#if NUM_DIR_LIGHTS > 0\n\tDirectionalLight directionalLight;\n\tfor ( int i = 0; i < NUM_DIR_LIGHTS; i ++ ) {\n\t\tdirectionalLight = directionalLights[ i ];\n\t\tshadow *= bool( directionalLight.shadow ) ? getShadow( directionalShadowMap[ i ], directionalLight.shadowMapSize, directionalLight.shadowBias, directionalLight.shadowRadius, vDirectionalShadowCoord[ i ] ) : 1.0;\n\t}\n\t#endif\n\t#if NUM_SPOT_LIGHTS > 0\n\tSpotLight spotLight;\n\tfor ( int i = 0; i < NUM_SPOT_LIGHTS; i ++ ) {\n\t\tspotLight = spotLights[ i ];\n\t\tshadow *= bool( spotLight.shadow ) ? getShadow( spotShadowMap[ i ], spotLight.shadowMapSize, spotLight.shadowBias, spotLight.shadowRadius, vSpotShadowCoord[ i ] ) : 1.0;\n\t}\n\t#endif\n\t#if NUM_POINT_LIGHTS > 0\n\tPointLight pointLight;\n\tfor ( int i = 0; i < NUM_POINT_LIGHTS; i ++ ) {\n\t\tpointLight = pointLights[ i ];\n\t\tshadow *= bool( pointLight.shadow ) ? getPointShadow( pointShadowMap[ i ], pointLight.shadowMapSize, pointLight.shadowBias, pointLight.shadowRadius, vPointShadowCoord[ i ] ) : 1.0;\n\t}\n\t#endif\n\t#if NUM_RECT_AREA_LIGHTS > 0\n\t#endif\n\t#endif\n\treturn shadow;\n}\n";

	var skinbase_vertex = "#ifdef USE_SKINNING\n\tmat4 boneMatX = getBoneMatrix( skinIndex.x );\n\tmat4 boneMatY = getBoneMatrix( skinIndex.y );\n\tmat4 boneMatZ = getBoneMatrix( skinIndex.z );\n\tmat4 boneMatW = getBoneMatrix( skinIndex.w );\n#endif";

	var skinning_pars_vertex = "#ifdef USE_SKINNING\n\tuniform mat4 bindMatrix;\n\tuniform mat4 bindMatrixInverse;\n\t#ifdef BONE_TEXTURE\n\t\tuniform sampler2D boneTexture;\n\t\tuniform int boneTextureWidth;\n\t\tuniform int boneTextureHeight;\n\t\tmat4 getBoneMatrix( const in float i ) {\n\t\t\tfloat j = i * 4.0;\n\t\t\tfloat x = mod( j, float( boneTextureWidth ) );\n\t\t\tfloat y = floor( j / float( boneTextureWidth ) );\n\t\t\tfloat dx = 1.0 / float( boneTextureWidth );\n\t\t\tfloat dy = 1.0 / float( boneTextureHeight );\n\t\t\ty = dy * ( y + 0.5 );\n\t\t\tvec4 v1 = texture2D( boneTexture, vec2( dx * ( x + 0.5 ), y ) );\n\t\t\tvec4 v2 = texture2D( boneTexture, vec2( dx * ( x + 1.5 ), y ) );\n\t\t\tvec4 v3 = texture2D( boneTexture, vec2( dx * ( x + 2.5 ), y ) );\n\t\t\tvec4 v4 = texture2D( boneTexture, vec2( dx * ( x + 3.5 ), y ) );\n\t\t\tmat4 bone = mat4( v1, v2, v3, v4 );\n\t\t\treturn bone;\n\t\t}\n\t#else\n\t\tuniform mat4 boneMatrices[ MAX_BONES ];\n\t\tmat4 getBoneMatrix( const in float i ) {\n\t\t\tmat4 bone = boneMatrices[ int(i) ];\n\t\t\treturn bone;\n\t\t}\n\t#endif\n#endif\n";

	var skinning_vertex = "#ifdef USE_SKINNING\n\tvec4 skinVertex = bindMatrix * vec4( transformed, 1.0 );\n\tvec4 skinned = vec4( 0.0 );\n\tskinned += boneMatX * skinVertex * skinWeight.x;\n\tskinned += boneMatY * skinVertex * skinWeight.y;\n\tskinned += boneMatZ * skinVertex * skinWeight.z;\n\tskinned += boneMatW * skinVertex * skinWeight.w;\n\tskinned  = bindMatrixInverse * skinned;\n#endif\n";

	var skinnormal_vertex = "#ifdef USE_SKINNING\n\tmat4 skinMatrix = mat4( 0.0 );\n\tskinMatrix += skinWeight.x * boneMatX;\n\tskinMatrix += skinWeight.y * boneMatY;\n\tskinMatrix += skinWeight.z * boneMatZ;\n\tskinMatrix += skinWeight.w * boneMatW;\n\tskinMatrix  = bindMatrixInverse * skinMatrix * bindMatrix;\n\tobjectNormal = vec4( skinMatrix * vec4( objectNormal, 0.0 ) ).xyz;\n#endif\n";

	var specularmap_fragment = "float specularStrength;\n#ifdef USE_SPECULARMAP\n\tvec4 texelSpecular = texture2D( specularMap, vUv );\n\tspecularStrength = texelSpecular.r;\n#else\n\tspecularStrength = 1.0;\n#endif";

	var specularmap_pars_fragment = "#ifdef USE_SPECULARMAP\n\tuniform sampler2D specularMap;\n#endif";

	var tonemapping_fragment = "#if defined( TONE_MAPPING )\n  gl_FragColor.rgb = toneMapping( gl_FragColor.rgb );\n#endif\n";

	var tonemapping_pars_fragment = "#define saturate(a) clamp( a, 0.0, 1.0 )\nuniform float toneMappingExposure;\nuniform float toneMappingWhitePoint;\nvec3 LinearToneMapping( vec3 color ) {\n  return toneMappingExposure * color;\n}\nvec3 ReinhardToneMapping( vec3 color ) {\n  color *= toneMappingExposure;\n  return saturate( color / ( vec3( 1.0 ) + color ) );\n}\n#define Uncharted2Helper( x ) max( ( ( x * ( 0.15 * x + 0.10 * 0.50 ) + 0.20 * 0.02 ) / ( x * ( 0.15 * x + 0.50 ) + 0.20 * 0.30 ) ) - 0.02 / 0.30, vec3( 0.0 ) )\nvec3 Uncharted2ToneMapping( vec3 color ) {\n  color *= toneMappingExposure;\n  return saturate( Uncharted2Helper( color ) / Uncharted2Helper( vec3( toneMappingWhitePoint ) ) );\n}\nvec3 OptimizedCineonToneMapping( vec3 color ) {\n  color *= toneMappingExposure;\n  color = max( vec3( 0.0 ), color - 0.004 );\n  return pow( ( color * ( 6.2 * color + 0.5 ) ) / ( color * ( 6.2 * color + 1.7 ) + 0.06 ), vec3( 2.2 ) );\n}\n";

	var uv_pars_fragment = "#if defined( USE_MAP ) || defined( USE_BUMPMAP ) || defined( USE_NORMALMAP ) || defined( USE_SPECULARMAP ) || defined( USE_ALPHAMAP ) || defined( USE_EMISSIVEMAP ) || defined( USE_ROUGHNESSMAP ) || defined( USE_METALNESSMAP )\n\tvarying vec2 vUv;\n#endif";

	var uv_pars_vertex = "#if defined( USE_MAP ) || defined( USE_BUMPMAP ) || defined( USE_NORMALMAP ) || defined( USE_SPECULARMAP ) || defined( USE_ALPHAMAP ) || defined( USE_EMISSIVEMAP ) || defined( USE_ROUGHNESSMAP ) || defined( USE_METALNESSMAP )\n\tvarying vec2 vUv;\n\tuniform vec4 offsetRepeat;\n#endif\n";

	var uv_vertex = "#if defined( USE_MAP ) || defined( USE_BUMPMAP ) || defined( USE_NORMALMAP ) || defined( USE_SPECULARMAP ) || defined( USE_ALPHAMAP ) || defined( USE_EMISSIVEMAP ) || defined( USE_ROUGHNESSMAP ) || defined( USE_METALNESSMAP )\n\tvUv = uv * offsetRepeat.zw + offsetRepeat.xy;\n#endif";

	var uv2_pars_fragment = "#if defined( USE_LIGHTMAP ) || defined( USE_AOMAP )\n\tvarying vec2 vUv2;\n#endif";

	var uv2_pars_vertex = "#if defined( USE_LIGHTMAP ) || defined( USE_AOMAP )\n\tattribute vec2 uv2;\n\tvarying vec2 vUv2;\n#endif";

	var uv2_vertex = "#if defined( USE_LIGHTMAP ) || defined( USE_AOMAP )\n\tvUv2 = uv2;\n#endif";

	var worldpos_vertex = "#if defined( USE_ENVMAP ) || defined( PHONG ) || defined( PHYSICAL ) || defined( LAMBERT ) || defined ( USE_SHADOWMAP )\n\t#ifdef USE_SKINNING\n\t\tvec4 worldPosition = modelMatrix * skinned;\n\t#else\n\t\tvec4 worldPosition = modelMatrix * vec4( transformed, 1.0 );\n\t#endif\n#endif\n";

	var cube_frag = "uniform samplerCube tCube;\nuniform float tFlip;\nuniform float opacity;\nvarying vec3 vWorldPosition;\n#include <common>\nvoid main() {\n\tgl_FragColor = textureCube( tCube, vec3( tFlip * vWorldPosition.x, vWorldPosition.yz ) );\n\tgl_FragColor.a *= opacity;\n}\n";

	var cube_vert = "varying vec3 vWorldPosition;\n#include <common>\nvoid main() {\n\tvWorldPosition = transformDirection( position, modelMatrix );\n\t#include <begin_vertex>\n\t#include <project_vertex>\n}\n";

	var depth_frag = "#if DEPTH_PACKING == 3200\n\tuniform float opacity;\n#endif\n#include <common>\n#include <packing>\n#include <uv_pars_fragment>\n#include <map_pars_fragment>\n#include <alphamap_pars_fragment>\n#include <logdepthbuf_pars_fragment>\n#include <clipping_planes_pars_fragment>\nvoid main() {\n\t#include <clipping_planes_fragment>\n\tvec4 diffuseColor = vec4( 1.0 );\n\t#if DEPTH_PACKING == 3200\n\t\tdiffuseColor.a = opacity;\n\t#endif\n\t#include <map_fragment>\n\t#include <alphamap_fragment>\n\t#include <alphatest_fragment>\n\t#include <logdepthbuf_fragment>\n\t#if DEPTH_PACKING == 3200\n\t\tgl_FragColor = vec4( vec3( gl_FragCoord.z ), opacity );\n\t#elif DEPTH_PACKING == 3201\n\t\tgl_FragColor = packDepthToRGBA( gl_FragCoord.z );\n\t#endif\n}\n";

	var depth_vert = "#include <common>\n#include <uv_pars_vertex>\n#include <displacementmap_pars_vertex>\n#include <morphtarget_pars_vertex>\n#include <skinning_pars_vertex>\n#include <logdepthbuf_pars_vertex>\n#include <clipping_planes_pars_vertex>\nvoid main() {\n\t#include <uv_vertex>\n\t#include <skinbase_vertex>\n\t#include <begin_vertex>\n\t#include <displacementmap_vertex>\n\t#include <morphtarget_vertex>\n\t#include <skinning_vertex>\n\t#include <project_vertex>\n\t#include <logdepthbuf_vertex>\n\t#include <clipping_planes_vertex>\n}\n";

	var distanceRGBA_frag = "uniform vec3 lightPos;\nvarying vec4 vWorldPosition;\n#include <common>\n#include <packing>\n#include <clipping_planes_pars_fragment>\nvoid main () {\n\t#include <clipping_planes_fragment>\n\tgl_FragColor = packDepthToRGBA( length( vWorldPosition.xyz - lightPos.xyz ) / 1000.0 );\n}\n";

	var distanceRGBA_vert = "varying vec4 vWorldPosition;\n#include <common>\n#include <morphtarget_pars_vertex>\n#include <skinning_pars_vertex>\n#include <clipping_planes_pars_vertex>\nvoid main() {\n\t#include <skinbase_vertex>\n\t#include <begin_vertex>\n\t#include <morphtarget_vertex>\n\t#include <skinning_vertex>\n\t#include <project_vertex>\n\t#include <worldpos_vertex>\n\t#include <clipping_planes_vertex>\n\tvWorldPosition = worldPosition;\n}\n";

	var equirect_frag = "uniform sampler2D tEquirect;\nuniform float tFlip;\nvarying vec3 vWorldPosition;\n#include <common>\nvoid main() {\n\tvec3 direction = normalize( vWorldPosition );\n\tvec2 sampleUV;\n\tsampleUV.y = saturate( tFlip * direction.y * -0.5 + 0.5 );\n\tsampleUV.x = atan( direction.z, direction.x ) * RECIPROCAL_PI2 + 0.5;\n\tgl_FragColor = texture2D( tEquirect, sampleUV );\n}\n";

	var equirect_vert = "varying vec3 vWorldPosition;\n#include <common>\nvoid main() {\n\tvWorldPosition = transformDirection( position, modelMatrix );\n\t#include <begin_vertex>\n\t#include <project_vertex>\n}\n";

	var linedashed_frag = "uniform vec3 diffuse;\nuniform float opacity;\nuniform float dashSize;\nuniform float totalSize;\nvarying float vLineDistance;\n#include <common>\n#include <color_pars_fragment>\n#include <fog_pars_fragment>\n#include <logdepthbuf_pars_fragment>\n#include <clipping_planes_pars_fragment>\nvoid main() {\n\t#include <clipping_planes_fragment>\n\tif ( mod( vLineDistance, totalSize ) > dashSize ) {\n\t\tdiscard;\n\t}\n\tvec3 outgoingLight = vec3( 0.0 );\n\tvec4 diffuseColor = vec4( diffuse, opacity );\n\t#include <logdepthbuf_fragment>\n\t#include <color_fragment>\n\toutgoingLight = diffuseColor.rgb;\n\tgl_FragColor = vec4( outgoingLight, diffuseColor.a );\n\t#include <premultiplied_alpha_fragment>\n\t#include <tonemapping_fragment>\n\t#include <encodings_fragment>\n\t#include <fog_fragment>\n}\n";

	var linedashed_vert = "uniform float scale;\nattribute float lineDistance;\nvarying float vLineDistance;\n#include <common>\n#include <color_pars_vertex>\n#include <logdepthbuf_pars_vertex>\n#include <clipping_planes_pars_vertex>\nvoid main() {\n\t#include <color_vertex>\n\tvLineDistance = scale * lineDistance;\n\tvec4 mvPosition = modelViewMatrix * vec4( position, 1.0 );\n\tgl_Position = projectionMatrix * mvPosition;\n\t#include <logdepthbuf_vertex>\n\t#include <clipping_planes_vertex>\n}\n";

	var meshbasic_frag = "uniform vec3 diffuse;\nuniform float opacity;\n#ifndef FLAT_SHADED\n\tvarying vec3 vNormal;\n#endif\n#include <common>\n#include <color_pars_fragment>\n#include <uv_pars_fragment>\n#include <uv2_pars_fragment>\n#include <map_pars_fragment>\n#include <alphamap_pars_fragment>\n#include <aomap_pars_fragment>\n#include <lightmap_pars_fragment>\n#include <envmap_pars_fragment>\n#include <fog_pars_fragment>\n#include <specularmap_pars_fragment>\n#include <logdepthbuf_pars_fragment>\n#include <clipping_planes_pars_fragment>\nvoid main() {\n\t#include <clipping_planes_fragment>\n\tvec4 diffuseColor = vec4( diffuse, opacity );\n\t#include <logdepthbuf_fragment>\n\t#include <map_fragment>\n\t#include <color_fragment>\n\t#include <alphamap_fragment>\n\t#include <alphatest_fragment>\n\t#include <specularmap_fragment>\n\tReflectedLight reflectedLight = ReflectedLight( vec3( 0.0 ), vec3( 0.0 ), vec3( 0.0 ), vec3( 0.0 ) );\n\t#ifdef USE_LIGHTMAP\n\t\treflectedLight.indirectDiffuse += texture2D( lightMap, vUv2 ).xyz * lightMapIntensity;\n\t#else\n\t\treflectedLight.indirectDiffuse += vec3( 1.0 );\n\t#endif\n\t#include <aomap_fragment>\n\treflectedLight.indirectDiffuse *= diffuseColor.rgb;\n\tvec3 outgoingLight = reflectedLight.indirectDiffuse;\n\t#include <normal_flip>\n\t#include <envmap_fragment>\n\tgl_FragColor = vec4( outgoingLight, diffuseColor.a );\n\t#include <premultiplied_alpha_fragment>\n\t#include <tonemapping_fragment>\n\t#include <encodings_fragment>\n\t#include <fog_fragment>\n}\n";

	var meshbasic_vert = "#include <common>\n#include <uv_pars_vertex>\n#include <uv2_pars_vertex>\n#include <envmap_pars_vertex>\n#include <color_pars_vertex>\n#include <morphtarget_pars_vertex>\n#include <skinning_pars_vertex>\n#include <logdepthbuf_pars_vertex>\n#include <clipping_planes_pars_vertex>\nvoid main() {\n\t#include <uv_vertex>\n\t#include <uv2_vertex>\n\t#include <color_vertex>\n\t#include <skinbase_vertex>\n\t#ifdef USE_ENVMAP\n\t#include <beginnormal_vertex>\n\t#include <morphnormal_vertex>\n\t#include <skinnormal_vertex>\n\t#include <defaultnormal_vertex>\n\t#endif\n\t#include <begin_vertex>\n\t#include <morphtarget_vertex>\n\t#include <skinning_vertex>\n\t#include <project_vertex>\n\t#include <logdepthbuf_vertex>\n\t#include <worldpos_vertex>\n\t#include <clipping_planes_vertex>\n\t#include <envmap_vertex>\n}\n";

	var meshlambert_frag = "uniform vec3 diffuse;\nuniform vec3 emissive;\nuniform float opacity;\nvarying vec3 vLightFront;\n#ifdef DOUBLE_SIDED\n\tvarying vec3 vLightBack;\n#endif\n#include <common>\n#include <packing>\n#include <color_pars_fragment>\n#include <uv_pars_fragment>\n#include <uv2_pars_fragment>\n#include <map_pars_fragment>\n#include <alphamap_pars_fragment>\n#include <aomap_pars_fragment>\n#include <lightmap_pars_fragment>\n#include <emissivemap_pars_fragment>\n#include <envmap_pars_fragment>\n#include <bsdfs>\n#include <lights_pars>\n#include <fog_pars_fragment>\n#include <shadowmap_pars_fragment>\n#include <shadowmask_pars_fragment>\n#include <specularmap_pars_fragment>\n#include <logdepthbuf_pars_fragment>\n#include <clipping_planes_pars_fragment>\nvoid main() {\n\t#include <clipping_planes_fragment>\n\tvec4 diffuseColor = vec4( diffuse, opacity );\n\tReflectedLight reflectedLight = ReflectedLight( vec3( 0.0 ), vec3( 0.0 ), vec3( 0.0 ), vec3( 0.0 ) );\n\tvec3 totalEmissiveRadiance = emissive;\n\t#include <logdepthbuf_fragment>\n\t#include <map_fragment>\n\t#include <color_fragment>\n\t#include <alphamap_fragment>\n\t#include <alphatest_fragment>\n\t#include <specularmap_fragment>\n\t#include <emissivemap_fragment>\n\treflectedLight.indirectDiffuse = getAmbientLightIrradiance( ambientLightColor );\n\t#include <lightmap_fragment>\n\treflectedLight.indirectDiffuse *= BRDF_Diffuse_Lambert( diffuseColor.rgb );\n\t#ifdef DOUBLE_SIDED\n\t\treflectedLight.directDiffuse = ( gl_FrontFacing ) ? vLightFront : vLightBack;\n\t#else\n\t\treflectedLight.directDiffuse = vLightFront;\n\t#endif\n\treflectedLight.directDiffuse *= BRDF_Diffuse_Lambert( diffuseColor.rgb ) * getShadowMask();\n\t#include <aomap_fragment>\n\tvec3 outgoingLight = reflectedLight.directDiffuse + reflectedLight.indirectDiffuse + totalEmissiveRadiance;\n\t#include <normal_flip>\n\t#include <envmap_fragment>\n\tgl_FragColor = vec4( outgoingLight, diffuseColor.a );\n\t#include <premultiplied_alpha_fragment>\n\t#include <tonemapping_fragment>\n\t#include <encodings_fragment>\n\t#include <fog_fragment>\n}\n";

	var meshlambert_vert = "#define LAMBERT\nvarying vec3 vLightFront;\n#ifdef DOUBLE_SIDED\n\tvarying vec3 vLightBack;\n#endif\n#include <common>\n#include <uv_pars_vertex>\n#include <uv2_pars_vertex>\n#include <envmap_pars_vertex>\n#include <bsdfs>\n#include <lights_pars>\n#include <color_pars_vertex>\n#include <morphtarget_pars_vertex>\n#include <skinning_pars_vertex>\n#include <shadowmap_pars_vertex>\n#include <logdepthbuf_pars_vertex>\n#include <clipping_planes_pars_vertex>\nvoid main() {\n\t#include <uv_vertex>\n\t#include <uv2_vertex>\n\t#include <color_vertex>\n\t#include <beginnormal_vertex>\n\t#include <morphnormal_vertex>\n\t#include <skinbase_vertex>\n\t#include <skinnormal_vertex>\n\t#include <defaultnormal_vertex>\n\t#include <begin_vertex>\n\t#include <morphtarget_vertex>\n\t#include <skinning_vertex>\n\t#include <project_vertex>\n\t#include <logdepthbuf_vertex>\n\t#include <clipping_planes_vertex>\n\t#include <worldpos_vertex>\n\t#include <envmap_vertex>\n\t#include <lights_lambert_vertex>\n\t#include <shadowmap_vertex>\n}\n";

	var meshphong_frag = "#define PHONG\nuniform vec3 diffuse;\nuniform vec3 emissive;\nuniform vec3 specular;\nuniform float shininess;\nuniform float opacity;\n#include <common>\n#include <packing>\n#include <color_pars_fragment>\n#include <uv_pars_fragment>\n#include <uv2_pars_fragment>\n#include <map_pars_fragment>\n#include <alphamap_pars_fragment>\n#include <aomap_pars_fragment>\n#include <lightmap_pars_fragment>\n#include <emissivemap_pars_fragment>\n#include <envmap_pars_fragment>\n#include <gradientmap_pars_fragment>\n#include <fog_pars_fragment>\n#include <bsdfs>\n#include <lights_pars>\n#include <lights_phong_pars_fragment>\n#include <shadowmap_pars_fragment>\n#include <bumpmap_pars_fragment>\n#include <normalmap_pars_fragment>\n#include <specularmap_pars_fragment>\n#include <logdepthbuf_pars_fragment>\n#include <clipping_planes_pars_fragment>\nvoid main() {\n\t#include <clipping_planes_fragment>\n\tvec4 diffuseColor = vec4( diffuse, opacity );\n\tReflectedLight reflectedLight = ReflectedLight( vec3( 0.0 ), vec3( 0.0 ), vec3( 0.0 ), vec3( 0.0 ) );\n\tvec3 totalEmissiveRadiance = emissive;\n\t#include <logdepthbuf_fragment>\n\t#include <map_fragment>\n\t#include <color_fragment>\n\t#include <alphamap_fragment>\n\t#include <alphatest_fragment>\n\t#include <specularmap_fragment>\n\t#include <normal_flip>\n\t#include <normal_fragment>\n\t#include <emissivemap_fragment>\n\t#include <lights_phong_fragment>\n\t#include <lights_template>\n\t#include <aomap_fragment>\n\tvec3 outgoingLight = reflectedLight.directDiffuse + reflectedLight.indirectDiffuse + reflectedLight.directSpecular + reflectedLight.indirectSpecular + totalEmissiveRadiance;\n\t#include <envmap_fragment>\n\tgl_FragColor = vec4( outgoingLight, diffuseColor.a );\n\t#include <premultiplied_alpha_fragment>\n\t#include <tonemapping_fragment>\n\t#include <encodings_fragment>\n\t#include <fog_fragment>\n}\n";

	var meshphong_vert = "#define PHONG\nvarying vec3 vViewPosition;\n#ifndef FLAT_SHADED\n\tvarying vec3 vNormal;\n#endif\n#include <common>\n#include <uv_pars_vertex>\n#include <uv2_pars_vertex>\n#include <displacementmap_pars_vertex>\n#include <envmap_pars_vertex>\n#include <color_pars_vertex>\n#include <morphtarget_pars_vertex>\n#include <skinning_pars_vertex>\n#include <shadowmap_pars_vertex>\n#include <logdepthbuf_pars_vertex>\n#include <clipping_planes_pars_vertex>\nvoid main() {\n\t#include <uv_vertex>\n\t#include <uv2_vertex>\n\t#include <color_vertex>\n\t#include <beginnormal_vertex>\n\t#include <morphnormal_vertex>\n\t#include <skinbase_vertex>\n\t#include <skinnormal_vertex>\n\t#include <defaultnormal_vertex>\n#ifndef FLAT_SHADED\n\tvNormal = normalize( transformedNormal );\n#endif\n\t#include <begin_vertex>\n\t#include <displacementmap_vertex>\n\t#include <morphtarget_vertex>\n\t#include <skinning_vertex>\n\t#include <project_vertex>\n\t#include <logdepthbuf_vertex>\n\t#include <clipping_planes_vertex>\n\tvViewPosition = - mvPosition.xyz;\n\t#include <worldpos_vertex>\n\t#include <envmap_vertex>\n\t#include <shadowmap_vertex>\n}\n";

	var meshphysical_frag = "#define PHYSICAL\nuniform vec3 diffuse;\nuniform vec3 emissive;\nuniform float roughness;\nuniform float metalness;\nuniform float opacity;\n#ifndef STANDARD\n\tuniform float clearCoat;\n\tuniform float clearCoatRoughness;\n#endif\nvarying vec3 vViewPosition;\n#ifndef FLAT_SHADED\n\tvarying vec3 vNormal;\n#endif\n#include <common>\n#include <packing>\n#include <color_pars_fragment>\n#include <uv_pars_fragment>\n#include <uv2_pars_fragment>\n#include <map_pars_fragment>\n#include <alphamap_pars_fragment>\n#include <aomap_pars_fragment>\n#include <lightmap_pars_fragment>\n#include <emissivemap_pars_fragment>\n#include <envmap_pars_fragment>\n#include <fog_pars_fragment>\n#include <bsdfs>\n#include <cube_uv_reflection_fragment>\n#include <lights_pars>\n#include <lights_physical_pars_fragment>\n#include <shadowmap_pars_fragment>\n#include <bumpmap_pars_fragment>\n#include <normalmap_pars_fragment>\n#include <roughnessmap_pars_fragment>\n#include <metalnessmap_pars_fragment>\n#include <logdepthbuf_pars_fragment>\n#include <clipping_planes_pars_fragment>\nvoid main() {\n\t#include <clipping_planes_fragment>\n\tvec4 diffuseColor = vec4( diffuse, opacity );\n\tReflectedLight reflectedLight = ReflectedLight( vec3( 0.0 ), vec3( 0.0 ), vec3( 0.0 ), vec3( 0.0 ) );\n\tvec3 totalEmissiveRadiance = emissive;\n\t#include <logdepthbuf_fragment>\n\t#include <map_fragment>\n\t#include <color_fragment>\n\t#include <alphamap_fragment>\n\t#include <alphatest_fragment>\n\t#include <specularmap_fragment>\n\t#include <roughnessmap_fragment>\n\t#include <metalnessmap_fragment>\n\t#include <normal_flip>\n\t#include <normal_fragment>\n\t#include <emissivemap_fragment>\n\t#include <lights_physical_fragment>\n\t#include <lights_template>\n\t#include <aomap_fragment>\n\tvec3 outgoingLight = reflectedLight.directDiffuse + reflectedLight.indirectDiffuse + reflectedLight.directSpecular + reflectedLight.indirectSpecular + totalEmissiveRadiance;\n\tgl_FragColor = vec4( outgoingLight, diffuseColor.a );\n\t#include <premultiplied_alpha_fragment>\n\t#include <tonemapping_fragment>\n\t#include <encodings_fragment>\n\t#include <fog_fragment>\n}\n";

	var meshphysical_vert = "#define PHYSICAL\nvarying vec3 vViewPosition;\n#ifndef FLAT_SHADED\n\tvarying vec3 vNormal;\n#endif\n#include <common>\n#include <uv_pars_vertex>\n#include <uv2_pars_vertex>\n#include <displacementmap_pars_vertex>\n#include <color_pars_vertex>\n#include <morphtarget_pars_vertex>\n#include <skinning_pars_vertex>\n#include <shadowmap_pars_vertex>\n#include <specularmap_pars_fragment>\n#include <logdepthbuf_pars_vertex>\n#include <clipping_planes_pars_vertex>\nvoid main() {\n\t#include <uv_vertex>\n\t#include <uv2_vertex>\n\t#include <color_vertex>\n\t#include <beginnormal_vertex>\n\t#include <morphnormal_vertex>\n\t#include <skinbase_vertex>\n\t#include <skinnormal_vertex>\n\t#include <defaultnormal_vertex>\n#ifndef FLAT_SHADED\n\tvNormal = normalize( transformedNormal );\n#endif\n\t#include <begin_vertex>\n\t#include <displacementmap_vertex>\n\t#include <morphtarget_vertex>\n\t#include <skinning_vertex>\n\t#include <project_vertex>\n\t#include <logdepthbuf_vertex>\n\t#include <clipping_planes_vertex>\n\tvViewPosition = - mvPosition.xyz;\n\t#include <worldpos_vertex>\n\t#include <shadowmap_vertex>\n}\n";

	var normal_frag = "#define NORMAL\nuniform float opacity;\n#if defined( FLAT_SHADED  ) || defined( USE_BUMPMAP ) || defined( USE_NORMALMAP )\n\tvarying vec3 vViewPosition;\n#endif\n#ifndef FLAT_SHADED\n\tvarying vec3 vNormal;\n#endif\n#include <packing>\n#include <uv_pars_fragment>\n#include <bumpmap_pars_fragment>\n#include <normalmap_pars_fragment>\n#include <logdepthbuf_pars_fragment>\nvoid main() {\n\t#include <logdepthbuf_fragment>\n\t#include <normal_flip>\n\t#include <normal_fragment>\n\tgl_FragColor = vec4( packNormalToRGB( normal ), opacity );\n\t#include <premultiplied_alpha_fragment>\n\t#include <encodings_fragment>\n}\n";

	var normal_vert = "#define NORMAL\n#if defined( FLAT_SHADED  ) || defined( USE_BUMPMAP ) || defined( USE_NORMALMAP )\n\tvarying vec3 vViewPosition;\n#endif\n#ifndef FLAT_SHADED\n\tvarying vec3 vNormal;\n#endif\n#include <uv_pars_vertex>\n#include <displacementmap_pars_vertex>\n#include <morphtarget_pars_vertex>\n#include <skinning_pars_vertex>\n#include <logdepthbuf_pars_vertex>\nvoid main() {\n\t#include <uv_vertex>\n\t#include <beginnormal_vertex>\n\t#include <morphnormal_vertex>\n\t#include <skinbase_vertex>\n\t#include <skinnormal_vertex>\n\t#include <defaultnormal_vertex>\n#ifndef FLAT_SHADED\n\tvNormal = normalize( transformedNormal );\n#endif\n\t#include <begin_vertex>\n\t#include <displacementmap_vertex>\n\t#include <morphtarget_vertex>\n\t#include <skinning_vertex>\n\t#include <project_vertex>\n\t#include <logdepthbuf_vertex>\n#if defined( FLAT_SHADED  ) || defined( USE_BUMPMAP ) || defined( USE_NORMALMAP )\n\tvViewPosition = - mvPosition.xyz;\n#endif\n}\n";

	var points_frag = "uniform vec3 diffuse;\nuniform float opacity;\n#include <common>\n#include <packing>\n#include <color_pars_fragment>\n#include <map_particle_pars_fragment>\n#include <fog_pars_fragment>\n#include <shadowmap_pars_fragment>\n#include <logdepthbuf_pars_fragment>\n#include <clipping_planes_pars_fragment>\nvoid main() {\n\t#include <clipping_planes_fragment>\n\tvec3 outgoingLight = vec3( 0.0 );\n\tvec4 diffuseColor = vec4( diffuse, opacity );\n\t#include <logdepthbuf_fragment>\n\t#include <map_particle_fragment>\n\t#include <color_fragment>\n\t#include <alphatest_fragment>\n\toutgoingLight = diffuseColor.rgb;\n\tgl_FragColor = vec4( outgoingLight, diffuseColor.a );\n\t#include <premultiplied_alpha_fragment>\n\t#include <tonemapping_fragment>\n\t#include <encodings_fragment>\n\t#include <fog_fragment>\n}\n";

	var points_vert = "uniform float size;\nuniform float scale;\n#include <common>\n#include <color_pars_vertex>\n#include <shadowmap_pars_vertex>\n#include <logdepthbuf_pars_vertex>\n#include <clipping_planes_pars_vertex>\nvoid main() {\n\t#include <color_vertex>\n\t#include <begin_vertex>\n\t#include <project_vertex>\n\t#ifdef USE_SIZEATTENUATION\n\t\tgl_PointSize = size * ( scale / - mvPosition.z );\n\t#else\n\t\tgl_PointSize = size;\n\t#endif\n\t#include <logdepthbuf_vertex>\n\t#include <clipping_planes_vertex>\n\t#include <worldpos_vertex>\n\t#include <shadowmap_vertex>\n}\n";

	var shadow_frag = "uniform float opacity;\n#include <common>\n#include <packing>\n#include <bsdfs>\n#include <lights_pars>\n#include <shadowmap_pars_fragment>\n#include <shadowmask_pars_fragment>\nvoid main() {\n\tgl_FragColor = vec4( 0.0, 0.0, 0.0, opacity * ( 1.0  - getShadowMask() ) );\n}\n";

	var shadow_vert = "#include <shadowmap_pars_vertex>\nvoid main() {\n\t#include <begin_vertex>\n\t#include <project_vertex>\n\t#include <worldpos_vertex>\n\t#include <shadowmap_vertex>\n}\n";

	var ShaderChunk = {
		alphamap_fragment: alphamap_fragment,
		alphamap_pars_fragment: alphamap_pars_fragment,
		alphatest_fragment: alphatest_fragment,
		aomap_fragment: aomap_fragment,
		aomap_pars_fragment: aomap_pars_fragment,
		begin_vertex: begin_vertex,
		beginnormal_vertex: beginnormal_vertex,
		bsdfs: bsdfs,
		bumpmap_pars_fragment: bumpmap_pars_fragment,
		clipping_planes_fragment: clipping_planes_fragment,
		clipping_planes_pars_fragment: clipping_planes_pars_fragment,
		clipping_planes_pars_vertex: clipping_planes_pars_vertex,
		clipping_planes_vertex: clipping_planes_vertex,
		color_fragment: color_fragment,
		color_pars_fragment: color_pars_fragment,
		color_pars_vertex: color_pars_vertex,
		color_vertex: color_vertex,
		common: common,
		cube_uv_reflection_fragment: cube_uv_reflection_fragment,
		defaultnormal_vertex: defaultnormal_vertex,
		displacementmap_pars_vertex: displacementmap_pars_vertex,
		displacementmap_vertex: displacementmap_vertex,
		emissivemap_fragment: emissivemap_fragment,
		emissivemap_pars_fragment: emissivemap_pars_fragment,
		encodings_fragment: encodings_fragment,
		encodings_pars_fragment: encodings_pars_fragment,
		envmap_fragment: envmap_fragment,
		envmap_pars_fragment: envmap_pars_fragment,
		envmap_pars_vertex: envmap_pars_vertex,
		envmap_vertex: envmap_vertex,
		fog_fragment: fog_fragment,
		fog_pars_fragment: fog_pars_fragment,
		gradientmap_pars_fragment: gradientmap_pars_fragment,
		lightmap_fragment: lightmap_fragment,
		lightmap_pars_fragment: lightmap_pars_fragment,
		lights_lambert_vertex: lights_lambert_vertex,
		lights_pars: lights_pars,
		lights_phong_fragment: lights_phong_fragment,
		lights_phong_pars_fragment: lights_phong_pars_fragment,
		lights_physical_fragment: lights_physical_fragment,
		lights_physical_pars_fragment: lights_physical_pars_fragment,
		lights_template: lights_template,
		logdepthbuf_fragment: logdepthbuf_fragment,
		logdepthbuf_pars_fragment: logdepthbuf_pars_fragment,
		logdepthbuf_pars_vertex: logdepthbuf_pars_vertex,
		logdepthbuf_vertex: logdepthbuf_vertex,
		map_fragment: map_fragment,
		map_pars_fragment: map_pars_fragment,
		map_particle_fragment: map_particle_fragment,
		map_particle_pars_fragment: map_particle_pars_fragment,
		metalnessmap_fragment: metalnessmap_fragment,
		metalnessmap_pars_fragment: metalnessmap_pars_fragment,
		morphnormal_vertex: morphnormal_vertex,
		morphtarget_pars_vertex: morphtarget_pars_vertex,
		morphtarget_vertex: morphtarget_vertex,
		normal_flip: normal_flip,
		normal_fragment: normal_fragment,
		normalmap_pars_fragment: normalmap_pars_fragment,
		packing: packing,
		premultiplied_alpha_fragment: premultiplied_alpha_fragment,
		project_vertex: project_vertex,
		roughnessmap_fragment: roughnessmap_fragment,
		roughnessmap_pars_fragment: roughnessmap_pars_fragment,
		shadowmap_pars_fragment: shadowmap_pars_fragment,
		shadowmap_pars_vertex: shadowmap_pars_vertex,
		shadowmap_vertex: shadowmap_vertex,
		shadowmask_pars_fragment: shadowmask_pars_fragment,
		skinbase_vertex: skinbase_vertex,
		skinning_pars_vertex: skinning_pars_vertex,
		skinning_vertex: skinning_vertex,
		skinnormal_vertex: skinnormal_vertex,
		specularmap_fragment: specularmap_fragment,
		specularmap_pars_fragment: specularmap_pars_fragment,
		tonemapping_fragment: tonemapping_fragment,
		tonemapping_pars_fragment: tonemapping_pars_fragment,
		uv_pars_fragment: uv_pars_fragment,
		uv_pars_vertex: uv_pars_vertex,
		uv_vertex: uv_vertex,
		uv2_pars_fragment: uv2_pars_fragment,
		uv2_pars_vertex: uv2_pars_vertex,
		uv2_vertex: uv2_vertex,
		worldpos_vertex: worldpos_vertex,

		cube_frag: cube_frag,
		cube_vert: cube_vert,
		depth_frag: depth_frag,
		depth_vert: depth_vert,
		distanceRGBA_frag: distanceRGBA_frag,
		distanceRGBA_vert: distanceRGBA_vert,
		equirect_frag: equirect_frag,
		equirect_vert: equirect_vert,
		linedashed_frag: linedashed_frag,
		linedashed_vert: linedashed_vert,
		meshbasic_frag: meshbasic_frag,
		meshbasic_vert: meshbasic_vert,
		meshlambert_frag: meshlambert_frag,
		meshlambert_vert: meshlambert_vert,
		meshphong_frag: meshphong_frag,
		meshphong_vert: meshphong_vert,
		meshphysical_frag: meshphysical_frag,
		meshphysical_vert: meshphysical_vert,
		normal_frag: normal_frag,
		normal_vert: normal_vert,
		points_frag: points_frag,
		points_vert: points_vert,
		shadow_frag: shadow_frag,
		shadow_vert: shadow_vert
	};

	/**
	 * @author mrdoob / http://mrdoob.com/
	 */

	function Color( r, g, b ) {

		if ( g === undefined && b === undefined ) {

			// r is THREE.Color, hex or string
			return this.set( r );

		}

		return this.setRGB( r, g, b );

	}

	Color.prototype = {

		constructor: Color,

		isColor: true,

		r: 1, g: 1, b: 1,

		set: function ( value ) {

			if ( value && value.isColor ) {

				this.copy( value );

			} else if ( typeof value === 'number' ) {

				this.setHex( value );

			} else if ( typeof value === 'string' ) {

				this.setStyle( value );

			}

			return this;

		},

		setScalar: function ( scalar ) {

			this.r = scalar;
			this.g = scalar;
			this.b = scalar;

			return this;

		},

		setHex: function ( hex ) {

			hex = Math.floor( hex );

			this.r = ( hex >> 16 & 255 ) / 255;
			this.g = ( hex >> 8 & 255 ) / 255;
			this.b = ( hex & 255 ) / 255;

			return this;

		},

		setRGB: function ( r, g, b ) {

			this.r = r;
			this.g = g;
			this.b = b;

			return this;

		},

		setHSL: function () {

			function hue2rgb( p, q, t ) {

				if ( t < 0 ) t += 1;
				if ( t > 1 ) t -= 1;
				if ( t < 1 / 6 ) return p + ( q - p ) * 6 * t;
				if ( t < 1 / 2 ) return q;
				if ( t < 2 / 3 ) return p + ( q - p ) * 6 * ( 2 / 3 - t );
				return p;

			}

			return function setHSL( h, s, l ) {

				// h,s,l ranges are in 0.0 - 1.0
				h = _Math.euclideanModulo( h, 1 );
				s = _Math.clamp( s, 0, 1 );
				l = _Math.clamp( l, 0, 1 );

				if ( s === 0 ) {

					this.r = this.g = this.b = l;

				} else {

					var p = l <= 0.5 ? l * ( 1 + s ) : l + s - ( l * s );
					var q = ( 2 * l ) - p;

					this.r = hue2rgb( q, p, h + 1 / 3 );
					this.g = hue2rgb( q, p, h );
					this.b = hue2rgb( q, p, h - 1 / 3 );

				}

				return this;

			};

		}(),

		setStyle: function ( style ) {

			function handleAlpha( string ) {

				if ( string === undefined ) return;

				if ( parseFloat( string ) < 1 ) {

					console.warn( 'THREE.Color: Alpha component of ' + style + ' will be ignored.' );

				}

			}


			var m;

			if ( m = /^((?:rgb|hsl)a?)\(\s*([^\)]*)\)/.exec( style ) ) {

				// rgb / hsl

				var color;
				var name = m[ 1 ];
				var components = m[ 2 ];

				switch ( name ) {

					case 'rgb':
					case 'rgba':

						if ( color = /^(\d+)\s*,\s*(\d+)\s*,\s*(\d+)\s*(,\s*([0-9]*\.?[0-9]+)\s*)?$/.exec( components ) ) {

							// rgb(255,0,0) rgba(255,0,0,0.5)
							this.r = Math.min( 255, parseInt( color[ 1 ], 10 ) ) / 255;
							this.g = Math.min( 255, parseInt( color[ 2 ], 10 ) ) / 255;
							this.b = Math.min( 255, parseInt( color[ 3 ], 10 ) ) / 255;

							handleAlpha( color[ 5 ] );

							return this;

						}

						if ( color = /^(\d+)\%\s*,\s*(\d+)\%\s*,\s*(\d+)\%\s*(,\s*([0-9]*\.?[0-9]+)\s*)?$/.exec( components ) ) {

							// rgb(100%,0%,0%) rgba(100%,0%,0%,0.5)
							this.r = Math.min( 100, parseInt( color[ 1 ], 10 ) ) / 100;
							this.g = Math.min( 100, parseInt( color[ 2 ], 10 ) ) / 100;
							this.b = Math.min( 100, parseInt( color[ 3 ], 10 ) ) / 100;

							handleAlpha( color[ 5 ] );

							return this;

						}

						break;

					case 'hsl':
					case 'hsla':

						if ( color = /^([0-9]*\.?[0-9]+)\s*,\s*(\d+)\%\s*,\s*(\d+)\%\s*(,\s*([0-9]*\.?[0-9]+)\s*)?$/.exec( components ) ) {

							// hsl(120,50%,50%) hsla(120,50%,50%,0.5)
							var h = parseFloat( color[ 1 ] ) / 360;
							var s = parseInt( color[ 2 ], 10 ) / 100;
							var l = parseInt( color[ 3 ], 10 ) / 100;

							handleAlpha( color[ 5 ] );

							return this.setHSL( h, s, l );

						}

						break;

				}

			} else if ( m = /^\#([A-Fa-f0-9]+)$/.exec( style ) ) {

				// hex color

				var hex = m[ 1 ];
				var size = hex.length;

				if ( size === 3 ) {

					// #ff0
					this.r = parseInt( hex.charAt( 0 ) + hex.charAt( 0 ), 16 ) / 255;
					this.g = parseInt( hex.charAt( 1 ) + hex.charAt( 1 ), 16 ) / 255;
					this.b = parseInt( hex.charAt( 2 ) + hex.charAt( 2 ), 16 ) / 255;

					return this;

				} else if ( size === 6 ) {

					// #ff0000
					this.r = parseInt( hex.charAt( 0 ) + hex.charAt( 1 ), 16 ) / 255;
					this.g = parseInt( hex.charAt( 2 ) + hex.charAt( 3 ), 16 ) / 255;
					this.b = parseInt( hex.charAt( 4 ) + hex.charAt( 5 ), 16 ) / 255;

					return this;

				}

			}

			if ( style && style.length > 0 ) {

				// color keywords
				var hex = ColorKeywords[ style ];

				if ( hex !== undefined ) {

					// red
					this.setHex( hex );

				} else {

					// unknown color
					console.warn( 'THREE.Color: Unknown color ' + style );

				}

			}

			return this;

		},

		clone: function () {

			return new this.constructor( this.r, this.g, this.b );

		},

		copy: function ( color ) {

			this.r = color.r;
			this.g = color.g;
			this.b = color.b;

			return this;

		},

		copyGammaToLinear: function ( color, gammaFactor ) {

			if ( gammaFactor === undefined ) gammaFactor = 2.0;

			this.r = Math.pow( color.r, gammaFactor );
			this.g = Math.pow( color.g, gammaFactor );
			this.b = Math.pow( color.b, gammaFactor );

			return this;

		},

		copyLinearToGamma: function ( color, gammaFactor ) {

			if ( gammaFactor === undefined ) gammaFactor = 2.0;

			var safeInverse = ( gammaFactor > 0 ) ? ( 1.0 / gammaFactor ) : 1.0;

			this.r = Math.pow( color.r, safeInverse );
			this.g = Math.pow( color.g, safeInverse );
			this.b = Math.pow( color.b, safeInverse );

			return this;

		},

		convertGammaToLinear: function () {

			var r = this.r, g = this.g, b = this.b;

			this.r = r * r;
			this.g = g * g;
			this.b = b * b;

			return this;

		},

		convertLinearToGamma: function () {

			this.r = Math.sqrt( this.r );
			this.g = Math.sqrt( this.g );
			this.b = Math.sqrt( this.b );

			return this;

		},

		getHex: function () {

			return ( this.r * 255 ) << 16 ^ ( this.g * 255 ) << 8 ^ ( this.b * 255 ) << 0;

		},

		getHexString: function () {

			return ( '000000' + this.getHex().toString( 16 ) ).slice( - 6 );

		},

		getHSL: function ( optionalTarget ) {

			// h,s,l ranges are in 0.0 - 1.0

			var hsl = optionalTarget || { h: 0, s: 0, l: 0 };

			var r = this.r, g = this.g, b = this.b;

			var max = Math.max( r, g, b );
			var min = Math.min( r, g, b );

			var hue, saturation;
			var lightness = ( min + max ) / 2.0;

			if ( min === max ) {

				hue = 0;
				saturation = 0;

			} else {

				var delta = max - min;

				saturation = lightness <= 0.5 ? delta / ( max + min ) : delta / ( 2 - max - min );

				switch ( max ) {

					case r: hue = ( g - b ) / delta + ( g < b ? 6 : 0 ); break;
					case g: hue = ( b - r ) / delta + 2; break;
					case b: hue = ( r - g ) / delta + 4; break;

				}

				hue /= 6;

			}

			hsl.h = hue;
			hsl.s = saturation;
			hsl.l = lightness;

			return hsl;

		},

		getStyle: function () {

			return 'rgb(' + ( ( this.r * 255 ) | 0 ) + ',' + ( ( this.g * 255 ) | 0 ) + ',' + ( ( this.b * 255 ) | 0 ) + ')';

		},

		offsetHSL: function ( h, s, l ) {

			var hsl = this.getHSL();

			hsl.h += h; hsl.s += s; hsl.l += l;

			this.setHSL( hsl.h, hsl.s, hsl.l );

			return this;

		},

		add: function ( color ) {

			this.r += color.r;
			this.g += color.g;
			this.b += color.b;

			return this;

		},

		addColors: function ( color1, color2 ) {

			this.r = color1.r + color2.r;
			this.g = color1.g + color2.g;
			this.b = color1.b + color2.b;

			return this;

		},

		addScalar: function ( s ) {

			this.r += s;
			this.g += s;
			this.b += s;

			return this;

		},

		sub: function( color ) {

			this.r = Math.max( 0, this.r - color.r );
			this.g = Math.max( 0, this.g - color.g );
			this.b = Math.max( 0, this.b - color.b );

			return this;

		},

		multiply: function ( color ) {

			this.r *= color.r;
			this.g *= color.g;
			this.b *= color.b;

			return this;

		},

		multiplyScalar: function ( s ) {

			this.r *= s;
			this.g *= s;
			this.b *= s;

			return this;

		},

		lerp: function ( color, alpha ) {

			this.r += ( color.r - this.r ) * alpha;
			this.g += ( color.g - this.g ) * alpha;
			this.b += ( color.b - this.b ) * alpha;

			return this;

		},

		equals: function ( c ) {

			return ( c.r === this.r ) && ( c.g === this.g ) && ( c.b === this.b );

		},

		fromArray: function ( array, offset ) {

			if ( offset === undefined ) offset = 0;

			this.r = array[ offset ];
			this.g = array[ offset + 1 ];
			this.b = array[ offset + 2 ];

			return this;

		},

		toArray: function ( array, offset ) {

			if ( array === undefined ) array = [];
			if ( offset === undefined ) offset = 0;

			array[ offset ] = this.r;
			array[ offset + 1 ] = this.g;
			array[ offset + 2 ] = this.b;

			return array;

		},

		toJSON: function () {

			return this.getHex();

		}

	};

	var ColorKeywords = { 'aliceblue': 0xF0F8FF, 'antiquewhite': 0xFAEBD7, 'aqua': 0x00FFFF, 'aquamarine': 0x7FFFD4, 'azure': 0xF0FFFF,
	'beige': 0xF5F5DC, 'bisque': 0xFFE4C4, 'black': 0x000000, 'blanchedalmond': 0xFFEBCD, 'blue': 0x0000FF, 'blueviolet': 0x8A2BE2,
	'brown': 0xA52A2A, 'burlywood': 0xDEB887, 'cadetblue': 0x5F9EA0, 'chartreuse': 0x7FFF00, 'chocolate': 0xD2691E, 'coral': 0xFF7F50,
	'cornflowerblue': 0x6495ED, 'cornsilk': 0xFFF8DC, 'crimson': 0xDC143C, 'cyan': 0x00FFFF, 'darkblue': 0x00008B, 'darkcyan': 0x008B8B,
	'darkgoldenrod': 0xB8860B, 'darkgray': 0xA9A9A9, 'darkgreen': 0x006400, 'darkgrey': 0xA9A9A9, 'darkkhaki': 0xBDB76B, 'darkmagenta': 0x8B008B,
	'darkolivegreen': 0x556B2F, 'darkorange': 0xFF8C00, 'darkorchid': 0x9932CC, 'darkred': 0x8B0000, 'darksalmon': 0xE9967A, 'darkseagreen': 0x8FBC8F,
	'darkslateblue': 0x483D8B, 'darkslategray': 0x2F4F4F, 'darkslategrey': 0x2F4F4F, 'darkturquoise': 0x00CED1, 'darkviolet': 0x9400D3,
	'deeppink': 0xFF1493, 'deepskyblue': 0x00BFFF, 'dimgray': 0x696969, 'dimgrey': 0x696969, 'dodgerblue': 0x1E90FF, 'firebrick': 0xB22222,
	'floralwhite': 0xFFFAF0, 'forestgreen': 0x228B22, 'fuchsia': 0xFF00FF, 'gainsboro': 0xDCDCDC, 'ghostwhite': 0xF8F8FF, 'gold': 0xFFD700,
	'goldenrod': 0xDAA520, 'gray': 0x808080, 'green': 0x008000, 'greenyellow': 0xADFF2F, 'grey': 0x808080, 'honeydew': 0xF0FFF0, 'hotpink': 0xFF69B4,
	'indianred': 0xCD5C5C, 'indigo': 0x4B0082, 'ivory': 0xFFFFF0, 'khaki': 0xF0E68C, 'lavender': 0xE6E6FA, 'lavenderblush': 0xFFF0F5, 'lawngreen': 0x7CFC00,
	'lemonchiffon': 0xFFFACD, 'lightblue': 0xADD8E6, 'lightcoral': 0xF08080, 'lightcyan': 0xE0FFFF, 'lightgoldenrodyellow': 0xFAFAD2, 'lightgray': 0xD3D3D3,
	'lightgreen': 0x90EE90, 'lightgrey': 0xD3D3D3, 'lightpink': 0xFFB6C1, 'lightsalmon': 0xFFA07A, 'lightseagreen': 0x20B2AA, 'lightskyblue': 0x87CEFA,
	'lightslategray': 0x778899, 'lightslategrey': 0x778899, 'lightsteelblue': 0xB0C4DE, 'lightyellow': 0xFFFFE0, 'lime': 0x00FF00, 'limegreen': 0x32CD32,
	'linen': 0xFAF0E6, 'magenta': 0xFF00FF, 'maroon': 0x800000, 'mediumaquamarine': 0x66CDAA, 'mediumblue': 0x0000CD, 'mediumorchid': 0xBA55D3,
	'mediumpurple': 0x9370DB, 'mediumseagreen': 0x3CB371, 'mediumslateblue': 0x7B68EE, 'mediumspringgreen': 0x00FA9A, 'mediumturquoise': 0x48D1CC,
	'mediumvioletred': 0xC71585, 'midnightblue': 0x191970, 'mintcream': 0xF5FFFA, 'mistyrose': 0xFFE4E1, 'moccasin': 0xFFE4B5, 'navajowhite': 0xFFDEAD,
	'navy': 0x000080, 'oldlace': 0xFDF5E6, 'olive': 0x808000, 'olivedrab': 0x6B8E23, 'orange': 0xFFA500, 'orangered': 0xFF4500, 'orchid': 0xDA70D6,
	'palegoldenrod': 0xEEE8AA, 'palegreen': 0x98FB98, 'paleturquoise': 0xAFEEEE, 'palevioletred': 0xDB7093, 'papayawhip': 0xFFEFD5, 'peachpuff': 0xFFDAB9,
	'peru': 0xCD853F, 'pink': 0xFFC0CB, 'plum': 0xDDA0DD, 'powderblue': 0xB0E0E6, 'purple': 0x800080, 'red': 0xFF0000, 'rosybrown': 0xBC8F8F,
	'royalblue': 0x4169E1, 'saddlebrown': 0x8B4513, 'salmon': 0xFA8072, 'sandybrown': 0xF4A460, 'seagreen': 0x2E8B57, 'seashell': 0xFFF5EE,
	'sienna': 0xA0522D, 'silver': 0xC0C0C0, 'skyblue': 0x87CEEB, 'slateblue': 0x6A5ACD, 'slategray': 0x708090, 'slategrey': 0x708090, 'snow': 0xFFFAFA,
	'springgreen': 0x00FF7F, 'steelblue': 0x4682B4, 'tan': 0xD2B48C, 'teal': 0x008080, 'thistle': 0xD8BFD8, 'tomato': 0xFF6347, 'turquoise': 0x40E0D0,
	'violet': 0xEE82EE, 'wheat': 0xF5DEB3, 'white': 0xFFFFFF, 'whitesmoke': 0xF5F5F5, 'yellow': 0xFFFF00, 'yellowgreen': 0x9ACD32 };

	/**
	 * @author alteredq / http://alteredqualia.com/
	 */

	function DataTexture( data, width, height, format, type, mapping, wrapS, wrapT, magFilter, minFilter, anisotropy, encoding ) {

		Texture.call( this, null, mapping, wrapS, wrapT, magFilter, minFilter, format, type, anisotropy, encoding );

		this.image = { data: data, width: width, height: height };

		this.magFilter = magFilter !== undefined ? magFilter : NearestFilter;
		this.minFilter = minFilter !== undefined ? minFilter : NearestFilter;

		this.generateMipmaps  = false;
		this.flipY = false;
		this.unpackAlignment = 1;

	}

	DataTexture.prototype = Object.create( Texture.prototype );
	DataTexture.prototype.constructor = DataTexture;

	DataTexture.prototype.isDataTexture = true;

	/**
	 * Uniforms library for shared webgl shaders
	 */

	var UniformsLib = {

		common: {

			diffuse: { value: new Color( 0xeeeeee ) },
			opacity: { value: 1.0 },

			map: { value: null },
			offsetRepeat: { value: new Vector4( 0, 0, 1, 1 ) },

			specularMap: { value: null },
			alphaMap: { value: null },

			envMap: { value: null },
			flipEnvMap: { value: - 1 },
			reflectivity: { value: 1.0 },
			refractionRatio: { value: 0.98 }

		},

		aomap: {

			aoMap: { value: null },
			aoMapIntensity: { value: 1 }

		},

		lightmap: {

			lightMap: { value: null },
			lightMapIntensity: { value: 1 }

		},

		emissivemap: {

			emissiveMap: { value: null }

		},

		bumpmap: {

			bumpMap: { value: null },
			bumpScale: { value: 1 }

		},

		normalmap: {

			normalMap: { value: null },
			normalScale: { value: new Vector2( 1, 1 ) }

		},

		displacementmap: {

			displacementMap: { value: null },
			displacementScale: { value: 1 },
			displacementBias: { value: 0 }

		},

		roughnessmap: {

			roughnessMap: { value: null }

		},

		metalnessmap: {

			metalnessMap: { value: null }

		},

		gradientmap: {

			gradientMap: { value: null }

		},

		fog: {

			fogDensity: { value: 0.00025 },
			fogNear: { value: 1 },
			fogFar: { value: 2000 },
			fogColor: { value: new Color( 0xffffff ) }

		},

		lights: {

			ambientLightColor: { value: [] },

			directionalLights: { value: [], properties: {
				direction: {},
				color: {},

				shadow: {},
				shadowBias: {},
				shadowRadius: {},
				shadowMapSize: {}
			} },

			directionalShadowMap: { value: [] },
			directionalShadowMatrix: { value: [] },

			spotLights: { value: [], properties: {
				color: {},
				position: {},
				direction: {},
				distance: {},
				coneCos: {},
				penumbraCos: {},
				decay: {},

				shadow: {},
				shadowBias: {},
				shadowRadius: {},
				shadowMapSize: {}
			} },

			spotShadowMap: { value: [] },
			spotShadowMatrix: { value: [] },

			pointLights: { value: [], properties: {
				color: {},
				position: {},
				decay: {},
				distance: {},

				shadow: {},
				shadowBias: {},
				shadowRadius: {},
				shadowMapSize: {}
			} },

			pointShadowMap: { value: [] },
			pointShadowMatrix: { value: [] },

			hemisphereLights: { value: [], properties: {
				direction: {},
				skyColor: {},
				groundColor: {}
			} },

	        // TODO (abelnation): RectAreaLight BRDF data needs to be moved from example to main src
	        rectAreaLights: { value: [], properties: {
	            color: {},
	            position: {},
	            width: {},
	            height: {},
	        } }

		},

		points: {

			diffuse: { value: new Color( 0xeeeeee ) },
			opacity: { value: 1.0 },
			size: { value: 1.0 },
			scale: { value: 1.0 },
			map: { value: null },
			offsetRepeat: { value: new Vector4( 0, 0, 1, 1 ) }

		}

	};

	/**
	 * @author alteredq / http://alteredqualia.com/
	 * @author mrdoob / http://mrdoob.com/
	 * @author mikael emtinger / http://gomo.se/
	 */

	var ShaderLib = {

		basic: {

			uniforms: UniformsUtils.merge( [
				UniformsLib.common,
				UniformsLib.aomap,
				UniformsLib.lightmap,
				UniformsLib.fog
			] ),

			vertexShader: ShaderChunk.meshbasic_vert,
			fragmentShader: ShaderChunk.meshbasic_frag

		},

		lambert: {

			uniforms: UniformsUtils.merge( [
				UniformsLib.common,
				UniformsLib.aomap,
				UniformsLib.lightmap,
				UniformsLib.emissivemap,
				UniformsLib.fog,
				UniformsLib.lights,
				{
					emissive: { value: new Color( 0x000000 ) }
				}
			] ),

			vertexShader: ShaderChunk.meshlambert_vert,
			fragmentShader: ShaderChunk.meshlambert_frag

		},

		phong: {

			uniforms: UniformsUtils.merge( [
				UniformsLib.common,
				UniformsLib.aomap,
				UniformsLib.lightmap,
				UniformsLib.emissivemap,
				UniformsLib.bumpmap,
				UniformsLib.normalmap,
				UniformsLib.displacementmap,
				UniformsLib.gradientmap,
				UniformsLib.fog,
				UniformsLib.lights,
				{
					emissive: { value: new Color( 0x000000 ) },
					specular: { value: new Color( 0x111111 ) },
					shininess: { value: 30 }
				}
			] ),

			vertexShader: ShaderChunk.meshphong_vert,
			fragmentShader: ShaderChunk.meshphong_frag

		},

		standard: {

			uniforms: UniformsUtils.merge( [
				UniformsLib.common,
				UniformsLib.aomap,
				UniformsLib.lightmap,
				UniformsLib.emissivemap,
				UniformsLib.bumpmap,
				UniformsLib.normalmap,
				UniformsLib.displacementmap,
				UniformsLib.roughnessmap,
				UniformsLib.metalnessmap,
				UniformsLib.fog,
				UniformsLib.lights,
				{
					emissive: { value: new Color( 0x000000 ) },
					roughness: { value: 0.5 },
					metalness: { value: 0 },
					envMapIntensity: { value: 1 } // temporary
				}
			] ),

			vertexShader: ShaderChunk.meshphysical_vert,
			fragmentShader: ShaderChunk.meshphysical_frag

		},

		points: {

			uniforms: UniformsUtils.merge( [
				UniformsLib.points,
				UniformsLib.fog
			] ),

			vertexShader: ShaderChunk.points_vert,
			fragmentShader: ShaderChunk.points_frag

		},

		dashed: {

			uniforms: UniformsUtils.merge( [
				UniformsLib.common,
				UniformsLib.fog,
				{
					scale: { value: 1 },
					dashSize: { value: 1 },
					totalSize: { value: 2 }
				}
			] ),

			vertexShader: ShaderChunk.linedashed_vert,
			fragmentShader: ShaderChunk.linedashed_frag

		},

		depth: {

			uniforms: UniformsUtils.merge( [
				UniformsLib.common,
				UniformsLib.displacementmap
			] ),

			vertexShader: ShaderChunk.depth_vert,
			fragmentShader: ShaderChunk.depth_frag

		},

		normal: {

			uniforms: UniformsUtils.merge( [
				UniformsLib.common,
				UniformsLib.bumpmap,
				UniformsLib.normalmap,
				UniformsLib.displacementmap,
				{
					opacity: { value: 1.0 }
				}
			] ),

			vertexShader: ShaderChunk.normal_vert,
			fragmentShader: ShaderChunk.normal_frag

		},

		/* -------------------------------------------------------------------------
		//	Cube map shader
		 ------------------------------------------------------------------------- */

		cube: {

			uniforms: {
				tCube: { value: null },
				tFlip: { value: - 1 },
				opacity: { value: 1.0 }
			},

			vertexShader: ShaderChunk.cube_vert,
			fragmentShader: ShaderChunk.cube_frag

		},

		/* -------------------------------------------------------------------------
		//	Cube map shader
		 ------------------------------------------------------------------------- */

		equirect: {

			uniforms: {
				tEquirect: { value: null },
				tFlip: { value: - 1 }
			},

			vertexShader: ShaderChunk.equirect_vert,
			fragmentShader: ShaderChunk.equirect_frag

		},

		distanceRGBA: {

			uniforms: {
				lightPos: { value: new Vector3() }
			},

			vertexShader: ShaderChunk.distanceRGBA_vert,
			fragmentShader: ShaderChunk.distanceRGBA_frag

		}

	};

	ShaderLib.physical = {

		uniforms: UniformsUtils.merge( [
			ShaderLib.standard.uniforms,
			{
				clearCoat: { value: 0 },
				clearCoatRoughness: { value: 0 }
			}
		] ),

		vertexShader: ShaderChunk.meshphysical_vert,
		fragmentShader: ShaderChunk.meshphysical_frag

	};

	/**
	 * @author bhouston / http://clara.io
	 */

	function Box2( min, max ) {

		this.min = ( min !== undefined ) ? min : new Vector2( + Infinity, + Infinity );
		this.max = ( max !== undefined ) ? max : new Vector2( - Infinity, - Infinity );

	}

	Box2.prototype = {

		constructor: Box2,

		set: function ( min, max ) {

			this.min.copy( min );
			this.max.copy( max );

			return this;

		},

		setFromPoints: function ( points ) {

			this.makeEmpty();

			for ( var i = 0, il = points.length; i < il; i ++ ) {

				this.expandByPoint( points[ i ] );

			}

			return this;

		},

		setFromCenterAndSize: function () {

			var v1 = new Vector2();

			return function setFromCenterAndSize( center, size ) {

				var halfSize = v1.copy( size ).multiplyScalar( 0.5 );
				this.min.copy( center ).sub( halfSize );
				this.max.copy( center ).add( halfSize );

				return this;

			};

		}(),

		clone: function () {

			return new this.constructor().copy( this );

		},

		copy: function ( box ) {

			this.min.copy( box.min );
			this.max.copy( box.max );

			return this;

		},

		makeEmpty: function () {

			this.min.x = this.min.y = + Infinity;
			this.max.x = this.max.y = - Infinity;

			return this;

		},

		isEmpty: function () {

			// this is a more robust check for empty than ( volume <= 0 ) because volume can get positive with two negative axes

			return ( this.max.x < this.min.x ) || ( this.max.y < this.min.y );

		},

		getCenter: function ( optionalTarget ) {

			var result = optionalTarget || new Vector2();
			return this.isEmpty() ? result.set( 0, 0 ) : result.addVectors( this.min, this.max ).multiplyScalar( 0.5 );

		},

		getSize: function ( optionalTarget ) {

			var result = optionalTarget || new Vector2();
			return this.isEmpty() ? result.set( 0, 0 ) : result.subVectors( this.max, this.min );

		},

		expandByPoint: function ( point ) {

			this.min.min( point );
			this.max.max( point );

			return this;

		},

		expandByVector: function ( vector ) {

			this.min.sub( vector );
			this.max.add( vector );

			return this;

		},

		expandByScalar: function ( scalar ) {

			this.min.addScalar( - scalar );
			this.max.addScalar( scalar );

			return this;

		},

		containsPoint: function ( point ) {

			return point.x < this.min.x || point.x > this.max.x ||
				point.y < this.min.y || point.y > this.max.y ? false : true;

		},

		containsBox: function ( box ) {

			return this.min.x <= box.min.x && box.max.x <= this.max.x &&
				this.min.y <= box.min.y && box.max.y <= this.max.y;

		},

		getParameter: function ( point, optionalTarget ) {

			// This can potentially have a divide by zero if the box
			// has a size dimension of 0.

			var result = optionalTarget || new Vector2();

			return result.set(
				( point.x - this.min.x ) / ( this.max.x - this.min.x ),
				( point.y - this.min.y ) / ( this.max.y - this.min.y )
			);

		},

		intersectsBox: function ( box ) {

			// using 6 splitting planes to rule out intersections.
			return box.max.x < this.min.x || box.min.x > this.max.x ||
				box.max.y < this.min.y || box.min.y > this.max.y ? false : true;

		},

		clampPoint: function ( point, optionalTarget ) {

			var result = optionalTarget || new Vector2();
			return result.copy( point ).clamp( this.min, this.max );

		},

		distanceToPoint: function () {

			var v1 = new Vector2();

			return function distanceToPoint( point ) {

				var clampedPoint = v1.copy( point ).clamp( this.min, this.max );
				return clampedPoint.sub( point ).length();

			};

		}(),

		intersect: function ( box ) {

			this.min.max( box.min );
			this.max.min( box.max );

			return this;

		},

		union: function ( box ) {

			this.min.min( box.min );
			this.max.max( box.max );

			return this;

		},

		translate: function ( offset ) {

			this.min.add( offset );
			this.max.add( offset );

			return this;

		},

		equals: function ( box ) {

			return box.min.equals( this.min ) && box.max.equals( this.max );

		}

	};

	/**
	 * @author mikael emtinger / http://gomo.se/
	 * @author alteredq / http://alteredqualia.com/
	 */

	function LensFlarePlugin( renderer, flares ) {

		var gl = renderer.context;
		var state = renderer.state;

		var vertexBuffer, elementBuffer;
		var shader, program, attributes, uniforms;

		var tempTexture, occlusionTexture;

		function init() {

			var vertices = new Float32Array( [
				- 1, - 1,  0, 0,
				 1, - 1,  1, 0,
				 1,  1,  1, 1,
				- 1,  1,  0, 1
			] );

			var faces = new Uint16Array( [
				0, 1, 2,
				0, 2, 3
			] );

			// buffers

			vertexBuffer     = gl.createBuffer();
			elementBuffer    = gl.createBuffer();

			gl.bindBuffer( gl.ARRAY_BUFFER, vertexBuffer );
			gl.bufferData( gl.ARRAY_BUFFER, vertices, gl.STATIC_DRAW );

			gl.bindBuffer( gl.ELEMENT_ARRAY_BUFFER, elementBuffer );
			gl.bufferData( gl.ELEMENT_ARRAY_BUFFER, faces, gl.STATIC_DRAW );

			// textures

			tempTexture      = gl.createTexture();
			occlusionTexture = gl.createTexture();

			state.bindTexture( gl.TEXTURE_2D, tempTexture );
			gl.texImage2D( gl.TEXTURE_2D, 0, gl.RGB, 16, 16, 0, gl.RGB, gl.UNSIGNED_BYTE, null );
			gl.texParameteri( gl.TEXTURE_2D, gl.TEXTURE_WRAP_S, gl.CLAMP_TO_EDGE );
			gl.texParameteri( gl.TEXTURE_2D, gl.TEXTURE_WRAP_T, gl.CLAMP_TO_EDGE );
			gl.texParameteri( gl.TEXTURE_2D, gl.TEXTURE_MAG_FILTER, gl.NEAREST );
			gl.texParameteri( gl.TEXTURE_2D, gl.TEXTURE_MIN_FILTER, gl.NEAREST );

			state.bindTexture( gl.TEXTURE_2D, occlusionTexture );
			gl.texImage2D( gl.TEXTURE_2D, 0, gl.RGBA, 16, 16, 0, gl.RGBA, gl.UNSIGNED_BYTE, null );
			gl.texParameteri( gl.TEXTURE_2D, gl.TEXTURE_WRAP_S, gl.CLAMP_TO_EDGE );
			gl.texParameteri( gl.TEXTURE_2D, gl.TEXTURE_WRAP_T, gl.CLAMP_TO_EDGE );
			gl.texParameteri( gl.TEXTURE_2D, gl.TEXTURE_MAG_FILTER, gl.NEAREST );
			gl.texParameteri( gl.TEXTURE_2D, gl.TEXTURE_MIN_FILTER, gl.NEAREST );

			shader = {

				vertexShader: [

					"uniform lowp int renderType;",

					"uniform vec3 screenPosition;",
					"uniform vec2 scale;",
					"uniform float rotation;",

					"uniform sampler2D occlusionMap;",

					"attribute vec2 position;",
					"attribute vec2 uv;",

					"varying vec2 vUV;",
					"varying float vVisibility;",

					"void main() {",

						"vUV = uv;",

						"vec2 pos = position;",

						"if ( renderType == 2 ) {",

							"vec4 visibility = texture2D( occlusionMap, vec2( 0.1, 0.1 ) );",
							"visibility += texture2D( occlusionMap, vec2( 0.5, 0.1 ) );",
							"visibility += texture2D( occlusionMap, vec2( 0.9, 0.1 ) );",
							"visibility += texture2D( occlusionMap, vec2( 0.9, 0.5 ) );",
							"visibility += texture2D( occlusionMap, vec2( 0.9, 0.9 ) );",
							"visibility += texture2D( occlusionMap, vec2( 0.5, 0.9 ) );",
							"visibility += texture2D( occlusionMap, vec2( 0.1, 0.9 ) );",
							"visibility += texture2D( occlusionMap, vec2( 0.1, 0.5 ) );",
							"visibility += texture2D( occlusionMap, vec2( 0.5, 0.5 ) );",

							"vVisibility =        visibility.r / 9.0;",
							"vVisibility *= 1.0 - visibility.g / 9.0;",
							"vVisibility *=       visibility.b / 9.0;",
							"vVisibility *= 1.0 - visibility.a / 9.0;",

							"pos.x = cos( rotation ) * position.x - sin( rotation ) * position.y;",
							"pos.y = sin( rotation ) * position.x + cos( rotation ) * position.y;",

						"}",

						"gl_Position = vec4( ( pos * scale + screenPosition.xy ).xy, screenPosition.z, 1.0 );",

					"}"

				].join( "\n" ),

				fragmentShader: [

					"uniform lowp int renderType;",

					"uniform sampler2D map;",
					"uniform float opacity;",
					"uniform vec3 color;",

					"varying vec2 vUV;",
					"varying float vVisibility;",

					"void main() {",

						// pink square

						"if ( renderType == 0 ) {",

							"gl_FragColor = vec4( 1.0, 0.0, 1.0, 0.0 );",

						// restore

						"} else if ( renderType == 1 ) {",

							"gl_FragColor = texture2D( map, vUV );",

						// flare

						"} else {",

							"vec4 texture = texture2D( map, vUV );",
							"texture.a *= opacity * vVisibility;",
							"gl_FragColor = texture;",
							"gl_FragColor.rgb *= color;",

						"}",

					"}"

				].join( "\n" )

			};

			program = createProgram( shader );

			attributes = {
				vertex: gl.getAttribLocation ( program, "position" ),
				uv:     gl.getAttribLocation ( program, "uv" )
			};

			uniforms = {
				renderType:     gl.getUniformLocation( program, "renderType" ),
				map:            gl.getUniformLocation( program, "map" ),
				occlusionMap:   gl.getUniformLocation( program, "occlusionMap" ),
				opacity:        gl.getUniformLocation( program, "opacity" ),
				color:          gl.getUniformLocation( program, "color" ),
				scale:          gl.getUniformLocation( program, "scale" ),
				rotation:       gl.getUniformLocation( program, "rotation" ),
				screenPosition: gl.getUniformLocation( program, "screenPosition" )
			};

		}

		/*
		 * Render lens flares
		 * Method: renders 16x16 0xff00ff-colored points scattered over the light source area,
		 *         reads these back and calculates occlusion.
		 */

		this.render = function ( scene, camera, viewport ) {

			if ( flares.length === 0 ) return;

			var tempPosition = new Vector3();

			var invAspect = viewport.w / viewport.z,
				halfViewportWidth = viewport.z * 0.5,
				halfViewportHeight = viewport.w * 0.5;

			var size = 16 / viewport.w,
				scale = new Vector2( size * invAspect, size );

			var screenPosition = new Vector3( 1, 1, 0 ),
				screenPositionPixels = new Vector2( 1, 1 );

			var validArea = new Box2();

			validArea.min.set( viewport.x, viewport.y );
			validArea.max.set( viewport.x + ( viewport.z - 16 ), viewport.y + ( viewport.w - 16 ) );

			if ( program === undefined ) {

				init();

			}

			gl.useProgram( program );

			state.initAttributes();
			state.enableAttribute( attributes.vertex );
			state.enableAttribute( attributes.uv );
			state.disableUnusedAttributes();

			// loop through all lens flares to update their occlusion and positions
			// setup gl and common used attribs/uniforms

			gl.uniform1i( uniforms.occlusionMap, 0 );
			gl.uniform1i( uniforms.map, 1 );

			gl.bindBuffer( gl.ARRAY_BUFFER, vertexBuffer );
			gl.vertexAttribPointer( attributes.vertex, 2, gl.FLOAT, false, 2 * 8, 0 );
			gl.vertexAttribPointer( attributes.uv, 2, gl.FLOAT, false, 2 * 8, 8 );

			gl.bindBuffer( gl.ELEMENT_ARRAY_BUFFER, elementBuffer );

			state.disable( gl.CULL_FACE );
			state.setDepthWrite( false );

			for ( var i = 0, l = flares.length; i < l; i ++ ) {

				size = 16 / viewport.w;
				scale.set( size * invAspect, size );

				// calc object screen position

				var flare = flares[ i ];

				tempPosition.set( flare.matrixWorld.elements[ 12 ], flare.matrixWorld.elements[ 13 ], flare.matrixWorld.elements[ 14 ] );

				tempPosition.applyMatrix4( camera.matrixWorldInverse );
				tempPosition.applyProjection( camera.projectionMatrix );

				// setup arrays for gl programs

				screenPosition.copy( tempPosition );

				// horizontal and vertical coordinate of the lower left corner of the pixels to copy

				screenPositionPixels.x = viewport.x + ( screenPosition.x * halfViewportWidth ) + halfViewportWidth - 8;
				screenPositionPixels.y = viewport.y + ( screenPosition.y * halfViewportHeight ) + halfViewportHeight - 8;

				// screen cull

				if ( validArea.containsPoint( screenPositionPixels ) === true ) {

					// save current RGB to temp texture

					state.activeTexture( gl.TEXTURE0 );
					state.bindTexture( gl.TEXTURE_2D, null );
					state.activeTexture( gl.TEXTURE1 );
					state.bindTexture( gl.TEXTURE_2D, tempTexture );
					gl.copyTexImage2D( gl.TEXTURE_2D, 0, gl.RGB, screenPositionPixels.x, screenPositionPixels.y, 16, 16, 0 );


					// render pink quad

					gl.uniform1i( uniforms.renderType, 0 );
					gl.uniform2f( uniforms.scale, scale.x, scale.y );
					gl.uniform3f( uniforms.screenPosition, screenPosition.x, screenPosition.y, screenPosition.z );

					state.disable( gl.BLEND );
					state.enable( gl.DEPTH_TEST );

					gl.drawElements( gl.TRIANGLES, 6, gl.UNSIGNED_SHORT, 0 );


					// copy result to occlusionMap

					state.activeTexture( gl.TEXTURE0 );
					state.bindTexture( gl.TEXTURE_2D, occlusionTexture );
					gl.copyTexImage2D( gl.TEXTURE_2D, 0, gl.RGBA, screenPositionPixels.x, screenPositionPixels.y, 16, 16, 0 );


					// restore graphics

					gl.uniform1i( uniforms.renderType, 1 );
					state.disable( gl.DEPTH_TEST );

					state.activeTexture( gl.TEXTURE1 );
					state.bindTexture( gl.TEXTURE_2D, tempTexture );
					gl.drawElements( gl.TRIANGLES, 6, gl.UNSIGNED_SHORT, 0 );


					// update object positions

					flare.positionScreen.copy( screenPosition );

					if ( flare.customUpdateCallback ) {

						flare.customUpdateCallback( flare );

					} else {

						flare.updateLensFlares();

					}

					// render flares

					gl.uniform1i( uniforms.renderType, 2 );
					state.enable( gl.BLEND );

					for ( var j = 0, jl = flare.lensFlares.length; j < jl; j ++ ) {

						var sprite = flare.lensFlares[ j ];

						if ( sprite.opacity > 0.001 && sprite.scale > 0.001 ) {

							screenPosition.x = sprite.x;
							screenPosition.y = sprite.y;
							screenPosition.z = sprite.z;

							size = sprite.size * sprite.scale / viewport.w;

							scale.x = size * invAspect;
							scale.y = size;

							gl.uniform3f( uniforms.screenPosition, screenPosition.x, screenPosition.y, screenPosition.z );
							gl.uniform2f( uniforms.scale, scale.x, scale.y );
							gl.uniform1f( uniforms.rotation, sprite.rotation );

							gl.uniform1f( uniforms.opacity, sprite.opacity );
							gl.uniform3f( uniforms.color, sprite.color.r, sprite.color.g, sprite.color.b );

							state.setBlending( sprite.blending, sprite.blendEquation, sprite.blendSrc, sprite.blendDst );
							renderer.setTexture2D( sprite.texture, 1 );

							gl.drawElements( gl.TRIANGLES, 6, gl.UNSIGNED_SHORT, 0 );

						}

					}

				}

			}

			// restore gl

			state.enable( gl.CULL_FACE );
			state.enable( gl.DEPTH_TEST );
			state.setDepthWrite( true );

			renderer.resetGLState();

		};

		function createProgram( shader ) {

			var program = gl.createProgram();

			var fragmentShader = gl.createShader( gl.FRAGMENT_SHADER );
			var vertexShader = gl.createShader( gl.VERTEX_SHADER );

			var prefix = "precision " + renderer.getPrecision() + " float;\n";

			gl.shaderSource( fragmentShader, prefix + shader.fragmentShader );
			gl.shaderSource( vertexShader, prefix + shader.vertexShader );

			gl.compileShader( fragmentShader );
			gl.compileShader( vertexShader );

			gl.attachShader( program, fragmentShader );
			gl.attachShader( program, vertexShader );

			gl.linkProgram( program );

			return program;

		}

	}

	/**
	 * @author mikael emtinger / http://gomo.se/
	 * @author alteredq / http://alteredqualia.com/
	 */

	function SpritePlugin( renderer, sprites ) {

		var gl = renderer.context;
		var state = renderer.state;

		var vertexBuffer, elementBuffer;
		var program, attributes, uniforms;

		var texture;

		// decompose matrixWorld

		var spritePosition = new Vector3();
		var spriteRotation = new Quaternion();
		var spriteScale = new Vector3();

		function init() {

			var vertices = new Float32Array( [
				- 0.5, - 0.5,  0, 0,
				  0.5, - 0.5,  1, 0,
				  0.5,   0.5,  1, 1,
				- 0.5,   0.5,  0, 1
			] );

			var faces = new Uint16Array( [
				0, 1, 2,
				0, 2, 3
			] );

			vertexBuffer  = gl.createBuffer();
			elementBuffer = gl.createBuffer();

			gl.bindBuffer( gl.ARRAY_BUFFER, vertexBuffer );
			gl.bufferData( gl.ARRAY_BUFFER, vertices, gl.STATIC_DRAW );

			gl.bindBuffer( gl.ELEMENT_ARRAY_BUFFER, elementBuffer );
			gl.bufferData( gl.ELEMENT_ARRAY_BUFFER, faces, gl.STATIC_DRAW );

			program = createProgram();

			attributes = {
				position:			gl.getAttribLocation ( program, 'position' ),
				uv:					gl.getAttribLocation ( program, 'uv' )
			};

			uniforms = {
				uvOffset:			gl.getUniformLocation( program, 'uvOffset' ),
				uvScale:			gl.getUniformLocation( program, 'uvScale' ),

				rotation:			gl.getUniformLocation( program, 'rotation' ),
				scale:				gl.getUniformLocation( program, 'scale' ),

				color:				gl.getUniformLocation( program, 'color' ),
				map:				gl.getUniformLocation( program, 'map' ),
				opacity:			gl.getUniformLocation( program, 'opacity' ),

				modelViewMatrix: 	gl.getUniformLocation( program, 'modelViewMatrix' ),
				projectionMatrix:	gl.getUniformLocation( program, 'projectionMatrix' ),

				fogType:			gl.getUniformLocation( program, 'fogType' ),
				fogDensity:			gl.getUniformLocation( program, 'fogDensity' ),
				fogNear:			gl.getUniformLocation( program, 'fogNear' ),
				fogFar:				gl.getUniformLocation( program, 'fogFar' ),
				fogColor:			gl.getUniformLocation( program, 'fogColor' ),

				alphaTest:			gl.getUniformLocation( program, 'alphaTest' )
			};

			var canvas = document.createElementNS( 'http://www.w3.org/1999/xhtml', 'canvas' );
			canvas.width = 8;
			canvas.height = 8;

			var context = canvas.getContext( '2d' );
			context.fillStyle = 'white';
			context.fillRect( 0, 0, 8, 8 );

			texture = new Texture( canvas );
			texture.needsUpdate = true;

		}

		this.render = function ( scene, camera ) {

			if ( sprites.length === 0 ) return;

			// setup gl

			if ( program === undefined ) {

				init();

			}

			gl.useProgram( program );

			state.initAttributes();
			state.enableAttribute( attributes.position );
			state.enableAttribute( attributes.uv );
			state.disableUnusedAttributes();

			state.disable( gl.CULL_FACE );
			state.enable( gl.BLEND );

			gl.bindBuffer( gl.ARRAY_BUFFER, vertexBuffer );
			gl.vertexAttribPointer( attributes.position, 2, gl.FLOAT, false, 2 * 8, 0 );
			gl.vertexAttribPointer( attributes.uv, 2, gl.FLOAT, false, 2 * 8, 8 );

			gl.bindBuffer( gl.ELEMENT_ARRAY_BUFFER, elementBuffer );

			gl.uniformMatrix4fv( uniforms.projectionMatrix, false, camera.projectionMatrix.elements );

			state.activeTexture( gl.TEXTURE0 );
			gl.uniform1i( uniforms.map, 0 );

			var oldFogType = 0;
			var sceneFogType = 0;
			var fog = scene.fog;

			if ( fog ) {

				gl.uniform3f( uniforms.fogColor, fog.color.r, fog.color.g, fog.color.b );

				if ( fog.isFog ) {

					gl.uniform1f( uniforms.fogNear, fog.near );
					gl.uniform1f( uniforms.fogFar, fog.far );

					gl.uniform1i( uniforms.fogType, 1 );
					oldFogType = 1;
					sceneFogType = 1;

				} else if ( fog.isFogExp2 ) {

					gl.uniform1f( uniforms.fogDensity, fog.density );

					gl.uniform1i( uniforms.fogType, 2 );
					oldFogType = 2;
					sceneFogType = 2;

				}

			} else {

				gl.uniform1i( uniforms.fogType, 0 );
				oldFogType = 0;
				sceneFogType = 0;

			}


			// update positions and sort

			for ( var i = 0, l = sprites.length; i < l; i ++ ) {

				var sprite = sprites[ i ];

				sprite.modelViewMatrix.multiplyMatrices( camera.matrixWorldInverse, sprite.matrixWorld );
				sprite.z = - sprite.modelViewMatrix.elements[ 14 ];

			}

			sprites.sort( painterSortStable );

			// render all sprites

			var scale = [];

			for ( var i = 0, l = sprites.length; i < l; i ++ ) {

				var sprite = sprites[ i ];
				var material = sprite.material;

				if ( material.visible === false ) continue;

				gl.uniform1f( uniforms.alphaTest, material.alphaTest );
				gl.uniformMatrix4fv( uniforms.modelViewMatrix, false, sprite.modelViewMatrix.elements );

				sprite.matrixWorld.decompose( spritePosition, spriteRotation, spriteScale );

				scale[ 0 ] = spriteScale.x;
				scale[ 1 ] = spriteScale.y;

				var fogType = 0;

				if ( scene.fog && material.fog ) {

					fogType = sceneFogType;

				}

				if ( oldFogType !== fogType ) {

					gl.uniform1i( uniforms.fogType, fogType );
					oldFogType = fogType;

				}

				if ( material.map !== null ) {

					gl.uniform2f( uniforms.uvOffset, material.map.offset.x, material.map.offset.y );
					gl.uniform2f( uniforms.uvScale, material.map.repeat.x, material.map.repeat.y );

				} else {

					gl.uniform2f( uniforms.uvOffset, 0, 0 );
					gl.uniform2f( uniforms.uvScale, 1, 1 );

				}

				gl.uniform1f( uniforms.opacity, material.opacity );
				gl.uniform3f( uniforms.color, material.color.r, material.color.g, material.color.b );

				gl.uniform1f( uniforms.rotation, material.rotation );
				gl.uniform2fv( uniforms.scale, scale );

				state.setBlending( material.blending, material.blendEquation, material.blendSrc, material.blendDst );
				state.setDepthTest( material.depthTest );
				state.setDepthWrite( material.depthWrite );

				if ( material.map ) {

					renderer.setTexture2D( material.map, 0 );

				} else {

					renderer.setTexture2D( texture, 0 );

				}

				gl.drawElements( gl.TRIANGLES, 6, gl.UNSIGNED_SHORT, 0 );

			}

			// restore gl

			state.enable( gl.CULL_FACE );

			renderer.resetGLState();

		};

		function createProgram() {

			var program = gl.createProgram();

			var vertexShader = gl.createShader( gl.VERTEX_SHADER );
			var fragmentShader = gl.createShader( gl.FRAGMENT_SHADER );

			gl.shaderSource( vertexShader, [

				'precision ' + renderer.getPrecision() + ' float;',

				'uniform mat4 modelViewMatrix;',
				'uniform mat4 projectionMatrix;',
				'uniform float rotation;',
				'uniform vec2 scale;',
				'uniform vec2 uvOffset;',
				'uniform vec2 uvScale;',

				'attribute vec2 position;',
				'attribute vec2 uv;',

				'varying vec2 vUV;',

				'void main() {',

					'vUV = uvOffset + uv * uvScale;',

					'vec2 alignedPosition = position * scale;',

					'vec2 rotatedPosition;',
					'rotatedPosition.x = cos( rotation ) * alignedPosition.x - sin( rotation ) * alignedPosition.y;',
					'rotatedPosition.y = sin( rotation ) * alignedPosition.x + cos( rotation ) * alignedPosition.y;',

					'vec4 finalPosition;',

					'finalPosition = modelViewMatrix * vec4( 0.0, 0.0, 0.0, 1.0 );',
					'finalPosition.xy += rotatedPosition;',
					'finalPosition = projectionMatrix * finalPosition;',

					'gl_Position = finalPosition;',

				'}'

			].join( '\n' ) );

			gl.shaderSource( fragmentShader, [

				'precision ' + renderer.getPrecision() + ' float;',

				'uniform vec3 color;',
				'uniform sampler2D map;',
				'uniform float opacity;',

				'uniform int fogType;',
				'uniform vec3 fogColor;',
				'uniform float fogDensity;',
				'uniform float fogNear;',
				'uniform float fogFar;',
				'uniform float alphaTest;',

				'varying vec2 vUV;',

				'void main() {',

					'vec4 texture = texture2D( map, vUV );',

					'if ( texture.a < alphaTest ) discard;',

					'gl_FragColor = vec4( color * texture.xyz, texture.a * opacity );',

					'if ( fogType > 0 ) {',

						'float depth = gl_FragCoord.z / gl_FragCoord.w;',
						'float fogFactor = 0.0;',

						'if ( fogType == 1 ) {',

							'fogFactor = smoothstep( fogNear, fogFar, depth );',

						'} else {',

							'const float LOG2 = 1.442695;',
							'fogFactor = exp2( - fogDensity * fogDensity * depth * depth * LOG2 );',
							'fogFactor = 1.0 - clamp( fogFactor, 0.0, 1.0 );',

						'}',

						'gl_FragColor = mix( gl_FragColor, vec4( fogColor, gl_FragColor.w ), fogFactor );',

					'}',

				'}'

			].join( '\n' ) );

			gl.compileShader( vertexShader );
			gl.compileShader( fragmentShader );

			gl.attachShader( program, vertexShader );
			gl.attachShader( program, fragmentShader );

			gl.linkProgram( program );

			return program;

		}

		function painterSortStable( a, b ) {

			if ( a.renderOrder !== b.renderOrder ) {

				return a.renderOrder - b.renderOrder;

			} else if ( a.z !== b.z ) {

				return b.z - a.z;

			} else {

				return b.id - a.id;

			}

		}

	}

	/**
	 * @author mrdoob / http://mrdoob.com/
	 * @author alteredq / http://alteredqualia.com/
	 */

	var materialId = 0;

	function Material() {

		Object.defineProperty( this, 'id', { value: materialId ++ } );

		this.uuid = _Math.generateUUID();

		this.name = '';
		this.type = 'Material';

		this.fog = true;
		this.lights = true;

		this.blending = NormalBlending;
		this.side = FrontSide;
		this.shading = SmoothShading; // THREE.FlatShading, THREE.SmoothShading
		this.vertexColors = NoColors; // THREE.NoColors, THREE.VertexColors, THREE.FaceColors

		this.opacity = 1;
		this.transparent = false;

		this.blendSrc = SrcAlphaFactor;
		this.blendDst = OneMinusSrcAlphaFactor;
		this.blendEquation = AddEquation;
		this.blendSrcAlpha = null;
		this.blendDstAlpha = null;
		this.blendEquationAlpha = null;

		this.depthFunc = LessEqualDepth;
		this.depthTest = true;
		this.depthWrite = true;

		this.clippingPlanes = null;
		this.clipIntersection = false;
		this.clipShadows = false;

		this.colorWrite = true;

		this.precision = null; // override the renderer's default precision for this material

		this.polygonOffset = false;
		this.polygonOffsetFactor = 0;
		this.polygonOffsetUnits = 0;

		this.alphaTest = 0;
		this.premultipliedAlpha = false;

		this.overdraw = 0; // Overdrawn pixels (typically between 0 and 1) for fixing antialiasing gaps in CanvasRenderer

		this.visible = true;

		this._needsUpdate = true;

	}

	Material.prototype = {

		constructor: Material,

		isMaterial: true,

		get needsUpdate() {

			return this._needsUpdate;

		},

		set needsUpdate( value ) {

			if ( value === true ) this.update();
			this._needsUpdate = value;

		},

		setValues: function ( values ) {

			if ( values === undefined ) return;

			for ( var key in values ) {

				var newValue = values[ key ];

				if ( newValue === undefined ) {

					console.warn( "THREE.Material: '" + key + "' parameter is undefined." );
					continue;

				}

				var currentValue = this[ key ];

				if ( currentValue === undefined ) {

					console.warn( "THREE." + this.type + ": '" + key + "' is not a property of this material." );
					continue;

				}

				if ( currentValue && currentValue.isColor ) {

					currentValue.set( newValue );

				} else if ( (currentValue && currentValue.isVector3) && (newValue && newValue.isVector3) ) {

					currentValue.copy( newValue );

				} else if ( key === 'overdraw' ) {

					// ensure overdraw is backwards-compatible with legacy boolean type
					this[ key ] = Number( newValue );

				} else {

					this[ key ] = newValue;

				}

			}

		},

		toJSON: function ( meta ) {

			var isRoot = meta === undefined;

			if ( isRoot ) {

				meta = {
					textures: {},
					images: {}
				};

			}

			var data = {
				metadata: {
					version: 4.4,
					type: 'Material',
					generator: 'Material.toJSON'
				}
			};

			// standard Material serialization
			data.uuid = this.uuid;
			data.type = this.type;

			if ( this.name !== '' ) data.name = this.name;

			if ( this.color && this.color.isColor ) data.color = this.color.getHex();

			if ( this.roughness !== undefined ) data.roughness = this.roughness;
			if ( this.metalness !== undefined ) data.metalness = this.metalness;

			if ( this.emissive && this.emissive.isColor ) data.emissive = this.emissive.getHex();
			if ( this.specular && this.specular.isColor ) data.specular = this.specular.getHex();
			if ( this.shininess !== undefined ) data.shininess = this.shininess;
			if ( this.clearCoat !== undefined ) data.clearCoat = this.clearCoat;
			if ( this.clearCoatRoughness !== undefined ) data.clearCoatRoughness = this.clearCoatRoughness;

			if ( this.map && this.map.isTexture ) data.map = this.map.toJSON( meta ).uuid;
			if ( this.alphaMap && this.alphaMap.isTexture ) data.alphaMap = this.alphaMap.toJSON( meta ).uuid;
			if ( this.lightMap && this.lightMap.isTexture ) data.lightMap = this.lightMap.toJSON( meta ).uuid;
			if ( this.bumpMap && this.bumpMap.isTexture ) {

				data.bumpMap = this.bumpMap.toJSON( meta ).uuid;
				data.bumpScale = this.bumpScale;

			}
			if ( this.normalMap && this.normalMap.isTexture ) {

				data.normalMap = this.normalMap.toJSON( meta ).uuid;
				data.normalScale = this.normalScale.toArray();

			}
			if ( this.displacementMap && this.displacementMap.isTexture ) {

				data.displacementMap = this.displacementMap.toJSON( meta ).uuid;
				data.displacementScale = this.displacementScale;
				data.displacementBias = this.displacementBias;

			}
			if ( this.roughnessMap && this.roughnessMap.isTexture ) data.roughnessMap = this.roughnessMap.toJSON( meta ).uuid;
			if ( this.metalnessMap && this.metalnessMap.isTexture ) data.metalnessMap = this.metalnessMap.toJSON( meta ).uuid;

			if ( this.emissiveMap && this.emissiveMap.isTexture ) data.emissiveMap = this.emissiveMap.toJSON( meta ).uuid;
			if ( this.specularMap && this.specularMap.isTexture ) data.specularMap = this.specularMap.toJSON( meta ).uuid;

			if ( this.envMap && this.envMap.isTexture ) {

				data.envMap = this.envMap.toJSON( meta ).uuid;
				data.reflectivity = this.reflectivity; // Scale behind envMap

			}

			if ( this.gradientMap && this.gradientMap.isTexture ) {

				data.gradientMap = this.gradientMap.toJSON( meta ).uuid;

			}

			if ( this.size !== undefined ) data.size = this.size;
			if ( this.sizeAttenuation !== undefined ) data.sizeAttenuation = this.sizeAttenuation;

			if ( this.blending !== NormalBlending ) data.blending = this.blending;
			if ( this.shading !== SmoothShading ) data.shading = this.shading;
			if ( this.side !== FrontSide ) data.side = this.side;
			if ( this.vertexColors !== NoColors ) data.vertexColors = this.vertexColors;

			if ( this.opacity < 1 ) data.opacity = this.opacity;
			if ( this.transparent === true ) data.transparent = this.transparent;

			data.depthFunc = this.depthFunc;
			data.depthTest = this.depthTest;
			data.depthWrite = this.depthWrite;

			if ( this.alphaTest > 0 ) data.alphaTest = this.alphaTest;
			if ( this.premultipliedAlpha === true ) data.premultipliedAlpha = this.premultipliedAlpha;
			if ( this.wireframe === true ) data.wireframe = this.wireframe;
			if ( this.wireframeLinewidth > 1 ) data.wireframeLinewidth = this.wireframeLinewidth;
			if ( this.wireframeLinecap !== 'round' ) data.wireframeLinecap = this.wireframeLinecap;
			if ( this.wireframeLinejoin !== 'round' ) data.wireframeLinejoin = this.wireframeLinejoin;

			data.skinning = this.skinning;
			data.morphTargets = this.morphTargets;

			// TODO: Copied from Object3D.toJSON

			function extractFromCache( cache ) {

				var values = [];

				for ( var key in cache ) {

					var data = cache[ key ];
					delete data.metadata;
					values.push( data );

				}

				return values;

			}

			if ( isRoot ) {

				var textures = extractFromCache( meta.textures );
				var images = extractFromCache( meta.images );

				if ( textures.length > 0 ) data.textures = textures;
				if ( images.length > 0 ) data.images = images;

			}

			return data;

		},

		clone: function () {

			return new this.constructor().copy( this );

		},

		copy: function ( source ) {

			this.name = source.name;

			this.fog = source.fog;
			this.lights = source.lights;

			this.blending = source.blending;
			this.side = source.side;
			this.shading = source.shading;
			this.vertexColors = source.vertexColors;

			this.opacity = source.opacity;
			this.transparent = source.transparent;

			this.blendSrc = source.blendSrc;
			this.blendDst = source.blendDst;
			this.blendEquation = source.blendEquation;
			this.blendSrcAlpha = source.blendSrcAlpha;
			this.blendDstAlpha = source.blendDstAlpha;
			this.blendEquationAlpha = source.blendEquationAlpha;

			this.depthFunc = source.depthFunc;
			this.depthTest = source.depthTest;
			this.depthWrite = source.depthWrite;

			this.colorWrite = source.colorWrite;

			this.precision = source.precision;

			this.polygonOffset = source.polygonOffset;
			this.polygonOffsetFactor = source.polygonOffsetFactor;
			this.polygonOffsetUnits = source.polygonOffsetUnits;

			this.alphaTest = source.alphaTest;

			this.premultipliedAlpha = source.premultipliedAlpha;

			this.overdraw = source.overdraw;

			this.visible = source.visible;
			this.clipShadows = source.clipShadows;
			this.clipIntersection = source.clipIntersection;

			var srcPlanes = source.clippingPlanes,
				dstPlanes = null;

			if ( srcPlanes !== null ) {

				var n = srcPlanes.length;
				dstPlanes = new Array( n );

				for ( var i = 0; i !== n; ++ i )
					dstPlanes[ i ] = srcPlanes[ i ].clone();

			}

			this.clippingPlanes = dstPlanes;

			return this;

		},

		update: function () {

			this.dispatchEvent( { type: 'update' } );

		},

		dispose: function () {

			this.dispatchEvent( { type: 'dispose' } );

		}

	};

	Object.assign( Material.prototype, EventDispatcher.prototype );

	/**
	 * @author alteredq / http://alteredqualia.com/
	 *
	 * parameters = {
	 *  defines: { "label" : "value" },
	 *  uniforms: { "parameter1": { value: 1.0 }, "parameter2": { value2: 2 } },
	 *
	 *  fragmentShader: <string>,
	 *  vertexShader: <string>,
	 *
	 *  wireframe: <boolean>,
	 *  wireframeLinewidth: <float>,
	 *
	 *  lights: <bool>,
	 *
	 *  skinning: <bool>,
	 *  morphTargets: <bool>,
	 *  morphNormals: <bool>
	 * }
	 */

	function ShaderMaterial( parameters ) {

		Material.call( this );

		this.type = 'ShaderMaterial';

		this.defines = {};
		this.uniforms = {};

		this.vertexShader = 'void main() {\n\tgl_Position = projectionMatrix * modelViewMatrix * vec4( position, 1.0 );\n}';
		this.fragmentShader = 'void main() {\n\tgl_FragColor = vec4( 1.0, 0.0, 0.0, 1.0 );\n}';

		this.linewidth = 1;

		this.wireframe = false;
		this.wireframeLinewidth = 1;

		this.fog = false; // set to use scene fog
		this.lights = false; // set to use scene lights
		this.clipping = false; // set to use user-defined clipping planes

		this.skinning = false; // set to use skinning attribute streams
		this.morphTargets = false; // set to use morph targets
		this.morphNormals = false; // set to use morph normals

		this.extensions = {
			derivatives: false, // set to use derivatives
			fragDepth: false, // set to use fragment depth values
			drawBuffers: false, // set to use draw buffers
			shaderTextureLOD: false // set to use shader texture LOD
		};

		// When rendered geometry doesn't include these attributes but the material does,
		// use these default values in WebGL. This avoids errors when buffer data is missing.
		this.defaultAttributeValues = {
			'color': [ 1, 1, 1 ],
			'uv': [ 0, 0 ],
			'uv2': [ 0, 0 ]
		};

		this.index0AttributeName = undefined;

		if ( parameters !== undefined ) {

			if ( parameters.attributes !== undefined ) {

				console.error( 'THREE.ShaderMaterial: attributes should now be defined in THREE.BufferGeometry instead.' );

			}

			this.setValues( parameters );

		}

	}

	ShaderMaterial.prototype = Object.create( Material.prototype );
	ShaderMaterial.prototype.constructor = ShaderMaterial;

	ShaderMaterial.prototype.isShaderMaterial = true;

	ShaderMaterial.prototype.copy = function ( source ) {

		Material.prototype.copy.call( this, source );

		this.fragmentShader = source.fragmentShader;
		this.vertexShader = source.vertexShader;

		this.uniforms = UniformsUtils.clone( source.uniforms );

		this.defines = source.defines;

		this.wireframe = source.wireframe;
		this.wireframeLinewidth = source.wireframeLinewidth;

		this.lights = source.lights;
		this.clipping = source.clipping;

		this.skinning = source.skinning;

		this.morphTargets = source.morphTargets;
		this.morphNormals = source.morphNormals;

		this.extensions = source.extensions;

		return this;

	};

	ShaderMaterial.prototype.toJSON = function ( meta ) {

		var data = Material.prototype.toJSON.call( this, meta );

		data.uniforms = this.uniforms;
		data.vertexShader = this.vertexShader;
		data.fragmentShader = this.fragmentShader;

		return data;

	};

	/**
	 * @author mrdoob / http://mrdoob.com/
	 * @author alteredq / http://alteredqualia.com/
	 * @author bhouston / https://clara.io
	 * @author WestLangley / http://github.com/WestLangley
	 *
	 * parameters = {
	 *
	 *  opacity: <float>,
	 *
	 *  map: new THREE.Texture( <Image> ),
	 *
	 *  alphaMap: new THREE.Texture( <Image> ),
	 *
	 *  displacementMap: new THREE.Texture( <Image> ),
	 *  displacementScale: <float>,
	 *  displacementBias: <float>,
	 *
	 *  wireframe: <boolean>,
	 *  wireframeLinewidth: <float>
	 * }
	 */

	function MeshDepthMaterial( parameters ) {

		Material.call( this );

		this.type = 'MeshDepthMaterial';

		this.depthPacking = BasicDepthPacking;

		this.skinning = false;
		this.morphTargets = false;

		this.map = null;

		this.alphaMap = null;

		this.displacementMap = null;
		this.displacementScale = 1;
		this.displacementBias = 0;

		this.wireframe = false;
		this.wireframeLinewidth = 1;

		this.fog = false;
		this.lights = false;

		this.setValues( parameters );

	}

	MeshDepthMaterial.prototype = Object.create( Material.prototype );
	MeshDepthMaterial.prototype.constructor = MeshDepthMaterial;

	MeshDepthMaterial.prototype.isMeshDepthMaterial = true;

	MeshDepthMaterial.prototype.copy = function ( source ) {

		Material.prototype.copy.call( this, source );

		this.depthPacking = source.depthPacking;

		this.skinning = source.skinning;
		this.morphTargets = source.morphTargets;

		this.map = source.map;

		this.alphaMap = source.alphaMap;

		this.displacementMap = source.displacementMap;
		this.displacementScale = source.displacementScale;
		this.displacementBias = source.displacementBias;

		this.wireframe = source.wireframe;
		this.wireframeLinewidth = source.wireframeLinewidth;

		return this;

	};

	/**
	 * @author bhouston / http://clara.io
	 * @author WestLangley / http://github.com/WestLangley
	 */

	function Box3( min, max ) {

		this.min = ( min !== undefined ) ? min : new Vector3( + Infinity, + Infinity, + Infinity );
		this.max = ( max !== undefined ) ? max : new Vector3( - Infinity, - Infinity, - Infinity );

	}

	Box3.prototype = {

		constructor: Box3,

		isBox3: true,

		set: function ( min, max ) {

			this.min.copy( min );
			this.max.copy( max );

			return this;

		},

		setFromArray: function ( array ) {

			var minX = + Infinity;
			var minY = + Infinity;
			var minZ = + Infinity;

			var maxX = - Infinity;
			var maxY = - Infinity;
			var maxZ = - Infinity;

			for ( var i = 0, l = array.length; i < l; i += 3 ) {

				var x = array[ i ];
				var y = array[ i + 1 ];
				var z = array[ i + 2 ];

				if ( x < minX ) minX = x;
				if ( y < minY ) minY = y;
				if ( z < minZ ) minZ = z;

				if ( x > maxX ) maxX = x;
				if ( y > maxY ) maxY = y;
				if ( z > maxZ ) maxZ = z;

			}

			this.min.set( minX, minY, minZ );
			this.max.set( maxX, maxY, maxZ );

		},

		setFromBufferAttribute: function ( attribute ) {

			var minX = + Infinity;
			var minY = + Infinity;
			var minZ = + Infinity;

			var maxX = - Infinity;
			var maxY = - Infinity;
			var maxZ = - Infinity;

			for ( var i = 0, l = attribute.count; i < l; i ++ ) {

				var x = attribute.getX( i );
				var y = attribute.getY( i );
				var z = attribute.getZ( i );

				if ( x < minX ) minX = x;
				if ( y < minY ) minY = y;
				if ( z < minZ ) minZ = z;

				if ( x > maxX ) maxX = x;
				if ( y > maxY ) maxY = y;
				if ( z > maxZ ) maxZ = z;

			}

			this.min.set( minX, minY, minZ );
			this.max.set( maxX, maxY, maxZ );

		},

		setFromPoints: function ( points ) {

			this.makeEmpty();

			for ( var i = 0, il = points.length; i < il; i ++ ) {

				this.expandByPoint( points[ i ] );

			}

			return this;

		},

		setFromCenterAndSize: function () {

			var v1 = new Vector3();

			return function setFromCenterAndSize( center, size ) {

				var halfSize = v1.copy( size ).multiplyScalar( 0.5 );

				this.min.copy( center ).sub( halfSize );
				this.max.copy( center ).add( halfSize );

				return this;

			};

		}(),

		setFromObject: function () {

			// Computes the world-axis-aligned bounding box of an object (including its children),
			// accounting for both the object's, and children's, world transforms

			var v1 = new Vector3();

			return function setFromObject( object ) {

				var scope = this;

				object.updateMatrixWorld( true );

				this.makeEmpty();

				object.traverse( function ( node ) {

					var i, l;

					var geometry = node.geometry;

					if ( geometry !== undefined ) {

						if ( geometry.isGeometry ) {

							var vertices = geometry.vertices;

							for ( i = 0, l = vertices.length; i < l; i ++ ) {

								v1.copy( vertices[ i ] );
								v1.applyMatrix4( node.matrixWorld );

								scope.expandByPoint( v1 );

							}

						} else if ( geometry.isBufferGeometry ) {

							var attribute = geometry.attributes.position;

							if ( attribute !== undefined ) {

								for ( i = 0, l = attribute.count; i < l; i ++ ) {

									v1.fromAttribute( attribute, i ).applyMatrix4( node.matrixWorld );

									scope.expandByPoint( v1 );

								}

							}

						}

					}

				} );

				return this;

			};

		}(),

		clone: function () {

			return new this.constructor().copy( this );

		},

		copy: function ( box ) {

			this.min.copy( box.min );
			this.max.copy( box.max );

			return this;

		},

		makeEmpty: function () {

			this.min.x = this.min.y = this.min.z = + Infinity;
			this.max.x = this.max.y = this.max.z = - Infinity;

			return this;

		},

		isEmpty: function () {

			// this is a more robust check for empty than ( volume <= 0 ) because volume can get positive with two negative axes

			return ( this.max.x < this.min.x ) || ( this.max.y < this.min.y ) || ( this.max.z < this.min.z );

		},

		getCenter: function ( optionalTarget ) {

			var result = optionalTarget || new Vector3();
			return this.isEmpty() ? result.set( 0, 0, 0 ) : result.addVectors( this.min, this.max ).multiplyScalar( 0.5 );

		},

		getSize: function ( optionalTarget ) {

			var result = optionalTarget || new Vector3();
			return this.isEmpty() ? result.set( 0, 0, 0 ) : result.subVectors( this.max, this.min );

		},

		expandByPoint: function ( point ) {

			this.min.min( point );
			this.max.max( point );

			return this;

		},

		expandByVector: function ( vector ) {

			this.min.sub( vector );
			this.max.add( vector );

			return this;

		},

		expandByScalar: function ( scalar ) {

			this.min.addScalar( - scalar );
			this.max.addScalar( scalar );

			return this;

		},

		containsPoint: function ( point ) {

			return point.x < this.min.x || point.x > this.max.x ||
				point.y < this.min.y || point.y > this.max.y ||
				point.z < this.min.z || point.z > this.max.z ? false : true;

		},

		containsBox: function ( box ) {

			return this.min.x <= box.min.x && box.max.x <= this.max.x &&
				this.min.y <= box.min.y && box.max.y <= this.max.y &&
				this.min.z <= box.min.z && box.max.z <= this.max.z;

		},

		getParameter: function ( point, optionalTarget ) {

			// This can potentially have a divide by zero if the box
			// has a size dimension of 0.

			var result = optionalTarget || new Vector3();

			return result.set(
				( point.x - this.min.x ) / ( this.max.x - this.min.x ),
				( point.y - this.min.y ) / ( this.max.y - this.min.y ),
				( point.z - this.min.z ) / ( this.max.z - this.min.z )
			);

		},

		intersectsBox: function ( box ) {

			// using 6 splitting planes to rule out intersections.
			return box.max.x < this.min.x || box.min.x > this.max.x ||
				box.max.y < this.min.y || box.min.y > this.max.y ||
				box.max.z < this.min.z || box.min.z > this.max.z ? false : true;

		},

		intersectsSphere: ( function () {

			var closestPoint;

			return function intersectsSphere( sphere ) {

				if ( closestPoint === undefined ) closestPoint = new Vector3();

				// Find the point on the AABB closest to the sphere center.
				this.clampPoint( sphere.center, closestPoint );

				// If that point is inside the sphere, the AABB and sphere intersect.
				return closestPoint.distanceToSquared( sphere.center ) <= ( sphere.radius * sphere.radius );

			};

		} )(),

		intersectsPlane: function ( plane ) {

			// We compute the minimum and maximum dot product values. If those values
			// are on the same side (back or front) of the plane, then there is no intersection.

			var min, max;

			if ( plane.normal.x > 0 ) {

				min = plane.normal.x * this.min.x;
				max = plane.normal.x * this.max.x;

			} else {

				min = plane.normal.x * this.max.x;
				max = plane.normal.x * this.min.x;

			}

			if ( plane.normal.y > 0 ) {

				min += plane.normal.y * this.min.y;
				max += plane.normal.y * this.max.y;

			} else {

				min += plane.normal.y * this.max.y;
				max += plane.normal.y * this.min.y;

			}

			if ( plane.normal.z > 0 ) {

				min += plane.normal.z * this.min.z;
				max += plane.normal.z * this.max.z;

			} else {

				min += plane.normal.z * this.max.z;
				max += plane.normal.z * this.min.z;

			}

			return ( min <= plane.constant && max >= plane.constant );

		},

		clampPoint: function ( point, optionalTarget ) {

			var result = optionalTarget || new Vector3();
			return result.copy( point ).clamp( this.min, this.max );

		},

		distanceToPoint: function () {

			var v1 = new Vector3();

			return function distanceToPoint( point ) {

				var clampedPoint = v1.copy( point ).clamp( this.min, this.max );
				return clampedPoint.sub( point ).length();

			};

		}(),

		getBoundingSphere: function () {

			var v1 = new Vector3();

			return function getBoundingSphere( optionalTarget ) {

				var result = optionalTarget || new Sphere();

				this.getCenter( result.center );

				result.radius = this.getSize( v1 ).length() * 0.5;

				return result;

			};

		}(),

		intersect: function ( box ) {

			this.min.max( box.min );
			this.max.min( box.max );

			// ensure that if there is no overlap, the result is fully empty, not slightly empty with non-inf/+inf values that will cause subsequence intersects to erroneously return valid values.
			if( this.isEmpty() ) this.makeEmpty();

			return this;

		},

		union: function ( box ) {

			this.min.min( box.min );
			this.max.max( box.max );

			return this;

		},

		applyMatrix4: function () {

			var points = [
				new Vector3(),
				new Vector3(),
				new Vector3(),
				new Vector3(),
				new Vector3(),
				new Vector3(),
				new Vector3(),
				new Vector3()
			];

			return function applyMatrix4( matrix ) {

				// transform of empty box is an empty box.
				if( this.isEmpty() ) return this;

				// NOTE: I am using a binary pattern to specify all 2^3 combinations below
				points[ 0 ].set( this.min.x, this.min.y, this.min.z ).applyMatrix4( matrix ); // 000
				points[ 1 ].set( this.min.x, this.min.y, this.max.z ).applyMatrix4( matrix ); // 001
				points[ 2 ].set( this.min.x, this.max.y, this.min.z ).applyMatrix4( matrix ); // 010
				points[ 3 ].set( this.min.x, this.max.y, this.max.z ).applyMatrix4( matrix ); // 011
				points[ 4 ].set( this.max.x, this.min.y, this.min.z ).applyMatrix4( matrix ); // 100
				points[ 5 ].set( this.max.x, this.min.y, this.max.z ).applyMatrix4( matrix ); // 101
				points[ 6 ].set( this.max.x, this.max.y, this.min.z ).applyMatrix4( matrix ); // 110
				points[ 7 ].set( this.max.x, this.max.y, this.max.z ).applyMatrix4( matrix );	// 111

				this.setFromPoints( points );

				return this;

			};

		}(),

		translate: function ( offset ) {

			this.min.add( offset );
			this.max.add( offset );

			return this;

		},

		equals: function ( box ) {

			return box.min.equals( this.min ) && box.max.equals( this.max );

		}

	};

	/**
	 * @author bhouston / http://clara.io
	 * @author mrdoob / http://mrdoob.com/
	 */

	function Sphere( center, radius ) {

		this.center = ( center !== undefined ) ? center : new Vector3();
		this.radius = ( radius !== undefined ) ? radius : 0;

	}

	Sphere.prototype = {

		constructor: Sphere,

		set: function ( center, radius ) {

			this.center.copy( center );
			this.radius = radius;

			return this;

		},

		setFromPoints: function () {

			var box = new Box3();

			return function setFromPoints( points, optionalCenter ) {

				var center = this.center;

				if ( optionalCenter !== undefined ) {

					center.copy( optionalCenter );

				} else {

					box.setFromPoints( points ).getCenter( center );

				}

				var maxRadiusSq = 0;

				for ( var i = 0, il = points.length; i < il; i ++ ) {

					maxRadiusSq = Math.max( maxRadiusSq, center.distanceToSquared( points[ i ] ) );

				}

				this.radius = Math.sqrt( maxRadiusSq );

				return this;

			};

		}(),

		clone: function () {

			return new this.constructor().copy( this );

		},

		copy: function ( sphere ) {

			this.center.copy( sphere.center );
			this.radius = sphere.radius;

			return this;

		},

		empty: function () {

			return ( this.radius <= 0 );

		},

		containsPoint: function ( point ) {

			return ( point.distanceToSquared( this.center ) <= ( this.radius * this.radius ) );

		},

		distanceToPoint: function ( point ) {

			return ( point.distanceTo( this.center ) - this.radius );

		},

		intersectsSphere: function ( sphere ) {

			var radiusSum = this.radius + sphere.radius;

			return sphere.center.distanceToSquared( this.center ) <= ( radiusSum * radiusSum );

		},

		intersectsBox: function ( box ) {

			return box.intersectsSphere( this );

		},

		intersectsPlane: function ( plane ) {

			// We use the following equation to compute the signed distance from
			// the center of the sphere to the plane.
			//
			// distance = q * n - d
			//
			// If this distance is greater than the radius of the sphere,
			// then there is no intersection.

			return Math.abs( this.center.dot( plane.normal ) - plane.constant ) <= this.radius;

		},

		clampPoint: function ( point, optionalTarget ) {

			var deltaLengthSq = this.center.distanceToSquared( point );

			var result = optionalTarget || new Vector3();

			result.copy( point );

			if ( deltaLengthSq > ( this.radius * this.radius ) ) {

				result.sub( this.center ).normalize();
				result.multiplyScalar( this.radius ).add( this.center );

			}

			return result;

		},

		getBoundingBox: function ( optionalTarget ) {

			var box = optionalTarget || new Box3();

			box.set( this.center, this.center );
			box.expandByScalar( this.radius );

			return box;

		},

		applyMatrix4: function ( matrix ) {

			this.center.applyMatrix4( matrix );
			this.radius = this.radius * matrix.getMaxScaleOnAxis();

			return this;

		},

		translate: function ( offset ) {

			this.center.add( offset );

			return this;

		},

		equals: function ( sphere ) {

			return sphere.center.equals( this.center ) && ( sphere.radius === this.radius );

		}

	};

	/**
	 * @author alteredq / http://alteredqualia.com/
	 * @author WestLangley / http://github.com/WestLangley
	 * @author bhouston / http://clara.io
	 * @author tschw
	 */

	function Matrix3() {

		this.elements = new Float32Array( [

			1, 0, 0,
			0, 1, 0,
			0, 0, 1

		] );

		if ( arguments.length > 0 ) {

			console.error( 'THREE.Matrix3: the constructor no longer reads arguments. use .set() instead.' );

		}

	}

	Matrix3.prototype = {

		constructor: Matrix3,

		isMatrix3: true,

		set: function ( n11, n12, n13, n21, n22, n23, n31, n32, n33 ) {

			var te = this.elements;

			te[ 0 ] = n11; te[ 1 ] = n21; te[ 2 ] = n31;
			te[ 3 ] = n12; te[ 4 ] = n22; te[ 5 ] = n32;
			te[ 6 ] = n13; te[ 7 ] = n23; te[ 8 ] = n33;

			return this;

		},

		identity: function () {

			this.set(

				1, 0, 0,
				0, 1, 0,
				0, 0, 1

			);

			return this;

		},

		clone: function () {

			return new this.constructor().fromArray( this.elements );

		},

		copy: function ( m ) {

			var me = m.elements;

			this.set(

				me[ 0 ], me[ 3 ], me[ 6 ],
				me[ 1 ], me[ 4 ], me[ 7 ],
				me[ 2 ], me[ 5 ], me[ 8 ]

			);

			return this;

		},

		setFromMatrix4: function( m ) {

			var me = m.elements;

			this.set(

				me[ 0 ], me[ 4 ], me[  8 ],
				me[ 1 ], me[ 5 ], me[  9 ],
				me[ 2 ], me[ 6 ], me[ 10 ]

			);

			return this;

		},

		applyToVector3Array: function () {

			var v1;

			return function applyToVector3Array( array, offset, length ) {

				if ( v1 === undefined ) v1 = new Vector3();
				if ( offset === undefined ) offset = 0;
				if ( length === undefined ) length = array.length;

				for ( var i = 0, j = offset; i < length; i += 3, j += 3 ) {

					v1.fromArray( array, j );
					v1.applyMatrix3( this );
					v1.toArray( array, j );

				}

				return array;

			};

		}(),

		applyToBufferAttribute: function () {

			var v1;

			return function applyToBufferAttribute( attribute ) {

				if ( v1 === undefined ) v1 = new Vector3();

				for ( var i = 0, l = attribute.count; i < l; i ++ ) {

					v1.x = attribute.getX( i );
					v1.y = attribute.getY( i );
					v1.z = attribute.getZ( i );

					v1.applyMatrix3( this );
```
