## Supplement File 1 for "Post Induction Hypotension prediction during general anesthesia using Machine Learning Techniques": lottie.min.js

```
(typeof navigator !== "undefined") && (function(root, factory) {
    if (typeof define === "function" && define.amd) {
        define(function() {
            return factory(root);
        });
    } else if (typeof module === "object" && module.exports) {
        module.exports = factory(root);
    } else {
        root.lottie = factory(root);
        root.bodymovin = root.lottie;
    }
}((window || {}), function(window) {
    "use strict";
    var svgNS = "http://www.w3.org/2000/svg";

var locationHref = '';

var initialDefaultFrame = -999999;

var subframeEnabled = true;
var expressionsPlugin;
var isSafari = /^((?!chrome|android).)*safari/i.test(navigator.userAgent);
var cachedColors = {};
var bm_rounder = Math.round;
var bm_rnd;
var bm_pow = Math.pow;
var bm_sqrt = Math.sqrt;
var bm_abs = Math.abs;
var bm_floor = Math.floor;
var bm_max = Math.max;
var bm_min = Math.min;
var blitter = 10;

var BMMath = {};
(function(){
    var propertyNames = ["abs", "acos", "acosh", "asin", "asinh", "atan", "atanh", "atan2", "ceil", "cbrt", "expm1", "clz32", "cos", "cosh", "exp", "floor", "fround", "hypot", "imul", "log", "log1p", "log2", "log10", "max", "min", "pow", "random", "round", "sign", "sin", "sinh", "sqrt", "tan", "tanh", "trunc", "E", "LN10", "LN2", "LOG10E", "LOG2E", "PI", "SQRT1_2", "SQRT2"];
    var i, len = propertyNames.length;
    for(i=0;i<len;i+=1){
        BMMath[propertyNames[i]] = Math[propertyNames[i]];
    }
}());

function ProjectInterface(){return {};}

BMMath.random = Math.random;
BMMath.abs = function(val){
    var tOfVal = typeof val;
    if(tOfVal === 'object' && val.length){
        var absArr = createSizedArray(val.length);
        var i, len = val.length;
        for(i=0;i<len;i+=1){
            absArr[i] = Math.abs(val[i]);
        }
        return absArr;
    }
    return Math.abs(val);

};
var defaultCurveSegments = 150;
var degToRads = Math.PI/180;
var roundCorner = 0.5519;

function roundValues(flag){
    if(flag){
        bm_rnd = Math.round;
    }else{
        bm_rnd = function(val){
            return val;
        };
    }
}
roundValues(false);

function styleDiv(element){
    element.style.position = 'absolute';
    element.style.top = 0;
    element.style.left = 0;
    element.style.display = 'block';
    element.style.transformOrigin = element.style.webkitTransformOrigin = '0 0';
    element.style.backfaceVisibility  = element.style.webkitBackfaceVisibility = 'visible';
    element.style.transformStyle = element.style.webkitTransformStyle = element.style.mozTransformStyle = "preserve-3d";
}

function BMEnterFrameEvent(n,c,t,d){
    this.type = n;
    this.currentTime = c;
    this.totalTime = t;
    this.direction = d < 0 ? -1:1;
}

function BMCompleteEvent(n,d){
    this.type = n;
    this.direction = d < 0 ? -1:1;
}

function BMCompleteLoopEvent(n,c,t,d){
    this.type = n;
    this.currentLoop = t;
    this.totalLoops = c;
    this.direction = d < 0 ? -1:1;
}

function BMSegmentStartEvent(n,f,t){
    this.type = n;
    this.firstFrame = f;
    this.totalFrames = t;
}

function BMDestroyEvent(n,t){
    this.type = n;
    this.target = t;
}

var createElementID = (function(){
    var _count = 0;
    return function createID() {
        return '__lottie_element_' + ++_count
    }
}())

function HSVtoRGB(h, s, v) {
    var r, g, b, i, f, p, q, t;
    i = Math.floor(h * 6);
    f = h * 6 - i;
    p = v * (1 - s);
    q = v * (1 - f * s);
    t = v * (1 - (1 - f) * s);
    switch (i % 6) {
        case 0: r = v; g = t; b = p; break;
        case 1: r = q; g = v; b = p; break;
        case 2: r = p; g = v; b = t; break;
        case 3: r = p; g = q; b = v; break;
        case 4: r = t; g = p; b = v; break;
        case 5: r = v; g = p; b = q; break;
    }
    return [ r,
        g,
         b ];
}

function RGBtoHSV(r, g, b) {
    var max = Math.max(r, g, b), min = Math.min(r, g, b),
        d = max - min,
        h,
        s = (max === 0 ? 0 : d / max),
        v = max / 255;

    switch (max) {
        case min: h = 0; break;
        case r: h = (g - b) + d * (g < b ? 6: 0); h /= 6 * d; break;
        case g: h = (b - r) + d * 2; h /= 6 * d; break;
        case b: h = (r - g) + d * 4; h /= 6 * d; break;
    }

    return [
         h,
         s,
         v
    ];
}

function addSaturationToRGB(color,offset){
    var hsv = RGBtoHSV(color[0]*255,color[1]*255,color[2]*255);
    hsv[1] += offset;
    if (hsv[1] > 1) {
        hsv[1] = 1;
    }
    else if (hsv[1] <= 0) {
        hsv[1] = 0;
    }
    return HSVtoRGB(hsv[0],hsv[1],hsv[2]);
}

function addBrightnessToRGB(color,offset){
    var hsv = RGBtoHSV(color[0]*255,color[1]*255,color[2]*255);
    hsv[2] += offset;
    if (hsv[2] > 1) {
        hsv[2] = 1;
    }
    else if (hsv[2] < 0) {
        hsv[2] = 0;
    }
    return HSVtoRGB(hsv[0],hsv[1],hsv[2]);
}

function addHueToRGB(color,offset) {
    var hsv = RGBtoHSV(color[0]*255,color[1]*255,color[2]*255);
    hsv[0] += offset/360;
    if (hsv[0] > 1) {
        hsv[0] -= 1;
    }
    else if (hsv[0] < 0) {
        hsv[0] += 1;
    }
    return HSVtoRGB(hsv[0],hsv[1],hsv[2]);
}

var rgbToHex = (function(){
    var colorMap = [];
    var i;
    var hex;
    for(i=0;i<256;i+=1){
        hex = i.toString(16);
        colorMap[i] = hex.length == 1 ? '0' + hex : hex;
    }

    return function(r, g, b) {
        if(r<0){
            r = 0;
        }
        if(g<0){
            g = 0;
        }
        if(b<0){
            b = 0;
        }
        return '#' + colorMap[r] + colorMap[g] + colorMap[b];
    };
}());
function BaseEvent(){}
BaseEvent.prototype = {
	triggerEvent: function (eventName, args) {
	    if (this._cbs[eventName]) {
	        var len = this._cbs[eventName].length;
	        for (var i = 0; i < len; i++){
	            this._cbs[eventName][i](args);
	        }
	    }
	},
	addEventListener: function (eventName, callback) {
	    if (!this._cbs[eventName]){
	        this._cbs[eventName] = [];
	    }
	    this._cbs[eventName].push(callback);

		return function() {
			this.removeEventListener(eventName, callback);
		}.bind(this);
	},
	removeEventListener: function (eventName,callback){
	    if (!callback){
	        this._cbs[eventName] = null;
	    }else if(this._cbs[eventName]){
	        var i = 0, len = this._cbs[eventName].length;
	        while(i<len){
	            if(this._cbs[eventName][i] === callback){
	                this._cbs[eventName].splice(i,1);
	                i -=1;
	                len -= 1;
	            }
	            i += 1;
	        }
	        if(!this._cbs[eventName].length){
	            this._cbs[eventName] = null;
	        }
	    }
	}
};
var createTypedArray = (function(){
	function createRegularArray(type, len){
		var i = 0, arr = [], value;
		switch(type) {
			case 'int16':
			case 'uint8c':
				value = 1;
				break;
			default:
				value = 1.1;
				break;
		}
		for(i = 0; i < len; i += 1) {
			arr.push(value);
		}
		return arr;
	}
	function createTypedArray(type, len){
		if(type === 'float32') {
			return new Float32Array(len);
		} else if(type === 'int16') {
			return new Int16Array(len);
		} else if(type === 'uint8c') {
			return new Uint8ClampedArray(len);
		}
	}
	if(typeof Uint8ClampedArray === 'function' && typeof Float32Array === 'function') {
		return createTypedArray;
	} else {
		return createRegularArray;
	}
}());

function createSizedArray(len) {
	return Array.apply(null,{length:len});
}
function createNS(type) {
	//return {appendChild:function(){},setAttribute:function(){},style:{}}
	return document.createElementNS(svgNS, type);
}
function createTag(type) {
	//return {appendChild:function(){},setAttribute:function(){},style:{}}
	return document.createElement(type);
}
function DynamicPropertyContainer(){};
DynamicPropertyContainer.prototype = {
	addDynamicProperty: function(prop) {
		if(this.dynamicProperties.indexOf(prop) === -1) {
	        this.dynamicProperties.push(prop);
	        this.container.addDynamicProperty(this);
	    	this._isAnimated = true;
	    }
	},
	iterateDynamicProperties: function(){
	    this._mdf = false;
	    var i, len = this.dynamicProperties.length;
	    for(i=0;i<len;i+=1){
	        this.dynamicProperties[i].getValue();
	        if(this.dynamicProperties[i]._mdf) {
	            this._mdf = true;
	        }
	    }
	},
	initDynamicPropertyContainer: function(container){
	    this.container = container;
	    this.dynamicProperties = [];
	    this._mdf = false;
	    this._isAnimated = false;
	}
}
var getBlendMode = (function() {

	var blendModeEnums = {
        0:'source-over',
        1:'multiply',
        2:'screen',
        3:'overlay',
        4:'darken',
        5:'lighten',
        6:'color-dodge',
        7:'color-burn',
        8:'hard-light',
        9:'soft-light',
        10:'difference',
        11:'exclusion',
        12:'hue',
        13:'saturation',
        14:'color',
        15:'luminosity'
    }

	return function(mode) {
		return blendModeEnums[mode] || '';
	}
}())
/*!
 Transformation Matrix v2.0
 (c) Epistemex 2014-2015
 www.epistemex.com
 By Ken Fyrstenberg
 Contributions by leeoniya.
 License: MIT, header required.
 */

/**
 * 2D transformation matrix object initialized with identity matrix.
 *
 * The matrix can synchronize a canvas context by supplying the context
 * as an argument, or later apply current absolute transform to an
 * existing context.
 *
 * All values are handled as floating point values.
 *
 * @param {CanvasRenderingContext2D} [context] - Optional context to sync with Matrix
 * @prop {number} a - scale x
 * @prop {number} b - shear y
 * @prop {number} c - shear x
 * @prop {number} d - scale y
 * @prop {number} e - translate x
 * @prop {number} f - translate y
 * @prop {CanvasRenderingContext2D|null} [context=null] - set or get current canvas context
 * @constructor
 */

var Matrix = (function(){

    var _cos = Math.cos;
    var _sin = Math.sin;
    var _tan = Math.tan;
    var _rnd = Math.round;

    function reset(){
        this.props[0] = 1;
        this.props[1] = 0;
        this.props[2] = 0;
        this.props[3] = 0;
        this.props[4] = 0;
        this.props[5] = 1;
        this.props[6] = 0;
        this.props[7] = 0;
        this.props[8] = 0;
        this.props[9] = 0;
        this.props[10] = 1;
        this.props[11] = 0;
        this.props[12] = 0;
        this.props[13] = 0;
        this.props[14] = 0;
        this.props[15] = 1;
        return this;
    }

    function rotate(angle) {
        if(angle === 0){
            return this;
        }
        var mCos = _cos(angle);
        var mSin = _sin(angle);
        return this._t(mCos, -mSin,  0, 0, mSin,  mCos, 0, 0, 0,  0,  1, 0, 0, 0, 0, 1);
    }

    function rotateX(angle){
        if(angle === 0){
            return this;
        }
        var mCos = _cos(angle);
        var mSin = _sin(angle);
        return this._t(1, 0, 0, 0, 0, mCos, -mSin, 0, 0, mSin,  mCos, 0, 0, 0, 0, 1);
    }

    function rotateY(angle){
        if(angle === 0){
            return this;
        }
        var mCos = _cos(angle);
        var mSin = _sin(angle);
        return this._t(mCos,  0,  mSin, 0, 0, 1, 0, 0, -mSin,  0,  mCos, 0, 0, 0, 0, 1);
    }

    function rotateZ(angle){
        if(angle === 0){
            return this;
        }
        var mCos = _cos(angle);
        var mSin = _sin(angle);
        return this._t(mCos, -mSin,  0, 0, mSin,  mCos, 0, 0, 0,  0,  1, 0, 0, 0, 0, 1);
    }

    function shear(sx,sy){
        return this._t(1, sy, sx, 1, 0, 0);
    }

    function skew(ax, ay){
        return this.shear(_tan(ax), _tan(ay));
    }

    function skewFromAxis(ax, angle){
        var mCos = _cos(angle);
        var mSin = _sin(angle);
        return this._t(mCos, mSin,  0, 0, -mSin,  mCos, 0, 0, 0,  0,  1, 0, 0, 0, 0, 1)
            ._t(1, 0,  0, 0, _tan(ax),  1, 0, 0, 0,  0,  1, 0, 0, 0, 0, 1)
            ._t(mCos, -mSin,  0, 0, mSin,  mCos, 0, 0, 0,  0,  1, 0, 0, 0, 0, 1);
        //return this._t(mCos, mSin, -mSin, mCos, 0, 0)._t(1, 0, _tan(ax), 1, 0, 0)._t(mCos, -mSin, mSin, mCos, 0, 0);
    }

    function scale(sx, sy, sz) {
        if(!sz && sz !== 0) {
            sz = 1;
        }
        if(sx === 1 && sy === 1 && sz === 1){
            return this;
        }
        return this._t(sx, 0, 0, 0, 0, sy, 0, 0, 0, 0, sz, 0, 0, 0, 0, 1);
    }

    function setTransform(a, b, c, d, e, f, g, h, i, j, k, l, m, n, o, p) {
        this.props[0] = a;
        this.props[1] = b;
        this.props[2] = c;
        this.props[3] = d;
        this.props[4] = e;
        this.props[5] = f;
        this.props[6] = g;
        this.props[7] = h;
        this.props[8] = i;
        this.props[9] = j;
        this.props[10] = k;
        this.props[11] = l;
        this.props[12] = m;
        this.props[13] = n;
        this.props[14] = o;
        this.props[15] = p;
        return this;
    }

    function translate(tx, ty, tz) {
        tz = tz || 0;
        if(tx !== 0 || ty !== 0 || tz !== 0){
            return this._t(1,0,0,0,0,1,0,0,0,0,1,0,tx,ty,tz,1);
        }
        return this;
    }

    function transform(a2, b2, c2, d2, e2, f2, g2, h2, i2, j2, k2, l2, m2, n2, o2, p2) {

        var _p = this.props;

        if(a2 === 1 && b2 === 0 && c2 === 0 && d2 === 0 && e2 === 0 && f2 === 1 && g2 === 0 && h2 === 0 && i2 === 0 && j2 === 0 && k2 === 1 && l2 === 0){
            //NOTE: commenting this condition because TurboFan deoptimizes code when present
            //if(m2 !== 0 || n2 !== 0 || o2 !== 0){
                _p[12] = _p[12] * a2 + _p[15] * m2;
                _p[13] = _p[13] * f2 + _p[15] * n2;
                _p[14] = _p[14] * k2 + _p[15] * o2;
                _p[15] = _p[15] * p2;
            //}
            this._identityCalculated = false;
            return this;
        }

        var a1 = _p[0];
        var b1 = _p[1];
        var c1 = _p[2];
        var d1 = _p[3];
        var e1 = _p[4];
        var f1 = _p[5];
        var g1 = _p[6];
        var h1 = _p[7];
        var i1 = _p[8];
        var j1 = _p[9];
        var k1 = _p[10];
        var l1 = _p[11];
        var m1 = _p[12];
        var n1 = _p[13];
        var o1 = _p[14];
        var p1 = _p[15];

        /* matrix order (canvas compatible):
         * ace
         * bdf
         * 001
         */
        _p[0] = a1 * a2 + b1 * e2 + c1 * i2 + d1 * m2;
        _p[1] = a1 * b2 + b1 * f2 + c1 * j2 + d1 * n2 ;
        _p[2] = a1 * c2 + b1 * g2 + c1 * k2 + d1 * o2 ;
        _p[3] = a1 * d2 + b1 * h2 + c1 * l2 + d1 * p2 ;

        _p[4] = e1 * a2 + f1 * e2 + g1 * i2 + h1 * m2 ;
        _p[5] = e1 * b2 + f1 * f2 + g1 * j2 + h1 * n2 ;
        _p[6] = e1 * c2 + f1 * g2 + g1 * k2 + h1 * o2 ;
        _p[7] = e1 * d2 + f1 * h2 + g1 * l2 + h1 * p2 ;

        _p[8] = i1 * a2 + j1 * e2 + k1 * i2 + l1 * m2 ;
        _p[9] = i1 * b2 + j1 * f2 + k1 * j2 + l1 * n2 ;
        _p[10] = i1 * c2 + j1 * g2 + k1 * k2 + l1 * o2 ;
        _p[11] = i1 * d2 + j1 * h2 + k1 * l2 + l1 * p2 ;

        _p[12] = m1 * a2 + n1 * e2 + o1 * i2 + p1 * m2 ;
        _p[13] = m1 * b2 + n1 * f2 + o1 * j2 + p1 * n2 ;
        _p[14] = m1 * c2 + n1 * g2 + o1 * k2 + p1 * o2 ;
        _p[15] = m1 * d2 + n1 * h2 + o1 * l2 + p1 * p2 ;

        this._identityCalculated = false;
        return this;
    }

    function isIdentity() {
        if(!this._identityCalculated){
            this._identity = !(this.props[0] !== 1 || this.props[1] !== 0 || this.props[2] !== 0 || this.props[3] !== 0 || this.props[4] !== 0 || this.props[5] !== 1 || this.props[6] !== 0 || this.props[7] !== 0 || this.props[8] !== 0 || this.props[9] !== 0 || this.props[10] !== 1 || this.props[11] !== 0 || this.props[12] !== 0 || this.props[13] !== 0 || this.props[14] !== 0 || this.props[15] !== 1);
            this._identityCalculated = true;
        }
        return this._identity;
    }

    function equals(matr){
        var i = 0;
        while (i < 16) {
            if(matr.props[i] !== this.props[i]) {
                return false;
            }
            i+=1;
        }
        return true;
    }

    function clone(matr){
        var i;
        for(i=0;i<16;i+=1){
            matr.props[i] = this.props[i];
        }
    }

    function cloneFromProps(props){
        var i;
        for(i=0;i<16;i+=1){
            this.props[i] = props[i];
        }
    }

    function applyToPoint(x, y, z) {

        return {
            x: x * this.props[0] + y * this.props[4] + z * this.props[8] + this.props[12],
            y: x * this.props[1] + y * this.props[5] + z * this.props[9] + this.props[13],
            z: x * this.props[2] + y * this.props[6] + z * this.props[10] + this.props[14]
        };
        /*return {
         x: x * me.a + y * me.c + me.e,
         y: x * me.b + y * me.d + me.f
         };*/
    }
    function applyToX(x, y, z) {
        return x * this.props[0] + y * this.props[4] + z * this.props[8] + this.props[12];
    }
    function applyToY(x, y, z) {
        return x * this.props[1] + y * this.props[5] + z * this.props[9] + this.props[13];
    }
    function applyToZ(x, y, z) {
        return x * this.props[2] + y * this.props[6] + z * this.props[10] + this.props[14];
    }

    function inversePoint(pt) {
        var determinant = this.props[0] * this.props[5] - this.props[1] * this.props[4];
        var a = this.props[5]/determinant;
        var b = - this.props[1]/determinant;
        var c = - this.props[4]/determinant;
        var d = this.props[0]/determinant;
        var e = (this.props[4] * this.props[13] - this.props[5] * this.props[12])/determinant;
        var f = - (this.props[0] * this.props[13] - this.props[1] * this.props[12])/determinant;
        return [pt[0] * a + pt[1] * c + e, pt[0] * b + pt[1] * d + f, 0];
    }

    function inversePoints(pts){
        var i, len = pts.length, retPts = [];
        for(i=0;i<len;i+=1){
            retPts[i] = inversePoint(pts[i]);
        }
        return retPts;
    }

    function applyToTriplePoints(pt1, pt2, pt3) {
        var arr = createTypedArray('float32', 6);
        if(this.isIdentity()) {
            arr[0] = pt1[0];
            arr[1] = pt1[1];
            arr[2] = pt2[0];
            arr[3] = pt2[1];
            arr[4] = pt3[0];
            arr[5] = pt3[1];
        } else {
            var p0 = this.props[0], p1 = this.props[1], p4 = this.props[4], p5 = this.props[5], p12 = this.props[12], p13 = this.props[13];
            arr[0] = pt1[0] * p0 + pt1[1] * p4 + p12;
            arr[1] = pt1[0] * p1 + pt1[1] * p5 + p13;
            arr[2] = pt2[0] * p0 + pt2[1] * p4 + p12;
            arr[3] = pt2[0] * p1 + pt2[1] * p5 + p13;
            arr[4] = pt3[0] * p0 + pt3[1] * p4 + p12;
            arr[5] = pt3[0] * p1 + pt3[1] * p5 + p13;
        }
        return arr;
    }

    function applyToPointArray(x,y,z){
        var arr;
        if(this.isIdentity()) {
            arr = [x,y,z];
        } else {
            arr = [x * this.props[0] + y * this.props[4] + z * this.props[8] + this.props[12],x * this.props[1] + y * this.props[5] + z * this.props[9] + this.props[13],x * this.props[2] + y * this.props[6] + z * this.props[10] + this.props[14]];
        }
        return arr;
    }

    function applyToPointStringified(x, y) {
        if(this.isIdentity()) {
            return x + ',' + y;
        }
        var _p = this.props;
        return Math.round((x * _p[0] + y * _p[4] + _p[12]) * 100) / 100+','+ Math.round((x * _p[1] + y * _p[5] + _p[13]) * 100) / 100;
    }

    function toCSS() {
        //Doesn't make much sense to add this optimization. If it is an identity matrix, it's very likely this will get called only once since it won't be keyframed.
        /*if(this.isIdentity()) {
            return '';
        }*/
        var i = 0;
        var props = this.props;
        var cssValue = 'matrix3d(';
        var v = 10000;
        while(i<16){
            cssValue += _rnd(props[i]*v)/v;
            cssValue += i === 15 ? ')':',';
            i += 1;
        }
        return cssValue;
    }

    function roundMatrixProperty(val) {
        var v = 10000;
        if((val < 0.000001 && val > 0) || (val > -0.000001 && val < 0)) {
            return _rnd(val * v) / v;
        }
        return val;
    }

    function to2dCSS() {
        //Doesn't make much sense to add this optimization. If it is an identity matrix, it's very likely this will get called only once since it won't be keyframed.
        /*if(this.isIdentity()) {
            return '';
        }*/
        var props = this.props;
        var _a = roundMatrixProperty(props[0]);
        var _b = roundMatrixProperty(props[1]);
        var _c = roundMatrixProperty(props[4]);
        var _d = roundMatrixProperty(props[5]);
        var _e = roundMatrixProperty(props[12]);
        var _f = roundMatrixProperty(props[13]);
        return "matrix(" + _a + ',' + _b + ',' + _c + ',' + _d + ',' + _e + ',' + _f + ")";
    }

    return function(){
        this.reset = reset;
        this.rotate = rotate;
        this.rotateX = rotateX;
        this.rotateY = rotateY;
        this.rotateZ = rotateZ;
        this.skew = skew;
        this.skewFromAxis = skewFromAxis;
        this.shear = shear;
        this.scale = scale;
        this.setTransform = setTransform;
        this.translate = translate;
        this.transform = transform;
        this.applyToPoint = applyToPoint;
        this.applyToX = applyToX;
        this.applyToY = applyToY;
        this.applyToZ = applyToZ;
        this.applyToPointArray = applyToPointArray;
        this.applyToTriplePoints = applyToTriplePoints;
        this.applyToPointStringified = applyToPointStringified;
        this.toCSS = toCSS;
        this.to2dCSS = to2dCSS;
        this.clone = clone;
        this.cloneFromProps = cloneFromProps;
        this.equals = equals;
        this.inversePoints = inversePoints;
        this.inversePoint = inversePoint;
        this._t = this.transform;
        this.isIdentity = isIdentity;
        this._identity = true;
        this._identityCalculated = false;

        this.props = createTypedArray('float32', 16);
        this.reset();
    };
}());

/*
 Copyright 2014 David Bau.

 Permission is hereby granted, free of charge, to any person obtaining
 a copy of this software and associated documentation files (the
 "Software"), to deal in the Software without restriction, including
 without limitation the rights to use, copy, modify, merge, publish,
 distribute, sublicense, and/or sell copies of the Software, and to
 permit persons to whom the Software is furnished to do so, subject to
 the following conditions:

 The above copyright notice and this permission notice shall be
 included in all copies or substantial portions of the Software.

 THE SOFTWARE IS PROVIDED "AS IS", WITHOUT WARRANTY OF ANY KIND,
 EXPRESS OR IMPLIED, INCLUDING BUT NOT LIMITED TO THE WARRANTIES OF
 MERCHANTABILITY, FITNESS FOR A PARTICULAR PURPOSE AND NONINFRINGEMENT.
 IN NO EVENT SHALL THE AUTHORS OR COPYRIGHT HOLDERS BE LIABLE FOR ANY
 CLAIM, DAMAGES OR OTHER LIABILITY, WHETHER IN AN ACTION OF CONTRACT,
 TORT OR OTHERWISE, ARISING FROM, OUT OF OR IN CONNECTION WITH THE
 SOFTWARE OR THE USE OR OTHER DEALINGS IN THE SOFTWARE.

 */

(function (pool, math) {
//
// The following constants are related to IEEE 754 limits.
//
    var global = this,
        width = 256,        // each RC4 output is 0 <= x < 256
        chunks = 6,         // at least six RC4 outputs for each double
        digits = 52,        // there are 52 significant digits in a double
        rngname = 'random', // rngname: name for Math.random and Math.seedrandom
        startdenom = math.pow(width, chunks),
        significance = math.pow(2, digits),
        overflow = significance * 2,
        mask = width - 1,
        nodecrypto;         // node.js crypto module, initialized at the bottom.

//
// seedrandom()
// This is the seedrandom function described above.
//
    function seedrandom(seed, options, callback) {
        var key = [];
        options = (options === true) ? { entropy: true } : (options || {});

        // Flatten the seed string or build one from local entropy if needed.
        var shortseed = mixkey(flatten(
            options.entropy ? [seed, tostring(pool)] :
                (seed === null) ? autoseed() : seed, 3), key);

        // Use the seed to initialize an ARC4 generator.
        var arc4 = new ARC4(key);

        // This function returns a random double in [0, 1) that contains
        // randomness in every bit of the mantissa of the IEEE 754 value.
        var prng = function() {
            var n = arc4.g(chunks),             // Start with a numerator n < 2 ^ 48
                d = startdenom,                 //   and denominator d = 2 ^ 48.
                x = 0;                          //   and no 'extra last byte'.
            while (n < significance) {          // Fill up all significant digits by
                n = (n + x) * width;              //   shifting numerator and
                d *= width;                       //   denominator and generating a
                x = arc4.g(1);                    //   new least-significant-byte.
            }
            while (n >= overflow) {             // To avoid rounding up, before adding
                n /= 2;                           //   last byte, shift everything
                d /= 2;                           //   right using integer math until
                x >>>= 1;                         //   we have exactly the desired bits.
            }
            return (n + x) / d;                 // Form the number within [0, 1).
        };

        prng.int32 = function() { return arc4.g(4) | 0; };
        prng.quick = function() { return arc4.g(4) / 0x100000000; };
        prng.double = prng;

        // Mix the randomness into accumulated entropy.
        mixkey(tostring(arc4.S), pool);

        // Calling convention: what to return as a function of prng, seed, is_math.
        return (options.pass || callback ||
        function(prng, seed, is_math_call, state) {
            if (state) {
                // Load the arc4 state from the given state if it has an S array.
                if (state.S) { copy(state, arc4); }
                // Only provide the .state method if requested via options.state.
                prng.state = function() { return copy(arc4, {}); };
            }

            // If called as a method of Math (Math.seedrandom()), mutate
            // Math.random because that is how seedrandom.js has worked since v1.0.
            if (is_math_call) { math[rngname] = prng; return seed; }

            // Otherwise, it is a newer calling convention, so return the
            // prng directly.
            else return prng;
        })(
            prng,
            shortseed,
            'global' in options ? options.global : (this == math),
            options.state);
    }
    math['seed' + rngname] = seedrandom;

//
// ARC4
//
// An ARC4 implementation.  The constructor takes a key in the form of
// an array of at most (width) integers that should be 0 <= x < (width).
//
// The g(count) method returns a pseudorandom integer that concatenates
// the next (count) outputs from ARC4.  Its return value is a number x
// that is in the range 0 <= x < (width ^ count).
//
    function ARC4(key) {
        var t, keylen = key.length,
            me = this, i = 0, j = me.i = me.j = 0, s = me.S = [];

        // The empty key [] is treated as [0].
        if (!keylen) { key = [keylen++]; }

        // Set up S using the standard key scheduling algorithm.
        while (i < width) {
            s[i] = i++;
        }
        for (i = 0; i < width; i++) {
            s[i] = s[j = mask & (j + key[i % keylen] + (t = s[i]))];
            s[j] = t;
        }

        // The "g" method returns the next (count) outputs as one number.
        me.g = function(count) {
            // Using instance members instead of closure state nearly doubles speed.
            var t, r = 0,
                i = me.i, j = me.j, s = me.S;
            while (count--) {
                t = s[i = mask & (i + 1)];
                r = r * width + s[mask & ((s[i] = s[j = mask & (j + t)]) + (s[j] = t))];
            }
            me.i = i; me.j = j;
            return r;
            // For robust unpredictability, the function call below automatically
            // discards an initial batch of values.  This is called RC4-drop[256].
            // See http://google.com/search?q=rsa+fluhrer+response&btnI
        };
    }

//
// copy()
// Copies internal state of ARC4 to or from a plain object.
//
    function copy(f, t) {
        t.i = f.i;
        t.j = f.j;
        t.S = f.S.slice();
        return t;
    }

//
// flatten()
// Converts an object tree to nested arrays of strings.
//
    function flatten(obj, depth) {
        var result = [], typ = (typeof obj), prop;
        if (depth && typ == 'object') {
            for (prop in obj) {
                try { result.push(flatten(obj[prop], depth - 1)); } catch (e) {}
            }
        }
        return (result.length ? result : typ == 'string' ? obj : obj + '\0');
    }

//
// mixkey()
// Mixes a string seed into a key that is an array of integers, and
// returns a shortened string seed that is equivalent to the result key.
//
    function mixkey(seed, key) {
        var stringseed = seed + '', smear, j = 0;
        while (j < stringseed.length) {
            key[mask & j] =
                mask & ((smear ^= key[mask & j] * 19) + stringseed.charCodeAt(j++));
        }
        return tostring(key);
    }

//
// autoseed()
// Returns an object for autoseeding, using window.crypto and Node crypto
// module if available.
//
    function autoseed() {
        try {
            if (nodecrypto) { return tostring(nodecrypto.randomBytes(width)); }
            var out = new Uint8Array(width);
            (global.crypto || global.msCrypto).getRandomValues(out);
            return tostring(out);
        } catch (e) {
            var browser = global.navigator,
                plugins = browser && browser.plugins;
            return [+new Date(), global, plugins, global.screen, tostring(pool)];
        }
    }

//
// tostring()
// Converts an array of charcodes to a string
//
    function tostring(a) {
        return String.fromCharCode.apply(0, a);
    }

//
// When seedrandom.js is loaded, we immediately mix a few bits
// from the built-in RNG into the entropy pool.  Because we do
// not want to interfere with deterministic PRNG state later,
// seedrandom will not call math.random on its own again after
// initialization.
//
    mixkey(math.random(), pool);

//
// Nodejs and AMD support: export the implementation as a module using
// either convention.
//

// End anonymous scope, and pass initial values.
})(
    [],     // pool: entropy pool starts empty
    BMMath    // math: package containing random, pow, and seedrandom
);
var BezierFactory = (function(){
    /**
     * BezierEasing - use bezier curve for transition easing function
     * by Gaëtan Renaudeau 2014 - 2015 – MIT License
     *
     * Credits: is based on Firefox's nsSMILKeySpline.cpp
     * Usage:
     * var spline = BezierEasing([ 0.25, 0.1, 0.25, 1.0 ])
     * spline.get(x) => returns the easing value | x must be in [0, 1] range
     *
     */

        var ob = {};
    ob.getBezierEasing = getBezierEasing;
    var beziers = {};

    function getBezierEasing(a,b,c,d,nm){
        var str = nm || ('bez_' + a+'_'+b+'_'+c+'_'+d).replace(/\./g, 'p');
        if(beziers[str]){
            return beziers[str];
        }
        var bezEasing = new BezierEasing([a,b,c,d]);
        beziers[str] = bezEasing;
        return bezEasing;
    }

// These values are established by empiricism with tests (tradeoff: performance VS precision)
    var NEWTON_ITERATIONS = 4;
    var NEWTON_MIN_SLOPE = 0.001;
    var SUBDIVISION_PRECISION = 0.0000001;
    var SUBDIVISION_MAX_ITERATIONS = 10;

    var kSplineTableSize = 11;
    var kSampleStepSize = 1.0 / (kSplineTableSize - 1.0);

    var float32ArraySupported = typeof Float32Array === "function";

    function A (aA1, aA2) { return 1.0 - 3.0 * aA2 + 3.0 * aA1; }
    function B (aA1, aA2) { return 3.0 * aA2 - 6.0 * aA1; }
    function C (aA1)      { return 3.0 * aA1; }

// Returns x(t) given t, x1, and x2, or y(t) given t, y1, and y2.
    function calcBezier (aT, aA1, aA2) {
        return ((A(aA1, aA2)*aT + B(aA1, aA2))*aT + C(aA1))*aT;
    }

// Returns dx/dt given t, x1, and x2, or dy/dt given t, y1, and y2.
    function getSlope (aT, aA1, aA2) {
        return 3.0 * A(aA1, aA2)*aT*aT + 2.0 * B(aA1, aA2) * aT + C(aA1);
    }

    function binarySubdivide (aX, aA, aB, mX1, mX2) {
        var currentX, currentT, i = 0;
        do {
            currentT = aA + (aB - aA) / 2.0;
            currentX = calcBezier(currentT, mX1, mX2) - aX;
            if (currentX > 0.0) {
                aB = currentT;
            } else {
                aA = currentT;
            }
        } while (Math.abs(currentX) > SUBDIVISION_PRECISION && ++i < SUBDIVISION_MAX_ITERATIONS);
        return currentT;
    }

    function newtonRaphsonIterate (aX, aGuessT, mX1, mX2) {
        for (var i = 0; i < NEWTON_ITERATIONS; ++i) {
            var currentSlope = getSlope(aGuessT, mX1, mX2);
            if (currentSlope === 0.0) return aGuessT;
            var currentX = calcBezier(aGuessT, mX1, mX2) - aX;
            aGuessT -= currentX / currentSlope;
        }
        return aGuessT;
    }

    /**
     * points is an array of [ mX1, mY1, mX2, mY2 ]
     */
    function BezierEasing (points) {
        this._p = points;
        this._mSampleValues = float32ArraySupported ? new Float32Array(kSplineTableSize) : new Array(kSplineTableSize);
        this._precomputed = false;

        this.get = this.get.bind(this);
    }

    BezierEasing.prototype = {

        get: function (x) {
            var mX1 = this._p[0],
                mY1 = this._p[1],
                mX2 = this._p[2],
                mY2 = this._p[3];
            if (!this._precomputed) this._precompute();
            if (mX1 === mY1 && mX2 === mY2) return x; // linear
            // Because JavaScript number are imprecise, we should guarantee the extremes are right.
            if (x === 0) return 0;
            if (x === 1) return 1;
            return calcBezier(this._getTForX(x), mY1, mY2);
        },

        // Private part

        _precompute: function () {
            var mX1 = this._p[0],
                mY1 = this._p[1],
                mX2 = this._p[2],
                mY2 = this._p[3];
            this._precomputed = true;
            if (mX1 !== mY1 || mX2 !== mY2)
                this._calcSampleValues();
        },

        _calcSampleValues: function () {
            var mX1 = this._p[0],
                mX2 = this._p[2];
            for (var i = 0; i < kSplineTableSize; ++i) {
                this._mSampleValues[i] = calcBezier(i * kSampleStepSize, mX1, mX2);
            }
        },

        /**
         * getTForX chose the fastest heuristic to determine the percentage value precisely from a given X projection.
         */
        _getTForX: function (aX) {
            var mX1 = this._p[0],
                mX2 = this._p[2],
                mSampleValues = this._mSampleValues;

            var intervalStart = 0.0;
            var currentSample = 1;
            var lastSample = kSplineTableSize - 1;

            for (; currentSample !== lastSample && mSampleValues[currentSample] <= aX; ++currentSample) {
                intervalStart += kSampleStepSize;
            }
            --currentSample;

            // Interpolate to provide an initial guess for t
            var dist = (aX - mSampleValues[currentSample]) / (mSampleValues[currentSample+1] - mSampleValues[currentSample]);
            var guessForT = intervalStart + dist * kSampleStepSize;

            var initialSlope = getSlope(guessForT, mX1, mX2);
            if (initialSlope >= NEWTON_MIN_SLOPE) {
                return newtonRaphsonIterate(aX, guessForT, mX1, mX2);
            } else if (initialSlope === 0.0) {
                return guessForT;
            } else {
                return binarySubdivide(aX, intervalStart, intervalStart + kSampleStepSize, mX1, mX2);
            }
        }
    };

    return ob;

}());
(function () {
    var lastTime = 0;
    var vendors = ['ms', 'moz', 'webkit', 'o'];
    for(var x = 0; x < vendors.length && !window.requestAnimationFrame; ++x) {
        window.requestAnimationFrame = window[vendors[x] + 'RequestAnimationFrame'];
        window.cancelAnimationFrame = window[vendors[x] + 'CancelAnimationFrame'] || window[vendors[x] + 'CancelRequestAnimationFrame'];
    }
    if(!window.requestAnimationFrame)
        window.requestAnimationFrame = function (callback, element) {
            var currTime = new Date().getTime();
            var timeToCall = Math.max(0, 16 - (currTime - lastTime));
            var id = setTimeout(function () {
                    callback(currTime + timeToCall);
                },
                timeToCall);
            lastTime = currTime + timeToCall;
            return id;
        };
    if(!window.cancelAnimationFrame)
        window.cancelAnimationFrame = function (id) {
            clearTimeout(id);
        };
}());

function extendPrototype(sources,destination){
    var i, len = sources.length, sourcePrototype;
    for (i = 0;i < len;i += 1) {
        sourcePrototype = sources[i].prototype;
        for (var attr in sourcePrototype) {
            if (sourcePrototype.hasOwnProperty(attr)) destination.prototype[attr] = sourcePrototype[attr];
        }
    }
}

function getDescriptor(object, prop) {
    return Object.getOwnPropertyDescriptor(object, prop);
}

function createProxyFunction(prototype) {
	function ProxyFunction(){}
	ProxyFunction.prototype = prototype;
	return ProxyFunction;
}
function bezFunction(){

    var easingFunctions = [];
    var math = Math;

    function pointOnLine2D(x1,y1, x2,y2, x3,y3){
        var det1 = (x1*y2) + (y1*x3) + (x2*y3) - (x3*y2) - (y3*x1) - (x2*y1);
        return det1 > -0.001 && det1 < 0.001;
    }

    function pointOnLine3D(x1,y1,z1, x2,y2,z2, x3,y3,z3){
        if(z1 === 0 && z2 === 0 && z3 === 0) {
            return pointOnLine2D(x1,y1, x2,y2, x3,y3);
        }
        var dist1 = Math.sqrt(Math.pow(x2 - x1, 2) + Math.pow(y2 - y1, 2) + Math.pow(z2 - z1, 2));
        var dist2 = Math.sqrt(Math.pow(x3 - x1, 2) + Math.pow(y3 - y1, 2) + Math.pow(z3 - z1, 2));
        var dist3 = Math.sqrt(Math.pow(x3 - x2, 2) + Math.pow(y3 - y2, 2) + Math.pow(z3 - z2, 2));
        var diffDist;
        if(dist1 > dist2){
            if(dist1 > dist3){
                diffDist = dist1 - dist2 - dist3;
            } else {
                diffDist = dist3 - dist2 - dist1;
            }
        } else if(dist3 > dist2){
            diffDist = dist3 - dist2 - dist1;
        } else {
            diffDist = dist2 - dist1 - dist3;
        }
        return diffDist > -0.0001 && diffDist < 0.0001;
    }

    var getBezierLength = (function(){

        return function(pt1,pt2,pt3,pt4){
            var curveSegments = defaultCurveSegments;
            var k;
            var i, len;
            var ptCoord,perc,addedLength = 0;
            var ptDistance;
            var point = [],lastPoint = [];
            var lengthData = bezier_length_pool.newElement();
            len = pt3.length;
            for(k=0;k<curveSegments;k+=1){
                perc = k/(curveSegments-1);
                ptDistance = 0;
                for(i=0;i<len;i+=1){
                    ptCoord = bm_pow(1-perc,3)*pt1[i]+3*bm_pow(1-perc,2)*perc*pt3[i]+3*(1-perc)*bm_pow(perc,2)*pt4[i]+bm_pow(perc,3)*pt2[i];
                    point[i] = ptCoord;
                    if(lastPoint[i] !== null){
                        ptDistance += bm_pow(point[i] - lastPoint[i],2);
                    }
                    lastPoint[i] = point[i];
                }
                if(ptDistance){
                    ptDistance = bm_sqrt(ptDistance);
                    addedLength += ptDistance;
                }
                lengthData.percents[k] = perc;
                lengthData.lengths[k] = addedLength;
            }
            lengthData.addedLength = addedLength;
            return lengthData;
        };
    }());

    function getSegmentsLength(shapeData) {
        var segmentsLength = segments_length_pool.newElement();
        var closed = shapeData.c;
        var pathV = shapeData.v;
        var pathO = shapeData.o;
        var pathI = shapeData.i;
        var i, len = shapeData._length;
        var lengths = segmentsLength.lengths;
        var totalLength = 0;
        for(i=0;i<len-1;i+=1){
            lengths[i] = getBezierLength(pathV[i],pathV[i+1],pathO[i],pathI[i+1]);
            totalLength += lengths[i].addedLength;
        }
        if(closed && len){
            lengths[i] = getBezierLength(pathV[i],pathV[0],pathO[i],pathI[0]);
            totalLength += lengths[i].addedLength;
        }
        segmentsLength.totalLength = totalLength;
        return segmentsLength;
    }

    function BezierData(length){
        this.segmentLength = 0;
        this.points = new Array(length);
    }

    function PointData(partial,point){
        this.partialLength = partial;
        this.point = point;
    }

    var buildBezierData = (function(){

        var storedData = {};

        return function (pt1, pt2, pt3, pt4){
            var bezierName = (pt1[0]+'_'+pt1[1]+'_'+pt2[0]+'_'+pt2[1]+'_'+pt3[0]+'_'+pt3[1]+'_'+pt4[0]+'_'+pt4[1]).replace(/\./g, 'p');
            if(!storedData[bezierName]){
                var curveSegments = defaultCurveSegments;
                var k, i, len;
                var ptCoord,perc,addedLength = 0;
                var ptDistance;
                var point,lastPoint = null;
                if (pt1.length === 2 && (pt1[0] != pt2[0] || pt1[1] != pt2[1]) && pointOnLine2D(pt1[0],pt1[1],pt2[0],pt2[1],pt1[0]+pt3[0],pt1[1]+pt3[1]) && pointOnLine2D(pt1[0],pt1[1],pt2[0],pt2[1],pt2[0]+pt4[0],pt2[1]+pt4[1])){
                    curveSegments = 2;
                }
                var bezierData = new BezierData(curveSegments);
                len = pt3.length;
                for (k = 0; k < curveSegments; k += 1) {
                    point = createSizedArray(len);
                    perc = k / (curveSegments - 1);
                    ptDistance = 0;
                    for (i = 0; i < len; i += 1){
                        ptCoord = bm_pow(1-perc,3)*pt1[i]+3*bm_pow(1-perc,2)*perc*(pt1[i] + pt3[i])+3*(1-perc)*bm_pow(perc,2)*(pt2[i] + pt4[i])+bm_pow(perc,3)*pt2[i];
                        point[i] = ptCoord;
                        if(lastPoint !== null){
                            ptDistance += bm_pow(point[i] - lastPoint[i],2);
                        }
                    }
                    ptDistance = bm_sqrt(ptDistance);
                    addedLength += ptDistance;
                    bezierData.points[k] = new PointData(ptDistance, point);
                    lastPoint = point;
                }
                bezierData.segmentLength = addedLength;
                storedData[bezierName] = bezierData;
            }
            return storedData[bezierName];
        };
    }());

    function getDistancePerc(perc,bezierData){
        var percents = bezierData.percents;
        var lengths = bezierData.lengths;
        var len = percents.length;
        var initPos = bm_floor((len-1)*perc);
        var lengthPos = perc*bezierData.addedLength;
        var lPerc = 0;
        if(initPos === len - 1 || initPos === 0 || lengthPos === lengths[initPos]){
            return percents[initPos];
        }else{
            var dir = lengths[initPos] > lengthPos ? -1 : 1;
            var flag = true;
            while(flag){
                if(lengths[initPos] <= lengthPos && lengths[initPos+1] > lengthPos){
                    lPerc = (lengthPos - lengths[initPos]) / (lengths[initPos+1] - lengths[initPos]);
                    flag = false;
                }else{
                    initPos += dir;
                }
                if(initPos < 0 || initPos >= len - 1){
                    //FIX for TypedArrays that don't store floating point values with enough accuracy
                    if(initPos === len - 1) {
                        return percents[initPos];
                    }
                    flag = false;
                }
            }
            return percents[initPos] + (percents[initPos+1] - percents[initPos])*lPerc;
        }
    }

    function getPointInSegment(pt1, pt2, pt3, pt4, percent, bezierData) {
        var t1 = getDistancePerc(percent,bezierData);
        var u0 = 1;
        var u1 = 1 - t1;
        var ptX = Math.round((u1*u1*u1* pt1[0] + (t1*u1*u1 + u1*t1*u1 + u1*u1*t1)* pt3[0] + (t1*t1*u1 + u1*t1*t1 + t1*u1*t1)*pt4[0] + t1*t1*t1* pt2[0])* 1000) / 1000;
        var ptY = Math.round((u1*u1*u1* pt1[1] + (t1*u1*u1 + u1*t1*u1 + u1*u1*t1)* pt3[1] + (t1*t1*u1 + u1*t1*t1 + t1*u1*t1)*pt4[1] + t1*t1*t1* pt2[1])* 1000) / 1000;
        return [ptX, ptY];
    }

    function getSegmentArray() {

    }

    var bezier_segment_points = createTypedArray('float32', 8);

    function getNewSegment(pt1,pt2,pt3,pt4,startPerc,endPerc, bezierData){

        startPerc = startPerc < 0 ? 0 : startPerc > 1 ? 1 : startPerc;
        var t0 = getDistancePerc(startPerc,bezierData);
        endPerc = endPerc > 1 ? 1 : endPerc;
        var t1 = getDistancePerc(endPerc,bezierData);
        var i, len = pt1.length;
        var u0 = 1 - t0;
        var u1 = 1 - t1;
        var u0u0u0 = u0*u0*u0;
        var t0u0u0_3 = t0*u0*u0*3;
        var t0t0u0_3 = t0*t0*u0*3;
        var t0t0t0 = t0*t0*t0;
        //
        var u0u0u1 = u0*u0*u1;
        var t0u0u1_3 = t0*u0*u1 + u0*t0*u1 + u0*u0*t1;
        var t0t0u1_3 = t0*t0*u1 + u0*t0*t1 + t0*u0*t1;
        var t0t0t1 = t0*t0*t1;
        //
        var u0u1u1 = u0*u1*u1;
        var t0u1u1_3 = t0*u1*u1 + u0*t1*u1 + u0*u1*t1;
        var t0t1u1_3 = t0*t1*u1 + u0*t1*t1 + t0*u1*t1;
        var t0t1t1 = t0*t1*t1;
        //
        var u1u1u1 = u1*u1*u1;
        var t1u1u1_3 = t1*u1*u1 + u1*t1*u1 + u1*u1*t1;
        var t1t1u1_3 = t1*t1*u1 + u1*t1*t1 + t1*u1*t1;
        var t1t1t1 = t1*t1*t1;
        for(i=0;i<len;i+=1){
            bezier_segment_points[i * 4] = Math.round((u0u0u0 * pt1[i] + t0u0u0_3 * pt3[i] + t0t0u0_3 * pt4[i] + t0t0t0 * pt2[i]) * 1000) / 1000;
            bezier_segment_points[i * 4 + 1] = Math.round((u0u0u1 * pt1[i] + t0u0u1_3 * pt3[i] + t0t0u1_3 * pt4[i] + t0t0t1 * pt2[i]) * 1000) / 1000;
            bezier_segment_points[i * 4 + 2] = Math.round((u0u1u1 * pt1[i] + t0u1u1_3 * pt3[i] + t0t1u1_3 * pt4[i] + t0t1t1 * pt2[i]) * 1000) / 1000;
            bezier_segment_points[i * 4 + 3] = Math.round((u1u1u1 * pt1[i] + t1u1u1_3 * pt3[i] + t1t1u1_3 * pt4[i] + t1t1t1 * pt2[i]) * 1000) / 1000;
        }

        return bezier_segment_points;
    }

    return {
        getSegmentsLength : getSegmentsLength,
        getNewSegment : getNewSegment,
        getPointInSegment : getPointInSegment,
        buildBezierData : buildBezierData,
        pointOnLine2D : pointOnLine2D,
        pointOnLine3D : pointOnLine3D
    };
}

var bez = bezFunction();
function dataFunctionManager(){

    //var tCanvasHelper = createTag('canvas').getContext('2d');

    function completeLayers(layers, comps, fontManager){
        var layerData;
        var animArray, lastFrame;
        var i, len = layers.length;
        var j, jLen, k, kLen;
        for(i=0;i<len;i+=1){
            layerData = layers[i];
            if(!('ks' in layerData) || layerData.completed){
                continue;
            }
            layerData.completed = true;
            if(layerData.tt){
                layers[i-1].td = layerData.tt;
            }
            animArray = [];
            lastFrame = -1;
            if(layerData.hasMask){
                var maskProps = layerData.masksProperties;
                jLen = maskProps.length;
                for(j=0;j<jLen;j+=1){
                    if(maskProps[j].pt.k.i){
                        convertPathsToAbsoluteValues(maskProps[j].pt.k);
                    }else{
                        kLen = maskProps[j].pt.k.length;
                        for(k=0;k<kLen;k+=1){
                            if(maskProps[j].pt.k[k].s){
                                convertPathsToAbsoluteValues(maskProps[j].pt.k[k].s[0]);
                            }
                            if(maskProps[j].pt.k[k].e){
                                convertPathsToAbsoluteValues(maskProps[j].pt.k[k].e[0]);
                            }
                        }
                    }
                }
            }
            if(layerData.ty===0){
                layerData.layers = findCompLayers(layerData.refId, comps);
                completeLayers(layerData.layers,comps, fontManager);
            }else if(layerData.ty === 4){
                completeShapes(layerData.shapes);
            }else if(layerData.ty == 5){
                completeText(layerData, fontManager);
            }
        }
    }

    function findCompLayers(id,comps){
        var i = 0, len = comps.length;
        while(i<len){
            if(comps[i].id === id){
                if(!comps[i].layers.__used) {
                    comps[i].layers.__used = true;
                    return comps[i].layers;
                }
                return JSON.parse(JSON.stringify(comps[i].layers));
            }
            i += 1;
        }
    }

    function completeShapes(arr){
        var i, len = arr.length;
        var j, jLen;
        var hasPaths = false;
        for(i=len-1;i>=0;i-=1){
            if(arr[i].ty == 'sh'){
                if(arr[i].ks.k.i){
                    convertPathsToAbsoluteValues(arr[i].ks.k);
                }else{
                    jLen = arr[i].ks.k.length;
                    for(j=0;j<jLen;j+=1){
                        if(arr[i].ks.k[j].s){
                            convertPathsToAbsoluteValues(arr[i].ks.k[j].s[0]);
                        }
                        if(arr[i].ks.k[j].e){
                            convertPathsToAbsoluteValues(arr[i].ks.k[j].e[0]);
                        }
                    }
                }
                hasPaths = true;
            }else if(arr[i].ty == 'gr'){
                completeShapes(arr[i].it);
            }
        }
        /*if(hasPaths){
            //mx: distance
            //ss: sensitivity
            //dc: decay
            arr.splice(arr.length-1,0,{
                "ty": "ms",
                "mx":20,
                "ss":10,
                 "dc":0.001,
                "maxDist":200
            });
        }*/
    }

    function convertPathsToAbsoluteValues(path){
        var i, len = path.i.length;
        for(i=0;i<len;i+=1){
            path.i[i][0] += path.v[i][0];
            path.i[i][1] += path.v[i][1];
            path.o[i][0] += path.v[i][0];
            path.o[i][1] += path.v[i][1];
        }
    }

    function checkVersion(minimum,animVersionString){
        var animVersion = animVersionString ? animVersionString.split('.') : [100,100,100];
        if(minimum[0]>animVersion[0]){
            return true;
        } else if(animVersion[0] > minimum[0]){
            return false;
        }
        if(minimum[1]>animVersion[1]){
            return true;
        } else if(animVersion[1] > minimum[1]){
            return false;
        }
        if(minimum[2]>animVersion[2]){
            return true;
        } else if(animVersion[2] > minimum[2]){
            return false;
        }
    }

    var checkText = (function(){
        var minimumVersion = [4,4,14];

        function updateTextLayer(textLayer){
            var documentData = textLayer.t.d;
            textLayer.t.d = {
                k: [
                    {
                        s:documentData,
                        t:0
                    }
                ]
            };
        }

        function iterateLayers(layers){
            var i, len = layers.length;
            for(i=0;i<len;i+=1){
                if(layers[i].ty === 5){
                    updateTextLayer(layers[i]);
                }
            }
        }

        return function (animationData){
            if(checkVersion(minimumVersion,animationData.v)){
                iterateLayers(animationData.layers);
                if(animationData.assets){
                    var i, len = animationData.assets.length;
                    for(i=0;i<len;i+=1){
                        if(animationData.assets[i].layers){
                            iterateLayers(animationData.assets[i].layers);

                        }
                    }
                }
            }
        };
    }());

    var checkChars = (function() {
        var minimumVersion = [4,7,99];
        return function (animationData){
            if(animationData.chars && !checkVersion(minimumVersion,animationData.v)){
                var i, len = animationData.chars.length, j, jLen, k, kLen;
                var pathData, paths;
                for(i = 0; i < len; i += 1) {
                    if(animationData.chars[i].data && animationData.chars[i].data.shapes) {
                        paths = animationData.chars[i].data.shapes[0].it;
                        jLen = paths.length;

                        for(j = 0; j < jLen; j += 1) {
                            pathData = paths[j].ks.k;
                            if(!pathData.__converted) {
                                convertPathsToAbsoluteValues(paths[j].ks.k);
                                pathData.__converted = true;
                            }
                        }
                    }
                }
            }
        };
    }());

    var checkColors = (function(){
        var minimumVersion = [4,1,9];

        function iterateShapes(shapes){
            var i, len = shapes.length;
            var j, jLen;
            for(i=0;i<len;i+=1){
                if(shapes[i].ty === 'gr'){
                    iterateShapes(shapes[i].it);
                }else if(shapes[i].ty === 'fl' || shapes[i].ty === 'st'){
                    if(shapes[i].c.k && shapes[i].c.k[0].i){
                        jLen = shapes[i].c.k.length;
                        for(j=0;j<jLen;j+=1){
                            if(shapes[i].c.k[j].s){
                                shapes[i].c.k[j].s[0] /= 255;
                                shapes[i].c.k[j].s[1] /= 255;
                                shapes[i].c.k[j].s[2] /= 255;
                                shapes[i].c.k[j].s[3] /= 255;
                            }
                            if(shapes[i].c.k[j].e){
                                shapes[i].c.k[j].e[0] /= 255;
                                shapes[i].c.k[j].e[1] /= 255;
                                shapes[i].c.k[j].e[2] /= 255;
                                shapes[i].c.k[j].e[3] /= 255;
                            }
                        }
                    } else {
                        shapes[i].c.k[0] /= 255;
                        shapes[i].c.k[1] /= 255;
                        shapes[i].c.k[2] /= 255;
                        shapes[i].c.k[3] /= 255;
                    }
                }
            }
        }

        function iterateLayers(layers){
            var i, len = layers.length;
            for(i=0;i<len;i+=1){
                if(layers[i].ty === 4){
                    iterateShapes(layers[i].shapes);
                }
            }
        }

        return function (animationData){
            if(checkVersion(minimumVersion,animationData.v)){
                iterateLayers(animationData.layers);
                if(animationData.assets){
                    var i, len = animationData.assets.length;
                    for(i=0;i<len;i+=1){
                        if(animationData.assets[i].layers){
                            iterateLayers(animationData.assets[i].layers);

                        }
                    }
                }
            }
        };
    }());

    var checkShapes = (function(){
        var minimumVersion = [4,4,18];


        function completeShapes(arr){
            var i, len = arr.length;
            var j, jLen;
            var hasPaths = false;
            for(i=len-1;i>=0;i-=1){
                if(arr[i].ty == 'sh'){
                    if(arr[i].ks.k.i){
                        arr[i].ks.k.c = arr[i].closed;
                    }else{
                        jLen = arr[i].ks.k.length;
                        for(j=0;j<jLen;j+=1){
                            if(arr[i].ks.k[j].s){
                                arr[i].ks.k[j].s[0].c = arr[i].closed;
                            }
                            if(arr[i].ks.k[j].e){
                                arr[i].ks.k[j].e[0].c = arr[i].closed;
                            }
                        }
                    }
                    hasPaths = true;
                }else if(arr[i].ty == 'gr'){
                    completeShapes(arr[i].it);
                }
            }
        }

        function iterateLayers(layers){
            var layerData;
            var i, len = layers.length;
            var j, jLen, k, kLen;
            for(i=0;i<len;i+=1){
                layerData = layers[i];
                if(layerData.hasMask){
                    var maskProps = layerData.masksProperties;
                    jLen = maskProps.length;
                    for(j=0;j<jLen;j+=1){
                        if(maskProps[j].pt.k.i){
                            maskProps[j].pt.k.c = maskProps[j].cl;
                        }else{
                            kLen = maskProps[j].pt.k.length;
                            for(k=0;k<kLen;k+=1){
                                if(maskProps[j].pt.k[k].s){
                                    maskProps[j].pt.k[k].s[0].c = maskProps[j].cl;
                                }
                                if(maskProps[j].pt.k[k].e){
                                    maskProps[j].pt.k[k].e[0].c = maskProps[j].cl;
                                }
                            }
                        }
                    }
                }
                if(layerData.ty === 4){
                    completeShapes(layerData.shapes);
                }
            }
        }

        return function (animationData){
            if(checkVersion(minimumVersion,animationData.v)){
                iterateLayers(animationData.layers);
                if(animationData.assets){
                    var i, len = animationData.assets.length;
                    for(i=0;i<len;i+=1){
                        if(animationData.assets[i].layers){
                            iterateLayers(animationData.assets[i].layers);

                        }
                    }
                }
            }
        };
    }());

    function completeData(animationData, fontManager){
        if(animationData.__complete){
            return;
        }
        checkColors(animationData);
        checkText(animationData);
        checkChars(animationData);
        checkShapes(animationData);
        completeLayers(animationData.layers, animationData.assets, fontManager);
        animationData.__complete = true;
        //blitAnimation(animationData, animationData.assets, fontManager);
    }

    function completeText(data, fontManager){
        if(data.t.a.length === 0 && !('m' in data.t.p)){
            data.singleShape = true;
        }
    }

    var moduleOb = {};
    moduleOb.completeData = completeData;

    return moduleOb;
}

var dataManager = dataFunctionManager();
var FontManager = (function(){

    var maxWaitingTime = 5000;
    var emptyChar = {
        w: 0,
        size:0,
        shapes:[]
    };
    var combinedCharacters = [];
    //Hindi characters
    combinedCharacters = combinedCharacters.concat([2304, 2305, 2306, 2307, 2362, 2363, 2364, 2364, 2366
    , 2367, 2368, 2369, 2370, 2371, 2372, 2373, 2374, 2375, 2376, 2377, 2378, 2379
    , 2380, 2381, 2382, 2383, 2387, 2388, 2389, 2390, 2391, 2402, 2403]);

    function setUpNode(font, family){
        var parentNode = createTag('span');
        parentNode.style.fontFamily    = family;
        var node = createTag('span');
        // Characters that vary significantly among different fonts
        node.innerHTML = 'giItT1WQy@!-/#';
        // Visible - so we can measure it - but not on the screen
        parentNode.style.position      = 'absolute';
        parentNode.style.left          = '-10000px';
        parentNode.style.top           = '-10000px';
        // Large font size makes even subtle changes obvious
        parentNode.style.fontSize      = '300px';
        // Reset any font properties
        parentNode.style.fontVariant   = 'normal';
        parentNode.style.fontStyle     = 'normal';
        parentNode.style.fontWeight    = 'normal';
        parentNode.style.letterSpacing = '0';
        parentNode.appendChild(node);
        document.body.appendChild(parentNode);

        // Remember width with no applied web font
        var width = node.offsetWidth;
        node.style.fontFamily = font + ', '+family;
        return {node:node, w:width, parent:parentNode};
    }

    function checkLoadedFonts() {
        var i, len = this.fonts.length;
        var node, w;
        var loadedCount = len;
        for(i=0;i<len; i+= 1){
            if(this.fonts[i].loaded){
                loadedCount -= 1;
                continue;
            }
            if(this.fonts[i].fOrigin === 'n' || this.fonts[i].origin === 0){
                this.fonts[i].loaded = true;
            } else{
                node = this.fonts[i].monoCase.node;
                w = this.fonts[i].monoCase.w;
                if(node.offsetWidth !== w){
                    loadedCount -= 1;
                    this.fonts[i].loaded = true;
                }else{
                    node = this.fonts[i].sansCase.node;
                    w = this.fonts[i].sansCase.w;
                    if(node.offsetWidth !== w){
                        loadedCount -= 1;
                        this.fonts[i].loaded = true;
                    }
                }
                if(this.fonts[i].loaded){
                    this.fonts[i].sansCase.parent.parentNode.removeChild(this.fonts[i].sansCase.parent);
                    this.fonts[i].monoCase.parent.parentNode.removeChild(this.fonts[i].monoCase.parent);
                }
            }
        }

        if(loadedCount !== 0 && Date.now() - this.initTime < maxWaitingTime){
            setTimeout(this.checkLoadedFonts.bind(this),20);
        }else{
            setTimeout(function(){this.isLoaded = true;}.bind(this),0);

        }
    }

    function createHelper(def, fontData){
        var tHelper = createNS('text');
        tHelper.style.fontSize = '100px';
        //tHelper.style.fontFamily = fontData.fFamily;
        tHelper.setAttribute('font-family', fontData.fFamily);
        tHelper.setAttribute('font-style', fontData.fStyle);
        tHelper.setAttribute('font-weight', fontData.fWeight);
        tHelper.textContent = '1';
        if(fontData.fClass){
            tHelper.style.fontFamily = 'inherit';
            tHelper.setAttribute('class', fontData.fClass);
        } else {
            tHelper.style.fontFamily = fontData.fFamily;
        }
        def.appendChild(tHelper);
        var tCanvasHelper = createTag('canvas').getContext('2d');
        tCanvasHelper.font = fontData.fWeight + ' ' + fontData.fStyle + ' 100px '+ fontData.fFamily;
        //tCanvasHelper.font = ' 100px '+ fontData.fFamily;
        return tHelper;
    }

    function addFonts(fontData, defs){
        if(!fontData){
            this.isLoaded = true;
            return;
        }
        if(this.chars){
            this.isLoaded = true;
            this.fonts = fontData.list;
            return;
        }


        var fontArr = fontData.list;
        var i, len = fontArr.length;
        var _pendingFonts = len;
        for(i=0; i<len; i+= 1){
            var shouldLoadFont = true;
            var loadedSelector;
            var j;
            fontArr[i].loaded = false;
            fontArr[i].monoCase = setUpNode(fontArr[i].fFamily,'monospace');
            fontArr[i].sansCase = setUpNode(fontArr[i].fFamily,'sans-serif');
            if(!fontArr[i].fPath) {
                fontArr[i].loaded = true;
                _pendingFonts -= 1;
            }else if(fontArr[i].fOrigin === 'p' || fontArr[i].origin === 3){
                loadedSelector = document.querySelectorAll('style[f-forigin="p"][f-family="'+ fontArr[i].fFamily +'"], style[f-origin="3"][f-family="'+ fontArr[i].fFamily +'"]');

                if (loadedSelector.length > 0) {
                    shouldLoadFont = false;
                }

                if (shouldLoadFont) {
                    var s = createTag('style');
                    s.setAttribute('f-forigin', fontArr[i].fOrigin);
                    s.setAttribute('f-origin', fontArr[i].origin);
                    s.setAttribute('f-family', fontArr[i].fFamily);
                    s.type = "text/css";
                    s.innerHTML = "@font-face {" + "font-family: "+fontArr[i].fFamily+"; font-style: normal; src: url('"+fontArr[i].fPath+"');}";
                    defs.appendChild(s);
                }
            } else if(fontArr[i].fOrigin === 'g' || fontArr[i].origin === 1){
                loadedSelector = document.querySelectorAll('link[f-forigin="g"], link[f-origin="1"]');

                for (j = 0; j < loadedSelector.length; j++) {
                    if (loadedSelector[j].href.indexOf(fontArr[i].fPath) !== -1) {
                        // Font is already loaded
                        shouldLoadFont = false;
                    }
                }

                if (shouldLoadFont) {
                    var l = createTag('link');
                    l.setAttribute('f-forigin', fontArr[i].fOrigin);
                    l.setAttribute('f-origin', fontArr[i].origin);
                    l.type = "text/css";
                    l.rel = "stylesheet";
                    l.href = fontArr[i].fPath;
                    document.body.appendChild(l);
                }
            } else if(fontArr[i].fOrigin === 't' || fontArr[i].origin === 2){
                loadedSelector = document.querySelectorAll('script[f-forigin="t"], script[f-origin="2"]');

                for (j = 0; j < loadedSelector.length; j++) {
                    if (fontArr[i].fPath === loadedSelector[j].src) {
                        // Font is already loaded
                        shouldLoadFont = false;
                    }
                }

                if (shouldLoadFont) {
                    var sc = createTag('link');
                    sc.setAttribute('f-forigin', fontArr[i].fOrigin);
                    sc.setAttribute('f-origin', fontArr[i].origin);
                    sc.setAttribute('rel','stylesheet');
                    sc.setAttribute('href',fontArr[i].fPath);
                    defs.appendChild(sc);
                }
            }
            fontArr[i].helper = createHelper(defs,fontArr[i]);
            fontArr[i].cache = {};
            this.fonts.push(fontArr[i]);
        }
        if (_pendingFonts === 0) {
            this.isLoaded = true;
        } else {
            //On some cases even if the font is loaded, it won't load correctly when measuring text on canvas.
            //Adding this timeout seems to fix it
           setTimeout(this.checkLoadedFonts.bind(this), 100);
        }
    }

    function addChars(chars){
        if(!chars){
            return;
        }
        if(!this.chars){
            this.chars = [];
        }
        var i, len = chars.length;
        var j, jLen = this.chars.length, found;
        for(i=0;i<len;i+=1){
            j = 0;
            found = false;
            while(j<jLen){
                if(this.chars[j].style === chars[i].style && this.chars[j].fFamily === chars[i].fFamily && this.chars[j].ch === chars[i].ch){
                    found = true;
                }
                j += 1;
            }
            if(!found){
                this.chars.push(chars[i]);
                jLen += 1;
            }
        }
    }

    function getCharData(char, style, font){
        var i = 0, len = this.chars.length;
        while( i < len) {
            if(this.chars[i].ch === char && this.chars[i].style === style && this.chars[i].fFamily === font){
                return this.chars[i];
            }
            i+= 1;
        }
        if(console && console.warn) {
            console.warn('Missing character from exported characters list: ', char, style, font);
        }
        return emptyChar;
    }

    function measureText(char, fontName, size) {
        var fontData = this.getFontByName(fontName);
        var index = char.charCodeAt(0);
        if(!fontData.cache[index + 1]) {
            var tHelper = fontData.helper;
            //Canvas version
            //fontData.cache[index] = tHelper.measureText(char).width / 100;
            //SVG version
            //console.log(tHelper.getBBox().width)
            if (char === ' ') {
                tHelper.textContent = '|' + char + '|';
                var doubleSize = tHelper.getComputedTextLength();
                tHelper.textContent = '||';
                var singleSize = tHelper.getComputedTextLength();
                fontData.cache[index + 1] = (doubleSize - singleSize)/100;
            } else {
                tHelper.textContent = char;
                fontData.cache[index + 1] = (tHelper.getComputedTextLength())/100;
            }
        }
        return fontData.cache[index + 1] * size;
    }

    function getFontByName(name){
        var i = 0, len = this.fonts.length;
        while(i<len){
            if(this.fonts[i].fName === name) {
                return this.fonts[i];
            }
            i += 1;
        }
        return this.fonts[0];
    }

    function getCombinedCharacterCodes() {
        return combinedCharacters;
    }

    function loaded() {
        return this.isLoaded;
    }

    var Font = function(){
        this.fonts = [];
        this.chars = null;
        this.typekitLoaded = 0;
        this.isLoaded = false;
        this.initTime = Date.now();
    };
    //TODO: for now I'm adding these methods to the Class and not the prototype. Think of a better way to implement it.
    Font.getCombinedCharacterCodes = getCombinedCharacterCodes;

    Font.prototype.addChars = addChars;
    Font.prototype.addFonts = addFonts;
    Font.prototype.getCharData = getCharData;
    Font.prototype.getFontByName = getFontByName;
    Font.prototype.measureText = measureText;
    Font.prototype.checkLoadedFonts = checkLoadedFonts;
    Font.prototype.loaded = loaded;

    return Font;

}());
var PropertyFactory = (function(){

    var initFrame = initialDefaultFrame;
    var math_abs = Math.abs;

    function interpolateValue(frameNum, caching) {
        var offsetTime = this.offsetTime;
        var newValue;
        if (this.propType === 'multidimensional') {
            newValue = createTypedArray('float32', this.pv.length);
        }
        var iterationIndex = caching.lastIndex;
        var i = iterationIndex;
        var len = this.keyframes.length - 1, flag = true;
        var keyData, nextKeyData;

        while (flag) {
            keyData = this.keyframes[i];
            nextKeyData = this.keyframes[i + 1];
            if (i === len - 1 && frameNum >= nextKeyData.t - offsetTime){
                if(keyData.h){
                    keyData = nextKeyData;
                }
                iterationIndex = 0;
                break;
            }
            if ((nextKeyData.t - offsetTime) > frameNum){
                iterationIndex = i;
                break;
            }
            if (i < len - 1){
                i += 1;
            } else {
                iterationIndex = 0;
                flag = false;
            }
        }

        var k, kLen, perc, jLen, j, fnc;
        var nextKeyTime = nextKeyData.t - offsetTime;
        var keyTime = keyData.t - offsetTime;
        var endValue;
        if (keyData.to) {
            if (!keyData.bezierData) {
                keyData.bezierData = bez.buildBezierData(keyData.s, nextKeyData.s || keyData.e, keyData.to, keyData.ti);
            }
            var bezierData = keyData.bezierData;
            if (frameNum >= nextKeyTime || frameNum < keyTime) {
                var ind = frameNum >= nextKeyTime ? bezierData.points.length - 1 : 0;
                kLen = bezierData.points[ind].point.length;
                for (k = 0; k < kLen; k += 1) {
                    newValue[k] = bezierData.points[ind].point[k];
                }
                // caching._lastKeyframeIndex = -1;
            } else {
                if (keyData.__fnct) {
                    fnc = keyData.__fnct;
                } else {
                    fnc = BezierFactory.getBezierEasing(keyData.o.x, keyData.o.y, keyData.i.x, keyData.i.y, keyData.n).get;
                    keyData.__fnct = fnc;
                }
                perc = fnc((frameNum - keyTime) / (nextKeyTime - keyTime));
                var distanceInLine = bezierData.segmentLength*perc;

                var segmentPerc;
                var addedLength =  (caching.lastFrame < frameNum && caching._lastKeyframeIndex === i) ? caching._lastAddedLength : 0;
                j =  (caching.lastFrame < frameNum && caching._lastKeyframeIndex === i) ? caching._lastPoint : 0;
                flag = true;
                jLen = bezierData.points.length;
                while (flag) {
                    addedLength += bezierData.points[j].partialLength;
                    if (distanceInLine === 0 || perc === 0 || j === bezierData.points.length - 1) {
                        kLen = bezierData.points[j].point.length;
                        for (k = 0; k < kLen; k += 1) {
                            newValue[k] = bezierData.points[j].point[k];
                        }
                        break;
                    } else if (distanceInLine >= addedLength && distanceInLine < addedLength + bezierData.points[j + 1].partialLength) {
                        segmentPerc = (distanceInLine - addedLength) / bezierData.points[j + 1].partialLength;
                        kLen = bezierData.points[j].point.length;
                        for (k = 0; k < kLen; k += 1) {
                            newValue[k] = bezierData.points[j].point[k] + (bezierData.points[j + 1].point[k] - bezierData.points[j].point[k]) * segmentPerc;
                        }
                        break;
                    }
                    if (j < jLen - 1){
                        j += 1;
                    } else {
                        flag = false;
                    }
                }
                caching._lastPoint = j;
                caching._lastAddedLength = addedLength - bezierData.points[j].partialLength;
                caching._lastKeyframeIndex = i;
            }
        } else {
            var outX, outY, inX, inY, keyValue;
            len = keyData.s.length;
            endValue = nextKeyData.s || keyData.e;
            if (this.sh && keyData.h !== 1) {
                if (frameNum >= nextKeyTime) {
                    newValue[0] = endValue[0];
                    newValue[1] = endValue[1];
                    newValue[2] = endValue[2];
                } else if (frameNum <= keyTime) {
                    newValue[0] = keyData.s[0];
                    newValue[1] = keyData.s[1];
                    newValue[2] = keyData.s[2];
                } else {
                    var quatStart = createQuaternion(keyData.s);
                    var quatEnd = createQuaternion(endValue);
                    var time = (frameNum - keyTime) / (nextKeyTime - keyTime);
                    quaternionToEuler(newValue, slerp(quatStart, quatEnd, time));
                }

            } else {
                for(i = 0; i < len; i += 1) {
                    if (keyData.h !== 1) {
                        if (frameNum >= nextKeyTime) {
                            perc = 1;
                        } else if(frameNum < keyTime) {
                            perc = 0;
                        } else {
                            if(keyData.o.x.constructor === Array) {
                                if (!keyData.__fnct) {
                                    keyData.__fnct = [];
                                }
                                if (!keyData.__fnct[i]) {
                                    outX = (typeof keyData.o.x[i] === 'undefined') ? keyData.o.x[0] : keyData.o.x[i];
                                    outY = (typeof keyData.o.y[i] === 'undefined') ? keyData.o.y[0] : keyData.o.y[i];
                                    inX = (typeof keyData.i.x[i] === 'undefined') ? keyData.i.x[0] : keyData.i.x[i];
                                    inY = (typeof keyData.i.y[i] === 'undefined') ? keyData.i.y[0] : keyData.i.y[i];
                                    fnc = BezierFactory.getBezierEasing(outX, outY, inX, inY).get;
                                    keyData.__fnct[i] = fnc;
                                } else {
                                    fnc = keyData.__fnct[i];
                                }
                            } else {
                                if (!keyData.__fnct) {
                                    outX = keyData.o.x;
                                    outY = keyData.o.y;
                                    inX = keyData.i.x;
                                    inY = keyData.i.y;
                                    fnc = BezierFactory.getBezierEasing(outX, outY, inX, inY).get;
                                    keyData.__fnct = fnc;
                                } else {
                                    fnc = keyData.__fnct;
                                }
                            }
                            perc = fnc((frameNum - keyTime) / (nextKeyTime - keyTime ));
                        }
                    }

                    endValue = nextKeyData.s || keyData.e;
                    keyValue = keyData.h === 1 ? keyData.s[i] : keyData.s[i] + (endValue[i] - keyData.s[i]) * perc;

                    if (len === 1) {
                        newValue = keyValue;
                    } else {
                        newValue[i] = keyValue;
                    }
                }
            }
        }
        caching.lastIndex = iterationIndex;
        return newValue;
    }

    //based on @Toji's https://github.com/toji/gl-matrix/
    function slerp(a, b, t) {
        var out = [];
        var ax = a[0], ay = a[1], az = a[2], aw = a[3],
        bx = b[0], by = b[1], bz = b[2], bw = b[3]

        var omega, cosom, sinom, scale0, scale1;

        cosom = ax * bx + ay * by + az * bz + aw * bw;
        if (cosom < 0.0) {
            cosom = -cosom;
            bx = -bx;
            by = -by;
            bz = -bz;
            bw = -bw;
        }
        if ((1.0 - cosom) > 0.000001) {
            omega = Math.acos(cosom);
            sinom = Math.sin(omega);
            scale0 = Math.sin((1.0 - t) * omega) / sinom;
            scale1 = Math.sin(t * omega) / sinom;
        } else {
            scale0 = 1.0 - t;
            scale1 = t;
        }
        out[0] = scale0 * ax + scale1 * bx;
        out[1] = scale0 * ay + scale1 * by;
        out[2] = scale0 * az + scale1 * bz;
        out[3] = scale0 * aw + scale1 * bw;

        return out;
    }

    function quaternionToEuler(out, quat) {
        var qx = quat[0];
        var qy = quat[1];
        var qz = quat[2];
        var qw = quat[3];
        var heading = Math.atan2(2*qy*qw-2*qx*qz , 1 - 2*qy*qy - 2*qz*qz)
        var attitude = Math.asin(2*qx*qy + 2*qz*qw)
        var bank = Math.atan2(2*qx*qw-2*qy*qz , 1 - 2*qx*qx - 2*qz*qz);
        out[0] = heading/degToRads;
        out[1] = attitude/degToRads;
        out[2] = bank/degToRads;
    }

    function createQuaternion(values) {
        var heading = values[0] * degToRads;
        var attitude = values[1] * degToRads;
        var bank = values[2] * degToRads;
        var c1 = Math.cos(heading / 2);
        var c2 = Math.cos(attitude / 2);
        var c3 = Math.cos(bank / 2);
        var s1 = Math.sin(heading / 2);
        var s2 = Math.sin(attitude / 2);
        var s3 = Math.sin(bank / 2);
        var w = c1 * c2 * c3 - s1 * s2 * s3;
        var x = s1 * s2 * c3 + c1 * c2 * s3;
        var y = s1 * c2 * c3 + c1 * s2 * s3;
        var z = c1 * s2 * c3 - s1 * c2 * s3;

        return [x,y,z,w];
    }

    function getValueAtCurrentTime(){
        var frameNum = this.comp.renderedFrame - this.offsetTime;
        var initTime = this.keyframes[0].t - this.offsetTime;
        var endTime = this.keyframes[this.keyframes.length- 1].t-this.offsetTime;
        if(!(frameNum === this._caching.lastFrame || (this._caching.lastFrame !== initFrame && ((this._caching.lastFrame >= endTime && frameNum >= endTime) || (this._caching.lastFrame < initTime && frameNum < initTime))))){
            if(this._caching.lastFrame >= frameNum) {
                this._caching._lastKeyframeIndex = -1;
                this._caching.lastIndex = 0;
            }

            var renderResult = this.interpolateValue(frameNum, this._caching);
            this.pv = renderResult;
        }
        this._caching.lastFrame = frameNum;
        return this.pv;
    }

    function setVValue(val) {
        var multipliedValue;
        if(this.propType === 'unidimensional') {
            multipliedValue = val * this.mult;
            if(math_abs(this.v - multipliedValue) > 0.00001) {
                this.v = multipliedValue;
                this._mdf = true;
            }
        } else {
            var i = 0, len = this.v.length;
            while (i < len) {
                multipliedValue = val[i] * this.mult;
                if (math_abs(this.v[i] - multipliedValue) > 0.00001) {
                    this.v[i] = multipliedValue;
                    this._mdf = true;
                }
                i += 1;
            }
        }
    }

    function processEffectsSequence() {
        if(this.elem.globalData.frameId === this.frameId || !this.effectsSequence.length) {
            return;
        }
        if(this.lock) {
            this.setVValue(this.pv);
            return;
        }
        this.lock = true;
        this._mdf = this._isFirstFrame;
        var multipliedValue;
        var i, len = this.effectsSequence.length;
        var finalValue = this.kf ? this.pv : this.data.k;
        for(i = 0; i < len; i += 1) {
            finalValue = this.effectsSequence[i](finalValue);
        }
        this.setVValue(finalValue);
        this._isFirstFrame = false;
        this.lock = false;
        this.frameId = this.elem.globalData.frameId;
    }

    function addEffect(effectFunction) {
        this.effectsSequence.push(effectFunction);
        this.container.addDynamicProperty(this);
    }

    function ValueProperty(elem, data, mult, container){
        this.propType = 'unidimensional';
        this.mult = mult || 1;
        this.data = data;
        this.v = mult ? data.k * mult : data.k;
        this.pv = data.k;
        this._mdf = false;
        this.elem = elem;
        this.container = container;
        this.comp = elem.comp;
        this.k = false;
        this.kf = false;
        this.vel = 0;
        this.effectsSequence = [];
        this._isFirstFrame = true;
        this.getValue = processEffectsSequence;
        this.setVValue = setVValue;
        this.addEffect = addEffect;
    }

    function MultiDimensionalProperty(elem, data, mult, container) {
        this.propType = 'multidimensional';
        this.mult = mult || 1;
        this.data = data;
        this._mdf = false;
        this.elem = elem;
        this.container = container;
        this.comp = elem.comp;
        this.k = false;
        this.kf = false;
        this.frameId = -1;
        var i, len = data.k.length;
        this.v = createTypedArray('float32', len);
        this.pv = createTypedArray('float32', len);
        var arr = createTypedArray('float32', len);
        this.vel = createTypedArray('float32', len);
        for (i = 0; i < len; i += 1) {
            this.v[i] = data.k[i] * this.mult;
            this.pv[i] = data.k[i];
        }
        this._isFirstFrame = true;
        this.effectsSequence = [];
        this.getValue = processEffectsSequence;
        this.setVValue = setVValue;
        this.addEffect = addEffect;
    }

    function KeyframedValueProperty(elem, data, mult, container) {
        this.propType = 'unidimensional';
        this.keyframes = data.k;
        this.offsetTime = elem.data.st;
        this.frameId = -1;
        this._caching = {lastFrame: initFrame, lastIndex: 0, value: 0, _lastKeyframeIndex: -1};
        this.k = true;
        this.kf = true;
        this.data = data;
        this.mult = mult || 1;
        this.elem = elem;
        this.container = container;
        this.comp = elem.comp;
        this.v = initFrame;
        this.pv = initFrame;
        this._isFirstFrame = true;
        this.getValue = processEffectsSequence;
        this.setVValue = setVValue;
        this.interpolateValue = interpolateValue;
        this.effectsSequence = [getValueAtCurrentTime.bind(this)];
        this.addEffect = addEffect;
    }

    function KeyframedMultidimensionalProperty(elem, data, mult, container){
        this.propType = 'multidimensional';
        var i, len = data.k.length;
        var s, e,to,ti;
        for (i = 0; i < len - 1; i += 1) {
            if (data.k[i].to && data.k[i].s && data.k[i].e) {
                s = data.k[i].s;
                e = data.k[i].e;
                to = data.k[i].to;
                ti = data.k[i].ti;
                if((s.length === 2 && !(s[0] === e[0] && s[1] === e[1]) && bez.pointOnLine2D(s[0],s[1],e[0],e[1],s[0] + to[0],s[1] + to[1]) && bez.pointOnLine2D(s[0],s[1],e[0],e[1],e[0] + ti[0],e[1] + ti[1])) || (s.length === 3 && !(s[0] === e[0] && s[1] === e[1] && s[2] === e[2]) && bez.pointOnLine3D(s[0],s[1],s[2],e[0],e[1],e[2],s[0] + to[0],s[1] + to[1],s[2] + to[2]) && bez.pointOnLine3D(s[0],s[1],s[2],e[0],e[1],e[2],e[0] + ti[0],e[1] + ti[1],e[2] + ti[2]))){
                    data.k[i].to = null;
                    data.k[i].ti = null;
                }
                if(s[0] === e[0] && s[1] === e[1] && to[0] === 0 && to[1] === 0 && ti[0] === 0 && ti[1] === 0) {
                    if(s.length === 2 || (s[2] === e[2] && to[2] === 0 && ti[2] === 0)) {
                        data.k[i].to = null;
                        data.k[i].ti = null;
                    }
                }
            }
        }
        this.effectsSequence = [getValueAtCurrentTime.bind(this)];
        this.keyframes = data.k;
        this.offsetTime = elem.data.st;
        this.k = true;
        this.kf = true;
        this._isFirstFrame = true;
        this.mult = mult || 1;
        this.elem = elem;
        this.container = container;
        this.comp = elem.comp;
        this.getValue = processEffectsSequence;
        this.setVValue = setVValue;
        this.interpolateValue = interpolateValue;
        this.frameId = -1;
        var arrLen = data.k[0].s.length;
        this.v = createTypedArray('float32', arrLen);
        this.pv = createTypedArray('float32', arrLen);
        for (i = 0; i < arrLen; i += 1) {
            this.v[i] = initFrame;
            this.pv[i] = initFrame;
        }
        this._caching={lastFrame:initFrame,lastIndex:0,value:createTypedArray('float32', arrLen)};
        this.addEffect = addEffect;
    }

    function getProp(elem,data,type, mult, container) {
        var p;
        if(!data.k.length){
            p = new ValueProperty(elem,data, mult, container);
        }else if(typeof(data.k[0]) === 'number'){
            p = new MultiDimensionalProperty(elem,data, mult, container);
        }else{
            switch(type){
                case 0:
                    p = new KeyframedValueProperty(elem,data,mult, container);
                    break;
                case 1:
                    p = new KeyframedMultidimensionalProperty(elem,data,mult, container);
                    break;
            }
        }
        if(p.effectsSequence.length){
            container.addDynamicProperty(p);
        }
        return p;
    }

    var ob = {
        getProp: getProp
    };
    return ob;
}());
var TransformPropertyFactory = (function() {

    function applyToMatrix(mat) {
        var _mdf = this._mdf;
        this.iterateDynamicProperties();
        this._mdf = this._mdf || _mdf;
        if (this.a) {
            mat.translate(-this.a.v[0], -this.a.v[1], this.a.v[2]);
        }
        if (this.s) {
            mat.scale(this.s.v[0], this.s.v[1], this.s.v[2]);
        }
        if (this.sk) {
            mat.skewFromAxis(-this.sk.v, this.sa.v);
        }
        if (this.r) {
            mat.rotate(-this.r.v);
        } else {
            mat.rotateZ(-this.rz.v).rotateY(this.ry.v).rotateX(this.rx.v).rotateZ(-this.or.v[2]).rotateY(this.or.v[1]).rotateX(this.or.v[0]);
        }
        if (this.data.p.s) {
            if (this.data.p.z) {
                mat.translate(this.px.v, this.py.v, -this.pz.v);
            } else {
                mat.translate(this.px.v, this.py.v, 0);
            }
        } else {
            mat.translate(this.p.v[0], this.p.v[1], -this.p.v[2]);
        }
    }
    function processKeys(forceRender){
        if (this.elem.globalData.frameId === this.frameId) {
            return;
        }
        if(this._isDirty) {
            this.precalculateMatrix();
            this._isDirty = false;
        }

        this.iterateDynamicProperties();

        if (this._mdf || forceRender) {
            this.v.cloneFromProps(this.pre.props);
            if (this.appliedTransformations < 1) {
                this.v.translate(-this.a.v[0], -this.a.v[1], this.a.v[2]);
            }
            if(this.appliedTransformations < 2) {
                this.v.scale(this.s.v[0], this.s.v[1], this.s.v[2]);
            }
            if (this.sk && this.appliedTransformations < 3) {
                this.v.skewFromAxis(-this.sk.v, this.sa.v);
            }
            if (this.r && this.appliedTransformations < 4) {
                this.v.rotate(-this.r.v);
            } else if (!this.r && this.appliedTransformations < 4){
                this.v.rotateZ(-this.rz.v).rotateY(this.ry.v).rotateX(this.rx.v).rotateZ(-this.or.v[2]).rotateY(this.or.v[1]).rotateX(this.or.v[0]);
            }
            if (this.autoOriented) {
                var v1,v2, frameRate = this.elem.globalData.frameRate;
                if(this.p && this.p.keyframes && this.p.getValueAtTime) {
                    if (this.p._caching.lastFrame+this.p.offsetTime <= this.p.keyframes[0].t) {
                        v1 = this.p.getValueAtTime((this.p.keyframes[0].t + 0.01) / frameRate,0);
                        v2 = this.p.getValueAtTime(this.p.keyframes[0].t / frameRate, 0);
                    } else if(this.p._caching.lastFrame+this.p.offsetTime >= this.p.keyframes[this.p.keyframes.length - 1].t) {
                        v1 = this.p.getValueAtTime((this.p.keyframes[this.p.keyframes.length - 1].t / frameRate), 0);
                        v2 = this.p.getValueAtTime((this.p.keyframes[this.p.keyframes.length - 1].t - 0.01) / frameRate, 0);
                    } else {
                        v1 = this.p.pv;
                        v2 = this.p.getValueAtTime((this.p._caching.lastFrame+this.p.offsetTime - 0.01) / frameRate, this.p.offsetTime);
                    }
                } else if(this.px && this.px.keyframes && this.py.keyframes && this.px.getValueAtTime && this.py.getValueAtTime) {
                    v1 = [];
                    v2 = [];
                    var px = this.px, py = this.py, frameRate;
                    if (px._caching.lastFrame+px.offsetTime <= px.keyframes[0].t) {
                        v1[0] = px.getValueAtTime((px.keyframes[0].t + 0.01) / frameRate,0);
                        v1[1] = py.getValueAtTime((py.keyframes[0].t + 0.01) / frameRate,0);
                        v2[0] = px.getValueAtTime((px.keyframes[0].t) / frameRate,0);
                        v2[1] = py.getValueAtTime((py.keyframes[0].t) / frameRate,0);
                    } else if(px._caching.lastFrame+px.offsetTime >= px.keyframes[px.keyframes.length - 1].t) {
                        v1[0] = px.getValueAtTime((px.keyframes[px.keyframes.length - 1].t / frameRate),0);
                        v1[1] = py.getValueAtTime((py.keyframes[py.keyframes.length - 1].t / frameRate),0);
                        v2[0] = px.getValueAtTime((px.keyframes[px.keyframes.length - 1].t - 0.01) / frameRate,0);
                        v2[1] = py.getValueAtTime((py.keyframes[py.keyframes.length - 1].t - 0.01) / frameRate,0);
                    } else {
                        v1 = [px.pv, py.pv];
                        v2[0] = px.getValueAtTime((px._caching.lastFrame+px.offsetTime - 0.01) / frameRate,px.offsetTime);
                        v2[1] = py.getValueAtTime((py._caching.lastFrame+py.offsetTime - 0.01) / frameRate,py.offsetTime);
                    }
                }
                this.v.rotate(-Math.atan2(v1[1] - v2[1], v1[0] - v2[0]));
            }
            if(this.data.p && this.data.p.s){
                if(this.data.p.z) {
                    this.v.translate(this.px.v, this.py.v, -this.pz.v);
                } else {
                    this.v.translate(this.px.v, this.py.v, 0);
                }
            }else{
                this.v.translate(this.p.v[0],this.p.v[1],-this.p.v[2]);
            }
        }
        this.frameId = this.elem.globalData.frameId;
    }

    function precalculateMatrix() {
        if(!this.a.k) {
            this.pre.translate(-this.a.v[0], -this.a.v[1], this.a.v[2]);
            this.appliedTransformations = 1;
        } else {
            return;
        }
        if(!this.s.effectsSequence.length) {
            this.pre.scale(this.s.v[0], this.s.v[1], this.s.v[2]);
            this.appliedTransformations = 2;
        } else {
            return;
        }
        if(this.sk) {
            if(!this.sk.effectsSequence.length && !this.sa.effectsSequence.length) {
                this.pre.skewFromAxis(-this.sk.v, this.sa.v);
            this.appliedTransformations = 3;
            } else {
                return;
            }
        }
        if (this.r) {
            if(!this.r.effectsSequence.length) {
                this.pre.rotate(-this.r.v);
                this.appliedTransformations = 4;
            } else {
                return;
            }
        } else if(!this.rz.effectsSequence.length && !this.ry.effectsSequence.length && !this.rx.effectsSequence.length && !this.or.effectsSequence.length) {
            this.pre.rotateZ(-this.rz.v).rotateY(this.ry.v).rotateX(this.rx.v).rotateZ(-this.or.v[2]).rotateY(this.or.v[1]).rotateX(this.or.v[0]);
            this.appliedTransformations = 4;
        }
    }

    function autoOrient(){
        //
        //var prevP = this.getValueAtTime();
    }

    function addDynamicProperty(prop) {
        this._addDynamicProperty(prop);
        this.elem.addDynamicProperty(prop);
        this._isDirty = true;
    }

    function TransformProperty(elem,data,container){
        this.elem = elem;
        this.frameId = -1;
        this.propType = 'transform';
        this.data = data;
        this.v = new Matrix();
        //Precalculated matrix with non animated properties
        this.pre = new Matrix();
        this.appliedTransformations = 0;
        this.initDynamicPropertyContainer(container || elem);
        if(data.p && data.p.s){
            this.px = PropertyFactory.getProp(elem,data.p.x,0,0,this);
            this.py = PropertyFactory.getProp(elem,data.p.y,0,0,this);
            if(data.p.z){
                this.pz = PropertyFactory.getProp(elem,data.p.z,0,0,this);
            }
        }else{
            this.p = PropertyFactory.getProp(elem,data.p || {k:[0,0,0]},1,0,this);
        }
        if(data.rx) {
            this.rx = PropertyFactory.getProp(elem, data.rx, 0, degToRads, this);
            this.ry = PropertyFactory.getProp(elem, data.ry, 0, degToRads, this);
            this.rz = PropertyFactory.getProp(elem, data.rz, 0, degToRads, this);
            if(data.or.k[0].ti) {
                var i, len = data.or.k.length;
                for(i=0;i<len;i+=1) {
                    data.or.k[i].to = data.or.k[i].ti = null;
                }
            }
            this.or = PropertyFactory.getProp(elem, data.or, 1, degToRads, this);
            //sh Indicates it needs to be capped between -180 and 180
            this.or.sh = true;
        } else {
            this.r = PropertyFactory.getProp(elem, data.r || {k: 0}, 0, degToRads, this);
        }
        if(data.sk){
            this.sk = PropertyFactory.getProp(elem, data.sk, 0, degToRads, this);
            this.sa = PropertyFactory.getProp(elem, data.sa, 0, degToRads, this);
        }
        this.a = PropertyFactory.getProp(elem,data.a || {k:[0,0,0]},1,0,this);
        this.s = PropertyFactory.getProp(elem,data.s || {k:[100,100,100]},1,0.01,this);
        // Opacity is not part of the transform properties, that's why it won't use this.dynamicProperties. That way transforms won't get updated if opacity changes.
        if(data.o){
            this.o = PropertyFactory.getProp(elem,data.o,0,0.01,elem);
        } else {
            this.o = {_mdf:false,v:1};
        }
        this._isDirty = true;
        if(!this.dynamicProperties.length){
            this.getValue(true);
        }
    }

    TransformProperty.prototype = {
        applyToMatrix: applyToMatrix,
        getValue: processKeys,
        precalculateMatrix: precalculateMatrix,
        autoOrient: autoOrient
    }

    extendPrototype([DynamicPropertyContainer], TransformProperty);
    TransformProperty.prototype.addDynamicProperty = addDynamicProperty;
    TransformProperty.prototype._addDynamicProperty = DynamicPropertyContainer.prototype.addDynamicProperty;

    function getTransformProperty(elem,data,container){
        return new TransformProperty(elem,data,container);
    }

    return {
        getTransformProperty: getTransformProperty
    };

}());
function ShapePath(){
	this.c = false;
	this._length = 0;
	this._maxLength = 8;
	this.v = createSizedArray(this._maxLength);
	this.o = createSizedArray(this._maxLength);
	this.i = createSizedArray(this._maxLength);
}

ShapePath.prototype.setPathData = function(closed, len) {
	this.c = closed;
	this.setLength(len);
	var i = 0;
	while(i < len){
		this.v[i] = point_pool.newElement();
		this.o[i] = point_pool.newElement();
		this.i[i] = point_pool.newElement();
		i += 1;
	}
};

ShapePath.prototype.setLength = function(len) {
	while(this._maxLength < len) {
		this.doubleArrayLength();
	}
	this._length = len;
};

ShapePath.prototype.doubleArrayLength = function() {
	this.v = this.v.concat(createSizedArray(this._maxLength));
	this.i = this.i.concat(createSizedArray(this._maxLength));
	this.o = this.o.concat(createSizedArray(this._maxLength));
	this._maxLength *= 2;
};

ShapePath.prototype.setXYAt = function(x, y, type, pos, replace) {
	var arr;
	this._length = Math.max(this._length, pos + 1);
	if(this._length >= this._maxLength) {
		this.doubleArrayLength();
	}
	switch(type){
		case 'v':
			arr = this.v;
			break;
		case 'i':
			arr = this.i;
			break;
		case 'o':
			arr = this.o;
			break;
	}
	if(!arr[pos] || (arr[pos] && !replace)){
		arr[pos] = point_pool.newElement();
	}
	arr[pos][0] = x;
	arr[pos][1] = y;
};

ShapePath.prototype.setTripleAt = function(vX,vY,oX,oY,iX,iY,pos, replace) {
	this.setXYAt(vX,vY,'v',pos, replace);
	this.setXYAt(oX,oY,'o',pos, replace);
	this.setXYAt(iX,iY,'i',pos, replace);
};

ShapePath.prototype.reverse = function() {
	var newPath = new ShapePath();
	newPath.setPathData(this.c, this._length);
	var vertices = this.v, outPoints = this.o, inPoints = this.i;
	var init = 0;
	if (this.c) {
		newPath.setTripleAt(vertices[0][0], vertices[0][1], inPoints[0][0], inPoints[0][1], outPoints[0][0], outPoints[0][1], 0, false);
        init = 1;
    }
    var cnt = this._length - 1;
    var len = this._length;

    var i;
    for (i = init; i < len; i += 1) {
    	newPath.setTripleAt(vertices[cnt][0], vertices[cnt][1], inPoints[cnt][0], inPoints[cnt][1], outPoints[cnt][0], outPoints[cnt][1], i, false);
        cnt -= 1;
    }
    return newPath;
};
var ShapePropertyFactory = (function(){

    var initFrame = -999999;

    function interpolateShape(frameNum, previousValue, caching) {
        var iterationIndex = caching.lastIndex;
        var keyPropS,keyPropE,isHold, j, k, jLen, kLen, perc, vertexValue;
        var kf = this.keyframes;
        if(frameNum < kf[0].t-this.offsetTime){
            keyPropS = kf[0].s[0];
            isHold = true;
            iterationIndex = 0;
        }else if(frameNum >= kf[kf.length - 1].t-this.offsetTime){
            keyPropS = kf[kf.length - 1].s ? kf[kf.length - 1].s[0] : kf[kf.length - 2].e[0];
            /*if(kf[kf.length - 1].s){
                keyPropS = kf[kf.length - 1].s[0];
            }else{
                keyPropS = kf[kf.length - 2].e[0];
            }*/
            isHold = true;
        }else{
            var i = iterationIndex;
            var len = kf.length- 1,flag = true,keyData,nextKeyData;
            while(flag){
                keyData = kf[i];
                nextKeyData = kf[i+1];
                if((nextKeyData.t - this.offsetTime) > frameNum){
                    break;
                }
                if(i < len - 1){
                    i += 1;
                }else{
                    flag = false;
                }
            }
            isHold = keyData.h === 1;
            iterationIndex = i;
            if(!isHold){
                if(frameNum >= nextKeyData.t-this.offsetTime){
                    perc = 1;
                }else if(frameNum < keyData.t-this.offsetTime){
                    perc = 0;
                }else{
                    var fnc;
                    if(keyData.__fnct){
                        fnc = keyData.__fnct;
                    }else{
                        fnc = BezierFactory.getBezierEasing(keyData.o.x,keyData.o.y,keyData.i.x,keyData.i.y).get;
                        keyData.__fnct = fnc;
                    }
                    perc = fnc((frameNum-(keyData.t-this.offsetTime))/((nextKeyData.t-this.offsetTime)-(keyData.t-this.offsetTime)));
                }
                keyPropE = nextKeyData.s ? nextKeyData.s[0] : keyData.e[0];
            }
            keyPropS = keyData.s[0];
        }
        jLen = previousValue._length;
        kLen = keyPropS.i[0].length;
        caching.lastIndex = iterationIndex;

        for(j=0;j<jLen;j+=1){
            for(k=0;k<kLen;k+=1){
                vertexValue = isHold ? keyPropS.i[j][k] :  keyPropS.i[j][k]+(keyPropE.i[j][k]-keyPropS.i[j][k])*perc;
                previousValue.i[j][k] = vertexValue;
                vertexValue = isHold ? keyPropS.o[j][k] :  keyPropS.o[j][k]+(keyPropE.o[j][k]-keyPropS.o[j][k])*perc;
                previousValue.o[j][k] = vertexValue;
                vertexValue = isHold ? keyPropS.v[j][k] :  keyPropS.v[j][k]+(keyPropE.v[j][k]-keyPropS.v[j][k])*perc;
                previousValue.v[j][k] = vertexValue;
            }
        }
    }

    function interpolateShapeCurrentTime(){
        var frameNum = this.comp.renderedFrame - this.offsetTime;
        var initTime = this.keyframes[0].t - this.offsetTime;
        var endTime = this.keyframes[this.keyframes.length - 1].t - this.offsetTime;
        var lastFrame = this._caching.lastFrame;
        if(!(lastFrame !== initFrame && ((lastFrame < initTime && frameNum < initTime) || (lastFrame > endTime && frameNum > endTime)))){
            ////
            this._caching.lastIndex = lastFrame < frameNum ? this._caching.lastIndex : 0;
            this.interpolateShape(frameNum, this.pv, this._caching);
            ////
        }
        this._caching.lastFrame = frameNum;
        return this.pv;
    }

    function resetShape(){
        this.paths = this.localShapeCollection;
    }

    function shapesEqual(shape1, shape2) {
        if(shape1._length !== shape2._length || shape1.c !== shape2.c){
            return false;
        }
        var i, len = shape1._length;
        for(i = 0; i < len; i += 1) {
            if(shape1.v[i][0] !== shape2.v[i][0]
            || shape1.v[i][1] !== shape2.v[i][1]
            || shape1.o[i][0] !== shape2.o[i][0]
            || shape1.o[i][1] !== shape2.o[i][1]
            || shape1.i[i][0] !== shape2.i[i][0]
            || shape1.i[i][1] !== shape2.i[i][1]) {
                return false;
            }
        }
        return true;
    }

    function setVValue(newPath) {
        if(!shapesEqual(this.v, newPath)) {
            this.v = shape_pool.clone(newPath);
            this.localShapeCollection.releaseShapes();
            this.localShapeCollection.addShape(this.v);
            this._mdf = true;
            this.paths = this.localShapeCollection;
        }
    }

    function processEffectsSequence() {
        if(this.elem.globalData.frameId === this.frameId || !this.effectsSequence.length) {
            return;
        }
        if(this.lock) {
            this.setVValue(this.pv);
            return;
        }
        this.lock = true;
        this._mdf = false;
        var finalValue = this.kf ? this.pv : this.data.ks ? this.data.ks.k : this.data.pt.k;
        var i, len = this.effectsSequence.length;
        for(i = 0; i < len; i += 1) {
            finalValue = this.effectsSequence[i](finalValue);
        }
        this.setVValue(finalValue);
        this.lock = false;
        this.frameId = this.elem.globalData.frameId;
    };

    function ShapeProperty(elem, data, type){
        this.propType = 'shape';
        this.comp = elem.comp;
        this.container = elem;
        this.elem = elem;
        this.data = data;
        this.k = false;
        this.kf = false;
        this._mdf = false;
        var pathData = type === 3 ? data.pt.k : data.ks.k;
        this.v = shape_pool.clone(pathData);
        this.pv = shape_pool.clone(this.v);
        this.localShapeCollection = shapeCollection_pool.newShapeCollection();
        this.paths = this.localShapeCollection;
        this.paths.addShape(this.v);
        this.reset = resetShape;
        this.effectsSequence = [];
    }

    function addEffect(effectFunction) {
        this.effectsSequence.push(effectFunction);
        this.container.addDynamicProperty(this);
    }

    ShapeProperty.prototype.interpolateShape = interpolateShape;
    ShapeProperty.prototype.getValue = processEffectsSequence;
    ShapeProperty.prototype.setVValue = setVValue;
    ShapeProperty.prototype.addEffect = addEffect;

    function KeyframedShapeProperty(elem,data,type){
        this.propType = 'shape';
        this.comp = elem.comp;
        this.elem = elem;
        this.container = elem;
        this.offsetTime = elem.data.st;
        this.keyframes = type === 3 ? data.pt.k : data.ks.k;
        this.k = true;
        this.kf = true;
        var i, len = this.keyframes[0].s[0].i.length;
        var jLen = this.keyframes[0].s[0].i[0].length;
        this.v = shape_pool.newElement();
        this.v.setPathData(this.keyframes[0].s[0].c, len);
        this.pv = shape_pool.clone(this.v);
        this.localShapeCollection = shapeCollection_pool.newShapeCollection();
        this.paths = this.localShapeCollection;
        this.paths.addShape(this.v);
        this.lastFrame = initFrame;
        this.reset = resetShape;
        this._caching = {lastFrame: initFrame, lastIndex: 0};
        this.effectsSequence = [interpolateShapeCurrentTime.bind(this)];
    }
    KeyframedShapeProperty.prototype.getValue = processEffectsSequence;
    KeyframedShapeProperty.prototype.interpolateShape = interpolateShape;
    KeyframedShapeProperty.prototype.setVValue = setVValue;
    KeyframedShapeProperty.prototype.addEffect = addEffect;

    var EllShapeProperty = (function(){

        var cPoint = roundCorner;

        function EllShapeProperty(elem,data) {
            /*this.v = {
                v: createSizedArray(4),
                i: createSizedArray(4),
                o: createSizedArray(4),
                c: true
            };*/
            this.v = shape_pool.newElement();
            this.v.setPathData(true, 4);
            this.localShapeCollection = shapeCollection_pool.newShapeCollection();
            this.paths = this.localShapeCollection;
            this.localShapeCollection.addShape(this.v);
            this.d = data.d;
            this.elem = elem;
            this.comp = elem.comp;
            this.frameId = -1;
            this.initDynamicPropertyContainer(elem);
            this.p = PropertyFactory.getProp(elem,data.p,1,0,this);
            this.s = PropertyFactory.getProp(elem,data.s,1,0,this);
            if(this.dynamicProperties.length){
                this.k = true;
            }else{
                this.k = false;
                this.convertEllToPath();
            }
        };

        EllShapeProperty.prototype = {
            reset: resetShape,
            getValue: function (){
                if(this.elem.globalData.frameId === this.frameId){
                    return;
                }
                this.frameId = this.elem.globalData.frameId;
                this.iterateDynamicProperties();

                if(this._mdf){
                    this.convertEllToPath();
                }
            },
            convertEllToPath: function() {
                var p0 = this.p.v[0], p1 = this.p.v[1], s0 = this.s.v[0]/2, s1 = this.s.v[1]/2;
                var _cw = this.d !== 3;
                var _v = this.v;
                _v.v[0][0] = p0;
                _v.v[0][1] = p1 - s1;
                _v.v[1][0] = _cw ? p0 + s0 : p0 - s0;
                _v.v[1][1] = p1;
                _v.v[2][0] = p0;
                _v.v[2][1] = p1 + s1;
                _v.v[3][0] = _cw ? p0 - s0 : p0 + s0;
                _v.v[3][1] = p1;
                _v.i[0][0] = _cw ? p0 - s0 * cPoint : p0 + s0 * cPoint;
                _v.i[0][1] = p1 - s1;
                _v.i[1][0] = _cw ? p0 + s0 : p0 - s0;
                _v.i[1][1] = p1 - s1 * cPoint;
                _v.i[2][0] = _cw ? p0 + s0 * cPoint : p0 - s0 * cPoint;
                _v.i[2][1] = p1 + s1;
                _v.i[3][0] = _cw ? p0 - s0 : p0 + s0;
                _v.i[3][1] = p1 + s1 * cPoint;
                _v.o[0][0] = _cw ? p0 + s0 * cPoint : p0 - s0 * cPoint;
                _v.o[0][1] = p1 - s1;
                _v.o[1][0] = _cw ? p0 + s0 : p0 - s0;
                _v.o[1][1] = p1 + s1 * cPoint;
                _v.o[2][0] = _cw ? p0 - s0 * cPoint : p0 + s0 * cPoint;
                _v.o[2][1] = p1 + s1;
                _v.o[3][0] = _cw ? p0 - s0 : p0 + s0;
                _v.o[3][1] = p1 - s1 * cPoint;
            }
        }

        extendPrototype([DynamicPropertyContainer], EllShapeProperty);

        return EllShapeProperty;
    }());

    var StarShapeProperty = (function() {

        function StarShapeProperty(elem,data) {
            this.v = shape_pool.newElement();
            this.v.setPathData(true, 0);
            this.elem = elem;
            this.comp = elem.comp;
            this.data = data;
            this.frameId = -1;
            this.d = data.d;
            this.initDynamicPropertyContainer(elem);
            if(data.sy === 1){
                this.ir = PropertyFactory.getProp(elem,data.ir,0,0,this);
                this.is = PropertyFactory.getProp(elem,data.is,0,0.01,this);
                this.convertToPath = this.convertStarToPath;
            } else {
                this.convertToPath = this.convertPolygonToPath;
            }
            this.pt = PropertyFactory.getProp(elem,data.pt,0,0,this);
            this.p = PropertyFactory.getProp(elem,data.p,1,0,this);
            this.r = PropertyFactory.getProp(elem,data.r,0,degToRads,this);
            this.or = PropertyFactory.getProp(elem,data.or,0,0,this);
            this.os = PropertyFactory.getProp(elem,data.os,0,0.01,this);
            this.localShapeCollection = shapeCollection_pool.newShapeCollection();
            this.localShapeCollection.addShape(this.v);
            this.paths = this.localShapeCollection;
            if(this.dynamicProperties.length){
                this.k = true;
            }else{
                this.k = false;
                this.convertToPath();
            }
        };

        StarShapeProperty.prototype = {
            reset: resetShape,
            getValue: function() {
                if(this.elem.globalData.frameId === this.frameId){
                    return;
                }
                this.frameId = this.elem.globalData.frameId;
                this.iterateDynamicProperties();
                if(this._mdf){
                    this.convertToPath();
                }
            },
            convertStarToPath: function() {
                var numPts = Math.floor(this.pt.v)*2;
                var angle = Math.PI*2/numPts;
                /*this.v.v.length = numPts;
                this.v.i.length = numPts;
                this.v.o.length = numPts;*/
                var longFlag = true;
                var longRad = this.or.v;
                var shortRad = this.ir.v;
                var longRound = this.os.v;
                var shortRound = this.is.v;
                var longPerimSegment = 2*Math.PI*longRad/(numPts*2);
                var shortPerimSegment = 2*Math.PI*shortRad/(numPts*2);
                var i, rad,roundness,perimSegment, currentAng = -Math.PI/ 2;
                currentAng += this.r.v;
                var dir = this.data.d === 3 ? -1 : 1;
                this.v._length = 0;
                for(i=0;i<numPts;i+=1){
                    rad = longFlag ? longRad : shortRad;
                    roundness = longFlag ? longRound : shortRound;
                    perimSegment = longFlag ? longPerimSegment : shortPerimSegment;
                    var x = rad * Math.cos(currentAng);
                    var y = rad * Math.sin(currentAng);
                    var ox = x === 0 && y === 0 ? 0 : y/Math.sqrt(x*x + y*y);
                    var oy = x === 0 && y === 0 ? 0 : -x/Math.sqrt(x*x + y*y);
                    x +=  + this.p.v[0];
                    y +=  + this.p.v[1];
                    this.v.setTripleAt(x,y,x-ox*perimSegment*roundness*dir,y-oy*perimSegment*roundness*dir,x+ox*perimSegment*roundness*dir,y+oy*perimSegment*roundness*dir, i, true);

                    /*this.v.v[i] = [x,y];
                    this.v.i[i] = [x+ox*perimSegment*roundness*dir,y+oy*perimSegment*roundness*dir];
                    this.v.o[i] = [x-ox*perimSegment*roundness*dir,y-oy*perimSegment*roundness*dir];
                    this.v._length = numPts;*/
                    longFlag = !longFlag;
                    currentAng += angle*dir;
                }
            },
            convertPolygonToPath: function() {
                var numPts = Math.floor(this.pt.v);
                var angle = Math.PI*2/numPts;
                var rad = this.or.v;
                var roundness = this.os.v;
                var perimSegment = 2*Math.PI*rad/(numPts*4);
                var i, currentAng = -Math.PI/ 2;
                var dir = this.data.d === 3 ? -1 : 1;
                currentAng += this.r.v;
                this.v._length = 0;
                for(i=0;i<numPts;i+=1){
                    var x = rad * Math.cos(currentAng);
                    var y = rad * Math.sin(currentAng);
                    var ox = x === 0 && y === 0 ? 0 : y/Math.sqrt(x*x + y*y);
                    var oy = x === 0 && y === 0 ? 0 : -x/Math.sqrt(x*x + y*y);
                    x +=  + this.p.v[0];
                    y +=  + this.p.v[1];
                    this.v.setTripleAt(x,y,x-ox*perimSegment*roundness*dir,y-oy*perimSegment*roundness*dir,x+ox*perimSegment*roundness*dir,y+oy*perimSegment*roundness*dir, i, true);
                    currentAng += angle*dir;
                }
                this.paths.length = 0;
                this.paths[0] = this.v;
            }

        }
        extendPrototype([DynamicPropertyContainer], StarShapeProperty);

        return StarShapeProperty;
    }());

    var RectShapeProperty = (function() {

         function RectShapeProperty(elem,data) {
            this.v = shape_pool.newElement();
            this.v.c = true;
            this.localShapeCollection = shapeCollection_pool.newShapeCollection();
            this.localShapeCollection.addShape(this.v);
            this.paths = this.localShapeCollection;
            this.elem = elem;
            this.comp = elem.comp;
            this.frameId = -1;
            this.d = data.d;
            this.initDynamicPropertyContainer(elem);
            this.p = PropertyFactory.getProp(elem,data.p,1,0,this);
            this.s = PropertyFactory.getProp(elem,data.s,1,0,this);
            this.r = PropertyFactory.getProp(elem,data.r,0,0,this);
            if(this.dynamicProperties.length){
                this.k = true;
            }else{
                this.k = false;
                this.convertRectToPath();
            }
        };

        RectShapeProperty.prototype = {
            convertRectToPath: function (){
                var p0 = this.p.v[0], p1 = this.p.v[1], v0 = this.s.v[0]/2, v1 = this.s.v[1]/2;
                var round = bm_min(v0,v1,this.r.v);
                var cPoint = round*(1-roundCorner);
                this.v._length = 0;

                if(this.d === 2 || this.d === 1) {
                    this.v.setTripleAt(p0+v0, p1-v1+round,p0+v0, p1-v1+round,p0+v0,p1-v1+cPoint,0, true);
                    this.v.setTripleAt(p0+v0, p1+v1-round,p0+v0, p1+v1-cPoint,p0+v0, p1+v1-round,1, true);
                    if(round!== 0){
                        this.v.setTripleAt(p0+v0-round, p1+v1,p0+v0-round,p1+v1,p0+v0-cPoint,p1+v1,2, true);
                        this.v.setTripleAt(p0-v0+round,p1+v1,p0-v0+cPoint,p1+v1,p0-v0+round,p1+v1,3, true);
                        this.v.setTripleAt(p0-v0,p1+v1-round,p0-v0,p1+v1-round,p0-v0,p1+v1-cPoint,4, true);
                        this.v.setTripleAt(p0-v0,p1-v1+round,p0-v0,p1-v1+cPoint,p0-v0,p1-v1+round,5, true);
                        this.v.setTripleAt(p0-v0+round,p1-v1,p0-v0+round,p1-v1,p0-v0+cPoint,p1-v1,6, true);
                        this.v.setTripleAt(p0+v0-round,p1-v1,p0+v0-cPoint,p1-v1,p0+v0-round,p1-v1,7, true);
                    } else {
                        this.v.setTripleAt(p0-v0,p1+v1,p0-v0+cPoint,p1+v1,p0-v0,p1+v1,2);
                        this.v.setTripleAt(p0-v0,p1-v1,p0-v0,p1-v1+cPoint,p0-v0,p1-v1,3);
                    }
                }else{
                    this.v.setTripleAt(p0+v0,p1-v1+round,p0+v0,p1-v1+cPoint,p0+v0,p1-v1+round,0, true);
                    if(round!== 0){
                        this.v.setTripleAt(p0+v0-round,p1-v1,p0+v0-round,p1-v1,p0+v0-cPoint,p1-v1,1, true);
                        this.v.setTripleAt(p0-v0+round,p1-v1,p0-v0+cPoint,p1-v1,p0-v0+round,p1-v1,2, true);
                        this.v.setTripleAt(p0-v0,p1-v1+round,p0-v0,p1-v1+round,p0-v0,p1-v1+cPoint,3, true);
                        this.v.setTripleAt(p0-v0,p1+v1-round,p0-v0,p1+v1-cPoint,p0-v0,p1+v1-round,4, true);
                        this.v.setTripleAt(p0-v0+round,p1+v1,p0-v0+round,p1+v1,p0-v0+cPoint,p1+v1,5, true);
                        this.v.setTripleAt(p0+v0-round,p1+v1,p0+v0-cPoint,p1+v1,p0+v0-round,p1+v1,6, true);
                        this.v.setTripleAt(p0+v0,p1+v1-round,p0+v0,p1+v1-round,p0+v0,p1+v1-cPoint,7, true);
                    } else {
                        this.v.setTripleAt(p0-v0,p1-v1,p0-v0+cPoint,p1-v1,p0-v0,p1-v1,1, true);
                        this.v.setTripleAt(p0-v0,p1+v1,p0-v0,p1+v1-cPoint,p0-v0,p1+v1,2, true);
                        this.v.setTripleAt(p0+v0,p1+v1,p0+v0-cPoint,p1+v1,p0+v0,p1+v1,3, true);

                    }
                }
            },
            getValue: function(frameNum){
                if(this.elem.globalData.frameId === this.frameId){
                    return;
                }
                this.frameId = this.elem.globalData.frameId;
                this.iterateDynamicProperties();
                if(this._mdf){
                    this.convertRectToPath();
                }

            },
            reset: resetShape
        }
        extendPrototype([DynamicPropertyContainer], RectShapeProperty);

        return RectShapeProperty;
    }());

    function getShapeProp(elem,data,type){
        var prop;
        if(type === 3 || type === 4){
            var dataProp = type === 3 ? data.pt : data.ks;
            var keys = dataProp.k;
            if(keys.length){
                prop = new KeyframedShapeProperty(elem, data, type);
            }else{
                prop = new ShapeProperty(elem, data, type);
            }
        }else if(type === 5){
            prop = new RectShapeProperty(elem, data);
        }else if(type === 6){
            prop = new EllShapeProperty(elem, data);
        }else if(type === 7){
            prop = new StarShapeProperty(elem, data);
        }
        if(prop.k){
            elem.addDynamicProperty(prop);
        }
        return prop;
    }

    function getConstructorFunction() {
        return ShapeProperty;
    }

    function getKeyframedConstructorFunction() {
        return KeyframedShapeProperty;
    }

    var ob = {};
    ob.getShapeProp = getShapeProp;
    ob.getConstructorFunction = getConstructorFunction;
    ob.getKeyframedConstructorFunction = getKeyframedConstructorFunction;
    return ob;
}());
var ShapeModifiers = (function(){
    var ob = {};
    var modifiers = {};
    ob.registerModifier = registerModifier;
    ob.getModifier = getModifier;

    function registerModifier(nm,factory){
        if(!modifiers[nm]){
            modifiers[nm] = factory;
        }
    }

    function getModifier(nm,elem, data){
        return new modifiers[nm](elem, data);
    }

    return ob;
}());

function ShapeModifier(){}
ShapeModifier.prototype.initModifierProperties = function(){};
ShapeModifier.prototype.addShapeToModifier = function(){};
ShapeModifier.prototype.addShape = function(data){
    if(!this.closed){
        var shapeData = {shape:data.sh, data: data, localShapeCollection:shapeCollection_pool.newShapeCollection()};
        this.shapes.push(shapeData);
        this.addShapeToModifier(shapeData);
        if(this._isAnimated) {
            data.setAsAnimated();
        }
    }
};
ShapeModifier.prototype.init = function(elem,data){
    this.shapes = [];
    this.elem = elem;
    this.initDynamicPropertyContainer(elem);
    this.initModifierProperties(elem,data);
    this.frameId = initialDefaultFrame;
    this.closed = false;
    this.k = false;
    if(this.dynamicProperties.length){
        this.k = true;
    }else{
        this.getValue(true);
    }
};
ShapeModifier.prototype.processKeys = function(){
    if(this.elem.globalData.frameId === this.frameId){
        return;
    }
    this.frameId = this.elem.globalData.frameId;
    this.iterateDynamicProperties();
};

extendPrototype([DynamicPropertyContainer], ShapeModifier);
function TrimModifier(){
}
extendPrototype([ShapeModifier], TrimModifier);
TrimModifier.prototype.initModifierProperties = function(elem, data) {
    this.s = PropertyFactory.getProp(elem, data.s, 0, 0.01, this);
    this.e = PropertyFactory.getProp(elem, data.e, 0, 0.01, this);
    this.o = PropertyFactory.getProp(elem, data.o, 0, 0, this);
    this.sValue = 0;
    this.eValue = 0;
    this.getValue = this.processKeys;
    this.m = data.m;
    this._isAnimated = !!this.s.effectsSequence.length || !!this.e.effectsSequence.length || !!this.o.effectsSequence.length;
};

TrimModifier.prototype.addShapeToModifier = function(shapeData){
    shapeData.pathsData = [];
};

TrimModifier.prototype.calculateShapeEdges = function(s, e, shapeLength, addedLength, totalModifierLength) {
    var segments = [];
    if (e <= 1) {
        segments.push({
            s: s,
            e: e
        });
    } else if (s >= 1) {
        segments.push({
            s: s - 1,
            e: e - 1
        });
    } else {
        segments.push({
            s: s,
            e: 1
        });
        segments.push({
            s: 0,
            e: e - 1
        });
    }
    var shapeSegments = [];
    var i, len = segments.length, segmentOb;
    for (i = 0; i < len; i += 1) {
        segmentOb = segments[i];
        if (segmentOb.e * totalModifierLength < addedLength || segmentOb.s * totalModifierLength > addedLength + shapeLength) {

        } else {
            var shapeS, shapeE;
            if (segmentOb.s * totalModifierLength <= addedLength) {
                shapeS = 0;
            } else {
                shapeS = (segmentOb.s * totalModifierLength - addedLength) / shapeLength;
            }
            if(segmentOb.e * totalModifierLength >= addedLength + shapeLength) {
                shapeE = 1;
            } else {
                shapeE = ((segmentOb.e * totalModifierLength - addedLength) / shapeLength);
            }
            shapeSegments.push([shapeS, shapeE]);
        }
    }
    if (!shapeSegments.length) {
        shapeSegments.push([0, 0]);
    }
    return shapeSegments;
};

TrimModifier.prototype.releasePathsData = function(pathsData) {
    var i, len = pathsData.length;
    for (i = 0; i < len; i += 1) {
        segments_length_pool.release(pathsData[i]);
    }
    pathsData.length = 0;
    return pathsData;
};

TrimModifier.prototype.processShapes = function(_isFirstFrame) {
    var s, e;
    if (this._mdf || _isFirstFrame) {
        var o = (this.o.v % 360) / 360;
        if (o < 0) {
            o += 1;
        }
        s = (this.s.v > 1 ? 1 : this.s.v < 0 ? 0 : this.s.v) + o;
        e = (this.e.v > 1 ? 1 : this.e.v < 0 ? 0 : this.e.v) + o;
        if (s === e) {

        }
        if (s > e) {
            var _s = s;
            s = e;
            e = _s;
        }
        s = Math.round(s * 10000) * 0.0001;
        e = Math.round(e * 10000) * 0.0001;
        this.sValue = s;
        this.eValue = e;
    } else {
        s = this.sValue;
        e = this.eValue;
    }
    var shapePaths;
    var i, len = this.shapes.length, j, jLen;
    var pathsData, pathData, totalShapeLength, totalModifierLength = 0;

    if (e === s) {
        for (i = 0; i < len; i += 1) {
            this.shapes[i].localShapeCollection.releaseShapes();
            this.shapes[i].shape._mdf = true;
            this.shapes[i].shape.paths = this.shapes[i].localShapeCollection;
        }
    } else if (!((e === 1 && s === 0) || (e===0 && s === 1))){
        var segments = [], shapeData, localShapeCollection;
        for (i = 0; i < len; i += 1) {
            shapeData = this.shapes[i];
            // if shape hasn't changed and trim properties haven't changed, cached previous path can be used
            if (!shapeData.shape._mdf && !this._mdf && !_isFirstFrame && this.m !== 2) {
                shapeData.shape.paths = shapeData.localShapeCollection;
            } else {
                shapePaths = shapeData.shape.paths;
                jLen = shapePaths._length;
                totalShapeLength = 0;
                if (!shapeData.shape._mdf && shapeData.pathsData.length) {
                    totalShapeLength = shapeData.totalShapeLength;
                } else {
                    pathsData = this.releasePathsData(shapeData.pathsData);
                    for (j = 0; j < jLen; j += 1) {
                        pathData = bez.getSegmentsLength(shapePaths.shapes[j]);
                        pathsData.push(pathData);
                        totalShapeLength += pathData.totalLength;
                    }
                    shapeData.totalShapeLength = totalShapeLength;
                    shapeData.pathsData = pathsData;
                }

                totalModifierLength += totalShapeLength;
                shapeData.shape._mdf = true;
            }
        }
        var shapeS = s, shapeE = e, addedLength = 0, edges;
        for (i = len - 1; i >= 0; i -= 1) {
            shapeData = this.shapes[i];
            if (shapeData.shape._mdf) {
                localShapeCollection = shapeData.localShapeCollection;
                localShapeCollection.releaseShapes();
                //if m === 2 means paths are trimmed individually so edges need to be found for this specific shape relative to whoel group
                if (this.m === 2 && len > 1) {
                    edges = this.calculateShapeEdges(s, e, shapeData.totalShapeLength, addedLength, totalModifierLength);
                    addedLength += shapeData.totalShapeLength;
                } else {
                    edges = [[shapeS, shapeE]];
                }
                jLen = edges.length;
                for (j = 0; j < jLen; j += 1) {
                    shapeS = edges[j][0];
                    shapeE = edges[j][1];
                    segments.length = 0;
                    if (shapeE <= 1) {
                        segments.push({
                            s:shapeData.totalShapeLength * shapeS,
                            e:shapeData.totalShapeLength * shapeE
                        });
                    } else if (shapeS >= 1) {
                        segments.push({
                            s:shapeData.totalShapeLength * (shapeS - 1),
                            e:shapeData.totalShapeLength * (shapeE - 1)
                        });
                    } else {
                        segments.push({
                            s:shapeData.totalShapeLength * shapeS,
                            e:shapeData.totalShapeLength
                        });
                        segments.push({
                            s:0,
                            e:shapeData.totalShapeLength * (shapeE - 1)
                        });
                    }
                    var newShapesData = this.addShapes(shapeData,segments[0]);
                    if (segments[0].s !== segments[0].e) {
                        if (segments.length > 1) {
                            var lastShapeInCollection = shapeData.shape.paths.shapes[shapeData.shape.paths._length - 1];
                            if (lastShapeInCollection.c) {
                                var lastShape = newShapesData.pop();
                                this.addPaths(newShapesData, localShapeCollection);
                                newShapesData = this.addShapes(shapeData, segments[1], lastShape);
                            } else {
                                this.addPaths(newShapesData, localShapeCollection);
                                newShapesData = this.addShapes(shapeData, segments[1]);
                            }
                        }
                        this.addPaths(newShapesData, localShapeCollection);
                    }

                }
                shapeData.shape.paths = localShapeCollection;
            }
        }
    } else if (this._mdf) {
        for (i = 0; i < len; i += 1) {
            //Releasign Trim Cached paths data when no trim applied in case shapes are modified inbetween.
            //Don't remove this even if it's losing cached info.
            this.shapes[i].pathsData.length = 0;
            this.shapes[i].shape._mdf = true;
        }
    }
};

TrimModifier.prototype.addPaths = function(newPaths, localShapeCollection) {
    var i, len = newPaths.length;
    for (i = 0; i < len; i += 1) {
        localShapeCollection.addShape(newPaths[i]);
    }
};

TrimModifier.prototype.addSegment = function(pt1, pt2, pt3, pt4, shapePath, pos, newShape) {
    shapePath.setXYAt(pt2[0], pt2[1], 'o', pos);
    shapePath.setXYAt(pt3[0], pt3[1], 'i', pos + 1);
    if(newShape){
        shapePath.setXYAt(pt1[0], pt1[1], 'v', pos);
    }
    shapePath.setXYAt(pt4[0], pt4[1], 'v', pos + 1);
};

TrimModifier.prototype.addSegmentFromArray = function(points, shapePath, pos, newShape) {
    shapePath.setXYAt(points[1], points[5], 'o', pos);
    shapePath.setXYAt(points[2], points[6], 'i', pos + 1);
    if(newShape){
        shapePath.setXYAt(points[0], points[4], 'v', pos);
    }
    shapePath.setXYAt(points[3], points[7], 'v', pos + 1);
};

TrimModifier.prototype.addShapes = function(shapeData, shapeSegment, shapePath) {
    var pathsData = shapeData.pathsData;
    var shapePaths = shapeData.shape.paths.shapes;
    var i, len = shapeData.shape.paths._length, j, jLen;
    var addedLength = 0;
    var currentLengthData,segmentCount;
    var lengths;
    var segment;
    var shapes = [];
    var initPos;
    var newShape = true;
    if (!shapePath) {
        shapePath = shape_pool.newElement();
        segmentCount = 0;
        initPos = 0;
    } else {
        segmentCount = shapePath._length;
        initPos = shapePath._length;
    }
    shapes.push(shapePath);
    for (i = 0; i < len; i += 1) {
        lengths = pathsData[i].lengths;
        shapePath.c = shapePaths[i].c;
        jLen = shapePaths[i].c ? lengths.length : lengths.length + 1;
        for (j = 1; j < jLen; j +=1) {
            currentLengthData = lengths[j-1];
            if (addedLength + currentLengthData.addedLength < shapeSegment.s) {
                addedLength += currentLengthData.addedLength;
                shapePath.c = false;
            } else if(addedLength > shapeSegment.e) {
                shapePath.c = false;
                break;
            } else {
                if (shapeSegment.s <= addedLength && shapeSegment.e >= addedLength + currentLengthData.addedLength) {
                    this.addSegment(shapePaths[i].v[j - 1], shapePaths[i].o[j - 1], shapePaths[i].i[j], shapePaths[i].v[j], shapePath, segmentCount, newShape);
                    newShape = false;
                } else {
                    segment = bez.getNewSegment(shapePaths[i].v[j - 1], shapePaths[i].v[j], shapePaths[i].o[j - 1], shapePaths[i].i[j], (shapeSegment.s - addedLength)/currentLengthData.addedLength,(shapeSegment.e - addedLength)/currentLengthData.addedLength, lengths[j-1]);
                    this.addSegmentFromArray(segment, shapePath, segmentCount, newShape);
                    // this.addSegment(segment.pt1, segment.pt3, segment.pt4, segment.pt2, shapePath, segmentCount, newShape);
                    newShape = false;
                    shapePath.c = false;
                }
                addedLength += currentLengthData.addedLength;
                segmentCount += 1;
            }
        }
        if (shapePaths[i].c && lengths.length) {
            currentLengthData = lengths[j - 1];
            if (addedLength <= shapeSegment.e) {
                var segmentLength = lengths[j - 1].addedLength;
                if (shapeSegment.s <= addedLength && shapeSegment.e >= addedLength + segmentLength) {
                    this.addSegment(shapePaths[i].v[j - 1], shapePaths[i].o[j - 1], shapePaths[i].i[0], shapePaths[i].v[0], shapePath, segmentCount, newShape);
                    newShape = false;
                } else {
                    segment = bez.getNewSegment(shapePaths[i].v[j - 1], shapePaths[i].v[0], shapePaths[i].o[j - 1], shapePaths[i].i[0], (shapeSegment.s - addedLength) / segmentLength, (shapeSegment.e - addedLength) / segmentLength, lengths[j - 1]);
                    this.addSegmentFromArray(segment, shapePath, segmentCount, newShape);
                    // this.addSegment(segment.pt1, segment.pt3, segment.pt4, segment.pt2, shapePath, segmentCount, newShape);
                    newShape = false;
                    shapePath.c = false;
                }
            } else {
                shapePath.c = false;
            }
            addedLength += currentLengthData.addedLength;
            segmentCount += 1;
        }
        if (shapePath._length) {
            shapePath.setXYAt(shapePath.v[initPos][0], shapePath.v[initPos][1], 'i', initPos);
            shapePath.setXYAt(shapePath.v[shapePath._length - 1][0], shapePath.v[shapePath._length - 1][1],'o', shapePath._length - 1);
        }
        if (addedLength > shapeSegment.e) {
            break;
        }
        if (i < len - 1) {
            shapePath = shape_pool.newElement();
            newShape = true;
            shapes.push(shapePath);
            segmentCount = 0;
        }
    }
    return shapes;
};


ShapeModifiers.registerModifier('tm', TrimModifier);
function RoundCornersModifier(){}
extendPrototype([ShapeModifier],RoundCornersModifier);
RoundCornersModifier.prototype.initModifierProperties = function(elem,data){
    this.getValue = this.processKeys;
    this.rd = PropertyFactory.getProp(elem,data.r,0,null,this);
    this._isAnimated = !!this.rd.effectsSequence.length;
};

RoundCornersModifier.prototype.processPath = function(path, round){
    var cloned_path = shape_pool.newElement();
    cloned_path.c = path.c;
    var i, len = path._length;
    var currentV,currentI,currentO,closerV, newV,newO,newI,distance,newPosPerc,index = 0;
    var vX,vY,oX,oY,iX,iY;
    for(i=0;i<len;i+=1){
        currentV = path.v[i];
        currentO = path.o[i];
        currentI = path.i[i];
        if(currentV[0]===currentO[0] && currentV[1]===currentO[1] && currentV[0]===currentI[0] && currentV[1]===currentI[1]){
            if((i===0 || i === len - 1) && !path.c){
                cloned_path.setTripleAt(currentV[0],currentV[1],currentO[0],currentO[1],currentI[0],currentI[1],index);
                /*cloned_path.v[index] = currentV;
                cloned_path.o[index] = currentO;
                cloned_path.i[index] = currentI;*/
                index += 1;
            } else {
                if(i===0){
                    closerV = path.v[len-1];
                } else {
                    closerV = path.v[i-1];
                }
                distance = Math.sqrt(Math.pow(currentV[0]-closerV[0],2)+Math.pow(currentV[1]-closerV[1],2));
                newPosPerc = distance ? Math.min(distance/2,round)/distance : 0;
                vX = iX = currentV[0]+(closerV[0]-currentV[0])*newPosPerc;
                vY = iY = currentV[1]-(currentV[1]-closerV[1])*newPosPerc;
                oX = vX-(vX-currentV[0])*roundCorner;
                oY = vY-(vY-currentV[1])*roundCorner;
                cloned_path.setTripleAt(vX,vY,oX,oY,iX,iY,index);
                index += 1;

                if(i === len - 1){
                    closerV = path.v[0];
                } else {
                    closerV = path.v[i+1];
                }
                distance = Math.sqrt(Math.pow(currentV[0]-closerV[0],2)+Math.pow(currentV[1]-closerV[1],2));
                newPosPerc = distance ? Math.min(distance/2,round)/distance : 0;
                vX = oX = currentV[0]+(closerV[0]-currentV[0])*newPosPerc;
                vY = oY = currentV[1]+(closerV[1]-currentV[1])*newPosPerc;
                iX = vX-(vX-currentV[0])*roundCorner;
                iY = vY-(vY-currentV[1])*roundCorner;
                cloned_path.setTripleAt(vX,vY,oX,oY,iX,iY,index);
                index += 1;
            }
        } else {
            cloned_path.setTripleAt(path.v[i][0],path.v[i][1],path.o[i][0],path.o[i][1],path.i[i][0],path.i[i][1],index);
            index += 1;
        }
    }
    return cloned_path;
};

RoundCornersModifier.prototype.processShapes = function(_isFirstFrame){
    var shapePaths;
    var i, len = this.shapes.length;
    var j, jLen;
    var rd = this.rd.v;

    if(rd !== 0){
        var shapeData, newPaths, localShapeCollection;
        for(i=0;i<len;i+=1){
            shapeData = this.shapes[i];
            newPaths = shapeData.shape.paths;
            localShapeCollection = shapeData.localShapeCollection;
            if(!(!shapeData.shape._mdf && !this._mdf && !_isFirstFrame)){
                localShapeCollection.releaseShapes();
                shapeData.shape._mdf = true;
                shapePaths = shapeData.shape.paths.shapes;
                jLen = shapeData.shape.paths._length;
                for(j=0;j<jLen;j+=1){
                    localShapeCollection.addShape(this.processPath(shapePaths[j],rd));
                }
            }
            shapeData.shape.paths = shapeData.localShapeCollection;
        }

    }
    if(!this.dynamicProperties.length){
        this._mdf = false;
    }
};

ShapeModifiers.registerModifier('rd',RoundCornersModifier);
function RepeaterModifier(){}
extendPrototype([ShapeModifier], RepeaterModifier);

RepeaterModifier.prototype.initModifierProperties = function(elem,data){
    this.getValue = this.processKeys;
    this.c = PropertyFactory.getProp(elem,data.c,0,null,this);
    this.o = PropertyFactory.getProp(elem,data.o,0,null,this);
    this.tr = TransformPropertyFactory.getTransformProperty(elem,data.tr,this);
    this.so = PropertyFactory.getProp(elem,data.tr.so,0,0.01,this);
    this.eo = PropertyFactory.getProp(elem,data.tr.eo,0,0.01,this);
    this.data = data;
    if(!this.dynamicProperties.length){
        this.getValue(true);
    }
    this._isAnimated = !!this.dynamicProperties.length;
    this.pMatrix = new Matrix();
    this.rMatrix = new Matrix();
    this.sMatrix = new Matrix();
    this.tMatrix = new Matrix();
    this.matrix = new Matrix();
};

RepeaterModifier.prototype.applyTransforms = function(pMatrix, rMatrix, sMatrix, transform, perc, inv){
    var dir = inv ? -1 : 1;
    var scaleX = transform.s.v[0] + (1 - transform.s.v[0]) * (1 - perc);
    var scaleY = transform.s.v[1] + (1 - transform.s.v[1]) * (1 - perc);
    pMatrix.translate(transform.p.v[0] * dir * perc, transform.p.v[1] * dir * perc, transform.p.v[2]);
    rMatrix.translate(-transform.a.v[0], -transform.a.v[1], transform.a.v[2]);
    rMatrix.rotate(-transform.r.v * dir * perc);
    rMatrix.translate(transform.a.v[0], transform.a.v[1], transform.a.v[2]);
    sMatrix.translate(-transform.a.v[0], -transform.a.v[1], transform.a.v[2]);
    sMatrix.scale(inv ? 1/scaleX : scaleX, inv ? 1/scaleY : scaleY);
    sMatrix.translate(transform.a.v[0], transform.a.v[1], transform.a.v[2]);
};

RepeaterModifier.prototype.init = function(elem, arr, pos, elemsData) {
    this.elem = elem;
    this.arr = arr;
    this.pos = pos;
    this.elemsData = elemsData;
    this._currentCopies = 0;
    this._elements = [];
    this._groups = [];
    this.frameId = -1;
    this.initDynamicPropertyContainer(elem);
    this.initModifierProperties(elem,arr[pos]);
    var cont = 0;
    while(pos>0){
        pos -= 1;
        //this._elements.unshift(arr.splice(pos,1)[0]);
        this._elements.unshift(arr[pos]);
        cont += 1;
    }
    if(this.dynamicProperties.length){
        this.k = true;
    }else{
        this.getValue(true);
    }
};

RepeaterModifier.prototype.resetElements = function(elements){
    var i, len = elements.length;
    for(i = 0; i < len; i += 1) {
        elements[i]._processed = false;
        if(elements[i].ty === 'gr'){
            this.resetElements(elements[i].it);
        }
    }
};

RepeaterModifier.prototype.cloneElements = function(elements){
    var i, len = elements.length;
    var newElements = JSON.parse(JSON.stringify(elements));
    this.resetElements(newElements);
    return newElements;
};

RepeaterModifier.prototype.changeGroupRender = function(elements, renderFlag) {
    var i, len = elements.length;
    for(i = 0; i < len; i += 1) {
        elements[i]._render = renderFlag;
        if(elements[i].ty === 'gr') {
            this.changeGroupRender(elements[i].it, renderFlag);
        }
    }
};

RepeaterModifier.prototype.processShapes = function(_isFirstFrame) {
    var items, itemsTransform, i, dir, cont;
    if(this._mdf || _isFirstFrame){
        var copies = Math.ceil(this.c.v);
        if(this._groups.length < copies){
            while(this._groups.length < copies){
                var group = {
                    it:this.cloneElements(this._elements),
                    ty:'gr'
                };
                group.it.push({"a":{"a":0,"ix":1,"k":[0,0]},"nm":"Transform","o":{"a":0,"ix":7,"k":100},"p":{"a":0,"ix":2,"k":[0,0]},"r":{"a":1,"ix":6,"k":[{s:0,e:0,t:0},{s:0,e:0,t:1}]},"s":{"a":0,"ix":3,"k":[100,100]},"sa":{"a":0,"ix":5,"k":0},"sk":{"a":0,"ix":4,"k":0},"ty":"tr"});

                this.arr.splice(0,0,group);
                this._groups.splice(0,0,group);
                this._currentCopies += 1;
            }
            this.elem.reloadShapes();
        }
        cont = 0;
        var renderFlag;
        for(i = 0; i  <= this._groups.length - 1; i += 1){
            renderFlag = cont < copies;
            this._groups[i]._render = renderFlag;
            this.changeGroupRender(this._groups[i].it, renderFlag);
            cont += 1;
        }

        this._currentCopies = copies;
        ////

        var offset = this.o.v;
        var offsetModulo = offset%1;
        var roundOffset = offset > 0 ? Math.floor(offset) : Math.ceil(offset);
        var k;
        var tMat = this.tr.v.props;
        var pProps = this.pMatrix.props;
        var rProps = this.rMatrix.props;
        var sProps = this.sMatrix.props;
        this.pMatrix.reset();
        this.rMatrix.reset();
        this.sMatrix.reset();
        this.tMatrix.reset();
        this.matrix.reset();
        var iteration = 0;

        if(offset > 0) {
            while(iteration<roundOffset){
                this.applyTransforms(this.pMatrix, this.rMatrix, this.sMatrix, this.tr, 1, false);
                iteration += 1;
            }
            if(offsetModulo){
                this.applyTransforms(this.pMatrix, this.rMatrix, this.sMatrix, this.tr, offsetModulo, false);
                iteration += offsetModulo;
            }
        } else if(offset < 0) {
            while(iteration>roundOffset){
                this.applyTransforms(this.pMatrix, this.rMatrix, this.sMatrix, this.tr, 1, true);
                iteration -= 1;
            }
            if(offsetModulo){
                this.applyTransforms(this.pMatrix, this.rMatrix, this.sMatrix, this.tr, - offsetModulo, true);
                iteration -= offsetModulo;
            }
        }
        i = this.data.m === 1 ? 0 : this._currentCopies - 1;
        dir = this.data.m === 1 ? 1 : -1;
        cont = this._currentCopies;
        var j, jLen;
        while(cont){
            items = this.elemsData[i].it;
            itemsTransform = items[items.length - 1].transform.mProps.v.props;
            jLen = itemsTransform.length;
            items[items.length - 1].transform.mProps._mdf = true;
            items[items.length - 1].transform.op._mdf = true;
            items[items.length - 1].transform.op.v = this.so.v + (this.eo.v - this.so.v) * (i / (this._currentCopies - 1));
            if(iteration !== 0){
                if((i !== 0 && dir === 1) || (i !== this._currentCopies - 1 && dir === -1)){
                    this.applyTransforms(this.pMatrix, this.rMatrix, this.sMatrix, this.tr, 1, false);
                }
                this.matrix.transform(rProps[0],rProps[1],rProps[2],rProps[3],rProps[4],rProps[5],rProps[6],rProps[7],rProps[8],rProps[9],rProps[10],rProps[11],rProps[12],rProps[13],rProps[14],rProps[15]);
                this.matrix.transform(sProps[0],sProps[1],sProps[2],sProps[3],sProps[4],sProps[5],sProps[6],sProps[7],sProps[8],sProps[9],sProps[10],sProps[11],sProps[12],sProps[13],sProps[14],sProps[15]);
                this.matrix.transform(pProps[0],pProps[1],pProps[2],pProps[3],pProps[4],pProps[5],pProps[6],pProps[7],pProps[8],pProps[9],pProps[10],pProps[11],pProps[12],pProps[13],pProps[14],pProps[15]);

                for(j=0;j<jLen;j+=1) {
                    itemsTransform[j] = this.matrix.props[j];
                }
                this.matrix.reset();
            } else {
                this.matrix.reset();
                for(j=0;j<jLen;j+=1) {
                    itemsTransform[j] = this.matrix.props[j];
                }
            }
            iteration += 1;
            cont -= 1;
            i += dir;
        }
    } else {
        cont = this._currentCopies;
        i = 0;
        dir = 1;
        while(cont){
            items = this.elemsData[i].it;
            itemsTransform = items[items.length - 1].transform.mProps.v.props;
            items[items.length - 1].transform.mProps._mdf = false;
            items[items.length - 1].transform.op._mdf = false;
            cont -= 1;
            i += dir;
        }
    }
};

RepeaterModifier.prototype.addShape = function(){};

ShapeModifiers.registerModifier('rp',RepeaterModifier);
function ShapeCollection(){
	this._length = 0;
	this._maxLength = 4;
	this.shapes = createSizedArray(this._maxLength);
}

ShapeCollection.prototype.addShape = function(shapeData){
	if(this._length === this._maxLength){
		this.shapes = this.shapes.concat(createSizedArray(this._maxLength));
		this._maxLength *= 2;
	}
	this.shapes[this._length] = shapeData;
	this._length += 1;
};

ShapeCollection.prototype.releaseShapes = function(){
	var i;
	for(i = 0; i < this._length; i += 1) {
		shape_pool.release(this.shapes[i]);
	}
	this._length = 0;
};
function DashProperty(elem, data, renderer, container) {
    this.elem = elem;
    this.frameId = -1;
    this.dataProps = createSizedArray(data.length);
    this.renderer = renderer;
    this.k = false;
    this.dashStr = '';
    this.dashArray = createTypedArray('float32',  data.length ? data.length - 1 : 0);
    this.dashoffset = createTypedArray('float32',  1);
    this.initDynamicPropertyContainer(container);
    var i, len = data.length || 0, prop;
    for(i = 0; i < len; i += 1) {
        prop = PropertyFactory.getProp(elem,data[i].v,0, 0, this);
        this.k = prop.k || this.k;
        this.dataProps[i] = {n:data[i].n,p:prop};
    }
    if(!this.k){
        this.getValue(true);
    }
    this._isAnimated = this.k;
}

DashProperty.prototype.getValue = function(forceRender) {
    if(this.elem.globalData.frameId === this.frameId && !forceRender){
        return;
    }
    this.frameId = this.elem.globalData.frameId;
    this.iterateDynamicProperties();
    this._mdf = this._mdf || forceRender;
    if (this._mdf) {
        var i = 0, len = this.dataProps.length;
        if(this.renderer === 'svg') {
            this.dashStr = '';
        }
        for(i=0;i<len;i+=1){
            if(this.dataProps[i].n != 'o'){
                if(this.renderer === 'svg') {
                    this.dashStr += ' ' + this.dataProps[i].p.v;
                }else{
                    this.dashArray[i] = this.dataProps[i].p.v;
                }
            }else{
                this.dashoffset[0] = this.dataProps[i].p.v;
            }
        }
    }
};
extendPrototype([DynamicPropertyContainer], DashProperty);
function GradientProperty(elem,data,container){
    this.data = data;
    this.c = createTypedArray('uint8c', data.p*4);
    var cLength = data.k.k[0].s ? (data.k.k[0].s.length - data.p*4) : data.k.k.length - data.p*4;
    this.o = createTypedArray('float32', cLength);
    this._cmdf = false;
    this._omdf = false;
    this._collapsable = this.checkCollapsable();
    this._hasOpacity = cLength;
    this.initDynamicPropertyContainer(container);
    this.prop = PropertyFactory.getProp(elem,data.k,1,null,this);
    this.k = this.prop.k;
    this.getValue(true);
}

GradientProperty.prototype.comparePoints = function(values, points) {
    var i = 0, len = this.o.length/2, diff;
    while(i < len) {
        diff = Math.abs(values[i*4] - values[points*4 + i*2]);
        if(diff > 0.01){
            return false;
        }
        i += 1;
    }
    return true;
};

GradientProperty.prototype.checkCollapsable = function() {
    if (this.o.length/2 !== this.c.length/4) {
        return false;
    }
    if (this.data.k.k[0].s) {
        var i = 0, len = this.data.k.k.length;
        while (i < len) {
            if (!this.comparePoints(this.data.k.k[i].s, this.data.p)) {
                return false;
            }
            i += 1;
        }
    } else if(!this.comparePoints(this.data.k.k, this.data.p)) {
        return false;
    }
    return true;
};

GradientProperty.prototype.getValue = function(forceRender){
    this.prop.getValue();
    this._mdf = false;
    this._cmdf = false;
    this._omdf = false;
    if(this.prop._mdf || forceRender){
        var i, len = this.data.p*4;
        var mult, val;
        for(i=0;i<len;i+=1){
            mult = i%4 === 0 ? 100 : 255;
            val = Math.round(this.prop.v[i]*mult);
            if(this.c[i] !== val){
                this.c[i] = val;
                this._cmdf = !forceRender;
            }
        }
        if(this.o.length){
            len = this.prop.v.length;
            for(i=this.data.p*4;i<len;i+=1){
                mult = i%2 === 0 ? 100 : 1;
                val = i%2 === 0 ?  Math.round(this.prop.v[i]*100):this.prop.v[i];
                if(this.o[i-this.data.p*4] !== val){
                    this.o[i-this.data.p*4] = val;
                    this._omdf = !forceRender;
                }
            }
        }
        this._mdf = !forceRender;
    }
};

extendPrototype([DynamicPropertyContainer], GradientProperty);
var buildShapeString = function(pathNodes, length, closed, mat) {
	if(length === 0) {
            return '';
        }
        var _o = pathNodes.o;
        var _i = pathNodes.i;
        var _v = pathNodes.v;
        var i, shapeString = " M" + mat.applyToPointStringified(_v[0][0], _v[0][1]);
        for(i = 1; i < length; i += 1) {
            shapeString += " C" + mat.applyToPointStringified(_o[i - 1][0], _o[i - 1][1]) + " " + mat.applyToPointStringified(_i[i][0], _i[i][1]) + " " + mat.applyToPointStringified(_v[i][0], _v[i][1]);
        }
        if (closed && length) {
            shapeString += " C" + mat.applyToPointStringified(_o[i - 1][0], _o[i - 1][1]) + " " + mat.applyToPointStringified(_i[0][0], _i[0][1]) + " " + mat.applyToPointStringified(_v[0][0], _v[0][1]);
            shapeString += 'z';
        }
        return shapeString;
}
var ImagePreloader = (function(){

    var proxyImage = (function(){
        var canvas = createTag('canvas');
        canvas.width = 1;
        canvas.height = 1;
        var ctx = canvas.getContext('2d');
        ctx.fillStyle = '#FF0000';
        ctx.fillRect(0, 0, 1, 1);
        return canvas;
    }())

    function imageLoaded(){
        this.loadedAssets += 1;
        if(this.loadedAssets === this.totalImages){
            if(this.imagesLoadedCb) {
                this.imagesLoadedCb(null);
            }
        }
    }

    function getAssetsPath(assetData, assetsPath, original_path) {
        var path = '';
        if (assetData.e) {
            path = assetData.p;
        } else if(assetsPath) {
            var imagePath = assetData.p;
            if (imagePath.indexOf('images/') !== -1) {
                imagePath = imagePath.split('/')[1];
            }
            path = assetsPath + imagePath;
        } else {
            path = original_path;
            path += assetData.u ? assetData.u : '';
            path += assetData.p;
        }
        return path;
    }

    function createImageData(assetData) {
        var path = getAssetsPath(assetData, this.assetsPath, this.path);
        var img = createTag('img');
        img.crossOrigin = 'anonymous';
        img.addEventListener('load', this._imageLoaded.bind(this), false);
        img.addEventListener('error', function() {
            ob.img = proxyImage;
            this._imageLoaded();
        }.bind(this), false);
        img.src = path;
        var ob = {
            img: img,
            assetData: assetData
        }
        return ob;
    }

    function loadAssets(assets, cb){
        this.imagesLoadedCb = cb;
        var i, len = assets.length;
        for (i = 0; i < len; i += 1) {
            if(!assets[i].layers){
                this.totalImages += 1;
                this.images.push(this._createImageData(assets[i]));
            }
        }
    }

    function setPath(path){
        this.path = path || '';
    }

    function setAssetsPath(path){
        this.assetsPath = path || '';
    }

    function getImage(assetData) {
        var i = 0, len = this.images.length;
        while (i < len) {
            if (this.images[i].assetData === assetData) {
                return this.images[i].img;
            }
            i += 1;
        }
    }

    function destroy() {
        this.imagesLoadedCb = null;
        this.images.length = 0;
    }

    function loaded() {
        return this.totalImages === this.loadedAssets;
    }

    return function ImagePreloader(){
        this.loadAssets = loadAssets;
        this.setAssetsPath = setAssetsPath;
        this.setPath = setPath;
        this.loaded = loaded;
        this.destroy = destroy;
        this.getImage = getImage;
        this._createImageData = createImageData;
        this._imageLoaded = imageLoaded;
        this.assetsPath = '';
        this.path = '';
        this.totalImages = 0;
        this.loadedAssets = 0;
        this.imagesLoadedCb = null;
        this.images = [];
    };
}());
var featureSupport = (function(){
	var ob = {
		maskType: true
	};
	if (/MSIE 10/i.test(navigator.userAgent) || /MSIE 9/i.test(navigator.userAgent) || /rv:11.0/i.test(navigator.userAgent) || /Edge\/\d./i.test(navigator.userAgent)) {
	   ob.maskType = false;
	}
	return ob;
}());
var filtersFactory = (function(){
	var ob = {};
	ob.createFilter = createFilter;
	ob.createAlphaToLuminanceFilter = createAlphaToLuminanceFilter;

	function createFilter(filId){
        	var fil = createNS('filter');
        	fil.setAttribute('id',filId);
                fil.setAttribute('filterUnits','objectBoundingBox');
                fil.setAttribute('x','0%');
                fil.setAttribute('y','0%');
                fil.setAttribute('width','100%');
                fil.setAttribute('height','100%');
                return fil;
	}

	function createAlphaToLuminanceFilter(){
                var feColorMatrix = createNS('feColorMatrix');
                feColorMatrix.setAttribute('type','matrix');
                feColorMatrix.setAttribute('color-interpolation-filters','sRGB');
                feColorMatrix.setAttribute('values','0 0 0 1 0  0 0 0 1 0  0 0 0 1 0  0 0 0 1 1');
                return feColorMatrix;
	}

	return ob;
}());
var assetLoader = (function(){

	function formatResponse(xhr) {
		if(xhr.response && typeof xhr.response === 'object') {
			return xhr.response;
		} else if(xhr.response && typeof xhr.response === 'string') {
			return JSON.parse(xhr.response);
		} else if(xhr.responseText) {
			return JSON.parse(xhr.responseText);
		}
	}

	function loadAsset(path, callback, errorCallback) {
		var response;
		var xhr = new XMLHttpRequest();
		xhr.open('GET', path, true);
		// set responseType after calling open or IE will break.
		xhr.responseType = "json";
	    xhr.send();
	    xhr.onreadystatechange = function () {
	        if (xhr.readyState == 4) {
	            if(xhr.status == 200){
	            	response = formatResponse(xhr);
	            	callback(response);
	            }else{
	                try{
	            		response = formatResponse(xhr);
	            		callback(response);
	                }catch(err){
	                	if(errorCallback) {
	                		errorCallback(err);
	                	}
	                }
	            }
	        }
	    };
	}
	return {
		load: loadAsset
	}
}())

function TextAnimatorProperty(textData, renderType, elem){
    this._isFirstFrame = true;
	this._hasMaskedPath = false;
	this._frameId = -1;
	this._textData = textData;
	this._renderType = renderType;
    this._elem = elem;
	this._animatorsData = createSizedArray(this._textData.a.length);
	this._pathData = {};
	this._moreOptions = {
		alignment: {}
	};
	this.renderedLetters = [];
    this.lettersChangedFlag = false;
    this.initDynamicPropertyContainer(elem);

}

TextAnimatorProperty.prototype.searchProperties = function(){
    var i, len = this._textData.a.length, animatorProps;
    var getProp = PropertyFactory.getProp;
    for(i=0;i<len;i+=1){
        animatorProps = this._textData.a[i];
        this._animatorsData[i] = new TextAnimatorDataProperty(this._elem, animatorProps, this);
    }
    if(this._textData.p && 'm' in this._textData.p){
        this._pathData = {
            f: getProp(this._elem,this._textData.p.f,0,0,this),
            l: getProp(this._elem,this._textData.p.l,0,0,this),
            r: this._textData.p.r,
            m: this._elem.maskManager.getMaskProperty(this._textData.p.m)
        };
        this._hasMaskedPath = true;
    } else {
        this._hasMaskedPath = false;
    }
    this._moreOptions.alignment = getProp(this._elem,this._textData.m.a,1,0,this);
};

TextAnimatorProperty.prototype.getMeasures = function(documentData, lettersChangedFlag){
    this.lettersChangedFlag = lettersChangedFlag;
    if(!this._mdf && !this._isFirstFrame && !lettersChangedFlag && (!this._hasMaskedPath || !this._pathData.m._mdf)) {
        return;
    }
    this._isFirstFrame = false;
    var alignment = this._moreOptions.alignment.v;
    var animators = this._animatorsData;
    var textData = this._textData;
    var matrixHelper = this.mHelper;
    var renderType = this._renderType;
    var renderedLettersCount = this.renderedLetters.length;
    var data = this.data;
    var xPos,yPos;
    var i, len;
    var letters = documentData.l, pathInfo, currentLength, currentPoint, segmentLength, flag, pointInd, segmentInd, prevPoint, points, segments, partialLength, totalLength, perc, tanAngle, mask;
    if(this._hasMaskedPath) {
        mask = this._pathData.m;
        if(!this._pathData.n || this._pathData._mdf){
            var paths = mask.v;
            if(this._pathData.r){
                paths = paths.reverse();
            }
            // TODO: release bezier data cached from previous pathInfo: this._pathData.pi
            pathInfo = {
                tLength: 0,
                segments: []
            };
            len = paths._length - 1;
            var bezierData;
            totalLength = 0;
            for (i = 0; i < len; i += 1) {
                bezierData = bez.buildBezierData(paths.v[i]
                    , paths.v[i + 1]
                    , [paths.o[i][0] - paths.v[i][0], paths.o[i][1] - paths.v[i][1]]
                    , [paths.i[i + 1][0] - paths.v[i + 1][0], paths.i[i + 1][1] - paths.v[i + 1][1]]);
                pathInfo.tLength += bezierData.segmentLength;
                pathInfo.segments.push(bezierData);
                totalLength += bezierData.segmentLength;
            }
            i = len;
            if (mask.v.c) {
                bezierData = bez.buildBezierData(paths.v[i]
                    , paths.v[0]
                    , [paths.o[i][0] - paths.v[i][0], paths.o[i][1] - paths.v[i][1]]
                    , [paths.i[0][0] - paths.v[0][0], paths.i[0][1] - paths.v[0][1]]);
                pathInfo.tLength += bezierData.segmentLength;
                pathInfo.segments.push(bezierData);
                totalLength += bezierData.segmentLength;
            }
            this._pathData.pi = pathInfo;
        }
        pathInfo = this._pathData.pi;

        currentLength = this._pathData.f.v;
        segmentInd = 0;
        pointInd = 1;
        segmentLength = 0;
        flag = true;
        segments = pathInfo.segments;
        if (currentLength < 0 && mask.v.c) {
            if (pathInfo.tLength < Math.abs(currentLength)) {
                currentLength = -Math.abs(currentLength) % pathInfo.tLength;
            }
            segmentInd = segments.length - 1;
            points = segments[segmentInd].points;
            pointInd = points.length - 1;
            while (currentLength < 0) {
                currentLength += points[pointInd].partialLength;
                pointInd -= 1;
                if (pointInd < 0) {
                    segmentInd -= 1;
                    points = segments[segmentInd].points;
                    pointInd = points.length - 1;
                }
            }

        }
        points = segments[segmentInd].points;
        prevPoint = points[pointInd - 1];
        currentPoint = points[pointInd];
        partialLength = currentPoint.partialLength;
    }


    len = letters.length;
    xPos = 0;
    yPos = 0;
    var yOff = documentData.finalSize * 1.2 * 0.714;
    var firstLine = true;
    var animatorProps, animatorSelector;
    var j, jLen;
    var letterValue;

    jLen = animators.length;
    var lastLetter;

    var mult, ind = -1, offf, xPathPos, yPathPos;
    var initPathPos = currentLength,initSegmentInd = segmentInd, initPointInd = pointInd, currentLine = -1;
    var elemOpacity;
    var sc,sw,fc,k;
    var lineLength = 0;
    var letterSw, letterSc, letterFc, letterM = '', letterP = this.defaultPropsArray, letterO;

    //
    if(documentData.j === 2 || documentData.j === 1) {
        var animatorJustifyOffset = 0;
        var animatorFirstCharOffset = 0;
        var justifyOffsetMult = documentData.j === 2 ? -0.5 : -1;
        var lastIndex = 0;
        var isNewLine = true;

        for (i = 0; i < len; i += 1) {
            if (letters[i].n) {
                if(animatorJustifyOffset) {
                    animatorJustifyOffset += animatorFirstCharOffset;
                }
                while (lastIndex < i) {
                    letters[lastIndex].animatorJustifyOffset = animatorJustifyOffset;
                    lastIndex += 1;
                }
                animatorJustifyOffset = 0;
                isNewLine = true;
            } else {
                for (j = 0; j < jLen; j += 1) {
                    animatorProps = animators[j].a;
                    if (animatorProps.t.propType) {
                        if (isNewLine && documentData.j === 2) {
                            animatorFirstCharOffset += animatorProps.t.v * justifyOffsetMult;
                        }
                        animatorSelector = animators[j].s;
                        mult = animatorSelector.getMult(letters[i].anIndexes[j], textData.a[j].s.totalChars);
                        if (mult.length) {
                            animatorJustifyOffset += animatorProps.t.v*mult[0] * justifyOffsetMult;
                        } else {
                            animatorJustifyOffset += animatorProps.t.v*mult * justifyOffsetMult;
                        }
                    }
                }
                isNewLine = false;
            }
        }
        if(animatorJustifyOffset) {
            animatorJustifyOffset += animatorFirstCharOffset;
        }
        while(lastIndex < i) {
            letters[lastIndex].animatorJustifyOffset = animatorJustifyOffset;
            lastIndex += 1;
        }
    }
    //

    for( i = 0; i < len; i += 1) {

        matrixHelper.reset();
        elemOpacity = 1;
        if(letters[i].n) {
            xPos = 0;
            yPos += documentData.yOffset;
            yPos += firstLine ? 1 : 0;
            currentLength = initPathPos ;
            firstLine = false;
            lineLength = 0;
            if(this._hasMaskedPath) {
                segmentInd = initSegmentInd;
                pointInd = initPointInd;
                points = segments[segmentInd].points;
                prevPoint = points[pointInd - 1];
                currentPoint = points[pointInd];
                partialLength = currentPoint.partialLength;
                segmentLength = 0;
            }
            letterO = letterSw = letterFc = letterM = '';
            letterP = this.defaultPropsArray;
        }else{
            if(this._hasMaskedPath) {
                if(currentLine !== letters[i].line){
                    switch(documentData.j){
                        case 1:
                            currentLength += totalLength - documentData.lineWidths[letters[i].line];
                            break;
                        case 2:
                            currentLength += (totalLength - documentData.lineWidths[letters[i].line])/2;
                            break;
                    }
                    currentLine = letters[i].line;
                }
                if (ind !== letters[i].ind) {
                    if (letters[ind]) {
                        currentLength += letters[ind].extra;
                    }
                    currentLength += letters[i].an / 2;
                    ind = letters[i].ind;
                }
                currentLength += alignment[0] * letters[i].an / 200;
                var animatorOffset = 0;
                for (j = 0; j < jLen; j += 1) {
                    animatorProps = animators[j].a;
                    if (animatorProps.p.propType) {
                        animatorSelector = animators[j].s;
                        mult = animatorSelector.getMult(letters[i].anIndexes[j],textData.a[j].s.totalChars);
                        if(mult.length){
                            animatorOffset += animatorProps.p.v[0] * mult[0];
                        } else{
                            animatorOffset += animatorProps.p.v[0] * mult;
                        }

                    }
                    if (animatorProps.a.propType) {
                        animatorSelector = animators[j].s;
                        mult = animatorSelector.getMult(letters[i].anIndexes[j],textData.a[j].s.totalChars);
                        if(mult.length){
                            animatorOffset += animatorProps.a.v[0] * mult[0];
                        } else{
                            animatorOffset += animatorProps.a.v[0] * mult;
                        }

                    }
                }
                flag = true;
                while (flag) {
                    if (segmentLength + partialLength >= currentLength + animatorOffset || !points) {
                        perc = (currentLength + animatorOffset - segmentLength) / currentPoint.partialLength;
                        xPathPos = prevPoint.point[0] + (currentPoint.point[0] - prevPoint.point[0]) * perc;
                        yPathPos = prevPoint.point[1] + (currentPoint.point[1] - prevPoint.point[1]) * perc;
                        matrixHelper.translate(-alignment[0]*letters[i].an/200, -(alignment[1] * yOff / 100));
                        flag = false;
                    } else if (points) {
                        segmentLength += currentPoint.partialLength;
                        pointInd += 1;
                        if (pointInd >= points.length) {
                            pointInd = 0;
                            segmentInd += 1;
                            if (!segments[segmentInd]) {
                                if (mask.v.c) {
                                    pointInd = 0;
                                    segmentInd = 0;
                                    points = segments[segmentInd].points;
                                } else {
                                    segmentLength -= currentPoint.partialLength;
                                    points = null;
                                }
                            } else {
                                points = segments[segmentInd].points;
                            }
                        }
                        if (points) {
                            prevPoint = currentPoint;
                            currentPoint = points[pointInd];
                            partialLength = currentPoint.partialLength;
                        }
                    }
                }
                offf = letters[i].an / 2 - letters[i].add;
                matrixHelper.translate(-offf, 0, 0);
            } else {
                offf = letters[i].an/2 - letters[i].add;
                matrixHelper.translate(-offf,0,0);

                // Grouping alignment
                matrixHelper.translate(-alignment[0]*letters[i].an/200, -alignment[1]*yOff/100, 0);
            }

            lineLength += letters[i].l/2;
            for(j=0;j<jLen;j+=1){
                animatorProps = animators[j].a;
                if (animatorProps.t.propType) {
                    animatorSelector = animators[j].s;
                    mult = animatorSelector.getMult(letters[i].anIndexes[j],textData.a[j].s.totalChars);
                    //This condition is to prevent applying tracking to first character in each line. Might be better to use a boolean "isNewLine"
                    if(xPos !== 0 || documentData.j !== 0) {
                        if(this._hasMaskedPath) {
                            if(mult.length) {
                                currentLength += animatorProps.t.v*mult[0];
                            } else {
                                currentLength += animatorProps.t.v*mult;
                            }
                        }else{
                            if(mult.length) {
                                xPos += animatorProps.t.v*mult[0];
                            } else {
                                xPos += animatorProps.t.v*mult;
                            }
                        }
                    }
                }
            }
            lineLength += letters[i].l/2;
            if(documentData.strokeWidthAnim) {
                sw = documentData.sw || 0;
            }
            if(documentData.strokeColorAnim) {
                if(documentData.sc){
                    sc = [documentData.sc[0], documentData.sc[1], documentData.sc[2]];
                }else{
                    sc = [0,0,0];
                }
            }
            if(documentData.fillColorAnim && documentData.fc) {
                fc = [documentData.fc[0], documentData.fc[1], documentData.fc[2]];
            }
            for(j=0;j<jLen;j+=1){
                animatorProps = animators[j].a;
                if (animatorProps.a.propType) {
                    animatorSelector = animators[j].s;
                    mult = animatorSelector.getMult(letters[i].anIndexes[j],textData.a[j].s.totalChars);

                    if(mult.length){
                        matrixHelper.translate(-animatorProps.a.v[0]*mult[0], -animatorProps.a.v[1]*mult[1], animatorProps.a.v[2]*mult[2]);
                    } else {
                        matrixHelper.translate(-animatorProps.a.v[0]*mult, -animatorProps.a.v[1]*mult, animatorProps.a.v[2]*mult);
                    }
                }
            }
            for(j=0;j<jLen;j+=1){
                animatorProps = animators[j].a;
                if (animatorProps.s.propType) {
                    animatorSelector = animators[j].s;
                    mult = animatorSelector.getMult(letters[i].anIndexes[j],textData.a[j].s.totalChars);
                    if(mult.length){
                        matrixHelper.scale(1+((animatorProps.s.v[0]-1)*mult[0]),1+((animatorProps.s.v[1]-1)*mult[1]),1);
                    } else {
                        matrixHelper.scale(1+((animatorProps.s.v[0]-1)*mult),1+((animatorProps.s.v[1]-1)*mult),1);
                    }
                }
            }
            for(j=0;j<jLen;j+=1) {
                animatorProps = animators[j].a;
                animatorSelector = animators[j].s;
                mult = animatorSelector.getMult(letters[i].anIndexes[j],textData.a[j].s.totalChars);
                if (animatorProps.sk.propType) {
                    if(mult.length) {
                        matrixHelper.skewFromAxis(-animatorProps.sk.v * mult[0], animatorProps.sa.v * mult[1]);
                    } else {
                        matrixHelper.skewFromAxis(-animatorProps.sk.v * mult, animatorProps.sa.v * mult);
                    }
                }
                if (animatorProps.r.propType) {
                    if(mult.length) {
                        matrixHelper.rotateZ(-animatorProps.r.v * mult[2]);
                    } else {
                        matrixHelper.rotateZ(-animatorProps.r.v * mult);
                    }
                }
                if (animatorProps.ry.propType) {

                    if(mult.length) {
                        matrixHelper.rotateY(animatorProps.ry.v*mult[1]);
                    }else{
                        matrixHelper.rotateY(animatorProps.ry.v*mult);
                    }
                }
                if (animatorProps.rx.propType) {
                    if(mult.length) {
                        matrixHelper.rotateX(animatorProps.rx.v*mult[0]);
                    } else {
                        matrixHelper.rotateX(animatorProps.rx.v*mult);
                    }
                }
                if (animatorProps.o.propType) {
                    if(mult.length) {
                        elemOpacity += ((animatorProps.o.v)*mult[0] - elemOpacity)*mult[0];
                    } else {
                        elemOpacity += ((animatorProps.o.v)*mult - elemOpacity)*mult;
                    }
                }
                if (documentData.strokeWidthAnim && animatorProps.sw.propType) {
                    if(mult.length) {
                        sw += animatorProps.sw.v*mult[0];
                    } else {
                        sw += animatorProps.sw.v*mult;
                    }
                }
                if (documentData.strokeColorAnim && animatorProps.sc.propType) {
                    for(k=0;k<3;k+=1){
                        if(mult.length) {
                            sc[k] = sc[k] + (animatorProps.sc.v[k] - sc[k])*mult[0];
                        } else {
                            sc[k] = sc[k] + (animatorProps.sc.v[k] - sc[k])*mult;
                        }
                    }
                }
                if (documentData.fillColorAnim && documentData.fc) {
                    if(animatorProps.fc.propType){
                        for(k=0;k<3;k+=1){
                            if(mult.length) {
                                fc[k] = fc[k] + (animatorProps.fc.v[k] - fc[k])*mult[0];
                            } else {
                                fc[k] = fc[k] + (animatorProps.fc.v[k] - fc[k])*mult;
                            }
                        }
                    }
                    if(animatorProps.fh.propType){
                        if(mult.length) {
                            fc = addHueToRGB(fc,animatorProps.fh.v*mult[0]);
                        } else {
                            fc = addHueToRGB(fc,animatorProps.fh.v*mult);
                        }
                    }
                    if(animatorProps.fs.propType){
                        if(mult.length) {
                            fc = addSaturationToRGB(fc,animatorProps.fs.v*mult[0]);
                        } else {
                            fc = addSaturationToRGB(fc,animatorProps.fs.v*mult);
                        }
                    }
                    if(animatorProps.fb.propType){
                        if(mult.length) {
                            fc = addBrightnessToRGB(fc,animatorProps.fb.v*mult[0]);
                        } else {
                            fc = addBrightnessToRGB(fc,animatorProps.fb.v*mult);
                        }
                    }
                }
            }

            for(j=0;j<jLen;j+=1){
                animatorProps = animators[j].a;

                if (animatorProps.p.propType) {
                    animatorSelector = animators[j].s;
                    mult = animatorSelector.getMult(letters[i].anIndexes[j],textData.a[j].s.totalChars);
                    if(this._hasMaskedPath) {
                        if(mult.length) {
                            matrixHelper.translate(0, animatorProps.p.v[1] * mult[0], -animatorProps.p.v[2] * mult[1]);
                        } else {
                            matrixHelper.translate(0, animatorProps.p.v[1] * mult, -animatorProps.p.v[2] * mult);
                        }
                    }else{
                        if(mult.length) {
                            matrixHelper.translate(animatorProps.p.v[0] * mult[0], animatorProps.p.v[1] * mult[1], -animatorProps.p.v[2] * mult[2]);
                        } else {
                            matrixHelper.translate(animatorProps.p.v[0] * mult, animatorProps.p.v[1] * mult, -animatorProps.p.v[2] * mult);

                        }
                    }
                }
            }
            if(documentData.strokeWidthAnim){
                letterSw = sw < 0 ? 0 : sw;
            }
            if(documentData.strokeColorAnim){
                letterSc = 'rgb('+Math.round(sc[0]*255)+','+Math.round(sc[1]*255)+','+Math.round(sc[2]*255)+')';
            }
            if(documentData.fillColorAnim && documentData.fc){
                letterFc = 'rgb('+Math.round(fc[0]*255)+','+Math.round(fc[1]*255)+','+Math.round(fc[2]*255)+')';
            }

            if(this._hasMaskedPath) {
                matrixHelper.translate(0,-documentData.ls);

                matrixHelper.translate(0, alignment[1]*yOff/100 + yPos,0);
                if (textData.p.p) {
                    tanAngle = (currentPoint.point[1] - prevPoint.point[1]) / (currentPoint.point[0] - prevPoint.point[0]);
                    var rot = Math.atan(tanAngle) * 180 / Math.PI;
                    if (currentPoint.point[0] < prevPoint.point[0]) {
                        rot += 180;
                    }
                    matrixHelper.rotate(-rot * Math.PI / 180);
                }
                matrixHelper.translate(xPathPos, yPathPos, 0);
                currentLength -= alignment[0]*letters[i].an/200;
                if(letters[i+1] && ind !== letters[i+1].ind){
                    currentLength += letters[i].an / 2;
                    currentLength += documentData.tr/1000*documentData.finalSize;
                }
            }else{

                matrixHelper.translate(xPos,yPos,0);

                if(documentData.ps){
                    //matrixHelper.translate(documentData.ps[0],documentData.ps[1],0);
                    matrixHelper.translate(documentData.ps[0],documentData.ps[1] + documentData.ascent,0);
                }
                switch(documentData.j){
                    case 1:
                        matrixHelper.translate(letters[i].animatorJustifyOffset + documentData.justifyOffset + (documentData.boxWidth - documentData.lineWidths[letters[i].line]),0,0);
                        break;
                    case 2:
                        matrixHelper.translate(letters[i].animatorJustifyOffset + documentData.justifyOffset + (documentData.boxWidth - documentData.lineWidths[letters[i].line])/2,0,0);
                        break;
                }
                matrixHelper.translate(0,-documentData.ls);
                matrixHelper.translate(offf,0,0);
                matrixHelper.translate(alignment[0]*letters[i].an/200,alignment[1]*yOff/100,0);
                xPos += letters[i].l + documentData.tr/1000*documentData.finalSize;
            }
            if(renderType === 'html'){
                letterM = matrixHelper.toCSS();
            }else if(renderType === 'svg'){
                letterM = matrixHelper.to2dCSS();
            }else{
                letterP = [matrixHelper.props[0],matrixHelper.props[1],matrixHelper.props[2],matrixHelper.props[3],matrixHelper.props[4],matrixHelper.props[5],matrixHelper.props[6],matrixHelper.props[7],matrixHelper.props[8],matrixHelper.props[9],matrixHelper.props[10],matrixHelper.props[11],matrixHelper.props[12],matrixHelper.props[13],matrixHelper.props[14],matrixHelper.props[15]];
            }
            letterO = elemOpacity;
        }

        if(renderedLettersCount <= i) {
            letterValue = new LetterProps(letterO,letterSw,letterSc,letterFc,letterM,letterP);
            this.renderedLetters.push(letterValue);
            renderedLettersCount += 1;
            this.lettersChangedFlag = true;
        } else {
            letterValue = this.renderedLetters[i];
            this.lettersChangedFlag = letterValue.update(letterO, letterSw, letterSc, letterFc, letterM, letterP) || this.lettersChangedFlag;
        }
    }
};

TextAnimatorProperty.prototype.getValue = function(){
	if(this._elem.globalData.frameId === this._frameId){
        return;
    }
    this._frameId = this._elem.globalData.frameId;
    this.iterateDynamicProperties();
};

TextAnimatorProperty.prototype.mHelper = new Matrix();
TextAnimatorProperty.prototype.defaultPropsArray = [];
extendPrototype([DynamicPropertyContainer], TextAnimatorProperty);
function TextAnimatorDataProperty(elem, animatorProps, container) {
	var defaultData = {propType:false};
	var getProp = PropertyFactory.getProp;
	var textAnimator_animatables = animatorProps.a;
	this.a = {
		r: textAnimator_animatables.r ? getProp(elem, textAnimator_animatables.r, 0, degToRads, container) : defaultData,
		rx: textAnimator_animatables.rx ? getProp(elem, textAnimator_animatables.rx, 0, degToRads, container) : defaultData,
		ry: textAnimator_animatables.ry ? getProp(elem, textAnimator_animatables.ry, 0, degToRads, container) : defaultData,
		sk: textAnimator_animatables.sk ? getProp(elem, textAnimator_animatables.sk, 0, degToRads, container) : defaultData,
		sa: textAnimator_animatables.sa ? getProp(elem, textAnimator_animatables.sa, 0, degToRads, container) : defaultData,
		s: textAnimator_animatables.s ? getProp(elem, textAnimator_animatables.s, 1, 0.01, container) : defaultData,
		a: textAnimator_animatables.a ? getProp(elem, textAnimator_animatables.a, 1, 0, container) : defaultData,
		o: textAnimator_animatables.o ? getProp(elem, textAnimator_animatables.o, 0, 0.01, container) : defaultData,
		p: textAnimator_animatables.p ? getProp(elem,textAnimator_animatables.p, 1, 0, container) : defaultData,
		sw: textAnimator_animatables.sw ? getProp(elem, textAnimator_animatables.sw, 0, 0, container) : defaultData,
		sc: textAnimator_animatables.sc ? getProp(elem, textAnimator_animatables.sc, 1, 0, container) : defaultData,
		fc: textAnimator_animatables.fc ? getProp(elem, textAnimator_animatables.fc, 1, 0, container) : defaultData,
		fh: textAnimator_animatables.fh ? getProp(elem, textAnimator_animatables.fh, 0, 0, container) : defaultData,
		fs: textAnimator_animatables.fs ? getProp(elem, textAnimator_animatables.fs, 0, 0.01, container) : defaultData,
		fb: textAnimator_animatables.fb ? getProp(elem, textAnimator_animatables.fb, 0, 0.01, container) : defaultData,
		t: textAnimator_animatables.t ? getProp(elem, textAnimator_animatables.t, 0, 0, container) : defaultData
	};

	this.s = TextSelectorProp.getTextSelectorProp(elem,animatorProps.s, container);
    this.s.t = animatorProps.s.t;
}
function LetterProps(o, sw, sc, fc, m, p){
    this.o = o;
    this.sw = sw;
    this.sc = sc;
    this.fc = fc;
    this.m = m;
    this.p = p;
    this._mdf = {
    	o: true,
    	sw: !!sw,
    	sc: !!sc,
    	fc: !!fc,
    	m: true,
    	p: true
    };
}

LetterProps.prototype.update = function(o, sw, sc, fc, m, p) {
	this._mdf.o = false;
	this._mdf.sw = false;
	this._mdf.sc = false;
	this._mdf.fc = false;
	this._mdf.m = false;
	this._mdf.p = false;
	var updated = false;

	if(this.o !== o) {
		this.o = o;
		this._mdf.o = true;
		updated = true;
	}
	if(this.sw !== sw) {
		this.sw = sw;
		this._mdf.sw = true;
		updated = true;
	}
	if(this.sc !== sc) {
		this.sc = sc;
		this._mdf.sc = true;
		updated = true;
	}
	if(this.fc !== fc) {
		this.fc = fc;
		this._mdf.fc = true;
		updated = true;
	}
	if(this.m !== m) {
		this.m = m;
		this._mdf.m = true;
		updated = true;
	}
	if(p.length && (this.p[0] !== p[0] || this.p[1] !== p[1] || this.p[4] !== p[4] || this.p[5] !== p[5] || this.p[12] !== p[12] || this.p[13] !== p[13])) {
		this.p = p;
		this._mdf.p = true;
		updated = true;
	}
	return updated;
};
function TextProperty(elem, data){
	this._frameId = initialDefaultFrame;
	this.pv = '';
	this.v = '';
	this.kf = false;
	this._isFirstFrame = true;
	this._mdf = false;
    this.data = data;
	this.elem = elem;
    this.comp = this.elem.comp;
	this.keysIndex = 0;
    this.canResize = false;
    this.minimumFontSize = 1;
    this.effectsSequence = [];
	this.currentData = {
		ascent: 0,
        boxWidth: this.defaultBoxWidth,
        f: '',
        fStyle: '',
        fWeight: '',
        fc: '',
        j: '',
        justifyOffset: '',
        l: [],
        lh: 0,
        lineWidths: [],
        ls: '',
        of: '',
        s: '',
        sc: '',
        sw: 0,
        t: 0,
        tr: 0,
        sz:0,
        ps:null,
        fillColorAnim: false,
        strokeColorAnim: false,
        strokeWidthAnim: false,
        yOffset: 0,
        finalSize:0,
        finalText:[],
        finalLineHeight: 0,
        __complete: false

	};
    this.copyData(this.currentData, this.data.d.k[0].s);

    if(!this.searchProperty()) {
        this.completeTextData(this.currentData);
    }
}

TextProperty.prototype.defaultBoxWidth = [0,0];

TextProperty.prototype.copyData = function(obj, data) {
    for(var s in data) {
        if(data.hasOwnProperty(s)) {
            obj[s] = data[s];
        }
    }
    return obj;
}

TextProperty.prototype.setCurrentData = function(data){
    if(!data.__complete) {
        this.completeTextData(data);
    }
    this.currentData = data;
    this.currentData.boxWidth = this.currentData.boxWidth || this.defaultBoxWidth;
    this._mdf = true;
};

TextProperty.prototype.searchProperty = function() {
    return this.searchKeyframes();
};

TextProperty.prototype.searchKeyframes = function() {
    this.kf = this.data.d.k.length > 1;
    if(this.kf) {
        this.addEffect(this.getKeyframeValue.bind(this));
    }
    return this.kf;
}

TextProperty.prototype.addEffect = function(effectFunction) {
	this.effectsSequence.push(effectFunction);
    this.elem.addDynamicProperty(this);
};

TextProperty.prototype.getValue = function(_finalValue) {
    if((this.elem.globalData.frameId === this.frameId || !this.effectsSequence.length) && !_finalValue) {
        return;
    }
    this.currentData.t = this.data.d.k[this.keysIndex].s.t;
    var currentValue = this.currentData;
    var currentIndex = this.keysIndex;
    if(this.lock) {
        this.setCurrentData(this.currentData);
        return;
    }
    this.lock = true;
    this._mdf = false;
    var multipliedValue;
    var i, len = this.effectsSequence.length;
    var finalValue = _finalValue || this.data.d.k[this.keysIndex].s;
    for(i = 0; i < len; i += 1) {
        //Checking if index changed to prevent creating a new object every time the expression updates.
        if(currentIndex !== this.keysIndex) {
            finalValue = this.effectsSequence[i](finalValue, finalValue.t);
        } else {
            finalValue = this.effectsSequence[i](this.currentData, finalValue.t);
        }
    }
    if(currentValue !== finalValue) {
        this.setCurrentData(finalValue);
    }
    this.pv = this.v = this.currentData;
    this.lock = false;
    this.frameId = this.elem.globalData.frameId;
}

TextProperty.prototype.getKeyframeValue = function() {
    var textKeys = this.data.d.k, textDocumentData;
    var frameNum = this.elem.comp.renderedFrame;
    var i = 0, len = textKeys.length;
    while(i <= len - 1) {
        textDocumentData = textKeys[i].s;
        if(i === len - 1 || textKeys[i+1].t > frameNum){
            break;
        }
        i += 1;
    }
    if(this.keysIndex !== i) {
        this.keysIndex = i;
    }
    return this.data.d.k[this.keysIndex].s;
};

TextProperty.prototype.buildFinalText = function(text) {
    var combinedCharacters = FontManager.getCombinedCharacterCodes();
    var charactersArray = [];
    var i = 0, len = text.length;
    while (i < len) {
        if (combinedCharacters.indexOf(text.charCodeAt(i)) !== -1) {
            charactersArray[charactersArray.length - 1] += text.charAt(i);
        } else {
            charactersArray.push(text.charAt(i));
        }
        i += 1;
    }
    return charactersArray;
}

TextProperty.prototype.completeTextData = function(documentData) {
    documentData.__complete = true;
    var fontManager = this.elem.globalData.fontManager;
    var data = this.data;
    var letters = [];
    var i, len;
    var newLineFlag, index = 0, val;
    var anchorGrouping = data.m.g;
    var currentSize = 0, currentPos = 0, currentLine = 0, lineWidths = [];
    var lineWidth = 0;
    var maxLineWidth = 0;
    var j, jLen;
    var fontData = fontManager.getFontByName(documentData.f);
    var charData, cLength = 0;
    var styles = fontData.fStyle ? fontData.fStyle.split(' ') : [];

    var fWeight = 'normal', fStyle = 'normal';
    len = styles.length;
    var styleName;
    for(i=0;i<len;i+=1){
        styleName = styles[i].toLowerCase();
        switch(styleName) {
            case 'italic':
            fStyle = 'italic';
            break;
            case 'bold':
            fWeight = '700';
            break;
            case 'black':
            fWeight = '900';
            break;
            case 'medium':
            fWeight = '500';
            break;
            case 'regular':
            case 'normal':
            fWeight = '400';
            break;
            case 'light':
            case 'thin':
            fWeight = '200';
            break;
        }
    }
    documentData.fWeight = fontData.fWeight || fWeight;
    documentData.fStyle = fStyle;
    len = documentData.t.length;
    documentData.finalSize = documentData.s;
    documentData.finalText = this.buildFinalText(documentData.t);
    documentData.finalLineHeight = documentData.lh;
    var trackingOffset = documentData.tr/1000*documentData.finalSize;
    var charCode;
    if(documentData.sz){
        var flag = true;
        var boxWidth = documentData.sz[0];
        var boxHeight = documentData.sz[1];
        var currentHeight, finalText;
        while(flag) {
            finalText = this.buildFinalText(documentData.t);
            currentHeight = 0;
            lineWidth = 0;
            len = finalText.length;
            trackingOffset = documentData.tr/1000*documentData.finalSize;
            var lastSpaceIndex = -1;
            for(i=0;i<len;i+=1){
                charCode = finalText[i].charCodeAt(0);
                newLineFlag = false;
                if(finalText[i] === ' '){
                    lastSpaceIndex = i;
                }else if(charCode === 13 || charCode === 3){
                    lineWidth = 0;
                    newLineFlag = true;
                    currentHeight += documentData.finalLineHeight || documentData.finalSize*1.2;
                }
                if(fontManager.chars){
                    charData = fontManager.getCharData(finalText[i], fontData.fStyle, fontData.fFamily);
                    cLength = newLineFlag ? 0 : charData.w*documentData.finalSize/100;
                }else{
                    //tCanvasHelper.font = documentData.s + 'px '+ fontData.fFamily;
                    cLength = fontManager.measureText(finalText[i], documentData.f, documentData.finalSize);
                }
                if(lineWidth + cLength > boxWidth && finalText[i] !== ' '){
                    if(lastSpaceIndex === -1){
                        len += 1;
                    } else {
                        i = lastSpaceIndex;
                    }
                    currentHeight += documentData.finalLineHeight || documentData.finalSize*1.2;
                    finalText.splice(i, lastSpaceIndex === i ? 1 : 0,"\r");
                    //finalText = finalText.substr(0,i) + "\r" + finalText.substr(i === lastSpaceIndex ? i + 1 : i);
                    lastSpaceIndex = -1;
                    lineWidth = 0;
                }else {
                    lineWidth += cLength;
                    lineWidth += trackingOffset;
                }
            }
            currentHeight += fontData.ascent*documentData.finalSize/100;
            if(this.canResize && documentData.finalSize > this.minimumFontSize && boxHeight < currentHeight) {
                documentData.finalSize -= 1;
                documentData.finalLineHeight = documentData.finalSize * documentData.lh / documentData.s;
            } else {
                documentData.finalText = finalText;
                len = documentData.finalText.length;
                flag = false;
            }
        }

    }
    lineWidth = - trackingOffset;
    cLength = 0;
    var uncollapsedSpaces = 0;
    var currentChar;
    for (i = 0;i < len ;i += 1) {
        newLineFlag = false;
        currentChar = documentData.finalText[i];
        charCode = currentChar.charCodeAt(0);
        if (currentChar === ' '){
            val = '\u00A0';
        } else if (charCode === 13 || charCode === 3) {
            uncollapsedSpaces = 0;
            lineWidths.push(lineWidth);
            maxLineWidth = lineWidth > maxLineWidth ? lineWidth : maxLineWidth;
            lineWidth = - 2 * trackingOffset;
            val = '';
            newLineFlag = true;
            currentLine += 1;
        }else{
            val = documentData.finalText[i];
        }
        if(fontManager.chars){
            charData = fontManager.getCharData(currentChar, fontData.fStyle, fontManager.getFontByName(documentData.f).fFamily);
            cLength = newLineFlag ? 0 : charData.w*documentData.finalSize/100;
        }else{
            //var charWidth = fontManager.measureText(val, documentData.f, documentData.finalSize);
            //tCanvasHelper.font = documentData.finalSize + 'px '+ fontManager.getFontByName(documentData.f).fFamily;
            cLength = fontManager.measureText(val, documentData.f, documentData.finalSize);
        }

        //
        if(currentChar === ' '){
            uncollapsedSpaces += cLength + trackingOffset;
        } else {
            lineWidth += cLength + trackingOffset + uncollapsedSpaces;
            uncollapsedSpaces = 0;
        }
        letters.push({l:cLength,an:cLength,add:currentSize,n:newLineFlag, anIndexes:[], val: val, line: currentLine, animatorJustifyOffset: 0});
        if(anchorGrouping == 2){
            currentSize += cLength;
            if(val === '' || val === '\u00A0' || i === len - 1){
                if(val === '' || val === '\u00A0'){
                    currentSize -= cLength;
                }
                while(currentPos<=i){
                    letters[currentPos].an = currentSize;
                    letters[currentPos].ind = index;
                    letters[currentPos].extra = cLength;
                    currentPos += 1;
                }
                index += 1;
                currentSize = 0;
            }
        }else if(anchorGrouping == 3){
            currentSize += cLength;
            if(val === '' || i === len - 1){
                if(val === ''){
                    currentSize -= cLength;
                }
                while(currentPos<=i){
                    letters[currentPos].an = currentSize;
                    letters[currentPos].ind = index;
                    letters[currentPos].extra = cLength;
                    currentPos += 1;
                }
                currentSize = 0;
                index += 1;
            }
        }else{
            letters[index].ind = index;
            letters[index].extra = 0;
            index += 1;
        }
    }
    documentData.l = letters;
    maxLineWidth = lineWidth > maxLineWidth ? lineWidth : maxLineWidth;
    lineWidths.push(lineWidth);
    if(documentData.sz){
        documentData.boxWidth = documentData.sz[0];
        documentData.justifyOffset = 0;
    }else{
        documentData.boxWidth = maxLineWidth;
        switch(documentData.j){
            case 1:
                documentData.justifyOffset = - documentData.boxWidth;
                break;
            case 2:
                documentData.justifyOffset = - documentData.boxWidth/2;
                break;
            default:
                documentData.justifyOffset = 0;
        }
    }
    documentData.lineWidths = lineWidths;

    var animators = data.a, animatorData, letterData;
    jLen = animators.length;
    var based, ind, indexes = [];
    for(j=0;j<jLen;j+=1){
        animatorData = animators[j];
        if(animatorData.a.sc){
            documentData.strokeColorAnim = true;
        }
        if(animatorData.a.sw){
            documentData.strokeWidthAnim = true;
        }
        if(animatorData.a.fc || animatorData.a.fh || animatorData.a.fs || animatorData.a.fb){
            documentData.fillColorAnim = true;
        }
        ind = 0;
        based = animatorData.s.b;
        for(i=0;i<len;i+=1){
            letterData = letters[i];
            letterData.anIndexes[j] = ind;
            if((based == 1 && letterData.val !== '') || (based == 2 && letterData.val !== '' && letterData.val !== '\u00A0') || (based == 3 && (letterData.n || letterData.val == '\u00A0' || i == len - 1)) || (based == 4 && (letterData.n || i == len - 1))){
                if(animatorData.s.rn === 1){
                    indexes.push(ind);
                }
                ind += 1;
            }
        }
        data.a[j].s.totalChars = ind;
        var currentInd = -1, newInd;
        if(animatorData.s.rn === 1){
            for(i = 0; i < len; i += 1){
                letterData = letters[i];
                if(currentInd != letterData.anIndexes[j]){
                    currentInd = letterData.anIndexes[j];
                    newInd = indexes.splice(Math.floor(Math.random()*indexes.length),1)[0];
                }
                letterData.anIndexes[j] = newInd;
            }
        }
    }
    documentData.yOffset = documentData.finalLineHeight || documentData.finalSize*1.2;
    documentData.ls = documentData.ls || 0;
    documentData.ascent = fontData.ascent*documentData.finalSize/100;
};

TextProperty.prototype.updateDocumentData = function(newData, index) {
	index = index === undefined ? this.keysIndex : index;
    var dData = this.copyData({}, this.data.d.k[index].s);
    dData = this.copyData(dData, newData);
    this.data.d.k[index].s = dData;
    this.recalculate(index);
    this.elem.addDynamicProperty(this);
};

TextProperty.prototype.recalculate = function(index) {
    var dData = this.data.d.k[index].s;
    dData.__complete = false;
    this.keysIndex = 0;
    this._isFirstFrame = true;
    this.getValue(dData);
}

TextProperty.prototype.canResizeFont = function(_canResize) {
    this.canResize = _canResize;
    this.recalculate(this.keysIndex);
    this.elem.addDynamicProperty(this);
};

TextProperty.prototype.setMinimumFontSize = function(_fontValue) {
    this.minimumFontSize = Math.floor(_fontValue) || 1;
    this.recalculate(this.keysIndex);
    this.elem.addDynamicProperty(this);
};

var TextSelectorProp = (function(){
    var max = Math.max;
    var min = Math.min;
    var floor = Math.floor;

    function TextSelectorProp(elem,data){
        this._currentTextLength = -1;
        this.k = false;
        this.data = data;
        this.elem = elem;
        this.comp = elem.comp;
        this.finalS = 0;
        this.finalE = 0;
        this.initDynamicPropertyContainer(elem);
        this.s = PropertyFactory.getProp(elem,data.s || {k:0},0,0,this);
        if('e' in data){
            this.e = PropertyFactory.getProp(elem,data.e,0,0,this);
        }else{
            this.e = {v:100};
        }
        this.o = PropertyFactory.getProp(elem,data.o || {k:0},0,0,this);
        this.xe = PropertyFactory.getProp(elem,data.xe || {k:0},0,0,this);
        this.ne = PropertyFactory.getProp(elem,data.ne || {k:0},0,0,this);
        this.a = PropertyFactory.getProp(elem,data.a,0,0.01,this);
        if(!this.dynamicProperties.length){
            this.getValue();
        }
    }

    TextSelectorProp.prototype = {
        getMult: function(ind) {
            if(this._currentTextLength !== this.elem.textProperty.currentData.l.length) {
                this.getValue();
            }
            //var easer = bez.getEasingCurve(this.ne.v/100,0,1-this.xe.v/100,1);
            var easer = BezierFactory.getBezierEasing(this.ne.v/100,0,1-this.xe.v/100,1).get;
            var mult = 0;
            var s = this.finalS;
            var e = this.finalE;
            var type = this.data.sh;
            if(type == 2){
                if(e === s){
                    mult = ind >= e ? 1 : 0;
                }else{
                    mult = max(0,min(0.5/(e-s) + (ind-s)/(e-s),1));
                }
                mult = easer(mult);
            }else if(type == 3){
                if(e === s){
                    mult = ind >= e ? 0 : 1;
                }else{
                    mult = 1 - max(0,min(0.5/(e-s) + (ind-s)/(e-s),1));
                }

                mult = easer(mult);
            }else if(type == 4){
                if(e === s){
                    mult = 0;
                }else{
                    mult = max(0,min(0.5/(e-s) + (ind-s)/(e-s),1));
                    if(mult<0.5){
                        mult *= 2;
                    }else{
                        mult = 1 - 2*(mult-0.5);
                    }
                }
                mult = easer(mult);
            }else if(type == 5){
                if(e === s){
                    mult = 0;
                }else{
                    var tot = e - s;
                    /*ind += 0.5;
                    mult = -4/(tot*tot)*(ind*ind)+(4/tot)*ind;*/
                    ind = min(max(0,ind+0.5-s),e-s);
                    var x = -tot/2+ind;
                    var a = tot/2;
                    mult = Math.sqrt(1 - (x*x)/(a*a));
                }
                mult = easer(mult);
            }else if(type == 6){
                if(e === s){
                    mult = 0;
                }else{
                    ind = min(max(0,ind+0.5-s),e-s);
                    mult = (1+(Math.cos((Math.PI+Math.PI*2*(ind)/(e-s)))))/2;
                    /*
                     ind = Math.min(Math.max(s,ind),e-1);
                     mult = (1+(Math.cos((Math.PI+Math.PI*2*(ind-s)/(e-1-s)))))/2;
                     mult = Math.max(mult,(1/(e-1-s))/(e-1-s));*/
                }
                mult = easer(mult);
            }else {
                if(ind >= floor(s)){
                    if(ind-s < 0){
                        mult = 1 - (s - ind);
                    }else{
                        mult = max(0,min(e-ind,1));
                    }
                }
                mult = easer(mult);
            }
            return mult*this.a.v;
        },
        getValue: function(newCharsFlag) {
            this.iterateDynamicProperties();
            this._mdf = newCharsFlag || this._mdf;
            this._currentTextLength = this.elem.textProperty.currentData.l.length || 0;
            if(newCharsFlag && this.data.r === 2) {
                this.e.v = this._currentTextLength;
            }
            var divisor = this.data.r === 2 ? 1 : 100 / this.data.totalChars;
            var o = this.o.v/divisor;
            var s = this.s.v/divisor + o;
            var e = (this.e.v/divisor) + o;
            if(s>e){
                var _s = s;
                s = e;
                e = _s;
            }
            this.finalS = s;
            this.finalE = e;
        }
    }
    extendPrototype([DynamicPropertyContainer], TextSelectorProp);

    function getTextSelectorProp(elem, data,arr) {
        return new TextSelectorProp(elem, data, arr);
    }

    return {
        getTextSelectorProp: getTextSelectorProp
    };
}());


var pool_factory = (function() {
	return function(initialLength, _create, _release, _clone) {

		var _length = 0;
		var _maxLength = initialLength;
		var pool = createSizedArray(_maxLength);

		var ob = {
			newElement: newElement,
			release: release
		};

		function newElement(){
			var element;
			if(_length){
				_length -= 1;
				element = pool[_length];
			} else {
				element = _create();
			}
			return element;
		}

		function release(element) {
			if(_length === _maxLength) {
				pool = pooling.double(pool);
				_maxLength = _maxLength*2;
			}
			if (_release) {
				_release(element);
			}
			pool[_length] = element;
			_length += 1;
		}

		function clone() {
			var clonedElement = newElement();
			return _clone(clonedElement);
		}

		return ob;
	};
}());

var pooling = (function(){

	function double(arr){
		return arr.concat(createSizedArray(arr.length));
	}

	return {
		double: double
	};
}());
var point_pool = (function(){

	function create() {
		return createTypedArray('float32', 2);
	}
	return pool_factory(8, create);
}());
var shape_pool = (function(){

	function create() {
		return new ShapePath();
	}

	function release(shapePath) {
		var len = shapePath._length, i;
		for(i = 0; i < len; i += 1) {
			point_pool.release(shapePath.v[i]);
			point_pool.release(shapePath.i[i]);
			point_pool.release(shapePath.o[i]);
			shapePath.v[i] = null;
			shapePath.i[i] = null;
			shapePath.o[i] = null;
		}
		shapePath._length = 0;
		shapePath.c = false;
	}

	function clone(shape) {
		var cloned = factory.newElement();
		var i, len = shape._length === undefined ? shape.v.length : shape._length;
		cloned.setLength(len);
		cloned.c = shape.c;
		var pt;

		for(i = 0; i < len; i += 1) {
			cloned.setTripleAt(shape.v[i][0],shape.v[i][1],shape.o[i][0],shape.o[i][1],shape.i[i][0],shape.i[i][1], i);
		}
		return cloned;
	}

	var factory = pool_factory(4, create, release);
	factory.clone = clone;

	return factory;
}());
var shapeCollection_pool = (function(){
	var ob = {
		newShapeCollection: newShapeCollection,
		release: release
	};

	var _length = 0;
	var _maxLength = 4;
	var pool = createSizedArray(_maxLength);

	function newShapeCollection(){
		var shapeCollection;
		if(_length){
			_length -= 1;
			shapeCollection = pool[_length];
		} else {
			shapeCollection = new ShapeCollection();
		}
		return shapeCollection;
	}

	function release(shapeCollection) {
		var i, len = shapeCollection._length;
		for(i = 0; i < len; i += 1) {
			shape_pool.release(shapeCollection.shapes[i]);
		}
		shapeCollection._length = 0;

		if(_length === _maxLength) {
			pool = pooling.double(pool);
			_maxLength = _maxLength*2;
		}
		pool[_length] = shapeCollection;
		_length += 1;
	}

	return ob;
}());
var segments_length_pool = (function(){

	function create() {
		return {
			lengths: [],
			totalLength: 0
		};
	}

	function release(element) {
		var i, len = element.lengths.length;
		for(i=0;i<len;i+=1) {
			bezier_length_pool.release(element.lengths[i]);
		}
		element.lengths.length = 0;
	}

	return pool_factory(8, create, release);
}());
var bezier_length_pool = (function(){

	function create() {
		return {
            addedLength: 0,
            percents: createTypedArray('float32', defaultCurveSegments),
            lengths: createTypedArray('float32', defaultCurveSegments),
        };
	}
	return pool_factory(8, create);
}());
function BaseRenderer(){}
BaseRenderer.prototype.checkLayers = function(num){
    var i, len = this.layers.length, data;
    this.completeLayers = true;
    for (i = len - 1; i >= 0; i--) {
        if (!this.elements[i]) {
            data = this.layers[i];
            if(data.ip - data.st <= (num - this.layers[i].st) && data.op - data.st > (num - this.layers[i].st))
            {
                this.buildItem(i);
            }
        }
        this.completeLayers = this.elements[i] ? this.completeLayers:false;
    }
    this.checkPendingElements();
};

BaseRenderer.prototype.createItem = function(layer){
    switch(layer.ty){
        case 2:
            return this.createImage(layer);
        case 0:
            return this.createComp(layer);
        case 1:
            return this.createSolid(layer);
        case 3:
            return this.createNull(layer);
        case 4:
            return this.createShape(layer);
        case 5:
            return this.createText(layer);
        case 13:
            return this.createCamera(layer);
    }
    return this.createNull(layer);
};

BaseRenderer.prototype.createCamera = function(){
    throw new Error('You\'re using a 3d camera. Try the html renderer.');
};

BaseRenderer.prototype.buildAllItems = function(){
    var i, len = this.layers.length;
    for(i=0;i<len;i+=1){
        this.buildItem(i);
    }
    this.checkPendingElements();
};

BaseRenderer.prototype.includeLayers = function(newLayers){
    this.completeLayers = false;
    var i, len = newLayers.length;
    var j, jLen = this.layers.length;
    for(i=0;i<len;i+=1){
        j = 0;
        while(j<jLen){
            if(this.layers[j].id == newLayers[i].id){
                this.layers[j] = newLayers[i];
                break;
            }
            j += 1;
        }
    }
};

BaseRenderer.prototype.setProjectInterface = function(pInterface){
    this.globalData.projectInterface = pInterface;
};

BaseRenderer.prototype.initItems = function(){
    if(!this.globalData.progressiveLoad){
        this.buildAllItems();
    }
};
BaseRenderer.prototype.buildElementParenting = function(element, parentName, hierarchy) {
    var elements = this.elements;
    var layers = this.layers;
    var i=0, len = layers.length;
    while (i < len) {
        if (layers[i].ind == parentName) {
            if (!elements[i] || elements[i] === true) {
                this.buildItem(i);
                this.addPendingElement(element);
            } else {
                hierarchy.push(elements[i]);
                elements[i].setAsParent();
                if(layers[i].parent !== undefined) {
                    this.buildElementParenting(element, layers[i].parent, hierarchy);
                } else {
                    element.setHierarchy(hierarchy);
                }
            }
        }
        i += 1;
    }
};

BaseRenderer.prototype.addPendingElement = function(element){
    this.pendingElements.push(element);
};

BaseRenderer.prototype.searchExtraCompositions = function(assets){
    var i, len = assets.length;
    for(i=0;i<len;i+=1){
        if(assets[i].xt){
            var comp = this.createComp(assets[i]);
            comp.initExpressions();
            this.globalData.projectInterface.registerComposition(comp);
        }
    }
};

BaseRenderer.prototype.setupGlobalData = function(animData, fontsContainer) {
    this.globalData.fontManager = new FontManager();
    this.globalData.fontManager.addChars(animData.chars);
    this.globalData.fontManager.addFonts(animData.fonts, fontsContainer);
    this.globalData.getAssetData = this.animationItem.getAssetData.bind(this.animationItem);
    this.globalData.getAssetsPath = this.animationItem.getAssetsPath.bind(this.animationItem);
    this.globalData.imageLoader = this.animationItem.imagePreloader;
    this.globalData.frameId = 0;
    this.globalData.frameRate = animData.fr;
    this.globalData.nm = animData.nm;
    this.globalData.compSize = {
        w: animData.w,
        h: animData.h
    }
}
function SVGRenderer(animationItem, config){
    this.animationItem = animationItem;
    this.layers = null;
    this.renderedFrame = -1;
    this.svgElement = createNS('svg');
    var ariaLabel = '';
    if (config && config.title) {
        var titleElement = createNS('title');
        var titleId = createElementID();
        titleElement.setAttribute('id', titleId);
        titleElement.textContent = config.title;
        this.svgElement.appendChild(titleElement);
        ariaLabel += titleId;
    }
    if (config && config.description) {
        var descElement = createNS('desc');
        var descId = createElementID();
        descElement.setAttribute('id', descId);
        descElement.textContent = config.description;
        this.svgElement.appendChild(descElement);
        ariaLabel += ' ' + descId;
    }
    if (ariaLabel) {
        this.svgElement.setAttribute('aria-labelledby', ariaLabel)
    }
    var defs = createNS( 'defs');
    this.svgElement.appendChild(defs);
    var maskElement = createNS('g');
    this.svgElement.appendChild(maskElement);
    this.layerElement = maskElement;
    this.renderConfig = {
        preserveAspectRatio: (config && config.preserveAspectRatio) || 'xMidYMid meet',
        imagePreserveAspectRatio: (config && config.imagePreserveAspectRatio) || 'xMidYMid slice',
        progressiveLoad: (config && config.progressiveLoad) || false,
        hideOnTransparent: (config && config.hideOnTransparent === false) ? false : true,
        viewBoxOnly: (config && config.viewBoxOnly) || false,
        viewBoxSize: (config && config.viewBoxSize) || false,
        className: (config && config.className) || ''
    };

    this.globalData = {
        _mdf: false,
        frameNum: -1,
        defs: defs,
        renderConfig: this.renderConfig
    };
    this.elements = [];
    this.pendingElements = [];
    this.destroyed = false;
    this.rendererType = 'svg';

}

extendPrototype([BaseRenderer],SVGRenderer);

SVGRenderer.prototype.createNull = function (data) {
    return new NullElement(data,this.globalData,this);
};

SVGRenderer.prototype.createShape = function (data) {
    return new SVGShapeElement(data,this.globalData,this);
};

SVGRenderer.prototype.createText = function (data) {
    return new SVGTextElement(data,this.globalData,this);

};

SVGRenderer.prototype.createImage = function (data) {
    return new IImageElement(data,this.globalData,this);
};

SVGRenderer.prototype.createComp = function (data) {
    return new SVGCompElement(data,this.globalData,this);

};

SVGRenderer.prototype.createSolid = function (data) {
    return new ISolidElement(data,this.globalData,this);
};

SVGRenderer.prototype.configAnimation = function(animData){
    this.svgElement.setAttribute('xmlns','http://www.w3.org/2000/svg');
    if(this.renderConfig.viewBoxSize) {
        this.svgElement.setAttribute('viewBox',this.renderConfig.viewBoxSize);
    } else {
        this.svgElement.setAttribute('viewBox','0 0 '+animData.w+' '+animData.h);
    }

    if(!this.renderConfig.viewBoxOnly) {
        this.svgElement.setAttribute('width',animData.w);
        this.svgElement.setAttribute('height',animData.h);
        this.svgElement.style.width = '100%';
        this.svgElement.style.height = '100%';
        this.svgElement.style.transform = 'translate3d(0,0,0)';
    }
    if(this.renderConfig.className) {
        this.svgElement.setAttribute('class', this.renderConfig.className);
    }
    this.svgElement.setAttribute('preserveAspectRatio',this.renderConfig.preserveAspectRatio);
    //this.layerElement.style.transform = 'translate3d(0,0,0)';
    //this.layerElement.style.transformOrigin = this.layerElement.style.mozTransformOrigin = this.layerElement.style.webkitTransformOrigin = this.layerElement.style['-webkit-transform'] = "0px 0px 0px";
    this.animationItem.wrapper.appendChild(this.svgElement);
    //Mask animation
    var defs = this.globalData.defs;

    this.setupGlobalData(animData, defs);
    this.globalData.progressiveLoad = this.renderConfig.progressiveLoad;
    this.data = animData;

    var maskElement = createNS( 'clipPath');
    var rect = createNS('rect');
    rect.setAttribute('width',animData.w);
    rect.setAttribute('height',animData.h);
    rect.setAttribute('x',0);
    rect.setAttribute('y',0);
    var maskId = createElementID();
    maskElement.setAttribute('id', maskId);
    maskElement.appendChild(rect);
    this.layerElement.setAttribute("clip-path", "url(" + locationHref + "#"+maskId+")");

    defs.appendChild(maskElement);
    this.layers = animData.layers;
    this.elements = createSizedArray(animData.layers.length);
};


SVGRenderer.prototype.destroy = function () {
    this.animationItem.wrapper.innerHTML = '';
    this.layerElement = null;
    this.globalData.defs = null;
    var i, len = this.layers ? this.layers.length : 0;
    for (i = 0; i < len; i++) {
        if(this.elements[i]){
            this.elements[i].destroy();
        }
    }
    this.elements.length = 0;
    this.destroyed = true;
    this.animationItem = null;
};

SVGRenderer.prototype.updateContainerSize = function () {
};

SVGRenderer.prototype.buildItem  = function(pos){
    var elements = this.elements;
    if(elements[pos] || this.layers[pos].ty == 99){
        return;
    }
    elements[pos] = true;
    var element = this.createItem(this.layers[pos]);

    elements[pos] = element;
    if(expressionsPlugin){
        if(this.layers[pos].ty === 0){
            this.globalData.projectInterface.registerComposition(element);
        }
        element.initExpressions();
    }
    this.appendElementInPos(element,pos);
    if(this.layers[pos].tt){
        if(!this.elements[pos - 1] || this.elements[pos - 1] === true){
            this.buildItem(pos - 1);
            this.addPendingElement(element);
        } else {
            element.setMatte(elements[pos - 1].layerId);
        }
    }
};

SVGRenderer.prototype.checkPendingElements  = function(){
    while(this.pendingElements.length){
        var element = this.pendingElements.pop();
        element.checkParenting();
        if(element.data.tt){
            var i = 0, len = this.elements.length;
            while(i<len){
                if(this.elements[i] === element){
                    element.setMatte(this.elements[i - 1].layerId);
                    break;
                }
                i += 1;
            }
        }
    }
};

SVGRenderer.prototype.renderFrame = function(num){
    if(this.renderedFrame === num || this.destroyed){
        return;
    }
    if(num === null){
        num = this.renderedFrame;
    }else{
        this.renderedFrame = num;
    }
    // console.log('-------');
    // console.log('FRAME ',num);
    this.globalData.frameNum = num;
    this.globalData.frameId += 1;
    this.globalData.projectInterface.currentFrame = num;
    this.globalData._mdf = false;
    var i, len = this.layers.length;
    if(!this.completeLayers){
        this.checkLayers(num);
    }
    for (i = len - 1; i >= 0; i--) {
        if(this.completeLayers || this.elements[i]){
            this.elements[i].prepareFrame(num - this.layers[i].st);
        }
    }
    if(this.globalData._mdf) {
        for (i = 0; i < len; i += 1) {
            if(this.completeLayers || this.elements[i]){
                this.elements[i].renderFrame();
            }
        }
    }
};

SVGRenderer.prototype.appendElementInPos = function(element, pos){
    var newElement = element.getBaseElement();
    if(!newElement){
        return;
    }
    var i = 0;
    var nextElement;
    while(i<pos){
        if(this.elements[i] && this.elements[i]!== true && this.elements[i].getBaseElement()){
            nextElement = this.elements[i].getBaseElement();
        }
        i += 1;
    }
    if(nextElement){
        this.layerElement.insertBefore(newElement, nextElement);
    } else {
        this.layerElement.appendChild(newElement);
    }
};

SVGRenderer.prototype.hide = function(){
    this.layerElement.style.display = 'none';
};

SVGRenderer.prototype.show = function(){
    this.layerElement.style.display = 'block';
};

function CanvasRenderer(animationItem, config){
    this.animationItem = animationItem;
    this.renderConfig = {
        clearCanvas: (config && config.clearCanvas !== undefined) ? config.clearCanvas : true,
        context: (config && config.context) || null,
        progressiveLoad: (config && config.progressiveLoad) || false,
        preserveAspectRatio: (config && config.preserveAspectRatio) || 'xMidYMid meet',
        imagePreserveAspectRatio: (config && config.imagePreserveAspectRatio) || 'xMidYMid slice',
        className: (config && config.className) || ''
    };
    this.renderConfig.dpr = (config && config.dpr) || 1;
    if (this.animationItem.wrapper) {
        this.renderConfig.dpr = (config && config.dpr) || window.devicePixelRatio || 1;
    }
    this.renderedFrame = -1;
    this.globalData = {
        frameNum: -1,
        _mdf: false,
        renderConfig: this.renderConfig,
        currentGlobalAlpha: -1
    };
    this.contextData = new CVContextData();
    this.elements = [];
    this.pendingElements = [];
    this.transformMat = new Matrix();
    this.completeLayers = false;
    this.rendererType = 'canvas';
}
extendPrototype([BaseRenderer],CanvasRenderer);

CanvasRenderer.prototype.createShape = function (data) {
    return new CVShapeElement(data, this.globalData, this);
};

CanvasRenderer.prototype.createText = function (data) {
    return new CVTextElement(data, this.globalData, this);
};

CanvasRenderer.prototype.createImage = function (data) {
    return new CVImageElement(data, this.globalData, this);
};

CanvasRenderer.prototype.createComp = function (data) {
    return new CVCompElement(data, this.globalData, this);
};

CanvasRenderer.prototype.createSolid = function (data) {
    return new CVSolidElement(data, this.globalData, this);
};

CanvasRenderer.prototype.createNull = SVGRenderer.prototype.createNull;

CanvasRenderer.prototype.ctxTransform = function(props){
    if(props[0] === 1 && props[1] === 0 && props[4] === 0 && props[5] === 1 && props[12] === 0 && props[13] === 0){
        return;
    }
    if(!this.renderConfig.clearCanvas){
        this.canvasContext.transform(props[0],props[1],props[4],props[5],props[12],props[13]);
        return;
    }
    this.transformMat.cloneFromProps(props);
    var cProps = this.contextData.cTr.props;
    this.transformMat.transform(cProps[0],cProps[1],cProps[2],cProps[3],cProps[4],cProps[5],cProps[6],cProps[7],cProps[8],cProps[9],cProps[10],cProps[11],cProps[12],cProps[13],cProps[14],cProps[15]);
    //this.contextData.cTr.transform(props[0],props[1],props[2],props[3],props[4],props[5],props[6],props[7],props[8],props[9],props[10],props[11],props[12],props[13],props[14],props[15]);
    this.contextData.cTr.cloneFromProps(this.transformMat.props);
    var trProps = this.contextData.cTr.props;
    this.canvasContext.setTransform(trProps[0],trProps[1],trProps[4],trProps[5],trProps[12],trProps[13]);
};

CanvasRenderer.prototype.ctxOpacity = function(op){
    /*if(op === 1){
        return;
    }*/
    if(!this.renderConfig.clearCanvas){
        this.canvasContext.globalAlpha *= op < 0 ? 0 : op;
        this.globalData.currentGlobalAlpha = this.contextData.cO;
        return;
    }
    this.contextData.cO *= op < 0 ? 0 : op;
    if(this.globalData.currentGlobalAlpha !== this.contextData.cO) {
        this.canvasContext.globalAlpha = this.contextData.cO;
        this.globalData.currentGlobalAlpha = this.contextData.cO;
    }
};

CanvasRenderer.prototype.reset = function(){
    if(!this.renderConfig.clearCanvas){
        this.canvasContext.restore();
        return;
    }
    this.contextData.reset();
};

CanvasRenderer.prototype.save = function(actionFlag){
    if(!this.renderConfig.clearCanvas){
        this.canvasContext.save();
        return;
    }
    if(actionFlag){
        this.canvasContext.save();
    }
    var props = this.contextData.cTr.props;
    if(this.contextData._length <= this.contextData.cArrPos) {
        this.contextData.duplicate();
    }
    var i, arr = this.contextData.saved[this.contextData.cArrPos];
    for (i = 0; i < 16; i += 1) {
        arr[i] = props[i];
    }
    this.contextData.savedOp[this.contextData.cArrPos] = this.contextData.cO;
    this.contextData.cArrPos += 1;
};

CanvasRenderer.prototype.restore = function(actionFlag){
    if(!this.renderConfig.clearCanvas){
        this.canvasContext.restore();
        return;
    }
    if(actionFlag){
        this.canvasContext.restore();
        this.globalData.blendMode = 'source-over';
    }
    this.contextData.cArrPos -= 1;
    var popped = this.contextData.saved[this.contextData.cArrPos];
    var i,arr = this.contextData.cTr.props;
    for(i=0;i<16;i+=1){
        arr[i] = popped[i];
    }
    this.canvasContext.setTransform(popped[0],popped[1],popped[4],popped[5],popped[12],popped[13]);
    popped = this.contextData.savedOp[this.contextData.cArrPos];
    this.contextData.cO = popped;
    if(this.globalData.currentGlobalAlpha !== popped) {
        this.canvasContext.globalAlpha = popped;
        this.globalData.currentGlobalAlpha = popped;
    }
};

CanvasRenderer.prototype.configAnimation = function(animData){
    if(this.animationItem.wrapper){
        this.animationItem.container = createTag('canvas');
        this.animationItem.container.style.width = '100%';
        this.animationItem.container.style.height = '100%';
        //this.animationItem.container.style.transform = 'translate3d(0,0,0)';
        //this.animationItem.container.style.webkitTransform = 'translate3d(0,0,0)';
        this.animationItem.container.style.transformOrigin = this.animationItem.container.style.mozTransformOrigin = this.animationItem.container.style.webkitTransformOrigin = this.animationItem.container.style['-webkit-transform'] = "0px 0px 0px";
        this.animationItem.wrapper.appendChild(this.animationItem.container);
        this.canvasContext = this.animationItem.container.getContext('2d');
        if(this.renderConfig.className) {
            this.animationItem.container.setAttribute('class', this.renderConfig.className);
        }
    }else{
        this.canvasContext = this.renderConfig.context;
    }
    this.data = animData;
    this.layers = animData.layers;
    this.transformCanvas = {
        w: animData.w,
        h:animData.h,
        sx:0,
        sy:0,
        tx:0,
        ty:0
    };
    this.setupGlobalData(animData, document.body);
    this.globalData.canvasContext = this.canvasContext;
    this.globalData.renderer = this;
    this.globalData.isDashed = false;
    this.globalData.progressiveLoad = this.renderConfig.progressiveLoad;
    this.globalData.transformCanvas = this.transformCanvas;
    this.elements = createSizedArray(animData.layers.length);

    this.updateContainerSize();
};

CanvasRenderer.prototype.updateContainerSize = function () {
    this.reset();
    var elementWidth,elementHeight;
    if(this.animationItem.wrapper && this.animationItem.container){
        elementWidth = this.animationItem.wrapper.offsetWidth;
        elementHeight = this.animationItem.wrapper.offsetHeight;
        this.animationItem.container.setAttribute('width',elementWidth * this.renderConfig.dpr );
        this.animationItem.container.setAttribute('height',elementHeight * this.renderConfig.dpr);
    }else{
        elementWidth = this.canvasContext.canvas.width * this.renderConfig.dpr;
        elementHeight = this.canvasContext.canvas.height * this.renderConfig.dpr;
    }
    var elementRel,animationRel;
    if(this.renderConfig.preserveAspectRatio.indexOf('meet') !== -1 || this.renderConfig.preserveAspectRatio.indexOf('slice') !== -1){
        var par = this.renderConfig.preserveAspectRatio.split(' ');
        var fillType = par[1] || 'meet';
        var pos = par[0] || 'xMidYMid';
        var xPos = pos.substr(0,4);
        var yPos = pos.substr(4);
        elementRel = elementWidth/elementHeight;
        animationRel = this.transformCanvas.w/this.transformCanvas.h;
        if(animationRel>elementRel && fillType === 'meet' || animationRel<elementRel && fillType === 'slice'){
            this.transformCanvas.sx = elementWidth/(this.transformCanvas.w/this.renderConfig.dpr);
            this.transformCanvas.sy = elementWidth/(this.transformCanvas.w/this.renderConfig.dpr);
        }else{
            this.transformCanvas.sx = elementHeight/(this.transformCanvas.h / this.renderConfig.dpr);
            this.transformCanvas.sy = elementHeight/(this.transformCanvas.h / this.renderConfig.dpr);
        }

        if(xPos === 'xMid' && ((animationRel<elementRel && fillType==='meet') || (animationRel>elementRel && fillType === 'slice'))){
            this.transformCanvas.tx = (elementWidth-this.transformCanvas.w*(elementHeight/this.transformCanvas.h))/2*this.renderConfig.dpr;
        } else if(xPos === 'xMax' && ((animationRel<elementRel && fillType==='meet') || (animationRel>elementRel && fillType === 'slice'))){
            this.transformCanvas.tx = (elementWidth-this.transformCanvas.w*(elementHeight/this.transformCanvas.h))*this.renderConfig.dpr;
        } else {
            this.transformCanvas.tx = 0;
        }
        if(yPos === 'YMid' && ((animationRel>elementRel && fillType==='meet') || (animationRel<elementRel && fillType === 'slice'))){
            this.transformCanvas.ty = ((elementHeight-this.transformCanvas.h*(elementWidth/this.transformCanvas.w))/2)*this.renderConfig.dpr;
        } else if(yPos === 'YMax' && ((animationRel>elementRel && fillType==='meet') || (animationRel<elementRel && fillType === 'slice'))){
            this.transformCanvas.ty = ((elementHeight-this.transformCanvas.h*(elementWidth/this.transformCanvas.w)))*this.renderConfig.dpr;
        } else {
            this.transformCanvas.ty = 0;
        }

    }else if(this.renderConfig.preserveAspectRatio == 'none'){
        this.transformCanvas.sx = elementWidth/(this.transformCanvas.w/this.renderConfig.dpr);
        this.transformCanvas.sy = elementHeight/(this.transformCanvas.h/this.renderConfig.dpr);
        this.transformCanvas.tx = 0;
        this.transformCanvas.ty = 0;
    }else{
        this.transformCanvas.sx = this.renderConfig.dpr;
        this.transformCanvas.sy = this.renderConfig.dpr;
        this.transformCanvas.tx = 0;
        this.transformCanvas.ty = 0;
    }
    this.transformCanvas.props = [this.transformCanvas.sx,0,0,0,0,this.transformCanvas.sy,0,0,0,0,1,0,this.transformCanvas.tx,this.transformCanvas.ty,0,1];
    /*var i, len = this.elements.length;
    for(i=0;i<len;i+=1){
        if(this.elements[i] && this.elements[i].data.ty === 0){
            this.elements[i].resize(this.globalData.transformCanvas);
        }
    }*/
    this.ctxTransform(this.transformCanvas.props);
    this.canvasContext.beginPath();
    this.canvasContext.rect(0,0,this.transformCanvas.w,this.transformCanvas.h);
    this.canvasContext.closePath();
    this.canvasContext.clip();

    this.renderFrame(this.renderedFrame, true);
};

CanvasRenderer.prototype.destroy = function () {
    if(this.renderConfig.clearCanvas) {
        this.animationItem.wrapper.innerHTML = '';
    }
    var i, len = this.layers ? this.layers.length : 0;
    for (i = len - 1; i >= 0; i-=1) {
        if(this.elements[i]) {
            this.elements[i].destroy();
        }
    }
    this.elements.length = 0;
    this.globalData.canvasContext = null;
    this.animationItem.container = null;
    this.destroyed = true;
};

CanvasRenderer.prototype.renderFrame = function(num, forceRender){
    if((this.renderedFrame === num && this.renderConfig.clearCanvas === true && !forceRender) || this.destroyed || num === -1){
        return;
    }
    this.renderedFrame = num;
    this.globalData.frameNum = num - this.animationItem._isFirstFrame;
    this.globalData.frameId += 1;
    this.globalData._mdf = !this.renderConfig.clearCanvas || forceRender;
    this.globalData.projectInterface.currentFrame = num;

     // console.log('--------');
     // console.log('NEW: ',num);
    var i, len = this.layers.length;
    if(!this.completeLayers){
        this.checkLayers(num);
    }

    for (i = 0; i < len; i++) {
        if(this.completeLayers || this.elements[i]){
            this.elements[i].prepareFrame(num - this.layers[i].st);
        }
    }
    if(this.globalData._mdf) {
        if(this.renderConfig.clearCanvas === true){
            this.canvasContext.clearRect(0, 0, this.transformCanvas.w, this.transformCanvas.h);
        }else{
            this.save();
        }
        for (i = len - 1; i >= 0; i-=1) {
            if(this.completeLayers || this.elements[i]){
                this.elements[i].renderFrame();
            }
        }
        if(this.renderConfig.clearCanvas !== true){
            this.restore();
        }
    }
};

CanvasRenderer.prototype.buildItem = function(pos){
    var elements = this.elements;
    if(elements[pos] || this.layers[pos].ty == 99){
        return;
    }
    var element = this.createItem(this.layers[pos], this,this.globalData);
    elements[pos] = element;
    element.initExpressions();
    /*if(this.layers[pos].ty === 0){
        element.resize(this.globalData.transformCanvas);
    }*/
};

CanvasRenderer.prototype.checkPendingElements  = function(){
    while(this.pendingElements.length){
        var element = this.pendingElements.pop();
        element.checkParenting();
    }
};

CanvasRenderer.prototype.hide = function(){
    this.animationItem.container.style.display = 'none';
};

CanvasRenderer.prototype.show = function(){
    this.animationItem.container.style.display = 'block';
};

function HybridRenderer(animationItem, config){
    this.animationItem = animationItem;
    this.layers = null;
    this.renderedFrame = -1;
    this.renderConfig = {
        className: (config && config.className) || '',
        imagePreserveAspectRatio: (config && config.imagePreserveAspectRatio) || 'xMidYMid slice',
        hideOnTransparent: (config && config.hideOnTransparent === false) ? false : true
    };
    this.globalData = {
        _mdf: false,
        frameNum: -1,
        renderConfig: this.renderConfig
    };
    this.pendingElements = [];
    this.elements = [];
    this.threeDElements = [];
    this.destroyed = false;
    this.camera = null;
    this.supports3d = true;
    this.rendererType = 'html';

}

extendPrototype([BaseRenderer],HybridRenderer);

HybridRenderer.prototype.buildItem = SVGRenderer.prototype.buildItem;

HybridRenderer.prototype.checkPendingElements  = function(){
    while(this.pendingElements.length){
        var element = this.pendingElements.pop();
        element.checkParenting();
    }
};

HybridRenderer.prototype.appendElementInPos = function(element, pos){
    var newDOMElement = element.getBaseElement();
    if(!newDOMElement){
        return;
    }
    var layer = this.layers[pos];
    if(!layer.ddd || !this.supports3d){
        if(this.threeDElements) {
            this.addTo3dContainer(newDOMElement,pos);
        } else {
            var i = 0;
            var nextDOMElement, nextLayer, tmpDOMElement;
            while(i<pos){
                if(this.elements[i] && this.elements[i]!== true && this.elements[i].getBaseElement){
                    nextLayer = this.elements[i];
                    tmpDOMElement = this.layers[i].ddd ? this.getThreeDContainerByPos(i) : nextLayer.getBaseElement();
                    nextDOMElement = tmpDOMElement || nextDOMElement;
                }
                i += 1;
            }
            if(nextDOMElement){
                if(!layer.ddd || !this.supports3d){
                    this.layerElement.insertBefore(newDOMElement, nextDOMElement);
                }
            } else {
                if(!layer.ddd || !this.supports3d){
                    this.layerElement.appendChild(newDOMElement);
                }
            }
        }

    } else {
        this.addTo3dContainer(newDOMElement,pos);
    }
};

HybridRenderer.prototype.createShape = function (data) {
    if(!this.supports3d){
        return new SVGShapeElement(data, this.globalData, this);
    }
    return new HShapeElement(data, this.globalData, this);
};

HybridRenderer.prototype.createText = function (data) {
    if(!this.supports3d){
        return new SVGTextElement(data, this.globalData, this);
    }
    return new HTextElement(data, this.globalData, this);
};

HybridRenderer.prototype.createCamera = function (data) {
    this.camera = new HCameraElement(data, this.globalData, this);
    return this.camera;
};

HybridRenderer.prototype.createImage = function (data) {
    if(!this.supports3d){
        return new IImageElement(data, this.globalData, this);
    }
    return new HImageElement(data, this.globalData, this);
};

HybridRenderer.prototype.createComp = function (data) {
    if(!this.supports3d){
        return new SVGCompElement(data, this.globalData, this);
    }
    return new HCompElement(data, this.globalData, this);

};

HybridRenderer.prototype.createSolid = function (data) {
    if(!this.supports3d){
        return new ISolidElement(data, this.globalData, this);
    }
    return new HSolidElement(data, this.globalData, this);
};

HybridRenderer.prototype.createNull = SVGRenderer.prototype.createNull;

HybridRenderer.prototype.getThreeDContainerByPos = function(pos){
    var i = 0, len = this.threeDElements.length;
    while(i<len) {
        if(this.threeDElements[i].startPos <= pos && this.threeDElements[i].endPos >= pos) {
            return this.threeDElements[i].perspectiveElem;
        }
        i += 1;
    }
};

HybridRenderer.prototype.createThreeDContainer = function(pos, type){
    var perspectiveElem = createTag('div');
    styleDiv(perspectiveElem);
    var container = createTag('div');
    styleDiv(container);
    if(type === '3d') {
        perspectiveElem.style.width = this.globalData.compSize.w+'px';
        perspectiveElem.style.height = this.globalData.compSize.h+'px';
        perspectiveElem.style.transformOrigin = perspectiveElem.style.mozTransformOrigin = perspectiveElem.style.webkitTransformOrigin = "50% 50%";
        container.style.transform = container.style.webkitTransform = 'matrix3d(1,0,0,0,0,1,0,0,0,0,1,0,0,0,0,1)';
    }

    perspectiveElem.appendChild(container);
    //this.resizerElem.appendChild(perspectiveElem);
    var threeDContainerData = {
        container:container,
        perspectiveElem:perspectiveElem,
        startPos: pos,
        endPos: pos,
        type: type
    };
    this.threeDElements.push(threeDContainerData);
    return threeDContainerData;
};

HybridRenderer.prototype.build3dContainers = function(){
    var i, len = this.layers.length;
    var lastThreeDContainerData;
    var currentContainer = '';
    for(i=0;i<len;i+=1){
        if(this.layers[i].ddd && this.layers[i].ty !== 3){
            if(currentContainer !== '3d'){
                currentContainer = '3d';
                lastThreeDContainerData = this.createThreeDContainer(i,'3d');
            }
            lastThreeDContainerData.endPos = Math.max(lastThreeDContainerData.endPos,i);
        } else {
            if(currentContainer !== '2d'){
                currentContainer = '2d';
                lastThreeDContainerData = this.createThreeDContainer(i,'2d');
            }
            lastThreeDContainerData.endPos = Math.max(lastThreeDContainerData.endPos,i);
        }
    }
    len = this.threeDElements.length;
    for(i = len - 1; i >= 0; i --) {
        this.resizerElem.appendChild(this.threeDElements[i].perspectiveElem);
    }
};

HybridRenderer.prototype.addTo3dContainer = function(elem,pos){
    var i = 0, len = this.threeDElements.length;
    while(i<len){
        if(pos <= this.threeDElements[i].endPos){
            var j = this.threeDElements[i].startPos;
            var nextElement;
            while(j<pos){
                if(this.elements[j] && this.elements[j].getBaseElement){
                    nextElement = this.elements[j].getBaseElement();
                }
                j += 1;
            }
            if(nextElement){
                this.threeDElements[i].container.insertBefore(elem, nextElement);
            } else {
                this.threeDElements[i].container.appendChild(elem);
            }
            break;
        }
        i += 1;
    }
};

HybridRenderer.prototype.configAnimation = function(animData){
    var resizerElem = createTag('div');
    var wrapper = this.animationItem.wrapper;
    resizerElem.style.width = animData.w+'px';
    resizerElem.style.height = animData.h+'px';
    this.resizerElem = resizerElem;
    styleDiv(resizerElem);
    resizerElem.style.transformStyle = resizerElem.style.webkitTransformStyle = resizerElem.style.mozTransformStyle = "flat";
    if(this.renderConfig.className) {
      resizerElem.setAttribute('class', this.renderConfig.className);
    }
    wrapper.appendChild(resizerElem);

    resizerElem.style.overflow = 'hidden';
    var svg = createNS('svg');
    svg.setAttribute('width','1');
    svg.setAttribute('height','1');
    styleDiv(svg);
    this.resizerElem.appendChild(svg);
    var defs = createNS('defs');
    svg.appendChild(defs);
    this.data = animData;
    //Mask animation
    this.setupGlobalData(animData, svg);
    this.globalData.defs = defs;
    this.layers = animData.layers;
    this.layerElement = this.resizerElem;
    this.build3dContainers();
    this.updateContainerSize();
};

HybridRenderer.prototype.destroy = function () {
    this.animationItem.wrapper.innerHTML = '';
    this.animationItem.container = null;
    this.globalData.defs = null;
    var i, len = this.layers ? this.layers.length : 0;
    for (i = 0; i < len; i++) {
        this.elements[i].destroy();
    }
    this.elements.length = 0;
    this.destroyed = true;
    this.animationItem = null;
};

HybridRenderer.prototype.updateContainerSize = function () {
    var elementWidth = this.animationItem.wrapper.offsetWidth;
    var elementHeight = this.animationItem.wrapper.offsetHeight;
    var elementRel = elementWidth/elementHeight;
    var animationRel = this.globalData.compSize.w/this.globalData.compSize.h;
    var sx,sy,tx,ty;
    if(animationRel>elementRel){
        sx = elementWidth/(this.globalData.compSize.w);
        sy = elementWidth/(this.globalData.compSize.w);
        tx = 0;
        ty = ((elementHeight-this.globalData.compSize.h*(elementWidth/this.globalData.compSize.w))/2);
    }else{
        sx = elementHeight/(this.globalData.compSize.h);
        sy = elementHeight/(this.globalData.compSize.h);
        tx = (elementWidth-this.globalData.compSize.w*(elementHeight/this.globalData.compSize.h))/2;
        ty = 0;
    }
    this.resizerElem.style.transform = this.resizerElem.style.webkitTransform = 'matrix3d(' + sx + ',0,0,0,0,'+sy+',0,0,0,0,1,0,'+tx+','+ty+',0,1)';
};

HybridRenderer.prototype.renderFrame = SVGRenderer.prototype.renderFrame;

HybridRenderer.prototype.hide = function(){
    this.resizerElem.style.display = 'none';
};

HybridRenderer.prototype.show = function(){
    this.resizerElem.style.display = 'block';
};

HybridRenderer.prototype.initItems = function(){
    this.buildAllItems();
    if(this.camera){
        this.camera.setup();
    } else {
        var cWidth = this.globalData.compSize.w;
        var cHeight = this.globalData.compSize.h;
        var i, len = this.threeDElements.length;
        for(i=0;i<len;i+=1){
            this.threeDElements[i].perspectiveElem.style.perspective = this.threeDElements[i].perspectiveElem.style.webkitPerspective = Math.sqrt(Math.pow(cWidth,2) + Math.pow(cHeight,2)) + 'px';
        }
    }
};

HybridRenderer.prototype.searchExtraCompositions = function(assets){
    var i, len = assets.length;
    var floatingContainer = createTag('div');
    for(i=0;i<len;i+=1){
        if(assets[i].xt){
            var comp = this.createComp(assets[i],floatingContainer,this.globalData.comp,null);
            comp.initExpressions();
            this.globalData.projectInterface.registerComposition(comp);
        }
    }
};

function MaskElement(data,element,globalData) {
    this.data = data;
    this.element = element;
    this.globalData = globalData;
    this.storedData = [];
    this.masksProperties = this.data.masksProperties || [];
    this.maskElement = null;
    var defs = this.globalData.defs;
    var i, len = this.masksProperties ? this.masksProperties.length : 0;
    this.viewData = createSizedArray(len);
    this.solidPath = '';


    var path, properties = this.masksProperties;
    var count = 0;
    var currentMasks = [];
    var j, jLen;
    var layerId = createElementID();
    var rect, expansor, feMorph,x;
    var maskType = 'clipPath', maskRef = 'clip-path';
    for (i = 0; i < len; i++) {

        if((properties[i].mode !== 'a' && properties[i].mode !== 'n')|| properties[i].inv || properties[i].o.k !== 100){
            maskType = 'mask';
            maskRef = 'mask';
        }

        if((properties[i].mode == 's' || properties[i].mode == 'i') && count === 0){
            rect = createNS( 'rect');
            rect.setAttribute('fill', '#ffffff');
            rect.setAttribute('width', this.element.comp.data.w || 0);
            rect.setAttribute('height', this.element.comp.data.h || 0);
            currentMasks.push(rect);
        } else {
            rect = null;
        }

        path = createNS( 'path');
        if(properties[i].mode == 'n') {
            // TODO move this to a factory or to a constructor
            this.viewData[i] = {
                op: PropertyFactory.getProp(this.element,properties[i].o,0,0.01,this.element),
                prop: ShapePropertyFactory.getShapeProp(this.element,properties[i],3),
                elem: path,
                lastPath: ''
            };
            defs.appendChild(path);
            continue;
        }
        count += 1;

        path.setAttribute('fill', properties[i].mode === 's' ? '#000000':'#ffffff');
        path.setAttribute('clip-rule','nonzero');
        var filterID;

        if (properties[i].x.k !== 0) {
            maskType = 'mask';
            maskRef = 'mask';
            x = PropertyFactory.getProp(this.element,properties[i].x,0,null,this.element);
            filterID = createElementID();
            expansor = createNS('filter');
            expansor.setAttribute('id',filterID);
            feMorph = createNS('feMorphology');
            feMorph.setAttribute('operator','dilate');
            feMorph.setAttribute('in','SourceGraphic');
            feMorph.setAttribute('radius','0');
            expansor.appendChild(feMorph);
            defs.appendChild(expansor);
            path.setAttribute('stroke', properties[i].mode === 's' ? '#000000':'#ffffff');
        } else {
            feMorph = null;
            x = null;
        }

        // TODO move this to a factory or to a constructor
        this.storedData[i] = {
             elem: path,
             x: x,
             expan: feMorph,
            lastPath: '',
            lastOperator:'',
            filterId:filterID,
            lastRadius:0
        };
        if(properties[i].mode == 'i'){
            jLen = currentMasks.length;
            var g = createNS('g');
            for(j=0;j<jLen;j+=1){
                g.appendChild(currentMasks[j]);
            }
            var mask = createNS('mask');
            mask.setAttribute('mask-type','alpha');
            mask.setAttribute('id',layerId+'_'+count);
            mask.appendChild(path);
            defs.appendChild(mask);
            g.setAttribute('mask','url(' + locationHref + '#'+layerId+'_'+count+')');

            currentMasks.length = 0;
            currentMasks.push(g);
        }else{
            currentMasks.push(path);
        }
        if(properties[i].inv && !this.solidPath){
            this.solidPath = this.createLayerSolidPath();
        }
        // TODO move this to a factory or to a constructor
        this.viewData[i] = {
            elem: path,
            lastPath: '',
            op: PropertyFactory.getProp(this.element,properties[i].o,0,0.01,this.element),
            prop:ShapePropertyFactory.getShapeProp(this.element,properties[i],3),
            invRect: rect
        };
        if(!this.viewData[i].prop.k){
            this.drawPath(properties[i],this.viewData[i].prop.v,this.viewData[i]);
        }
    }

    this.maskElement = createNS( maskType);

    len = currentMasks.length;
    for(i=0;i<len;i+=1){
        this.maskElement.appendChild(currentMasks[i]);
    }

    if(count > 0){
        this.maskElement.setAttribute('id', layerId);
        this.element.maskedElement.setAttribute(maskRef, "url(" + locationHref + "#" + layerId + ")");
        defs.appendChild(this.maskElement);
    }
    if (this.viewData.length) {
        this.element.addRenderableComponent(this);
    }

}

MaskElement.prototype.getMaskProperty = function(pos){
    return this.viewData[pos].prop;
};

MaskElement.prototype.renderFrame = function (isFirstFrame) {
    var finalMat = this.element.finalTransform.mat;
    var i, len = this.masksProperties.length;
    for (i = 0; i < len; i++) {
        if(this.viewData[i].prop._mdf || isFirstFrame){
            this.drawPath(this.masksProperties[i],this.viewData[i].prop.v,this.viewData[i]);
        }
        if(this.viewData[i].op._mdf || isFirstFrame){
            this.viewData[i].elem.setAttribute('fill-opacity',this.viewData[i].op.v);
        }
        if(this.masksProperties[i].mode !== 'n'){
            if(this.viewData[i].invRect && (this.element.finalTransform.mProp._mdf || isFirstFrame)){
                this.viewData[i].invRect.setAttribute('x', -finalMat.props[12]);
                this.viewData[i].invRect.setAttribute('y', -finalMat.props[13]);
            }
            if(this.storedData[i].x && (this.storedData[i].x._mdf || isFirstFrame)){
                var feMorph = this.storedData[i].expan;
                if(this.storedData[i].x.v < 0){
                    if(this.storedData[i].lastOperator !== 'erode'){
                        this.storedData[i].lastOperator = 'erode';
                        this.storedData[i].elem.setAttribute('filter','url(' + locationHref + '#'+this.storedData[i].filterId+')');
                    }
                    feMorph.setAttribute('radius',-this.storedData[i].x.v);
                }else{
                    if(this.storedData[i].lastOperator !== 'dilate'){
                        this.storedData[i].lastOperator = 'dilate';
                        this.storedData[i].elem.setAttribute('filter',null);
                    }
                    this.storedData[i].elem.setAttribute('stroke-width', this.storedData[i].x.v*2);

                }
            }
        }
    }
};

MaskElement.prototype.getMaskelement = function () {
    return this.maskElement;
};

MaskElement.prototype.createLayerSolidPath = function(){
    var path = 'M0,0 ';
    path += ' h' + this.globalData.compSize.w ;
    path += ' v' + this.globalData.compSize.h ;
    path += ' h-' + this.globalData.compSize.w ;
    path += ' v-' + this.globalData.compSize.h + ' ';
    return path;
};

MaskElement.prototype.drawPath = function(pathData,pathNodes,viewData){
    var pathString = " M"+pathNodes.v[0][0]+','+pathNodes.v[0][1];
    var i, len;
    len = pathNodes._length;
    for(i=1;i<len;i+=1){
        //pathString += " C"+pathNodes.o[i-1][0]+','+pathNodes.o[i-1][1] + " "+pathNodes.i[i][0]+','+pathNodes.i[i][1] + " "+pathNodes.v[i][0]+','+pathNodes.v[i][1];
        pathString += " C"+pathNodes.o[i-1][0]+','+pathNodes.o[i-1][1] + " "+pathNodes.i[i][0]+','+pathNodes.i[i][1] + " "+pathNodes.v[i][0]+','+pathNodes.v[i][1];
    }
        //pathString += " C"+pathNodes.o[i-1][0]+','+pathNodes.o[i-1][1] + " "+pathNodes.i[0][0]+','+pathNodes.i[0][1] + " "+pathNodes.v[0][0]+','+pathNodes.v[0][1];
    if(pathNodes.c && len > 1){
        pathString += " C"+pathNodes.o[i-1][0]+','+pathNodes.o[i-1][1] + " "+pathNodes.i[0][0]+','+pathNodes.i[0][1] + " "+pathNodes.v[0][0]+','+pathNodes.v[0][1];
    }
    //pathNodes.__renderedString = pathString;

    if(viewData.lastPath !== pathString){
        var pathShapeValue = '';
        if(viewData.elem){
            if(pathNodes.c){
                pathShapeValue = pathData.inv ? this.solidPath + pathString : pathString;
            }
            viewData.elem.setAttribute('d',pathShapeValue);
        }
        viewData.lastPath = pathString;
    }
};

MaskElement.prototype.destroy = function(){
    this.element = null;
    this.globalData = null;
    this.maskElement = null;
    this.data = null;
    this.masksProperties = null;
};

/**
 * @file
 * Handles AE's layer parenting property.
 *
 */

function HierarchyElement(){}

HierarchyElement.prototype = {
	/**
     * @function
     * Initializes hierarchy properties
     *
     */
	initHierarchy: function() {
		//element's parent list
	    this.hierarchy = [];
	    //if element is parent of another layer _isParent will be true
	    this._isParent = false;
	    this.checkParenting();
	},
	/**
     * @function
     * Sets layer's hierarchy.
     * @param {array} hierarch
     * layer's parent list
     *
     */
	setHierarchy: function(hierarchy){
	    this.hierarchy = hierarchy;
	},
	/**
     * @function
     * Sets layer as parent.
     *
     */
	setAsParent: function() {
	    this._isParent = true;
	},
	/**
     * @function
     * Searches layer's parenting chain
     *
     */
	checkParenting: function(){
	    if (this.data.parent !== undefined){
	        this.comp.buildElementParenting(this, this.data.parent, []);
	    }
	}
};
/**
 * @file
 * Handles element's layer frame update.
 * Checks layer in point and out point
 *
 */

function FrameElement(){}

FrameElement.prototype = {
    /**
     * @function
     * Initializes frame related properties.
     *
     */
    initFrame: function(){
        //set to true when inpoint is rendered
        this._isFirstFrame = false;
        //list of animated properties
        this.dynamicProperties = [];
        // If layer has been modified in current tick this will be true
        this._mdf = false;
    },
    /**
     * @function
     * Calculates all dynamic values
     *
     * @param {number} num
     * current frame number in Layer's time
     * @param {boolean} isVisible
     * if layers is currently in range
     *
     */
    prepareProperties: function(num, isVisible) {
        var i, len = this.dynamicProperties.length;
        for (i = 0;i < len; i += 1) {
            if (isVisible || (this._isParent && this.dynamicProperties[i].propType === 'transform')) {
                this.dynamicProperties[i].getValue();
                if (this.dynamicProperties[i]._mdf) {
                    this.globalData._mdf = true;
                    this._mdf = true;
                }
            }
        }
    },
    addDynamicProperty: function(prop) {
        if(this.dynamicProperties.indexOf(prop) === -1) {
            this.dynamicProperties.push(prop);
        }
    }
};
function TransformElement(){}

TransformElement.prototype = {
    initTransform: function() {
        this.finalTransform = {
            mProp: this.data.ks ? TransformPropertyFactory.getTransformProperty(this, this.data.ks, this) : {o:0},
            _matMdf: false,
            _opMdf: false,
            mat: new Matrix()
        };
        if (this.data.ao) {
            this.finalTransform.mProp.autoOriented = true;
        }

        //TODO: check TYPE 11: Guided elements
        if (this.data.ty !== 11) {
            //this.createElements();
        }
    },
    renderTransform: function() {

        this.finalTransform._opMdf = this.finalTransform.mProp.o._mdf || this._isFirstFrame;
        this.finalTransform._matMdf = this.finalTransform.mProp._mdf || this._isFirstFrame;

        if (this.hierarchy) {
            var mat;
            var finalMat = this.finalTransform.mat;
            var i = 0, len = this.hierarchy.length;
            //Checking if any of the transformation matrices in the hierarchy chain has changed.
            if (!this.finalTransform._matMdf) {
                while (i < len) {
                    if (this.hierarchy[i].finalTransform.mProp._mdf) {
                        this.finalTransform._matMdf = true;
                        break;
                    }
                    i += 1;
                }
            }

            if (this.finalTransform._matMdf) {
                mat = this.finalTransform.mProp.v.props;
                finalMat.cloneFromProps(mat);
                for (i = 0; i < len; i += 1) {
                    mat = this.hierarchy[i].finalTransform.mProp.v.props;
                    finalMat.transform(mat[0], mat[1], mat[2], mat[3], mat[4], mat[5], mat[6], mat[7], mat[8], mat[9], mat[10], mat[11], mat[12], mat[13], mat[14], mat[15]);
                }
            }
        }
    },
    globalToLocal: function(pt) {
        var transforms = [];
        transforms.push(this.finalTransform);
        var flag = true;
        var comp = this.comp;
        while (flag) {
            if (comp.finalTransform) {
                if (comp.data.hasMask) {
                    transforms.splice(0, 0, comp.finalTransform);
                }
                comp = comp.comp;
            } else {
                flag = false;
            }
        }
        var i, len = transforms.length,ptNew;
        for (i = 0; i < len; i += 1) {
            ptNew = transforms[i].mat.applyToPointArray(0, 0, 0);
            //ptNew = transforms[i].mat.applyToPointArray(pt[0],pt[1],pt[2]);
            pt = [pt[0] - ptNew[0], pt[1] - ptNew[1], 0];
        }
        return pt;
    },
    mHelper: new Matrix()
};
function RenderableElement(){

}

RenderableElement.prototype = {
    initRenderable: function() {
        //layer's visibility related to inpoint and outpoint. Rename isVisible to isInRange
        this.isInRange = false;
        //layer's display state
        this.hidden = false;
        // If layer's transparency equals 0, it can be hidden
        this.isTransparent = false;
        //list of animated components
        this.renderableComponents = [];
    },
    addRenderableComponent: function(component) {
        if(this.renderableComponents.indexOf(component) === -1) {
            this.renderableComponents.push(component);
        }
    },
    removeRenderableComponent: function(component) {
        if(this.renderableComponents.indexOf(component) !== -1) {
            this.renderableComponents.splice(this.renderableComponents.indexOf(component), 1);
        }
    },
    prepareRenderableFrame: function(num) {
        this.checkLayerLimits(num);
    },
    checkTransparency: function(){
        if(this.finalTransform.mProp.o.v <= 0) {
            if(!this.isTransparent && this.globalData.renderConfig.hideOnTransparent){
                this.isTransparent = true;
                this.hide();
            }
        } else if(this.isTransparent) {
            this.isTransparent = false;
            this.show();
        }
    },
    /**
     * @function
     * Initializes frame related properties.
     *
     * @param {number} num
     * current frame number in Layer's time
     *
     */
    checkLayerLimits: function(num) {
        if(this.data.ip - this.data.st <= num && this.data.op - this.data.st > num)
        {
            if(this.isInRange !== true){
                this.globalData._mdf = true;
                this._mdf = true;
                this.isInRange = true;
                this.show();
            }
        } else {
            if(this.isInRange !== false){
                this.globalData._mdf = true;
                this.isInRange = false;
                this.hide();
            }
        }
    },
    renderRenderable: function() {
        var i, len = this.renderableComponents.length;
        for(i = 0; i < len; i += 1) {
            this.renderableComponents[i].renderFrame(this._isFirstFrame);
        }
        /*this.maskManager.renderFrame(this.finalTransform.mat);
        this.renderableEffectsManager.renderFrame(this._isFirstFrame);*/
    },
    sourceRectAtTime: function(){
        return {
            top:0,
            left:0,
            width:100,
            height:100
        };
    },
    getLayerSize: function(){
        if(this.data.ty === 5){
            return {w:this.data.textData.width,h:this.data.textData.height};
        }else{
            return {w:this.data.width,h:this.data.height};
        }
    }
};
function RenderableDOMElement() {}

(function(){
    var _prototype = {
        initElement: function(data,globalData,comp) {
            this.initFrame();
            this.initBaseData(data, globalData, comp);
            this.initTransform(data, globalData, comp);
            this.initHierarchy();
            this.initRenderable();
            this.initRendererElement();
            this.createContainerElements();
            this.createRenderableComponents();
            this.createContent();
            this.hide();
        },
        hide: function(){
            if (!this.hidden && (!this.isInRange || this.isTransparent)) {
                var elem = this.baseElement || this.layerElement;
                elem.style.display = 'none';
                this.hidden = true;
            }
        },
        show: function(){
            if (this.isInRange && !this.isTransparent){
                if (!this.data.hd) {
                    var elem = this.baseElement || this.layerElement;
                    elem.style.display = 'block';
                }
                this.hidden = false;
                this._isFirstFrame = true;
            }
        },
        renderFrame: function() {
            //If it is exported as hidden (data.hd === true) no need to render
            //If it is not visible no need to render
            if (this.data.hd || this.hidden) {
                return;
            }
            this.renderTransform();
            this.renderRenderable();
            this.renderElement();
            this.renderInnerContent();
            if (this._isFirstFrame) {
                this._isFirstFrame = false;
            }
        },
        renderInnerContent: function() {},
        prepareFrame: function(num) {
            this._mdf = false;
            this.prepareRenderableFrame(num);
            this.prepareProperties(num, this.isInRange);
            this.checkTransparency();
        },
        destroy: function(){
            this.innerElem =  null;
            this.destroyBaseElement();
        }
    };
    extendPrototype([RenderableElement, createProxyFunction(_prototype)], RenderableDOMElement);
}());
function ProcessedElement(element, position) {
	this.elem = element;
	this.pos = position;
}
function SVGStyleData(data, level) {
	this.data = data;
	this.type = data.ty;
	this.d = '';
	this.lvl = level;
	this._mdf = false;
	this.closed = data.hd === true;
	this.pElem = createNS('path');
	this.msElem = null;
}

SVGStyleData.prototype.reset = function() {
	this.d = '';
	this._mdf = false;
};
function SVGShapeData(transformers, level, shape) {
    this.caches = [];
    this.styles = [];
    this.transformers = transformers;
    this.lStr = '';
    this.sh = shape;
    this.lvl = level;
    //TODO find if there are some cases where _isAnimated can be false.
    // For now, since shapes add up with other shapes. They have to be calculated every time.
    // One way of finding out is checking if all styles associated to this shape depend only of this shape
    this._isAnimated = !!shape.k;
    // TODO: commenting this for now since all shapes are animated
    var i = 0, len = transformers.length;
    while(i < len) {
    	if(transformers[i].mProps.dynamicProperties.length) {
    		this._isAnimated = true;
    		break;
    	}
    	i += 1;
    }
}

SVGShapeData.prototype.setAsAnimated = function() {
    this._isAnimated = true;
}
function SVGTransformData(mProps, op, container) {
	this.transform = {
		mProps: mProps,
		op: op,
		container: container
	};
	this.elements = [];
    this._isAnimated = this.transform.mProps.dynamicProperties.length || this.transform.op.effectsSequence.length;
}
function SVGStrokeStyleData(elem, data, styleOb){
	this.initDynamicPropertyContainer(elem);
	this.getValue = this.iterateDynamicProperties;
	this.o = PropertyFactory.getProp(elem,data.o,0,0.01,this);
	this.w = PropertyFactory.getProp(elem,data.w,0,null,this);
	this.d = new DashProperty(elem,data.d||{},'svg',this);
	this.c = PropertyFactory.getProp(elem,data.c,1,255,this);
	this.style = styleOb;
    this._isAnimated = !!this._isAnimated;
}

extendPrototype([DynamicPropertyContainer], SVGStrokeStyleData);
function SVGFillStyleData(elem, data, styleOb){
	this.initDynamicPropertyContainer(elem);
	this.getValue = this.iterateDynamicProperties;
	this.o = PropertyFactory.getProp(elem,data.o,0,0.01,this);
	this.c = PropertyFactory.getProp(elem,data.c,1,255,this);
	this.style = styleOb;
}

extendPrototype([DynamicPropertyContainer], SVGFillStyleData);
function SVGGradientFillStyleData(elem, data, styleOb){
    this.initDynamicPropertyContainer(elem);
    this.getValue = this.iterateDynamicProperties;
    this.initGradientData(elem, data, styleOb);
}

SVGGradientFillStyleData.prototype.initGradientData = function(elem, data, styleOb){
    this.o = PropertyFactory.getProp(elem,data.o,0,0.01,this);
    this.s = PropertyFactory.getProp(elem,data.s,1,null,this);
    this.e = PropertyFactory.getProp(elem,data.e,1,null,this);
    this.h = PropertyFactory.getProp(elem,data.h||{k:0},0,0.01,this);
    this.a = PropertyFactory.getProp(elem,data.a||{k:0},0,degToRads,this);
    this.g = new GradientProperty(elem,data.g,this);
    this.style = styleOb;
    this.stops = [];
    this.setGradientData(styleOb.pElem, data);
    this.setGradientOpacity(data, styleOb);
    this._isAnimated = !!this._isAnimated;

};

SVGGradientFillStyleData.prototype.setGradientData = function(pathElement,data){

    var gradientId = createElementID();
    var gfill = createNS(data.t === 1 ? 'linearGradient' : 'radialGradient');
    gfill.setAttribute('id',gradientId);
    gfill.setAttribute('spreadMethod','pad');
    gfill.setAttribute('gradientUnits','userSpaceOnUse');
    var stops = [];
    var stop, j, jLen;
    jLen = data.g.p*4;
    for(j=0;j<jLen;j+=4){
        stop = createNS('stop');
        gfill.appendChild(stop);
        stops.push(stop);
    }
    pathElement.setAttribute( data.ty === 'gf' ? 'fill':'stroke','url(' + locationHref + '#'+gradientId+')');

    this.gf = gfill;
    this.cst = stops;
};

SVGGradientFillStyleData.prototype.setGradientOpacity = function(data, styleOb){
    if(this.g._hasOpacity && !this.g._collapsable){
        var stop, j, jLen;
        var mask = createNS("mask");
        var maskElement = createNS( 'path');
        mask.appendChild(maskElement);
        var opacityId = createElementID();
        var maskId = createElementID();
        mask.setAttribute('id',maskId);
        var opFill = createNS(data.t === 1 ? 'linearGradient' : 'radialGradient');
        opFill.setAttribute('id',opacityId);
        opFill.setAttribute('spreadMethod','pad');
        opFill.setAttribute('gradientUnits','userSpaceOnUse');
        jLen = data.g.k.k[0].s ? data.g.k.k[0].s.length : data.g.k.k.length;
        var stops = this.stops;
        for(j=data.g.p*4;j<jLen;j+=2){
            stop = createNS('stop');
            stop.setAttribute('stop-color','rgb(255,255,255)');
            opFill.appendChild(stop);
            stops.push(stop);
        }
        maskElement.setAttribute( data.ty === 'gf' ? 'fill':'stroke','url(' + locationHref + '#'+opacityId+')');
        this.of = opFill;
        this.ms = mask;
        this.ost = stops;
        this.maskId = maskId;
        styleOb.msElem = maskElement;
    }
};

extendPrototype([DynamicPropertyContainer], SVGGradientFillStyleData);
function SVGGradientStrokeStyleData(elem, data, styleOb){
	this.initDynamicPropertyContainer(elem);
	this.getValue = this.iterateDynamicProperties;
	this.w = PropertyFactory.getProp(elem,data.w,0,null,this);
	this.d = new DashProperty(elem,data.d||{},'svg',this);
    this.initGradientData(elem, data, styleOb);
    this._isAnimated = !!this._isAnimated;
}

extendPrototype([SVGGradientFillStyleData, DynamicPropertyContainer], SVGGradientStrokeStyleData);
function ShapeGroupData() {
	this.it = [];
    this.prevViewData = [];
    this.gr = createNS('g');
}
var SVGElementsRenderer = (function() {
	var _identityMatrix = new Matrix();
	var _matrixHelper = new Matrix();

	var ob = {
		createRenderFunction: createRenderFunction
	}

	function createRenderFunction(data) {
	    var ty = data.ty;
	    switch(data.ty) {
	        case 'fl':
	        return renderFill;
	        case 'gf':
	        return renderGradient;
	        case 'gs':
	        return renderGradientStroke;
	        case 'st':
	        return renderStroke;
	        case 'sh':
	        case 'el':
	        case 'rc':
	        case 'sr':
	        return renderPath;
	        case 'tr':
	        return renderContentTransform;
	    }
	}

	function renderContentTransform(styleData, itemData, isFirstFrame) {
	    if(isFirstFrame || itemData.transform.op._mdf){
	        itemData.transform.container.setAttribute('opacity',itemData.transform.op.v);
	    }
	    if(isFirstFrame || itemData.transform.mProps._mdf){
	        itemData.transform.container.setAttribute('transform',itemData.transform.mProps.v.to2dCSS());
	    }
	}

	function renderPath(styleData, itemData, isFirstFrame) {
	    var j, jLen,pathStringTransformed,redraw,pathNodes,l, lLen = itemData.styles.length;
	    var lvl = itemData.lvl;
	    var paths, mat, props, iterations, k;
	    for(l=0;l<lLen;l+=1){
	        redraw = itemData.sh._mdf || isFirstFrame;
	        if(itemData.styles[l].lvl < lvl){
	            mat = _matrixHelper.reset();
	            iterations = lvl - itemData.styles[l].lvl;
	            k = itemData.transformers.length-1;
	            while(!redraw && iterations > 0) {
	                redraw = itemData.transformers[k].mProps._mdf || redraw;
	                iterations --;
	                k --;
	            }
	            if(redraw) {
	                iterations = lvl - itemData.styles[l].lvl;
	                k = itemData.transformers.length-1;
	                while(iterations > 0) {
	                    props = itemData.transformers[k].mProps.v.props;
	                    mat.transform(props[0],props[1],props[2],props[3],props[4],props[5],props[6],props[7],props[8],props[9],props[10],props[11],props[12],props[13],props[14],props[15]);
	                    iterations --;
	                    k --;
	                }
	            }
	        } else {
	            mat = _identityMatrix;
	        }
	        paths = itemData.sh.paths;
	        jLen = paths._length;
	        if(redraw){
	            pathStringTransformed = '';
	            for(j=0;j<jLen;j+=1){
	                pathNodes = paths.shapes[j];
	                if(pathNodes && pathNodes._length){
	                    pathStringTransformed += buildShapeString(pathNodes, pathNodes._length, pathNodes.c, mat);
	                }
	            }
	            itemData.caches[l] = pathStringTransformed;
	        } else {
	            pathStringTransformed = itemData.caches[l];
	        }
	        itemData.styles[l].d += styleData.hd === true ? '' : pathStringTransformed;
	        itemData.styles[l]._mdf = redraw || itemData.styles[l]._mdf;
	    }
	}

	function renderFill (styleData,itemData, isFirstFrame){
	    var styleElem = itemData.style;

	    if(itemData.c._mdf || isFirstFrame){
	        styleElem.pElem.setAttribute('fill','rgb('+bm_floor(itemData.c.v[0])+','+bm_floor(itemData.c.v[1])+','+bm_floor(itemData.c.v[2])+')');
	    }
	    if(itemData.o._mdf || isFirstFrame){
	        styleElem.pElem.setAttribute('fill-opacity',itemData.o.v);
	    }
	};

	function renderGradientStroke (styleData, itemData, isFirstFrame) {
	    renderGradient(styleData, itemData, isFirstFrame);
	    renderStroke(styleData, itemData, isFirstFrame);
	}

	function renderGradient(styleData, itemData, isFirstFrame) {
	    var gfill = itemData.gf;
	    var hasOpacity = itemData.g._hasOpacity;
	    var pt1 = itemData.s.v, pt2 = itemData.e.v;

	    if (itemData.o._mdf || isFirstFrame) {
	        var attr = styleData.ty === 'gf' ? 'fill-opacity' : 'stroke-opacity';
	        itemData.style.pElem.setAttribute(attr, itemData.o.v);
	    }
	    if (itemData.s._mdf || isFirstFrame) {
	        var attr1 = styleData.t === 1 ? 'x1' : 'cx';
	        var attr2 = attr1 === 'x1' ? 'y1' : 'cy';
	        gfill.setAttribute(attr1, pt1[0]);
	        gfill.setAttribute(attr2, pt1[1]);
	        if (hasOpacity && !itemData.g._collapsable) {
	            itemData.of.setAttribute(attr1, pt1[0]);
	            itemData.of.setAttribute(attr2, pt1[1]);
	        }
	    }
	    var stops, i, len, stop;
	    if (itemData.g._cmdf || isFirstFrame) {
	        stops = itemData.cst;
	        var cValues = itemData.g.c;
	        len = stops.length;
	        for (i = 0; i < len; i += 1){
	            stop = stops[i];
	            stop.setAttribute('offset', cValues[i * 4] + '%');
	            stop.setAttribute('stop-color','rgb('+ cValues[i * 4 + 1] + ',' + cValues[i * 4 + 2] + ','+cValues[i * 4 + 3] + ')');
	        }
	    }
	    if (hasOpacity && (itemData.g._omdf || isFirstFrame)) {
	        var oValues = itemData.g.o;
	        if(itemData.g._collapsable) {
	            stops = itemData.cst;
	        } else {
	            stops = itemData.ost;
	        }
	        len = stops.length;
	        for (i = 0; i < len; i += 1) {
	            stop = stops[i];
	            if(!itemData.g._collapsable) {
	                stop.setAttribute('offset', oValues[i * 2] + '%');
	            }
	            stop.setAttribute('stop-opacity', oValues[i * 2 + 1]);
	        }
	    }
	    if (styleData.t === 1) {
	        if (itemData.e._mdf  || isFirstFrame) {
	            gfill.setAttribute('x2', pt2[0]);
	            gfill.setAttribute('y2', pt2[1]);
	            if (hasOpacity && !itemData.g._collapsable) {
	                itemData.of.setAttribute('x2', pt2[0]);
	                itemData.of.setAttribute('y2', pt2[1]);
	            }
	        }
	    } else {
	        var rad;
	        if (itemData.s._mdf || itemData.e._mdf || isFirstFrame) {
	            rad = Math.sqrt(Math.pow(pt1[0] - pt2[0], 2) + Math.pow(pt1[1] - pt2[1], 2));
	            gfill.setAttribute('r', rad);
	            if(hasOpacity && !itemData.g._collapsable){
	                itemData.of.setAttribute('r', rad);
	            }
	        }
	        if (itemData.e._mdf || itemData.h._mdf || itemData.a._mdf || isFirstFrame) {
	            if (!rad) {
	                rad = Math.sqrt(Math.pow(pt1[0] - pt2[0], 2) + Math.pow(pt1[1] - pt2[1], 2));
	            }
	            var ang = Math.atan2(pt2[1] - pt1[1], pt2[0] - pt1[0]);

	            var percent = itemData.h.v >= 1 ? 0.99 : itemData.h.v <= -1 ? -0.99: itemData.h.v;
	            var dist = rad * percent;
	            var x = Math.cos(ang + itemData.a.v) * dist + pt1[0];
	            var y = Math.sin(ang + itemData.a.v) * dist + pt1[1];
	            gfill.setAttribute('fx', x);
	            gfill.setAttribute('fy', y);
	            if (hasOpacity && !itemData.g._collapsable) {
	                itemData.of.setAttribute('fx', x);
	                itemData.of.setAttribute('fy', y);
	            }
	        }
	        //gfill.setAttribute('fy','200');
	    }
	};

	function renderStroke(styleData, itemData, isFirstFrame) {
	    var styleElem = itemData.style;
	    var d = itemData.d;
	    if (d && (d._mdf || isFirstFrame) && d.dashStr) {
	        styleElem.pElem.setAttribute('stroke-dasharray', d.dashStr);
	        styleElem.pElem.setAttribute('stroke-dashoffset', d.dashoffset[0]);
	    }
	    if(itemData.c && (itemData.c._mdf || isFirstFrame)){
	        styleElem.pElem.setAttribute('stroke','rgb(' + bm_floor(itemData.c.v[0]) + ',' + bm_floor(itemData.c.v[1]) + ',' + bm_floor(itemData.c.v[2]) + ')');
	    }
	    if(itemData.o._mdf || isFirstFrame){
	        styleElem.pElem.setAttribute('stroke-opacity', itemData.o.v);
	    }
	    if(itemData.w._mdf || isFirstFrame){
	        styleElem.pElem.setAttribute('stroke-width', itemData.w.v);
	        if(styleElem.msElem){
	            styleElem.msElem.setAttribute('stroke-width', itemData.w.v);
	        }
	    }
	};

	return ob;
}())
function ShapeTransformManager() {
	this.sequences = {};
	this.sequenceList = [];
    this.transform_key_count = 0;
}

ShapeTransformManager.prototype = {
	addTransformSequence: function(transforms) {
		var i, len = transforms.length;
		var key = '_';
		for(i = 0; i < len; i += 1) {
			key += transforms[i].transform.key + '_';
		}
		var sequence = this.sequences[key];
		if(!sequence) {
			sequence = {
				transforms: [].concat(transforms),
				finalTransform: new Matrix(),
				_mdf: false
			};
			this.sequences[key] = sequence;
			this.sequenceList.push(sequence);
		}
		return sequence;
	},
	processSequence: function(sequence, isFirstFrame) {
		var i = 0, len = sequence.transforms.length, _mdf = isFirstFrame;
		while (i < len && !isFirstFrame) {
			if (sequence.transforms[i].transform.mProps._mdf) {
				_mdf = true;
				break;
			}
			i += 1
		}
		if (_mdf) {
			var props;
			sequence.finalTransform.reset();
			for (i = len - 1; i >= 0; i -= 1) {
		        props = sequence.transforms[i].transform.mProps.v.props;
		        sequence.finalTransform.transform(props[0],props[1],props[2],props[3],props[4],props[5],props[6],props[7],props[8],props[9],props[10],props[11],props[12],props[13],props[14],props[15]);
			}
		}
		sequence._mdf = _mdf;

	},
	processSequences: function(isFirstFrame) {
		var i, len = this.sequenceList.length;
		for (i = 0; i < len; i += 1) {
			this.processSequence(this.sequenceList[i], isFirstFrame);
		}

	},
	getNewKey: function() {
		return '_' + this.transform_key_count++;
	}
}
function CVShapeData(element, data, styles, transformsManager) {
    this.styledShapes = [];
    this.tr = [0,0,0,0,0,0];
    var ty = 4;
    if(data.ty == 'rc'){
        ty = 5;
    }else if(data.ty == 'el'){
        ty = 6;
    }else if(data.ty == 'sr'){
        ty = 7;
    }
    this.sh = ShapePropertyFactory.getShapeProp(element,data,ty,element);
    var i , len = styles.length,styledShape;
    for (i = 0; i < len; i += 1) {
        if (!styles[i].closed) {
            styledShape = {
                transforms: transformsManager.addTransformSequence(styles[i].transforms),
                trNodes: []
            }
            this.styledShapes.push(styledShape);
            styles[i].elements.push(styledShape);
        }
    }
}

CVShapeData.prototype.setAsAnimated = SVGShapeData.prototype.setAsAnimated;
function BaseElement(){
}

BaseElement.prototype = {
    checkMasks: function(){
        if(!this.data.hasMask){
            return false;
        }
        var i = 0, len = this.data.masksProperties.length;
        while(i<len) {
            if((this.data.masksProperties[i].mode !== 'n' && this.data.masksProperties[i].cl !== false)) {
                return true;
            }
            i += 1;
        }
        return false;
    },
    initExpressions: function(){
        this.layerInterface = LayerExpressionInterface(this);
        if(this.data.hasMask && this.maskManager) {
            this.layerInterface.registerMaskInterface(this.maskManager);
        }
        var effectsInterface = EffectsExpressionInterface.createEffectsInterface(this,this.layerInterface);
        this.layerInterface.registerEffectsInterface(effectsInterface);

        if(this.data.ty === 0 || this.data.xt){
            this.compInterface = CompExpressionInterface(this);
        } else if(this.data.ty === 4){
            this.layerInterface.shapeInterface = ShapeExpressionInterface(this.shapesData,this.itemsData,this.layerInterface);
            this.layerInterface.content = this.layerInterface.shapeInterface;
        } else if(this.data.ty === 5){
            this.layerInterface.textInterface = TextExpressionInterface(this);
            this.layerInterface.text = this.layerInterface.textInterface;
        }
    },
    setBlendMode: function(){
        var blendModeValue = getBlendMode(this.data.bm);
        var elem = this.baseElement || this.layerElement;

        elem.style['mix-blend-mode'] = blendModeValue;
    },
    initBaseData: function(data, globalData, comp){
        this.globalData = globalData;
        this.comp = comp;
        this.data = data;
        this.layerId = createElementID();

        //Stretch factor for old animations missing this property.
        if(!this.data.sr){
            this.data.sr = 1;
        }
        // effects manager
        this.effectsManager = new EffectsManager(this.data,this,this.dynamicProperties);

    },
    getType: function(){
        return this.type;
    }
    ,sourceRectAtTime: function(){}
}
function NullElement(data,globalData,comp){
    this.initFrame();
	this.initBaseData(data, globalData, comp);
    this.initFrame();
    this.initTransform(data, globalData, comp);
    this.initHierarchy();
}

NullElement.prototype.prepareFrame = function(num) {
    this.prepareProperties(num, true);
};

NullElement.prototype.renderFrame = function() {
};

NullElement.prototype.getBaseElement = function() {
	return null;
};

NullElement.prototype.destroy = function() {
};

NullElement.prototype.sourceRectAtTime = function() {
};

NullElement.prototype.hide = function() {
};

extendPrototype([BaseElement,TransformElement,HierarchyElement,FrameElement], NullElement);

function SVGBaseElement(){
}

SVGBaseElement.prototype = {
    initRendererElement: function() {
        this.layerElement = createNS('g');
    },
    createContainerElements: function(){
        this.matteElement = createNS('g');
        this.transformedElement = this.layerElement;
        this.maskedElement = this.layerElement;
        this._sizeChanged = false;
        var layerElementParent = null;
        //If this layer acts as a mask for the following layer
        var filId, fil, gg;
        if (this.data.td) {
            if (this.data.td == 3 || this.data.td == 1) {
                var masker = createNS('mask');
                masker.setAttribute('id', this.layerId);
                masker.setAttribute('mask-type', this.data.td == 3 ? 'luminance' : 'alpha');
                masker.appendChild(this.layerElement);
                layerElementParent = masker;
                this.globalData.defs.appendChild(masker);
                // This is only for IE and Edge when mask if of type alpha
                if (!featureSupport.maskType && this.data.td == 1) {
                    masker.setAttribute('mask-type', 'luminance');
                    filId = createElementID();
                    fil = filtersFactory.createFilter(filId);
                    this.globalData.defs.appendChild(fil);
                    fil.appendChild(filtersFactory.createAlphaToLuminanceFilter());
                    gg = createNS('g');
                    gg.appendChild(this.layerElement);
                    layerElementParent = gg;
                    masker.appendChild(gg);
                    gg.setAttribute('filter','url(' + locationHref + '#' + filId + ')');
                }
            } else if(this.data.td == 2) {
                var maskGroup = createNS('mask');
                maskGroup.setAttribute('id', this.layerId);
                maskGroup.setAttribute('mask-type','alpha');
                var maskGrouper = createNS('g');
                maskGroup.appendChild(maskGrouper);
                filId = createElementID();
                fil = filtersFactory.createFilter(filId);
                ////

                // This solution doesn't work on Android when meta tag with viewport attribute is set
                /*var feColorMatrix = createNS('feColorMatrix');
                feColorMatrix.setAttribute('type', 'matrix');
                feColorMatrix.setAttribute('color-interpolation-filters', 'sRGB');
                feColorMatrix.setAttribute('values','1 0 0 0 0 0 1 0 0 0 0 0 1 0 0 0 0 0 -1 1');
                fil.appendChild(feColorMatrix);*/
                ////
                var feCTr = createNS('feComponentTransfer');
                feCTr.setAttribute('in','SourceGraphic');
                fil.appendChild(feCTr);
                var feFunc = createNS('feFuncA');
                feFunc.setAttribute('type','table');
                feFunc.setAttribute('tableValues','1.0 0.0');
                feCTr.appendChild(feFunc);
                ////
                this.globalData.defs.appendChild(fil);
                var alphaRect = createNS('rect');
                alphaRect.setAttribute('width',  this.comp.data.w);
                alphaRect.setAttribute('height', this.comp.data.h);
                alphaRect.setAttribute('x','0');
                alphaRect.setAttribute('y','0');
                alphaRect.setAttribute('fill','#ffffff');
                alphaRect.setAttribute('opacity','0');
                maskGrouper.setAttribute('filter', 'url(' + locationHref + '#'+filId+')');
                maskGrouper.appendChild(alphaRect);
                maskGrouper.appendChild(this.layerElement);
                layerElementParent = maskGrouper;
                if (!featureSupport.maskType) {
                    maskGroup.setAttribute('mask-type', 'luminance');
                    fil.appendChild(filtersFactory.createAlphaToLuminanceFilter());
                    gg = createNS('g');
                    maskGrouper.appendChild(alphaRect);
                    gg.appendChild(this.layerElement);
                    layerElementParent = gg;
                    maskGrouper.appendChild(gg);
                }
                this.globalData.defs.appendChild(maskGroup);
            }
        } else if (this.data.tt) {
            this.matteElement.appendChild(this.layerElement);
            layerElementParent = this.matteElement;
            this.baseElement = this.matteElement;
        } else {
            this.baseElement = this.layerElement;
        }
        if (this.data.ln) {
            this.layerElement.setAttribute('id', this.data.ln);
        }
        if (this.data.cl) {
            this.layerElement.setAttribute('class', this.data.cl);
        }
        //Clipping compositions to hide content that exceeds boundaries. If collapsed transformations is on, component should not be clipped
        if (this.data.ty === 0 && !this.data.hd) {
            var cp = createNS( 'clipPath');
            var pt = createNS('path');
            pt.setAttribute('d','M0,0 L' + this.data.w + ',0' + ' L' + this.data.w + ',' + this.data.h + ' L0,' + this.data.h + 'z');
            var clipId = createElementID();
            cp.setAttribute('id',clipId);
            cp.appendChild(pt);
            this.globalData.defs.appendChild(cp);

            if (this.checkMasks()) {
                var cpGroup = createNS('g');
                cpGroup.setAttribute('clip-path','url(' + locationHref + '#'+clipId + ')');
                cpGroup.appendChild(this.layerElement);
                this.transformedElement = cpGroup;
                if (layerElementParent) {
                    layerElementParent.appendChild(this.transformedElement);
                } else {
                    this.baseElement = this.transformedElement;
                }
            } else {
                this.layerElement.setAttribute('clip-path','url(' + locationHref + '#'+clipId+')');
            }

        }
        if (this.data.bm !== 0) {
            this.setBlendMode();
        }

    },
    renderElement: function() {
        if (this.finalTransform._matMdf) {
            this.transformedElement.setAttribute('transform', this.finalTransform.mat.to2dCSS());
        }
        if (this.finalTransform._opMdf) {
            this.transformedElement.setAttribute('opacity', this.finalTransform.mProp.o.v);
        }
    },
    destroyBaseElement: function() {
        this.layerElement = null;
        this.matteElement = null;
        this.maskManager.destroy();
    },
    getBaseElement: function() {
        if (this.data.hd) {
            return null;
        }
        return this.baseElement;
    },
    createRenderableComponents: function() {
        this.maskManager = new MaskElement(this.data, this, this.globalData);
        this.renderableEffectsManager = new SVGEffects(this);
    },
    setMatte: function(id) {
        if (!this.matteElement) {
            return;
        }
        this.matteElement.setAttribute("mask", "url(" + locationHref + "#" + id + ")");
    }
};
function IShapeElement(){
}

IShapeElement.prototype = {
    addShapeToModifiers: function(data) {
        var i, len = this.shapeModifiers.length;
        for(i=0;i<len;i+=1){
            this.shapeModifiers[i].addShape(data);
        }
    },
    isShapeInAnimatedModifiers: function(data) {
        var i = 0, len = this.shapeModifiers.length;
        while(i < len) {
            if(this.shapeModifiers[i].isAnimatedWithShape(data)) {
                return true;
            }
        }
        return false;
    },
    renderModifiers: function() {
        if(!this.shapeModifiers.length){
            return;
        }
        var i, len = this.shapes.length;
        for(i=0;i<len;i+=1){
            this.shapes[i].sh.reset();
        }

        len = this.shapeModifiers.length;
        for(i=len-1;i>=0;i-=1){
            this.shapeModifiers[i].processShapes(this._isFirstFrame);
        }
    },
    lcEnum: {
        '1': 'butt',
        '2': 'round',
        '3': 'square'
    },
    ljEnum: {
        '1': 'miter',
        '2': 'round',
        '3': 'bevel'
    },
    searchProcessedElement: function(elem){
        var elements = this.processedElements;
        var i = 0, len = elements.length;
        while (i < len) {
            if (elements[i].elem === elem) {
                return elements[i].pos;
            }
            i += 1;
        }
        return 0;
    },
    addProcessedElement: function(elem, pos){
        var elements = this.processedElements;
        var i = elements.length;
        while(i) {
            i -= 1;
            if (elements[i].elem === elem) {
                elements[i].pos = pos;
                return;
            }
        }
        elements.push(new ProcessedElement(elem, pos));
    },
    prepareFrame: function(num) {
        this.prepareRenderableFrame(num);
        this.prepareProperties(num, this.isInRange);
    }
};
function ITextElement(){
}

ITextElement.prototype.initElement = function(data,globalData,comp){
    this.lettersChangedFlag = true;
    this.initFrame();
    this.initBaseData(data, globalData, comp);
    this.textProperty = new TextProperty(this, data.t, this.dynamicProperties);
    this.textAnimator = new TextAnimatorProperty(data.t, this.renderType, this);
    this.initTransform(data, globalData, comp);
    this.initHierarchy();
    this.initRenderable();
    this.initRendererElement();
    this.createContainerElements();
    this.createRenderableComponents();
    this.createContent();
    this.hide();
    this.textAnimator.searchProperties(this.dynamicProperties);
};

ITextElement.prototype.prepareFrame = function(num) {
    this._mdf = false;
    this.prepareRenderableFrame(num);
    this.prepareProperties(num, this.isInRange);
    if(this.textProperty._mdf || this.textProperty._isFirstFrame) {
        this.buildNewText();
        this.textProperty._isFirstFrame = false;
        this.textProperty._mdf = false;
    }
};

ITextElement.prototype.createPathShape = function(matrixHelper, shapes) {
    var j,jLen = shapes.length;
    var k, kLen, pathNodes;
    var shapeStr = '';
    for(j=0;j<jLen;j+=1){
        pathNodes = shapes[j].ks.k;
        shapeStr += buildShapeString(pathNodes, pathNodes.i.length, true, matrixHelper);
    }
    return shapeStr;
};

ITextElement.prototype.updateDocumentData = function(newData, index) {
    this.textProperty.updateDocumentData(newData, index);
};

ITextElement.prototype.canResizeFont = function(_canResize) {
    this.textProperty.canResizeFont(_canResize);
};

ITextElement.prototype.setMinimumFontSize = function(_fontSize) {
    this.textProperty.setMinimumFontSize(_fontSize);
};

ITextElement.prototype.applyTextPropertiesToMatrix = function(documentData, matrixHelper, lineNumber, xPos, yPos) {
    if(documentData.ps){
        matrixHelper.translate(documentData.ps[0],documentData.ps[1] + documentData.ascent,0);
    }
    matrixHelper.translate(0,-documentData.ls,0);
    switch(documentData.j){
        case 1:
            matrixHelper.translate(documentData.justifyOffset + (documentData.boxWidth - documentData.lineWidths[lineNumber]),0,0);
            break;
        case 2:
            matrixHelper.translate(documentData.justifyOffset + (documentData.boxWidth - documentData.lineWidths[lineNumber] )/2,0,0);
            break;
    }
    matrixHelper.translate(xPos, yPos, 0);
};


ITextElement.prototype.buildColor = function(colorData) {
    return 'rgb(' + Math.round(colorData[0]*255) + ',' + Math.round(colorData[1]*255) + ',' + Math.round(colorData[2]*255) + ')';
};

ITextElement.prototype.emptyProp = new LetterProps();

ITextElement.prototype.destroy = function(){

};
function ICompElement(){}

extendPrototype([BaseElement, TransformElement, HierarchyElement, FrameElement, RenderableDOMElement], ICompElement);

ICompElement.prototype.initElement = function(data,globalData,comp) {
    this.initFrame();
    this.initBaseData(data, globalData, comp);
    this.initTransform(data, globalData, comp);
    this.initRenderable();
    this.initHierarchy();
    this.initRendererElement();
    this.createContainerElements();
    this.createRenderableComponents();
    if(this.data.xt || !globalData.progressiveLoad){
        this.buildAllItems();
    }
    this.hide();
};

/*ICompElement.prototype.hide = function(){
    if(!this.hidden){
        this.hideElement();
        var i,len = this.elements.length;
        for( i = 0; i < len; i+=1 ){
            if(this.elements[i]){
                this.elements[i].hide();
            }
        }
    }
};*/

ICompElement.prototype.prepareFrame = function(num){
    this._mdf = false;
    this.prepareRenderableFrame(num);
    this.prepareProperties(num, this.isInRange);
    if(!this.isInRange && !this.data.xt){
        return;
    }

    if (!this.tm._placeholder) {
        var timeRemapped = this.tm.v;
        if(timeRemapped === this.data.op){
            timeRemapped = this.data.op - 1;
        }
        this.renderedFrame = timeRemapped;
    } else {
        this.renderedFrame = num/this.data.sr;
    }
    var i,len = this.elements.length;
    if(!this.completeLayers){
        this.checkLayers(this.renderedFrame);
    }
    //This iteration needs to be backwards because of how expressions connect between each other
    for( i = len - 1; i >= 0; i -= 1 ){
        if(this.completeLayers || this.elements[i]){
            this.elements[i].prepareFrame(this.renderedFrame - this.layers[i].st);
            if(this.elements[i]._mdf) {
                this._mdf = true;
            }
        }
    }
};

ICompElement.prototype.renderInnerContent = function() {
    var i,len = this.layers.length;
    for( i = 0; i < len; i += 1 ){
        if(this.completeLayers || this.elements[i]){
            this.elements[i].renderFrame();
        }
    }
};

ICompElement.prototype.setElements = function(elems){
    this.elements = elems;
};

ICompElement.prototype.getElements = function(){
    return this.elements;
};

ICompElement.prototype.destroyElements = function(){
    var i,len = this.layers.length;
    for( i = 0; i < len; i+=1 ){
        if(this.elements[i]){
            this.elements[i].destroy();
        }
    }
};

ICompElement.prototype.destroy = function(){
    this.destroyElements();
    this.destroyBaseElement();
};

function IImageElement(data,globalData,comp){
    this.assetData = globalData.getAssetData(data.refId);
    this.initElement(data,globalData,comp);
    this.sourceRect = {top:0,left:0,width:this.assetData.w,height:this.assetData.h};
}

extendPrototype([BaseElement,TransformElement,SVGBaseElement,HierarchyElement,FrameElement,RenderableDOMElement], IImageElement);

IImageElement.prototype.createContent = function(){

    var assetPath = this.globalData.getAssetsPath(this.assetData);

    this.innerElem = createNS('image');
    this.innerElem.setAttribute('width',this.assetData.w+"px");
    this.innerElem.setAttribute('height',this.assetData.h+"px");
    this.innerElem.setAttribute('preserveAspectRatio',this.assetData.pr || this.globalData.renderConfig.imagePreserveAspectRatio);
    this.innerElem.setAttributeNS('http://www.w3.org/1999/xlink','href',assetPath);

    this.layerElement.appendChild(this.innerElem);
};

IImageElement.prototype.sourceRectAtTime = function() {
	return this.sourceRect;
}
function ISolidElement(data,globalData,comp){
    this.initElement(data,globalData,comp);
}
extendPrototype([IImageElement], ISolidElement);

ISolidElement.prototype.createContent = function(){

    var rect = createNS('rect');
    ////rect.style.width = this.data.sw;
    ////rect.style.height = this.data.sh;
    ////rect.style.fill = this.data.sc;
    rect.setAttribute('width',this.data.sw);
    rect.setAttribute('height',this.data.sh);
    rect.setAttribute('fill',this.data.sc);
    this.layerElement.appendChild(rect);
};
function SVGCompElement(data,globalData,comp){
    this.layers = data.layers;
    this.supports3d = true;
    this.completeLayers = false;
    this.pendingElements = [];
    this.elements = this.layers ? createSizedArray(this.layers.length) : [];
    //this.layerElement = createNS('g');
    this.initElement(data,globalData,comp);
    this.tm = data.tm ? PropertyFactory.getProp(this,data.tm,0,globalData.frameRate,this) : {_placeholder:true};
}

extendPrototype([SVGRenderer, ICompElement, SVGBaseElement], SVGCompElement);
function SVGTextElement(data,globalData,comp){
    this.textSpans = [];
    this.renderType = 'svg';
    this.initElement(data,globalData,comp);
}

extendPrototype([BaseElement,TransformElement,SVGBaseElement,HierarchyElement,FrameElement,RenderableDOMElement,ITextElement], SVGTextElement);

SVGTextElement.prototype.createContent = function(){

    if (this.data.singleShape && !this.globalData.fontManager.chars) {
        this.textContainer = createNS('text');
    }
};

SVGTextElement.prototype.buildTextContents = function(textArray) {
    var i = 0, len = textArray.length;
    var textContents = [], currentTextContent = '';
    while (i < len) {
        if(textArray[i] === String.fromCharCode(13) || textArray[i] === String.fromCharCode(3)) {
            textContents.push(currentTextContent);
            currentTextContent = '';
        } else {
            currentTextContent += textArray[i];
        }
        i += 1;
    }
    textContents.push(currentTextContent);
    return textContents;
}

SVGTextElement.prototype.buildNewText = function(){
    var i, len;

    var documentData = this.textProperty.currentData;
    this.renderedLetters = createSizedArray(documentData ? documentData.l.length : 0);
    if(documentData.fc) {
        this.layerElement.setAttribute('fill', this.buildColor(documentData.fc));
    }else{
        this.layerElement.setAttribute('fill', 'rgba(0,0,0,0)');
    }
    if(documentData.sc){
        this.layerElement.setAttribute('stroke', this.buildColor(documentData.sc));
        this.layerElement.setAttribute('stroke-width', documentData.sw);
    }
    this.layerElement.setAttribute('font-size', documentData.finalSize);
    var fontData = this.globalData.fontManager.getFontByName(documentData.f);
    if(fontData.fClass){
        this.layerElement.setAttribute('class',fontData.fClass);
    } else {
        this.layerElement.setAttribute('font-family', fontData.fFamily);
        var fWeight = documentData.fWeight, fStyle = documentData.fStyle;
        this.layerElement.setAttribute('font-style', fStyle);
        this.layerElement.setAttribute('font-weight', fWeight);
    }
    this.layerElement.setAttribute('arial-label', documentData.t);

    var letters = documentData.l || [];
    var usesGlyphs = !!this.globalData.fontManager.chars;
    len = letters.length;

    var tSpan;
    var matrixHelper = this.mHelper;
    var shapes, shapeStr = '', singleShape = this.data.singleShape;
    var xPos = 0, yPos = 0, firstLine = true;
    var trackingOffset = documentData.tr/1000*documentData.finalSize;
    if(singleShape && !usesGlyphs && !documentData.sz) {
        var tElement = this.textContainer;
        var justify = 'start';
        switch(documentData.j) {
            case 1:
                justify = 'end';
                break;
            case 2:
                justify = 'middle';
                break;
        }
        tElement.setAttribute('text-anchor',justify);
        tElement.setAttribute('letter-spacing',trackingOffset);
        var textContent = this.buildTextContents(documentData.finalText);
        len = textContent.length;
        yPos = documentData.ps ? documentData.ps[1] + documentData.ascent : 0;
        for ( i = 0; i < len; i += 1) {
            tSpan = this.textSpans[i] || createNS('tspan');
            tSpan.textContent = textContent[i];
            tSpan.setAttribute('x', 0);
            tSpan.setAttribute('y', yPos);
            tSpan.style.display = 'inherit';
            tElement.appendChild(tSpan);
            this.textSpans[i] = tSpan;
            yPos += documentData.finalLineHeight;
        }

        this.layerElement.appendChild(tElement);
    } else {
        var cachedSpansLength = this.textSpans.length;
        var shapeData, charData;
        for (i = 0; i < len; i += 1) {
            if(!usesGlyphs || !singleShape || i === 0){
                tSpan = cachedSpansLength > i ? this.textSpans[i] : createNS(usesGlyphs?'path':'text');
                if (cachedSpansLength <= i) {
                    tSpan.setAttribute('stroke-linecap', 'butt');
                    tSpan.setAttribute('stroke-linejoin','round');
                    tSpan.setAttribute('stroke-miterlimit','4');
                    this.textSpans[i] = tSpan;
                    this.layerElement.appendChild(tSpan);
                }
                tSpan.style.display = 'inherit';
            }

            matrixHelper.reset();
            matrixHelper.scale(documentData.finalSize / 100, documentData.finalSize / 100);
            if (singleShape) {
                if(letters[i].n) {
                    xPos = -trackingOffset;
                    yPos += documentData.yOffset;
                    yPos += firstLine ? 1 : 0;
                    firstLine = false;
                }
                this.applyTextPropertiesToMatrix(documentData, matrixHelper, letters[i].line, xPos, yPos);
                xPos += letters[i].l || 0;
                //xPos += letters[i].val === ' ' ? 0 : trackingOffset;
                xPos += trackingOffset;
            }
            if(usesGlyphs) {
                charData = this.globalData.fontManager.getCharData(documentData.finalText[i], fontData.fStyle, this.globalData.fontManager.getFontByName(documentData.f).fFamily);
                shapeData = charData && charData.data || {};
                shapes = shapeData.shapes ? shapeData.shapes[0].it : [];
                if(!singleShape){
                    tSpan.setAttribute('d',this.createPathShape(matrixHelper,shapes));
                } else {
                    shapeStr += this.createPathShape(matrixHelper,shapes);
                }
            } else {
                if(singleShape) {
                    tSpan.setAttribute("transform", "translate(" + matrixHelper.props[12] + "," + matrixHelper.props[13] + ")");
                }
                tSpan.textContent = letters[i].val;
                tSpan.setAttributeNS("http://www.w3.org/XML/1998/namespace", "xml:space","preserve");
            }
            //
        }
        if (singleShape && tSpan) {
            tSpan.setAttribute('d',shapeStr);
        }
    }
    while (i < this.textSpans.length){
        this.textSpans[i].style.display = 'none';
        i += 1;
    }

    this._sizeChanged = true;
};

SVGTextElement.prototype.sourceRectAtTime = function(time){
    this.prepareFrame(this.comp.renderedFrame - this.data.st);
    this.renderInnerContent();
    if(this._sizeChanged){
        this._sizeChanged = false;
        var textBox = this.layerElement.getBBox();
        this.bbox = {
            top: textBox.y,
            left: textBox.x,
            width: textBox.width,
            height: textBox.height
        };
    }
    return this.bbox;
};

SVGTextElement.prototype.renderInnerContent = function(){

    if(!this.data.singleShape){
        this.textAnimator.getMeasures(this.textProperty.currentData, this.lettersChangedFlag);
        if(this.lettersChangedFlag || this.textAnimator.lettersChangedFlag){
            this._sizeChanged = true;
            var  i,len;
            var renderedLetters = this.textAnimator.renderedLetters;

            var letters = this.textProperty.currentData.l;

            len = letters.length;
            var renderedLetter, textSpan;
            for(i=0;i<len;i+=1){
                if(letters[i].n){
                    continue;
                }
                renderedLetter = renderedLetters[i];
                textSpan = this.textSpans[i];
                if(renderedLetter._mdf.m) {
                    textSpan.setAttribute('transform',renderedLetter.m);
                }
                if(renderedLetter._mdf.o) {
                    textSpan.setAttribute('opacity',renderedLetter.o);
                }
                if(renderedLetter._mdf.sw){
                    textSpan.setAttribute('stroke-width',renderedLetter.sw);
                }
                if(renderedLetter._mdf.sc){
                    textSpan.setAttribute('stroke',renderedLetter.sc);
                }
                if(renderedLetter._mdf.fc){
                    textSpan.setAttribute('fill',renderedLetter.fc);
                }
            }
        }
    }
};
function SVGShapeElement(data,globalData,comp){
    //List of drawable elements
    this.shapes = [];
    // Full shape data
    this.shapesData = data.shapes;
    //List of styles that will be applied to shapes
    this.stylesList = [];
    //List of modifiers that will be applied to shapes
    this.shapeModifiers = [];
    //List of items in shape tree
    this.itemsData = [];
    //List of items in previous shape tree
    this.processedElements = [];
    // List of animated components
    this.animatedContents = [];
    this.initElement(data,globalData,comp);
    //Moving any property that doesn't get too much access after initialization because of v8 way of handling more than 10 properties.
    // List of elements that have been created
    this.prevViewData = [];
    //Moving any property that doesn't get too much access after initialization because of v8 way of handling more than 10 properties.
}

extendPrototype([BaseElement,TransformElement,SVGBaseElement,IShapeElement,HierarchyElement,FrameElement,RenderableDOMElement], SVGShapeElement);

SVGShapeElement.prototype.initSecondaryElement = function() {
};

SVGShapeElement.prototype.identityMatrix = new Matrix();

SVGShapeElement.prototype.buildExpressionInterface = function(){};

SVGShapeElement.prototype.createContent = function(){
    this.searchShapes(this.shapesData,this.itemsData,this.prevViewData,this.layerElement, 0, [], true);
    this.filterUniqueShapes();
};

/*
This method searches for multiple shapes that affect a single element and one of them is animated
*/
SVGShapeElement.prototype.filterUniqueShapes = function(){
    var i, len = this.shapes.length, shape;
    var j, jLen = this.stylesList.length;
    var style, count = 0;
    var tempShapes = [];
    var areAnimated = false;
    for(j = 0; j < jLen; j += 1) {
        style = this.stylesList[j];
        areAnimated = false;
        tempShapes.length = 0;
        for(i = 0; i < len; i += 1) {
            shape = this.shapes[i];
            if(shape.styles.indexOf(style) !== -1) {
                tempShapes.push(shape);
                areAnimated = shape._isAnimated || areAnimated;
            }
        }
        if(tempShapes.length > 1 && areAnimated) {
            this.setShapesAsAnimated(tempShapes);
        }
    }
}

SVGShapeElement.prototype.setShapesAsAnimated = function(shapes){
    var i, len = shapes.length;
    for(i = 0; i < len; i += 1) {
        shapes[i].setAsAnimated();
    }
}

SVGShapeElement.prototype.createStyleElement = function(data, level){
    //TODO: prevent drawing of hidden styles
    var elementData;
    var styleOb = new SVGStyleData(data, level);

    var pathElement = styleOb.pElem;
    if(data.ty === 'st') {
        elementData = new SVGStrokeStyleData(this, data, styleOb);
    } else if(data.ty === 'fl') {
        elementData = new SVGFillStyleData(this, data, styleOb);
    } else if(data.ty === 'gf' || data.ty === 'gs') {
        var gradientConstructor = data.ty === 'gf' ? SVGGradientFillStyleData : SVGGradientStrokeStyleData;
        elementData = new gradientConstructor(this, data, styleOb);
        this.globalData.defs.appendChild(elementData.gf);
        if (elementData.maskId) {
            this.globalData.defs.appendChild(elementData.ms);
            this.globalData.defs.appendChild(elementData.of);
            pathElement.setAttribute('mask','url(' + locationHref + '#' + elementData.maskId + ')');
        }
    }

    if(data.ty === 'st' || data.ty === 'gs') {
        pathElement.setAttribute('stroke-linecap', this.lcEnum[data.lc] || 'round');
        pathElement.setAttribute('stroke-linejoin',this.ljEnum[data.lj] || 'round');
        pathElement.setAttribute('fill-opacity','0');
        if(data.lj === 1) {
            pathElement.setAttribute('stroke-miterlimit',data.ml);
        }
    }

    if(data.r === 2) {
        pathElement.setAttribute('fill-rule', 'evenodd');
    }

    if(data.ln){
        pathElement.setAttribute('id',data.ln);
    }
    if(data.cl){
        pathElement.setAttribute('class',data.cl);
    }
    if(data.bm){
        pathElement.style['mix-blend-mode'] = getBlendMode(data.bm);
    }
    this.stylesList.push(styleOb);
    this.addToAnimatedContents(data, elementData);
    return elementData;
};

SVGShapeElement.prototype.createGroupElement = function(data) {
    var elementData = new ShapeGroupData();
    if(data.ln){
        elementData.gr.setAttribute('id',data.ln);
    }
    if(data.cl){
        elementData.gr.setAttribute('class',data.cl);
    }
    if(data.bm){
        elementData.gr.style['mix-blend-mode'] = getBlendMode(data.bm);
    }
    return elementData;
};

SVGShapeElement.prototype.createTransformElement = function(data, container) {
    var transformProperty = TransformPropertyFactory.getTransformProperty(this,data,this);
    var elementData = new SVGTransformData(transformProperty, transformProperty.o, container);
    this.addToAnimatedContents(data, elementData);
    return elementData;
};

SVGShapeElement.prototype.createShapeElement = function(data, ownTransformers, level) {
    var ty = 4;
    if(data.ty === 'rc'){
        ty = 5;
    }else if(data.ty === 'el'){
        ty = 6;
    }else if(data.ty === 'sr'){
        ty = 7;
    }
    var shapeProperty = ShapePropertyFactory.getShapeProp(this,data,ty,this);
    var elementData = new SVGShapeData(ownTransformers, level, shapeProperty);
    this.shapes.push(elementData);
    this.addShapeToModifiers(elementData);
    this.addToAnimatedContents(data, elementData);
    return elementData;
};

SVGShapeElement.prototype.addToAnimatedContents = function(data, element) {
    var i = 0, len = this.animatedContents.length;
    while(i < len) {
        if(this.animatedContents[i].element === element) {
            return;
        }
        i += 1;
    }
    this.animatedContents.push({
        fn: SVGElementsRenderer.createRenderFunction(data),
        element: element,
        data: data
    });
};

SVGShapeElement.prototype.setElementStyles = function(elementData){
    var arr = elementData.styles;
    var j, jLen = this.stylesList.length;
    for (j = 0; j < jLen; j += 1) {
        if (!this.stylesList[j].closed) {
            arr.push(this.stylesList[j]);
        }
    }
};

SVGShapeElement.prototype.reloadShapes = function(){
    this._isFirstFrame = true;
    var i, len = this.itemsData.length;
    for( i = 0; i < len; i += 1) {
        this.prevViewData[i] = this.itemsData[i];
    }
    this.searchShapes(this.shapesData,this.itemsData,this.prevViewData,this.layerElement, 0, [], true);
    this.filterUniqueShapes();
    len = this.dynamicProperties.length;
    for(i = 0; i < len; i += 1) {
        this.dynamicProperties[i].getValue();
    }
    this.renderModifiers();
};

SVGShapeElement.prototype.searchShapes = function(arr,itemsData,prevViewData,container, level, transformers, render){
    var ownTransformers = [].concat(transformers);
    var i, len = arr.length - 1;
    var j, jLen;
    var ownStyles = [], ownModifiers = [], styleOb, currentTransform, modifier, processedPos;
    for(i=len;i>=0;i-=1){
        processedPos = this.searchProcessedElement(arr[i]);
        if(!processedPos){
            arr[i]._render = render;
        } else {
            itemsData[i] = prevViewData[processedPos - 1];
        }
        if(arr[i].ty == 'fl' || arr[i].ty == 'st' || arr[i].ty == 'gf' || arr[i].ty == 'gs'){
            if(!processedPos){
                itemsData[i] = this.createStyleElement(arr[i], level);
            } else {
                itemsData[i].style.closed = false;
            }
            if(arr[i]._render){
                container.appendChild(itemsData[i].style.pElem);
            }
            ownStyles.push(itemsData[i].style);
        }else if(arr[i].ty == 'gr'){
            if(!processedPos){
                itemsData[i] = this.createGroupElement(arr[i]);
            } else {
                jLen = itemsData[i].it.length;
                for(j=0;j<jLen;j+=1){
                    itemsData[i].prevViewData[j] = itemsData[i].it[j];
                }
            }
            this.searchShapes(arr[i].it,itemsData[i].it,itemsData[i].prevViewData,itemsData[i].gr, level + 1, ownTransformers, render);
            if(arr[i]._render){
                container.appendChild(itemsData[i].gr);
            }
        }else if(arr[i].ty == 'tr'){
            if(!processedPos){
                itemsData[i] = this.createTransformElement(arr[i], container);
            }
            currentTransform = itemsData[i].transform;
            ownTransformers.push(currentTransform);
        }else if(arr[i].ty == 'sh' || arr[i].ty == 'rc' || arr[i].ty == 'el' || arr[i].ty == 'sr'){
            if(!processedPos){
                itemsData[i] = this.createShapeElement(arr[i], ownTransformers, level);
            }
            this.setElementStyles(itemsData[i]);

        }else if(arr[i].ty == 'tm' || arr[i].ty == 'rd' || arr[i].ty == 'ms'){
            if(!processedPos){
                modifier = ShapeModifiers.getModifier(arr[i].ty);
                modifier.init(this,arr[i]);
                itemsData[i] = modifier;
                this.shapeModifiers.push(modifier);
            } else {
                modifier = itemsData[i];
                modifier.closed = false;
            }
            ownModifiers.push(modifier);
        }else if(arr[i].ty == 'rp'){
            if(!processedPos){
                modifier = ShapeModifiers.getModifier(arr[i].ty);
                itemsData[i] = modifier;
                modifier.init(this,arr,i,itemsData);
                this.shapeModifiers.push(modifier);
                render = false;
            }else{
                modifier = itemsData[i];
                modifier.closed = true;
            }
            ownModifiers.push(modifier);
        }
        this.addProcessedElement(arr[i], i + 1);
    }
    len = ownStyles.length;
    for(i=0;i<len;i+=1){
        ownStyles[i].closed = true;
    }
    len = ownModifiers.length;
    for(i=0;i<len;i+=1){
        ownModifiers[i].closed = true;
    }
};

SVGShapeElement.prototype.renderInnerContent = function() {
    this.renderModifiers();
    var i, len = this.stylesList.length;
    for(i=0;i<len;i+=1){
        this.stylesList[i].reset();
    }
    this.renderShape();

    for (i = 0; i < len; i += 1) {
        if (this.stylesList[i]._mdf || this._isFirstFrame) {
            if(this.stylesList[i].msElem){
                this.stylesList[i].msElem.setAttribute('d', this.stylesList[i].d);
                //Adding M0 0 fixes same mask bug on all browsers
                this.stylesList[i].d = 'M0 0' + this.stylesList[i].d;
            }
            this.stylesList[i].pElem.setAttribute('d', this.stylesList[i].d || 'M0 0');
        }
    }
};

SVGShapeElement.prototype.renderShape = function() {
    var i, len = this.animatedContents.length;
    var animatedContent;
    for(i = 0; i < len; i += 1) {
        animatedContent = this.animatedContents[i];
        if((this._isFirstFrame || animatedContent.element._isAnimated) && animatedContent.data !== true) {
            animatedContent.fn(animatedContent.data, animatedContent.element, this._isFirstFrame);
        }
    }
}

SVGShapeElement.prototype.destroy = function(){
    this.destroyBaseElement();
    this.shapesData = null;
    this.itemsData = null;
};

function SVGTintFilter(filter, filterManager){
    this.filterManager = filterManager;
    var feColorMatrix = createNS('feColorMatrix');
    feColorMatrix.setAttribute('type','matrix');
    feColorMatrix.setAttribute('color-interpolation-filters','linearRGB');
    feColorMatrix.setAttribute('values','0.3333 0.3333 0.3333 0 0 0.3333 0.3333 0.3333 0 0 0.3333 0.3333 0.3333 0 0 0 0 0 1 0');
    feColorMatrix.setAttribute('result','f1');
    filter.appendChild(feColorMatrix);
    feColorMatrix = createNS('feColorMatrix');
    feColorMatrix.setAttribute('type','matrix');
    feColorMatrix.setAttribute('color-interpolation-filters','sRGB');
    feColorMatrix.setAttribute('values','1 0 0 0 0 0 1 0 0 0 0 0 1 0 0 0 0 0 1 0');
    feColorMatrix.setAttribute('result','f2');
    filter.appendChild(feColorMatrix);
    this.matrixFilter = feColorMatrix;
    if(filterManager.effectElements[2].p.v !== 100 || filterManager.effectElements[2].p.k){
        var feMerge = createNS('feMerge');
        filter.appendChild(feMerge);
        var feMergeNode;
        feMergeNode = createNS('feMergeNode');
        feMergeNode.setAttribute('in','SourceGraphic');
        feMerge.appendChild(feMergeNode);
        feMergeNode = createNS('feMergeNode');
        feMergeNode.setAttribute('in','f2');
        feMerge.appendChild(feMergeNode);
    }
}

SVGTintFilter.prototype.renderFrame = function(forceRender){
    if(forceRender || this.filterManager._mdf){
        var colorBlack = this.filterManager.effectElements[0].p.v;
        var colorWhite = this.filterManager.effectElements[1].p.v;
        var opacity = this.filterManager.effectElements[2].p.v/100;
        this.matrixFilter.setAttribute('values',(colorWhite[0]- colorBlack[0])+' 0 0 0 '+ colorBlack[0] +' '+ (colorWhite[1]- colorBlack[1]) +' 0 0 0 '+ colorBlack[1] +' '+ (colorWhite[2]- colorBlack[2]) +' 0 0 0 '+ colorBlack[2] +' 0 0 0 ' + opacity + ' 0');
    }
};
function SVGFillFilter(filter, filterManager){
    this.filterManager = filterManager;
    var feColorMatrix = createNS('feColorMatrix');
    feColorMatrix.setAttribute('type','matrix');
    feColorMatrix.setAttribute('color-interpolation-filters','sRGB');
    feColorMatrix.setAttribute('values','1 0 0 0 0 0 1 0 0 0 0 0 1 0 0 0 0 0 1 0');
    filter.appendChild(feColorMatrix);
    this.matrixFilter = feColorMatrix;
}
SVGFillFilter.prototype.renderFrame = function(forceRender){
    if(forceRender || this.filterManager._mdf){
        var color = this.filterManager.effectElements[2].p.v;
        var opacity = this.filterManager.effectElements[6].p.v;
        this.matrixFilter.setAttribute('values','0 0 0 0 '+color[0]+' 0 0 0 0 '+color[1]+' 0 0 0 0 '+color[2]+' 0 0 0 '+opacity+' 0');
    }
};
function SVGStrokeEffect(elem, filterManager){
    this.initialized = false;
    this.filterManager = filterManager;
    this.elem = elem;
    this.paths = [];
}

SVGStrokeEffect.prototype.initialize = function(){

    var elemChildren = this.elem.layerElement.children || this.elem.layerElement.childNodes;
    var path,groupPath, i, len;
    if(this.filterManager.effectElements[1].p.v === 1){
        len = this.elem.maskManager.masksProperties.length;
        i = 0;
    } else {
        i = this.filterManager.effectElements[0].p.v - 1;
        len = i + 1;
    }
    groupPath = createNS('g');
    groupPath.setAttribute('fill','none');
    groupPath.setAttribute('stroke-linecap','round');
    groupPath.setAttribute('stroke-dashoffset',1);
    for(i;i<len;i+=1){
        path = createNS('path');
        groupPath.appendChild(path);
        this.paths.push({p:path,m:i});
    }
    if(this.filterManager.effectElements[10].p.v === 3){
        var mask = createNS('mask');
        var id = createElementID();
        mask.setAttribute('id',id);
        mask.setAttribute('mask-type','alpha');
        mask.appendChild(groupPath);
        this.elem.globalData.defs.appendChild(mask);
        var g = createNS('g');
        g.setAttribute('mask','url(' + locationHref + '#'+id+')');
        while (elemChildren[0]) {
            g.appendChild(elemChildren[0]);
        }
        this.elem.layerElement.appendChild(g);
        this.masker = mask;
        groupPath.setAttribute('stroke','#fff');
    } else if(this.filterManager.effectElements[10].p.v === 1 || this.filterManager.effectElements[10].p.v === 2){
        if(this.filterManager.effectElements[10].p.v === 2){
            elemChildren = this.elem.layerElement.children || this.elem.layerElement.childNodes;
            while(elemChildren.length){
                this.elem.layerElement.removeChild(elemChildren[0]);
            }
        }
        this.elem.layerElement.appendChild(groupPath);
        this.elem.layerElement.removeAttribute('mask');
        groupPath.setAttribute('stroke','#fff');
    }
    this.initialized = true;
    this.pathMasker = groupPath;
};

SVGStrokeEffect.prototype.renderFrame = function(forceRender){
    if(!this.initialized){
        this.initialize();
    }
    var i, len = this.paths.length;
    var mask, path;
    for(i=0;i<len;i+=1){
        if(this.paths[i].m === -1) {
            continue;
        }
        mask = this.elem.maskManager.viewData[this.paths[i].m];
        path = this.paths[i].p;
        if(forceRender || this.filterManager._mdf || mask.prop._mdf){
            path.setAttribute('d',mask.lastPath);
        }
        if(forceRender || this.filterManager.effectElements[9].p._mdf || this.filterManager.effectElements[4].p._mdf || this.filterManager.effectElements[7].p._mdf || this.filterManager.effectElements[8].p._mdf || mask.prop._mdf){
            var dasharrayValue;
            if(this.filterManager.effectElements[7].p.v !== 0 || this.filterManager.effectElements[8].p.v !== 100){
                var s = Math.min(this.filterManager.effectElements[7].p.v,this.filterManager.effectElements[8].p.v)/100;
                var e = Math.max(this.filterManager.effectElements[7].p.v,this.filterManager.effectElements[8].p.v)/100;
                var l = path.getTotalLength();
                dasharrayValue = '0 0 0 ' + l*s + ' ';
                var lineLength = l*(e-s);
                var segment = 1+this.filterManager.effectElements[4].p.v*2*this.filterManager.effectElements[9].p.v/100;
                var units = Math.floor(lineLength/segment);
                var j;
                for(j=0;j<units;j+=1){
                    dasharrayValue += '1 ' + this.filterManager.effectElements[4].p.v*2*this.filterManager.effectElements[9].p.v/100 + ' ';
                }
                dasharrayValue += '0 ' + l*10 + ' 0 0';
            } else {
                dasharrayValue = '1 ' + this.filterManager.effectElements[4].p.v*2*this.filterManager.effectElements[9].p.v/100;
            }
            path.setAttribute('stroke-dasharray',dasharrayValue);
        }
    }
    if(forceRender || this.filterManager.effectElements[4].p._mdf){
        this.pathMasker.setAttribute('stroke-width',this.filterManager.effectElements[4].p.v*2);
    }

    if(forceRender || this.filterManager.effectElements[6].p._mdf){
        this.pathMasker.setAttribute('opacity',this.filterManager.effectElements[6].p.v);
    }
    if(this.filterManager.effectElements[10].p.v === 1 || this.filterManager.effectElements[10].p.v === 2){
        if(forceRender || this.filterManager.effectElements[3].p._mdf){
            var color = this.filterManager.effectElements[3].p.v;
            this.pathMasker.setAttribute('stroke','rgb('+bm_floor(color[0]*255)+','+bm_floor(color[1]*255)+','+bm_floor(color[2]*255)+')');
        }
    }
};
function SVGTritoneFilter(filter, filterManager){
    this.filterManager = filterManager;
    var feColorMatrix = createNS('feColorMatrix');
    feColorMatrix.setAttribute('type','matrix');
    feColorMatrix.setAttribute('color-interpolation-filters','linearRGB');
    feColorMatrix.setAttribute('values','0.3333 0.3333 0.3333 0 0 0.3333 0.3333 0.3333 0 0 0.3333 0.3333 0.3333 0 0 0 0 0 1 0');
    feColorMatrix.setAttribute('result','f1');
    filter.appendChild(feColorMatrix);
    var feComponentTransfer = createNS('feComponentTransfer');
    feComponentTransfer.setAttribute('color-interpolation-filters','sRGB');
    filter.appendChild(feComponentTransfer);
    this.matrixFilter = feComponentTransfer;
    var feFuncR = createNS('feFuncR');
    feFuncR.setAttribute('type','table');
    feComponentTransfer.appendChild(feFuncR);
    this.feFuncR = feFuncR;
    var feFuncG = createNS('feFuncG');
    feFuncG.setAttribute('type','table');
    feComponentTransfer.appendChild(feFuncG);
    this.feFuncG = feFuncG;
    var feFuncB = createNS('feFuncB');
    feFuncB.setAttribute('type','table');
    feComponentTransfer.appendChild(feFuncB);
    this.feFuncB = feFuncB;
}

SVGTritoneFilter.prototype.renderFrame = function(forceRender){
    if(forceRender || this.filterManager._mdf){
        var color1 = this.filterManager.effectElements[0].p.v;
        var color2 = this.filterManager.effectElements[1].p.v;
        var color3 = this.filterManager.effectElements[2].p.v;
        var tableR = color3[0] + ' ' + color2[0] + ' ' + color1[0];
        var tableG = color3[1] + ' ' + color2[1] + ' ' + color1[1];
        var tableB = color3[2] + ' ' + color2[2] + ' ' + color1[2];
        this.feFuncR.setAttribute('tableValues', tableR);
        this.feFuncG.setAttribute('tableValues', tableG);
        this.feFuncB.setAttribute('tableValues', tableB);
        //var opacity = this.filterManager.effectElements[2].p.v/100;
        //this.matrixFilter.setAttribute('values',(colorWhite[0]- colorBlack[0])+' 0 0 0 '+ colorBlack[0] +' '+ (colorWhite[1]- colorBlack[1]) +' 0 0 0 '+ colorBlack[1] +' '+ (colorWhite[2]- colorBlack[2]) +' 0 0 0 '+ colorBlack[2] +' 0 0 0 ' + opacity + ' 0');
    }
};
function SVGProLevelsFilter(filter, filterManager){
    this.filterManager = filterManager;
    var effectElements = this.filterManager.effectElements;
    var feComponentTransfer = createNS('feComponentTransfer');
    var feFuncR, feFuncG, feFuncB;

    if(effectElements[10].p.k || effectElements[10].p.v !== 0 || effectElements[11].p.k || effectElements[11].p.v !== 1 || effectElements[12].p.k || effectElements[12].p.v !== 1 || effectElements[13].p.k || effectElements[13].p.v !== 0 || effectElements[14].p.k || effectElements[14].p.v !== 1){
        this.feFuncR = this.createFeFunc('feFuncR', feComponentTransfer);
    }
    if(effectElements[17].p.k || effectElements[17].p.v !== 0 || effectElements[18].p.k || effectElements[18].p.v !== 1 || effectElements[19].p.k || effectElements[19].p.v !== 1 || effectElements[20].p.k || effectElements[20].p.v !== 0 || effectElements[21].p.k || effectElements[21].p.v !== 1){
        this.feFuncG = this.createFeFunc('feFuncG', feComponentTransfer);
    }
    if(effectElements[24].p.k || effectElements[24].p.v !== 0 || effectElements[25].p.k || effectElements[25].p.v !== 1 || effectElements[26].p.k || effectElements[26].p.v !== 1 || effectElements[27].p.k || effectElements[27].p.v !== 0 || effectElements[28].p.k || effectElements[28].p.v !== 1){
        this.feFuncB = this.createFeFunc('feFuncB', feComponentTransfer);
    }
    if(effectElements[31].p.k || effectElements[31].p.v !== 0 || effectElements[32].p.k || effectElements[32].p.v !== 1 || effectElements[33].p.k || effectElements[33].p.v !== 1 || effectElements[34].p.k || effectElements[34].p.v !== 0 || effectElements[35].p.k || effectElements[35].p.v !== 1){
        this.feFuncA = this.createFeFunc('feFuncA', feComponentTransfer);
    }

    if(this.feFuncR || this.feFuncG || this.feFuncB || this.feFuncA){
        feComponentTransfer.setAttribute('color-interpolation-filters','sRGB');
        filter.appendChild(feComponentTransfer);
        feComponentTransfer = createNS('feComponentTransfer');
    }

    if(effectElements[3].p.k || effectElements[3].p.v !== 0 || effectElements[4].p.k || effectElements[4].p.v !== 1 || effectElements[5].p.k || effectElements[5].p.v !== 1 || effectElements[6].p.k || effectElements[6].p.v !== 0 || effectElements[7].p.k || effectElements[7].p.v !== 1){

        feComponentTransfer.setAttribute('color-interpolation-filters','sRGB');
        filter.appendChild(feComponentTransfer);
        this.feFuncRComposed = this.createFeFunc('feFuncR', feComponentTransfer);
        this.feFuncGComposed = this.createFeFunc('feFuncG', feComponentTransfer);
        this.feFuncBComposed = this.createFeFunc('feFuncB', feComponentTransfer);
    }
}

SVGProLevelsFilter.prototype.createFeFunc = function(type, feComponentTransfer) {
    var feFunc = createNS(type);
    feFunc.setAttribute('type','table');
    feComponentTransfer.appendChild(feFunc);
    return feFunc;
};

SVGProLevelsFilter.prototype.getTableValue = function(inputBlack, inputWhite, gamma, outputBlack, outputWhite) {
    var cnt = 0;
    var segments = 256;
    var perc;
    var min = Math.min(inputBlack, inputWhite);
    var max = Math.max(inputBlack, inputWhite);
    var table = Array.call(null,{length:segments});
    var colorValue;
    var pos = 0;
    var outputDelta = outputWhite - outputBlack;
    var inputDelta = inputWhite - inputBlack;
    while(cnt <= 256) {
        perc = cnt/256;
        if(perc <= min){
            colorValue = inputDelta < 0 ? outputWhite : outputBlack;
        } else if(perc >= max){
            colorValue = inputDelta < 0 ? outputBlack : outputWhite;
        } else {
            colorValue = (outputBlack + outputDelta * Math.pow((perc - inputBlack) / inputDelta, 1 / gamma));
        }
        table[pos++] = colorValue;
        cnt += 256/(segments-1);
    }
    return table.join(' ');
};

SVGProLevelsFilter.prototype.renderFrame = function(forceRender){
    if(forceRender || this.filterManager._mdf){
        var val, cnt, perc, bezier;
        var effectElements = this.filterManager.effectElements;
        if(this.feFuncRComposed && (forceRender || effectElements[3].p._mdf || effectElements[4].p._mdf || effectElements[5].p._mdf || effectElements[6].p._mdf || effectElements[7].p._mdf)){
            val = this.getTableValue(effectElements[3].p.v,effectElements[4].p.v,effectElements[5].p.v,effectElements[6].p.v,effectElements[7].p.v);
            this.feFuncRComposed.setAttribute('tableValues',val);
            this.feFuncGComposed.setAttribute('tableValues',val);
            this.feFuncBComposed.setAttribute('tableValues',val);
        }


        if(this.feFuncR && (forceRender || effectElements[10].p._mdf || effectElements[11].p._mdf || effectElements[12].p._mdf || effectElements[13].p._mdf || effectElements[14].p._mdf)){
            val = this.getTableValue(effectElements[10].p.v,effectElements[11].p.v,effectElements[12].p.v,effectElements[13].p.v,effectElements[14].p.v);
            this.feFuncR.setAttribute('tableValues',val);
        }

        if(this.feFuncG && (forceRender || effectElements[17].p._mdf || effectElements[18].p._mdf || effectElements[19].p._mdf || effectElements[20].p._mdf || effectElements[21].p._mdf)){
            val = this.getTableValue(effectElements[17].p.v,effectElements[18].p.v,effectElements[19].p.v,effectElements[20].p.v,effectElements[21].p.v);
            this.feFuncG.setAttribute('tableValues',val);
        }

        if(this.feFuncB && (forceRender || effectElements[24].p._mdf || effectElements[25].p._mdf || effectElements[26].p._mdf || effectElements[27].p._mdf || effectElements[28].p._mdf)){
            val = this.getTableValue(effectElements[24].p.v,effectElements[25].p.v,effectElements[26].p.v,effectElements[27].p.v,effectElements[28].p.v);
            this.feFuncB.setAttribute('tableValues',val);
        }

        if(this.feFuncA && (forceRender || effectElements[31].p._mdf || effectElements[32].p._mdf || effectElements[33].p._mdf || effectElements[34].p._mdf || effectElements[35].p._mdf)){
            val = this.getTableValue(effectElements[31].p.v,effectElements[32].p.v,effectElements[33].p.v,effectElements[34].p.v,effectElements[35].p.v);
            this.feFuncA.setAttribute('tableValues',val);
        }

    }
};
function SVGDropShadowEffect(filter, filterManager){
    filter.setAttribute('x','-100%');
    filter.setAttribute('y','-100%');
    filter.setAttribute('width','400%');
    filter.setAttribute('height','400%');
    this.filterManager = filterManager;

    var feGaussianBlur = createNS('feGaussianBlur');
    feGaussianBlur.setAttribute('in','SourceAlpha');
    feGaussianBlur.setAttribute('result','drop_shadow_1');
    feGaussianBlur.setAttribute('stdDeviation','0');
    this.feGaussianBlur = feGaussianBlur;
    filter.appendChild(feGaussianBlur);

    var feOffset = createNS('feOffset');
    feOffset.setAttribute('dx','25');
    feOffset.setAttribute('dy','0');
    feOffset.setAttribute('in','drop_shadow_1');
    feOffset.setAttribute('result','drop_shadow_2');
    this.feOffset = feOffset;
    filter.appendChild(feOffset);
    var feFlood = createNS('feFlood');
    feFlood.setAttribute('flood-color','#00ff00');
    feFlood.setAttribute('flood-opacity','1');
    feFlood.setAttribute('result','drop_shadow_3');
    this.feFlood = feFlood;
    filter.appendChild(feFlood);

    var feComposite = createNS('feComposite');
    feComposite.setAttribute('in','drop_shadow_3');
    feComposite.setAttribute('in2','drop_shadow_2');
    feComposite.setAttribute('operator','in');
    feComposite.setAttribute('result','drop_shadow_4');
    filter.appendChild(feComposite);


    var feMerge = createNS('feMerge');
    filter.appendChild(feMerge);
    var feMergeNode;
    feMergeNode = createNS('feMergeNode');
    feMerge.appendChild(feMergeNode);
    feMergeNode = createNS('feMergeNode');
    feMergeNode.setAttribute('in','SourceGraphic');
    this.feMergeNode = feMergeNode;
    this.feMerge = feMerge;
    this.originalNodeAdded = false;
    feMerge.appendChild(feMergeNode);
}

SVGDropShadowEffect.prototype.renderFrame = function(forceRender){
    if(forceRender || this.filterManager._mdf){
        if(forceRender || this.filterManager.effectElements[4].p._mdf){
            this.feGaussianBlur.setAttribute('stdDeviation', this.filterManager.effectElements[4].p.v / 4);
        }
        if(forceRender || this.filterManager.effectElements[0].p._mdf){
            var col = this.filterManager.effectElements[0].p.v;
            this.feFlood.setAttribute('flood-color',rgbToHex(Math.round(col[0]*255),Math.round(col[1]*255),Math.round(col[2]*255)));
        }
        if(forceRender || this.filterManager.effectElements[1].p._mdf){
            this.feFlood.setAttribute('flood-opacity',this.filterManager.effectElements[1].p.v/255);
        }
        if(forceRender || this.filterManager.effectElements[2].p._mdf || this.filterManager.effectElements[3].p._mdf){
            var distance = this.filterManager.effectElements[3].p.v;
            var angle = (this.filterManager.effectElements[2].p.v - 90) * degToRads;
            var x = distance * Math.cos(angle);
            var y = distance * Math.sin(angle);
            this.feOffset.setAttribute('dx', x);
            this.feOffset.setAttribute('dy', y);
        }
        /*if(forceRender || this.filterManager.effectElements[5].p._mdf){
            if(this.filterManager.effectElements[5].p.v === 1 && this.originalNodeAdded) {
                this.feMerge.removeChild(this.feMergeNode);
                this.originalNodeAdded = false;
            } else if(this.filterManager.effectElements[5].p.v === 0 && !this.originalNodeAdded) {
                this.feMerge.appendChild(this.feMergeNode);
                this.originalNodeAdded = true;
            }
        }*/
    }
};
var _svgMatteSymbols = [];

function SVGMatte3Effect(filterElem, filterManager, elem){
    this.initialized = false;
    this.filterManager = filterManager;
    this.filterElem = filterElem;
    this.elem = elem;
    elem.matteElement = createNS('g');
    elem.matteElement.appendChild(elem.layerElement);
    elem.matteElement.appendChild(elem.transformedElement);
    elem.baseElement = elem.matteElement;
}

SVGMatte3Effect.prototype.findSymbol = function(mask) {
    var i = 0, len = _svgMatteSymbols.length;
    while(i < len) {
        if(_svgMatteSymbols[i] === mask) {
            return _svgMatteSymbols[i];
        }
        i += 1;
    }
    return null;
};

SVGMatte3Effect.prototype.replaceInParent = function(mask, symbolId) {
    var parentNode = mask.layerElement.parentNode;
    if(!parentNode) {
        return;
    }
    var children = parentNode.children;
    var i = 0, len = children.length;
    while (i < len) {
        if (children[i] === mask.layerElement) {
            break;
        }
        i += 1;
    }
    var nextChild;
    if (i <= len - 2) {
        nextChild = children[i + 1];
    }
    var useElem = createNS('use');
    useElem.setAttribute('href', '#' + symbolId);
    if(nextChild) {
        parentNode.insertBefore(useElem, nextChild);
    } else {
        parentNode.appendChild(useElem);
    }
};

SVGMatte3Effect.prototype.setElementAsMask = function(elem, mask) {
    if(!this.findSymbol(mask)) {
        var symbolId = createElementID();
        var masker = createNS('mask');
        masker.setAttribute('id', mask.layerId);
        masker.setAttribute('mask-type', 'alpha');
        _svgMatteSymbols.push(mask);
        var defs = elem.globalData.defs;
        defs.appendChild(masker);
        var symbol = createNS('symbol');
        symbol.setAttribute('id', symbolId);
        this.replaceInParent(mask, symbolId);
        symbol.appendChild(mask.layerElement);
        defs.appendChild(symbol);
        var useElem = createNS('use');
        useElem.setAttribute('href', '#' + symbolId);
        masker.appendChild(useElem);
        mask.data.hd = false;
        mask.show();
    }
    elem.setMatte(mask.layerId);
};

SVGMatte3Effect.prototype.initialize = function() {
    var ind = this.filterManager.effectElements[0].p.v;
    var elements = this.elem.comp.elements;
    var i = 0, len = elements.length;
    while (i < len) {
    	if (elements[i] && elements[i].data.ind === ind) {
    		this.setElementAsMask(this.elem, elements[i]);
    	}
    	i += 1;
    }
    this.initialized = true;
};

SVGMatte3Effect.prototype.renderFrame = function() {
	if(!this.initialized) {
		this.initialize();
	}
};
function SVGEffects(elem){
    var i, len = elem.data.ef ? elem.data.ef.length : 0;
    var filId = createElementID();
    var fil = filtersFactory.createFilter(filId);
    var count = 0;
    this.filters = [];
    var filterManager;
    for(i=0;i<len;i+=1){
        filterManager = null;
        if(elem.data.ef[i].ty === 20){
            count += 1;
            filterManager = new SVGTintFilter(fil, elem.effectsManager.effectElements[i]);
        }else if(elem.data.ef[i].ty === 21){
            count += 1;
            filterManager = new SVGFillFilter(fil, elem.effectsManager.effectElements[i]);
        }else if(elem.data.ef[i].ty === 22){
            filterManager = new SVGStrokeEffect(elem, elem.effectsManager.effectElements[i]);
        }else if(elem.data.ef[i].ty === 23){
            count += 1;
            filterManager = new SVGTritoneFilter(fil, elem.effectsManager.effectElements[i]);
        }else if(elem.data.ef[i].ty === 24){
            count += 1;
            filterManager = new SVGProLevelsFilter(fil, elem.effectsManager.effectElements[i]);
        }else if(elem.data.ef[i].ty === 25){
            count += 1;
            filterManager = new SVGDropShadowEffect(fil, elem.effectsManager.effectElements[i]);
        }else if(elem.data.ef[i].ty === 28){
            //count += 1;
            filterManager = new SVGMatte3Effect(fil, elem.effectsManager.effectElements[i], elem);
        }
        if(filterManager) {
            this.filters.push(filterManager);
        }
    }
    if(count){
        elem.globalData.defs.appendChild(fil);
        elem.layerElement.setAttribute('filter','url(' + locationHref + '#'+filId+')');
    }
    if (this.filters.length) {
        elem.addRenderableComponent(this);
    }
}

SVGEffects.prototype.renderFrame = function(_isFirstFrame){
    var i, len = this.filters.length;
    for(i=0;i<len;i+=1){
        this.filters[i].renderFrame(_isFirstFrame);
    }
};
function CVContextData() {
	this.saved = [];
    this.cArrPos = 0;
    this.cTr = new Matrix();
    this.cO = 1;
    var i, len = 15;
    this.savedOp = createTypedArray('float32', len);
    for(i=0;i<len;i+=1){
        this.saved[i] = createTypedArray('float32', 16);
    }
    this._length = len;
}

CVContextData.prototype.duplicate = function() {
	var newLength = this._length * 2;
	var currentSavedOp = this.savedOp;
    this.savedOp = createTypedArray('float32', newLength);
    this.savedOp.set(currentSavedOp);
    var i = 0;
    for(i = this._length; i < newLength; i += 1) {
        this.saved[i] = createTypedArray('float32', 16);
    }
    this._length = newLength;
};

CVContextData.prototype.reset = function() {
	this.cArrPos = 0;
	this.cTr.reset();
    this.cO = 1;
};
function CVBaseElement(){
}

CVBaseElement.prototype = {
    createElements: function(){},
    initRendererElement: function(){},
    createContainerElements: function(){
        this.canvasContext = this.globalData.canvasContext;
        this.renderableEffectsManager = new CVEffects(this);
    },
    createContent: function(){},
    setBlendMode: function(){
        var globalData = this.globalData;
        if(globalData.blendMode !== this.data.bm) {
            globalData.blendMode = this.data.bm;
            var blendModeValue = getBlendMode(this.data.bm);
            globalData.canvasContext.globalCompositeOperation = blendModeValue;
        }
    },
    createRenderableComponents: function(){
        this.maskManager = new CVMaskElement(this.data, this);
    },
    hideElement: function(){
        if (!this.hidden && (!this.isInRange || this.isTransparent)) {
            this.hidden = true;
        }
    },
    showElement: function(){
        if (this.isInRange && !this.isTransparent){
            this.hidden = false;
            this._isFirstFrame = true;
            this.maskManager._isFirstFrame = true;
        }
    },
    renderFrame: function() {
        if (this.hidden || this.data.hd) {
            return;
        }
        this.renderTransform();
        this.renderRenderable();
        this.setBlendMode();
        this.globalData.renderer.save();
        this.globalData.renderer.ctxTransform(this.finalTransform.mat.props);
        this.globalData.renderer.ctxOpacity(this.finalTransform.mProp.o.v);
        this.renderInnerContent();
        this.globalData.renderer.restore();
        if(this.maskManager.hasMasks) {
            this.globalData.renderer.restore(true);
        }
        if (this._isFirstFrame) {
            this._isFirstFrame = false;
        }
    },
    destroy: function(){
        this.canvasContext = null;
        this.data = null;
        this.globalData = null;
        this.maskManager.destroy();
    },
    mHelper: new Matrix()
};
CVBaseElement.prototype.hide = CVBaseElement.prototype.hideElement;
CVBaseElement.prototype.show = CVBaseElement.prototype.showElement;

function CVImageElement(data, globalData, comp){
    this.failed = false;
    this.assetData = globalData.getAssetData(data.refId);
    this.img = globalData.imageLoader.getImage(this.assetData);
    this.initElement(data,globalData,comp);
}
extendPrototype([BaseElement, TransformElement, CVBaseElement, HierarchyElement, FrameElement, RenderableElement], CVImageElement);

CVImageElement.prototype.initElement = SVGShapeElement.prototype.initElement;
CVImageElement.prototype.prepareFrame = IImageElement.prototype.prepareFrame;

CVImageElement.prototype.createContent = function(){

    if (this.img.width && (this.assetData.w !== this.img.width || this.assetData.h !== this.img.height)) {
        var canvas = createTag('canvas');
        canvas.width = this.assetData.w;
        canvas.height = this.assetData.h;
        var ctx = canvas.getContext('2d');

        var imgW = this.img.width;
        var imgH = this.img.height;
        var imgRel = imgW / imgH;
        var canvasRel = this.assetData.w/this.assetData.h;
        var widthCrop, heightCrop;
        var par = this.assetData.pr || this.globalData.renderConfig.imagePreserveAspectRatio;
        if((imgRel > canvasRel && par === 'xMidYMid slice') || (imgRel < canvasRel && par !== 'xMidYMid slice')) {
            heightCrop = imgH;
            widthCrop = heightCrop*canvasRel;
        } else {
            widthCrop = imgW;
            heightCrop = widthCrop/canvasRel;
        }
        ctx.drawImage(this.img,(imgW-widthCrop)/2,(imgH-heightCrop)/2,widthCrop,heightCrop,0,0,this.assetData.w,this.assetData.h);
        this.img = canvas;
    }

};

CVImageElement.prototype.renderInnerContent = function(parentMatrix){
    if (this.failed) {
        return;
    }
    this.canvasContext.drawImage(this.img, 0, 0);
};

CVImageElement.prototype.destroy = function(){
    this.img = null;
};
function CVCompElement(data, globalData, comp) {
    this.completeLayers = false;
    this.layers = data.layers;
    this.pendingElements = [];
    this.elements = createSizedArray(this.layers.length);
    this.initElement(data, globalData, comp);
    this.tm = data.tm ? PropertyFactory.getProp(this,data.tm,0,globalData.frameRate, this) : {_placeholder:true};
}

extendPrototype([CanvasRenderer, ICompElement, CVBaseElement], CVCompElement);

CVCompElement.prototype.renderInnerContent = function() {
    var i,len = this.layers.length;
    for( i = len - 1; i >= 0; i -= 1 ){
        if(this.completeLayers || this.elements[i]){
            this.elements[i].renderFrame();
        }
    }
};

CVCompElement.prototype.destroy = function(){
    var i,len = this.layers.length;
    for( i = len - 1; i >= 0; i -= 1 ){
        if(this.elements[i]) {
            this.elements[i].destroy();
        }
    }
    this.layers = null;
    this.elements = null;
};

function CVMaskElement(data,element){
    this.data = data;
    this.element = element;
    this.masksProperties = this.data.masksProperties || [];
    this.viewData = createSizedArray(this.masksProperties.length);
    var i, len = this.masksProperties.length, hasMasks = false;
    for (i = 0; i < len; i++) {
        if(this.masksProperties[i].mode !== 'n'){
            hasMasks = true;
        }
        this.viewData[i] = ShapePropertyFactory.getShapeProp(this.element,this.masksProperties[i],3);
    }
    this.hasMasks = hasMasks;
    if(hasMasks) {
        this.element.addRenderableComponent(this);
    }
}

CVMaskElement.prototype.renderFrame = function () {
    if(!this.hasMasks){
        return;
    }
    var transform = this.element.finalTransform.mat;
    var ctx = this.element.canvasContext;
    var i, len = this.masksProperties.length;
    var pt,pts,data;
    ctx.beginPath();
    for (i = 0; i < len; i++) {
        if(this.masksProperties[i].mode !== 'n'){
            if (this.masksProperties[i].inv) {
                ctx.moveTo(0, 0);
                ctx.lineTo(this.element.globalData.compSize.w, 0);
                ctx.lineTo(this.element.globalData.compSize.w, this.element.globalData.compSize.h);
                ctx.lineTo(0, this.element.globalData.compSize.h);
                ctx.lineTo(0, 0);
            }
            data = this.viewData[i].v;
            pt = transform.applyToPointArray(data.v[0][0],data.v[0][1],0);
            ctx.moveTo(pt[0], pt[1]);
            var j, jLen = data._length;
            for (j = 1; j < jLen; j++) {
                pts = transform.applyToTriplePoints(data.o[j - 1], data.i[j], data.v[j]);
                ctx.bezierCurveTo(pts[0], pts[1], pts[2], pts[3], pts[4], pts[5]);
            }
            pts = transform.applyToTriplePoints(data.o[j - 1], data.i[0], data.v[0]);
            ctx.bezierCurveTo(pts[0], pts[1], pts[2], pts[3], pts[4], pts[5]);
        }
    }
    this.element.globalData.renderer.save(true);
    ctx.clip();
};

CVMaskElement.prototype.getMaskProperty = MaskElement.prototype.getMaskProperty;

CVMaskElement.prototype.destroy = function(){
    this.element = null;
};
function CVShapeElement(data, globalData, comp) {
    this.shapes = [];
    this.shapesData = data.shapes;
    this.stylesList = [];
    this.itemsData = [];
    this.prevViewData = [];
    this.shapeModifiers = [];
    this.processedElements = [];
    this.transformsManager = new ShapeTransformManager();
    this.initElement(data, globalData, comp);
}

extendPrototype([BaseElement,TransformElement,CVBaseElement,IShapeElement,HierarchyElement,FrameElement,RenderableElement], CVShapeElement);

CVShapeElement.prototype.initElement = RenderableDOMElement.prototype.initElement;

CVShapeElement.prototype.transformHelper = {opacity:1,_opMdf:false};

CVShapeElement.prototype.dashResetter = [];

CVShapeElement.prototype.createContent = function(){
    this.searchShapes(this.shapesData,this.itemsData,this.prevViewData, true, []);
};

CVShapeElement.prototype.createStyleElement = function(data, transforms) {
    var styleElem = {
        data: data,
        type: data.ty,
        preTransforms: this.transformsManager.addTransformSequence(transforms),
        transforms: [],
        elements: [],
        closed: data.hd === true
    };
    var elementData = {};
    if(data.ty == 'fl' || data.ty == 'st'){
        elementData.c = PropertyFactory.getProp(this,data.c,1,255,this);
        if(!elementData.c.k){
            styleElem.co = 'rgb('+bm_floor(elementData.c.v[0])+','+bm_floor(elementData.c.v[1])+','+bm_floor(elementData.c.v[2])+')';
        }
    } else if (data.ty === 'gf' || data.ty === 'gs') {
        elementData.s = PropertyFactory.getProp(this,data.s,1,null,this);
        elementData.e = PropertyFactory.getProp(this,data.e,1,null,this);
        elementData.h = PropertyFactory.getProp(this,data.h||{k:0},0,0.01,this);
        elementData.a = PropertyFactory.getProp(this,data.a||{k:0},0,degToRads,this);
        elementData.g = new GradientProperty(this,data.g,this);
    }
    elementData.o = PropertyFactory.getProp(this,data.o,0,0.01,this);
    if(data.ty == 'st' || data.ty == 'gs') {
        styleElem.lc = this.lcEnum[data.lc] || 'round';
        styleElem.lj = this.ljEnum[data.lj] || 'round';
        if(data.lj == 1) {
            styleElem.ml = data.ml;
        }
        elementData.w = PropertyFactory.getProp(this,data.w,0,null,this);
        if(!elementData.w.k){
            styleElem.wi = elementData.w.v;
        }
        if(data.d){
            var d = new DashProperty(this,data.d,'canvas', this);
            elementData.d = d;
            if(!elementData.d.k){
                styleElem.da = elementData.d.dashArray;
                styleElem.do = elementData.d.dashoffset[0];
            }
        }
    } else {
        styleElem.r = data.r === 2 ? 'evenodd' : 'nonzero';
    }
    this.stylesList.push(styleElem);
    elementData.style = styleElem;
    return elementData;
};

CVShapeElement.prototype.createGroupElement = function(data) {
    var elementData = {
        it: [],
        prevViewData: []
    };
    return elementData;
};

CVShapeElement.prototype.createTransformElement = function(data) {
    var elementData = {
        transform : {
            opacity: 1,
            _opMdf:false,
            key: this.transformsManager.getNewKey(),
            op: PropertyFactory.getProp(this,data.o,0,0.01,this),
            mProps: TransformPropertyFactory.getTransformProperty(this,data,this)
        }
    };
    return elementData;
};

CVShapeElement.prototype.createShapeElement = function(data) {
    var elementData = new CVShapeData(this, data, this.stylesList, this.transformsManager);

    this.shapes.push(elementData);
    this.addShapeToModifiers(elementData);
    return elementData;
};

CVShapeElement.prototype.reloadShapes = function() {
    this._isFirstFrame = true;
    var i, len = this.itemsData.length;
    for (i = 0; i < len; i += 1) {
        this.prevViewData[i] = this.itemsData[i];
    }
    this.searchShapes(this.shapesData,this.itemsData,this.prevViewData, true, []);
    len = this.dynamicProperties.length;
    for (i = 0; i < len; i += 1) {
        this.dynamicProperties[i].getValue();
    }
    this.renderModifiers();
    this.transformsManager.processSequences(this._isFirstFrame);
};

CVShapeElement.prototype.addTransformToStyleList = function(transform) {
    var i, len = this.stylesList.length;
    for (i = 0; i < len; i += 1) {
        if(!this.stylesList[i].closed) {
            this.stylesList[i].transforms.push(transform);
        }
    }
}

CVShapeElement.prototype.removeTransformFromStyleList = function() {
    var i, len = this.stylesList.length;
    for (i = 0; i < len; i += 1) {
        if(!this.stylesList[i].closed) {
            this.stylesList[i].transforms.pop();
        }
    }
}

CVShapeElement.prototype.closeStyles = function(styles) {
    var i, len = styles.length, j, jLen;
    for (i = 0; i < len; i += 1) {
        styles[i].closed = true;
    }
}

CVShapeElement.prototype.searchShapes = function(arr,itemsData, prevViewData, shouldRender, transforms){
    var i, len = arr.length - 1;
    var j, jLen;
    var ownStyles = [], ownModifiers = [], processedPos, modifier, currentTransform;
    var ownTransforms = [].concat(transforms);
    for(i=len;i>=0;i-=1){
        processedPos = this.searchProcessedElement(arr[i]);
        if(!processedPos){
            arr[i]._shouldRender = shouldRender;
        } else {
            itemsData[i] = prevViewData[processedPos - 1];
        }
        if(arr[i].ty == 'fl' || arr[i].ty == 'st'|| arr[i].ty == 'gf'|| arr[i].ty == 'gs'){
            if(!processedPos){
                itemsData[i] = this.createStyleElement(arr[i], ownTransforms);
            } else {
                itemsData[i].style.closed = false;
            }

            ownStyles.push(itemsData[i].style);
        }else if(arr[i].ty == 'gr'){
            if(!processedPos){
                itemsData[i] = this.createGroupElement(arr[i]);
            } else {
                jLen = itemsData[i].it.length;
                for(j=0;j<jLen;j+=1){
                    itemsData[i].prevViewData[j] = itemsData[i].it[j];
                }
            }
            this.searchShapes(arr[i].it,itemsData[i].it,itemsData[i].prevViewData, shouldRender, ownTransforms);
        }else if(arr[i].ty == 'tr'){
            if(!processedPos){
                currentTransform = this.createTransformElement(arr[i]);
                itemsData[i] = currentTransform;
            }
            ownTransforms.push(itemsData[i]);
            this.addTransformToStyleList(itemsData[i]);
        }else if(arr[i].ty == 'sh' || arr[i].ty == 'rc' || arr[i].ty == 'el' || arr[i].ty == 'sr'){
            if(!processedPos){
                itemsData[i] = this.createShapeElement(arr[i]);
            }

        }else if(arr[i].ty == 'tm' || arr[i].ty == 'rd'){
            if(!processedPos){
                modifier = ShapeModifiers.getModifier(arr[i].ty);
                modifier.init(this,arr[i]);
                itemsData[i] = modifier;
                this.shapeModifiers.push(modifier);
            } else {
                modifier = itemsData[i];
                modifier.closed = false;
            }
            ownModifiers.push(modifier);
        } else if(arr[i].ty == 'rp'){
            if(!processedPos){
                modifier = ShapeModifiers.getModifier(arr[i].ty);
                itemsData[i] = modifier;
                modifier.init(this,arr,i,itemsData);
                this.shapeModifiers.push(modifier);
                shouldRender = false;
            }else{
                modifier = itemsData[i];
                modifier.closed = true;
            }
            ownModifiers.push(modifier);
        }
        this.addProcessedElement(arr[i], i + 1);
    }
    this.removeTransformFromStyleList();
    this.closeStyles(ownStyles);
    len = ownModifiers.length;
    for(i=0;i<len;i+=1){
        ownModifiers[i].closed = true;
    }
};

CVShapeElement.prototype.renderInnerContent = function() {
    this.transformHelper.opacity = 1;
    this.transformHelper._opMdf = false;
    this.renderModifiers();
    this.transformsManager.processSequences(this._isFirstFrame);
    this.renderShape(this.transformHelper,this.shapesData,this.itemsData,true);
};

CVShapeElement.prototype.renderShapeTransform = function(parentTransform, groupTransform) {
    var props, groupMatrix;
    if(parentTransform._opMdf || groupTransform.op._mdf || this._isFirstFrame) {
        groupTransform.opacity = parentTransform.opacity;
        groupTransform.opacity *= groupTransform.op.v;
        groupTransform._opMdf = true;
    }
};

CVShapeElement.prototype.drawLayer = function() {
    var i, len = this.stylesList.length;
    var j, jLen, k, kLen,elems,nodes, renderer = this.globalData.renderer, ctx = this.globalData.canvasContext, type, currentStyle;
    for(i=0;i<len;i+=1){
        currentStyle = this.stylesList[i];
        type = currentStyle.type;

        //Skipping style when
        //Stroke width equals 0
        //style should not be rendered (extra unused repeaters)
        //current opacity equals 0
        //global opacity equals 0
        if(((type === 'st' || type === 'gs') && currentStyle.wi === 0) || !currentStyle.data._shouldRender || currentStyle.coOp === 0 || this.globalData.currentGlobalAlpha === 0){
            continue;
        }
        renderer.save();
        elems = currentStyle.elements;
        if(type === 'st' || type === 'gs'){
            ctx.strokeStyle = type === 'st' ? currentStyle.co : currentStyle.grd;
            ctx.lineWidth = currentStyle.wi;
            ctx.lineCap = currentStyle.lc;
            ctx.lineJoin = currentStyle.lj;
            ctx.miterLimit = currentStyle.ml || 0;
        } else {
            ctx.fillStyle = type === 'fl' ? currentStyle.co : currentStyle.grd;
        }
        renderer.ctxOpacity(currentStyle.coOp);
        if(type !== 'st' && type !== 'gs'){
            ctx.beginPath();
        }
        renderer.ctxTransform(currentStyle.preTransforms.finalTransform.props);
        jLen = elems.length;
        for(j=0;j<jLen;j+=1){
            if(type === 'st' || type === 'gs'){
                ctx.beginPath();
                if(currentStyle.da){
                    ctx.setLineDash(currentStyle.da);
                    ctx.lineDashOffset = currentStyle.do;
                }
            }
            nodes = elems[j].trNodes;
            kLen = nodes.length;

            for(k=0;k<kLen;k+=1){
                if(nodes[k].t == 'm'){
                    ctx.moveTo(nodes[k].p[0],nodes[k].p[1]);
                }else if(nodes[k].t == 'c'){
                    ctx.bezierCurveTo(nodes[k].pts[0],nodes[k].pts[1],nodes[k].pts[2],nodes[k].pts[3],nodes[k].pts[4],nodes[k].pts[5]);
                }else{
                    ctx.closePath();
                }
            }
            if(type === 'st' || type === 'gs'){
                ctx.stroke();
                if(currentStyle.da){
                    ctx.setLineDash(this.dashResetter);
                }
            }
        }
        if(type !== 'st' && type !== 'gs'){
            ctx.fill(currentStyle.r);
        }
        renderer.restore();
    }
};

CVShapeElement.prototype.renderShape = function(parentTransform,items,data,isMain){
    var i, len = items.length - 1;
    var groupTransform;
    groupTransform = parentTransform;
    for(i=len;i>=0;i-=1){
        if(items[i].ty == 'tr'){
            groupTransform = data[i].transform;
            this.renderShapeTransform(parentTransform, groupTransform);
        }else if(items[i].ty == 'sh' || items[i].ty == 'el' || items[i].ty == 'rc' || items[i].ty == 'sr'){
            this.renderPath(items[i],data[i]);
        }else if(items[i].ty == 'fl'){
            this.renderFill(items[i],data[i],groupTransform);
        }else if(items[i].ty == 'st'){
            this.renderStroke(items[i],data[i],groupTransform);
        }else if(items[i].ty == 'gf' || items[i].ty == 'gs'){
            this.renderGradientFill(items[i],data[i],groupTransform);
        }else if(items[i].ty == 'gr'){
            this.renderShape(groupTransform,items[i].it,data[i].it);
        }else if(items[i].ty == 'tm'){
            //
        }
    }
    if(isMain){
        this.drawLayer();
    }

};

CVShapeElement.prototype.renderStyledShape = function(styledShape, shape){
    if(this._isFirstFrame || shape._mdf || styledShape.transforms._mdf) {
        var shapeNodes = styledShape.trNodes;
        var paths = shape.paths;
        var i, len, j, jLen = paths._length;
        shapeNodes.length = 0;
        var groupTransformMat = styledShape.transforms.finalTransform;
        for (j = 0; j < jLen; j += 1) {
            var pathNodes = paths.shapes[j];
            if(pathNodes && pathNodes.v){
                len = pathNodes._length;
                for (i = 1; i < len; i += 1) {
                    if (i === 1) {
                        shapeNodes.push({
                            t: 'm',
                            p: groupTransformMat.applyToPointArray(pathNodes.v[0][0], pathNodes.v[0][1], 0)
                        });
                    }
                    shapeNodes.push({
                        t: 'c',
                        pts: groupTransformMat.applyToTriplePoints(pathNodes.o[i - 1], pathNodes.i[i], pathNodes.v[i])
                    });
                }
                if (len === 1) {
                    shapeNodes.push({
                        t: 'm',
                        p: groupTransformMat.applyToPointArray(pathNodes.v[0][0], pathNodes.v[0][1], 0)
                    });
                }
                if (pathNodes.c && len) {
                    shapeNodes.push({
                        t: 'c',
                        pts: groupTransformMat.applyToTriplePoints(pathNodes.o[i - 1], pathNodes.i[0], pathNodes.v[0])
                    });
                    shapeNodes.push({
                        t: 'z'
                    });
                }
            }
        }
        styledShape.trNodes = shapeNodes;
    }
}

CVShapeElement.prototype.renderPath = function(pathData,itemData){
    if(pathData.hd !== true && pathData._shouldRender) {
        var i, len = itemData.styledShapes.length;
        for (i = 0; i < len; i += 1) {
            this.renderStyledShape(itemData.styledShapes[i], itemData.sh);
        }
    }
};

CVShapeElement.prototype.renderFill = function(styleData,itemData, groupTransform){
    var styleElem = itemData.style;

    if (itemData.c._mdf || this._isFirstFrame) {
        styleElem.co = 'rgb('
        + bm_floor(itemData.c.v[0]) + ','
        + bm_floor(itemData.c.v[1]) + ','
        + bm_floor(itemData.c.v[2]) + ')';
    }
    if (itemData.o._mdf || groupTransform._opMdf || this._isFirstFrame) {
        styleElem.coOp = itemData.o.v * groupTransform.opacity;
    }
};

CVShapeElement.prototype.renderGradientFill = function(styleData,itemData, groupTransform){
    var styleElem = itemData.style;
    if(!styleElem.grd || itemData.g._mdf || itemData.s._mdf || itemData.e._mdf || (styleData.t !== 1 && (itemData.h._mdf || itemData.a._mdf))) {
        var ctx = this.globalData.canvasContext;
        var grd;
        var pt1 = itemData.s.v, pt2 = itemData.e.v;
        if (styleData.t === 1) {
            grd = ctx.createLinearGradient(pt1[0], pt1[1], pt2[0], pt2[1]);
        } else {
            var rad = Math.sqrt(Math.pow(pt1[0] - pt2[0], 2) + Math.pow(pt1[1] - pt2[1], 2));
            var ang = Math.atan2(pt2[1] - pt1[1], pt2[0] - pt1[0]);

            var percent = itemData.h.v >= 1 ? 0.99 : itemData.h.v <= -1 ? -0.99: itemData.h.v;
            var dist = rad * percent;
            var x = Math.cos(ang + itemData.a.v) * dist + pt1[0];
            var y = Math.sin(ang + itemData.a.v) * dist + pt1[1];
            var grd = ctx.createRadialGradient(x, y, 0, pt1[0], pt1[1], rad);
        }

        var i, len = styleData.g.p;
        var cValues = itemData.g.c;
        var opacity = 1;

        for (i = 0; i < len; i += 1){
            if(itemData.g._hasOpacity && itemData.g._collapsable) {
                opacity = itemData.g.o[i*2 + 1];
            }
            grd.addColorStop(cValues[i * 4] / 100,'rgba('+ cValues[i * 4 + 1] + ',' + cValues[i * 4 + 2] + ','+cValues[i * 4 + 3] + ',' + opacity + ')');
        }
        styleElem.grd = grd;
    }
    styleElem.coOp = itemData.o.v*groupTransform.opacity;

};

CVShapeElement.prototype.renderStroke = function(styleData,itemData, groupTransform){
    var styleElem = itemData.style;
    var d = itemData.d;
    if(d && (d._mdf  || this._isFirstFrame)){
        styleElem.da = d.dashArray;
        styleElem.do = d.dashoffset[0];
    }
    if(itemData.c._mdf || this._isFirstFrame){
        styleElem.co = 'rgb('+bm_floor(itemData.c.v[0])+','+bm_floor(itemData.c.v[1])+','+bm_floor(itemData.c.v[2])+')';
    }
    if(itemData.o._mdf || groupTransform._opMdf || this._isFirstFrame){
        styleElem.coOp = itemData.o.v*groupTransform.opacity;
    }
    if(itemData.w._mdf || this._isFirstFrame){
        styleElem.wi = itemData.w.v;
    }
};


CVShapeElement.prototype.destroy = function(){
    this.shapesData = null;
    this.globalData = null;
    this.canvasContext = null;
    this.stylesList.length = 0;
    this.itemsData.length = 0;
};


function CVSolidElement(data, globalData, comp) {
    this.initElement(data,globalData,comp);
}
extendPrototype([BaseElement, TransformElement, CVBaseElement, HierarchyElement, FrameElement, RenderableElement], CVSolidElement);

CVSolidElement.prototype.initElement = SVGShapeElement.prototype.initElement;
CVSolidElement.prototype.prepareFrame = IImageElement.prototype.prepareFrame;

CVSolidElement.prototype.renderInnerContent = function() {
    var ctx = this.canvasContext;
    ctx.fillStyle = this.data.sc;
    ctx.fillRect(0, 0, this.data.sw, this.data.sh);
    //
};
function CVTextElement(data, globalData, comp){
    this.textSpans = [];
    this.yOffset = 0;
    this.fillColorAnim = false;
    this.strokeColorAnim = false;
    this.strokeWidthAnim = false;
    this.stroke = false;
    this.fill = false;
    this.justifyOffset = 0;
    this.currentRender = null;
    this.renderType = 'canvas';
    this.values = {
        fill: 'rgba(0,0,0,0)',
        stroke: 'rgba(0,0,0,0)',
        sWidth: 0,
        fValue: ''
    };
    this.initElement(data,globalData,comp);
}
extendPrototype([BaseElement,TransformElement,CVBaseElement,HierarchyElement,FrameElement,RenderableElement,ITextElement], CVTextElement);

CVTextElement.prototype.tHelper = createTag('canvas').getContext('2d');

CVTextElement.prototype.buildNewText = function(){
    var documentData = this.textProperty.currentData;
    this.renderedLetters = createSizedArray(documentData.l ? documentData.l.length : 0);

    var hasFill = false;
    if(documentData.fc) {
        hasFill = true;
        this.values.fill = this.buildColor(documentData.fc);
    }else{
        this.values.fill = 'rgba(0,0,0,0)';
    }
    this.fill = hasFill;
    var hasStroke = false;
    if(documentData.sc){
        hasStroke = true;
        this.values.stroke = this.buildColor(documentData.sc);
        this.values.sWidth = documentData.sw;
    }
    var fontData = this.globalData.fontManager.getFontByName(documentData.f);
    var i, len;
    var letters = documentData.l;
    var matrixHelper = this.mHelper;
    this.stroke = hasStroke;
    this.values.fValue = documentData.finalSize + 'px '+ this.globalData.fontManager.getFontByName(documentData.f).fFamily;
    len = documentData.finalText.length;
    //this.tHelper.font = this.values.fValue;
    var charData, shapeData, k, kLen, shapes, j, jLen, pathNodes, commands, pathArr, singleShape = this.data.singleShape;
    var trackingOffset = documentData.tr/1000*documentData.finalSize;
    var xPos = 0, yPos = 0, firstLine = true;
    var cnt = 0;
    for (i = 0; i < len; i += 1) {
        charData = this.globalData.fontManager.getCharData(documentData.finalText[i], fontData.fStyle, this.globalData.fontManager.getFontByName(documentData.f).fFamily);
        shapeData = charData && charData.data || {};
        matrixHelper.reset();
        if(singleShape && letters[i].n) {
            xPos = -trackingOffset;
            yPos += documentData.yOffset;
            yPos += firstLine ? 1 : 0;
            firstLine = false;
        }

        shapes = shapeData.shapes ? shapeData.shapes[0].it : [];
        jLen = shapes.length;
        matrixHelper.scale(documentData.finalSize/100,documentData.finalSize/100);
        if(singleShape){
            this.applyTextPropertiesToMatrix(documentData, matrixHelper, letters[i].line, xPos, yPos);
        }
        commands = createSizedArray(jLen);
        for(j=0;j<jLen;j+=1){
            kLen = shapes[j].ks.k.i.length;
            pathNodes = shapes[j].ks.k;
            pathArr = [];
            for(k=1;k<kLen;k+=1){
                if(k==1){
                    pathArr.push(matrixHelper.applyToX(pathNodes.v[0][0],pathNodes.v[0][1],0),matrixHelper.applyToY(pathNodes.v[0][0],pathNodes.v[0][1],0));
                }
                pathArr.push(matrixHelper.applyToX(pathNodes.o[k-1][0],pathNodes.o[k-1][1],0),matrixHelper.applyToY(pathNodes.o[k-1][0],pathNodes.o[k-1][1],0),matrixHelper.applyToX(pathNodes.i[k][0],pathNodes.i[k][1],0),matrixHelper.applyToY(pathNodes.i[k][0],pathNodes.i[k][1],0),matrixHelper.applyToX(pathNodes.v[k][0],pathNodes.v[k][1],0),matrixHelper.applyToY(pathNodes.v[k][0],pathNodes.v[k][1],0));
            }
            pathArr.push(matrixHelper.applyToX(pathNodes.o[k-1][0],pathNodes.o[k-1][1],0),matrixHelper.applyToY(pathNodes.o[k-1][0],pathNodes.o[k-1][1],0),matrixHelper.applyToX(pathNodes.i[0][0],pathNodes.i[0][1],0),matrixHelper.applyToY(pathNodes.i[0][0],pathNodes.i[0][1],0),matrixHelper.applyToX(pathNodes.v[0][0],pathNodes.v[0][1],0),matrixHelper.applyToY(pathNodes.v[0][0],pathNodes.v[0][1],0));
            commands[j] = pathArr;
        }
        if(singleShape){
            xPos += letters[i].l;
            xPos += trackingOffset;
        }
        if(this.textSpans[cnt]){
            this.textSpans[cnt].elem = commands;
        } else {
            this.textSpans[cnt] = {elem: commands};
        }
        cnt +=1;
    }
};

CVTextElement.prototype.renderInnerContent = function(){
    var ctx = this.canvasContext;
    var finalMat = this.finalTransform.mat.props;
    ctx.font = this.values.fValue;
    ctx.lineCap = 'butt';
    ctx.lineJoin = 'miter';
    ctx.miterLimit = 4;

    if(!this.data.singleShape){
        this.textAnimator.getMeasures(this.textProperty.currentData, this.lettersChangedFlag);
    }

    var  i,len, j, jLen, k, kLen;
    var renderedLetters = this.textAnimator.renderedLetters;

    var letters = this.textProperty.currentData.l;

    len = letters.length;
    var renderedLetter;
    var lastFill = null, lastStroke = null, lastStrokeW = null, commands, pathArr;
    for(i=0;i<len;i+=1){
        if(letters[i].n){
            continue;
        }
        renderedLetter = renderedLetters[i];
        if(renderedLetter){
            this.globalData.renderer.save();
            this.globalData.renderer.ctxTransform(renderedLetter.p);
            this.globalData.renderer.ctxOpacity(renderedLetter.o);
        }
        if(this.fill){
            if(renderedLetter && renderedLetter.fc){
                if(lastFill !== renderedLetter.fc){
                    lastFill = renderedLetter.fc;
                    ctx.fillStyle = renderedLetter.fc;
                }
            }else if(lastFill !== this.values.fill){
                lastFill = this.values.fill;
                ctx.fillStyle = this.values.fill;
            }
            commands = this.textSpans[i].elem;
            jLen = commands.length;
            this.globalData.canvasContext.beginPath();
            for(j=0;j<jLen;j+=1) {
                pathArr = commands[j];
                kLen = pathArr.length;
                this.globalData.canvasContext.moveTo(pathArr[0], pathArr[1]);
                for (k = 2; k < kLen; k += 6) {
                    this.globalData.canvasContext.bezierCurveTo(pathArr[k], pathArr[k + 1], pathArr[k + 2], pathArr[k + 3], pathArr[k + 4], pathArr[k + 5]);
                }
            }
            this.globalData.canvasContext.closePath();
            this.globalData.canvasContext.fill();
            ///ctx.fillText(this.textSpans[i].val,0,0);
        }
        if(this.stroke){
            if(renderedLetter && renderedLetter.sw){
                if(lastStrokeW !== renderedLetter.sw){
                    lastStrokeW = renderedLetter.sw;
                    ctx.lineWidth = renderedLetter.sw;
                }
            }else if(lastStrokeW !== this.values.sWidth){
                lastStrokeW = this.values.sWidth;
                ctx.lineWidth = this.values.sWidth;
            }
            if(renderedLetter && renderedLetter.sc){
                if(lastStroke !== renderedLetter.sc){
                    lastStroke = renderedLetter.sc;
                    ctx.strokeStyle = renderedLetter.sc;
                }
            }else if(lastStroke !== this.values.stroke){
                lastStroke = this.values.stroke;
                ctx.strokeStyle = this.values.stroke;
            }
            commands = this.textSpans[i].elem;
            jLen = commands.length;
            this.globalData.canvasContext.beginPath();
            for(j=0;j<jLen;j+=1) {
                pathArr = commands[j];
                kLen = pathArr.length;
                this.globalData.canvasContext.moveTo(pathArr[0], pathArr[1]);
                for (k = 2; k < kLen; k += 6) {
                    this.globalData.canvasContext.bezierCurveTo(pathArr[k], pathArr[k + 1], pathArr[k + 2], pathArr[k + 3], pathArr[k + 4], pathArr[k + 5]);
                }
            }
            this.globalData.canvasContext.closePath();
            this.globalData.canvasContext.stroke();
            ///ctx.strokeText(letters[i].val,0,0);
        }
        if(renderedLetter) {
            this.globalData.renderer.restore();
        }
    }
};
function CVEffects() {

}
CVEffects.prototype.renderFrame = function(){};
function HBaseElement(data,globalData,comp){}
HBaseElement.prototype = {
    checkBlendMode: function(){},
    initRendererElement: function(){
        this.baseElement = createTag(this.data.tg || 'div');
        if(this.data.hasMask) {
            this.svgElement = createNS('svg');
            this.layerElement = createNS('g');
            this.maskedElement = this.layerElement;
            this.svgElement.appendChild(this.layerElement);
            this.baseElement.appendChild(this.svgElement);
        } else {
            this.layerElement = this.baseElement;
        }
        styleDiv(this.baseElement);
    },
    createContainerElements: function(){
        this.renderableEffectsManager = new CVEffects(this);
        this.transformedElement = this.baseElement;
        this.maskedElement = this.layerElement;
        if (this.data.ln) {
            this.layerElement.setAttribute('id',this.data.ln);
        }
        if (this.data.cl) {
            this.layerElement.setAttribute('class', this.data.cl);
        }
        if (this.data.bm !== 0) {
            this.setBlendMode();
        }
    },
    renderElement: function() {
        if(this.finalTransform._matMdf){
            this.transformedElement.style.transform = this.transformedElement.style.webkitTransform = this.finalTransform.mat.toCSS();
        }
        if(this.finalTransform._opMdf){
            this.transformedElement.style.opacity = this.finalTransform.mProp.o.v;
        }
    },
    renderFrame: function() {
        //If it is exported as hidden (data.hd === true) no need to render
        //If it is not visible no need to render
        if (this.data.hd || this.hidden) {
            return;
        }
        this.renderTransform();
        this.renderRenderable();
        this.renderElement();
        this.renderInnerContent();
        if (this._isFirstFrame) {
            this._isFirstFrame = false;
        }
    },
    destroy: function(){
        this.layerElement = null;
        this.transformedElement = null;
        if(this.matteElement) {
            this.matteElement = null;
        }
        if(this.maskManager) {
            this.maskManager.destroy();
            this.maskManager = null;
        }
    },
    createRenderableComponents: function(){
        this.maskManager = new MaskElement(this.data, this, this.globalData);
    },
    addEffects: function(){
    },
    setMatte: function(){}
};
HBaseElement.prototype.getBaseElement = SVGBaseElement.prototype.getBaseElement;
HBaseElement.prototype.destroyBaseElement = HBaseElement.prototype.destroy;
HBaseElement.prototype.buildElementParenting = HybridRenderer.prototype.buildElementParenting;
function HSolidElement(data,globalData,comp){
    this.initElement(data,globalData,comp);
}
extendPrototype([BaseElement,TransformElement,HBaseElement,HierarchyElement,FrameElement,RenderableDOMElement], HSolidElement);

HSolidElement.prototype.createContent = function(){
    var rect;
    if(this.data.hasMask){
        rect = createNS('rect');
        rect.setAttribute('width',this.data.sw);
        rect.setAttribute('height',this.data.sh);
        rect.setAttribute('fill',this.data.sc);
        this.svgElement.setAttribute('width',this.data.sw);
        this.svgElement.setAttribute('height',this.data.sh);
    } else {
        rect = createTag('div');
        rect.style.width = this.data.sw + 'px';
        rect.style.height = this.data.sh + 'px';
        rect.style.backgroundColor = this.data.sc;
    }
    this.layerElement.appendChild(rect);
};

function HCompElement(data,globalData,comp){
    this.layers = data.layers;
    this.supports3d = !data.hasMask;
    this.completeLayers = false;
    this.pendingElements = [];
    this.elements = this.layers ? createSizedArray(this.layers.length) : [];
    this.initElement(data,globalData,comp);
    this.tm = data.tm ? PropertyFactory.getProp(this,data.tm,0,globalData.frameRate,this) : {_placeholder:true};
}

extendPrototype([HybridRenderer, ICompElement, HBaseElement], HCompElement);
HCompElement.prototype._createBaseContainerElements = HCompElement.prototype.createContainerElements;

HCompElement.prototype.createContainerElements = function(){
    this._createBaseContainerElements();
    //divElement.style.clip = 'rect(0px, '+this.data.w+'px, '+this.data.h+'px, 0px)';
    if(this.data.hasMask){
        this.svgElement.setAttribute('width',this.data.w);
        this.svgElement.setAttribute('height',this.data.h);
        this.transformedElement = this.baseElement;
    } else {
        this.transformedElement = this.layerElement;
    }
};

HCompElement.prototype.addTo3dContainer = function(elem,pos) {
    var j = 0;
    var nextElement;
    while(j<pos){
        if(this.elements[j] && this.elements[j].getBaseElement){
            nextElement = this.elements[j].getBaseElement();
        }
        j += 1;
    }
    if(nextElement){
        this.layerElement.insertBefore(elem, nextElement);
    } else {
        this.layerElement.appendChild(elem);
    }
}

function HShapeElement(data,globalData,comp){
    //List of drawable elements
    this.shapes = [];
    // Full shape data
    this.shapesData = data.shapes;
    //List of styles that will be applied to shapes
    this.stylesList = [];
    //List of modifiers that will be applied to shapes
    this.shapeModifiers = [];
    //List of items in shape tree
    this.itemsData = [];
    //List of items in previous shape tree
    this.processedElements = [];
    // List of animated components
    this.animatedContents = [];
    this.shapesContainer = createNS('g');
    this.initElement(data,globalData,comp);
    //Moving any property that doesn't get too much access after initialization because of v8 way of handling more than 10 properties.
    // List of elements that have been created
    this.prevViewData = [];
    this.currentBBox = {
        x:999999,
        y: -999999,
        h: 0,
        w: 0
    };
}
extendPrototype([BaseElement,TransformElement,HSolidElement,SVGShapeElement,HBaseElement,HierarchyElement,FrameElement,RenderableElement], HShapeElement);
HShapeElement.prototype._renderShapeFrame = HShapeElement.prototype.renderInnerContent;

HShapeElement.prototype.createContent = function(){
    var cont;
    this.baseElement.style.fontSize = 0;
    if (this.data.hasMask) {
        this.layerElement.appendChild(this.shapesContainer);
        cont = this.svgElement;
    } else {
        cont = createNS('svg');
        var size = this.comp.data ? this.comp.data : this.globalData.compSize;
        cont.setAttribute('width',size.w);
        cont.setAttribute('height',size.h);
        cont.appendChild(this.shapesContainer);
        this.layerElement.appendChild(cont);
    }

    this.searchShapes(this.shapesData,this.itemsData,this.prevViewData,this.shapesContainer,0, [], true);
    this.filterUniqueShapes();
    this.shapeCont = cont;
};

HShapeElement.prototype.getTransformedPoint = function(transformers, point) {
    var i, len = transformers.length;
    for(i = 0; i < len; i += 1) {
        point = transformers[i].mProps.v.applyToPointArray(point[0], point[1], 0);
    }
    return point;
}

HShapeElement.prototype.calculateShapeBoundingBox = function(item, boundingBox) {
    var shape = item.sh.v;
    var transformers = item.transformers;
    var i, len = shape._length, vPoint, oPoint, nextIPoint, nextVPoint, bounds;
    if (len <= 1) {
        return;
    }
    for (i = 0; i < len - 1; i += 1) {
        vPoint = this.getTransformedPoint(transformers, shape.v[i]);
        oPoint = this.getTransformedPoint(transformers, shape.o[i]);
        nextIPoint = this.getTransformedPoint(transformers, shape.i[i + 1]);
        nextVPoint = this.getTransformedPoint(transformers, shape.v[i + 1]);
        this.checkBounds(vPoint, oPoint, nextIPoint, nextVPoint, boundingBox);
    }
    if(shape.c) {
        vPoint = this.getTransformedPoint(transformers, shape.v[i]);
        oPoint = this.getTransformedPoint(transformers, shape.o[i]);
        nextIPoint = this.getTransformedPoint(transformers, shape.i[0]);
        nextVPoint = this.getTransformedPoint(transformers, shape.v[0]);
        this.checkBounds(vPoint, oPoint, nextIPoint, nextVPoint, boundingBox);
    }
}

HShapeElement.prototype.checkBounds = function(vPoint, oPoint, nextIPoint, nextVPoint, boundingBox) {
    this.getBoundsOfCurve(vPoint, oPoint, nextIPoint, nextVPoint);
    var bounds = this.shapeBoundingBox;
    boundingBox.x = bm_min(bounds.left, boundingBox.x);
    boundingBox.xMax = bm_max(bounds.right, boundingBox.xMax);
    boundingBox.y = bm_min(bounds.top, boundingBox.y);
    boundingBox.yMax = bm_max(bounds.bottom, boundingBox.yMax);
}

HShapeElement.prototype.shapeBoundingBox = {
    left:0,
    right:0,
    top:0,
    bottom:0,
}

HShapeElement.prototype.tempBoundingBox = {
    x:0,
    xMax:0,
    y:0,
    yMax:0,
    width:0,
    height:0
}

HShapeElement.prototype.getBoundsOfCurve = function(p0, p1, p2, p3) {

    var bounds = [[p0[0],p3[0]], [p0[1],p3[1]]];

    for (var a, b, c, t, b2ac, t1, t2, i = 0; i < 2; ++i) {

      b = 6 * p0[i] - 12 * p1[i] + 6 * p2[i];
      a = -3 * p0[i] + 9 * p1[i] - 9 * p2[i] + 3 * p3[i];
      c = 3 * p1[i] - 3 * p0[i];

      b = b | 0;
      a = a | 0;
      c = c | 0;

      if (a === 0) {

        if (b === 0) {
          continue;
        }

        t = -c / b;

        if (0 < t && t < 1) {
          bounds[i].push(this.calculateF(t,p0,p1,p2,p3,i));
        }
        continue;
      }

      b2ac = b * b - 4 * c * a;

      if (b2ac < 0) {
        continue;
      }

      t1 = (-b + bm_sqrt(b2ac))/(2 * a);
      if (0 < t1 && t1 < 1) bounds[i].push(this.calculateF(t1,p0,p1,p2,p3,i));

      t2 = (-b - bm_sqrt(b2ac))/(2 * a);
      if (0 < t2 && t2 < 1) bounds[i].push(this.calculateF(t2,p0,p1,p2,p3,i));

    }

    this.shapeBoundingBox.left = bm_min.apply(null, bounds[0]);
    this.shapeBoundingBox.top = bm_min.apply(null, bounds[1]);
    this.shapeBoundingBox.right = bm_max.apply(null, bounds[0]);
    this.shapeBoundingBox.bottom = bm_max.apply(null, bounds[1]);
  };

  HShapeElement.prototype.calculateF = function(t, p0, p1, p2, p3, i) {
    return bm_pow(1-t, 3) * p0[i]
        + 3 * bm_pow(1-t, 2) * t * p1[i]
        + 3 * (1-t) * bm_pow(t, 2) * p2[i]
        + bm_pow(t, 3) * p3[i];
  }

HShapeElement.prototype.calculateBoundingBox = function(itemsData, boundingBox) {
    var i, len = itemsData.length, path;
    for(i = 0; i < len; i += 1) {
        if(itemsData[i] && itemsData[i].sh) {
            this.calculateShapeBoundingBox(itemsData[i], boundingBox)
        } else if(itemsData[i] && itemsData[i].it) {
            this.calculateBoundingBox(itemsData[i].it, boundingBox)
        }
    }
}

HShapeElement.prototype.currentBoxContains = function(box) {
    return this.currentBBox.x <= box.x
    && this.currentBBox.y <= box.y
    && this.currentBBox.width + this.currentBBox.x >= box.x + box.width
    && this.currentBBox.height + this.currentBBox.y >= box.y + box.height
}

HShapeElement.prototype.renderInnerContent = function() {
    this._renderShapeFrame();

    if(!this.hidden && (this._isFirstFrame || this._mdf)) {
        var tempBoundingBox = this.tempBoundingBox;
        var max = 999999;
        tempBoundingBox.x = max;
        tempBoundingBox.xMax = -max;
        tempBoundingBox.y = max;
        tempBoundingBox.yMax = -max;
        this.calculateBoundingBox(this.itemsData, tempBoundingBox);
        tempBoundingBox.width = tempBoundingBox.xMax < tempBoundingBox.x ? 0 : tempBoundingBox.xMax - tempBoundingBox.x;
        tempBoundingBox.height = tempBoundingBox.yMax < tempBoundingBox.y ? 0 : tempBoundingBox.yMax - tempBoundingBox.y;
        //var tempBoundingBox = this.shapeCont.getBBox();
        if(this.currentBoxContains(tempBoundingBox)) {
            return;
        }
        var changed = false;
        if(this.currentBBox.w !== tempBoundingBox.width){
            this.currentBBox.w = tempBoundingBox.width;
            this.shapeCont.setAttribute('width',tempBoundingBox.width);
            changed = true;
        }
        if(this.currentBBox.h !== tempBoundingBox.height){
            this.currentBBox.h = tempBoundingBox.height;
            this.shapeCont.setAttribute('height',tempBoundingBox.height);
            changed = true;
        }
        if(changed  || this.currentBBox.x !== tempBoundingBox.x  || this.currentBBox.y !== tempBoundingBox.y){
            this.currentBBox.w = tempBoundingBox.width;
            this.currentBBox.h = tempBoundingBox.height;
            this.currentBBox.x = tempBoundingBox.x;
            this.currentBBox.y = tempBoundingBox.y;

            this.shapeCont.setAttribute('viewBox',this.currentBBox.x+' '+this.currentBBox.y+' '+this.currentBBox.w+' '+this.currentBBox.h);
            this.shapeCont.style.transform = this.shapeCont.style.webkitTransform = 'translate(' + this.currentBBox.x + 'px,' + this.currentBBox.y + 'px)';
        }
    }

};
function HTextElement(data,globalData,comp){
    this.textSpans = [];
    this.textPaths = [];
    this.currentBBox = {
        x:999999,
        y: -999999,
        h: 0,
        w: 0
    };
    this.renderType = 'svg';
    this.isMasked = false;
    this.initElement(data,globalData,comp);

}
extendPrototype([BaseElement,TransformElement,HBaseElement,HierarchyElement,FrameElement,RenderableDOMElement,ITextElement], HTextElement);

HTextElement.prototype.createContent = function(){
    this.isMasked = this.checkMasks();
    if(this.isMasked){
        this.renderType = 'svg';
        this.compW = this.comp.data.w;
        this.compH = this.comp.data.h;
        this.svgElement.setAttribute('width',this.compW);
        this.svgElement.setAttribute('height',this.compH);
        var g = createNS('g');
        this.maskedElement.appendChild(g);
        this.innerElem = g;
    } else {
        this.renderType = 'html';
        this.innerElem = this.layerElement;
    }

    this.checkParenting();

};

HTextElement.prototype.buildNewText = function(){
    var documentData = this.textProperty.currentData;
    this.renderedLetters = createSizedArray(documentData.l ? documentData.l.length : 0);
    var innerElemStyle = this.innerElem.style;
    innerElemStyle.color = innerElemStyle.fill = documentData.fc ? this.buildColor(documentData.fc) : 'rgba(0,0,0,0)';
    if(documentData.sc){
        innerElemStyle.stroke = this.buildColor(documentData.sc);
        innerElemStyle.strokeWidth = documentData.sw+'px';
    }
    var fontData = this.globalData.fontManager.getFontByName(documentData.f);
    if(!this.globalData.fontManager.chars){
        innerElemStyle.fontSize = documentData.finalSize+'px';
        innerElemStyle.lineHeight = documentData.finalSize+'px';
        if(fontData.fClass){
            this.innerElem.className = fontData.fClass;
        } else {
            innerElemStyle.fontFamily = fontData.fFamily;
            var fWeight = documentData.fWeight, fStyle = documentData.fStyle;
            innerElemStyle.fontStyle = fStyle;
            innerElemStyle.fontWeight = fWeight;
        }
    }
    var i, len;

    var letters = documentData.l;
    len = letters.length;
    var tSpan,tParent,tCont;
    var matrixHelper = this.mHelper;
    var shapes, shapeStr = '';
    var cnt = 0;
    for (i = 0;i < len ;i += 1) {
        if(this.globalData.fontManager.chars){
            if(!this.textPaths[cnt]){
                tSpan = createNS('path');
                tSpan.setAttribute('stroke-linecap', 'butt');
                tSpan.setAttribute('stroke-linejoin','round');
                tSpan.setAttribute('stroke-miterlimit','4');
            } else {
                tSpan = this.textPaths[cnt];
            }
            if(!this.isMasked){
                if(this.textSpans[cnt]){
                    tParent = this.textSpans[cnt];
                    tCont = tParent.children[0];
                } else {

                    tParent = createTag('div');
                    tCont = createNS('svg');
                    tCont.appendChild(tSpan);
                    styleDiv(tParent);
                }
            }
        }else{
            if(!this.isMasked){
                if(this.textSpans[cnt]){
                    tParent = this.textSpans[cnt];
                    tSpan = this.textPaths[cnt];
                } else {
                    tParent = createTag('span');
                    styleDiv(tParent);
                    tSpan = createTag('span');
                    styleDiv(tSpan);
                    tParent.appendChild(tSpan);
                }
            } else {
                tSpan = this.textPaths[cnt] ? this.textPaths[cnt] : createNS('text');
            }
        }
        //tSpan.setAttribute('visibility', 'hidden');
        if(this.globalData.fontManager.chars){
            var charData = this.globalData.fontManager.getCharData(documentData.finalText[i], fontData.fStyle, this.globalData.fontManager.getFontByName(documentData.f).fFamily);
            var shapeData;
            if(charData){
                shapeData = charData.data;
            } else {
                shapeData = null;
            }
            matrixHelper.reset();
            if(shapeData && shapeData.shapes){
                shapes = shapeData.shapes[0].it;
                matrixHelper.scale(documentData.finalSize/100,documentData.finalSize/100);
                shapeStr = this.createPathShape(matrixHelper,shapes);
                tSpan.setAttribute('d',shapeStr);
            }
            if(!this.isMasked){
                this.innerElem.appendChild(tParent);
                if(shapeData && shapeData.shapes){

                    //document.body.appendChild is needed to get exact measure of shape
                    document.body.appendChild(tCont);
                    var boundingBox = tCont.getBBox();
                    tCont.setAttribute('width',boundingBox.width + 2);
                    tCont.setAttribute('height',boundingBox.height + 2);
                    tCont.setAttribute('viewBox',(boundingBox.x-1)+' '+ (boundingBox.y-1)+' '+ (boundingBox.width+2)+' '+ (boundingBox.height+2));
                    tCont.style.transform = tCont.style.webkitTransform = 'translate(' + (boundingBox.x-1) + 'px,' + (boundingBox.y-1) + 'px)';

                    letters[i].yOffset = boundingBox.y-1;

                } else{
                    tCont.setAttribute('width',1);
                    tCont.setAttribute('height',1);
                }
                    tParent.appendChild(tCont);
            }else{
                this.innerElem.appendChild(tSpan);
            }
        }else{
            tSpan.textContent = letters[i].val;
            tSpan.setAttributeNS("http://www.w3.org/XML/1998/namespace", "xml:space","preserve");
            if(!this.isMasked){
                this.innerElem.appendChild(tParent);
                //
                tSpan.style.transform = tSpan.style.webkitTransform = 'translate3d(0,'+ -documentData.finalSize/1.2+'px,0)';
            } else {
                this.innerElem.appendChild(tSpan);
            }
        }
        //
        if(!this.isMasked){
            this.textSpans[cnt] = tParent;
        }else{
            this.textSpans[cnt] = tSpan;
        }
        this.textSpans[cnt].style.display = 'block';
        this.textPaths[cnt] = tSpan;
        cnt += 1;
    }
    while(cnt < this.textSpans.length){
        this.textSpans[cnt].style.display = 'none';
        cnt += 1;
    }
};

HTextElement.prototype.renderInnerContent = function() {

    if(this.data.singleShape){
        if(!this._isFirstFrame && !this.lettersChangedFlag){
            return;
        } else {
            // Todo Benchmark if using this is better than getBBox
             if(this.isMasked && this.finalTransform._matMdf){
                 this.svgElement.setAttribute('viewBox',-this.finalTransform.mProp.p.v[0]+' '+ -this.finalTransform.mProp.p.v[1]+' '+this.compW+' '+this.compH);
                this.svgElement.style.transform = this.svgElement.style.webkitTransform = 'translate(' + -this.finalTransform.mProp.p.v[0] + 'px,' + -this.finalTransform.mProp.p.v[1] + 'px)';
             }
        }
    }

    this.textAnimator.getMeasures(this.textProperty.currentData, this.lettersChangedFlag);
    if(!this.lettersChangedFlag && !this.textAnimator.lettersChangedFlag){
        return;
    }
    var  i,len, count = 0;
    var renderedLetters = this.textAnimator.renderedLetters;

    var letters = this.textProperty.currentData.l;

    len = letters.length;
    var renderedLetter, textSpan, textPath;
    for(i=0;i<len;i+=1){
        if(letters[i].n){
            count += 1;
            continue;
        }
        textSpan = this.textSpans[i];
        textPath = this.textPaths[i];
        renderedLetter = renderedLetters[count];
        count += 1;
        if(renderedLetter._mdf.m) {
            if(!this.isMasked){
                textSpan.style.transform = textSpan.style.webkitTransform = renderedLetter.m;
            }else{
                textSpan.setAttribute('transform',renderedLetter.m);
            }
        }
        ////textSpan.setAttribute('opacity',renderedLetter.o);
        textSpan.style.opacity = renderedLetter.o;
        if(renderedLetter.sw && renderedLetter._mdf.sw){
            textPath.setAttribute('stroke-width',renderedLetter.sw);
        }
        if(renderedLetter.sc && renderedLetter._mdf.sc){
```
