## Supplement File 1 for "Post Induction Hypotension prediction during general anesthesia using Machine Learning Techniques": preview-export-min.js

```
!function(e,t){"object"==typeof module&&"object"==typeof module.exports?module.exports=e.document?t(e,!0):function(e){if(!e.document)throw new Error("jQuery requires a window with a document");return t(e)}:t(e)}("undefined"!=typeof window?window:this,function(h,e){function t(e,t){return t.toUpperCase()}var n=[],l=n.slice,g=n.concat,s=n.push,o=n.indexOf,r={},i=r.toString,m=r.hasOwnProperty,v={},y=h.document,a="2.1.4",T=function(e,t){return new T.fn.init(e,t)},c=/^[\s\uFEFF\xA0]+|[\s\uFEFF\xA0]+$/g,u=/^-ms-/,p=/-([\da-z])/gi;function f(e){var t="length"in e&&e.length,n=T.type(e);return"function"!==n&&!T.isWindow(e)&&(!(1!==e.nodeType||!t)||("array"===n||0===t||"number"==typeof t&&0<t&&t-1 in e))}T.fn=T.prototype={jquery:a,constructor:T,selector:"",length:0,toArray:function(){return l.call(this)},get:function(e){return null!=e?e<0?this[e+this.length]:this[e]:l.call(this)},pushStack:function(e){var t=T.merge(this.constructor(),e);return t.prevObject=this,t.context=this.context,t},each:function(e,t){return T.each(this,e,t)},map:function(n){return this.pushStack(T.map(this,function(e,t){return n.call(e,t,e)}))},slice:function(){return this.pushStack(l.apply(this,arguments))},first:function(){return this.eq(0)},last:function(){return this.eq(-1)},eq:function(e){var t=this.length,n=+e+(e<0?t:0);return this.pushStack(0<=n&&n<t?[this[n]]:[])},end:function(){return this.prevObject||this.constructor(null)},push:s,sort:n.sort,splice:n.splice},T.extend=T.fn.extend=function(){var e,t,n,r,o,i,a=arguments[0]||{},s=1,c=arguments.length,u=!1;for("boolean"==typeof a&&(u=a,a=arguments[s]||{},s++),"object"==typeof a||T.isFunction(a)||(a={}),s===c&&(a=this,s--);s<c;s++)if(null!=(e=arguments[s]))for(t in e)n=a[t],a!==(r=e[t])&&(u&&r&&(T.isPlainObject(r)||(o=T.isArray(r)))?(i=o?(o=!1,n&&T.isArray(n)?n:[]):n&&T.isPlainObject(n)?n:{},a[t]=T.extend(u,i,r)):void 0!==r&&(a[t]=r));return a},T.extend({expando:"jQuery"+(a+Math.random()).replace(/\D/g,""),isReady:!0,error:function(e){throw new Error(e)},noop:function(){},isFunction:function(e){return"function"===T.type(e)},isArray:Array.isArray,isWindow:function(e){return null!=e&&e===e.window},isNumeric:function(e){return!T.isArray(e)&&0<=e-parseFloat(e)+1},isPlainObject:function(e){return"object"===T.type(e)&&!e.nodeType&&!T.isWindow(e)&&!(e.constructor&&!m.call(e.constructor.prototype,"isPrototypeOf"))},isEmptyObject:function(e){var t;for(t in e)return!1;return!0},type:function(e){return null==e?e+"":"object"==typeof e||"function"==typeof e?r[i.call(e)]||"object":typeof e},globalEval:function(e){var t,n=eval;(e=T.trim(e))&&(1===e.indexOf("use strict")?((t=y.createElement("script")).text=e,y.head.appendChild(t).parentNode.removeChild(t)):n(e))},camelCase:function(e){return e.replace(u,"ms-").replace(p,t)},nodeName:function(e,t){return e.nodeName&&e.nodeName.toLowerCase()===t.toLowerCase()},each:function(e,t,n){var r=0,o=e.length,i=f(e);if(n){if(i)for(;r<o&&!1!==t.apply(e[r],n);r++);else for(r in e)if(!1===t.apply(e[r],n))break}else if(i)for(;r<o&&!1!==t.call(e[r],r,e[r]);r++);else for(r in e)if(!1===t.call(e[r],r,e[r]))break;return e},trim:function(e){return null==e?"":(e+"").replace(c,"")},makeArray:function(e,t){var n=t||[];return null!=e&&(f(Object(e))?T.merge(n,"string"==typeof e?[e]:e):s.call(n,e)),n},inArray:function(e,t,n){return null==t?-1:o.call(t,e,n)},merge:function(e,t){for(var n=+t.length,r=0,o=e.length;r<n;r++)e[o++]=t[r];return e.length=o,e},grep:function(e,t,n){for(var r=[],o=0,i=e.length,a=!n;o<i;o++)!t(e[o],o)!=a&&r.push(e[o]);return r},map:function(e,t,n){var r,o=0,i=e.length,a=[];if(f(e))for(;o<i;o++)null!=(r=t(e[o],o,n))&&a.push(r);else for(o in e)null!=(r=t(e[o],o,n))&&a.push(r);return g.apply([],a)},guid:1,proxy:function(e,t){var n,r,o;return"string"==typeof t&&(n=e[t],t=e,e=n),T.isFunction(e)?(r=l.call(arguments,2),(o=function(){return e.apply(t||this,r.concat(l.call(arguments)))}).guid=e.guid=e.guid||T.guid++,o):void 0},now:Date.now,support:v}),T.each("Boolean Number String Function Array Date RegExp Object Error".split(" "),function(e,t){r["[object "+t+"]"]=t.toLowerCase()});var d=function(n){function p(e,t,n){var r="0x"+t-65536;return r!=r||n?t:r<0?String.fromCharCode(65536+r):String.fromCharCode(r>>10|55296,1023&r|56320)}function r(){v()}var e,h,b,i,o,g,f,m,w,u,l,v,k,a,y,x,s,d,T,C="sizzle"+1*new Date,S=n.document,_=0,j=0,c=ie(),$=ie(),A=ie(),E=function(e,t){return e===t&&(l=!0),0},N={}.hasOwnProperty,t=[],O=t.pop,D=t.push,L=t.push,F=t.slice,P=function(e,t){for(var n=0,r=e.length;n<r;n++)if(e[n]===t)return n;return-1},I="checked|selected|async|autofocus|autoplay|controls|defer|disabled|hidden|ismap|loop|multiple|open|readonly|required|scoped",q="[\\x20\\t\\r\\n\\f]",z="(?:\\\\.|[\\w-]|[^\\x00-\\xa0])+",H=z.replace("w","w#"),M="\\["+q+"*("+z+")(?:"+q+"*([*^$|!~]?=)"+q+"*(?:'((?:\\\\.|[^\\\\'])*)'|\"((?:\\\\.|[^\\\\\"])*)\"|("+H+"))|)"+q+"*\\]",B=":("+z+")(?:\\((('((?:\\\\.|[^\\\\'])*)'|\"((?:\\\\.|[^\\\\\"])*)\")|((?:\\\\.|[^\\\\()[\\]]|"+M+")*)|.*)\\)|)",W=new RegExp(q+"+","g"),R=new RegExp("^"+q+"+|((?:^|[^\\\\])(?:\\\\.)*)"+q+"+$","g"),V=new RegExp("^"+q+"*,"+q+"*"),U=new RegExp("^"+q+"*([>+~]|"+q+")"+q+"*"),X=new RegExp("="+q+"*([^\\]'\"]*?)"+q+"*\\]","g"),Z=new RegExp(B),J=new RegExp("^"+H+"$"),Y={ID:new RegExp("^#("+z+")"),CLASS:new RegExp("^\\.("+z+")"),TAG:new RegExp("^("+z.replace("w","w*")+")"),ATTR:new RegExp("^"+M),PSEUDO:new RegExp("^"+B),CHILD:new RegExp("^:(only|first|last|nth|nth-last)-(child|of-type)(?:\\("+q+"*(even|odd|(([+-]|)(\\d*)n|)"+q+"*(?:([+-]|)"+q+"*(\\d+)|))"+q+"*\\)|)","i"),bool:new RegExp("^(?:"+I+")$","i"),needsContext:new RegExp("^"+q+"*[>+~]|:(even|odd|eq|gt|lt|nth|first|last)(?:\\("+q+"*((?:-\\d)?\\d*)"+q+"*\\)|)(?=[^-]|$)","i")},G=/^(?:input|select|textarea|button)$/i,K=/^h\d$/i,Q=/^[^{]+\{\s*\[native \w/,ee=/^(?:#([\w-]+)|(\w+)|\.([\w-]+))$/,te=/[+~]/,ne=/'|\\/g,re=new RegExp("\\\\([\\da-f]{1,6}"+q+"?|("+q+")|.)","ig");try{L.apply(t=F.call(S.childNodes),S.childNodes),t[S.childNodes.length].nodeType}catch(e){L={apply:t.length?function(e,t){D.apply(e,F.call(t))}:function(e,t){for(var n=e.length,r=0;e[n++]=t[r++];);e.length=n-1}}}function oe(e,t,n,r){var o,i,a,s,c,u,l,p,f,d;if((t?t.ownerDocument||t:S)!==k&&v(t),n=n||[],s=(t=t||k).nodeType,"string"!=typeof e||!e||1!==s&&9!==s&&11!==s)return n;if(!r&&y){if(11!==s&&(o=ee.exec(e)))if(a=o[1]){if(9===s){if(!(i=t.getElementById(a))||!i.parentNode)return n;if(i.id===a)return n.push(i),n}else if(t.ownerDocument&&(i=t.ownerDocument.getElementById(a))&&T(t,i)&&i.id===a)return n.push(i),n}else{if(o[2])return L.apply(n,t.getElementsByTagName(e)),n;if((a=o[3])&&h.getElementsByClassName)return L.apply(n,t.getElementsByClassName(a)),n}if(h.qsa&&(!x||!x.test(e))){if(p=l=C,f=t,d=1!==s&&e,1===s&&"object"!==t.nodeName.toLowerCase()){for(u=g(e),(l=t.getAttribute("id"))?p=l.replace(ne,"\\$&"):t.setAttribute("id",p),p="[id='"+p+"'] ",c=u.length;c--;)u[c]=p+ge(u[c]);f=te.test(e)&&de(t.parentNode)||t,d=u.join(",")}if(d)try{return L.apply(n,f.querySelectorAll(d)),n}catch(e){}finally{l||t.removeAttribute("id")}}}return m(e.replace(R,"$1"),t,n,r)}function ie(){var r=[];return function e(t,n){return r.push(t+" ")>b.cacheLength&&delete e[r.shift()],e[t+" "]=n}}function ae(e){return e[C]=!0,e}function se(e){var t=k.createElement("div");try{return!!e(t)}catch(e){return!1}finally{t.parentNode&&t.parentNode.removeChild(t),t=null}}function ce(e,t){for(var n=e.split("|"),r=e.length;r--;)b.attrHandle[n[r]]=t}function ue(e,t){var n=t&&e,r=n&&1===e.nodeType&&1===t.nodeType&&(~t.sourceIndex||1<<31)-(~e.sourceIndex||1<<31);if(r)return r;if(n)for(;n=n.nextSibling;)if(n===t)return-1;return e?1:-1}function le(t){return function(e){return"input"===e.nodeName.toLowerCase()&&e.type===t}}function pe(n){return function(e){var t=e.nodeName.toLowerCase();return("input"===t||"button"===t)&&e.type===n}}function fe(a){return ae(function(i){return i=+i,ae(function(e,t){for(var n,r=a([],e.length,i),o=r.length;o--;)e[n=r[o]]&&(e[n]=!(t[n]=e[n]))})})}function de(e){return e&&void 0!==e.getElementsByTagName&&e}for(e in h=oe.support={},o=oe.isXML=function(e){var t=e&&(e.ownerDocument||e).documentElement;return!!t&&"HTML"!==t.nodeName},v=oe.setDocument=function(e){var t,n,c=e?e.ownerDocument||e:S;return c!==k&&9===c.nodeType&&c.documentElement?(a=(k=c).documentElement,(n=c.defaultView)&&n!==n.top&&(n.addEventListener?n.addEventListener("unload",r,!1):n.attachEvent&&n.attachEvent("onunload",r)),y=!o(c),h.attributes=se(function(e){return e.className="i",!e.getAttribute("className")}),h.getElementsByTagName=se(function(e){return e.appendChild(c.createComment("")),!e.getElementsByTagName("*").length}),h.getElementsByClassName=Q.test(c.getElementsByClassName),h.getById=se(function(e){return a.appendChild(e).id=C,!c.getElementsByName||!c.getElementsByName(C).length}),h.getById?(b.find.ID=function(e,t){if(void 0!==t.getElementById&&y){var n=t.getElementById(e);return n&&n.parentNode?[n]:[]}},b.filter.ID=function(e){var t=e.replace(re,p);return function(e){return e.getAttribute("id")===t}}):(delete b.find.ID,b.filter.ID=function(e){var n=e.replace(re,p);return function(e){var t=void 0!==e.getAttributeNode&&e.getAttributeNode("id");return t&&t.value===n}}),b.find.TAG=h.getElementsByTagName?function(e,t){return void 0!==t.getElementsByTagName?t.getElementsByTagName(e):h.qsa?t.querySelectorAll(e):void 0}:function(e,t){var n,r=[],o=0,i=t.getElementsByTagName(e);if("*"!==e)return i;for(;n=i[o++];)1===n.nodeType&&r.push(n);return r},b.find.CLASS=h.getElementsByClassName&&function(e,t){return y?t.getElementsByClassName(e):void 0},s=[],x=[],(h.qsa=Q.test(c.querySelectorAll))&&(se(function(e){a.appendChild(e).innerHTML="<a id='"+C+"'></a><select id='"+C+"-\f]' msallowcapture=''><option selected=''></option></select>",e.querySelectorAll("[msallowcapture^='']").length&&x.push("[*^$]="+q+"*(?:''|\"\")"),e.querySelectorAll("[selected]").length||x.push("\\["+q+"*(?:value|"+I+")"),e.querySelectorAll("[id~="+C+"-]").length||x.push("~="),e.querySelectorAll(":checked").length||x.push(":checked"),e.querySelectorAll("a#"+C+"+*").length||x.push(".#.+[+~]")}),se(function(e){var t=c.createElement("input");t.setAttribute("type","hidden"),e.appendChild(t).setAttribute("name","D"),e.querySelectorAll("[name=d]").length&&x.push("name"+q+"*[*^$|!~]?="),e.querySelectorAll(":enabled").length||x.push(":enabled",":disabled"),e.querySelectorAll("*,:x"),x.push(",.*:")})),(h.matchesSelector=Q.test(d=a.matches||a.webkitMatchesSelector||a.mozMatchesSelector||a.oMatchesSelector||a.msMatchesSelector))&&se(function(e){h.disconnectedMatch=d.call(e,"div"),d.call(e,"[s!='']:x"),s.push("!=",B)}),x=x.length&&new RegExp(x.join("|")),s=s.length&&new RegExp(s.join("|")),t=Q.test(a.compareDocumentPosition),T=t||Q.test(a.contains)?function(e,t){var n=9===e.nodeType?e.documentElement:e,r=t&&t.parentNode;return e===r||!(!r||1!==r.nodeType||!(n.contains?n.contains(r):e.compareDocumentPosition&&16&e.compareDocumentPosition(r)))}:function(e,t){if(t)for(;t=t.parentNode;)if(t===e)return!0;return!1},E=t?function(e,t){if(e===t)return l=!0,0;var n=!e.compareDocumentPosition-!t.compareDocumentPosition;return n||(1&(n=(e.ownerDocument||e)===(t.ownerDocument||t)?e.compareDocumentPosition(t):1)||!h.sortDetached&&t.compareDocumentPosition(e)===n?e===c||e.ownerDocument===S&&T(S,e)?-1:t===c||t.ownerDocument===S&&T(S,t)?1:u?P(u,e)-P(u,t):0:4&n?-1:1)}:function(e,t){if(e===t)return l=!0,0;var n,r=0,o=e.parentNode,i=t.parentNode,a=[e],s=[t];if(!o||!i)return e===c?-1:t===c?1:o?-1:i?1:u?P(u,e)-P(u,t):0;if(o===i)return ue(e,t);for(n=e;n=n.parentNode;)a.unshift(n);for(n=t;n=n.parentNode;)s.unshift(n);for(;a[r]===s[r];)r++;return r?ue(a[r],s[r]):a[r]===S?-1:s[r]===S?1:0},c):k},oe.matches=function(e,t){return oe(e,null,null,t)},oe.matchesSelector=function(e,t){if((e.ownerDocument||e)!==k&&v(e),t=t.replace(X,"='$1']"),!(!h.matchesSelector||!y||s&&s.test(t)||x&&x.test(t)))try{var n=d.call(e,t);if(n||h.disconnectedMatch||e.document&&11!==e.document.nodeType)return n}catch(e){}return 0<oe(t,k,null,[e]).length},oe.contains=function(e,t){return(e.ownerDocument||e)!==k&&v(e),T(e,t)},oe.attr=function(e,t){(e.ownerDocument||e)!==k&&v(e);var n=b.attrHandle[t.toLowerCase()],r=n&&N.call(b.attrHandle,t.toLowerCase())?n(e,t,!y):void 0;return void 0!==r?r:h.attributes||!y?e.getAttribute(t):(r=e.getAttributeNode(t))&&r.specified?r.value:null},oe.error=function(e){throw new Error("Syntax error, unrecognized expression: "+e)},oe.uniqueSort=function(e){var t,n=[],r=0,o=0;if(l=!h.detectDuplicates,u=!h.sortStable&&e.slice(0),e.sort(E),l){for(;t=e[o++];)t===e[o]&&(r=n.push(o));for(;r--;)e.splice(n[r],1)}return u=null,e},i=oe.getText=function(e){var t,n="",r=0,o=e.nodeType;if(o){if(1===o||9===o||11===o){if("string"==typeof e.textContent)return e.textContent;for(e=e.firstChild;e;e=e.nextSibling)n+=i(e)}else if(3===o||4===o)return e.nodeValue}else for(;t=e[r++];)n+=i(t);return n},(b=oe.selectors={cacheLength:50,createPseudo:ae,match:Y,attrHandle:{},find:{},relative:{">":{dir:"parentNode",first:!0}," ":{dir:"parentNode"},"+":{dir:"previousSibling",first:!0},"~":{dir:"previousSibling"}},preFilter:{ATTR:function(e){return e[1]=e[1].replace(re,p),e[3]=(e[3]||e[4]||e[5]||"").replace(re,p),"~="===e[2]&&(e[3]=" "+e[3]+" "),e.slice(0,4)},CHILD:function(e){return e[1]=e[1].toLowerCase(),"nth"===e[1].slice(0,3)?(e[3]||oe.error(e[0]),e[4]=+(e[4]?e[5]+(e[6]||1):2*("even"===e[3]||"odd"===e[3])),e[5]=+(e[7]+e[8]||"odd"===e[3])):e[3]&&oe.error(e[0]),e},PSEUDO:function(e){var t,n=!e[6]&&e[2];return Y.CHILD.test(e[0])?null:(e[3]?e[2]=e[4]||e[5]||"":n&&Z.test(n)&&(t=g(n,!0))&&(t=n.indexOf(")",n.length-t)-n.length)&&(e[0]=e[0].slice(0,t),e[2]=n.slice(0,t)),e.slice(0,3))}},filter:{TAG:function(e){var t=e.replace(re,p).toLowerCase();return"*"===e?function(){return!0}:function(e){return e.nodeName&&e.nodeName.toLowerCase()===t}},CLASS:function(e){var t=c[e+" "];return t||(t=new RegExp("(^|"+q+")"+e+"("+q+"|$)"))&&c(e,function(e){return t.test("string"==typeof e.className&&e.className||void 0!==e.getAttribute&&e.getAttribute("class")||"")})},ATTR:function(n,r,o){return function(e){var t=oe.attr(e,n);return null==t?"!="===r:!r||(t+="","="===r?t===o:"!="===r?t!==o:"^="===r?o&&0===t.indexOf(o):"*="===r?o&&-1<t.indexOf(o):"$="===r?o&&t.slice(-o.length)===o:"~="===r?-1<(" "+t.replace(W," ")+" ").indexOf(o):"|="===r&&(t===o||t.slice(0,o.length+1)===o+"-"))}},CHILD:function(d,e,t,h,g){var m="nth"!==d.slice(0,3),v="last"!==d.slice(-4),y="of-type"===e;return 1===h&&0===g?function(e){return!!e.parentNode}:function(e,t,n){var r,o,i,a,s,c,u=m!=v?"nextSibling":"previousSibling",l=e.parentNode,p=y&&e.nodeName.toLowerCase(),f=!n&&!y;if(l){if(m){for(;u;){for(i=e;i=i[u];)if(y?i.nodeName.toLowerCase()===p:1===i.nodeType)return!1;c=u="only"===d&&!c&&"nextSibling"}return!0}if(c=[v?l.firstChild:l.lastChild],v&&f){for(s=(r=(o=l[C]||(l[C]={}))[d]||[])[0]===_&&r[1],a=r[0]===_&&r[2],i=s&&l.childNodes[s];i=++s&&i&&i[u]||(a=s=0)||c.pop();)if(1===i.nodeType&&++a&&i===e){o[d]=[_,s,a];break}}else if(f&&(r=(e[C]||(e[C]={}))[d])&&r[0]===_)a=r[1];else for(;(i=++s&&i&&i[u]||(a=s=0)||c.pop())&&((y?i.nodeName.toLowerCase()!==p:1!==i.nodeType)||!++a||(f&&((i[C]||(i[C]={}))[d]=[_,a]),i!==e)););return(a-=g)===h||a%h==0&&0<=a/h}}},PSEUDO:function(e,i){var t,a=b.pseudos[e]||b.setFilters[e.toLowerCase()]||oe.error("unsupported pseudo: "+e);return a[C]?a(i):1<a.length?(t=[e,e,"",i],b.setFilters.hasOwnProperty(e.toLowerCase())?ae(function(e,t){for(var n,r=a(e,i),o=r.length;o--;)e[n=P(e,r[o])]=!(t[n]=r[o])}):function(e){return a(e,0,t)}):a}},pseudos:{not:ae(function(e){var r=[],o=[],s=f(e.replace(R,"$1"));return s[C]?ae(function(e,t,n,r){for(var o,i=s(e,null,r,[]),a=e.length;a--;)(o=i[a])&&(e[a]=!(t[a]=o))}):function(e,t,n){return r[0]=e,s(r,null,n,o),r[0]=null,!o.pop()}}),has:ae(function(t){return function(e){return 0<oe(t,e).length}}),contains:ae(function(t){return t=t.replace(re,p),function(e){return-1<(e.textContent||e.innerText||i(e)).indexOf(t)}}),lang:ae(function(n){return J.test(n||"")||oe.error("unsupported lang: "+n),n=n.replace(re,p).toLowerCase(),function(e){var t;do{if(t=y?e.lang:e.getAttribute("xml:lang")||e.getAttribute("lang"))return(t=t.toLowerCase())===n||0===t.indexOf(n+"-")}while((e=e.parentNode)&&1===e.nodeType);return!1}}),target:function(e){var t=n.location&&n.location.hash;return t&&t.slice(1)===e.id},root:function(e){return e===a},focus:function(e){return e===k.activeElement&&(!k.hasFocus||k.hasFocus())&&!!(e.type||e.href||~e.tabIndex)},enabled:function(e){return!1===e.disabled},disabled:function(e){return!0===e.disabled},checked:function(e){var t=e.nodeName.toLowerCase();return"input"===t&&!!e.checked||"option"===t&&!!e.selected},selected:function(e){return e.parentNode&&e.parentNode.selectedIndex,!0===e.selected},empty:function(e){for(e=e.firstChild;e;e=e.nextSibling)if(e.nodeType<6)return!1;return!0},parent:function(e){return!b.pseudos.empty(e)},header:function(e){return K.test(e.nodeName)},input:function(e){return G.test(e.nodeName)},button:function(e){var t=e.nodeName.toLowerCase();return"input"===t&&"button"===e.type||"button"===t},text:function(e){var t;return"input"===e.nodeName.toLowerCase()&&"text"===e.type&&(null==(t=e.getAttribute("type"))||"text"===t.toLowerCase())},first:fe(function(){return[0]}),last:fe(function(e,t){return[t-1]}),eq:fe(function(e,t,n){return[n<0?n+t:n]}),even:fe(function(e,t){for(var n=0;n<t;n+=2)e.push(n);return e}),odd:fe(function(e,t){for(var n=1;n<t;n+=2)e.push(n);return e}),lt:fe(function(e,t,n){for(var r=n<0?n+t:n;0<=--r;)e.push(r);return e}),gt:fe(function(e,t,n){for(var r=n<0?n+t:n;++r<t;)e.push(r);return e})}}).pseudos.nth=b.pseudos.eq,{radio:!0,checkbox:!0,file:!0,password:!0,image:!0})b.pseudos[e]=le(e);for(e in{submit:!0,reset:!0})b.pseudos[e]=pe(e);function he(){}function ge(e){for(var t=0,n=e.length,r="";t<n;t++)r+=e[t].value;return r}function me(a,e,t){var s=e.dir,c=t&&"parentNode"===s,u=j++;return e.first?function(e,t,n){for(;e=e[s];)if(1===e.nodeType||c)return a(e,t,n)}:function(e,t,n){var r,o,i=[_,u];if(n){for(;e=e[s];)if((1===e.nodeType||c)&&a(e,t,n))return!0}else for(;e=e[s];)if(1===e.nodeType||c){if((r=(o=e[C]||(e[C]={}))[s])&&r[0]===_&&r[1]===u)return i[2]=r[2];if((o[s]=i)[2]=a(e,t,n))return!0}}}function ve(o){return 1<o.length?function(e,t,n){for(var r=o.length;r--;)if(!o[r](e,t,n))return!1;return!0}:o[0]}function ye(e,t,n,r,o){for(var i,a=[],s=0,c=e.length,u=null!=t;s<c;s++)!(i=e[s])||n&&!n(i,r,o)||(a.push(i),u&&t.push(s));return a}function xe(d,h,g,m,v,e){return m&&!m[C]&&(m=xe(m)),v&&!v[C]&&(v=xe(v,e)),ae(function(e,t,n,r){var o,i,a,s=[],c=[],u=t.length,l=e||function(e,t,n){for(var r=0,o=t.length;r<o;r++)oe(e,t[r],n);return n}(h||"*",n.nodeType?[n]:n,[]),p=!d||!e&&h?l:ye(l,s,d,n,r),f=g?v||(e?d:u||m)?[]:t:p;if(g&&g(p,f,n,r),m)for(o=ye(f,c),m(o,[],n,r),i=o.length;i--;)(a=o[i])&&(f[c[i]]=!(p[c[i]]=a));if(e){if(v||d){if(v){for(o=[],i=f.length;i--;)(a=f[i])&&o.push(p[i]=a);v(null,f=[],o,r)}for(i=f.length;i--;)(a=f[i])&&-1<(o=v?P(e,a):s[i])&&(e[o]=!(t[o]=a))}}else f=ye(f===t?f.splice(u,f.length):f),v?v(null,t,f,r):L.apply(t,f)})}function be(e){for(var o,t,n,r=e.length,i=b.relative[e[0].type],a=i||b.relative[" "],s=i?1:0,c=me(function(e){return e===o},a,!0),u=me(function(e){return-1<P(o,e)},a,!0),l=[function(e,t,n){var r=!i&&(n||t!==w)||((o=t).nodeType?c(e,t,n):u(e,t,n));return o=null,r}];s<r;s++)if(t=b.relative[e[s].type])l=[me(ve(l),t)];else{if((t=b.filter[e[s].type].apply(null,e[s].matches))[C]){for(n=++s;n<r&&!b.relative[e[n].type];n++);return xe(1<s&&ve(l),1<s&&ge(e.slice(0,s-1).concat({value:" "===e[s-2].type?"*":""})).replace(R,"$1"),t,s<n&&be(e.slice(s,n)),n<r&&be(e=e.slice(n)),n<r&&ge(e))}l.push(t)}return ve(l)}return he.prototype=b.filters=b.pseudos,b.setFilters=new he,g=oe.tokenize=function(e,t){var n,r,o,i,a,s,c,u=$[e+" "];if(u)return t?0:u.slice(0);for(a=e,s=[],c=b.preFilter;a;){for(i in n&&!(r=V.exec(a))||(r&&(a=a.slice(r[0].length)||a),s.push(o=[])),n=!1,(r=U.exec(a))&&(n=r.shift(),o.push({value:n,type:r[0].replace(R," ")}),a=a.slice(n.length)),b.filter)!(r=Y[i].exec(a))||c[i]&&!(r=c[i](r))||(n=r.shift(),o.push({value:n,type:i,matches:r}),a=a.slice(n.length));if(!n)break}return t?a.length:a?oe.error(e):$(e,s).slice(0)},f=oe.compile=function(e,t){var n,r=[],o=[],i=A[e+" "];if(!i){for(t||(t=g(e)),n=t.length;n--;)(i=be(t[n]))[C]?r.push(i):o.push(i);(i=A(e,function(m,v){function e(e,t,n,r,o){var i,a,s,c=0,u="0",l=e&&[],p=[],f=w,d=e||x&&b.find.TAG("*",o),h=_+=null==f?1:Math.random()||.1,g=d.length;for(o&&(w=t!==k&&t);u!==g&&null!=(i=d[u]);u++){if(x&&i){for(a=0;s=m[a++];)if(s(i,t,n)){r.push(i);break}o&&(_=h)}y&&((i=!s&&i)&&c--,e&&l.push(i))}if(c+=u,y&&u!==c){for(a=0;s=v[a++];)s(l,p,t,n);if(e){if(0<c)for(;u--;)l[u]||p[u]||(p[u]=O.call(r));p=ye(p)}L.apply(r,p),o&&!e&&0<p.length&&1<c+v.length&&oe.uniqueSort(r)}return o&&(_=h,w=f),l}var y=0<v.length,x=0<m.length;return y?ae(e):e}(o,r))).selector=e}return i},m=oe.select=function(e,t,n,r){var o,i,a,s,c,u="function"==typeof e&&e,l=!r&&g(e=u.selector||e);if(n=n||[],1===l.length){if(2<(i=l[0]=l[0].slice(0)).length&&"ID"===(a=i[0]).type&&h.getById&&9===t.nodeType&&y&&b.relative[i[1].type]){if(!(t=(b.find.ID(a.matches[0].replace(re,p),t)||[])[0]))return n;u&&(t=t.parentNode),e=e.slice(i.shift().value.length)}for(o=Y.needsContext.test(e)?0:i.length;o--&&(a=i[o],!b.relative[s=a.type]);)if((c=b.find[s])&&(r=c(a.matches[0].replace(re,p),te.test(i[0].type)&&de(t.parentNode)||t))){if(i.splice(o,1),!(e=r.length&&ge(i)))return L.apply(n,r),n;break}}return(u||f(e,l))(r,t,!y,n,te.test(e)&&de(t.parentNode)||t),n},h.sortStable=C.split("").sort(E).join("")===C,h.detectDuplicates=!!l,v(),h.sortDetached=se(function(e){return 1&e.compareDocumentPosition(k.createElement("div"))}),se(function(e){return e.innerHTML="<a href='#'></a>","#"===e.firstChild.getAttribute("href")})||ce("type|href|height|width",function(e,t,n){return n?void 0:e.getAttribute(t,"type"===t.toLowerCase()?1:2)}),h.attributes&&se(function(e){return e.innerHTML="<input/>",e.firstChild.setAttribute("value",""),""===e.firstChild.getAttribute("value")})||ce("value",function(e,t,n){return n||"input"!==e.nodeName.toLowerCase()?void 0:e.defaultValue}),se(function(e){return null==e.getAttribute("disabled")})||ce(I,function(e,t,n){var r;return n?void 0:!0===e[t]?t.toLowerCase():(r=e.getAttributeNode(t))&&r.specified?r.value:null}),oe}(h);T.find=d,T.expr=d.selectors,T.expr[":"]=T.expr.pseudos,T.unique=d.uniqueSort,T.text=d.getText,T.isXMLDoc=d.isXML,T.contains=d.contains;var x=T.expr.match.needsContext,b=/^<(\w+)\s*\/?>(?:<\/\1>|)$/,w=/^.[^:#\[\.,]*$/;function k(e,n,r){if(T.isFunction(n))return T.grep(e,function(e,t){return!!n.call(e,t,e)!==r});if(n.nodeType)return T.grep(e,function(e){return e===n!==r});if("string"==typeof n){if(w.test(n))return T.filter(n,e,r);n=T.filter(n,e)}return T.grep(e,function(e){return 0<=o.call(n,e)!==r})}T.filter=function(e,t,n){var r=t[0];return n&&(e=":not("+e+")"),1===t.length&&1===r.nodeType?T.find.matchesSelector(r,e)?[r]:[]:T.find.matches(e,T.grep(t,function(e){return 1===e.nodeType}))},T.fn.extend({find:function(e){var t,n=this.length,r=[],o=this;if("string"!=typeof e)return this.pushStack(T(e).filter(function(){for(t=0;t<n;t++)if(T.contains(o[t],this))return!0}));for(t=0;t<n;t++)T.find(e,o[t],r);return(r=this.pushStack(1<n?T.unique(r):r)).selector=this.selector?this.selector+" "+e:e,r},filter:function(e){return this.pushStack(k(this,e||[],!1))},not:function(e){return this.pushStack(k(this,e||[],!0))},is:function(e){return!!k(this,"string"==typeof e&&x.test(e)?T(e):e||[],!1).length}});var C,S=/^(?:\s*(<[\w\W]+>)[^>]*|#([\w-]*))$/;(T.fn.init=function(e,t){var n,r;if(!e)return this;if("string"!=typeof e)return e.nodeType?(this.context=this[0]=e,this.length=1,this):T.isFunction(e)?void 0!==C.ready?C.ready(e):e(T):(void 0!==e.selector&&(this.selector=e.selector,this.context=e.context),T.makeArray(e,this));if(!(n="<"===e[0]&&">"===e[e.length-1]&&3<=e.length?[null,e,null]:S.exec(e))||!n[1]&&t)return!t||t.jquery?(t||C).find(e):this.constructor(t).find(e);if(n[1]){if(t=t instanceof T?t[0]:t,T.merge(this,T.parseHTML(n[1],t&&t.nodeType?t.ownerDocument||t:y,!0)),b.test(n[1])&&T.isPlainObject(t))for(n in t)T.isFunction(this[n])?this[n](t[n]):this.attr(n,t[n]);return this}return(r=y.getElementById(n[2]))&&r.parentNode&&(this.length=1,this[0]=r),this.context=y,this.selector=e,this}).prototype=T.fn,C=T(y);var _=/^(?:parents|prev(?:Until|All))/,j={children:!0,contents:!0,next:!0,prev:!0};function $(e,t){for(;(e=e[t])&&1!==e.nodeType;);return e}T.extend({dir:function(e,t,n){for(var r=[],o=void 0!==n;(e=e[t])&&9!==e.nodeType;)if(1===e.nodeType){if(o&&T(e).is(n))break;r.push(e)}return r},sibling:function(e,t){for(var n=[];e;e=e.nextSibling)1===e.nodeType&&e!==t&&n.push(e);return n}}),T.fn.extend({has:function(e){var t=T(e,this),n=t.length;return this.filter(function(){for(var e=0;e<n;e++)if(T.contains(this,t[e]))return!0})},closest:function(e,t){for(var n,r=0,o=this.length,i=[],a=x.test(e)||"string"!=typeof e?T(e,t||this.context):0;r<o;r++)for(n=this[r];n&&n!==t;n=n.parentNode)if(n.nodeType<11&&(a?-1<a.index(n):1===n.nodeType&&T.find.matchesSelector(n,e))){i.push(n);break}return this.pushStack(1<i.length?T.unique(i):i)},index:function(e){return e?"string"==typeof e?o.call(T(e),this[0]):o.call(this,e.jquery?e[0]:e):this[0]&&this[0].parentNode?this.first().prevAll().length:-1},add:function(e,t){return this.pushStack(T.unique(T.merge(this.get(),T(e,t))))},addBack:function(e){return this.add(null==e?this.prevObject:this.prevObject.filter(e))}}),T.each({parent:function(e){var t=e.parentNode;return t&&11!==t.nodeType?t:null},parents:function(e){return T.dir(e,"parentNode")},parentsUntil:function(e,t,n){return T.dir(e,"parentNode",n)},next:function(e){return $(e,"nextSibling")},prev:function(e){return $(e,"previousSibling")},nextAll:function(e){return T.dir(e,"nextSibling")},prevAll:function(e){return T.dir(e,"previousSibling")},nextUntil:function(e,t,n){return T.dir(e,"nextSibling",n)},prevUntil:function(e,t,n){return T.dir(e,"previousSibling",n)},siblings:function(e){return T.sibling((e.parentNode||{}).firstChild,e)},children:function(e){return T.sibling(e.firstChild)},contents:function(e){return e.contentDocument||T.merge([],e.childNodes)}},function(r,o){T.fn[r]=function(e,t){var n=T.map(this,o,e);return"Until"!==r.slice(-5)&&(t=e),t&&"string"==typeof t&&(n=T.filter(t,n)),1<this.length&&(j[r]||T.unique(n),_.test(r)&&n.reverse()),this.pushStack(n)}});var A,E=/\S+/g,N={};function O(){y.removeEventListener("DOMContentLoaded",O,!1),h.removeEventListener("load",O,!1),T.ready()}T.Callbacks=function(o){o="string"==typeof o?N[o]||function(e){var n=N[e]={};return T.each(e.match(E)||[],function(e,t){n[t]=!0}),n}(o):T.extend({},o);var t,n,r,i,a,s,c=[],u=!o.once&&[],l=function(e){for(t=o.memory&&e,n=!0,s=i||0,i=0,a=c.length,r=!0;c&&s<a;s++)if(!1===c[s].apply(e[0],e[1])&&o.stopOnFalse){t=!1;break}r=!1,c&&(u?u.length&&l(u.shift()):t?c=[]:p.disable())},p={add:function(){if(c){var e=c.length;!function r(e){T.each(e,function(e,t){var n=T.type(t);"function"===n?o.unique&&p.has(t)||c.push(t):t&&t.length&&"string"!==n&&r(t)})}(arguments),r?a=c.length:t&&(i=e,l(t))}return this},remove:function(){return c&&T.each(arguments,function(e,t){for(var n;-1<(n=T.inArray(t,c,n));)c.splice(n,1),r&&(n<=a&&a--,n<=s&&s--)}),this},has:function(e){return e?-1<T.inArray(e,c):!(!c||!c.length)},empty:function(){return c=[],a=0,this},disable:function(){return c=u=t=void 0,this},disabled:function(){return!c},lock:function(){return u=void 0,t||p.disable(),this},locked:function(){return!u},fireWith:function(e,t){return!c||n&&!u||(t=[e,(t=t||[]).slice?t.slice():t],r?u.push(t):l(t)),this},fire:function(){return p.fireWith(this,arguments),this},fired:function(){return!!n}};return p},T.extend({Deferred:function(e){var i=[["resolve","done",T.Callbacks("once memory"),"resolved"],["reject","fail",T.Callbacks("once memory"),"rejected"],["notify","progress",T.Callbacks("memory")]],o="pending",a={state:function(){return o},always:function(){return s.done(arguments).fail(arguments),this},then:function(){var o=arguments;return T.Deferred(function(r){T.each(i,function(e,t){var n=T.isFunction(o[e])&&o[e];s[t[1]](function(){var e=n&&n.apply(this,arguments);e&&T.isFunction(e.promise)?e.promise().done(r.resolve).fail(r.reject).progress(r.notify):r[t[0]+"With"](this===a?r.promise():this,n?[e]:arguments)})}),o=null}).promise()},promise:function(e){return null!=e?T.extend(e,a):a}},s={};return a.pipe=a.then,T.each(i,function(e,t){var n=t[2],r=t[3];a[t[1]]=n.add,r&&n.add(function(){o=r},i[1^e][2].disable,i[2][2].lock),s[t[0]]=function(){return s[t[0]+"With"](this===s?a:this,arguments),this},s[t[0]+"With"]=n.fireWith}),a.promise(s),e&&e.call(s,s),s},when:function(e){function t(t,n,r){return function(e){n[t]=this,r[t]=1<arguments.length?l.call(arguments):e,r===o?u.notifyWith(n,r):--c||u.resolveWith(n,r)}}var o,n,r,i=0,a=l.call(arguments),s=a.length,c=1!==s||e&&T.isFunction(e.promise)?s:0,u=1===c?e:T.Deferred();if(1<s)for(o=new Array(s),n=new Array(s),r=new Array(s);i<s;i++)a[i]&&T.isFunction(a[i].promise)?a[i].promise().done(t(i,r,a)).fail(u.reject).progress(t(i,n,o)):--c;return c||u.resolveWith(r,a),u.promise()}}),T.fn.ready=function(e){return T.ready.promise().done(e),this},T.extend({isReady:!1,readyWait:1,holdReady:function(e){e?T.readyWait++:T.ready(!0)},ready:function(e){(!0===e?--T.readyWait:T.isReady)||((T.isReady=!0)!==e&&0<--T.readyWait||(A.resolveWith(y,[T]),T.fn.triggerHandler&&(T(y).triggerHandler("ready"),T(y).off("ready"))))}}),T.ready.promise=function(e){return A||(A=T.Deferred(),"complete"===y.readyState?setTimeout(T.ready):(y.addEventListener("DOMContentLoaded",O,!1),h.addEventListener("load",O,!1))),A.promise(e)},T.ready.promise();var D=T.access=function(e,t,n,r,o,i,a){var s=0,c=e.length,u=null==n;if("object"===T.type(n))for(s in o=!0,n)T.access(e,t,s,n[s],!0,i,a);else if(void 0!==r&&(o=!0,T.isFunction(r)||(a=!0),u&&(t=a?(t.call(e,r),null):(u=t,function(e,t,n){return u.call(T(e),n)})),t))for(;s<c;s++)t(e[s],n,a?r:r.call(e[s],s,t(e[s],n)));return o?e:u?t.call(e):c?t(e[0],n):i};function L(){Object.defineProperty(this.cache={},0,{get:function(){return{}}}),this.expando=T.expando+L.uid++}T.acceptData=function(e){return 1===e.nodeType||9===e.nodeType||!+e.nodeType},L.uid=1,L.accepts=T.acceptData,L.prototype={key:function(t){if(!L.accepts(t))return 0;var n={},r=t[this.expando];if(!r){r=L.uid++;try{n[this.expando]={value:r},Object.defineProperties(t,n)}catch(e){n[this.expando]=r,T.extend(t,n)}}return this.cache[r]||(this.cache[r]={}),r},set:function(e,t,n){var r,o=this.key(e),i=this.cache[o];if("string"==typeof t)i[t]=n;else if(T.isEmptyObject(i))T.extend(this.cache[o],t);else for(r in t)i[r]=t[r];return i},get:function(e,t){var n=this.cache[this.key(e)];return void 0===t?n:n[t]},access:function(e,t,n){var r;return void 0===t||t&&"string"==typeof t&&void 0===n?void 0!==(r=this.get(e,t))?r:this.get(e,T.camelCase(t)):(this.set(e,t,n),void 0!==n?n:t)},remove:function(e,t){var n,r,o,i=this.key(e),a=this.cache[i];if(void 0===t)this.cache[i]={};else{n=(r=T.isArray(t)?t.concat(t.map(T.camelCase)):(o=T.camelCase(t),t in a?[t,o]:(r=o)in a?[r]:r.match(E)||[])).length;for(;n--;)delete a[r[n]]}},hasData:function(e){return!T.isEmptyObject(this.cache[e[this.expando]]||{})},discard:function(e){e[this.expando]&&delete this.cache[e[this.expando]]}};var F=new L,P=new L,I=/^(?:\{[\w\W]*\}|\[[\w\W]*\])$/,q=/([A-Z])/g;function z(e,t,n){var r;if(void 0===n&&1===e.nodeType)if(r="data-"+t.replace(q,"-$1").toLowerCase(),"string"==typeof(n=e.getAttribute(r))){try{n="true"===n||"false"!==n&&("null"===n?null:+n+""===n?+n:I.test(n)?T.parseJSON(n):n)}catch(e){}P.set(e,t,n)}else n=void 0;return n}T.extend({hasData:function(e){return P.hasData(e)||F.hasData(e)},data:function(e,t,n){return P.access(e,t,n)},removeData:function(e,t){P.remove(e,t)},_data:function(e,t,n){return F.access(e,t,n)},_removeData:function(e,t){F.remove(e,t)}}),T.fn.extend({data:function(r,e){var t,n,o,i=this[0],a=i&&i.attributes;if(void 0!==r)return"object"==typeof r?this.each(function(){P.set(this,r)}):D(this,function(t){var e,n=T.camelCase(r);if(i&&void 0===t){if(void 0!==(e=P.get(i,r)))return e;if(void 0!==(e=P.get(i,n)))return e;if(void 0!==(e=z(i,n,void 0)))return e}else this.each(function(){var e=P.get(this,n);P.set(this,n,t),-1!==r.indexOf("-")&&void 0!==e&&P.set(this,r,t)})},null,e,1<arguments.length,null,!0);if(this.length&&(o=P.get(i),1===i.nodeType&&!F.get(i,"hasDataAttrs"))){for(t=a.length;t--;)a[t]&&(0===(n=a[t].name).indexOf("data-")&&(n=T.camelCase(n.slice(5)),z(i,n,o[n])));F.set(i,"hasDataAttrs",!0)}return o},removeData:function(e){return this.each(function(){P.remove(this,e)})}}),T.extend({queue:function(e,t,n){var r;return e?(t=(t||"fx")+"queue",r=F.get(e,t),n&&(!r||T.isArray(n)?r=F.access(e,t,T.makeArray(n)):r.push(n)),r||[]):void 0},dequeue:function(e,t){t=t||"fx";var n=T.queue(e,t),r=n.length,o=n.shift(),i=T._queueHooks(e,t);"inprogress"===o&&(o=n.shift(),r--),o&&("fx"===t&&n.unshift("inprogress"),delete i.stop,o.call(e,function(){T.dequeue(e,t)},i)),!r&&i&&i.empty.fire()},_queueHooks:function(e,t){var n=t+"queueHooks";return F.get(e,n)||F.access(e,n,{empty:T.Callbacks("once memory").add(function(){F.remove(e,[t+"queue",n])})})}}),T.fn.extend({queue:function(t,n){var e=2;return"string"!=typeof t&&(n=t,t="fx",e--),arguments.length<e?T.queue(this[0],t):void 0===n?this:this.each(function(){var e=T.queue(this,t,n);T._queueHooks(this,t),"fx"===t&&"inprogress"!==e[0]&&T.dequeue(this,t)})},dequeue:function(e){return this.each(function(){T.dequeue(this,e)})},clearQueue:function(e){return this.queue(e||"fx",[])},promise:function(e,t){function n(){--o||i.resolveWith(a,[a])}var r,o=1,i=T.Deferred(),a=this,s=this.length;for("string"!=typeof e&&(t=e,e=void 0),e=e||"fx";s--;)(r=F.get(a[s],e+"queueHooks"))&&r.empty&&(o++,r.empty.add(n));return n(),i.promise(t)}});function H(e,t){return e=t||e,"none"===T.css(e,"display")||!T.contains(e.ownerDocument,e)}var M,B,W=/[+-]?(?:\d*\.|)\d+(?:[eE][+-]?\d+|)/.source,R=["Top","Right","Bottom","Left"],V=/^(?:checkbox|radio)$/i;M=y.createDocumentFragment().appendChild(y.createElement("div")),(B=y.createElement("input")).setAttribute("type","radio"),B.setAttribute("checked","checked"),B.setAttribute("name","t"),M.appendChild(B),v.checkClone=M.cloneNode(!0).cloneNode(!0).lastChild.checked,M.innerHTML="<textarea>x</textarea>",v.noCloneChecked=!!M.cloneNode(!0).lastChild.defaultValue;var U="undefined";v.focusinBubbles="onfocusin"in h;var X=/^key/,Z=/^(?:mouse|pointer|contextmenu)|click/,J=/^(?:focusinfocus|focusoutblur)$/,Y=/^([^.]*)(?:\.(.+)|)$/;function G(){return!0}function K(){return!1}function Q(){try{return y.activeElement}catch(e){}}T.event={global:{},add:function(t,e,n,r,o){var i,a,s,c,u,l,p,f,d,h,g,m=F.get(t);if(m)for(n.handler&&(n=(i=n).handler,o=i.selector),n.guid||(n.guid=T.guid++),(c=m.events)||(c=m.events={}),(a=m.handle)||(a=m.handle=function(e){return typeof T!=U&&T.event.triggered!==e.type?T.event.dispatch.apply(t,arguments):void 0}),u=(e=(e||"").match(E)||[""]).length;u--;)d=g=(s=Y.exec(e[u])||[])[1],h=(s[2]||"").split(".").sort(),d&&(p=T.event.special[d]||{},d=(o?p.delegateType:p.bindType)||d,p=T.event.special[d]||{},l=T.extend({type:d,origType:g,data:r,handler:n,guid:n.guid,selector:o,needsContext:o&&T.expr.match.needsContext.test(o),namespace:h.join(".")},i),(f=c[d])||((f=c[d]=[]).delegateCount=0,p.setup&&!1!==p.setup.call(t,r,h,a)||t.addEventListener&&t.addEventListener(d,a,!1)),p.add&&(p.add.call(t,l),l.handler.guid||(l.handler.guid=n.guid)),o?f.splice(f.delegateCount++,0,l):f.push(l),T.event.global[d]=!0)},remove:function(e,t,n,r,o){var i,a,s,c,u,l,p,f,d,h,g,m=F.hasData(e)&&F.get(e);if(m&&(c=m.events)){for(u=(t=(t||"").match(E)||[""]).length;u--;)if(d=g=(s=Y.exec(t[u])||[])[1],h=(s[2]||"").split(".").sort(),d){for(p=T.event.special[d]||{},f=c[d=(r?p.delegateType:p.bindType)||d]||[],s=s[2]&&new RegExp("(^|\\.)"+h.join("\\.(?:.*\\.|)")+"(\\.|$)"),a=i=f.length;i--;)l=f[i],!o&&g!==l.origType||n&&n.guid!==l.guid||s&&!s.test(l.namespace)||r&&r!==l.selector&&("**"!==r||!l.selector)||(f.splice(i,1),l.selector&&f.delegateCount--,p.remove&&p.remove.call(e,l));a&&!f.length&&(p.teardown&&!1!==p.teardown.call(e,h,m.handle)||T.removeEvent(e,d,m.handle),delete c[d])}else for(d in c)T.event.remove(e,d+t[u],n,r,!0);T.isEmptyObject(c)&&(delete m.handle,F.remove(e,"events"))}},trigger:function(e,t,n,r){var o,i,a,s,c,u,l,p=[n||y],f=m.call(e,"type")?e.type:e,d=m.call(e,"namespace")?e.namespace.split("."):[];if(i=a=n=n||y,3!==n.nodeType&&8!==n.nodeType&&!J.test(f+T.event.triggered)&&(0<=f.indexOf(".")&&(f=(d=f.split(".")).shift(),d.sort()),c=f.indexOf(":")<0&&"on"+f,(e=e[T.expando]?e:new T.Event(f,"object"==typeof e&&e)).isTrigger=r?2:3,e.namespace=d.join("."),e.namespace_re=e.namespace?new RegExp("(^|\\.)"+d.join("\\.(?:.*\\.|)")+"(\\.|$)"):null,e.result=void 0,e.target||(e.target=n),t=null==t?[e]:T.makeArray(t,[e]),l=T.event.special[f]||{},r||!l.trigger||!1!==l.trigger.apply(n,t))){if(!r&&!l.noBubble&&!T.isWindow(n)){for(s=l.delegateType||f,J.test(s+f)||(i=i.parentNode);i;i=i.parentNode)p.push(i),a=i;a===(n.ownerDocument||y)&&p.push(a.defaultView||a.parentWindow||h)}for(o=0;(i=p[o++])&&!e.isPropagationStopped();)e.type=1<o?s:l.bindType||f,(u=(F.get(i,"events")||{})[e.type]&&F.get(i,"handle"))&&u.apply(i,t),(u=c&&i[c])&&u.apply&&T.acceptData(i)&&(e.result=u.apply(i,t),!1===e.result&&e.preventDefault());return e.type=f,r||e.isDefaultPrevented()||l._default&&!1!==l._default.apply(p.pop(),t)||!T.acceptData(n)||c&&T.isFunction(n[f])&&!T.isWindow(n)&&((a=n[c])&&(n[c]=null),n[T.event.triggered=f](),T.event.triggered=void 0,a&&(n[c]=a)),e.result}},dispatch:function(e){e=T.event.fix(e);var t,n,r,o,i,a=[],s=l.call(arguments),c=(F.get(this,"events")||{})[e.type]||[],u=T.event.special[e.type]||{};if((s[0]=e).delegateTarget=this,!u.preDispatch||!1!==u.preDispatch.call(this,e)){for(a=T.event.handlers.call(this,e,c),t=0;(o=a[t++])&&!e.isPropagationStopped();)for(e.currentTarget=o.elem,n=0;(i=o.handlers[n++])&&!e.isImmediatePropagationStopped();)e.namespace_re&&!e.namespace_re.test(i.namespace)||(e.handleObj=i,e.data=i.data,void 0!==(r=((T.event.special[i.origType]||{}).handle||i.handler).apply(o.elem,s))&&!1===(e.result=r)&&(e.preventDefault(),e.stopPropagation()));return u.postDispatch&&u.postDispatch.call(this,e),e.result}},handlers:function(e,t){var n,r,o,i,a=[],s=t.delegateCount,c=e.target;if(s&&c.nodeType&&(!e.button||"click"!==e.type))for(;c!==this;c=c.parentNode||this)if(!0!==c.disabled||"click"!==e.type){for(r=[],n=0;n<s;n++)void 0===r[o=(i=t[n]).selector+" "]&&(r[o]=i.needsContext?0<=T(o,this).index(c):T.find(o,this,null,[c]).length),r[o]&&r.push(i);r.length&&a.push({elem:c,handlers:r})}return s<t.length&&a.push({elem:this,handlers:t.slice(s)}),a},props:"altKey bubbles cancelable ctrlKey currentTarget eventPhase metaKey relatedTarget shiftKey target timeStamp view which".split(" "),fixHooks:{},keyHooks:{props:"char charCode key keyCode".split(" "),filter:function(e,t){return null==e.which&&(e.which=null!=t.charCode?t.charCode:t.keyCode),e}},mouseHooks:{props:"button buttons clientX clientY offsetX offsetY pageX pageY screenX screenY toElement".split(" "),filter:function(e,t){var n,r,o,i=t.button;return null==e.pageX&&null!=t.clientX&&(r=(n=e.target.ownerDocument||y).documentElement,o=n.body,e.pageX=t.clientX+(r&&r.scrollLeft||o&&o.scrollLeft||0)-(r&&r.clientLeft||o&&o.clientLeft||0),e.pageY=t.clientY+(r&&r.scrollTop||o&&o.scrollTop||0)-(r&&r.clientTop||o&&o.clientTop||0)),e.which||void 0===i||(e.which=1&i?1:2&i?3:4&i?2:0),e}},fix:function(e){if(e[T.expando])return e;var t,n,r,o=e.type,i=e,a=this.fixHooks[o];for(a||(this.fixHooks[o]=a=Z.test(o)?this.mouseHooks:X.test(o)?this.keyHooks:{}),r=a.props?this.props.concat(a.props):this.props,e=new T.Event(i),t=r.length;t--;)e[n=r[t]]=i[n];return e.target||(e.target=y),3===e.target.nodeType&&(e.target=e.target.parentNode),a.filter?a.filter(e,i):e},special:{load:{noBubble:!0},focus:{trigger:function(){return this!==Q()&&this.focus?(this.focus(),!1):void 0},delegateType:"focusin"},blur:{trigger:function(){return this===Q()&&this.blur?(this.blur(),!1):void 0},delegateType:"focusout"},click:{trigger:function(){return"checkbox"===this.type&&this.click&&T.nodeName(this,"input")?(this.click(),!1):void 0},_default:function(e){return T.nodeName(e.target,"a")}},beforeunload:{postDispatch:function(e){void 0!==e.result&&e.originalEvent&&(e.originalEvent.returnValue=e.result)}}},simulate:function(e,t,n,r){var o=T.extend(new T.Event,n,{type:e,isSimulated:!0,originalEvent:{}});r?T.event.trigger(o,null,t):T.event.dispatch.call(t,o),o.isDefaultPrevented()&&n.preventDefault()}},T.removeEvent=function(e,t,n){e.removeEventListener&&e.removeEventListener(t,n,!1)},T.Event=function(e,t){return this instanceof T.Event?(e&&e.type?(this.originalEvent=e,this.type=e.type,this.isDefaultPrevented=e.defaultPrevented||void 0===e.defaultPrevented&&!1===e.returnValue?G:K):this.type=e,t&&T.extend(this,t),this.timeStamp=e&&e.timeStamp||T.now(),void(this[T.expando]=!0)):new T.Event(e,t)},T.Event.prototype={isDefaultPrevented:K,isPropagationStopped:K,isImmediatePropagationStopped:K,preventDefault:function(){var e=this.originalEvent;this.isDefaultPrevented=G,e&&e.preventDefault&&e.preventDefault()},stopPropagation:function(){var e=this.originalEvent;this.isPropagationStopped=G,e&&e.stopPropagation&&e.stopPropagation()},stopImmediatePropagation:function(){var e=this.originalEvent;this.isImmediatePropagationStopped=G,e&&e.stopImmediatePropagation&&e.stopImmediatePropagation(),this.stopPropagation()}},T.each({mouseenter:"mouseover",mouseleave:"mouseout",pointerenter:"pointerover",pointerleave:"pointerout"},function(e,o){T.event.special[e]={delegateType:o,bindType:o,handle:function(e){var t,n=e.relatedTarget,r=e.handleObj;return n&&(n===this||T.contains(this,n))||(e.type=r.origType,t=r.handler.apply(this,arguments),e.type=o),t}}}),v.focusinBubbles||T.each({focus:"focusin",blur:"focusout"},function(n,r){function o(e){T.event.simulate(r,e.target,T.event.fix(e),!0)}T.event.special[r]={setup:function(){var e=this.ownerDocument||this,t=F.access(e,r);t||e.addEventListener(n,o,!0),F.access(e,r,(t||0)+1)},teardown:function(){var e=this.ownerDocument||this,t=F.access(e,r)-1;t?F.access(e,r,t):(e.removeEventListener(n,o,!0),F.remove(e,r))}}}),T.fn.extend({on:function(e,t,n,r,o){var i,a;if("object"==typeof e){for(a in"string"!=typeof t&&(n=n||t,t=void 0),e)this.on(a,t,n,e[a],o);return this}if(null==n&&null==r?(r=t,n=t=void 0):null==r&&("string"==typeof t?(r=n,n=void 0):(r=n,n=t,t=void 0)),!1===r)r=K;else if(!r)return this;return 1===o&&(i=r,(r=function(e){return T().off(e),i.apply(this,arguments)}).guid=i.guid||(i.guid=T.guid++)),this.each(function(){T.event.add(this,e,r,n,t)})},one:function(e,t,n,r){return this.on(e,t,n,r,1)},off:function(e,t,n){var r,o;if(e&&e.preventDefault&&e.handleObj)return r=e.handleObj,T(e.delegateTarget).off(r.namespace?r.origType+"."+r.namespace:r.origType,r.selector,r.handler),this;if("object"!=typeof e)return!1!==t&&"function"!=typeof t||(n=t,t=void 0),!1===n&&(n=K),this.each(function(){T.event.remove(this,e,n,t)});for(o in e)this.off(o,t,e[o]);return this},trigger:function(e,t){return this.each(function(){T.event.trigger(e,t,this)})},triggerHandler:function(e,t){var n=this[0];return n?T.event.trigger(e,t,n,!0):void 0}});var ee=/<(?!area|br|col|embed|hr|img|input|link|meta|param)(([\w:]+)[^>]*)\/>/gi,te=/<([\w:]+)/,ne=/<|&#?\w+;/,re=/<(?:script|style|link)/i,oe=/checked\s*(?:[^=]|=\s*.checked.)/i,ie=/^$|\/(?:java|ecma)script/i,ae=/^true\/(.*)/,se=/^\s*<!(?:\[CDATA\[|--)|(?:\]\]|--)>\s*$/g,ce={option:[1,"<select multiple='multiple'>","</select>"],thead:[1,"<table>","</table>"],col:[2,"<table><colgroup>","</colgroup></table>"],tr:[2,"<table><tbody>","</tbody></table>"],td:[3,"<table><tbody><tr>","</tr></tbody></table>"],_default:[0,"",""]};function ue(e,t){return T.nodeName(e,"table")&&T.nodeName(11!==t.nodeType?t:t.firstChild,"tr")?e.getElementsByTagName("tbody")[0]||e.appendChild(e.ownerDocument.createElement("tbody")):e}function le(e){return e.type=(null!==e.getAttribute("type"))+"/"+e.type,e}function pe(e){var t=ae.exec(e.type);return t?e.type=t[1]:e.removeAttribute("type"),e}function fe(e,t){for(var n=0,r=e.length;n<r;n++)F.set(e[n],"globalEval",!t||F.get(t[n],"globalEval"))}function de(e,t){var n,r,o,i,a,s,c,u;if(1===t.nodeType){if(F.hasData(e)&&(i=F.access(e),a=F.set(t,i),u=i.events))for(o in delete a.handle,a.events={},u)for(n=0,r=u[o].length;n<r;n++)T.event.add(t,o,u[o][n]);P.hasData(e)&&(s=P.access(e),c=T.extend({},s),P.set(t,c))}}function he(e,t){var n=e.getElementsByTagName?e.getElementsByTagName(t||"*"):e.querySelectorAll?e.querySelectorAll(t||"*"):[];return void 0===t||t&&T.nodeName(e,t)?T.merge([e],n):n}ce.optgroup=ce.option,ce.tbody=ce.tfoot=ce.colgroup=ce.caption=ce.thead,ce.th=ce.td,T.extend({clone:function(e,t,n){var r,o,i,a,s,c,u,l=e.cloneNode(!0),p=T.contains(e.ownerDocument,e);if(!(v.noCloneChecked||1!==e.nodeType&&11!==e.nodeType||T.isXMLDoc(e)))for(a=he(l),r=0,o=(i=he(e)).length;r<o;r++)s=i[r],c=a[r],void 0,"input"===(u=c.nodeName.toLowerCase())&&V.test(s.type)?c.checked=s.checked:"input"!==u&&"textarea"!==u||(c.defaultValue=s.defaultValue);if(t)if(n)for(i=i||he(e),a=a||he(l),r=0,o=i.length;r<o;r++)de(i[r],a[r]);else de(e,l);return 0<(a=he(l,"script")).length&&fe(a,!p&&he(e,"script")),l},buildFragment:function(e,t,n,r){for(var o,i,a,s,c,u,l=t.createDocumentFragment(),p=[],f=0,d=e.length;f<d;f++)if((o=e[f])||0===o)if("object"===T.type(o))T.merge(p,o.nodeType?[o]:o);else if(ne.test(o)){for(i=i||l.appendChild(t.createElement("div")),a=(te.exec(o)||["",""])[1].toLowerCase(),s=ce[a]||ce._default,i.innerHTML=s[1]+o.replace(ee,"<$1></$2>")+s[2],u=s[0];u--;)i=i.lastChild;T.merge(p,i.childNodes),(i=l.firstChild).textContent=""}else p.push(t.createTextNode(o));for(l.textContent="",f=0;o=p[f++];)if((!r||-1===T.inArray(o,r))&&(c=T.contains(o.ownerDocument,o),i=he(l.appendChild(o),"script"),c&&fe(i),n))for(u=0;o=i[u++];)ie.test(o.type||"")&&n.push(o);return l},cleanData:function(e){for(var t,n,r,o,i=T.event.special,a=0;void 0!==(n=e[a]);a++){if(T.acceptData(n)&&((o=n[F.expando])&&(t=F.cache[o]))){if(t.events)for(r in t.events)i[r]?T.event.remove(n,r):T.removeEvent(n,r,t.handle);F.cache[o]&&delete F.cache[o]}delete P.cache[n[P.expando]]}}}),T.fn.extend({text:function(e){return D(this,function(e){return void 0===e?T.text(this):this.empty().each(function(){1!==this.nodeType&&11!==this.nodeType&&9!==this.nodeType||(this.textContent=e)})},null,e,arguments.length)},append:function(){return this.domManip(arguments,function(e){1!==this.nodeType&&11!==this.nodeType&&9!==this.nodeType||ue(this,e).appendChild(e)})},prepend:function(){return this.domManip(arguments,function(e){if(1===this.nodeType||11===this.nodeType||9===this.nodeType){var t=ue(this,e);t.insertBefore(e,t.firstChild)}})},before:function(){return this.domManip(arguments,function(e){this.parentNode&&this.parentNode.insertBefore(e,this)})},after:function(){return this.domManip(arguments,function(e){this.parentNode&&this.parentNode.insertBefore(e,this.nextSibling)})},remove:function(e,t){for(var n,r=e?T.filter(e,this):this,o=0;null!=(n=r[o]);o++)t||1!==n.nodeType||T.cleanData(he(n)),n.parentNode&&(t&&T.contains(n.ownerDocument,n)&&fe(he(n,"script")),n.parentNode.removeChild(n));return this},empty:function(){for(var e,t=0;null!=(e=this[t]);t++)1===e.nodeType&&(T.cleanData(he(e,!1)),e.textContent="");return this},clone:function(e,t){return e=null!=e&&e,t=null==t?e:t,this.map(function(){return T.clone(this,e,t)})},html:function(e){return D(this,function(e){var t=this[0]||{},n=0,r=this.length;if(void 0===e&&1===t.nodeType)return t.innerHTML;if("string"==typeof e&&!re.test(e)&&!ce[(te.exec(e)||["",""])[1].toLowerCase()]){e=e.replace(ee,"<$1></$2>");try{for(;n<r;n++)1===(t=this[n]||{}).nodeType&&(T.cleanData(he(t,!1)),t.innerHTML=e);t=0}catch(e){}}t&&this.empty().append(e)},null,e,arguments.length)},replaceWith:function(){var t=arguments[0];return this.domManip(arguments,function(e){t=this.parentNode,T.cleanData(he(this)),t&&t.replaceChild(e,this)}),t&&(t.length||t.nodeType)?this:this.remove()},detach:function(e){return this.remove(e,!0)},domManip:function(n,r){n=g.apply([],n);var e,t,o,i,a,s,c=0,u=this.length,l=this,p=u-1,f=n[0],d=T.isFunction(f);if(d||1<u&&"string"==typeof f&&!v.checkClone&&oe.test(f))return this.each(function(e){var t=l.eq(e);d&&(n[0]=f.call(this,e,t.html())),t.domManip(n,r)});if(u&&(t=(e=T.buildFragment(n,this[0].ownerDocument,!1,this)).firstChild,1===e.childNodes.length&&(e=t),t)){for(i=(o=T.map(he(e,"script"),le)).length;c<u;c++)a=e,c!==p&&(a=T.clone(a,!0,!0),i&&T.merge(o,he(a,"script"))),r.call(this[c],a,c);if(i)for(s=o[o.length-1].ownerDocument,T.map(o,pe),c=0;c<i;c++)a=o[c],ie.test(a.type||"")&&!F.access(a,"globalEval")&&T.contains(s,a)&&(a.src?T._evalUrl&&T._evalUrl(a.src):T.globalEval(a.textContent.replace(se,"")))}return this}}),T.each({appendTo:"append",prependTo:"prepend",insertBefore:"before",insertAfter:"after",replaceAll:"replaceWith"},function(e,a){T.fn[e]=function(e){for(var t,n=[],r=T(e),o=r.length-1,i=0;i<=o;i++)t=i===o?this:this.clone(!0),T(r[i])[a](t),s.apply(n,t.get());return this.pushStack(n)}});var ge,me={};function ve(e,t){var n,r=T(t.createElement(e)).appendTo(t.body),o=h.getDefaultComputedStyle&&(n=h.getDefaultComputedStyle(r[0]))?n.display:T.css(r[0],"display");return r.detach(),o}function ye(e){var t=y,n=me[e];return n||("none"!==(n=ve(e,t))&&n||((t=(ge=(ge||T("<iframe frameborder='0' width='0' height='0'/>")).appendTo(t.documentElement))[0].contentDocument).write(),t.close(),n=ve(e,t),ge.detach()),me[e]=n),n}var xe=/^margin/,be=new RegExp("^("+W+")(?!px)[a-z%]+$","i"),we=function(e){return e.ownerDocument.defaultView&&e.ownerDocument.defaultView.opener?e.ownerDocument.defaultView.getComputedStyle(e,null):h.getComputedStyle(e,null)};function ke(e,t,n){var r,o,i,a,s=e.style;return(n=n||we(e))&&(a=n.getPropertyValue(t)||n[t]),n&&(""!==a||T.contains(e.ownerDocument,e)||(a=T.style(e,t)),be.test(a)&&xe.test(t)&&(r=s.width,o=s.minWidth,i=s.maxWidth,s.minWidth=s.maxWidth=s.width=a,a=n.width,s.width=r,s.minWidth=o,s.maxWidth=i)),void 0!==a?a+"":a}function Te(e,t){return{get:function(){return e()?void delete this.get:(this.get=t).apply(this,arguments)}}}!function(){var t,n,r=y.documentElement,o=y.createElement("div"),i=y.createElement("div");if(i.style){function e(){i.style.cssText="-webkit-box-sizing:border-box;-moz-box-sizing:border-box;box-sizing:border-box;display:block;margin-top:1%;top:1%;border:1px;padding:1px;width:4px;position:absolute",i.innerHTML="",r.appendChild(o);var e=h.getComputedStyle(i,null);t="1%"!==e.top,n="4px"===e.width,r.removeChild(o)}i.style.backgroundClip="content-box",i.cloneNode(!0).style.backgroundClip="",v.clearCloneStyle="content-box"===i.style.backgroundClip,o.style.cssText="border:0;width:0;height:0;top:0;left:-9999px;margin-top:1px;position:absolute",o.appendChild(i),h.getComputedStyle&&T.extend(v,{pixelPosition:function(){return e(),t},boxSizingReliable:function(){return null==n&&e(),n},reliableMarginRight:function(){var e,t=i.appendChild(y.createElement("div"));return t.style.cssText=i.style.cssText="-webkit-box-sizing:content-box;-moz-box-sizing:content-box;box-sizing:content-box;display:block;margin:0;border:0;padding:0",t.style.marginRight=t.style.width="0",i.style.width="1px",r.appendChild(o),e=!parseFloat(h.getComputedStyle(t,null).marginRight),r.removeChild(o),i.removeChild(t),e}})}}(),T.swap=function(e,t,n,r){var o,i,a={};for(i in t)a[i]=e.style[i],e.style[i]=t[i];for(i in o=n.apply(e,r||[]),t)e.style[i]=a[i];return o};var Ce=/^(none|table(?!-c[ea]).+)/,Se=new RegExp("^("+W+")(.*)$","i"),_e=new RegExp("^([+-])=("+W+")","i"),je={position:"absolute",visibility:"hidden",display:"block"},$e={letterSpacing:"0",fontWeight:"400"},Ae=["Webkit","O","Moz","ms"];function Ee(e,t){if(t in e)return t;for(var n=t[0].toUpperCase()+t.slice(1),r=t,o=Ae.length;o--;)if((t=Ae[o]+n)in e)return t;return r}function Ne(e,t,n){var r=Se.exec(t);return r?Math.max(0,r[1]-(n||0))+(r[2]||"px"):t}function Oe(e,t,n,r,o){for(var i=n===(r?"border":"content")?4:"width"===t?1:0,a=0;i<4;i+=2)"margin"===n&&(a+=T.css(e,n+R[i],!0,o)),r?("content"===n&&(a-=T.css(e,"padding"+R[i],!0,o)),"margin"!==n&&(a-=T.css(e,"border"+R[i]+"Width",!0,o))):(a+=T.css(e,"padding"+R[i],!0,o),"padding"!==n&&(a+=T.css(e,"border"+R[i]+"Width",!0,o)));return a}function De(e,t,n){var r=!0,o="width"===t?e.offsetWidth:e.offsetHeight,i=we(e),a="border-box"===T.css(e,"boxSizing",!1,i);if(o<=0||null==o){if(((o=ke(e,t,i))<0||null==o)&&(o=e.style[t]),be.test(o))return o;r=a&&(v.boxSizingReliable()||o===e.style[t]),o=parseFloat(o)||0}return o+Oe(e,t,n||(a?"border":"content"),r,i)+"px"}function Le(e,t){for(var n,r,o,i=[],a=0,s=e.length;a<s;a++)(r=e[a]).style&&(i[a]=F.get(r,"olddisplay"),n=r.style.display,t?(i[a]||"none"!==n||(r.style.display=""),""===r.style.display&&H(r)&&(i[a]=F.access(r,"olddisplay",ye(r.nodeName)))):(o=H(r),"none"===n&&o||F.set(r,"olddisplay",o?n:T.css(r,"display"))));for(a=0;a<s;a++)(r=e[a]).style&&(t&&"none"!==r.style.display&&""!==r.style.display||(r.style.display=t?i[a]||"":"none"));return e}function Fe(e,t,n,r,o){return new Fe.prototype.init(e,t,n,r,o)}T.extend({cssHooks:{opacity:{get:function(e,t){if(t){var n=ke(e,"opacity");return""===n?"1":n}}}},cssNumber:{columnCount:!0,fillOpacity:!0,flexGrow:!0,flexShrink:!0,fontWeight:!0,lineHeight:!0,opacity:!0,order:!0,orphans:!0,widows:!0,zIndex:!0,zoom:!0},cssProps:{float:"cssFloat"},style:function(e,t,n,r){if(e&&3!==e.nodeType&&8!==e.nodeType&&e.style){var o,i,a,s=T.camelCase(t),c=e.style;return t=T.cssProps[s]||(T.cssProps[s]=Ee(c,s)),a=T.cssHooks[t]||T.cssHooks[s],void 0===n?a&&"get"in a&&void 0!==(o=a.get(e,!1,r))?o:c[t]:("string"===(i=typeof n)&&(o=_e.exec(n))&&(n=(o[1]+1)*o[2]+parseFloat(T.css(e,t)),i="number"),void(null!=n&&n==n&&("number"!==i||T.cssNumber[s]||(n+="px"),v.clearCloneStyle||""!==n||0!==t.indexOf("background")||(c[t]="inherit"),a&&"set"in a&&void 0===(n=a.set(e,n,r))||(c[t]=n))))}},css:function(e,t,n,r){var o,i,a,s=T.camelCase(t);return t=T.cssProps[s]||(T.cssProps[s]=Ee(e.style,s)),(a=T.cssHooks[t]||T.cssHooks[s])&&"get"in a&&(o=a.get(e,!0,n)),void 0===o&&(o=ke(e,t,r)),"normal"===o&&t in $e&&(o=$e[t]),""===n||n?(i=parseFloat(o),!0===n||T.isNumeric(i)?i||0:o):o}}),T.each(["height","width"],function(e,o){T.cssHooks[o]={get:function(e,t,n){return t?Ce.test(T.css(e,"display"))&&0===e.offsetWidth?T.swap(e,je,function(){return De(e,o,n)}):De(e,o,n):void 0},set:function(e,t,n){var r=n&&we(e);return Ne(0,t,n?Oe(e,o,n,"border-box"===T.css(e,"boxSizing",!1,r),r):0)}}}),T.cssHooks.marginRight=Te(v.reliableMarginRight,function(e,t){return t?T.swap(e,{display:"inline-block"},ke,[e,"marginRight"]):void 0}),T.each({margin:"",padding:"",border:"Width"},function(o,i){T.cssHooks[o+i]={expand:function(e){for(var t=0,n={},r="string"==typeof e?e.split(" "):[e];t<4;t++)n[o+R[t]+i]=r[t]||r[t-2]||r[0];return n}},xe.test(o)||(T.cssHooks[o+i].set=Ne)}),T.fn.extend({css:function(e,t){return D(this,function(e,t,n){var r,o,i={},a=0;if(T.isArray(t)){for(r=we(e),o=t.length;a<o;a++)i[t[a]]=T.css(e,t[a],!1,r);return i}return void 0!==n?T.style(e,t,n):T.css(e,t)},e,t,1<arguments.length)},show:function(){return Le(this,!0)},hide:function(){return Le(this)},toggle:function(e){return"boolean"==typeof e?e?this.show():this.hide():this.each(function(){H(this)?T(this).show():T(this).hide()})}}),((T.Tween=Fe).prototype={constructor:Fe,init:function(e,t,n,r,o,i){this.elem=e,this.prop=n,this.easing=o||"swing",this.options=t,this.start=this.now=this.cur(),this.end=r,this.unit=i||(T.cssNumber[n]?"":"px")},cur:function(){var e=Fe.propHooks[this.prop];return e&&e.get?e.get(this):Fe.propHooks._default.get(this)},run:function(e){var t,n=Fe.propHooks[this.prop];return this.options.duration?this.pos=t=T.easing[this.easing](e,this.options.duration*e,0,1,this.options.duration):this.pos=t=e,this.now=(this.end-this.start)*t+this.start,this.options.step&&this.options.step.call(this.elem,this.now,this),n&&n.set?n.set(this):Fe.propHooks._default.set(this),this}}).init.prototype=Fe.prototype,(Fe.propHooks={_default:{get:function(e){var t;return null==e.elem[e.prop]||e.elem.style&&null!=e.elem.style[e.prop]?(t=T.css(e.elem,e.prop,""))&&"auto"!==t?t:0:e.elem[e.prop]},set:function(e){T.fx.step[e.prop]?T.fx.step[e.prop](e):e.elem.style&&(null!=e.elem.style[T.cssProps[e.prop]]||T.cssHooks[e.prop])?T.style(e.elem,e.prop,e.now+e.unit):e.elem[e.prop]=e.now}}}).scrollTop=Fe.propHooks.scrollLeft={set:function(e){e.elem.nodeType&&e.elem.parentNode&&(e.elem[e.prop]=e.now)}},T.easing={linear:function(e){return e},swing:function(e){return.5-Math.cos(e*Math.PI)/2}},T.fx=Fe.prototype.init,T.fx.step={};var Pe,Ie,qe,ze,He,Me=/^(?:toggle|show|hide)$/,Be=new RegExp("^(?:([+-])=|)("+W+")([a-z%]*)$","i"),We=/queueHooks$/,Re=[function(t,e,n){var r,o,i,a,s,c,u,l=this,p={},f=t.style,d=t.nodeType&&H(t),h=F.get(t,"fxshow");for(r in n.queue||(null==(s=T._queueHooks(t,"fx")).unqueued&&(s.unqueued=0,c=s.empty.fire,s.empty.fire=function(){s.unqueued||c()}),s.unqueued++,l.always(function(){l.always(function(){s.unqueued--,T.queue(t,"fx").length||s.empty.fire()})})),1===t.nodeType&&("height"in e||"width"in e)&&(n.overflow=[f.overflow,f.overflowX,f.overflowY],u=T.css(t,"display"),"inline"===("none"===u?F.get(t,"olddisplay")||ye(t.nodeName):u)&&"none"===T.css(t,"float")&&(f.display="inline-block")),n.overflow&&(f.overflow="hidden",l.always(function(){f.overflow=n.overflow[0],f.overflowX=n.overflow[1],f.overflowY=n.overflow[2]})),e)if(o=e[r],Me.exec(o)){if(delete e[r],i=i||"toggle"===o,o===(d?"hide":"show")){if("show"!==o||!h||void 0===h[r])continue;d=!0}p[r]=h&&h[r]||T.style(t,r)}else u=void 0;if(T.isEmptyObject(p))"inline"===("none"===u?ye(t.nodeName):u)&&(f.display=u);else for(r in h?"hidden"in h&&(d=h.hidden):h=F.access(t,"fxshow",{}),i&&(h.hidden=!d),d?T(t).show():l.done(function(){T(t).hide()}),l.done(function(){var e;for(e in F.remove(t,"fxshow"),p)T.style(t,e,p[e])}),p)a=Ze(d?h[r]:0,r,l),r in h||(h[r]=a.start,d&&(a.end=a.start,a.start="width"===r||"height"===r?1:0))}],Ve={"*":[function(e,t){var n=this.createTween(e,t),r=n.cur(),o=Be.exec(t),i=o&&o[3]||(T.cssNumber[e]?"":"px"),a=(T.cssNumber[e]||"px"!==i&&+r)&&Be.exec(T.css(n.elem,e)),s=1,c=20;if(a&&a[3]!==i)for(i=i||a[3],o=o||[],a=+r||1;a/=s=s||".5",T.style(n.elem,e,a+i),s!==(s=n.cur()/r)&&1!==s&&--c;);return o&&(a=n.start=+a||+r||0,n.unit=i,n.end=o[1]?a+(o[1]+1)*o[2]:+o[2]),n}]};function Ue(){return setTimeout(function(){Pe=void 0}),Pe=T.now()}function Xe(e,t){var n,r=0,o={height:e};for(t=t?1:0;r<4;r+=2-t)o["margin"+(n=R[r])]=o["padding"+n]=e;return t&&(o.opacity=o.width=e),o}function Ze(e,t,n){for(var r,o=(Ve[t]||[]).concat(Ve["*"]),i=0,a=o.length;i<a;i++)if(r=o[i].call(n,t,e))return r}function Je(i,e,t){var n,a,r=0,o=Re.length,s=T.Deferred().always(function(){delete c.elem}),c=function(){if(a)return!1;for(var e=Pe||Ue(),t=Math.max(0,u.startTime+u.duration-e),n=1-(t/u.duration||0),r=0,o=u.tweens.length;r<o;r++)u.tweens[r].run(n);return s.notifyWith(i,[u,n,t]),n<1&&o?t:(s.resolveWith(i,[u]),!1)},u=s.promise({elem:i,props:T.extend({},e),opts:T.extend(!0,{specialEasing:{}},t),originalProperties:e,originalOptions:t,startTime:Pe||Ue(),duration:t.duration,tweens:[],createTween:function(e,t){var n=T.Tween(i,u.opts,e,t,u.opts.specialEasing[e]||u.opts.easing);return u.tweens.push(n),n},stop:function(e){var t=0,n=e?u.tweens.length:0;if(a)return this;for(a=!0;t<n;t++)u.tweens[t].run(1);return e?s.resolveWith(i,[u,e]):s.rejectWith(i,[u,e]),this}}),l=u.props;for(function(e,t){var n,r,o,i,a;for(n in e)if(o=t[r=T.camelCase(n)],i=e[n],T.isArray(i)&&(o=i[1],i=e[n]=i[0]),n!==r&&(e[r]=i,delete e[n]),(a=T.cssHooks[r])&&"expand"in a)for(n in i=a.expand(i),delete e[r],i)n in e||(e[n]=i[n],t[n]=o);else t[r]=o}(l,u.opts.specialEasing);r<o;r++)if(n=Re[r].call(u,i,l,u.opts))return n;return T.map(l,Ze,u),T.isFunction(u.opts.start)&&u.opts.start.call(i,u),T.fx.timer(T.extend(c,{elem:i,anim:u,queue:u.opts.queue})),u.progress(u.opts.progress).done(u.opts.done,u.opts.complete).fail(u.opts.fail).always(u.opts.always)}T.Animation=T.extend(Je,{tweener:function(e,t){for(var n,r=0,o=(e=T.isFunction(e)?(t=e,["*"]):e.split(" ")).length;r<o;r++)n=e[r],Ve[n]=Ve[n]||[],Ve[n].unshift(t)},prefilter:function(e,t){t?Re.unshift(e):Re.push(e)}}),T.speed=function(e,t,n){var r=e&&"object"==typeof e?T.extend({},e):{complete:n||!n&&t||T.isFunction(e)&&e,duration:e,easing:n&&t||t&&!T.isFunction(t)&&t};return r.duration=T.fx.off?0:"number"==typeof r.duration?r.duration:r.duration in T.fx.speeds?T.fx.speeds[r.duration]:T.fx.speeds._default,null!=r.queue&&!0!==r.queue||(r.queue="fx"),r.old=r.complete,r.complete=function(){T.isFunction(r.old)&&r.old.call(this),r.queue&&T.dequeue(this,r.queue)},r},T.fn.extend({fadeTo:function(e,t,n,r){return this.filter(H).css("opacity",0).show().end().animate({opacity:t},e,n,r)},animate:function(t,e,n,r){function o(){var e=Je(this,T.extend({},t),a);(i||F.get(this,"finish"))&&e.stop(!0)}var i=T.isEmptyObject(t),a=T.speed(e,n,r);return o.finish=o,i||!1===a.queue?this.each(o):this.queue(a.queue,o)},stop:function(o,e,i){function a(e){var t=e.stop;delete e.stop,t(i)}return"string"!=typeof o&&(i=e,e=o,o=void 0),e&&!1!==o&&this.queue(o||"fx",[]),this.each(function(){var e=!0,t=null!=o&&o+"queueHooks",n=T.timers,r=F.get(this);if(t)r[t]&&r[t].stop&&a(r[t]);else for(t in r)r[t]&&r[t].stop&&We.test(t)&&a(r[t]);for(t=n.length;t--;)n[t].elem!==this||null!=o&&n[t].queue!==o||(n[t].anim.stop(i),e=!1,n.splice(t,1));!e&&i||T.dequeue(this,o)})},finish:function(a){return!1!==a&&(a=a||"fx"),this.each(function(){var e,t=F.get(this),n=t[a+"queue"],r=t[a+"queueHooks"],o=T.timers,i=n?n.length:0;for(t.finish=!0,T.queue(this,a,[]),r&&r.stop&&r.stop.call(this,!0),e=o.length;e--;)o[e].elem===this&&o[e].queue===a&&(o[e].anim.stop(!0),o.splice(e,1));for(e=0;e<i;e++)n[e]&&n[e].finish&&n[e].finish.call(this);delete t.finish})}}),T.each(["toggle","show","hide"],function(e,r){var o=T.fn[r];T.fn[r]=function(e,t,n){return null==e||"boolean"==typeof e?o.apply(this,arguments):this.animate(Xe(r,!0),e,t,n)}}),T.each({slideDown:Xe("show"),slideUp:Xe("hide"),slideToggle:Xe("toggle"),fadeIn:{opacity:"show"},fadeOut:{opacity:"hide"},fadeToggle:{opacity:"toggle"}},function(e,r){T.fn[e]=function(e,t,n){return this.animate(r,e,t,n)}}),T.timers=[],T.fx.tick=function(){var e,t=0,n=T.timers;for(Pe=T.now();t<n.length;t++)(e=n[t])()||n[t]!==e||n.splice(t--,1);n.length||T.fx.stop(),Pe=void 0},T.fx.timer=function(e){T.timers.push(e),e()?T.fx.start():T.timers.pop()},T.fx.interval=13,T.fx.start=function(){Ie||(Ie=setInterval(T.fx.tick,T.fx.interval))},T.fx.stop=function(){clearInterval(Ie),Ie=null},T.fx.speeds={slow:600,fast:200,_default:400},T.fn.delay=function(r,e){return r=T.fx&&T.fx.speeds[r]||r,e=e||"fx",this.queue(e,function(e,t){var n=setTimeout(e,r);t.stop=function(){clearTimeout(n)}})},qe=y.createElement("input"),ze=y.createElement("select"),He=ze.appendChild(y.createElement("option")),qe.type="checkbox",v.checkOn=""!==qe.value,v.optSelected=He.selected,ze.disabled=!0,v.optDisabled=!He.disabled,(qe=y.createElement("input")).value="t",qe.type="radio",v.radioValue="t"===qe.value;var Ye,Ge=T.expr.attrHandle;T.fn.extend({attr:function(e,t){return D(this,T.attr,e,t,1<arguments.length)},removeAttr:function(e){return this.each(function(){T.removeAttr(this,e)})}}),T.extend({attr:function(e,t,n){var r,o,i=e.nodeType;if(e&&3!==i&&8!==i&&2!==i)return typeof e.getAttribute==U?T.prop(e,t,n):(1===i&&T.isXMLDoc(e)||(t=t.toLowerCase(),r=T.attrHooks[t]||(T.expr.match.bool.test(t)?Ye:void 0)),void 0===n?r&&"get"in r&&null!==(o=r.get(e,t))?o:null==(o=T.find.attr(e,t))?void 0:o:null!==n?r&&"set"in r&&void 0!==(o=r.set(e,n,t))?o:(e.setAttribute(t,n+""),n):void T.removeAttr(e,t))},removeAttr:function(e,t){var n,r,o=0,i=t&&t.match(E);if(i&&1===e.nodeType)for(;n=i[o++];)r=T.propFix[n]||n,T.expr.match.bool.test(n)&&(e[r]=!1),e.removeAttribute(n)},attrHooks:{type:{set:function(e,t){if(!v.radioValue&&"radio"===t&&T.nodeName(e,"input")){var n=e.value;return e.setAttribute("type",t),n&&(e.value=n),t}}}}}),Ye={set:function(e,t,n){return!1===t?T.removeAttr(e,n):e.setAttribute(n,n),n}},T.each(T.expr.match.bool.source.match(/\w+/g),function(e,t){var i=Ge[t]||T.find.attr;Ge[t]=function(e,t,n){var r,o;return n||(o=Ge[t],Ge[t]=r,r=null!=i(e,t,n)?t.toLowerCase():null,Ge[t]=o),r}});var Ke=/^(?:input|select|textarea|button)$/i;T.fn.extend({prop:function(e,t){return D(this,T.prop,e,t,1<arguments.length)},removeProp:function(e){return this.each(function(){delete this[T.propFix[e]||e]})}}),T.extend({propFix:{for:"htmlFor",class:"className"},prop:function(e,t,n){var r,o,i=e.nodeType;if(e&&3!==i&&8!==i&&2!==i)return(1!==i||!T.isXMLDoc(e))&&(t=T.propFix[t]||t,o=T.propHooks[t]),void 0!==n?o&&"set"in o&&void 0!==(r=o.set(e,n,t))?r:e[t]=n:o&&"get"in o&&null!==(r=o.get(e,t))?r:e[t]},propHooks:{tabIndex:{get:function(e){return e.hasAttribute("tabindex")||Ke.test(e.nodeName)||e.href?e.tabIndex:-1}}}}),v.optSelected||(T.propHooks.selected={get:function(e){var t=e.parentNode;return t&&t.parentNode&&t.parentNode.selectedIndex,null}}),T.each(["tabIndex","readOnly","maxLength","cellSpacing","cellPadding","rowSpan","colSpan","useMap","frameBorder","contentEditable"],function(){T.propFix[this.toLowerCase()]=this});var Qe=/[\t\r\n\f]/g;T.fn.extend({addClass:function(t){var e,n,r,o,i,a,s="string"==typeof t&&t,c=0,u=this.length;if(T.isFunction(t))return this.each(function(e){T(this).addClass(t.call(this,e,this.className))});if(s)for(e=(t||"").match(E)||[];c<u;c++)if(r=1===(n=this[c]).nodeType&&(n.className?(" "+n.className+" ").replace(Qe," "):" ")){for(i=0;o=e[i++];)r.indexOf(" "+o+" ")<0&&(r+=o+" ");a=T.trim(r),n.className!==a&&(n.className=a)}return this},removeClass:function(t){var e,n,r,o,i,a,s=0===arguments.length||"string"==typeof t&&t,c=0,u=this.length;if(T.isFunction(t))return this.each(function(e){T(this).removeClass(t.call(this,e,this.className))});if(s)for(e=(t||"").match(E)||[];c<u;c++)if(r=1===(n=this[c]).nodeType&&(n.className?(" "+n.className+" ").replace(Qe," "):"")){for(i=0;o=e[i++];)for(;0<=r.indexOf(" "+o+" ");)r=r.replace(" "+o+" "," ");a=t?T.trim(r):"",n.className!==a&&(n.className=a)}return this},toggleClass:function(o,t){var i=typeof o;return"boolean"==typeof t&&"string"==i?t?this.addClass(o):this.removeClass(o):this.each(T.isFunction(o)?function(e){T(this).toggleClass(o.call(this,e,this.className,t),t)}:function(){if("string"==i)for(var e,t=0,n=T(this),r=o.match(E)||[];e=r[t++];)n.hasClass(e)?n.removeClass(e):n.addClass(e);else i!=U&&"boolean"!=i||(this.className&&F.set(this,"__className__",this.className),this.className=this.className||!1===o?"":F.get(this,"__className__")||"")})},hasClass:function(e){for(var t=" "+e+" ",n=0,r=this.length;n<r;n++)if(1===this[n].nodeType&&0<=(" "+this[n].className+" ").replace(Qe," ").indexOf(t))return!0;return!1}});var et=/\r/g;T.fn.extend({val:function(n){var r,e,o,t=this[0];return arguments.length?(o=T.isFunction(n),this.each(function(e){var t;1===this.nodeType&&(null==(t=o?n.call(this,e,T(this).val()):n)?t="":"number"==typeof t?t+="":T.isArray(t)&&(t=T.map(t,function(e){return null==e?"":e+""})),(r=T.valHooks[this.type]||T.valHooks[this.nodeName.toLowerCase()])&&"set"in r&&void 0!==r.set(this,t,"value")||(this.value=t))})):t?(r=T.valHooks[t.type]||T.valHooks[t.nodeName.toLowerCase()])&&"get"in r&&void 0!==(e=r.get(t,"value"))?e:"string"==typeof(e=t.value)?e.replace(et,""):null==e?"":e:void 0}}),T.extend({valHooks:{option:{get:function(e){var t=T.find.attr(e,"value");return null!=t?t:T.trim(T.text(e))}},select:{get:function(e){for(var t,n,r=e.options,o=e.selectedIndex,i="select-one"===e.type||o<0,a=i?null:[],s=i?o+1:r.length,c=o<0?s:i?o:0;c<s;c++)if(!(!(n=r[c]).selected&&c!==o||(v.optDisabled?n.disabled:null!==n.getAttribute("disabled"))||n.parentNode.disabled&&T.nodeName(n.parentNode,"optgroup"))){if(t=T(n).val(),i)return t;a.push(t)}return a},set:function(e,t){for(var n,r,o=e.options,i=T.makeArray(t),a=o.length;a--;)((r=o[a]).selected=0<=T.inArray(r.value,i))&&(n=!0);return n||(e.selectedIndex=-1),i}}}}),T.each(["radio","checkbox"],function(){T.valHooks[this]={set:function(e,t){return T.isArray(t)?e.checked=0<=T.inArray(T(e).val(),t):void 0}},v.checkOn||(T.valHooks[this].get=function(e){return null===e.getAttribute("value")?"on":e.value})}),T.each("blur focus focusin focusout load resize scroll unload click dblclick mousedown mouseup mousemove mouseover mouseout mouseenter mouseleave change select submit keydown keypress keyup error contextmenu".split(" "),function(e,n){T.fn[n]=function(e,t){return 0<arguments.length?this.on(n,null,e,t):this.trigger(n)}}),T.fn.extend({hover:function(e,t){return this.mouseenter(e).mouseleave(t||e)},bind:function(e,t,n){return this.on(e,null,t,n)},unbind:function(e,t){return this.off(e,null,t)},delegate:function(e,t,n,r){return this.on(t,e,n,r)},undelegate:function(e,t,n){return 1===arguments.length?this.off(e,"**"):this.off(t,e||"**",n)}});var tt=T.now(),nt=/\?/;T.parseJSON=function(e){return JSON.parse(e+"")},T.parseXML=function(e){var t;if(!e||"string"!=typeof e)return null;try{t=(new DOMParser).parseFromString(e,"text/xml")}catch(e){t=void 0}return t&&!t.getElementsByTagName("parsererror").length||T.error("Invalid XML: "+e),t};var rt=/#.*$/,ot=/([?&])_=[^&]*/,it=/^(.*?):[ \t]*([^\r\n]*)$/gm,at=/^(?:GET|HEAD)$/,st=/^\/\//,ct=/^([\w.+-]+:)(?:\/\/(?:[^\/?#]*@|)([^\/?#:]*)(?::(\d+)|)|)/,ut={},lt={},pt="*/".concat("*"),ft=h.location.href,dt=ct.exec(ft.toLowerCase())||[];function ht(i){return function(e,t){"string"!=typeof e&&(t=e,e="*");var n,r=0,o=e.toLowerCase().match(E)||[];if(T.isFunction(t))for(;n=o[r++];)"+"===n[0]?(n=n.slice(1)||"*",(i[n]=i[n]||[]).unshift(t)):(i[n]=i[n]||[]).push(t)}}function gt(t,o,i,a){var s={},c=t===lt;function u(e){var r;return s[e]=!0,T.each(t[e]||[],function(e,t){var n=t(o,i,a);return"string"!=typeof n||c||s[n]?c?!(r=n):void 0:(o.dataTypes.unshift(n),u(n),!1)}),r}return u(o.dataTypes[0])||!s["*"]&&u("*")}function mt(e,t){var n,r,o=T.ajaxSettings.flatOptions||{};for(n in t)void 0!==t[n]&&((o[n]?e:r||(r={}))[n]=t[n]);return r&&T.extend(!0,e,r),e}T.extend({active:0,lastModified:{},etag:{},ajaxSettings:{url:ft,type:"GET",isLocal:/^(?:about|app|app-storage|.+-extension|file|res|widget):$/.test(dt[1]),global:!0,processData:!0,async:!0,contentType:"application/x-www-form-urlencoded; charset=UTF-8",accepts:{"*":pt,text:"text/plain",html:"text/html",xml:"application/xml, text/xml",json:"application/json, text/javascript"},contents:{xml:/xml/,html:/html/,json:/json/},responseFields:{xml:"responseXML",text:"responseText",json:"responseJSON"},converters:{"* text":String,"text html":!0,"text json":T.parseJSON,"text xml":T.parseXML},flatOptions:{url:!0,context:!0}},ajaxSetup:function(e,t){return t?mt(mt(e,T.ajaxSettings),t):mt(T.ajaxSettings,e)},ajaxPrefilter:ht(ut),ajaxTransport:ht(lt),ajax:function(e,t){"object"==typeof e&&(t=e,e=void 0),t=t||{};var l,p,f,n,d,r,h,o,g=T.ajaxSetup({},t),m=g.context||g,v=g.context&&(m.nodeType||m.jquery)?T(m):T.event,y=T.Deferred(),x=T.Callbacks("once memory"),b=g.statusCode||{},i={},a={},w=0,s="canceled",k={readyState:0,getResponseHeader:function(e){var t;if(2===w){if(!n)for(n={};t=it.exec(f);)n[t[1].toLowerCase()]=t[2];t=n[e.toLowerCase()]}return null==t?null:t},getAllResponseHeaders:function(){return 2===w?f:null},setRequestHeader:function(e,t){var n=e.toLowerCase();return w||(e=a[n]=a[n]||e,i[e]=t),this},overrideMimeType:function(e){return w||(g.mimeType=e),this},statusCode:function(e){var t;if(e)if(w<2)for(t in e)b[t]=[b[t],e[t]];else k.always(e[k.status]);return this},abort:function(e){var t=e||s;return l&&l.abort(t),c(0,t),this}};if(y.promise(k).complete=x.add,k.success=k.done,k.error=k.fail,g.url=((e||g.url||ft)+"").replace(rt,"").replace(st,dt[1]+"//"),g.type=t.method||t.type||g.method||g.type,g.dataTypes=T.trim(g.dataType||"*").toLowerCase().match(E)||[""],null==g.crossDomain&&(r=ct.exec(g.url.toLowerCase()),g.crossDomain=!(!r||r[1]===dt[1]&&r[2]===dt[2]&&(r[3]||("http:"===r[1]?"80":"443"))===(dt[3]||("http:"===dt[1]?"80":"443")))),g.data&&g.processData&&"string"!=typeof g.data&&(g.data=T.param(g.data,g.traditional)),gt(ut,g,t,k),2===w)return k;for(o in(h=T.event&&g.global)&&0==T.active++&&T.event.trigger("ajaxStart"),g.type=g.type.toUpperCase(),g.hasContent=!at.test(g.type),p=g.url,g.hasContent||(g.data&&(p=g.url+=(nt.test(p)?"&":"?")+g.data,delete g.data),!1===g.cache&&(g.url=ot.test(p)?p.replace(ot,"$1_="+tt++):p+(nt.test(p)?"&":"?")+"_="+tt++)),g.ifModified&&(T.lastModified[p]&&k.setRequestHeader("If-Modified-Since",T.lastModified[p]),T.etag[p]&&k.setRequestHeader("If-None-Match",T.etag[p])),(g.data&&g.hasContent&&!1!==g.contentType||t.contentType)&&k.setRequestHeader("Content-Type",g.contentType),k.setRequestHeader("Accept",g.dataTypes[0]&&g.accepts[g.dataTypes[0]]?g.accepts[g.dataTypes[0]]+("*"!==g.dataTypes[0]?", "+pt+"; q=0.01":""):g.accepts["*"]),g.headers)k.setRequestHeader(o,g.headers[o]);if(g.beforeSend&&(!1===g.beforeSend.call(m,k,g)||2===w))return k.abort();for(o in s="abort",{success:1,error:1,complete:1})k[o](g[o]);if(l=gt(lt,g,t,k)){k.readyState=1,h&&v.trigger("ajaxSend",[k,g]),g.async&&0<g.timeout&&(d=setTimeout(function(){k.abort("timeout")},g.timeout));try{w=1,l.send(i,c)}catch(e){if(!(w<2))throw e;c(-1,e)}}else c(-1,"No Transport");function c(e,t,n,r){var o,i,a,s,c,u=t;2!==w&&(w=2,d&&clearTimeout(d),l=void 0,f=r||"",k.readyState=0<e?4:0,o=200<=e&&e<300||304===e,n&&(s=function(e,t,n){for(var r,o,i,a,s=e.contents,c=e.dataTypes;"*"===c[0];)c.shift(),void 0===r&&(r=e.mimeType||t.getResponseHeader("Content-Type"));if(r)for(o in s)if(s[o]&&s[o].test(r)){c.unshift(o);break}if(c[0]in n)i=c[0];else{for(o in n){if(!c[0]||e.converters[o+" "+c[0]]){i=o;break}a||(a=o)}i=i||a}return i?(i!==c[0]&&c.unshift(i),n[i]):void 0}(g,k,n)),s=function(e,t,n,r){var o,i,a,s,c,u={},l=e.dataTypes.slice();if(l[1])for(a in e.converters)u[a.toLowerCase()]=e.converters[a];for(i=l.shift();i;)if(e.responseFields[i]&&(n[e.responseFields[i]]=t),!c&&r&&e.dataFilter&&(t=e.dataFilter(t,e.dataType)),c=i,i=l.shift())if("*"===i)i=c;else if("*"!==c&&c!==i){if(!(a=u[c+" "+i]||u["* "+i]))for(o in u)if((s=o.split(" "))[1]===i&&(a=u[c+" "+s[0]]||u["* "+s[0]])){!0===a?a=u[o]:!0!==u[o]&&(i=s[0],l.unshift(s[1]));break}if(!0!==a)if(a&&e.throws)t=a(t);else try{t=a(t)}catch(e){return{state:"parsererror",error:a?e:"No conversion from "+c+" to "+i}}}return{state:"success",data:t}}(g,s,k,o),o?(g.ifModified&&((c=k.getResponseHeader("Last-Modified"))&&(T.lastModified[p]=c),(c=k.getResponseHeader("etag"))&&(T.etag[p]=c)),204===e||"HEAD"===g.type?u="nocontent":304===e?u="notmodified":(u=s.state,i=s.data,o=!(a=s.error))):(a=u,!e&&u||(u="error",e<0&&(e=0))),k.status=e,k.statusText=(t||u)+"",o?y.resolveWith(m,[i,u,k]):y.rejectWith(m,[k,u,a]),k.statusCode(b),b=void 0,h&&v.trigger(o?"ajaxSuccess":"ajaxError",[k,g,o?i:a]),x.fireWith(m,[k,u]),h&&(v.trigger("ajaxComplete",[k,g]),--T.active||T.event.trigger("ajaxStop")))}return k},getJSON:function(e,t,n){return T.get(e,t,n,"json")},getScript:function(e,t){return T.get(e,void 0,t,"script")}}),T.each(["get","post"],function(e,o){T[o]=function(e,t,n,r){return T.isFunction(t)&&(r=r||n,n=t,t=void 0),T.ajax({url:e,type:o,dataType:r,data:t,success:n})}}),T._evalUrl=function(e){return T.ajax({url:e,type:"GET",dataType:"script",async:!1,global:!1,throws:!0})},T.fn.extend({wrapAll:function(t){var e;return T.isFunction(t)?this.each(function(e){T(this).wrapAll(t.call(this,e))}):(this[0]&&(e=T(t,this[0].ownerDocument).eq(0).clone(!0),this[0].parentNode&&e.insertBefore(this[0]),e.map(function(){for(var e=this;e.firstElementChild;)e=e.firstElementChild;return e}).append(this)),this)},wrapInner:function(n){return this.each(T.isFunction(n)?function(e){T(this).wrapInner(n.call(this,e))}:function(){var e=T(this),t=e.contents();t.length?t.wrapAll(n):e.append(n)})},wrap:function(t){var n=T.isFunction(t);return this.each(function(e){T(this).wrapAll(n?t.call(this,e):t)})},unwrap:function(){return this.parent().each(function(){T.nodeName(this,"body")||T(this).replaceWith(this.childNodes)}).end()}}),T.expr.filters.hidden=function(e){return e.offsetWidth<=0&&e.offsetHeight<=0},T.expr.filters.visible=function(e){return!T.expr.filters.hidden(e)};var vt=/%20/g,yt=/\[\]$/,xt=/\r?\n/g,bt=/^(?:submit|button|image|reset|file)$/i,wt=/^(?:input|select|textarea|keygen)/i;function kt(n,e,r,o){var t;if(T.isArray(e))T.each(e,function(e,t){r||yt.test(n)?o(n,t):kt(n+"["+("object"==typeof t?e:"")+"]",t,r,o)});else if(r||"object"!==T.type(e))o(n,e);else for(t in e)kt(n+"["+t+"]",e[t],r,o)}T.param=function(e,t){function n(e,t){t=T.isFunction(t)?t():null==t?"":t,o[o.length]=encodeURIComponent(e)+"="+encodeURIComponent(t)}var r,o=[];if(void 0===t&&(t=T.ajaxSettings&&T.ajaxSettings.traditional),T.isArray(e)||e.jquery&&!T.isPlainObject(e))T.each(e,function(){n(this.name,this.value)});else for(r in e)kt(r,e[r],t,n);return o.join("&").replace(vt,"+")},T.fn.extend({serialize:function(){return T.param(this.serializeArray())},serializeArray:function(){return this.map(function(){var e=T.prop(this,"elements");return e?T.makeArray(e):this}).filter(function(){var e=this.type;return this.name&&!T(this).is(":disabled")&&wt.test(this.nodeName)&&!bt.test(e)&&(this.checked||!V.test(e))}).map(function(e,t){var n=T(this).val();return null==n?null:T.isArray(n)?T.map(n,function(e){return{name:t.name,value:e.replace(xt,"\r\n")}}):{name:t.name,value:n.replace(xt,"\r\n")}}).get()}}),T.ajaxSettings.xhr=function(){try{return new XMLHttpRequest}catch(e){}};var Tt=0,Ct={},St={0:200,1223:204},_t=T.ajaxSettings.xhr();h.attachEvent&&h.attachEvent("onunload",function(){for(var e in Ct)Ct[e]()}),v.cors=!!_t&&"withCredentials"in _t,v.ajax=_t=!!_t,T.ajaxTransport(function(i){var a;return v.cors||_t&&!i.crossDomain?{send:function(e,t){var n,r=i.xhr(),o=++Tt;if(r.open(i.type,i.url,i.async,i.username,i.password),i.xhrFields)for(n in i.xhrFields)r[n]=i.xhrFields[n];for(n in i.mimeType&&r.overrideMimeType&&r.overrideMimeType(i.mimeType),i.crossDomain||e["X-Requested-With"]||(e["X-Requested-With"]="XMLHttpRequest"),e)r.setRequestHeader(n,e[n]);a=function(e){return function(){a&&(delete Ct[o],a=r.onload=r.onerror=null,"abort"===e?r.abort():"error"===e?t(r.status,r.statusText):t(St[r.status]||r.status,r.statusText,"string"==typeof r.responseText?{text:r.responseText}:void 0,r.getAllResponseHeaders()))}},r.onload=a(),r.onerror=a("error"),a=Ct[o]=a("abort");try{r.send(i.hasContent&&i.data||null)}catch(e){if(a)throw e}},abort:function(){a&&a()}}:void 0}),T.ajaxSetup({accepts:{script:"text/javascript, application/javascript, application/ecmascript, application/x-ecmascript"},contents:{script:/(?:java|ecma)script/},converters:{"text script":function(e){return T.globalEval(e),e}}}),T.ajaxPrefilter("script",function(e){void 0===e.cache&&(e.cache=!1),e.crossDomain&&(e.type="GET")}),T.ajaxTransport("script",function(n){var r,o;if(n.crossDomain)return{send:function(e,t){r=T("<script>").prop({async:!0,charset:n.scriptCharset,src:n.url}).on("load error",o=function(e){r.remove(),o=null,e&&t("error"===e.type?404:200,e.type)}),y.head.appendChild(r[0])},abort:function(){o&&o()}}});var jt=[],$t=/(=)\?(?=&|$)|\?\?/;T.ajaxSetup({jsonp:"callback",jsonpCallback:function(){var e=jt.pop()||T.expando+"_"+tt++;return this[e]=!0,e}}),T.ajaxPrefilter("json jsonp",function(e,t,n){var r,o,i,a=!1!==e.jsonp&&($t.test(e.url)?"url":"string"==typeof e.data&&!(e.contentType||"").indexOf("application/x-www-form-urlencoded")&&$t.test(e.data)&&"data");return a||"jsonp"===e.dataTypes[0]?(r=e.jsonpCallback=T.isFunction(e.jsonpCallback)?e.jsonpCallback():e.jsonpCallback,a?e[a]=e[a].replace($t,"$1"+r):!1!==e.jsonp&&(e.url+=(nt.test(e.url)?"&":"?")+e.jsonp+"="+r),e.converters["script json"]=function(){return i||T.error(r+" was not called"),i[0]},e.dataTypes[0]="json",o=h[r],h[r]=function(){i=arguments},n.always(function(){h[r]=o,e[r]&&(e.jsonpCallback=t.jsonpCallback,jt.push(r)),i&&T.isFunction(o)&&o(i[0]),i=o=void 0}),"script"):void 0}),T.parseHTML=function(e,t,n){if(!e||"string"!=typeof e)return null;"boolean"==typeof t&&(n=t,t=!1),t=t||y;var r=b.exec(e),o=!n&&[];return r?[t.createElement(r[1])]:(r=T.buildFragment([e],t,o),o&&o.length&&T(o).remove(),T.merge([],r.childNodes))};var At=T.fn.load;T.fn.load=function(e,t,n){if("string"!=typeof e&&At)return At.apply(this,arguments);var r,o,i,a=this,s=e.indexOf(" ");return 0<=s&&(r=T.trim(e.slice(s)),e=e.slice(0,s)),T.isFunction(t)?(n=t,t=void 0):t&&"object"==typeof t&&(o="POST"),0<a.length&&T.ajax({url:e,type:o,dataType:"html",data:t}).done(function(e){i=arguments,a.html(r?T("<div>").append(T.parseHTML(e)).find(r):e)}).complete(n&&function(e,t){a.each(n,i||[e.responseText,t,e])}),this},T.each(["ajaxStart","ajaxStop","ajaxComplete","ajaxError","ajaxSuccess","ajaxSend"],function(e,t){T.fn[t]=function(e){return this.on(t,e)}}),T.expr.filters.animated=function(t){return T.grep(T.timers,function(e){return t===e.elem}).length};var Et=h.document.documentElement;function Nt(e){return T.isWindow(e)?e:9===e.nodeType&&e.defaultView}T.offset={setOffset:function(e,t,n){var r,o,i,a,s,c,u=T.css(e,"position"),l=T(e),p={};"static"===u&&(e.style.position="relative"),s=l.offset(),i=T.css(e,"top"),c=T.css(e,"left"),o=("absolute"===u||"fixed"===u)&&-1<(i+c).indexOf("auto")?(a=(r=l.position()).top,r.left):(a=parseFloat(i)||0,parseFloat(c)||0),T.isFunction(t)&&(t=t.call(e,n,s)),null!=t.top&&(p.top=t.top-s.top+a),null!=t.left&&(p.left=t.left-s.left+o),"using"in t?t.using.call(e,p):l.css(p)}},T.fn.extend({offset:function(t){if(arguments.length)return void 0===t?this:this.each(function(e){T.offset.setOffset(this,t,e)});var e,n,r=this[0],o={top:0,left:0},i=r&&r.ownerDocument;return i?(e=i.documentElement,T.contains(e,r)?(typeof r.getBoundingClientRect!=U&&(o=r.getBoundingClientRect()),n=Nt(i),{top:o.top+n.pageYOffset-e.clientTop,left:o.left+n.pageXOffset-e.clientLeft}):o):void 0},position:function(){if(this[0]){var e,t,n=this[0],r={top:0,left:0};return"fixed"===T.css(n,"position")?t=n.getBoundingClientRect():(e=this.offsetParent(),t=this.offset(),T.nodeName(e[0],"html")||(r=e.offset()),r.top+=T.css(e[0],"borderTopWidth",!0),r.left+=T.css(e[0],"borderLeftWidth",!0)),{top:t.top-r.top-T.css(n,"marginTop",!0),left:t.left-r.left-T.css(n,"marginLeft",!0)}}},offsetParent:function(){return this.map(function(){for(var e=this.offsetParent||Et;e&&!T.nodeName(e,"html")&&"static"===T.css(e,"position");)e=e.offsetParent;return e||Et})}}),T.each({scrollLeft:"pageXOffset",scrollTop:"pageYOffset"},function(t,o){var i="pageYOffset"===o;T.fn[t]=function(e){return D(this,function(e,t,n){var r=Nt(e);return void 0===n?r?r[o]:e[t]:void(r?r.scrollTo(i?h.pageXOffset:n,i?n:h.pageYOffset):e[t]=n)},t,e,arguments.length,null)}}),T.each(["top","left"],function(e,n){T.cssHooks[n]=Te(v.pixelPosition,function(e,t){return t?(t=ke(e,n),be.test(t)?T(e).position()[n]+"px":t):void 0})}),T.each({Height:"height",Width:"width"},function(i,a){T.each({padding:"inner"+i,content:a,"":"outer"+i},function(r,e){T.fn[e]=function(e,t){var n=arguments.length&&(r||"boolean"!=typeof e),o=r||(!0===e||!0===t?"margin":"border");return D(this,function(e,t,n){var r;return T.isWindow(e)?e.document.documentElement["client"+i]:9===e.nodeType?(r=e.documentElement,Math.max(e.body["scroll"+i],r["scroll"+i],e.body["offset"+i],r["offset"+i],r["client"+i])):void 0===n?T.css(e,t,o):T.style(e,t,n,o)},a,n?e:void 0,n,null)}})}),T.fn.size=function(){return this.length},T.fn.andSelf=T.fn.addBack,"function"==typeof define&&define.amd&&define("jquery",[],function(){return T});var Ot=h.jQuery,Dt=h.$;return T.noConflict=function(e){return h.$===T&&(h.$=Dt),e&&h.jQuery===T&&(h.jQuery=Ot),T},typeof e==U&&(h.jQuery=h.$=T),T});"use strict";function _typeof(e){return(_typeof="function"==typeof Symbol&&"symbol"==typeof Symbol.iterator?function(e){return typeof e}:function(e){return e&&"function"==typeof Symbol&&e.constructor===Symbol&&e!==Symbol.prototype?"symbol":typeof e})(e)}function _typeof(e){return(_typeof="function"==typeof Symbol&&"symbol"==typeof Symbol.iterator?function(e){return typeof e}:function(e){return e&&"function"==typeof Symbol&&e.constructor===Symbol&&e!==Symbol.prototype?"symbol":typeof e})(e)}window.Kvs=function(){function e(e){e=e||{},this.namespace=e.namespace||"kvs",e.seconds&&(this.seconds=e.seconds||2592e3)}return e.prototype.get=function(e){if(e=this.namespace+"-"+e,!this.isLocalStorageSupported()||void 0===localStorage[e])return null;var t=JSON.parse(localStorage[e]),n=(new Date).getTime();if(void 0!==t.meta.exp&&t.meta.exp<n)return delete localStorage[e],null;switch(t.meta.type){case"string":return t.data;case"undefined":case"null":return t.data;case"object":default:return JSON.parse(t.data)}},e.prototype.set=function(e,t,n){if(this.isLocalStorageSupported()){var r={meta:{type:_typeof(t)}};switch(void 0===n&&void 0===this.seconds||(r.meta.exp=(new Date).getTime()+1e3*parseInt(n||this.seconds)),r.meta.type){case"string":r.data=t;break;case"object":default:r.data=JSON.stringify(t)}localStorage[this.namespace+"-"+e]=JSON.stringify(r)}},e.prototype.delete=function(e){this.isLocalStorageSupported()&&delete localStorage[this.namespace+"-"+e]},e.prototype.remove=function(e){if(this.isLocalStorageSupported()){var t=this.get(e);return this.delete(e),t}},e.prototype.isLocalStorageSupported=function(){try{var e=window.localStorage;return e.setItem("test","1"),e.removeItem("test"),!0}catch(e){return!1}},e}(),window.prx=window.prx||{},window.sd=window.sd||{},window.evt=window.evt||{},window.pee=window.pee||{},window.proto&&(window.prxy=window.prxy||window.proto.data),prx.kvs=new Kvs({namespace:"proto"}),prx.utils=window.prx.utils||{},prx.utils.dynamic_properties=["listitems","buttons","options","tabs"],prx.utils.XSSJSWhitelistForFroala={b:["style"],br:[],div:["style"],em:["style"],i:["style"],span:["style","class"],strong:["style"],strike:["style"],s:["style"],u:["style"],font:[]},prx.utils.setCookie=function(e,t,n){var r=new Date;r.setDate(r.getDate()+n);var o=escape(t)+(null==n?"":"; expires="+r.toUTCString());document.cookie=e+"="+o},prx.utils.getCookie=function(e){var t,n,r,o=document.cookie.split(";");for(t=0;t<o.length;t++)if(n=o[t].substr(0,o[t].indexOf("=")),r=o[t].substr(o[t].indexOf("=")+1),(n=n.replace(/^\s+|\s+$/g,""))==e)return unescape(r)},prx.utils.deleteCookie=function(e){document.cookie=e+"=; expires=Thu, 01 Jan 1970 00:00:01 GMT;"},prx.utils.getGuid=function(){return"xxxxxxxx-xxxx-4xxx-yxxx-xxxxxxxxxxxx".replace(/[xy]/g,function(e){var t=16*Math.random()|0;return("x"==e?t:3&t|8).toString(16)})},prx.utils.isGuid=function(e){return/^[0-9a-fA-F]{8}-[0-9a-fA-F]{4}-[0-9a-fA-F]{4}-[0-9a-fA-F]{4}-?[0-9a-fA-F]{12}$/.test(e)},prx.utils.getColor=function(e){if("string"==typeof e){if(!/^[0-9a-f]{3,6}$/i.test(e))try{e=JSON.parse(e)}catch(e){}"number"==typeof e&&(e=""+e)}return"object"==_typeof(e)?e:void 0===e||null==e?"":"none"==(e=e.trim())?"rgba(0,0,0,0)":0==e?"rgba(0,0,0,1)":("rgba"==e.substring(0,4)&&3<(e.match(/,/g)||[]).length&&(e=e.replace(/,([^,]*)$/,".$1")),e==e.replace(/[^A-F0-9]/gi,"")?"#"+e:e)},prx.utils.getBgCss=function(e){return prx.gradients.getBgCss(e)},prx.utils.getBgCssByProperty=function(e,t){return prx.gradients.getBgCssByProperty(e,t)},prx.utils.getBgCssByValue=function(e){return prx.gradients.getBgCssByValue(e)},prx.utils.checkColorPickerInputOnUpdate=function(e){var t=/[#a-f0-9]/i;return!!(t.test(e)||/^rgb\((?:(?:25[0-5]|2[0-4][0-9]|[01]?[0-9][0-9]?)\s?,\s?){2}(?:25[0-5]|2[0-4][0-9]|[01]?[0-9][0-9]?)\)/.test(e)||/rgba\((\d{1,3}),\s*(\d{1,3}),\s*(\d{1,3}),\s*(\d*(?:\.\d+)?)\)/gi.test(e)||/^hsl\(\s*(0|[1-9]\d?|[12]\d\d|3[0-5]\d)\s*,\s*((0|[1-9]\d?|100)%)\s*,\s*((0|[1-9]\d?|100)%)\s*\)$/.test(e))&&(t.test(e)&&"#"!==e.substring(0,1)?"#"+e:e)},prx.utils.getOpacity=function(e){if(void 0===e)return"";return/^[A-Z0-9,.%]+$/.test(e)?e:1},prx.utils.checkURL=function(e){return/^(http|https)/i.test(e)?e:"url"},prx.utils.isURL=function(e){return e===prx.utils.checkURL(e)},prx.utils.getNormalizedFilename=function(e){return void 0===e?"":e=e.replace(/[<,>,:,"",/,\,|,?,*]/gi,"-")},prx.utils.escapeHTML=function(e){return"string"!=typeof e?e:e.replace(/&/g,"&amp;").replace(/</g,"&lt;").replace(/>/g,"&gt;").replace(/"/g,"&quot;").replace(/'/g,"&#039;").replace(/\\/g,"&#92;")},prx.utils.unEscapeHTML=function(e){return"string"!=typeof e?e:e.replace(/&amp;/g,"&").replace(/&lt;/g,"<").replace(/&gt;/g,">").replace(/&quot;/g,'"').replace(/&#039;/g,"'").replace(/&#92;/g,"\\")},prx.utils.escapeEditableProperties=function(e){return"string"!=typeof e?e:e.replace(/&/g,"&amp;").replace(/(?!<br>|<br\/>|<br \/>)</g,"&lt;").replace(/"/g,"&quot;").replace(/'/g,"&#039;")},prx.utils.escapeForRegEx=function(e){return"string"!=typeof e?e:e.replace(/"/g,"\\\\").replace(/\//g,"\\/").replace(/\*/g,"\\*").replace(/\+/g,"\\+").replace(/\[/g,"\\[").replace(/\]/g,"\\]").replace(/\-/g,"\\-").replace(/\(/g,"\\(").replace(/\)/g,"\\)")},prx.utils.escapeRichText=function(e,t){return void 0===t&&(t=prx.utils.XSSJSWhitelistForFroala),"string"!=typeof e?e:e=(e=filterXSS(e,{whiteList:t,stripIgnoreTag:!0,stripIgnoreTagBody:["script"]})).replace(/&#8203;/gi,"")},prx.utils.getEventPageXY=function(e){var t={x:0,y:0};if("touchstart"==e.type||"touchmove"==e.type||"touchend"==e.type||"touchcancel"==e.type){var n=e.originalEvent.touches[0]||e.originalEvent.changedTouches[0];t.x=n.pageX,t.y=n.pageY}else"click"!=e.type&&"mousedown"!=e.type&&"mouseup"!=e.type&&"mousemove"!=e.type&&"mouseover"!=e.type&&"mouseout"!=e.type&&"mouseenter"!=e.type&&"mouseleave"!=e.type||(t.x=e.pageX,t.y=e.pageY);return t},prx.utils.getAssetsServer=function(e){if(e){for(var t=prx.nodejsassetsserver.maxsubdomains,n=0,r=0;r<e.length;r++)n+=e.charCodeAt(r);return(t<(n=n%(t-1)+2)||n<1)&&(n=2),prx.nodejsassetsserver.subdomainprefix+n+prx.nodejsassetsserver.host}return prx.nodejsassetsserver.subdomainprefix+"1"+prx.nodejsassetsserver.host},prx.utils.forceWebkitRerender=function(e){var t=e.get(0),n=t.style.display;t.style.display="none",t.offsetHeight,t.style.display=n},prx.utils.multilineEllipsis=function(e,t,a,s,c){c||(c=25),!0!==s&&(s=!1);var n=$(e).clone(!0,!0);$(t,n).each(function(e,t){var n="",r=0,o="";if(0!==$(t).outerHeight())for(;$(t).outerHeight()>a;){o=$(t).text(),s&&-1!=o.indexOf(".")&&(n=o.substring(o.lastIndexOf(".")),o=o.substring(0,o.lastIndexOf(".")),r=n.length);var i=o.replace(/\W*\s(\S)*$/,"...");if(o.length==i.length){i=i.substr(0,c-3-r)+"..."+n,$(t).text(i);break}o+=n,$(t).text(i)}else(o=$(t).text()).length>c&&(s&&-1!=o.indexOf(".")&&(n=o.substring(o.lastIndexOf(".")+1),o=o.substring(0,o.lastIndexOf(".")),r=n.length),o=o.substr(0,c-3-r)+"..."+n,$(t).text(o))}),$(e).replaceWith(n)},prx.utils.bindScreenshotsFunctionality=function(e){$(e).find(".thumbnail-img").on("error",function(){$(this).hide(),$(this).siblings(".dummy-img").hide(),$(this).siblings(".loading-placeholder-img").show()}),$(e).find(".thumbnail-img").on("load",function(){$(this).siblings(".loading-placeholder-img").hide(),$(this).show(),$(this).parents(".containerLI, .screenLI").hasClass("has-multiple-states")&&$(this).siblings(".dummy-img").show()})},prx.utils.arraysAreIdentical=function(e,t){if(e.length!==t.length)return!1;for(var n=0,r=e.length;n<r;n++)if(e[n]!==t[n])return!1;return!0},prx.utils.getNewBezierEase=function(e,t,n,r){return new Ease(BezierEasing(e,t,n,r))},prx.utils.debounceFunction=function(r,o,i){var a;return function(){var e=this,t=arguments,n=i&&!a;clearTimeout(a),a=setTimeout(function(){a=null,i||r.apply(e,t)},o),n&&r.apply(e,t)}},prx.utils.raf=function(e){window.requestAnimationFrame(e)},prx.utils.isValidUrl=function(e){return new RegExp(/^(?:(?:https?|ftp):\/\/)(?:\S+(?::\S*)?@)?(?:(?!(?:10|127)(?:\.\d{1,3}){3})(?!(?:169\.254|192\.168)(?:\.\d{1,3}){2})(?!172\.(?:1[6-9]|2\d|3[0-1])(?:\.\d{1,3}){2})(?:[1-9]\d?|1\d\d|2[01]\d|22[0-3])(?:\.(?:1?\d{1,2}|2[0-4]\d|25[0-5])){2}(?:\.(?:[1-9]\d?|1\d\d|2[0-4]\d|25[0-4]))|(?:(?:[a-z\u00a1-\uffff0-9]-*)*[a-z\u00a1-\uffff0-9]+)(?:\.(?:[a-z\u00a1-\uffff0-9]-*)*[a-z\u00a1-\uffff0-9]+)*(?:\.(?:[a-z\u00a1-\uffff]{2,}))\.?)(?::\d{2,5})?(?:[/?#]\S*)?$/i).test(e)},prx.utils.isFaceTimeUrl=function(e){return!(!e.startsWith("facetime:")&&!e.startsWith("facetime-audio:"))&&!(-1<(e=e.replace(/^facetime:/,"").replace(/^facetime-audio:/,"")).indexOf(":"))},prx.utils.isValidUserPreset=function(e){return!(!/^[0-9a-fA-F]{8}-[0-9a-fA-F]{4}-[0-9a-fA-F]{4}-[0-9a-fA-F]{4}-?[0-9a-fA-F]{12}$/.test(e)||void 0===prx.easing._customEasings[e])},prx.utils.isSpringrk4=function(e){return"springrk4("===e.substr(0,10)},prx.utils.getDefaultDefaultTransition=function(){return{duration:250,delay:0,easing:"out"}},prx.utils.isSafariVorGreater=function(e){if("undefined"==typeof navigator||"undefined"!=typeof navigator&&void 0===navigator.userAgent)return!1;if(-1==navigator.userAgent.indexOf("Safari")||-1!==navigator.userAgent.indexOf("Chrome"))return!1;var t=navigator.userAgent.substring(navigator.userAgent.indexOf("Version")).split("/")[1].replace("Safari","").trim();return-1<prx.utils.compareVersions(t,e)},prx.utils.compareVersions=function(e,t){var n=/^v?(?:\d+)(\.(?:[x*]|\d+)(\.(?:[x*]|\d+)(\.(?:[x*]|\d+))?(?:-[\da-z\-]+(?:\.[\da-z\-]+)*)?(?:\+[\da-z\-]+(?:\.[\da-z\-]+)*)?)?)?$/i;return function(e,t){if(!d(e)||!d(t))return-2;for(var n=p(e),r=p(t),o=0;o<Math.max(n.length-1,r.length-1);o++){var i=parseInt(n[o]||0,10),a=parseInt(r[o]||0,10);if(a<i)return 1;if(i<a)return-1}var s=n[n.length-1],c=r[r.length-1];if(s&&c){var u=s.split(".").map(f),l=c.split(".").map(f);for(o=0;o<Math.max(u.length,l.length);o++){if(void 0===u[o]||"string"==typeof l[o]&&"number"==typeof u[o])return-1;if(void 0===l[o]||"string"==typeof u[o]&&"number"==typeof l[o])return 1;if(u[o]>l[o])return 1;if(l[o]>u[o])return-1}}else if(s||c)return s?-1:1;return 0}(e,t);function p(e){var t=e.replace(/^v/,"").replace(/\+.*$/,""),n=function(e,t){return-1===e.indexOf(t)?e.length:e.indexOf(t)}(t,"-"),r=t.substring(0,n).split(".");return r.push(t.substring(n+1)),r}function f(e){return isNaN(Number(e))?e:Number(e)}function d(e){return"string"==typeof e&&!!n.test(e)}},prx.utils.isMac=function(){return-1!=navigator.appVersion.indexOf("Mac")},prx.utils.isWindows=function(){return-1!==window.navigator.userAgent.indexOf("Windows")},prx.crossmsg={},prx.crossmsg.ini=function(){window.addEventListener("message",function(e){prx.crossmsg.checkrequestdomain(e)&&prx.crossmsg.onmsg(e.data)},!1)},prx.crossmsg.getdomain=function(){return prx.export2html?"*":prx.url.crossmsg},prx.crossmsg.checkrequestdomain=function(e){var t=e.origin||e.originalEvent.origin;return!prx.export2html&&t==prx.url.crossmsg||!!prx.export2html},prx.crossmsg.send=function(e,t){prx.crossmsg.gettargetframe(t).postMessage(e,prx.crossmsg.getdomain())},prx.unsupportedBrowsers={},prx.unsupportedBrowsers.getIEVersion=function(){var e=navigator.userAgent,t=e.indexOf("MSIE ");if(0<t)return parseInt(e.substring(t+5,e.indexOf(".",t)),10);if(0<e.indexOf("Trident/")){var n=e.indexOf("rv:");return parseInt(e.substring(n+3,e.indexOf(".",n)),10)}var r=e.indexOf("Edge/");return 0<r?parseInt(e.substring(r+5,e.indexOf(".",r)),10):void 0},prx.unsupportedBrowsers.checkVersion=function(e){if(null!=prx.unsupportedBrowsers.getIEVersion())prx.unsupportedBrowsers.showWarning("ie",e,!0);else{var t=navigator.userAgent||navigator.vendor||window.opera;/Mobile|iP(hone|od|ad)|Android|BlackBerry|IEMobile|Kindle|NetFront|Silk-Accelerated|(hpw|web)OS|Fennec|Minimo|Opera M(obi|ini)|Blazer|Dolfin|Dolphin|Skyfire|Zune/.test(t)&&prx.unsupportedBrowsers.showWarning("mobile",e,!1)}},prx.unsupportedBrowsers.showWarning=function(e,t,n){var r=$('<div id="bad-browser-prompt" class="top-header-alert link"><span id="warning-text"></span><span class="close-btn embed-icon-close-x"></span></div>');switch(t){case"spaces":case"export":case"prto":case"player":var o="You are using a browser that is not supported by Proto.io. We recommend that you use one of the browsers listed <a id='link' href='https://support.proto.io/hc/en-us/articles/220036207-The-Proto-io-user-interfaces' target='_blank'>here</a> to use the Player.";r.find("#warning-text").html(o),$("body").prepend(r);break;case"editor":if("ie"==e)o="You are using a browser that is not supported by Proto.io. We recommend that you use one of the browsers listed <a id='link' href='https://support.proto.io/hc/en-us/articles/220036207-The-Proto-io-user-interfaces' target='_blank'>here</a> to use the Editor.";else{o="You are accessing the Proto.io Editor from a device that is not supported. Some features will not work. We recommend that you switch to a desktop or laptop computer to use the Editor.";r.addClass("error")}r.find("#warning-text").html(o),$("#editor-top-panel").before(r),prx.layout.helper.toggleTopInfoBar(r.outerHeight(),!0)}$("#bad-browser-prompt").click(function(){1==n&&window.open("https://support.proto.io/hc/en-us/articles/220036207-The-Proto-io-user-interfaces","_blank")}),$("#bad-browser-prompt a").click(function(e){e.stopPropagation()}),$("#bad-browser-prompt .close-btn").click(function(){prx.unsupportedBrowsers.removeWarning(t)})},prx.unsupportedBrowsers.removeWarning=function(e){$("#bad-browser-prompt").remove(),"editor"==e&&prx.layout.helper.toggleTopInfoBar(0,!1)},window.prx=window.prx||{},prx.embed=0,prx.export2html=0,prx.letterboxmode=!1,prx.goToPage=function(e){var t=!0,n=prx.inAnnotationMode;prx.inAnnotationMode&&(t=prx.disableAnnotations()),t&&(prx.highlightPage(e),prx.crossmsg.send({action:"change-page",data:{pageid:e}}),$("#menu-screen-browser [data-screen-id="+parseInt(e)+"]").attr("data-orientation")!=prx.orientation&&prx.switchOrientation(!1),n&&prx.iniAnnotations())},prx.setActivePage=function(e){prx.activePage={},prx.activePage.id=e,void 0!==prx.data&&void 0!==prx.data.pages&&(prx.activePage.title=prx.data.pages[prx.getPageIndexFromId(e)].title)},prx.getPageIndexFromId=function(e){for(var t=0;t<prx.data.pages.length;t++)if(parseInt(prx.data.pages[t].id)==parseInt(e))return t;return-1};var getColumnIndex=function(e,t){t=t.toLowerCase();for(var n=0;n<e.length;n++)if(e[n].toLowerCase()==t)return n;return!1},getMonthsArray=function(){return["January","February","March","April","May","June","July","August","September","October","November","December"]},getScreenIndexFromId=function(e){for(var t=-1,n=0;n<prx.data.pages.length;n++)prx.data.pages[n].id==e&&(t=n);return t},innerPrx=function(){return $("#pio-playerframe").get(0).contentWindow.prx};function setCookie(e,t,n){var r=new Date;r.setDate(r.getDate()+n);var o=escape(t)+(null==n?"":"; expires="+r.toUTCString());document.cookie=e+"="+o}function getCookie(e){var t,n,r,o=document.cookie.split(";");for(t=0;t<o.length;t++)if(n=o[t].substr(0,o[t].indexOf("=")),r=o[t].substr(o[t].indexOf("=")+1),(n=n.replace(/^\s+|\s+$/g,""))==e)return unescape(r)}function deleteCookie(e){document.cookie=e+"=; expires=Thu, 01 Jan 1970 00:00:01 GMT;"}function _defineProperty(e,t,n){return t in e?Object.defineProperty(e,t,{value:n,enumerable:!0,configurable:!0,writable:!0}):e[t]=n,e}prx.iniBgColor=function(){var e=$("body").css("background-color");prx.checkBackgroundColorBrightness(e)},prx.checkBackgroundColorBrightness=function(e){if(-1!=e.indexOf("#"))var t=e.substring(1),n=(i=parseInt(t,16))>>16&255,r=i>>8&255,o=i>>0&255;else{var i=(t=-1!==e.indexOf("rgba")?e.substring(4):e.substring(3)).slice(1,-1).split(",");n=parseInt(i[0].trim()),r=parseInt(i[1].trim()),o=parseInt(i[2].trim())}var a=.2989*n+.587*r+.114*o,s=$(".sidebar-action-buttons > .header-action-button, .sidebar-action-buttons > #zoom, .sidebar-action-buttons > #zoom .ui-slider-handle, #branding, #screen-nav-arrow-left, #screen-nav-arrow-right");a<120?s.addClass("dark-bg"):s.removeClass("dark-bg")},prx.utils.playerEngineEnterFullscreen=function(){$("body").addClass("player-engine-fullscreen")},prx.utils.playerEngineExitFullscreen=function(){$("body").removeClass("player-engine-fullscreen")},prx.utils.missedFonts=function(){var e;prx.spaces||prx.export2html||prx.letterboxmode||prx.embed||(prx.user&&prx.user.editor_access&&(e="https://support.proto.io/hc/en-us/articles/360007433231"),prx.toaster.create("Some fonts used in this project are not available. A default font is displayed instead.",{type:"warning",link:e,linkLabel:"Learn more",title:"Missing Fonts"}))},prx.utils.onmobile=function(){var e=navigator.userAgent||navigator.vendor||window.opera;return/(android|bb\d+|meego).+mobile|avantgo|bada\/|blackberry|blazer|compal|elaine|fennec|hiptop|iemobile|ip(hone|od|ad)|iris|kindle|lge |maemo|midp|mmp|mobile.+firefox|netfront|opera m(ob|in)i|palm( os)?|phone|p(ixi|re)\/|plucker|pocket|psp|series(4|6)0|symbian|treo|up\.(browser|link)|vodafone|wap|windows ce|xda|xiino/i.test(e)||/1207|6310|6590|3gso|4thp|50[1-6]i|770s|802s|a wa|abac|ac(er|oo|s\-)|ai(ko|rn)|al(av|ca|co)|amoi|an(ex|ny|yw)|aptu|ar(ch|go)|as(te|us)|attw|au(di|\-m|r |s )|avan|be(ck|ll|nq)|bi(lb|rd)|bl(ac|az)|br(e|v)w|bumb|bw\-(n|u)|c55\/|capi|ccwa|cdm\-|cell|chtm|cldc|cmd\-|co(mp|nd)|craw|da(it|ll|ng)|dbte|dc\-s|devi|dica|dmob|do(c|p)o|ds(12|\-d)|el(49|ai)|em(l2|ul)|er(ic|k0)|esl8|ez([4-7]0|os|wa|ze)|fetc|fly(\-|_)|g1 u|g560|gene|gf\-5|g\-mo|go(\.w|od)|gr(ad|un)|haie|hcit|hd\-(m|p|t)|hei\-|hi(pt|ta)|hp( i|ip)|hs\-c|ht(c(\-| |_|a|g|p|s|t)|tp)|hu(aw|tc)|i\-(20|go|ma)|i230|iac( |\-|\/)|ibro|idea|ig01|ikom|im1k|inno|ipaq|iris|ja(t|v)a|jbro|jemu|jigs|kddi|keji|kgt( |\/)|klon|kpt |kwc\-|kyo(c|k)|le(no|xi)|lg( g|\/(k|l|u)|50|54|\-[a-w])|libw|lynx|m1\-w|m3ga|m50\/|ma(te|ui|xo)|mc(01|21|ca)|m\-cr|me(rc|ri)|mi(o8|oa|ts)|mmef|mo(01|02|bi|de|do|t(\-| |o|v)|zz)|mt(50|p1|v )|mwbp|mywa|n10[0-2]|n20[2-3]|n30(0|2)|n50(0|2|5)|n7(0(0|1)|10)|ne((c|m)\-|on|tf|wf|wg|wt)|nok(6|i)|nzph|o2im|op(ti|wv)|oran|owg1|p800|pan(a|d|t)|pdxg|pg(13|\-([1-8]|c))|phil|pire|pl(ay|uc)|pn\-2|po(ck|rt|se)|prox|psio|pt\-g|qa\-a|qc(07|12|21|32|60|\-[2-7]|i\-)|qtek|r380|r600|raks|rim9|ro(ve|zo)|s55\/|sa(ge|ma|mm|ms|ny|va)|sc(01|h\-|oo|p\-)|sdk\/|se(c(\-|0|1)|47|mc|nd|ri)|sgh\-|shar|sie(\-|m)|sk\-0|sl(45|id)|sm(al|ar|b3|it|t5)|so(ft|ny)|sp(01|h\-|v\-|v )|sy(01|mb)|t2(18|50)|t6(00|10|18)|ta(gt|lk)|tcl\-|tdg\-|tel(i|m)|tim\-|t\-mo|to(pl|sh)|ts(70|m\-|m3|m5)|tx\-9|up(\.b|g1|si)|utst|v400|v750|veri|vi(rg|te)|vk(40|5[0-3]|\-v)|vm40|voda|vulc|vx(52|53|60|61|70|80|81|83|85|98)|w3c(\-| )|webc|whit|wi(g |nc|nw)|wmlb|wonu|x700|yas\-|your|zeto|zte\-/i.test(e.substr(0,4))},prx.utils.isUserTestingPage=function(e){var t=["usertesting","uxarmy","validately","userzoom","lookback","userlytics"];for(var n in t)if(e.includes(t[n]))return!0;return!1},prx.changeZoomLevel=function(e){var t=1<arguments.length&&void 0!==arguments[1]&&arguments[1];prx._zoom=e/100;var n={"-moz-transform":"scale("+e/100+")","-webkit-transform":"scale("+e/100+")",transform:"scale("+e/100+")"};$("#zoom .ui-slider-handle").html(parseInt(e)+"%"),$(".body_generic").css(n),1!==prx._zoom?$(".body_generic").addClass("zoomed"):$(".body_generic").removeClass("zoomed"),prx.positionFrame(),prx.inAnnotationMode&&(prx.resizeAnnotationsOverlay(),prx.positionAnnotationPopup()),0<$("#zoom .ui-slider-handle").length&&(prx._zoomcookie&&setCookie("zoomLevel-"+prx.project.id,e),$("#zoom").slider("value",e),prx.temporarilyShowZoomLevel()),prx._autozoom=!1,t||prx.crossmsg.send({action:"zoom-level-changed",data:{zoom:prx._zoom}})},prx.normalizeZoomLevel=function(e){if("string"==typeof e)if("+"==e.substr(0,1)){var t=parseInt(e.substr(1));e=Math.floor(Math.round(100*prx._zoom)/t)*t+t}else if("-"==e.substr(0,1)){t=parseInt(e.substr(1));e=Math.round(100*prx._zoom)%t!=0?Math.floor(Math.round(100*prx._zoom)/t)*t:100*prx._zoom-t}return 0<$("#zoom .ui-slider-handle").length?Math.min(Math.max(e,$("#zoom").slider("option","min")),$("#zoom").slider("option","max")):Math.floor(e)},prx.resetZoomMinValue=function(){0<$("#zoom .ui-slider-handle").length&&$("#zoom").slider("option","min",20)},prx.fitToScreenOnPageStart=function(){if(prx._zoomcookie)var e=getCookie("zoomLevel-"+prx.project.id);var t=prx.getAutoZoomValue();prx.embed||void 0===e?(t<100&&(prx.changeZoomLevel(t,!0),prx._zoomcookie&&deleteCookie("zoomLevel-"+prx.project.id)),prx._autozoom=!0):(prx.changeZoomLevel(e,!0),e==t&&(prx._autozoom=!0)),prx.positionFrame()},prx.getAutoZoomValue=function(){var e=$(".frame_generic").outerWidth()/prx._zoom,t=$(".frame_generic").outerHeight()/prx._zoom,n=$(window).width()-parseInt($("#maincontent").css("padding-left"))-parseInt($("#maincontent").css("padding-right"))-parseInt($("#maincontent").css("border-right-width")),r=$(window).height()-parseInt($("#maincontent").css("padding-top"))-parseInt($("#maincontent").css("padding-bottom"))-$(".top-header-alert").innerHeight();if(prx.embed||prx.spaces&&(!prx.spaces||prx.hasSpacesThemeHands()))o=r/t*100,i=n/e*100;else var o=5*Math.floor(r/t*100/5),i=5*Math.floor(n/e*100/5);if(prx.hasSpacesThemeHands())var a=o;else a=Math.min(o,i,100);return prx.normalizeZoomLevel(a)},prx.applySkin=function(){prx.applyBackground();var e=prx.getCustomSkinUrl();if($(".body_generic_inner").css("background-image","none"),!1===e)return!1;var t=$('<img src="'+e+'"/>');t.on("load",function(){"custom"==prx.skin&&!0===prx.device.hasCustomSkinNotch?$(".detail-top").css("background-image","url("+e+")"):$(".body_generic_inner").css("background-image","url("+e+")"),prx.positionCanvasInSkin(t),prx.applySkinCallback()})},prx.applyBackground=function(){(prx.hasSpacesTheme()||"apple-watch-42mm-2"==prx.skin||"apple-watch-38mm-2"==prx.skin|"apple-watch-40mm-2"==prx.skin||"apple-watch-44mm-2"==prx.skin)&&$("#pio-playerframe").css("background-color","#000000")},prx.positionCanvasInSkin=function(e){var t=e.get(0).naturalHeight,n=e.get(0).naturalWidth;"custom"==prx.skin&&!0===prx.device.hasCustomSkinNotch?($(".detail-top").width(n).height(t),$(".detail-top").css({"background-size":n+"px "+t+"px","margin-top":"-"+(t-$("#pio-playerframe").height())/2+"px","margin-left":"-"+(n-$("#pio-playerframe").width())/2+"px"})):($(".body_generic_inner").width(n).height(t),$("#pio-playerframe").css({"margin-top":(t-$("#pio-playerframe").height())/2+"px","margin-left":(n-$("#pio-playerframe").width())/2+"px"}))},prx.applySkinCallback=function(){prx.positionFrame(),prx.fitToScreenOnPageStart()},prx.getCustomSkinUrl=function(){return("custom"==prx.skin||"shared"==prx.skin)&&void 0!==prx.device&&prx.utils.getAssetsServer()+"/device/"+("shared"==prx.skin?"shared/":"")+prx.device.device_s3bucket+"/skin_"+prx.orientation+".png?ac="+prx.account},prx.hasSpacesTheme=function(){return!1},prx.hasSpacesThemeHands=function(){return!1},prx.positionFrame=function(){$("#maincontent").css({display:"block","margin-left":"0px","margin-top":"0px"}),$(".body_generic").css({width:"auto",position:"relative","margin-left":"0px"}),$(".frame_generic").css({width:"auto",height:"auto",margin:"auto"}),0<$(".body_generic_inner[data-img-width]").length&&($(".body_generic_inner").width($(".body_generic_inner").attr("data-img-width")).height($(".body_generic_inner").attr("data-img-height")).css({"background-position":"center center","margin-left":prx.hasSpacesTheme()?prx.theme.orientations[prx.orientation].excessLeft/prx.theme.orientations[prx.orientation].scale:0}),$("#maincontent").removeClass("forced-no-skin")),0<$("#pio-playerframe[data-canvas-x]").length&&$("#pio-playerframe").css("margin-left",$("#pio-playerframe").attr("data-canvas-x")+"px");var e,t=$(".frame_generic").width()*prx._zoom,n=$(window).width()-parseInt($("#maincontent").css("border-right-width"))-parseInt($("#maincontent").css("padding-left"))-parseInt($("#maincontent").css("padding-right")),r=$(".frame_generic").height()*prx._zoom,o=$(window).height()-2*parseInt($("#maincontent").css("padding-top"))-$(".top-header-alert").innerHeight();if($(".frame_generic").width(($(".body_generic_inner").innerWidth()+$(".body_generic").innerWidth()-$(".body_generic").width()+parseInt($(".body_generic").css("border-left-width"))+parseInt($(".body_generic").css("border-right-width")))*prx._zoom).height(($(".body_generic_inner").innerHeight()+$(".body_generic").innerHeight()-$(".body_generic").height()+parseInt($(".body_generic").css("border-top-width"))+parseInt($(".body_generic").css("border-bottom-width")))*prx._zoom),$(".body_generic").css({width:1/prx._zoom*100+"%",position:"absolute"}),prx.hasSpacesTheme()&&t+prx.theme.orientations[prx.orientation].excessLeft>n){2,prx._zoom;var i=n/prx._zoom;i=Math.max(i,prx.theme.orientations[prx.orientation].width*prx.theme.orientations[prx.orientation].scale+150/prx._zoom);prx.theme.orientations[prx.orientation].excessLeft;var a=i/2-prx.theme.orientations[prx.orientation].width*prx.theme.orientations[prx.orientation].scale/2-prx.theme.orientations[prx.orientation].x*prx.theme.orientations[prx.orientation].scale;$(".body_generic_inner").css({"margin-left":"0px",width:i+"px","background-position":a+"px center"}),$(".body_generic").css({"margin-left":"0px",width:i+"px"}),$(".frame_generic").width(n),$("#pio-playerframe").css("margin-left",(i-$("#pio-playerframe").width())/2+"px"),t=i}n<t?($("#maincontent").css({display:"block"}),$("#maincontent").css({"margin-left":-2*$("#maincontent").offset().left+"px"}).get(0).scrollLeft=0,r<o&&($("#maincontent").css({"margin-top":(o-r)/2+"px"}).get(0).scrollTop=0)):($("#maincontent").css({display:"-webkit-box"}),$("#maincontent").css((_defineProperty(e={display:"-webkit-flexbox"},"display","-webkit-flex"),_defineProperty(e,"display","-ms-flex"),_defineProperty(e,"display","flex"),e)))},prx.iniOrientation=function(){$("#change-orientation").click(function(e){return prx.switchOrientation(!0),!1})},prx.switchOrientation=function(e){if(void 0!==prx.device){var t=$("#pio-playerframe"),n="portrait"==prx.orientation?"landscape":"portrait";if(!prx.orientations[n])return!1;prx.orientation=n;var r=prx.device[prx.orientation];t.height(r[1]).width(r[0]),$("#maincontent").toggleClass("portrait").toggleClass("landscape"),$(".body_generic").toggleClass("skin-orientation-portrait").toggleClass("skin-orientation-landscape"),prx.resetZoomMinValue(),prx.positionFrame(),prx.applySkin(),prx._autozoom?(prx.device.defaultOrientation!=prx.orientation?prx.changeZoomLevel(Math.min(prx.getAutoZoomValue(),100*prx._zoom)):prx.changeZoomLevel(prx.getAutoZoomValue()),prx._autozoom=!0):prx.changeZoomLevel($("#zoom").slider("option","value"));try{e?prx.crossmsg.send({action:"change-orientation"}):prx.crossmsg.send({action:"change-body-orientation"})}catch(e){}}},prx.checkLocalChromeForWarning=function(){return!0},prx.iniRefreshPreview=function(){$("#refresh-preview").click(function(e){try{prx.crossmsg.send({action:"show-loader"})}catch(e){}if(prx.prto){var t=window.location.href;t.indexOf("?")<0?t+="?":t+="&",t+="get_frame_url=1",$.ajax(t,{error:function(e){console.error("An error occurred while getting data from server "+e),prx.crossmsg.send({action:"hide-loader"})},success:function(e){e.success?$("#pio-playerframe").attr("src",e.url):(console.error("Not a valid server response, could not refresh"),prx.crossmsg.send({action:"hide-loader"}))},dataType:"json"})}else $("#pio-playerframe").attr("src",$("#pio-playerframe").attr("src"));prx.screenshots.ini(),prx.iniScreenBrowser({sort:prx.sort,pages:prx.data.pages,startscreen:prx.startscreen}),void 0!==prx.spaces&&prx.spaces&&void 0!==prx.hasSpacesTheme&&prx.hasSpacesTheme()&&$("#pio-playerframe").css("background-color","transparent")})},prx.crossmsg=prx.crossmsg||{},prx.crossmsg.gettargetframe=function(e){if(!0===e)return window.opener;try{return $("#pio-playerframe").get(0).contentWindow}catch(e){}},prx.crossmsg.onmsg=function(e){try{switch(e.action){case"player-ready":prx.iniPlayerReady(e.data);break;case"switch-orientation":prx.switchOrientation();break;case"scroll-to-top":prx.scrollToTop();break;case"highlight-page":prx.highlightPage(e.data.pageid);break;case"show-actions":prx.showActionsActionsInPreview(e.data.activeStateIds);break;case"hide-actions":prx.hideActionsActionsInPreview();break;case"show-click-effect":prx.showClickEffectActionsInPreview();break;case"hide-click-effect":prx.hideClickEffectActionsInPreview();break;case"go-to-previous-screen":prx.goToPreviousScreen();break;case"go-to-next-screen":prx.goToNextScreen();break;case"player-engine-enter-fullscreen":prx.utils.playerEngineEnterFullscreen();break;case"player-engine-exit-fullscreen":prx.utils.playerEngineExitFullscreen();break;case"missed-fonts":prx.utils.missedFonts();break;case"inputblur":var t=$("iframe").get(0),n=t.style.display;t.style.display="none",t.offsetHeight,t.style.display=n;break;case"ready-to-load-data":var r=new URL(window.location.href);r.searchParams.delete("hc"),window.history.replaceState({},document.title,r.toString()),prx.crossmsg.send({action:"ready-to-load-data"},!0);break;case"project-data-saved":try{var o=$("#pio-playerframe");o.contents().find("#canvas-loader").fadeOut(),o.attr("src",prx.engineIFrameURL)}catch(e){}}}catch(e){}},prx.iniPlayerReady=function(e){prx.applySkin(),prx.device=e.device},prx.data={},prx.sort="",prx._zoom=1,prx._autozoom=!0,prx._zoomcookie=!1,prx.inAnnotationMode=!1,prx.url={},prx.embed=1,$(document).ready(function(){prx.iniOrientation(),prx.iniRefreshPreview(),prx.positionFrame(),prx.fitToScreenOnPageStart(),prx.applySkin(),prx.crossmsg.ini()}),$(window).resize(function(){prx.unsupportedBrowsers.checkVersion("embed"),prx.positionFrame(),prx._autozoom&&(prx.changeZoomLevel(prx.getAutoZoomValue()),prx._autozoom=!0)}),function(e,d){var r="__lodash_hash_undefined__",n=9007199254740991,h="[object Arguments]",g="[object Boolean]",m="[object Date]",v="[object Function]",y="[object GeneratorFunction]",x="[object Map]",b="[object Number]",w="[object Object]",o="[object Promise]",k="[object RegExp]",T="[object Set]",C="[object String]",S="[object Symbol]",i="[object WeakMap]",_="[object ArrayBuffer]",j="[object DataView]",$="[object Float32Array]",A="[object Float64Array]",E="[object Int8Array]",N="[object Int16Array]",O="[object Int32Array]",D="[object Uint8Array]",L="[object Uint8ClampedArray]",F="[object Uint16Array]",P="[object Uint32Array]",I=/\w*$/,t=/^\[object .+?Constructor\]$/,a=/^(?:0|[1-9]\d*)$/,q={};q[h]=q["[object Array]"]=q[_]=q[j]=q[g]=q[m]=q[$]=q[A]=q[E]=q[N]=q[O]=q[x]=q[b]=q[w]=q[k]=q[T]=q[C]=q[S]=q[D]=q[L]=q[F]=q[P]=!0,q["[object Error]"]=q[v]=q[i]=!1;var s="object"==typeof global&&global&&global.Object===Object&&global,c="object"==typeof self&&self&&self.Object===Object&&self,u=s||c||Function("return this")(),l="object"==typeof exports&&exports&&!exports.nodeType&&exports,p=l&&"object"==typeof module&&module&&!module.nodeType&&module,f=p&&p.exports===l;function z(e,t){return e.set(t[0],t[1]),e}function H(e,t){return e.add(t),e}function M(e,t,n,r){var o=-1,i=e?e.length:0;for(r&&i&&(n=e[++o]);++o<i;)n=t(n,e[o],o,e);return n}function B(e){var t=!1;if(null!=e&&"function"!=typeof e.toString)try{t=!!(e+"")}catch(e){}return t}function W(e){var n=-1,r=Array(e.size);return e.forEach(function(e,t){r[++n]=[t,e]}),r}function R(t,n){return function(e){return t(n(e))}}function V(e){var t=-1,n=Array(e.size);return e.forEach(function(e){n[++t]=e}),n}var U,X=Array.prototype,Z=Function.prototype,J=Object.prototype,Y=u["__core-js_shared__"],G=(U=/[^.]+$/.exec(Y&&Y.keys&&Y.keys.IE_PROTO||""))?"Symbol(src)_1."+U:"",K=Z.toString,Q=J.hasOwnProperty,ee=J.toString,te=RegExp("^"+K.call(Q).replace(/[\\^$.*+?()[\]{}|]/g,"\\$&").replace(/hasOwnProperty|(function).*?(?=\\\()| for .+?(?=\\\])/g,"$1.*?")+"$"),ne=f?u.Buffer:d,re=u.Symbol,oe=u.Uint8Array,ie=R(Object.getPrototypeOf,Object),ae=Object.create,se=J.propertyIsEnumerable,ce=X.splice,ue=Object.getOwnPropertySymbols,le=ne?ne.isBuffer:d,pe=R(Object.keys,Object),fe=Ie(u,"DataView"),de=Ie(u,"Map"),he=Ie(u,"Promise"),ge=Ie(u,"Set"),me=Ie(u,"WeakMap"),ve=Ie(Object,"create"),ye=Be(fe),xe=Be(de),be=Be(he),we=Be(ge),ke=Be(me),Te=re?re.prototype:d,Ce=Te?Te.valueOf:d;function Se(e){var t=-1,n=e?e.length:0;for(this.clear();++t<n;){var r=e[t];this.set(r[0],r[1])}}function _e(e){var t=-1,n=e?e.length:0;for(this.clear();++t<n;){var r=e[t];this.set(r[0],r[1])}}function je(e){var t=-1,n=e?e.length:0;for(this.clear();++t<n;){var r=e[t];this.set(r[0],r[1])}}function $e(e){this.__data__=new _e(e)}function Ae(e,t){var n=Re(e)||function(e){return function(e){return function(e){return!!e&&"object"==typeof e}(e)&&Ve(e)}(e)&&Q.call(e,"callee")&&(!se.call(e,"callee")||ee.call(e)==h)}(e)?function(e,t){for(var n=-1,r=Array(e);++n<e;)r[n]=t(n);return r}(e.length,String):[],r=n.length,o=!!r;for(var i in e)!t&&!Q.call(e,i)||o&&("length"==i||He(i,r))||n.push(i);return n}function Ee(e,t,n){var r=e[t];Q.call(e,t)&&We(r,n)&&(n!==d||t in e)||(e[t]=n)}function Ne(e,t){for(var n=e.length;n--;)if(We(e[n][0],t))return n;return-1}function Oe(n,r,o,i,e,t,a){var s;if(i&&(s=t?i(n,e,t,a):i(n)),s!==d)return s;if(!Ze(n))return n;var c=Re(n);if(c){if(s=function(e){var t=e.length,n=e.constructor(t);t&&"string"==typeof e[0]&&Q.call(e,"index")&&(n.index=e.index,n.input=e.input);return n}(n),!r)return function(e,t){var n=-1,r=e.length;t||(t=Array(r));for(;++n<r;)t[n]=e[n];return t}(n,s)}else{var u=ze(n),l=u==v||u==y;if(Ue(n))return function(e,t){if(t)return e.slice();var n=new e.constructor(e.length);return e.copy(n),n}(n,r);if(u==w||u==h||l&&!t){if(B(n))return t?n:{};if(s=function(e){return"function"!=typeof e.constructor||Me(e)?{}:function(e){return Ze(e)?ae(e):{}}(ie(e))}(l?{}:n),!r)return function(e,t){return Fe(e,qe(e),t)}(n,function(e,t){return e&&Fe(t,Je(t),e)}(s,n))}else{if(!q[u])return t?n:{};s=function(e,t,n,r){var o=e.constructor;switch(t){case _:return Le(e);case g:case m:return new o(+e);case j:return function(e,t){var n=t?Le(e.buffer):e.buffer;return new e.constructor(n,e.byteOffset,e.byteLength)}(e,r);case $:case A:case E:case N:case O:case D:case L:case F:case P:return function(e,t){var n=t?Le(e.buffer):e.buffer;return new e.constructor(n,e.byteOffset,e.length)}(e,r);case x:return function(e,t,n){return M(t?n(W(e),!0):W(e),z,new e.constructor)}(e,r,n);case b:case C:return new o(e);case k:return function(e){var t=new e.constructor(e.source,I.exec(e));return t.lastIndex=e.lastIndex,t}(e);case T:return function(e,t,n){return M(t?n(V(e),!0):V(e),H,new e.constructor)}(e,r,n);case S:return function(e){return Ce?Object(Ce.call(e)):{}}(e)}}(n,u,Oe,r)}}a||(a=new $e);var p=a.get(n);if(p)return p;if(a.set(n,s),!c)var f=o?function(e){return function(e,t,n){var r=t(e);return Re(e)?r:function(e,t){for(var n=-1,r=t.length,o=e.length;++n<r;)e[o+n]=t[n];return e}(r,n(e))}(e,Je,qe)}(n):Je(n);return function(e,t){for(var n=-1,r=e?e.length:0;++n<r&&!1!==t(e[n],n,e););}(f||n,function(e,t){f&&(e=n[t=e]),Ee(s,t,Oe(e,r,o,i,t,n,a))}),s}function De(e){return!(!Ze(e)||function(e){return!!G&&G in e}(e))&&(Xe(e)||B(e)?te:t).test(Be(e))}function Le(e){var t=new e.constructor(e.byteLength);return new oe(t).set(new oe(e)),t}function Fe(e,t,n,r){n||(n={});for(var o=-1,i=t.length;++o<i;){var a=t[o],s=r?r(n[a],e[a],a,n,e):d;Ee(n,a,s===d?e[a]:s)}return n}function Pe(e,t){var n=e.__data__;return function(e){var t=typeof e;return"string"==t||"number"==t||"symbol"==t||"boolean"==t?"__proto__"!==e:null===e}(t)?n["string"==typeof t?"string":"hash"]:n.map}function Ie(e,t){var n=function(e,t){return null==e?d:e[t]}(e,t);return De(n)?n:d}Se.prototype.clear=function(){this.__data__=ve?ve(null):{}},Se.prototype.delete=function(e){return this.has(e)&&delete this.__data__[e]},Se.prototype.get=function(e){var t=this.__data__;if(ve){var n=t[e];return n===r?d:n}return Q.call(t,e)?t[e]:d},Se.prototype.has=function(e){var t=this.__data__;return ve?t[e]!==d:Q.call(t,e)},Se.prototype.set=function(e,t){return this.__data__[e]=ve&&t===d?r:t,this},_e.prototype.clear=function(){this.__data__=[]},_e.prototype.delete=function(e){var t=this.__data__,n=Ne(t,e);return!(n<0)&&(n==t.length-1?t.pop():ce.call(t,n,1),!0)},_e.prototype.get=function(e){var t=this.__data__,n=Ne(t,e);return n<0?d:t[n][1]},_e.prototype.has=function(e){return-1<Ne(this.__data__,e)},_e.prototype.set=function(e,t){var n=this.__data__,r=Ne(n,e);return r<0?n.push([e,t]):n[r][1]=t,this},je.prototype.clear=function(){this.__data__={hash:new Se,map:new(de||_e),string:new Se}},je.prototype.delete=function(e){return Pe(this,e).delete(e)},je.prototype.get=function(e){return Pe(this,e).get(e)},je.prototype.has=function(e){return Pe(this,e).has(e)},je.prototype.set=function(e,t){return Pe(this,e).set(e,t),this},$e.prototype.clear=function(){this.__data__=new _e},$e.prototype.delete=function(e){return this.__data__.delete(e)},$e.prototype.get=function(e){return this.__data__.get(e)},$e.prototype.has=function(e){return this.__data__.has(e)},$e.prototype.set=function(e,t){var n=this.__data__;if(n instanceof _e){var r=n.__data__;if(!de||r.length<199)return r.push([e,t]),this;n=this.__data__=new je(r)}return n.set(e,t),this};var qe=ue?R(ue,Object):function(){return[]},ze=function(e){return ee.call(e)};function He(e,t){return!!(t=null==t?n:t)&&("number"==typeof e||a.test(e))&&-1<e&&e%1==0&&e<t}function Me(e){var t=e&&e.constructor;return e===("function"==typeof t&&t.prototype||J)}function Be(e){if(null!=e){try{return K.call(e)}catch(e){}try{return e+""}catch(e){}}return""}function We(e,t){return e===t||e!=e&&t!=t}(fe&&ze(new fe(new ArrayBuffer(1)))!=j||de&&ze(new de)!=x||he&&ze(he.resolve())!=o||ge&&ze(new ge)!=T||me&&ze(new me)!=i)&&(ze=function(e){var t=ee.call(e),n=t==w?e.constructor:d,r=n?Be(n):d;if(r)switch(r){case ye:return j;case xe:return x;case be:return o;case we:return T;case ke:return i}return t}),e.cloneDeep=function(e){return Oe(e,!0,!0)};var Re=Array.isArray;function Ve(e){return null!=e&&function(e){return"number"==typeof e&&-1<e&&e%1==0&&e<=n}(e.length)&&!Xe(e)}var Ue=le||function(){return!1};function Xe(e){var t=Ze(e)?ee.call(e):"";return t==v||t==y}function Ze(e){var t=typeof e;return!!e&&("object"==t||"function"==t)}function Je(e){return Ve(e)?Ae(e):function(e){if(!Me(e))return pe(e);var t=[];for(var n in Object(e))Q.call(e,n)&&"constructor"!=n&&t.push(n);return t}(e)}}(window.lodash=window.lodash||{}),prx.embed=0,prx.export2html=1,prx.getCustomSkinUrl=function(){return("custom"==prx.skin||"shared"==prx.skin)&&"src/images/skin_"+prx.orientation+".png"},$(document).ready(function(){prx.iniBgColor(),prx.unsupportedBrowsers.checkVersion("export"),setTimeout(function(){prx.applySkin()},1e3)});
```
