## Supplementary figures and images for "Post Induction Hypotension prediction during general anesthesia using Machine Learning Techniques"

### 08d3b5585c3baf0fd8ac52e8c44a8630.png

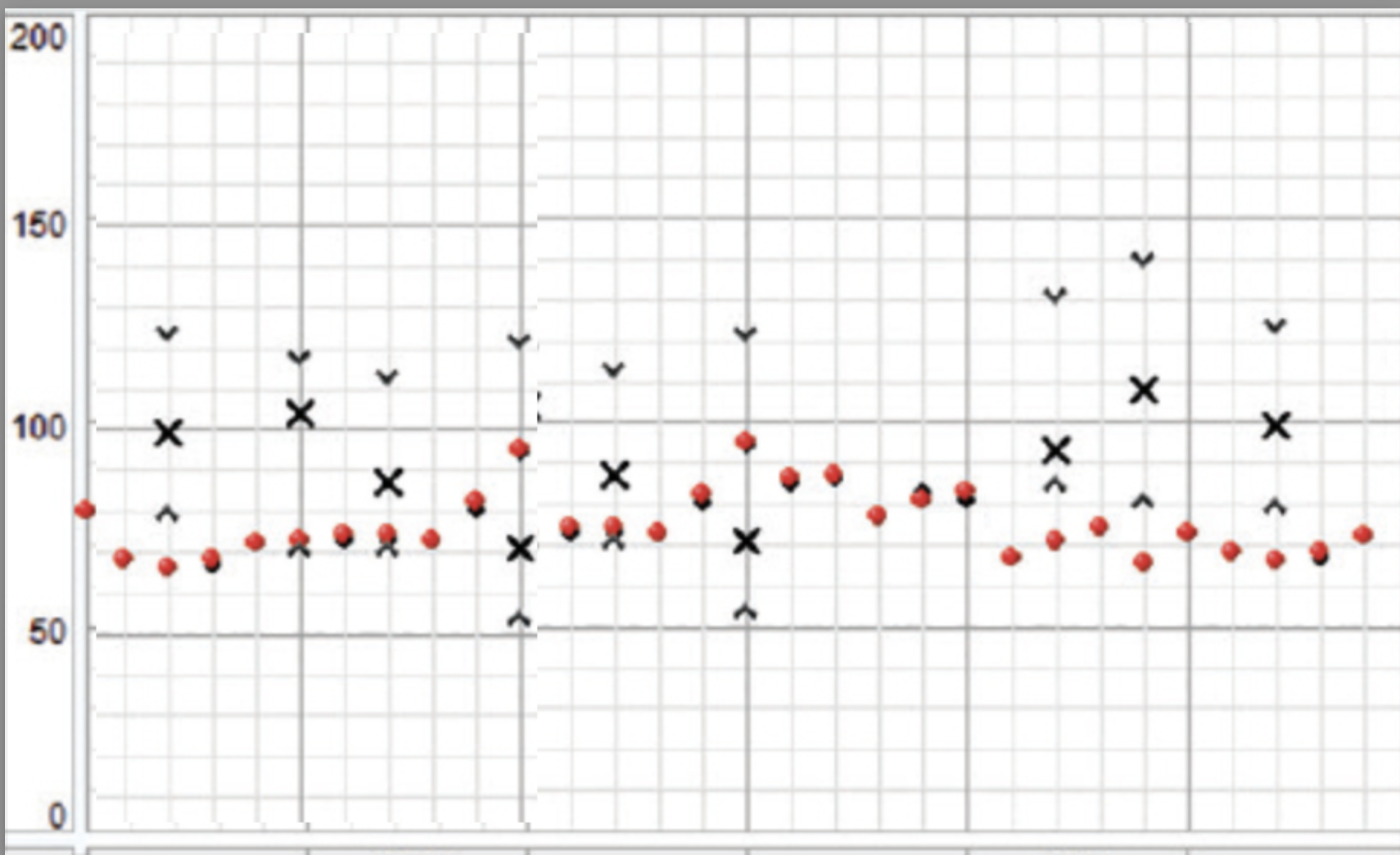

### 68d047acd614373a75ca2548bba92b44.png

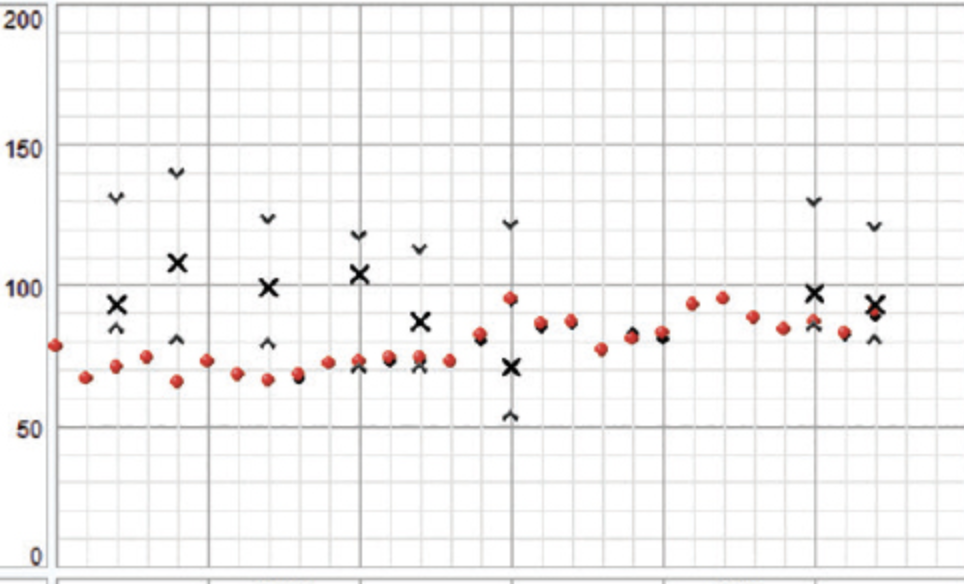

### 7902aa96fae82cf51263d633901fa229.png

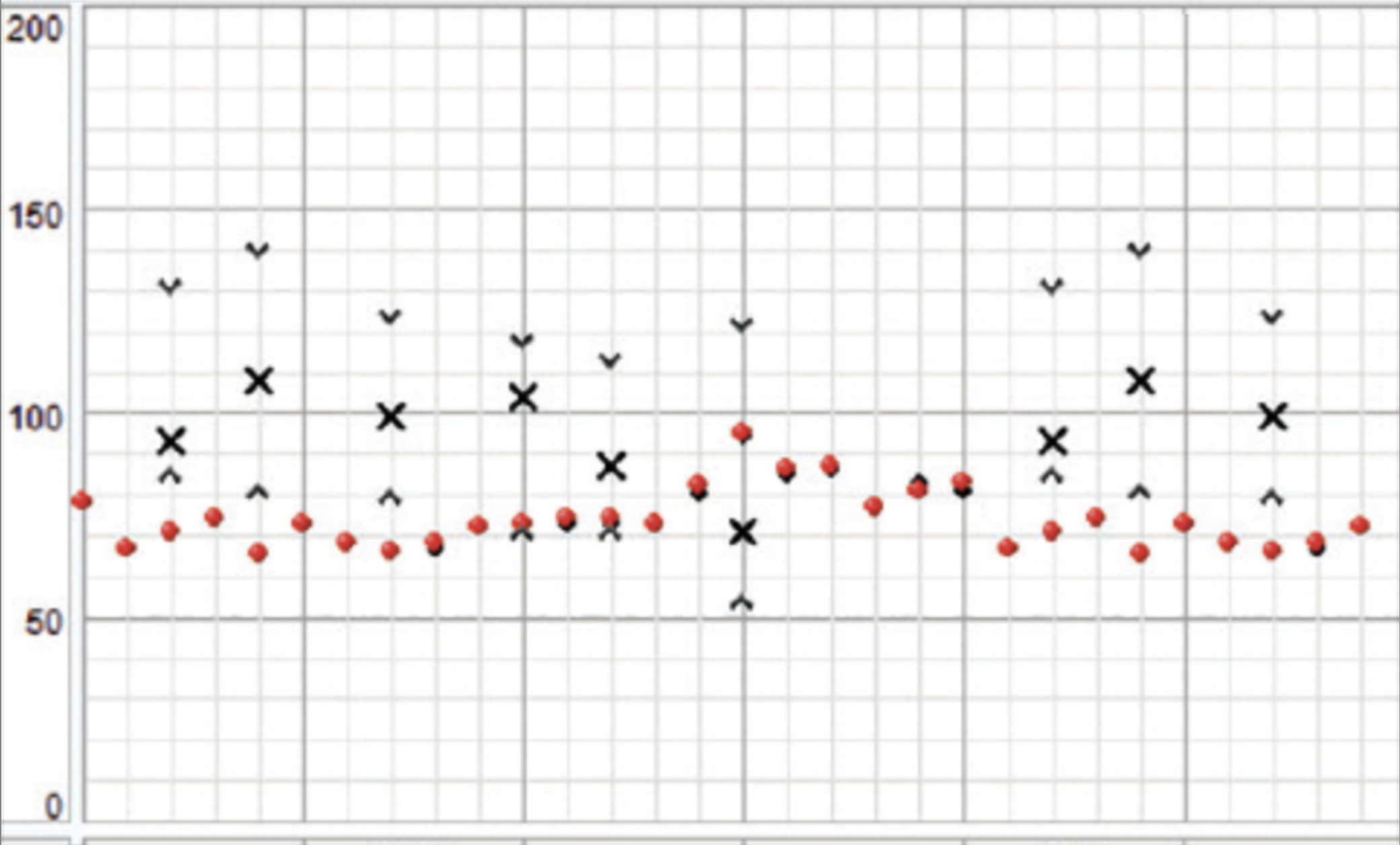

### actions-sprite-1.png

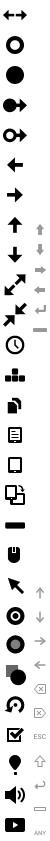

### android-icon-hires.png

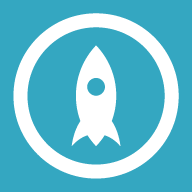

### android-icon.png

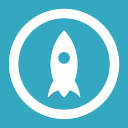

### appicon.png

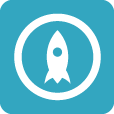

### apple-touch-icon-retina180.png

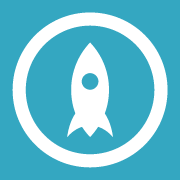

### apple-touch-icon.png

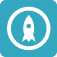

### startup-568.png

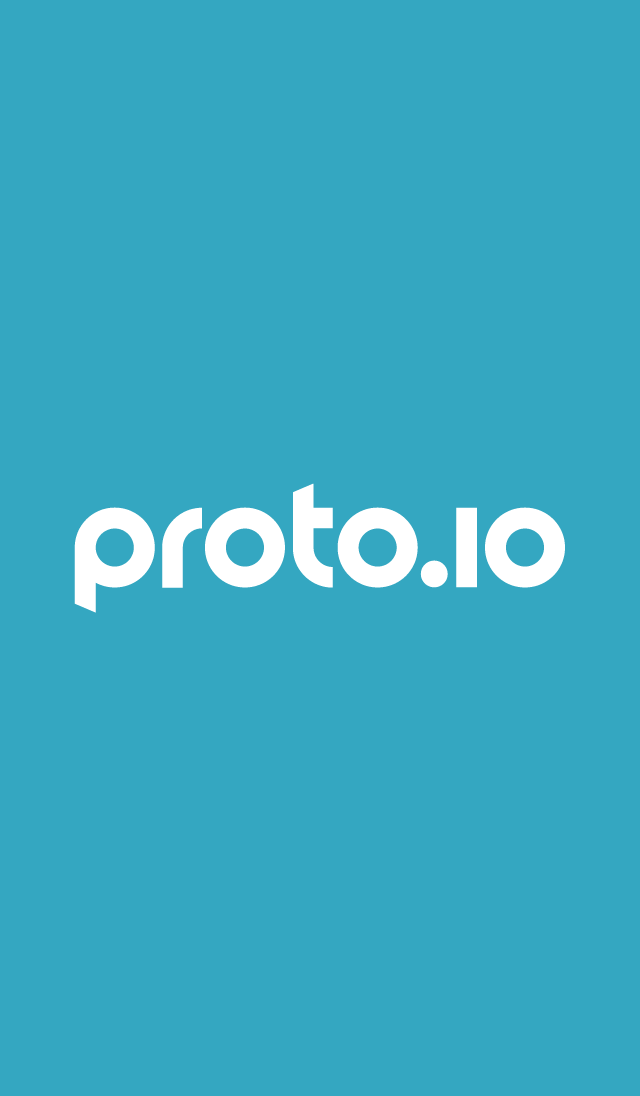

### startup-ipad-landscape-retina.png

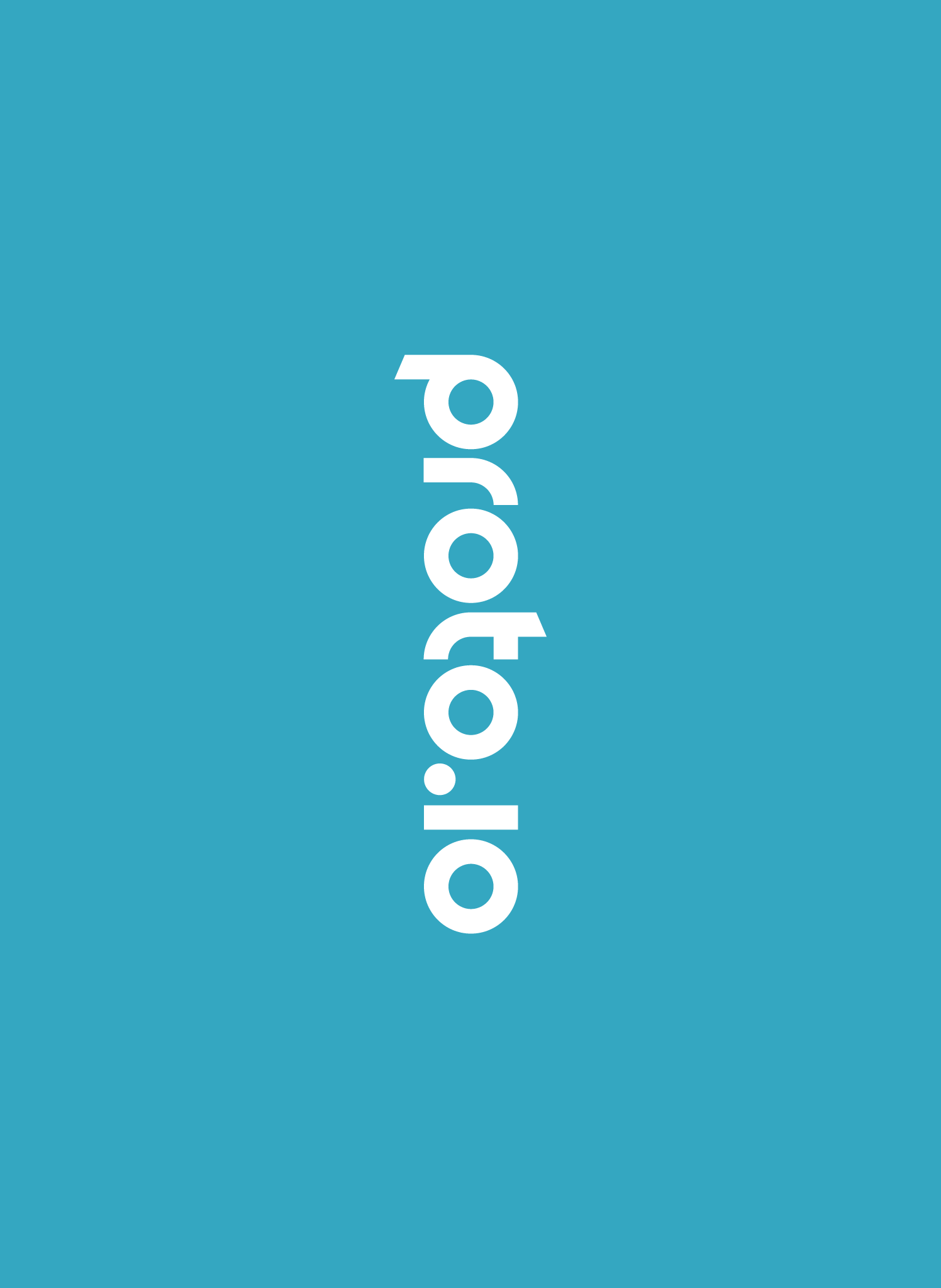

### startup-ipad-landscape.png

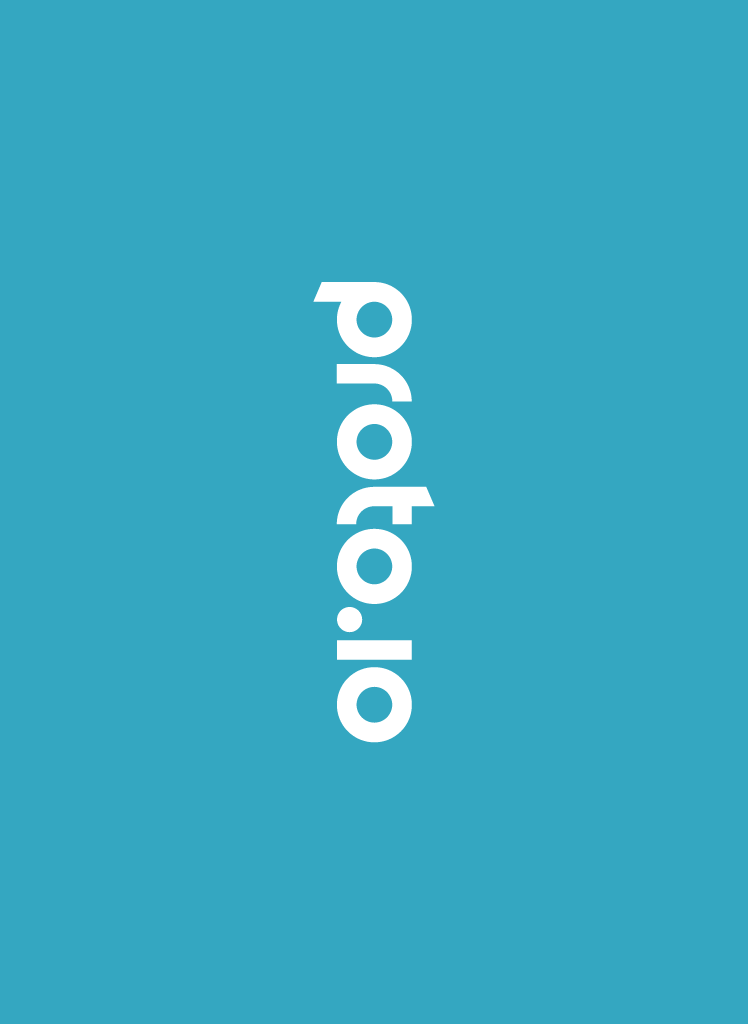

### startup-ipad-pro-landscape.png

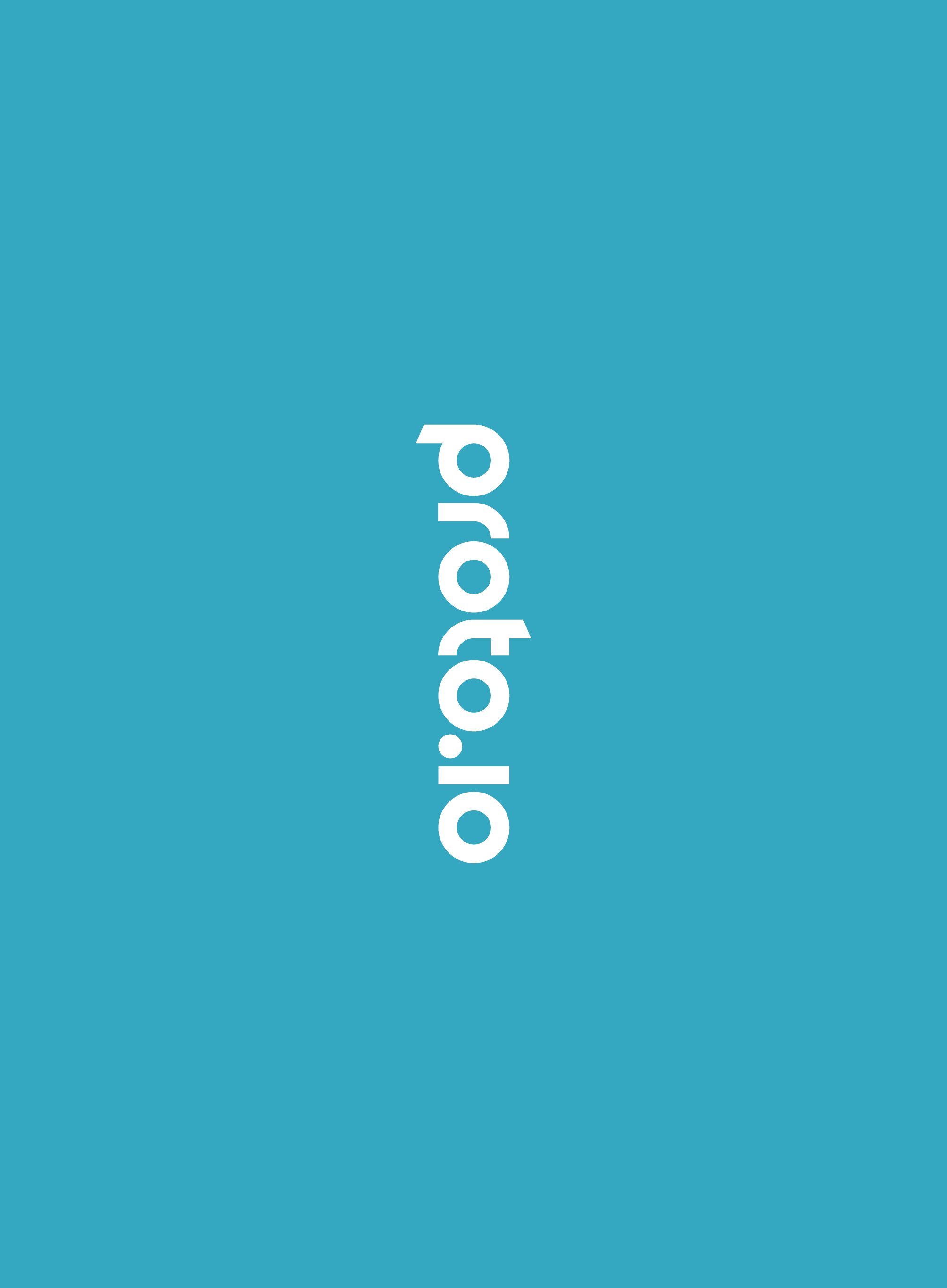

### startup-ipad-pro.png

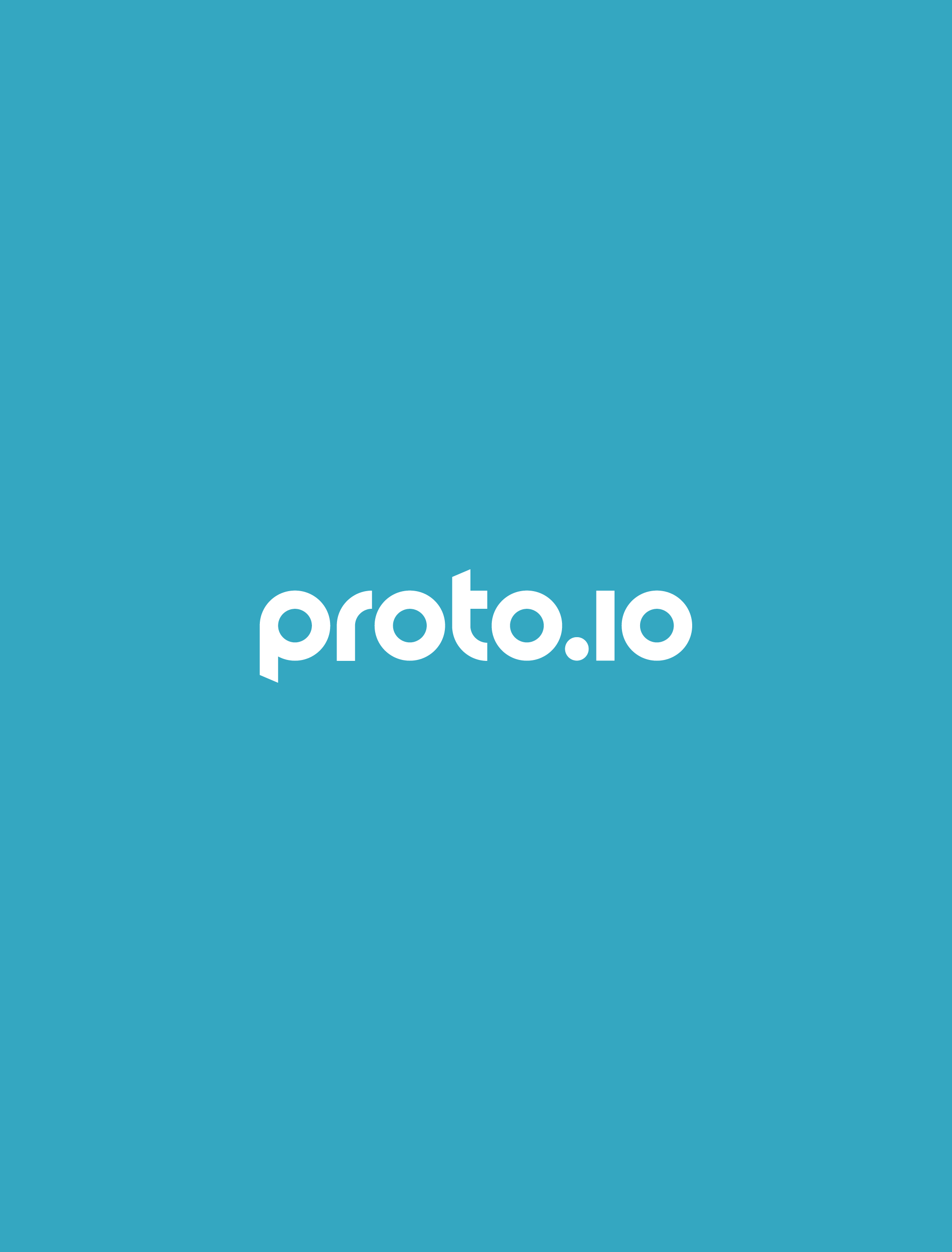

### startup-ipad-retina.png

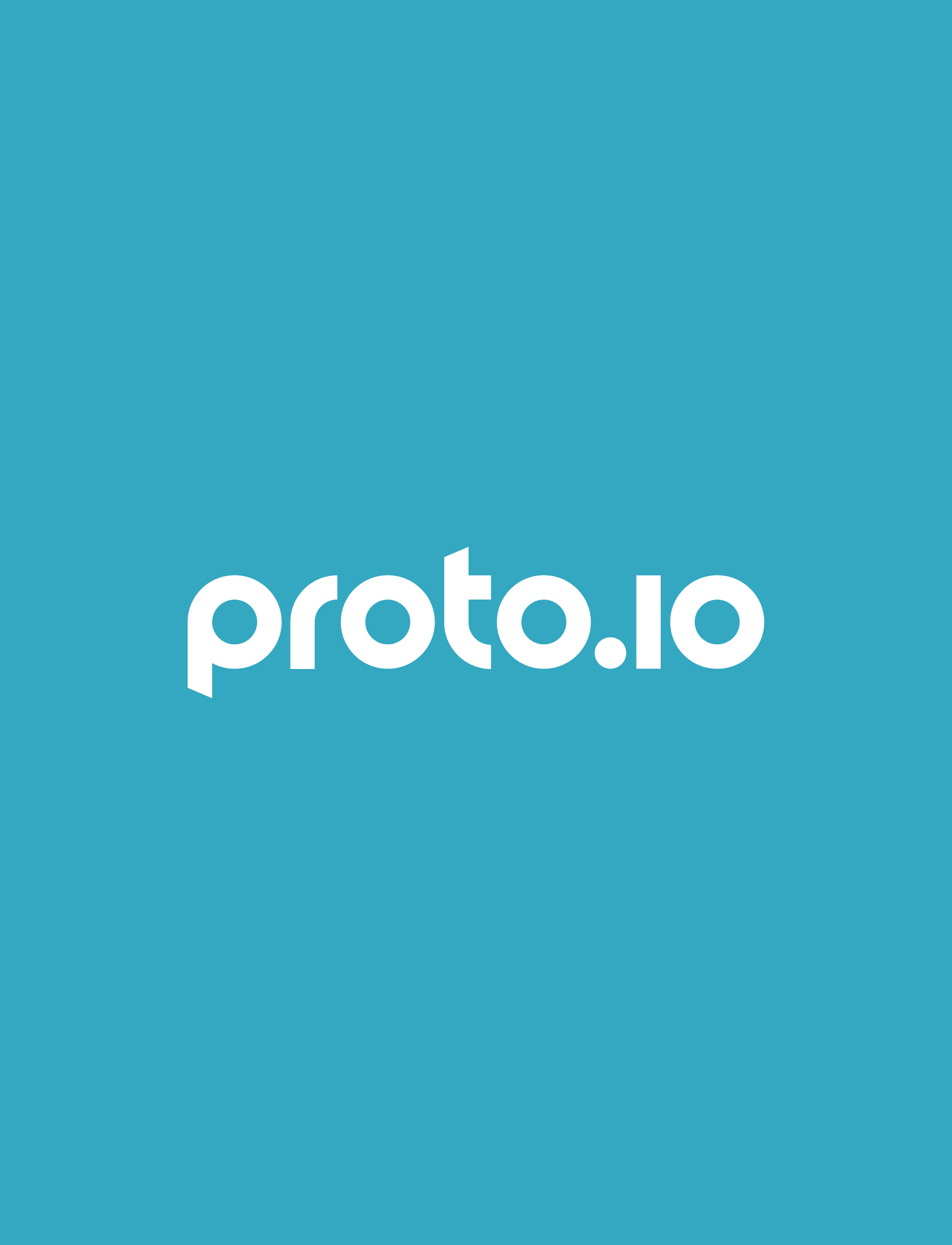

### startup-ipad.png

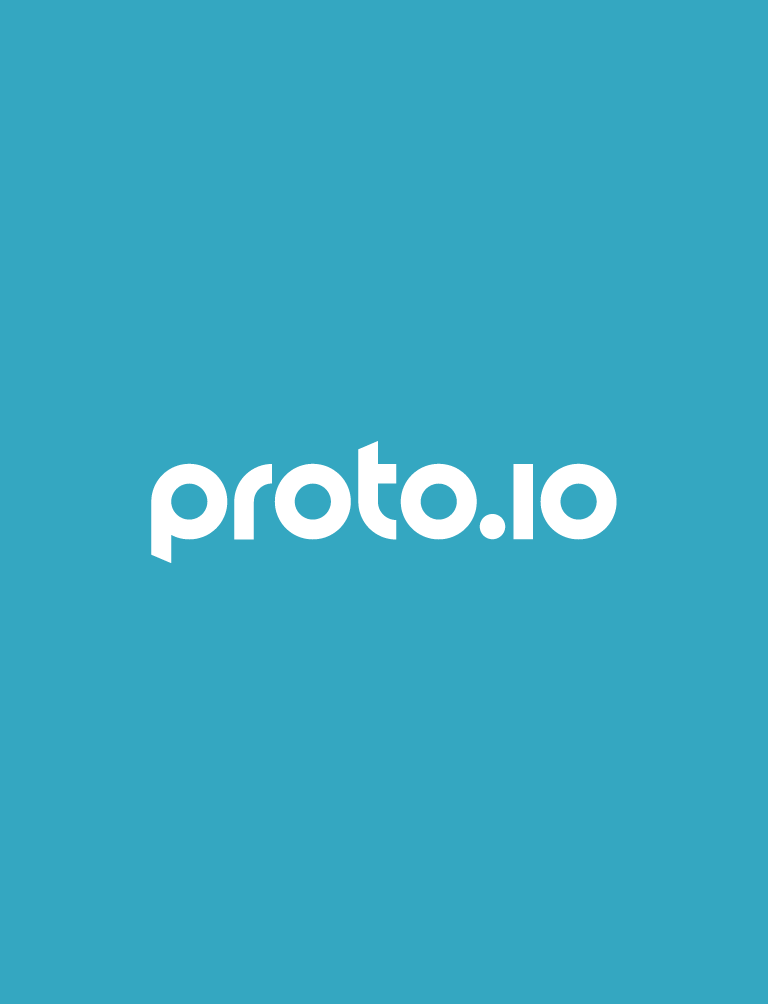

### startup-retina-6.png

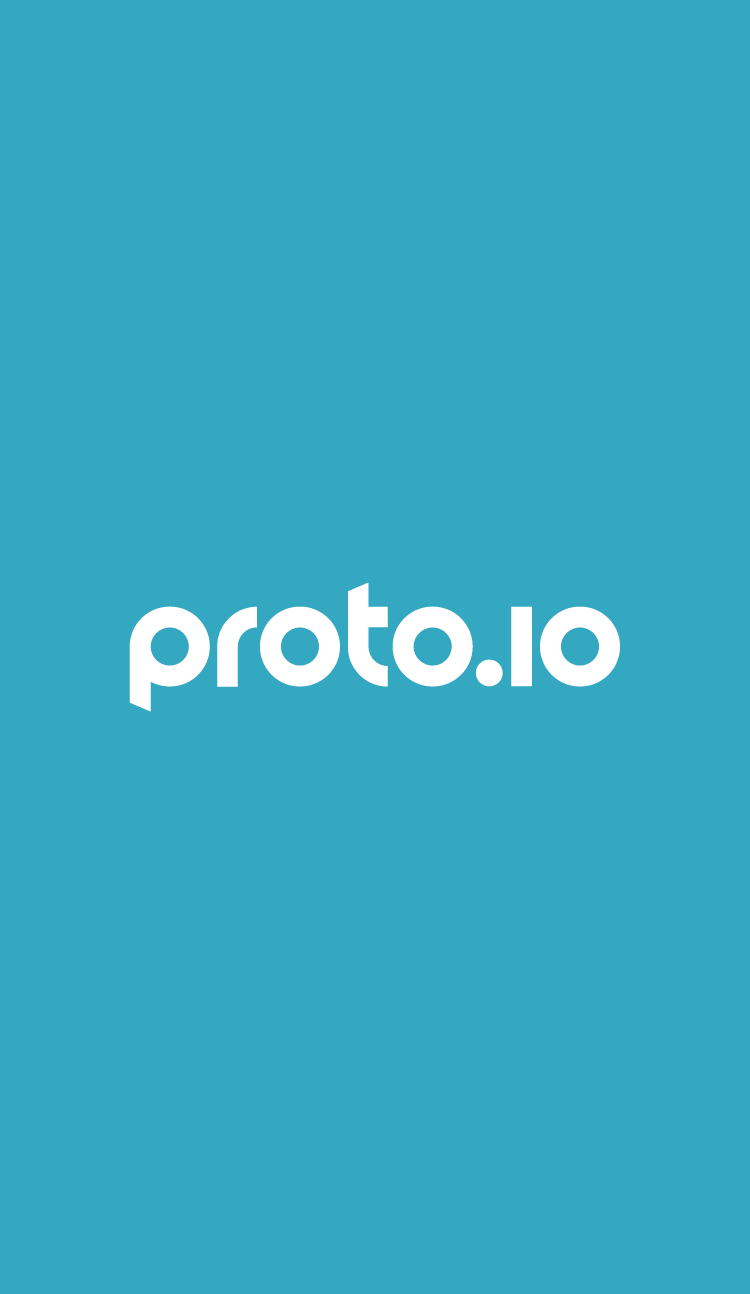

### startup-retina-6plus-landscape.png

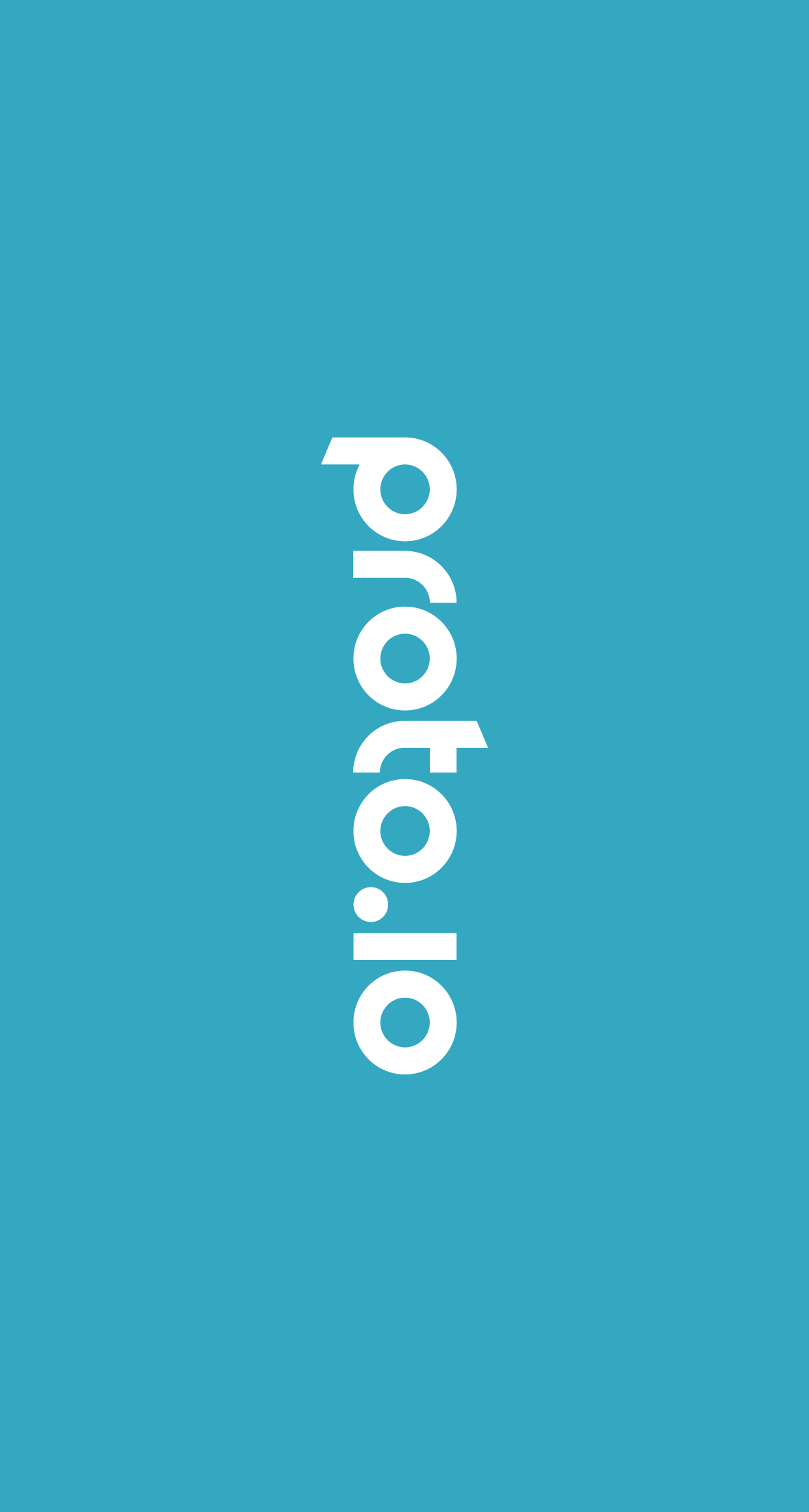

### startup-retina-6plus.png

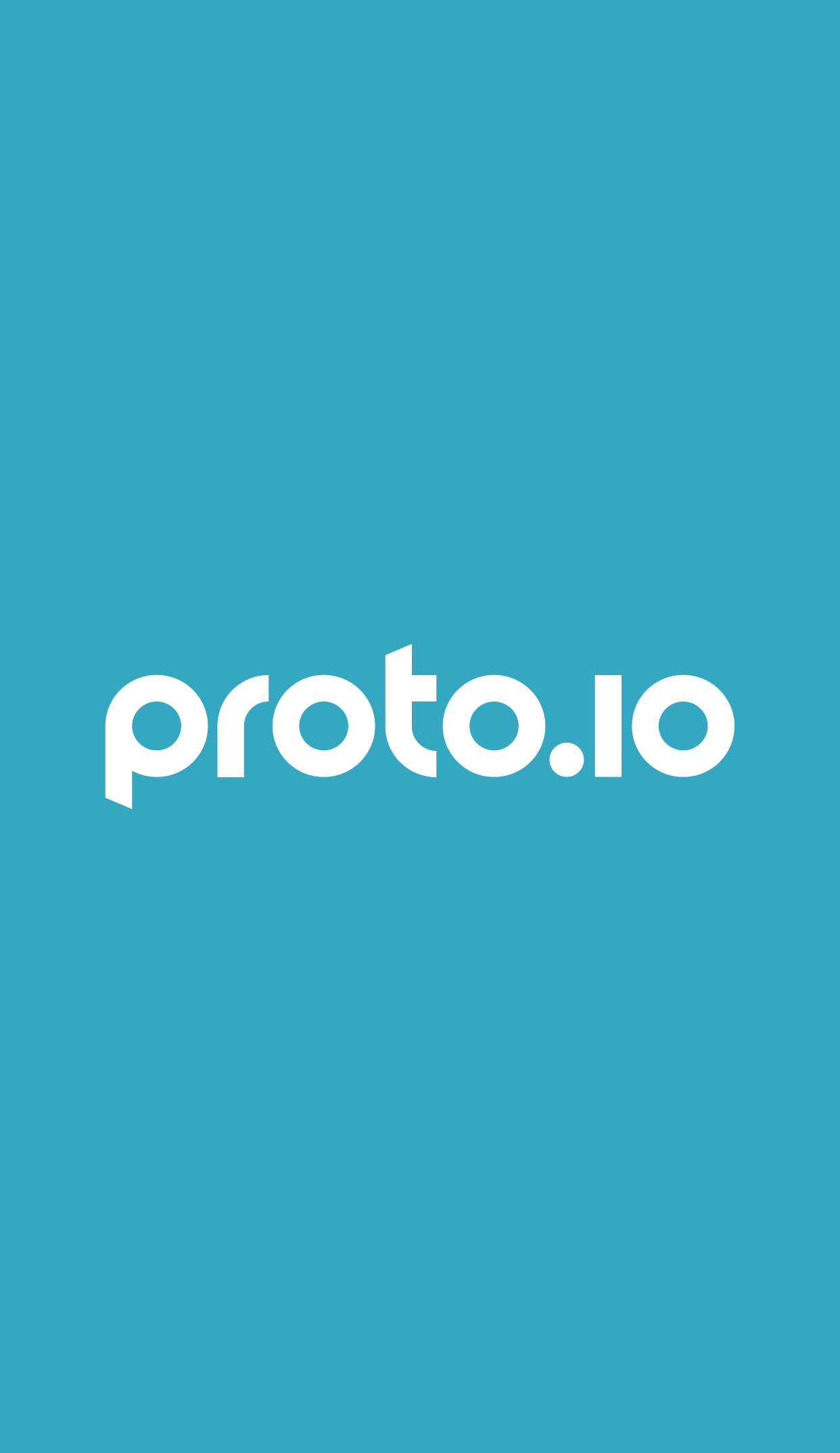

### startup-retina.png

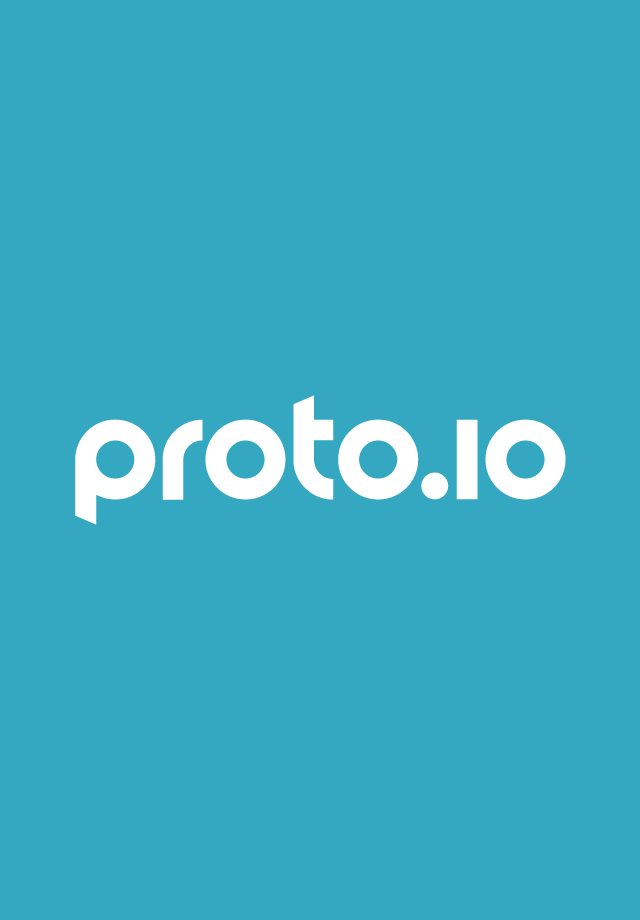

### startup.png

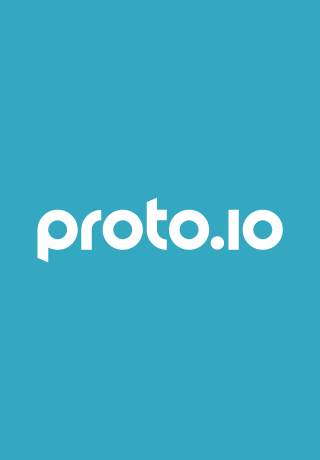
